# Does immune recognition of SARS-CoV2 epitopes vary between different ethnic groups?

**DOI:** 10.1101/2021.05.24.21257707

**Authors:** Tungadri Bose, Namrata Pant, Nishal Kumar Pinna, Subhrajit Bhar, Anirban Dutta, Sharmila S. Mande

## Abstract

The SARS-CoV2 mediated Covid-19 pandemic has impacted humankind at an unprecedented scale. While substantial research efforts have focused towards understand the mechanisms of viral infection and developing vaccines/ therapeutics, factors affecting the susceptibility to SARS-CoV2 infection and manifestation of Covid-19 remain less explored. Given that the Human Leukocyte Antigen (HLA) system is known to vary among ethnic populations, it is likely to affect the recognition of the virus, and in turn, the susceptibility to Covid-19. To understand this, we used bioinformatic tools to probe all SARS-CoV2 peptides which could elicit T-cell response in humans. We also tried to answer the intriguing question of whether these potential epitopes were equally immunogenic across ethnicities, by studying the distribution of HLA alleles among different populations and their share of cognate epitopes. We provide evidence that the newer mutations in SARS-CoV2 are unlikely to alter the T-cell mediated immunogenic responses among the studied ethnic populations. The work presented herein is expected to bolster our understanding of the pandemic, by providing insights into differential immunological response of ethnic populations to the virus as well as by gauging the possible effects of mutations in SARS-CoV2 on efficacy of potential epitope-based vaccines through evaluating ∼40000 viral genomes.

## INTRODUCTION

The ongoing outbreak of severe acute respiratory syndrome (SARS), commonly known as Covid-19, has already infected over 157 million and has led to the death of 3,200,000 people worldwide (“WHO Coronavirus Disease (COVID-19) Dashboard,” n.d.). SARS-CoV2, a newly identified lineage B *Betacoronavirus* has been held responsible for this global pandemic (Letko et al., 2020). While the current pandemic is unprecedented in scale as compared to the earlier coronavirus outbreaks (Pathan et al., 2020), they are quite alike considering the ongoing quest for an effective preventive/ treatment regime to counter the disease (Ayukekbong et al., 2020). A significant amount of research has been directed at understanding various facets to the pathogenesis. These include the viral attachment and entry (Hoffmann et al., 2020; Letko et al., 2020), formation of the coronavirus replication/ transcription complex (Y. Chen et al., 2020; Krichel et al., 2020), replication, transcription and translation of viral proteins (Hillen et al., 2020; Satarker and Nampoothiri, 2020; Wang et al., 2020), virion assembly and release (Kumar et al., 2020; Li et al., 2020) and the commonalities and differences of this disease with seasonal flu and previous SARS infections (Song et al., 2020; Xu et al., 2020). In spite of these efforts, factors affecting susceptibility to infection of SARS-CoV2 and manifestation of Covid-19 are yet to be properly understood and demands more attention. It may be noted that SARS-CoV2’s rate of genomic mutation is similar to most RNA viruses, (Pathan et al., 2020), and recent evidences seem to suggest that the mortality rates among Covid-19 patients is strongly associated with the mutation profile of the infecting virus (Toyoshima et al., 2020; “WHO | Variant analysis of SARS-CoV-2 genomes,” n.d.). While a pre-existing medical condition is likely to increase the risk of severity of Covid-19 infection (CDC, 2020), yet another factor influencing the susceptibility to the disease could be the genetic makeup of an individual. The Human Leukocyte Antigens (HLA) system, a major determinant of our ability to detect and neutralize an invading pathogen, is encoded by the Major Histocompatibility Complex (MHC) genes located on chromosome-6. MHCs, which are further categorized into classes-I and II, are highly polymorphic and are known to vary significantly among individuals of different ethnicities. The outcome of an infection event is therefore dependent on both the genotype of the virus as well as the host cell surface (MHC) molecules destined to present viral antigenic peptides to the human T-cell receptor (TCR) of T-lymphocytes (also called killer T-cells or CD8-positive cytotoxic T-cells) and T-helper cells (also called CD4-positive T cells) (Murray and McMichael, 1992; van Montfoort et al., 2014).

In this work, we have investigated how the genetic variations across ethnicities is likely to influence the ability of their immune system in timely recognition of the virus, and in turn, their susceptibility to Covid-19. To this end, state-of-the art bioinformatic tools were used to (a) identify the probable antigenic peptides on the SARS-CoV2 proteomes, and (b) identify HLA alleles which could recognize and present these epitopes to the T-cells, along with the prevalence of these alleles in different ethnic groups. In addition, a large set of fully sequenced SARS-CoV2 genomes were analyzed to probe the possible effect of viral genetic variations on antigenic recognition. Whether the variations in the viral genome over time are likely to change the susceptibility of an ethnic group to Covid-19 infection was evaluated. In this context it may be noted that, given the computational challenges of identifying B-cell epitopes with adequate confidence, the current study did not focus on this aspect of human immunity. Results presented in this work also assumes additional importance when viewed in the context of a recent publication which highlighted the prospect and benefits of considering non-spike proteins for future Covid-19 vaccine designs (Peng et al., 2020).

## RESULTS

### Identification of SARS-CoV2 Epitopes

One of the key mechanism of identification of an invading pathogen by the host immune system involves MHC class-I and MHC class-II proteins presenting the pathogenic protein fragments (epitopes) on the surface of the CD8-positive cytotoxic T-cells and CD4-positive T-helper cells respectively. Given this, efforts were first made to identify SARS-CoV2 epitopes that could be recognized by the HLA allelic variants. A total of 326 epitopic regions from all the proteins encoded by the SARS-CoV2 reference genome (GenBank Accession no. MN908947) were predicted to bind to 130 HLA allelic variants (Supplementary Table 1) with reasonable confidence (see Materials and Methods). This comprised of 276 CD8 (MHC class-I) and 50 CD4 (MHC class-II) epitopes. From the list of predicted epitopes and the corresponding HLA alleles, it was observed that some of the HLA alleles could recognize higher number of SARS-CoV2 epitopes and thus might play a more significant role in immune response. Of the 108 MHC class-I associated HLA alleles, that were predicted to be involved in the antigenic recognition of SARS-CoV2 proteins in (CD8-positive) cytotoxic T-lymphocyte cells, the highest number of epitopes were identified by HLA-A*02:11, HLA.A*26:02, HLA.B*15:17, HLA.A*24:03 and HLA.B*35.41 (50, 32, 32, 28 and 19 epitopes respectively). In contrast, among the MHC class-II associated 22 HLA alleles involved in the antigen recognition of SARS-CoV2 proteins in (CD4-positive) T-helper cells, HLA-DRB1*01:01, HLA.DRB*115:01, HLA.DRB*115:06 and HLA.DRB*101:02 were predicted to present the highest number of epitopes (16, 13, 13, and 7 epitopes respectively). It was also noted that few of the epitopes could be recognized by multiple HLA alleles (Supplementary Table 1). It therefore appeared that the potential of a population/ ethnic group to cope with Covid-19 infection could be determined by their MHC gene pool.

### Distribution of SARS-CoV2 Cognate HLA Alleles Across Ethnicities

The diversity in the allelic makeup of 26 different ethnic groups constituting five super-populations were studied using data available from the 1000 genomes project (ftp://ftp.1000genomes.ebi.ac.uk/vol1/ftp/data_collections/HLA_types/) (see Materials and Methods). Only those HLA alleles which were predicted to recognize one or more SARS-CoV2 epitopes were considered for the presented analyses. In terms of MHC class-I associated HLA alleles (henceforth termed as MHC-I alleles), while Ad Mixed American (AMR) and South Asians (SAS) demonstrated the highest diversities, Europeans (EUR) were observed to be least diverse (Fig. 1). In contrast, EUR had the highest richness of MHC class-II associated HLA alleles (henceforth termed as MHC-II alleles), alongside AMR (Fig. 1). East Asians (EAS) and SAS had the least MHC-II allelic diversity. The richness and diversity of both MHC-I and MHC-II alleles were found to be moderate among the African (AFR) super-population. In general, the HLA allele richness among super-populations was seen to be more diverse in case of MHC-I as compared to MHC-II (Supplementary Table 2). The ethnicities with least MHC-I and MCH-II HLA allelic richness included Japanese from Tokyo (JPT), Chinese Dai residing in Xishuangbanna (CDX), Southern Han Chinese (CHS), Mende ethnic group from Sierra Leone (MSL) and Yoruba from Ibadan (YRI). In contrast, Colombians from Medellin (CLM) exhibited high richness of both classes of MHC alleles. Most others, like the Kinh from Ho Chi Minh City (KHV) and Sri Lankan Tamils from the UK (STU) demonstrated contrasting characteristics for richness of MHC-I and MHC-II alleles.

**Fig. 1:**
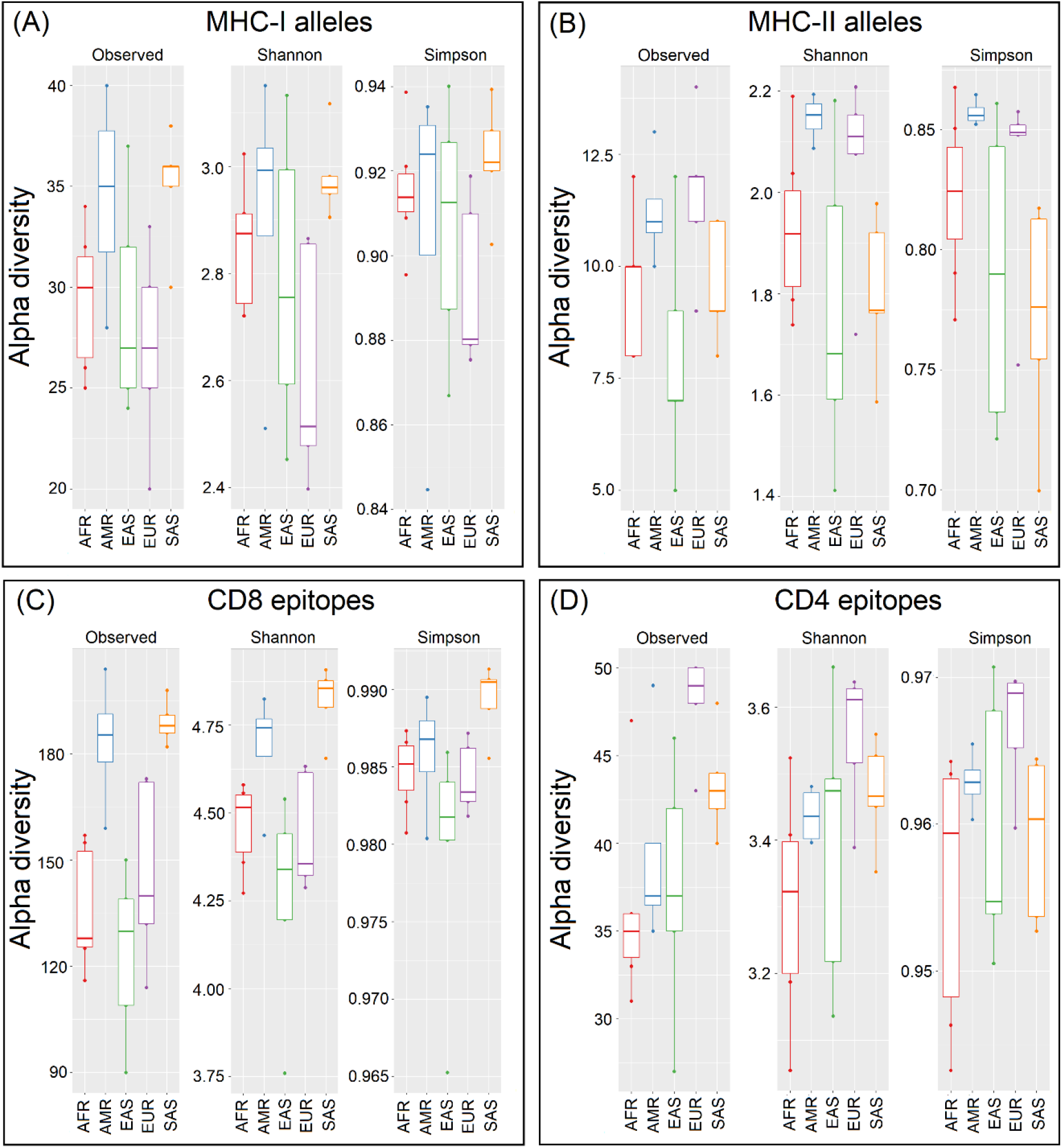
Diversity of HLA alleles and cognate epitopes across super-populations. Richness (total observed numbers), Shannon diversity and Simpson index indicating the heterogeneity of (A) MHC-I and (B) MHC-II alleles encoded by gene pool of studied super-populations as well as the corresponding (C) CD8-specific and (D) CD4-specific SARS-CoV2 epitopes recognized by the HLA-system in these super-populations. The super-populations are abbreviated as AFR-Africans; AMR-Ad mixed Americans; EAS-East Asians; EUR-Europeans; SAS-South east Asians.

As represented in the Euler diagram of HLA alleles in Fig. 2 (details in Supplementary Table 3), 22 MHC-I and 10 MHC-II alleles were omnipresent in all the super-populations (see Materials and Methods). In addition to this, SAS and EAS shared 17 MHC-I alleles, 10 of which were found to be unique to this two super-populations. Certain alleles specific to each of the super-populations were also found. As expected, intra-population variations existed and the occurrence of each of the (SARS-CoV2 associated) MHC-I and MHC-II alleles were not found to be uniform among the samples constituting the ethnic groups (Supplementary Table 2). To account for this, frequency of occurrence for every MHC allele in each of the ethnic groups was computed (see Materials and Methods). Results obtained were used to construct heat maps (Supplementary Figures 1 and 2) for gauging the distribution of the potent MHC alleles which could play a role in immune response against SARS-CoV2. While some of the alleles were seen to be more frequent across ethnicities, it was interesting to note that the alleles with the highest antigenic recognition abilities against SARS-CoV2 were often under-represented across different ethnicities. For instance, the most common HLA alleles observed across ethnicities included MHC-I alleles HLA.C*04:01, HLA.A*01:01, HLA.A*02:01 and MHC-II alleles HLA.DRB*103:01, HLA.DRB*111:01, HLA.DRB*101:01. However, amongst them only HLA.DRB*101:01 (MHC-II) was noted to possess high antigenic recognition ability. In contrast MHC-I alleles (like HLA.A*02:11, HLA.A*26:02 and HLA.B*15:17) and MHC-II alleles (like HLA.DRB*115:01 and HLA.DRB*115:06) with high antigen recognition capability were seen to be less common across ethnicities and in most cases were also sparsely distributed even among samples from same ethnicity. In summary, noticeable variability in the potential of the HLA alleles to recognize and present the SARS-CoV2 epitopes to the immune cells was observed. We subsequently investigated if this could have a bearing on the level of antigenic recognition among different ethnic groups.

**Fig. 2:**
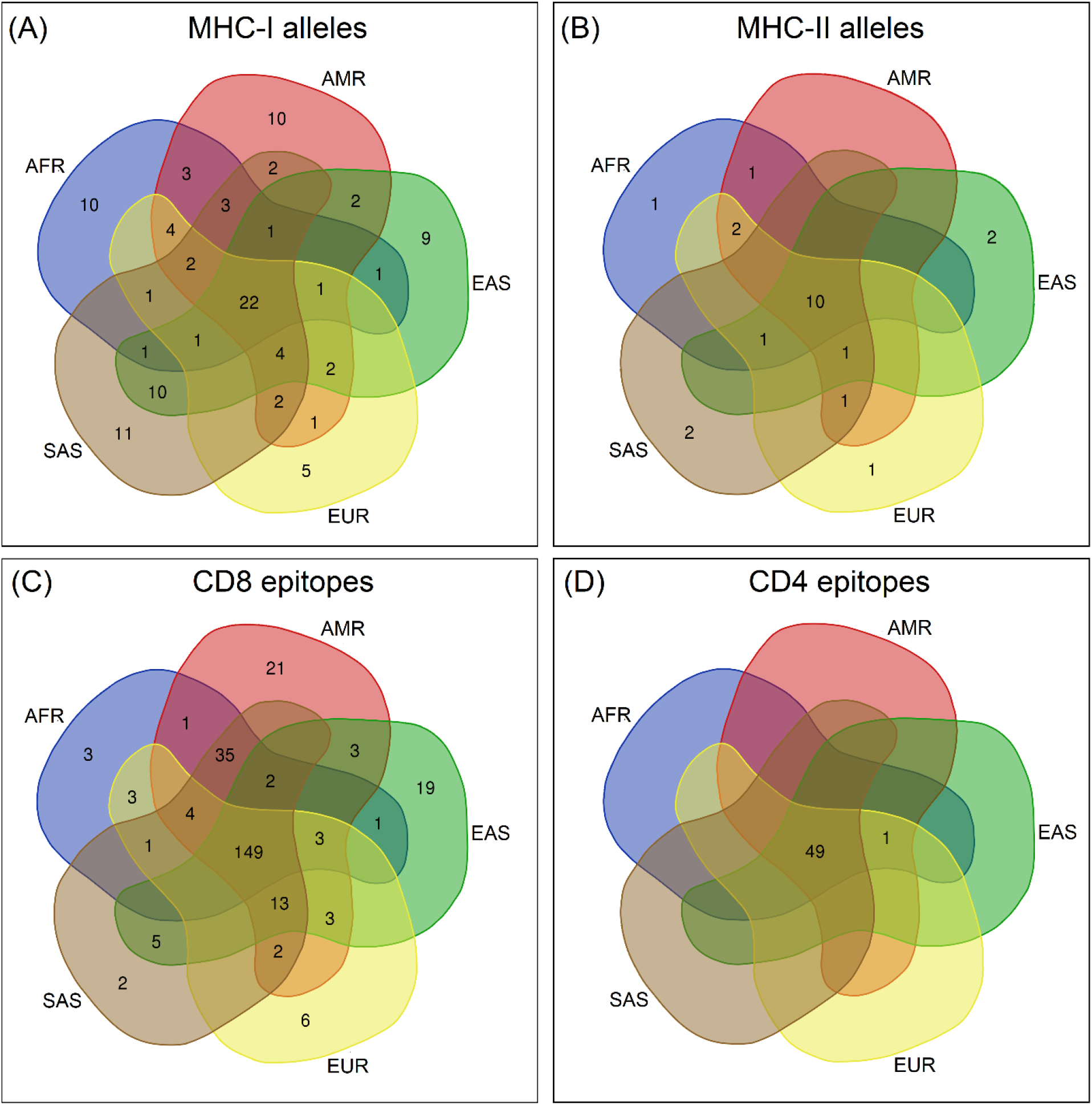
HLA alleles and cognate epitopes shared across super-populations. Euler diagrams representing the intersecting and unique sets of (A) MHC-I and (B) MHC-II alleles observed in different super-populations as well as the sets of corresponding (C) CD8-specific and (D) CD4-specific SARS-CoV2 epitopes recognized by the HLA-system in these super-populations. The super-populations are abbreviated as AFR-Africans; AMR-Ad mixed Americans; EAS-East Asians; EUR-Europeans; SAS-South east Asians.

### SARS-CoV2 Epitope Recognition Capability of HLA alleles Across Ethnicities

The overall trend observed with respect to diversity of the predicted CD8 and CD4 epitopes were comparable to those of their corresponding alleles (MHC-I and MHC-II alleles respectively) across ethnicities (Fig. 1). However, certain subtle differences, specifically with respect to their relative richness, were observed. For example, the change in relative richness (number of observed alleles or epitopes for an ethnicity) among African (AFR) to East Asian (EAS) super-populations in the plots associated with MHC-I alleles and the corresponding CD8 epitopes was apparent. Similarly, a change in richness of MHC-II alleles and CD4 epitopes in case of Ad Mixed American (AMR) to EAS super-populations was also visible. The richness of the antigenic recognition potential among European (EUR) ethnicities was seen to be quite diverse, particularly for CD8 epitopes (Supplementary Table 2). Most notably, the African Caribbean residing in Barbados (ACB) had a marked difference in their antigenic recognition potential of MHC-II molecules as compared to the rest of the AFR ethnicities. Finnish in Finland (FIN), British from England and Scotland (GBR) and Utah residents of northern and western European ancestry (CEU) had a marked difference in CD8 epitopic richness, as compared to the rest of the Europeans. The richness of CD4 epitopes among the South Asian (SAS) ethnicities varied considerably, with Chinese Dai residing in Xishuangbanna (CDX) having the lowest richness among all the ethnic groups. It was speculated that this change in the relative richness could probably be due to the variations in (a) the distribution of HLA alleles among the ethnic populations, and (b) the disparity in the capacity of some of the HLA alleles to recognize multiple epitopes. To further introspect into this aspect, the richness of MHC-I and MHC-II alleles and their recognition potentials for CD8 and CD4 epitopes among the 26 different ethnic groups were probed in tandem (Fig. 3). As evident from the mirror-plot (Fig. 3), probing in terms of the ratio of the richness of HLA alleles present (represented along negative x-axis) to the richness of epitopes recognized (positive x-axis) revealed interesting patterns among the various ethnicities. Most EAS and AFR ethnicities, except for Yoruba from Ibadan (YRI) and Japanese from Tokyo (JPT), had lower epitope recognition capabilities with respect to the number of MHC-I alleles present. For MHC-II alleles, while this ratio was found to be the least among the AMR ethnicities, EAS sub-populations (ethnicities) showed wide variations, with JPT and Southern Han Chinese (CHS) exhibiting the highest values among all ethnicities.

**Fig. 3:**
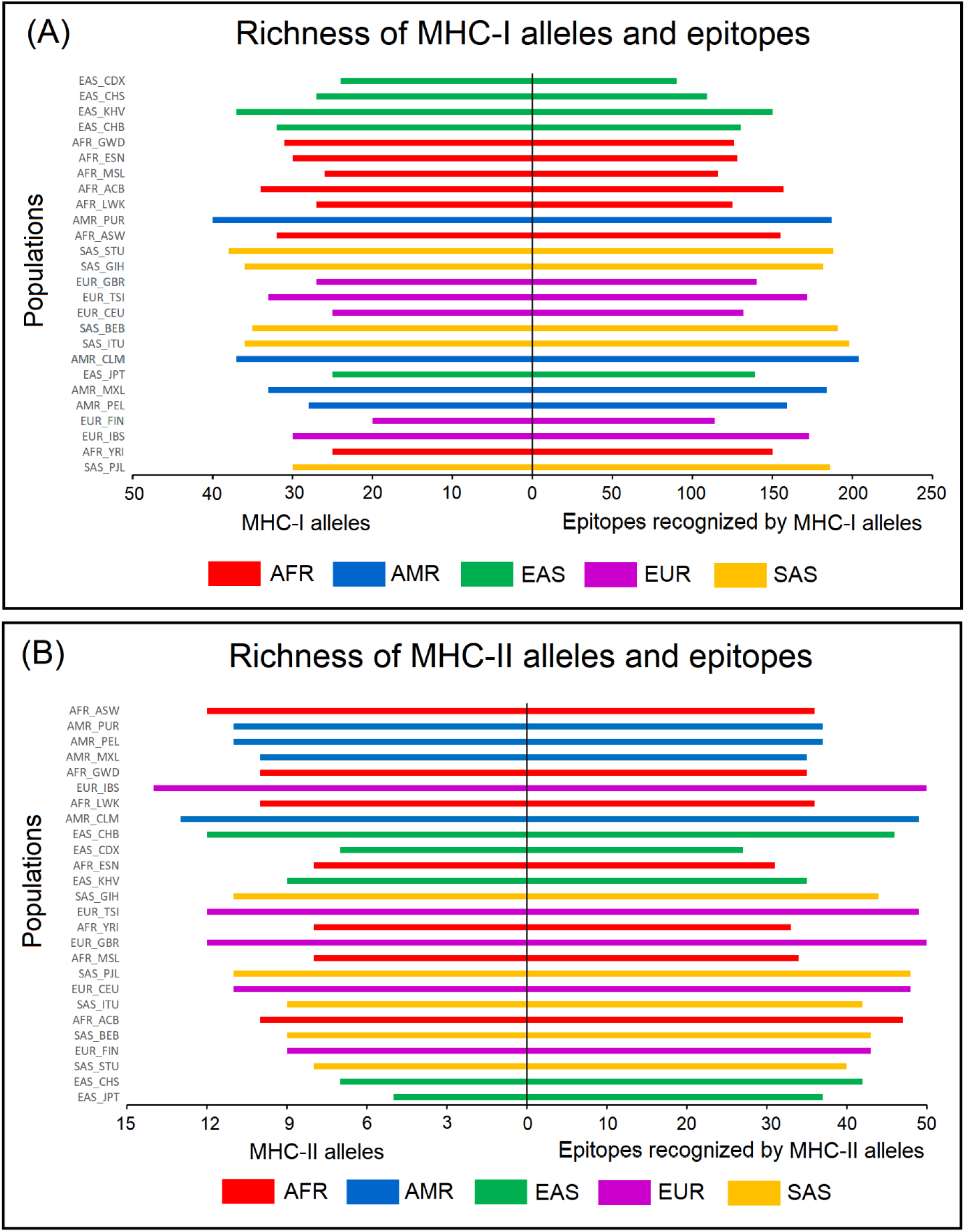
Comparative view of HLA allelic richness and cognate epitopes across populations. Mirror-plots indicating the richness of MHC alleles in different populations (along the left half of the plots) along with the richness of SARS-CoV2 epitopes predicted to be recognized by these HLA alleles (along the right half of the plots). (A) MHC-I alleles and predicted cognate CD8-specific epitopes and (B) MHC-II alleles and predicted cognate CD4-specific epitopes, across 26 different populations (ethnic groups). In each of the plots, the ethnicities were sorted along the vertical axis based on the ratio of the richness of MHC alleles to the richness of epitopes recognized. Super-population names are abbreviated as AFR-Africans; AMR-Ad mixed Americans; EAS-East Asians; EUR-Europeans; SAS-South east Asians, and prefixed to each of the population names in the plots, as well as represented with horizontal bars of specific colors. The abbreviations used for names of different populations/ ethnic groups are in accordance with those originally reported by the 1000 genomes project and the International Genome Sample Resource (IGSR) (https://www.internationalgenome.org/faq/which-populations-are-part-your-study/).

As indicated in the Fig. 2 (details in Supplementary Table 3), 149 CD8 and 49 CD4 epitopes were found to be common across five super-populations. In contrast to the MHC-I alleles (wherein SAS and EAS had the highest commonalities), SAS and AMR were found to have the highest overlap in terms of recognizing CD8 epitopes. Additionally, AMR and EAS were found to have the potential to identify a significant number of epitopes which were not identified by the MHC-I complexes of the other super-populations. Most notably, the CD4 epitopes were seen to be equally recognized among all super-populations. The trends in recognition of epitopes by different alleles present across various ethnicities provided further insights (Supplementary Figures 3 and 4). The antigenic peptide FLLPSLATV (Epitope 3 from nsp6) appeared to be the most recognized CD8-specific epitope across ethnicities. Notably, this epitope could be recognized by 14 HLA allelic variants, the highest among all the CD8-specific SARS-CoV2 antigens recognized in this study (Supplementary Figure 3 and Supplementary Table 1). However, for both the CD8 and CD4 epitopes, there were no observable correlations between the epitopes which were recognized by higher number of HLA variants and those which were most common across ethnicities. For example, while the CD4-dependent antigenic peptides ESPFVMMSAPPAQYE and KDQVILLNKHIDAYK (Epitopes 277 and 309 respectively) were recognized by four HLA variants each, the maximum among the CD4-specific SARS-CoV2 antigens recognized in this study (Supplementary Figure 4 and Supplementary Table 1), Epitope309 was not as common across different ethnicities as Epitope277. The above observations indicate that there can be certain discernable differences at an overall population level, with respect to the CD4 and CD8 cell mediated immune response against SARS-CoV2.

### Immune Sensitivity to SARS-CoV2 for Individuals Belonging to Various Ethnic Groups

The potential of each of the individual subjects (comprising the 26 ethnic groups and five super-populations) to recognize the SARS-CoV2 epitopes was determined (Supplementary Table 4). Based on this, the overall epitope recognition potential of the ethnicities and super-populations were computed. Supplementary Figures 5 and 6 depicts the average count of epitopes recognized by individuals representing super-population and ethnic groups respectively. East Asians (EAS) and Africans (AFR) reported low potentials to recognize both CD8 and CD4 epitopes. In contrast, ethnicities comprising the European (EUR) and South Asian (SAS) super-populations, showed high potentials to recognize all forms of SARS-CoV2 epitopes. The Peruvians from Lima (PEL) exhibited an interesting pattern with extremely low CD4, but very high CD8 epitope recognition capacity. Based on the above observations, we further probed if there were any statistically significant differences between the epitope recognition potential among the various ethnicities and super-populations (see Materials and Methods). The p-values for the Kruskal-Wallis test among all the super-populations and ethnic groups were seen to be 0.005257 and 0.004022 at 95% confidence interval, thereby implying significant difference in epitope recognition potential in at least one of the five super-populations and 26 ethnic groups. Indeed, the results obtained from Wilcoxon signed-rank test (see Materials and Methods) indicated substantial differences in the recognition potentials of both CD8 and CD4 epitopes among super-populations as well as ethnicities except for AMR super-population (Supplementary Table 5). Further, to check for any major differences in the epitope recognition potentials between individuals of different ethnicities, Dunn’s test was performed (see Materials and Methods). Most notably, among the SAS ethnicities, GIH - Gujarati Indian from Houston, Texas and PJL - Punjabi from Lahore, Pakistan showed significant mean rank differences from STU - Sri Lankan Tamil from the UK (Table 1 and Supplementary Table 6). PUR - Puerto Ricans from Puerto Rico and PEL - Peruvians from Lima, Peru (among AMR) and JPT - Japanese in Tokyo, Japan and CHB - Han Chinese in Beijing, China (among EAS) also demonstrated appreciable differences. In contrast, despite diverse origins, the mean rank differences between certain ethnic groups (such as, SAS_STU versus EUR_IBS) were found to be extremely low (Table 1 and Supplementary Table 6). While the prevalence of specific HLA alleles in the gene pool of a population or an ethnic group provides an overall idea about how well recognized the SARS-CoV2 epitopes would be in that population, the immune response of each of the individuals would be independently governed by their own allelic make-up. The above results provide an average estimate of (CD4 and CD8 cell mediated) immune sensitivity to SARS-CoV2 for individuals belonging to population and highlights inter-ethnic differences therein.

**Table 1:** Dunn’s test results depicting variation in the SARS-CoV2 epitope recognition potential between different ethnicities. Differences in the epitope recognition potential among individuals of different ethnic groups have been presented in terms of mean rank differences and p-values of the test (see Supplementary Table 6 for complete table with all the ethnic groups). Populations (or ethnic groups) exhibiting significant differences (with p-values < 0.05) have been highlighted. Positive mean rank difference indicates the first ethnic group in a compared pair has higher epitope recognition potential and vice-versa.

| Comparisons | Mean Rank Difference | P-values |
| --- | --- | --- |
| Ethnic group belonging to same super-population exhibiting high mean rank difference (5 maximum values) |  |  |
| SAS_STU vs SAS_GIH | -338.08 | **2.77E-03** |
| SAS_STU vs SAS_PJL | -267.99 | **1.97E-02** |
| AMR_PUR vs AMR_PEL | 237.82 | 5.30E-02 |
| AMR_PEL vs AMR_MXL | -229.11 | 9.80E-02 |
| EAS_JPT vs EAS_CHB | -222.67 | 6.02E-02 |
| Ethnic group belonging to different super-population exhibiting low mean rank difference (5 minimum values) |  |  |
| SAS_STU vs EUR_IBS | 0.29 | 9.98E-01 |
| EAS_CDX vs AMR_PEL | 6.29 | 9.63E-01 |
| SAS_BEB vs EUR_FIN | -13.61 | 9.16E-01 |
| EAS_JPT vs AFR_LWK | 15.77 | 9.02E-01 |
| AMR_PEL vs AFR_ASW | 21.65 | 8.83E-01 |

Note: Super-population names are abbreviated as AFR- Africans; AMR- Ad mixed Americans; EAS- East Asians; EUR- Europeans; SAS- South east Asians, and prefixed to each of the population names in the table. The abbreviations used for names of different populations/ ethnic groups are in accordance with those originally reported by the 1000 genomes project and the International Genome Sample Resource (IGSR) (https://www.internationalgenome.org/faq/which-populations-are-part-your-study/).

### Impact of Genetic Variability of SARS-CoV2 on Immune Recognition

In addition to the HLA makeup, yet another factor which could determine the fate of the infection process is the genetic variation in the SARS-CoV2 genomes. Genetic variation could lead to epitope modifications, which in turn, might alter the binding affinity/ recognition of the viral epitopes by MHC-I/II alleles. To accommodate for this factor, the epitope signature of 40,342 SARS-CoV2 genomes, were generated (see Materials and Methods) based on the 326 epitopes predicted for the reference genome and their 2989 variants (Table 2 and Supplementary Table 7). Apart from the original 326 epitopes, only 12 of the 2989 epitope variants were seen to be present in more than 0.5% of the studied SARS-CoV2 genomes. Moreover, some of the variants were found to exclusively co-occur among a sub-set of strains isolated from certain geographies, most prominently among those isolated from India or United Kingdom. Even more significantly, a particular epitope variant (SEVGPEHSL at position 376 on ORF1a) was noted to be exclusively absent among the SARS-CoV2 genome sequences isolated and sequenced in Iran (Supplementary Table 7). It was further observed that while the more frequent epitope variants (those observed in more than 0.5% of the studied SARS-CoV2 genomes) exhibited a mixed pattern of altered immunological behaviors, the human MHC-I/II alleles did not have a high binding affinity/ recognition potential for a large proportion (1346 out of 2651) of the lesser frequent SARS-CoV2 epitope variants (Table 2).

**Table 2:** Overview of variations in the epitopic regions in different SARS-CoV2 strains. Summary of the different parameters associated with the epitope variance with the 40342 SARS-CoV2 strains analyzed in the study has been tabulated.

| **Parameter** | **Statistics** |
| --- | --- |
| Number of genomes analyzed | 40342 |
| Number of epitopes in the reference genome (NCBI Genbank accession no: MN908947) | 326 |
| Number of additional epitopes (variants) identified among the analyzed genomes | 2663 |
| Total number of epitopes (reference epitopes + variants) | 2989 |
| Epitope variants present in at-least 0.5% of the studied genomes - more frequent epitope variants | 12 |
| Epitope variants present in less than 0.5% of the studied genomes - less frequent epitope variants | 2651 |
| Epitope variants with no change in epitope recognition by HLA alleles with respect to the reference epitopes | 896 |
| Epitope variants recognized by higher number of HLA alleles with respect to the reference epitopes | 417 |
| Epitope variants recognized by fewer number of HLA alleles with respect to the reference epitopes | 406 |
| Epitope variants not recognized by HLA system | 944 |
| Number of less frequent epitope variants not well recognized by HLA system when compared to the reference epitope | 1346 |
| Number of more frequent epitope variants not well recognized by HLA system when compared to the reference epitope | 4 |
| Number of more frequent epitope variants better recognized by HLA system when compared to the reference epitope | 4 |

While it would be interesting to understand the effect of these variations in the viral genome on the antigenic recognition potentials among different ethnicities no major effects were expected, given that only 12 epitope variants were present in at least 0.5% of the studied SARS-CoV2 genomes. Even in the hypothetical scenario, wherein every population/ ethnicity is simultaneously exposed to all the 40,342 viral variants, the individual antigenic recognition potential would be driven by the originally identified 326 epitopes, and not by the lesser frequent epitope variants. Considering the continuously evolving genome of SARS-CoV2, which entails selection/ retention of specific epitope variants over time in the newly emerging genomes, an analysis was performed to check if there were any temporal changes in the susceptibility to Covid-19 in any of the ethnicities (see Materials and Methods). Even in this case (Fig. 4), it was noted that temporal variations in the SARS-CoV2 genome, while resulting in certain changes in its epitope signature, did not appreciably alter the epitope recognition ability among ethnic groups, at least at a population level (also see Supplementary Figure 7). Since ethnic groups (and people from same geography) are known to predominantly encoded for certain HLA gene variations, it may be perceived that the SARS-CoV2 epitope recognition potential of an ethnic group is not likely to change owing to the temporal variations in the viral genome. Further, it will be appreciated that although the said changes in the viral genomes might alter the biological functioning of the viral genes, yet they remain largely inconsequential with respect to immune determination in the host.

**Fig. 4:**
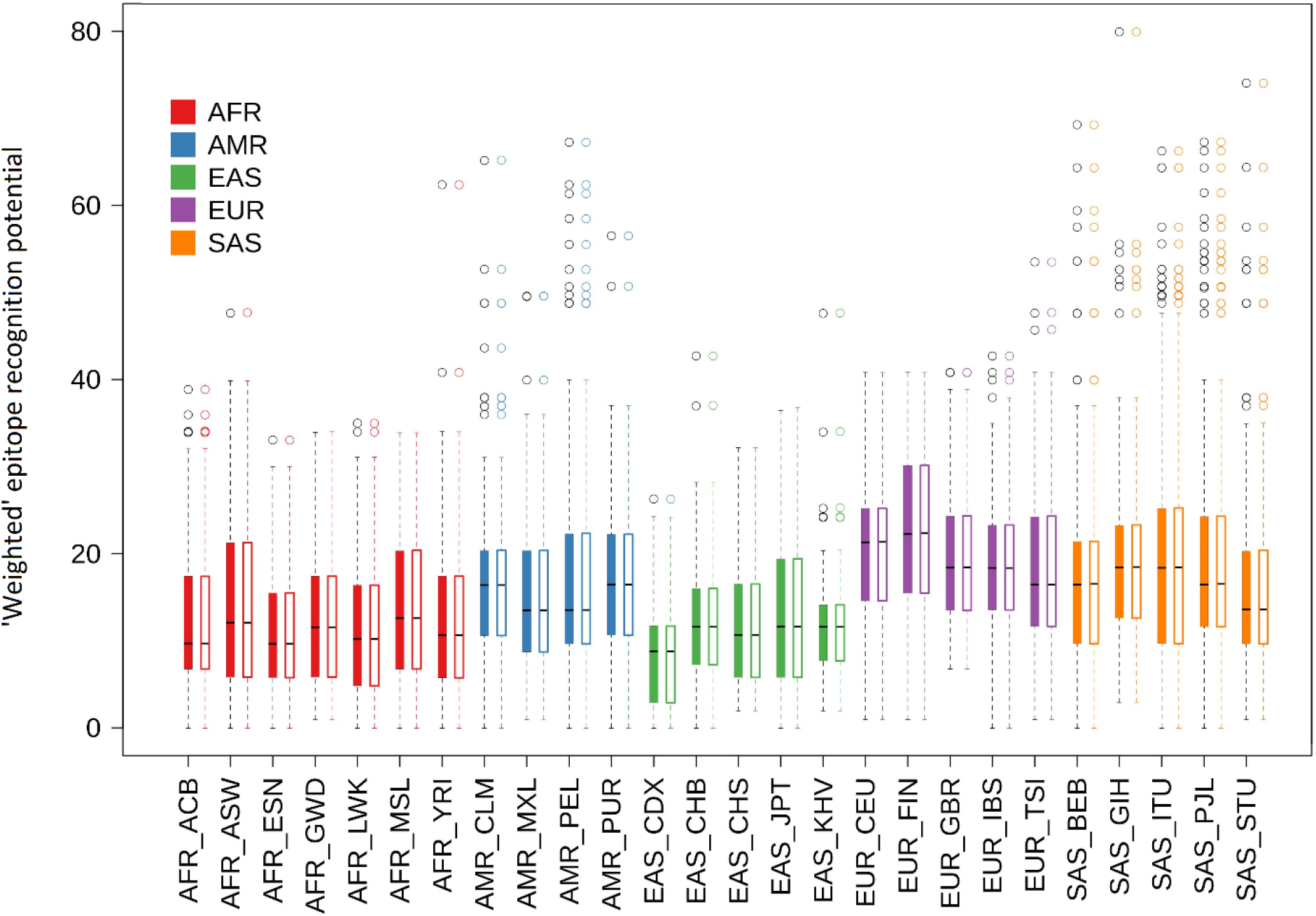
Average epitope recognition potential against evolving SARS-CoV2 variants. Representation of the average of predicted SARS-CoV2 ‘weighted’ epitope recognition potential of individuals from different populations (ethnic groups), considering different SARS-CoV2 genome variants observed across geographies. For each population (ethnic-group), there are two box-plots indicating their average ‘weighted’ epitope recognition potential (see Materials and Methods). For each population, the color-filled box on the left corresponds to the epitopes predicted from the first 10,000 SARS-CoV2 genomes (out of a total 40,342 genomes analyzed in the study) when arranged in the chronological order of their collection dates. The corresponding unfilled boxes on the right corresponds to the epitopes predicted from the last 10,000 SARS-CoV2 genomes based on their collection dates. Although the SARS-CoV2 genomes may be evolving over time, no observable variation in the way its epitope repertoire is recognized by individuals at an overall population level could be noted. Out identification capacity among different ethnic groups. Each data-point on the plots represents the average of the number of epitopes in a given SARS-CoV2 genome that could be identified by all the individuals in an ethnic population. Dark black lines, for each of the ethnic groups, indicate the mean value for these data-points for the studied 40,342 SARS-CoV2 genomes. Super-population names are abbreviated as AFR-Africans; AMR-Ad mixed Americans; EAS-East Asians; EUR-Europeans; SAS-South east Asians, and prefixed to each of the population names in the plots, as well as represented with boxes drawn in specific colors. The abbreviations used for names of different populations/ ethnic groups are in accordance with those originally reported by the 1000 genomes project and the International Genome Sample Resource (IGSR) (https://www.internationalgenome.org/faq/which-populations-are-part-your-study/).

## DISCUSSIONS

There have been three major coronavirus associated SARS outbreaks in the last 20 years. While there have been extensive research regarding the pathophysiology of these viruses (Y. Chen et al., 2020; Hillen et al., 2020; Hoffmann et al., 2020; Krichel et al., 2020; Kumar et al., 2020; Letko et al., 2020; Li et al., 2020; Satarker and Nampoothiri, 2020; Song et al., 2020; Wang et al., 2020; Xu et al., 2020), as yet we have limited knowledge into the factors affecting susceptibility to SARS-CoV2 infection and manifestation of Covid-19. Some scientists have opined that the mortality rate in Covid-19 could be linked to the genomic profile of the infecting virus (Toyoshima et al., 2020; “WHO | Variant analysis of SARS-CoV-2 genomes,” n.d.) which seem to mutate at a rate similar to most RNA viruses. Further, an underlying/ pre-existing medical condition might be considered to be an added risk towards increasing the severity/ complications associated with Covid-19 infection (CDC, 2020). In addition, the host genetic makeup could play a decisive role in disease manifestation. In this context, polymorphic genes like the Human Leukocyte Antigens (HLA) system which is known to vary significantly among individuals of different ethnicities assumes importance. Thus, the outcome of an infection event is multi-factorial and at least dependent on (a) genotype of the virus and (b) host cell surface (MHC) molecules destined to present viral antigenic peptides to the human immune cells.

To understand this, the antigenic peptides from over 40,000 SARS-CoV2 genomes were predicted for all major HLA alleles, as documented in the 1000 genomes project (ftp://ftp.1000genomes.ebi.ac.uk/vol1/ftp/data_collections/HLA_types/). The state-of-the-art tools NetMHCpan 4.1 and NetMHCIIpan 4.0 were used for the purpose in the current work (Reynisson et al., 2020). It may be noted that most of other available tools and web-services for predicting MHC-I and MHC-II epitopes (Jespersen et al., 2017; Paul et al., 2016; Saha and Raghava, 2004; Zhang et al., 2009) were not equipped to handle the entire HLA allelic set that had been reported in the 1000 genomes project data. Further, most of the other tools also did not have a provision to identify variable length MHC-I epitopes. Consequently, some of the earlier studies associate with *in silico* identification of SARS-CoV2 epitopes (Abdelmageed et al., 2020; Dong et al., 2020; Lin et al., 2020; Naz et al., 2020; TOPUZOĞULLARI et al., 2020) are likely to have suffered from the mentioned limitations associated with these tools (Jespersen et al., 2017; Paul et al., 2016; Saha and Raghava, 2004; Zhang et al., 2009). In other words, these studies might not have arrived at a list of epitopes as comprehensive as the one mentioned in the current study. Further, studies following a combinatorial approach (H.-Z. Chen et al., 2020; Zaheer et al., 2020) are also expected to miss some of the epitopes reported in this study. It may however be noted that like most previous studies, the current study refrained from predicting potential B-cell epitopes, due to the computational complexities involved in the process and may be considered a limitation of this exercise. Most B-cell epitopes are conformational in nature and comprise of discontinuous amino acid stretches which are difficult to be identified (with adequate confidence) from genomic information alone (Wang et al., 2011).

A total of 276 CD8 (involving 108 MHC class-I alleles) and 50 CD4 (involving 22 MHC class-II alleles) SARS-CoV2 epitopes were predicted in the current study. The highest number of SARS-CoV2 epitopes were presented by the MHC class-I associated HLA alleles HLA-A*02:11, HLA.A*26:02, HLA.B*15:17, HLA.A*24:03, HLA.B*35.41, and MHC class-II associated HLA alleles HLA-DRB1*01:01, HLA.DRB*115:01, HLA.DRB*115:06, HLA.DRB*101:02. The diversity of both HLA alleles encoded in the genome as well as the epitopes recognized by their HLA systems was observed to vary across the super-populations (Fig. 1) and among the ethnic groups (Supplementary Table 2) comprising these super-populations. While the super-populations were seen to cluster in terms of the MHC-I genes encoded in the genome, such patterns could not be observed for MHC-II genes (Supplementary Figures 1-2). In addition, certain HLA alleles were found to be more efficient in recognizing SARS-CoV2 epitopes as compared to others. Given the skewed distribution of these HLA alleles among a few of the ethnic groups, contrasting characteristics with respect to the ratio of the number of epitopes to the number of HLA alleles involved in their recognition, among each of the ethnic groups were observed, even among ethnicities from the same super-population (Fig. 3). For example, GIH and PJL showed significant mean rank differences from STU among the ethnicities from the SAS super-population (Table 1 and Supplementary Table 6). Conversely, in a few cases, the mean rank differences between certain ethnic groups from different super-populations (such as, SAS_STU versus EUR_IBS) were found to be extremely low (Table 1 and Supplementary Table 6).

In this study, an attempt was further made to gauge the variations in the SARS-CoV2 epitope regions resulting from temporal changes in the viral genome and its effect on the epitope recognition potential across the ethnicities worldwide. The SARS-CoV2 genomic variation data which was obtained for different geographies over a six month period indicated that although the SARS-CoV2 genomic variations altered the overall predicted SARS-COV2 epitope signature profile (Table 2 and Supplementary Table 7), most of the alternate epitopes were not as common and occurred in ≤ 0.5% of the analyzed genomes. Further, the overall capacity of an ethnic group to recognize SARS-CoV2 did not seem to change either (a) on encountering a new viral strain (Fig. 4) or (b) over the course of the pandemic (Supplementary Figure 7). Moreover, a minimal set of SARS-CoV2 epitopes was identified, which can be recognized by the HLA repertoire of individuals from all ethnicities (Table 3). Candidates from this set of predicted antigenic peptides could provide an opportunity to design newer vaccines against Covid-19 (refer to Supplementary File 1). While most of the currently available Covid-19 vaccines are designed to target the viral spike protein, a recent study while inspecting T-cell memory from patients recovering from Covid-19 infection commented on the prospect of considering non-spike proteins within future Covid-19 vaccine designs (Peng et al., 2020). Moreover, the outcome of this study can also aid in disease prognosis and designing of personalized therapy regimes for Covid-19 patients. The idea of providing personalized therapy to Covid-19 patients, especially those requiring critical care is already under clinical consideration (Cacciapuoti et al., 2020; Fang and Schooley, 2020; Garcia-Vidal et al., 2020). In addition, results presented herein could also prove beneficial in understanding/ countering a future flu/ pneumonia outbreak involving a similar lineage B *Betacoronavirus*. Given that the world has already witnessed two such major outbreaks in less than 20 years, it would be prudent to prepare blueprints of a more potent coronavirus vaccine, particularly against the lineage B *Betacoronavirus*, before the next outbreak.

**Table 3:**
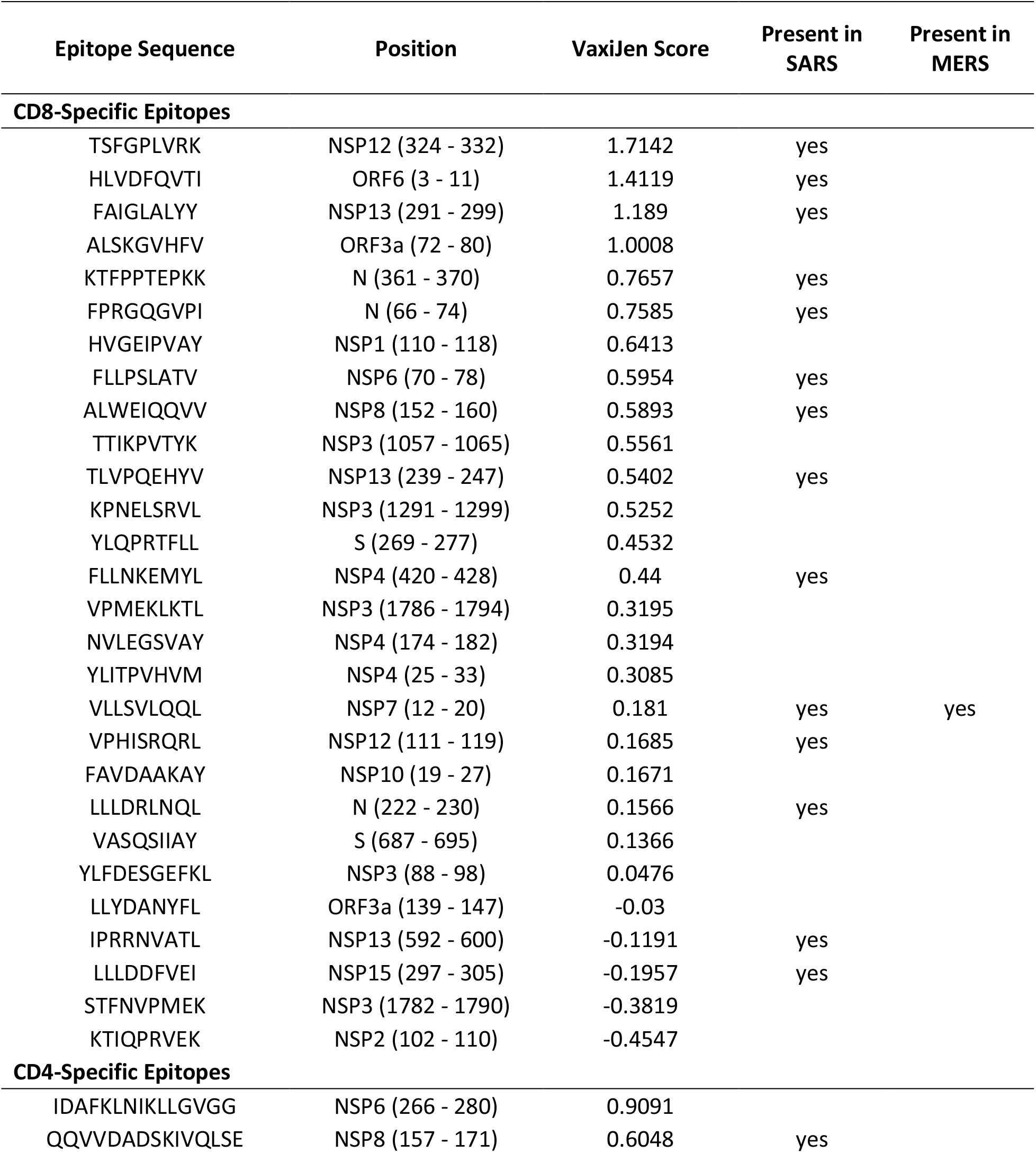
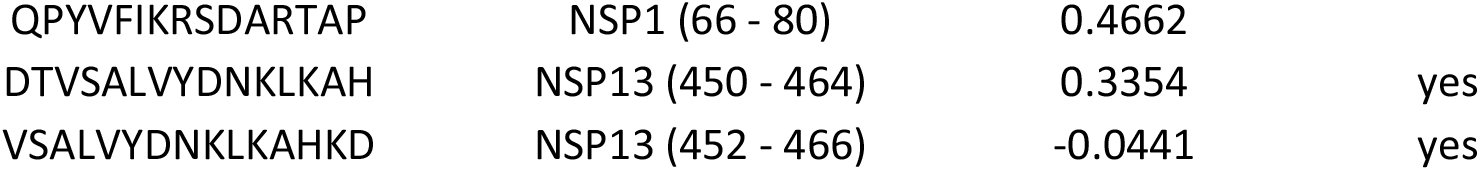
List of SASR-CoV2 antigenic peptides for designing a potential multi-epitope vaccine. The table comprises of 28 CD8 and 5 CD4-specific peptides that are suggested as potential multi-epitope vaccine candidates as determined by their ability to be recognized by the HLA repertoire of the individuals of all the five super-populations. The position of these peptides on the respective SARS-CoV2 proteins is also mentioned. The list is sorted based on descending order of VaxiJen scores. The presence of these peptides in SARS coronavirus and MERS coronavirus proteomes is also indicated.

The context and results of the presented work is somewhat aligned to another intriguing aspect of the prevailing pandemic with respect to disease severity and case fatality rate (CFR). Appreciable spatial and temporal variations in CFR have been observed across geographies and suitable explanation(s) for these variations have eluded researchers, given the wide array of possible confounding factors. A key observation in this regard pertains to the economic status of a country. Death rates due to Covid-19 has been observed to be positively associated with GDP (per-capita) in multiple studies (Cao et al., 2020; Sorci et al., 2020). Higher death rates despite (expected) better access to healthcare in high-income populations seems outright counter intuitive. While some scientists have tried to explain these observations citing higher life-expectancy and consequently an older population in high-income countries, who would be more susceptible to Covid-19, others have hinted at the possibility of the “so called hygiene-hypothesis” at play (Bloomfield et al., 2016; Chatterjee et al., 2021). The results of the current study unravel yet another interesting aspect in this context pertaining to the genetic diversity. Severe cases of Covid-19 disease have been seen to be characterized by higher levels of inflammatory cytokines and CD8+ T cell exhaustion (Yang et al., 2020). Reports have also indicated that in case of milder Covid-19 infections, not leading to death and other complications, higher proportions of SARS-CoV2 specific CD8+ T cells have been identified (Peng et al., 2020). Our results indicate that the heavily affected European population (EUR) tends to harbor a larger fraction of MHC-II alleles in their gene-pool, which are specific to SARS-CoV2 epitopes. On the other hand, the South Asian (SAS) population, having a relatively lower fatality rate, exhibited a relatively larger proportion of MHC-I alleles specific to the SARS-CoV2 epitopes. It is worth further investigation whether a larger pool of MHC-I alleles presenting SARS-CoV2 antigens to CD8+ T cells in the SAS population indeed can be linked to the apparently lower fatality rate in this region. A recent study while reporting an inverse associations between MHC-I epitopes and mortality rates also indicated at this possibility (Wilson et al., 2021). Moreover, any possible relationship of the larger pool of the MHC-II alleles in the EUR population with CD4+ T Cell activation, cytokine production leading to a disproportionate immune response (or a so called “cytokine-storm”) will also be intriguing to explore. While the current work is limited by the number of representative individuals genotyped for each population in the 1000 genomes project, the results nonetheless provide an overview of possible trends in protective immunity and T-cell responses against SARS-CoV2 across different geographies/ ethnicities.

Overall, to our knowledge, this is the first ever account to capture the effects of the evolution of SARS-CoV2 genome on its potential interactions with the human HLA gene products in a global perspective. This assumes immense importance in the context of vaccine development and its efficacy against the newer lineages of SARS-CoV2. It is however important to note that the nature of the findings was determined by (a) the availability of data in public repositories and (b) the efficiency of the available bioinformatic tools. As a result, insights into some of the aspects of immune response, such as, if there are any B-cell specific conformational epitopes in SARS-CoV2, expression levels of the MHC alleles in response to Covid-19 infection and its variations across ethnicities, age groups, co-morbidities, etc., if any, could not be obtained. Nonetheless, the observations presented in this study are expected to highlight the need of understanding the crosstalk of the pathogen with the components of HLA system. This, we believe, would have far reaching consequences in our coexistence with SARS-CoV2 and other RNA viruses which we may encounter in future.

## MATERIALS AND METHODS

### Data Acquisition

Full length sequences of all SARS-CoV2 proteins (reference sequences) were obtained from NCBI (GenBank Accession no. MN908947). Further, individual non-structural protein sequences were also obtained from NCBI (NCBI Accession no. YP_009725300.1 to YP_009725312.1). These protein sequences were used for predicting antigenic peptides (epitopes) on the viral proteome. The human HLA allelic profiles were obtained from the 1000 genomes project (ftp://ftp.1000genomes.ebi.ac.uk/vol1/ftp/data_collections/HLA_types/). This data corresponded to the distribution of major MHC-I and MHC-II HLA types, viz. HLA-A, HLA-B, HLA-C, HLA-DRB and HLA-DQB from 2693 individuals (Samples) who belonged to five super-populations (Regions) and 26 ethnicities (Populations) as described in the 1000 genomes project (Abi-Rached et al., 2018). Further, high quality fully sequenced genome sequences of SARS-CoV2 were obtained from GISAID (https://www.gisaid.org/) (Elbe and Buckland-Merrett, 2017; Shu and McCauley, 2017). Genomic data and corresponding metadata for 40342 genomes (which were deposited to GISAID till 11^th^ June 2020) have been used in this analysis (see Supplementary Table 10 for details).

### Epitope Prediction

NetMHCpan 4.1 (http://www.cbs.dtu.dk/services/NetMHCpan/) and NetMHCIIpan 4.0 (http://www.cbs.dtu.dk/services/NetMHCIIpan/) were used with default parameters for identifying strong and weak binders (i.e., 0.5 and 2 for NetMHCpan and 1 and 5 for NetMHCIIpan) (Reynisson et al., 2020) to generate epitopes from SARS-CoV2 protein sequences against 226 MHC-I and 75 MHC-II alleles reported in 1000 genomes project. Variable length (8 to 14-mer) CD8 epitopes (mapping to MHC-I alleles) and 15-mer CD4 epitopes (corresponding to MHC-II alleles) were thus obtained which were further filtered for subsequent use. These epitopes were then filtered based on the prediction scores. In case multiple overlapping epitopes were recognized by the same HLA allele, the epitope HLA allele pairs with the higher prediction score was retained. Next, all predictions with scores < 0.95 were discarded to retain only epitope HLA allele pairs with high binding affinity (recognition capacity). This list comprised of 326 (276 CD8 and 50 CD4) epitopes having high prediction scores for 130 (108 MHC-I and 22 MHC-II) HLA alleles and was used for all further analysis.

### Computing Allele and Epitope Distribution Across Populations

Based on the data obtained from the 1000 genomes project, the overall distribution of MHC-I and MHC-II alleles among the 26 ethnic groups and five super-populations were determined. In addition, the distribution of CD8 and CD4 epitopes recognized by individuals (Samples) across these ethnicities and super-populations were also ascertained from the filtered epitope prediction results (Supplementary Tables 4 and 5). These were used to compute various diversity indices (viz. richness, Shannon index and Simpson index) for MHC-I and MHC-II alleles as well as CD8 and CD4 epitopes. While the phyloseq package (from R version 3.6) was used to calculate diversity, the ggplot2 package in R version 3.6 was used for the visualization. The data was also used to generate Euler plots to depict the distribution of alleles across super-populations as well as the epitopes which would be recognized by these alleles. For this purpose, an online Euler diagram generation tool (http://bioinformatics.psb.ugent.be/webtools/Venn/) was used. Moreover, heatmaps to visually inspect the frequency of distribution of HLA alleles (both MHC-I and MHC-II) and the identified epitopes across the 26 ethnic groups were created using the heatmap.2 function of gplots library in R version 3.6 (Supplementary Figures 1-4). For this purpose, ‘frequency’ of an allele (or an identified epitope) in a given ethnicity was computed as number of individuals (Samples) from that ethnicity which encoded for the allele (or could recognize the epitope) divided by the total number of individuals (Samples) representing the ethnic group. The heatmaps were further augmented with a colour-key along the vertical axis depicting (a) the number of epitopes recognized by the HLA alleles (Supplementary Figures 1 and 2), and (b) the number of HLA alleles which could identify a given epitope (Supplementary Figures 3 and 4) according to the data provided in Supplementary Table 2. In addition, the data represented in the heat maps were also used to hierarchically cluster the ethnic groups based on the HLA alleles they encode and the epitopes they could identify (Supplementary Figures 1-4). The hclust package in R version 3.6 was used for this purpose.

### Statistical Analysis

In order to probe for any statistical differences between the number of epitopes recognized by the individuals (epitope recognition potential) among the various ethnicities and super-populations, Kruskal-Wallis test was conducted after grouping the individuals (Samples) into (a) super-populations and (b) ethnic levels. Additionally, Wilcoxon signed-rank test was also performed to test whether mean ranks of the epitope recognition potential of a super-population (and/or ethnicity) differed with respect to that of all other super-populations (and/or ethnicities) combined (Supplementary Table 5). The above tests were conducted both for the CD8 and CD4 epitopes individually as well as in combination. The p-values were corrected for multiple testing using Benjamini-Hochberg (BH) correction method. Further, the specific super-population/ ethnic group pairs with significant differences were identified using Dunn’s test with Benjamini-Hochberg (BH) correction (Supplementary Table 6). The DescTools package in R version 3.6 was used for this purpose.

### Epitope Variations Among SARS-CoV2 Genomes

Nextstrain/augur pipeline (accessed on 21^st^ April, 2020 from https://github.com/nextstrain/ncov) was used to align 40342 SARS-CoV2 genome sequences downloaded from GISAID using Wuhan-Hu-1/2019, as a reference sequence using MAFFT (Katoh and Standley, 2013). Conforming to the Nextstrain protocol, 130 nucleotides from 5’-end and 50 nucleotides from the 3’-end as well as single nucleotide positions 18529, 29849, 29851, 29853 were masked from multiple sequence alignment (MSA). An initial maximum likelihood (raw) phylogenetic tree (GTR model) was built using IQ-TREE (Nguyen et al., 2015) and further refined using default parameters in the pipeline. The tree was then processed to construct a TimeTree having the ancestor of the following two genomes, Wuhan-Hu-1/2019 and Wuhan/WH01/2019, at its root. Finally, using augur’s translation step, a translated MSA of all 14 SARS-CoV2 proteins were retrieved. The 326 epitopic regions (w.r.t. reference genome Wuhan/Hu-1/2019) were cropped out from the translated MSA, and amino acid variations occurring in each epitope across 40342 SARS-CoV2 isolates were obtained. A total of 2989 variations could be identified in the 326 reference epitopes.

### Analysis of the Genetic Variability of SARS-CoV2 on Immune Recognition

A ‘weighted’ epitope recognition potential for every individual was calculated using the cumulated value of the prediction scores (≥ 0.95) of the epitopes present in SARS-CoV2 interacting with alleles of the individual. This was calculated using both the CD8 epitopes interacting with MHCI alleles and CD4 epitopes interacting with MHCII alleles. Subsequently, we calculated the ‘weighted’ epitope recognition potential for all the 2693 individuals against 40342 variants of SARS-CoV2 genomes in the present study. To evaluate any possible effect of mutation occurring/accumulating in the SARS-CoV2 genome over-time on immune recognition of the virus among different ethnicities, the mean ‘weighted’ epitope recognition potential for each individual belonging to an ethnic group was calculated with epitopes predicted from the first 10,000 and last 10,000 SARS-CoV2 genomes arranged in the chronological order of their collection dates (Fig. 4). Further, we also tried to estimate the variations of ‘weighted’ epitope recognition potential within each ethnic group for all the 40342 variants of SARS-CoV2 genomes (Supplementary Figure 7), where the mean ‘weighted’ epitope recognition potential (of all the individuals belonging to an ethnic group) for each genome was calculated and arranged in the chronological order of their collection dates (in months).

## Data Availability

All data are available as supplementary to the manuscript.

## AUTHOR CONTRIBUTIONS

TB, AD and SSM conceptualized the work and designed the protocol. NP, NKP and SB performed all the analyses. TB, NP, and AD wrote the manuscript. SSM supervised the project and reviewed the manuscript. All authors read and approved the final version of the manuscript.

## ACKNOWLEDGEMENT

The presented work is based on SARS-CoV2 genome sequence data retrieved from the GISAID repository (www.gisaid.org). We also gratefully acknowledge the originating laboratories responsible for obtaining the specimens and the submitting laboratories where genetic sequence data were generated and shared via the GISAID Initiative (Supplementary Table 10). All submitters of data may be contacted directly via GISAID.

## Supplementary Table Legends

**Supplementary Table 1: Epitopes predicted from the SARS-CoV2 reference proteome**. 326 epitopes predicted by NetMHCpan and NetMHCIIpan from the SARS-CoV2 reference proteome along with corresponding HLA alleles involved in their recognition are presented in the table. Only those epitopes identified with prediction scores ≥ 0.95 are reported. For each epitope the number of alleles interacting with it, along with the best scoring allele and the corresponding prediction score are also mentioned.

**Supplementary Table 2: Observed richness of HLA alleles and epitopes**. Table denoting observed richness of MHC-I and MHC-II alleles among the 26 ethnic populations belonging to five super-populations, as well as the observed richness of CD8-specific and CD4-specific SARS-CoV2 epitopes recognized by the respective HLA alleles.

**Supplementary Table 3: Intersecting and unique sets of HLA alleles and SARS-CoV2 epitopes**. (A) Intersecting and unique sets between the MHC-I and MHC-II alleles in the five super-populations. (B) Intersecting and unique sets between CD8-specific and CD4-specific SARS-CoV2 epitopes recognized by the HLA alleles prevalent in the five super-populations. Super-population names are abbreviated as AFR-Africans; AMR-Ad mixed Americans; EAS-East Asians; EUR-Europeans; SAS-South east Asians.

**Supplementary Table 4: Individual HLA allelic makeup and corresponding SARS-CoV2 epitope recognition potential**. The table depicts individual HLA allelic makeup (HLA-A, HLA-B, HLA-C HLA-DQ and HLA-DR) of 2693 individuals representing 26 ethnic groups and five super-populations as retrieved from 1000 genome project data. The number of epitopes that can be recognized by each individual allele based on their allelic makeup are also indicated along with a breakup of the epitopes recognized by each of alleles. Super-population names are abbreviated as AFR-Africans; AMR-Ad mixed Americans; EAS-East Asians; EUR-Europeans; SAS-South east Asians. The abbreviations used for names of different populations/ ethnic groups are in accordance with those originally reported by the 1000 genomes project and the International Genome Sample Resource (IGSR) (https://www.internationalgenome.org/faq/which-populations-are-part-your-study/).

**Supplementary Table 5: Wilcoxon signed-rank test results depicting variations in the SARS-CoV2 epitope recognition potential of ethnic groups and super populations**. (A) Differences in the epitope recognition potential of a specific ethnic group w.r.t. the overall epitope recognition potential of all other ethnic groups taken together are depicted. Significant differences are highlighted (Wilcoxon signed rank test BH corrected p-value < 0.05). (**B)** Differences in the epitope recognition potential of a specific super-population w.r.t. the overall epitope recognition potential of all other super populations taken together are depicted. Significant differences are highlighted (Wilcoxon signed rank test BH corrected p-value < 0.05). The p-values obtained from Wilcoxon signed rank test were corrected for multiple testing using Benjamini-Hochberg (BH) method.

**Supplementary Table 6: Pair-wise comparison of epitope recognition potential between different ethnic groups and super populations**. (A) Differences in epitope recognition potentials between individuals representing any two specific ethnic groups are represented in terms of mean rank differences. (B) Differences in epitope recognition potentials between individuals representing any two specific super-populations are represented in terms of mean rank differences. Significant differences have been highlighted (Dunn’s test p-values < 0.05). Populations (or ethnic groups) exhibiting significant differences (with p-values < 0.05) have been highlighted. Super-population names are abbreviated as AFR-Africans; AMR-Ad mixed Americans; EAS-East Asians; EUR-Europeans; SAS-South east Asians, and prefixed to each of the population names in the table. The abbreviations used for names of different populations/ ethnic groups are in accordance with those originally reported by the 1000 genomes project and the International Genome Sample Resource (IGSR) (https://www.internationalgenome.org/faq/which-populations-are-part-your-study/).

**Supplementary Table 7: Distribution of SARS-CoV2 epitope variants across different geographies**. Table enlisting the SARS-CoV2 epitope variations along with their distribution frequencies in the genomes isolated and sequenced in different geographies. Epitope sequences along with their positions in the respective protein are also mentioned.

**Supplementary Table 8: Multi-epitope peptide vaccine candidates for SARS-CoV2**. Table enlisting (A) CD8 specific and (B) CD4 specific SARS-CoV2 epitopes that may be considered for designing multi-epitope peptide vaccines. The super population(s) where the epitope have been predicted to be well recognized as well as the VaxiJen scores for the respective epitopes are indicated. Epitopes (and their variants) which were found to be present in atleast 10 studied SARS-CoV2 genomes and were recognized by the HLA-system of at least 5% of individuals constituting the five super-populations are presented in the table. The presence of these peptides in SARS coronavirus and MERS coronavirus proteomes is also indicated.

**Supplementary Table 9: SARS and MERS protein sequences analyzed**. Proteins from SARS and MERS coronavirus that were analyzed to check the presence of epitopes similar to the identified vaccine candidates (epitopes) in SARS-COV2.

**Supplementary Table 10: Acknowledgement Table**. GISAID Accession IDs of the SARS-CoV2 genomes, along with the Originating laboratories responsible for obtaining the specimens and the Submitting laboratories where genetic sequence data were generated and shared via the GISAID Initiative, on which this research is based.

## Supplementary Figure Legends

**Supplementary Figure 1: Distribution of MHC-I alleles among the ethnic groups**. Heat-map depicting the distribution of MHC-I alleles among the 26 ethnic groups involved in this study. Intensity of the colour indicates frequency of a particular allele in an ethnic group. Both the ethnic groups and the MHC-I alleles have been hierarchically clustered. An additional colour-key along the vertical axis indicates the number of SARS-CoV2 epitopes recognized by the HLA allele. Along the horizontal axis ethnic groups have been tagged with different colours based on their affiliations to respective super-populations.

**Supplementary Figure 2: Distribution of MHC-II alleles among the ethnic groups**. Heat-map depicting the distribution of MHC-II alleles among the 26 ethnic groups involved in this study. Intensity of the colour indicates frequency of a particular allele in an ethnic group. Both the ethnic groups and the MHC-II alleles have been hierarchically clustered. An additional colour-key along the vertical axis indicates the number of SARS-CoV2 epitopes recognized by the HLA allele. Along the horizontal axis ethnic groups have been tagged with different colours based on their affiliations to respective super-populations.

**Supplementary Figure 3: Distribution of CD8-specific epitopes recognized by the HLA-system among the ethnic groups**. Heat-map depicting the distribution of CD8-specific epitopes that could be recognized by the HLA alleles prevalent among the 26 ethnic groups involved in this study. Both the ethnic groups and the CD8-specific epitopes have been hierarchically clustered. An additional colour-key along the vertical axis indicates the number of human HLA-types capable of recognizing the epitope. Along the horizontal axis ethnic groups have been tagged with different colours based on their affiliations to respective super-populations.

**Supplementary Figure 4: Distribution of CD4-specific epitopes recognized by the HLA-system among the ethnic groups**. Heat-map depicting the distribution of CD4-specific epitopes that could be recognized by the HLA alleles prevalent among the 26 ethnic groups involved in this study. Both the ethnic groups and the CD4-specific epitopes have been hierarchically clustered. An additional colour-key along the vertical axis indicates the number of human HLA-types capable of recognizing the epitope. Along the horizontal axis ethnic groups have been tagged with different colours based on their affiliations to respective super-populations.

**Supplementary Figure 5: Average count of SARS-CoV2 epitopes for each super-population**. Average count of the number of SARS-CoV2 epitopes identified by the samples comprising each of the super-populations: (A) total epitopes, (B) CD8-specific epitopes and (C) CD4-specific epitopes. 95% confidence interval of the computed means are indicated in the bar plot. Bars marked with asterisk (*) represents that mean counts of epitopes identified by the particular super population is significantly different from rest of the populations combined together.

**Supplementary Figure 6: Average count of SARS-CoV2 epitopes for each ethnic group**. Average count of the number of SARS-CoV2 epitopes identified by the samples comprising each of the ethnic groups: (A) total epitopes, (B) CD8-specific epitopes and (C) CD4-specific epitopes. 95% confidence interval of the computed means are indicated in the bar plot. Bars marked with asterisk (*) represents that mean counts of epitopes identified by the particular ethnic group is significantly different from rest of the ethnic groups combined together.

**Supplementary Figure 7: Evolution of SARS-CoV2 genomes and epitope recognition capabilities**. Representation of the mean SARS-CoV2 epitope identification capacity among different ethnic groups with respect to SARS-Cov2 genomes sequenced till 11^th^ June 2020, as obtained from the GISAID (https://www.gisaid.org/). Each data-point on the plots represents the average of the number of epitopes in a given SARS-CoV2 genome that could be identified by all the individuals in an ethnic population. Dark black lines, for each of the ethnic groups, indicate the mean value for the SARS-CoV2 genomes isolated, sequenced and deposited to GISAID (A) in December 2019, (B) in January 2020, (C) in February 2020, (D) in March 2020, (E) in April 2020 and (F) in May-June 2020.

**Supplementary Figure 8: Distribution of epitopes shortlisted as vaccine peptides across super-populations**. Euler representation of (a) CD8-specific and (b) CD4-specific SARS-CoV2 epitopes recognized by the HLA alleles in the five super-populations. Epitopes observed in at least 10 SARS-CoV2 genomes and those potentially recognized by at least 5% of the individuals representing a super-population have been considered. The CD8-specific and CD4-specific epitopes at the intersection of the five super-populations could serve as potential vaccine candidates.

