## Supplementary Figure for "Does immune recognition of SARS-CoV2 epitopes vary between different ethnic groups?"

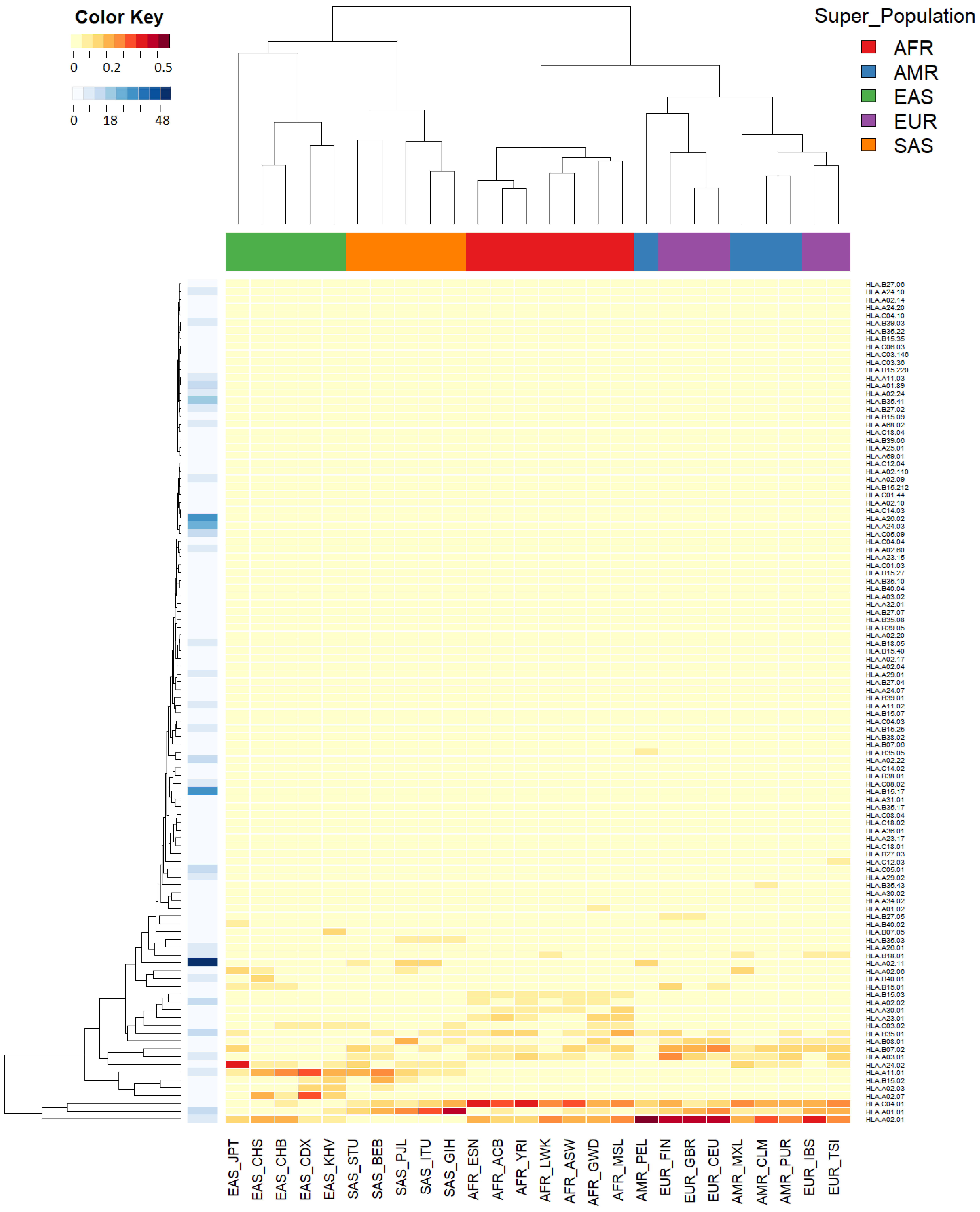


**Supplementary Figure 1: Distribution of MHC-I alleles among the ethnic groups.** Heat-map depicting the distribution of MHC-I alleles among the 26 ethnic groups involved in this study. Intensity of the colour indicates frequency of a particular allele in an ethnic group. Both the ethnic groups and the MHC-I alleles have been hierarchically clustered. An additional colour-key along the vertical axis indicates the number of SARS-CoV2 epitopes recognized by the HLA allele. Along the horizontal axis ethnic groups have been tagged with different colours based on their affiliations to respective super-populations.

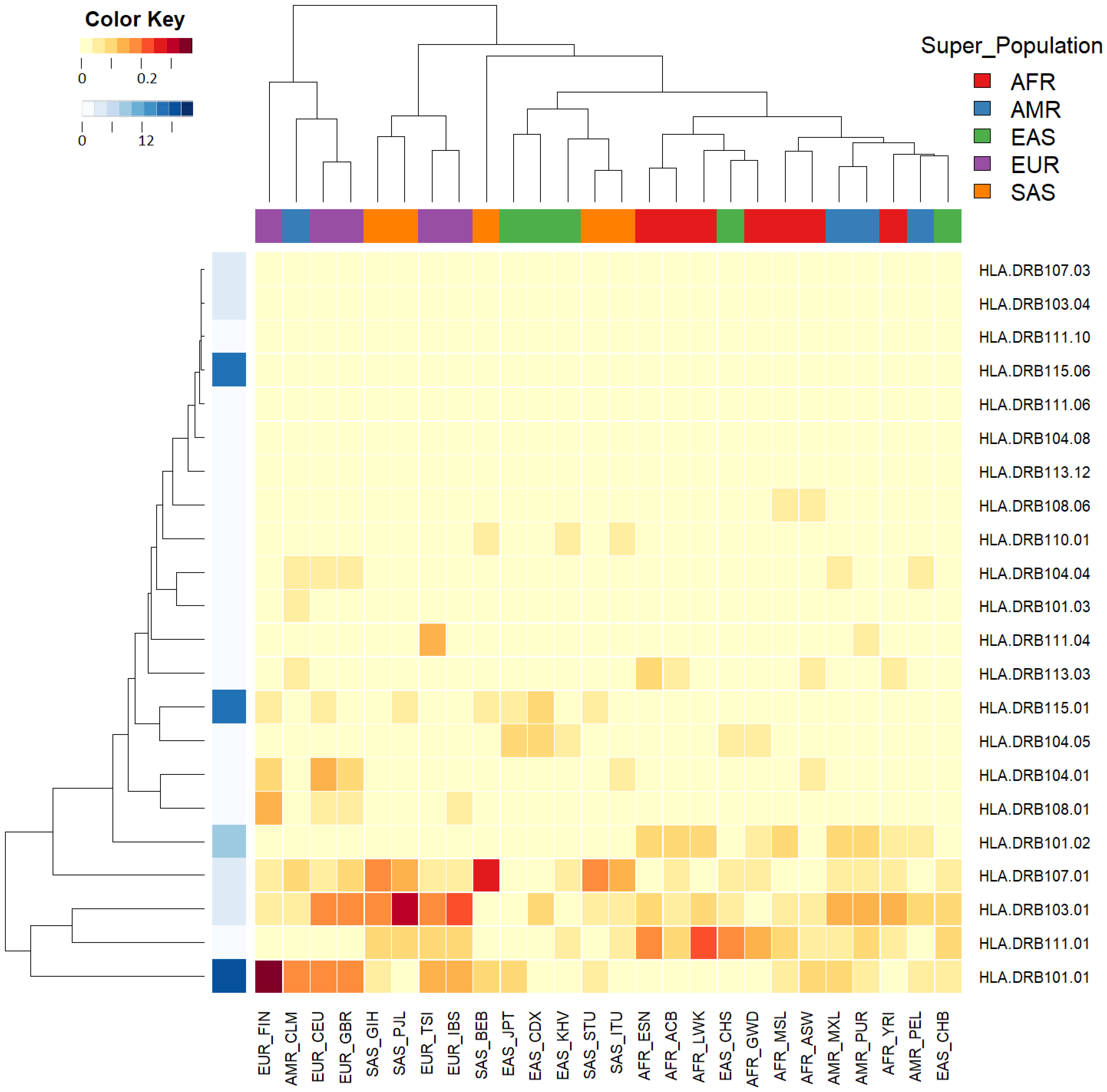


**Supplementary Figure 2: Distribution of MHC-II alleles among the ethnic groups.** Heat-map depicting the distribution of MHC-II alleles among the 26 ethnic groups involved in this study. Intensity of the colour indicates frequency of a particular allele in an ethnic group. Both the ethnic groups and the MHC-II alleles have been hierarchically clustered. An additional colour-key along the vertical axis indicates the number of SARS-CoV2 epitopes recognized by the HLA allele. Along the horizontal axis ethnic groups have been tagged with different colours based on their affiliations to respective super-populations.

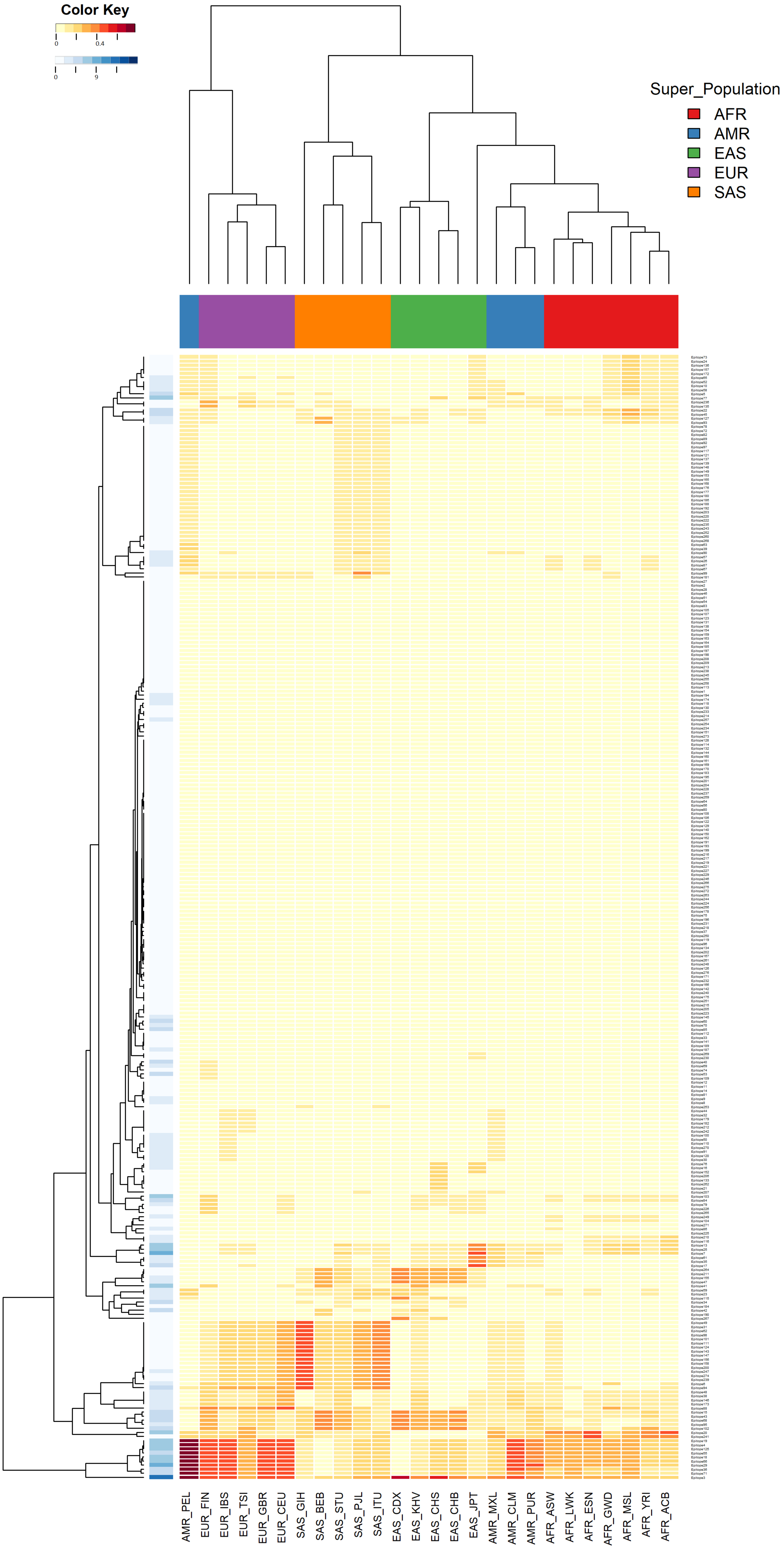


**Supplementary Figure 3: Distribution of CD8-specific epitopes recognized by the HLA-system among the ethnic groups.** Heat-map depicting the distribution of CD8-specific epitopes that could be recognized by the HLA alleles prevalent among the 26 ethnic groups involved in this study. Both the ethnic groups and the CD8-specific epitopes have been hierarchically clustered. An additional colour-key along the vertical axis indicates the number of human HLA-types capable of recognizing the epitope. Along the horizontal axis ethnic groups have been tagged with different colours based on their affiliations to respective super-populations.

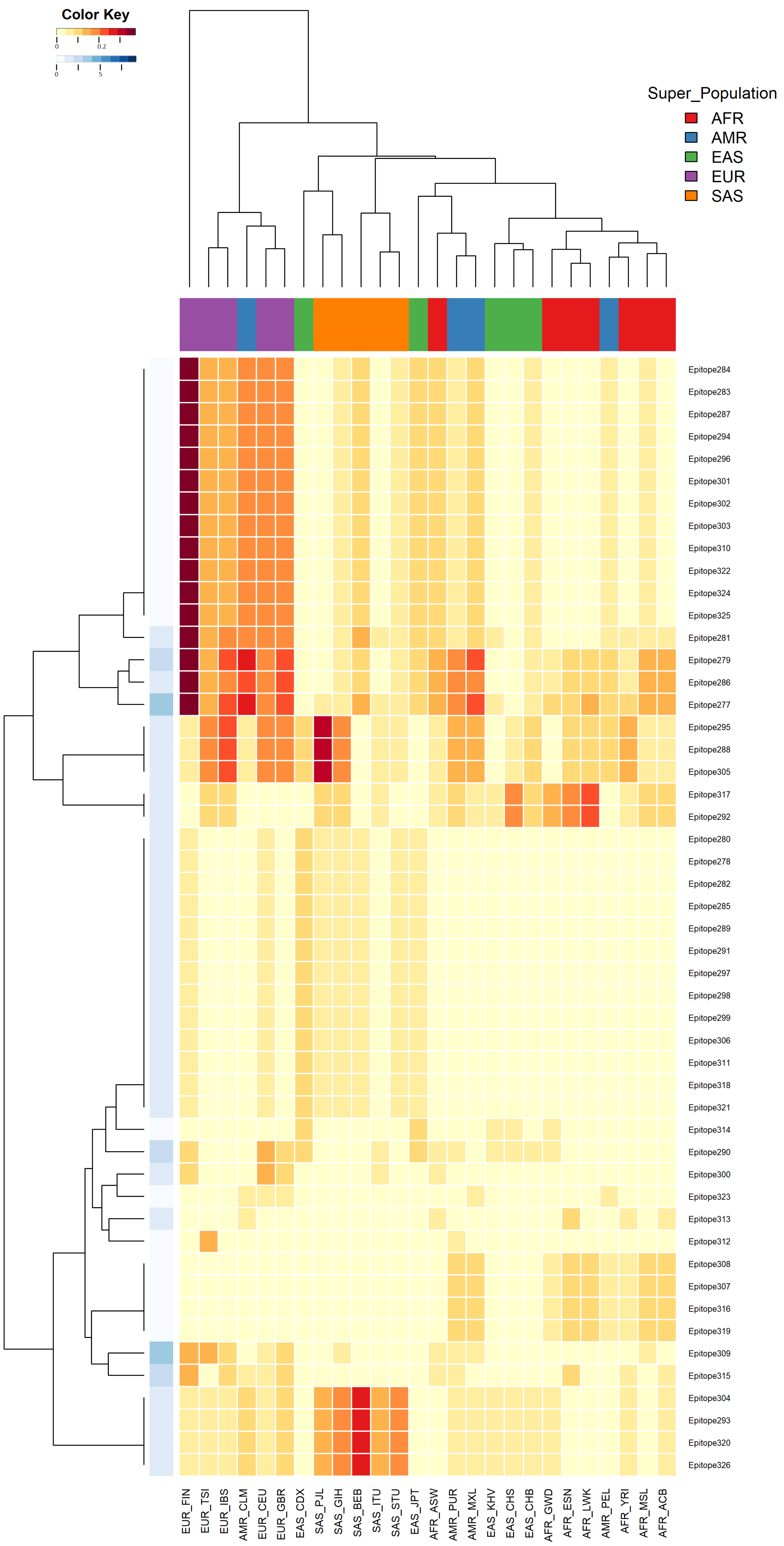


**Supplementary Figure 4: Distribution of CD4-specific epitopes recognized by the HLA-system among the ethnic groups.** Heat-map depicting the distribution of CD4-specific epitopes that could be recognized by the HLA alleles prevalent among the 26 ethnic groups involved in this study. Both the ethnic groups and the CD4-specific epitopes have been hierarchically clustered. An additional colour-key along the vertical axis indicates the number of human HLA-types capable of recognizing the epitope. Along the horizontal axis ethnic groups have been tagged with different colours based on their affiliations to respective super-populations

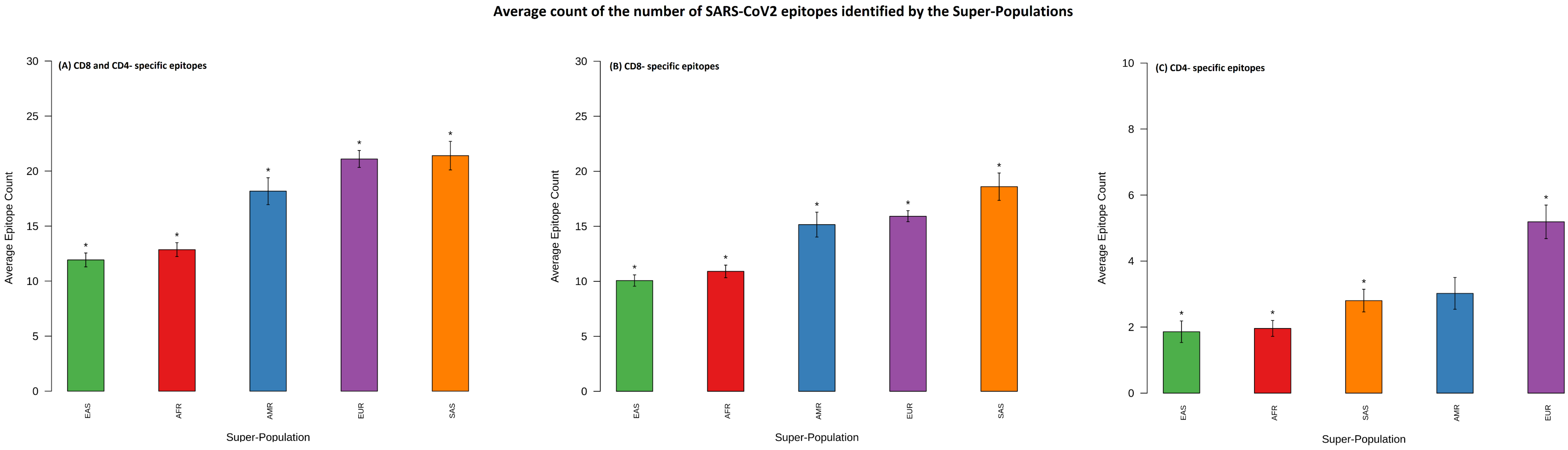


**Supplementary Figure 5: Average count of SARS-CoV2 epitopes for each super-population.** Average count of the number of SARS-CoV2 epitopes identified by the samples comprising each of the super-populations: (A) total epitopes, (B) CD8-specific epitopes and (C) CD4-specific epitopes. 95% confidence interval of the computed means are indicated in the bar plot. Bars marked with asterisk (*) represents that mean counts of epitopes identified by the particular super-population is significantly different from rest of the populations combined together.

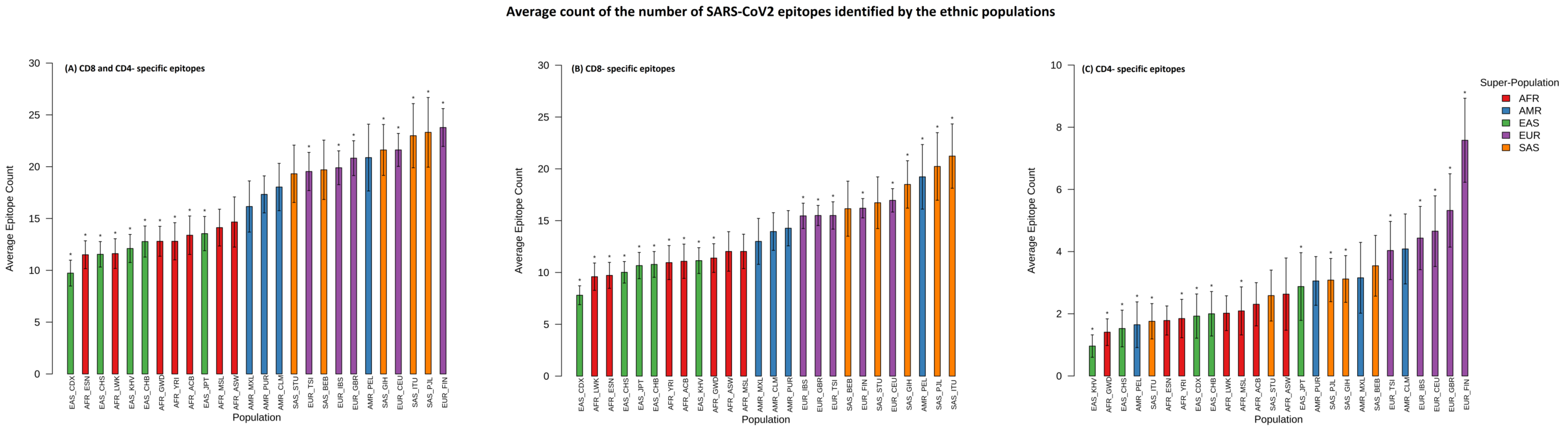


**Supplementary Figure 6: Average count of SARS-CoV2 epitopes for each ethnic group.** Average count of the number of SARS-CoV2 epitopes identified by the samples comprising each of the ethnic groups: (A) total epitopes, (B) CD8-specific epitopes and (C) CD4-specific epitopes. 95% confidence interval of the computed means are indicated in the bar plot. Bars marked with asterisk (*) represents that mean counts of epitopes identified by the particular ethnic group is significantly different from rest of the ethnic groups combined together.

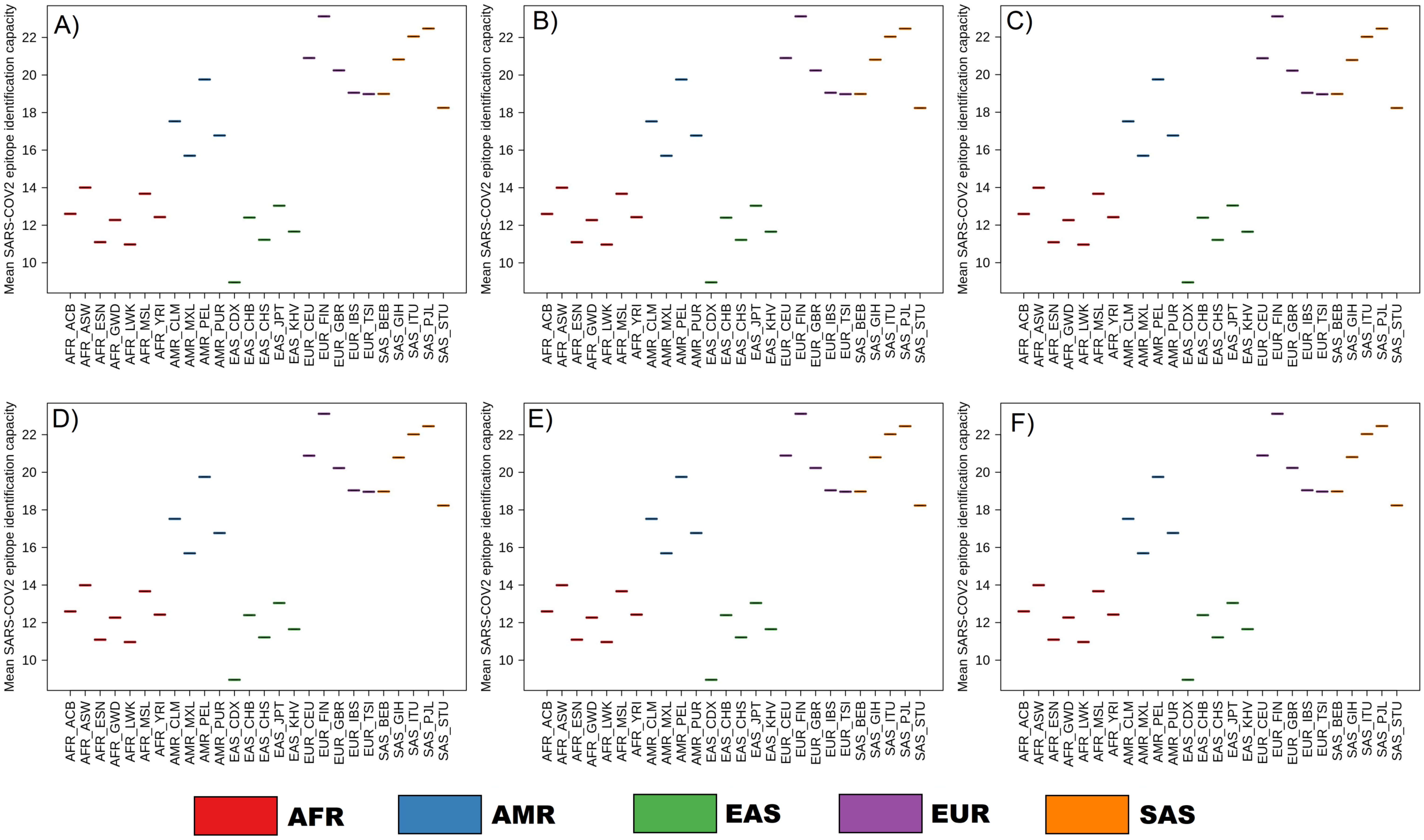


**Supplementary Figure 7: Evolution of SARS-CoV2 genomes and epitope recognition capabilities.** Representation of the mean SARS-CoV2 epitope identification capacity among different ethnic groups with respect to SARS-Cov2 genomes sequenced till 11$^{th}$ June 2020, as obtained from the GISAID (https://www.gisaid.org/). Each data-point on the plots represents the average of the number of epitopes in a given SARS-CoV2 genome that could be identified by all the individuals in an ethnic group. Dark black lines, for each of the ethnic groups, indicate the mean value for the SARS-CoV2 genomes isolated, sequenced and deposited to GISAID (A) in December 2019, (B) in January 2020, (C) in February 2020, (D) in March 2020, (E) in April 2020 and (F) in May-June 2020.

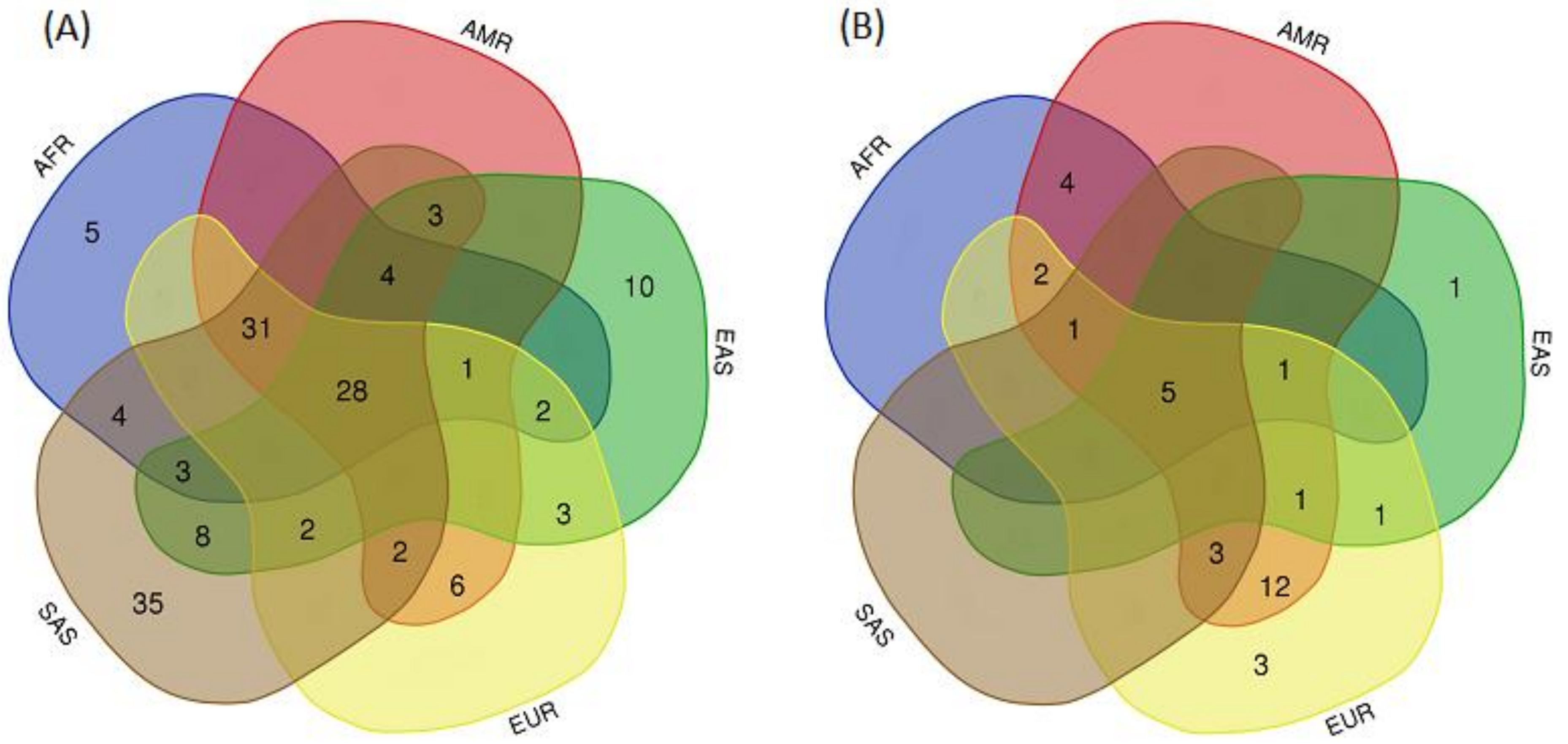


**Supplementary Figure 8: Distribution of epitopes shortlisted as vaccine peptides across super-populations.** Euler representation of (a) CD8-specific and (b) CD4-specific SARS-CoV2 epitopes recognized by the HLA alleles in the five super-populations. Epitopes observed in at least 10 SARS-CoV2 genomes and those potentially recognized by at least 5% of the individuals representing a super-population have been considered. The CD8-specific and CD4-specific epitopes at the intersection of the five super-populations could serve as potential vaccine candidates.
