## Supplementary File for "Does immune recognition of SARS-CoV2 epitopes vary between different ethnic groups?"

### Supplementary File 1

**Title:** Does immune recognition of SARS-CoV2 epitopes vary between different ethnic groups?

Tungadri Bose[$]*, Namrata Pant[$], Nishal Kumar Pinna, Subhrajit Bhar, Anirban Dutta*, Sharmila S. Mande*

TCS Research, Tata Consultancy Services Limited, Pune, India

[$]Equal contributors

*Corresponding authors

#### Identification of viral peptides as potential vaccine candidates

Since the temporal changes in the SARS-CoV2 genome did not seemed to appreciably change the epitope recognition ability of the host, we further investigated if a minimal set of antigenic peptides could be identified which may be utilized to apriori sensitize the human immune system, thereby providing resistance to Covid-19 infection at a global scale. Based on the 326 epitopes (and their variants) that were recognized (with a predictions scores ≥ 0.95) by at least 5% of the individuals in each of the ethnicities, euler plots (Supplementary Figure 8) for identifying the minimal set of SARS-CoV2 epitopes (both CD8 and CD4) were generated. A total of 28 CD8 and 5 CD4-specific epitopes were found to match our criteria (Table 3, Supplementary Table 8). Of these, 12 CD8 and 2 CD4-specific epitopes were also confirmed as potential vaccine subunits by the software tool Vaxijen (1) with a threshold ≥ 0.5. Further, several of the identified epitopes were found to map (with 100% identity and 100% overlap) with the SARS proteome, thereby indicating at the possibility of using these epitopes to target multiple lineage B *Betacoronavirus* strains (Table 3). However, the overlap of these epitopes to the MERS proteome was extremely limited. The list of SARS and MERS protein sequences used in this study have been listed in Supplementary Table 9. In addition to the above-mentioned predicted epitopes, 31 CD8-specific epitopes were also found, which can be explored further as potential generic vaccine candidates, except for East Asian (EAS) ethnicities (Supplementary Table 8). Among these EAS ethnicities 10 additional CD8-specific potential vaccine candidates were found. These were unique to EAS and could possibly provide adequate protection to individuals of this super-population. Similar patterns among the CD4-specific epitopes were also noted (Supplementary Table 8). Overall, results indicated that sensitizing the immune system with any of these (or a cocktail of these) predicted epitopes mentioned in Supplementary Table 8 could potentially provide protection to Covid-19 infection at a global scale.
