## Supplementary Table for "Does immune recognition of SARS-CoV2 epitopes vary between different ethnic groups?"

**Supplementary Table 10: Acknowledgement Table.** GISAID Accession IDs of the SARS-CoV2 genomes, along with the Originating laboratories responsible for obtaining the specimens and the Submitting laboratories where genetic sequence data were generated and shared via the GISAID Initiative, on which this research is based.

We gratefully acknowledge the following Originating laboratories responsible for obtaining the specimens and the Submitting laboratories where genetic sequence data were generated and shared via the GISAID Initiative, on which this research is based. All submitters of data may be contacted directly via www.gisaid.org

| EPI_Accession_ID | Originating Laboratory | Submitting Laboratory |
|---|---|---|
| EPI_ISL_402119 | National Institute for Viral Disease Control and Prevention, China CDC | National Institute for Viral Disease Control and Prevention, China CDC |
| EPI_ISL_402120 | National Institute for Viral Disease Control and Prevention, China CDC | National Institute for Viral Disease Control and Prevention, China CDC |
| EPI_ISL_402121 | National Institute for Viral Disease Control and Prevention, China CDC | National Institute for Viral Disease Control and Prevention, China CDC |
| EPI_ISL_402123 | Institute of Pathogen Biology, Chinese Academy of Medical Sciences & Peking Union Medical College | Institute of Pathogen Biology, Chinese Academy of Medical Sciences & Peking Union Medical College |
| EPI_ISL_402124 | Wuhan Jinyintan Hospital | Wuhan Institute of Virology, Chinese Academy of Sciences |
| EPI_ISL_402125 | unknown | National Institute for Communicable Disease Control and Prevention (ICDC) Chinese Center for Disease Control and Prevention (China CDC) |
| EPI_ISL_402127 | Wuhan Jinyintan Hospital | Wuhan Institute of Virology, Chinese Academy of Sciences |
| EPI_ISL_402128 | Wuhan Jinyintan Hospital | Wuhan Institute of Virology, Chinese Academy of Sciences |
| EPI_ISL_402129 | Wuhan Jinyintan Hospital | Wuhan Institute of Virology, Chinese Academy of Sciences |
| EPI_ISL_402130 | Wuhan Jinyintan Hospital | Wuhan Institute of Virology, Chinese Academy of Sciences |
| EPI_ISL_402132 | Wuhan Jinyintan Hospital | Hubei Provincial Center for Disease Control and Prevention |

| EPI_Accession_ID | Originating Laboratory | Submitting Laboratory |
|---|---|---|
| EPI_ISL_403929 | Institute of Pathogen Biology, Chinese Academy of Medical Sciences & Peking Union Medical College | Institute of Pathogen Biology, Chinese Academy of Medical Sciences & Peking Union Medical College |
| EPI_ISL_403930 | Institute of Pathogen Biology, Chinese Academy of Medical Sciences & Peking Union Medical College | Institute of Pathogen Biology, Chinese Academy of Medical Sciences & Peking Union Medical College |
| EPI_ISL_403931 | Institute of Pathogen Biology, Chinese Academy of Medical Sciences & Peking Union Medical College | Institute of Pathogen Biology, Chinese Academy of Medical Sciences & Peking Union Medical College |
| EPI_ISL_403932 | Guangdong Provincial Center for Diseases Control and Prevention; Guangdong Provincial Public Health | Department of Microbiology, Guangdong Provincial Center for Diseases Control and Prevention |
| EPI_ISL_403933 | Guangdong Provincial Center for Diseases Control and Prevention; Guangdong Provincial Public Health | Department of Microbiology, Guangdong Provincial Center for Diseases Control and Prevention |
| EPI_ISL_403934 | Guangdong Provincial Center for Diseases Control and Prevention; Guangdong Provincial Public Health | Department of Microbiology, Guangdong Provincial Center for Diseases Control and Prevention |
| EPI_ISL_403935 | Guangdong Provincial Center for Diseases Control and Prevention; Guangdong Provincial Public Health | Department of Microbiology, Guangdong Provincial Center for Diseases Control and Prevention |
| EPI_ISL_403936 | Guangdong Provincial Center for Diseases Control and Prevention; Guangdong Provincial Public Health | Department of Microbiology, Guangdong Provincial Center for Diseases Control and Prevention |
| EPI_ISL_403937 | Guangdong Provincial Center for Diseases Control and Prevention; Guangdong Provincial Public Health | Department of Microbiology, Guangdong Provincial Center for Diseases Control and Prevention |
| EPI_ISL_403962 | Bamrasnaradura Hospital | 1. Department of Medical Sciences, Ministry of Public Health, Thailand 2. Thai Red Cross Emerging Infectious Diseases - Health Science Centre 3. Department of Disease Control, Ministry of Public Health, Thailand |
| EPI_ISL_403963 | Bamrasnaradura Hospital | 1. Department of Medical Sciences, Ministry of Public Health, Thailand 2. Thai Red Cross Emerging Infectious Diseases - Health Science Centre 3. Department of Disease Control, Ministry of Public Health, Thailand |
| EPI_ISL_404227 | Zhejiang Provincial Center for Disease Control and Prevention | Department of Microbiology, Zhejiang Provincial Center for Disease Control and Prevention |
| EPI_ISL_404228 | Zhejiang Provincial Center for Disease Control and Prevention | Department of Microbiology, Zhejiang Provincial Center for Disease Control and Prevention |
| EPI_ISL_404253 | IL Department of Public Health Chicago Laboratory | Pathogen Discovery, Respiratory Viruses Branch, Division of Viral Diseases, Centers for Disease Control and Prevention |

| EPI_Accession_ID | Originating Laboratory | Submitting Laboratory |
|---|---|---|
| EPI_ISL_404895 | Providence Regional Medical Center | Division of Viral Diseases, Centers for Disease Control and Prevention |
| EPI_ISL_405839 | The University of Hong Kong - Shenzhen Hospital | Li Ka Shing Faculty of Medicine, The University of Hong Kong |
| EPI_ISL_406030 | The University of Hong Kong - Shenzhen Hospital | Li Ka Shing Faculty of Medicine, The University of Hong Kong |
| EPI_ISL_406031 | Centers for Disease Control, R.O.C. (Taiwan) | Centers for Disease Control, R.O.C. (Taiwan) |
| EPI_ISL_406034 | California Department of Public Health | Pathogen Discovery, Respiratory Viruses Branch, Division of Viral Diseases, Centers for Disease Control and Prevention |
| EPI_ISL_406036 | California Department of Public Health | Pathogen Discovery, Respiratory Viruses Branch, Division of Viral Diseases, Centers for Disease Control and Prevention |
| EPI_ISL_406223 | Arizona Department of Health Services | Pathogen Discovery, Respiratory Viruses Branch, Division of Viral Diseases, Centers for Disease Control and Prevention |
| EPI_ISL_406531 | Guangdong Provincial Center for Diseases Control and Prevention; Guangdong Provinical Public Health | Guangdong Provincial Center for Disease Control and Prevention |
| EPI_ISL_406533 | Guangdong Provincial Center for Diseases Control and Prevention; Guangdong Provinical Public Health | Guangdong Provincial Center for Diseases Control and Prevention |
| EPI_ISL_406534 | Guangdong Provincial Center for Diseases Control and Prevention; Guangdong Provincial Public Health | Guangdong Provincial Center for Diseases Control and Prevention |
| EPI_ISL_406535 | Guangdong Provincial Center for Diseases Control and Prevention; Guangdong Provincial Public Health | Guangdong Provincial Center for Diseases Control and Prevention |
| EPI_ISL_406536 | Guangdong Provincial Center for Diseases Control and Prevention; Guangdong Provincial Public Health | Guangdong Provincial Center for Diseases Control and Prevention |
| EPI_ISL_406538 | Guangdong Provincial Center for Diseases Control and Prevention;Guangdong Provincial Institute of Public Health | Guangdong Provincial Center for Diseases Control and Prevention |
| EPI_ISL_406593 | Shenzhen Key Laboratory of Pathogen and Immunity, National Clinical Research Center for Infectious Disease, Shenzhen Third People's Hospital | Shenzhen Key Laboratory of Pathogen and Immunity, National Clinical Research Center for Infectious Disease, Shenzhen Third People's Hospital |

| EPI_Accession_ID | Originating Laboratory | Submitting Laboratory |
|---|---|---|
| EPI_ISL_406594 | Shenzhen Key Laboratory of Pathogen and Immunity, National Clinical Research Center for Infectious Disease, Shenzhen Third People's Hospital | Shenzhen Key Laboratory of Pathogen and Immunity, National Clinical Research Center for Infectious Disease, Shenzhen Third People's Hospital |
| EPI_ISL_406596 | Department of Infectious and Tropical Diseases, Bichat Claude Bernard Hospital, Paris | National Reference Center for Viruses of Respiratory Infections, Institut Pasteur, Paris |
| EPI_ISL_406597 | Department of Infectious and Tropical Diseases, Bichat Claude Bernard Hospital, Paris | National Reference Center for Viruses of Respiratory Infections, Institut Pasteur, Paris |
| EPI_ISL_406716 | unknown | State Key Laboratory of Virology, Wuhan University |
| EPI_ISL_406717 | unknown | State Key Laboratory of Virology, Wuhan University |
| EPI_ISL_406798 | General Hospital of Central Theater Command of People's Liberation Army of China | BGI & Institute of Microbiology, Chinese Academy of Sciences & Shandong First Medical University & Shandong Academy of Medical Sciences & General Hospital of Central Theater Command of People's Liberation Army of China |
| EPI_ISL_406800 | General Hospital of Central Theater Command of People's Liberation Army of China | BGI & Institute of Microbiology, Chinese Academy of Sciences & Shandong First Medical University & Shandong Academy of Medical Sciences & General Hospital of Central Theater Command of People's Liberation Army of China |
| EPI_ISL_406801 | General Hospital of Central Theater Command of People's Liberation Army of China | BGI & Institute of Microbiology, Chinese Academy of Sciences & Shandong First Medical University & Shandong Academy of Medical Sciences & General Hospital of Central Theater Command of People's Liberation Army of China |
| EPI_ISL_406844 | Monash Medical Centre | Collaboration between the University of Melbourne at The Peter Doherty Institute for Infection and Immunity, and the Victorian Infectious Disease Reference Laboratory |
| EPI_ISL_406862 | CharitÃ© UniversitÃ¤tsmedizin Berlin, Institute of Virology; Institut fÃ¼r Mikrobiologie der Bundeswehr, Munich | CharitÃ© UniversitÃ¤tsmedizin Berlin, Institute of Virology |
| EPI_ISL_406970 | Hangzhou Center for Disease and Control Microbiology Lab | Hangzhou Center for Disease and Control Microbiology Lab |

| EPI_Accession_ID | Originating Laboratory | Submitting Laboratory |
|---|---|---|
| EPI_ISL_406973 | Singapore General Hospital | National Public Health Laboratory |
| EPI_ISL_407071 | Respiratory Virus Unit, Microbiology Services Colindale, Public Health England | Respiratory Virus Unit, Microbiology Services Colindale, Public Health England |
| EPI_ISL_407073 | Respiratory Virus Unit, Microbiology Services Colindale, Public Health England | Respiratory Virus Unit, Microbiology Services Colindale, Public Health England |
| EPI_ISL_407079 | Lapland Central Hospital | Department of Virology, University of Helsinki and Helsinki University Hospital, Helsinki, Finland |
| EPI_ISL_407084 | Department of Virology III, National Institute of Infectious Diseases | Pathogen Genomics Center, National Institute of Infectious Diseases |
| EPI_ISL_407193 | Korea Centers for Disease Control & Prevention (KCDC) Center for Laboratory Control of Infectious Diseases Division of Viral Diseases | Korea Centers for Disease Control & Prevention (KCDC) Center for Laboratory Control of Infectious Diseases Division of Viral Diseases |
| EPI_ISL_407313 | Hangzhou Center for Disease Control and Prevention | Hangzhou Center for Disease Control and Prevention |
| EPI_ISL_407893 | Centre for Infectious Diseases and Microbiology Laboratory Services | NSW Health Pathology - Institute of Clinical Pathology and Medical Research; Westmead Hospital; University of Sydney |
| EPI_ISL_407894 | Pathology Queensland | Public Health Virology Laboratory |
| EPI_ISL_407896 | Pathology Queensland | Public Health Virology Laboratory |
| EPI_ISL_407976 | KU Leuven, Clinical and Epidemiological Virology | KU Leuven, Clinical and Epidemiological Virology |
| EPI_ISL_407987 | Singapore General Hospital | Programme in Emerging Infectious Diseases, Duke-NUS Medical School |
| EPI_ISL_407988 | National Centre for Infectious Diseases | Programme in Emerging Infectious Diseases, Duke-NUS Medical School |
| EPI_ISL_408008 | California Department of Health | Pathogen Discovery, Respiratory Viruses Branch, Division of Viral Diseases, Centers for Disease Control and Prevention |
| EPI_ISL_408009 | California Department of Health | Pathogen Discovery, Respiratory Viruses Branch, Division of Viral Diseases, Centers for Disease Control and Prevention |
| EPI_ISL_408010 | California Department of Health | Pathogen Discovery, Respiratory Viruses Branch, Division of Viral Diseases, Centers for Disease Control and Prevention |

| EPI_Accession_ID | Originating Laboratory | Submitting Laboratory |
|---|---|---|
| EPI_ISL_408430 | Department of Infectious and Tropical Diseases, Bichat Claude Bernard Hospital, Paris | National Reference Center for Viruses of Respiratory Infections, Institut Pasteur, Paris |
| EPI_ISL_408431 | Sorbonne UniversitÃ©, Inserm et Assistance Publique-HÃ´pitaux de Paris (PitiÃ© SalpÃ©triÃ¨re) | National Reference Center for Viruses of Respiratory Infections, Institut Pasteur, Paris |
| EPI_ISL_408478 | Yongchuan District Center for Disease Control and Prevention | Chongqing Municipal Center for Disease Control and Prevention |
| EPI_ISL_408479 | Zhongxian Center for Disease Control and Prevention | Chongqing Municipal Center for Disease Control and Prevention |
| EPI_ISL_408480 | National Institute for Viral Disease Control and Prevention, China CDC | National Institute for Viral Disease Control & Prevention, CCDC |
| EPI_ISL_408481 | National Institute for Viral Disease Control and Prevention, China CDC | National Institute for Viral Disease Control & Prevention, CCDC |
| EPI_ISL_408482 | National Institute for Viral Disease Control and Prevention, China CDC | National Institute for Viral Disease Control & Prevention, CCDC |
| EPI_ISL_408484 | National Institute for Viral Disease Control and Prevention, China CDC | National Institute for Viral Disease Control & Prevention, CCDC |
| EPI_ISL_408486 | National Institute for Viral Disease Control and Prevention, China CDC | National Institute for Viral Disease Control & Prevention, CCDC |
| EPI_ISL_408488 | National Institute for Viral Disease Control and Prevention, China CDC | National Institute for Viral Disease Control & Prevention, CCDC |
| EPI_ISL_408489 | Department of Laboratory Medicine, National Taiwan University Hospital | Microbial Genomics Core Lab, National Taiwan University Centers of Genomic and Precision Medicine |
| EPI_ISL_408511 | Institute of Viral Disease Control and Prevention, China CDC | Institute of Viral Disease Control and Prevention, China CDC |
| EPI_ISL_408514 | Institute of Viral Disease Control and Prevention, China CDC | Institute of Viral Disease Control and Prevention, China CDC |
| EPI_ISL_408515 | Institute of Viral Disease Control and Prevention, China CDC | Institute of Viral Disease Control and Prevention, China CDC |
| EPI_ISL_408665 | Dept. of Virology III, National Institute of Infectious Diseases | Pathogen Genomics Center, National Institute of Infectious Diseases |
| EPI_ISL_408666 | Dept. of Virology III, National Institute of Infectious Diseases | Pathogen Genomics Center, National Institute of Infectious Diseases |

| EPI_Accession_ID | Originating Laboratory | Submitting Laboratory |
|---|---|---|
| EPI_ISL_408667 | Dept. of Virology III, National Institute of Infectious Diseases | Pathogen Genomics Center, National Institute of Infectious Diseases |
| EPI_ISL_408668 | National Influenza Center - National Institute of Hygiene and Epidemiology (NIHE) | National Influenza Center - National Institute of Hygiene and Epidemiology (NIHE) |
| EPI_ISL_408669 | Dept. of Virology III, National Institute of Infectious Diseases | Pathogen Genomics Center, National Institute of Infectious Diseases |
| EPI_ISL_408670 | Wisconsin Department of Health Services | Pathogen Discovery, Respiratory Viruses Branch, Division of Viral Diseases, Centers for Disease Control and Prevention |
| EPI_ISL_408976 | Centre for Infectious Diseases and Microbiology Laboratory Services | NSW Health Pathology - Institute of Clinical Pathology and Medical Research; Westmead Hospital; University of Sydney |
| EPI_ISL_408977 | Serology, Virology and OTDS Laboratories (SAViD), NSW Health Pathology Randwick | NSW Health Pathology - Institute of Clinical Pathology and Medical Research; Centre for Infectious Diseases and Microbiology Laboratory Services; Westmead Hospital; University of Sydney |
| EPI_ISL_408978 | Wuhan Fourth Hospital | Beijing Genomics Institute (BGI) |
| EPI_ISL_409067 | Massachusetts Department of Public Health | Pathogen Discovery, Respiratory Viruses Branch, Division of Viral Diseases, Centers for Disease Control and Prevention |
| EPI_ISL_410044 | California Department of Public Health | Pathogen Discovery, Respiratory Viruses Branch, Division of Viral Diseases, Centers for Disease Control and Prevention |
| EPI_ISL_410045 | IL Department of Public Health Chicago Laboratory | Pathogen Discovery, Respiratory Viruses Branch, Division of Viral Diseases, Centers for Disease Control and Prevention |
| EPI_ISL_410218 | Department of Laboratory Medicine, National Taiwan University Hospital | Microbial Genomics Core Lab, National Taiwan University Centers of Genomic and Precision Medicine |
| EPI_ISL_410301 | National Influenza Centre, National Public Health Laboratory, Kathmandu, Nepal | The University of Hong Kong |
| EPI_ISL_410486 | CNR Virus des Infections Respiratoires - France SUD | CNR Virus des Infections Respiratoires - France SUD |
| EPI_ISL_410531 | Dept. of Pathology, National Institute of Infectious Diseases | Pathogen Genomics Center, National Institute of Infectious Diseases |

| EPI_Accession_ID | Originating Laboratory | Submitting Laboratory |
|---|---|---|
| EPI_ISL_410532 | Dept. of Pathology, National Institute of Infectious Diseases | Pathogen Genomics Center, National Institute of Infectious Diseases |
| EPI_ISL_410535 | National Centre for Infectious Diseases | Programme in Emerging Infectious Diseases, Duke-NUS Medical School |
| EPI_ISL_410536 | Singapore General Hospital, Molecular Laboratory, Division of Pathology | Programme in Emerging Infectious Diseases, Duke-NUS Medical School |
| EPI_ISL_410537 | Singapore General Hospital, Molecular Laboratory, Division of Pathology | Programme in Emerging Infectious Diseases, Duke-NUS Medical School |
| EPI_ISL_410546 | INMI Lazzaro Spallanzani IRCCS | Laboratory of Virology, INMI Lazzaro Spallanzani IRCCS |
| EPI_ISL_410713 | National Public Health Laboratory, National Centre for Infectious Diseases | National Public Health Laboratory, National Centre for Infectious Diseases |
| EPI_ISL_410714 | National Public Health Laboratory, National Centre for Infectious Diseases | National Public Health Laboratory, National Centre for Infectious Diseases |
| EPI_ISL_410715 | National Public Health Laboratory, National Centre for Infectious Diseases | National Public Health Laboratory, National Centre for Infectious Diseases |
| EPI_ISL_410716 | National Public Health Laboratory, National Centre for Infectious Diseases | National Centre for Infectious Diseases, National Centre for Infectious Diseases |
| EPI_ISL_410717 | Pathology Queensland | Public Health Virology Laboratory |
| EPI_ISL_410718 | Pathology Queensland | Public Health Virology Laboratory |
| EPI_ISL_410719 | National Public Health Laboratory | National Public Health Laboratory |
| EPI_ISL_411060 | Fujian Center for Disease Control and Prevention | Fujian Center for Disease Control and Prevention |
| EPI_ISL_411066 | Fujian Center for Disease Control and Prevention | Fujian Center for Disease Control and Prevention |
| EPI_ISL_411219 | Department of Infectious and Tropical Diseases, Bichat Claude Bernard Hospital, Paris | Laboratoire Virpath, CIRI U111, UCBL1, INSERM, CNRS, ENS Lyon |
| EPI_ISL_411902 | Virology Unit, Institut Pasteur du Cambodge. | Virology Unit, Institut Pasteur du Cambodge (Sequencing done by: Jessica E Manning/Jennifer A Bohl at Malaria and Vector Research Research Laboratory, National Institute of Allergy and Infectious Diseases and Vida Ahyong from Chan-Zuckerberg Biohub) |
| EPI_ISL_411915 | Laboratory Medicine | Department of Laboratory Medicine, Lin-Kou Chang Gung Memorial Hospital, Taoyuan, Taiwan. |
| EPI_ISL_411926 | Taiwan Centers for Disease Control | Taiwan Centers for Disease Control |

| EPI_Accession_ID | Originating Laboratory | Submitting Laboratory |
|---|---|---|
| EPI_ISL_411927 | Taiwan Centers for Disease Control | Taiwan Centers for Disease Control |
| EPI_ISL_411950 | NHC Key laboratory of Enteric Pathogenic Microbiology, Institute of Pathogenic Microbiology | Jiangsu Provincial Center for Disease Control & Prevention |
| EPI_ISL_411951 | unknown | Unit for Laboratory Development and Technology Transfer, Public Health Agency of Sweden |
| EPI_ISL_411952 | NHC Key laboratory of Enteric Pathogenic Microbiology, Institute of Pathogenic Microbiology | Jiangsu Provincial Center for Disease Control & Prevention |
| EPI_ISL_411953 | NHC Key laboratory of Enteric Pathogenic Microbiology, Institute of Pathogenic Microbiology | Jiangsu Provincial Center for Disease Control & Prevention |
| EPI_ISL_411954 | California Department of Public Health | Pathogen Discovery, Respiratory Viruses Branch, Division of Viral Diseases, Centers for Disease Control and Prevention |
| EPI_ISL_411955 | California Department of Public Health | Pathogen Discovery, Respiratory Viruses Branch, Division of Viral Diseases, Centers for Disease Control and Prevention |
| EPI_ISL_411956 | Texas Department of State Health Services | Pathogen Discovery, Respiratory Viruses Branch, Division of Viral Diseases, Centers for Disease Control and Prevention |
| EPI_ISL_411957 | unknown | Key Laboratory of Human Diseases, Comparative Medicine, Institute of Laboratory Animal Science |
| EPI_ISL_412028 | Hong Kong Department of Health | School of Public Health, The University of Hon g Kong |
| EPI_ISL_412029 | Hong Kong Department of Health | The University of Hong Kong |
| EPI_ISL_412030 | Hong Kong Department of Health | School of Public Health, The University of Hon g Kong |
| EPI_ISL_412116 | Respiratory Virus Unit, Microbiology Services Colindale, Public Health England | Respiratory Virus Unit, Microbiology Services Colindale, Public Health England |
| EPI_ISL_412862 | California Department of Public Health | Pathogen Discovery, Respiratory Viruses Branch, Division of Viral Diseases, Centers for Disease Control and Prevention |
| EPI_ISL_412869 | Division of Viral Diseases, Center for Laboratory Control of Infectious Diseases, Korea Centers for Diseases Control and Prevention | Division of Viral Diseases, Center for Laboratory Control of Infectious Diseases, Korea Centers for Diseases Control and Prevention |

| EPI_Accession_ID | Originating Laboratory | Submitting Laboratory |
|---|---|---|
| EPI_ISL_412870 | Division of Viral Diseases, Center for Laboratory Control of Infectious Diseases, Korea Centers for Diseases Control and Prevention | Division of Viral Diseases, Center for Laboratory Control of Infectious Diseases, Korea Centers for Diseases Control and Prevention |
| EPI_ISL_412871 | Division of Viral Diseases, Center for Laboratory Control of Infectious Diseases, Korea Centers for Diseases Control and Prevention | Division of Viral Diseases, Center for Laboratory Control of Infectious Diseases, Korea Centers for Diseases Control and Prevention |
| EPI_ISL_412872 | Division of Viral Diseases, Center for Laboratory Control of Infectious Diseases, Korea Centers for Diseases Control and Prevention | Division of Viral Diseases, Center for Laboratory Control of Infectious Diseases, Korea Centers for Diseases Control and Prevention |
| EPI_ISL_412873 | Division of Viral Diseases, Center for Laboratory Control of Infectious Diseases, Korea Centers for Diseases Control and Prevention | Division of Viral Diseases, Center for Laboratory Control of Infectious Diseases, Korea Centers for Diseases Control and Prevention |
| EPI_ISL_412898 | Wuhan Jinyintan Hospital | Hubei Provincial Center for Disease Control and Prevention |
| EPI_ISL_412899 | Wuhan Jinyintan Hospital | Hubei Provincial Center for Disease Control and Prevention |
| EPI_ISL_412912 | State Health Office Baden-Wuerttemberg | CharitÃ© UniversitÃ¤tsmedizin Berlin, Institute of Virology |
| EPI_ISL_412964 | Hospital Israelita Albert Einstein | Instituto Adolfo Lutz Interdisciplinary Procedures Center Strategic Laboratory |
| EPI_ISL_412965 | BCCDC Public Health Laboratory | BCCDC Public Health Laboratory |
| EPI_ISL_412966 | unknown | Technology Centre, Guangzhou Customs |
| EPI_ISL_412967 | unknown | Technology Centre, Guangzhou Customs |
| EPI_ISL_412968 | unknown | Takayuki Hishiki Kanagawa Prefectural Institute of Public Health, Department of Microbiology |
| EPI_ISL_412969 | unknown | Takayuki Hishiki Kanagawa Prefectural Institute of Public Health, Department of Microbiology |
| EPI_ISL_412970 | Washington State Department of Health | Seattle Flu Study |
| EPI_ISL_412971 | HUS Diagnostiikkakeskus, Hallinto | Department of Virology Faculty of Medicine, Medicum University of Helsinki |
| EPI_ISL_412972 | Instituto Nacional de Enfermedades Respiratorias | Instituto de Diagnostico y Referencia Epidemiologicos (INDRE) |

| EPI_Accession_ID | Originating Laboratory | Submitting Laboratory |
|---|---|---|
| EPI_ISL_412973 | Department of Infectious Diseases, Istituto Superiore di SanitÃ , Roma , Italy | Virology Laboratory, Scientific Department, Army Medical Center |
| EPI_ISL_412974 | Department of Infectious Diseases, Istituto Superiore di SanitÃ , Rome, Italy | Virology Laboratory, Scientific Department, Army Medical Center |
| EPI_ISL_412975 | Centre for Infectious Diseases and Microbiology Laboratory Services | NSW Health Pathology - Institute of Clinical Pathology and Medical Research; Westmead Hospital; University of Sydney |
| EPI_ISL_412978 | The Central Hospital Of Wuhan | Hubei Provincial Center for Disease Control and Prevention |
| EPI_ISL_412979 | Union Hospital of Tongji Medical College, Huazhong University of Science and Technology | Hubei Provincial Center for Disease Control and Prevention |
| EPI_ISL_412980 | Union Hospital of Tongji Medical College, Huazhong University of Science and Technology | Hubei Provincial Center for Disease Control and Prevention |
| EPI_ISL_412981 | CR&WISCO GENERAL HOSPITAL | Hubei Provincial Center for Disease Control and Prevention |
| EPI_ISL_412982 | Wuhan Lung Hospital | Hubei Provincial Center for Disease Control and Prevention |
| EPI_ISL_412983 | Tianmen Center for Disease Control and Prevention | Hubei Provincial Center for Disease Control and Prevention |
| EPI_ISL_413014 | Public Health Ontario Laboratory | Ontario Agency for Health Protection and Promotion (OAHPP) |
| EPI_ISL_413015 | Public Health Ontario Laboratory | National Microbiology Laboratory |
| EPI_ISL_413016 | Hospital Israelita Albert Einstein | Instituto Adolfo Lutz, Interdisciplinary Procedures Center, Strategic Laboratory |
| EPI_ISL_413017 | Department of Microbiology, Institute for Viral Diseases, College of Medicine, Korea University | Department of Microbiology, Institute for Viral Diseases, College of Medicine, Korea University |
| EPI_ISL_413018 | Department of Microbiology, Institute for Viral Diseases, College of Medicine, Korea University | Department of Microbiology, Institute for Viral Diseases, College of Medicine, Korea University |
| EPI_ISL_413019 | Department of Internal Medicine, Triemli Hospital | Institute of Medical Virology, University of Zurich |
| EPI_ISL_413020 | Department of Internal Medicine, Triemli Hospital | Institute of Medical Virology, University of Zurich |
| EPI_ISL_413021 | Klinik Hirslanden Zurich | Institute of Medical Virology, University of Zurich |
| EPI_ISL_413022 | Division of Infectious Diseases, University Hospital Zurich | Institute of Medical Virology, University of Zurich |

| EPI_Accession_ID | Originating Laboratory | Submitting Laboratory |
|---|---|---|
| EPI_ISL_413023 | Division of Infectious Diseases, University Hospital Zurich | Institute of Medical Virology, University of Zurich |
| EPI_ISL_413024 | Division of Infectious Diseases, University Hospital Zurich | Institute of Medical Virology, University of Zurich |
| EPI_ISL_413025 | Harborview Medical Center | UW Virology Lab |
| EPI_ISL_413213 | Centre for Infectious Diseases and Microbiology Laboratory Services | NSW Health Pathology - Institute of Clinical Pathology and Medical Research; Westmead Hospital; University of Sydney |
| EPI_ISL_413214 | Centre for Infectious Diseases and Microbiology Laboratory Services | NSW Health Pathology - Institute of Clinical Pathology and Medical Research; Westmead Hospital; University of Sydney |
| EPI_ISL_413221 | West of Scotland Specialist Virology Centre, NHSGGC | MRC-University of Glasgow Centre for Virus Research |
| EPI_ISL_413455 | Washington State Public Health Lab | University of Washington Virology Lab |
| EPI_ISL_413456 | Seattle Flu Study | Seattle Flu Study |
| EPI_ISL_413457 | Washington State Public Health Lab | UW Virology Lab |
| EPI_ISL_413458 | Washington State Public Health Lab | UW Virology Lab |
| EPI_ISL_413459 | Department of Pathology, Toshima Hospital | Pathogen Genomics Center, National Institute of Infectious Diseases |
| EPI_ISL_413485 | Department of microbiology laboratory,Anhui Provincial Center for Disease Control and Prevention | Department of microbiology laboratory,Anhui Provincial Center for Disease Control and Prevention |
| EPI_ISL_413486 | Valley Medical Center | University of Washington Virology Lab |
| EPI_ISL_413487 | Harborview Medical Center | University of Washington Virology Lab |
| EPI_ISL_413488 | Center of Medical Microbiology, Virology, and Hospital Hygiene, University of Duesseldorf | Center of Medical Microbiology, Virology, and Hospital Hygiene, University of Duesseldorf |
| EPI_ISL_413489 | Laboratorio di Microbiologia e Virologia, UniversitÃ Vita-Salute San Raffaele, Milano | Laboratorio di Microbiologia e Virologia, UniversitÃ Vita-Salute San Raffaele, Milano |
| EPI_ISL_413490 | Auckland Hospital | Institute of Environmental Science and Research (ESR) |
| EPI_ISL_413513 | Division of Infectious Diseases, Department of Internal Medicine, Korea University College of Medicine | Department of Microbiology, Institute for Viral Diseases, College of Medicine, Korea University |
| EPI_ISL_413514 | Department of Microbiology, Institute for Viral Diseases, College of Medicine, Korea University | Department of Microbiology, Institute for Viral Diseases, College of Medicine, Korea University |
| EPI_ISL_413515 | Division of Infectious Diseases, Department of Internal Medicine, Korea University College of Medicine | Department of Microbiology, Institute for Viral Diseases, College of Medicine, Korea University |

| EPI_Accession_ID | Originating Laboratory | Submitting Laboratory |
|---|---|---|
| EPI_ISL_413516 | Department of Microbiology, Institute for Viral Diseases, College of Medicine, Korea University | Department of Microbiology, Institute for Viral Diseases, College of Medicine, Korea University |
| EPI_ISL_413518 | unknown | Infectious Disease Control Center |
| EPI_ISL_413519 | unknown | Infectious Disease Control Center |
| EPI_ISL_413520 | unknown | Infectious Disease Control Center |
| EPI_ISL_413521 | unknown | Infectious Disease Control Center |
| EPI_ISL_413522 | Indian Council of Medical Research - National Institute of Virology | National Influenza Center, Indian Council of Medical Research - National Institute of Virology |
| EPI_ISL_413523 | Indian Council of Medical Research-National Institute of Virology | National Influenza Center, Indian Council of Medical Research-National Institute of Virology |
| EPI_ISL_413550 | Centre for Human and Zoonotic Virology (CHAZVY), College of Medicine University of Lagos/Lagos University Teaching Hospital (LUTH), part of the Laboratory Network of the Nigeria Centre for Disease Control (NCDC) | African Centre of Excellence for Genomics of Infectious Diseases (ACEGID), Redeemer's University, Ede, Osun State, Nigeria |
| EPI_ISL_413555 | Wales Specialist Virology Centre | Public Health Wales Microbiology Cardiff |
| EPI_ISL_413556 | Wales Specialist Virology Centre | Public Health Wales Microbiology Cardiff |
| EPI_ISL_413557 | California Department of Public Health | Chiu Laboratory, University of California, San Francisco |
| EPI_ISL_413558 | California Department of Public Health | Chiu Laboratory, University of California, San Francisco |
| EPI_ISL_413559 | California Department of Public Health | Chiu Laboratory, University of California, San Francisco |
| EPI_ISL_413560 | Seattle Flu Study | Seattle Flu Study |
| EPI_ISL_413561 | California Department of Public Health | Chiu Laboratory, University of California, San Francisco |
| EPI_ISL_413563 | UW Virology Lab | UW Virology Lab |
| EPI_ISL_413564 | MHC West-Brabant | Erasmus Medical Center |
| EPI_ISL_413565 | Foundation Pamm | Erasmus Medical Center |
| EPI_ISL_413566 | MHC Gooi & Vechtstreek | Erasmus Medical Center |
| EPI_ISL_413568 | MHC Drente | Erasmus Medical Center |
| EPI_ISL_413569 | RIVM | Erasmus Medical Center |
| EPI_ISL_413570 | RIVM | Erasmus Medical Center |
| EPI_ISL_413571 | MHC Brabant Zuidoost | Erasmus Medical Center |
| EPI_ISL_413572 | MHC Kennemerland | Erasmus Medical Center |
| EPI_ISL_413573 | Dienst Gezondheid & Jeugd Zuid-Holland Zuid | Erasmus Medical Center |
| EPI_ISL_413574 | MHC West-Brabant | Erasmus Medical Center |

| EPI_Accession_ID | Originating Laboratory | Submitting Laboratory |
|---|---|---|
| EPI_ISL_413576 | RIVM | Erasmus Medical Center |
| EPI_ISL_413577 | MHC Gooi & Vechtstreek | Erasmus Medical Center |
| EPI_ISL_413578 | ErasmusMC | Erasmus Medical Center |
| EPI_ISL_413579 | MHC Haaglanden | Erasmus Medical Center |
| EPI_ISL_413581 | RIVM | Erasmus Medical Center |
| EPI_ISL_413583 | MHC Rotterdam-Rijnmond | Erasmus Medical Center |
| EPI_ISL_413584 | unknown | Erasmus Medical Center |
| EPI_ISL_413586 | Foundation Elisabeth-Tweesteden Ziekenhuis | Erasmus Medical Center |
| EPI_ISL_413587 | Foundation Elisabeth-Tweesteden Ziekenhuis | Erasmus Medical Center |
| EPI_ISL_413588 | MHC Utrecht | Erasmus Medical Center |
| EPI_ISL_413589 | MHC Utrecht | Erasmus Medical Center |
| EPI_ISL_413591 | MHC Flevoland | Erasmus Medical Center |
| EPI_ISL_413592 | Department of Laboratory Medicine, National Taiwan University Hospital | Microbial Genomics Core Lab, National Taiwan University Centers of Genomic and Precision Medicine |
| EPI_ISL_413593 | Laboratoire National de SantÃ© | Erasmus Medical Center |
| EPI_ISL_413594 | Centre for Infectious Diseases and Microbiology Laboratory Services | NSW Health Pathology - Institute of Clinical Pathology and Medical Research; Westmead Hospital; University of Sydney |
| EPI_ISL_413595 | Centre for Infectious Diseases and Microbiology Laboratory Services | NSW Health Pathology - Institute of Clinical Pathology and Medical Research; Westmead Hospital; University of Sydney |
| EPI_ISL_413596 | Centre for Infectious Diseases and Microbiology - Public Health | NSW Health Pathology - Institute of Clinical Pathology and Medical Research; Westmead Hospital; University of Sydney |
| EPI_ISL_413597 | Centre for Infectious Diseases and Microbiology- Public Health | NSW Health Pathology - Institute of Clinical Pathology and Medical Research; Westmead Hospital; University of Sydney |
| EPI_ISL_413598 | Centre for Infectious Diseases and Microbiology - Public Health | NSW Health Pathology - Institute of Clinical Pathology and Medical Research; Westmead Hospital; University of Sydney |
| EPI_ISL_413599 | Centre for Infectious Diseases and Microbiology - Public Health | NSW Health Pathology - Institute of Clinical Pathology and Medical Research; Westmead Hospital; University of Sydney |

| EPI_Accession_ID | Originating Laboratory | Submitting Laboratory |
|---|---|---|
| EPI_ISL_413600 | Centre for Infectious Diseases and Microbiology - Public Health | NSW Health Pathology - Institute of Clinical Pathology and Medical Research; Westmead Hospital; University of Sydney |
| EPI_ISL_413601 | UW Virology Lab | UW Virology Lab |
| EPI_ISL_413602 | Department of Virology and Immunology, University of Helsinki and Helsinki University Hospital, Huslab Finland | Department of Virology, Faculty of Medicine, University of Helsinki, Helsinki, Finland |
| EPI_ISL_413603 | Department of Virology and Immunology, University of Helsinki and Helsinki University Hospital, Huslab Finland | Department of Virology, Faculty of Medicine, University of Helsinki, Helsinki, Finland |
| EPI_ISL_413604 | Department of Virology and Immunology, University of Helsinki and Helsinki University Hospital, Huslab Finland | Department of Virology, Faculty of Medicine, University of Helsinki, Helsinki, Finland |
| EPI_ISL_413605 | Department of Virology and Immunology, University of Helsinki and Helsinki University Hospital, Huslab Finland | Department of Virology, Faculty of Medicine, University of Helsinki, Helsinki, Finland |
| EPI_ISL_413647 | Centro Hospital do Porto, E.P.E. - H. Geral de Santo Antonio | Instituto Nacional de Saude (INSA) |
| EPI_ISL_413648 | Centro Hospitalar e UniversitÃ¡rio de Sao Joao, Porto | Instituto Nacional de Saude (INSA) |
| EPI_ISL_413649 | UW Virology Lab | UW Virology Lab |
| EPI_ISL_413650 | UW Virology Lab | UW Virology Lab |
| EPI_ISL_413651 | UW Virology Lab | UW Virology Lab |
| EPI_ISL_413652 | UW Virology Lab | UW Virology Lab |
| EPI_ISL_413653 | UW Virology Lab | UW Virology Lab |
| EPI_ISL_413850 | Guangdong Provincial Institution of Public Health, Guangdong Provinical Center for Disease Control and Prevention | Guangdong Provincial Institution of Public Health |
| EPI_ISL_413851 | Guangdong Provincial Institution of Public Health, Guangdong Provinical Center for Disease Control and Prevention | Guangdong Provincial Institution of Public Health |
| EPI_ISL_413852 | Guangdong Provincial Institution of Public Health, Guangdong Provinical Center for Disease Control and Prevention | Guangdong Provincial Institution of Public Health |

| EPI_Accession_ID | Originating Laboratory | Submitting Laboratory |
|---|---|---|
| EPI_ISL_413853 | Guangdong Provincial Institution of Public Health, Guangdong Provinical Center for Disease Control and Prevention | Guangdong Provincial Institution of Public Health |
| EPI_ISL_413854 | Guangdong Provincial Institution of Public Health, Guangdong Provinical Center for Disease Control and Prevention | Guangdong Provincial Institution of Public Health |
| EPI_ISL_413855 | Guangdong Provincial Institution of Public Health, Guangdong Provinical Center for Disease Control and Prevention | Guangdong Provincial Institution of Public Health |
| EPI_ISL_413856 | Guangdong Provincial Institution of Public Health, Guangdong Provinical Center for Disease Control and Prevention | Guangdong Provincial Institution of Public Health |
| EPI_ISL_413857 | Guangdong Provincial Institution of Public Health, Guangdong Provinical Center for Disease Control and Prevention | Guangdong Provincial Institution of Public Health |
| EPI_ISL_413858 | Guangdong Provincial Institution of Public Health, Guangdong Provinical Center for Disease Control and Prevention | Guangdong Provincial Institution of Public Health |
| EPI_ISL_413859 | Guangdong Provincial Institution of Public Health, Guangdong Provinical Center for Disease Control and Prevention | Guangdong Provincial Institution of Public Health |
| EPI_ISL_413860 | Guangdong Provincial Institution of Public Health, Guangdong Provinical Center for Disease Control and Prevention | Guangdong Provincial Institution of Public Health |
| EPI_ISL_413861 | Guangdong Provincial Institution of Public Health, Guangdong Provinical Center for Disease Control and Prevention | Guangdong Provincial Institution of Public Health |
| EPI_ISL_413862 | Guangdong Provincial Institution of Public Health, Guangdong Provinical Center for Disease Control and Prevention | Guangdong Provincial Institution of Public Health |
| EPI_ISL_413863 | Guangdong Provincial Institution of Public Health, Guangdong Provinical Center for Disease Control and Prevention | Guangdong Provincial Institution of Public Health |

| EPI_Accession_ID | Originating Laboratory | Submitting Laboratory |
|---|---|---|
| EPI_ISL_413864 | Guangdong Provincial Institution of Public Health, Guangdong Provinical Center for Disease Control and Prevention | Guangdong Provincial Institution of Public Health |
| EPI_ISL_413866 | Guangdong Provincial Institution of Public Health, Guangdong Provinical Center for Disease Control and Prevention | Guangdong Provincial Institution of Public Health |
| EPI_ISL_413867 | Guangdong Provincial Institution of Public Health, Guangdong Provinical Center for Disease Control and Prevention | Guangdong Provincial Institution of Public Health |
| EPI_ISL_413868 | Guangdong Provincial Institution of Public Health, Guangdong Provinical Center for Disease Control and Prevention | Guangdong Provincial Institution of Public Health |
| EPI_ISL_413869 | Guangdong Provincial Institution of Public Health, Guangdong Provinical Center for Disease Control and Prevention | Guangdong Provincial Institution of Public Health |
| EPI_ISL_413870 | Guangdong Provincial Institution of Public Health, Guangdong Provinical Center for Disease Control and Prevention | Guangdong Provincial Institution of Public Health |
| EPI_ISL_413871 | Guangdong Provincial Institution of Public Health, Guangdong Provinical Center for Disease Control and Prevention | Guangdong Provincial Institution of Public Health |
| EPI_ISL_413872 | Guangdong Provincial Institution of Public Health, Guangdong Provinical Center for Disease Control and Prevention | Guangdong Provincial Institution of Public Health |
| EPI_ISL_413873 | Guangdong Provincial Institution of Public Health, Guangdong Provinical Center for Disease Control and Prevention | Guangdong Provincial Institution of Public Health |
| EPI_ISL_413874 | Guangdong Provincial Institution of Public Health, Guangdong Provinical Center for Disease Control and Prevention | Guangdong Provincial Institution of Public Health |
| EPI_ISL_413875 | Guangdong Provincial Institution of Public Health, Guangdong Provinical Center for Disease Control and Prevention | Guangdong Provincial Institution of Public Health |

| EPI_Accession_ID | Originating Laboratory | Submitting Laboratory |
|---|---|---|
| EPI_ISL_413878 | Guangdong Provincial Institution of Public Health, Guangdong Provinical Center for Disease Control and Prevention | Guangdong Provincial Institution of Public Health |
| EPI_ISL_413879 | Guangdong Provincial Institution of Public Health, Guangdong Provinical Center for Disease Control and Prevention | Guangdong Provincial Institution of Public Health |
| EPI_ISL_413880 | Guangdong Provincial Institution of Public Health, Guangdong Provinical Center for Disease Control and Prevention | Guangdong Provincial Institution of Public Health |
| EPI_ISL_413882 | Guangdong Provincial Institution of Public Health, Guangdong Provinical Center for Disease Control and Prevention | Guangdong Provincial Institution of Public Health |
| EPI_ISL_413883 | Guangdong Provincial Institution of Public Health, Guangdong Provinical Center for Disease Control and Prevention | Guangdong Provincial Institution of Public Health |
| EPI_ISL_413884 | Guangdong Provincial Institution of Public Health, Guangdong Provinical Center for Disease Control and Prevention | Guangdong Provincial Institution of Public Health |
| EPI_ISL_413888 | Guangdong Provincial Institution of Public Health, Guangdong Provinical Center for Disease Control and Prevention | Guangdong Provincial Institution of Public Health |
| EPI_ISL_413889 | Guangdong Provincial Institution of Public Health, Guangdong Provinical Center for Disease Control and Prevention | Guangdong Provincial Institution of Public Health |
| EPI_ISL_413892 | Guangdong Provincial Institution of Public Health, Guangdong Provinical Center for Disease Control and Prevention | Guangdong Provincial Institution of Public Health |
| EPI_ISL_413893 | Guangdong Provincial Institution of Public Health, Guangdong Provinical Center for Disease Control and Prevention | Guangdong Provincial Institution of Public Health |
| EPI_ISL_413894 | Guangdong Provincial Institution of Public Health, Guangdong Provinical Center for Disease Control and Prevention | Guangdong Provincial Institution of Public Health |

| EPI_Accession_ID | Originating Laboratory | Submitting Laboratory |
|---|---|---|
| EPI_ISL_413896 | Guangdong Provincial Institution of Public Health, Guangdong Provinical Center for Disease Control and Prevention | Guangdong Provincial Institution of Public Health |
| EPI_ISL_413897 | Guangdong Provincial Institution of Public Health, Guangdong Provinical Center for Disease Control and Prevention | Guangdong Provincial Institution of Public Health |
| EPI_ISL_413898 | Guangdong Provincial Institution of Public Health, Guangdong Provinical Center for Disease Control and Prevention | Guangdong Provincial Institution of Public Health |
| EPI_ISL_413900 | Guangdong Provincial Institution of Public Health, Guangdong Provinical Center for Disease Control and Prevention | Guangdong Provincial Institution of Public Health |
| EPI_ISL_413902 | Guangdong Provincial Institution of Public Health, Guangdong Provinical Center for Disease Control and Prevention | Guangdong Provincial Institution of Public Health |
| EPI_ISL_413922 | California Department of Public Health | Chiu Laboratory, University of California, San Francisco |
| EPI_ISL_413924 | California Department of Public Health | Chiu Laboratory, University of California, San Francisco |
| EPI_ISL_413925 | California Department of Public Health | Chiu Laboratory, University of California, San Francisco |
| EPI_ISL_413926 | California Department of Public Health | Chiu Laboratory, University of California, San Francisco |
| EPI_ISL_413928 | California Department of Public Health | Chiu Laboratory, University of California, San Francisco |
| EPI_ISL_413930 | California Department of Public Health | Chiu Laboratory UCSF-Abbott Viral Diagnostics and Discovery Center University of California, San Francisco |
| EPI_ISL_413931 | California Department of Public Health | Chiu Laboratory, University of California, San Francisco |
| EPI_ISL_413996 | Laboratoire de Virologie, HUG | Swiss National Reference Centre for Influenza |
| EPI_ISL_413997 | Laboratoire de Virologie, HUG | Swiss National Reference Centre for Influenza |
| EPI_ISL_413999 | Laboratoire de Virologie, HUG | Swiss National Reference Centre for Influenza |
| EPI_ISL_414005 | Respiratory Virus Unit, Microbiology Services Colindale, Public Health England | Respiratory Virus Unit, Microbiology Services Colindale, Public Health England |
| EPI_ISL_414006 | Respiratory Virus Unit, Microbiology Services Colindale, Public Health England | Respiratory Virus Unit, Microbiology Services Colindale, Public Health England |
| EPI_ISL_414007 | Respiratory Virus Unit, Microbiology Services Colindale, Public Health England | Respiratory Virus Unit, Microbiology Services Colindale, Public Health England |

| EPI_Accession_ID | Originating Laboratory | Submitting Laboratory |
|---|---|---|
| EPI_ISL_414008 | Respiratory Virus Unit, Microbiology Services Colindale, Public Health England | Respiratory Virus Unit, Microbiology Services Colindale, Public Health England |
| EPI_ISL_414009 | Respiratory Virus Unit, Microbiology Services Colindale, Public Health England | Respiratory Virus Unit, Microbiology Services Colindale, Public Health England |
| EPI_ISL_414010 | Respiratory Virus Unit, Microbiology Services Colindale, Public Health England | Respiratory Virus Unit, Microbiology Services Colindale, Public Health England |
| EPI_ISL_414011 | Respiratory Virus Unit, Microbiology Services Colindale, Public Health England | Respiratory Virus Unit, Microbiology Services Colindale, Public Health England |
| EPI_ISL_414012 | Respiratory Virus Unit, Microbiology Services Colindale, Public Health England | Respiratory Virus Unit, Microbiology Services Colindale, Public Health England |
| EPI_ISL_414013 | Respiratory Virus Unit, Microbiology Services Colindale, Public Health England | Respiratory Virus Unit, Microbiology Services Colindale, Public Health England |
| EPI_ISL_414014 | Hospital Israelita Albert Einstein | Instituto Adolfo Lutz, Interdiciplinary Procedures Center, Strategic Laboratory |
| EPI_ISL_414015 | Hospital SÃ£o Joaquim Beneficencia Portuguesa | Instituto Adolfo Lutz, Interdiciplinary Procedures Center, Strategic Laboratory |
| EPI_ISL_414016 | Hospital SÃ£o Joaquim Beneficencia Portuguesa | Instituto Adolfo Lutz, Interdiciplinary Procedures Center, Strategic Laboratory |
| EPI_ISL_414017 | Hospital SÃ£o Joaquim Beneficencia Portuguesa | Instituto Adolfo Lutz, Interdiciplinary Procedures Center, Strategic Laboratory |
| EPI_ISL_414019 | Laboratoire de Virologie, HUG | Swiss National Reference Centre for Influenza |
| EPI_ISL_414020 | Laboratoire de Virologie, HUG | Swiss National Reference Centre for Influenza |
| EPI_ISL_414021 | Laboratoire de Virologie, HUG | Swiss National Reference Centre for Influenza |
| EPI_ISL_414022 | Laboratoire de Virologie, HUG | Swiss National Reference Centre for Influenza |
| EPI_ISL_414023 | Laboratoire de Virologie, HUG | Swiss National Reference Centre for Influenza |
| EPI_ISL_414024 | West of Scotland Specialist Virology Centre, NHSGGC | MRC-University of Glasgow Centre for Virus Research |
| EPI_ISL_414025 | West of Scotland Specialist Virology Centre, NHSGGC | MRC-University of Glasgow Centre for Virus Research |
| EPI_ISL_414026 | West of Scotland Specialist Virology Centre, NHSGGC | MRC-University of Glasgow Centre for Virus Research |
| EPI_ISL_414027 | West of Scotland Specialist Virology Centre, NHSGGC | MRC-University of Glasgow Centre for Virus Research |
| EPI_ISL_414040 | Respiratory Virus Unit, Microbiology Services Colindale, Public Health England | Respiratory Virus Unit, Microbiology Services Colindale, Public Health England |
| EPI_ISL_414041 | Respiratory Virus Unit, Microbiology Services Colindale, Public Health England | Respiratory Virus Unit, Microbiology Services Colindale, Public Health England |

| EPI_Accession_ID | Originating Laboratory | Submitting Laboratory |
|---|---|---|
| EPI_ISL_414042 | Respiratory Virus Unit, Microbiology Services Colindale, Public Health England | Respiratory Virus Unit, Microbiology Services Colindale, Public Health England |
| EPI_ISL_414043 | Respiratory Virus Unit, Microbiology Services Colindale, Public Health England | Respiratory Virus Unit, Microbiology Services Colindale, Public Health England |
| EPI_ISL_414044 | Respiratory Virus Unit, Microbiology Services Colindale, Public Health England | Respiratory Virus Unit, Microbiology Services Colindale, Public Health England |
| EPI_ISL_414045 | LACEN RJ - LaboratÃ³rio Central de SaÃºde PÃºblica Noel Nutels | Instituto Oswaldo Cruz FIOCRUZ - Laboratory of Respiratory Viruses and Measles (LVRS) |
| EPI_ISL_414363 | UW Virology Lab | UW Virology Lab |
| EPI_ISL_414364 | UW Virology Lab | UW Virology Lab |
| EPI_ISL_414365 | UW Virology Lab | UW Virology Lab |
| EPI_ISL_414366 | UW Virology Lab | UW Virology Lab |
| EPI_ISL_414367 | UW Virology Lab | UW Virology Lab |
| EPI_ISL_414368 | UW Virology Lab | UW Virology Lab |
| EPI_ISL_414369 | UW Virology Lab | UW Virology Lab |
| EPI_ISL_414378 | National Centre for Infectious Diseases | Programme in Emerging Infectious Diseases, Duke-NUS Medical School |
| EPI_ISL_414379 | National Centre for Infectious Diseases | Programme in Emerging Infectious Diseases, Duke-NUS Medical School |
| EPI_ISL_414380 | National Centre for Infectious Diseases | Programme in Emerging Infectious Diseases, Duke-NUS Medical School |
| EPI_ISL_414414 | Pathology Queensland | Public Health Virology Laboratory |
| EPI_ISL_414423 | Dutch COVID-19 response team | Erasmus Medical Center |
| EPI_ISL_414424 | Dutch COVID-19 response team | Erasmus Medical Center |
| EPI_ISL_414425 | Dutch COVID-19 response team | Erasmus Medical Center |
| EPI_ISL_414426 | Dutch COVID-19 response team | Erasmus Medical Center |
| EPI_ISL_414427 | Dutch COVID-19 response team | Erasmus Medical Center |
| EPI_ISL_414428 | Dutch COVID-19 response team | Erasmus Medical Center |
| EPI_ISL_414429 | Dutch COVID-19 response team | Erasmus Medical Center |
| EPI_ISL_414431 | Dutch COVID-19 response team | Erasmus Medical Center |
| EPI_ISL_414432 | Dutch COVID-19 response team | Erasmus Medical Center |
| EPI_ISL_414433 | Dutch COVID-19 response team | Erasmus Medical Center |
| EPI_ISL_414434 | Dutch COVID-19 response team | Erasmus Medical Center |

| EPI_Accession_ID | Originating Laboratory | Submitting Laboratory |
|---|---|---|
| EPI_ISL_414435 | Dutch COVID-19 response team | Erasmus Medical Center |
| EPI_ISL_414436 | Dutch COVID-19 response team | Erasmus Medical Center |
| EPI_ISL_414437 | Dutch COVID-19 response team | Erasmus Medical Center |
| EPI_ISL_414438 | Dutch COVID-19 response team | Erasmus Medical Center |
| EPI_ISL_414439 | Dutch COVID-19 response team | Erasmus Medical Center |
| EPI_ISL_414440 | Dutch COVID-19 response team | Erasmus Medical Center |
| EPI_ISL_414441 | Dutch COVID-19 response team | Erasmus Medical Center |
| EPI_ISL_414442 | Dutch COVID-19 response team | Erasmus Medical Center |
| EPI_ISL_414443 | Dutch COVID-19 response team | Erasmus Medical Center |
| EPI_ISL_414444 | Dutch COVID-19 response team | Erasmus Medical Center |
| EPI_ISL_414445 | Dutch COVID-19 response team | Erasmus Medical Center |
| EPI_ISL_414446 | Dutch COVID-19 response team | Erasmus Medical Center |
| EPI_ISL_414448 | Dutch COVID-19 response team | Erasmus Medical Center |
| EPI_ISL_414449 | Dutch COVID-19 response team | Erasmus Medical Center |
| EPI_ISL_414450 | Dutch COVID-19 response team | Erasmus Medical Center |
| EPI_ISL_414451 | Dutch COVID-19 response team | Erasmus Medical Center |
| EPI_ISL_414452 | Dutch COVID-19 response team | Erasmus Medical Center |
| EPI_ISL_414453 | Dutch COVID-19 response team | Erasmus Medical Center |
| EPI_ISL_414454 | Dutch COVID-19 response team | Erasmus Medical Center |
| EPI_ISL_414455 | Dutch COVID-19 response team | Erasmus Medical Center |
| EPI_ISL_414456 | Dutch COVID-19 response team | Erasmus Medical Center |
| EPI_ISL_414457 | Dutch COVID-19 response team | Erasmus Medical Center |
| EPI_ISL_414458 | Dutch COVID-19 response team | Erasmus Medical Center |
| EPI_ISL_414459 | Dutch COVID-19 response team | Erasmus Medical Center |
| EPI_ISL_414460 | Dutch COVID-19 response team | Erasmus Medical Center |
| EPI_ISL_414461 | Dutch COVID-19 response team | Erasmus Medical Center |
| EPI_ISL_414462 | Dutch COVID-19 response team | Erasmus Medical Center |
| EPI_ISL_414463 | Dutch COVID-19 response team | Erasmus Medical Center |
| EPI_ISL_414464 | Dutch COVID-19 response team | Erasmus Medical Center |
| EPI_ISL_414465 | Dutch COVID-19 response team | Erasmus Medical Center |
| EPI_ISL_414466 | Dutch COVID-19 response team | Erasmus Medical Center |
| EPI_ISL_414467 | Dutch COVID-19 response team | Erasmus Medical Center |
| EPI_ISL_414468 | Dutch COVID-19 response team | Erasmus Medical Center |

| EPI_Accession_ID | Originating Laboratory | Submitting Laboratory |
|---|---|---|
| EPI_ISL_414469 | Dutch COVID-19 response team | Erasmus Medical Center |
| EPI_ISL_414470 | Dutch COVID-19 response team | Erasmus Medical Center |
| EPI_ISL_414471 | Dutch COVID-19 response team | Erasmus Medical Center |
| EPI_ISL_414476 | MSHS Clinical Microbiology Laboratories | MSHS Pathogen Surveillance Program |
| EPI_ISL_414477 | The National Institute of Public Health Center for Epidemiology and Microbiology | State Veterinary Institute Prague |
| EPI_ISL_414486 | Wales Specialist Virology Centre | Public Health Wales Microbiology Cardiff |
| EPI_ISL_414488 | Wales Specialist Virology Centre | Public Health Wales Microbiology Cardiff |
| EPI_ISL_414495 | Servicio MicrobiologÃ-a. Hospital ClÃ-nico Universitario. Valencia. | Sequencing and Bioinformatics Service. Molecular Epidemiology Laboratory. FISABIO-Public Health |
| EPI_ISL_414496 | Servicio Microbiologia. Hospital Clinico Universitario. Valencia. | Sequencing and Bioinformatics Service. Molecular Epidemiology Laboratory. FISABIO-Public Health |
| EPI_ISL_414497 | Center of Medical Microbiology, Virology, and Hospital Hygiene, University of Duesseldorf | Center of Medical Microbiology, Virology, and Hospital Hygiene, University of Duesseldorf |
| EPI_ISL_414498 | Center of Medical Microbiology, Virology, and Hospital Hygiene, University of Duesseldorf | Center of Medical Microbiology, Virology, and Hospital Hygiene, University of Duesseldorf |
| EPI_ISL_414499 | Center of Medical Microbiology, Virology, and Hospital Hygiene, University of Duesseldorf | Center of Medical Microbiology, Virology, and Hospital Hygiene, University of Duesseldorf |
| EPI_ISL_414500 | Virology Department, Sheffield Teaching Hospitals NHS Foundation Trust | Department of Infection, Immunity and Cardiovascular Disease, The Florey Institute, The Medical School, University of Sheffield |
| EPI_ISL_414501 | Virology Department, Sheffield Teaching Hospitals NHS Foundation Trust | Department of Infection, Immunity and Cardiovascular Disease, The Florey Institute, The Medical School, University of Sheffield |
| EPI_ISL_414504 | Center of Medical Microbiology, Virology, and Hospital Hygiene, University of Duesseldorf | Center of Medical Microbiology, Virology, and Hospital Hygiene, University of Duesseldorf |
| EPI_ISL_414505 | Center of Medical Microbiology, Virology, and Hospital Hygiene, University of Duesseldorf | Center of Medical Microbiology, Virology, and Hospital Hygiene, University of Duesseldorf |
| EPI_ISL_414506 | Center of Medical Microbiology, Virology, and Hospital Hygiene, University of Duesseldorf | Center of Medical Microbiology, Virology, and Hospital Hygiene, University of Duesseldorf |
| EPI_ISL_414507 | Center of Medical Microbiology, Virology, and Hospital Hygiene, University of Duesseldorf | Center of Medical Microbiology, Virology, and Hospital Hygiene, University of Duesseldorf |

| EPI_Accession_ID | Originating Laboratory | Submitting Laboratory |
|---|---|---|
| EPI_ISL_414508 | Center of Medical Microbiology, Virology, and Hospital Hygiene, University of Duesseldorf | Center of Medical Microbiology, Virology, and Hospital Hygiene, University of Duesseldorf |
| EPI_ISL_414509 | Center of Medical Microbiology, Virology, and Hospital Hygiene, University of Duesseldorf | Center of Medical Microbiology, Virology, and Hospital Hygiene, University of Duesseldorf |
| EPI_ISL_414510 | unknown | Key Laboratory of Medical Molecular Virology (MOE/NHC/CAMS) |
| EPI_ISL_414517 | Hong Kong Department of Health | School of Public Health, The University of Hong Kong |
| EPI_ISL_414518 | Tai Lung Veterinary Laboratory, Agriculture, Fisheries and Conservation Department | School of Public Health, The University of Hong Kong |
| EPI_ISL_414519 | Hong Kong Department of Health | School of Public Health, The University of Hong Kong |
| EPI_ISL_414520 | Bundeswehr Institute of Microbiology | Bundeswehr Institute of Microbiology |
| EPI_ISL_414521 | Bundeswehr Institute of Microbiology | Bundeswehr Institute of Microbiology |
| EPI_ISL_414522 | Respiratory Virus Unit, Microbiology Services Colindale, Public Health England | Respiratory Virus Unit, Microbiology Services Colindale, Public Health England |
| EPI_ISL_414523 | Respiratory Virus Unit, Microbiology Services Colindale, Public Health England | Respiratory Virus Unit, Microbiology Services Colindale, Public Health England |
| EPI_ISL_414524 | Respiratory Virus Unit, Microbiology Services Colindale, Public Health England | Respiratory Virus Unit, Microbiology Services Colindale, Public Health England |
| EPI_ISL_414525 | Respiratory Virus Unit, Microbiology Services Colindale, Public Health England | Respiratory Virus Unit, Microbiology Services Colindale, Public Health England |
| EPI_ISL_414526 | Respiratory Virus Unit, Microbiology Services Colindale, Public Health England | Respiratory Virus Unit, Microbiology Services Colindale, Public Health England |
| EPI_ISL_414527 | Hong Kong Department of Health | School of Public Health, The University of Hong Kong |
| EPI_ISL_414528 | Hong Kong Department of Health | School of Public Health, The University of Hong Kong |
| EPI_ISL_414529 | Dutch COVID-19 response team | Erasmus Medical Center |
| EPI_ISL_414530 | Dutch COVID-19 response team | Erasmus Medical Center |
| EPI_ISL_414531 | Dutch COVID-19 response team | Erasmus Medical Center |
| EPI_ISL_414532 | Dutch COVID-19 response team | Erasmus Medical Center |
| EPI_ISL_414533 | Dutch COVID-19 response team | Erasmus Medical Center |
| EPI_ISL_414534 | Dutch COVID-19 response team | Erasmus Medical Center |
| EPI_ISL_414535 | Dutch COVID-19 response team | Erasmus Medical Center |
| EPI_ISL_414536 | Dutch COVID-19 response team | Erasmus Medical Center |
| EPI_ISL_414537 | Dutch COVID-19 response team | Erasmus Medical Center |

| EPI_Accession_ID | Originating Laboratory | Submitting Laboratory |
|---|---|---|
| EPI_ISL_414538 | Dutch COVID-19 response team | Erasmus Medical Center |
| EPI_ISL_414539 | Dutch COVID-19 response team | Erasmus Medical Center |
| EPI_ISL_414540 | Dutch COVID-19 response team | Erasmus Medical Center |
| EPI_ISL_414541 | Dutch COVID-19 response team | Erasmus Medical Center |
| EPI_ISL_414544 | Dutch COVID-19 response team | Erasmus Medical Center |
| EPI_ISL_414545 | Dutch COVID-19 response team | Erasmus Medical Center |
| EPI_ISL_414546 | Dutch COVID-19 response team | Erasmus Medical Center |
| EPI_ISL_414547 | Dutch COVID-19 response team | Erasmus Medical Center |
| EPI_ISL_414548 | Dutch COVID-19 response team | Erasmus Medical Center |
| EPI_ISL_414549 | Dutch COVID-19 response team | Erasmus Medical Center |
| EPI_ISL_414550 | Dutch COVID-19 response team | Erasmus Medical Center |
| EPI_ISL_414551 | Dutch COVID-19 response team | Erasmus Medical Center |
| EPI_ISL_414552 | Dutch COVID-19 response team | Erasmus Medical Center |
| EPI_ISL_414553 | Dutch COVID-19 response team | Erasmus Medical Center |
| EPI_ISL_414554 | Dutch COVID-19 response team | Erasmus Medical Center |
| EPI_ISL_414555 | Dutch COVID-19 response team | Erasmus Medical Center |
| EPI_ISL_414556 | Dutch COVID-19 response team | Erasmus Medical Center |
| EPI_ISL_414557 | Dutch COVID-19 response team | Erasmus Medical Center |
| EPI_ISL_414558 | Dutch COVID-19 response team | Erasmus Medical Center |
| EPI_ISL_414559 | Dutch COVID-19 response team | Erasmus Medical Center |
| EPI_ISL_414560 | Dutch COVID-19 response team | Erasmus Medical Center |
| EPI_ISL_414561 | Dutch COVID-19 response team | Erasmus Medical Center |
| EPI_ISL_414562 | Dutch COVID-19 response team | Erasmus Medical Center |
| EPI_ISL_414563 | Dutch COVID-19 response team | Erasmus Medical Center |
| EPI_ISL_414564 | Dutch COVID-19 response team | Erasmus Medical Center |
| EPI_ISL_414565 | Dutch COVID-19 response team | Erasmus Medical Center |
| EPI_ISL_414566 | Dutch COVID-19 response team | Erasmus Medical Center |
| EPI_ISL_414569 | Hong Kong Department of Health | School of Public Health, The University of Hong Kong |
| EPI_ISL_414571 | Hong Kong Department of Health | School of Public Health, The University of Hong Kong |
| EPI_ISL_414574 | Center of Medical Microbiology, Virology, and Hospital Hygiene, University of Duesseldorf | Center of Medical Microbiology, Virology, and Hospital Hygiene, University of Duesseldorf |
| EPI_ISL_414577 | Hospital de Talca, Chile | Instituto de Salud Publica de Chile |
| EPI_ISL_414578 | Hospital de Talca, Chile | Instituto de Salud Publica de Chile |

| EPI_Accession_ID | Originating Laboratory | Submitting Laboratory |
|---|---|---|
| EPI_ISL_414579 | Clinica Alemana de Santiago, Chile | Instituto de Salud Publica de Chile |
| EPI_ISL_414580 | Clinica Santa Maria, Santiago, Chile | Instituto de Salud Publica de Chile |
| EPI_ISL_414584 | UCD National Virus Reference Laboratory | UCD National Virus Reference Laboratory |
| EPI_ISL_414585 | UCD National Virus Reference Laboratory | UCD National Virus Reference Laboratory |
| EPI_ISL_414586 | UCD National Virus Reference Laboratory | UCD National Virus Reference Laboratory |
| EPI_ISL_414587 | UCD National Virus Reference Laboratory | UCD National Virus Reference Laboratory |
| EPI_ISL_414589 | Minnesota Department of Health, Public Health Laboratory | Minnesota Department of Health, Public Health Laboratory |
| EPI_ISL_414590 | Minnesota Department of Health, Public Health Laboratory | Minnesota Department of Health, Public Health Laboratory |
| EPI_ISL_414591 | UW Virology Lab | UW Virology Lab |
| EPI_ISL_414592 | UW Virology Lab | UW Virology Lab |
| EPI_ISL_414593 | UW Virology Lab | UW Virology Lab |
| EPI_ISL_414594 | UW Virology Lab | UW Virology Lab |
| EPI_ISL_414595 | UW Virology Lab | UW Virology Lab |
| EPI_ISL_414596 | UW Virology Lab | UW Virology Lab |
| EPI_ISL_414597 | UW Virology Lab | UW Virology Lab |
| EPI_ISL_414598 | Servicio Microbiologia, Hospital Clinico Universitario, Valencia | Sequencing and Bioinformatics Service and Molecular Epidemiology Research Group. FISABIO-Public Health. |
| EPI_ISL_414616 | UW Virology Lab | UW Virology Lab |
| EPI_ISL_414617 | UW Virology Lab | UW Virology Lab |
| EPI_ISL_414618 | UW Virology Lab | UW Virology Lab |
| EPI_ISL_414619 | UW Virology Lab | UW Virology Lab |
| EPI_ISL_414620 | UW Virology Lab | UW Virology Lab |
| EPI_ISL_414621 | UW Virology Lab | UW Virology Lab |
| EPI_ISL_414622 | UW Virology Lab | UW Virology Lab |
| EPI_ISL_414623 | Laboratoire de Virologie Institut de Virologie - INSERM U 1109 HÃ´pitaux Universitaires de Strasbourg | National Reference Center for Viruses of Respiratory Infections, Institut Pasteur, Paris |
| EPI_ISL_414624 | Centre Hositalier Universitaire de Rouen Laboratoire de Virologie | National Reference Center for Viruses of Respiratory Infections, Institut Pasteur, Paris |
| EPI_ISL_414625 | Centre Hospitalier RÃ©gional Universitaire de Nantes Laboratoire de Virologie | National Reference Center for Viruses of Respiratory Infections, Institut Pasteur, Paris |

| EPI_Accession_ID | Originating Laboratory | Submitting Laboratory |
|---|---|---|
| EPI_ISL_414626 | unknown | National Reference Center for Viruses of Respiratory Infections, Institut Pasteur, Paris |
| EPI_ISL_414627 | Centre Hospitalier CompiÃ¨gne Laboratoire de Biologie | National Reference Center for Viruses of Respiratory Infections, Institut Pasteur, Paris |
| EPI_ISL_414629 | Centre Hospitalier CompiÃ¨gne Laboratoire de Biologie | National Reference Center for Viruses of Respiratory Infections, Institut Pasteur, Paris |
| EPI_ISL_414630 | Centre Hospitalier CompiÃ¨gne Laboratoire de Biologie | National Reference Center for Viruses of Respiratory Infections, Institut Pasteur, Paris |
| EPI_ISL_414631 | HÃ´pital Robert DebrÃ© Laboratoire de Virologie | National Reference Center for Viruses of Respiratory Infections, Institut Pasteur, Paris |
| EPI_ISL_414632 | HÃ´pital Robert DebrÃ© Laboratoire de Virologie | National Reference Center for Viruses of Respiratory Infections, Institut Pasteur, Paris |
| EPI_ISL_414633 | Centre Hospitalier RenÃ© Dubois Laboratoire de Microbiologie - BÃ¢t A | National Reference Center for Viruses of Respiratory Infections, Institut Pasteur, Paris |
| EPI_ISL_414634 | Centre Hospitalier CompiÃ¨gne Laboratoire de Biologie | National Reference Center for Viruses of Respiratory Infections, Institut Pasteur, Paris |
| EPI_ISL_414635 | Centre Hospitalier CompiÃ¨gne Laboratoire de Biologie | National Reference Center for Viruses of Respiratory Infections, Institut Pasteur, Paris |
| EPI_ISL_414636 | Centre Hospitalier CompiÃ¨gne Laboratoire de Biologie | National Reference Center for Viruses of Respiratory Infections, Institut Pasteur, Paris |
| EPI_ISL_414637 | Centre Hospitalier CompiÃ¨gne Laboratoire de Biologie | National Reference Center for Viruses of Respiratory Infections, Institut Pasteur, Paris |
| EPI_ISL_414638 | Centre Hospitalier CompiÃ¨gne Laboratoire de Biologie | National Reference Center for Viruses of Respiratory Infections, Institut Pasteur, Paris |
| EPI_ISL_414639 | NYU Langone Health | Departments of Pathology and Medicine, New York University School of Medicine |
| EPI_ISL_414640 | Department of Virology and Immunology, University of Helsinki and Helsinki University Hospital, Huslab Finland | Department of Virology, Faculty of Medicine, University of Helsinki, Helsinki, Finland |
| EPI_ISL_414641 | Department of Virology and Immunology, University of Helsinki and Helsinki University Hospital, Huslab Finland | Department of Virology, Faculty of Medicine, University of Helsinki, Helsinki, Finland |

| EPI_Accession_ID | Originating Laboratory | Submitting Laboratory |
|---|---|---|
| EPI_ISL_414642 | Department of Virology and Immunology, University of Helsinki and Helsinki University Hospital, Huslab Finland | Department of Virology, Faculty of Medicine, University of Helsinki, Helsinki, Finland |
| EPI_ISL_414643 | Department of Virology and Immunology, University of Helsinki and Helsinki University Hospital, Huslab Finland | Department of Virology, Faculty of Medicine, University of Helsinki, Helsinki, Finland |
| EPI_ISL_414644 | Department of Virology and Immunology, University of Helsinki and Helsinki University Hospital, Huslab Finland | Department of Virology, Faculty of Medicine, University of Helsinki, Helsinki, Finland |
| EPI_ISL_414645 | Department of Virology and Immunology, University of Helsinki and Helsinki University Hospital, Huslab Finland | Department of Virology, Faculty of Medicine, University of Helsinki, Helsinki, Finland |
| EPI_ISL_414646 | Department of Virology and Immunology, University of Helsinki and Helsinki University Hospital, Huslab Finland | Department of Virology, Faculty of Medicine, University of Helsinki, Helsinki, Finland |
| EPI_ISL_414647 | Viral Respiratory Lab, National Institute for Biomedical Research (INRB) | Pathogen Sequencing Lab, National Institute for Biomedical Research (INRB) |
| EPI_ISL_414648 | Andersen Lab, The Scripps Research Institute | Andersen Lab, The Scripps Research Institute |
| EPI_ISL_414686 | State Key Laboratory of Respiratory Disease, National Clinical Research Center for Respiratory Disease, Guangzhou Institute of Respiratory Health, the First Affiliated Hospital of Guangzhou Medical University | The First Affiliated Hospital of Guangzhou Medical University & BGI-Shenzhen |
| EPI_ISL_414688 | State Key Laboratory of Respiratory Disease, National Clinical Research Center for Respiratory Disease, Guangzhou Institute of Respiratory Health, the First Affiliated Hospital of Guangzhou Medical University | The First Affiliated Hospital of Guangzhou Medical University & BGI-Shenzhen |
| EPI_ISL_414689 | State Key Laboratory of Respiratory Disease, National Clinical Research Center for Respiratory Disease, Guangzhou Institute of Respiratory Health, the First Affiliated Hospital of Guangzhou Medical University | The First Affiliated Hospital of Guangzhou Medical University & BGI-Shenzhen |

| EPI_Accession_ID | Originating Laboratory | Submitting Laboratory |
|---|---|---|
| EPI_ISL_414691 | State Key Laboratory of Respiratory Disease, National Clinical Research Center for Respiratory Disease, Guangzhou Institute of Respiratory Health, the First Affiliated Hospital of Guangzhou Medical University | The First Affiliated Hospital of Guangzhou Medical University & BGI-Shenzhen |
| EPI_ISL_414692 | State Key Laboratory of Respiratory Disease, National Clinical Research Center for Respiratory Disease, Guangzhou Institute of Respiratory Health, the First Affiliated Hospital of Guangzhou Medical University | The First Affiliated Hospital of Guangzhou Medical University & BGI-Shenzhen |
| EPI_ISL_414936 | Shandong Provincial Center for Disease Control and Prevention | Beijing Institute of Microbiology and Epidemiology |
| EPI_ISL_414937 | Shandong Provincial Center for Disease Control and Prevention | Beijing Institute of Microbiology and Epidemiology |
| EPI_ISL_414938 | Shandong Provincial Center for Disease Control and Prevention | Beijing Institute of Microbiology and Epidemiology |
| EPI_ISL_414939 | Shandong Provincial Center for Disease Control and Prevention | Beijing Institute of Microbiology and Epidemiology |
| EPI_ISL_414940 | Shandong Provincial Center for Disease Control and Prevention | Beijing Institute of Microbiology and Epidemiology |
| EPI_ISL_414941 | Shandong Provincial Center for Disease Control and Prevention | Beijing Institute of Microbiology and Epidemiology |
| EPI_ISL_414949 | Regional Virus Laboratory, Belfast | Public Health Wales Microbiology Cardiff |
| EPI_ISL_415041 | Wales Specialist Virology Centre | Public Health Wales Microbiology Cardiff |
| EPI_ISL_415105 | LaboratÃ³rio Central de SaÃºde PÃºblica Professor GonÃ§alo Moniz â€“ LACEN/BA | Instituto Oswaldo Cruz FIOCRUZ - Laboratory of Respiratory Viruses and Measles (LVRS) |
| EPI_ISL_415128 | LACEN/ES - LaboratÃ³rio Central de SaÃºde PÃºblica do EspÃ-rito Santo | Instituto Oswaldo Cruz FIOCRUZ - Laboratory of Respiratory Viruses and Measles (LVRS) |
| EPI_ISL_415129 | Respiratory Virus Unit, Microbiology Services Colindale, Public Health England | Respiratory Virus Unit, Microbiology Services Colindale, Public Health England |
| EPI_ISL_415130 | Respiratory Virus Unit, Microbiology Services Colindale, Public Health England | Respiratory Virus Unit, Microbiology Services Colindale, Public Health England |
| EPI_ISL_415131 | Respiratory Virus Unit, Microbiology Services Colindale, Public Health England | Respiratory Virus Unit, Microbiology Services Colindale, Public Health England |

| EPI_Accession_ID | Originating Laboratory | Submitting Laboratory |
|---|---|---|
| EPI_ISL_415132 | Respiratory Virus Unit, Microbiology Services Colindale, Public Health England | Respiratory Virus Unit, Microbiology Services Colindale, Public Health England |
| EPI_ISL_415133 | Respiratory Virus Unit, Microbiology Services Colindale, Public Health England | Respiratory Virus Unit, Microbiology Services Colindale, Public Health England |
| EPI_ISL_415134 | Respiratory Virus Unit, Microbiology Services Colindale, Public Health England | Respiratory Virus Unit, Microbiology Services Colindale, Public Health England |
| EPI_ISL_415135 | Respiratory Virus Unit, Microbiology Services Colindale, Public Health England | Respiratory Virus Unit, Microbiology Services Colindale, Public Health England |
| EPI_ISL_415136 | Respiratory Virus Unit, Microbiology Services Colindale, Public Health England | Respiratory Virus Unit, Microbiology Services Colindale, Public Health England |
| EPI_ISL_415137 | Respiratory Virus Unit, Microbiology Services Colindale, Public Health England | Respiratory Virus Unit, Microbiology Services Colindale, Public Health England |
| EPI_ISL_415138 | Respiratory Virus Unit, Microbiology Services Colindale, Public Health England | Respiratory Virus Unit, Microbiology Services Colindale, Public Health England |
| EPI_ISL_415139 | Respiratory Virus Unit, Microbiology Services Colindale, Public Health England | Respiratory Virus Unit, Microbiology Services Colindale, Public Health England |
| EPI_ISL_415140 | Respiratory Virus Unit, Microbiology Services Colindale, Public Health England | Respiratory Virus Unit, Microbiology Services Colindale, Public Health England |
| EPI_ISL_415141 | Respiratory Virus Unit, Microbiology Services Colindale, Public Health England | Respiratory Virus Unit, Microbiology Services Colindale, Public Health England |
| EPI_ISL_415142 | Respiratory Virus Unit, Microbiology Services Colindale, Public Health England | Respiratory Virus Unit, Microbiology Services Colindale, Public Health England |
| EPI_ISL_415143 | Respiratory Virus Unit, Microbiology Services Colindale, Public Health England | Respiratory Virus Unit, Microbiology Services Colindale, Public Health England |
| EPI_ISL_415144 | Respiratory Virus Unit, Microbiology Services Colindale, Public Health England | Respiratory Virus Unit, Microbiology Services Colindale, Public Health England |
| EPI_ISL_415145 | Respiratory Virus Unit, Microbiology Services Colindale, Public Health England | Respiratory Virus Unit, Microbiology Services Colindale, Public Health England |
| EPI_ISL_415146 | Respiratory Virus Unit, Microbiology Services Colindale, Public Health England | Respiratory Virus Unit, Microbiology Services Colindale, Public Health England |
| EPI_ISL_415147 | Respiratory Virus Unit, Microbiology Services Colindale, Public Health England | Respiratory Virus Unit, Microbiology Services Colindale, Public Health England |

| EPI_Accession_ID | Originating Laboratory | Submitting Laboratory |
|---|---|---|
| EPI_ISL_415149 | Respiratory Virus Unit, Microbiology Services Colindale, Public Health England | Respiratory Virus Unit, Microbiology Services Colindale, Public Health England |
| EPI_ISL_415150 | Respiratory Virus Unit, Microbiology Services Colindale, Public Health England | Respiratory Virus Unit, Microbiology Services Colindale, Public Health England |
| EPI_ISL_415151 | MSHS Clinical Microbiology Laboratories | MSHS Pathogen Surveillance Program |
| EPI_ISL_415152 | Gorgas Memorial Institute for Health Studies | Gorgas Memorial Institute for Health Studies |
| EPI_ISL_415153 | KU Leuven, Clinical and Epidemiological Virology | KU Leuven, Clinical and Epidemiological Virology |
| EPI_ISL_415154 | KU Leuven, Clincal and Epidemiological Virology | KU Leuven, Clincal and Epidemiological Virology |
| EPI_ISL_415155 | KU Leuven, Clinical and Epidemiological Virology | KU Leuven, Clinical and Epidemiological Virology |
| EPI_ISL_415156 | KU Leuven, Clinical and Epidemiological Virology | KU Leuven, Clinical and Epidemiological Virology |
| EPI_ISL_415157 | KU Leuven, Clinical and Epidemiological Virology | KU Leuven, Clinical and Epidemiological Virology |
| EPI_ISL_415158 | KU Leuven, Clinical and Epidemiological Virology | KU Leuven, Clinical and Epidemiological Virology |
| EPI_ISL_415159 | KU Leuven, Clinical and Epidemiological Virology | KU Leuven, Clinical and Epidemiological Virology |
| EPI_ISL_415453 | Wales Specialist Virology Centre | Public Health Wales Microbiology Cardiff |
| EPI_ISL_415454 | HÃ´pitaux universitaires de GenÃ¨ve Laboratoire de Virologie | HÃ´pitaux universitaires de GenÃ¨ve Laboratoire de Virologie |
| EPI_ISL_415455 | HÃ´pitaux universitaires de GenÃ¨ve Laboratoire de Virologie | HÃ´pitaux universitaires de GenÃ¨ve Laboratoire de Virologie |
| EPI_ISL_415456 | HÃ´pitaux universitaires de GenÃ¨ve Laboratoire de Virologie | HÃ´pitaux universitaires de GenÃ¨ve Laboratoire de Virologie |
| EPI_ISL_415457 | HÃ´pitaux universitaires de GenÃ¨ve Laboratoire de Virologie | HÃ´pitaux universitaires de GenÃ¨ve Laboratoire de Virologie |
| EPI_ISL_415458 | HÃ´pitaux universitaires de GenÃ¨ve Laboratoire de Virologie | HÃ´pitaux universitaires de GenÃ¨ve Laboratoire de Virologie |
| EPI_ISL_415459 | HÃ´pitaux universitaires de GenÃ¨ve Laboratoire de Virologie | HÃ´pitaux universitaires de GenÃ¨ve Laboratoire de Virologie |
| EPI_ISL_415460 | Dutch COVID-19 response team | Erasmus Medical Center |
| EPI_ISL_415461 | Dutch COVID-19 response team | Erasmus Medical Center |
| EPI_ISL_415462 | Dutch COVID-19 response team | Erasmus Medical Center |
| EPI_ISL_415463 | Dutch COVID-19 response team | Erasmus Medical Center |
| EPI_ISL_415465 | Dutch COVID-19 response team | Erasmus Medical Center |
| EPI_ISL_415466 | Dutch COVID-19 response team | Erasmus Medical Center |
| EPI_ISL_415467 | Dutch COVID-19 response team | Erasmus Medical Center |

| EPI_Accession_ID | Originating Laboratory | Submitting Laboratory |
|---|---|---|
| EPI_ISL_415468 | Dutch COVID-19 response team | Erasmus Medical Center |
| EPI_ISL_415469 | Dutch COVID-19 response team | Erasmus Medical Center |
| EPI_ISL_415470 | Dutch COVID-19 response team | Erasmus Medical Center |
| EPI_ISL_415471 | Dutch COVID-19 response team | Erasmus Medical Center |
| EPI_ISL_415472 | Dutch COVID-19 response team | Erasmus Medical Center |
| EPI_ISL_415473 | Dutch COVID-19 response team | Erasmus Medical Center |
| EPI_ISL_415474 | Dutch COVID-19 response team | Erasmus Medical Center |
| EPI_ISL_415475 | Dutch COVID-19 response team | Erasmus Medical Center |
| EPI_ISL_415476 | Dutch COVID-19 response team | Erasmus Medical Center |
| EPI_ISL_415477 | Dutch COVID-19 response team | Erasmus Medical Center |
| EPI_ISL_415478 | Dutch COVID-19 response team | Erasmus Medical Center |
| EPI_ISL_415479 | Dutch COVID-19 response team | Erasmus Medical Center |
| EPI_ISL_415480 | Dutch COVID-19 response team | Erasmus Medical Center |
| EPI_ISL_415481 | Dutch COVID-19 response team | Erasmus Medical Center |
| EPI_ISL_415482 | Dutch COVID-19 response team | Erasmus Medical Center |
| EPI_ISL_415483 | Dutch COVID-19 response team | Erasmus Medical Center |
| EPI_ISL_415484 | Dutch COVID-19 response team | Erasmus Medical Center |
| EPI_ISL_415485 | Dutch COVID-19 response team | Erasmus Medical Center |
| EPI_ISL_415486 | Dutch COVID-19 response team | Erasmus Medical Center |
| EPI_ISL_415487 | Dutch COVID-19 response team | Erasmus Medical Center |
| EPI_ISL_415488 | Dutch COVID-19 response team | Erasmus Medical Center |
| EPI_ISL_415489 | Dutch COVID-19 response team | Erasmus Medical Center |
| EPI_ISL_415490 | Dutch COVID-19 response team | Erasmus Medical Center |
| EPI_ISL_415491 | Dutch COVID-19 response team | Erasmus Medical Center |
| EPI_ISL_415492 | Dutch COVID-19 response team | Erasmus Medical Center |
| EPI_ISL_415495 | Dutch COVID-19 response team | Erasmus Medical Center |
| EPI_ISL_415496 | Dutch COVID-19 response team | Erasmus Medical Center |
| EPI_ISL_415497 | Dutch COVID-19 response team | Erasmus Medical Center |
| EPI_ISL_415498 | Dutch COVID-19 response team | Erasmus Medical Center |
| EPI_ISL_415500 | Dutch COVID-19 response team | Erasmus Medical Center |
| EPI_ISL_415501 | Dutch COVID-19 response team | Erasmus Medical Center |
| EPI_ISL_415502 | Dutch COVID-19 response team | Erasmus Medical Center |
| EPI_ISL_415503 | Dutch COVID-19 response team | Erasmus Medical Center |

| EPI_Accession_ID | Originating Laboratory | Submitting Laboratory |
|---|---|---|
| EPI_ISL_415504 | Dutch COVID-19 response team | Erasmus Medical Center |
| EPI_ISL_415505 | Dutch COVID-19 response team | Erasmus Medical Center |
| EPI_ISL_415506 | Dutch COVID-19 response team | Erasmus Medical Center |
| EPI_ISL_415508 | Dutch COVID-19 response team | Erasmus Medical Center |
| EPI_ISL_415510 | Dutch COVID-19 response team | Erasmus Medical Center |
| EPI_ISL_415511 | Dutch COVID-19 response team | Erasmus Medical Center |
| EPI_ISL_415512 | Dutch COVID-19 response team | Erasmus Medical Center |
| EPI_ISL_415513 | Dutch COVID-19 response team | Erasmus Medical Center |
| EPI_ISL_415514 | Dutch COVID-19 response team | Erasmus Medical Center |
| EPI_ISL_415515 | Dutch COVID-19 response team | Erasmus Medical Center |
| EPI_ISL_415516 | Dutch COVID-19 response team | Erasmus Medical Center |
| EPI_ISL_415524 | Dutch COVID-19 response team | Erasmus Medical Center |
| EPI_ISL_415525 | Dutch COVID-19 response team | Erasmus Medical Center |
| EPI_ISL_415526 | Dutch COVID-19 response team | Erasmus Medical Center |
| EPI_ISL_415527 | Dutch COVID-19 response team | Erasmus Medical Center |
| EPI_ISL_415528 | Dutch COVID-19 response team | Erasmus Medical Center |
| EPI_ISL_415529 | Dutch COVID-19 response team | Erasmus Medical Center |
| EPI_ISL_415530 | Dutch COVID-19 response team | Erasmus Medical Center |
| EPI_ISL_415531 | Dutch COVID-19 response team | Erasmus Medical Center |
| EPI_ISL_415532 | Dutch COVID-19 response team | Erasmus Medical Center |
| EPI_ISL_415533 | Dutch COVID-19 response team | Erasmus Medical Center |
| EPI_ISL_415534 | Dutch COVID-19 response team | Erasmus Medical Center |
| EPI_ISL_415535 | Dutch COVID-19 response team | Erasmus Medical Center |
| EPI_ISL_415536 | Wales Specialist Virology Centre | Public Health Wales Microbiology Cardiff |
| EPI_ISL_415537 | Wales Specialist Virology Centre | Public Health Wales Microbiology Cardiff |
| EPI_ISL_415538 | Wales Specialist Virology Centre | Public Health Wales Microbiology Cardiff |
| EPI_ISL_415539 | Utah Public Health Laboratory | Utah Public Health Laboratory |
| EPI_ISL_415541 | Utah Public Health Laboratory | Utah Public Health Laboratory |
| EPI_ISL_415542 | Utah Public Health Laboratory | Utah Public Health Laboratory |
| EPI_ISL_415543 | Utah Public Health Laboratory | Utah Public Health Laboratory |
| EPI_ISL_415544 | Utah Public Health Laboratory | Utah Public Health Laboratory |
| EPI_ISL_415577 | BCCDC Public Health Laboratory | BCCDC Public Health Laboratory |
| EPI_ISL_415578 | BCCDC Public Health Laboratory | BCCDC Public Health Laboratory |

| EPI_Accession_ID | Originating Laboratory | Submitting Laboratory |
|---|---|---|
| EPI_ISL_415579 | BCCDC Public Health Laboratory | BCCDC Public Health Laboratory |
| EPI_ISL_415580 | BCCDC Public Health Laboratory | BCCDC Public Health Laboratory |
| EPI_ISL_415581 | BCCDC Public Health Laboratory | BCCDC Public Health Laboratory |
| EPI_ISL_415582 | BCCDC Public Health Laboratory | BCCDC Public Health Laboratory |
| EPI_ISL_415584 | BCCDC Public Health Laboratory | BCCDC Public Health Laboratory |
| EPI_ISL_415585 | BCCDC Public Health Laboratory | BCCDC Public Health Laboratory |
| EPI_ISL_415586 | BCCDC Public Health Laboratory | BCCDC Public Health Laboratory |
| EPI_ISL_415587 | BCCDC Public Health Laboratory | BCCDC Public Health Laboratory |
| EPI_ISL_415588 | BCCDC Public Health Laboratory | BCCDC Public Health Laboratory |
| EPI_ISL_415589 | BCCDC Public Health Laboratory | BCCDC Public Health Laboratory |
| EPI_ISL_415590 | BCCDC Public Health Laboratory | BCCDC Public Health Laboratory |
| EPI_ISL_415591 | UW Virology Lab | UW Virology Lab |
| EPI_ISL_415592 | UW Virology Lab | UW Virology Lab |
| EPI_ISL_415594 | UW Virology Lab | UW Virology Lab |
| EPI_ISL_415595 | UW Virology Lab | UW Virology Lab |
| EPI_ISL_415596 | UW Virology Lab | UW Virology Lab |
| EPI_ISL_415597 | UW Virology Lab | UW Virology Lab |
| EPI_ISL_415598 | UW Virology Lab | UW Virology Lab |
| EPI_ISL_415599 | UW Virology Lab | UW Virology Lab |
| EPI_ISL_415600 | UW Virology Lab | UW Virology Lab |
| EPI_ISL_415601 | UW Virology Lab | UW Virology Lab |
| EPI_ISL_415602 | UW Virology Lab | UW Virology Lab |
| EPI_ISL_415603 | UW Virology Lab | UW Virology Lab |
| EPI_ISL_415604 | UW Virology Lab | UW Virology Lab |
| EPI_ISL_415606 | UW Virology Lab | UW Virology Lab |
| EPI_ISL_415607 | UW Virology Lab | UW Virology Lab |
| EPI_ISL_415608 | UW Virology Lab | UW Virology Lab |
| EPI_ISL_415609 | UW Virology Lab | UW Virology Lab |
| EPI_ISL_415611 | UW Virology Lab | UW Virology Lab |
| EPI_ISL_415612 | UW Virology Lab | UW Virology Lab |
| EPI_ISL_415614 | UW Virology Lab | UW Virology Lab |
| EPI_ISL_415615 | UW Virology Lab | UW Virology Lab |
| EPI_ISL_415616 | UW Virology Lab | UW Virology Lab |

| EPI_Accession_ID | Originating Laboratory | Submitting Laboratory |
|---|---|---|
| EPI_ISL_415617 | UW Virology Lab | UW Virology Lab |
| EPI_ISL_415619 | UW Virology Lab | UW Virology Lab |
| EPI_ISL_415620 | UW Virology Lab | UW Virology Lab |
| EPI_ISL_415621 | UW Virology Lab | UW Virology Lab |
| EPI_ISL_415622 | UW Virology Lab | UW Virology Lab |
| EPI_ISL_415623 | UW Virology Lab | UW Virology Lab |
| EPI_ISL_415624 | UW Virology Lab | UW Virology Lab |
| EPI_ISL_415625 | UW Virology Lab | UW Virology Lab |
| EPI_ISL_415626 | UW Virology Lab | UW Virology Lab |
| EPI_ISL_415627 | UW Virology Lab | UW Virology Lab |
| EPI_ISL_415628 | West of Scotland Specialist Virology Centre, NHSGGC | MRC-University of Glasgow Centre for Virus Research |
| EPI_ISL_415629 | Virology Department, Royal Infirmary of Edinburgh, NHS Lothian | Virology Department, Royal Infirmary of Edinburgh, NHS Lothian |
| EPI_ISL_415630 | West of Scotland Specialist Virology Centre, NHSGGC | MRC-University of Glasgow Centre for Virus Research |
| EPI_ISL_415631 | West of Scotland Specialist Virology Centre, NHSGGC | MRC-University of Glasgow Centre for Virus Research |
| EPI_ISL_415632 | Virology Department, Royal Infirmary of Edinburgh, NHS Lothian | Virology Department, Royal Infirmary of Edinburgh, NHS Lothian |
| EPI_ISL_415633 | Virology Department, Royal Infirmary of Edinburgh, NHS Lothian | Virology Department, Royal Infirmary of Edinburgh, NHS Lothian |
| EPI_ISL_415634 | Virology Department, Royal Infirmary of Edinburgh, NHS Lothian | Virology Department, Royal Infirmary of Edinburgh, NHS Lothian |
| EPI_ISL_415635 | Virology Department, Royal Infirmary of Edinburgh, NHS Lothian | Virology Department, Royal Infirmary of Edinburgh, NHS Lothian |
| EPI_ISL_415636 | Virology Department, Royal Infirmary of Edinburgh, NHS Lothian | Virology Department, Royal Infirmary of Edinburgh, NHS Lothian |
| EPI_ISL_415637 | Virology Department, Royal Infirmary of Edinburgh, NHS Lothian | Virology Department, Royal Infirmary of Edinburgh, NHS Lothian |
| EPI_ISL_415638 | Virology Department, Royal Infirmary of Edinburgh, NHS Lothian | Virology Department, Royal Infirmary of Edinburgh, NHS Lothian |
| EPI_ISL_415639 | Virology Department, Royal Infirmary of Edinburgh, NHS Lothian | Virology Department, Royal Infirmary of Edinburgh, NHS Lothian |
| EPI_ISL_415640 | Virology Department, Royal Infirmary of Edinburgh, NHS Lothian | Virology Department, Royal Infirmary of Edinburgh, NHS Lothian |

| EPI_Accession_ID | Originating Laboratory | Submitting Laboratory |
|---|---|---|
| EPI_ISL_415641 | R. G. Lugar Center for Public Health Research, National Center for Disease Control and Public Health (NCDC) of Georgia. | R. G. Lugar Center for Public Health Research, National Center for Disease Control and Public Health (NCDC) of Georgia. |
| EPI_ISL_415642 | R. G. Lugar Center for Public Health Research, National Center for Disease Control and Public Health (NCDC) of Georgia. | R. G. Lugar Center for Public Health Research, National Center for Disease Control and Public Health (NCDC) of Georgia. |
| EPI_ISL_415643 | R. G. Lugar Center for Public Health Research, National Center for Disease Control and Public Health (NCDC) of Georgia. | R. G. Lugar Center for Public Health Research, National Center for Disease Control and Public Health (NCDC) of Georgia. |
| EPI_ISL_415644 | R. G. Lugar Center for Public Health Research, National Center for Disease Control and Public Health (NCDC) of Georgia. | R. G. Lugar Center for Public Health Research, National Center for Disease Control and Public Health (NCDC) of Georgia. |
| EPI_ISL_415646 | Department of Virus and Microbiological Special diagnostics, Statens Serum Institut, Copenhagen, Denmark. | ViFU |
| EPI_ISL_415647 | Department of Virus and Microbiological Special diagnostics, Statens Serum Institut, Copenhagen, Denmark. | Statens Serum Institute |
| EPI_ISL_415648 | Department of Virus and Microbiological Special diagnostics, Statens Serum Institut, Copenhagen, Denmark. | ViFU |
| EPI_ISL_415649 | unknown | National Reference Center for Viruses of Respiratory Infections, Institut Pasteur, Paris |
| EPI_ISL_415650 | HÃ´pital Instruction des ArmÃ©es - BEGIN | National Reference Center for Viruses of Respiratory Infections, Institut Pasteur, Paris |
| EPI_ISL_415651 | Unknown | National Reference Center for Viruses of Respiratory Infections, Institut Pasteur, Paris |
| EPI_ISL_415652 | unknown | National Reference Center for Viruses of Respiratory Infections, Institut Pasteur, Paris |
| EPI_ISL_415653 | Centre Hospitalier CompiÃ¨gne Laboratoire de Biologie | National Reference Center for Viruses of Respiratory Infections, Institut Pasteur, Paris |
| EPI_ISL_415654 | Centre Hospitalier CompiÃ¨gne Laboratoire de Biologie | National Reference Center for Viruses of Respiratory Infections, Institut Pasteur, Paris |

| EPI_Accession_ID | Originating Laboratory | Submitting Laboratory |
|---|---|---|
| EPI_ISL_415655 | Wales Specialist Virology Centre | Public Health Wales Microbiology Cardiff |
| EPI_ISL_415656 | Wales Specialist Virology Centre | Public Health Wales Microbiology Cardiff |
| EPI_ISL_415657 | Wales Specialist Virology Centre | Public Health Wales Microbiology Cardiff |
| EPI_ISL_415658 | Laboratory of Molecular Virology, Pontificia Universidad CatÃ³lica de Chile | MSHS Pathogen Surveillance Program |
| EPI_ISL_415659 | Wales Specialist Virology Centre | Public Health Wales Microbiology Cardiff |
| EPI_ISL_415660 | Laboratory of Molecular Virology, Pontificia Universidad CatÃ³lica de Chile | MSHS Pathogen Surveillance Program |
| EPI_ISL_415661 | Laboratory of Molecular Virology, Pontificia Universidad CatÃ³lica de Chile | MSHS Pathogen Surveillance Program |
| EPI_ISL_415698 | HÃ´pitaux universitaires de GenÃ¨ve Laboratoire de Virologie | HÃ´pitaux universitaires de GenÃ¨ve Laboratoire de Virologie |
| EPI_ISL_415699 | HÃ´pitaux universitaires de GenÃ¨ve Laboratoire de Virologie | HÃ´pitaux universitaires de GenÃ¨ve Laboratoire de Virologie |
| EPI_ISL_415700 | HÃ´pitaux universitaires de GenÃ¨ve Laboratoire de Virologie | HÃ´pitaux universitaires de GenÃ¨ve Laboratoire de Virologie |
| EPI_ISL_415701 | HÃ´pitaux universitaires de GenÃ¨ve Laboratoire de Virologie | HÃ´pitaux universitaires de GenÃ¨ve Laboratoire de Virologie |
| EPI_ISL_415702 | HÃ´pitaux universitaires de GenÃ¨ve Laboratoire de Virologie | HÃ´pitaux universitaires de GenÃ¨ve Laboratoire de Virologie |
| EPI_ISL_415703 | HÃ´pitaux universitaires de GenÃ¨ve Laboratoire de Virologie | HÃ´pitaux universitaires de GenÃ¨ve Laboratoire de Virologie |
| EPI_ISL_415704 | HÃ´pitaux universitaires de GenÃ¨ve Laboratoire de Virologie | HÃ´pitaux universitaires de GenÃ¨ve Laboratoire de Virologie |
| EPI_ISL_415705 | HÃ´pitaux universitaires de GenÃ¨ve Laboratoire de Virologie | HÃ´pitaux universitaires de GenÃ¨ve Laboratoire de Virologie |
| EPI_ISL_415706 | HÃ´pitaux universitaires de GenÃ¨ve Laboratoire de Virologie | HÃ´pitaux universitaires de GenÃ¨ve Laboratoire de Virologie |
| EPI_ISL_415707 | HÃ´pitaux universitaires de GenÃ¨ve Laboratoire de Virologie | HÃ´pitaux universitaires de GenÃ¨ve Laboratoire de Virologie |
| EPI_ISL_415708 | HÃ´pitaux universitaires de GenÃ¨ve Laboratoire de Virologie | HÃ´pitaux universitaires de GenÃ¨ve Laboratoire de Virologie |

| EPI_Accession_ID | Originating Laboratory | Submitting Laboratory |
|---|---|---|
| EPI_ISL_415709 | State Key Laboratory for Diagnosis and Treatment of Infectious Diseases, National Clinical Research Center for Infectious Diseases, First Affiliated Hospital, Zhejiang University School of Medicine, Hangzhou, China. 310003 | State Key Laboratory for Diagnosis and Treatment of Infectious Diseases, National Clinical Research Center for Infectious Diseases, First Affiliated Hospital, Zhejiang University School of Medicine, Hangzhou, China. 310003 |
| EPI_ISL_415710 | WHO National Influenza Centre Russian Federation | WHO National Influenza Centre Russian Federation |
| EPI_ISL_415711 | State Key Laboratory for Diagnosis and Treatment of Infectious Diseases, National Clinical Research Center for Infectious Diseases, First Affiliated Hospital, Zhejiang University School of Medicine, Hangzhou, China. 310003 | State Key Laboratory for Diagnosis and Treatment of Infectious Diseases, National Clinical Research Center for Infectious Diseases, First Affiliated Hospital, Zhejiang University School of Medicine, Hangzhou, China. 310003 |
| EPI_ISL_415741 | Laboratory Medicine | Department of Laboratory Medicine, Lin-Kou Chang Gung Memorial Hospital, Taoyuan, Taiwan |
| EPI_ISL_415742 | Laboratory Medicine | Department of Laboratory Medicine, Lin-Kou Chang Gung Memorial Hospital, Taoyuan, Taiwan |
| EPI_ISL_415743 | Laboratory Medicine | Department of Laboratory Medicine, Lin-Kou Chang Gung Memorial Hospital, Taoyuan, Taiwan |
| EPI_ISL_415787 | Laboratorio de Referencia Nacional de Virus Respiratorio. Instituto Nacional de Salud. Peru | Laboratorio de Referencia Nacional de Biotecnologia y Biologia Molecular.Instituto Nacional de Salud.Peru |
| EPI_ISL_415916 | Wales Specialist Virology Centre | Public Health Wales Microbiology Cardiff |
| EPI_ISL_415918 | Wales Specialist Virology Centre | Public Health Wales Microbiology Cardiff |
| EPI_ISL_415919 | Wales Specialist Virology Centre | Public Health Wales Microbiology Cardiff |
| EPI_ISL_415920 | Wales Specialist Virology Centre | Public Health Wales Microbiology Cardiff |
| EPI_ISL_415977 | Wales Specialist Virology Centre | Public Health Wales Microbiology Cardiff |
| EPI_ISL_415978 | Wales Specialist Virology Centre | Public Health Wales Microbiology Cardiff |
| EPI_ISL_415991 | Wales Specialist Virology Centre | Public Health Wales Microbiology Cardiff |
| EPI_ISL_416024 | Wales Specialist Virology Centre | Public Health Wales Microbiology Cardiff |
| EPI_ISL_416026 | Wales Specialist Virology Centre | Public Health Wales Microbiology Cardiff |
| EPI_ISL_416027 | Wales Specialist Virology Centre | Public Health Wales Microbiology Cardiff |
| EPI_ISL_416028 | National Influenza Center - Instituto Adolfo Lutz | Instituto Adolfo Lutz, Interdiciplinary Procedures Center, Strategic Laboratory |
| EPI_ISL_416029 | LaboiratÃ³rio Fleury | Instituto Adolfo Lutz, Interdiciplinary Procedures Center, Strategic Laboratory |

| EPI_Accession_ID | Originating Laboratory | Submitting Laboratory |
|---|---|---|
| EPI_ISL_416031 | National Influenza Center - Instituto Adolfo Lutz | Instituto Adolfo Lutz, Interdiciplinary Procedures Center, Strategic Laboratory |
| EPI_ISL_416032 | National Influenza Center - Instituto Adolfo Lutz | Instituto Adolfo Lutz, Interdiciplinary Procedures Center, Strategic Laboratory |
| EPI_ISL_416033 | Hospital Israelita Albert Einstein | Instituto Adolfo Lutz, Interdiciplinary Procedures Center, Strategic Laboratory |
| EPI_ISL_416034 | Hospital Israelita Albert Einstein | Instituto Adolfo Lutz, Interdiciplinary Procedures Center, Strategic Laboratory |
| EPI_ISL_416035 | National Influenza Center - Instituto Adolfo Lutz | Instituto Adolfo Lutz, Interdiciplinary Procedures Center, Strategic Laboratory |
| EPI_ISL_416036 | National Influenza Center - Instituto Adolfo Lutz | Instituto Adolfo Lutz, Interdiciplinary Procedures Center, Strategic Laboratory |
| EPI_ISL_416042 | State Key Laboratory for Diagnosis and Treatment of Infectious Diseases, National Clinical Research Center for Infectious Diseases, First Affiliated Hospital, Zhejiang University School of Medicine, Hangzhou, China. 310003 | State Key Laboratory for Diagnosis and Treatment of Infectious Diseases, National Clinical Research Center for Infectious Diseases, First Affiliated Hospital, Zhejiang University School of Medicine, Hangzhou, China. 310003 |
| EPI_ISL_416044 | State Key Laboratory for Diagnosis and Treatment of Infectious Diseases, National Clinical Research Center for Infectious Diseases, First Affiliated Hospital, Zhejiang University School of Medicine, Hangzhou, China 310003 | State Key Laboratory for Diagnosis and Treatment of Infectious Diseases, National Clinical Research Center for Infectious Diseases, First Affiliated Hospital, Zhejiang University School of Medicine, Hangzhou, China 310003 |
| EPI_ISL_416046 | State Key Laboratory for Diagnosis and Treatment of Infectious Diseases, National Clinical Research Center for Infectious Diseases, First Affiliated Hospital, Zhejiang University School of Medicine, Hangzhou, China 310003 | State Key Laboratory for Diagnosis and Treatment of Infectious Diseases, National Clinical Research Center for Infectious Diseases, First Affiliated Hospital, Zhejiang University School of Medicine, Hangzhou, China 310003 |
| EPI_ISL_416047 | State Key Laboratory for Diagnosis and Treatment of Infectious Diseases, National Clinical Research Center for Infectious Diseases, First Affiliated Hospital, Zhejiang University School of Medicine, Hangzhou, China 310003 | State Key Laboratory for Diagnosis and Treatment of Infectious Diseases, National Clinical Research Center for Infectious Diseases, First Affiliated Hospital, Zhejiang University School of Medicine, Hangzhou, China 310003 |

| EPI_Accession_ID | Originating Laboratory | Submitting Laboratory |
|---|---|---|
| EPI_ISL_416140 | Department of Virus and Microbiological Special diagnostics, Statens Serum Institut, Copenhagen, Denmark. | Statens Serum Institute |
| EPI_ISL_416141 | Department of Virus and Microbiological Special diagnostics, Statens Serum Institut, Copenhagen, Denmark. | Statens Serum Institute |
| EPI_ISL_416142 | Department of Virus and Microbiological Special diagnostics, Statens Serum Institut, Copenhagen, Denmark. | Statens Serum Institute |
| EPI_ISL_416143 | Department of Virus and Microbiological Special diagnostics, Statens Serum Institut, Copenhagen, Denmark. | ViFU |
| EPI_ISL_416144 | Department of Virus and Microbiological Special diagnostics, Statens Serum Institut, Copenhagen, Denmark. | ViFU |
| EPI_ISL_416153 | Department of Virus and Microbiological Special diagnostics, Statens Serum Institut, Copenhagen, Denmark. | ViFU |
| EPI_ISL_416314 | Department of Microbiology, Faculty of Medicine, The Chinese University of Hong Kong, Hong Kong SAR, China | Department of Microbiology, Faculty of Medicine, Chinese University of Hong Kong, Hong Kong SAR, China |
| EPI_ISL_416316 | Shanghai Public Health Clinical Center, Shanghai Medical College, Fudan University | National Research Center for Translational Medicine (Shanghai), Ruijin Hospital affiliated to Shanghai Jiao Tong University School of Medicine & Shanghai Public Health Clinical Center |
| EPI_ISL_416317 | Shanghai Public Health Clinical Center, Shanghai Medical College, Fudan University | National Research Center for Translational Medicine (Shanghai), Ruijin Hospital affiliated to Shanghai Jiao Tong University School of Medicine & Shanghai Public Health Clinical Center |
| EPI_ISL_416318 | Shanghai Public Health Clinical Center, Shanghai Medical College, Fudan University | National Research Center for Translational Medicine (Shanghai), Ruijin Hospital affiliated to Shanghai Jiao Tong University School of Medicine & Shanghai Public Health Clinical Center |

| EPI_Accession_ID | Originating Laboratory | Submitting Laboratory |
|---|---|---|
| EPI_ISL_416319 | Shanghai Public Health Clinical Center, Shanghai Medical College, Fudan University | National Research Center for Translational Medicine (Shanghai), Ruijin Hospital affiliated to Shanghai Jiao Tong University School of Medicine & Shanghai Public Health Clinical Center |
| EPI_ISL_416320 | Shanghai Public Health Clinical Center, Shanghai Medical College, Fudan University | National Research Center for Translational Medicine (Shanghai), Ruijin Hospital affiliated to Shanghai Jiao Tong University School of Medicine & Shanghai Public Health Clinical Center |
| EPI_ISL_416321 | Shanghai Public Health Clinical Center, Shanghai Medical College, Fudan University | National Research Center for Translational Medicine (Shanghai), Ruijin Hospital affiliated to Shanghai Jiao Tong University School of Medicine & Shanghai Public Health Clinical Center |
| EPI_ISL_416322 | Shanghai Public Health Clinical Center, Shanghai Medical College, Fudan University | National Research Center for Translational Medicine (Shanghai), Ruijin Hospital affiliated to Shanghai Jiao Tong University School of Medicine & Shanghai Public Health Clinical Center |
| EPI_ISL_416323 | Shanghai Public Health Clinical Center, Shanghai Medical College, Fudan University | National Research Center for Translational Medicine (Shanghai), Ruijin Hospital affiliated to Shanghai Jiao Tong University School of Medicine & Shanghai Public Health Clinical Center |
| EPI_ISL_416324 | Shanghai Public Health Clinical Center, Shanghai Medical College, Fudan University | National Research Center for Translational Medicine (Shanghai), Ruijin Hospital affiliated to Shanghai Jiao Tong University School of Medicine & Shanghai Public Health Clinical Center |
| EPI_ISL_416325 | Shanghai Public Health Clinical Center, Shanghai Medical College, Fudan University | National Research Center for Translational Medicine (Shanghai), Ruijin Hospital affiliated to Shanghai Jiao Tong University School of Medicine & Shanghai Public Health Clinical Center |
| EPI_ISL_416326 | Shanghai Public Health Clinical Center, Shanghai Medical College, Fudan University | National Research Center for Translational Medicine (Shanghai), Ruijin Hospital affiliated to Shanghai Jiao Tong University School of Medicine & Shanghai Public Health Clinical Center |

| EPI_Accession_ID | Originating Laboratory | Submitting Laboratory |
|---|---|---|
| EPI_ISL_416327 | Shanghai Public Health Clinical Center, Shanghai Medical College, Fudan University | National Research Center for Translational Medicine (Shanghai), Ruijin Hospital affiliated to Shanghai Jiao Tong University School of Medicine & Shanghai Public Health Clinical Center |
| EPI_ISL_416328 | Shanghai Public Health Clinical Center, Shanghai Medical College, Fudan University | National Research Center for Translational Medicine (Shanghai), Ruijin Hospital affiliated to Shanghai Jiao Tong University School of Medicine & Shanghai Public Health Clinical Center |
| EPI_ISL_416329 | Shanghai Public Health Clinical Center, Shanghai Medical College, Fudan University | National Research Center for Translational Medicine (Shanghai), Ruijin Hospital affiliated to Shanghai Jiao Tong University School of Medicine & Shanghai Public Health Clinical Center |
| EPI_ISL_416330 | Shanghai Public Health Clinical Center, Shanghai Medical College, Fudan University | National Research Center for Translational Medicine (Shanghai), Ruijin Hospital affiliated to Shanghai Jiao Tong University School of Medicine & Shanghai Public Health Clinical Center |
| EPI_ISL_416331 | Shanghai Public Health Clinical Center, Shanghai Medical College, Fudan University | National Research Center for Translational Medicine (Shanghai), Ruijin Hospital affiliated to Shanghai Jiao Tong University School of Medicine & Shanghai Public Health Clinical Center |
| EPI_ISL_416332 | Shanghai Public Health Clinical Center, Shanghai Medical College, Fudan University | National Research Center for Translational Medicine (Shanghai), Ruijin Hospital affiliated to Shanghai Jiao Tong University School of Medicine & Shanghai Public Health Clinical Center |
| EPI_ISL_416333 | Shanghai Public Health Clinical Center, Shanghai Medical College, Fudan University | National Research Center for Translational Medicine (Shanghai), Ruijin Hospital affiliated to Shanghai Jiao Tong University School of Medicine & Shanghai Public Health Clinical Center |
| EPI_ISL_416334 | Shanghai Public Health Clinical Center, Shanghai Medical College, Fudan University | National Research Center for Translational Medicine (Shanghai), Ruijin Hospital affiliated to Shanghai Jiao Tong University School of Medicine & Shanghai Public Health Clinical Center |

| EPI_Accession_ID | Originating Laboratory | Submitting Laboratory |
|---|---|---|
| EPI_ISL_416335 | Shanghai Public Health Clinical Center, Shanghai Medical College, Fudan University | National Research Center for Translational Medicine (Shanghai), Ruijin Hospital affiliated to Shanghai Jiao Tong University School of Medicine & Shanghai Public Health Clinical Center |
| EPI_ISL_416336 | Shanghai Public Health Clinical Center, Shanghai Medical College, Fudan University | National Research Center for Translational Medicine (Shanghai), Ruijin Hospital affiliated to Shanghai Jiao Tong University School of Medicine & Shanghai Public Health Clinical Center |
| EPI_ISL_416337 | Shanghai Public Health Clinical Center, Shanghai Medical College, Fudan University | National Research Center for Translational Medicine (Shanghai), Ruijin Hospital affiliated to Shanghai Jiao Tong University School of Medicine & Shanghai Public Health Clinical Center |
| EPI_ISL_416338 | Shanghai Public Health Clinical Center, Shanghai Medical College, Fudan University | National Research Center for Translational Medicine (Shanghai), Ruijin Hospital affiliated to Shanghai Jiao Tong University School of Medicine & Shanghai Public Health Clinical Center |
| EPI_ISL_416339 | Shanghai Public Health Clinical Center, Shanghai Medical College, Fudan University | National Research Center for Translational Medicine (Shanghai), Ruijin Hospital affiliated to Shanghai Jiao Tong University School of Medicine & Shanghai Public Health Clinical Center |
| EPI_ISL_416340 | Shanghai Public Health Clinical Center, Shanghai Medical College, Fudan University | National Research Center for Translational Medicine (Shanghai), Ruijin Hospital affiliated to Shanghai Jiao Tong University School of Medicine & Shanghai Public Health Clinical Center |
| EPI_ISL_416341 | Shanghai Public Health Clinical Center, Shanghai Medical College, Fudan University | National Research Center for Translational Medicine (Shanghai), Ruijin Hospital affiliated to Shanghai Jiao Tong University School of Medicine & Shanghai Public Health Clinical Center |
| EPI_ISL_416342 | Shanghai Public Health Clinical Center, Shanghai Medical College, Fudan University | National Research Center for Translational Medicine (Shanghai), Ruijin Hospital affiliated to Shanghai Jiao Tong University School of Medicine & Shanghai Public Health Clinical Center |

| EPI_Accession_ID | Originating Laboratory | Submitting Laboratory |
|---|---|---|
| EPI_ISL_416343 | Shanghai Public Health Clinical Center, Shanghai Medical College, Fudan University | National Research Center for Translational Medicine (Shanghai), Ruijin Hospital affiliated to Shanghai Jiao Tong University School of Medicine & Shanghai Public Health Clinical Center |
| EPI_ISL_416344 | Shanghai Public Health Clinical Center, Shanghai Medical College, Fudan University | National Research Center for Translational Medicine (Shanghai), Ruijin Hospital affiliated to Shanghai Jiao Tong University School of Medicine & Shanghai Public Health Clinical Center |
| EPI_ISL_416345 | Shanghai Public Health Clinical Center, Shanghai Medical College, Fudan University | National Research Center for Translational Medicine (Shanghai), Ruijin Hospital affiliated to Shanghai Jiao Tong University School of Medicine & Shanghai Public Health Clinical Center |
| EPI_ISL_416346 | Shanghai Public Health Clinical Center, Shanghai Medical College, Fudan University | National Research Center for Translational Medicine (Shanghai), Ruijin Hospital affiliated to Shanghai Jiao Tong University School of Medicine & Shanghai Public Health Clinical Center |
| EPI_ISL_416347 | Shanghai Public Health Clinical Center, Shanghai Medical College, Fudan University | National Research Center for Translational Medicine (Shanghai), Ruijin Hospital affiliated to Shanghai Jiao Tong University School of Medicine & Shanghai Public Health Clinical Center |
| EPI_ISL_416348 | Shanghai Public Health Clinical Center, Shanghai Medical College, Fudan University | National Research Center for Translational Medicine (Shanghai), Ruijin Hospital affiliated to Shanghai Jiao Tong University School of Medicine & Shanghai Public Health Clinical Center |
| EPI_ISL_416349 | Shanghai Public Health Clinical Center, Shanghai Medical College, Fudan University | National Research Center for Translational Medicine (Shanghai), Ruijin Hospital affiliated to Shanghai Jiao Tong University School of Medicine & Shanghai Public Health Clinical Center |
| EPI_ISL_416350 | Shanghai Public Health Clinical Center, Shanghai Medical College, Fudan University | National Research Center for Translational Medicine (Shanghai), Ruijin Hospital affiliated to Shanghai Jiao Tong University School of Medicine & Shanghai Public Health Clinical Center |

| EPI_Accession_ID | Originating Laboratory | Submitting Laboratory |
| --- | --- | --- |
| EPI_ISL_416351 | Shanghai Public Health Clinical Center, Shanghai Medical College, Fudan University | National Research Center for Translational Medicine (Shanghai), Ruijin Hospital affiliated to Shanghai Jiao Tong University School of Medicine & Shanghai Public Health Clinical Center |
| EPI_ISL_416352 | Shanghai Public Health Clinical Center, Shanghai Medical College, Fudan University | National Research Center for Translational Medicine (Shanghai), Ruijin Hospital affiliated to Shanghai Jiao Tong University School of Medicine & Shanghai Public Health Clinical Center |
| EPI_ISL_416353 | Shanghai Public Health Clinical Center, Shanghai Medical College, Fudan University | National Research Center for Translational Medicine (Shanghai), Ruijin Hospital affiliated to Shanghai Jiao Tong University School of Medicine & Shanghai Public Health Clinical Center |
| EPI_ISL_416354 | Shanghai Public Health Clinical Center, Shanghai Medical College, Fudan University | National Research Center for Translational Medicine (Shanghai), Ruijin Hospital affiliated to Shanghai Jiao Tong University School of Medicine & Shanghai Public Health Clinical Center |
| EPI_ISL_416355 | Shanghai Public Health Clinical Center, Shanghai Medical College, Fudan University | National Research Center for Translational Medicine (Shanghai), Ruijin Hospital affiliated to Shanghai Jiao Tong University School of Medicine & Shanghai Public Health Clinical Center |
| EPI_ISL_416356 | Shanghai Public Health Clinical Center, Shanghai Medical College, Fudan University | National Research Center for Translational Medicine (Shanghai), Ruijin Hospital affiliated to Shanghai Jiao Tong University School of Medicine & Shanghai Public Health Clinical Center |
| EPI_ISL_416357 | Shanghai Public Health Clinical Center, Shanghai Medical College, Fudan University | National Research Center for Translational Medicine (Shanghai), Ruijin Hospital affiliated to Shanghai Jiao Tong University School of Medicine & Shanghai Public Health Clinical Center |
| EPI_ISL_416358 | Shanghai Public Health Clinical Center, Shanghai Medical College, Fudan University | National Research Center for Translational Medicine (Shanghai), Ruijin Hospital affiliated to Shanghai Jiao Tong University School of Medicine & Shanghai Public Health Clinical Center |

| EPI_Accession_ID | Originating Laboratory | Submitting Laboratory |
|---|---|---|
| EPI_ISL_416359 | Shanghai Public Health Clinical Center, Shanghai Medical College, Fudan University | National Research Center for Translational Medicine (Shanghai), Ruijin Hospital affiliated to Shanghai Jiao Tong University School of Medicine & Shanghai Public Health Clinical Center |
| EPI_ISL_416360 | Shanghai Public Health Clinical Center, Shanghai Medical College, Fudan University | National Research Center for Translational Medicine (Shanghai), Ruijin Hospital affiliated to Shanghai Jiao Tong University School of Medicine & Shanghai Public Health Clinical Center |
| EPI_ISL_416361 | Shanghai Public Health Clinical Center, Shanghai Medical College, Fudan University | National Research Center for Translational Medicine (Shanghai), Ruijin Hospital affiliated to Shanghai Jiao Tong University School of Medicine & Shanghai Public Health Clinical Center |
| EPI_ISL_416362 | Shanghai Public Health Clinical Center, Shanghai Medical College, Fudan University | National Research Center for Translational Medicine (Shanghai), Ruijin Hospital affiliated to Shanghai Jiao Tong University School of Medicine & Shanghai Public Health Clinical Center |
| EPI_ISL_416363 | Shanghai Public Health Clinical Center, Shanghai Medical College, Fudan University | National Research Center for Translational Medicine (Shanghai), Ruijin Hospital affiliated to Shanghai Jiao Tong University School of Medicine & Shanghai Public Health Clinical Center |
| EPI_ISL_416364 | Shanghai Public Health Clinical Center, Shanghai Medical College, Fudan University | National Research Center for Translational Medicine (Shanghai), Ruijin Hospital affiliated to Shanghai Jiao Tong University School of Medicine & Shanghai Public Health Clinical Center |
| EPI_ISL_416365 | Shanghai Public Health Clinical Center, Shanghai Medical College, Fudan University | National Research Center for Translational Medicine (Shanghai), Ruijin Hospital affiliated to Shanghai Jiao Tong University School of Medicine & Shanghai Public Health Clinical Center |
| EPI_ISL_416366 | Shanghai Public Health Clinical Center, Shanghai Medical College, Fudan University | National Research Center for Translational Medicine (Shanghai), Ruijin Hospital affiliated to Shanghai Jiao Tong University School of Medicine & Shanghai Public Health Clinical Center |

| EPI_Accession_ID | Originating Laboratory | Submitting Laboratory |
| --- | --- | --- |
| EPI_ISL_416367 | Shanghai Public Health Clinical Center, Shanghai Medical College, Fudan University | National Research Center for Translational Medicine (Shanghai), Ruijin Hospital affiliated to Shanghai Jiao Tong University School of Medicine & Shanghai Public Health Clinical Center |
| EPI_ISL_416368 | Shanghai Public Health Clinical Center, Shanghai Medical College, Fudan University | National Research Center for Translational Medicine (Shanghai), Ruijin Hospital affiliated to Shanghai Jiao Tong University School of Medicine & Shanghai Public Health Clinical Center |
| EPI_ISL_416369 | Shanghai Public Health Clinical Center, Shanghai Medical College, Fudan University | National Research Center for Translational Medicine (Shanghai), Ruijin Hospital affiliated to Shanghai Jiao Tong University School of Medicine & Shanghai Public Health Clinical Center |
| EPI_ISL_416370 | Shanghai Public Health Clinical Center, Shanghai Medical College, Fudan University | National Research Center for Translational Medicine (Shanghai), Ruijin Hospital affiliated to Shanghai Jiao Tong University School of Medicine & Shanghai Public Health Clinical Center |
| EPI_ISL_416371 | Shanghai Public Health Clinical Center, Shanghai Medical College, Fudan University | National Research Center for Translational Medicine (Shanghai), Ruijin Hospital affiliated to Shanghai Jiao Tong University School of Medicine & Shanghai Public Health Clinical Center |
| EPI_ISL_416372 | Shanghai Public Health Clinical Center, Shanghai Medical College, Fudan University | National Research Center for Translational Medicine (Shanghai), Ruijin Hospital affiliated to Shanghai Jiao Tong University School of Medicine & Shanghai Public Health Clinical Center |
| EPI_ISL_416373 | Shanghai Public Health Clinical Center, Shanghai Medical College, Fudan University | National Research Center for Translational Medicine (Shanghai), Ruijin Hospital affiliated to Shanghai Jiao Tong University School of Medicine & Shanghai Public Health Clinical Center |
| EPI_ISL_416374 | Shanghai Public Health Clinical Center, Shanghai Medical College, Fudan University | National Research Center for Translational Medicine (Shanghai), Ruijin Hospital affiliated to Shanghai Jiao Tong University School of Medicine & Shanghai Public Health Clinical Center |

| EPI_Accession_ID | Originating Laboratory | Submitting Laboratory |
|---|---|---|
| EPI_ISL_416375 | Shanghai Public Health Clinical Center, Shanghai Medical College, Fudan University | National Research Center for Translational Medicine (Shanghai), Ruijin Hospital affiliated to Shanghai Jiao Tong University School of Medicine & Shanghai Public Health Clinical Center |
| EPI_ISL_416376 | Shanghai Public Health Clinical Center, Shanghai Medical College, Fudan University | National Research Center for Translational Medicine (Shanghai), Ruijin Hospital affiliated to Shanghai Jiao Tong University School of Medicine & Shanghai Public Health Clinical Center |
| EPI_ISL_416377 | Shanghai Public Health Clinical Center, Shanghai Medical College, Fudan University | National Research Center for Translational Medicine (Shanghai), Ruijin Hospital affiliated to Shanghai Jiao Tong University School of Medicine & Shanghai Public Health Clinical Center |
| EPI_ISL_416378 | Shanghai Public Health Clinical Center, Shanghai Medical College, Fudan University | National Research Center for Translational Medicine (Shanghai), Ruijin Hospital affiliated to Shanghai Jiao Tong University School of Medicine & Shanghai Public Health Clinical Center |
| EPI_ISL_416379 | Shanghai Public Health Clinical Center, Shanghai Medical College, Fudan University | National Research Center for Translational Medicine (Shanghai), Ruijin Hospital affiliated to Shanghai Jiao Tong University School of Medicine & Shanghai Public Health Clinical Center |
| EPI_ISL_416380 | Shanghai Public Health Clinical Center, Shanghai Medical College, Fudan University | National Research Center for Translational Medicine (Shanghai), Ruijin Hospital affiliated to Shanghai Jiao Tong University School of Medicine & Shanghai Public Health Clinical Center |
| EPI_ISL_416381 | Shanghai Public Health Clinical Center, Shanghai Medical College, Fudan University | National Research Center for Translational Medicine (Shanghai), Ruijin Hospital affiliated to Shanghai Jiao Tong University School of Medicine & Shanghai Public Health Clinical Center |
| EPI_ISL_416382 | Shanghai Public Health Clinical Center, Shanghai Medical College, Fudan University | National Research Center for Translational Medicine (Shanghai), Ruijin Hospital affiliated to Shanghai Jiao Tong University School of Medicine & Shanghai Public Health Clinical Center |

| EPI_Accession_ID | Originating Laboratory | Submitting Laboratory |
|---|---|---|
| EPI_ISL_416383 | Shanghai Public Health Clinical Center, Shanghai Medical College, Fudan University | National Research Center for Translational Medicine (Shanghai), Ruijin Hospital affiliated to Shanghai Jiao Tong University School of Medicine & Shanghai Public Health Clinical Center |
| EPI_ISL_416384 | Shanghai Public Health Clinical Center, Shanghai Medical College, Fudan University | National Research Center for Translational Medicine (Shanghai), Ruijin Hospital affiliated to Shanghai Jiao Tong University School of Medicine & Shanghai Public Health Clinical Center |
| EPI_ISL_416385 | Shanghai Public Health Clinical Center, Shanghai Medical College, Fudan University | National Research Center for Translational Medicine (Shanghai), Ruijin Hospital affiliated to Shanghai Jiao Tong University School of Medicine & Shanghai Public Health Clinical Center |
| EPI_ISL_416386 | Shanghai Public Health Clinical Center, Shanghai Medical College, Fudan University | National Research Center for Translational Medicine (Shanghai), Ruijin Hospital affiliated to Shanghai Jiao Tong University School of Medicine & Shanghai Public Health Clinical Center |
| EPI_ISL_416387 | Shanghai Public Health Clinical Center, Shanghai Medical College, Fudan University | National Research Center for Translational Medicine (Shanghai), Ruijin Hospital affiliated to Shanghai Jiao Tong University School of Medicine & Shanghai Public Health Clinical Center |
| EPI_ISL_416388 | Shanghai Public Health Clinical Center, Shanghai Medical College, Fudan University | National Research Center for Translational Medicine (Shanghai), Ruijin Hospital affiliated to Shanghai Jiao Tong University School of Medicine & Shanghai Public Health Clinical Center |
| EPI_ISL_416389 | Shanghai Public Health Clinical Center, Shanghai Medical College, Fudan University | National Research Center for Translational Medicine (Shanghai), Ruijin Hospital affiliated to Shanghai Jiao Tong University School of Medicine & Shanghai Public Health Clinical Center |
| EPI_ISL_416390 | Shanghai Public Health Clinical Center, Shanghai Medical College, Fudan University | National Research Center for Translational Medicine (Shanghai), Ruijin Hospital affiliated to Shanghai Jiao Tong University School of Medicine & Shanghai Public Health Clinical Center |

| EPI_Accession_ID | Originating Laboratory | Submitting Laboratory |
|---|---|---|
| EPI_ISL_416391 | Shanghai Public Health Clinical Center, Shanghai Medical College, Fudan University | National Research Center for Translational Medicine (Shanghai), Ruijin Hospital affiliated to Shanghai Jiao Tong University School of Medicine & Shanghai Public Health Clinical Center |
| EPI_ISL_416392 | Shanghai Public Health Clinical Center, Shanghai Medical College, Fudan University | National Research Center for Translational Medicine (Shanghai), Ruijin Hospital affiliated to Shanghai Jiao Tong University School of Medicine & Shanghai Public Health Clinical Center |
| EPI_ISL_416393 | Shanghai Public Health Clinical Center, Shanghai Medical College, Fudan University | National Research Center for Translational Medicine (Shanghai), Ruijin Hospital affiliated to Shanghai Jiao Tong University School of Medicine & Shanghai Public Health Clinical Center |
| EPI_ISL_416394 | Shanghai Public Health Clinical Center, Shanghai Medical College, Fudan University | National Research Center for Translational Medicine (Shanghai), Ruijin Hospital affiliated to Shanghai Jiao Tong University School of Medicine & Shanghai Public Health Clinical Center |
| EPI_ISL_416395 | Shanghai Public Health Clinical Center, Shanghai Medical College, Fudan University | National Research Center for Translational Medicine (Shanghai), Ruijin Hospital affiliated to Shanghai Jiao Tong University School of Medicine & Shanghai Public Health Clinical Center |
| EPI_ISL_416396 | Shanghai Public Health Clinical Center, Shanghai Medical College, Fudan University | National Research Center for Translational Medicine (Shanghai), Ruijin Hospital affiliated to Shanghai Jiao Tong University School of Medicine & Shanghai Public Health Clinical Center |
| EPI_ISL_416397 | Shanghai Public Health Clinical Center, Shanghai Medical College, Fudan University | National Research Center for Translational Medicine (Shanghai), Ruijin Hospital affiliated to Shanghai Jiao Tong University School of Medicine & Shanghai Public Health Clinical Center |
| EPI_ISL_416398 | Shanghai Public Health Clinical Center, Shanghai Medical College, Fudan University | National Research Center for Translational Medicine (Shanghai), Ruijin Hospital affiliated to Shanghai Jiao Tong University School of Medicine & Shanghai Public Health Clinical Center |

| EPI_Accession_ID | Originating Laboratory | Submitting Laboratory |
|---|---|---|
| EPI_ISL_416399 | Shanghai Public Health Clinical Center, Shanghai Medical College, Fudan University | National Research Center for Translational Medicine (Shanghai), Ruijin Hospital affiliated to Shanghai Jiao Tong University School of Medicine & Shanghai Public Health Clinical Center |
| EPI_ISL_416400 | Shanghai Public Health Clinical Center, Shanghai Medical College, Fudan University | National Research Center for Translational Medicine (Shanghai), Ruijin Hospital affiliated to Shanghai Jiao Tong University School of Medicine & Shanghai Public Health Clinical Center |
| EPI_ISL_416401 | Shanghai Public Health Clinical Center, Shanghai Medical College, Fudan University | National Research Center for Translational Medicine (Shanghai), Ruijin Hospital affiliated to Shanghai Jiao Tong University School of Medicine & Shanghai Public Health Clinical Center |
| EPI_ISL_416402 | Shanghai Public Health Clinical Center, Shanghai Medical College, Fudan University | National Research Center for Translational Medicine (Shanghai), Ruijin Hospital affiliated to Shanghai Jiao Tong University School of Medicine & Shanghai Public Health Clinical Center |
| EPI_ISL_416403 | Shanghai Public Health Clinical Center, Shanghai Medical College, Fudan University | National Research Center for Translational Medicine (Shanghai), Ruijin Hospital affiliated to Shanghai Jiao Tong University School of Medicine & Shanghai Public Health Clinical Center |
| EPI_ISL_416404 | Shanghai Public Health Clinical Center, Shanghai Medical College, Fudan University | National Research Center for Translational Medicine (Shanghai), Ruijin Hospital affiliated to Shanghai Jiao Tong University School of Medicine & Shanghai Public Health Clinical Center |
| EPI_ISL_416405 | Shanghai Public Health Clinical Center, Shanghai Medical College, Fudan University | National Research Center for Translational Medicine (Shanghai), Ruijin Hospital affiliated to Shanghai Jiao Tong University School of Medicine & Shanghai Public Health Clinical Center |
| EPI_ISL_416406 | Shanghai Public Health Clinical Center, Shanghai Medical College, Fudan University | National Research Center for Translational Medicine (Shanghai), Ruijin Hospital affiliated to Shanghai Jiao Tong University School of Medicine & Shanghai Public Health Clinical Center |

| EPI_Accession_ID | Originating Laboratory | Submitting Laboratory |
|---|---|---|
| EPI_ISL_416407 | Shanghai Public Health Clinical Center, Shanghai Medical College, Fudan University | National Research Center for Translational Medicine (Shanghai), Ruijin Hospital affiliated to Shanghai Jiao Tong University School of Medicine & Shanghai Public Health Clinical Center |
| EPI_ISL_416408 | Shanghai Public Health Clinical Center, Shanghai Medical College, Fudan University | National Research Center for Translational Medicine (Shanghai), Ruijin Hospital affiliated to Shanghai Jiao Tong University School of Medicine & Shanghai Public Health Clinical Center |
| EPI_ISL_416409 | Shanghai Public Health Clinical Center, Shanghai Medical College, Fudan University | National Research Center for Translational Medicine (Shanghai), Ruijin Hospital affiliated to Shanghai Jiao Tong University School of Medicine & Shanghai Public Health Clinical Center |
| EPI_ISL_416410 | Victorian Infectious Diseases Reference Laboratory (VIDRL) | Victorian Infectious Diseases Reference Laboratory and Microbiological Diagnostic Unit Public Health Laboratory, Doherty Institute |
| EPI_ISL_416411 | Victorian Infectious Diseases Reference Laboratory (VIDRL) | Victorian Infectious Diseases Reference Laboratory and Microbiological Diagnostic Unit Public Health Laboratory, Doherty Institute |
| EPI_ISL_416412 | Victorian Infectious Diseases Reference Laboratory (VIDRL) | Victorian Infectious Diseases Reference Laboratory and Microbiological Diagnostic Unit Public Health Laboratory, Doherty Institute |
| EPI_ISL_416413 | Victorian Infectious Diseases Reference Laboratory (VIDRL) | Victorian Infectious Diseases Reference Laboratory and Microbiological Diagnostic Unit Public Health Laboratory, Doherty Institute |
| EPI_ISL_416414 | Victorian Infectious Diseases Reference Laboratory (VIDRL) | Victorian Infectious Diseases Reference Laboratory and Microbiological Diagnostic Unit Public Health Laboratory, Doherty Institute |
| EPI_ISL_416415 | Victorian Infectious Diseases Reference Laboratory (VIDRL) | Victorian Infectious Diseases Reference Laboratory and Microbiological Diagnostic Unit Public Health Laboratory, Doherty Institute |
| EPI_ISL_416417 | Connecticut State Department of Public Health | Grubaugh Lab - Yale School of Public Health |
| EPI_ISL_416418 | Connecticut State Department of Public Health | Grubaugh Lab - Yale School of Public Health |
| EPI_ISL_416419 | Connecticut State Department of Public Health | Grubaugh Lab - Yale School of Public Health |

| EPI_Accession_ID | Originating Laboratory | Submitting Laboratory |
|---|---|---|
| EPI_ISL_416420 | Yale Clinical Virology Laboratory | Grubaugh Lab - Yale School of Public Health |
| EPI_ISL_416421 | Yale Clinical Virology Laboratory | Grubaugh Lab - Yale School of Public Health |
| EPI_ISL_416422 | Yale Clinical Virology Laboratory | Grubaugh Lab - Yale School of Public Health |
| EPI_ISL_416423 | Yale Clinical Virology Laboratory | Grubaugh Lab - Yale School of Public Health |
| EPI_ISL_416424 | Yale Clinical Virology Laboratory | Grubaugh Lab - Yale School of Public Health |
| EPI_ISL_416425 | State Key Laboratory for Diagnosis and Treatment of Infectious Diseases, National Clinical Research Center for Infectious Diseases, First Affiliated Hospital, Zhejiang University School of Medicine, Hangzhou, China 310003 | State Key Laboratory for Diagnosis and Treatment of Infectious Diseases, National Clinical Research Center for Infectious Diseases, First Affiliated Hospital, Zhejiang University School of Medicine, Hangzhou, China 310003 |
| EPI_ISL_416426 | Virological Research Group, SzentÃ¡gothai Research Centre, University of PÃ©cs | Bioinformatics Research Group, SzentÃ¡gothai Research Centre, University of PÃ©cs |
| EPI_ISL_416427 | National Influenza Center, National Institute of Hygiene and Epidemiology (NIHE) | National Influenza Center, National Institute of Hygiene and Epidemiology (NIHE) |
| EPI_ISL_416428 | National Influenza Center, National Institute of Hygiene and Epidemiology (NIHE) | National Influenza Center, National Institute of Hygiene and Epidemiology (NIHE) |
| EPI_ISL_416429 | National Influenza Center, National Institute of Hygiene and Epidemiology (NIHE) | National Influenza Center, National Institute of Hygiene and Epidemiology (NIHE) |
| EPI_ISL_416430 | National Influenza Center, National Institute of Hygiene and Epidemiology (NIHE) | National Influenza Center, National Institute of Hygiene and Epidemiology (NIHE) |
| EPI_ISL_416431 | National Influenza Center, National Institute of Hygiene and Epidemiology (NIHE) | National Influenza Center, National Institute of Hygiene and Epidemiology (NIHE) |
| EPI_ISL_416432 | Clinical Microbiology Lab | Infectious Disease Research Department, King Abdullah International Medical Research Center (KAIMRC) |
| EPI_ISL_416433 | UW Virology Lab | UW Virology Lab |
| EPI_ISL_416434 | UW Virology Lab | UW Virology Lab |
| EPI_ISL_416435 | UW Virology Lab | UW Virology Lab |
| EPI_ISL_416436 | UW Virology Lab | UW Virology Lab |
| EPI_ISL_416437 | UW Virology Lab | UW Virology Lab |
| EPI_ISL_416438 | UW Virology Lab | UW Virology Lab |
| EPI_ISL_416439 | UW Virology Lab | UW Virology Lab |
| EPI_ISL_416440 | UW Virology Lab | UW Virology Lab |

| EPI_Accession_ID | Originating Laboratory | Submitting Laboratory |
|---|---|---|
| EPI_ISL_416441 | UW Virology Lab | UW Virology Lab |
| EPI_ISL_416442 | UW Virology Lab | UW Virology Lab |
| EPI_ISL_416443 | UW Virology Lab | UW Virology Lab |
| EPI_ISL_416444 | UW Virology Lab | UW Virology Lab |
| EPI_ISL_416445 | UW Virology Lab | UW Virology Lab |
| EPI_ISL_416446 | UW Virology Lab | UW Virology Lab |
| EPI_ISL_416447 | UW Virology Lab | UW Virology Lab |
| EPI_ISL_416448 | UW Virology Lab | UW Virology Lab |
| EPI_ISL_416449 | UW Virology Lab | UW Virology Lab |
| EPI_ISL_416450 | UW Virology Lab | UW Virology Lab |
| EPI_ISL_416451 | UW Virology Lab | UW Virology Lab |
| EPI_ISL_416452 | UW Virology Lab | UW Virology Lab |
| EPI_ISL_416453 | UW Virology Lab | UW Virology Lab |
| EPI_ISL_416454 | UW Virology Lab | UW Virology Lab |
| EPI_ISL_416455 | UW Virology Lab | UW Virology Lab |
| EPI_ISL_416456 | UW Virology Lab | UW Virology Lab |
| EPI_ISL_416457 | Andersen Lab, The Scripps Research Institute | Andersen Lab, The Scripps Research Institute |
| EPI_ISL_416458 | Virology laboratory Ministry of Health Kuwait sequenced at Dasman Diabetes Institute | Dasman Diabetes Institute |
| EPI_ISL_416460 | Seattle Flu Study | Seattle Flu Study |
| EPI_ISL_416461 | Seattle Flu Study | Seattle Flu Study |
| EPI_ISL_416462 | Seattle Flu Study | Seattle Flu Study |
| EPI_ISL_416463 | Seattle Flu Study | Seattle Flu Study |
| EPI_ISL_416465 | Seattle Flu Study | Seattle Flu Study |
| EPI_ISL_416466 | Seattle Flu Study | Seattle Flu Study |
| EPI_ISL_416467 | KU Leuven, Clinical and Epidemiological Virology | KU Leuven, Clinical and Epidemiological Virology |
| EPI_ISL_416468 | KU Leuven, Clinical and Epidemiological Virology | KU Leuven, Clinical and Epidemiological Virology |
| EPI_ISL_416469 | KU Leuven, Clinical and Epidemiological Virology | KU Leuven, Clinical and Epidemiological Virology |
| EPI_ISL_416470 | KU Leuven, Clinical and Epidemiological Virology | KU Leuven, Clinical and Epidemiological Virology |
| EPI_ISL_416471 | KU Leuven, Clinical and Epidemiological Virology | KU Leuven, Clinical and Epidemiological Virology |
| EPI_ISL_416472 | KU Leuven, Clinical and Epidemiological Virology | KU Leuven, Clinical and Epidemiological Virology |

| EPI_Accession_ID | Originating Laboratory | Submitting Laboratory |
|---|---|---|
| EPI_ISL_416473 | State Key Laboratory for Diagnosis and Treatment of Infectious Diseases, National Clinical Research Center for Infectious Diseases, First Affiliated Hospital, Zhejiang University School of Medicine, Hangzhou, China 310003 | State Key Laboratory for Diagnosis and Treatment of Infectious Diseases, National Clinical Research Center for Infectious Diseases, First Affiliated Hospital, Zhejiang University School of Medicine, Hangzhou, China 310003 |
| EPI_ISL_416474 | State Key Laboratory for Diagnosis and Treatment of Infectious Diseases, National Clinical Research Center for Infectious Diseases, First Affiliated Hospital, Zhejiang University School of Medicine, Hangzhou, China 310003 | State Key Laboratory for Diagnosis and Treatment of Infectious Diseases, National Clinical Research Center for Infectious Diseases, First Affiliated Hospital, Zhejiang University School of Medicine, Hangzhou, China 310003 |
| EPI_ISL_416475 | KU Leuven, Clinical and Epidemiological Virology | KU Leuven, Clinical and Epidemiological Virology |
| EPI_ISL_416476 | KU Leuven, Clinical and Epidemiological Virology | KU Leuven, Clinical and Epidemiological Virology |
| EPI_ISL_416477 | R. G. Lugar Center for Public Health Research, National Center for Disease Control and Public Health (NCDC) of Georgia. | R. G. Lugar Center for Public Health Research, National Center for Disease Control and Public Health (NCDC) of Georgia. |
| EPI_ISL_416478 | R. G. Lugar Center for Public Health Research, National Center for Disease Control and Public Health (NCDC) of Georgia. | R. G. Lugar Center for Public Health Research, National Center for Disease Control and Public Health (NCDC) of Georgia. |
| EPI_ISL_416479 | R. G. Lugar Center for Public Health Research, National Center for Disease Control and Public Health (NCDC) of Georgia. | R. G. Lugar Center for Public Health Research, National Center for Disease Control and Public Health (NCDC) of Georgia. |
| EPI_ISL_416480 | R. G. Lugar Center for Public Health Research, National Center for Disease Control and Public Health (NCDC) of Georgia. | R. G. Lugar Center for Public Health Research, National Center for Disease Control and Public Health (NCDC) of Georgia. |
| EPI_ISL_416481 | R. G. Lugar Center for Public Health Research, National Center for Disease Control and Public Health (NCDC) of Georgia. | R. G. Lugar Center for Public Health Research, National Center for Disease Control and Public Health (NCDC) of Georgia. |
| EPI_ISL_416482 | R. G. Lugar Center for Public Health Research, National Center for Disease Control and Public Health (NCDC) of Georgia. | R. G. Lugar Center for Public Health Research, National Center for Disease Control and Public Health (NCDC) of Georgia. |
| EPI_ISL_416483 | Servicio de MicrobiologÃ-a. Consorcio Hospital General Universitario de Valencia | Sequencing and Bioinformatics Service and Molecular Epidemiology Research Group. FISABIO-Public Health |

| EPI_Accession_ID | Originating Laboratory | Submitting Laboratory |
|---|---|---|
| EPI_ISL_416484 | Servicio de MicrobiologÃ-a. Consorcio Hospital General Universitario de Valencia | Sequencing and Bioinformatics Service and Molecular Epidemiology Research Group. FISABIO-Public Health |
| EPI_ISL_416485 | Servicio de Microbiologia. Consorcio Hospital General Universitario de Valencia | Sequencing and Bioinformatics Service and Molecular Epidemiology Research Group. FISABIO-Public Health |
| EPI_ISL_416486 | Servicio de MicrobiologÃ-a. Consorcio Hospital General Universitario de Valencia | Sequencing and Bioinformatics Service and Molecular Epidemiology Research Group. FISABIO-Public Health |
| EPI_ISL_416487 | Servicio de MicrobiologÃ-a. Consorcio Hospital General Universitario de Valencia | Sequencing and Bioinformatics Service and Molecular Epidemiology Research Group. FISABIO-Public Health |
| EPI_ISL_416488 | ViroGenetics - BSL3 Laboratory of Virology; Human Genome Variation Research Group & Genomics Centre MCB; Bioinformatics Research Group Department of Virology | ViroGenetics - BSL3 Laboratory of Virology; Human Genome Variation Research Group & Genomics Centre MCB; Bioinformatics Research Group Department of Virology |
| EPI_ISL_416489 | University of Wisconsin-Madison AIDS Vaccine Research Laboratories | University of Wisconsin-Madison AIDS Vaccine Research Laboratories |
| EPI_ISL_416491 | University of Wisconsin-Madison AIDS Vaccine Research Laboratories | University of Wisconsin-Madison AIDS Vaccine Research Laboratories |
| EPI_ISL_416492 | University of Wisconsin-Madison AIDS Vaccine Research Laboratories | University of Wisconsin-Madison AIDS Vaccine Research Laboratories |
| EPI_ISL_416493 | CH Jean de Navarre Laboratoire de Biologie | National Reference Center for Viruses of Respiratory Infections, Institut Pasteur, Paris |
| EPI_ISL_416494 | Centre Hositalier Universitaire de Rouen Laboratoire de Virologie | National Reference Center for Viruses of Respiratory Infections, Institut Pasteur, Paris |
| EPI_ISL_416495 | Centre Hospitalier CompiÃ¨gne Laboratoire de Biologie | National Reference Center for Viruses of Respiratory Infections, Institut Pasteur, Paris |
| EPI_ISL_416496 | Centre Hospitalier CompiÃ¨gne Laboratoire de Biologie | National Reference Center for Viruses of Respiratory Infections, Institut Pasteur, Paris |
| EPI_ISL_416497 | Centre Hospitalier CompiÃ¨gne Laboratoire de Biologie | National Reference Center for Viruses of Respiratory Infections, Institut Pasteur, Paris |
| EPI_ISL_416498 | Institut MÃ©dico lÃ©gal- Hop R. PoincarÃ© | National Reference Center for Viruses of Respiratory Infections, Institut Pasteur, Paris |
| EPI_ISL_416499 | LABM GH nord Essonne | National Reference Center for Viruses of Respiratory Infections, Institut Pasteur, Paris |

| EPI_Accession_ID | Originating Laboratory | Submitting Laboratory |
|---|---|---|
| EPI_ISL_416500 | LABM GH nord Essonne | National Reference Center for Viruses of Respiratory Infections, Institut Pasteur, Paris |
| EPI_ISL_416501 | Hopital franco britannique - Service des Urgences | National Reference Center for Viruses of Respiratory Infections, Institut Pasteur, Paris |
| EPI_ISL_416502 | CHRU Pontchaillou - Laboratoire de Virologie | National Reference Center for Viruses of Respiratory Infections, Institut Pasteur, Paris |
| EPI_ISL_416503 | CHRU Pontchaillou - Laboratoire de Virologie | National Reference Center for Viruses of Respiratory Infections, Institut Pasteur, Paris |
| EPI_ISL_416504 | CHRU Pontchaillou - Laboratoire de Virologie | National Reference Center for Viruses of Respiratory Infections, Institut Pasteur, Paris |
| EPI_ISL_416505 | CHRU Pontchaillou - Laboratoire de Virologie | National Reference Center for Viruses of Respiratory Infections, Institut Pasteur, Paris |
| EPI_ISL_416506 | CHRU Pontchaillou - Laboratoire de Virologie | National Reference Center for Viruses of Respiratory Infections, Institut Pasteur, Paris |
| EPI_ISL_416507 | CHRU Pontchaillou - Laboratoire de Virologie | National Reference Center for Viruses of Respiratory Infections, Institut Pasteur, Paris |
| EPI_ISL_416508 | CHRU Pontchaillou - Laboratoire de Virologie | National Reference Center for Viruses of Respiratory Infections, Institut Pasteur, Paris |
| EPI_ISL_416509 | CHRU Pontchaillou - Laboratoire de Virologie | National Reference Center for Viruses of Respiratory Infections, Institut Pasteur, Paris |
| EPI_ISL_416510 | CHRU Pontchaillou - Laboratoire de Virologie | National Reference Center for Viruses of Respiratory Infections, Institut Pasteur, Paris |
| EPI_ISL_416511 | CHRU Pontchaillou - Laboratoire de Virologie | National Reference Center for Viruses of Respiratory Infections, Institut Pasteur, Paris |
| EPI_ISL_416512 | CHRU Pontchaillou - Laboratoire de Virologie | National Reference Center for Viruses of Respiratory Infections, Institut Pasteur, Paris |
| EPI_ISL_416513 | CHRU Pontchaillou - Laboratoire de Virologie | National Reference Center for Viruses of Respiratory Infections, Institut Pasteur, Paris |
| EPI_ISL_416514 | Victorian Infectious Diseases Reference Laboratory (VIDRL) | Victorian Infectious Diseases Reference Laboratory and Microbiological Diagnostic Unit Public Health Laboratory, Doherty Institute |

| EPI_Accession_ID | Originating Laboratory | Submitting Laboratory |
|---|---|---|
| EPI_ISL_416515 | Victorian Infectious Diseases Reference Laboratory (VIDRL) | Victorian Infectious Diseases Reference Laboratory and Microbiological Diagnostic Unit Public Health Laboratory, Doherty Institute |
| EPI_ISL_416516 | Victorian Infectious Diseases Reference Laboratory (VIDRL) | Victorian Infectious Diseases Reference Laboratory and Microbiological Diagnostic Unit Public Health Laboratory, Doherty Institute |
| EPI_ISL_416517 | Victorian Infectious Diseases Reference Laboratory (VIDRL) | Victorian Infectious Diseases Reference Laboratory and Microbiological Diagnostic Unit Public Health Laboratory, Doherty Institute |
| EPI_ISL_416518 | Victorian Infectious Diseases Reference Laboratory (VIDRL) | Victorian Infectious Diseases Reference Laboratory and Microbiological Diagnostic Unit Public Health Laboratory, Doherty Institute |
| EPI_ISL_416519 | Auckland Hospital | Institute of Environmental Science and Research (ESR) |
| EPI_ISL_416520 | Auckland Hospital | Institute of Environmental Science and Research (ESR) |
| EPI_ISL_416521 | Public Health Laboratory | Public Health Laboratory, Saudi CDC |
| EPI_ISL_416522 | Public Health Laboratory, Saudi CDC | Public Health Laboratory, Saudi CDC |
| EPI_ISL_416523 | University of Wisconsin-Madison AIDS Vaccine Research Laboratories | University of Wisconsin-Madison AIDS Vaccine Research Laboratories |
| EPI_ISL_416524 | Saitama Medical University Hospital | Saitama Medical University |
| EPI_ISL_416525 | Saitama Medical University | Saitama Medical University |
| EPI_ISL_416526 | Auckland Hospital | Institute of Environmental Science and Research (ESR) |
| EPI_ISL_416538 | Wellington Hospital | Institute of Environmental Science and Research (ESR) |
| EPI_ISL_416539 | Wellington Hospital | Institute of Environmental Science and Research (ESR) |
| EPI_ISL_416541 | Dasman Diabetes Institute and Virology Laboratory Ministry of Health | Dasman Diabetes Institute |
| EPI_ISL_416542 | Dasman Diabetes Institute | Dasman Diabetes Institute |
| EPI_ISL_416543 | Dasman Diabetes Institute | Dasman Diabetes Institute |
| EPI_ISL_416635 | UW Virology Lab | UW Virology Lab |
| EPI_ISL_416636 | UW Virology Lab | UW Virology Lab |
| EPI_ISL_416637 | UW Virology Lab | UW Virology Lab |
| EPI_ISL_416638 | UW Virology Lab | UW Virology Lab |
| EPI_ISL_416639 | UW Virology Lab | UW Virology Lab |
| EPI_ISL_416640 | UW Virology Lab | UW Virology Lab |

| EPI_Accession_ID | Originating Laboratory | Submitting Laboratory |
| --- | --- | --- |
| EPI_ISL_416641 | UW Virology Lab | UW Virology Lab |
| EPI_ISL_416642 | UW Virology Lab | UW Virology Lab |
| EPI_ISL_416643 | UW Virology Lab | UW Virology Lab |
| EPI_ISL_416644 | UW Virology Lab | UW Virology Lab |
| EPI_ISL_416645 | UW Virology Lab | UW Virology Lab |
| EPI_ISL_416646 | UW Virology Lab | UW Virology Lab |
| EPI_ISL_416647 | UW Virology Lab | UW Virology Lab |
| EPI_ISL_416648 | UW Virology Lab | UW Virology Lab |
| EPI_ISL_416649 | UW Virology Lab | UW Virology Lab |
| EPI_ISL_416650 | UW Virology Lab | UW Virology Lab |
| EPI_ISL_416651 | UW Virology Lab | UW Virology Lab |
| EPI_ISL_416652 | UW Virology Lab | UW Virology Lab |
| EPI_ISL_416653 | UW Virology Lab | UW Virology Lab |
| EPI_ISL_416654 | UW Virology Lab | UW Virology Lab |
| EPI_ISL_416655 | UW Virology Lab | UW Virology Lab |
| EPI_ISL_416656 | UW Virology Lab | UW Virology Lab |
| EPI_ISL_416658 | UW Virology Lab | UW Virology Lab |
| EPI_ISL_416659 | UW Virology Lab | UW Virology Lab |
| EPI_ISL_416660 | UW Virology Lab | UW Virology Lab |
| EPI_ISL_416661 | UW Virology Lab | UW Virology Lab |
| EPI_ISL_416662 | UW Virology Lab | UW Virology Lab |
| EPI_ISL_416663 | UW Virology Lab | UW Virology Lab |
| EPI_ISL_416664 | UW Virology Lab | UW Virology Lab |
| EPI_ISL_416665 | UW Virology Lab | UW Virology Lab |
| EPI_ISL_416666 | UW Virology Lab | UW Virology Lab |
| EPI_ISL_416667 | UW Virology Lab | UW Virology Lab |
| EPI_ISL_416668 | UW Virology Lab | UW Virology Lab |
| EPI_ISL_416669 | UW Virology Lab | UW Virology Lab |
| EPI_ISL_416670 | UW Virology Lab | UW Virology Lab |
| EPI_ISL_416671 | UW Virology Lab | UW Virology Lab |
| EPI_ISL_416672 | UW Virology Lab | UW Virology Lab |
| EPI_ISL_416673 | UW Virology Lab | UW Virology Lab |
| EPI_ISL_416674 | UW Virology Lab | UW Virology Lab |

| EPI_Accession_ID | Originating Laboratory | Submitting Laboratory |
|---|---|---|
| EPI_ISL_416675 | UW Virology Lab | UW Virology Lab |
| EPI_ISL_416676 | UW Virology Lab | UW Virology Lab |
| EPI_ISL_416677 | UW Virology Lab | UW Virology Lab |
| EPI_ISL_416678 | UW Virology Lab | UW Virology Lab |
| EPI_ISL_416679 | UW Virology Lab | UW Virology Lab |
| EPI_ISL_416680 | UW Virology Lab | UW Virology Lab |
| EPI_ISL_416681 | UW Virology Lab | UW Virology Lab |
| EPI_ISL_416682 | UW Virology Lab | UW Virology Lab |
| EPI_ISL_416683 | UW Virology Lab | UW Virology Lab |
| EPI_ISL_416684 | UW Virology Lab | UW Virology Lab |
| EPI_ISL_416685 | UW Virology Lab | UW Virology Lab |
| EPI_ISL_416686 | UW Virology Lab | UW Virology Lab |
| EPI_ISL_416687 | UW Virology Lab | UW Virology Lab |
| EPI_ISL_416688 | UW Virology Lab | UW Virology Lab |
| EPI_ISL_416689 | UW Virology Lab | UW Virology Lab |
| EPI_ISL_416690 | UW Virology Lab | UW Virology Lab |
| EPI_ISL_416691 | UW Virology Lab | UW Virology Lab |
| EPI_ISL_416692 | UW Virology Lab | UW Virology Lab |
| EPI_ISL_416693 | UW Virology Lab | UW Virology Lab |
| EPI_ISL_416694 | UW Virology Lab | UW Virology Lab |
| EPI_ISL_416695 | UW Virology Lab | UW Virology Lab |
| EPI_ISL_416696 | UW Virology Lab | UW Virology Lab |
| EPI_ISL_416697 | UW Virology Lab | UW Virology Lab |
| EPI_ISL_416698 | UW Virology Lab | UW Virology Lab |
| EPI_ISL_416699 | UW Virology Lab | UW Virology Lab |
| EPI_ISL_416700 | UW Virology Lab | UW Virology Lab |
| EPI_ISL_416701 | UW Virology Lab | UW Virology Lab |
| EPI_ISL_416702 | UW Virology Lab | UW Virology Lab |
| EPI_ISL_416703 | UW Virology Lab | UW Virology Lab |
| EPI_ISL_416704 | UW Virology Lab | UW Virology Lab |
| EPI_ISL_416705 | UW Virology Lab | UW Virology Lab |
| EPI_ISL_416706 | UW Virology Lab | UW Virology Lab |
| EPI_ISL_416707 | UW Virology Lab | UW Virology Lab |

| EPI_Accession_ID | Originating Laboratory | Submitting Laboratory |
|---|---|---|
| EPI_ISL_416708 | UW Virology Lab | UW Virology Lab |
| EPI_ISL_416709 | UW Virology Lab | UW Virology Lab |
| EPI_ISL_416710 | UW Virology Lab | UW Virology Lab |
| EPI_ISL_416711 | UW Virology Lab | UW Virology Lab |
| EPI_ISL_416712 | UW Virology Lab | UW Virology Lab |
| EPI_ISL_416713 | UW Virology Lab | UW Virology Lab |
| EPI_ISL_416714 | UW Virology Lab | UW Virology Lab |
| EPI_ISL_416715 | UW Virology Lab | UW Virology Lab |
| EPI_ISL_416716 | UW Virology Lab | UW Virology Lab |
| EPI_ISL_416717 | UW Virology Lab | UW Virology Lab |
| EPI_ISL_416718 | UW Virology Lab | UW Virology Lab |
| EPI_ISL_416719 | UW Virology Lab | UW Virology Lab |
| EPI_ISL_416720 | UW Virology Lab | UW Virology Lab |
| EPI_ISL_416721 | UW Virology Lab | UW Virology Lab |
| EPI_ISL_416722 | UW Virology Lab | UW Virology Lab |
| EPI_ISL_416723 | UW Virology Lab | UW Virology Lab |
| EPI_ISL_416724 | UW Virology Lab | UW Virology Lab |
| EPI_ISL_416725 | UW Virology Lab | UW Virology Lab |
| EPI_ISL_416726 | UW Virology Lab | UW Virology Lab |
| EPI_ISL_416727 | UW Virology Lab | UW Virology Lab |
| EPI_ISL_416728 | UW Virology Lab | UW Virology Lab |
| EPI_ISL_416729 | UW Virology Lab | UW Virology Lab |
| EPI_ISL_416730 | Virology Department, Sheffield Teaching Hospitals NHS Foundation Trust | Department of Infection, Immunity and Cardiovascular Disease, The Florey Institute, The Medical School, University of Sheffield |
| EPI_ISL_416731 | Virology Department, Sheffield Teaching Hospitals NHS Foundation Trust | Department of Infection, Immunity and Cardiovascular Disease, The Florey Institute, The Medical School, University of Sheffield |
| EPI_ISL_416732 | Virology Department, Sheffield Teaching Hospitals NHS Foundation Trust | Department of Infection, Immunity and Cardiovascular Disease, The Florey Institute, The Medical School, University of Sheffield |

| EPI_Accession_ID | Originating Laboratory | Submitting Laboratory |
|---|---|---|
| EPI_ISL_416733 | Virology Department, Sheffield Teaching Hospitals NHS Foundation Trust | Department of Infection, Immunity and Cardiovascular Disease, The Florey Institute, The Medical School, University of Sheffield |
| EPI_ISL_416734 | Virology Department, Sheffield Teaching Hospitals NHS Foundation Trust | Department of Infection, Immunity and Cardiovascular Disease, The Florey Institute, The Medical School, University of Sheffield |
| EPI_ISL_416735 | Virology Department, Sheffield Teaching Hospitals NHS Foundation Trust | Department of Infection, Immunity and Cardiovascular Disease, The Florey Institute, The Medical School, University of Sheffield |
| EPI_ISL_416736 | Virology Department, Sheffield Teaching Hospitals NHS Foundation Trust | Department of Infection, Immunity and Cardiovascular Disease, The Florey Institute, The Medical School, University of Sheffield |
| EPI_ISL_416737 | Virology Department, Sheffield Teaching Hospitals NHS Foundation Trust | Department of Infection, Immunity and Cardiovascular Disease, The Florey Institute, The Medical School, University of Sheffield |
| EPI_ISL_416738 | Virology Department, Sheffield Teaching Hospitals NHS Foundation Trust | Department of Infection, Immunity and Cardiovascular Disease, The Florey Institute, The Medical School, University of Sheffield |
| EPI_ISL_416739 | Virology Department, Sheffield Teaching Hospitals NHS Foundation Trust | Department of Infection, Immunity and Cardiovascular Disease, The Florey Institute, The Medical School, University of Sheffield |
| EPI_ISL_416740 | Virology Department, Sheffield Teaching Hospitals NHS Foundation Trust | Department of Infection, Immunity and Cardiovascular Disease, The Florey Institute, The Medical School, University of Sheffield |
| EPI_ISL_416742 | NRL for Influenza, Centrum Epidemiology and Microbiology of National Institute of Public Health, Czech Republic | Charite Universitaetsmedizin Berlin, Institute of Virology |
| EPI_ISL_416743 | NRL for Influenza, Centrum Epidemiology and Microbiology of National Institute of Public Health, Czech Republic | Charite Universitaetsmedizin Berlin, Institute of Virology |
| EPI_ISL_416744 | Virological Research Group, SzentÃ¡gothai Research Centre | Bioinformatics Research Group, SzentÃ¡gothai Research Centre |
| EPI_ISL_416745 | CNR Virus des Infections Respiratoires - France SUD | CNR Virus des Infections Respiratoires - France SUD |

| EPI_Accession_ID | Originating Laboratory | Submitting Laboratory |
|---|---|---|
| EPI_ISL_416746 | CNR Virus des Infections Respiratoires - France SUD | CNR Virus des Infections Respiratoires - France SUD |
| EPI_ISL_416747 | Institut des Agents Infectieux (IAI) Hospices Civils de Lyon | CNR Virus des Infections Respiratoires - France SUD |
| EPI_ISL_416748 | Institut des Agents Infectieux (IAI) Hospices Civils de Lyon | CNR Virus des Infections Respiratoires - France SUD |
| EPI_ISL_416749 | Centre Hospitalier de Valence | CNR Virus des Infections Respiratoires - France SUD |
| EPI_ISL_416750 | Institut des Agents Infectieux (IAI) Hospices Civils de Lyon | CNR Virus des Infections Respiratoires - France SUD |
| EPI_ISL_416751 | CHU Gabriel Montpied | CNR Virus des Infections Respiratoires - France SUD |
| EPI_ISL_416752 | CHU Gabriel Montpied | CNR Virus des Infections Respiratoires - France SUD |
| EPI_ISL_416753 | Institut des Agents Infectieux (IAI) Hospices Civils de Lyon | CNR Virus des Infections Respiratoires - France SUD |
| EPI_ISL_416754 | Institut des Agents Infectieux (IAI) Hospices Civils de Lyon | CNR Virus des Infections Respiratoires - France SUD |
| EPI_ISL_416756 | Institut des Agents Infectieux (IAI) Hospices Civils de Lyon | CNR Virus des Infections Respiratoires - France SUD |
| EPI_ISL_416757 | Centre Hospitalier de Bourg en Bresse | CNR Virus des Infections Respiratoires - France SUD |
| EPI_ISL_416758 | Institut des Agents Infectieux (IAI) Hospices Civils de Lyon | CNR Virus des Infections Respiratoires - France SUD |
| EPI_ISL_416829 | National Public Health Laboratory | Malaysia Genome Institute |
| EPI_ISL_416830 | NYU Langone Health | Department of Pathology and Medicine, New York University School of Medicine |
| EPI_ISL_416831 | NYU Langone Health | Department of Pathology and Medicine, New York University School of Medicine |
| EPI_ISL_416832 | NYU Langone Health | Department of Pathology and Medicine, New York University School of Medicine |
| EPI_ISL_416866 | National Public Health Laboratory | Malaysia Genome Institute |
| EPI_ISL_416884 | National Public Health Laboratory | Malaysia Genome Institute |
| EPI_ISL_416885 | National Public Health Laboratory | Malaysia Genome Institute |
| EPI_ISL_416886 | National Public Health Laboratory | Malaysia Genome Institute |
| EPI_ISL_416907 | National Public Health Laboratory | Malaysia Genome Institute |
| EPI_ISL_416994 | COMPLEJO ASISTENCIAL UNIVERSITARIO DE BURGOS | Instituto de Salud Carlos III |
| EPI_ISL_416997 | Department of Clinical Microbiology | GIGA Medical Genomics |

| EPI_Accession_ID | Originating Laboratory | Submitting Laboratory |
|---|---|---|
| EPI_ISL_417004 | Department of Clinical Microbiology | GIGA Medical Genomics |
| EPI_ISL_417005 | Department of Clinical Microbiology | GIGA Medical Genomics |
| EPI_ISL_417006 | Department of Clinical Microbiology | GIGA Medical Genomics |
| EPI_ISL_417007 | HOSPITAL SANTA MARIA NAI | Instituto de Salud Carlos III |
| EPI_ISL_417008 | Department of Clinical Microbiology | GIGA Medical Genomics |
| EPI_ISL_417009 | Department of Clinical Microbiology | GIGA Medical Genomics |
| EPI_ISL_417010 | FUNDACION JIMENEZ DIAZ | Instituto de Salud Carlos III |
| EPI_ISL_417011 | Department of Clinical Microbiology | GIGA Medical Genomics |
| EPI_ISL_417012 | Department of Clinical Microbiology | GIGA Medical Genomics |
| EPI_ISL_417013 | Department of Clinical Microbiology | GIGA Medical Genomics |
| EPI_ISL_417014 | Department of Clinical Microbiology | GIGA Medical Genomics |
| EPI_ISL_417015 | Department of Clinical Microbiology | GIGA Medical Genomics |
| EPI_ISL_417016 | Department of Clinical Microbiology | GIGA Medical Genomics |
| EPI_ISL_417017 | Department of Clinical Microbiology | GIGA Medical Genomics |
| EPI_ISL_417018 | Department of Clinical Microbiology | GIGA Medical Genomics |
| EPI_ISL_417019 | Department of Clinical Microbiology | GIGA Medical Genomics |
| EPI_ISL_417020 | Department of Clinical Microbiology | GIGA Medical Genomics |
| EPI_ISL_417021 | Department of Clinical Microbiology | GIGA Medical Genomics |
| EPI_ISL_417022 | Department of Clinical Microbiology | GIGA Medical Genomics |
| EPI_ISL_417023 | Department of Clinical Microbiology | GIGA Medical Genomics |
| EPI_ISL_417025 | Department of Clinical Microbiology | GIGA Medical Genomics |
| EPI_ISL_417026 | Utah Public Health Laboratory | Utah Public Health Laboratory |
| EPI_ISL_417027 | Utah Public Health Laboratory | Utah Public Health Laboratory |
| EPI_ISL_417028 | Utah Public Health Laboratory | Utah Public Health Laboratory |
| EPI_ISL_417030 | Centre for Infectious Diseases and Microbiology Laboratory Services | NSW Health Pathology - Institute of Clinical Pathology and Medical Research; Westmead Hospital; University of Sydney |
| EPI_ISL_417031 | Pathology Queensland | Public Health Virology Laboratory |
| EPI_ISL_417032 | Rockhampton Base Hospital | Public Health Virology Laboratory |
| EPI_ISL_417033 | Sullivan Nicolaides Pathology | Public Health Virology Laboratory |
| EPI_ISL_417034 | Laboratorio de Ecologia de Doencas Transmissiveis na Amazonia, Instituto Leonidas e Maria Deane - Fiocruz Amazonia | Laboratorio de Ecologia de Doencas Transmissiveis na Amazonia, Instituto Leonidas e Maria Deane - Fiocruz Amazonia |

| EPI_Accession_ID | Originating Laboratory | Submitting Laboratory |
|---|---|---|
| EPI_ISL_417064 | Prince of Wales Hospital | Hong Kong Department of Health |
| EPI_ISL_417065 | Washington State Department of Health | Seattle Flu Study |
| EPI_ISL_417066 | Washington State Department of Health | Seattle Flu Study |
| EPI_ISL_417067 | Washington State Department of Health | Seattle Flu Study |
| EPI_ISL_417068 | Washington State Department of Health | Seattle Flu Study |
| EPI_ISL_417069 | Washington State Department of Health | Seattle Flu Study |
| EPI_ISL_417070 | Washington State Department of Health | Seattle Flu Study |
| EPI_ISL_417071 | Washington State Department of Health | Seattle Flu Study |
| EPI_ISL_417072 | Washington State Department of Health | Seattle Flu Study |
| EPI_ISL_417073 | Washington State Department of Health | Seattle Flu Study |
| EPI_ISL_417074 | Washington State Department of Health | Seattle Flu Study |
| EPI_ISL_417075 | Washington State Department of Health | Seattle Flu Study |
| EPI_ISL_417076 | Washington State Department of Health | Seattle Flu Study |
| EPI_ISL_417077 | Washington State Department of Health | Seattle Flu Study |
| EPI_ISL_417078 | Washington State Department of Health | Seattle Flu Study |
| EPI_ISL_417079 | Washington State Department of Health | Seattle Flu Study |
| EPI_ISL_417080 | Washington State Department of Health | Seattle Flu Study |
| EPI_ISL_417081 | Washington State Department of Health | Seattle Flu Study |
| EPI_ISL_417082 | Washington State Department of Health | Seattle Flu Study |
| EPI_ISL_417083 | Washington State Department of Health | Seattle Flu Study |
| EPI_ISL_417084 | Washington State Department of Health | Seattle Flu Study |
| EPI_ISL_417085 | Washington State Department of Health | Seattle Flu Study |
| EPI_ISL_417086 | Washington State Department of Health | Seattle Flu Study |
| EPI_ISL_417087 | Washington State Department of Health | Seattle Flu Study |
| EPI_ISL_417088 | Washington State Department of Health | Seattle Flu Study |
| EPI_ISL_417089 | Washington State Department of Health | Seattle Flu Study |
| EPI_ISL_417090 | Washington State Department of Health | Seattle Flu Study |
| EPI_ISL_417091 | Washington State Department of Health | Seattle Flu Study |
| EPI_ISL_417092 | Washington State Department of Health | Seattle Flu Study |
| EPI_ISL_417093 | Washington State Department of Health | Seattle Flu Study |
| EPI_ISL_417094 | Washington State Department of Health | Seattle Flu Study |
| EPI_ISL_417095 | Washington State Department of Health | Seattle Flu Study |
| EPI_ISL_417096 | Washington State Department of Health | Seattle Flu Study |

| EPI_Accession_ID | Originating Laboratory | Submitting Laboratory |
|---|---|---|
| EPI_ISL_417097 | Washington State Department of Health | Seattle Flu Study |
| EPI_ISL_417098 | Washington State Department of Health | Seattle Flu Study |
| EPI_ISL_417099 | Washington State Department of Health | Seattle Flu Study |
| EPI_ISL_417100 | Washington State Department of Health | Seattle Flu Study |
| EPI_ISL_417101 | Washington State Department of Health | Seattle Flu Study |
| EPI_ISL_417102 | Washington State Department of Health | Seattle Flu Study |
| EPI_ISL_417103 | Washington State Department of Health | Seattle Flu Study |
| EPI_ISL_417104 | Washington State Department of Health | Seattle Flu Study |
| EPI_ISL_417105 | Washington State Department of Health | Seattle Flu Study |
| EPI_ISL_417106 | Washington State Department of Health | Seattle Flu Study |
| EPI_ISL_417107 | Washington State Department of Health | Seattle Flu Study |
| EPI_ISL_417108 | Washington State Department of Health | Seattle Flu Study |
| EPI_ISL_417109 | Washington State Department of Health | Seattle Flu Study |
| EPI_ISL_417110 | Washington State Department of Health | Seattle Flu Study |
| EPI_ISL_417111 | Washington State Department of Health | Seattle Flu Study |
| EPI_ISL_417112 | Washington State Department of Health | Seattle Flu Study |
| EPI_ISL_417113 | Washington State Department of Health | Seattle Flu Study |
| EPI_ISL_417114 | Washington State Department of Health | Seattle Flu Study |
| EPI_ISL_417115 | Washington State Department of Health | Seattle Flu Study |
| EPI_ISL_417116 | Washington State Department of Health | Seattle Flu Study |
| EPI_ISL_417117 | Washington State Department of Health | Seattle Flu Study |
| EPI_ISL_417118 | Washington State Department of Health | Seattle Flu Study |
| EPI_ISL_417119 | Washington State Department of Health | Seattle Flu Study |
| EPI_ISL_417120 | Washington State Department of Health | Seattle Flu Study |
| EPI_ISL_417121 | Washington State Department of Health | Seattle Flu Study |
| EPI_ISL_417122 | Washington State Department of Health | Seattle Flu Study |
| EPI_ISL_417123 | Washington State Department of Health | Seattle Flu Study |
| EPI_ISL_417124 | Washington State Department of Health | Seattle Flu Study |
| EPI_ISL_417125 | Washington State Department of Health | Seattle Flu Study |
| EPI_ISL_417126 | Washington State Department of Health | Seattle Flu Study |
| EPI_ISL_417127 | Washington State Department of Health | Seattle Flu Study |
| EPI_ISL_417128 | Washington State Department of Health | Seattle Flu Study |
| EPI_ISL_417129 | Washington State Department of Health | Seattle Flu Study |

| EPI_Accession_ID | Originating Laboratory | Submitting Laboratory |
|---|---|---|
| EPI_ISL_417130 | Washington State Department of Health | Seattle Flu Study |
| EPI_ISL_417131 | Washington State Department of Health | Seattle Flu Study |
| EPI_ISL_417132 | Washington State Department of Health | Seattle Flu Study |
| EPI_ISL_417133 | Washington State Department of Health | Seattle Flu Study |
| EPI_ISL_417134 | Washington State Department of Health | Seattle Flu Study |
| EPI_ISL_417135 | Washington State Department of Health | Seattle Flu Study |
| EPI_ISL_417136 | Washington State Department of Health | Seattle Flu Study |
| EPI_ISL_417137 | Washington State Department of Health | Seattle Flu Study |
| EPI_ISL_417138 | Washington State Department of Health | Seattle Flu Study |
| EPI_ISL_417139 | Washington State Department of Health | Seattle Flu Study |
| EPI_ISL_417140 | Washington State Department of Health | Seattle Flu Study |
| EPI_ISL_417141 | Washington State Department of Health | Seattle Flu Study |
| EPI_ISL_417142 | Washington State Department of Health | Seattle Flu Study |
| EPI_ISL_417143 | Washington State Department of Health | Seattle Flu Study |
| EPI_ISL_417144 | Washington State Department of Health | Seattle Flu Study |
| EPI_ISL_417145 | Washington State Department of Health | Seattle Flu Study |
| EPI_ISL_417146 | Washington State Department of Health | Seattle Flu Study |
| EPI_ISL_417147 | Washington State Department of Health | Seattle Flu Study |
| EPI_ISL_417148 | Washington State Department of Health | Seattle Flu Study |
| EPI_ISL_417149 | Washington State Department of Health | Seattle Flu Study |
| EPI_ISL_417150 | Washington State Department of Health | Seattle Flu Study |
| EPI_ISL_417151 | Washington State Department of Health | Seattle Flu Study |
| EPI_ISL_417152 | Washington State Department of Health | Seattle Flu Study |
| EPI_ISL_417153 | Washington State Department of Health | Seattle Flu Study |
| EPI_ISL_417154 | Washington State Department of Health | Seattle Flu Study |
| EPI_ISL_417155 | Washington State Department of Health | Seattle Flu Study |
| EPI_ISL_417156 | Washington State Department of Health | Seattle Flu Study |
| EPI_ISL_417157 | Washington State Department of Health | Seattle Flu Study |
| EPI_ISL_417158 | Washington State Department of Health | Seattle Flu Study |
| EPI_ISL_417159 | Washington State Department of Health | Seattle Flu Study |
| EPI_ISL_417160 | Washington State Department of Health | Seattle Flu Study |
| EPI_ISL_417161 | Washington State Department of Health | Seattle Flu Study |
| EPI_ISL_417162 | Washington State Department of Health | Seattle Flu Study |

| EPI_Accession_ID | Originating Laboratory | Submitting Laboratory |
|---|---|---|
| EPI_ISL_417163 | Seattle Flu Study | Seattle Flu Study |
| EPI_ISL_417164 | Seattle Flu Study | Seattle Flu Study |
| EPI_ISL_417165 | Seattle Flu Study | Seattle Flu Study |
| EPI_ISL_417166 | Washington State Department of Health | Seattle Flu Study |
| EPI_ISL_417167 | Washington State Department of Health | Seattle Flu Study |
| EPI_ISL_417168 | Washington State Department of Health | Seattle Flu Study |
| EPI_ISL_417169 | Washington State Department of Health | Seattle Flu Study |
| EPI_ISL_417170 | Washington State Department of Health | Seattle Flu Study |
| EPI_ISL_417171 | Washington State Department of Health | Seattle Flu Study |
| EPI_ISL_417172 | Washington State Department of Health | Seattle Flu Study |
| EPI_ISL_417173 | Washington State Department of Health | Seattle Flu Study |
| EPI_ISL_417174 | Washington State Department of Health | Seattle Flu Study |
| EPI_ISL_417175 | Washington State Department of Health | Seattle Flu Study |
| EPI_ISL_417176 | Department of Pathology, Princess Margaret Hospital | Department of Health Technology and Informatics, Faculty of Health and Social Science, The Hong Kong Polytechnic University |
| EPI_ISL_417177 | Department of Pathology, Princess Margaret Hospital | Department of Health Technology and Informatics, Faculty of Health and Social Science, The Hong Kong Polytechnic University |
| EPI_ISL_417178 | Department of Pathology, Princess Margaret Hospital | Department of Health Technology and Informatics, Faculty of Health and Social Science, The Hong Kong Polytechnic University |
| EPI_ISL_417179 | Department of Pathology, Princess Margaret Hospital | Department of Health Technology and Informatics, Faculty of Health and Social Science, The Hong Kong Polytechnic University |
| EPI_ISL_417180 | Department of Pathology, United Christian Hospital | Department of Health Technology and Informatics, Faculty of Health and Social Science, The Hong Kong Polytechnic University |
| EPI_ISL_417181 | Department of Pathology, United Christian Hospital | Department of Health Technology and Informatics, Faculty of Health and Social Science, The Hong Kong Polytechnic University |

| EPI_Accession_ID | Originating Laboratory | Submitting Laboratory |
|---|---|---|
| EPI_ISL_417182 | Department of Pathology, United Christian Hospital | Department of Health Technology and Informatics, Faculty of Health and Social Science, The Hong Kong Polytechnic University |
| EPI_ISL_417183 | Department of Clinical Pathology, Pamela Youde Nethersole Eastern Hospital | Department of Health Technology and Informatics, Faculty of Health and Social Science, The Hong Kong Polytechnic University |
| EPI_ISL_417184 | Department of Clinical Pathology, Pamela Youde Nethersole Eastern Hospital | Department of Health Technology and Informatics, Faculty of Health and Social Science, The Hong Kong Polytechnic University |
| EPI_ISL_417185 | Department of Pathology, United Christian Hospital | Department of Health Technology and Informatics, Faculty of Health and Social Science, The Hong Kong Polytechnic University |
| EPI_ISL_417186 | National Institute for Communicable Diseases of the National Health Laboratory Service | National Institute for Communicable Diseases of the National Health Laboratory Service |
| EPI_ISL_417187 | Department of Clinical Pathology, Pamela Youde Nethersole Eastern Hospital | Department of Health Technology and Informatics, Faculty of Health and Social Science, The Hong Kong Polytechnic University |
| EPI_ISL_417188 | Department of Clinical Pathology, Pamela Youde Nethersole Eastern Hospital | Department of Health Technology and Informatics, Faculty of Health and Social Science, The Hong Kong Polytechnic University |
| EPI_ISL_417189 | Minnesota Department of Health, Public Health Laboratory | Minnesota Department of Health, Public Health Laboratory |
| EPI_ISL_417190 | Department of Clinical Pathology, Pamela Youde Nethersole Eastern Hospital | Department of Health Technology and Informatics, Faculty of Health and Social Science, The Hong Kong Polytechnic University |
| EPI_ISL_417191 | Minnesota Department of Health, Public Health Laboratory | Minnesota Department of Health, Public Health Laboratory |
| EPI_ISL_417192 | Minnesota Department of Health, Public Health Laboratory | Minnesota Department of Health, Public Health Laboratory |
| EPI_ISL_417193 | Department of Clinical Pathology, Pamela Youde Nethersole Eastern Hospital | Department of Health Technology and Informatics, Faculty of Health and Social Science, The Hong Kong Polytechnic University |

| EPI_Accession_ID | Originating Laboratory | Submitting Laboratory |
|---|---|---|
| EPI_ISL_417194 | Minnesota Department of Health, Public Health Laboratory | Minnesota Department of Health, Public Health Laboratory |
| EPI_ISL_417195 | Department of Clinical Pathology, Pamela Youde Nethersole Eastern Hospital | Department of Health Technology and Informatics, Faculty of Health and Social Science, The Hong Kong Polytechnic University |
| EPI_ISL_417196 | Minnesota Department of Health, Public Health Laboratory | Minnesota Department of Health, Public Health Laboratory |
| EPI_ISL_417197 | Department of Clinical Pathology, Pamela Youde Nethersole Eastern Hospital | Department of Health Technology and Informatics, Faculty of Health and Social Science, The Hong Kong Polytechnic University |
| EPI_ISL_417198 | Minnesota Department of Health, Public Health Laboratory | Minnesota Department of Health, Public Health Laboratory |
| EPI_ISL_417199 | Department of Clinical Pathology, Pamela Youde Nethersole Eastern Hospital | Department of Health Technology and Informatics, Faculty of Health and Social Science, The Hong Kong Polytechnic University |
| EPI_ISL_417200 | University of Wisconsin-Madison AIDS Vaccine Research Laboratories | University of Wisconsin-Madison AIDS Vaccine Research Laboratories |
| EPI_ISL_417201 | University of Wisconsin-Madison AIDS Vaccine Research Laboratories | University of Wisconsin-Madison AIDS Vaccine Research Laboratories |
| EPI_ISL_417202 | University of Wisconsin-Madison AIDS Vaccine Research Laboratories | University of Wisconsin-Madison AIDS Vaccine Research Laboratories |
| EPI_ISL_417203 | University of Wisconsin-Madison AIDS Vaccine Research Laboratories | University of Wisconsin-Madison AIDS Vaccine Research Laboratories |
| EPI_ISL_417204 | University of Wisconsin-Madison AIDS Vaccine Research Laboratories | University of Wisconsin-Madison AIDS Vaccine Research Laboratories |
| EPI_ISL_417205 | Servicio de MicrobiologÃ-a. Consorcio Hospital General Universitario de Valencia | Sequencing and Bioinformatics Service and Molecular Epidemiology Research Group. FISABIO-Public Health |
| EPI_ISL_417206 | Servicio de Microbiologia. Consorcio Hospital General Universitario de Valencia | Sequencing and Bioinformatics Service and Molecular Epidemiology Research Group. FISABIO-Public Health |
| EPI_ISL_417211 | Dunedin Hospital | University of Otago |
| EPI_ISL_417212 | Dunedin Hospital | University of Otago |
| EPI_ISL_417213 | Respiratory Virus Unit, Microbiology Services Colindale, Public Health England | Respiratory Virus Unit, Microbiology Services Colindale, Public Health England |

| EPI_Accession_ID | Originating Laboratory | Submitting Laboratory |
|---|---|---|
| EPI_ISL_417214 | Respiratory Virus Unit, Microbiology Services Colindale, Public Health England | Respiratory Virus Unit, Microbiology Services Colindale, Public Health England |
| EPI_ISL_417215 | Respiratory Virus Unit, Microbiology Services Colindale, Public Health England | Respiratory Virus Unit, Microbiology Services Colindale, Public Health England |
| EPI_ISL_417216 | Respiratory Virus Unit, Microbiology Services Colindale, Public Health England | Respiratory Virus Unit, Microbiology Services Colindale, Public Health England |
| EPI_ISL_417217 | Respiratory Virus Unit, Microbiology Services Colindale, Public Health England | Respiratory Virus Unit, Microbiology Services Colindale, Public Health England |
| EPI_ISL_417218 | Respiratory Virus Unit, Microbiology Services Colindale, Public Health England | Respiratory Virus Unit, Microbiology Services Colindale, Public Health England |
| EPI_ISL_417219 | Respiratory Virus Unit, Microbiology Services Colindale, Public Health England | Respiratory Virus Unit, Microbiology Services Colindale, Public Health England |
| EPI_ISL_417220 | Respiratory Virus Unit, Microbiology Services Colindale, Public Health England | Respiratory Virus Unit, Microbiology Services Colindale, Public Health England |
| EPI_ISL_417221 | Respiratory Virus Unit, Microbiology Services Colindale, Public Health England | Respiratory Virus Unit, Microbiology Services Colindale, Public Health England |
| EPI_ISL_417222 | Respiratory Virus Unit, Microbiology Services Colindale, Public Health England | Respiratory Virus Unit, Microbiology Services Colindale, Public Health England |
| EPI_ISL_417223 | Respiratory Virus Unit, Microbiology Services Colindale, Public Health England | Respiratory Virus Unit, Microbiology Services Colindale, Public Health England |
| EPI_ISL_417224 | Respiratory Virus Unit, Microbiology Services Colindale, Public Health England | Respiratory Virus Unit, Microbiology Services Colindale, Public Health England |
| EPI_ISL_417225 | Respiratory Virus Unit, Microbiology Services Colindale, Public Health England | Respiratory Virus Unit, Microbiology Services Colindale, Public Health England |
| EPI_ISL_417226 | Respiratory Virus Unit, Microbiology Services Colindale, Public Health England | Respiratory Virus Unit, Microbiology Services Colindale, Public Health England |
| EPI_ISL_417227 | Respiratory Virus Unit, Microbiology Services Colindale, Public Health England | Respiratory Virus Unit, Microbiology Services Colindale, Public Health England |
| EPI_ISL_417228 | Respiratory Virus Unit, Microbiology Services Colindale, Public Health England | Respiratory Virus Unit, Microbiology Services Colindale, Public Health England |
| EPI_ISL_417229 | Respiratory Virus Unit, Microbiology Services Colindale, Public Health England | Respiratory Virus Unit, Microbiology Services Colindale, Public Health England |

| EPI_Accession_ID | Originating Laboratory | Submitting Laboratory |
|---|---|---|
| EPI_ISL_417230 | Respiratory Virus Unit, Microbiology Services Colindale, Public Health England | Respiratory Virus Unit, Microbiology Services Colindale, Public Health England |
| EPI_ISL_417231 | Respiratory Virus Unit, Microbiology Services Colindale, Public Health England | Respiratory Virus Unit, Microbiology Services Colindale, Public Health England |
| EPI_ISL_417232 | Respiratory Virus Unit, Microbiology Services Colindale, Public Health England | Respiratory Virus Unit, Microbiology Services Colindale, Public Health England |
| EPI_ISL_417233 | Respiratory Virus Unit, Microbiology Services Colindale, Public Health England | Respiratory Virus Unit, Microbiology Services Colindale, Public Health England |
| EPI_ISL_417234 | Respiratory Virus Unit, Microbiology Services Colindale, Public Health England | Respiratory Virus Unit, Microbiology Services Colindale, Public Health England |
| EPI_ISL_417235 | Respiratory Virus Unit, Microbiology Services Colindale, Public Health England | Respiratory Virus Unit, Microbiology Services Colindale, Public Health England |
| EPI_ISL_417236 | Respiratory Virus Unit, Microbiology Services Colindale, Public Health England | Respiratory Virus Unit, Microbiology Services Colindale, Public Health England |
| EPI_ISL_417237 | Respiratory Virus Unit, Microbiology Services Colindale, Public Health England | Respiratory Virus Unit, Microbiology Services Colindale, Public Health England |
| EPI_ISL_417238 | Respiratory Virus Unit, Microbiology Services Colindale, Public Health England | Respiratory Virus Unit, Microbiology Services Colindale, Public Health England |
| EPI_ISL_417239 | Respiratory Virus Unit, Microbiology Services Colindale, Public Health England | Respiratory Virus Unit, Microbiology Services Colindale, Public Health England |
| EPI_ISL_417240 | Respiratory Virus Unit, Microbiology Services Colindale, Public Health England | Respiratory Virus Unit, Microbiology Services Colindale, Public Health England |
| EPI_ISL_417241 | Respiratory Virus Unit, Microbiology Services Colindale, Public Health England | Respiratory Virus Unit, Microbiology Services Colindale, Public Health England |
| EPI_ISL_417242 | Respiratory Virus Unit, Microbiology Services Colindale, Public Health England | Respiratory Virus Unit, Microbiology Services Colindale, Public Health England |
| EPI_ISL_417243 | Respiratory Virus Unit, Microbiology Services Colindale, Public Health England | Respiratory Virus Unit, Microbiology Services Colindale, Public Health England |
| EPI_ISL_417244 | Respiratory Virus Unit, Microbiology Services Colindale, Public Health England | Respiratory Virus Unit, Microbiology Services Colindale, Public Health England |
| EPI_ISL_417245 | Respiratory Virus Unit, Microbiology Services Colindale, Public Health England | Respiratory Virus Unit, Microbiology Services Colindale, Public Health England |

| EPI_Accession_ID | Originating Laboratory | Submitting Laboratory |
| --- | --- | --- |
| EPI_ISL_417246 | Respiratory Virus Unit, Microbiology Services Colindale, Public Health England | Respiratory Virus Unit, Microbiology Services Colindale, Public Health England |
| EPI_ISL_417247 | Respiratory Virus Unit, Microbiology Services Colindale, Public Health England | Respiratory Virus Unit, Microbiology Services Colindale, Public Health England |
| EPI_ISL_417248 | Respiratory Virus Unit, Microbiology Services Colindale, Public Health England | Respiratory Virus Unit, Microbiology Services Colindale, Public Health England |
| EPI_ISL_417249 | Respiratory Virus Unit, Microbiology Services Colindale, Public Health England | Respiratory Virus Unit, Microbiology Services Colindale, Public Health England |
| EPI_ISL_417250 | Respiratory Virus Unit, Microbiology Services Colindale, Public Health England | Respiratory Virus Unit, Microbiology Services Colindale, Public Health England |
| EPI_ISL_417251 | Respiratory Virus Unit, Microbiology Services Colindale, Public Health England | Respiratory Virus Unit, Microbiology Services Colindale, Public Health England |
| EPI_ISL_417252 | Respiratory Virus Unit, Microbiology Services Colindale, Public Health England | Respiratory Virus Unit, Microbiology Services Colindale, Public Health England |
| EPI_ISL_417253 | Respiratory Virus Unit, Microbiology Services Colindale, Public Health England | Respiratory Virus Unit, Microbiology Services Colindale, Public Health England |
| EPI_ISL_417254 | Respiratory Virus Unit, Microbiology Services Colindale, Public Health England | Respiratory Virus Unit, Microbiology Services Colindale, Public Health England |
| EPI_ISL_417255 | Respiratory Virus Unit, Microbiology Services Colindale, Public Health England | Respiratory Virus Unit, Microbiology Services Colindale, Public Health England |
| EPI_ISL_417256 | Respiratory Virus Unit, Microbiology Services Colindale, Public Health England | Respiratory Virus Unit, Microbiology Services Colindale, Public Health England |
| EPI_ISL_417257 | Respiratory Virus Unit, Microbiology Services Colindale, Public Health England | Respiratory Virus Unit, Microbiology Services Colindale, Public Health England |
| EPI_ISL_417258 | Respiratory Virus Unit, Microbiology Services Colindale, Public Health England | Respiratory Virus Unit, Microbiology Services Colindale, Public Health England |
| EPI_ISL_417260 | Respiratory Virus Unit, Microbiology Services Colindale, Public Health England | Respiratory Virus Unit, Microbiology Services Colindale, Public Health England |
| EPI_ISL_417261 | Respiratory Virus Unit, Microbiology Services Colindale, Public Health England | Respiratory Virus Unit, Microbiology Services Colindale, Public Health England |
| EPI_ISL_417262 | Respiratory Virus Unit, Microbiology Services Colindale, Public Health England | Respiratory Virus Unit, Microbiology Services Colindale, Public Health England |

| EPI_Accession_ID | Originating Laboratory | Submitting Laboratory |
|---|---|---|
| EPI_ISL_417264 | Respiratory Virus Unit, Microbiology Services Colindale, Public Health England | Respiratory Virus Unit, Microbiology Services Colindale, Public Health England |
| EPI_ISL_417265 | Respiratory Virus Unit, Microbiology Services Colindale, Public Health England | Respiratory Virus Unit, Microbiology Services Colindale, Public Health England |
| EPI_ISL_417266 | Respiratory Virus Unit, Microbiology Services Colindale, Public Health England | Respiratory Virus Unit, Microbiology Services Colindale, Public Health England |
| EPI_ISL_417267 | Respiratory Virus Unit, Microbiology Services Colindale, Public Health England | Respiratory Virus Unit, Microbiology Services Colindale, Public Health England |
| EPI_ISL_417268 | Respiratory Virus Unit, Microbiology Services Colindale, Public Health England | Respiratory Virus Unit, Microbiology Services Colindale, Public Health England |
| EPI_ISL_417269 | Respiratory Virus Unit, Microbiology Services Colindale, Public Health England | Respiratory Virus Unit, Microbiology Services Colindale, Public Health England |
| EPI_ISL_417270 | Respiratory Virus Unit, Microbiology Services Colindale, Public Health England | Respiratory Virus Unit, Microbiology Services Colindale, Public Health England |
| EPI_ISL_417271 | Respiratory Virus Unit, Microbiology Services Colindale, Public Health England | Respiratory Virus Unit, Microbiology Services Colindale, Public Health England |
| EPI_ISL_417272 | Respiratory Virus Unit, Microbiology Services Colindale, Public Health England | Respiratory Virus Unit, Microbiology Services Colindale, Public Health England |
| EPI_ISL_417273 | Respiratory Virus Unit, Microbiology Services Colindale, Public Health England | Respiratory Virus Unit, Microbiology Services Colindale, Public Health England |
| EPI_ISL_417274 | Respiratory Virus Unit, Microbiology Services Colindale, Public Health England | Respiratory Virus Unit, Microbiology Services Colindale, Public Health England |
| EPI_ISL_417275 | Respiratory Virus Unit, Microbiology Services Colindale, Public Health England | Respiratory Virus Unit, Microbiology Services Colindale, Public Health England |
| EPI_ISL_417276 | Respiratory Virus Unit, Microbiology Services Colindale, Public Health England | Respiratory Virus Unit, Microbiology Services Colindale, Public Health England |
| EPI_ISL_417277 | Respiratory Virus Unit, Microbiology Services Colindale, Public Health England | Respiratory Virus Unit, Microbiology Services Colindale, Public Health England |
| EPI_ISL_417278 | Respiratory Virus Unit, Microbiology Services Colindale, Public Health England | Respiratory Virus Unit, Microbiology Services Colindale, Public Health England |
| EPI_ISL_417279 | Respiratory Virus Unit, Microbiology Services Colindale, Public Health England | Respiratory Virus Unit, Microbiology Services Colindale, Public Health England |

| EPI_Accession_ID | Originating Laboratory | Submitting Laboratory |
|---|---|---|
| EPI_ISL_417280 | Respiratory Virus Unit, Microbiology Services Colindale, Public Health England | Respiratory Virus Unit, Microbiology Services Colindale, Public Health England |
| EPI_ISL_417281 | Respiratory Virus Unit, Microbiology Services Colindale, Public Health England | Respiratory Virus Unit, Microbiology Services Colindale, Public Health England |
| EPI_ISL_417282 | Respiratory Virus Unit, Microbiology Services Colindale, Public Health England | Respiratory Virus Unit, Microbiology Services Colindale, Public Health England |
| EPI_ISL_417283 | Respiratory Virus Unit, Microbiology Services Colindale, Public Health England | Respiratory Virus Unit, Microbiology Services Colindale, Public Health England |
| EPI_ISL_417284 | Respiratory Virus Unit, Microbiology Services Colindale, Public Health England | Respiratory Virus Unit, Microbiology Services Colindale, Public Health England |
| EPI_ISL_417285 | Respiratory Virus Unit, Microbiology Services Colindale, Public Health England | Respiratory Virus Unit, Microbiology Services Colindale, Public Health England |
| EPI_ISL_417286 | Respiratory Virus Unit, Microbiology Services Colindale, Public Health England | Respiratory Virus Unit, Microbiology Services Colindale, Public Health England |
| EPI_ISL_417287 | Respiratory Virus Unit, Microbiology Services Colindale, Public Health England | Respiratory Virus Unit, Microbiology Services Colindale, Public Health England |
| EPI_ISL_417288 | Respiratory Virus Unit, Microbiology Services Colindale, Public Health England | Respiratory Virus Unit, Microbiology Services Colindale, Public Health England |
| EPI_ISL_417289 | Respiratory Virus Unit, Microbiology Services Colindale, Public Health England | Respiratory Virus Unit, Microbiology Services Colindale, Public Health England |
| EPI_ISL_417290 | Respiratory Virus Unit, Microbiology Services Colindale, Public Health England | Respiratory Virus Unit, Microbiology Services Colindale, Public Health England |
| EPI_ISL_417291 | Respiratory Virus Unit, Microbiology Services Colindale, Public Health England | Respiratory Virus Unit, Microbiology Services Colindale, Public Health England |
| EPI_ISL_417292 | Respiratory Virus Unit, Microbiology Services Colindale, Public Health England | Respiratory Virus Unit, Microbiology Services Colindale, Public Health England |
| EPI_ISL_417293 | Respiratory Virus Unit, Microbiology Services Colindale, Public Health England | Respiratory Virus Unit, Microbiology Services Colindale, Public Health England |
| EPI_ISL_417294 | Respiratory Virus Unit, Microbiology Services Colindale, Public Health England | Respiratory Virus Unit, Microbiology Services Colindale, Public Health England |
| EPI_ISL_417295 | Respiratory Virus Unit, Microbiology Services Colindale, Public Health England | Respiratory Virus Unit, Microbiology Services Colindale, Public Health England |

| EPI_Accession_ID | Originating Laboratory | Submitting Laboratory |
|---|---|---|
| EPI_ISL_417296 | Respiratory Virus Unit, Microbiology Services Colindale, Public Health England | Respiratory Virus Unit, Microbiology Services Colindale, Public Health England |
| EPI_ISL_417297 | Respiratory Virus Unit, Microbiology Services Colindale, Public Health England | Respiratory Virus Unit, Microbiology Services Colindale, Public Health England |
| EPI_ISL_417298 | Respiratory Virus Unit, Microbiology Services Colindale, Public Health England | Respiratory Virus Unit, Microbiology Services Colindale, Public Health England |
| EPI_ISL_417299 | Respiratory Virus Unit, Microbiology Services Colindale, Public Health England | Respiratory Virus Unit, Microbiology Services Colindale, Public Health England |
| EPI_ISL_417300 | Respiratory Virus Unit, Microbiology Services Colindale, Public Health England | Respiratory Virus Unit, Microbiology Services Colindale, Public Health England |
| EPI_ISL_417301 | Respiratory Virus Unit, Microbiology Services Colindale, Public Health England | Respiratory Virus Unit, Microbiology Services Colindale, Public Health England |
| EPI_ISL_417302 | Respiratory Virus Unit, Microbiology Services Colindale, Public Health England | Respiratory Virus Unit, Microbiology Services Colindale, Public Health England |
| EPI_ISL_417303 | Respiratory Virus Unit, Microbiology Services Colindale, Public Health England | Respiratory Virus Unit, Microbiology Services Colindale, Public Health England |
| EPI_ISL_417304 | Respiratory Virus Unit, Microbiology Services Colindale, Public Health England | Respiratory Virus Unit, Microbiology Services Colindale, Public Health England |
| EPI_ISL_417305 | Respiratory Virus Unit, Microbiology Services Colindale, Public Health England | Respiratory Virus Unit, Microbiology Services Colindale, Public Health England |
| EPI_ISL_417306 | Respiratory Virus Unit, Microbiology Services Colindale, Public Health England | Respiratory Virus Unit, Microbiology Services Colindale, Public Health England |
| EPI_ISL_417307 | Respiratory Virus Unit, Microbiology Services Colindale, Public Health England | Respiratory Virus Unit, Microbiology Services Colindale, Public Health England |
| EPI_ISL_417308 | Respiratory Virus Unit, Microbiology Services Colindale, Public Health England | Respiratory Virus Unit, Microbiology Services Colindale, Public Health England |
| EPI_ISL_417309 | Respiratory Virus Unit, Microbiology Services Colindale, Public Health England | Respiratory Virus Unit, Microbiology Services Colindale, Public Health England |
| EPI_ISL_417310 | Respiratory Virus Unit, Microbiology Services Colindale, Public Health England | Respiratory Virus Unit, Microbiology Services Colindale, Public Health England |
| EPI_ISL_417311 | Respiratory Virus Unit, Microbiology Services Colindale, Public Health England | Respiratory Virus Unit, Microbiology Services Colindale, Public Health England |

| EPI_Accession_ID | Originating Laboratory | Submitting Laboratory |
|---|---|---|
| EPI_ISL_417312 | Respiratory Virus Unit, Microbiology Services Colindale, Public Health England | Respiratory Virus Unit, Microbiology Services Colindale, Public Health England |
| EPI_ISL_417313 | Respiratory Virus Unit, Microbiology Services Colindale, Public Health England | Respiratory Virus Unit, Microbiology Services Colindale, Public Health England |
| EPI_ISL_417314 | Respiratory Virus Unit, Microbiology Services Colindale, Public Health England | Respiratory Virus Unit, Microbiology Services Colindale, Public Health England |
| EPI_ISL_417315 | Respiratory Virus Unit, Microbiology Services Colindale, Public Health England | Respiratory Virus Unit, Microbiology Services Colindale, Public Health England |
| EPI_ISL_417317 | Santa Clara County Public Health Department | Chiu Laboratory, University of California, San Francisco |
| EPI_ISL_417318 | Santa Clara County Public Health Department | Chiu Laboratory, University of California, San Francisco |
| EPI_ISL_417319 | Santa Clara County Public Health Department | Chiu Laboratory, University of California, San Francisco |
| EPI_ISL_417320 | Santa Clara County Public Health Department | Chiu Laboratory, University of California, San Francisco |
| EPI_ISL_417323 | California Department of Public Health | Chiu Laboratory, University of California, San Francisco |
| EPI_ISL_417324 | California Department of Public Health | Chiu Laboratory, University of California, San Francisco |
| EPI_ISL_417325 | California Department of Public Health | Chiu Laboratory, University of California, San Francisco |
| EPI_ISL_417327 | California Department of Public Health | Chiu Laboratory, University of California, San Francisco |
| EPI_ISL_417328 | California Department of Public Health | Chiu Laboratory, University of California, San Francisco |
| EPI_ISL_417330 | Chiu Laboratory, University of California, San Francisco | Chiu Laboratory, University of California, San Francisco |
| EPI_ISL_417331 | Chiu Laboratory, University of California, San Francisco | Chiu Laboratory, University of California, San Francisco |
| EPI_ISL_417332 | Chiu Laboratory, University of California, San Francisco | Chiu Laboratory, University of California, San Francisco |
| EPI_ISL_417333 | Institut des Agents Infectieux (IAI), Hospices Civils de Lyon | CNR Virus des Infections Respiratoires - France SUD |
| EPI_ISL_417334 | Institut des Agents Infectieux (IAI), Hospices Civils de Lyon | CNR Virus des Infections Respiratoires - France SUD |
| EPI_ISL_417335 | Institut des Agents Infectieux (IAI), Hospices Civils de Lyon | CNR Virus des Infections Respiratoires - France SUD |
| EPI_ISL_417336 | Institut des Agents Infectieux (IAI), Hospices Civils de Lyon | CNR Virus des Infections Respiratoires - France SUD |
| EPI_ISL_417337 | Institut des Agents Infectieux (IAI), Hospices Civils de Lyon | CNR Virus des Infections Respiratoires - France SUD |
| EPI_ISL_417338 | Centre Hospitalier de Macon | CNR Virus des Infections Respiratoires - France SUD |
| EPI_ISL_417339 | Institut des Agents Infectieux (IAI), Hospices Civils de Lyon | CNR Virus des Infections Respiratoires - France SUD |

| EPI_Accession_ID | Originating Laboratory | Submitting Laboratory |
|---|---|---|
| EPI_ISL_417340 | Centre Hospitalier de Bourg en Bresse | CNR Virus des Infections Respiratoires - France SUD |
| EPI_ISL_417341 | UW Virology Lab | UW Virology Lab |
| EPI_ISL_417342 | UW Virology Lab | UW Virology Lab |
| EPI_ISL_417343 | UW Virology Lab | UW Virology Lab |
| EPI_ISL_417344 | UW Virology Lab | UW Virology Lab |
| EPI_ISL_417345 | UW Virology Lab | UW Virology Lab |
| EPI_ISL_417346 | UW Virology Lab | UW Virology Lab |
| EPI_ISL_417347 | UW Virology Lab | UW Virology Lab |
| EPI_ISL_417348 | UW Virology Lab | UW Virology Lab |
| EPI_ISL_417349 | UW Virology Lab | UW Virology Lab |
| EPI_ISL_417350 | UW Virology Lab | UW Virology Lab |
| EPI_ISL_417351 | UW Virology Lab | UW Virology Lab |
| EPI_ISL_417352 | UW Virology Lab | UW Virology Lab |
| EPI_ISL_417353 | UW Virology Lab | UW Virology Lab |
| EPI_ISL_417354 | UW Virology Lab | UW Virology Lab |
| EPI_ISL_417355 | UW Virology Lab | UW Virology Lab |
| EPI_ISL_417356 | UW Virology Lab | UW Virology Lab |
| EPI_ISL_417357 | UW Virology Lab | UW Virology Lab |
| EPI_ISL_417358 | UW Virology Lab | UW Virology Lab |
| EPI_ISL_417359 | UW Virology Lab | UW Virology Lab |
| EPI_ISL_417360 | UW Virology Lab | UW Virology Lab |
| EPI_ISL_417361 | UW Virology Lab | UW Virology Lab |
| EPI_ISL_417362 | UW Virology Lab | UW Virology Lab |
| EPI_ISL_417363 | UW Virology Lab | UW Virology Lab |
| EPI_ISL_417364 | UW Virology Lab | UW Virology Lab |
| EPI_ISL_417365 | UW Virology Lab | UW Virology Lab |
| EPI_ISL_417366 | UW Virology Lab | UW Virology Lab |
| EPI_ISL_417367 | UW Virology Lab | UW Virology Lab |
| EPI_ISL_417368 | UW Virology Lab | UW Virology Lab |
| EPI_ISL_417369 | UW Virology Lab | UW Virology Lab |
| EPI_ISL_417370 | UW Virology Lab | UW Virology Lab |
| EPI_ISL_417371 | UW Virology Lab | UW Virology Lab |
| EPI_ISL_417372 | UW Virology Lab | UW Virology Lab |

| EPI_Accession_ID | Originating Laboratory | Submitting Laboratory |
|---|---|---|
| EPI_ISL_417373 | UW Virology Lab | UW Virology Lab |
| EPI_ISL_417374 | UW Virology Lab | UW Virology Lab |
| EPI_ISL_417375 | UW Virology Lab | UW Virology Lab |
| EPI_ISL_417376 | UW Virology Lab | UW Virology Lab |
| EPI_ISL_417377 | UW Virology Lab | UW Virology Lab |
| EPI_ISL_417378 | UW Virology Lab | UW Virology Lab |
| EPI_ISL_417380 | UW Virology Lab | UW Virology Lab |
| EPI_ISL_417381 | UW Virology Lab | UW Virology Lab |
| EPI_ISL_417382 | UW Virology Lab | UW Virology Lab |
| EPI_ISL_417383 | Centre for Infectious Diseases and Microbiology Public Health | NSW Health Pathology - Institute of Clinical Pathology and Medical Research; Westmead Hospital; University of Sydney |
| EPI_ISL_417384 | Centre for Infectious Diseases and Microbiology Public Health | NSW Health Pathology - Institute of Clinical Pathology and Medical Research; Westmead Hospital; University of Sydney |
| EPI_ISL_417385 | Centre for Infectious Diseases and Microbiology Public Health | NSW Health Pathology - Institute of Clinical Pathology and Medical Research; Westmead Hospital; University of Sydney |
| EPI_ISL_417386 | Centre for Infectious Diseases and Microbiology Public Health | NSW Health Pathology - Institute of Clinical Pathology and Medical Research; Westmead Hospital; University of Sydney |
| EPI_ISL_417387 | Centre for Infectious Diseases and Microbiology Public Health | NSW Health Pathology - Institute of Clinical Pathology and Medical Research; Westmead Hospital; University of Sydney |
| EPI_ISL_417388 | Centre for Infectious Diseases and Microbiology Public Health | NSW Health Pathology - Institute of Clinical Pathology and Medical Research; Westmead Hospital; University of Sydney |
| EPI_ISL_417389 | Centre for Infectious Diseases and Microbiology Public Health | NSW Health Pathology - Institute of Clinical Pathology and Medical Research; Westmead Hospital; University of Sydney |
| EPI_ISL_417390 | Centre for Infectious Diseases and Microbiology Public Health | NSW Health Pathology - Institute of Clinical Pathology and Medical Research; Westmead Hospital; University of Sydney |

| EPI_Accession_ID | Originating Laboratory | Submitting Laboratory |
|---|---|---|
| EPI_ISL_417391 | Centre for Infectious Diseases and Microbiology Public Health | NSW Health Pathology - Institute of Clinical Pathology and Medical Research; Westmead Hospital; University of Sydney |
| EPI_ISL_417392 | Centre for Infectious Diseases and Microbiology Public Health | NSW Health Pathology - Institute of Clinical Pathology and Medical Research; Westmead Hospital; University of Sydney |
| EPI_ISL_417393 | Centre for Infectious Diseases and Microbiology Public Health | NSW Health Pathology - Institute of Clinical Pathology and Medical Research; Westmead Hospital; University of Sydney |
| EPI_ISL_417394 | Centre for Infectious Diseases and Microbiology Public Health | NSW Health Pathology - Institute of Clinical Pathology and Medical Research; Westmead Hospital; University of Sydney |
| EPI_ISL_417395 | Centre for Infectious Diseases and Microbiology Public Health | NSW Health Pathology - Institute of Clinical Pathology and Medical Research; Westmead Hospital; University of Sydney |
| EPI_ISL_417396 | Centre for Infectious Diseases and Microbiology Public Health | NSW Health Pathology - Institute of Clinical Pathology and Medical Research; Westmead Hospital; University of Sydney |
| EPI_ISL_417397 | Centre for Infectious Diseases and Microbiology Public Health | NSW Health Pathology - Institute of Clinical Pathology and Medical Research; Westmead Hospital; University of Sydney |
| EPI_ISL_417398 | Centre for Infectious Diseases and Microbiology Public Health | NSW Health Pathology - Institute of Clinical Pathology and Medical Research; Westmead Hospital; University of Sydney |
| EPI_ISL_417399 | Centre for Infectious Diseases and Microbiology Public Health | NSW Health Pathology - Institute of Clinical Pathology and Medical Research; Westmead Hospital; University of Sydney |
| EPI_ISL_417400 | Centre for Infectious Diseases and Microbiology Public Health | NSW Health Pathology - Institute of Clinical Pathology and Medical Research; Westmead Hospital; University of Sydney |
| EPI_ISL_417401 | Centre for Infectious Diseases and Microbiology Public Health | NSW Health Pathology - Institute of Clinical Pathology and Medical Research; Westmead Hospital; University of Sydney |

| EPI_Accession_ID | Originating Laboratory | Submitting Laboratory |
|---|---|---|
| EPI_ISL_417402 | Centre for Infectious Diseases and Microbiology Public Health | NSW Health Pathology - Institute of Clinical Pathology and Medical Research; Westmead Hospital; University of Sydney |
| EPI_ISL_417403 | Centre for Infectious Diseases and Microbiology Public Health | NSW Health Pathology - Institute of Clinical Pathology and Medical Research; Westmead Hospital; University of Sydney |
| EPI_ISL_417404 | Centre for Infectious Diseases and Microbiology Public Health | NSW Health Pathology - Institute of Clinical Pathology and Medical Research; Westmead Hospital; University of Sydney |
| EPI_ISL_417405 | Centre for Infectious Diseases and Microbiology Public Health | NSW Health Pathology - Institute of Clinical Pathology and Medical Research; Westmead Hospital; University of Sydney |
| EPI_ISL_417406 | Centre for Infectious Diseases and Microbiology Public Health | NSW Health Pathology - Institute of Clinical Pathology and Medical Research; Westmead Hospital; University of Sydney |
| EPI_ISL_417407 | Centre for Infectious Diseases and Microbiology Public Health | NSW Health Pathology - Institute of Clinical Pathology and Medical Research; Westmead Hospital; University of Sydney |
| EPI_ISL_417408 | Centre for Infectious Diseases and Microbiology Public Health | NSW Health Pathology - Institute of Clinical Pathology and Medical Research; Westmead Hospital; University of Sydney |
| EPI_ISL_417409 | Centre for Infectious Diseases and Microbiology Public Health | NSW Health Pathology - Institute of Clinical Pathology and Medical Research; Westmead Hospital; University of Sydney |
| EPI_ISL_417410 | Centre for Infectious Diseases and Microbiology Public Health | NSW Health Pathology - Institute of Clinical Pathology and Medical Research; Westmead Hospital; University of Sydney |
| EPI_ISL_417411 | Centre for Infectious Diseases and Microbiology Public Health | NSW Health Pathology - Institute of Clinical Pathology and Medical Research; Westmead Hospital; University of Sydney |
| EPI_ISL_417412 | Centre for Infectious Diseases and Microbiology Public Health | NSW Health Pathology - Institute of Clinical Pathology and Medical Research; Westmead Hospital; University of Sydney |

| EPI_Accession_ID | Originating Laboratory | Submitting Laboratory |
|---|---|---|
| EPI_ISL_417418 | Laboratory of Molecular Virology International Center fro Genetic Engineering and Biotechnology (ICGEB) | ARGO Open Lab Platform for Genome sequencing |
| EPI_ISL_417419 | Laboratory of Molecular Virology International Center for Genetic Engineering and Biotechnology (ICGEB) | ARGO Open Lab Platform for Genome sequencing |
| EPI_ISL_417420 | Jiangxi province Center for Disease Control and Prevention | Jiangxi province Center for Disease Control and Prevention |
| EPI_ISL_417421 | Laboratory of Molecular Virology International Center for Genetic Engineering and Biotechnology (ICGEB) | ARGO Open Lab Platform for Genome sequencing |
| EPI_ISL_417422 | KU Leuven, Clinical and Epidemiological Virology | KU Leuven, Clinical and Epidemiological Virology |
| EPI_ISL_417423 | Laboratory of Molecular Virology International Center for Genetic Engineering and Biotechnology (ICGEB) | ARGO Open Lab Platform for Genome sequencing |
| EPI_ISL_417424 | KU Leuven, Clinical and Epidemiological Virology | KU Leuven, Clinical and Epidemiological Virology |
| EPI_ISL_417425 | KU Leuven, Clinical and Epidemiological Virology | KU Leuven, Clinical and Epidemiological Virology |
| EPI_ISL_417426 | KU Leuven, Clinical and Epidemiological Virology | KU Leuven, Clinical and Epidemiological Virology |
| EPI_ISL_417427 | KU Leuven, Clinical and Epidemiological Virology | KU Leuven, Clinical and Epidemiological Virology |
| EPI_ISL_417428 | KU Leuven, Clinical and Epidemiological Virology | KU Leuven, Clinical and Epidemiological Virology |
| EPI_ISL_417429 | KU Leuven, Clinical and Epidemiological Virology | KU Leuven, Clinical and Epidemiological Virology |
| EPI_ISL_417430 | KU Leuven, Clinical and Epidemiological Virology | KU Leuven, Clinical and Epidemiological Virology |
| EPI_ISL_417432 | UCD National Virus Reference Laboratory | Public Health Wales Microbiology Cardiff |
| EPI_ISL_417433 | Viral Respiratory Lab, National Institute for Biomedical Research (INRB) | Pathogen Sequencing Lab, National Institute for Biomedical Research (INRB) |
| EPI_ISL_417434 | Viral Respiratory Lab, National Institute for Biomedical Research (INRB) | Pathogen Sequencing Lab, National Institute for Biomedical Research (INRB) |
| EPI_ISL_417435 | Viral Respiratory Lab, National Institute for Biomedical Research (INRB) | Pathogen Sequencing Lab, National Institute for Biomedical Research (INRB) |
| EPI_ISL_417437 | Viral Respiratory Lab, National Institute for Biomedical Research (INRB) | Pathogen Sequencing Lab, National Institute for Biomedical Research (INRB) |
| EPI_ISL_417438 | Viral Respiratory Lab, National Institute for Biomedical Research (INRB) | Pathogen Sequencing Lab, National Institute for Biomedical Research (INRB) |
| EPI_ISL_417439 | Viral Respiratory Lab, National Institute for Biomedical Research (INRB) | Pathogen Sequencing Lab, National Institute for Biomedical Research (INRB) |
| EPI_ISL_417440 | Viral Respiratory Lab, National Institute for Biomedical Research (INRB) | Pathogen Sequencing Lab, National Institute for Biomedical Research (INRB) |

| EPI_Accession_ID | Originating Laboratory | Submitting Laboratory |
|---|---|---|
| EPI_ISL_417441 | Viral Respiratory Lab, National Institute for Biomedical Research (INRB) | Pathogen Sequencing Lab, National Institute for Biomedical Research (INRB) |
| EPI_ISL_417442 | Viral Respiratory Lab, National Institute for Biomedical Research (INRB) | Pathogen Sequencing Lab, National Institute for Biomedical Research (INRB) |
| EPI_ISL_417443 | State Key Laboratory for Emerging Infectious Diseases Department of Microbiology Li Ka Shing Faculty of Medicine The University of Hong Kong | State Key Laboratory for Emerging Infectious Diseases Department of Microbiology Li Ka Shing Faculty of Medicine The University of Hong Kong |
| EPI_ISL_417444 | unknown | Department of Healthcare Biotechnology |
| EPI_ISL_417445 | Laboratory of Infectious Diseases, Department of Biomedical and Clinical Sciences L. Sacco, University of Milan | Laboratory of Infectious Diseases, Department of Biomedical and Clinical Sciences L. Sacco, University of Milan |
| EPI_ISL_417447 | Laboratory of Infectious Diseases, Department of Biomedical and Clinical Sciences L. Sacco, University of Milan | Laboratory of Infectious Diseases, Department of Biomedical and Clinical Sciences L. Sacco, University of Milan |
| EPI_ISL_417448 | UW Virology Lab | UW Virology Lab |
| EPI_ISL_417449 | UW Virology Lab | UW Virology Lab |
| EPI_ISL_417450 | UW Virology Lab | UW Virology Lab |
| EPI_ISL_417451 | UW Virology Lab | UW Virology Lab |
| EPI_ISL_417452 | UW Virology Lab | UW Virology Lab |
| EPI_ISL_417453 | UW Virology Lab | UW Virology Lab |
| EPI_ISL_417454 | UW Virology Lab | UW Virology Lab |
| EPI_ISL_417455 | UW Virology Lab | UW Virology Lab |
| EPI_ISL_417456 | UW Virology Lab | UW Virology Lab |
| EPI_ISL_417457 | Center of Medical Microbiology, Virology, and Hospital Hygiene, University of Duesseldorf | Center of Medical Microbiology, Virology, and Hospital Hygiene, University of Duesseldorf |
| EPI_ISL_417458 | Center of Medical Microbiology, Virology, and Hospital Hygiene, University of Duesseldorf | Center of Medical Microbiology, Virology, and Hospital Hygiene, University of Duesseldorf |
| EPI_ISL_417459 | Center of Medical Microbiology, Virology, and Hospital Hygiene, University of Duesseldorf | Center of Medical Microbiology, Virology, and Hospital Hygiene, University of Duesseldorf |
| EPI_ISL_417460 | Center of Medical Microbiology, Virology, and Hospital Hygiene, University of Duesseldorf | Center of Medical Microbiology, Virology, and Hospital Hygiene, University of Duesseldorf |
| EPI_ISL_417461 | Center of Medical Microbiology, Virology, and Hospital Hygiene, University of Duesseldorf | Center of Medical Microbiology, Virology, and Hospital Hygiene, University of Duesseldorf |

| EPI_Accession_ID | Originating Laboratory | Submitting Laboratory |
|---|---|---|
| EPI_ISL_417462 | Center of Medical Microbiology, Virology, and Hospital Hygiene, University of Duesseldorf | Center of Medical Microbiology, Virology, and Hospital Hygiene, University of Duesseldorf |
| EPI_ISL_417463 | Center of Medical Microbiology, Virology, and Hospital Hygiene, University of Duesseldorf | Center of Medical Microbiology, Virology, and Hospital Hygiene, University of Duesseldorf |
| EPI_ISL_417464 | Center of Medical Microbiology, Virology, and Hospital Hygiene, University of Duesseldorf | Center of Medical Microbiology, Virology, and Hospital Hygiene, University of Duesseldorf |
| EPI_ISL_417465 | Center of Medical Microbiology, Virology, and Hospital Hygiene, University of Duesseldorf | Center of Medical Microbiology, Virology, and Hospital Hygiene, University of Duesseldorf |
| EPI_ISL_417466 | Center of Medical Microbiology, Virology, and Hospital Hygiene, University of Duesseldorf | Center of Medical Microbiology, Virology, and Hospital Hygiene, University of Duesseldorf |
| EPI_ISL_417467 | Center of Medical Microbiology, Virology, and Hospital Hygiene, University of Duesseldorf | Center of Medical Microbiology, Virology, and Hospital Hygiene, University of Duesseldorf |
| EPI_ISL_417468 | Center of Medical Microbiology, Virology, and Hospital Hygiene, University of Duesseldorf | Center of Medical Microbiology, Virology, and Hospital Hygiene, University of Duesseldorf |
| EPI_ISL_417469 | Minnesota Department of Health, Public Health Laboratory | Minnesota Department of Health, Public Health Laboratory |
| EPI_ISL_417470 | Minnesota Department of Health, Public Health Laboratory | Minnesota Department of Health, Public Health Laboratory |
| EPI_ISL_417471 | Minnesota Department of Health, Public Health Laboratory | Minnesota Department of Health, Public Health Laboratory |
| EPI_ISL_417472 | Minnesota Department of Health, Public Health Laboratory | Minnesota Department of Health, Public Health Laboratory |
| EPI_ISL_417473 | Minnesota Department of Health, Public Health Laboratory | Minnesota Department of Health, Public Health Laboratory |
| EPI_ISL_417474 | Minnesota Department of Health, Public Health Laboratory | Minnesota Department of Health, Public Health Laboratory |
| EPI_ISL_417475 | Minnesota Department of Health, Public Health Laboratory | Minnesota Department of Health, Public Health Laboratory |
| EPI_ISL_417476 | Minnesota Department of Health, Public Health Laboratory | Minnesota Department of Health, Public Health Laboratory |
| EPI_ISL_417477 | Minnesota Department of Health, Public Health Laboratory | Minnesota Department of Health, Public Health Laboratory |

| EPI_Accession_ID | Originating Laboratory | Submitting Laboratory |
|---|---|---|
| EPI_ISL_417478 | Minnesota Department of Health, Public Health Laboratory | Minnesota Department of Health, Public Health Laboratory |
| EPI_ISL_417479 | Minnesota Department of Health, Public Health Laboratory | Minnesota Department of Health, Public Health Laboratory |
| EPI_ISL_417480 | Minnesota Department of Health, Public Health Laboratory | Minnesota Department of Health, Public Health Laboratory |
| EPI_ISL_417481 | deCODE genetics | deCODE genetics |
| EPI_ISL_417482 | Institute of Microbiology, Universidad San Francisco de Quito | Institute of Microbiology, Universidad San Francisco de Quito |
| EPI_ISL_417483 | Oslo University Hospital, Department of Medical Microbiology | Norwegian Institute of Public Health |
| EPI_ISL_417484 | Oslo University Hospital, Department of Medical Microbiology | Norwegian Institute of Public Health, Department of Virology |
| EPI_ISL_417485 | University Hospital of Northern Norway, Department for Microbiology and Infectious Disease Control | Norwegian Institute of Public Health, Department of Virology |
| EPI_ISL_417486 | Hospital of Southern Norway - Kristiansand, Department of Medical Microbiology | Norwegian Institute of Public Health, Department of Virology |
| EPI_ISL_417488 | Oslo University Hospital, Department of Medical Microbiology | Norwegian Institute of Public Health, Department of Virology |
| EPI_ISL_417489 | Oslo University Hospital, Department of Medical Microbiology | Norwegian Institute of Public Health, Department of Virology |
| EPI_ISL_417490 | Oslo University Hospital, Department of Medical Microbiology | Norwegian Institute of Public Health, Department of Virology |
| EPI_ISL_417491 | Virology Laboratory, Department of Biomedical Sciences and Public Health, University Politecnica delle Marche | Virology and Legal Medicine Laboratories, Department of Biomedical Sciences and Public Health, University Politecnica delle Marche |
| EPI_ISL_417492 | Minnesota Department of Health, Public Health Laboratory | Minnesota Department of Health, Public Health Laboratory |
| EPI_ISL_417493 | Minnesota Department of Health, Public Health Laboratory | Minnesota Department of Health, Public Health Laboratory |
| EPI_ISL_417494 | Minnesota Department of Health, Public Health Laboratory | Minnesota Department of Health, Public Health Laboratory |

| EPI_Accession_ID | Originating Laboratory | Submitting Laboratory |
|---|---|---|
| EPI_ISL_417495 | Minnesota Department of Health, Public Health Laboratory | Minnesota Department of Health, Public Health Laboratory |
| EPI_ISL_417496 | Minnesota Department of Health, Public Health Laboratory | Minnesota Department of Health, Public Health Laboratory |
| EPI_ISL_417497 | Minnesota Department of Health, Public Health Laboratory | Minnesota Department of Health, Public Health Laboratory |
| EPI_ISL_417498 | Minnesota Department of Health, Public Health Laboratory | Minnesota Department of Health, Public Health Laboratory |
| EPI_ISL_417499 | Minnesota Department of Health, Public Health Laboratory | Minnesota Department of Health, Public Health Laboratory |
| EPI_ISL_417500 | Minnesota Department of Health, Public Health Laboratory | Minnesota Department of Health, Public Health Laboratory |
| EPI_ISL_417501 | Minnesota Department of Health, Public Health Laboratory | Minnesota Department of Health, Public Health Laboratory |
| EPI_ISL_417502 | Minnesota Department of Health, Public Health Laboratory | Minnesota Department of Health, Public Health Laboratory |
| EPI_ISL_417503 | Minnesota Department of Health, Public Health Laboratory | Minnesota Department of Health, Public Health Laboratory |
| EPI_ISL_417504 | University of Wisconsin-Madison AIDS Vaccine Research Laboratories | University of Wisconsin-Madison AIDS Vaccine Research Laboratories |
| EPI_ISL_417505 | University of Wisconsin-Madison AIDS Vaccine Research Laboratories | University of Wisconsin-Madison AIDS Vaccine Research Laboratories |
| EPI_ISL_417506 | University of Wisconsin-Madison AIDS Vaccine Research Laboratories | University of Wisconsin-Madison AIDS Vaccine Research Laboratories |
| EPI_ISL_417507 | University of Wisconsin-Madison AIDS Vaccine Research Laboratories | University of Wisconsin-Madison AIDS Vaccine Research Laboratories |
| EPI_ISL_417508 | University of Wisconsin-Madison AIDS Vaccine Research Laboratories | University of Wisconsin-Madison AIDS Vaccine Research Laboratories |
| EPI_ISL_417509 | University of Wisconsin-Madison AIDS Vaccine Research Laboratories | University of Wisconsin-Madison AIDS Vaccine Research Laboratories |
| EPI_ISL_417510 | University of Wisconsin-Madison AIDS Vaccine Research Laboratories | University of Wisconsin-Madison AIDS Vaccine Research Laboratories |

| EPI_Accession_ID | Originating Laboratory | Submitting Laboratory |
|---|---|---|
| EPI_ISL_417512 | University of Wisconsin-Madison AIDS Vaccine Research Laboratories | University of Wisconsin-Madison AIDS Vaccine Research Laboratories |
| EPI_ISL_417513 | University of Wisconsin-Madison AIDS Vaccine Research Laboratories | University of Wisconsin-Madison AIDS Vaccine Research Laboratories |
| EPI_ISL_417514 | University of Wisconsin-Madison AIDS Vaccine Research Laboratories | University of Wisconsin-Madison AIDS Vaccine Research Laboratories |
| EPI_ISL_417515 | University of Wisconsin-Madison AIDS Vaccine Research Laboratories | University of Wisconsin-Madison AIDS Vaccine Research Laboratories |
| EPI_ISL_417516 | University of Wisconsin-Madison AIDS Vaccine Research Laboratories | University of Wisconsin-Madison AIDS Vaccine Research Laboratories |
| EPI_ISL_417517 | University of Wisconsin-Madison AIDS Vaccine Research Laboratories | University of Wisconsin-Madison AIDS Vaccine Research Laboratories |
| EPI_ISL_417518 | Laboratory Medicine | Department of Laboratory Medicine, Lin-Kou Chang Gung Memorial Hospital, Taoyuan, Taiwan |
| EPI_ISL_417519 | Laboratory Medicine | Department of Laboratory Medicine, Lin-Kou Chang Gung Memorial Hospital, Taoyuan, Taiwan |
| EPI_ISL_417520 | Laboratory Medicine | Department of Laboratory Medicine, Lin-Kou Chang Gung Memorial Hospital, Taoyuan, Taiwan |
| EPI_ISL_417521 | Laboratory Medicine | Department of Laboratory Medicine, Lin-Kou Chang Gung Memorial Hospital, Taoyuan, Taiwan |
| EPI_ISL_417522 | Laboratory Medicine | Department of Laboratory Medicine, Lin-Kou Chang Gung Memorial Hospital, Taoyuan, Taiwan |
| EPI_ISL_417523 | Laboratory Medicine | Department of Laboratory Medicine, Lin-Kou Chang Gung Memorial Hospital, Taoyuan, Taiwan |
| EPI_ISL_417524 | Laboratory Medicine | Department of Laboratory Medicine, Lin-Kou Chang Gung Memorial Hospital, Taoyuan, Taiwan |
| EPI_ISL_417525 | Laboratory Medicine | Department of Laboratory Medicine, Lin-Kou Chang Gung Memorial Hospital, Taoyuan, Taiwan |
| EPI_ISL_417526 | Laboratoire Nationale de SantÃ©, Microbiology, Virology | Laboratoire Nationale de SantÃ©, Microbiology, Epidemiology and Microbial Genomics |
| EPI_ISL_417527 | Laboratoire Nationale de SantÃ©, Microbiology, Virology | Laboratoire Nationale de SantÃ©, Microbiology, Epidemiology and Microbial Genomics |

| EPI_Accession_ID | Originating Laboratory | Submitting Laboratory |
|---|---|---|
| EPI_ISL_417528 | Laboratoire Nationale de SantÃ©, Microbiology, Virology | Laboratoire Nationale de SantÃ©, Microbiology, Epidemiology and Microbial Genomics |
| EPI_ISL_417529 | Laboratoire Nationale de SantÃ©, Microbiology, Virology | Laboratoire Nationale de SantÃ©, Microbiology, Epidemiology and Microbial Genomics |
| EPI_ISL_417530 | Laboratoire Nationale de SantÃ©, Microbiology, Virology | Laboratoire Nationale de SantÃ©, Microbiology, Epidemiology and Microbial Genomics |
| EPI_ISL_417531 | Laboratoire Nationale de SantÃ©, Microbiology, Virology | Laboratoire Nationale de SantÃ©, Microbiology, Epidemiology and Microbial Genomics |
| EPI_ISL_417532 | Laboratoire Nationale de SantÃ©, Microbiology, Virology | Laboratoire Nationale de SantÃ©, Microbiology, Epidemiology and Microbial Genomics |
| EPI_ISL_417533 | Laboratoire Nationale de SantÃ©, Microbiology, Virology | Laboratoire Nationale de SantÃ©, Microbiology, Epidemiology and Microbial Genomics |
| EPI_ISL_417534 | Laboratoire Nationale de SantÃ©, Microbiology, Virology | Laboratoire Nationale de SantÃ©, Microbiology, Epidemiology and Microbial Genomics |
| EPI_ISL_417535 | deCODE genetics | deCODE genetics |
| EPI_ISL_417536 | deCODE genetics | deCODE genetics |
| EPI_ISL_417537 | deCODE genetics | deCODE genetics |
| EPI_ISL_417538 | deCODE genetics | deCODE genetics |
| EPI_ISL_417539 | deCODE genetics | deCODE genetics |
| EPI_ISL_417540 | deCODE genetics | deCODE genetics |
| EPI_ISL_417541 | deCODE genetics | deCODE genetics |
| EPI_ISL_417542 | deCODE genetics | deCODE genetics |
| EPI_ISL_417543 | deCODE genetics | deCODE genetics |
| EPI_ISL_417544 | deCODE genetics | deCODE genetics |
| EPI_ISL_417545 | deCODE genetics | deCODE genetics |
| EPI_ISL_417546 | deCODE genetics | deCODE genetics |
| EPI_ISL_417547 | deCODE genetics | deCODE genetics |
| EPI_ISL_417548 | deCODE genetics | deCODE genetics |
| EPI_ISL_417549 | deCODE genetics | deCODE genetics |
| EPI_ISL_417550 | The National University Hospital of Iceland | deCODE genetics |
| EPI_ISL_417551 | The National University Hospital of Iceland | deCODE genetics |
| EPI_ISL_417552 | deCODE genetics | deCODE genetics |
| EPI_ISL_417553 | The National University Hospital of Iceland | deCODE genetics |

| EPI_Accession_ID | Originating Laboratory | Submitting Laboratory |
|---|---|---|
| EPI_ISL_417554 | The National University Hospital of Iceland | deCODE genetics |
| EPI_ISL_417555 | The National University Hospital of Iceland | deCODE genetics |
| EPI_ISL_417556 | The National University Hospital of Iceland | deCODE genetics |
| EPI_ISL_417557 | The National University Hospital of Iceland | deCODE genetics |
| EPI_ISL_417558 | The National University Hospital of Iceland | deCODE genetics |
| EPI_ISL_417559 | The National University Hospital of Iceland | deCODE genetics |
| EPI_ISL_417560 | The National University Hospital of Iceland | deCODE genetics |
| EPI_ISL_417561 | The National University Hospital of Iceland | deCODE genetics |
| EPI_ISL_417562 | The National University Hospital of Iceland | deCODE genetics |
| EPI_ISL_417563 | The National University Hospital of Iceland | deCODE genetics |
| EPI_ISL_417564 | The National University Hospital of Iceland | deCODE genetics |
| EPI_ISL_417565 | The National University Hospital of Iceland | deCODE genetics |
| EPI_ISL_417566 | The National University Hospital of Iceland | deCODE genetics |
| EPI_ISL_417567 | The National University Hospital of Iceland | deCODE genetics |
| EPI_ISL_417568 | The National University Hospital of Iceland | deCODE genetics |
| EPI_ISL_417569 | The National University Hospital of Iceland | deCODE genetics |
| EPI_ISL_417570 | The National University Hospital of Iceland | deCODE genetics |
| EPI_ISL_417571 | The National University Hospital of Iceland | deCODE genetics |
| EPI_ISL_417572 | The National University Hospital of Iceland | deCODE genetics |
| EPI_ISL_417573 | The National University Hospital of Iceland | deCODE genetics |
| EPI_ISL_417574 | The National University Hospital of Iceland | deCODE genetics |
| EPI_ISL_417575 | The National University Hospital of Iceland | deCODE genetics |
| EPI_ISL_417576 | The National University Hospital of Iceland | deCODE genetics |
| EPI_ISL_417577 | The National University Hospital of Iceland | deCODE genetics |
| EPI_ISL_417578 | The National University Hospital of Iceland | deCODE genetics |
| EPI_ISL_417579 | The National University Hospital of Iceland | deCODE genetics |
| EPI_ISL_417580 | The National University Hospital of Iceland | deCODE genetics |
| EPI_ISL_417581 | The National University Hospital of Iceland | deCODE genetics |
| EPI_ISL_417582 | The National University Hospital of Iceland | deCODE genetics |
| EPI_ISL_417583 | The National University Hospital of Iceland | deCODE genetics |
| EPI_ISL_417584 | The National University Hospital of Iceland | deCODE genetics |
| EPI_ISL_417585 | The National University Hospital of Iceland | deCODE genetics |
| EPI_ISL_417586 | The National University Hospital of Iceland | deCODE genetics |

| EPI_Accession_ID | Originating Laboratory | Submitting Laboratory |
|---|---|---|
| EPI_ISL_417587 | The National University Hospital of Iceland | deCODE genetics |
| EPI_ISL_417588 | The National University Hospital of Iceland | deCODE genetics |
| EPI_ISL_417589 | The National University Hospital of Iceland | deCODE genetics |
| EPI_ISL_417590 | deCODE genetics | deCODE genetics |
| EPI_ISL_417591 | The National University Hospital of Iceland | deCODE genetics |
| EPI_ISL_417592 | The National University Hospital of Iceland | deCODE genetics |
| EPI_ISL_417593 | The National University Hospital of Iceland | deCODE genetics |
| EPI_ISL_417594 | The National University Hospital of Iceland | deCODE genetics |
| EPI_ISL_417595 | The National University Hospital of Iceland | deCODE genetics |
| EPI_ISL_417596 | The National University Hospital of Iceland | deCODE genetics |
| EPI_ISL_417597 | The National University Hospital of Iceland | deCODE genetics |
| EPI_ISL_417598 | The National University Hospital of Iceland | deCODE genetics |
| EPI_ISL_417599 | The National University Hospital of Iceland | deCODE genetics |
| EPI_ISL_417600 | The National University Hospital of Iceland | deCODE genetics |
| EPI_ISL_417601 | The National University Hospital of Iceland | deCODE genetics |
| EPI_ISL_417603 | The National University Hospital of Iceland | deCODE genetics |
| EPI_ISL_417604 | The National University Hospital of Iceland | deCODE genetics |
| EPI_ISL_417605 | The National University Hospital of Iceland | deCODE genetics |
| EPI_ISL_417606 | The National University Hospital of Iceland | deCODE genetics |
| EPI_ISL_417607 | The National University Hospital of Iceland | deCODE genetics |
| EPI_ISL_417608 | The National University Hospital of Iceland | deCODE genetics |
| EPI_ISL_417609 | The National University Hospital of Iceland | deCODE genetics |
| EPI_ISL_417610 | The National University Hospital of Iceland | deCODE genetics |
| EPI_ISL_417611 | The National University Hospital of Iceland | deCODE genetics |
| EPI_ISL_417612 | The National University Hospital of Iceland | deCODE genetics |
| EPI_ISL_417613 | The National University Hospital of Iceland | deCODE genetics |
| EPI_ISL_417614 | The National University Hospital of Iceland | deCODE genetics |
| EPI_ISL_417615 | The National University Hospital of Iceland | deCODE genetics |
| EPI_ISL_417617 | The National University Hospital of Iceland | deCODE genetics |
| EPI_ISL_417618 | deCODE genetics | deCODE genetics |
| EPI_ISL_417619 | The National University Hospital of Iceland | deCODE genetics |
| EPI_ISL_417620 | The National University Hospital of Iceland | deCODE genetics |
| EPI_ISL_417621 | The National University Hospital of Iceland | deCODE genetics |

| EPI_Accession_ID | Originating Laboratory | Submitting Laboratory |
|---|---|---|
| EPI_ISL_417622 | The National University Hospital of Iceland | deCODE genetics |
| EPI_ISL_417623 | The National University Hospital of Iceland | deCODE genetics |
| EPI_ISL_417624 | The National University Hospital of Iceland | deCODE genetics |
| EPI_ISL_417625 | The National University Hospital of Iceland | deCODE genetics |
| EPI_ISL_417626 | The National University Hospital of Iceland | deCODE genetics |
| EPI_ISL_417627 | The National University Hospital of Iceland | deCODE genetics |
| EPI_ISL_417628 | The National University Hospital of Iceland | deCODE genetics |
| EPI_ISL_417629 | The National University Hospital of Iceland | deCODE genetics |
| EPI_ISL_417630 | The National University Hospital of Iceland | deCODE genetics |
| EPI_ISL_417631 | The National University Hospital of Iceland | deCODE genetics |
| EPI_ISL_417632 | The National University Hospital of Iceland | deCODE genetics |
| EPI_ISL_417633 | The National University Hospital of Iceland | deCODE genetics |
| EPI_ISL_417634 | The National University Hospital of Iceland | deCODE genetics |
| EPI_ISL_417635 | The National University Hospital of Iceland | deCODE genetics |
| EPI_ISL_417636 | The National University Hospital of Iceland | deCODE genetics |
| EPI_ISL_417637 | The National University Hospital of Iceland | deCODE genetics |
| EPI_ISL_417638 | The National University Hospital of Iceland | deCODE genetics |
| EPI_ISL_417639 | The National University Hospital of Iceland | deCODE genetics |
| EPI_ISL_417640 | The National University Hospital of Iceland | deCODE genetics |
| EPI_ISL_417641 | The National University Hospital of Iceland | deCODE genetics |
| EPI_ISL_417642 | The National University Hospital of Iceland | deCODE genetics |
| EPI_ISL_417643 | The National University Hospital of Iceland | deCODE genetics |
| EPI_ISL_417644 | The National University Hospital of Iceland | deCODE genetics |
| EPI_ISL_417645 | The National University Hospital of Iceland | deCODE genetics |
| EPI_ISL_417646 | The National University Hospital of Iceland | deCODE genetics |
| EPI_ISL_417647 | The National University Hospital of Iceland | deCODE genetics |
| EPI_ISL_417648 | The National University Hospital of Iceland | deCODE genetics |
| EPI_ISL_417649 | The National University Hospital of Iceland | deCODE genetics |
| EPI_ISL_417650 | The National University Hospital of Iceland | deCODE genetics |
| EPI_ISL_417651 | The National University Hospital of Iceland | deCODE genetics |
| EPI_ISL_417652 | The National University Hospital of Iceland | deCODE genetics |
| EPI_ISL_417653 | The National University Hospital of Iceland | deCODE genetics |
| EPI_ISL_417654 | The National University Hospital of Iceland | deCODE genetics |

| EPI_Accession_ID | Originating Laboratory | Submitting Laboratory |
|---|---|---|
| EPI_ISL_417655 | deCODE genetics | deCODE genetics |
| EPI_ISL_417656 | deCODE genetics | deCODE genetics |
| EPI_ISL_417657 | deCODE genetics | deCODE genetics |
| EPI_ISL_417658 | deCODE genetics | deCODE genetics |
| EPI_ISL_417659 | deCODE genetics | deCODE genetics |
| EPI_ISL_417660 | deCODE genetics | deCODE genetics |
| EPI_ISL_417661 | deCODE genetics | deCODE genetics |
| EPI_ISL_417662 | deCODE genetics | deCODE genetics |
| EPI_ISL_417663 | deCODE genetics | deCODE genetics |
| EPI_ISL_417664 | deCODE genetics | deCODE genetics |
| EPI_ISL_417665 | deCODE genetics | deCODE genetics |
| EPI_ISL_417666 | deCODE genetics | deCODE genetics |
| EPI_ISL_417667 | deCODE genetics | deCODE genetics |
| EPI_ISL_417668 | deCODE genetics | deCODE genetics |
| EPI_ISL_417669 | deCODE genetics | deCODE genetics |
| EPI_ISL_417670 | deCODE genetics | deCODE genetics |
| EPI_ISL_417672 | deCODE genetics | deCODE genetics |
| EPI_ISL_417673 | The National University Hospital of Iceland | deCODE genetics |
| EPI_ISL_417674 | The National University Hospital of Iceland | deCODE genetics |
| EPI_ISL_417675 | The National University Hospital of Iceland | deCODE genetics |
| EPI_ISL_417676 | deCODE genetics | deCODE genetics |
| EPI_ISL_417677 | The National University Hospital of Iceland | deCODE genetics |
| EPI_ISL_417678 | The National University Hospital of Iceland | deCODE genetics |
| EPI_ISL_417679 | The National University Hospital of Iceland | deCODE genetics |
| EPI_ISL_417680 | The National University Hospital of Iceland | deCODE genetics |
| EPI_ISL_417681 | The National University Hospital of Iceland | deCODE genetics |
| EPI_ISL_417682 | The National University Hospital of Iceland | deCODE genetics |
| EPI_ISL_417683 | The National University Hospital of Iceland | deCODE genetics |
| EPI_ISL_417684 | The National University Hospital of Iceland | deCODE genetics |
| EPI_ISL_417685 | The National University Hospital of Iceland | deCODE genetics |
| EPI_ISL_417687 | The National University Hospital of Iceland | deCODE genetics |
| EPI_ISL_417688 | The National University Hospital of Iceland | deCODE genetics |
| EPI_ISL_417689 | The National University Hospital of Iceland | deCODE genetics |

| EPI_Accession_ID | Originating Laboratory | Submitting Laboratory |
|---|---|---|
| EPI_ISL_417690 | The National University Hospital of Iceland | deCODE genetics |
| EPI_ISL_417691 | The National University Hospital of Iceland | deCODE genetics |
| EPI_ISL_417692 | The National University Hospital of Iceland | deCODE genetics |
| EPI_ISL_417693 | The National University Hospital of Iceland | deCODE genetics |
| EPI_ISL_417694 | The National University Hospital of Iceland | deCODE genetics |
| EPI_ISL_417695 | The National University Hospital of Iceland | deCODE genetics |
| EPI_ISL_417696 | The National University Hospital of Iceland | deCODE genetics |
| EPI_ISL_417697 | The National University Hospital of Iceland | deCODE genetics |
| EPI_ISL_417698 | The National University Hospital of Iceland | deCODE genetics |
| EPI_ISL_417699 | The National University Hospital of Iceland | deCODE genetics |
| EPI_ISL_417700 | The National University Hospital of Iceland | deCODE genetics |
| EPI_ISL_417701 | The National University Hospital of Iceland | deCODE genetics |
| EPI_ISL_417702 | The National University Hospital of Iceland | deCODE genetics |
| EPI_ISL_417703 | The National University Hospital of Iceland | deCODE genetics |
| EPI_ISL_417704 | The National University Hospital of Iceland | deCODE genetics |
| EPI_ISL_417705 | The National University Hospital of Iceland | deCODE genetics |
| EPI_ISL_417706 | The National University Hospital of Iceland | deCODE genetics |
| EPI_ISL_417707 | The National University Hospital of Iceland | deCODE genetics |
| EPI_ISL_417708 | The National University Hospital of Iceland | deCODE genetics |
| EPI_ISL_417709 | The National University Hospital of Iceland | deCODE genetics |
| EPI_ISL_417710 | The National University Hospital of Iceland | deCODE genetics |
| EPI_ISL_417711 | The National University Hospital of Iceland | deCODE genetics |
| EPI_ISL_417712 | The National University Hospital of Iceland | deCODE genetics |
| EPI_ISL_417713 | The National University Hospital of Iceland | deCODE genetics |
| EPI_ISL_417714 | The National University Hospital of Iceland | deCODE genetics |
| EPI_ISL_417715 | The National University Hospital of Iceland | deCODE genetics |
| EPI_ISL_417716 | The National University Hospital of Iceland | deCODE genetics |
| EPI_ISL_417717 | The National University Hospital of Iceland | deCODE genetics |
| EPI_ISL_417718 | The National University Hospital of Iceland | deCODE genetics |
| EPI_ISL_417720 | The National University Hospital of Iceland | deCODE genetics |
| EPI_ISL_417721 | The National University Hospital of Iceland | deCODE genetics |
| EPI_ISL_417722 | The National University Hospital of Iceland | deCODE genetics |
| EPI_ISL_417723 | The National University Hospital of Iceland | deCODE genetics |

| EPI_Accession_ID | Originating Laboratory | Submitting Laboratory |
|---|---|---|
| EPI_ISL_417724 | The National University Hospital of Iceland | deCODE genetics |
| EPI_ISL_417725 | The National University Hospital of Iceland | deCODE genetics |
| EPI_ISL_417726 | The National University Hospital of Iceland | deCODE genetics |
| EPI_ISL_417727 | The National University Hospital of Iceland | deCODE genetics |
| EPI_ISL_417728 | The National University Hospital of Iceland | deCODE genetics |
| EPI_ISL_417729 | The National University Hospital of Iceland | deCODE genetics |
| EPI_ISL_417730 | The National University Hospital of Iceland | deCODE genetics |
| EPI_ISL_417731 | The National University Hospital of Iceland | deCODE genetics |
| EPI_ISL_417732 | The National University Hospital of Iceland | deCODE genetics |
| EPI_ISL_417733 | The National University Hospital of Iceland | deCODE genetics |
| EPI_ISL_417734 | The National University Hospital of Iceland | deCODE genetics |
| EPI_ISL_417735 | The National University Hospital of Iceland | deCODE genetics |
| EPI_ISL_417736 | The National University Hospital of Iceland | deCODE genetics |
| EPI_ISL_417737 | The National University Hospital of Iceland | deCODE genetics |
| EPI_ISL_417738 | The National University Hospital of Iceland | deCODE genetics |
| EPI_ISL_417739 | The National University Hospital of Iceland | deCODE genetics |
| EPI_ISL_417740 | The National University Hospital of Iceland | deCODE genetics |
| EPI_ISL_417741 | The National University Hospital of Iceland | deCODE genetics |
| EPI_ISL_417742 | The National University Hospital of Iceland | deCODE genetics |
| EPI_ISL_417743 | The National University Hospital of Iceland | deCODE genetics |
| EPI_ISL_417744 | The National University Hospital of Iceland | deCODE genetics |
| EPI_ISL_417745 | The National University Hospital of Iceland | deCODE genetics |
| EPI_ISL_417746 | The National University Hospital of Iceland | deCODE genetics |
| EPI_ISL_417747 | The National University Hospital of Iceland | deCODE genetics |
| EPI_ISL_417748 | The National University Hospital of Iceland | deCODE genetics |
| EPI_ISL_417749 | The National University Hospital of Iceland | deCODE genetics |
| EPI_ISL_417750 | The National University Hospital of Iceland | deCODE genetics |
| EPI_ISL_417751 | The National University Hospital of Iceland | deCODE genetics |
| EPI_ISL_417752 | The National University Hospital of Iceland | deCODE genetics |
| EPI_ISL_417753 | The National University Hospital of Iceland | deCODE genetics |
| EPI_ISL_417754 | The National University Hospital of Iceland | deCODE genetics |
| EPI_ISL_417755 | The National University Hospital of Iceland | deCODE genetics |
| EPI_ISL_417757 | The National University Hospital of Iceland | deCODE genetics |

| EPI_Accession_ID | Originating Laboratory | Submitting Laboratory |
|---|---|---|
| EPI_ISL_417758 | The National University Hospital of Iceland | deCODE genetics |
| EPI_ISL_417759 | The National University Hospital of Iceland | deCODE genetics |
| EPI_ISL_417760 | The National University Hospital of Iceland | deCODE genetics |
| EPI_ISL_417761 | The National University Hospital of Iceland | deCODE genetics |
| EPI_ISL_417762 | The National University Hospital of Iceland | deCODE genetics |
| EPI_ISL_417763 | The National University Hospital of Iceland | deCODE genetics |
| EPI_ISL_417764 | The National University Hospital of Iceland | deCODE genetics |
| EPI_ISL_417765 | The National University Hospital of Iceland | deCODE genetics |
| EPI_ISL_417766 | The National University Hospital of Iceland | deCODE genetics |
| EPI_ISL_417767 | The National University Hospital of Iceland | deCODE genetics |
| EPI_ISL_417768 | The National University Hospital of Iceland | deCODE genetics |
| EPI_ISL_417769 | The National University Hospital of Iceland | deCODE genetics |
| EPI_ISL_417770 | The National University Hospital of Iceland | deCODE genetics |
| EPI_ISL_417771 | The National University Hospital of Iceland | deCODE genetics |
| EPI_ISL_417772 | The National University Hospital of Iceland | deCODE genetics |
| EPI_ISL_417773 | The National University Hospital of Iceland | deCODE genetics |
| EPI_ISL_417774 | The National University Hospital of Iceland | deCODE genetics |
| EPI_ISL_417775 | The National University Hospital of Iceland | deCODE genetics |
| EPI_ISL_417776 | The National University Hospital of Iceland | deCODE genetics |
| EPI_ISL_417777 | The National University Hospital of Iceland | deCODE genetics |
| EPI_ISL_417779 | The National University Hospital of Iceland | deCODE genetics |
| EPI_ISL_417780 | The National University Hospital of Iceland | deCODE genetics |
| EPI_ISL_417781 | The National University Hospital of Iceland | deCODE genetics |
| EPI_ISL_417782 | The National University Hospital of Iceland | deCODE genetics |
| EPI_ISL_417783 | The National University Hospital of Iceland | deCODE genetics |
| EPI_ISL_417784 | The National University Hospital of Iceland | deCODE genetics |
| EPI_ISL_417785 | The National University Hospital of Iceland | deCODE genetics |
| EPI_ISL_417786 | The National University Hospital of Iceland | deCODE genetics |
| EPI_ISL_417787 | The National University Hospital of Iceland | deCODE genetics |
| EPI_ISL_417788 | The National University Hospital of Iceland | deCODE genetics |
| EPI_ISL_417789 | The National University Hospital of Iceland | deCODE genetics |
| EPI_ISL_417790 | The National University Hospital of Iceland | deCODE genetics |
| EPI_ISL_417791 | The National University Hospital of Iceland | deCODE genetics |

| EPI_Accession_ID | Originating Laboratory | Submitting Laboratory |
|---|---|---|
| EPI_ISL_417792 | The National University Hospital of Iceland | deCODE genetics |
| EPI_ISL_417793 | The National University Hospital of Iceland | deCODE genetics |
| EPI_ISL_417795 | The National University Hospital of Iceland | deCODE genetics |
| EPI_ISL_417796 | The National University Hospital of Iceland | deCODE genetics |
| EPI_ISL_417797 | The National University Hospital of Iceland | deCODE genetics |
| EPI_ISL_417798 | The National University Hospital of Iceland | deCODE genetics |
| EPI_ISL_417799 | The National University Hospital of Iceland | deCODE genetics |
| EPI_ISL_417800 | The National University Hospital of Iceland | deCODE genetics |
| EPI_ISL_417801 | The National University Hospital of Iceland | deCODE genetics |
| EPI_ISL_417802 | The National University Hospital of Iceland | deCODE genetics |
| EPI_ISL_417803 | The National University Hospital of Iceland | deCODE genetics |
| EPI_ISL_417804 | The National University Hospital of Iceland | deCODE genetics |
| EPI_ISL_417805 | The National University Hospital of Iceland | deCODE genetics |
| EPI_ISL_417806 | The National University Hospital of Iceland | deCODE genetics |
| EPI_ISL_417807 | The National University Hospital of Iceland | deCODE genetics |
| EPI_ISL_417808 | The National University Hospital of Iceland | deCODE genetics |
| EPI_ISL_417809 | The National University Hospital of Iceland | deCODE genetics |
| EPI_ISL_417810 | The National University Hospital of Iceland | deCODE genetics |
| EPI_ISL_417811 | The National University Hospital of Iceland | deCODE genetics |
| EPI_ISL_417812 | The National University Hospital of Iceland | deCODE genetics |
| EPI_ISL_417813 | The National University Hospital of Iceland | deCODE genetics |
| EPI_ISL_417814 | The National University Hospital of Iceland | deCODE genetics |
| EPI_ISL_417815 | The National University Hospital of Iceland | deCODE genetics |
| EPI_ISL_417816 | The National University Hospital of Iceland | deCODE genetics |
| EPI_ISL_417817 | The National University Hospital of Iceland | deCODE genetics |
| EPI_ISL_417818 | The National University Hospital of Iceland | deCODE genetics |
| EPI_ISL_417819 | The National University Hospital of Iceland | deCODE genetics |
| EPI_ISL_417820 | The National University Hospital of Iceland | deCODE genetics |
| EPI_ISL_417821 | The National University Hospital of Iceland | deCODE genetics |
| EPI_ISL_417822 | The National University Hospital of Iceland | deCODE genetics |
| EPI_ISL_417823 | The National University Hospital of Iceland | deCODE genetics |
| EPI_ISL_417824 | The National University Hospital of Iceland | deCODE genetics |
| EPI_ISL_417825 | The National University Hospital of Iceland | deCODE genetics |

| EPI_Accession_ID | Originating Laboratory | Submitting Laboratory |
|---|---|---|
| EPI_ISL_417826 | The National University Hospital of Iceland | deCODE genetics |
| EPI_ISL_417827 | The National University Hospital of Iceland | deCODE genetics |
| EPI_ISL_417828 | The National University Hospital of Iceland | deCODE genetics |
| EPI_ISL_417829 | The National University Hospital of Iceland | deCODE genetics |
| EPI_ISL_417830 | The National University Hospital of Iceland | deCODE genetics |
| EPI_ISL_417831 | The National University Hospital of Iceland | deCODE genetics |
| EPI_ISL_417832 | The National University Hospital of Iceland | deCODE genetics |
| EPI_ISL_417833 | The National University Hospital of Iceland | deCODE genetics |
| EPI_ISL_417834 | The National University Hospital of Iceland | deCODE genetics |
| EPI_ISL_417835 | The National University Hospital of Iceland | deCODE genetics |
| EPI_ISL_417836 | The National University Hospital of Iceland | deCODE genetics |
| EPI_ISL_417837 | The National University Hospital of Iceland | deCODE genetics |
| EPI_ISL_417838 | The National University Hospital of Iceland | deCODE genetics |
| EPI_ISL_417839 | The National University Hospital of Iceland | deCODE genetics |
| EPI_ISL_417840 | The National University Hospital of Iceland | deCODE genetics |
| EPI_ISL_417841 | The National University Hospital of Iceland | deCODE genetics |
| EPI_ISL_417842 | The National University Hospital of Iceland | deCODE genetics |
| EPI_ISL_417843 | The National University Hospital of Iceland | deCODE genetics |
| EPI_ISL_417844 | The National University Hospital of Iceland | deCODE genetics |
| EPI_ISL_417845 | The National University Hospital of Iceland | deCODE genetics |
| EPI_ISL_417846 | The National University Hospital of Iceland | deCODE genetics |
| EPI_ISL_417847 | The National University Hospital of Iceland | deCODE genetics |
| EPI_ISL_417848 | The National University Hospital of Iceland | deCODE genetics |
| EPI_ISL_417849 | The National University Hospital of Iceland | deCODE genetics |
| EPI_ISL_417850 | The National University Hospital of Iceland | deCODE genetics |
| EPI_ISL_417851 | The National University Hospital of Iceland | deCODE genetics |
| EPI_ISL_417852 | The National University Hospital of Iceland | deCODE genetics |
| EPI_ISL_417853 | The National University Hospital of Iceland | deCODE genetics |
| EPI_ISL_417854 | The National University Hospital of Iceland | deCODE genetics |
| EPI_ISL_417855 | The National University Hospital of Iceland | deCODE genetics |
| EPI_ISL_417856 | The National University Hospital of Iceland | deCODE genetics |
| EPI_ISL_417857 | The National University Hospital of Iceland | deCODE genetics |
| EPI_ISL_417860 | The National University Hospital of Iceland | deCODE genetics |

| EPI_Accession_ID | Originating Laboratory | Submitting Laboratory |
|---|---|---|
| EPI_ISL_417861 | The National University Hospital of Iceland | deCODE genetics |
| EPI_ISL_417862 | The National University Hospital of Iceland | deCODE genetics |
| EPI_ISL_417863 | The National University Hospital of Iceland | deCODE genetics |
| EPI_ISL_417864 | The National University Hospital of Iceland | deCODE genetics |
| EPI_ISL_417865 | The National University Hospital of Iceland | deCODE genetics |
| EPI_ISL_417866 | The National University Hospital of Iceland | deCODE genetics |
| EPI_ISL_417867 | The National University Hospital of Iceland | deCODE genetics |
| EPI_ISL_417868 | The National University Hospital of Iceland | deCODE genetics |
| EPI_ISL_417870 | The National University Hospital of Iceland | deCODE genetics |
| EPI_ISL_417871 | The National University Hospital of Iceland | deCODE genetics |
| EPI_ISL_417872 | The National University Hospital of Iceland | deCODE genetics |
| EPI_ISL_417873 | The National University Hospital of Iceland | deCODE genetics |
| EPI_ISL_417874 | The National University Hospital of Iceland | deCODE genetics |
| EPI_ISL_417875 | The National University Hospital of Iceland | deCODE genetics |
| EPI_ISL_417876 | The National University Hospital of Iceland | deCODE genetics |
| EPI_ISL_417877 | Institute of Virology, Biomedical Research Center of the Slovak Academy of Sciences, Bratislava; Public Health Authority of the Slovak Republic, Bratislava | Institute of Virology, Biomedical Research Center of the Slovak Academy of Sciences, Bratislava; Comenius University Science Park, Bratislava |
| EPI_ISL_417878 | Institute of Virology, Biomedical Research Center of the Slovak Academy of Sciences, Bratislava; Public Health Authority of the Slovak Republic, Bratislava | Institute of Virology, Biomedical Research Center of the Slovak Academy of Sciences, Bratislava; Comenius University Science Park, Bratislava |
| EPI_ISL_417879 | Institute of Virology, Biomedical Research Center of the Slovak Academy of Sciences, Bratislava; Public Health Authority of the Slovak Republic, Bratislava | Institute of Virology, Biomedical Research Center of the Slovak Academy of Sciences, Bratislava; Comenius University Science Park, Bratislava |
| EPI_ISL_417880 | Institute of Virology, Biomedical Research Center of the Slovak Academy of Sciences, Bratislava; Public Health Authority of the Slovak Republic, Bratislava | Institute of Virology, Biomedical Research Center of the Slovak Academy of Sciences, Bratislava; Comenius University Science Park, Bratislava |
| EPI_ISL_417917 | Department of Medical Microbiology, University Malaya Medical Centre | Department of Medical Microbiology |
| EPI_ISL_417918 | Department of Medical Microbiology, University Malaya Medical Centre | Department of Medical Microbiology, Faculty of Medicine, University of Malaya |
| EPI_ISL_417921 | INMI Lazzaro Spallanzani IRCCS | Laboratory of Virology, INMI Lazzaro Spallanzani IRCCS |
| EPI_ISL_417922 | INMI Lazzaro Spallanzani IRCCS | Laboratory of Virology, INMI Lazzaro Spallanzani IRCCS |

| EPI_Accession_ID | Originating Laboratory | Submitting Laboratory |
|---|---|---|
| EPI_ISL_417923 | INMI Lazzaro Spallanzani IRCCS | Laboratory of Virology, INMI Lazzaro Spallanzani IRCCS |
| EPI_ISL_417924 | SecretarÃ-a de Salud MedellÃ-n | Instituto Nacional de Salud, Universidad Cooperativa de Colombia, Instituto Alexander von Humboldt, Imperial College-London, London School of Hygiene & Tropical Medicine |
| EPI_ISL_417925 | LaboratÃ³rio Simili | Bioinformatics Laboratory / LNCC |
| EPI_ISL_417926 | LaboratÃ³rio Simili | Bioinformatics Laboratory / LNCC |
| EPI_ISL_417928 | LaboratÃ³rio Hermes Pardini | Bioinformatics Laboratory - LNCC |
| EPI_ISL_417929 | LaboratÃ³rio Hermes Pardini | Bioinformatics Laboratory - LNCC |
| EPI_ISL_417930 | LaboratÃ³rio Hermes Pardini | Bioinformatics Laboratory - LNCC |
| EPI_ISL_417931 | UCSF Clinical Microbiology Laboratory | Chan-Zuckerberg Biohub |
| EPI_ISL_417932 | UCSF Clinical Microbiology Laboratory | Chan-Zuckerberg Biohub |
| EPI_ISL_417933 | UCSF Clinical Microbiology Laboratory | Chan-Zuckerberg Biohub |
| EPI_ISL_417934 | LaboratÃ³rio Hermes Pardini | Bioinformatics Laboratory - LNCC |
| EPI_ISL_417935 | UCSF Clinical Microbiology Laboratory | Chan-Zuckerberg Biohub |
| EPI_ISL_417936 | LaboratÃ³rio Hermes Pardini | Bioinformatics Laboratory - LNCC |
| EPI_ISL_417937 | UCSF Clinical Microbiology Laboratory | Chan-Zuckerberg Biohub |
| EPI_ISL_417938 | UCSF Clinical Microbiology Laboratory | Chan-Zuckerberg Biohub |
| EPI_ISL_417939 | UCSF Clinical Microbiology Laboratory | Chan-Zuckerberg Biohub |
| EPI_ISL_417941 | Viral Respiratory Lab, National Institute for Biomedical Research (INRB) | Pathogen Sequencing Lab, National Institute for Biomedical Research (INRB) |
| EPI_ISL_417942 | Viral Respiratory Lab, National Institute for Biomedical Research (INRB) | Pathogen Sequencing Lab, National Institute for Biomedical Research (INRB) |
| EPI_ISL_417944 | Viral Respiratory Lab, National Institute for Biomedical Research (INRB) | Pathogen Sequencing Lab, National Institute for Biomedical Research (INRB) |
| EPI_ISL_417946 | Viral Respiratory Lab, National Institute for Biomedical Research (INRB) | Pathogen Sequencing Lab, National Institute for Biomedical Research (INRB) |
| EPI_ISL_417947 | Viral Respiratory Lab, National Institute for Biomedical Research (INRB) | Pathogen Sequencing Lab, National Institute for Biomedical Research (INRB) |
| EPI_ISL_417948 | Viral Respiratory Lab, National Institute for Biomedical Research (INRB) | Pathogen Sequencing Lab, National Institute for Biomedical Research (INRB) |
| EPI_ISL_417949 | LaboratÃ³rio Hermes Pardini | Bioinformatics Laboratory - LNCC |

| EPI_Accession_ID | Originating Laboratory | Submitting Laboratory |
|---|---|---|
| EPI_ISL_417950 | Viral Respiratory Lab, National Institute for Biomedical Research (INRB) | Pathogen Sequencing Lab, National Institute for Biomedical Research (INRB) |
| EPI_ISL_417952 | Hospital Universitario 12 de Octubre | Hospital Universitario La Paz |
| EPI_ISL_417954 | Hospital Universitario 12 de Octubre | Hospital Universitario La Paz |
| EPI_ISL_417955 | Viral Respiratory Lab, National Institute for Biomedical Research (INRB) | Pathogen Sequencing Lab, National Institute for Biomedical Research (INRB) |
| EPI_ISL_417956 | Hospital Universitario 12 de Octubre | Hospital Universitario La Paz |
| EPI_ISL_417957 | Hospital Universitario 12 de Octubre | Hospital Universitario La Paz |
| EPI_ISL_417958 | Utah Public Health Laboratory | Utah Public Health Laboratory |
| EPI_ISL_417959 | Utah Public Health Laboratory | Utah Public Health Laboratory |
| EPI_ISL_417960 | Utah Public Health Laboratory | Utah Public Health Laboratory |
| EPI_ISL_417961 | Hospital Universitario 12 de Octubre | Hospital Universitario La Paz |
| EPI_ISL_417963 | Hospital Universitario 12 de Octubre | Hospital Universitario La Paz |
| EPI_ISL_417964 | Utah Public Health Laboratory | Utah Public Health Laboratory |
| EPI_ISL_417966 | Utah Public Health Laboratory | Utah Public Health Laboratory |
| EPI_ISL_417967 | Hospital Universitario 12 de Octubre | Hospital Universitario La Paz |
| EPI_ISL_417969 | Hospital Universitario La Paz | Hospital Universitario La Paz |
| EPI_ISL_417970 | Utah Public Health Laboratory | Utah Public Health Laboratory |
| EPI_ISL_417971 | Utah Public Health Laboratory | Utah Public Health Laboratory |
| EPI_ISL_417972 | Hospital Universitario La Paz | Hospital Universitario La Paz |
| EPI_ISL_417973 | Utah Public Health Laboratory | Utah Public Health Laboratory |
| EPI_ISL_417974 | Utah Public Health Laboratory | Utah Public Health Laboratory |
| EPI_ISL_417975 | Hospital Universitario La Paz | Hospital Universitario La Paz |
| EPI_ISL_417976 | Utah Public Health Laboratory | Utah Public Health Laboratory |
| EPI_ISL_417977 | Utah Public Health Laboratory | Utah Public Health Laboratory |
| EPI_ISL_417978 | Hospital Universitario La Paz | Hospital Universitario La Paz |
| EPI_ISL_417979 | Hospital Universitario RamÃ³n y Cajal | Hospital Universitario La Paz |
| EPI_ISL_417980 | Hospital Universitario RamÃ³n y Cajal | Hospital Universitario La Paz |
| EPI_ISL_417981 | Hospital Universitario RamÃ³n y Cajal | Hospital Universitario La Paz |
| EPI_ISL_417982 | Universidade Federal do Rio de Janeiro | Bioinformatics Laboratory - LNCC |
| EPI_ISL_417984 | Universidade Federal do Rio de Janeiro | Bioinformatics Laboratory - LNCC |
| EPI_ISL_417986 | Centro Hospitalar e Universitario de Sao Joao, Porto | Instituto Nacional de Saude (INSA) |
| EPI_ISL_417987 | Centro Hospitalar e Universitario de Sao Joao, Porto | Instituto Nacional de Saude (INSA) |

| EPI_Accession_ID | Originating Laboratory | Submitting Laboratory |
|---|---|---|
| EPI_ISL_417988 | CHULC - H Curry Cabral | Instituto Nacional de Saude (INSA) |
| EPI_ISL_417989 | Centro Hospitalar e Universitario de Sao Joao, Porto | Instituto Nacional de Saude (INSA) |
| EPI_ISL_417990 | CHULC - H Curry Cabral | Instituto Nacional de Saude (INSA) |
| EPI_ISL_417992 | CHULC - H D Estefania | Instituto Nacional de Saude (INSA) |
| EPI_ISL_417993 | CHULC - H D Estefania | Instituto Nacional de Saude (INSA) |
| EPI_ISL_417994 | CHULC - H Curry Cabral | Instituto Nacional de Saude (INSA) |
| EPI_ISL_417995 | CHULC - H Curry Cabral | Instituto Nacional de Saude (INSA) |
| EPI_ISL_417996 | CHULC - H Curry Cabral | Instituto Nacional de Saude (INSA) |
| EPI_ISL_417997 | Centro Hospital do Porto, E.P.E. - H. Geral de Santo Antonio | Instituto Nacional de Saude (INSA) |
| EPI_ISL_417998 | Centro Hospital do Porto, E.P.E. - H. Geral de Santo Antonio | Instituto Nacional de Saude (INSA) |
| EPI_ISL_417999 | Centro Hospital do Porto, E.P.E. - H. Geral de Santo Antonio | Instituto Nacional de Saude (INSA) |
| EPI_ISL_418000 | ARS Algarve - LaboratÃ³rio Laura Ayres | Instituto Nacional de Saude (INSA) |
| EPI_ISL_418001 | ARS Algarve - LaboratÃ³rio Laura Ayres | Instituto Nacional de Saude (INSA) |
| EPI_ISL_418002 | CHU Coimbra | Instituto Nacional de Saude (INSA) |
| EPI_ISL_418003 | H Braga | Instituto Nacional de Saude (INSA) |
| EPI_ISL_418004 | ARS Algarve - LaboratÃ³rio Laura Ayres | Instituto Nacional de Saude (INSA) |
| EPI_ISL_418005 | CHU Coimbra - PediÃ¡trico | Instituto Nacional de Saude (INSA) |
| EPI_ISL_418006 | CHBarreiro Montijo | Instituto Nacional de Saude (INSA) |
| EPI_ISL_418007 | H Braga | Instituto Nacional de Saude (INSA) |
| EPI_ISL_418008 | H Braga | Instituto Nacional de Saude (INSA) |
| EPI_ISL_418009 | HSE Ilha Terceira - Angra do Heroismo | Instituto Nacional de Saude (INSA) |
| EPI_ISL_418010 | CHULC - H Curry Cabral | Instituto Nacional de Saude (INSA) |
| EPI_ISL_418011 | CHULC - H Curry Cabral | Instituto Nacional de Saude (INSA) |
| EPI_ISL_418012 | CHULC - H Curry Cabral | Instituto Nacional de Saude (INSA) |
| EPI_ISL_418013 | CHULC - H Curry Cabral | Instituto Nacional de Saude (INSA) |
| EPI_ISL_418014 | CHULC - H Curry Cabral | Instituto Nacional de Saude (INSA) |
| EPI_ISL_418015 | CHULC - H Curry Cabral | Instituto Nacional de Saude (INSA) |
| EPI_ISL_418016 | CHULC - H Curry Cabral | Instituto Nacional de Saude (INSA) |
| EPI_ISL_418017 | CHMT | Instituto Nacional de Saude (INSA) |
| EPI_ISL_418018 | H Garcia de Orta | Instituto Nacional de Saude (INSA) |

| EPI_Accession_ID | Originating Laboratory | Submitting Laboratory |
|---|---|---|
| EPI_ISL_418019 | H Braga | Instituto Nacional de Saude (INSA) |
| EPI_ISL_418020 | H Braga | Instituto Nacional de Saude (INSA) |
| EPI_ISL_418021 | H Braga | Instituto Nacional de Saude (INSA) |
| EPI_ISL_418022 | H Braga | Instituto Nacional de Saude (INSA) |
| EPI_ISL_418023 | H Evora | Instituto Nacional de Saude (INSA) |
| EPI_ISL_418024 | CHUA - Faro | Instituto Nacional de Saude (INSA) |
| EPI_ISL_418025 | H Santarem | Instituto Nacional de Saude (INSA) |
| EPI_ISL_418026 | H Dr. Nelio Mendonca - Funchal | Instituto Nacional de Saude (INSA) |
| EPI_ISL_418027 | CHTMAD | Instituto Nacional de Saude (INSA) |
| EPI_ISL_418028 | UW Virology Lab | UW Virology Lab |
| EPI_ISL_418029 | UW Virology Lab | UW Virology Lab |
| EPI_ISL_418030 | UW Virology Lab | UW Virology Lab |
| EPI_ISL_418031 | UW Virology Lab | UW Virology Lab |
| EPI_ISL_418032 | UW Virology Lab | UW Virology Lab |
| EPI_ISL_418033 | UW Virology Lab | UW Virology Lab |
| EPI_ISL_418034 | UW Virology Lab | UW Virology Lab |
| EPI_ISL_418035 | UW Virology Lab | UW Virology Lab |
| EPI_ISL_418036 | UW Virology Lab | UW Virology Lab |
| EPI_ISL_418037 | UW Virology Lab | UW Virology Lab |
| EPI_ISL_418038 | UW Virology Lab | UW Virology Lab |
| EPI_ISL_418039 | UW Virology Lab | UW Virology Lab |
| EPI_ISL_418040 | UW Virology Lab | UW Virology Lab |
| EPI_ISL_418041 | UW Virology Lab | UW Virology Lab |
| EPI_ISL_418042 | UW Virology Lab | UW Virology Lab |
| EPI_ISL_418043 | UW Virology Lab | UW Virology Lab |
| EPI_ISL_418044 | UW Virology Lab | UW Virology Lab |
| EPI_ISL_418045 | UW Virology Lab | UW Virology Lab |
| EPI_ISL_418046 | UW Virology Lab | UW Virology Lab |
| EPI_ISL_418047 | UW Virology Lab | UW Virology Lab |
| EPI_ISL_418048 | UW Virology Lab | UW Virology Lab |
| EPI_ISL_418049 | UW Virology Lab | UW Virology Lab |
| EPI_ISL_418050 | UW Virology Lab | UW Virology Lab |
| EPI_ISL_418051 | UW Virology Lab | UW Virology Lab |

| EPI_Accession_ID | Originating Laboratory | Submitting Laboratory |
|---|---|---|
| EPI_ISL_418052 | UW Virology Lab | UW Virology Lab |
| EPI_ISL_418053 | UW Virology Lab | UW Virology Lab |
| EPI_ISL_418054 | UW Virology Lab | UW Virology Lab |
| EPI_ISL_418055 | UW Virology Lab | UW Virology Lab |
| EPI_ISL_418056 | UW Virology Lab | UW Virology Lab |
| EPI_ISL_418057 | UW Virology Lab | UW Virology Lab |
| EPI_ISL_418058 | UW Virology Lab | UW Virology Lab |
| EPI_ISL_418059 | UW Virology Lab | UW Virology Lab |
| EPI_ISL_418060 | UW Virology Lab | UW Virology Lab |
| EPI_ISL_418061 | UW Virology Lab | UW Virology Lab |
| EPI_ISL_418062 | UW Virology Lab | UW Virology Lab |
| EPI_ISL_418063 | UW Virology Lab | UW Virology Lab |
| EPI_ISL_418064 | UW Virology Lab | UW Virology Lab |
| EPI_ISL_418065 | UW Virology Lab | UW Virology Lab |
| EPI_ISL_418066 | UW Virology Lab | UW Virology Lab |
| EPI_ISL_418067 | UW Virology Lab | UW Virology Lab |
| EPI_ISL_418068 | UW Virology Lab | UW Virology Lab |
| EPI_ISL_418069 | UW Virology Lab | UW Virology Lab |
| EPI_ISL_418070 | UW Virology Lab | UW Virology Lab |
| EPI_ISL_418071 | UW Virology Lab | UW Virology Lab |
| EPI_ISL_418072 | UW Virology Lab | UW Virology Lab |
| EPI_ISL_418073 | UW Virology Lab | UW Virology Lab |
| EPI_ISL_418074 | UW Virology Lab | UW Virology Lab |
| EPI_ISL_418075 | UW Virology Lab | UW Virology Lab |
| EPI_ISL_418076 | UW Virology Lab | UW Virology Lab |
| EPI_ISL_418077 | UW Virology Lab | UW Virology Lab |
| EPI_ISL_418078 | UW Virology Lab | UW Virology Lab |
| EPI_ISL_418079 | UW Virology Lab | UW Virology Lab |
| EPI_ISL_418080 | UW Virology Lab | UW Virology Lab |
| EPI_ISL_418081 | UW Virology Lab | UW Virology Lab |
| EPI_ISL_418082 | UW Virology Lab | UW Virology Lab |
| EPI_ISL_418083 | Wales Specialist Virology Centre | Public Health Wales Microbiology Cardiff |
| EPI_ISL_418084 | Wales Specialist Virology Centre | Public Health Wales Microbiology Cardiff |

| EPI_Accession_ID | Originating Laboratory | Submitting Laboratory |
|---|---|---|
| EPI_ISL_418085 | Wales Specialist Virology Centre | Public Health Wales Microbiology Cardiff |
| EPI_ISL_418086 | Wales Specialist Virology Centre | Public Health Wales Microbiology Cardiff |
| EPI_ISL_418087 | Wales Specialist Virology Centre | Public Health Wales Microbiology Cardiff |
| EPI_ISL_418088 | Wales Specialist Virology Centre | Public Health Wales Microbiology Cardiff |
| EPI_ISL_418089 | Wales Specialist Virology Centre | Public Health Wales Microbiology Cardiff |
| EPI_ISL_418090 | Wales Specialist Virology Centre | Public Health Wales Microbiology Cardiff |
| EPI_ISL_418091 | Wales Specialist Virology Centre | Public Health Wales Microbiology Cardiff |
| EPI_ISL_418092 | Wales Specialist Virology Centre | Public Health Wales Microbiology Cardiff |
| EPI_ISL_418093 | Wales Specialist Virology Centre | Public Health Wales Microbiology Cardiff |
| EPI_ISL_418094 | Wales Specialist Virology Centre | Public Health Wales Microbiology Cardiff |
| EPI_ISL_418095 | Wales Specialist Virology Centre | Public Health Wales Microbiology Cardiff |
| EPI_ISL_418096 | Wales Specialist Virology Centre | Public Health Wales Microbiology Cardiff |
| EPI_ISL_418097 | Wales Specialist Virology Centre | Public Health Wales Microbiology Cardiff |
| EPI_ISL_418098 | Wales Specialist Virology Centre | Public Health Wales Microbiology Cardiff |
| EPI_ISL_418099 | Wales Specialist Virology Centre | Public Health Wales Microbiology Cardiff |
| EPI_ISL_418100 | Wales Specialist Virology Centre | Public Health Wales Microbiology Cardiff |
| EPI_ISL_418101 | Wales Specialist Virology Centre | Public Health Wales Microbiology Cardiff |
| EPI_ISL_418102 | Wales Specialist Virology Centre | Public Health Wales Microbiology Cardiff |
| EPI_ISL_418103 | Wales Specialist Virology Centre | Public Health Wales Microbiology Cardiff |
| EPI_ISL_418104 | Wales Specialist Virology Centre | Public Health Wales Microbiology Cardiff |
| EPI_ISL_418105 | Wales Specialist Virology Centre | Public Health Wales Microbiology Cardiff |
| EPI_ISL_418106 | Wales Specialist Virology Centre | Public Health Wales Microbiology Cardiff |
| EPI_ISL_418107 | Wales Specialist Virology Centre | Public Health Wales Microbiology Cardiff |
| EPI_ISL_418108 | Wales Specialist Virology Centre | Public Health Wales Microbiology Cardiff |
| EPI_ISL_418109 | Wales Specialist Virology Centre | Public Health Wales Microbiology Cardiff |
| EPI_ISL_418110 | Wales Specialist Virology Centre | Public Health Wales Microbiology Cardiff |
| EPI_ISL_418111 | Wales Specialist Virology Centre | Public Health Wales Microbiology Cardiff |
| EPI_ISL_418112 | Wales Specialist Virology Centre | Public Health Wales Microbiology Cardiff |
| EPI_ISL_418113 | Wales Specialist Virology Centre | Public Health Wales Microbiology Cardiff |
| EPI_ISL_418114 | Wales Specialist Virology Centre | Public Health Wales Microbiology Cardiff |
| EPI_ISL_418115 | Wales Specialist Virology Centre | Public Health Wales Microbiology Cardiff |
| EPI_ISL_418116 | Wales Specialist Virology Centre | Public Health Wales Microbiology Cardiff |
| EPI_ISL_418117 | Wales Specialist Virology Centre | Public Health Wales Microbiology Cardiff |

| EPI_Accession_ID | Originating Laboratory | Submitting Laboratory |
|---|---|---|
| EPI_ISL_418118 | Wales Specialist Virology Centre | Public Health Wales Microbiology Cardiff |
| EPI_ISL_418119 | Wales Specialist Virology Centre | Public Health Wales Microbiology Cardiff |
| EPI_ISL_418120 | Wales Specialist Virology Centre | Public Health Wales Microbiology Cardiff |
| EPI_ISL_418121 | Wales Specialist Virology Centre | Public Health Wales Microbiology Cardiff |
| EPI_ISL_418122 | Wales Specialist Virology Centre | Public Health Wales Microbiology Cardiff |
| EPI_ISL_418123 | Wales Specialist Virology Centre | Public Health Wales Microbiology Cardiff |
| EPI_ISL_418124 | Wales Specialist Virology Centre | Public Health Wales Microbiology Cardiff |
| EPI_ISL_418125 | Wales Specialist Virology Centre | Public Health Wales Microbiology Cardiff |
| EPI_ISL_418126 | Wales Specialist Virology Centre | Public Health Wales Microbiology Cardiff |
| EPI_ISL_418127 | Wales Specialist Virology Centre | Public Health Wales Microbiology Cardiff |
| EPI_ISL_418128 | Wales Specialist Virology Centre | Public Health Wales Microbiology Cardiff |
| EPI_ISL_418129 | Wales Specialist Virology Centre | Public Health Wales Microbiology Cardiff |
| EPI_ISL_418130 | Wales Specialist Virology Centre | Public Health Wales Microbiology Cardiff |
| EPI_ISL_418131 | Wales Specialist Virology Centre | Public Health Wales Microbiology Cardiff |
| EPI_ISL_418132 | Wales Specialist Virology Centre | Public Health Wales Microbiology Cardiff |
| EPI_ISL_418133 | Wales Specialist Virology Centre | Public Health Wales Microbiology Cardiff |
| EPI_ISL_418134 | Wales Specialist Virology Centre | Public Health Wales Microbiology Cardiff |
| EPI_ISL_418135 | Wales Specialist Virology Centre | Public Health Wales Microbiology Cardiff |
| EPI_ISL_418136 | Wales Specialist Virology Centre | Public Health Wales Microbiology Cardiff |
| EPI_ISL_418137 | Wales Specialist Virology Centre | Public Health Wales Microbiology Cardiff |
| EPI_ISL_418138 | Wales Specialist Virology Centre | Public Health Wales Microbiology Cardiff |
| EPI_ISL_418139 | Wales Specialist Virology Centre | Public Health Wales Microbiology Cardiff |
| EPI_ISL_418140 | Wales Specialist Virology Centre | Public Health Wales Microbiology Cardiff |
| EPI_ISL_418141 | Wales Specialist Virology Centre | Public Health Wales Microbiology Cardiff |
| EPI_ISL_418142 | Wales Specialist Virology Centre | Public Health Wales Microbiology Cardiff |
| EPI_ISL_418143 | Wales Specialist Virology Centre | Public Health Wales Microbiology Cardiff |
| EPI_ISL_418144 | Wales Specialist Virology Centre | Public Health Wales Microbiology Cardiff |
| EPI_ISL_418145 | Wales Specialist Virology Centre | Public Health Wales Microbiology Cardiff |
| EPI_ISL_418146 | Wales Specialist Virology Centre | Public Health Wales Microbiology Cardiff |
| EPI_ISL_418147 | Wales Specialist Virology Centre | Public Health Wales Microbiology Cardiff |
| EPI_ISL_418148 | Wales Specialist Virology Centre | Public Health Wales Microbiology Cardiff |
| EPI_ISL_418149 | Wales Specialist Virology Centre | Public Health Wales Microbiology Cardiff |
| EPI_ISL_418150 | Wales Specialist Virology Centre | Public Health Wales Microbiology Cardiff |

| EPI_Accession_ID | Originating Laboratory | Submitting Laboratory |
|---|---|---|
| EPI_ISL_418151 | Wales Specialist Virology Centre | Public Health Wales Microbiology Cardiff |
| EPI_ISL_418152 | Wales Specialist Virology Centre | Public Health Wales Microbiology Cardiff |
| EPI_ISL_418153 | Wales Specialist Virology Centre | Public Health Wales Microbiology Cardiff |
| EPI_ISL_418154 | Wales Specialist Virology Centre | Public Health Wales Microbiology Cardiff |
| EPI_ISL_418155 | Wales Specialist Virology Centre | Public Health Wales Microbiology Cardiff |
| EPI_ISL_418156 | Wales Specialist Virology Centre | Public Health Wales Microbiology Cardiff |
| EPI_ISL_418157 | Wales Specialist Virology Centre | Public Health Wales Microbiology Cardiff |
| EPI_ISL_418158 | Wales Specialist Virology Centre | Public Health Wales Microbiology Cardiff |
| EPI_ISL_418159 | Wales Specialist Virology Centre | Public Health Wales Microbiology Cardiff |
| EPI_ISL_418160 | Wales Specialist Virology Centre | Public Health Wales Microbiology Cardiff |
| EPI_ISL_418161 | Wales Specialist Virology Centre | Public Health Wales Microbiology Cardiff |
| EPI_ISL_418162 | Wales Specialist Virology Centre | Public Health Wales Microbiology Cardiff |
| EPI_ISL_418163 | Wales Specialist Virology Centre | Public Health Wales Microbiology Cardiff |
| EPI_ISL_418164 | Wales Specialist Virology Centre | Public Health Wales Microbiology Cardiff |
| EPI_ISL_418165 | Wales Specialist Virology Centre | Public Health Wales Microbiology Cardiff |
| EPI_ISL_418182 | Hospital Universitario 12 de Octubre | Hospital Universitario La Paz |
| EPI_ISL_418183 | Virological Research Group, SzentÃ¡gothai Research Centre | Bioinformatics Research Group, SzentÃ¡gothai Research Centre |
| EPI_ISL_418184 | Gundersen Molecular Diagnostics Laboratory | Kabara Cancer Research Institute |
| EPI_ISL_418185 | Gundersen Molecular Diagnostic Laboratory | Kabara Cancer Research Institute |
| EPI_ISL_418186 | Gundersen Molecular Diagnostic Laboratory | Kabara Cancer Research Institute |
| EPI_ISL_418187 | Gundersen Molecular Diagnostics Laboratory | Kabara Cancer Research Institute |
| EPI_ISL_418188 | Gundersen Molecular Diagnostics Laboratory | Kabara Cancer Research Institute |
| EPI_ISL_418190 | NYU Langone Health | Department of Pathology and Medicine, New York University School of Medicine |
| EPI_ISL_418192 | NYU Langone Health | Department of Pathology and Medicine, New York University School of Medicine |
| EPI_ISL_418193 | NYU Langone Health | Department of Pathology and Medicine, New York University School of Medicine |
| EPI_ISL_418194 | NYU Langone Health | Department of Pathology and Medicine, New York University School of Medicine |
| EPI_ISL_418196 | NYU Langone Health | Department of Pathology and Medicine, New York University School of Medicine |

| EPI_Accession_ID | Originating Laboratory | Submitting Laboratory |
|---|---|---|
| EPI_ISL_418197 | NYU Langone Health | Department of Pathology and Medicine, New York University School of Medicine |
| EPI_ISL_418198 | NYU Langone Health | Department of Pathology and Medicine, New York University School of Medicine |
| EPI_ISL_418199 | NYU Langone Health | Department of Pathology and Medicine, New York University School of Medicine |
| EPI_ISL_418200 | NYU Langone Health | Department of Pathology and Medicine, New York University School of Medicine |
| EPI_ISL_418202 | NYU Langone Health | Department of Pathology and Medicine, New York University School of Medicine |
| EPI_ISL_418203 | NYU Langone Health | Department of Pathology and Medicine, New York University School of Medicine |
| EPI_ISL_418204 | NYU Langone Health | Department of Pathology and Medicine, New York University School of Medicine |
| EPI_ISL_418205 | NYU Langone Health | Department of Pathology and Medicine, New York University School of Medicine |
| EPI_ISL_418206 | Institut Pasteur Dakar | Institut Pasteur de Dakar |
| EPI_ISL_418207 | Institut Pasteur Dakar | Institut Pasteur de Dakar |
| EPI_ISL_418208 | Institut Pasteur Dakar | Institut Pasteur de Dakar |
| EPI_ISL_418209 | Institut Pasteur Dakar | Institut Pasteur de Dakar |
| EPI_ISL_418210 | Institut Pasteur Dakar | Institut Pasteur de Dakar |
| EPI_ISL_418211 | Institut Pasteur Dakar | Institut Pasteur de Dakar |
| EPI_ISL_418212 | Institut Pasteur Dakar | Institut Pasteur de Dakar |
| EPI_ISL_418213 | Institut Pasteur Dakar | Institut Pasteur de Dakar |
| EPI_ISL_418214 | Institut Pasteur Dakar | Institut Pasteur de Dakar |
| EPI_ISL_418215 | Instirut Pasteur Dakar | Institut Pasteur de Dakar |
| EPI_ISL_418216 | Institut Pasteur Dakar | Institut Pasteur de Dakar |
| EPI_ISL_418217 | Institut Pasteur Dakar | Institut Pasteur de Dakar |
| EPI_ISL_418218 | Centre Hospitalier CompiÃ¨gne Laboratoire de Biologie | National Reference Center for Viruses of Respiratory Infections, Institut Pasteur, Paris |
| EPI_ISL_418219 | CHU - HÃ´pital Cavale Blanche - Labo. de Virologie | National Reference Center for Viruses of Respiratory Infections, Institut Pasteur, Paris |

| EPI_Accession_ID | Originating Laboratory | Submitting Laboratory |
|---|---|---|
| EPI_ISL_418220 | Centre Hospitalier CompiÃ¨gne Laboratoire de Biologie | National Reference Center for Viruses of Respiratory Infections, Institut Pasteur, Paris |
| EPI_ISL_418221 | Centre Hospitalier CompiÃ¨gne Laboratoire de Biologie | National Reference Center for Viruses of Respiratory Infections, Institut Pasteur, Paris |
| EPI_ISL_418222 | CHRU Bretonneau - Serv. Bacterio-Virol. | National Reference Center for Viruses of Respiratory Infections, Institut Pasteur, Paris |
| EPI_ISL_418223 | Centre Hospitalier CompiÃ¨gne Laboratoire de Biologie | National Reference Center for Viruses of Respiratory Infections, Institut Pasteur, Paris |
| EPI_ISL_418224 | Centre Hospitalier CompiÃ¨gne Laboratoire de Biologie | National Reference Center for Viruses of Respiratory Infections, Institut Pasteur, Paris |
| EPI_ISL_418225 | Centre Hospitalier CompiÃ¨gne Laboratoire de Biologie | National Reference Center for Viruses of Respiratory Infections, Institut Pasteur, Paris |
| EPI_ISL_418226 | EHPAD - RÃ©sidences les CÃ¨dres | National Reference Center for Viruses of Respiratory Infections, Institut Pasteur, Paris |
| EPI_ISL_418227 | Centre Hospitalier CompiÃ¨gne Laboratoire de Biologie | National Reference Center for Viruses of Respiratory Infections, Institut Pasteur, Paris |
| EPI_ISL_418228 | Centre Hospitalier CompiÃ¨gne Laboratoire de Biologie | National Reference Center for Viruses of Respiratory Infections, Institut Pasteur, Paris |
| EPI_ISL_418229 | Hopital franco britannique - Laboratoire | National Reference Center for Viruses of Respiratory Infections, Institut Pasteur, Paris |
| EPI_ISL_418230 | Clinique AVERAY LA BROUSTE, Med. Polyvalente | National Reference Center for Viruses of Respiratory Infections, Institut Pasteur, Paris |
| EPI_ISL_418231 | Centre Hospitalier CompiÃ¨gne Laboratoire de Biologie | National Reference Center for Viruses of Respiratory Infections, Institut Pasteur, Paris |
| EPI_ISL_418232 | Service des Urgences | National Reference Center for Viruses of Respiratory Infections, Institut Pasteur, Paris |
| EPI_ISL_418233 | Service des Urgences | National Reference Center for Viruses of Respiratory Infections, Institut Pasteur, Paris |
| EPI_ISL_418234 | LABM GH nord Essonne | National Reference Center for Viruses of Respiratory Infections, Institut Pasteur, Paris |
| EPI_ISL_418235 | Cabinet mÃ©dical | National Reference Center for Viruses of Respiratory Infections, Institut Pasteur, Paris |

| EPI_Accession_ID | Originating Laboratory | Submitting Laboratory |
| --- | --- | --- |
| EPI_ISL_418236 | Centre Hospitalier CompiÃ¨gne Laboratoire de Biologie | National Reference Center for Viruses of Respiratory Infections, Institut Pasteur, Paris |
| EPI_ISL_418237 | Centre Hospitalier CompiÃ¨gne Laboratoire de Biologie | National Reference Center for Viruses of Respiratory Infections, Institut Pasteur, Paris |
| EPI_ISL_418238 | Centre Hospitalier CompiÃ¨gne Laboratoire de Biologie | National Reference Center for Viruses of Respiratory Infections, Institut Pasteur, Paris |
| EPI_ISL_418239 | Centre Hospitalier CompiÃ¨gne Laboratoire de Biologie | National Reference Center for Viruses of Respiratory Infections, Institut Pasteur, Paris |
| EPI_ISL_418240 | LABM GH nord Essonne | National Reference Center for Viruses of Respiratory Infections, Institut Pasteur, Paris |
| EPI_ISL_418241 | NIC Viral Respiratory Unit - Institut Pasteur of Algeria | National Reference Center for Viruses of Respiratory Infections, Institut Pasteur, Paris |
| EPI_ISL_418242 | NIC Viral Respiratory Unit - Institut Pasteur of Algeria | National Reference Center for Viruses of Respiratory Infections, Institut Pasteur, Paris |
| EPI_ISL_418243 | HOSPITAL UNIVERSITARIO VIRGEN DE LAS NIEVES | Instituto de Salud Carlos III |
| EPI_ISL_418244 | HOSPITAL UNIVERSITARIO VIRGEN DE LAS NIEVES | Instituto de Salud Carlos III |
| EPI_ISL_418245 | Hospital General y Universitario de Guadalajara | Instituto de Salud Carlos III |
| EPI_ISL_418246 | Hospital General y Universitario de Guadalajara | Instituto de Salud Carlos III |
| EPI_ISL_418247 | HOSPITAL GENERAL DE SEGOVIA | Instituto de Salud Carlos III |
| EPI_ISL_418248 | COMPLEJO ASISTENCIAL UNIVERSITARIO DE BURGOS | Instituto de Salud Carlos III |
| EPI_ISL_418249 | COMPLEJO ASISTENCIAL UNIVERSITARIO DE BURGOS | Instituto de Salud Carlos III |
| EPI_ISL_418251 | HOSPITAL UNIVERSITARIO LA PAZ | Instituto de Salud Carlos III |
| EPI_ISL_418252 | FUNDACION JIMENEZ DIAZ | Instituto de Salud Carlos III |
| EPI_ISL_418253 | HOSPITAL TXAGORRITXU | Instituto de Salud Carlos III |
| EPI_ISL_418254 | NYU Langone Health | Department of Pathology and Medicine, New York University School of Medicine |
| EPI_ISL_418255 | Presidio Ospedaliero "S. Spirito" - PESCARA | Istituto Zooprofilattico Sperimentale dell'Abruzzo e Molise "G. Caporale" |
| EPI_ISL_418256 | Ospedale â€œSan Liberatoreâ€�di Atri | Istituto Zooprofilattico Sperimentale dell'Abruzzo e Molise "G. Caporale" |
| EPI_ISL_418257 | Ospedale Civile Giuseppe Mazzini, Teramo | Istituto Zooprofilattico Sperimentale dell'Abruzzo e Molise "G.Caporale" |

| EPI_Accession_ID | Originating Laboratory | Submitting Laboratory |
|---|---|---|
| EPI_ISL_418258 | Presidio ospedaliero "Santo Spirito" | Istituto Zooprofilattico Sperimentale dell'Abruzzo e Molise "G. Caporale" |
| EPI_ISL_418259 | Presidio ospedaliero "Santo Spirito" | Istituto Zooprofilattico Sperimentale dell'Abruzzo e Molise "G. Caporale" |
| EPI_ISL_418260 | Ospedale Civile Giuseppe Mazzini | Istituto Zooprofilattico Sperimentale dell'Abruzzo e Molise "G. Caporale" |
| EPI_ISL_418261 | Ospedale Civile Giuseppe Mazzini | Istituto Zooprofilattico Sperimentale dell'Abruzzo e Molise "G. Caporale" |
| EPI_ISL_418262 | Instituto Nacional de Salud | Instituto Nacional de Salud Universidad Cooperativa de Colombia Instituto Alexander von Humboldt Imperial College-London London School of Hygiene & Tropical Medicine |
| EPI_ISL_418263 | Laboratory of Microbiology, Department of Medicine, National and Kapodistrian University of Athens, Greece | Laboratory of Biology, Department of Medicine, Democritus University of Thrace, Greece |
| EPI_ISL_418264 | Laboratory of Microbiology, Department of Medicine, National and Kapodistrian University of Athens, Greece | Laboratory of Biology, Department of Medicine, Democritus University of Thrace, Greece |
| EPI_ISL_418265 | Laboratory of Microbiology, Department of Medicine, National and Kapodistrian University of Athens, Greece | Laboratory of Biology, Department of Medicine, Democritus University of Thrace, Greece |
| EPI_ISL_418267 | unknown | Microbiology and Immunology department |
| EPI_ISL_418268 | Hospital Universitari Germans Trias i Pujol(HUGTiP)/FundaciÃ³ Lluita contra la SIDA (FLSida)/IRTA-CReSA | IrsiCaixa AIDS Research Lab |
| EPI_ISL_418269 | unknown | Microbiology and Immunology department |
| EPI_ISL_418270 | KU Leuven, Clinical and Epidemiological Virology | KU Leuven, Clinical and Epidemiological Virology |
| EPI_ISL_418271 | University Hospital Basel, Clinical Virology | University Hospital Basel, Labormedizin |
| EPI_ISL_418273 | University Hospital Basel, Clinical Virology | University Hospital Basel, Labormedizin |
| EPI_ISL_418274 | University Hospital Basel, Clinical Virology | University Hospital Basel, Clinical Bacteriology |
| EPI_ISL_418275 | University Hospital Basel, Clinical Virology | University Hospital Basel, Clinical Bacteriology |
| EPI_ISL_418277 | University Hospital Basel, Clinical Virology | University Hospital Basel, Clinical Bacteriology |
| EPI_ISL_418278 | University Hospital Basel, Clinical Virology | University Hospital Basel, Clinical Bacteriology |

| EPI_Accession_ID | Originating Laboratory | Submitting Laboratory |
|---|---|---|
| EPI_ISL_418279 | University Hospital Basel, Clinical Virology | University Hospital Basel, Clinical Bacteriology |
| EPI_ISL_418280 | University Hospital Basel, Clinical Virology | University Hospital Basel, Clinical Bacteriology |
| EPI_ISL_418281 | University Hospital Basel, Clinical Virology | University Hospital Basel, Clinical Bacteriology |
| EPI_ISL_418282 | University Hospital Basel, Clinical Virology | University Hospital Basel, Clinical Bacteriology |
| EPI_ISL_418283 | University Hospital Basel, Clinical Virology | University Hospital Basel, Clinical Bacteriology |
| EPI_ISL_418284 | University Hospital Basel, Clinical Virology | University Hospital Basel, Clinical Bacteriology |
| EPI_ISL_418285 | University Hospital Basel, Clinical Virology | University Hospital Basel, Clinical Bacteriology |
| EPI_ISL_418286 | Virology Department, Sheffield Teaching Hospitals NHS Foundation Trust | Department of Infection, Immunity and Cardiovascular Disease, The Florey Institute, The Medical School, University of Sheffield |
| EPI_ISL_418287 | Virology Department, Sheffield Teaching Hospitals NHS Foundation Trust | Department of Infection, Immunity and Cardiovascular Disease, The Florey Institute, The Medical School, University of Sheffield |
| EPI_ISL_418288 | Virology Department, Sheffield Teaching Hospitals NHS Foundation Trust | Department of Infection, Immunity and Cardiovascular Disease, The Florey Institute, The Medical School, University of Sheffield |
| EPI_ISL_418289 | Virology Department, Sheffield Teaching Hospitals NHS Foundation Trust | Department of Infection, Immunity and Cardiovascular Disease, The Florey Institute, The Medical School, University of Sheffield |
| EPI_ISL_418290 | Virology Department, Sheffield Teaching Hospitals NHS Foundation Trust | Department of Infection, Immunity and Cardiovascular Disease, The Florey Institute, The Medical School, University of Sheffield |
| EPI_ISL_418291 | Virology Department, Sheffield Teaching Hospitals NHS Foundation Trust | Department of Infection, Immunity and Cardiovascular Disease, The Florey Institute, The Medical School, University of Sheffield |
| EPI_ISL_418292 | Virology Department, Sheffield Teaching Hospitals NHS Foundation Trust | Department of Infection, Immunity and Cardiovascular Disease, The Florey Institute, The Medical School, University of Sheffield |
| EPI_ISL_418293 | Virology Department, Sheffield Teaching Hospitals NHS Foundation Trust | Department of Infection, Immunity and Cardiovascular Disease, The Florey Institute, The Medical School, University of Sheffield |

| EPI_Accession_ID | Originating Laboratory | Submitting Laboratory |
|---|---|---|
| EPI_ISL_418294 | Virology Department, Sheffield Teaching Hospitals NHS Foundation Trust | Department of Infection, Immunity and Cardiovascular Disease, The Florey Institute, The Medical School, University of Sheffield |
| EPI_ISL_418295 | Virology Department, Sheffield Teaching Hospitals NHS Foundation Trust | Department of Infection, Immunity and Cardiovascular Disease, The Florey Institute, The Medical School, University of Sheffield |
| EPI_ISL_418296 | Virology Department, Sheffield Teaching Hospitals NHS Foundation Trust | Department of Infection, Immunity and Cardiovascular Disease, The Florey Institute, The Medical School, University of Sheffield |
| EPI_ISL_418297 | Virology Department, Sheffield Teaching Hospitals NHS Foundation Trust | Department of Infection, Immunity and Cardiovascular Disease, The Florey Institute, The Medical School, University of Sheffield |
| EPI_ISL_418298 | Virology Department, Sheffield Teaching Hospitals NHS Foundation Trust | Department of Infection, Immunity and Cardiovascular Disease, The Florey Institute, The Medical School, University of Sheffield |
| EPI_ISL_418299 | Virology Department, Sheffield Teaching Hospitals NHS Foundation Trust | Department of Infection, Immunity and Cardiovascular Disease, The Florey Institute, The Medical School, University of Sheffield |
| EPI_ISL_418300 | Virology Department, Sheffield Teaching Hospitals NHS Foundation Trust | Department of Infection, Immunity and Cardiovascular Disease, The Florey Institute, The Medical School, University of Sheffield |
| EPI_ISL_418301 | Virology Department, Sheffield Teaching Hospitals NHS Foundation Trust | Department of Infection, Immunity and Cardiovascular Disease, The Florey Institute, The Medical School, University of Sheffield |
| EPI_ISL_418302 | Virology Department, Sheffield Teaching Hospitals NHS Foundation Trust | Department of Infection, Immunity and Cardiovascular Disease, The Florey Institute, The Medical School, University of Sheffield |
| EPI_ISL_418303 | Virology Department, Sheffield Teaching Hospitals NHS Foundation Trust | Department of Infection, Immunity and Cardiovascular Disease, The Florey Institute, The Medical School, University of Sheffield |
| EPI_ISL_418304 | Virology Department, Sheffield Teaching Hospitals NHS Foundation Trust | Department of Infection, Immunity and Cardiovascular Disease, The Florey Institute, The Medical School, University of Sheffield |

| EPI_Accession_ID | Originating Laboratory | Submitting Laboratory |
|---|---|---|
| EPI_ISL_418305 | Virology Department, Sheffield Teaching Hospitals NHS Foundation Trust | Department of Infection, Immunity and Cardiovascular Disease, The Florey Institute, The Medical School, University of Sheffield |
| EPI_ISL_418306 | Virology Department, Sheffield Teaching Hospitals NHS Foundation Trust | Department of Infection, Immunity and Cardiovascular Disease, The Florey Institute, The Medical School, University of Sheffield |
| EPI_ISL_418307 | Virology Department, Sheffield Teaching Hospitals NHS Foundation Trust | Department of Infection, Immunity and Cardiovascular Disease, The Florey Institute, The Medical School, University of Sheffield |
| EPI_ISL_418308 | Virology Department, Sheffield Teaching Hospitals NHS Foundation Trust | Department of Infection, Immunity and Cardiovascular Disease, The Florey Institute, The Medical School, University of Sheffield |
| EPI_ISL_418309 | Virology Department, Sheffield Teaching Hospitals NHS Foundation Trust | Department of Infection, Immunity and Cardiovascular Disease, The Florey Institute, The Medical School, University of Sheffield |
| EPI_ISL_418310 | Virology Department, Sheffield Teaching Hospitals NHS Foundation Trust | Department of Infection, Immunity and Cardiovascular Disease, The Florey Institute, The Medical School, University of Sheffield |
| EPI_ISL_418311 | Virology Department, Sheffield Teaching Hospitals NHS Foundation Trust | Department of Infection, Immunity and Cardiovascular Disease, The Florey Institute, The Medical School, University of Sheffield |
| EPI_ISL_418312 | Virology Department, Sheffield Teaching Hospitals NHS Foundation Trust | Department of Infection, Immunity and Cardiovascular Disease, The Florey Institute, The Medical School, University of Sheffield |
| EPI_ISL_418313 | Virology Department, Sheffield Teaching Hospitals NHS Foundation Trust | Department of Infection, Immunity and Cardiovascular Disease, The Florey Institute, The Medical School, University of Sheffield |
| EPI_ISL_418314 | Virology Department, Sheffield Teaching Hospitals NHS Foundation Trust | Department of Infection, Immunity and Cardiovascular Disease, The Florey Institute, The Medical School, University of Sheffield |
| EPI_ISL_418315 | Virology Department, Sheffield Teaching Hospitals NHS Foundation Trust | Department of Infection, Immunity and Cardiovascular Disease, The Florey Institute, The Medical School, University of Sheffield |

| EPI_Accession_ID | Originating Laboratory | Submitting Laboratory |
|---|---|---|
| EPI_ISL_418316 | Virology Department, Sheffield Teaching Hospitals NHS Foundation Trust | Department of Infection, Immunity and Cardiovascular Disease, The Florey Institute, The Medical School, University of Sheffield |
| EPI_ISL_418317 | Virology Department, Sheffield Teaching Hospitals NHS Foundation Trust | Department of Infection, Immunity and Cardiovascular Disease, The Florey Institute, The Medical School, University of Sheffield |
| EPI_ISL_418318 | Virology Department, Sheffield Teaching Hospitals NHS Foundation Trust | Department of Infection, Immunity and Cardiovascular Disease, The Florey Institute, The Medical School, University of Sheffield |
| EPI_ISL_418319 | Virology Department, Sheffield Teaching Hospitals NHS Foundation Trust | Department of Infection, Immunity and Cardiovascular Disease, The Florey Institute, The Medical School, University of Sheffield |
| EPI_ISL_418320 | Virology Department, Sheffield Teaching Hospitals NHS Foundation Trust | Department of Infection, Immunity and Cardiovascular Disease, The Florey Institute, The Medical School, University of Sheffield |
| EPI_ISL_418321 | Virology Department, Sheffield Teaching Hospitals NHS Foundation Trust | Department of Infection, Immunity and Cardiovascular Disease, The Florey Institute, The Medical School, University of Sheffield |
| EPI_ISL_418322 | Public Health Ontario Laboratories | Public Health Ontario Laboratories |
| EPI_ISL_418323 | Public Health Ontario Laboratories | Public Health Ontario Laboratories |
| EPI_ISL_418324 | Public Health Ontario Laboratories | Public Health Ontario Laboratories |
| EPI_ISL_418325 | Public Health Ontario Laboratories | Public Health Ontario Laboratories |
| EPI_ISL_418326 | Public Health Ontario Laboratories | Public Health Ontario Laboratories |
| EPI_ISL_418327 | Public Health Ontario Laboratories | Public Health Ontario Laboratories |
| EPI_ISL_418328 | Public Health Ontario Laboratories | Public Health Ontario Laboratories |
| EPI_ISL_418329 | Public Health Ontario Laboratories | Public Health Ontario Laboratories |
| EPI_ISL_418330 | Public Health Ontario Laboratories | Public Health Ontario Laboratories |
| EPI_ISL_418331 | Public Health Ontario Laboratories | Public Health Ontario Laboratories |
| EPI_ISL_418332 | Public Health Ontario Laboratories | Public Health Ontario Laboratories |
| EPI_ISL_418335 | Public Health Ontario Laboratories | Public Health Ontario Laboratories |
| EPI_ISL_418336 | Public Health Ontario Laboratories | Public Health Ontario Laboratories |
| EPI_ISL_418337 | Public Health Ontario Laboratories | Public Health Ontario Laboratories |
| EPI_ISL_418338 | Public Health Ontario Laboratories | Public Health Ontario Laboratories |

| EPI_Accession_ID | Originating Laboratory | Submitting Laboratory |
|---|---|---|
| EPI_ISL_418339 | Public Health Ontario Laboratories | Public Health Ontario Laboratories |
| EPI_ISL_418340 | Public Health Ontario Laboratories | Public Health Ontario Laboratories |
| EPI_ISL_418341 | Public Health Ontario Laboratories | Public Health Ontario Laboratories |
| EPI_ISL_418342 | Public Health Ontario Laboratories | Public Health Ontario Laboratories |
| EPI_ISL_418343 | Public Health Ontario Laboratories | Public Health Ontario Laboratories |
| EPI_ISL_418344 | Public Health Ontario Laboratories | Public Health Ontario Laboratories |
| EPI_ISL_418345 | Public Health Ontario Laboratories | Public Health Ontario Laboratories |
| EPI_ISL_418346 | Public Health Ontario Laboratories | Public Health Ontario Laboratories |
| EPI_ISL_418347 | Public Health Ontario Laboratories | Public Health Ontario Laboratories |
| EPI_ISL_418348 | Public Health Ontario Laboratories | Public Health Ontario Laboratories |
| EPI_ISL_418349 | Public Health Ontario Laboratories | Public Health Ontario Laboratories |
| EPI_ISL_418350 | Public Health Ontario Laboratories | Public Health Ontario Laboratories |
| EPI_ISL_418351 | Public Health Ontario Laboratories | Public Health Ontario Laboratories |
| EPI_ISL_418352 | Public Health Ontario Laboratories | Public Health Ontario Laboratories |
| EPI_ISL_418353 | Public Health Ontario Laboratories | Public Health Ontario Laboratories |
| EPI_ISL_418354 | Public Health Ontario Laboratories | Public Health Ontario Laboratories |
| EPI_ISL_418355 | Public Health Ontario Laboratories | Public Health Ontario Laboratories |
| EPI_ISL_418356 | Public Health Ontario Laboratories | Public Health Ontario Laboratories |
| EPI_ISL_418357 | Public Health Ontario Laboratories | Public Health Ontario Laboratories |
| EPI_ISL_418358 | Public Health Ontario Laboratories | Public Health Ontario Laboratories |
| EPI_ISL_418359 | Public Health Ontario Laboratories | Public Health Ontario Laboratories |
| EPI_ISL_418360 | Public Health Ontario Laboratories | Public Health Ontario Laboratories |
| EPI_ISL_418361 | Public Health Ontario Laboratories | Public Health Ontario Laboratories |
| EPI_ISL_418362 | Public Health Ontario Laboratories | Public Health Ontario Laboratories |
| EPI_ISL_418363 | Public Health Ontario Laboratories | Public Health Ontario Laboratories |
| EPI_ISL_418364 | Public Health Ontario Laboratories | Public Health Ontario Laboratories |
| EPI_ISL_418365 | Public Health Ontario Laboratories | Public Health Ontario Laboratories |
| EPI_ISL_418366 | Public Health Ontario Laboratories | Public Health Ontario Laboratories |
| EPI_ISL_418367 | Public Health Ontario Laboratories | Public Health Ontario Laboratories |
| EPI_ISL_418368 | Public Health Ontario Laboratories | Public Health Ontario Laboratories |
| EPI_ISL_418369 | Public Health Ontario Laboratories | Public Health Ontario Laboratories |
| EPI_ISL_418370 | Public Health Ontario Laboratories | Public Health Ontario Laboratories |
| EPI_ISL_418371 | Public Health Ontario Laboratories | Public Health Ontario Laboratories |

| EPI_Accession_ID | Originating Laboratory | Submitting Laboratory |
|---|---|---|
| EPI_ISL_418372 | Public Health Ontario Laboratories | Public Health Ontario Laboratories |
| EPI_ISL_418373 | Public Health Ontario Laboratories | Public Health Ontario Laboratories |
| EPI_ISL_418374 | Public Health Ontario Laboratories | Public Health Ontario Laboratories |
| EPI_ISL_418375 | Public Health Ontario Laboratories | Public Health Ontario Laboratories |
| EPI_ISL_418376 | Public Health Ontario Laboratories | Public Health Ontario Laboratories |
| EPI_ISL_418377 | Public Health Ontario Laboratories | Public Health Ontario Laboratories |
| EPI_ISL_418378 | Public Health Ontario Laboratories | Public Health Ontario Laboratories |
| EPI_ISL_418379 | Public Health Ontario Laboratories | Public Health Ontario Laboratories |
| EPI_ISL_418380 | Public Health Ontario Laboratories | Public Health Ontario Laboratories |
| EPI_ISL_418381 | Public Health Ontario Laboratories | Public Health Ontario Laboratories |
| EPI_ISL_418382 | Public Health Ontario Laboratories | Public Health Ontario Laboratories |
| EPI_ISL_418383 | Public Health Ontario Laboratories | Public Health Ontario Laboratories |
| EPI_ISL_418385 | Department of Virology and Immunology, University of Helsinki and Helsinki University Hospital, Huslab Finland | Department of Virology, Faculty of Medicine, University of Helsinki, Helsinki, Finland |
| EPI_ISL_418386 | Department of Virology and Immunology, University of Helsinki and Helsinki University Hospital, Huslab Finland | Department of Virology, Faculty of Medicine, University of Helsinki, Helsinki, Finland |
| EPI_ISL_418387 | Department of Virology and Immunology, University of Helsinki and Helsinki University Hospital, Huslab Finland | Department of Virology, Faculty of Medicine, University of Helsinki, Helsinki, Finland |
| EPI_ISL_418388 | Department of Virology and Immunology, University of Helsinki and Helsinki University Hospital, Huslab Finland | Department of Virology, Faculty of Medicine, University of Helsinki, Helsinki, Finland |
| EPI_ISL_418389 | Department of Virology and Immunology, University of Helsinki and Helsinki University Hospital, Huslab Finland | Department of Virology, Faculty of Medicine, University of Helsinki, Helsinki, Finland |
| EPI_ISL_418390 | Department of Virology and Immunology, University of Helsinki and Helsinki University Hospital, Huslab Finland | Department of Virology, Faculty of Medicine, University of Helsinki, Helsinki, Finland |
| EPI_ISL_418391 | Department of Virology and Immunology, University of Helsinki and Helsinki University Hospital, Huslab Finland | Department of Virology, Faculty of Medicine, University of Helsinki, Helsinki, Finland |

| EPI_Accession_ID | Originating Laboratory | Submitting Laboratory |
|---|---|---|
| EPI_ISL_418392 | Department of Virology and Immunology, University of Helsinki and Helsinki University Hospital, Huslab Finland | Department of Virology, Faculty of Medicine, University of Helsinki, Helsinki, Finland |
| EPI_ISL_418393 | Department of Virology and Immunology, University of Helsinki and Helsinki University Hospital, Huslab Finland | Department of Virology, Faculty of Medicine, University of Helsinki, Helsinki, Finland |
| EPI_ISL_418394 | Department of Virology and Immunology, University of Helsinki and Helsinki University Hospital, Huslab Finland | Department of Virology, Faculty of Medicine, University of Helsinki, Helsinki, Finland |
| EPI_ISL_418395 | Department of Virology and Immunology, University of Helsinki and Helsinki University Hospital, Huslab Finland | Department of Virology, Faculty of Medicine, University of Helsinki, Helsinki, Finland |
| EPI_ISL_418396 | Department of Virology and Immunology, University of Helsinki and Helsinki University Hospital, Huslab Finland | Department of Virology, Faculty of Medicine, University of Helsinki, Helsinki, Finland |
| EPI_ISL_418397 | Department of Virology and Immunology, University of Helsinki and Helsinki University Hospital, Huslab Finland | Department of Virology, Faculty of Medicine, University of Helsinki, Helsinki, Finland |
| EPI_ISL_418398 | Department of Virology and Immunology, University of Helsinki and Helsinki University Hospital, Huslab Finland | Department of Virology, Faculty of Medicine, University of Helsinki, Helsinki, Finland |
| EPI_ISL_418399 | Department of Virology and Immunology, University of Helsinki and Helsinki University Hospital, Huslab Finland | Department of Virology, Faculty of Medicine, University of Helsinki, Helsinki, Finland |
| EPI_ISL_418400 | Department of Virology and Immunology, University of Helsinki and Helsinki University Hospital, Huslab Finland | Department of Virology, Faculty of Medicine, University of Helsinki, Helsinki, Finland |
| EPI_ISL_418401 | Department of Virology and Immunology, University of Helsinki and Helsinki University Hospital, Huslab Finland | Department of Virology, Faculty of Medicine, University of Helsinki, Helsinki, Finland |
| EPI_ISL_418402 | Department of Virology and Immunology, University of Helsinki and Helsinki University Hospital, Huslab Finland | Department of Virology, Faculty of Medicine, University of Helsinki, Helsinki, Finland |

| EPI_Accession_ID | Originating Laboratory | Submitting Laboratory |
|---|---|---|
| EPI_ISL_418403 | Department of Virology and Immunology, University of Helsinki and Helsinki University Hospital, Huslab Finland | Department of Virology, Faculty of Medicine, University of Helsinki, Helsinki, Finland |
| EPI_ISL_418404 | Department of Virology and Immunology, University of Helsinki and Helsinki University Hospital, Huslab Finland | Department of Virology, Faculty of Medicine, University of Helsinki, Helsinki, Finland |
| EPI_ISL_418405 | Department of Virology and Immunology, University of Helsinki and Helsinki University Hospital, Huslab Finland | Department of Virology, Faculty of Medicine, University of Helsinki, Helsinki, Finland |
| EPI_ISL_418406 | Department of Virology and Immunology, University of Helsinki and Helsinki University Hospital, Huslab Finland | Department of Virology, Faculty of Medicine, University of Helsinki, Helsinki, Finland |
| EPI_ISL_418407 | Department of Virology and Immunology, University of Helsinki and Helsinki University Hospital, Huslab Finland | Department of Virology, Faculty of Medicine, University of Helsinki, Helsinki, Finland |
| EPI_ISL_418408 | Department of Virology and Immunology, University of Helsinki and Helsinki University Hospital, Huslab Finland | Department of Virology, Faculty of Medicine, University of Helsinki, Helsinki, Finland |
| EPI_ISL_418409 | Department of Virology and Immunology, University of Helsinki and Helsinki University Hospital, Huslab Finland | Department of Virology, Faculty of Medicine, University of Helsinki, Helsinki, Finland |
| EPI_ISL_418410 | Department of Virology and Immunology, University of Helsinki and Helsinki University Hospital, Huslab Finland | Department of Virology, Faculty of Medicine, University of Helsinki, Helsinki, Finland |
| EPI_ISL_418411 | Department of Virology and Immunology, University of Helsinki and Helsinki University Hospital, Huslab Finland | Department of Virology, Faculty of Medicine, University of Helsinki, Helsinki, Finland |
| EPI_ISL_418412 | Centre Hospitalier des Vals d'Ardeche | CNR Virus des Infections Respiratoires - France SUD |
| EPI_ISL_418413 | Centre Hospitalier de Macon | CNR Virus des Infections Respiratoires - France SUD |
| EPI_ISL_418414 | Centre Hospitalier de Valence | CNR Virus des Infections Respiratoires - France SUD |
| EPI_ISL_418415 | Centre Hospitalier de Valence | CNR Virus des Infections Respiratoires - France SUD |
| EPI_ISL_418416 | GH Les Portes du Sud | CNR Virus des Infections Respiratoires - France SUD |
| EPI_ISL_418417 | Centre Hospitalier de Valence | CNR Virus des Infections Respiratoires - France SUD |

| EPI_Accession_ID | Originating Laboratory | Submitting Laboratory |
|---|---|---|
| EPI_ISL_418418 | Centre Hospitalier Saint Joseph Saint Luc | CNR Virus des Infections Respiratoires - France SUD |
| EPI_ISL_418419 | Centre Hospitalier Saint Joseph Saint Luc | CNR Virus des Infections Respiratoires - France SUD |
| EPI_ISL_418420 | Institut des Agents Infectieux (IAI), Hospices Civils de Lyon | CNR Virus des Infections Respiratoires - France SUD |
| EPI_ISL_418421 | Institut des Agents Infectieux (IAI), Hospices Civils de Lyon | CNR Virus des Infections Respiratoires - France SUD |
| EPI_ISL_418422 | Institut des Agents Infectieux (IAI), Hospices Civils de Lyon | CNR Virus des Infections Respiratoires - France SUD |
| EPI_ISL_418423 | Institut des Agents Infectieux (IAI), Hospices Civils de Lyon | CNR Virus des Infections Respiratoires - France SUD |
| EPI_ISL_418424 | Institut des Agents Infectieux (IAI), Hospices Civils de Lyon | CNR Virus des Infections Respiratoires - France SUD |
| EPI_ISL_418425 | Institut des Agents Infectieux (IAI), Hospices Civils de Lyon | CNR Virus des Infections Respiratoires - France SUD |
| EPI_ISL_418426 | Centre Hospitalier de Bourg en Bresse | CNR Virus des Infections Respiratoires - France SUD |
| EPI_ISL_418427 | Hopital PrivÃ© de l'Est Lyonnais | CNR Virus des Infections Respiratoires - France SUD |
| EPI_ISL_418428 | Centre Hospitalier Lucien Hussel | CNR Virus des Infections Respiratoires - France SUD |
| EPI_ISL_418429 | Institut des Agents Infectieux (IAI), Hospices Civils de Lyon | CNR Virus des Infections Respiratoires - France SUD |
| EPI_ISL_418430 | Institut des Agents Infectieux (IAI), Hospices Civils de Lyon | CNR Virus des Infections Respiratoires - France SUD |
| EPI_ISL_418431 | Institut des Agents Infectieux (IAI), Hospices Civils de Lyon | CNR Virus des Infections Respiratoires - France SUD |
| EPI_ISL_418432 | Institut des Agents Infectieux (IAI), Hospices Civils de Lyon | CNR Virus des Infections Respiratoires - France SUD |
| EPI_ISL_418433 | University Hospital Basel, Clinical Virology | University Hospital Basel, Clinical Bacteriology |
| EPI_ISL_418434 | University Hospital Basel, Clinical Virology | University Hospital Basel, Clinical Bacteriology |
| EPI_ISL_418435 | University Hospital Basel, Clinical Virology | University Hospital Basel, Clinical Bacteriology |
| EPI_ISL_418436 | University Hospital Basel, Clinical Virology | University Hospital Basel, Clinical Virology |
| EPI_ISL_418437 | University Hospital Basel, Clinical Virology | University Hospital Basel, Clinical Bacteriology |
| EPI_ISL_418438 | University Hospital Basel, Clinical Virology | University Hospital Basel, Clinical Bacteriology |
| EPI_ISL_418439 | University Hospital Basel, Clinical Virology | University Hospital Basel, Clinical Bacteriology |
| EPI_ISL_418440 | University Hospital Basel, Clinical Virology | University Hospital Basel, Clinical Bacteriology |

| EPI_Accession_ID | Originating Laboratory | Submitting Laboratory |
|---|---|---|
| EPI_ISL_418441 | Hangzhou Center for Disease Control and Prevention | Inspection Center of Hangzhou Center for Disease Control and Prevention |
| EPI_ISL_418442 | Hangzhou Center for Disease Control and Prevention | Inspection Center of Hangzhou Center for Disease Control and Prevention |
| EPI_ISL_418502 | Hangzhou Center for Disease Control and Prevention | Inspection Center of Hangzhou Center for Disease Control and Prevention |
| EPI_ISL_418503 | Hangzhou Center for Disease Control and Prevention | Inspection Center of Hangzhou Center for Disease Control and Prevention |
| EPI_ISL_418504 | Hangzhou Center for Disease Control and Prevention | Inspection Center of Hangzhou Center for Disease Control and Prevention |
| EPI_ISL_418506 | Hangzhou Center for Disease Control and Prevention | Inspection Center of Hanghzou Center for Disease Control and Prevention |
| EPI_ISL_418507 | Hangzhou Center for Disease Control and Prevention | Inspection Center of Hangzhou Center for Disease Control and Prevention |
| EPI_ISL_418508 | Hangzhou Center for Disease Control and Prevention | Inspection Center of Hangzhou Center for Disease Control and Prevention |
| EPI_ISL_418509 | Hangzhou Center for Disease Control and Prevention | Inspection Center of Hangzhou Center for Disease Control and Prevention |
| EPI_ISL_418510 | Hangzhou Center for Disease Control and Prevention | Insepction Center of Hangzhou Center for Disease Control and Prevention |
| EPI_ISL_418511 | Hangzhou Center for Disease Control and Prevention | Inspection Center of Hangzhou Center for Disease Control and Prevention |
| EPI_ISL_418512 | Hangzhou Center for Disease Control and Prevention | Inspection Center of Hangzhou Center for Disease Control and Prevention |
| EPI_ISL_418513 | Hangzhou Center for Disease Control and Prevention | Inspection Center of Hangzhou Center for Disease Control and Prevention |
| EPI_ISL_418514 | Hangzhou Center for Disease Control and Prevention | Inspection Center of Hangzhou Center for Disease Control and Prevention |
| EPI_ISL_418515 | Hangzhou Center for Disease Control and Prevention | Inspection Center of Hangzhou Center for Disease Control and Prevention |
| EPI_ISL_418516 | UCD National Virus Reference Laboratory | UCD National Virus Reference Laboratory |
| EPI_ISL_418548 | UCD National Virus Reference Laboratory | UCD National Virus Reference Laboratory |
| EPI_ISL_418580 | UCD National Virus Reference Laboratory | UCD National Virus Reference Laboratory |

| EPI_Accession_ID | Originating Laboratory | Submitting Laboratory |
|---|---|---|
| EPI_ISL_418581 | UCD National Virus Reference Laboratory | UCD National Virus Reference Laboratory |
| EPI_ISL_418582 | UCD National Virus Reference Laboratory | UCD National Virus Reference Laboratory |
| EPI_ISL_418583 | UCD National Virus Reference Laboratory | UCD National Virus Reference Laboratory |
| EPI_ISL_418584 | UCD National Virus Reference Laboratory | UCD National Virus Reference Laboratory |
| EPI_ISL_418623 | Department of Clinical Microbiology | GIGA Medical Genomics |
| EPI_ISL_418624 | Department of Clinical Microbiology | GIGA Medical Genomics |
| EPI_ISL_418625 | Department of Clinical Microbiology | GIGA Medical Genomics |
| EPI_ISL_418626 | Department of Clinical Microbiology | GIGA Medical Genomics |
| EPI_ISL_418627 | Department of Clinical Microbiology | GIGA Medical Genomics |
| EPI_ISL_418628 | Department of Clinical Microbiology | GIGA Medical Genomics |
| EPI_ISL_418629 | Department of Clinical Microbiology | GIGA Medical Genomics |
| EPI_ISL_418630 | Department of Clinical Microbiology | GIGA Medical Genomics |
| EPI_ISL_418631 | Department of Clinical Microbiology | GIGA Medical Genomics |
| EPI_ISL_418632 | Department of Clinical Microbiology | GIGA Medical Genomics |
| EPI_ISL_418633 | Department of Clinical Microbiology | GIGA Medical Genomics |
| EPI_ISL_418634 | Department of Clinical Microbiology | GIGA Medical Genomics |
| EPI_ISL_418635 | Department of Clinical Microbiology | GIGA Medical Genomics |
| EPI_ISL_418636 | Department of Clinical Microbiology | GIGA Medical Genomics |
| EPI_ISL_418637 | Department of Clinical Microbiology | GIGA Medical Genomics |
| EPI_ISL_418638 | Department of Clinical Microbiology | GIGA Medical Genomics |
| EPI_ISL_418639 | Department of Clinical Microbiology | GIGA Medical Genomics |
| EPI_ISL_418640 | Department of Clinical Microbiology | GIGA Medical Genomics |
| EPI_ISL_418641 | Department of Clinical Microbiology | GIGA Medical Genomics |
| EPI_ISL_418642 | Department of Clinical Microbiology | GIGA Medical Genomics |
| EPI_ISL_418643 | Department of Clinical Microbiology | GIGA Medical Genomics |
| EPI_ISL_418644 | Department of Clinical Microbiology | GIGA Medical Genomics |
| EPI_ISL_418645 | Department of Clinical Microbiology | GIGA Medical Genomics |
| EPI_ISL_418646 | Department of Clinical Microbiology | GIGA Medical Genomics |
| EPI_ISL_418647 | Department of Clinical Microbiology | GIGA Medical Genomics |
| EPI_ISL_418648 | Department of Clinical Microbiology | GIGA Medical Genomics |
| EPI_ISL_418649 | Department of Clinical Microbiology | GIGA Medical Genomics |
| EPI_ISL_418650 | Department of Clinical Microbiology | GIGA Medical Genomics |
| EPI_ISL_418651 | Department of Clinical Microbiology | GIGA Medical Genomics |

| EPI_Accession_ID | Originating Laboratory | Submitting Laboratory |
|---|---|---|
| EPI_ISL_418652 | Department of Clinical Microbiology | GIGA Medical Genomics |
| EPI_ISL_418653 | Department of Clinical Microbiology | GIGA Medical Genomics |
| EPI_ISL_418654 | Department of Clinical Microbiology | GIGA Medical Genomics |
| EPI_ISL_418655 | Department of Clinical Microbiology | GIGA Medical Genomics |
| EPI_ISL_418656 | Department of Clinical Microbiology | GIGA Medical Genomics |
| EPI_ISL_418657 | Department of Clinical Microbiology | GIGA Medical Genomics |
| EPI_ISL_418658 | Department of Clinical Microbiology | GIGA Medical Genomics |
| EPI_ISL_418659 | Department of Clinical Microbiology | GIGA Medical Genomics |
| EPI_ISL_418660 | Department of Clinical Microbiology | GIGA Medical Genomics |
| EPI_ISL_418661 | Department of Clinical Microbiology | GIGA Medical Genomics |
| EPI_ISL_418662 | Department of Clinical Microbiology | GIGA Medical Genomics |
| EPI_ISL_418663 | Department of Clinical Microbiology | GIGA Medical Genomics |
| EPI_ISL_418664 | Department of Clinical Microbiology | GIGA Medical Genomics |
| EPI_ISL_418665 | Department of Clinical Microbiology | GIGA Medical Genomics |
| EPI_ISL_418666 | Department of Clinical Microbiology | GIGA Medical Genomics |
| EPI_ISL_418667 | Respiratory Virus Unit, Microbiology Services Colindale, Public Health England | Respiratory Virus Unit, Microbiology Services Colindale, Public Health England |
| EPI_ISL_418668 | Respiratory Virus Unit, Microbiology Services Colindale, Public Health England | Respiratory Virus Unit, Microbiology Services Colindale, Public Health England |
| EPI_ISL_418669 | Respiratory Virus Unit, Microbiology Services Colindale, Public Health England | Respiratory Virus Unit, Microbiology Services Colindale, Public Health England |
| EPI_ISL_418670 | Respiratory Virus Unit, Microbiology Services Colindale, Public Health England | Respiratory Virus Unit, Microbiology Services Colindale, Public Health England |
| EPI_ISL_418671 | Respiratory Virus Unit, Microbiology Services Colindale, Public Health England | Respiratory Virus Unit, Microbiology Services Colindale, Public Health England |
| EPI_ISL_418672 | Respiratory Virus Unit, Microbiology Services Colindale, Public Health England | Respiratory Virus Unit, Microbiology Services Colindale, Public Health England |
| EPI_ISL_418673 | Respiratory Virus Unit, Microbiology Services Colindale, Public Health England | Respiratory Virus Unit, Microbiology Services Colindale, Public Health England |
| EPI_ISL_418674 | Respiratory Virus Unit, Microbiology Services Colindale, Public Health England | Respiratory Virus Unit, Microbiology Services Colindale, Public Health England |
| EPI_ISL_418675 | Respiratory Virus Unit, Microbiology Services Colindale, Public Health England | Respiratory Virus Unit, Microbiology Services Colindale, Public Health England |

| EPI_Accession_ID | Originating Laboratory | Submitting Laboratory |
|---|---|---|
| EPI_ISL_418676 | Respiratory Virus Unit, Microbiology Services Colindale, Public Health England | Respiratory Virus Unit, Microbiology Services Colindale, Public Health England |
| EPI_ISL_418677 | Respiratory Virus Unit, Microbiology Services Colindale, Public Health England | Respiratory Virus Unit, Microbiology Services Colindale, Public Health England |
| EPI_ISL_418678 | Respiratory Virus Unit, Microbiology Services Colindale, Public Health England | Respiratory Virus Unit, Microbiology Services Colindale, Public Health England |
| EPI_ISL_418679 | Respiratory Virus Unit, Microbiology Services Colindale, Public Health England | Respiratory Virus Unit, Microbiology Services Colindale, Public Health England |
| EPI_ISL_418680 | Respiratory Virus Unit, Microbiology Services Colindale, Public Health England | Respiratory Virus Unit, Microbiology Services Colindale, Public Health England |
| EPI_ISL_418681 | Respiratory Virus Unit, Microbiology Services Colindale, Public Health England | Respiratory Virus Unit, Microbiology Services Colindale, Public Health England |
| EPI_ISL_418682 | Respiratory Virus Unit, Microbiology Services Colindale, Public Health England | Respiratory Virus Unit, Microbiology Services Colindale, Public Health England |
| EPI_ISL_418683 | Respiratory Virus Unit, Microbiology Services Colindale, Public Health England | Respiratory Virus Unit, Microbiology Services Colindale, Public Health England |
| EPI_ISL_418684 | Respiratory Virus Unit, Microbiology Services Colindale, Public Health England | Respiratory Virus Unit, Microbiology Services Colindale, Public Health England |
| EPI_ISL_418685 | Respiratory Virus Unit, Microbiology Services Colindale, Public Health England | Respiratory Virus Unit, Microbiology Services Colindale, Public Health England |
| EPI_ISL_418686 | Respiratory Virus Unit, Microbiology Services Colindale, Public Health England | Respiratory Virus Unit, Microbiology Services Colindale, Public Health England |
| EPI_ISL_418687 | Respiratory Virus Unit, Microbiology Services Colindale, Public Health England | Respiratory Virus Unit, Microbiology Services Colindale, Public Health England |
| EPI_ISL_418688 | Respiratory Virus Unit, Microbiology Services Colindale, Public Health England | Respiratory Virus Unit, Microbiology Services Colindale, Public Health England |
| EPI_ISL_418689 | Respiratory Virus Unit, Microbiology Services Colindale, Public Health England | Respiratory Virus Unit, Microbiology Services Colindale, Public Health England |
| EPI_ISL_418690 | Respiratory Virus Unit, Microbiology Services Colindale, Public Health England | Respiratory Virus Unit, Microbiology Services Colindale, Public Health England |
| EPI_ISL_418691 | Respiratory Virus Unit, Microbiology Services Colindale, Public Health England | Respiratory Virus Unit, Microbiology Services Colindale, Public Health England |

| EPI_Accession_ID | Originating Laboratory | Submitting Laboratory |
|---|---|---|
| EPI_ISL_418692 | Respiratory Virus Unit, Microbiology Services Colindale, Public Health England | Respiratory Virus Unit, Microbiology Services Colindale, Public Health England |
| EPI_ISL_418693 | Respiratory Virus Unit, Microbiology Services Colindale, Public Health England | Respiratory Virus Unit, Microbiology Services Colindale, Public Health England |
| EPI_ISL_418694 | Respiratory Virus Unit, Microbiology Services Colindale, Public Health England | Respiratory Virus Unit, Microbiology Services Colindale, Public Health England |
| EPI_ISL_418695 | Respiratory Virus Unit, Microbiology Services Colindale, Public Health England | Respiratory Virus Unit, Microbiology Services Colindale, Public Health England |
| EPI_ISL_418696 | Respiratory Virus Unit, Microbiology Services Colindale, Public Health England | Respiratory Virus Unit, Microbiology Services Colindale, Public Health England |
| EPI_ISL_418697 | Respiratory Virus Unit, Microbiology Services Colindale, Public Health England | Respiratory Virus Unit, Microbiology Services Colindale, Public Health England |
| EPI_ISL_418698 | Respiratory Virus Unit, Microbiology Services Colindale, Public Health England | Respiratory Virus Unit, Microbiology Services Colindale, Public Health England |
| EPI_ISL_418699 | Respiratory Virus Unit, Microbiology Services Colindale, Public Health England | Respiratory Virus Unit, Microbiology Services Colindale, Public Health England |
| EPI_ISL_418700 | Respiratory Virus Unit, Microbiology Services Colindale, Public Health England | Respiratory Virus Unit, Microbiology Services Colindale, Public Health England |
| EPI_ISL_418701 | Respiratory Virus Unit, Microbiology Services Colindale, Public Health England | Respiratory Virus Unit, Microbiology Services Colindale, Public Health England |
| EPI_ISL_418702 | Respiratory Virus Unit, Microbiology Services Colindale, Public Health England | Respiratory Virus Unit, Microbiology Services Colindale, Public Health England |
| EPI_ISL_418703 | Respiratory Virus Unit, Microbiology Services Colindale, Public Health England | Respiratory Virus Unit, Microbiology Services Colindale, Public Health England |
| EPI_ISL_418704 | Respiratory Virus Unit, Microbiology Services Colindale, Public Health England | Respiratory Virus Unit, Microbiology Services Colindale, Public Health England |
| EPI_ISL_418705 | Respiratory Virus Unit, Microbiology Services Colindale, Public Health England | Respiratory Virus Unit, Microbiology Services Colindale, Public Health England |
| EPI_ISL_418706 | Respiratory Virus Unit, Microbiology Services Colindale, Public Health England | Respiratory Virus Unit, Microbiology Services Colindale, Public Health England |
| EPI_ISL_418707 | Respiratory Virus Unit, Microbiology Services Colindale, Public Health England | Respiratory Virus Unit, Microbiology Services Colindale, Public Health England |

| EPI_Accession_ID | Originating Laboratory | Submitting Laboratory |
|---|---|---|
| EPI_ISL_418708 | Respiratory Virus Unit, Microbiology Services Colindale, Public Health England | Respiratory Virus Unit, Microbiology Services Colindale, Public Health England |
| EPI_ISL_418709 | Respiratory Virus Unit, Microbiology Services Colindale, Public Health England | Respiratory Virus Unit, Microbiology Services Colindale, Public Health England |
| EPI_ISL_418710 | Respiratory Virus Unit, Microbiology Services Colindale, Public Health England | Respiratory Virus Unit, Microbiology Services Colindale, Public Health England |
| EPI_ISL_418711 | Respiratory Virus Unit, Microbiology Services Colindale, Public Health England | Respiratory Virus Unit, Microbiology Services Colindale, Public Health England |
| EPI_ISL_418712 | Respiratory Virus Unit, Microbiology Services Colindale, Public Health England | Respiratory Virus Unit, Microbiology Services Colindale, Public Health England |
| EPI_ISL_418713 | Respiratory Virus Unit, Microbiology Services Colindale, Public Health England | Respiratory Virus Unit, Microbiology Services Colindale, Public Health England |
| EPI_ISL_418714 | Respiratory Virus Unit, Microbiology Services Colindale, Public Health England | Respiratory Virus Unit, Microbiology Services Colindale, Public Health England |
| EPI_ISL_418715 | Respiratory Virus Unit, Microbiology Services Colindale, Public Health England | Respiratory Virus Unit, Microbiology Services Colindale, Public Health England |
| EPI_ISL_418716 | Respiratory Virus Unit, Microbiology Services Colindale, Public Health England | Respiratory Virus Unit, Microbiology Services Colindale, Public Health England |
| EPI_ISL_418717 | Respiratory Virus Unit, Microbiology Services Colindale, Public Health England | Respiratory Virus Unit, Microbiology Services Colindale, Public Health England |
| EPI_ISL_418718 | Respiratory Virus Unit, Microbiology Services Colindale, Public Health England | Respiratory Virus Unit, Microbiology Services Colindale, Public Health England |
| EPI_ISL_418719 | Respiratory Virus Unit, Microbiology Services Colindale, Public Health England | Respiratory Virus Unit, Microbiology Services Colindale, Public Health England |
| EPI_ISL_418720 | Respiratory Virus Unit, Microbiology Services Colindale, Public Health England | Respiratory Virus Unit, Microbiology Services Colindale, Public Health England |
| EPI_ISL_418721 | Respiratory Virus Unit, Microbiology Services Colindale, Public Health England | Respiratory Virus Unit, Microbiology Services Colindale, Public Health England |
| EPI_ISL_418722 | Respiratory Virus Unit, Microbiology Services Colindale, Public Health England | Respiratory Virus Unit, Microbiology Services Colindale, Public Health England |
| EPI_ISL_418723 | Respiratory Virus Unit, Microbiology Services Colindale, Public Health England | Respiratory Virus Unit, Microbiology Services Colindale, Public Health England |

| EPI_Accession_ID | Originating Laboratory | Submitting Laboratory |
|---|---|---|
| EPI_ISL_418724 | Respiratory Virus Unit, Microbiology Services Colindale, Public Health England | Respiratory Virus Unit, Microbiology Services Colindale, Public Health England |
| EPI_ISL_418725 | Respiratory Virus Unit, Microbiology Services Colindale, Public Health England | Respiratory Virus Unit, Microbiology Services Colindale, Public Health England |
| EPI_ISL_418726 | Respiratory Virus Unit, Microbiology Services Colindale, Public Health England | Respiratory Virus Unit, Microbiology Services Colindale, Public Health England |
| EPI_ISL_418727 | Respiratory Virus Unit, Microbiology Services Colindale, Public Health England | Respiratory Virus Unit, Microbiology Services Colindale, Public Health England |
| EPI_ISL_418728 | Respiratory Virus Unit, Microbiology Services Colindale, Public Health England | Respiratory Virus Unit, Microbiology Services Colindale, Public Health England |
| EPI_ISL_418729 | Respiratory Virus Unit, Microbiology Services Colindale, Public Health England | Respiratory Virus Unit, Microbiology Services Colindale, Public Health England |
| EPI_ISL_418730 | Respiratory Virus Unit, Microbiology Services Colindale, Public Health England | Respiratory Virus Unit, Microbiology Services Colindale, Public Health England |
| EPI_ISL_418731 | Respiratory Virus Unit, Microbiology Services Colindale, Public Health England | Respiratory Virus Unit, Microbiology Services Colindale, Public Health England |
| EPI_ISL_418732 | Respiratory Virus Unit, Microbiology Services Colindale, Public Health England | Respiratory Virus Unit, Microbiology Services Colindale, Public Health England |
| EPI_ISL_418733 | Respiratory Virus Unit, Microbiology Services Colindale, Public Health England | Respiratory Virus Unit, Microbiology Services Colindale, Public Health England |
| EPI_ISL_418734 | Respiratory Virus Unit, Microbiology Services Colindale, Public Health England | Respiratory Virus Unit, Microbiology Services Colindale, Public Health England |
| EPI_ISL_418735 | Respiratory Virus Unit, Microbiology Services Colindale, Public Health England | Respiratory Virus Unit, Microbiology Services Colindale, Public Health England |
| EPI_ISL_418736 | Respiratory Virus Unit, Microbiology Services Colindale, Public Health England | Respiratory Virus Unit, Microbiology Services Colindale, Public Health England |
| EPI_ISL_418737 | Respiratory Virus Unit, Microbiology Services Colindale, Public Health England | Respiratory Virus Unit, Microbiology Services Colindale, Public Health England |
| EPI_ISL_418738 | Respiratory Virus Unit, Microbiology Services Colindale, Public Health England | Respiratory Virus Unit, Microbiology Services Colindale, Public Health England |
| EPI_ISL_418739 | Respiratory Virus Unit, Microbiology Services Colindale, Public Health England | Respiratory Virus Unit, Microbiology Services Colindale, Public Health England |

| EPI_Accession_ID | Originating Laboratory | Submitting Laboratory |
|---|---|---|
| EPI_ISL_418740 | Respiratory Virus Unit, Microbiology Services Colindale, Public Health England | Respiratory Virus Unit, Microbiology Services Colindale, Public Health England |
| EPI_ISL_418741 | Respiratory Virus Unit, Microbiology Services Colindale, Public Health England | Respiratory Virus Unit, Microbiology Services Colindale, Public Health England |
| EPI_ISL_418742 | Respiratory Virus Unit, Microbiology Services Colindale, Public Health England | Respiratory Virus Unit, Microbiology Services Colindale, Public Health England |
| EPI_ISL_418743 | Respiratory Virus Unit, Microbiology Services Colindale, Public Health England | Respiratory Virus Unit, Microbiology Services Colindale, Public Health England |
| EPI_ISL_418744 | Respiratory Virus Unit, Microbiology Services Colindale, Public Health England | Respiratory Virus Unit, Microbiology Services Colindale, Public Health England |
| EPI_ISL_418745 | Respiratory Virus Unit, Microbiology Services Colindale, Public Health England | Respiratory Virus Unit, Microbiology Services Colindale, Public Health England |
| EPI_ISL_418746 | Respiratory Virus Unit, Microbiology Services Colindale, Public Health England | Respiratory Virus Unit, Microbiology Services Colindale, Public Health England |
| EPI_ISL_418747 | Respiratory Virus Unit, Microbiology Services Colindale, Public Health England | Respiratory Virus Unit, Microbiology Services Colindale, Public Health England |
| EPI_ISL_418748 | Respiratory Virus Unit, Microbiology Services Colindale, Public Health England | Respiratory Virus Unit, Microbiology Services Colindale, Public Health England |
| EPI_ISL_418749 | Respiratory Virus Unit, Microbiology Services Colindale, Public Health England | Respiratory Virus Unit, Microbiology Services Colindale, Public Health England |
| EPI_ISL_418750 | Respiratory Virus Unit, Microbiology Services Colindale, Public Health England | Respiratory Virus Unit, Microbiology Services Colindale, Public Health England |
| EPI_ISL_418751 | Respiratory Virus Unit, Microbiology Services Colindale, Public Health England | Respiratory Virus Unit, Microbiology Services Colindale, Public Health England |
| EPI_ISL_418752 | Respiratory Virus Unit, Microbiology Services Colindale, Public Health England | Respiratory Virus Unit, Microbiology Services Colindale, Public Health England |
| EPI_ISL_418753 | Respiratory Virus Unit, Microbiology Services Colindale, Public Health England | Respiratory Virus Unit, Microbiology Services Colindale, Public Health England |
| EPI_ISL_418754 | Respiratory Virus Unit, Microbiology Services Colindale, Public Health England | Respiratory Virus Unit, Microbiology Services Colindale, Public Health England |
| EPI_ISL_418755 | Respiratory Virus Unit, Microbiology Services Colindale, Public Health England | Respiratory Virus Unit, Microbiology Services Colindale, Public Health England |

| EPI_Accession_ID | Originating Laboratory | Submitting Laboratory |
|---|---|---|
| EPI_ISL_418756 | Respiratory Virus Unit, Microbiology Services Colindale, Public Health England | Respiratory Virus Unit, Microbiology Services Colindale, Public Health England |
| EPI_ISL_418757 | Respiratory Virus Unit, Microbiology Services Colindale, Public Health England | Respiratory Virus Unit, Microbiology Services Colindale, Public Health England |
| EPI_ISL_418758 | Respiratory Virus Unit, Microbiology Services Colindale, Public Health England | Respiratory Virus Unit, Microbiology Services Colindale, Public Health England |
| EPI_ISL_418759 | Respiratory Virus Unit, Microbiology Services Colindale, Public Health England | Respiratory Virus Unit, Microbiology Services Colindale, Public Health England |
| EPI_ISL_418760 | Respiratory Virus Unit, Microbiology Services Colindale, Public Health England | Respiratory Virus Unit, Microbiology Services Colindale, Public Health England |
| EPI_ISL_418761 | Respiratory Virus Unit, Microbiology Services Colindale, Public Health England | Respiratory Virus Unit, Microbiology Services Colindale, Public Health England |
| EPI_ISL_418762 | Respiratory Virus Unit, Microbiology Services Colindale, Public Health England | Respiratory Virus Unit, Microbiology Services Colindale, Public Health England |
| EPI_ISL_418763 | Respiratory Virus Unit, Microbiology Services Colindale, Public Health England | Respiratory Virus Unit, Microbiology Services Colindale, Public Health England |
| EPI_ISL_418764 | Respiratory Virus Unit, Microbiology Services Colindale, Public Health England | Respiratory Virus Unit, Microbiology Services Colindale, Public Health England |
| EPI_ISL_418765 | Respiratory Virus Unit, Microbiology Services Colindale, Public Health England | Respiratory Virus Unit, Microbiology Services Colindale, Public Health England |
| EPI_ISL_418766 | Respiratory Virus Unit, Microbiology Services Colindale, Public Health England | Respiratory Virus Unit, Microbiology Services Colindale, Public Health England |
| EPI_ISL_418767 | Respiratory Virus Unit, Microbiology Services Colindale, Public Health England | Respiratory Virus Unit, Microbiology Services Colindale, Public Health England |
| EPI_ISL_418768 | Respiratory Virus Unit, Microbiology Services Colindale, Public Health England | Respiratory Virus Unit, Microbiology Services Colindale, Public Health England |
| EPI_ISL_418769 | Respiratory Virus Unit, Microbiology Services Colindale, Public Health England | Respiratory Virus Unit, Microbiology Services Colindale, Public Health England |
| EPI_ISL_418770 | Respiratory Virus Unit, Microbiology Services Colindale, Public Health England | Respiratory Virus Unit, Microbiology Services Colindale, Public Health England |
| EPI_ISL_418771 | WA State Department of Health | Pathogen Discovery, Respiratory Viruses Branch, Division of Viral Diseases, Centers for Disease Control and Prevention |

| EPI_Accession_ID | Originating Laboratory | Submitting Laboratory |
|---|---|---|
| EPI_ISL_418772 | WA State Department of Health | Pathogen Discovery, Respiratory Viruses Branch, Division of Viral Diseases, Centers for Disease Control and Prevention |
| EPI_ISL_418773 | WA State Department of Health | Pathogen Discovery, Respiratory Viruses Branch, Division of Viral Diseases, Centers for Disease Control and Prevention |
| EPI_ISL_418774 | WA State Department of Health | Pathogen Discovery, Respiratory Viruses Branch, Division of Viral Diseases, Centers for Disease Control and Prevention |
| EPI_ISL_418775 | WA State Department of Health | Pathogen Discovery, Respiratory Viruses Branch, Division of Viral Diseases, Centers for Disease Control and Prevention |
| EPI_ISL_418776 | WA State Department of Health | Pathogen Discovery, Respiratory Viruses Branch, Division of Viral Diseases, Centers for Disease Control and Prevention |
| EPI_ISL_418777 | WA State Department of Health | Pathogen Discovery, Respiratory Viruses Branch, Division of Viral Diseases, Centers for Disease Control and Prevention |
| EPI_ISL_418778 | WA State Department of Health | Pathogen Discovery, Respiratory Viruses Branch, Division of Viral Diseases, Centers for Disease Control and Prevention |
| EPI_ISL_418779 | WA State Department of Health | Pathogen Discovery, Respiratory Viruses Branch, Division of Viral Diseases, Centers for Disease Control and Prevention |
| EPI_ISL_418780 | WA State Department of Health | Pathogen Discovery, Respiratory Viruses Branch, Division of Viral Diseases, Centers for Disease Control and Prevention |
| EPI_ISL_418781 | WA State Department of Health | Pathogen Discovery, Respiratory Viruses Branch, Division of Viral Diseases, Centers for Disease Control and Prevention |
| EPI_ISL_418782 | WA State Department of Health | Pathogen Discovery, Respiratory Viruses Branch, Division of Viral Diseases, Centers for Disease Control and Prevention |

| EPI_Accession_ID | Originating Laboratory | Submitting Laboratory |
|---|---|---|
| EPI_ISL_418783 | WA State Department of Health | Pathogen Discovery, Respiratory Viruses Branch, Division of Viral Diseases, Centers for Disease Control and Prevention |
| EPI_ISL_418784 | WA State Department of Health | Pathogen Discovery, Respiratory Viruses Branch, Division of Viral Diseases, Centers for Disease Control and Prevention |
| EPI_ISL_418785 | WA State Department of Health | Pathogen Discovery, Respiratory Viruses Branch, Division of Viral Diseases, Centers for Disease Control and Prevention |
| EPI_ISL_418786 | WA State Department of Health | Pathogen Discovery, Respiratory Viruses Branch, Division of Viral Diseases, Centers for Disease Control and Prevention |
| EPI_ISL_418787 | WA State Department of Health | Pathogen Discovery, Respiratory Viruses Branch, Division of Viral Diseases, Centers for Disease Control and Prevention |
| EPI_ISL_418788 | WA State Department of Health | Pathogen Discovery, Respiratory Viruses Branch, Division of Viral Diseases, Centers for Disease Control and Prevention |
| EPI_ISL_418789 | WA State Department of Health | Pathogen Discovery, Respiratory Viruses Branch, Division of Viral Diseases, Centers for Disease Control and Prevention |
| EPI_ISL_418790 | WA State Department of Health | Pathogen Discovery, Respiratory Viruses Branch, Division of Viral Diseases, Centers for Disease Control and Prevention |
| EPI_ISL_418791 | WA State Department of Health | Pathogen Discovery, Respiratory Viruses Branch, Division of Viral Diseases, Centers for Disease Control and Prevention |
| EPI_ISL_418792 | KU Leuven, Clinical and Epidemiological Virology | KU Leuven, Clinical and Epidemiological Virology |
| EPI_ISL_418793 | KU Leuven, Clinical and Epidemiological Virology | KU Leuven, Clinical and Epidemiological Virology |
| EPI_ISL_418794 | KU Leuven, Clinical and Epidemiological Virology | KU Leuven, Clinical and Epidemiological Virology |
| EPI_ISL_418795 | KU Leuven, Clinical and Epidemiological Virology | KU Leuven, Clinical and Epidemiological Virology |
| EPI_ISL_418797 | KU Leuven, Clinical and Epidemiological Virology | KU Leuven, Clinical and Epidemiological Virology |
| EPI_ISL_418798 | KU Leuven, Clinical and Epidemiological Virology | KU Leuven, Clinical and Epidemiological Virology |

| EPI_Accession_ID | Originating Laboratory | Submitting Laboratory |
|---|---|---|
| EPI_ISL_418799 | Mater Pathology | Public Health Virology Laboratory |
| EPI_ISL_418800 | KU Leuven, Clinical and Epidemiological Virology | KU Leuven, Clinical and Epidemiological Virology |
| EPI_ISL_418801 | Mater Pathology | Public Health Virology Laboratory |
| EPI_ISL_418802 | Pathology Queensland | Public Health Virology Laboratory |
| EPI_ISL_418803 | Pathology Queensland | Public Health Virology Laboratory |
| EPI_ISL_418804 | Pathology Queensland | Public Health Virology Laboratory |
| EPI_ISL_418805 | KU Leuven, Clinical and Epidemiological Virology | KU Leuven, Clinical and Epidemiological Virology |
| EPI_ISL_418806 | KU Leuven, Clinical and Epidemiological Virology | KU Leuven, Clinical and Epidemiological Virology |
| EPI_ISL_418807 | Pathology Queensland | Public Health Virology Laboratory |
| EPI_ISL_418808 | Pathology Queensland | Public Health Virology Laboratory |
| EPI_ISL_418809 | University of Wisconsin - Madison: Influenza Research Institute | University of Wisconsin Madison, AIDS Vaccine Research Laboratories |
| EPI_ISL_418811 | Dr. Georges-L.-Dumont University Hospital Centre | National Microbiology Laboratory |
| EPI_ISL_418812 | Cadham Provincial Laboratory | National Microbiology Laboratory |
| EPI_ISL_418813 | Cadham Provincial Laboratory | National Microbiology Laboratory |
| EPI_ISL_418814 | Queen Elizabeth II Health Science Centre | National Microbiology Laboratory |
| EPI_ISL_418815 | Department of Clinical Pathology, Pamela Youde Nethersole Eastern Hospital | Department of Health Technology and Informatics, Faculty of Health and Social Science, The Hong Kong Polytechnic University |
| EPI_ISL_418816 | BCCDC Public Health Laboratory | BCCDC Public Health Laboratory |
| EPI_ISL_418817 | BCCDC Public Health Laboratory | BCCDC Public Health Laboratory |
| EPI_ISL_418818 | BCCDC Public Health Laboratory | BCCDC Public Health Laboratory |
| EPI_ISL_418819 | BCCDC Public Health Laboratory | BCCDC Public Health Laboratory |
| EPI_ISL_418820 | BCCDC Public Health Laboratory | BCCDC Public Health Laboratory |
| EPI_ISL_418821 | BCCDC Public Health Laboratory | BCCDC Public Health Laboratory |
| EPI_ISL_418822 | BCCDC Public Health Laboratory | BCCDC Public Health Laboratory |
| EPI_ISL_418823 | BCCDC Public Health Laboratory | BCCDC Public Health Laboratory |
| EPI_ISL_418824 | BCCDC Public Health Laboratory | BCCDC Public Health Laboratory |
| EPI_ISL_418825 | BCCDC Public Health Laboratory | BCCDC Public Health Laboratory |
| EPI_ISL_418826 | BCCDC Public Health Laboratory | BCCDC Public Health Laboratory |
| EPI_ISL_418827 | BCCDC Public Health Laboratory | BCCDC Public Health Laboratory |
| EPI_ISL_418828 | BCCDC Public Health Laboratory | BCCDC Public Health Laboratory |
| EPI_ISL_418829 | BCCDC Public Health Laboratory | BCCDC Public Health Laboratory |

| EPI_Accession_ID | Originating Laboratory | Submitting Laboratory |
|---|---|---|
| EPI_ISL_418830 | BCCDC Public Health Laboratory | BCCDC Public Health Laboratory |
| EPI_ISL_418831 | BCCDC Public Health Laboratory | BCCDC Public Health Laboratory |
| EPI_ISL_418832 | BCCDC Public Health Laboratory | BCCDC Public Health Laboratory |
| EPI_ISL_418833 | BCCDC Public Health Laboratory | BCCDC Public Health Laboratory |
| EPI_ISL_418834 | BCCDC Public Health Laboratory | BCCDC Public Health Laboratory |
| EPI_ISL_418835 | BCCDC Public Health Laboratory | BCCDC Public Health Laboratory |
| EPI_ISL_418836 | BCCDC Public Health Laboratory | BCCDC Public Health Laboratory |
| EPI_ISL_418837 | BCCDC Public Health Laboratory | BCCDC Public Health Laboratory |
| EPI_ISL_418838 | BCCDC Public Health Laboratory | BCCDC Public Health Laboratory |
| EPI_ISL_418839 | BCCDC Public Health Laboratory | BCCDC Public Health Laboratory |
| EPI_ISL_418840 | BCCDC Public Health Laboratory | BCCDC Public Health Laboratory |
| EPI_ISL_418841 | BCCDC Public Health Laboratory | BCCDC Public Health Laboratory |
| EPI_ISL_418842 | BCCDC Public Health Laboratory | BCCDC Public Health Laboratory |
| EPI_ISL_418843 | BCCDC Public Health Laboratory | BCCDC Public Health Laboratory |
| EPI_ISL_418844 | BCCDC Public Health Laboratory | BCCDC Public Health Laboratory |
| EPI_ISL_418845 | BCCDC Public Health Laboratory | BCCDC Public Health Laboratory |
| EPI_ISL_418846 | BCCDC Public Health Laboratory | BCCDC Public Health Laboratory |
| EPI_ISL_418847 | BCCDC Public Health Laboratory | BCCDC Public Health Laboratory |
| EPI_ISL_418848 | BCCDC Public Health Laboratory | BCCDC Public Health Laboratory |
| EPI_ISL_418849 | BCCDC Public Health Laboratory | BCCDC Public Health Laboratory |
| EPI_ISL_418850 | BCCDC Public Health Laboratory | BCCDC Public Health Laboratory |
| EPI_ISL_418851 | BCCDC Public Health Laboratory | BCCDC Public Health Laboratory |
| EPI_ISL_418852 | BCCDC Public Health Laboratory | BCCDC Public Health Laboratory |
| EPI_ISL_418853 | BCCDC Public Health Laboratory | BCCDC Public Health Laboratory |
| EPI_ISL_418854 | BCCDC Public Health Laboratory | BCCDC Public Health Laboratory |
| EPI_ISL_418855 | BCCDC Public Health Laboratory | BCCDC Public Health Laboratory |
| EPI_ISL_418856 | BCCDC Public Health Laboratory | BCCDC Public Health Laboratory |
| EPI_ISL_418857 | BCCDC Public Health Laboratory | BCCDC Public Health Laboratory |
| EPI_ISL_418858 | BCCDC Public Health Laboratory | BCCDC Public Health Laboratory |
| EPI_ISL_418859 | BCCDC Public Health Laboratory | BCCDC Public Health Laboratory |
| EPI_ISL_418860 | Hospital Universitari Vall d'Hebron (HUVH) - Vall d'Hebron Research Institute (VHIR) | Hospital Universitari Vall d'Hebron (HUVH) - Vall d'Hebron Research Institute (VHIR) |

| EPI_Accession_ID | Originating Laboratory | Submitting Laboratory |
|---|---|---|
| EPI_ISL_418861 | Hospital Universitari Vall d'Hebron (HUVH) - Vall d'Hebron Research Institute (VHIR) | Hospital Universitari Vall d'Hebron (HUVH) - Vall d'Hebron Research Institute (VHIR) |
| EPI_ISL_418863 | KU Leuven, Clinical and Epidemiological Virology | KU Leuven, Clinical and Epidemiological Virology |
| EPI_ISL_418864 | Virginia DCLS | Virginia DCLS |
| EPI_ISL_418866 | UW Virology Lab | UW Virology Lab |
| EPI_ISL_418867 | UW Virology Lab | UW Virology Lab |
| EPI_ISL_418868 | UW Virology Lab | UW Virology Lab |
| EPI_ISL_418869 | UW Virology Lab | UW Virology Lab |
| EPI_ISL_418870 | UW Virology Lab | UW Virology Lab |
| EPI_ISL_418871 | UW Virology Lab | UW Virology Lab |
| EPI_ISL_418872 | UW Virology Lab | UW Virology Lab |
| EPI_ISL_418873 | UW Virology Lab | UW Virology Lab |
| EPI_ISL_418875 | UW Virology Lab | UW Virology Lab |
| EPI_ISL_418876 | UW Virology Lab | UW Virology Lab |
| EPI_ISL_418877 | UW Virology Lab | UW Virology Lab |
| EPI_ISL_418878 | UW Virology Lab | UW Virology Lab |
| EPI_ISL_418879 | UW Virology Lab | UW Virology Lab |
| EPI_ISL_418880 | UW Virology Lab | UW Virology Lab |
| EPI_ISL_418881 | UW Virology Lab | UW Virology Lab |
| EPI_ISL_418882 | UW Virology Lab | UW Virology Lab |
| EPI_ISL_418883 | UW Virology Lab | UW Virology Lab |
| EPI_ISL_418884 | UW Virology Lab | UW Virology Lab |
| EPI_ISL_418885 | UW Virology Lab | UW Virology Lab |
| EPI_ISL_418886 | UW Virology Lab | UW Virology Lab |
| EPI_ISL_418887 | UW Virology Lab | UW Virology Lab |
| EPI_ISL_418888 | UW Virology Lab | UW Virology Lab |
| EPI_ISL_418889 | UW Virology Lab | UW Virology Lab |
| EPI_ISL_418890 | UW Virology Lab | UW Virology Lab |
| EPI_ISL_418891 | UW Virology Lab | UW Virology Lab |
| EPI_ISL_418892 | UW Virology Lab | UW Virology Lab |
| EPI_ISL_418893 | UW Virology Lab | UW Virology Lab |
| EPI_ISL_418894 | UW Virology Lab | UW Virology Lab |
| EPI_ISL_418895 | UW Virology Lab | UW Virology Lab |

| EPI_Accession_ID | Originating Laboratory | Submitting Laboratory |
|---|---|---|
| EPI_ISL_418896 | UW Virology Lab | UW Virology Lab |
| EPI_ISL_418897 | UW Virology Lab | UW Virology Lab |
| EPI_ISL_418898 | UW Virology Lab | UW Virology Lab |
| EPI_ISL_418899 | UW Virology Lab | UW Virology Lab |
| EPI_ISL_418900 | UW Virology Lab | UW Virology Lab |
| EPI_ISL_418901 | UW Virology Lab | UW Virology Lab |
| EPI_ISL_418902 | UW Virology Lab | UW Virology Lab |
| EPI_ISL_418903 | UW Virology Lab | UW Virology Lab |
| EPI_ISL_418904 | UW Virology Lab | UW Virology Lab |
| EPI_ISL_418905 | UW Virology Lab | UW Virology Lab |
| EPI_ISL_418906 | UW Virology Lab | UW Virology Lab |
| EPI_ISL_418907 | UW Virology Lab | UW Virology Lab |
| EPI_ISL_418908 | UW Virology Lab | UW Virology Lab |
| EPI_ISL_418909 | UW Virology Lab | UW Virology Lab |
| EPI_ISL_418910 | UW Virology Lab | UW Virology Lab |
| EPI_ISL_418911 | UW Virology Lab | UW Virology Lab |
| EPI_ISL_418912 | UW Virology Lab | UW Virology Lab |
| EPI_ISL_418913 | UW Virology Lab | UW Virology Lab |
| EPI_ISL_418914 | UW Virology Lab | UW Virology Lab |
| EPI_ISL_418915 | UW Virology Lab | UW Virology Lab |
| EPI_ISL_418916 | UW Virology Lab | UW Virology Lab |
| EPI_ISL_418917 | UW Virology Lab | UW Virology Lab |
| EPI_ISL_418918 | UW Virology Lab | UW Virology Lab |
| EPI_ISL_418919 | UW Virology Lab | UW Virology Lab |
| EPI_ISL_418920 | UW Virology Lab | UW Virology Lab |
| EPI_ISL_418921 | UW Virology Lab | UW Virology Lab |
| EPI_ISL_418922 | UW Virology Lab | UW Virology Lab |
| EPI_ISL_418923 | UW Virology Lab | UW Virology Lab |
| EPI_ISL_418924 | UW Virology Lab | UW Virology Lab |
| EPI_ISL_418926 | UW Virology Lab | UW Virology Lab |
| EPI_ISL_418927 | UW Virology Lab | UW Virology Lab |
| EPI_ISL_418928 | UW Virology Lab | UW Virology Lab |
| EPI_ISL_418929 | UW Virology Lab | UW Virology Lab |

| EPI_Accession_ID | Originating Laboratory | Submitting Laboratory |
|---|---|---|
| EPI_ISL_418930 | UW Virology Lab | UW Virology Lab |
| EPI_ISL_418931 | UW Virology Lab | UW Virology Lab |
| EPI_ISL_418932 | UW Virology Lab | UW Virology Lab |
| EPI_ISL_418933 | UW Virology Lab | UW Virology Lab |
| EPI_ISL_418934 | UW Virology Lab | UW Virology Lab |
| EPI_ISL_418935 | UW Virology Lab | UW Virology Lab |
| EPI_ISL_418936 | UW Virology Lab | UW Virology Lab |
| EPI_ISL_418937 | UW Virology Lab | UW Virology Lab |
| EPI_ISL_418938 | UW Virology Lab | UW Virology Lab |
| EPI_ISL_418939 | UW Virology Lab | UW Virology Lab |
| EPI_ISL_418940 | UW Virology Lab | UW Virology Lab |
| EPI_ISL_418942 | UW Virology Lab | UW Virology Lab |
| EPI_ISL_418943 | UW Virology Lab | UW Virology Lab |
| EPI_ISL_418944 | UW Virology Lab | UW Virology Lab |
| EPI_ISL_418945 | UW Virology Lab | UW Virology Lab |
| EPI_ISL_418946 | UW Virology Lab | UW Virology Lab |
| EPI_ISL_418947 | UW Virology Lab | UW Virology Lab |
| EPI_ISL_418948 | UW Virology Lab | UW Virology Lab |
| EPI_ISL_418949 | UW Virology Lab | UW Virology Lab |
| EPI_ISL_418950 | UW Virology Lab | UW Virology Lab |
| EPI_ISL_418951 | UW Virology Lab | UW Virology Lab |
| EPI_ISL_418952 | UW Virology Lab | UW Virology Lab |
| EPI_ISL_418953 | UW Virology Lab | UW Virology Lab |
| EPI_ISL_418954 | UW Virology Lab | UW Virology Lab |
| EPI_ISL_418955 | UW Virology Lab | UW Virology Lab |
| EPI_ISL_418956 | Virginia DCLS | Virginia DCLS |
| EPI_ISL_418957 | Virginia DCLS | Virginia DCLS |
| EPI_ISL_418958 | Virginia DCLS | Virginia DCLS |
| EPI_ISL_418959 | Universidade Federal do Rio de Janeiro - UFRJ | Bioinformatics Laboratory - LNCC |
| EPI_ISL_418962 | Utah Public Health Laboratory | Utah Public Health Laboratory |
| EPI_ISL_418963 | Utah Public Health Laboratory | Utah Public Health Laboratory |
| EPI_ISL_418964 | Utah Public Health Laboratory | Utah Public Health Laboratory |
| EPI_ISL_418965 | Utah Public Health Laboratory | Utah Public Health Laboratory |

| EPI_Accession_ID | Originating Laboratory | Submitting Laboratory |
|---|---|---|
| EPI_ISL_418967 | Utah Public Health Laboratory | Utah Public Health Laboratory |
| EPI_ISL_418968 | NYU Langone Health | Department of Pathology and Medicine, New York University School of Medicine |
| EPI_ISL_418969 | NYU Langone Health | Department of Pathology and Medicine, New York University School of Medicine |
| EPI_ISL_418970 | NYU Langone Health | Department of Pathology and Medicine, New York University School of Medicine |
| EPI_ISL_418971 | NYU Langone Health | Department of Pathology and Medicine, New York University School of Medicine |
| EPI_ISL_418972 | NYU Langone Health | Department of Pathology and Medicine, New York University School of Medicine |
| EPI_ISL_418973 | NYU Langone Health | Department of Pathology and Medicine, New York University School of Medicine |
| EPI_ISL_418974 | NYU Langone Health | Department of Pathology and Medicine, New York University School of Medicine |
| EPI_ISL_418975 | NYU Langone Health | Department of Pathology and Medicine, New York University School of Medicine |
| EPI_ISL_418976 | NYU Langone Health | Department of Pathology and Medicine, New York University School of Medicine |
| EPI_ISL_418977 | NYU Langone Health | Department of Pathology and Medicine, New York University School of Medicine |
| EPI_ISL_418978 | NYU Langone Health | Department of Pathology and Medicine, New York University School of Medicine |
| EPI_ISL_418979 | NYU Langone Health | Department of Pathology and Medicine, New York University School of Medicine |
| EPI_ISL_418980 | NYU Langone Health | Department of Pathology and Medicine, New York University School of Medicine |
| EPI_ISL_418981 | KU Leuven, Clinical and Epidemiological Virology | KU Leuven, Clinical and Epidemiological Virology |
| EPI_ISL_418982 | KU Leuven, Clinical and Epidemiological Virology | KU Leuven, Clinical and Epidemiological Virology |
| EPI_ISL_418983 | KU Leuven, Clinical and Epidemiological Virology | KU Leuven, Clinical and Epidemiological Virology |
| EPI_ISL_418984 | KU Leuven, Clinical and Epidemiological Virology | KU Leuven, Clinical and Epidemiological Virology |
| EPI_ISL_418985 | KU Leuven, Clinical and Epidemiological Virology | KU Leuven, Clinical and Epidemiological Virology |
| EPI_ISL_418986 | KU Leuven, Clinical and Epidemiological Virology | KU Leuven, Clinical and Epidemiological Virology |

| EPI_Accession_ID | Originating Laboratory | Submitting Laboratory |
|---|---|---|
| EPI_ISL_418987 | KU Leuven, Clinical and Epidemiological Virology | KU Leuven, Clinical and Epidemiological Virology |
| EPI_ISL_418988 | Institute information KU Leuven, Clinical and Epidemiological Virology | Institute information KU Leuven, Clinical and Epidemiological Virology |
| EPI_ISL_418989 | KU Leuven, Clinical and Epidemiological Virology | KU Leuven, Clinical and Epidemiological Virology |
| EPI_ISL_418990 | State Key Laboratory for Diagnosis and Treatment of Infectious Diseases, National Clinical Research Center for Infectious Diseases, First Affiliated Hospital, Zhejiang University School of Medicine, Hangzhou, China 310003 | State Key Laboratory for Diagnosis and Treatment of Infectious Diseases, National Clinical Research Center for Infectious Diseases, First Affiliated Hospital, Zhejiang University School of Medicine, Hangzhou, China 310003 |
| EPI_ISL_418991 | State Key Laboratory for Diagnosis and Treatment of Infectious Diseases, National Clinical Research Center for Infectious Diseases, First Affiliated Hospital, Zhejiang University School of Medicine, Hangzhou, China 310003 | State Key Laboratory for Diagnosis and Treatment of Infectious Diseases, National Clinical Research Center for Infectious Diseases, First Affiliated Hospital, Zhejiang University School of Medicine, Hangzhou, China 310003 |
| EPI_ISL_418992 | National Public Health Laboratory, National Centre for Infectious Diseases | National Public Health Laboratory, National Centre for Infectious Diseases |
| EPI_ISL_418993 | National Public Health Laboratory, National Centre for Infectious Diseases | National Public Health Laboratory, National Centre for Infectious Diseases |
| EPI_ISL_418994 | National Public Health Laboratory, National Centre for Infectious Diseases | National Public Health Laboratory, National Centre for Infectious Diseases |
| EPI_ISL_418995 | National Public Health Laboratory, National Centre for Infectious Diseases | National Public Health Laboratory, National Centre for Infectious Diseases |
| EPI_ISL_418996 | National Public Health Laboratory, National Centre for Infectious Diseases | National Public Health Laboratory, National Centre for Infectious Diseases |
| EPI_ISL_418997 | National Public Health Laboratory, National Centre for Infectious Diseases | National Public Health Laboratory, National Centre for Infectious Diseases |
| EPI_ISL_418998 | National Public Health Laboratory, National Centre for Infectious Diseases | National Public Health Laboratory, National Centre for Infectious Diseases |
| EPI_ISL_418999 | National Public Health Laboratory, National Centre for Infectious Diseases | National Public Health Laboratory, National Centre for Infectious Diseases |
| EPI_ISL_419000 | National Public Health Laboratory, National Centre for Infectious Diseases | National Public Health Laboratory, National Centre for Infectious Diseases |

| EPI_Accession_ID | Originating Laboratory | Submitting Laboratory |
|---|---|---|
| EPI_ISL_419001 | National Public Health Laboratory, National Centre for Infectious Diseases | National Public Health Laboratory, National Centre for Infectious Diseases |
| EPI_ISL_419168 | Centre Hospitalier de Valence | CNR Virus des Infections Respiratoires - France SUD |
| EPI_ISL_419169 | Institut des Agents Infectieux (IAI), Hospices Civils de Lyon | CNR Virus des Infections Respiratoires - France SUD |
| EPI_ISL_419170 | Institut des Agents Infectieux (IAI), Hospices Civils de Lyon | CNR Virus des Infections Respiratoires - France SUD |
| EPI_ISL_419171 | Institut des Agents Infectieux (IAI), Hospices Civils de Lyon | CNR Virus des Infections Respiratoires - France SUD |
| EPI_ISL_419172 | Institut des Agents Infectieux (IAI), Hospices Civils de Lyon | CNR Virus des Infections Respiratoires - France SUD |
| EPI_ISL_419173 | Institut des Agents Infectieux (IAI), Hospices Civils de Lyon | CNR Virus des Infections Respiratoires - France SUD |
| EPI_ISL_419174 | Centre Hospitalier de Macon | CNR Virus des Infections Respiratoires - France SUD |
| EPI_ISL_419175 | Centre Hospitalier de Macon | CNR Virus des Infections Respiratoires - France SUD |
| EPI_ISL_419176 | Centre Hospitalier de Macon | CNR Virus des Infections Respiratoires - France SUD |
| EPI_ISL_419177 | Institut des Agents Infectieux (IAI), Hospices Civils de Lyon | CNR Virus des Infections Respiratoires - France SUD |
| EPI_ISL_419178 | Institut des Agents Infectieux (IAI), Hospices Civils de Lyon | CNR Virus des Infections Respiratoires - France SUD |
| EPI_ISL_419179 | Institut des Agents Infectieux (IAI), Hospices Civils de Lyon | CNR Virus des Infections Respiratoires - France SUD |
| EPI_ISL_419180 | Institut des Agents Infectieux (IAI), Hospices Civils de Lyon | CNR Virus des Infections Respiratoires - France SUD |
| EPI_ISL_419181 | Institut des Agents Infectieux (IAI), Hospices Civils de Lyon | CNR Virus des Infections Respiratoires - France SUD |
| EPI_ISL_419182 | Institut des Agents Infectieux (IAI), Hospices Civils de Lyon | CNR Virus des Infections Respiratoires - France SUD |
| EPI_ISL_419183 | Centre Hospitalier de Bourg en Bresse | CNR Virus des Infections Respiratoires - France SUD |
| EPI_ISL_419184 | Institut des Agents Infectieux (IAI), Hospices Civils de Lyon | CNR Virus des Infections Respiratoires - France SUD |
| EPI_ISL_419185 | Centre Hospitalier de Bourg en Bresse | CNR Virus des Infections Respiratoires - France SUD |
| EPI_ISL_419186 | Centre Hospitalier de Bourg en Bresse | CNR Virus des Infections Respiratoires - France SUD |

| EPI_Accession_ID | Originating Laboratory | Submitting Laboratory |
|---|---|---|
| EPI_ISL_419187 | Centre Hospitalier de Macon | CNR Virus des Infections Respiratoires - France SUD |
| EPI_ISL_419188 | Centre Hospitalier de Macon | CNR Virus des Infections Respiratoires - France SUD |
| EPI_ISL_419210 | The Chaim Sheba Medical Center | Israel Institute for Biological Research |
| EPI_ISL_419211 | Central Virology Laboratory | Israel Institute for Biological Research |
| EPI_ISL_419213 | Department of Clinical Pathology, Pamela Youde Nethersole Eastern Hospital | Department of Health Technology and Informatics, Faculty of Health and Social Science, The Hong Kong Polytechnic University |
| EPI_ISL_419214 | Department of Clinical Pathology, Pamela Youde Nethersole Eastern Hospital | Department of Health Technology and Informatics, Faculty of Health and Social Science, The Hong Kong Polytechnic University |
| EPI_ISL_419215 | Department of Clinical Pathology, Pamela Youde Nethersole Eastern Hospital | Department of Health Technology and Informatics, Faculty of Health and Social Science, The Hong Kong Polytechnic University |
| EPI_ISL_419216 | Department of Clinical Pathology, Pamela Youde Nethersole Eastern Hospital | Department of Health Technology and Informatics, Faculty of Health and Social Science, The Hong Kong Polytechnic University |
| EPI_ISL_419217 | Department of Pathology, Princess Margaret Hospital | Department of Health Technology and Informatics, Faculty of Health and Social Science, The Hong Kong Polytechnic University |
| EPI_ISL_419218 | Department of Clinical Pathology, Pamela Youde Nethersole Eastern Hospital | Department of Health Technology and Informatics, Faculty of Health and Social Science, The Hong Kong Polytechnic University |
| EPI_ISL_419219 | Department of Clinical Pathology, Pamela Youde Nethersole Eastern Hospital | Department of Health Technology and Informatics, Faculty of Health and Social Science, The Hong Kong Polytechnic University |
| EPI_ISL_419221 | Department of Pathology, United Christian Hospital | Department of Health Technology and Informatics, Faculty of Health and Social Science, The Hong Kong Polytechnic University |
| EPI_ISL_419222 | Department of Pathology, Princess Margaret Hospital | Department of Health Technology and Informatics, Faculty of Health and Social Science, The Hong Kong Polytechnic University |

| EPI_Accession_ID | Originating Laboratory | Submitting Laboratory |
| --- | --- | --- |
| EPI_ISL_419223 | Department of Pathology, Princess Margaret Hospital | Department of Health Technology and Informatics, Faculty of Health and Social Science, The Hong Kong Polytechnic University |
| EPI_ISL_419224 | Department of Clinical Pathology, Pamela Youde Nethersole Eastern Hospital | Department of Health Technology and Informatics, Faculty of Health and Social Science, The Hong Kong Polytechnic University |
| EPI_ISL_419225 | Department of Clinical Pathology, Pamela Youde Nethersole Eastern Hospital | Department of Health Technology and Informatics, Faculty of Health and Social Science, The Hong Kong Polytechnic University |
| EPI_ISL_419226 | Department of Clinical Pathology, Pamela Youde Nethersole Eastern Hospital | Department of Health Technology and Informatics, Faculty of Health and Social Science, The Hong Kong Polytechnic University |
| EPI_ISL_419227 | Department of Clinical Pathology, Pamela Youde Nethersole Eastern Hospital | Department of Health Technology and Informatics, Faculty of Health and Social Science, The Hong Kong Polytechnic University |
| EPI_ISL_419228 | Department of Clinical Pathology, Pamela Youde Nethersole Eastern Hospital | Department of Health Technology and Informatics, Faculty of Health and Social Science, The Hong Kong Polytechnic University |
| EPI_ISL_419229 | Department of Clinical Pathology, Pamela Youde Nethersole Eastern Hospital | Department of Health Technology and Informatics, Faculty of Health and Social Science, The Hong Kong Polytechnic University |
| EPI_ISL_419230 | Hospital Universitario Virgen de las Nieves | Instituto de Salud Carlos III |
| EPI_ISL_419231 | Department of Clinical Pathology, Tuen Mun Hospital, 23 Tsing Chung Koon Road, Tuen Mun, N.T. | Department of Health Technology and Informatics, Faculty of Health and Social Science, The Hong Kong Polytechnic University |
| EPI_ISL_419232 | Department of Clinical Pathology, Pamela Youde Nethersole Eastern Hospital | Department of Health Technology and Informatics, Faculty of Health and Social Science, The Hong Kong Polytechnic University |
| EPI_ISL_419233 | Hospital Universitario de Canarias | Instituto de Salud Carlos III |
| EPI_ISL_419234 | Hospital San Pedro | Instituto de Salud Carlos III |
| EPI_ISL_419235 | Fundacion Jimenez Diaz | Instituto de Salud Carlos III |
| EPI_ISL_419236 | Fundacion Jimenez Diaz | Instituto de Salud Carlos III |
| EPI_ISL_419237 | Fundacion Jimenez Diaz | Instituto de Salud Carlos III |

| EPI_Accession_ID | Originating Laboratory | Submitting Laboratory |
|---|---|---|
| EPI_ISL_419238 | HOSPITAL DE CRUCES. | Instituto de Salud Carlos III |
| EPI_ISL_419240 | HOSPITAL TXAGORRITXU | Instituto de Salud Carlos III |
| EPI_ISL_419241 | Department of Clinical Pathology, Pamela Youde Nethersole Eastern Hospital | Department of Health Technology and Informatics, Faculty of Health and Social Science, The Hong Kong Polytechnic University |
| EPI_ISL_419242 | Department of Clinical Pathology, Tuen Mun Hospital | Department of Health Technology and Informatics, Faculty of Health and Social Science, The Hong Kong Polytechnic University |
| EPI_ISL_419243 | Department of Clinical Pathology, Pamela Youde Nethersole Eastern Hospital | Department of Health Technology and Informatics, Faculty of Health and Social Science, The Hong Kong Polytechnic University |
| EPI_ISL_419244 | Department of Clinical Pathology, Pamela Youde Nethersole Eastern Hospital | Department of Health Technology and Informatics, Faculty of Health and Social Science, The Hong Kong Polytechnic University |
| EPI_ISL_419245 | Department of Clinical Pathology, Pamela Youde Nethersole Eastern Hospital | Department of Health Technology and Informatics, Faculty of Health and Social Science, The Hong Kong Polytechnic University |
| EPI_ISL_419246 | Department of Clinical Pathology, Pamela Youde Nethersole Eastern Hospital | Department of Health Technology and Informatics, Faculty of Health and Social Science, The Hong Kong Polytechnic University |
| EPI_ISL_419247 | Department of Clinical Pathology, Pamela Youde Nethersole Eastern Hospital | Department of Health Technology and Informatics, Faculty of Health and Social Science, The Hong Kong Polytechnic University |
| EPI_ISL_419248 | Department of Clinical Pathology, Pamela Youde Nethersole Eastern Hospital | Department of Health Technology and Informatics, Faculty of Health and Social Science, The Hong Kong Polytechnic University |
| EPI_ISL_419249 | Department of Clinical Pathology, Pamela Youde Nethersole Eastern Hospital | Department of Health Technology and Informatics, Faculty of Health and Social Science, The Hong Kong Polytechnic University |
| EPI_ISL_419250 | Department of Clinical Pathology, Pamela Youde Nethersole Eastern Hospital | Department of Health Technology and Informatics, Faculty of Health and Social Science, The Hong Kong Polytechnic University |

| EPI_Accession_ID | Originating Laboratory | Submitting Laboratory |
|---|---|---|
| EPI_ISL_419251 | Department of Clinical Pathology, Pamela Youde Nethersole Eastern Hospital | Department of Health Technology and Informatics, Faculty of Health and Social Science, The Hong Kong Polytechnic University |
| EPI_ISL_419252 | Department of Clinical Pathology, Pamela Youde Nethersole Eastern Hospital | Department of Health Technology and Informatics, Faculty of Health and Social Science, The Hong Kong Polytechnic University |
| EPI_ISL_419253 | Department of Pathology, United Christian Hospital | Department of Health Technology and Informatics, Faculty of Health and Social Science, The Hong Kong Polytechnic University |
| EPI_ISL_419254 | INMI Lazzaro Spallanzani IRCCS | Laboratory of Virology, INMI Lazzaro Spallanzani IRCCS |
| EPI_ISL_419255 | INMI Lazzaro Spallanzani IRCCS | INMI Lazzaro Spallanzani IRCCS |
| EPI_ISL_419256 | Virginia DCLS | Virginia DCLS |
| EPI_ISL_419257 | Virginia DCLS | Virginia DCLS |
| EPI_ISL_419258 | Virginia DCLS | Virginia DCLS |
| EPI_ISL_419259 | Lab voor klinische biologie | Onderzoeksgroep Virologie |
| EPI_ISL_419260 | Virginia DCLS | Virginia DCLS |
| EPI_ISL_419261 | Virginia DCLS | Virginia DCLS |
| EPI_ISL_419262 | Virginia DCLS | Virginia DCLS |
| EPI_ISL_419263 | Virginia DCLS | Virginia DCLS |
| EPI_ISL_419264 | Lab voor klinische biologie | Onderzoeksgroep Virologie |
| EPI_ISL_419265 | Lab voor klinische biologie | Onderzoeksgroep Virologie |
| EPI_ISL_419266 | Lab voor klinische biologie | Onderzoeksgroep Virologie |
| EPI_ISL_419296 | Kochi Prefectural Institute of Public Health | Pathogen Genomics Center, National Institute of Infectious Diseases |
| EPI_ISL_419297 | Chiba Prefectural Institute of Public Health | Pathogen Genomics Center, National Institute of Infectious Diseases |
| EPI_ISL_419298 | Chiba Prefectural Institute of Public Health | Pathogen Genomics Center, National Institute of Infectious Diseases |
| EPI_ISL_419299 | Ishikawa Prefectural Institute of Public Health and Environmental Science | Pathogen Genomics Center, National Institute of Infectious Diseases |
| EPI_ISL_419300 | Ishikawa Prefectural Institute of Public Health and Environmental Science | Pathogen Genomics Center, National Institute of Infectious Diseases |

| EPI_Accession_ID | Originating Laboratory | Submitting Laboratory |
|---|---|---|
| EPI_ISL_419301 | Saitama Prefectural Institute of Public Health | Pathogen Genomics Center, National Institute of Infectious Diseases |
| EPI_ISL_419302 | Saitama Prefectural Institute of Public Health | Pathogen Genomics Center, National Institute of Infectious Diseases |
| EPI_ISL_419303 | Saitama Prefectural Institute of Public Health | Pathogen Genomics Center, National Institute of Infectious Diseases |
| EPI_ISL_419304 | Saitama Prefectural Institute of Public Health | Pathogen Genomics Center, National Institute of Infectious Diseases |
| EPI_ISL_419305 | Saitama Prefectural Institute of Public Health | Pathogen Genomics Center, National Institute of Infectious Diseases |
| EPI_ISL_419306 | Saitama Prefectural Institute of Public Health | Pathogen Genomics Center, National Institute of Infectious Diseases |
| EPI_ISL_419307 | Saitama Prefectural Institute of Public Health | Pathogen Genomics Center, National Institute of Infectious Diseases |
| EPI_ISL_419308 | Saitama Prefectural Institute of Public Health | Pathogen Genomics Center, National Institute of Infectious Diseases |
| EPI_ISL_419309 | Chiba Prefectural Institute of Public Health | Pathogen Genomics Center, National Institute of Infectious Diseases |
| EPI_ISL_419310 | Chiba Prefectural Institute of Public Health | Pathogen Genomics Center, National Institute of Infectious Diseases |
| EPI_ISL_419311 | Chiba Prefectural Institute of Public Health | Pathogen Genomics Center, National Institute of Infectious Diseases |
| EPI_ISL_419313 | Molecular Biology and Biotechnology Lab II | Molecular Biology and Biotechnology Lab II |
| EPI_ISL_419386 | Hospital Prof. Doutor Fernando Fonseca, EPE | Instituto Gulbenkian de CiÃªncia |
| EPI_ISL_419387 | Hospital Prof. Doutor Fernando Fonseca, EPE | Instituto Gulbenkian de CiÃªncia |
| EPI_ISL_419388 | Minnesota Department of Health, Public Health Laboratory | Minnesota Department of Health, Public Health Laboratory |
| EPI_ISL_419390 | Minnesota Department of Health, Public Health Laboratory | Minnesota Department of Health, Public Health Laboratory |
| EPI_ISL_419391 | Minnesota Department of Health, Public Health Laboratory | Minnesota Department of Health, Public Health Laboratory |
| EPI_ISL_419392 | Minnesota Department of Health, Public Health Laboratory | Minnesota Department of Health, Public Health Laboratory |

| EPI_Accession_ID | Originating Laboratory | Submitting Laboratory |
|---|---|---|
| EPI_ISL_419393 | Minnesota Department of Health, Public Health Laboratory | Minnesota Department of Health, Public Health Laboratory |
| EPI_ISL_419394 | Minnesota Department of Health, Public Health Laboratory | Minnesota Department of Health, Public Health Laboratory |
| EPI_ISL_419395 | Minnesota Department of Health, Public Health Laboratory | Minnesota Department of Health, Public Health Laboratory |
| EPI_ISL_419396 | Minnesota Department of Health, Public Health Laboratory | Minnesota Department of Health, Public Health Laboratory |
| EPI_ISL_419397 | Minnesota Department of Health, Public Health Laboratory | Minnesota Department of Health, Public Health Laboratory |
| EPI_ISL_419398 | Wales Specialist Virology Centre | Public Health Wales Microbiology Cardiff |
| EPI_ISL_419399 | Wales Specialist Virology Centre | Public Health Wales Microbiology Cardiff |
| EPI_ISL_419400 | Wales Specialist Virology Centre | Public Health Wales Microbiology Cardiff |
| EPI_ISL_419401 | Wales Specialist Virology Centre | Public Health Wales Microbiology Cardiff |
| EPI_ISL_419402 | Wales Specialist Virology Centre | Public Health Wales Microbiology Cardiff |
| EPI_ISL_419403 | Wales Specialist Virology Centre | Public Health Wales Microbiology Cardiff |
| EPI_ISL_419404 | Wales Specialist Virology Centre | Public Health Wales Microbiology Cardiff |
| EPI_ISL_419405 | Wales Specialist Virology Centre | Public Health Wales Microbiology Cardiff |
| EPI_ISL_419406 | Wales Specialist Virology Centre | Public Health Wales Microbiology Cardiff |
| EPI_ISL_419407 | Wales Specialist Virology Centre | Public Health Wales Microbiology Cardiff |
| EPI_ISL_419408 | Wales Specialist Virology Centre | Public Health Wales Microbiology Cardiff |
| EPI_ISL_419409 | Wales Specialist Virology Centre | Public Health Wales Microbiology Cardiff |
| EPI_ISL_419410 | Wales Specialist Virology Centre | Public Health Wales Microbiology Cardiff |
| EPI_ISL_419411 | Wales Specialist Virology Centre | Public Health Wales Microbiology Cardiff |
| EPI_ISL_419412 | Wales Specialist Virology Centre | Public Health Wales Microbiology Cardiff |
| EPI_ISL_419413 | Wales Specialist Virology Centre | Public Health Wales Microbiology Cardiff |
| EPI_ISL_419414 | Wales Specialist Virology Centre | Public Health Wales Microbiology Cardiff |
| EPI_ISL_419415 | Wales Specialist Virology Centre | Public Health Wales Microbiology Cardiff |
| EPI_ISL_419416 | Wales Specialist Virology Centre | Public Health Wales Microbiology Cardiff |
| EPI_ISL_419417 | Wales Specialist Virology Centre | Public Health Wales Microbiology Cardiff |
| EPI_ISL_419418 | Wales Specialist Virology Centre | Public Health Wales Microbiology Cardiff |
| EPI_ISL_419420 | Wales Specialist Virology Centre | Public Health Wales Microbiology Cardiff |
| EPI_ISL_419421 | Wales Specialist Virology Centre | Public Health Wales Microbiology Cardiff |

| EPI_Accession_ID | Originating Laboratory | Submitting Laboratory |
| --- | --- | --- |
| EPI_ISL_419422 | Wales Specialist Virology Centre | Public Health Wales Microbiology Cardiff |
| EPI_ISL_419423 | Wales Specialist Virology Centre | Public Health Wales Microbiology Cardiff |
| EPI_ISL_419424 | Wales Specialist Virology Centre | Public Health Wales Microbiology Cardiff |
| EPI_ISL_419425 | Wales Specialist Virology Centre | Public Health Wales Microbiology Cardiff |
| EPI_ISL_419426 | Wales Specialist Virology Centre | Public Health Wales Microbiology Cardiff |
| EPI_ISL_419427 | Wales Specialist Virology Centre | Public Health Wales Microbiology Cardiff |
| EPI_ISL_419428 | Wales Specialist Virology Centre | Public Health Wales Microbiology Cardiff |
| EPI_ISL_419429 | Wales Specialist Virology Centre | Public Health Wales Microbiology Cardiff |
| EPI_ISL_419430 | Wales Specialist Virology Centre | Public Health Wales Microbiology Cardiff |
| EPI_ISL_419431 | Wales Specialist Virology Centre | Public Health Wales Microbiology Cardiff |
| EPI_ISL_419432 | Wales Specialist Virology Centre | Public Health Wales Microbiology Cardiff |
| EPI_ISL_419433 | Wales Specialist Virology Centre | Public Health Wales Microbiology Cardiff |
| EPI_ISL_419434 | Wales Specialist Virology Centre | Public Health Wales Microbiology Cardiff |
| EPI_ISL_419435 | Wales Specialist Virology Centre | Public Health Wales Microbiology Cardiff |
| EPI_ISL_419436 | Wales Specialist Virology Centre | Public Health Wales Microbiology Cardiff |
| EPI_ISL_419437 | Wales Specialist Virology Centre | Public Health Wales Microbiology Cardiff |
| EPI_ISL_419438 | Wales Specialist Virology Centre | Public Health Wales Microbiology Cardiff |
| EPI_ISL_419439 | Wales Specialist Virology Centre | Public Health Wales Microbiology Cardiff |
| EPI_ISL_419440 | Wales Specialist Virology Centre | Public Health Wales Microbiology Cardiff |
| EPI_ISL_419441 | Wales Specialist Virology Centre | Public Health Wales Microbiology Cardiff |
| EPI_ISL_419442 | Wales Specialist Virology Centre | Public Health Wales Microbiology Cardiff |
| EPI_ISL_419443 | Wales Specialist Virology Centre | Public Health Wales Microbiology Cardiff |
| EPI_ISL_419444 | Wales Specialist Virology Centre | Public Health Wales Microbiology Cardiff |
| EPI_ISL_419445 | Wales Specialist Virology Centre | Public Health Wales Microbiology Cardiff |
| EPI_ISL_419446 | Wales Specialist Virology Centre | Public Health Wales Microbiology Cardiff |
| EPI_ISL_419447 | Wales Specialist Virology Centre | Public Health Wales Microbiology Cardiff |
| EPI_ISL_419448 | Wales Specialist Virology Centre | Public Health Wales Microbiology Cardiff |
| EPI_ISL_419449 | Wales Specialist Virology Centre | Public Health Wales Microbiology Cardiff |
| EPI_ISL_419450 | Wales Specialist Virology Centre | Public Health Wales Microbiology Cardiff |
| EPI_ISL_419452 | Wales Specialist Virology Centre | Public Health Wales Microbiology Cardiff |
| EPI_ISL_419453 | Wales Specialist Virology Centre | Public Health Wales Microbiology Cardiff |
| EPI_ISL_419454 | Wales Specialist Virology Centre | Public Health Wales Microbiology Cardiff |
| EPI_ISL_419455 | Wales Specialist Virology Centre | Public Health Wales Microbiology Cardiff |

| EPI_Accession_ID | Originating Laboratory | Submitting Laboratory |
|---|---|---|
| EPI_ISL_419457 | Wales Specialist Virology Centre | Public Health Wales Microbiology Cardiff |
| EPI_ISL_419458 | Wales Specialist Virology Centre | Public Health Wales Microbiology Cardiff |
| EPI_ISL_419459 | Wales Specialist Virology Centre | Public Health Wales Microbiology Cardiff |
| EPI_ISL_419460 | Wales Specialist Virology Centre | Public Health Wales Microbiology Cardiff |
| EPI_ISL_419461 | Wales Specialist Virology Centre | Public Health Wales Microbiology Cardiff |
| EPI_ISL_419462 | Wales Specialist Virology Centre | Public Health Wales Microbiology Cardiff |
| EPI_ISL_419463 | Wales Specialist Virology Centre | Public Health Wales Microbiology Cardiff |
| EPI_ISL_419464 | Wales Specialist Virology Centre | Public Health Wales Microbiology Cardiff |
| EPI_ISL_419465 | Wales Specialist Virology Centre | Public Health Wales Microbiology Cardiff |
| EPI_ISL_419466 | Wales Specialist Virology Centre | Public Health Wales Microbiology Cardiff |
| EPI_ISL_419467 | Wales Specialist Virology Centre | Public Health Wales Microbiology Cardiff |
| EPI_ISL_419468 | Wales Specialist Virology Centre | Public Health Wales Microbiology Cardiff |
| EPI_ISL_419469 | Wales Specialist Virology Centre | Public Health Wales Microbiology Cardiff |
| EPI_ISL_419470 | Wales Specialist Virology Centre | Public Health Wales Microbiology Cardiff |
| EPI_ISL_419471 | Wales Specialist Virology Centre | Public Health Wales Microbiology Cardiff |
| EPI_ISL_419472 | Wales Specialist Virology Centre | Public Health Wales Microbiology Cardiff |
| EPI_ISL_419473 | Wales Specialist Virology Centre | Public Health Wales Microbiology Cardiff |
| EPI_ISL_419474 | Wales Specialist Virology Centre | Public Health Wales Microbiology Cardiff |
| EPI_ISL_419475 | Wales Specialist Virology Centre | Public Health Wales Microbiology Cardiff |
| EPI_ISL_419476 | Wales Specialist Virology Centre | Public Health Wales Microbiology Cardiff |
| EPI_ISL_419477 | Wales Specialist Virology Centre | Public Health Wales Microbiology Cardiff |
| EPI_ISL_419478 | Wales Specialist Virology Centre | Public Health Wales Microbiology Cardiff |
| EPI_ISL_419479 | Wales Specialist Virology Centre | Public Health Wales Microbiology Cardiff |
| EPI_ISL_419480 | Wales Specialist Virology Centre | Public Health Wales Microbiology Cardiff |
| EPI_ISL_419481 | Wales Specialist Virology Centre | Public Health Wales Microbiology Cardiff |
| EPI_ISL_419482 | Wales Specialist Virology Centre | Public Health Wales Microbiology Cardiff |
| EPI_ISL_419483 | Wales Specialist Virology Centre | Public Health Wales Microbiology Cardiff |
| EPI_ISL_419484 | Wales Specialist Virology Centre | Public Health Wales Microbiology Cardiff |
| EPI_ISL_419485 | Wales Specialist Virology Centre | Public Health Wales Microbiology Cardiff |
| EPI_ISL_419486 | Wales Specialist Virology Centre | Public Health Wales Microbiology Cardiff |
| EPI_ISL_419487 | Wales Specialist Virology Centre | Public Health Wales Microbiology Cardiff |
| EPI_ISL_419488 | Wales Specialist Virology Centre | Public Health Wales Microbiology Cardiff |
| EPI_ISL_419489 | Wales Specialist Virology Centre | Public Health Wales Microbiology Cardiff |

| EPI_Accession_ID | Originating Laboratory | Submitting Laboratory |
|---|---|---|
| EPI_ISL_419490 | Wales Specialist Virology Centre | Public Health Wales Microbiology Cardiff |
| EPI_ISL_419491 | Wales Specialist Virology Centre | Public Health Wales Microbiology Cardiff |
| EPI_ISL_419492 | Wales Specialist Virology Centre | Public Health Wales Microbiology Cardiff |
| EPI_ISL_419493 | Wales Specialist Virology Centre | Public Health Wales Microbiology Cardiff |
| EPI_ISL_419494 | Wales Specialist Virology Centre | Public Health Wales Microbiology Cardiff |
| EPI_ISL_419495 | Wales Specialist Virology Centre | Public Health Wales Microbiology Cardiff |
| EPI_ISL_419496 | Wales Specialist Virology Centre | Public Health Wales Microbiology Cardiff |
| EPI_ISL_419497 | Wales Specialist Virology Centre | Public Health Wales Microbiology Cardiff |
| EPI_ISL_419498 | Wales Specialist Virology Centre | Public Health Wales Microbiology Cardiff |
| EPI_ISL_419499 | Wales Specialist Virology Centre | Public Health Wales Microbiology Cardiff |
| EPI_ISL_419500 | Wales Specialist Virology Centre | Public Health Wales Microbiology Cardiff |
| EPI_ISL_419502 | Wales Specialist Virology Centre | Public Health Wales Microbiology Cardiff |
| EPI_ISL_419503 | Wales Specialist Virology Centre | Public Health Wales Microbiology Cardiff |
| EPI_ISL_419504 | Wales Specialist Virology Centre | Public Health Wales Microbiology Cardiff |
| EPI_ISL_419505 | Wales Specialist Virology Centre | Public Health Wales Microbiology Cardiff |
| EPI_ISL_419506 | Wales Specialist Virology Centre | Public Health Wales Microbiology Cardiff |
| EPI_ISL_419507 | Wales Specialist Virology Centre | Public Health Wales Microbiology Cardiff |
| EPI_ISL_419508 | Wales Specialist Virology Centre | Public Health Wales Microbiology Cardiff |
| EPI_ISL_419509 | Wales Specialist Virology Centre | Public Health Wales Microbiology Cardiff |
| EPI_ISL_419510 | Wales Specialist Virology Centre | Public Health Wales Microbiology Cardiff |
| EPI_ISL_419511 | Wales Specialist Virology Centre | Public Health Wales Microbiology Cardiff |
| EPI_ISL_419512 | Yale COVID-19 Biorepository | Grubaugh Lab - Yale School of Public Health |
| EPI_ISL_419513 | Yale COVID-19 Biorepository | Grubaugh Lab - Yale School of Public Health |
| EPI_ISL_419514 | Yale COVID-19 Biorepository | Grubaugh Lab - Yale School of Public Health |
| EPI_ISL_419515 | Yale COVID-19 Biorepository | Grubaugh Lab - Yale School of Public Health |
| EPI_ISL_419516 | Yale COVID-19 Biorepository | Grubaugh Lab - Yale School of Public Health |
| EPI_ISL_419517 | Yale Clinical Virology Laboratory | Grubaugh Lab - Yale School of Public Health |
| EPI_ISL_419518 | Yale Clinical Virology Laboratory | Grubaugh Lab - Yale School of Public Health |
| EPI_ISL_419519 | Yale Clinical Virology Laboratory | Grubaugh Lab - Yale School of Public Health |
| EPI_ISL_419520 | Yale Clinical Virology Laboratory | Grubaugh Lab - Yale School of Public Health |
| EPI_ISL_419521 | Yale Clinical Virology Laboratory | Grubaugh Lab - Yale School of Public Health |
| EPI_ISL_419522 | Yale Clinical Virology Laboratory | Grubaugh Lab - Yale School of Public Health |
| EPI_ISL_419523 | Yale Clinical Virology Laboratory | Grubaugh Lab - Yale School of Public Health |

| EPI_Accession_ID | Originating Laboratory | Submitting Laboratory |
|---|---|---|
| EPI_ISL_419524 | Yale Clinical Virology Laboratory | Grubaugh Lab - Yale School of Public Health |
| EPI_ISL_419525 | Yale Clinical Virology Laboratory | Grubaugh Lab - Yale School of Public Health |
| EPI_ISL_419526 | Yale Clinical Virology Laboratory | Grubaugh Lab - Yale School of Public Health |
| EPI_ISL_419527 | Yale Clinical Virology Laboratory | Grubaugh Lab - Yale School of Public Health |
| EPI_ISL_419528 | Yale Clinical Virology Laboratory | Grubaugh Lab - Yale School of Public Health |
| EPI_ISL_419541 | Center of Medical Microbiology, Virology, and Hospital Hygiene, University of Duesseldorf | Center of Medical Microbiology, Virology, and Hospital Hygiene, University of Duesseldorf |
| EPI_ISL_419542 | Center of Medical Microbiology, Virology, and Hospital Hygiene, University of Duesseldorf | Center of Medical Microbiology, Virology, and Hospital Hygiene, University of Duesseldorf |
| EPI_ISL_419543 | Center of Medical Microbiology, Virology, and Hospital Hygiene, University of Duesseldorf | Center of Medical Microbiology, Virology, and Hospital Hygiene, University of Duesseldorf |
| EPI_ISL_419544 | Center of Medical Microbiology, Virology, and Hospital Hygiene, University of Duesseldorf | Center of Medical Microbiology, Virology, and Hospital Hygiene, University of Duesseldorf |
| EPI_ISL_419545 | Center of Medical Microbiology, Virology, and Hospital Hygiene, University of Duesseldorf | Center of Medical Microbiology, Virology, and Hospital Hygiene, University of Duesseldorf |
| EPI_ISL_419546 | Center of Medical Microbiology, Virology, and Hospital Hygiene, University of Duesseldorf | Center of Medical Microbiology, Virology, and Hospital Hygiene, University of Duesseldorf |
| EPI_ISL_419547 | Center of Medical Microbiology, Virology, and Hospital Hygiene, University of Duesseldorf | Center of Medical Microbiology, Virology, and Hospital Hygiene, University of Duesseldorf |
| EPI_ISL_419548 | Center of Medical Microbiology, Virology, and Hospital Hygiene, University of Duesseldorf | Center of Medical Microbiology, Virology, and Hospital Hygiene, University of Duesseldorf |
| EPI_ISL_419549 | Center of Medical Microbiology, Virology, and Hospital Hygiene, University of Duesseldorf | Center of Medical Microbiology, Virology, and Hospital Hygiene, University of Duesseldorf |
| EPI_ISL_419550 | Center of Medical Microbiology, Virology, and Hospital Hygiene, University of Duesseldorf | Center of Medical Microbiology, Virology, and Hospital Hygiene, University of Duesseldorf |
| EPI_ISL_419551 | Center of Medical Microbiology, Virology, and Hospital Hygiene, University of Duesseldorf | Center of Medical Microbiology, Virology, and Hospital Hygiene, University of Duesseldorf |
| EPI_ISL_419552 | Center of Medical Microbiology, Virology, and Hospital Hygiene, University of Duesseldorf | Center of Medical Microbiology, Virology, and Hospital Hygiene, University of Duesseldorf |
| EPI_ISL_419553 | RI State Health Laboratories | Pathogen Discovery, Respiratory Viruses Branch, Division of Viral Diseases, Centers for Disease Control and Prevention |

| EPI_Accession_ID | Originating Laboratory | Submitting Laboratory |
|---|---|---|
| EPI_ISL_419554 | California Department of Public Health | Pathogen Discovery, Respiratory Viruses Branch, Division of Viral Diseases, Centers for Disease Control and Prevention |
| EPI_ISL_419555 | WA State Department of Health | Pathogen Discovery, Respiratory Viruses Branch, Division of Viral Diseases, Centers for Disease Control and Prevention |
| EPI_ISL_419556 | GA Department of Public Health Laboratory | Pathogen Discovery, Respiratory Viruses Branch, Division of Viral Diseases, Centers for Disease Control and Prevention |
| EPI_ISL_419557 | GA Department of Public Health Laboratory | Pathogen Discovery, Respiratory Viruses Branch, Division of Viral Diseases, Centers for Disease Control and Prevention |
| EPI_ISL_419558 | OR State PHL-Virology/Immunology Section | Pathogen Discovery, Respiratory Viruses Branch, Division of Viral Diseases, Centers for Disease Control and Prevention |
| EPI_ISL_419559 | FL Bureau of Public Health Laboratories-Tampa | Pathogen Discovery, Respiratory Viruses Branch, Division of Viral Diseases, Centers for Disease Control and Prevention |
| EPI_ISL_419560 | FL Bureau of Public Health Laboratories-Tampa | Pathogen Discovery, Respiratory Viruses Branch, Division of Viral Diseases, Centers for Disease Control and Prevention |
| EPI_ISL_419562 | Laboratoire National de SantÃ©, Microbiology, Virology | Laboratoire National de SantÃ©, Microbiology, Epidemiology and Microbial Genomics |
| EPI_ISL_419563 | Laboratoire National de SantÃ©, Microbiology, Virology | Laboratoire National de SantÃ©, Microbiology, Epidemiology and Microbial Genomics |
| EPI_ISL_419564 | Laboratoire National de SantÃ©, Microbiology, Virology | Laboratoire National de SantÃ©, Microbiology, Epidemiology and Microbial Genomics |
| EPI_ISL_419565 | Laboratoire National de SantÃ©, Microbiology, Virology | Laboratoire National de SantÃ©, Microbiology, Epidemiology and Microbial Genomics |
| EPI_ISL_419566 | Laboratoire National de SantÃ©, Microbiology, Virology | Laboratoire National de SantÃ©, Microbiology, Epidemiology and Microbial Genomics |
| EPI_ISL_419567 | Laboratoire National de SantÃ©, Microbiology, Virology | Laboratoire National de SantÃ©, Microbiology, Epidemiology and Microbial Genomics |

| EPI_Accession_ID | Originating Laboratory | Submitting Laboratory |
|---|---|---|
| EPI_ISL_419568 | Laboratoire National de SantÃ©, Microbiology, Virology | Laboratoire National de SantÃ©, Microbiology, Epidemiology and Microbial Genomics |
| EPI_ISL_419569 | Laboratoire National de SantÃ©, Microbiology, Virology | Laboratoire National de SantÃ©, Microbiology, Epidemiology and Microbial Genomics |
| EPI_ISL_419570 | Laboratoire National de SantÃ©, Microbiology, Virology | Laboratoire National de SantÃ©, Microbiology, Epidemiology and Microbial Genomics |
| EPI_ISL_419571 | Laboratoire National de SantÃ©, Microbiology, Virology | Laboratoire National de SantÃ©, Microbiology, Epidemiology and Microbial Genomics |
| EPI_ISL_419572 | Laboratoire National de SantÃ©, Microbiology, Virology | Laboratoire National de SantÃ©, Microbiology, Epidemiology and Microbial Genomics |
| EPI_ISL_419573 | Laboratoire National de SantÃ©, Microbiology, Virology | Laboratoire National de SantÃ©, Microbiology, Epidemiology and Microbial Genomics |
| EPI_ISL_419574 | Laboratoire National de SantÃ©, Microbiology, Virology | Laboratoire National de SantÃ©, Microbiology, Epidemiology and Microbial Genomics |
| EPI_ISL_419575 | Laboratoire National de SantÃ©, Microbiology, Virology | Laboratoire National de SantÃ©, Microbiology, Epidemiology and Microbial Genomics |
| EPI_ISL_419576 | Laboratoire National de SantÃ©, Microbiology, Virology | Laboratoire National de SantÃ©, Microbiology, Epidemiology and Microbial Genomics |
| EPI_ISL_419577 | Laboratoire National de SantÃ©, Microbiology, Virology | Laboratoire National de SantÃ©, Microbiology, Epidemiology and Microbial Genomics |
| EPI_ISL_419578 | Laboratoire National de SantÃ©, Microbiology, Virology | Laboratoire National de SantÃ©, Microbiology, Epidemiology and Microbial Genomics |
| EPI_ISL_419579 | Laboratoire National de SantÃ©, Microbiology, Virology | Laboratoire National de SantÃ©, Microbiology, Epidemiology and Microbial Genomics |
| EPI_ISL_419580 | Laboratoire National de SantÃ©, Microbiology, Virology | Laboratoire National de SantÃ©, Microbiology, Epidemiology and Microbial Genomics |
| EPI_ISL_419581 | Laboratoire National de SantÃ©, Microbiology, Virology | Laboratoire National de SantÃ©, Microbiology, Epidemiology and Microbial Genomics |
| EPI_ISL_419582 | Laboratoire National de SantÃ©, Microbiology, Virology | Laboratoire National de SantÃ©, Microbiology, Epidemiology and Microbial Genomics |
| EPI_ISL_419583 | Laboratoire National de SantÃ©, Microbiology, Virology | Laboratoire National de SantÃ©, Microbiology, Epidemiology and Microbial Genomics |

| EPI_Accession_ID | Originating Laboratory | Submitting Laboratory |
|---|---|---|
| EPI_ISL_419584 | Laboratoire National de SantÃ©, Microbiology, Virology | Laboratoire National de SantÃ©, Microbiology, Epidemiology and Microbial Genomics |
| EPI_ISL_419585 | Laboratoire National de SantÃ©, Microbiology, Virology | Laboratoire National de SantÃ©, Microbiology, Epidemiology and Microbial Genomics |
| EPI_ISL_419586 | Laboratoire National de SantÃ©, Microbiology, Virology | Laboratoire National de SantÃ©, Microbiology, Epidemiology and Microbial Genomics |
| EPI_ISL_419587 | Laboratoire National de SantÃ©, Microbiology, Virology | Laboratoire National de SantÃ©, Microbiology, Epidemiology and Microbial Genomics |
| EPI_ISL_419588 | Laboratoire National de SantÃ©, Microbiology, Virology | Laboratoire National de SantÃ©, Microbiology, Epidemiology and Microbial Genomics |
| EPI_ISL_419589 | Laboratoire National de SantÃ©, Microbiology, Virology | Laboratoire National de SantÃ©, Microbiology, Epidemiology and Microbial Genomics |
| EPI_ISL_419590 | Laboratoire National de SantÃ©, Microbiology, Virology | Laboratoire National de SantÃ©, Microbiology, Epidemiology and Microbial Genomics |
| EPI_ISL_419591 | Laboratoire National de SantÃ©, Microbiology, Virology | Laboratoire National de SantÃ©, Microbiology, Epidemiology and Microbial Genomics |
| EPI_ISL_419592 | Laboratoire National de SantÃ©, Microbiology, Virology | Laboratoire National de SantÃ©, Microbiology, Epidemiology and Microbial Genomics |
| EPI_ISL_419593 | Laboratoire National de SantÃ©, Microbiology, Virology | Laboratoire National de SantÃ©, Microbiology, Epidemiology and Microbial Genomics |
| EPI_ISL_419594 | Laboratoire National de SantÃ©, Microbiology, Virology | Laboratoire National de SantÃ©, Microbiology, Epidemiology and Microbial Genomics |
| EPI_ISL_419595 | Laboratoire National de SantÃ©, Microbiology, Virology | Laboratoire National de SantÃ©, Microbiology, Epidemiology and Microbial Genomics |
| EPI_ISL_419596 | Laboratoire National de SantÃ©, Microbiology, Virology | Laboratoire National de SantÃ©, Microbiology, Epidemiology and Microbial Genomics |
| EPI_ISL_419597 | Laboratoire National de SantÃ©, Microbiology, Virology | Laboratoire National de SantÃ©, Microbiology, Epidemiology and Microbial Genomics |
| EPI_ISL_419598 | Laboratoire National de SantÃ©, Microbiology, Virology | Laboratoire National de SantÃ©, Microbiology, Epidemiology and Microbial Genomics |
| EPI_ISL_419599 | Laboratoire National de SantÃ©, Microbiology, Virology | Laboratoire National de SantÃ©, Microbiology, Epidemiology and Microbial Genomics |

| EPI_Accession_ID | Originating Laboratory | Submitting Laboratory |
| --- | --- | --- |
| EPI_ISL_419600 | Laboratoire National de SantÃ©, Microbiology, Virology | Laboratoire National de SantÃ©, Microbiology, Epidemiology and Microbial Genomics |
| EPI_ISL_419601 | Laboratoire National de SantÃ©, Microbiology, Virology | Laboratoire National de SantÃ©, Microbiology, Epidemiology and Microbial Genomics |
| EPI_ISL_419602 | Laboratoire National de SantÃ©, Microbiology, Virology | Laboratoire National de SantÃ©, Microbiology, Epidemiology and Microbial Genomics |
| EPI_ISL_419603 | Laboratoire National de SantÃ©, Microbiology, Virology | Laboratoire National de SantÃ©, Microbiology, Epidemiology and Microbial Genomics |
| EPI_ISL_419604 | Laboratoire National de SantÃ©, Microbiology, Virology | Laboratoire National de SantÃ©, Microbiology, Epidemiology and Microbial Genomics |
| EPI_ISL_419605 | Laboratoire National de SantÃ©, Microbiology, Virology | Laboratoire National de SantÃ©, Microbiology, Epidemiology and Microbial Genomics |
| EPI_ISL_419606 | Laboratoire National de SantÃ©, Microbiology, Virology | Laboratoire National de SantÃ©, Microbiology, Epidemiology and Microbial Genomics |
| EPI_ISL_419607 | Laboratoire National de SantÃ©, Microbiology, Virology | Laboratoire National de SantÃ©, Microbiology, Epidemiology and Microbial Genomics |
| EPI_ISL_419651 | Gundersen Molecular Diagnostics Laboratory | Kabara Cancer Research Institute |
| EPI_ISL_419652 | Gundersen Molecular Diagnostics Laboratory | Kabara Cancer Research Institute |
| EPI_ISL_419654 | Center for Virology, Medical University of Vienna | Bergthaler laboratory, CeMM Research Center for Molecular Medicine of the Austrian Academy of Sciences |
| EPI_ISL_419655 | Center for Virology, Medical University of Vienna | Bergthaler laboratory, CeMM Research Center for Molecular Medicine of the Austrian Academy of Sciences |
| EPI_ISL_419656 | Center for Virology, Medical University of Vienna | Bergthaler laboratory, CeMM Research Center for Molecular Medicine of the Austrian Academy of Sciences |
| EPI_ISL_419657 | Center for Virology, Medical University of Vienna | Bergthaler laboratory, CeMM Research Center for Molecular Medicine of the Austrian Academy of Sciences |
| EPI_ISL_419658 | Center for Virology, Medical University of Vienna | Bergthaler laboratory, CeMM Research Center for Molecular Medicine of the Austrian Academy of Sciences |

| EPI_Accession_ID | Originating Laboratory | Submitting Laboratory |
|---|---|---|
| EPI_ISL_419659 | Center for Virology, Medical University of Vienna | Bergthaler laboratory, CeMM Research Center for Molecular Medicine of the Austrian Academy of Sciences |
| EPI_ISL_419660 | Center for Virology, Medical University of Vienna | Bergthaler laboratory, CeMM Research Center for Molecular Medicine of the Austrian Academy of Sciences |
| EPI_ISL_419661 | Center for Virology, Medical University of Vienna | Bergthaler laboratory, CeMM Research Center for Molecular Medicine of the Austrian Academy of Sciences |
| EPI_ISL_419662 | Center for Virology, Medical University of Vienna | Bergthaler laboratory, CeMM Research Center for Molecular Medicine of the Austrian Academy of Sciences |
| EPI_ISL_419663 | Center for Virology, Medical University of Vienna | Bergthaler laboratory, CeMM Research Center for Molecular Medicine of the Austrian Academy of Sciences |
| EPI_ISL_419664 | Center for Virology, Medical University of Vienna | Bergthaler laboratory, CeMM Research Center for Molecular Medicine of the Austrian Academy of Sciences |
| EPI_ISL_419665 | Center for Virology, Medical University of Vienna | Bergthaler laboratory, CeMM Research Center for Molecular Medicine of the Austrian Academy of Sciences |
| EPI_ISL_419666 | Center for Virology, Medical University of Vienna | Bergthaler laboratory, CeMM Research Center for Molecular Medicine of the Austrian Academy of Sciences |
| EPI_ISL_419667 | Center for Virology, Medical University of Vienna | Bergthaler laboratory, CeMM Research Center for Molecular Medicine of the Austrian Academy of Sciences |
| EPI_ISL_419668 | Center for Virology, Medical University of Vienna | Bergthaler laboratory, CeMM Research Center for Molecular Medicine of the Austrian Academy of Sciences |
| EPI_ISL_419669 | Center for Virology, Medical University of Vienna | Bergthaler laboratory, CeMM Research Center for Molecular Medicine of the Austrian Academy of Sciences |

| EPI_Accession_ID | Originating Laboratory | Submitting Laboratory |
|---|---|---|
| EPI_ISL_419670 | Center for Virology, Medical University of Vienna | Bergthaler laboratory, CeMM Research Center for Molecular Medicine of the Austrian Academy of Sciences |
| EPI_ISL_419671 | Center for Virology, Medical University of Vienna | Bergthaler laboratory, CeMM Research Center for Molecular Medicine of the Austrian Academy of Sciences |
| EPI_ISL_419672 | Center for Virology, Medical University of Vienna | Bergthaler laboratory, CeMM Research Center for Molecular Medicine of the Austrian Academy of Sciences |
| EPI_ISL_419673 | Center for Virology, Medical University of Vienna | Bergthaler laboratory, CeMM Research Center for Molecular Medicine of the Austrian Academy of Sciences |
| EPI_ISL_419674 | Center for Virology, Medical University of Vienna | Bergthaler laboratory, CeMM Research Center for Molecular Medicine of the Austrian Academy of Sciences |
| EPI_ISL_419675 | Servicio de MicrobiologÃ-a. Consorcio Hospital General Universitario de Valencia | Sequencing and Bioinformatics Service and Molecular Epidemiology Research Group. FISABIO-Public Health |
| EPI_ISL_419676 | Servicio de MicrobiologÃ-a. Consorcio Hospital General Universitario de Valencia | Sequencing and Bioinformatics Service and Molecular Epidemiology Research Group. FISABIO-Public Health |
| EPI_ISL_419677 | Servicio de MicrobiologÃ-a. Consorcio Hospital General Universitario de Valencia | Sequencing and Bioinformatics Service and Molecular Epidemiology Research Group. FISABIO-Public Health |
| EPI_ISL_419678 | Servicio de MicrobiologÃ-a. Consorcio Hospital General Universitario de Valencia | Sequencing and Bioinformatics Service and Molecular Epidemiology Research Group. FISABIO-Public Health |
| EPI_ISL_419679 | Servicio de MicrobiologÃ-a. Consorcio Hospital General Universitario de Valencia | Sequencing and Bioinformatics Service and Molecular Epidemiology Research Group. FISABIO-Public Health |
| EPI_ISL_419680 | Servicio de MicrobiologÃ-a. Consorcio Hospital General Universitario de Valencia | Sequencing and Bioinformatics Service and Molecular Epidemiology Research Group. FISABIO-Public Health |
| EPI_ISL_419681 | Servicio de MicrobiologÃ-a. Consorcio Hospital General Universitario de Valencia | Sequencing and Bioinformatics Service and Molecular Epidemiology Research Group. FISABIO-Public Health |
| EPI_ISL_419682 | Servicio de MicrobiologÃ-a. Consorcio Hospital General Universitario de Valencia | Sequencing and Bioinformatics Service and Molecular Epidemiology Research Group. FISABIO-Public Health |
| EPI_ISL_419683 | Servicio de MicrobiologÃ-a. Consorcio Hospital General Universitario de Valencia | Sequencing and Bioinformatics Service and Molecular Epidemiology Research Group. FISABIO-Public Health |

| EPI_Accession_ID | Originating Laboratory | Submitting Laboratory |
|---|---|---|
| EPI_ISL_419684 | Servicio de MicrobiologÃ-a. Consorcio Hospital General Universitario de Valencia | Sequencing and Bioinformatics Service and Molecular Epidemiology Research Group. FISABIO-Public Health |
| EPI_ISL_419685 | Servicio de MicrobiologÃ-a. Consorcio Hospital General Universitario de Valencia | Sequencing and Bioinformatics Service and Molecular Epidemiology Research Group. FISABIO-Public Health |
| EPI_ISL_419686 | Servicio de MicrobiologÃ-a. Consorcio Hospital General Universitario de Valencia | Sequencing and Bioinformatics Service and Molecular Epidemiology Research Group. FISABIO-Public Health |
| EPI_ISL_419687 | Servicio de MicrobiologÃ-a. Consorcio Hospital General Universitario de Valencia | Sequencing and Bioinformatics Service and Molecular Epidemiology Research Group. FISABIO-Public Health |
| EPI_ISL_419688 | Servicio de MicrobiologÃ-a. Consorcio Hospital General Universitario de Valencia | Sequencing and Bioinformatics Service and Molecular Epidemiology Research Group. FISABIO-Public Health |
| EPI_ISL_419689 | Servicio de MicrobiologÃ-a. Consorcio Hospital General Universitario de Valencia | Sequencing and Bioinformatics Service and Molecular Epidemiology Research Group. FISABIO-Public Health |
| EPI_ISL_419690 | Servicio de MicrobiologÃ-a. Consorcio Hospital General Universitario de Valencia | Sequencing and Bioinformatics Service and Molecular Epidemiology Research Group. FISABIO-Public Health |
| EPI_ISL_419696 | NYU Langone Health | Departments of Pathology and Medicine, New York University School of Medicine |
| EPI_ISL_419697 | NYU Langone Health | Departments of Pathology and Medicine, New York University School of Medicine |
| EPI_ISL_419698 | NYU Langone Health | Departments of Pathology and Medicine, New York University School of Medicine |
| EPI_ISL_419699 | NYU Langone Health | Departments of Pathology and Medicine, New York University School of Medicine |
| EPI_ISL_419700 | NYU Langone Health | Departments of Pathology and Medicine, New York University School of Medicine |
| EPI_ISL_419701 | NYU Langone Health | Departments of Pathology and Medicine, New York University School of Medicine |
| EPI_ISL_419702 | NYU Langone Health | Departments of Pathology and Medicine, New York University School of Medicine |
| EPI_ISL_419703 | NYU Langone Health | Departments of Pathology and Medicine, New York University School of Medicine |
| EPI_ISL_419704 | NYU Langone Health | Departments of Pathology and Medicine, New York University School of Medicine |

| EPI_Accession_ID | Originating Laboratory | Submitting Laboratory |
|---|---|---|
| EPI_ISL_419705 | NYU Langone Health | Departments of Pathology and Medicine, New York University School of Medicine |
| EPI_ISL_419706 | Virginia DCLS | Virginia DCLS |
| EPI_ISL_419708 | Virginia DCLS | Virginia DCLS |
| EPI_ISL_419709 | HOSPITAL TXAGORRITXU | Instituto de Salud Carlos III |
| EPI_ISL_419710 | Virginia DCLS | Virginia DCLS |
| EPI_ISL_419711 | Virginia DCLS | Virginia DCLS |
| EPI_ISL_419712 | Virginia DCLS | Virginia DCLS |
| EPI_ISL_419713 | Virginia DCLS | Virginia DCLS |
| EPI_ISL_419714 | Microbiological Diagnostic Unit Public Health Laboratory | Microbiological Diagnostic Unit Public Health Laboratory |
| EPI_ISL_419715 | Microbiological Diagnostic Unit Public Health Laboratory | Microbiological Diagnostic Unit Public Health Laboratory |
| EPI_ISL_419716 | Microbiological Diagnostic Unit Public Health Laboratory | Microbiological Diagnostic Unit Public Health Laboratory |
| EPI_ISL_419717 | Microbiological Diagnostic Unit Public Health Laboratory | Microbiological Diagnostic Unit Public Health Laboratory |
| EPI_ISL_419718 | Microbiological Diagnostic Unit Public Health Laboratory | Microbiological Diagnostic Unit Public Health Laboratory |
| EPI_ISL_419719 | Microbiological Diagnostic Unit Public Health Laboratory | Microbiological Diagnostic Unit Public Health Laboratory |
| EPI_ISL_419720 | Microbiological Diagnostic Unit Public Health Laboratory | Microbiological Diagnostic Unit Public Health Laboratory |
| EPI_ISL_419721 | Microbiological Diagnostic Unit Public Health Laboratory | Microbiological Diagnostic Unit Public Health Laboratory |
| EPI_ISL_419722 | Microbiological Diagnostic Unit Public Health Laboratory | Microbiological Diagnostic Unit Public Health Laboratory |
| EPI_ISL_419723 | Microbiological Diagnostic Unit Public Health Laboratory | Microbiological Diagnostic Unit Public Health Laboratory |
| EPI_ISL_419724 | Microbiological Diagnostic Unit Public Health Laboratory | Microbiological Diagnostic Unit Public Health Laboratory |
| EPI_ISL_419725 | Microbiological Diagnostic Unit Public Health Laboratory | Microbiological Diagnostic Unit Public Health Laboratory |

| EPI_Accession_ID | Originating Laboratory | Submitting Laboratory |
|---|---|---|
| EPI_ISL_419726 | Microbiological Diagnostic Unit Public Health Laboratory | Microbiological Diagnostic Unit Public Health Laboratory |
| EPI_ISL_419727 | Microbiological Diagnostic Unit Public Health Laboratory | Microbiological Diagnostic Unit Public Health Laboratory |
| EPI_ISL_419728 | Microbiological Diagnostic Unit Public Health Laboratory | Microbiological Diagnostic Unit Public Health Laboratory |
| EPI_ISL_419729 | Microbiological Diagnostic Unit Public Health Laboratory | Microbiological Diagnostic Unit Public Health Laboratory |
| EPI_ISL_419730 | Microbiological Diagnostic Unit Public Health Laboratory | Microbiological Diagnostic Unit Public Health Laboratory |
| EPI_ISL_419731 | Microbiological Diagnostic Unit Public Health Laboratory | Microbiological Diagnostic Unit Public Health Laboratory |
| EPI_ISL_419732 | Microbiological Diagnostic Unit Public Health Laboratory | Microbiological Diagnostic Unit Public Health Laboratory |
| EPI_ISL_419733 | Victorian Infectious Diseases Reference Laboratory (VIDRL) | Victorian Infectious Diseases Reference Laboratory and Microbiological Diagnostic Unit Public Health Laboratory, Doherty Institute |
| EPI_ISL_419734 | Victorian Infectious Diseases Reference Laboratory (VIDRL) | Victorian Infectious Diseases Reference Laboratory and Microbiological Diagnostic Unit Public Health Laboratory, Doherty Institute |
| EPI_ISL_419735 | Victorian Infectious Diseases Reference Laboratory (VIDRL) | Victorian Infectious Diseases Reference Laboratory and Microbiological Diagnostic Unit Public Health Laboratory, Doherty Institute |
| EPI_ISL_419736 | Victorian Infectious Diseases Reference Laboratory (VIDRL) | Victorian Infectious Diseases Reference Laboratory and Microbiological Diagnostic Unit Public Health Laboratory, Doherty Institute |
| EPI_ISL_419737 | Victorian Infectious Diseases Reference Laboratory (VIDRL) | Victorian Infectious Diseases Reference Laboratory and Microbiological Diagnostic Unit Public Health Laboratory, Doherty Institute |
| EPI_ISL_419738 | Victorian Infectious Diseases Reference Laboratory (VIDRL) | Victorian Infectious Diseases Reference Laboratory and Microbiological Diagnostic Unit Public Health Laboratory, Doherty Institute |

| EPI_Accession_ID | Originating Laboratory | Submitting Laboratory |
|---|---|---|
| EPI_ISL_419739 | Victorian Infectious Diseases Reference Laboratory (VIDRL) | Victorian Infectious Diseases Reference Laboratory and Microbiological Diagnostic Unit Public Health Laboratory, Doherty Institute |
| EPI_ISL_419740 | Victorian Infectious Diseases Reference Laboratory (VIDRL) | Victorian Infectious Diseases Reference Laboratory and Microbiological Diagnostic Unit Public Health Laboratory, Doherty Institute |
| EPI_ISL_419741 | Victorian Infectious Diseases Reference Laboratory (VIDRL) | Victorian Infectious Diseases Reference Laboratory and Microbiological Diagnostic Unit Public Health Laboratory, Doherty Institute |
| EPI_ISL_419742 | Victorian Infectious Diseases Reference Laboratory (VIDRL) | Victorian Infectious Diseases Reference Laboratory and Microbiological Diagnostic Unit Public Health Laboratory, Doherty Institute |
| EPI_ISL_419743 | Victorian Infectious Diseases Reference Laboratory (VIDRL) | Victorian Infectious Diseases Reference Laboratory and Microbiological Diagnostic Unit Public Health Laboratory, Doherty Institute |
| EPI_ISL_419744 | Victorian Infectious Diseases Reference Laboratory (VIDRL) | Victorian Infectious Diseases Reference Laboratory and Microbiological Diagnostic Unit Public Health Laboratory, Doherty Institute |
| EPI_ISL_419745 | Victorian Infectious Diseases Reference Laboratory (VIDRL) | Victorian Infectious Diseases Reference Laboratory and Microbiological Diagnostic Unit Public Health Laboratory, Doherty Institute |
| EPI_ISL_419746 | Victorian Infectious Diseases Reference Laboratory (VIDRL) | Victorian Infectious Diseases Reference Laboratory and Microbiological Diagnostic Unit Public Health Laboratory, Doherty Institute |
| EPI_ISL_419747 | Victorian Infectious Diseases Reference Laboratory (VIDRL) | Victorian Infectious Diseases Reference Laboratory and Microbiological Diagnostic Unit Public Health Laboratory, Doherty Institute |
| EPI_ISL_419748 | Victorian Infectious Diseases Reference Laboratory (VIDRL) | Victorian Infectious Diseases Reference Laboratory and Microbiological Diagnostic Unit Public Health Laboratory, Doherty Institute |
| EPI_ISL_419749 | Victorian Infectious Diseases Reference Laboratory (VIDRL) | Victorian Infectious Diseases Reference Laboratory and Microbiological Diagnostic Unit Public Health Laboratory, Doherty Institute |

| EPI_Accession_ID | Originating Laboratory | Submitting Laboratory |
|---|---|---|
| EPI_ISL_419750 | Victorian Infectious Diseases Reference Laboratory (VIDRL) | Victorian Infectious Diseases Reference Laboratory and Microbiological Diagnostic Unit Public Health Laboratory, Doherty Institute |
| EPI_ISL_419751 | Victorian Infectious Diseases Reference Laboratory (VIDRL) | Victorian Infectious Diseases Reference Laboratory and Microbiological Diagnostic Unit Public Health Laboratory, Doherty Institute |
| EPI_ISL_419752 | Victorian Infectious Diseases Reference Laboratory (VIDRL) | Victorian Infectious Diseases Reference Laboratory and Microbiological Diagnostic Unit Public Health Laboratory, Doherty Institute |
| EPI_ISL_419753 | Victorian Infectious Diseases Reference Laboratory (VIDRL) | Victorian Infectious Diseases Reference Laboratory and Microbiological Diagnostic Unit Public Health Laboratory, Doherty Institute |
| EPI_ISL_419754 | Victorian Infectious Diseases Reference Laboratory (VIDRL) | Victorian Infectious Diseases Reference Laboratory and Microbiological Diagnostic Unit Public Health Laboratory, Doherty Institute |
| EPI_ISL_419755 | Victorian Infectious Diseases Reference Laboratory (VIDRL) | Victorian Infectious Diseases Reference Laboratory and Microbiological Diagnostic Unit Public Health Laboratory, Doherty Institute |
| EPI_ISL_419756 | Victorian Infectious Diseases Reference Laboratory (VIDRL) | Victorian Infectious Diseases Reference Laboratory and Microbiological Diagnostic Unit Public Health Laboratory, Doherty Institute |
| EPI_ISL_419757 | Victorian Infectious Diseases Reference Laboratory (VIDRL) | Victorian Infectious Diseases Reference Laboratory and Microbiological Diagnostic Unit Public Health Laboratory, Doherty Institute |
| EPI_ISL_419758 | Victorian Infectious Diseases Reference Laboratory (VIDRL) | Victorian Infectious Diseases Reference Laboratory and Microbiological Diagnostic Unit Public Health Laboratory, Doherty Institute |
| EPI_ISL_419759 | Victorian Infectious Diseases Reference Laboratory (VIDRL) | Victorian Infectious Diseases Reference Laboratory and Microbiological Diagnostic Unit Public Health Laboratory, Doherty Institute |
| EPI_ISL_419760 | Victorian Infectious Diseases Reference Laboratory (VIDRL) | Victorian Infectious Diseases Reference Laboratory and Microbiological Diagnostic Unit Public Health Laboratory, Doherty Institute |

| EPI_Accession_ID | Originating Laboratory | Submitting Laboratory |
|---|---|---|
| EPI_ISL_419761 | Victorian Infectious Diseases Reference Laboratory (VIDRL) | Victorian Infectious Diseases Reference Laboratory and Microbiological Diagnostic Unit Public Health Laboratory, Doherty Institute |
| EPI_ISL_419762 | Victorian Infectious Diseases Reference Laboratory (VIDRL) | Victorian Infectious Diseases Reference Laboratory and Microbiological Diagnostic Unit Public Health Laboratory, Doherty Institute |
| EPI_ISL_419763 | Victorian Infectious Diseases Reference Laboratory (VIDRL) | Victorian Infectious Diseases Reference Laboratory and Microbiological Diagnostic Unit Public Health Laboratory, Doherty Institute |
| EPI_ISL_419764 | Victorian Infectious Diseases Reference Laboratory (VIDRL) | Victorian Infectious Diseases Reference Laboratory and Microbiological Diagnostic Unit Public Health Laboratory, Doherty Institute |
| EPI_ISL_419765 | Victorian Infectious Diseases Reference Laboratory (VIDRL) | Victorian Infectious Diseases Reference Laboratory and Microbiological Diagnostic Unit Public Health Laboratory, Doherty Institute |
| EPI_ISL_419766 | Victorian Infectious Diseases Reference Laboratory (VIDRL) | Victorian Infectious Diseases Reference Laboratory and Microbiological Diagnostic Unit Public Health Laboratory, Doherty Institute |
| EPI_ISL_419767 | Victorian Infectious Diseases Reference Laboratory (VIDRL) | Victorian Infectious Diseases Reference Laboratory and Microbiological Diagnostic Unit Public Health Laboratory, Doherty Institute |
| EPI_ISL_419768 | Victorian Infectious Diseases Reference Laboratory (VIDRL) | Victorian Infectious Diseases Reference Laboratory and Microbiological Diagnostic Unit Public Health Laboratory, Doherty Institute |
| EPI_ISL_419769 | Victorian Infectious Diseases Reference Laboratory (VIDRL) | Victorian Infectious Diseases Reference Laboratory and Microbiological Diagnostic Unit Public Health Laboratory, Doherty Institute |
| EPI_ISL_419770 | Victorian Infectious Diseases Reference Laboratory (VIDRL) | Victorian Infectious Diseases Reference Laboratory and Microbiological Diagnostic Unit Public Health Laboratory, Doherty Institute |
| EPI_ISL_419771 | Victorian Infectious Diseases Reference Laboratory (VIDRL) | Victorian Infectious Diseases Reference Laboratory and Microbiological Diagnostic Unit Public Health Laboratory, Doherty Institute |

| EPI_Accession_ID | Originating Laboratory | Submitting Laboratory |
|---|---|---|
| EPI_ISL_419772 | Victorian Infectious Diseases Reference Laboratory (VIDRL) | Victorian Infectious Diseases Reference Laboratory and Microbiological Diagnostic Unit Public Health Laboratory, Doherty Institute |
| EPI_ISL_419773 | Victorian Infectious Diseases Reference Laboratory (VIDRL) | Victorian Infectious Diseases Reference Laboratory and Microbiological Diagnostic Unit Public Health Laboratory, Doherty Institute |
| EPI_ISL_419774 | Victorian Infectious Diseases Reference Laboratory (VIDRL) | Victorian Infectious Diseases Reference Laboratory and Microbiological Diagnostic Unit Public Health Laboratory, Doherty Institute |
| EPI_ISL_419775 | Victorian Infectious Diseases Reference Laboratory (VIDRL) | Victorian Infectious Diseases Reference Laboratory and Microbiological Diagnostic Unit Public Health Laboratory, Doherty Institute |
| EPI_ISL_419776 | Victorian Infectious Diseases Reference Laboratory (VIDRL) | Victorian Infectious Diseases Reference Laboratory and Microbiological Diagnostic Unit Public Health Laboratory, Doherty Institute |
| EPI_ISL_419777 | Victorian Infectious Diseases Reference Laboratory (VIDRL) | Victorian Infectious Diseases Reference Laboratory and Microbiological Diagnostic Unit Public Health Laboratory, Doherty Institute |
| EPI_ISL_419778 | Victorian Infectious Diseases Reference Laboratory (VIDRL) | Victorian Infectious Diseases Reference Laboratory and Microbiological Diagnostic Unit Public Health Laboratory, Doherty Institute |
| EPI_ISL_419779 | Victorian Infectious Diseases Reference Laboratory (VIDRL) | Victorian Infectious Diseases Reference Laboratory and Microbiological Diagnostic Unit Public Health Laboratory, Doherty Institute |
| EPI_ISL_419780 | Victorian Infectious Diseases Reference Laboratory (VIDRL) | Victorian Infectious Diseases Reference Laboratory and Microbiological Diagnostic Unit Public Health Laboratory, Doherty Institute |
| EPI_ISL_419781 | Victorian Infectious Diseases Reference Laboratory (VIDRL) | Victorian Infectious Diseases Reference Laboratory and Microbiological Diagnostic Unit Public Health Laboratory, Doherty Institute |
| EPI_ISL_419782 | Victorian Infectious Diseases Reference Laboratory (VIDRL) | Victorian Infectious Diseases Reference Laboratory and Microbiological Diagnostic Unit Public Health Laboratory, Doherty Institute |

| EPI_Accession_ID | Originating Laboratory | Submitting Laboratory |
| --- | --- | --- |
| EPI_ISL_419783 | Victorian Infectious Diseases Reference Laboratory (VIDRL) | Victorian Infectious Diseases Reference Laboratory and Microbiological Diagnostic Unit Public Health Laboratory, Doherty Institute |
| EPI_ISL_419784 | Victorian Infectious Diseases Reference Laboratory (VIDRL) | Victorian Infectious Diseases Reference Laboratory and Microbiological Diagnostic Unit Public Health Laboratory, Doherty Institute |
| EPI_ISL_419785 | Victorian Infectious Diseases Reference Laboratory (VIDRL) | Victorian Infectious Diseases Reference Laboratory and Microbiological Diagnostic Unit Public Health Laboratory, Doherty Institute |
| EPI_ISL_419786 | Victorian Infectious Diseases Reference Laboratory (VIDRL) | Victorian Infectious Diseases Reference Laboratory and Microbiological Diagnostic Unit Public Health Laboratory, Doherty Institute |
| EPI_ISL_419787 | Victorian Infectious Diseases Reference Laboratory (VIDRL) | Victorian Infectious Diseases Reference Laboratory and Microbiological Diagnostic Unit Public Health Laboratory, Doherty Institute |
| EPI_ISL_419788 | Victorian Infectious Diseases Reference Laboratory (VIDRL) | Victorian Infectious Diseases Reference Laboratory and Microbiological Diagnostic Unit Public Health Laboratory, Doherty Institute |
| EPI_ISL_419789 | Victorian Infectious Diseases Reference Laboratory (VIDRL) | Victorian Infectious Diseases Reference Laboratory and Microbiological Diagnostic Unit Public Health Laboratory, Doherty Institute |
| EPI_ISL_419790 | Victorian Infectious Diseases Reference Laboratory (VIDRL) | Victorian Infectious Diseases Reference Laboratory and Microbiological Diagnostic Unit Public Health Laboratory, Doherty Institute |
| EPI_ISL_419791 | Victorian Infectious Diseases Reference Laboratory (VIDRL) | Victorian Infectious Diseases Reference Laboratory and Microbiological Diagnostic Unit Public Health Laboratory, Doherty Institute |
| EPI_ISL_419792 | Victorian Infectious Diseases Reference Laboratory (VIDRL) | Victorian Infectious Diseases Reference Laboratory and Microbiological Diagnostic Unit Public Health Laboratory, Doherty Institute |
| EPI_ISL_419793 | Victorian Infectious Diseases Reference Laboratory (VIDRL) | Victorian Infectious Diseases Reference Laboratory and Microbiological Diagnostic Unit Public Health Laboratory, Doherty Institute |

| EPI_Accession_ID | Originating Laboratory | Submitting Laboratory |
|---|---|---|
| EPI_ISL_419794 | Victorian Infectious Diseases Reference Laboratory (VIDRL) | Victorian Infectious Diseases Reference Laboratory and Microbiological Diagnostic Unit Public Health Laboratory, Doherty Institute |
| EPI_ISL_419795 | Victorian Infectious Diseases Reference Laboratory (VIDRL) | Victorian Infectious Diseases Reference Laboratory and Microbiological Diagnostic Unit Public Health Laboratory, Doherty Institute |
| EPI_ISL_419796 | Victorian Infectious Diseases Reference Laboratory (VIDRL) | Victorian Infectious Diseases Reference Laboratory and Microbiological Diagnostic Unit Public Health Laboratory, Doherty Institute |
| EPI_ISL_419797 | Victorian Infectious Diseases Reference Laboratory (VIDRL) | Victorian Infectious Diseases Reference Laboratory and Microbiological Diagnostic Unit Public Health Laboratory, Doherty Institute |
| EPI_ISL_419798 | Victorian Infectious Diseases Reference Laboratory (VIDRL) | Victorian Infectious Diseases Reference Laboratory and Microbiological Diagnostic Unit Public Health Laboratory, Doherty Institute |
| EPI_ISL_419799 | Victorian Infectious Diseases Reference Laboratory (VIDRL) | Victorian Infectious Diseases Reference Laboratory and Microbiological Diagnostic Unit Public Health Laboratory, Doherty Institute |
| EPI_ISL_419800 | Victorian Infectious Diseases Reference Laboratory (VIDRL) | Victorian Infectious Diseases Reference Laboratory and Microbiological Diagnostic Unit Public Health Laboratory, Doherty Institute |
| EPI_ISL_419801 | Victorian Infectious Diseases Reference Laboratory (VIDRL) | Victorian Infectious Diseases Reference Laboratory and Microbiological Diagnostic Unit Public Health Laboratory, Doherty Institute |
| EPI_ISL_419802 | Victorian Infectious Diseases Reference Laboratory (VIDRL) | Victorian Infectious Diseases Reference Laboratory and Microbiological Diagnostic Unit Public Health Laboratory, Doherty Institute |
| EPI_ISL_419803 | Victorian Infectious Diseases Reference Laboratory (VIDRL) | Victorian Infectious Diseases Reference Laboratory and Microbiological Diagnostic Unit Public Health Laboratory, Doherty Institute |
| EPI_ISL_419804 | Victorian Infectious Diseases Reference Laboratory (VIDRL) | Victorian Infectious Diseases Reference Laboratory and Microbiological Diagnostic Unit Public Health Laboratory, Doherty Institute |

| EPI_Accession_ID | Originating Laboratory | Submitting Laboratory |
|---|---|---|
| EPI_ISL_419805 | Victorian Infectious Diseases Reference Laboratory (VIDRL) | Victorian Infectious Diseases Reference Laboratory and Microbiological Diagnostic Unit Public Health Laboratory, Doherty Institute |
| EPI_ISL_419806 | Victorian Infectious Diseases Reference Laboratory (VIDRL) | Victorian Infectious Diseases Reference Laboratory and Microbiological Diagnostic Unit Public Health Laboratory, Doherty Institute |
| EPI_ISL_419807 | Victorian Infectious Diseases Reference Laboratory (VIDRL) | Victorian Infectious Diseases Reference Laboratory and Microbiological Diagnostic Unit Public Health Laboratory, Doherty Institute |
| EPI_ISL_419808 | Victorian Infectious Diseases Reference Laboratory (VIDRL) | Victorian Infectious Diseases Reference Laboratory and Microbiological Diagnostic Unit Public Health Laboratory, Doherty Institute |
| EPI_ISL_419809 | Victorian Infectious Diseases Reference Laboratory (VIDRL) | Victorian Infectious Diseases Reference Laboratory and Microbiological Diagnostic Unit Public Health Laboratory, Doherty Institute |
| EPI_ISL_419810 | Victorian Infectious Diseases Reference Laboratory (VIDRL) | Victorian Infectious Diseases Reference Laboratory and Microbiological Diagnostic Unit Public Health Laboratory, Doherty Institute |
| EPI_ISL_419811 | Victorian Infectious Diseases Reference Laboratory (VIDRL) | Victorian Infectious Diseases Reference Laboratory and Microbiological Diagnostic Unit Public Health Laboratory, Doherty Institute |
| EPI_ISL_419812 | Victorian Infectious Diseases Reference Laboratory (VIDRL) | Victorian Infectious Diseases Reference Laboratory and Microbiological Diagnostic Unit Public Health Laboratory, Doherty Institute |
| EPI_ISL_419813 | Victorian Infectious Diseases Reference Laboratory (VIDRL) | Victorian Infectious Diseases Reference Laboratory and Microbiological Diagnostic Unit Public Health Laboratory, Doherty Institute |
| EPI_ISL_419814 | Victorian Infectious Diseases Reference Laboratory (VIDRL) | Victorian Infectious Diseases Reference Laboratory and Microbiological Diagnostic Unit Public Health Laboratory, Doherty Institute |
| EPI_ISL_419815 | Victorian Infectious Diseases Reference Laboratory (VIDRL) | Victorian Infectious Diseases Reference Laboratory and Microbiological Diagnostic Unit Public Health Laboratory, Doherty Institute |

| EPI_Accession_ID | Originating Laboratory | Submitting Laboratory |
|---|---|---|
| EPI_ISL_419816 | Victorian Infectious Diseases Reference Laboratory (VIDRL) | Victorian Infectious Diseases Reference Laboratory and Microbiological Diagnostic Unit Public Health Laboratory, Doherty Institute |
| EPI_ISL_419817 | Victorian Infectious Diseases Reference Laboratory (VIDRL) | Victorian Infectious Diseases Reference Laboratory and Microbiological Diagnostic Unit Public Health Laboratory, Doherty Institute |
| EPI_ISL_419818 | Victorian Infectious Diseases Reference Laboratory (VIDRL) | Victorian Infectious Diseases Reference Laboratory and Microbiological Diagnostic Unit Public Health Laboratory, Doherty Institute |
| EPI_ISL_419819 | Victorian Infectious Diseases Reference Laboratory (VIDRL) | Victorian Infectious Diseases Reference Laboratory and Microbiological Diagnostic Unit Public Health Laboratory, Doherty Institute |
| EPI_ISL_419820 | Victorian Infectious Diseases Reference Laboratory (VIDRL) | Victorian Infectious Diseases Reference Laboratory and Microbiological Diagnostic Unit Public Health Laboratory, Doherty Institute |
| EPI_ISL_419821 | Victorian Infectious Diseases Reference Laboratory (VIDRL) | Victorian Infectious Diseases Reference Laboratory and Microbiological Diagnostic Unit Public Health Laboratory, Doherty Institute |
| EPI_ISL_419822 | Victorian Infectious Diseases Reference Laboratory (VIDRL) | Victorian Infectious Diseases Reference Laboratory and Microbiological Diagnostic Unit Public Health Laboratory, Doherty Institute |
| EPI_ISL_419823 | Microbiological Diagnostic Unit Public Health Laboratory | Microbiological Diagnostic Unit Public Health Laboratory |
| EPI_ISL_419824 | Microbiological Diagnostic Unit Public Health Laboratory | Microbiological Diagnostic Unit Public Health Laboratory |
| EPI_ISL_419825 | Microbiological Diagnostic Unit Public Health Laboratory | Microbiological Diagnostic Unit Public Health Laboratory |
| EPI_ISL_419826 | Victorian Infectious Diseases Reference Laboratory (VIDRL) | Victorian Infectious Diseases Reference Laboratory and Microbiological Diagnostic Unit Public Health Laboratory, Doherty Institute |
| EPI_ISL_419827 | Microbiological Diagnostic Unit Public Health Laboratory | Microbiological Diagnostic Unit Public Health Laboratory |

| EPI_Accession_ID | Originating Laboratory | Submitting Laboratory |
|---|---|---|
| EPI_ISL_419828 | Microbiological Diagnostic Unit Public Health Laboratory | Microbiological Diagnostic Unit Public Health Laboratory |
| EPI_ISL_419829 | Microbiological Diagnostic Unit Public Health Laboratory | Microbiological Diagnostic Unit Public Health Laboratory |
| EPI_ISL_419830 | Microbiological Diagnostic Unit Public Health Laboratory | Microbiological Diagnostic Unit Public Health Laboratory |
| EPI_ISL_419831 | Royal Darwin Hospital | Victorian Infectious Diseases Reference Laboratory and Microbiological Diagnostic Unit Public Health Laboratory, Doherty Institute |
| EPI_ISL_419832 | Royal Darwin Hospital | Victorian Infectious Diseases Reference Laboratory and Microbiological Diagnostic Unit Public Health Laboratory, Doherty Institute |
| EPI_ISL_419833 | Royal Darwin Hospital | Victorian Infectious Diseases Reference Laboratory and Microbiological Diagnostic Unit Public Health Laboratory, Doherty Institute |
| EPI_ISL_419834 | Victorian Infectious Diseases Reference Laboratory (VIDRL) | Victorian Infectious Diseases Reference Laboratory and Microbiological Diagnostic Unit Public Health Laboratory, Doherty Institute |
| EPI_ISL_419835 | Royal Darwin Hospital | Victorian Infectious Diseases Reference Laboratory and Microbiological Diagnostic Unit Public Health Laboratory, Doherty Institute |
| EPI_ISL_419836 | Victorian Infectious Diseases Reference Laboratory (VIDRL) | Victorian Infectious Diseases Reference Laboratory and Microbiological Diagnostic Unit Public Health Laboratory, Doherty Institute |
| EPI_ISL_419837 | Victorian Infectious Diseases Reference Laboratory (VIDRL) | Victorian Infectious Diseases Reference Laboratory and Microbiological Diagnostic Unit Public Health Laboratory, Doherty Institute |
| EPI_ISL_419838 | Victorian Infectious Diseases Reference Laboratory (VIDRL) | Victorian Infectious Diseases Reference Laboratory and Microbiological Diagnostic Unit Public Health Laboratory, Doherty Institute |
| EPI_ISL_419839 | Victorian Infectious Diseases Reference Laboratory (VIDRL) | Victorian Infectious Diseases Reference Laboratory and Microbiological Diagnostic Unit Public Health Laboratory, Doherty Institute |

| EPI_Accession_ID | Originating Laboratory | Submitting Laboratory |
|---|---|---|
| EPI_ISL_419840 | Victorian Infectious Diseases Reference Laboratory (VIDRL) | Victorian Infectious Diseases Reference Laboratory and Microbiological Diagnostic Unit Public Health Laboratory, Doherty Institute |
| EPI_ISL_419841 | Victorian Infectious Diseases Reference Laboratory (VIDRL) | Victorian Infectious Diseases Reference Laboratory and Microbiological Diagnostic Unit Public Health Laboratory, Doherty Institute |
| EPI_ISL_419842 | Victorian Infectious Diseases Reference Laboratory (VIDRL) | Victorian Infectious Diseases Reference Laboratory and Microbiological Diagnostic Unit Public Health Laboratory, Doherty Institute |
| EPI_ISL_419843 | Victorian Infectious Diseases Reference Laboratory (VIDRL) | Victorian Infectious Diseases Reference Laboratory and Microbiological Diagnostic Unit Public Health Laboratory, Doherty Institute |
| EPI_ISL_419844 | Victorian Infectious Diseases Reference Laboratory (VIDRL) | Victorian Infectious Diseases Reference Laboratory and Microbiological Diagnostic Unit Public Health Laboratory, Doherty Institute |
| EPI_ISL_419845 | Victorian Infectious Diseases Reference Laboratory (VIDRL) | Victorian Infectious Diseases Reference Laboratory and Microbiological Diagnostic Unit Public Health Laboratory, Doherty Institute |
| EPI_ISL_419846 | Victorian Infectious Diseases Reference Laboratory (VIDRL) | Victorian Infectious Diseases Reference Laboratory and Microbiological Diagnostic Unit Public Health Laboratory, Doherty Institute |
| EPI_ISL_419847 | Victorian Infectious Diseases Reference Laboratory (VIDRL) | Victorian Infectious Diseases Reference Laboratory and Microbiological Diagnostic Unit Public Health Laboratory, Doherty Institute |
| EPI_ISL_419848 | Victorian Infectious Diseases Reference Laboratory (VIDRL) | Victorian Infectious Diseases Reference Laboratory and Microbiological Diagnostic Unit Public Health Laboratory, Doherty Institute |
| EPI_ISL_419849 | Victorian Infectious Diseases Reference Laboratory (VIDRL) | Victorian Infectious Diseases Reference Laboratory and Microbiological Diagnostic Unit Public Health Laboratory, Doherty Institute |
| EPI_ISL_419850 | Victorian Infectious Diseases Reference Laboratory (VIDRL) | Victorian Infectious Diseases Reference Laboratory and Microbiological Diagnostic Unit Public Health Laboratory, Doherty Institute |

| EPI_Accession_ID | Originating Laboratory | Submitting Laboratory |
|---|---|---|
| EPI_ISL_419851 | Victorian Infectious Diseases Reference Laboratory (VIDRL) | Victorian Infectious Diseases Reference Laboratory and Microbiological Diagnostic Unit Public Health Laboratory, Doherty Institute |
| EPI_ISL_419852 | Victorian Infectious Diseases Reference Laboratory (VIDRL) | Victorian Infectious Diseases Reference Laboratory and Microbiological Diagnostic Unit Public Health Laboratory, Doherty Institute |
| EPI_ISL_419853 | Victorian Infectious Diseases Reference Laboratory (VIDRL) | Victorian Infectious Diseases Reference Laboratory and Microbiological Diagnostic Unit Public Health Laboratory, Doherty Institute |
| EPI_ISL_419854 | Victorian Infectious Diseases Reference Laboratory (VIDRL) | Victorian Infectious Diseases Reference Laboratory and Microbiological Diagnostic Unit Public Health Laboratory, Doherty Institute |
| EPI_ISL_419855 | Victorian Infectious Diseases Reference Laboratory (VIDRL) | Victorian Infectious Diseases Reference Laboratory and Microbiological Diagnostic Unit Public Health Laboratory, Doherty Institute |
| EPI_ISL_419856 | Victorian Infectious Diseases Reference Laboratory (VIDRL) | Victorian Infectious Diseases Reference Laboratory and Microbiological Diagnostic Unit Public Health Laboratory, Doherty Institute |
| EPI_ISL_419857 | Victorian Infectious Diseases Reference Laboratory (VIDRL) | Victorian Infectious Diseases Reference Laboratory and Microbiological Diagnostic Unit Public Health Laboratory, Doherty Institute |
| EPI_ISL_419858 | Victorian Infectious Diseases Reference Laboratory (VIDRL) | Victorian Infectious Diseases Reference Laboratory and Microbiological Diagnostic Unit Public Health Laboratory, Doherty Institute |
| EPI_ISL_419859 | Victorian Infectious Diseases Reference Laboratory (VIDRL) | Victorian Infectious Diseases Reference Laboratory and Microbiological Diagnostic Unit Public Health Laboratory, Doherty Institute |
| EPI_ISL_419860 | Victorian Infectious Diseases Reference Laboratory (VIDRL) | Victorian Infectious Diseases Reference Laboratory and Microbiological Diagnostic Unit Public Health Laboratory, Doherty Institute |
| EPI_ISL_419861 | Victorian Infectious Diseases Reference Laboratory (VIDRL) | Victorian Infectious Diseases Reference Laboratory and Microbiological Diagnostic Unit Public Health Laboratory, Doherty Institute |

| EPI_Accession_ID | Originating Laboratory | Submitting Laboratory |
| --- | --- | --- |
| EPI_ISL_419862 | Victorian Infectious Diseases Reference Laboratory (VIDRL) | Victorian Infectious Diseases Reference Laboratory and Microbiological Diagnostic Unit Public Health Laboratory, Doherty Institute |
| EPI_ISL_419863 | Victorian Infectious Diseases Reference Laboratory (VIDRL) | Victorian Infectious Diseases Reference Laboratory and Microbiological Diagnostic Unit Public Health Laboratory, Doherty Institute |
| EPI_ISL_419864 | Victorian Infectious Diseases Reference Laboratory (VIDRL) | Victorian Infectious Diseases Reference Laboratory and Microbiological Diagnostic Unit Public Health Laboratory, Doherty Institute |
| EPI_ISL_419865 | Victorian Infectious Diseases Reference Laboratory (VIDRL) | Victorian Infectious Diseases Reference Laboratory and Microbiological Diagnostic Unit Public Health Laboratory, Doherty Institute |
| EPI_ISL_419866 | Victorian Infectious Diseases Reference Laboratory (VIDRL) | Victorian Infectious Diseases Reference Laboratory and Microbiological Diagnostic Unit Public Health Laboratory, Doherty Institute |
| EPI_ISL_419867 | Victorian Infectious Diseases Reference Laboratory (VIDRL) | Victorian Infectious Diseases Reference Laboratory and Microbiological Diagnostic Unit Public Health Laboratory, Doherty Institute |
| EPI_ISL_419868 | Victorian Infectious Diseases Reference Laboratory (VIDRL) | Victorian Infectious Diseases Reference Laboratory and Microbiological Diagnostic Unit Public Health Laboratory, Doherty Institute |
| EPI_ISL_419869 | Victorian Infectious Diseases Reference Laboratory (VIDRL) | Victorian Infectious Diseases Reference Laboratory and Microbiological Diagnostic Unit Public Health Laboratory, Doherty Institute |
| EPI_ISL_419870 | Victorian Infectious Diseases Reference Laboratory (VIDRL) | Victorian Infectious Diseases Reference Laboratory and Microbiological Diagnostic Unit Public Health Laboratory, Doherty Institute |
| EPI_ISL_419871 | Victorian Infectious Diseases Reference Laboratory (VIDRL) | Victorian Infectious Diseases Reference Laboratory and Microbiological Diagnostic Unit Public Health Laboratory, Doherty Institute |
| EPI_ISL_419872 | Victorian Infectious Diseases Reference Laboratory (VIDRL) | Victorian Infectious Diseases Reference Laboratory and Microbiological Diagnostic Unit Public Health Laboratory, Doherty Institute |

| EPI_Accession_ID | Originating Laboratory | Submitting Laboratory |
|---|---|---|
| EPI_ISL_419873 | Victorian Infectious Diseases Reference Laboratory (VIDRL) | Victorian Infectious Diseases Reference Laboratory and Microbiological Diagnostic Unit Public Health Laboratory, Doherty Institute |
| EPI_ISL_419874 | Victorian Infectious Diseases Reference Laboratory (VIDRL) | Victorian Infectious Diseases Reference Laboratory and Microbiological Diagnostic Unit Public Health Laboratory, Doherty Institute |
| EPI_ISL_419875 | Victorian Infectious Diseases Reference Laboratory (VIDRL) | Victorian Infectious Diseases Reference Laboratory and Microbiological Diagnostic Unit Public Health Laboratory, Doherty Institute |
| EPI_ISL_419876 | Victorian Infectious Diseases Reference Laboratory (VIDRL) | Victorian Infectious Diseases Reference Laboratory and Microbiological Diagnostic Unit Public Health Laboratory, Doherty Institute |
| EPI_ISL_419877 | Victorian Infectious Diseases Reference Laboratory (VIDRL) | Victorian Infectious Diseases Reference Laboratory and Microbiological Diagnostic Unit Public Health Laboratory, Doherty Institute |
| EPI_ISL_419878 | Victorian Infectious Diseases Reference Laboratory (VIDRL) | Victorian Infectious Diseases Reference Laboratory and Microbiological Diagnostic Unit Public Health Laboratory, Doherty Institute |
| EPI_ISL_419879 | Victorian Infectious Diseases Reference Laboratory (VIDRL) | Victorian Infectious Diseases Reference Laboratory and Microbiological Diagnostic Unit Public Health Laboratory, Doherty Institute |
| EPI_ISL_419880 | Victorian Infectious Diseases Reference Laboratory (VIDRL) | Victorian Infectious Diseases Reference Laboratory and Microbiological Diagnostic Unit Public Health Laboratory, Doherty Institute |
| EPI_ISL_419881 | Victorian Infectious Diseases Reference Laboratory (VIDRL) | Victorian Infectious Diseases Reference Laboratory and Microbiological Diagnostic Unit Public Health Laboratory, Doherty Institute |
| EPI_ISL_419882 | Victorian Infectious Diseases Reference Laboratory (VIDRL) | Victorian Infectious Diseases Reference Laboratory and Microbiological Diagnostic Unit Public Health Laboratory, Doherty Institute |
| EPI_ISL_419883 | Victorian Infectious Diseases Reference Laboratory (VIDRL) | Victorian Infectious Diseases Reference Laboratory and Microbiological Diagnostic Unit Public Health Laboratory, Doherty Institute |

| EPI_Accession_ID | Originating Laboratory | Submitting Laboratory |
|---|---|---|
| EPI_ISL_419884 | Victorian Infectious Diseases Reference Laboratory (VIDRL) | Victorian Infectious Diseases Reference Laboratory and Microbiological Diagnostic Unit Public Health Laboratory, Doherty Institute |
| EPI_ISL_419885 | Victorian Infectious Diseases Reference Laboratory (VIDRL) | Victorian Infectious Diseases Reference Laboratory and Microbiological Diagnostic Unit Public Health Laboratory, Doherty Institute |
| EPI_ISL_419886 | Victorian Infectious Diseases Reference Laboratory (VIDRL) | Victorian Infectious Diseases Reference Laboratory and Microbiological Diagnostic Unit Public Health Laboratory, Doherty Institute |
| EPI_ISL_419887 | Victorian Infectious Diseases Reference Laboratory (VIDRL) | Victorian Infectious Diseases Reference Laboratory and Microbiological Diagnostic Unit Public Health Laboratory, Doherty Institute |
| EPI_ISL_419888 | Victorian Infectious Diseases Reference Laboratory (VIDRL) | Victorian Infectious Diseases Reference Laboratory and Microbiological Diagnostic Unit Public Health Laboratory, Doherty Institute |
| EPI_ISL_419889 | Victorian Infectious Diseases Reference Laboratory (VIDRL) | Victorian Infectious Diseases Reference Laboratory and Microbiological Diagnostic Unit Public Health Laboratory, Doherty Institute |
| EPI_ISL_419890 | Victorian Infectious Diseases Reference Laboratory (VIDRL) | Victorian Infectious Diseases Reference Laboratory and Microbiological Diagnostic Unit Public Health Laboratory, Doherty Institute |
| EPI_ISL_419891 | Victorian Infectious Diseases Reference Laboratory (VIDRL) | Victorian Infectious Diseases Reference Laboratory and Microbiological Diagnostic Unit Public Health Laboratory, Doherty Institute |
| EPI_ISL_419892 | Victorian Infectious Diseases Reference Laboratory (VIDRL) | Victorian Infectious Diseases Reference Laboratory and Microbiological Diagnostic Unit Public Health Laboratory, Doherty Institute |
| EPI_ISL_419893 | Victorian Infectious Diseases Reference Laboratory (VIDRL) | Victorian Infectious Diseases Reference Laboratory and Microbiological Diagnostic Unit Public Health Laboratory, Doherty Institute |
| EPI_ISL_419894 | Victorian Infectious Diseases Reference Laboratory (VIDRL) | Victorian Infectious Diseases Reference Laboratory and Microbiological Diagnostic Unit Public Health Laboratory, Doherty Institute |

| EPI_Accession_ID | Originating Laboratory | Submitting Laboratory |
|---|---|---|
| EPI_ISL_419895 | Victorian Infectious Diseases Reference Laboratory (VIDRL) | Victorian Infectious Diseases Reference Laboratory and Microbiological Diagnostic Unit Public Health Laboratory, Doherty Institute |
| EPI_ISL_419896 | Victorian Infectious Diseases Reference Laboratory (VIDRL) | Victorian Infectious Diseases Reference Laboratory and Microbiological Diagnostic Unit Public Health Laboratory, Doherty Institute |
| EPI_ISL_419897 | Victorian Infectious Diseases Reference Laboratory (VIDRL) | Victorian Infectious Diseases Reference Laboratory and Microbiological Diagnostic Unit Public Health Laboratory, Doherty Institute |
| EPI_ISL_419898 | Victorian Infectious Diseases Reference Laboratory (VIDRL) | Victorian Infectious Diseases Reference Laboratory and Microbiological Diagnostic Unit Public Health Laboratory, Doherty Institute |
| EPI_ISL_419899 | Victorian Infectious Diseases Reference Laboratory (VIDRL) | Victorian Infectious Diseases Reference Laboratory and Microbiological Diagnostic Unit Public Health Laboratory, Doherty Institute |
| EPI_ISL_419900 | Victorian Infectious Diseases Reference Laboratory (VIDRL) | Victorian Infectious Diseases Reference Laboratory and Microbiological Diagnostic Unit Public Health Laboratory, Doherty Institute |
| EPI_ISL_419901 | Victorian Infectious Diseases Reference Laboratory (VIDRL) | Victorian Infectious Diseases Reference Laboratory and Microbiological Diagnostic Unit Public Health Laboratory, Doherty Institute |
| EPI_ISL_419902 | Victorian Infectious Diseases Reference Laboratory (VIDRL) | Victorian Infectious Diseases Reference Laboratory and Microbiological Diagnostic Unit Public Health Laboratory, Doherty Institute |
| EPI_ISL_419903 | Victorian Infectious Diseases Reference Laboratory (VIDRL) | Victorian Infectious Diseases Reference Laboratory and Microbiological Diagnostic Unit Public Health Laboratory, Doherty Institute |
| EPI_ISL_419904 | Victorian Infectious Diseases Reference Laboratory (VIDRL) | Victorian Infectious Diseases Reference Laboratory and Microbiological Diagnostic Unit Public Health Laboratory, Doherty Institute |
| EPI_ISL_419905 | Victorian Infectious Diseases Reference Laboratory (VIDRL) | Victorian Infectious Diseases Reference Laboratory and Microbiological Diagnostic Unit Public Health Laboratory, Doherty Institute |

| EPI_Accession_ID | Originating Laboratory | Submitting Laboratory |
|---|---|---|
| EPI_ISL_419906 | Victorian Infectious Diseases Reference Laboratory (VIDRL) | Victorian Infectious Diseases Reference Laboratory and Microbiological Diagnostic Unit Public Health Laboratory, Doherty Institute |
| EPI_ISL_419907 | Victorian Infectious Diseases Reference Laboratory (VIDRL) | Victorian Infectious Diseases Reference Laboratory and Microbiological Diagnostic Unit Public Health Laboratory, Doherty Institute |
| EPI_ISL_419908 | Victorian Infectious Diseases Reference Laboratory (VIDRL) | Victorian Infectious Diseases Reference Laboratory and Microbiological Diagnostic Unit Public Health Laboratory, Doherty Institute |
| EPI_ISL_419909 | Victorian Infectious Diseases Reference Laboratory (VIDRL) | Victorian Infectious Diseases Reference Laboratory and Microbiological Diagnostic Unit Public Health Laboratory, Doherty Institute |
| EPI_ISL_419910 | Victorian Infectious Diseases Reference Laboratory (VIDRL) | Victorian Infectious Diseases Reference Laboratory and Microbiological Diagnostic Unit Public Health Laboratory, Doherty Institute |
| EPI_ISL_419911 | Victorian Infectious Diseases Reference Laboratory (VIDRL) | Victorian Infectious Diseases Reference Laboratory and Microbiological Diagnostic Unit Public Health Laboratory, Doherty Institute |
| EPI_ISL_419912 | Victorian Infectious Diseases Reference Laboratory (VIDRL) | Victorian Infectious Diseases Reference Laboratory and Microbiological Diagnostic Unit Public Health Laboratory, Doherty Institute |
| EPI_ISL_419913 | Victorian Infectious Diseases Reference Laboratory (VIDRL) | Victorian Infectious Diseases Reference Laboratory and Microbiological Diagnostic Unit Public Health Laboratory, Doherty Institute |
| EPI_ISL_419914 | Victorian Infectious Diseases Reference Laboratory (VIDRL) | Victorian Infectious Diseases Reference Laboratory and Microbiological Diagnostic Unit Public Health Laboratory, Doherty Institute |
| EPI_ISL_419915 | Victorian Infectious Diseases Reference Laboratory (VIDRL) | Victorian Infectious Diseases Reference Laboratory and Microbiological Diagnostic Unit Public Health Laboratory, Doherty Institute |
| EPI_ISL_419916 | Victorian Infectious Diseases Reference Laboratory (VIDRL) | Victorian Infectious Diseases Reference Laboratory and Microbiological Diagnostic Unit Public Health Laboratory, Doherty Institute |

| EPI_Accession_ID | Originating Laboratory | Submitting Laboratory |
|---|---|---|
| EPI_ISL_419917 | Victorian Infectious Diseases Reference Laboratory (VIDRL) | Victorian Infectious Diseases Reference Laboratory and Microbiological Diagnostic Unit Public Health Laboratory, Doherty Institute |
| EPI_ISL_419918 | Victorian Infectious Diseases Reference Laboratory (VIDRL) | Victorian Infectious Diseases Reference Laboratory and Microbiological Diagnostic Unit Public Health Laboratory, Doherty Institute |
| EPI_ISL_419919 | Victorian Infectious Diseases Reference Laboratory (VIDRL) | Victorian Infectious Diseases Reference Laboratory and Microbiological Diagnostic Unit Public Health Laboratory, Doherty Institute |
| EPI_ISL_419920 | Victorian Infectious Diseases Reference Laboratory (VIDRL) | Victorian Infectious Diseases Reference Laboratory and Microbiological Diagnostic Unit Public Health Laboratory, Doherty Institute |
| EPI_ISL_419921 | Victorian Infectious Diseases Reference Laboratory (VIDRL) | Victorian Infectious Diseases Reference Laboratory and Microbiological Diagnostic Unit Public Health Laboratory, Doherty Institute |
| EPI_ISL_419922 | Victorian Infectious Diseases Reference Laboratory (VIDRL) | Victorian Infectious Diseases Reference Laboratory and Microbiological Diagnostic Unit Public Health Laboratory, Doherty Institute |
| EPI_ISL_419923 | Victorian Infectious Diseases Reference Laboratory (VIDRL) | Victorian Infectious Diseases Reference Laboratory and Microbiological Diagnostic Unit Public Health Laboratory, Doherty Institute |
| EPI_ISL_419924 | Victorian Infectious Diseases Reference Laboratory (VIDRL) | Victorian Infectious Diseases Reference Laboratory and Microbiological Diagnostic Unit Public Health Laboratory, Doherty Institute |
| EPI_ISL_419925 | Victorian Infectious Diseases Reference Laboratory (VIDRL) | Victorian Infectious Diseases Reference Laboratory and Microbiological Diagnostic Unit Public Health Laboratory, Doherty Institute |
| EPI_ISL_419926 | Victorian Infectious Diseases Reference Laboratory (VIDRL) | Victorian Infectious Diseases Reference Laboratory and Microbiological Diagnostic Unit Public Health Laboratory, Doherty Institute |
| EPI_ISL_419927 | Victorian Infectious Diseases Reference Laboratory (VIDRL) | Victorian Infectious Diseases Reference Laboratory and Microbiological Diagnostic Unit Public Health Laboratory, Doherty Institute |

| EPI_Accession_ID | Originating Laboratory | Submitting Laboratory |
|---|---|---|
| EPI_ISL_419928 | Victorian Infectious Diseases Reference Laboratory (VIDRL) | Victorian Infectious Diseases Reference Laboratory and Microbiological Diagnostic Unit Public Health Laboratory, Doherty Institute |
| EPI_ISL_419929 | Victorian Infectious Diseases Reference Laboratory (VIDRL) | Victorian Infectious Diseases Reference Laboratory and Microbiological Diagnostic Unit Public Health Laboratory, Doherty Institute |
| EPI_ISL_419930 | Victorian Infectious Diseases Reference Laboratory (VIDRL) | Victorian Infectious Diseases Reference Laboratory and Microbiological Diagnostic Unit Public Health Laboratory, Doherty Institute |
| EPI_ISL_419931 | Victorian Infectious Diseases Reference Laboratory (VIDRL) | Victorian Infectious Diseases Reference Laboratory and Microbiological Diagnostic Unit Public Health Laboratory, Doherty Institute |
| EPI_ISL_419932 | Victorian Infectious Diseases Reference Laboratory (VIDRL) | Victorian Infectious Diseases Reference Laboratory and Microbiological Diagnostic Unit Public Health Laboratory, Doherty Institute |
| EPI_ISL_419933 | Victorian Infectious Diseases Reference Laboratory (VIDRL) | Victorian Infectious Diseases Reference Laboratory and Microbiological Diagnostic Unit Public Health Laboratory, Doherty Institute |
| EPI_ISL_419934 | Victorian Infectious Diseases Reference Laboratory (VIDRL) | Victorian Infectious Diseases Reference Laboratory and Microbiological Diagnostic Unit Public Health Laboratory, Doherty Institute |
| EPI_ISL_419935 | Victorian Infectious Diseases Reference Laboratory (VIDRL) | Victorian Infectious Diseases Reference Laboratory and Microbiological Diagnostic Unit Public Health Laboratory, Doherty Institute |
| EPI_ISL_419936 | Victorian Infectious Diseases Reference Laboratory (VIDRL) | Victorian Infectious Diseases Reference Laboratory and Microbiological Diagnostic Unit Public Health Laboratory, Doherty Institute |
| EPI_ISL_419937 | Victorian Infectious Diseases Reference Laboratory (VIDRL) | Victorian Infectious Diseases Reference Laboratory and Microbiological Diagnostic Unit Public Health Laboratory, Doherty Institute |
| EPI_ISL_419938 | Victorian Infectious Diseases Reference Laboratory (VIDRL) | Victorian Infectious Diseases Reference Laboratory and Microbiological Diagnostic Unit Public Health Laboratory, Doherty Institute |

| EPI_Accession_ID | Originating Laboratory | Submitting Laboratory |
|---|---|---|
| EPI_ISL_419939 | Victorian Infectious Diseases Reference Laboratory (VIDRL) | Victorian Infectious Diseases Reference Laboratory and Microbiological Diagnostic Unit Public Health Laboratory, Doherty Institute |
| EPI_ISL_419940 | Victorian Infectious Diseases Reference Laboratory (VIDRL) | Victorian Infectious Diseases Reference Laboratory and Microbiological Diagnostic Unit Public Health Laboratory, Doherty Institute |
| EPI_ISL_419941 | Victorian Infectious Diseases Reference Laboratory (VIDRL) | Victorian Infectious Diseases Reference Laboratory and Microbiological Diagnostic Unit Public Health Laboratory, Doherty Institute |
| EPI_ISL_419942 | Victorian Infectious Diseases Reference Laboratory (VIDRL) | Victorian Infectious Diseases Reference Laboratory and Microbiological Diagnostic Unit Public Health Laboratory, Doherty Institute |
| EPI_ISL_419943 | Victorian Infectious Diseases Reference Laboratory (VIDRL) | Victorian Infectious Diseases Reference Laboratory and Microbiological Diagnostic Unit Public Health Laboratory, Doherty Institute |
| EPI_ISL_419944 | Victorian Infectious Diseases Reference Laboratory (VIDRL) | Victorian Infectious Diseases Reference Laboratory and Microbiological Diagnostic Unit Public Health Laboratory, Doherty Institute |
| EPI_ISL_419945 | Victorian Infectious Diseases Reference Laboratory (VIDRL) | Victorian Infectious Diseases Reference Laboratory and Microbiological Diagnostic Unit Public Health Laboratory, Doherty Institute |
| EPI_ISL_419946 | Victorian Infectious Diseases Reference Laboratory (VIDRL) | Victorian Infectious Diseases Reference Laboratory and Microbiological Diagnostic Unit Public Health Laboratory, Doherty Institute |
| EPI_ISL_419947 | Victorian Infectious Diseases Reference Laboratory (VIDRL) | Victorian Infectious Diseases Reference Laboratory and Microbiological Diagnostic Unit Public Health Laboratory, Doherty Institute |
| EPI_ISL_419948 | Victorian Infectious Diseases Reference Laboratory (VIDRL) | Victorian Infectious Diseases Reference Laboratory and Microbiological Diagnostic Unit Public Health Laboratory, Doherty Institute |
| EPI_ISL_419949 | Victorian Infectious Diseases Reference Laboratory (VIDRL) | Victorian Infectious Diseases Reference Laboratory and Microbiological Diagnostic Unit Public Health Laboratory, Doherty Institute |

| EPI_Accession_ID | Originating Laboratory | Submitting Laboratory |
|---|---|---|
| EPI_ISL_419950 | Victorian Infectious Diseases Reference Laboratory (VIDRL) | Victorian Infectious Diseases Reference Laboratory and Microbiological Diagnostic Unit Public Health Laboratory, Doherty Institute |
| EPI_ISL_419951 | Victorian Infectious Diseases Reference Laboratory (VIDRL) | Victorian Infectious Diseases Reference Laboratory and Microbiological Diagnostic Unit Public Health Laboratory, Doherty Institute |
| EPI_ISL_419952 | Victorian Infectious Diseases Reference Laboratory (VIDRL) | Victorian Infectious Diseases Reference Laboratory and Microbiological Diagnostic Unit Public Health Laboratory, Doherty Institute |
| EPI_ISL_419953 | Victorian Infectious Diseases Reference Laboratory (VIDRL) | Victorian Infectious Diseases Reference Laboratory and Microbiological Diagnostic Unit Public Health Laboratory, Doherty Institute |
| EPI_ISL_419954 | Victorian Infectious Diseases Reference Laboratory (VIDRL) | Victorian Infectious Diseases Reference Laboratory and Microbiological Diagnostic Unit Public Health Laboratory, Doherty Institute |
| EPI_ISL_419955 | Victorian Infectious Diseases Reference Laboratory (VIDRL) | Victorian Infectious Diseases Reference Laboratory and Microbiological Diagnostic Unit Public Health Laboratory, Doherty Institute |
| EPI_ISL_419956 | Victorian Infectious Diseases Reference Laboratory (VIDRL) | Victorian Infectious Diseases Reference Laboratory and Microbiological Diagnostic Unit Public Health Laboratory, Doherty Institute |
| EPI_ISL_419957 | Victorian Infectious Diseases Reference Laboratory (VIDRL) | Victorian Infectious Diseases Reference Laboratory and Microbiological Diagnostic Unit Public Health Laboratory, Doherty Institute |
| EPI_ISL_419958 | Victorian Infectious Diseases Reference Laboratory (VIDRL) | Victorian Infectious Diseases Reference Laboratory and Microbiological Diagnostic Unit Public Health Laboratory, Doherty Institute |
| EPI_ISL_419959 | Victorian Infectious Diseases Reference Laboratory (VIDRL) | Victorian Infectious Diseases Reference Laboratory and Microbiological Diagnostic Unit Public Health Laboratory, Doherty Institute |
| EPI_ISL_419960 | Victorian Infectious Diseases Reference Laboratory (VIDRL) | Victorian Infectious Diseases Reference Laboratory and Microbiological Diagnostic Unit Public Health Laboratory, Doherty Institute |

| EPI_Accession_ID | Originating Laboratory | Submitting Laboratory |
|---|---|---|
| EPI_ISL_419961 | Victorian Infectious Diseases Reference Laboratory (VIDRL) | Victorian Infectious Diseases Reference Laboratory and Microbiological Diagnostic Unit Public Health Laboratory, Doherty Institute |
| EPI_ISL_419962 | Victorian Infectious Diseases Reference Laboratory (VIDRL) | Victorian Infectious Diseases Reference Laboratory and Microbiological Diagnostic Unit Public Health Laboratory, Doherty Institute |
| EPI_ISL_419963 | Victorian Infectious Diseases Reference Laboratory (VIDRL) | Victorian Infectious Diseases Reference Laboratory and Microbiological Diagnostic Unit Public Health Laboratory, Doherty Institute |
| EPI_ISL_419964 | Victorian Infectious Diseases Reference Laboratory (VIDRL) | Victorian Infectious Diseases Reference Laboratory and Microbiological Diagnostic Unit Public Health Laboratory, Doherty Institute |
| EPI_ISL_419965 | Victorian Infectious Diseases Reference Laboratory (VIDRL) | Victorian Infectious Diseases Reference Laboratory and Microbiological Diagnostic Unit Public Health Laboratory, Doherty Institute |
| EPI_ISL_419966 | Victorian Infectious Diseases Reference Laboratory (VIDRL) | Victorian Infectious Diseases Reference Laboratory and Microbiological Diagnostic Unit Public Health Laboratory, Doherty Institute |
| EPI_ISL_419967 | Victorian Infectious Diseases Reference Laboratory (VIDRL) | Victorian Infectious Diseases Reference Laboratory and Microbiological Diagnostic Unit Public Health Laboratory, Doherty Institute |
| EPI_ISL_419968 | Victorian Infectious Diseases Reference Laboratory (VIDRL) | Victorian Infectious Diseases Reference Laboratory and Microbiological Diagnostic Unit Public Health Laboratory, Doherty Institute |
| EPI_ISL_419969 | Victorian Infectious Diseases Reference Laboratory (VIDRL) | Victorian Infectious Diseases Reference Laboratory and Microbiological Diagnostic Unit Public Health Laboratory, Doherty Institute |
| EPI_ISL_419970 | Victorian Infectious Diseases Reference Laboratory (VIDRL) | Victorian Infectious Diseases Reference Laboratory and Microbiological Diagnostic Unit Public Health Laboratory, Doherty Institute |
| EPI_ISL_419971 | Victorian Infectious Diseases Reference Laboratory (VIDRL) | Victorian Infectious Diseases Reference Laboratory and Microbiological Diagnostic Unit Public Health Laboratory, Doherty Institute |

| EPI_Accession_ID | Originating Laboratory | Submitting Laboratory |
| --- | --- | --- |
| EPI_ISL_419972 | Victorian Infectious Diseases Reference Laboratory (VIDRL) | Victorian Infectious Diseases Reference Laboratory and Microbiological Diagnostic Unit Public Health Laboratory, Doherty Institute |
| EPI_ISL_419973 | Victorian Infectious Diseases Reference Laboratory (VIDRL) | Victorian Infectious Diseases Reference Laboratory and Microbiological Diagnostic Unit Public Health Laboratory, Doherty Institute |
| EPI_ISL_419974 | Victorian Infectious Diseases Reference Laboratory (VIDRL) | Victorian Infectious Diseases Reference Laboratory and Microbiological Diagnostic Unit Public Health Laboratory, Doherty Institute |
| EPI_ISL_419975 | Victorian Infectious Diseases Reference Laboratory (VIDRL) | Victorian Infectious Diseases Reference Laboratory and Microbiological Diagnostic Unit Public Health Laboratory, Doherty Institute |
| EPI_ISL_419976 | Victorian Infectious Diseases Reference Laboratory (VIDRL) | Victorian Infectious Diseases Reference Laboratory and Microbiological Diagnostic Unit Public Health Laboratory, Doherty Institute |
| EPI_ISL_419977 | Victorian Infectious Diseases Reference Laboratory (VIDRL) | Victorian Infectious Diseases Reference Laboratory and Microbiological Diagnostic Unit Public Health Laboratory, Doherty Institute |
| EPI_ISL_419978 | Victorian Infectious Diseases Reference Laboratory (VIDRL) | Victorian Infectious Diseases Reference Laboratory and Microbiological Diagnostic Unit Public Health Laboratory, Doherty Institute |
| EPI_ISL_419979 | Victorian Infectious Diseases Reference Laboratory (VIDRL) | Victorian Infectious Diseases Reference Laboratory and Microbiological Diagnostic Unit Public Health Laboratory, Doherty Institute |
| EPI_ISL_419980 | Microbiological Diagnostic Unit Public Health Laboratory | Microbiological Diagnostic Unit Public Health Laboratory |
| EPI_ISL_419981 | Microbiological Diagnostic Unit Public Health Laboratory | Microbiological Diagnostic Unit Public Health Laboratory |
| EPI_ISL_419982 | Victorian Infectious Diseases Reference Laboratory (VIDRL) | Victorian Infectious Diseases Reference Laboratory and Microbiological Diagnostic Unit Public Health Laboratory, Doherty Institute |

| EPI_Accession_ID | Originating Laboratory | Submitting Laboratory |
|---|---|---|
| EPI_ISL_419983 | Victorian Infectious Diseases Reference Laboratory (VIDRL) | Victorian Infectious Diseases Reference Laboratory and Microbiological Diagnostic Unit Public Health Laboratory, Doherty Institute |
| EPI_ISL_419984 | Victorian Infectious Diseases Reference Laboratory (VIDRL) | Victorian Infectious Diseases Reference Laboratory and Microbiological Diagnostic Unit Public Health Laboratory, Doherty Institute |
| EPI_ISL_419985 | Victorian Infectious Diseases Reference Laboratory (VIDRL) | Victorian Infectious Diseases Reference Laboratory and Microbiological Diagnostic Unit Public Health Laboratory, Doherty Institute |
| EPI_ISL_419986 | Victorian Infectious Diseases Reference Laboratory (VIDRL) | Victorian Infectious Diseases Reference Laboratory and Microbiological Diagnostic Unit Public Health Laboratory, Doherty Institute |
| EPI_ISL_419987 | Victorian Infectious Diseases Reference Laboratory (VIDRL) | Victorian Infectious Diseases Reference Laboratory and Microbiological Diagnostic Unit Public Health Laboratory, Doherty Institute |
| EPI_ISL_419988 | Victorian Infectious Diseases Reference Laboratory (VIDRL) | Victorian Infectious Diseases Reference Laboratory and Microbiological Diagnostic Unit Public Health Laboratory, Doherty Institute |
| EPI_ISL_419989 | Victorian Infectious Diseases Reference Laboratory (VIDRL) | Victorian Infectious Diseases Reference Laboratory and Microbiological Diagnostic Unit Public Health Laboratory, Doherty Institute |
| EPI_ISL_419990 | Victorian Infectious Diseases Reference Laboratory (VIDRL) | Victorian Infectious Diseases Reference Laboratory and Microbiological Diagnostic Unit Public Health Laboratory, Doherty Institute |
| EPI_ISL_419991 | Victorian Infectious Diseases Reference Laboratory (VIDRL) | Victorian Infectious Diseases Reference Laboratory and Microbiological Diagnostic Unit Public Health Laboratory, Doherty Institute |
| EPI_ISL_419992 | Victorian Infectious Diseases Reference Laboratory (VIDRL) | Victorian Infectious Diseases Reference Laboratory and Microbiological Diagnostic Unit Public Health Laboratory, Doherty Institute |
| EPI_ISL_419993 | Victorian Infectious Diseases Reference Laboratory (VIDRL) | Victorian Infectious Diseases Reference Laboratory and Microbiological Diagnostic Unit Public Health Laboratory, Doherty Institute |

| EPI_Accession_ID | Originating Laboratory | Submitting Laboratory |
|---|---|---|
| EPI_ISL_419994 | Victorian Infectious Diseases Reference Laboratory (VIDRL) | Victorian Infectious Diseases Reference Laboratory and Microbiological Diagnostic Unit Public Health Laboratory, Doherty Institute |
| EPI_ISL_419995 | Victorian Infectious Diseases Reference Laboratory (VIDRL) | Victorian Infectious Diseases Reference Laboratory and Microbiological Diagnostic Unit Public Health Laboratory, Doherty Institute |
| EPI_ISL_419996 | Victorian Infectious Diseases Reference Laboratory (VIDRL) | Victorian Infectious Diseases Reference Laboratory and Microbiological Diagnostic Unit Public Health Laboratory, Doherty Institute |
| EPI_ISL_419997 | Victorian Infectious Diseases Reference Laboratory (VIDRL) | Victorian Infectious Diseases Reference Laboratory and Microbiological Diagnostic Unit Public Health Laboratory, Doherty Institute |
| EPI_ISL_419998 | Victorian Infectious Diseases Reference Laboratory (VIDRL) | Victorian Infectious Diseases Reference Laboratory and Microbiological Diagnostic Unit Public Health Laboratory, Doherty Institute |
| EPI_ISL_419999 | Microbiological Diagnostic Unit Public Health Laboratory | Microbiological Diagnostic Unit Public Health Laboratory |
| EPI_ISL_420000 | Microbiological Diagnostic Unit Public Health Laboratory | Microbiological Diagnostic Unit Public Health Laboratory |
| EPI_ISL_420001 | Microbiological Diagnostic Unit Public Health Laboratory | Microbiological Diagnostic Unit Public Health Laboratory |
| EPI_ISL_420002 | Microbiological Diagnostic Unit Public Health Laboratory | Microbiological Diagnostic Unit Public Health Laboratory |
| EPI_ISL_420003 | Microbiological Diagnostic Unit Public Health Laboratory | Microbiological Diagnostic Unit Public Health Laboratory |
| EPI_ISL_420004 | Victorian Infectious Diseases Reference Laboratory (VIDRL) | Victorian Infectious Diseases Reference Laboratory and Microbiological Diagnostic Unit Public Health Laboratory, Doherty Institute |
| EPI_ISL_420005 | Victorian Infectious Diseases Reference Laboratory (VIDRL) | Victorian Infectious Diseases Reference Laboratory and Microbiological Diagnostic Unit Public Health Laboratory, Doherty Institute |
| EPI_ISL_420006 | Microbiological Diagnostic Unit Public Health Laboratory | Microbiological Diagnostic Unit Public Health Laboratory |

| EPI_Accession_ID | Originating Laboratory | Submitting Laboratory |
|---|---|---|
| EPI_ISL_420007 | Microbiological Diagnostic Unit Public Health Laboratory | Microbiological Diagnostic Unit Public Health Laboratory |
| EPI_ISL_420008 | Microbiological Diagnostic Unit Public Health Laboratory | Microbiological Diagnostic Unit Public Health Laboratory |
| EPI_ISL_420009 | Microbiological Diagnostic Unit Public Health Laboratory | Microbiological Diagnostic Unit Public Health Laboratory |
| EPI_ISL_420010 | Microbiological Diagnostic Unit Public Health Laboratory | Microbiological Diagnostic Unit Public Health Laboratory |
| EPI_ISL_420011 | Microbiological Diagnostic Unit Public Health Laboratory | Microbiological Diagnostic Unit Public Health Laboratory |
| EPI_ISL_420012 | Microbiological Diagnostic Unit Public Health Laboratory | Microbiological Diagnostic Unit Public Health Laboratory |
| EPI_ISL_420013 | Microbiological Diagnostic Unit Public Health Laboratory | Microbiological Diagnostic Unit Public Health Laboratory |
| EPI_ISL_420014 | Microbiological Diagnostic Unit Public Health Laboratory | Microbiological Diagnostic Unit Public Health Laboratory |
| EPI_ISL_420015 | Microbiological Diagnostic Unit Public Health Laboratory | Microbiological Diagnostic Unit Public Health Laboratory |
| EPI_ISL_420016 | Microbiological Diagnostic Unit Public Health Laboratory | Microbiological Diagnostic Unit Public Health Laboratory |
| EPI_ISL_420017 | Microbiological Diagnostic Unit Public Health Laboratory | Microbiological Diagnostic Unit Public Health Laboratory |
| EPI_ISL_420018 | Virginia DCLS | Virginia DCLS |
| EPI_ISL_420019 | Virginia DCLS | Virginia DCLS |
| EPI_ISL_420020 | Virginia DCLS | Virginia DCLS |
| EPI_ISL_420021 | Virginia DCLS | Virginia DCLS |
| EPI_ISL_420022 | Virginia DCLS | Virginia DCLS |
| EPI_ISL_420023 | Virginia DCLS | Virginia DCLS |
| EPI_ISL_420024 | Virginia DCLS | Virginia DCLS |
| EPI_ISL_420025 | Virginia DCLS | Virginia DCLS |
| EPI_ISL_420026 | Virginia DCLS | Virginia DCLS |
| EPI_ISL_420027 | Virginia DCLS | Virginia DCLS |
| EPI_ISL_420028 | Virginia DCLS | Virginia DCLS |

| EPI_Accession_ID | Originating Laboratory | Submitting Laboratory |
|---|---|---|
| EPI_ISL_420029 | Virginia DCLS | Virginia DCLS |
| EPI_ISL_420030 | Viral Respiratory Lab, National Institute for Biomedical Research (INRB) | Pathogen Sequencing Lab, National Institute for Biomedical Research (INRB) |
| EPI_ISL_420031 | Viral Respiratory Lab, National Institute for Biomedical Research (INRB) | Pathogen Sequencing Lab, National Institute for Biomedical Research (INRB) |
| EPI_ISL_420032 | Viral Respiratory Lab, National Institute for Biomedical Research (INRB) | Pathogen Sequencing Lab, National Institute for Biomedical Research (INRB) |
| EPI_ISL_420033 | Viral Respiratory Lab, National Institute for Biomedical Research (INRB) | Pathogen Sequencing Lab, National Institute for Biomedical Research (INRB) |
| EPI_ISL_420034 | Viral Respiratory Lab, National Institute for Biomedical Research (INRB) | Pathogen Sequencing Lab, National Institute for Biomedical Research (INRB) |
| EPI_ISL_420035 | Viral Respiratory Lab, National Institute for Biomedical Research (INRB) | Pathogen Sequencing Lab, National Institute for Biomedical Research (INRB) |
| EPI_ISL_420036 | Victorian Infectious Diseases Reference Laboratory (VIDRL) | Victorian Infectious Diseases Reference Laboratory and Microbiological Diagnostic Unit Public Health Laboratory, Doherty Institute |
| EPI_ISL_420037 | NIC Viral Respiratory Unit - Institut Pasteur of Algeria | National Reference Center for Viruses of Respiratory Infections, Institut Pasteur, Paris |
| EPI_ISL_420038 | Sentinelles network | National Reference Center for Viruses of Respiratory Infections, Institut Pasteur, Paris |
| EPI_ISL_420039 | L'Air du Temps | National Reference Center for Viruses of Respiratory Infections, Institut Pasteur, Paris |
| EPI_ISL_420040 | L'Air du Temps | National Reference Center for Viruses of Respiratory Infections, Institut Pasteur, Paris |
| EPI_ISL_420041 | CH CompiÃ¨gne Laboratoire de Biologie | National Reference Center for Viruses of Respiratory Infections, Institut Pasteur, Paris |
| EPI_ISL_420042 | Service de Biologie clinique | National Reference Center for Viruses of Respiratory Infections, Institut Pasteur, Paris |
| EPI_ISL_420043 | CMIP | National Reference Center for Viruses of Respiratory Infections, Institut Pasteur, Paris |
| EPI_ISL_420044 | CH Jean de Navarre Laboratoire de Biologie | National Reference Center for Viruses of Respiratory Infections, Institut Pasteur, Paris |

| EPI_Accession_ID | Originating Laboratory | Submitting Laboratory |
|---|---|---|
| EPI_ISL_420045 | Sentinelles network | National Reference Center for Viruses of Respiratory Infections, Institut Pasteur, Paris |
| EPI_ISL_420046 | RÃ©sidence Villa Caroline | National Reference Center for Viruses of Respiratory Infections, Institut Pasteur, Paris |
| EPI_ISL_420047 | RÃ©sidence Villa Caroline | National Reference Center for Viruses of Respiratory Infections, Institut Pasteur, Paris |
| EPI_ISL_420048 | Service de Biologie MÃ©dicale - BP 125 | National Reference Center for Viruses of Respiratory Infections, Institut Pasteur, Paris |
| EPI_ISL_420049 | CH CompiÃ¨gne Laboratoire de Biologie | National Reference Center for Viruses of Respiratory Infections, Institut Pasteur, Paris |
| EPI_ISL_420050 | CH CompiÃ¨gne Laboratoire de Biologie | National Reference Center for Viruses of Respiratory Infections, Institut Pasteur, Paris |
| EPI_ISL_420051 | RÃ©sidence Eleusis | National Reference Center for Viruses of Respiratory Infections, Institut Pasteur, Paris |
| EPI_ISL_420052 | RÃ©sidence les Marines | National Reference Center for Viruses of Respiratory Infections, Institut Pasteur, Paris |
| EPI_ISL_420053 | CH Jean de Navarre Laboratoire de Biologie | National Reference Center for Viruses of Respiratory Infections, Institut Pasteur, Paris |
| EPI_ISL_420054 | RÃ©sidence de maintenon | National Reference Center for Viruses of Respiratory Infections, Institut Pasteur, Paris |
| EPI_ISL_420055 | Sentinelles network | National Reference Center for Viruses of Respiratory Infections, Institut Pasteur, Paris |
| EPI_ISL_420056 | CH CompiÃ¨gne Laboratoire de Biologie | National Reference Center for Viruses of Respiratory Infections, Institut Pasteur, Paris |
| EPI_ISL_420057 | CH CompiÃ¨gne Laboratoire de Biologie | National Reference Center for Viruses of Respiratory Infections, Institut Pasteur, Paris |
| EPI_ISL_420058 | Service de Biologie MÃ©dicale - BP 125 | National Reference Center for Viruses of Respiratory Infections, Institut Pasteur, Paris |
| EPI_ISL_420059 | Service de Biologie MÃ©dicale - BP 125 | National Reference Center for Viruses of Respiratory Infections, Institut Pasteur, Paris |
| EPI_ISL_420060 | Service de Biologie MÃ©dicale - BP 125 | National Reference Center for Viruses of Respiratory Infections, Institut Pasteur, Paris |

| EPI_Accession_ID | Originating Laboratory | Submitting Laboratory |
|---|---|---|
| EPI_ISL_420061 | CMIP | National Reference Center for Viruses of Respiratory Infections, Institut Pasteur, Paris |
| EPI_ISL_420062 | Service de Biologie MÃ©dicale - BP 125 | National Reference Center for Viruses of Respiratory Infections, Institut Pasteur, Paris |
| EPI_ISL_420063 | Labo BM - Site de Juvisy - Hopital GÃ©nÃ©ral | National Reference Center for Viruses of Respiratory Infections, Institut Pasteur, Paris |
| EPI_ISL_420064 | Service de Biologie MÃ©dicale - BP 125 | National Reference Center for Viruses of Respiratory Infections, Institut Pasteur, Paris |
| EPI_ISL_420069 | Institut Pasteur Dakar | Institut Pasteur de Dakar |
| EPI_ISL_420070 | Institut Pasteur Dakar | Institut Pasteur de Dakar |
| EPI_ISL_420071 | Institut Pasteur Dakar | Institut Pasteur de Dakar |
| EPI_ISL_420072 | Institut Pasteur Dakar | Institut Pasteur de Dakar |
| EPI_ISL_420073 | Institut Pasteur Dakar | Institut Pasteur de Dakar |
| EPI_ISL_420074 | Institut Pasteur Dakar | Institut Pasteur de Dakar |
| EPI_ISL_420076 | Institut Pasteur Dakar | Institut Pasteur de Dakar |
| EPI_ISL_420077 | Institut Pasteur Dakar | Institut Pasteur de Dakar |
| EPI_ISL_420078 | Institut Pasteur Dakar | Institut Pasteur de Dakar |
| EPI_ISL_420079 | Institut Pasteur Dakar | Institut Pasteur de Dakar |
| EPI_ISL_420080 | WHO National Influenza Centre Russian Federation | WHO National Influenza Centre Russian Federation |
| EPI_ISL_420081 | WHO National Influenza Centre Russian Federation | WHO National Influenza Centre Russian Federation |
| EPI_ISL_420082 | Centers for Disease Control, R.O.C. (Taiwan) | Centers for Disease Control, R.O.C. (Taiwan) |
| EPI_ISL_420083 | Centers for Disease Control, R.O.C. (Taiwan) | Centers for Disease Control, R.O.C. (Taiwan) |
| EPI_ISL_420084 | Centers for Disease Control, R.O.C. (Taiwan) | Centers for Disease Control, R.O.C. (Taiwan) |
| EPI_ISL_420085 | Centers for Disease Control, R.O.C. (Taiwan) | Centers for Disease Control, R.O.C. (Taiwan) |
| EPI_ISL_420086 | Yale Clinical Virology Laboratory | Grubaugh Lab - Yale School of Public Health |
| EPI_ISL_420087 | Yale Clinical Virology Laboratory | Grubaugh Lab - Yale School of Public Health |
| EPI_ISL_420088 | Yale Clinical Virology Laboratory | Grubaugh Lab - Yale School of Public Health |
| EPI_ISL_420089 | Yale Clinical Virology Laboratory | Grubaugh Lab - Yale School of Public Health |
| EPI_ISL_420090 | Yale Clinical Virology Laboratory | Grubaugh Lab - Yale School of Public Health |
| EPI_ISL_420091 | Yale Clinical Virology Laboratory | Grubaugh Lab - Yale School of Public Health |
| EPI_ISL_420092 | Yale Clinical Virology Laboratory | Grubaugh Lab - Yale School of Public Health |
| EPI_ISL_420093 | Yale Clinical Virology Laboratory | Grubaugh Lab - Yale School of Public Health |
| EPI_ISL_420094 | Yale Clinical Virology Laboratory | Grubaugh Lab - Yale School of Public Health |

| EPI_Accession_ID | Originating Laboratory | Submitting Laboratory |
|---|---|---|
| EPI_ISL_420095 | Yale Clinical Virology Laboratory | Grubaugh Lab - Yale School of Public Health |
| EPI_ISL_420096 | Yale Clinical Virology Laboratory | Grubaugh Lab - Yale School of Public Health |
| EPI_ISL_420097 | Yale Clinical Virology Laboratory | Grubaugh Lab - Yale School of Public Health |
| EPI_ISL_420098 | Yale Clinical Virology Laboratory | Grubaugh Lab - Yale School of Public Health |
| EPI_ISL_420099 | National Centre for Infectious Diseases | Programme in Emerging Infectious Diseases, Duke-NUS Medical School |
| EPI_ISL_420100 | National Centre for Infectious Diseases | Programme in Emerging Infectious Diseases, Duke-NUS Medical School |
| EPI_ISL_420101 | National Centre for Infectious Diseases | Programme in Emerging Infectious Diseases, Duke-NUS Medical School |
| EPI_ISL_420102 | National Centre for Infectious Diseases | Programme in Emerging Infectious Diseases, Duke-NUS Medical School |
| EPI_ISL_420103 | National Centre for Infectious Diseases | Programme in Emerging Infectious Diseases, Duke-NUS Medical School |
| EPI_ISL_420104 | National Centre for Infectious Diseases | Programme in Emerging Infectious Diseases, Duke-NUS Medical School |
| EPI_ISL_420105 | National Centre for Infectious Diseases | Programme in Emerging Infectious Diseases, Duke-NUS Medical School |
| EPI_ISL_420106 | National Centre for Infectious Diseases | Programme in Emerging Infectious Diseases, Duke-NUS Medical School |
| EPI_ISL_420107 | National Centre for Infectious Diseases | Programme in Emerging Infectious Diseases, Duke-NUS Medical School |
| EPI_ISL_420108 | National Centre for Infectious Diseases | Programme in Emerging Infectious Diseases, Duke-NUS Medical School |
| EPI_ISL_420109 | National Centre for Infectious Diseases | Programme in Emerging Infectious Diseases, Duke-NUS Medical School |
| EPI_ISL_420110 | National Centre for Infectious Diseases | Programme in Emerging Infectious Diseases, Duke-NUS Medical School |
| EPI_ISL_420111 | National Centre for Infectious Diseases | Programme in Emerging Infectious Diseases, Duke-NUS Medical School |
| EPI_ISL_420112 | Servicio de MicrobiologÃ-a. Consorcio Hospital General Universitario de Valencia | Sequencing and Bioinformatics Service and Molecular Epidemiology Research Group. FISABIO-Public Health |

| EPI_Accession_ID | Originating Laboratory | Submitting Laboratory |
|---|---|---|
| EPI_ISL_420113 | Servicio de MicrobiologÃ-a. Consorcio Hospital General Universitario de Valencia | Sequencing and Bioinformatics Service and Molecular Epidemiology Research Group. FISABIO-Public Health |
| EPI_ISL_420114 | Servicio de MicrobiologÃ-a. Consorcio Hospital General Universitario de Valencia | Sequencing and Bioinformatics Service and Molecular Epidemiology Research Group. FISABIO-Public Health |
| EPI_ISL_420115 | Servicio de MicrobiologÃ-a. Consorcio Hospital General Universitario de Valencia | Sequencing and Bioinformatics Service and Molecular Epidemiology Research Group. FISABIO-Public Health |
| EPI_ISL_420116 | Servicio de MicrobiologÃ-a. Consorcio Hospital General Universitario de Valencia | Sequencing and Bioinformatics Service and Molecular Epidemiology Research Group. FISABIO-Public Health |
| EPI_ISL_420117 | Servicio de MicrobiologÃ-a. Consorcio Hospital General Universitario de Valencia | Sequencing and Bioinformatics Service and Molecular Epidemiology Research Group. FISABIO-Public Health |
| EPI_ISL_420118 | Servicio de MicrobiologÃ-a. Consorcio Hospital General Universitario de Valencia | Sequencing and Bioinformatics Service and Molecular Epidemiology Research Group. FISABIO-Public Health |
| EPI_ISL_420119 | Servicio de MicrobiologÃ-a. Consorcio Hospital General Universitario de Valencia | Sequencing and Bioinformatics Service and Molecular Epidemiology Research Group. FISABIO-Public Health |
| EPI_ISL_420120 | Servicio de MicrobiologÃ-a. Consorcio Hospital General Universitario de Valencia | Sequencing and Bioinformatics Service and Molecular Epidemiology Research Group. FISABIO-Public Health |
| EPI_ISL_420121 | Servicio de MicrobiologÃ-a. Consorcio Hospital General Universitario de Valencia | Sequencing and Bioinformatics Service and Molecular Epidemiology Research Group. FISABIO-Public Health |
| EPI_ISL_420122 | Servicio de MicrobiologÃ-a. Consorcio Hospital General Universitario de Valencia | Sequencing and Bioinformatics Service and Molecular Epidemiology Research Group. FISABIO-Public Health |
| EPI_ISL_420123 | Servicio de MicrobiologÃ-a. Consorcio Hospital General Universitario de Valencia | Sequencing and Bioinformatics Service and Molecular Epidemiology Research Group. FISABIO-Public Health |
| EPI_ISL_420124 | Servicio de MicrobiologÃ-a. Consorcio Hospital General Universitario de Valencia | Sequencing and Bioinformatics Service and Molecular Epidemiology Research Group. FISABIO-Public Health |
| EPI_ISL_420125 | Servicio de Microbiologia. Hospital Clinico Universitario de Valencia | Sequencing and Bioinformatics Service and Molecular Epidemiology Research Group. FISABIO-Public Health |
| EPI_ISL_420126 | Servicio de Microbiologia. Hospital Clinico Universitario de Valencia | Sequencing and Bioinformatics Service and Molecular Epidemiology Research Group. FISABIO-Public Health |
| EPI_ISL_420127 | Servicio de Microbiologia. Hospital Clinico Universitario de Valencia | Sequencing and Bioinformatics Service and Molecular Epidemiology Research Group. FISABIO-Public Health |
| EPI_ISL_420128 | Servicio de Microbiologia. Hospital Clinico Universitario de Valencia | Sequencing and Bioinformatics Service and Molecular Epidemiology Research Group. FISABIO-Public Health |

| EPI_Accession_ID | Originating Laboratory | Submitting Laboratory |
|---|---|---|
| EPI_ISL_420129 | Servicio de Microbiologia. Hospital Clinico Universitario de Valencia | Sequencing and Bioinformatics Service and Molecular Epidemiology Research Group. FISABIO-Public Health |
| EPI_ISL_420130 | Servicio de Microbiologia. Hospital Clinico Universitario de Valencia | Sequencing and Bioinformatics Service and Molecular Epidemiology Research Group. FISABIO-Public Health |
| EPI_ISL_420131 | Servicio de Microbiologia. Hospital Clinico Universitario de Valencia | Sequencing and Bioinformatics Service and Molecular Epidemiology Research Group. FISABIO-Public Health |
| EPI_ISL_420132 | Servicio de Microbiologia. Hospital Clinico Universitario de Valencia | Sequencing and Bioinformatics Service and Molecular Epidemiology Research Group. FISABIO-Public Health |
| EPI_ISL_420134 | Akershus University Hospital, Department for Microbiology and Infectious Disease Control | Norwegian Institute of Public Health, Department of Virology |
| EPI_ISL_420135 | Oslo University Hospital, Department of Medical Microbiology | Norwegian Institute of Public Health, Department of Virology |
| EPI_ISL_420136 | Akershus University Hospital, Department for Microbiology and Infectious Disease Control | Norwegian Institute of Public Health, Department of Virology |
| EPI_ISL_420137 | Vestfold Hospital, Tonsberg Department of Microbiology | Norwegian Institute of Public Health, Department of Virology |
| EPI_ISL_420138 | University Hospital of Northern Norway, Department for Microbiology and Infectious Disease Control | Norwegian Institute of Public Health, Department of Virology |
| EPI_ISL_420139 | Akershus University Hospital, Department for Microbiology and Infectious Disease Control | Norwegian Institute of Public Health, Department of Virology |
| EPI_ISL_420140 | Department for Virology, Molecular Biology and Genome Research, R. G. Lugar Center for Public Health Research, National Center for Disease Control and Public Health (NCDC) of Georgia. | Department for Virology, Molecular Biology and Genome Research, R. G. Lugar Center for Public Health Research, National Center for Disease Control and Public Health (NCDC) of Georgia. |
| EPI_ISL_420141 | Furst Medical Laboratory | Norwegian Institute of Public Health, Department of Virology |
| EPI_ISL_420142 | Department for Virology, Molecular Biology and Genome Research, R. G. Lugar Center for Public Health Research, National Center for Disease Control and Public Health (NCDC) of Georgia. | Department for Virology, Molecular Biology and Genome Research, R. G. Lugar Center for Public Health Research, National Center for Disease Control and Public Health (NCDC) of Georgia. |
| EPI_ISL_420143 | Unilabs Laboratory Medicine | Norwegian Institute of Public Health, Department of Virology |

| EPI_Accession_ID | Originating Laboratory | Submitting Laboratory |
|---|---|---|
| EPI_ISL_420144 | Department for Virology, Molecular Biology and Genome Research, R. G. Lugar Center for Public Health Research, National Center for Disease Control and Public Health (NCDC) of Georgia. | Department for Virology, Molecular Biology and Genome Research, R. G. Lugar Center for Public Health Research, National Center for Disease Control and Public Health (NCDC) of Georgia. |
| EPI_ISL_420145 | Forde Hospital Department of Microbiology | Norwegian Institute of Public Health, Department of Virology |
| EPI_ISL_420146 | Furst Medical Laboratory | Norwegian Institute of Public Health, Department of Virology |
| EPI_ISL_420147 | Hospital of Southern Norway - Kristiansand, Department of Medical Microbiology | Norwegian Institute of Public Health, Department of Virology |
| EPI_ISL_420148 | Ostfold Hospital Trust -Kalnes Centre for Laboratory Medicine Section for gene technology and infection serology | Norwegian Institute of Public Health, Department of Virology |
| EPI_ISL_420149 | Stavanger University Hospital, Department of Medical Microbiology | Norwegian Institute of Public Health, Department of Virology |
| EPI_ISL_420150 | Oslo University Hospital, Department of Medical Microbiology | Norwegian Institute of Public Health, Department of Virology |
| EPI_ISL_420151 | Nordland Hospital - Bodo, Laboratory Department, Molecular Biology Unit | Norwegian Institute of Public Health, Department of Virology |
| EPI_ISL_420152 | University Hospital of Northern Norway, Department for Microbiology and Infectious Disease Control | Norwegian Institute of Public Health, Department of Virology |
| EPI_ISL_420153 | University Hospital of Northern Norway, Department for Microbiology and Infectious Disease Control | Norwegian Institute of Public Health, Department of Virology |
| EPI_ISL_420154 | Virology Department, Sheffield Teaching Hospitals NHS Foundation Trust | Department of Infection, Immunity and Cardiovascular Disease, The Florey Institute, The Medical School, University of Sheffield |
| EPI_ISL_420155 | Virology Department, Sheffield Teaching Hospitals NHS Foundation Trust | Department of Infection, Immunity and Cardiovascular Disease, The Florey Institute, The Medical School, University of Sheffield |
| EPI_ISL_420156 | Virology Department, Sheffield Teaching Hospitals NHS Foundation Trust | Department of Infection, Immunity and Cardiovascular Disease, The Florey Institute, The Medical School, University of Sheffield |

| EPI_Accession_ID | Originating Laboratory | Submitting Laboratory |
|---|---|---|
| EPI_ISL_420157 | Virology Department, Sheffield Teaching Hospitals NHS Foundation Trust | Department of Infection, Immunity and Cardiovascular Disease, The Florey Institute, The Medical School, University of Sheffield |
| EPI_ISL_420158 | Virology Department, Sheffield Teaching Hospitals NHS Foundation Trust | Department of Infection, Immunity and Cardiovascular Disease, The Florey Institute, The Medical School, University of Sheffield |
| EPI_ISL_420159 | Virology Department, Sheffield Teaching Hospitals NHS Foundation Trust | Department of Infection, Immunity and Cardiovascular Disease, The Florey Institute, The Medical School, University of Sheffield |
| EPI_ISL_420160 | Virology Department, Sheffield Teaching Hospitals NHS Foundation Trust | Department of Infection, Immunity and Cardiovascular Disease, The Florey Institute, The Medical School, University of Sheffield |
| EPI_ISL_420161 | Virology Department, Sheffield Teaching Hospitals NHS Foundation Trust | Department of Infection, Immunity and Cardiovascular Disease, The Florey Institute, The Medical School, University of Sheffield |
| EPI_ISL_420162 | Virology Department, Sheffield Teaching Hospitals NHS Foundation Trust | Department of Infection, Immunity and Cardiovascular Disease, The Florey Institute, The Medical School, University of Sheffield |
| EPI_ISL_420163 | Virology Department, Sheffield Teaching Hospitals NHS Foundation Trust | Department of Infection, Immunity and Cardiovascular Disease, The Florey Institute, The Medical School, University of Sheffield |
| EPI_ISL_420164 | Virology Department, Sheffield Teaching Hospitals NHS Foundation Trust | Department of Infection, Immunity and Cardiovascular Disease, The Florey Institute, The Medical School, University of Sheffield |
| EPI_ISL_420165 | Virology Department, Sheffield Teaching Hospitals NHS Foundation Trust | Department of Infection, Immunity and Cardiovascular Disease, The Florey Institute, The Medical School, University of Sheffield |
| EPI_ISL_420166 | Virology Department, Sheffield Teaching Hospitals NHS Foundation Trust | Department of Infection, Immunity and Cardiovascular Disease, The Florey Institute, The Medical School, University of Sheffield |
| EPI_ISL_420167 | Virology Department, Sheffield Teaching Hospitals NHS Foundation Trust | Department of Infection, Immunity and Cardiovascular Disease, The Florey Institute, The Medical School, University of Sheffield |

| EPI_Accession_ID | Originating Laboratory | Submitting Laboratory |
|---|---|---|
| EPI_ISL_420168 | Virology Department, Sheffield Teaching Hospitals NHS Foundation Trust | Department of Infection, Immunity and Cardiovascular Disease, The Florey Institute, The Medical School, University of Sheffield |
| EPI_ISL_420169 | Virology Department, Sheffield Teaching Hospitals NHS Foundation Trust | Department of Infection, Immunity and Cardiovascular Disease, The Florey Institute, The Medical School, University of Sheffield |
| EPI_ISL_420170 | Virology Department, Sheffield Teaching Hospitals NHS Foundation Trust | Department of Infection, Immunity and Cardiovascular Disease, The Florey Institute, The Medical School, University of Sheffield |
| EPI_ISL_420171 | Virology Department, Sheffield Teaching Hospitals NHS Foundation Trust | Department of Infection, Immunity and Cardiovascular Disease, The Florey Institute, The Medical School, University of Sheffield |
| EPI_ISL_420172 | Virology Department, Sheffield Teaching Hospitals NHS Foundation Trust | Department of Infection, Immunity and Cardiovascular Disease, The Florey Institute, The Medical School, University of Sheffield |
| EPI_ISL_420173 | Virology Department, Sheffield Teaching Hospitals NHS Foundation Trust | Department of Infection, Immunity and Cardiovascular Disease, The Florey Institute, The Medical School, University of Sheffield |
| EPI_ISL_420174 | Virology Department, Sheffield Teaching Hospitals NHS Foundation Trust | Department of Infection, Immunity and Cardiovascular Disease, The Florey Institute, The Medical School, University of Sheffield |
| EPI_ISL_420175 | Virology Department, Sheffield Teaching Hospitals NHS Foundation Trust | Department of Infection, Immunity and Cardiovascular Disease, The Florey Institute, The Medical School, University of Sheffield |
| EPI_ISL_420176 | Virology Department, Sheffield Teaching Hospitals NHS Foundation Trust | Department of Infection, Immunity and Cardiovascular Disease, The Florey Institute, The Medical School, University of Sheffield |
| EPI_ISL_420177 | Virology Department, Sheffield Teaching Hospitals NHS Foundation Trust | Department of Infection, Immunity and Cardiovascular Disease, The Florey Institute, The Medical School, University of Sheffield |
| EPI_ISL_420178 | Virology Department, Sheffield Teaching Hospitals NHS Foundation Trust | Department of Infection, Immunity and Cardiovascular Disease, The Florey Institute, The Medical School, University of Sheffield |

| EPI_Accession_ID | Originating Laboratory | Submitting Laboratory |
|---|---|---|
| EPI_ISL_420179 | Virology Department, Sheffield Teaching Hospitals NHS Foundation Trust | Department of Infection, Immunity and Cardiovascular Disease, The Florey Institute, The Medical School, University of Sheffield |
| EPI_ISL_420180 | Virology Department, Sheffield Teaching Hospitals NHS Foundation Trust | Department of Infection, Immunity and Cardiovascular Disease, The Florey Institute, The Medical School, University of Sheffield |
| EPI_ISL_420181 | Virology Department, Sheffield Teaching Hospitals NHS Foundation Trust | Department of Infection, Immunity and Cardiovascular Disease, The Florey Institute, The Medical School, University of Sheffield |
| EPI_ISL_420182 | Virology Department, Sheffield Teaching Hospitals NHS Foundation Trust | Department of Infection, Immunity and Cardiovascular Disease, The Florey Institute, The Medical School, University of Sheffield |
| EPI_ISL_420183 | Virology Department, Sheffield Teaching Hospitals NHS Foundation Trust | Department of Infection, Immunity and Cardiovascular Disease, The Florey Institute, The Medical School, University of Sheffield |
| EPI_ISL_420184 | Virology Department, Sheffield Teaching Hospitals NHS Foundation Trust | Department of Infection, Immunity and Cardiovascular Disease, The Florey Institute, The Medical School, University of Sheffield |
| EPI_ISL_420185 | Virology Department, Sheffield Teaching Hospitals NHS Foundation Trust | Department of Infection, Immunity and Cardiovascular Disease, The Florey Institute, The Medical School, University of Sheffield |
| EPI_ISL_420186 | Virology Department, Sheffield Teaching Hospitals NHS Foundation Trust | Department of Infection, Immunity and Cardiovascular Disease, The Florey Institute, The Medical School, University of Sheffield |
| EPI_ISL_420187 | Virology Department, Sheffield Teaching Hospitals NHS Foundation Trust | Department of Infection, Immunity and Cardiovascular Disease, The Florey Institute, The Medical School, University of Sheffield |
| EPI_ISL_420188 | Virology Department, Sheffield Teaching Hospitals NHS Foundation Trust | Department of Infection, Immunity and Cardiovascular Disease, The Florey Institute, The Medical School, University of Sheffield |
| EPI_ISL_420189 | Virology Department, Sheffield Teaching Hospitals NHS Foundation Trust | Department of Infection, Immunity and Cardiovascular Disease, The Florey Institute, The Medical School, University of Sheffield |

| EPI_Accession_ID | Originating Laboratory | Submitting Laboratory |
|---|---|---|
| EPI_ISL_420190 | Virology Department, Sheffield Teaching Hospitals NHS Foundation Trust | Department of Infection, Immunity and Cardiovascular Disease, The Florey Institute, The Medical School, University of Sheffield |
| EPI_ISL_420191 | Virology Department, Sheffield Teaching Hospitals NHS Foundation Trust | Department of Infection, Immunity and Cardiovascular Disease, The Florey Institute, The Medical School, University of Sheffield |
| EPI_ISL_420192 | Virology Department, Sheffield Teaching Hospitals NHS Foundation Trust | Department of Infection, Immunity and Cardiovascular Disease, The Florey Institute, The Medical School, University of Sheffield |
| EPI_ISL_420193 | Virology Department, Sheffield Teaching Hospitals NHS Foundation Trust | Department of Infection, Immunity and Cardiovascular Disease, The Florey Institute, The Medical School, University of Sheffield |
| EPI_ISL_420194 | Virology Department, Sheffield Teaching Hospitals NHS Foundation Trust | Department of Infection, Immunity and Cardiovascular Disease, The Florey Institute, The Medical School, University of Sheffield |
| EPI_ISL_420195 | Virology Department, Sheffield Teaching Hospitals NHS Foundation Trust | Department of Infection, Immunity and Cardiovascular Disease, The Florey Institute, The Medical School, University of Sheffield |
| EPI_ISL_420196 | Virology Department, Sheffield Teaching Hospitals NHS Foundation Trust | Department of Infection, Immunity and Cardiovascular Disease, The Florey Institute, The Medical School, University of Sheffield |
| EPI_ISL_420197 | Virology Department, Sheffield Teaching Hospitals NHS Foundation Trust | Department of Infection, Immunity and Cardiovascular Disease, The Florey Institute, The Medical School, University of Sheffield |
| EPI_ISL_420198 | Virology Department, Sheffield Teaching Hospitals NHS Foundation Trust | Department of Infection, Immunity and Cardiovascular Disease, The Florey Institute, The Medical School, University of Sheffield |
| EPI_ISL_420199 | Virology Department, Sheffield Teaching Hospitals NHS Foundation Trust | Department of Infection, Immunity and Cardiovascular Disease, The Florey Institute, The Medical School, University of Sheffield |
| EPI_ISL_420200 | Virology Department, Sheffield Teaching Hospitals NHS Foundation Trust | Department of Infection, Immunity and Cardiovascular Disease, The Florey Institute, The Medical School, University of Sheffield |

| EPI_Accession_ID | Originating Laboratory | Submitting Laboratory |
|---|---|---|
| EPI_ISL_420201 | Virology Department, Sheffield Teaching Hospitals NHS Foundation Trust | Department of Infection, Immunity and Cardiovascular Disease, The Florey Institute, The Medical School, University of Sheffield |
| EPI_ISL_420202 | Virology Department, Sheffield Teaching Hospitals NHS Foundation Trust | Department of Infection, Immunity and Cardiovascular Disease, The Florey Institute, The Medical School, University of Sheffield |
| EPI_ISL_420203 | Virology Department, Sheffield Teaching Hospitals NHS Foundation Trust | Department of Infection, Immunity and Cardiovascular Disease, The Florey Institute, The Medical School, University of Sheffield |
| EPI_ISL_420204 | Virology Department, Sheffield Teaching Hospitals NHS Foundation Trust | Department of Infection, Immunity and Cardiovascular Disease, The Florey Institute, The Medical School, University of Sheffield |
| EPI_ISL_420205 | Virology Department, Sheffield Teaching Hospitals NHS Foundation Trust | Department of Infection, Immunity and Cardiovascular Disease, The Florey Institute, The Medical School, University of Sheffield |
| EPI_ISL_420206 | Virology Department, Sheffield Teaching Hospitals NHS Foundation Trust | Department of Infection, Immunity and Cardiovascular Disease, The Florey Institute, The Medical School, University of Sheffield |
| EPI_ISL_420207 | Virology Department, Sheffield Teaching Hospitals NHS Foundation Trust | Department of Infection, Immunity and Cardiovascular Disease, The Florey Institute, The Medical School, University of Sheffield |
| EPI_ISL_420208 | Virology Department, Sheffield Teaching Hospitals NHS Foundation Trust | Department of Infection, Immunity and Cardiovascular Disease, The Florey Institute, The Medical School, University of Sheffield |
| EPI_ISL_420209 | Virology Department, Sheffield Teaching Hospitals NHS Foundation Trust | Department of Infection, Immunity and Cardiovascular Disease, The Florey Institute, The Medical School, University of Sheffield |
| EPI_ISL_420210 | Virology Department, Sheffield Teaching Hospitals NHS Foundation Trust | Department of Infection, Immunity and Cardiovascular Disease, The Florey Institute, The Medical School, University of Sheffield |
| EPI_ISL_420211 | Virology Department, Sheffield Teaching Hospitals NHS Foundation Trust | Department of Infection, Immunity and Cardiovascular Disease, The Florey Institute, The Medical School, University of Sheffield |

| EPI_Accession_ID | Originating Laboratory | Submitting Laboratory |
|---|---|---|
| EPI_ISL_420212 | Virology Department, Sheffield Teaching Hospitals NHS Foundation Trust | Department of Infection, Immunity and Cardiovascular Disease, The Florey Institute, The Medical School, University of Sheffield |
| EPI_ISL_420213 | Virology Department, Sheffield Teaching Hospitals NHS Foundation Trust | Department of Infection, Immunity and Cardiovascular Disease, The Florey Institute, The Medical School, University of Sheffield |
| EPI_ISL_420214 | Virology Department, Sheffield Teaching Hospitals NHS Foundation Trust | Department of Infection, Immunity and Cardiovascular Disease, The Florey Institute, The Medical School, University of Sheffield |
| EPI_ISL_420215 | Virology Department, Sheffield Teaching Hospitals NHS Foundation Trust | Department of Infection, Immunity and Cardiovascular Disease, The Florey Institute, The Medical School, University of Sheffield |
| EPI_ISL_420216 | Virology Department, Sheffield Teaching Hospitals NHS Foundation Trust | Department of Infection, Immunity and Cardiovascular Disease, The Florey Institute, The Medical School, University of Sheffield |
| EPI_ISL_420217 | Virology Department, Sheffield Teaching Hospitals NHS Foundation Trust | Department of Infection, Immunity and Cardiovascular Disease, The Florey Institute, The Medical School, University of Sheffield |
| EPI_ISL_420218 | Virology Department, Sheffield Teaching Hospitals NHS Foundation Trust | Department of Infection, Immunity and Cardiovascular Disease, The Florey Institute, The Medical School, University of Sheffield |
| EPI_ISL_420219 | Virology Department, Sheffield Teaching Hospitals NHS Foundation Trust | Department of Infection, Immunity and Cardiovascular Disease, The Florey Institute, The Medical School, University of Sheffield |
| EPI_ISL_420220 | Virology Department, Sheffield Teaching Hospitals NHS Foundation Trust | Department of Infection, Immunity and Cardiovascular Disease, The Florey Institute, The Medical School, University of Sheffield |
| EPI_ISL_420221 | Virology Department, Sheffield Teaching Hospitals NHS Foundation Trust | Department of Infection, Immunity and Cardiovascular Disease, The Florey Institute, The Medical School, University of Sheffield |
| EPI_ISL_420222 | Virology Department, Sheffield Teaching Hospitals NHS Foundation Trust | Department of Infection, Immunity and Cardiovascular Disease, The Florey Institute, The Medical School, University of Sheffield |

| EPI_Accession_ID | Originating Laboratory | Submitting Laboratory |
|---|---|---|
| EPI_ISL_420223 | Virology Department, Sheffield Teaching Hospitals NHS Foundation Trust | Department of Infection, Immunity and Cardiovascular Disease, The Florey Institute, The Medical School, University of Sheffield |
| EPI_ISL_420224 | Virology Department, Sheffield Teaching Hospitals NHS Foundation Trust | Department of Infection, Immunity and Cardiovascular Disease, The Florey Institute, The Medical School, University of Sheffield |
| EPI_ISL_420225 | Virology Department, Sheffield Teaching Hospitals NHS Foundation Trust | Department of Infection, Immunity and Cardiovascular Disease, The Florey Institute, The Medical School, University of Sheffield |
| EPI_ISL_420226 | Virology Department, Sheffield Teaching Hospitals NHS Foundation Trust | Department of Infection, Immunity and Cardiovascular Disease, The Florey Institute, The Medical School, University of Sheffield |
| EPI_ISL_420227 | Virology Department, Sheffield Teaching Hospitals NHS Foundation Trust | Department of Infection, Immunity and Cardiovascular Disease, The Florey Institute, The Medical School, University of Sheffield |
| EPI_ISL_420228 | Virology Department, Sheffield Teaching Hospitals NHS Foundation Trust | Department of Infection, Immunity and Cardiovascular Disease, The Florey Institute, The Medical School, University of Sheffield |
| EPI_ISL_420229 | Virology Department, Sheffield Teaching Hospitals NHS Foundation Trust | Department of Infection, Immunity and Cardiovascular Disease, The Florey Institute, The Medical School, University of Sheffield |
| EPI_ISL_420230 | Virology Department, Sheffield Teaching Hospitals NHS Foundation Trust | Department of Infection, Immunity and Cardiovascular Disease, The Florey Institute, The Medical School, University of Sheffield |
| EPI_ISL_420231 | Virology Department, Sheffield Teaching Hospitals NHS Foundation Trust | Department of Infection, Immunity and Cardiovascular Disease, The Florey Institute, The Medical School, University of Sheffield |
| EPI_ISL_420232 | Virology Department, Sheffield Teaching Hospitals NHS Foundation Trust | Department of Infection, Immunity and Cardiovascular Disease, The Florey Institute, The Medical School, University of Sheffield |
| EPI_ISL_420233 | Virology Department, Sheffield Teaching Hospitals NHS Foundation Trust | Department of Infection, Immunity and Cardiovascular Disease, The Florey Institute, The Medical School, University of Sheffield |

| EPI_Accession_ID | Originating Laboratory | Submitting Laboratory |
|---|---|---|
| EPI_ISL_420234 | Virology Department, Sheffield Teaching Hospitals NHS Foundation Trust | Department of Infection, Immunity and Cardiovascular Disease, The Florey Institute, The Medical School, University of Sheffield |
| EPI_ISL_420235 | Virology Department, Sheffield Teaching Hospitals NHS Foundation Trust | Department of Infection, Immunity and Cardiovascular Disease, The Florey Institute, The Medical School, University of Sheffield |
| EPI_ISL_420236 | Virology Department, Sheffield Teaching Hospitals NHS Foundation Trust | Department of Infection, Immunity and Cardiovascular Disease, The Florey Institute, The Medical School, University of Sheffield |
| EPI_ISL_420237 | Virology Department, Sheffield Teaching Hospitals NHS Foundation Trust | Department of Infection, Immunity and Cardiovascular Disease, The Florey Institute, The Medical School, University of Sheffield |
| EPI_ISL_420238 | Virology Department, Sheffield Teaching Hospitals NHS Foundation Trust | Department of Infection, Immunity and Cardiovascular Disease, The Florey Institute, The Medical School, University of Sheffield |
| EPI_ISL_420239 | Virology Department, Sheffield Teaching Hospitals NHS Foundation Trust | Department of Infection, Immunity and Cardiovascular Disease, The Florey Institute, The Medical School, University of Sheffield |
| EPI_ISL_420240 | Virology Department, Sheffield Teaching Hospitals NHS Foundation Trust | Department of Infection, Immunity and Cardiovascular Disease, The Florey Institute, The Medical School, University of Sheffield |
| EPI_ISL_420241 | Virology Department, Sheffield Teaching Hospitals NHS Foundation Trust | Department of Infection, Immunity and Cardiovascular Disease, The Florey Institute, The Medical School, University of Sheffield |
| EPI_ISL_420242 | Virology Department, Sheffield Teaching Hospitals NHS Foundation Trust | Department of Infection, Immunity and Cardiovascular Disease, The Florey Institute, The Medical School, University of Sheffield |
| EPI_ISL_420243 | Virology Department, Sheffield Teaching Hospitals NHS Foundation Trust | Department of Infection, Immunity and Cardiovascular Disease, The Florey Institute, The Medical School, University of Sheffield |
| EPI_ISL_420244 | Virology Department, Sheffield Teaching Hospitals NHS Foundation Trust | Department of Infection, Immunity and Cardiovascular Disease, The Florey Institute, The Medical School, University of Sheffield |

| EPI_Accession_ID | Originating Laboratory | Submitting Laboratory |
|---|---|---|
| EPI_ISL_420245 | Virology Department, Sheffield Teaching Hospitals NHS Foundation Trust | Department of Infection, Immunity and Cardiovascular Disease, The Florey Institute, The Medical School, University of Sheffield |
| EPI_ISL_420246 | Virology Department, Sheffield Teaching Hospitals NHS Foundation Trust | Department of Infection, Immunity and Cardiovascular Disease, The Florey Institute, The Medical School, University of Sheffield |
| EPI_ISL_420247 | Virology Department, Sheffield Teaching Hospitals NHS Foundation Trust | Department of Infection, Immunity and Cardiovascular Disease, The Florey Institute, The Medical School, University of Sheffield |
| EPI_ISL_420248 | Virology Department, Sheffield Teaching Hospitals NHS Foundation Trust | Department of Infection, Immunity and Cardiovascular Disease, The Florey Institute, The Medical School, University of Sheffield |
| EPI_ISL_420249 | Virology Department, Sheffield Teaching Hospitals NHS Foundation Trust | Department of Infection, Immunity and Cardiovascular Disease, The Florey Institute, The Medical School, University of Sheffield |
| EPI_ISL_420250 | Virology Department, Sheffield Teaching Hospitals NHS Foundation Trust | Department of Infection, Immunity and Cardiovascular Disease, The Florey Institute, The Medical School, University of Sheffield |
| EPI_ISL_420251 | Virology Department, Sheffield Teaching Hospitals NHS Foundation Trust | Department of Infection, Immunity and Cardiovascular Disease, The Florey Institute, The Medical School, University of Sheffield |
| EPI_ISL_420252 | Virology Department, Sheffield Teaching Hospitals NHS Foundation Trust | Department of Infection, Immunity and Cardiovascular Disease, The Florey Institute, The Medical School, University of Sheffield |
| EPI_ISL_420253 | Virology Department, Sheffield Teaching Hospitals NHS Foundation Trust | Department of Infection, Immunity and Cardiovascular Disease, The Florey Institute, The Medical School, University of Sheffield |
| EPI_ISL_420254 | Virology Department, Sheffield Teaching Hospitals NHS Foundation Trust | Department of Infection, Immunity and Cardiovascular Disease, The Florey Institute, The Medical School, University of Sheffield |
| EPI_ISL_420255 | Virology Department, Sheffield Teaching Hospitals NHS Foundation Trust | Department of Infection, Immunity and Cardiovascular Disease, The Florey Institute, The Medical School, University of Sheffield |

| EPI_Accession_ID | Originating Laboratory | Submitting Laboratory |
|---|---|---|
| EPI_ISL_420256 | Virology Department, Sheffield Teaching Hospitals NHS Foundation Trust | Department of Infection, Immunity and Cardiovascular Disease, The Florey Institute, The Medical School, University of Sheffield |
| EPI_ISL_420257 | Virology Department, Sheffield Teaching Hospitals NHS Foundation Trust | Department of Infection, Immunity and Cardiovascular Disease, The Florey Institute, The Medical School, University of Sheffield |
| EPI_ISL_420258 | Virology Department, Sheffield Teaching Hospitals NHS Foundation Trust | Department of Infection, Immunity and Cardiovascular Disease, The Florey Institute, The Medical School, University of Sheffield |
| EPI_ISL_420259 | Virology Department, Sheffield Teaching Hospitals NHS Foundation Trust | Department of Infection, Immunity and Cardiovascular Disease, The Florey Institute, The Medical School, University of Sheffield |
| EPI_ISL_420260 | Virology Department, Sheffield Teaching Hospitals NHS Foundation Trust | Department of Infection, Immunity and Cardiovascular Disease, The Florey Institute, The Medical School, University of Sheffield |
| EPI_ISL_420261 | Virology Department, Sheffield Teaching Hospitals NHS Foundation Trust | Department of Infection, Immunity and Cardiovascular Disease, The Florey Institute, The Medical School, University of Sheffield |
| EPI_ISL_420262 | Virology Department, Sheffield Teaching Hospitals NHS Foundation Trust | Department of Infection, Immunity and Cardiovascular Disease, The Florey Institute, The Medical School, University of Sheffield |
| EPI_ISL_420263 | Virology Department, Sheffield Teaching Hospitals NHS Foundation Trust | Department of Infection, Immunity and Cardiovascular Disease, The Florey Institute, The Medical School, University of Sheffield |
| EPI_ISL_420264 | Virology Department, Sheffield Teaching Hospitals NHS Foundation Trust | Department of Infection, Immunity and Cardiovascular Disease, The Florey Institute, The Medical School, University of Sheffield |
| EPI_ISL_420265 | Virology Department, Sheffield Teaching Hospitals NHS Foundation Trust | Department of Infection, Immunity and Cardiovascular Disease, The Florey Institute, The Medical School, University of Sheffield |
| EPI_ISL_420266 | Virology Department, Sheffield Teaching Hospitals NHS Foundation Trust | Department of Infection, Immunity and Cardiovascular Disease, The Florey Institute, The Medical School, University of Sheffield |

| EPI_Accession_ID | Originating Laboratory | Submitting Laboratory |
|---|---|---|
| EPI_ISL_420267 | Virology Department, Sheffield Teaching Hospitals NHS Foundation Trust | Department of Infection, Immunity and Cardiovascular Disease, The Florey Institute, The Medical School, University of Sheffield |
| EPI_ISL_420268 | Virology Department, Sheffield Teaching Hospitals NHS Foundation Trust | Department of Infection, Immunity and Cardiovascular Disease, The Florey Institute, The Medical School, University of Sheffield |
| EPI_ISL_420269 | Virology Department, Sheffield Teaching Hospitals NHS Foundation Trust | Department of Infection, Immunity and Cardiovascular Disease, The Florey Institute, The Medical School, University of Sheffield |
| EPI_ISL_420270 | Virology Department, Sheffield Teaching Hospitals NHS Foundation Trust | Department of Infection, Immunity and Cardiovascular Disease, The Florey Institute, The Medical School, University of Sheffield |
| EPI_ISL_420271 | Virology Department, Sheffield Teaching Hospitals NHS Foundation Trust | Department of Infection, Immunity and Cardiovascular Disease, The Florey Institute, The Medical School, University of Sheffield |
| EPI_ISL_420272 | Virology Department, Sheffield Teaching Hospitals NHS Foundation Trust | Department of Infection, Immunity and Cardiovascular Disease, The Florey Institute, The Medical School, University of Sheffield |
| EPI_ISL_420273 | Virology Department, Sheffield Teaching Hospitals NHS Foundation Trust | Department of Infection, Immunity and Cardiovascular Disease, The Florey Institute, The Medical School, University of Sheffield |
| EPI_ISL_420274 | Virology Department, Sheffield Teaching Hospitals NHS Foundation Trust | Department of Infection, Immunity and Cardiovascular Disease, The Florey Institute, The Medical School, University of Sheffield |
| EPI_ISL_420275 | Virology Department, Sheffield Teaching Hospitals NHS Foundation Trust | Department of Infection, Immunity and Cardiovascular Disease, The Florey Institute, The Medical School, University of Sheffield |
| EPI_ISL_420276 | Virology Department, Sheffield Teaching Hospitals NHS Foundation Trust | Department of Infection, Immunity and Cardiovascular Disease, The Florey Institute, The Medical School, University of Sheffield |
| EPI_ISL_420277 | Virology Department, Sheffield Teaching Hospitals NHS Foundation Trust | Department of Infection, Immunity and Cardiovascular Disease, The Florey Institute, The Medical School, University of Sheffield |

| EPI_Accession_ID | Originating Laboratory | Submitting Laboratory |
|---|---|---|
| EPI_ISL_420278 | Virology Department, Sheffield Teaching Hospitals NHS Foundation Trust | Department of Infection, Immunity and Cardiovascular Disease, The Florey Institute, The Medical School, University of Sheffield |
| EPI_ISL_420279 | Virology Department, Sheffield Teaching Hospitals NHS Foundation Trust | Department of Infection, Immunity and Cardiovascular Disease, The Florey Institute, The Medical School, University of Sheffield |
| EPI_ISL_420280 | Virology Department, Sheffield Teaching Hospitals NHS Foundation Trust | Department of Infection, Immunity and Cardiovascular Disease, The Florey Institute, The Medical School, University of Sheffield |
| EPI_ISL_420281 | Virology Department, Sheffield Teaching Hospitals NHS Foundation Trust | Department of Infection, Immunity and Cardiovascular Disease, The Florey Institute, The Medical School, University of Sheffield |
| EPI_ISL_420282 | Virology Department, Sheffield Teaching Hospitals NHS Foundation Trust | Department of Infection, Immunity and Cardiovascular Disease, The Florey Institute, The Medical School, University of Sheffield |
| EPI_ISL_420283 | Virology Department, Sheffield Teaching Hospitals NHS Foundation Trust | Department of Infection, Immunity and Cardiovascular Disease, The Florey Institute, The Medical School, University of Sheffield |
| EPI_ISL_420284 | Virology Department, Sheffield Teaching Hospitals NHS Foundation Trust | Department of Infection, Immunity and Cardiovascular Disease, The Florey Institute, The Medical School, University of Sheffield |
| EPI_ISL_420285 | Virology Department, Sheffield Teaching Hospitals NHS Foundation Trust | Department of Infection, Immunity and Cardiovascular Disease, The Florey Institute, The Medical School, University of Sheffield |
| EPI_ISL_420286 | Virology Department, Sheffield Teaching Hospitals NHS Foundation Trust | Department of Infection, Immunity and Cardiovascular Disease, The Florey Institute, The Medical School, University of Sheffield |
| EPI_ISL_420287 | Virology Department, Sheffield Teaching Hospitals NHS Foundation Trust | Department of Infection, Immunity and Cardiovascular Disease, The Florey Institute, The Medical School, University of Sheffield |
| EPI_ISL_420288 | Virology Department, Sheffield Teaching Hospitals NHS Foundation Trust | Department of Infection, Immunity and Cardiovascular Disease, The Florey Institute, The Medical School, University of Sheffield |

| EPI_Accession_ID | Originating Laboratory | Submitting Laboratory |
|---|---|---|
| EPI_ISL_420289 | Virology Department, Sheffield Teaching Hospitals NHS Foundation Trust | Department of Infection, Immunity and Cardiovascular Disease, The Florey Institute, The Medical School, University of Sheffield |
| EPI_ISL_420290 | Virology Department, Sheffield Teaching Hospitals NHS Foundation Trust | Department of Infection, Immunity and Cardiovascular Disease, The Florey Institute, The Medical School, University of Sheffield |
| EPI_ISL_420291 | Virology Department, Sheffield Teaching Hospitals NHS Foundation Trust | Department of Infection, Immunity and Cardiovascular Disease, The Florey Institute, The Medical School, University of Sheffield |
| EPI_ISL_420292 | Virology Department, Sheffield Teaching Hospitals NHS Foundation Trust | Department of Infection, Immunity and Cardiovascular Disease, The Florey Institute, The Medical School, University of Sheffield |
| EPI_ISL_420293 | Wildlife Conservation Society, Bronx Zoo | Diagnostic Virology Laboratory, United States Department of Agriculture, National Veterinary Services Laboratories |
| EPI_ISL_420294 | Institute of Microbiology and Immunology, Faculty of Medicine, University of Ljubljana | Institute of Microbiology and Immunology, Faculty of Medicine, University of Ljubljana |
| EPI_ISL_420295 | Institute of Microbiology and Immunology, Faculty of Medicine, University of Ljubljana | Institute of Microbiology and Immunology, Faculty of Medicine, University of Ljubljana |
| EPI_ISL_420296 | NYU Langone Health | Departments of Pathology and Medicine, New York University School of Medicine |
| EPI_ISL_420297 | NYU Langone Health | Departments of Pathology and Medicine, New York University School of Medicine |
| EPI_ISL_420298 | NYU Langone Health | Departments of Pathology and Medicine, New York University School of Medicine |
| EPI_ISL_420299 | NYU Langone Health | Departments of Pathology and Medicine, New York University School of Medicine |
| EPI_ISL_420300 | NYU Langone Health | Departments of Pathology and Medicine, New York University School of Medicine |
| EPI_ISL_420301 | NYU Langone Health | Departments of Pathology and Medicine, New York University School of Medicine |
| EPI_ISL_420302 | NYU Langone Health | Departments of Pathology and Medicine, New York University School of Medicine |

| EPI_Accession_ID | Originating Laboratory | Submitting Laboratory |
|---|---|---|
| EPI_ISL_420303 | Alaska State Virology Laboratory | Alaska State Virology Laboratory |
| EPI_ISL_420304 | Alaska State Virology Laboratory | Alaska State Virology Laboratory |
| EPI_ISL_420305 | Alaska State Virology Laboratory | Alaska State Virology Laboratory |
| EPI_ISL_420306 | Alaska State Virology Laboratory | Alaska State Virology Laboratory |
| EPI_ISL_420307 | NYU Langone Health | Departments of Pathology and Medicine, New York University School of Medicine |
| EPI_ISL_420308 | NYU Langone Health | Departments of Pathology and Medicine, New York University School of Medicine |
| EPI_ISL_420309 | NYU Langone Health | Departments of Pathology and Medicine, New York University School of Medicine |
| EPI_ISL_420310 | University Hospital of Northern Norway, Department for Microbiology and Infectious Disease Control | Norwegian Institute of Public Health, Department of Virology |
| EPI_ISL_420311 | Akershus University Hospital, Department for Microbiology and Infectious Disease Control | Norwegian Institute of Public Health, Department of Virology |
| EPI_ISL_420312 | Akershus University Hospital, Department for Microbiology and Infectious Disease Control | Norwegian Institute of Public Health, Department of Virology |
| EPI_ISL_420313 | Furst Medical Laboratory | Norwegian Institute of Public Health, Department of Virology |
| EPI_ISL_420314 | KU Leuven, Clinical and Epidemiological Virology | KU Leuven, Clinical and Epidemiological Virology |
| EPI_ISL_420315 | KU Leuven, Clinical and Epidemiological Virology | KU Leuven, Clinical and Epidemiological Virology |
| EPI_ISL_420316 | KU Leuven, Clinical and Epidemiological Virology | KU Leuven, Clinical and Epidemiological Virology |
| EPI_ISL_420317 | KU Leuven, Clinical and Epidemiological Virology | KU Leuven, Clinical and Epidemiological Virology |
| EPI_ISL_420318 | KU Leuven, Clinical and Epidemiological Virology | KU Leuven, Clinical and Epidemiological Virology |
| EPI_ISL_420319 | KU Leuven, Clinical and Epidemiological Virology | KU Leuven, Clinical and Epidemiological Virology |
| EPI_ISL_420320 | KU Leuven, Clinical and Epidemiological Virology | KU Leuven, Clinical and Epidemiological Virology |
| EPI_ISL_420321 | KU Leuven, Clinical and Epidemiological Virology | KU Leuven, Clinical and Epidemiological Virology |
| EPI_ISL_420322 | KU Leuven, Clinical and Epidemiological Virology | KU Leuven, Clinical and Epidemiological Virology |
| EPI_ISL_420323 | KU Leuven, Clinical and Epidemiological Virology | KU Leuven, Clinical and Epidemiological Virology |
| EPI_ISL_420324 | KU Leuven, Clinical and Epidemiological Virology | KU Leuven, Clinical and Epidemiological Virology |
| EPI_ISL_420325 | KU Leuven, Clinical and Epidemiological Virology | KU Leuven, Clinical and Epidemiological Virology |
| EPI_ISL_420326 | KU Leuven, Clinical and Epidemiological Virology | KU Leuven, Clinical and Epidemiological Virology |
| EPI_ISL_420327 | KU Leuven, Clinical and Epidemiological Virology | KU Leuven, Clinical and Epidemiological Virology |
| EPI_ISL_420328 | KU Leuven, Clinical and Epidemiological Virology | KU Leuven, Clinical and Epidemiological Virology |

| EPI_Accession_ID | Originating Laboratory | Submitting Laboratory |
|---|---|---|
| EPI_ISL_420329 | KU Leuven, Clinical and Epidemiological Virology | KU Leuven, Clinical and Epidemiological Virology |
| EPI_ISL_420330 | KU Leuven, Clinical and Epidemiological Virology | KU Leuven, Clinical and Epidemiological Virology |
| EPI_ISL_420331 | KU Leuven, Clinical and Epidemiological Virology | KU Leuven, Clinical and Epidemiological Virology |
| EPI_ISL_420332 | KU Leuven, Clinical and Epidemiological Virology | KU Leuven, Clinical and Epidemiological Virology |
| EPI_ISL_420333 | KU Leuven, Clinical and Epidemiological Virology | KU Leuven, Clinical and Epidemiological Virology |
| EPI_ISL_420334 | KU Leuven, Clinical and Epidemiological Virology | KU Leuven, Clinical and Epidemiological Virology |
| EPI_ISL_420335 | KU Leuven, Clinical and Epidemiological Virology | KU Leuven, Clinical and Epidemiological Virology |
| EPI_ISL_420336 | KU Leuven, Clinical and Epidemiological Virology | KU Leuven, Clinical and Epidemiological Virology |
| EPI_ISL_420337 | KU Leuven, Clinical and Epidemiological Virology | KU Leuven, Clinical and Epidemiological Virology |
| EPI_ISL_420338 | KU Leuven, Clinical and Epidemiological Virology | KU Leuven, Clinical and Epidemiological Virology |
| EPI_ISL_420339 | KU Leuven, Clinical and Epidemiological Virology | KU Leuven, Clinical and Epidemiological Virology |
| EPI_ISL_420340 | KU Leuven, Clinical and Epidemiological Virology | KU Leuven, Clinical and Epidemiological Virology |
| EPI_ISL_420341 | KU Leuven, Clinical and Epidemiological Virology | KU Leuven, Clinical and Epidemiological Virology |
| EPI_ISL_420342 | KU Leuven, Clinical and Epidemiological Virology | KU Leuven, Clinical and Epidemiological Virology |
| EPI_ISL_420343 | KU Leuven, Clinical and Epidemiological Virology | KU Leuven, Clinical and Epidemiological Virology |
| EPI_ISL_420344 | KU Leuven, Clinical and Epidemiological Virology | KU Leuven, Clinical and Epidemiological Virology |
| EPI_ISL_420345 | KU Leuven, Clinical and Epidemiological Virology | KU Leuven, Clinical and Epidemiological Virology |
| EPI_ISL_420346 | KU Leuven, Clinical and Epidemiological Virology | KU Leuven, Clinical and Epidemiological Virology |
| EPI_ISL_420347 | KU Leuven, Clinical and Epidemiological Virology | KU Leuven, Clinical and Epidemiological Virology |
| EPI_ISL_420348 | KU Leuven, Clinical and Epidemiological Virology | KU Leuven, Clinical and Epidemiological Virology |
| EPI_ISL_420349 | KU Leuven, Clinical and Epidemiological Virology | KU Leuven, Clinical and Epidemiological Virology |
| EPI_ISL_420350 | KU Leuven, Clinical and Epidemiological Virology | KU Leuven, Clinical and Epidemiological Virology |
| EPI_ISL_420351 | KU Leuven, Clinical and Epidemiological Virology | KU Leuven, Clinical and Epidemiological Virology |
| EPI_ISL_420352 | KU Leuven, Clinical and Epidemiological Virology | KU Leuven, Clinical and Epidemiological Virology |
| EPI_ISL_420353 | KU Leuven, Clinical and Epidemiological Virology | KU Leuven, Clinical and Epidemiological Virology |
| EPI_ISL_420354 | KU Leuven, Clinical and Epidemiological Virology | KU Leuven, Clinical and Epidemiological Virology |
| EPI_ISL_420355 | KU Leuven, Clinical and Epidemiological Virology | KU Leuven, Clinical and Epidemiological Virology |
| EPI_ISL_420356 | KU Leuven, Clinical and Epidemiological Virology | KU Leuven, Clinical and Epidemiological Virology |
| EPI_ISL_420357 | KU Leuven, Clinical and Epidemiological Virology | KU Leuven, Clinical and Epidemiological Virology |
| EPI_ISL_420358 | KU Leuven, Clinical and Epidemiological Virology | KU Leuven, Clinical and Epidemiological Virology |
| EPI_ISL_420359 | KU Leuven, Clinical and Epidemiological Virology | KU Leuven, Clinical and Epidemiological Virology |
| EPI_ISL_420360 | KU Leuven, Clinical and Epidemiological Virology | KU Leuven, Clinical and Epidemiological Virology |
| EPI_ISL_420361 | KU Leuven, Clinical and Epidemiological Virology | KU Leuven, Clinical and Epidemiological Virology |

| EPI_Accession_ID | Originating Laboratory | Submitting Laboratory |
|---|---|---|
| EPI_ISL_420362 | KU Leuven, Clinical and Epidemiological Virology | KU Leuven, Clinical and Epidemiological Virology |
| EPI_ISL_420363 | KU Leuven, Clinical and Epidemiological Virology | KU Leuven, Clinical and Epidemiological Virology |
| EPI_ISL_420364 | KU Leuven, Clinical and Epidemiological Virology | KU Leuven, Clinical and Epidemiological Virology |
| EPI_ISL_420365 | KU Leuven, Clinical and Epidemiological Virology | KU Leuven, Clinical and Epidemiological Virology |
| EPI_ISL_420366 | KU Leuven, Clinical and Epidemiological Virology | KU Leuven, Clinical and Epidemiological Virology |
| EPI_ISL_420367 | KU Leuven, Clinical and Epidemiological Virology | KU Leuven, Clinical and Epidemiological Virology |
| EPI_ISL_420368 | KU Leuven, Clinical and Epidemiological Virology | KU Leuven, Clinical and Epidemiological Virology |
| EPI_ISL_420369 | KU Leuven, Clinical and Epidemiological Virology | KU Leuven, Clinical and Epidemiological Virology |
| EPI_ISL_420370 | KU Leuven, Clinical and Epidemiological Virology | KU Leuven, Clinical and Epidemiological Virology |
| EPI_ISL_420371 | KU Leuven, Clinical and Epidemiological Virology | KU Leuven, Clinical and Epidemiological Virology |
| EPI_ISL_420372 | KU Leuven, Clinical and Epidemiological Virology | KU Leuven, Clinical and Epidemiological Virology |
| EPI_ISL_420373 | KU Leuven, Clinical and Epidemiological Virology | KU Leuven, Clinical and Epidemiological Virology |
| EPI_ISL_420374 | KU Leuven, Clinical and Epidemiological Virology | KU Leuven, Clinical and Epidemiological Virology |
| EPI_ISL_420375 | KU Leuven, Clinical and Epidemiological Virology | KU Leuven, Clinical and Epidemiological Virology |
| EPI_ISL_420376 | KU Leuven, Clinical and Epidemiological Virology | KU Leuven, Clinical and Epidemiological Virology |
| EPI_ISL_420377 | KU Leuven, Clinical and Epidemiological Virology | KU Leuven, Clinical and Epidemiological Virology |
| EPI_ISL_420378 | KU Leuven, Clinical and Epidemiological Virology | KU Leuven, Clinical and Epidemiological Virology |
| EPI_ISL_420379 | KU Leuven, Clinical and Epidemiological Virology | KU Leuven, Clinical and Epidemiological Virology |
| EPI_ISL_420380 | KU Leuven, Clinical and Epidemiological Virology | KU Leuven, Clinical and Epidemiological Virology |
| EPI_ISL_420381 | KU Leuven, Clinical and Epidemiological Virology | KU Leuven, Clinical and Epidemiological Virology |
| EPI_ISL_420382 | KU Leuven, Clinical and Epidemiological Virology | KU Leuven, Clinical and Epidemiological Virology |
| EPI_ISL_420383 | KU Leuven, Clinical and Epidemiological Virology | KU Leuven, Clinical and Epidemiological Virology |
| EPI_ISL_420384 | KU Leuven, Clinical and Epidemiological Virology | KU Leuven, Clinical and Epidemiological Virology |
| EPI_ISL_420385 | KU Leuven, Clinical and Epidemiological Virology | KU Leuven, Clinical and Epidemiological Virology |
| EPI_ISL_420386 | KU Leuven, Clinical and Epidemiological Virology | KU Leuven, Clinical and Epidemiological Virology |
| EPI_ISL_420387 | KU Leuven, Clinical and Epidemiological Virology | KU Leuven, Clinical and Epidemiological Virology |
| EPI_ISL_420388 | KU Leuven, Clinical and Epidemiological Virology | KU Leuven, Clinical and Epidemiological Virology |
| EPI_ISL_420389 | KU Leuven, Clinical and Epidemiological Virology | KU Leuven, Clinical and Epidemiological Virology |
| EPI_ISL_420390 | KU Leuven, Clinical and Epidemiological Virology | KU Leuven, Clinical and Epidemiological Virology |
| EPI_ISL_420391 | KU Leuven, Clinical and Epidemiological Virology | KU Leuven, Clinical and Epidemiological Virology |
| EPI_ISL_420392 | KU Leuven, Clinical and Epidemiological Virology | KU Leuven, Clinical and Epidemiological Virology |
| EPI_ISL_420393 | KU Leuven, Clinical and Epidemiological Virology | KU Leuven, Clinical and Epidemiological Virology |
| EPI_ISL_420394 | KU Leuven, Clinical and Epidemiological Virology | KU Leuven, Clinical and Epidemiological Virology |

| EPI_Accession_ID | Originating Laboratory | Submitting Laboratory |
|---|---|---|
| EPI_ISL_420395 | KU Leuven, Clinical and Epidemiological Virology | KU Leuven, Clinical and Epidemiological Virology |
| EPI_ISL_420396 | KU Leuven, Clinical and Epidemiological Virology | KU Leuven, Clinical and Epidemiological Virology |
| EPI_ISL_420397 | KU Leuven, Clinical and Epidemiological Virology | KU Leuven, Clinical and Epidemiological Virology |
| EPI_ISL_420398 | KU Leuven, Clinical and Epidemiological Virology | KU Leuven, Clinical and Epidemiological Virology |
| EPI_ISL_420399 | KU Leuven, Clinical and Epidemiological Virology | KU Leuven, Clinical and Epidemiological Virology |
| EPI_ISL_420400 | KU Leuven, Clinical and Epidemiological Virology | KU Leuven, Clinical and Epidemiological Virology |
| EPI_ISL_420401 | KU Leuven, Clinical and Epidemiological Virology | KU Leuven, Clinical and Epidemiological Virology |
| EPI_ISL_420402 | KU Leuven, Clinical and Epidemiological Virology | KU Leuven, Clinical and Epidemiological Virology |
| EPI_ISL_420403 | KU Leuven, Clinical and Epidemiological Virology | KU Leuven, Clinical and Epidemiological Virology |
| EPI_ISL_420404 | KU Leuven, Clinical and Epidemiological Virology | KU Leuven, Clinical and Epidemiological Virology |
| EPI_ISL_420405 | KU Leuven, Clinical and Epidemiological Virology | KU Leuven, Clinical and Epidemiological Virology |
| EPI_ISL_420406 | KU Leuven, Clinical and Epidemiological Virology | KU Leuven, Clinical and Epidemiological Virology |
| EPI_ISL_420407 | KU Leuven, Clinical and Epidemiological Virology | KU Leuven, Clinical and Epidemiological Virology |
| EPI_ISL_420408 | KU Leuven, Clinical and Epidemiological Virology | KU Leuven, Clinical and Epidemiological Virology |
| EPI_ISL_420409 | KU Leuven, Clinical and Epidemiological Virology | KU Leuven, Clinical and Epidemiological Virology |
| EPI_ISL_420410 | KU Leuven, Clinical and Epidemiological Virology | KU Leuven, Clinical and Epidemiological Virology |
| EPI_ISL_420411 | KU Leuven, Clinical and Epidemiological Virology | KU Leuven, Clinical and Epidemiological Virology |
| EPI_ISL_420412 | KU Leuven, Clinical and Epidemiological Virology | KU Leuven, Clinical and Epidemiological Virology |
| EPI_ISL_420413 | KU Leuven, Clinical and Epidemiological Virology | KU Leuven, Clinical and Epidemiological Virology |
| EPI_ISL_420414 | KU Leuven, Clinical and Epidemiological Virology | KU Leuven, Clinical and Epidemiological Virology |
| EPI_ISL_420415 | KU Leuven, Clinical and Epidemiological Virology | KU Leuven, Clinical and Epidemiological Virology |
| EPI_ISL_420416 | KU Leuven, Clinical and Epidemiological Virology | KU Leuven, Clinical and Epidemiological Virology |
| EPI_ISL_420417 | KU Leuven, Clinical and Epidemiological Virology | KU Leuven, Clinical and Epidemiological Virology |
| EPI_ISL_420418 | KU Leuven, Clinical and Epidemiological Virology | KU Leuven, Clinical and Epidemiological Virology |
| EPI_ISL_420419 | KU Leuven, Clinical and Epidemiological Virology | KU Leuven, Clinical and Epidemiological Virology |
| EPI_ISL_420420 | KU Leuven, Clinical and Epidemiological Virology | KU Leuven, Clinical and Epidemiological Virology |
| EPI_ISL_420421 | KU Leuven, Clinical and Epidemiological Virology | KU Leuven, Clinical and Epidemiological Virology |
| EPI_ISL_420422 | KU Leuven, Clinical and Epidemiological Virology | KU Leuven, Clinical and Epidemiological Virology |
| EPI_ISL_420423 | KU Leuven, Clinical and Epidemiological Virology | KU Leuven, Clinical and Epidemiological Virology |
| EPI_ISL_420424 | KU Leuven, Clinical and Epidemiological Virology | KU Leuven, Clinical and Epidemiological Virology |
| EPI_ISL_420425 | KU Leuven, Clinical and Epidemiological Virology | KU Leuven, Clinical and Epidemiological Virology |
| EPI_ISL_420426 | KU Leuven, Clinical and Epidemiological Virology | KU Leuven, Clinical and Epidemiological Virology |
| EPI_ISL_420427 | KU Leuven, Clinical and Epidemiological Virology | KU Leuven, Clinical and Epidemiological Virology |

| EPI_Accession_ID | Originating Laboratory | Submitting Laboratory |
|---|---|---|
| EPI_ISL_420428 | KU Leuven, Clinical and Epidemiological Virology | KU Leuven, Clinical and Epidemiological Virology |
| EPI_ISL_420429 | KU Leuven, Clinical and Epidemiological Virology | KU Leuven, Clinical and Epidemiological Virology |
| EPI_ISL_420430 | KU Leuven, Clinical and Epidemiological Virology | KU Leuven, Clinical and Epidemiological Virology |
| EPI_ISL_420431 | KU Leuven, Clinical and Epidemiological Virology | KU Leuven, Clinical and Epidemiological Virology |
| EPI_ISL_420432 | KU Leuven, Clinical and Epidemiological Virology | KU Leuven, Clinical and Epidemiological Virology |
| EPI_ISL_420433 | KU Leuven, Clinical and Epidemiological Virology | KU Leuven, Clinical and Epidemiological Virology |
| EPI_ISL_420434 | KU Leuven, Clinical and Epidemiological Virology | KU Leuven, Clinical and Epidemiological Virology |
| EPI_ISL_420435 | KU Leuven, Clinical and Epidemiological Virology | KU Leuven, Clinical and Epidemiological Virology |
| EPI_ISL_420436 | KU Leuven, Clinical and Epidemiological Virology | KU Leuven, Clinical and Epidemiological Virology |
| EPI_ISL_420437 | KU Leuven, Clinical and Epidemiological Virology | KU Leuven, Clinical and Epidemiological Virology |
| EPI_ISL_420438 | KU Leuven, Clinical and Epidemiological Virology | KU Leuven, Clinical and Epidemiological Virology |
| EPI_ISL_420439 | KU Leuven, Clinical and Epidemiological Virology | KU Leuven, Clinical and Epidemiological Virology |
| EPI_ISL_420440 | KU Leuven, Clinical and Epidemiological Virology | KU Leuven, Clinical and Epidemiological Virology |
| EPI_ISL_420441 | KU Leuven, Clinical and Epidemiological Virology | KU Leuven, Clinical and Epidemiological Virology |
| EPI_ISL_420442 | KU Leuven, Clinical and Epidemiological Virology | KU Leuven, Clinical and Epidemiological Virology |
| EPI_ISL_420443 | KU Leuven, Clinical and Epidemiological Virology | KU Leuven, Clinical and Epidemiological Virology |
| EPI_ISL_420444 | KU Leuven, Clinical and Epidemiological Virology | KU Leuven, Clinical and Epidemiological Virology |
| EPI_ISL_420445 | KU Leuven, Clinical and Epidemiological Virology | KU Leuven, Clinical and Epidemiological Virology |
| EPI_ISL_420446 | KU Leuven, Clinical and Epidemiological Virology | KU Leuven, Clinical and Epidemiological Virology |
| EPI_ISL_420447 | KU Leuven, Clinical and Epidemiological Virology | KU Leuven, Clinical and Epidemiological Virology |
| EPI_ISL_420448 | KU Leuven, Clinical and Epidemiological Virology | KU Leuven, Clinical and Epidemiological Virology |
| EPI_ISL_420449 | KU Leuven, Clinical and Epidemiological Virology | KU Leuven, Clinical and Epidemiological Virology |
| EPI_ISL_420450 | KU Leuven, Clinical and Epidemiological Virology | KU Leuven, Clinical and Epidemiological Virology |
| EPI_ISL_420451 | KU Leuven, Clinical and Epidemiological Virology | KU Leuven, Clinical and Epidemiological Virology |
| EPI_ISL_420452 | KU Leuven, Clinical and Epidemiological Virology | KU Leuven, Clinical and Epidemiological Virology |
| EPI_ISL_420453 | KU Leuven, Clinical and Epidemiological Virology | KU Leuven, Clinical and Epidemiological Virology |
| EPI_ISL_420454 | KU Leuven, Clinical and Epidemiological Virology | KU Leuven, Clinical and Epidemiological Virology |
| EPI_ISL_420455 | Department of Clinical Pathology, Pamela Youde Nethersole Eastern Hospital | Department of Health Technology and Informatics, Faculty of Health and Social Science, The Hong Kong Polytechnic University |
| EPI_ISL_420456 | PathWest Laboratory Medicine WA | PathWest Laboratory Medicine WA |
| EPI_ISL_420457 | Respiratory Virus Unit, Microbiology Services Colindale, Public Health England | Respiratory Virus Unit, Microbiology Services Colindale, Public Health England |

| EPI_Accession_ID | Originating Laboratory | Submitting Laboratory |
|---|---|---|
| EPI_ISL_420458 | Respiratory Virus Unit, Microbiology Services Colindale, Public Health England | Respiratory Virus Unit, Microbiology Services Colindale, Public Health England |
| EPI_ISL_420459 | Respiratory Virus Unit, Microbiology Services Colindale, Public Health England | Respiratory Virus Unit, Microbiology Services Colindale, Public Health England |
| EPI_ISL_420460 | Respiratory Virus Unit, Microbiology Services Colindale, Public Health England | Respiratory Virus Unit, Microbiology Services Colindale, Public Health England |
| EPI_ISL_420461 | Respiratory Virus Unit, Microbiology Services Colindale, Public Health England | Respiratory Virus Unit, Microbiology Services Colindale, Public Health England |
| EPI_ISL_420462 | Respiratory Virus Unit, Microbiology Services Colindale, Public Health England | Respiratory Virus Unit, Microbiology Services Colindale, Public Health England |
| EPI_ISL_420463 | Respiratory Virus Unit, Microbiology Services Colindale, Public Health England | Respiratory Virus Unit, Microbiology Services Colindale, Public Health England |
| EPI_ISL_420464 | Respiratory Virus Unit, Microbiology Services Colindale, Public Health England | Respiratory Virus Unit, Microbiology Services Colindale, Public Health England |
| EPI_ISL_420465 | Respiratory Virus Unit, Microbiology Services Colindale, Public Health England | Respiratory Virus Unit, Microbiology Services Colindale, Public Health England |
| EPI_ISL_420466 | Respiratory Virus Unit, Microbiology Services Colindale, Public Health England | Respiratory Virus Unit, Microbiology Services Colindale, Public Health England |
| EPI_ISL_420467 | Respiratory Virus Unit, Microbiology Services Colindale, Public Health England | Respiratory Virus Unit, Microbiology Services Colindale, Public Health England |
| EPI_ISL_420468 | Respiratory Virus Unit, Microbiology Services Colindale, Public Health England | Respiratory Virus Unit, Microbiology Services Colindale, Public Health England |
| EPI_ISL_420469 | Respiratory Virus Unit, Microbiology Services Colindale, Public Health England | Respiratory Virus Unit, Microbiology Services Colindale, Public Health England |
| EPI_ISL_420470 | Respiratory Virus Unit, Microbiology Services Colindale, Public Health England | Respiratory Virus Unit, Microbiology Services Colindale, Public Health England |
| EPI_ISL_420471 | Respiratory Virus Unit, Microbiology Services Colindale, Public Health England | Respiratory Virus Unit, Microbiology Services Colindale, Public Health England |
| EPI_ISL_420472 | Respiratory Virus Unit, Microbiology Services Colindale, Public Health England | Respiratory Virus Unit, Microbiology Services Colindale, Public Health England |
| EPI_ISL_420473 | Respiratory Virus Unit, Microbiology Services Colindale, Public Health England | Respiratory Virus Unit, Microbiology Services Colindale, Public Health England |

| EPI_Accession_ID | Originating Laboratory | Submitting Laboratory |
|---|---|---|
| EPI_ISL_420474 | Respiratory Virus Unit, Microbiology Services Colindale, Public Health England | Respiratory Virus Unit, Microbiology Services Colindale, Public Health England |
| EPI_ISL_420475 | Respiratory Virus Unit, Microbiology Services Colindale, Public Health England | Respiratory Virus Unit, Microbiology Services Colindale, Public Health England |
| EPI_ISL_420476 | Respiratory Virus Unit, Microbiology Services Colindale, Public Health England | Respiratory Virus Unit, Microbiology Services Colindale, Public Health England |
| EPI_ISL_420477 | Respiratory Virus Unit, Microbiology Services Colindale, Public Health England | Respiratory Virus Unit, Microbiology Services Colindale, Public Health England |
| EPI_ISL_420478 | Respiratory Virus Unit, Microbiology Services Colindale, Public Health England | Respiratory Virus Unit, Microbiology Services Colindale, Public Health England |
| EPI_ISL_420479 | Respiratory Virus Unit, Microbiology Services Colindale, Public Health England | Respiratory Virus Unit, Microbiology Services Colindale, Public Health England |
| EPI_ISL_420480 | Respiratory Virus Unit, Microbiology Services Colindale, Public Health England | Respiratory Virus Unit, Microbiology Services Colindale, Public Health England |
| EPI_ISL_420481 | Respiratory Virus Unit, Microbiology Services Colindale, Public Health England | Respiratory Virus Unit, Microbiology Services Colindale, Public Health England |
| EPI_ISL_420482 | Respiratory Virus Unit, Microbiology Services Colindale, Public Health England | Respiratory Virus Unit, Microbiology Services Colindale, Public Health England |
| EPI_ISL_420483 | Respiratory Virus Unit, Microbiology Services Colindale, Public Health England | Respiratory Virus Unit, Microbiology Services Colindale, Public Health England |
| EPI_ISL_420484 | Respiratory Virus Unit, Microbiology Services Colindale, Public Health England | Respiratory Virus Unit, Microbiology Services Colindale, Public Health England |
| EPI_ISL_420485 | Respiratory Virus Unit, Microbiology Services Colindale, Public Health England | Respiratory Virus Unit, Microbiology Services Colindale, Public Health England |
| EPI_ISL_420486 | Respiratory Virus Unit, Microbiology Services Colindale, Public Health England | Respiratory Virus Unit, Microbiology Services Colindale, Public Health England |
| EPI_ISL_420487 | Respiratory Virus Unit, Microbiology Services Colindale, Public Health England | Respiratory Virus Unit, Microbiology Services Colindale, Public Health England |
| EPI_ISL_420488 | Respiratory Virus Unit, Microbiology Services Colindale, Public Health England | Respiratory Virus Unit, Microbiology Services Colindale, Public Health England |
| EPI_ISL_420489 | Respiratory Virus Unit, Microbiology Services Colindale, Public Health England | Respiratory Virus Unit, Microbiology Services Colindale, Public Health England |

| EPI_Accession_ID | Originating Laboratory | Submitting Laboratory |
|---|---|---|
| EPI_ISL_420490 | Respiratory Virus Unit, Microbiology Services Colindale, Public Health England | Respiratory Virus Unit, Microbiology Services Colindale, Public Health England |
| EPI_ISL_420491 | Respiratory Virus Unit, Microbiology Services Colindale, Public Health England | Respiratory Virus Unit, Microbiology Services Colindale, Public Health England |
| EPI_ISL_420492 | Respiratory Virus Unit, Microbiology Services Colindale, Public Health England | Respiratory Virus Unit, Microbiology Services Colindale, Public Health England |
| EPI_ISL_420493 | Respiratory Virus Unit, Microbiology Services Colindale, Public Health England | Respiratory Virus Unit, Microbiology Services Colindale, Public Health England |
| EPI_ISL_420494 | Respiratory Virus Unit, Microbiology Services Colindale, Public Health England | Respiratory Virus Unit, Microbiology Services Colindale, Public Health England |
| EPI_ISL_420495 | Respiratory Virus Unit, Microbiology Services Colindale, Public Health England | Respiratory Virus Unit, Microbiology Services Colindale, Public Health England |
| EPI_ISL_420496 | Respiratory Virus Unit, Microbiology Services Colindale, Public Health England | Respiratory Virus Unit, Microbiology Services Colindale, Public Health England |
| EPI_ISL_420497 | Respiratory Virus Unit, Microbiology Services Colindale, Public Health England | Respiratory Virus Unit, Microbiology Services Colindale, Public Health England |
| EPI_ISL_420498 | Respiratory Virus Unit, Microbiology Services Colindale, Public Health England | Respiratory Virus Unit, Microbiology Services Colindale, Public Health England |
| EPI_ISL_420499 | Respiratory Virus Unit, Microbiology Services Colindale, Public Health England | Respiratory Virus Unit, Microbiology Services Colindale, Public Health England |
| EPI_ISL_420500 | Respiratory Virus Unit, Microbiology Services Colindale, Public Health England | Respiratory Virus Unit, Microbiology Services Colindale, Public Health England |
| EPI_ISL_420501 | Respiratory Virus Unit, Microbiology Services Colindale, Public Health England | Respiratory Virus Unit, Microbiology Services Colindale, Public Health England |
| EPI_ISL_420502 | Respiratory Virus Unit, Microbiology Services Colindale, Public Health England | Respiratory Virus Unit, Microbiology Services Colindale, Public Health England |
| EPI_ISL_420503 | Respiratory Virus Unit, Microbiology Services Colindale, Public Health England | Respiratory Virus Unit, Microbiology Services Colindale, Public Health England |
| EPI_ISL_420504 | Respiratory Virus Unit, Microbiology Services Colindale, Public Health England | Respiratory Virus Unit, Microbiology Services Colindale, Public Health England |
| EPI_ISL_420505 | Respiratory Virus Unit, Microbiology Services Colindale, Public Health England | Respiratory Virus Unit, Microbiology Services Colindale, Public Health England |

| EPI_Accession_ID | Originating Laboratory | Submitting Laboratory |
|---|---|---|
| EPI_ISL_420506 | Respiratory Virus Unit, Microbiology Services Colindale, Public Health England | Respiratory Virus Unit, Microbiology Services Colindale, Public Health England |
| EPI_ISL_420507 | Respiratory Virus Unit, Microbiology Services Colindale, Public Health England | Respiratory Virus Unit, Microbiology Services Colindale, Public Health England |
| EPI_ISL_420508 | Respiratory Virus Unit, Microbiology Services Colindale, Public Health England | Respiratory Virus Unit, Microbiology Services Colindale, Public Health England |
| EPI_ISL_420509 | Respiratory Virus Unit, Microbiology Services Colindale, Public Health England | Respiratory Virus Unit, Microbiology Services Colindale, Public Health England |
| EPI_ISL_420510 | Respiratory Virus Unit, Microbiology Services Colindale, Public Health England | Respiratory Virus Unit, Microbiology Services Colindale, Public Health England |
| EPI_ISL_420511 | Respiratory Virus Unit, Microbiology Services Colindale, Public Health England | Respiratory Virus Unit, Microbiology Services Colindale, Public Health England |
| EPI_ISL_420512 | Respiratory Virus Unit, Microbiology Services Colindale, Public Health England | Respiratory Virus Unit, Microbiology Services Colindale, Public Health England |
| EPI_ISL_420513 | Respiratory Virus Unit, Microbiology Services Colindale, Public Health England | Respiratory Virus Unit, Microbiology Services Colindale, Public Health England |
| EPI_ISL_420514 | Respiratory Virus Unit, Microbiology Services Colindale, Public Health England | Respiratory Virus Unit, Microbiology Services Colindale, Public Health England |
| EPI_ISL_420515 | Respiratory Virus Unit, Microbiology Services Colindale, Public Health England | Respiratory Virus Unit, Microbiology Services Colindale, Public Health England |
| EPI_ISL_420516 | Respiratory Virus Unit, Microbiology Services Colindale, Public Health England | Respiratory Virus Unit, Microbiology Services Colindale, Public Health England |
| EPI_ISL_420517 | Respiratory Virus Unit, Microbiology Services Colindale, Public Health England | Respiratory Virus Unit, Microbiology Services Colindale, Public Health England |
| EPI_ISL_420518 | Respiratory Virus Unit, Microbiology Services Colindale, Public Health England | Respiratory Virus Unit, Microbiology Services Colindale, Public Health England |
| EPI_ISL_420519 | Respiratory Virus Unit, Microbiology Services Colindale, Public Health England | Respiratory Virus Unit, Microbiology Services Colindale, Public Health England |
| EPI_ISL_420520 | Respiratory Virus Unit, Microbiology Services Colindale, Public Health England | Respiratory Virus Unit, Microbiology Services Colindale, Public Health England |
| EPI_ISL_420521 | Respiratory Virus Unit, Microbiology Services Colindale, Public Health England | Respiratory Virus Unit, Microbiology Services Colindale, Public Health England |

| EPI_Accession_ID | Originating Laboratory | Submitting Laboratory |
|---|---|---|
| EPI_ISL_420522 | Respiratory Virus Unit, Microbiology Services Colindale, Public Health England | Respiratory Virus Unit, Microbiology Services Colindale, Public Health England |
| EPI_ISL_420523 | Respiratory Virus Unit, Microbiology Services Colindale, Public Health England | Respiratory Virus Unit, Microbiology Services Colindale, Public Health England |
| EPI_ISL_420524 | Respiratory Virus Unit, Microbiology Services Colindale, Public Health England | Respiratory Virus Unit, Microbiology Services Colindale, Public Health England |
| EPI_ISL_420525 | Respiratory Virus Unit, Microbiology Services Colindale, Public Health England | Respiratory Virus Unit, Microbiology Services Colindale, Public Health England |
| EPI_ISL_420526 | Respiratory Virus Unit, Microbiology Services Colindale, Public Health England | Respiratory Virus Unit, Microbiology Services Colindale, Public Health England |
| EPI_ISL_420527 | Respiratory Virus Unit, Microbiology Services Colindale, Public Health England | Respiratory Virus Unit, Microbiology Services Colindale, Public Health England |
| EPI_ISL_420528 | Respiratory Virus Unit, Microbiology Services Colindale, Public Health England | Respiratory Virus Unit, Microbiology Services Colindale, Public Health England |
| EPI_ISL_420529 | Respiratory Virus Unit, Microbiology Services Colindale, Public Health England | Respiratory Virus Unit, Microbiology Services Colindale, Public Health England |
| EPI_ISL_420530 | Respiratory Virus Unit, Microbiology Services Colindale, Public Health England | Respiratory Virus Unit, Microbiology Services Colindale, Public Health England |
| EPI_ISL_420531 | Department of Microbiology, PathWest QEII Medical Centre | Department of Microbiology, PathWest QEII Medical Centre |
| EPI_ISL_420532 | Department of Microbiology, PathWest QEII Medical Centre | Department of Microbiology, PathWest QEII Medical Centre |
| EPI_ISL_420533 | Department of Microbiology, PathWest QEII Medical Centre | Department of Microbiology, PathWest QEII Medical Centre |
| EPI_ISL_420534 | Department of Microbiology, PathWest QEII Medical Centre | Department of Microbiology, PathWest QEII Medical Centre |
| EPI_ISL_420536 | Department of Microbiology, PathWest QEII Medical Centre | Department of Microbiology, PathWest QEII Medical Centre |
| EPI_ISL_420537 | Department of Microbiology, PathWest QEII Medical Centre | Department of Microbiology, PathWest QEII Medical Centre |
| EPI_ISL_420538 | Department of Microbiology, PathWest QEII Medical Centre | Department of Microbiology, PathWest QEII Medical Centre |

| EPI_Accession_ID | Originating Laboratory | Submitting Laboratory |
|---|---|---|
| EPI_ISL_420539 | Department of Microbiology, PathWest QEII Medical Centre | Department of Microbiology, PathWest QEII Medical Centre |
| EPI_ISL_420541 | Institute of Microbiology and Immunology, Faculty of Medicine, University of Ljubljana | Institute of Microbiology and Immunology, Faculty of Medicine, University of Ljubljana |
| EPI_ISL_420542 | Respiratory Virus Unit, Microbiology Services Colindale, Public Health England | Respiratory Virus Unit, Microbiology Services Colindale, Public Health England |
| EPI_ISL_420543 | National Influenza Center, Indian Council of Medical Research - National Institute of Virology | Indian Council of Medical Research-National Institute of Virology, Microbial Containment Complex |
| EPI_ISL_420545 | National Influenza Center, Indian Council of Medical Research - National Institute of Virology | Indian Council of Medical Research-National Institute of Virology, Microbial Containment Complex |
| EPI_ISL_420547 | National Influenza Center, Indian Council of Medical Research - National Institute of Virology | Indian Council of Medical Research-National Institute of Virology, Microbial Containment Complex |
| EPI_ISL_420549 | National Influenza Center, Indian Council of Medical Research - National Institute of Virology | Indian Council of Medical Research-National Institute of Virology, Microbial Containment Complex |
| EPI_ISL_420551 | National Influenza Center, Indian Council of Medical Research - National Institute of Virology | Indian Council of Medical Research-National Institute of Virology, Microbial Containment Complex |
| EPI_ISL_420553 | National Influenza Center, Indian Council of Medical Research - National Institute of Virology | Indian Council of Medical Research-National Institute of Virology, Microbial Containment Complex |
| EPI_ISL_420555 | National Influenza Center, Indian Council of Medical Research - National Institute of Virology | Indian Council of Medical Research-National Institute of Virology, Microbial Containment Complex |
| EPI_ISL_420563 | Ospedale Civile Giuseppe Mazzini | Istituto Zooprofilattico Sperimentale dell'Abruzzo e Molise "G. Caporale" |
| EPI_ISL_420564 | Ospedale Civile Castel Di Sangro | Istituto Zooprofilattico Sperimentale dellâ€™Abruzzo e Molise â€œG. Caporaleâ€� |
| EPI_ISL_420565 | Ospedale Civile Giuseppe Mazzini | Istituto Zooprofilattico Sperimentale dellâ€™Abruzzo e Molise â€œG. Caporaleâ€� |
| EPI_ISL_420566 | Ospedale Regionale San Salvatore | Istituto Zooprofilattico Sperimentale dellâ€™Abruzzo e Molise â€œG. Caporaleâ€� |
| EPI_ISL_420567 | Ospedale Regionale San Salvatore | Istituto Zooprofilattico Sperimentale dellâ€™Abruzzo e Molise â€œG. Caporaleâ€� |
| EPI_ISL_420568 | Ospedale Civile Giuseppe Mazzini | Istituto Zooprofilattico Sperimentale dellâ€™Abruzzo e Molise â€œG. Caporaleâ€� |

| EPI_Accession_ID | Originating Laboratory | Submitting Laboratory |
|---|---|---|
| EPI_ISL_420569 | Ospedale Civile Giuseppe Mazzini | Istituto Zooprofilattico Sperimentale dellâ€™Abruzzo e Molise â€œG. Caporaleâ€� |
| EPI_ISL_420570 | NYU Langone Health | Departments of Pathology and Medicine, New York University School of Medicine |
| EPI_ISL_420571 | NYU Langone Health | Departments of Pathology and Medicine, New York University School of Medicine |
| EPI_ISL_420572 | NYU Langone Health | Departments of Pathology and Medicine, New York University School of Medicine |
| EPI_ISL_420573 | NYU Langone Health | Departments of Pathology and Medicine, New York University School of Medicine |
| EPI_ISL_420574 | NYU Langone Health | Departments of Pathology and Medicine, New York University School of Medicine |
| EPI_ISL_420575 | NYU Langone Health | Departments of Pathology and Medicine, New York University School of Medicine |
| EPI_ISL_420576 | NYU Langone Health | Departments of Pathology and Medicine, New York University School of Medicine |
| EPI_ISL_420577 | NYU Langone Health | Departments of Pathology and Medicine, New York University School of Medicine |
| EPI_ISL_420578 | NYU Langone Health | Departments of Pathology and Medicine, New York University School of Medicine |
| EPI_ISL_420579 | NYU Langone Health | Departments of Pathology and Medicine, New York University School of Medicine |
| EPI_ISL_420580 | NYU Langone Health | Departments of Pathology and Medicine, New York University School of Medicine |
| EPI_ISL_420581 | NYU Langone Health | Departments of Pathology and Medicine, New York University School of Medicine |
| EPI_ISL_420582 | NYU Langone Health | Departments of Pathology and Medicine, New York University School of Medicine |
| EPI_ISL_420583 | Ospedale Civile Giuseppe Mazzini | Istituto Zooprofilattico Sperimentale dellâ€™Abruzzo e Molise â€œG. Caporaleâ€� |
| EPI_ISL_420584 | NYU Langone Health | Departments of Pathology and Medicine, New York University School of Medicine |

| EPI_Accession_ID | Originating Laboratory | Submitting Laboratory |
|---|---|---|
| EPI_ISL_420585 | NYU Langone Health | Departments of Pathology and Medicine, New York University School of Medicine |
| EPI_ISL_420586 | NYU Langone Health | Departments of Pathology and Medicine, New York University School of Medicine |
| EPI_ISL_420587 | NYU Langone Health | Departments of Pathology and Medicine, New York University School of Medicine |
| EPI_ISL_420588 | NYU Langone Health | Departments of Pathology and Medicine, New York University School of Medicine |
| EPI_ISL_420589 | NYU Langone Health | Departments of Pathology and Medicine, New York University School of Medicine |
| EPI_ISL_420590 | NYU Langone Health | Departments of Pathology and Medicine, New York University School of Medicine |
| EPI_ISL_420591 | NYU Langone Health | Departments of Pathology and Medicine, New York University School of Medicine |
| EPI_ISL_420592 | Ospedale Civile Giuseppe Mazzini | Istituto Zooprofilattico Sperimentale dellâ€™Abruzzo e Molise â€œG. Caporaleâ€� |
| EPI_ISL_420598 | Servicio Virosis Respiratorias-Departamento VirologÃ-a-INEI | Instituto Nacional Enfermedades Infecciosas C.G.Malbran |
| EPI_ISL_420599 | Servicio Virosis Respiratorias-Departamento VirologÃ-a-INEI | Instituto Nacional Enfermedades Infecciosas C.G.Malbran |
| EPI_ISL_420600 | Servicio Virosis Respiratorias-Departamento VirologÃ-a-INEI | Instituto Nacional Enfermedades Infecciosas C.G.Malbran |
| EPI_ISL_420604 | Institut des Agents Infectieux (IAI), Hospices Civils de Lyon | CNR Virus des Infections Respiratoires - France SUD |
| EPI_ISL_420605 | Institut des Agents Infectieux (IAI), Hospices Civils de Lyon | CNR Virus des Infections Respiratoires - France SUD |
| EPI_ISL_420606 | Institut des Agents Infectieux (IAI), Hospices Civils de Lyon | CNR Virus des Infections Respiratoires - France SUD |
| EPI_ISL_420607 | Institut des Agents Infectieux (IAI), Hospices Civils de Lyon | CNR Virus des Infections Respiratoires - France SUD |
| EPI_ISL_420608 | Institut des Agents Infectieux (IAI), Hospices Civils de Lyon | CNR Virus des Infections Respiratoires - France SUD |

| EPI_Accession_ID | Originating Laboratory | Submitting Laboratory |
|---|---|---|
| EPI_ISL_420609 | Institut des Agents Infectieux (IAI), Hospices Civils de Lyon | CNR Virus des Infections Respiratoires - France SUD |
| EPI_ISL_420610 | Institut des Agents Infectieux (IAI), Hospices Civils de Lyon | CNR Virus des Infections Respiratoires - France SUD |
| EPI_ISL_420611 | Institut des Agents Infectieux (IAI), Hospices Civils de Lyon | CNR Virus des Infections Respiratoires - France SUD |
| EPI_ISL_420612 | Centre Hospitalier de Macon | CNR Virus des Infections Respiratoires - France SUD |
| EPI_ISL_420613 | Centre Hospitalier de Macon | CNR Virus des Infections Respiratoires - France SUD |
| EPI_ISL_420614 | Centre Hospitalier de Macon | CNR Virus des Infections Respiratoires - France SUD |
| EPI_ISL_420615 | Institut des Agents Infectieux (IAI), Hospices Civils de Lyon | CNR Virus des Infections Respiratoires - France SUD |
| EPI_ISL_420616 | Institut des Agents Infectieux (IAI), Hospices Civils de Lyon | CNR Virus des Infections Respiratoires - France SUD |
| EPI_ISL_420617 | Centre Hospitalier Saint Joseph Saint Luc | CNR Virus des Infections Respiratoires - France SUD |
| EPI_ISL_420618 | Institut des Agents Infectieux (IAI), Hospices Civils de Lyon | CNR Virus des Infections Respiratoires - France SUD |
| EPI_ISL_420619 | Institut des Agents Infectieux (IAI), Hospices Civils de Lyon | CNR Virus des Infections Respiratoires - France SUD |
| EPI_ISL_420620 | Centre Hospitalier de Bourg en Bresse | CNR Virus des Infections Respiratoires - France SUD |
| EPI_ISL_420621 | Institut des Agents Infectieux (IAI), Hospices Civils de Lyon | CNR Virus des Infections Respiratoires - France SUD |
| EPI_ISL_420622 | Institut des Agents Infectieux (IAI), Hospices Civils de Lyon | CNR Virus des Infections Respiratoires - France SUD |
| EPI_ISL_420623 | Institut des Agents Infectieux (IAI), Hospices Civils de Lyon | CNR Virus des Infections Respiratoires - France SUD |
| EPI_ISL_420624 | Institut des Agents Infectieux (IAI), Hospices Civils de Lyon | CNR Virus des Infections Respiratoires - France SUD |
| EPI_ISL_420625 | Institut des Agents Infectieux (IAI), Hospices Civils de Lyon | CNR Virus des Infections Respiratoires - France SUD |
| EPI_ISL_420628 | Virginia DCLS | Virginia DCLS |
| EPI_ISL_420629 | Virginia DCLS | Virginia DCLS |
| EPI_ISL_420630 | Virginia DCLS | Virginia DCLS |

| EPI_Accession_ID | Originating Laboratory | Submitting Laboratory |
|---|---|---|
| EPI_ISL_420631 | Respiratory Virus Unit, Microbiology Services Colindale, Public Health England | Respiratory Virus Unit, Microbiology Services Colindale, Public Health England |
| EPI_ISL_420632 | Respiratory Virus Unit, Microbiology Services Colindale, Public Health England | Respiratory Virus Unit, Microbiology Services Colindale, Public Health England |
| EPI_ISL_420633 | Respiratory Virus Unit, Microbiology Services Colindale, Public Health England | Respiratory Virus Unit, Microbiology Services Colindale, Public Health England |
| EPI_ISL_420634 | Respiratory Virus Unit, Microbiology Services Colindale, Public Health England | Respiratory Virus Unit, Microbiology Services Colindale, Public Health England |
| EPI_ISL_420635 | Respiratory Virus Unit, Microbiology Services Colindale, Public Health England | Respiratory Virus Unit, Microbiology Services Colindale, Public Health England |
| EPI_ISL_420636 | Respiratory Virus Unit, Microbiology Services Colindale, Public Health England | Respiratory Virus Unit, Microbiology Services Colindale, Public Health England |
| EPI_ISL_420637 | Respiratory Virus Unit, Microbiology Services Colindale, Public Health England | Respiratory Virus Unit, Microbiology Services Colindale, Public Health England |
| EPI_ISL_420638 | Respiratory Virus Unit, Microbiology Services Colindale, Public Health England | Respiratory Virus Unit, Microbiology Services Colindale, Public Health England |
| EPI_ISL_420639 | Respiratory Virus Unit, Microbiology Services Colindale, Public Health England | Respiratory Virus Unit, Microbiology Services Colindale, Public Health England |
| EPI_ISL_420640 | Respiratory Virus Unit, Microbiology Services Colindale, Public Health England | Respiratory Virus Unit, Microbiology Services Colindale, Public Health England |
| EPI_ISL_420641 | Respiratory Virus Unit, Microbiology Services Colindale, Public Health England | Respiratory Virus Unit, Microbiology Services Colindale, Public Health England |
| EPI_ISL_420642 | Respiratory Virus Unit, Microbiology Services Colindale, Public Health England | Respiratory Virus Unit, Microbiology Services Colindale, Public Health England |
| EPI_ISL_420643 | Respiratory Virus Unit, Microbiology Services Colindale, Public Health England | Respiratory Virus Unit, Microbiology Services Colindale, Public Health England |
| EPI_ISL_420644 | Respiratory Virus Unit, Microbiology Services Colindale, Public Health England | Respiratory Virus Unit, Microbiology Services Colindale, Public Health England |
| EPI_ISL_420645 | Respiratory Virus Unit, Microbiology Services Colindale, Public Health England | Respiratory Virus Unit, Microbiology Services Colindale, Public Health England |
| EPI_ISL_420646 | Respiratory Virus Unit, Microbiology Services Colindale, Public Health England | Respiratory Virus Unit, Microbiology Services Colindale, Public Health England |

| EPI_Accession_ID | Originating Laboratory | Submitting Laboratory |
|---|---|---|
| EPI_ISL_420647 | Respiratory Virus Unit, Microbiology Services Colindale, Public Health England | Respiratory Virus Unit, Microbiology Services Colindale, Public Health England |
| EPI_ISL_420648 | Respiratory Virus Unit, Microbiology Services Colindale, Public Health England | Respiratory Virus Unit, Microbiology Services Colindale, Public Health England |
| EPI_ISL_420649 | Respiratory Virus Unit, Microbiology Services Colindale, Public Health England | Respiratory Virus Unit, Microbiology Services Colindale, Public Health England |
| EPI_ISL_420650 | Respiratory Virus Unit, Microbiology Services Colindale, Public Health England | Respiratory Virus Unit, Microbiology Services Colindale, Public Health England |
| EPI_ISL_420651 | Respiratory Virus Unit, Microbiology Services Colindale, Public Health England | Respiratory Virus Unit, Microbiology Services Colindale, Public Health England |
| EPI_ISL_420652 | Respiratory Virus Unit, Microbiology Services Colindale, Public Health England | Respiratory Virus Unit, Microbiology Services Colindale, Public Health England |
| EPI_ISL_420653 | Respiratory Virus Unit, Microbiology Services Colindale, Public Health England | Respiratory Virus Unit, Microbiology Services Colindale, Public Health England |
| EPI_ISL_420654 | Respiratory Virus Unit, Microbiology Services Colindale, Public Health England | Respiratory Virus Unit, Microbiology Services Colindale, Public Health England |
| EPI_ISL_420655 | Respiratory Virus Unit, Microbiology Services Colindale, Public Health England | Respiratory Virus Unit, Microbiology Services Colindale, Public Health England |
| EPI_ISL_420656 | Respiratory Virus Unit, Microbiology Services Colindale, Public Health England | Respiratory Virus Unit, Microbiology Services Colindale, Public Health England |
| EPI_ISL_420657 | Respiratory Virus Unit, Microbiology Services Colindale, Public Health England | Respiratory Virus Unit, Microbiology Services Colindale, Public Health England |
| EPI_ISL_420658 | Respiratory Virus Unit, Microbiology Services Colindale, Public Health England | Respiratory Virus Unit, Microbiology Services Colindale, Public Health England |
| EPI_ISL_420659 | Respiratory Virus Unit, Microbiology Services Colindale, Public Health England | Respiratory Virus Unit, Microbiology Services Colindale, Public Health England |
| EPI_ISL_420660 | Respiratory Virus Unit, Microbiology Services Colindale, Public Health England | Respiratory Virus Unit, Microbiology Services Colindale, Public Health England |
| EPI_ISL_420661 | Respiratory Virus Unit, Microbiology Services Colindale, Public Health England | Respiratory Virus Unit, Microbiology Services Colindale, Public Health England |
| EPI_ISL_420662 | Respiratory Virus Unit, Microbiology Services Colindale, Public Health England | Respiratory Virus Unit, Microbiology Services Colindale, Public Health England |

| EPI_Accession_ID | Originating Laboratory | Submitting Laboratory |
|---|---|---|
| EPI_ISL_420663 | Respiratory Virus Unit, Microbiology Services Colindale, Public Health England | Respiratory Virus Unit, Microbiology Services Colindale, Public Health England |
| EPI_ISL_420664 | Respiratory Virus Unit, Microbiology Services Colindale, Public Health England | Respiratory Virus Unit, Microbiology Services Colindale, Public Health England |
| EPI_ISL_420665 | Respiratory Virus Unit, Microbiology Services Colindale, Public Health England | Respiratory Virus Unit, Microbiology Services Colindale, Public Health England |
| EPI_ISL_420666 | Respiratory Virus Unit, Microbiology Services Colindale, Public Health England | Respiratory Virus Unit, Microbiology Services Colindale, Public Health England |
| EPI_ISL_420667 | Respiratory Virus Unit, Microbiology Services Colindale, Public Health England | Respiratory Virus Unit, Microbiology Services Colindale, Public Health England |
| EPI_ISL_420668 | Respiratory Virus Unit, Microbiology Services Colindale, Public Health England | Respiratory Virus Unit, Microbiology Services Colindale, Public Health England |
| EPI_ISL_420669 | Respiratory Virus Unit, Microbiology Services Colindale, Public Health England | Respiratory Virus Unit, Microbiology Services Colindale, Public Health England |
| EPI_ISL_420670 | Respiratory Virus Unit, Microbiology Services Colindale, Public Health England | Respiratory Virus Unit, Microbiology Services Colindale, Public Health England |
| EPI_ISL_420671 | Respiratory Virus Unit, Microbiology Services Colindale, Public Health England | Respiratory Virus Unit, Microbiology Services Colindale, Public Health England |
| EPI_ISL_420672 | Respiratory Virus Unit, Microbiology Services Colindale, Public Health England | Respiratory Virus Unit, Microbiology Services Colindale, Public Health England |
| EPI_ISL_420673 | Respiratory Virus Unit, Microbiology Services Colindale, Public Health England | Respiratory Virus Unit, Microbiology Services Colindale, Public Health England |
| EPI_ISL_420674 | Respiratory Virus Unit, Microbiology Services Colindale, Public Health England | Respiratory Virus Unit, Microbiology Services Colindale, Public Health England |
| EPI_ISL_420675 | Respiratory Virus Unit, Microbiology Services Colindale, Public Health England | Respiratory Virus Unit, Microbiology Services Colindale, Public Health England |
| EPI_ISL_420676 | Respiratory Virus Unit, Microbiology Services Colindale, Public Health England | Respiratory Virus Unit, Microbiology Services Colindale, Public Health England |
| EPI_ISL_420677 | Respiratory Virus Unit, Microbiology Services Colindale, Public Health England | Respiratory Virus Unit, Microbiology Services Colindale, Public Health England |
| EPI_ISL_420678 | Respiratory Virus Unit, Microbiology Services Colindale, Public Health England | Respiratory Virus Unit, Microbiology Services Colindale, Public Health England |

| EPI_Accession_ID | Originating Laboratory | Submitting Laboratory |
| --- | --- | --- |
| EPI_ISL_420679 | Respiratory Virus Unit, Microbiology Services Colindale, Public Health England | Respiratory Virus Unit, Microbiology Services Colindale, Public Health England |
| EPI_ISL_420680 | Respiratory Virus Unit, Microbiology Services Colindale, Public Health England | Respiratory Virus Unit, Microbiology Services Colindale, Public Health England |
| EPI_ISL_420681 | Respiratory Virus Unit, Microbiology Services Colindale, Public Health England | Respiratory Virus Unit, Microbiology Services Colindale, Public Health England |
| EPI_ISL_420682 | Respiratory Virus Unit, Microbiology Services Colindale, Public Health England | Respiratory Virus Unit, Microbiology Services Colindale, Public Health England |
| EPI_ISL_420683 | Respiratory Virus Unit, Microbiology Services Colindale, Public Health England | Respiratory Virus Unit, Microbiology Services Colindale, Public Health England |
| EPI_ISL_420684 | Respiratory Virus Unit, Microbiology Services Colindale, Public Health England | Respiratory Virus Unit, Microbiology Services Colindale, Public Health England |
| EPI_ISL_420685 | Respiratory Virus Unit, Microbiology Services Colindale, Public Health England | Respiratory Virus Unit, Microbiology Services Colindale, Public Health England |
| EPI_ISL_420686 | Respiratory Virus Unit, Microbiology Services Colindale, Public Health England | Respiratory Virus Unit, Microbiology Services Colindale, Public Health England |
| EPI_ISL_420687 | Respiratory Virus Unit, Microbiology Services Colindale, Public Health England | Respiratory Virus Unit, Microbiology Services Colindale, Public Health England |
| EPI_ISL_420688 | Respiratory Virus Unit, Microbiology Services Colindale, Public Health England | Respiratory Virus Unit, Microbiology Services Colindale, Public Health England |
| EPI_ISL_420689 | Respiratory Virus Unit, Microbiology Services Colindale, Public Health England | Respiratory Virus Unit, Microbiology Services Colindale, Public Health England |
| EPI_ISL_420690 | Respiratory Virus Unit, Microbiology Services Colindale, Public Health England | Respiratory Virus Unit, Microbiology Services Colindale, Public Health England |
| EPI_ISL_420691 | Respiratory Virus Unit, Microbiology Services Colindale, Public Health England | Respiratory Virus Unit, Microbiology Services Colindale, Public Health England |
| EPI_ISL_420692 | Respiratory Virus Unit, Microbiology Services Colindale, Public Health England | Respiratory Virus Unit, Microbiology Services Colindale, Public Health England |
| EPI_ISL_420693 | Respiratory Virus Unit, Microbiology Services Colindale, Public Health England | Respiratory Virus Unit, Microbiology Services Colindale, Public Health England |
| EPI_ISL_420694 | Respiratory Virus Unit, Microbiology Services Colindale, Public Health England | Respiratory Virus Unit, Microbiology Services Colindale, Public Health England |

| EPI_Accession_ID | Originating Laboratory | Submitting Laboratory |
|---|---|---|
| EPI_ISL_420695 | Respiratory Virus Unit, Microbiology Services Colindale, Public Health England | Respiratory Virus Unit, Microbiology Services Colindale, Public Health England |
| EPI_ISL_420696 | Respiratory Virus Unit, Microbiology Services Colindale, Public Health England | Respiratory Virus Unit, Microbiology Services Colindale, Public Health England |
| EPI_ISL_420697 | Respiratory Virus Unit, Microbiology Services Colindale, Public Health England | Respiratory Virus Unit, Microbiology Services Colindale, Public Health England |
| EPI_ISL_420698 | Respiratory Virus Unit, Microbiology Services Colindale, Public Health England | Respiratory Virus Unit, Microbiology Services Colindale, Public Health England |
| EPI_ISL_420699 | Respiratory Virus Unit, Microbiology Services Colindale, Public Health England | Respiratory Virus Unit, Microbiology Services Colindale, Public Health England |
| EPI_ISL_420700 | Respiratory Virus Unit, Microbiology Services Colindale, Public Health England | Respiratory Virus Unit, Microbiology Services Colindale, Public Health England |
| EPI_ISL_420701 | Respiratory Virus Unit, Microbiology Services Colindale, Public Health England | Respiratory Virus Unit, Microbiology Services Colindale, Public Health England |
| EPI_ISL_420702 | Respiratory Virus Unit, Microbiology Services Colindale, Public Health England | Respiratory Virus Unit, Microbiology Services Colindale, Public Health England |
| EPI_ISL_420703 | Respiratory Virus Unit, Microbiology Services Colindale, Public Health England | Respiratory Virus Unit, Microbiology Services Colindale, Public Health England |
| EPI_ISL_420704 | Respiratory Virus Unit, Microbiology Services Colindale, Public Health England | Respiratory Virus Unit, Microbiology Services Colindale, Public Health England |
| EPI_ISL_420705 | Respiratory Virus Unit, Microbiology Services Colindale, Public Health England | Respiratory Virus Unit, Microbiology Services Colindale, Public Health England |
| EPI_ISL_420706 | Respiratory Virus Unit, Microbiology Services Colindale, Public Health England | Respiratory Virus Unit, Microbiology Services Colindale, Public Health England |
| EPI_ISL_420707 | Respiratory Virus Unit, Microbiology Services Colindale, Public Health England | Respiratory Virus Unit, Microbiology Services Colindale, Public Health England |
| EPI_ISL_420708 | Respiratory Virus Unit, Microbiology Services Colindale, Public Health England | Respiratory Virus Unit, Microbiology Services Colindale, Public Health England |
| EPI_ISL_420709 | Respiratory Virus Unit, Microbiology Services Colindale, Public Health England | Respiratory Virus Unit, Microbiology Services Colindale, Public Health England |
| EPI_ISL_420710 | Respiratory Virus Unit, Microbiology Services Colindale, Public Health England | Respiratory Virus Unit, Microbiology Services Colindale, Public Health England |

| EPI_Accession_ID | Originating Laboratory | Submitting Laboratory |
|---|---|---|
| EPI_ISL_420711 | Respiratory Virus Unit, Microbiology Services Colindale, Public Health England | Respiratory Virus Unit, Microbiology Services Colindale, Public Health England |
| EPI_ISL_420712 | Respiratory Virus Unit, Microbiology Services Colindale, Public Health England | Respiratory Virus Unit, Microbiology Services Colindale, Public Health England |
| EPI_ISL_420713 | Respiratory Virus Unit, Microbiology Services Colindale, Public Health England | Respiratory Virus Unit, Microbiology Services Colindale, Public Health England |
| EPI_ISL_420714 | Respiratory Virus Unit, Microbiology Services Colindale, Public Health England | Respiratory Virus Unit, Microbiology Services Colindale, Public Health England |
| EPI_ISL_420715 | Respiratory Virus Unit, Microbiology Services Colindale, Public Health England | Respiratory Virus Unit, Microbiology Services Colindale, Public Health England |
| EPI_ISL_420716 | Respiratory Virus Unit, Microbiology Services Colindale, Public Health England | Respiratory Virus Unit, Microbiology Services Colindale, Public Health England |
| EPI_ISL_420717 | Respiratory Virus Unit, Microbiology Services Colindale, Public Health England | Respiratory Virus Unit, Microbiology Services Colindale, Public Health England |
| EPI_ISL_420718 | Respiratory Virus Unit, Microbiology Services Colindale, Public Health England | Respiratory Virus Unit, Microbiology Services Colindale, Public Health England |
| EPI_ISL_420719 | Respiratory Virus Unit, Microbiology Services Colindale, Public Health England | Respiratory Virus Unit, Microbiology Services Colindale, Public Health England |
| EPI_ISL_420720 | Respiratory Virus Unit, Microbiology Services Colindale, Public Health England | Respiratory Virus Unit, Microbiology Services Colindale, Public Health England |
| EPI_ISL_420721 | Respiratory Virus Unit, Microbiology Services Colindale, Public Health England | Respiratory Virus Unit, Microbiology Services Colindale, Public Health England |
| EPI_ISL_420722 | Respiratory Virus Unit, Microbiology Services Colindale, Public Health England | Respiratory Virus Unit, Microbiology Services Colindale, Public Health England |
| EPI_ISL_420723 | Respiratory Virus Unit, Microbiology Services Colindale, Public Health England | Respiratory Virus Unit, Microbiology Services Colindale, Public Health England |
| EPI_ISL_420724 | Respiratory Virus Unit, Microbiology Services Colindale, Public Health England | Respiratory Virus Unit, Microbiology Services Colindale, Public Health England |
| EPI_ISL_420725 | Respiratory Virus Unit, Microbiology Services Colindale, Public Health England | Respiratory Virus Unit, Microbiology Services Colindale, Public Health England |
| EPI_ISL_420726 | Respiratory Virus Unit, Microbiology Services Colindale, Public Health England | Respiratory Virus Unit, Microbiology Services Colindale, Public Health England |

| EPI_Accession_ID | Originating Laboratory | Submitting Laboratory |
|---|---|---|
| EPI_ISL_420727 | Respiratory Virus Unit, Microbiology Services Colindale, Public Health England | Respiratory Virus Unit, Microbiology Services Colindale, Public Health England |
| EPI_ISL_420728 | Respiratory Virus Unit, Microbiology Services Colindale, Public Health England | Respiratory Virus Unit, Microbiology Services Colindale, Public Health England |
| EPI_ISL_420729 | Respiratory Virus Unit, Microbiology Services Colindale, Public Health England | Respiratory Virus Unit, Microbiology Services Colindale, Public Health England |
| EPI_ISL_420730 | Respiratory Virus Unit, Microbiology Services Colindale, Public Health England | Respiratory Virus Unit, Microbiology Services Colindale, Public Health England |
| EPI_ISL_420731 | Respiratory Virus Unit, Microbiology Services Colindale, Public Health England | Respiratory Virus Unit, Microbiology Services Colindale, Public Health England |
| EPI_ISL_420732 | Respiratory Virus Unit, Microbiology Services Colindale, Public Health England | Respiratory Virus Unit, Microbiology Services Colindale, Public Health England |
| EPI_ISL_420733 | Respiratory Virus Unit, Microbiology Services Colindale, Public Health England | Respiratory Virus Unit, Microbiology Services Colindale, Public Health England |
| EPI_ISL_420734 | Respiratory Virus Unit, Microbiology Services Colindale, Public Health England | Respiratory Virus Unit, Microbiology Services Colindale, Public Health England |
| EPI_ISL_420735 | Respiratory Virus Unit, Microbiology Services Colindale, Public Health England | Respiratory Virus Unit, Microbiology Services Colindale, Public Health England |
| EPI_ISL_420736 | Respiratory Virus Unit, Microbiology Services Colindale, Public Health England | Respiratory Virus Unit, Microbiology Services Colindale, Public Health England |
| EPI_ISL_420737 | Respiratory Virus Unit, Microbiology Services Colindale, Public Health England | Respiratory Virus Unit, Microbiology Services Colindale, Public Health England |
| EPI_ISL_420738 | Respiratory Virus Unit, Microbiology Services Colindale, Public Health England | Respiratory Virus Unit, Microbiology Services Colindale, Public Health England |
| EPI_ISL_420739 | Respiratory Virus Unit, Microbiology Services Colindale, Public Health England | Respiratory Virus Unit, Microbiology Services Colindale, Public Health England |
| EPI_ISL_420740 | Respiratory Virus Unit, Microbiology Services Colindale, Public Health England | Respiratory Virus Unit, Microbiology Services Colindale, Public Health England |
| EPI_ISL_420741 | Respiratory Virus Unit, Microbiology Services Colindale, Public Health England | Respiratory Virus Unit, Microbiology Services Colindale, Public Health England |
| EPI_ISL_420742 | Respiratory Virus Unit, Microbiology Services Colindale, Public Health England | Respiratory Virus Unit, Microbiology Services Colindale, Public Health England |

| EPI_Accession_ID | Originating Laboratory | Submitting Laboratory |
|---|---|---|
| EPI_ISL_420743 | Respiratory Virus Unit, Microbiology Services Colindale, Public Health England | Respiratory Virus Unit, Microbiology Services Colindale, Public Health England |
| EPI_ISL_420744 | Respiratory Virus Unit, Microbiology Services Colindale, Public Health England | Respiratory Virus Unit, Microbiology Services Colindale, Public Health England |
| EPI_ISL_420745 | Respiratory Virus Unit, Microbiology Services Colindale, Public Health England | Respiratory Virus Unit, Microbiology Services Colindale, Public Health England |
| EPI_ISL_420746 | Respiratory Virus Unit, Microbiology Services Colindale, Public Health England | Respiratory Virus Unit, Microbiology Services Colindale, Public Health England |
| EPI_ISL_420747 | Respiratory Virus Unit, Microbiology Services Colindale, Public Health England | Respiratory Virus Unit, Microbiology Services Colindale, Public Health England |
| EPI_ISL_420748 | Respiratory Virus Unit, Microbiology Services Colindale, Public Health England | Respiratory Virus Unit, Microbiology Services Colindale, Public Health England |
| EPI_ISL_420749 | Respiratory Virus Unit, Microbiology Services Colindale, Public Health England | Respiratory Virus Unit, Microbiology Services Colindale, Public Health England |
| EPI_ISL_420750 | Respiratory Virus Unit, Microbiology Services Colindale, Public Health England | Respiratory Virus Unit, Microbiology Services Colindale, Public Health England |
| EPI_ISL_420751 | Respiratory Virus Unit, Microbiology Services Colindale, Public Health England | Respiratory Virus Unit, Microbiology Services Colindale, Public Health England |
| EPI_ISL_420752 | Respiratory Virus Unit, Microbiology Services Colindale, Public Health England | Respiratory Virus Unit, Microbiology Services Colindale, Public Health England |
| EPI_ISL_420753 | Respiratory Virus Unit, Microbiology Services Colindale, Public Health England | Respiratory Virus Unit, Microbiology Services Colindale, Public Health England |
| EPI_ISL_420754 | Respiratory Virus Unit, Microbiology Services Colindale, Public Health England | Respiratory Virus Unit, Microbiology Services Colindale, Public Health England |
| EPI_ISL_420755 | Respiratory Virus Unit, Microbiology Services Colindale, Public Health England | Respiratory Virus Unit, Microbiology Services Colindale, Public Health England |
| EPI_ISL_420756 | Respiratory Virus Unit, Microbiology Services Colindale, Public Health England | Respiratory Virus Unit, Microbiology Services Colindale, Public Health England |
| EPI_ISL_420757 | Respiratory Virus Unit, Microbiology Services Colindale, Public Health England | Respiratory Virus Unit, Microbiology Services Colindale, Public Health England |
| EPI_ISL_420758 | Respiratory Virus Unit, Microbiology Services Colindale, Public Health England | Respiratory Virus Unit, Microbiology Services Colindale, Public Health England |

| EPI_Accession_ID | Originating Laboratory | Submitting Laboratory |
|---|---|---|
| EPI_ISL_420759 | Respiratory Virus Unit, Microbiology Services Colindale, Public Health England | Respiratory Virus Unit, Microbiology Services Colindale, Public Health England |
| EPI_ISL_420760 | Respiratory Virus Unit, Microbiology Services Colindale, Public Health England | Respiratory Virus Unit, Microbiology Services Colindale, Public Health England |
| EPI_ISL_420761 | Respiratory Virus Unit, Microbiology Services Colindale, Public Health England | Respiratory Virus Unit, Microbiology Services Colindale, Public Health England |
| EPI_ISL_420762 | Respiratory Virus Unit, Microbiology Services Colindale, Public Health England | Respiratory Virus Unit, Microbiology Services Colindale, Public Health England |
| EPI_ISL_420763 | Respiratory Virus Unit, Microbiology Services Colindale, Public Health England | Respiratory Virus Unit, Microbiology Services Colindale, Public Health England |
| EPI_ISL_420764 | Respiratory Virus Unit, Microbiology Services Colindale, Public Health England | Respiratory Virus Unit, Microbiology Services Colindale, Public Health England |
| EPI_ISL_420765 | Respiratory Virus Unit, Microbiology Services Colindale, Public Health England | Respiratory Virus Unit, Microbiology Services Colindale, Public Health England |
| EPI_ISL_420766 | Respiratory Virus Unit, Microbiology Services Colindale, Public Health England | Respiratory Virus Unit, Microbiology Services Colindale, Public Health England |
| EPI_ISL_420767 | Respiratory Virus Unit, Microbiology Services Colindale, Public Health England | Respiratory Virus Unit, Microbiology Services Colindale, Public Health England |
| EPI_ISL_420768 | Respiratory Virus Unit, Microbiology Services Colindale, Public Health England | Respiratory Virus Unit, Microbiology Services Colindale, Public Health England |
| EPI_ISL_420769 | Respiratory Virus Unit, Microbiology Services Colindale, Public Health England | Respiratory Virus Unit, Microbiology Services Colindale, Public Health England |
| EPI_ISL_420770 | Respiratory Virus Unit, Microbiology Services Colindale, Public Health England | Respiratory Virus Unit, Microbiology Services Colindale, Public Health England |
| EPI_ISL_420771 | Respiratory Virus Unit, Microbiology Services Colindale, Public Health England | Respiratory Virus Unit, Microbiology Services Colindale, Public Health England |
| EPI_ISL_420772 | Respiratory Virus Unit, Microbiology Services Colindale, Public Health England | Respiratory Virus Unit, Microbiology Services Colindale, Public Health England |
| EPI_ISL_420773 | Respiratory Virus Unit, Microbiology Services Colindale, Public Health England | Respiratory Virus Unit, Microbiology Services Colindale, Public Health England |
| EPI_ISL_420774 | Respiratory Virus Unit, Microbiology Services Colindale, Public Health England | Respiratory Virus Unit, Microbiology Services Colindale, Public Health England |

| EPI_Accession_ID | Originating Laboratory | Submitting Laboratory |
|---|---|---|
| EPI_ISL_420775 | Respiratory Virus Unit, Microbiology Services Colindale, Public Health England | Respiratory Virus Unit, Microbiology Services Colindale, Public Health England |
| EPI_ISL_420776 | Respiratory Virus Unit, Microbiology Services Colindale, Public Health England | Respiratory Virus Unit, Microbiology Services Colindale, Public Health England |
| EPI_ISL_420777 | Respiratory Virus Unit, Microbiology Services Colindale, Public Health England | Respiratory Virus Unit, Microbiology Services Colindale, Public Health England |
| EPI_ISL_420784 | AZ Department of Health Services | Pathogen Discovery, Respiratory Viruses Branch, Division of Viral Diseases, Centers for Disease Control and Prevention |
| EPI_ISL_420785 | FL Bureau of Health Laboratories Tampa | Pathogen Discovery, Respiratory Viruses Branch, Division of Viral Diseases, Centers for Disease Control and Prevention |
| EPI_ISL_420786 | GA Department of Public Health | Pathogen Discovery, Respiratory Viruses Branch, Division of Viral Diseases, Centers for Disease Control and Prevention |
| EPI_ISL_420787 | GA Department of Public Health | Pathogen Discovery, Respiratory Viruses Branch, Division of Viral Diseases, Centers for Disease Control and Prevention |
| EPI_ISL_420788 | GA Department of Public Health | Pathogen Discovery, Respiratory Viruses Branch, Division of Viral Diseases, Centers for Disease Control and Prevention |
| EPI_ISL_420789 | Illinois Department of Public Health Chicago Laboratory | Pathogen Discovery, Respiratory Viruses Branch, Division of Viral Diseases, Centers for Disease Control and Prevention |
| EPI_ISL_420790 | Illinois Department of Public Health Chicago Laboratory | Pathogen Discovery, Respiratory Viruses Branch, Division of Viral Diseases, Centers for Disease Control and Prevention |
| EPI_ISL_420791 | NH Department of Health and Human Services Public Health Labs | Pathogen Discovery, Respiratory Viruses Branch, Division of Viral Diseases, Centers for Disease Control and Prevention |
| EPI_ISL_420792 | NH Department of Health and Human Services Public Health Labs | Pathogen Discovery, Respiratory Viruses Branch, Division of Viral Diseases, Centers for Disease Control and Prevention |

| EPI_Accession_ID | Originating Laboratory | Submitting Laboratory |
|---|---|---|
| EPI_ISL_420793 | NYC Department of Health and Mental Hygiene | Pathogen Discovery, Respiratory Viruses Branch, Division of Viral Diseases, Centers for Disease Control and Prevention |
| EPI_ISL_420794 | Oregon State Public Health- Virology section | Pathogen Discovery, Respiratory Viruses Branch, Division of Viral Diseases, Centers for Disease Control and Prevention |
| EPI_ISL_420795 | RI State Health Laboratory | Pathogen Discovery, Respiratory Viruses Branch, Division of Viral Diseases, Centers for Disease Control and Prevention |
| EPI_ISL_420796 | Texas DSHS Lab Services | Pathogen Discovery, Respiratory Viruses Branch, Division of Viral Diseases, Centers for Disease Control and Prevention |
| EPI_ISL_420797 | Texas DSHS Lab Services | Pathogen Discovery, Respiratory Viruses Branch, Division of Viral Diseases, Centers for Disease Control and Prevention |
| EPI_ISL_420798 | Texas DSHS Lab Services | Pathogen Discovery, Respiratory Viruses Branch, Division of Viral Diseases, Centers for Disease Control and Prevention |
| EPI_ISL_420799 | Brian D. Allgood Army Community Hospital | Pathogen Discovery, Respiratory Viruses Branch, Division of Viral Diseases, Centers for Disease Control and Prevention |
| EPI_ISL_420800 | Brian D. Allgood Army Community Hospital | Pathogen Discovery, Respiratory Viruses Branch, Division of Viral Diseases, Centers for Disease Control and Prevention |
| EPI_ISL_420801 | Brian D. Allgood Army Community Hospital | Pathogen Discovery, Respiratory Viruses Branch, Division of Viral Diseases, Centers for Disease Control and Prevention |
| EPI_ISL_420803 | Utah Public Health Laboratory | Utah Public Health Laboratory |
| EPI_ISL_420804 | Utah Public Health Laboratory | Utah Public Health Laboratory |
| EPI_ISL_420805 | Utah Public Health Laboratory | Utah Public Health Laboratory |
| EPI_ISL_420806 | Utah Public Health Laboratory | Utah Public Health Laboratory |
| EPI_ISL_420807 | Utah Public Health Laboratory | Utah Public Health Laboratory |
| EPI_ISL_420808 | Utah Public Health Laboratory | Utah Public Health Laboratory |

| EPI_Accession_ID | Originating Laboratory | Submitting Laboratory |
|---|---|---|
| EPI_ISL_420809 | Utah Public Health Laboratory | Utah Public Health Laboratory |
| EPI_ISL_420810 | Utah Public Health Laboratory | Utah Public Health Laboratory |
| EPI_ISL_420811 | Utah Public Health Laboratory | Utah Public Health Laboratory |
| EPI_ISL_420812 | Utah Public Health Laboratory | Utah Public Health Laboratory |
| EPI_ISL_420813 | Utah Public Health Laboratory | Utah Public Health Laboratory |
| EPI_ISL_420814 | Utah Public Health Laboratory | Utah Public Health Laboratory |
| EPI_ISL_420815 | Utah Public Health Laboratory | Utah Public Health Laboratory |
| EPI_ISL_420816 | Utah Public Health Laboratory | Utah Public Health Laboratory |
| EPI_ISL_420817 | Utah Public Health Laboratory | Utah Public Health Laboratory |
| EPI_ISL_420818 | Utah Public Health Laboratory | Utah Public Health Laboratory |
| EPI_ISL_420819 | Utah Public Health Laboratory | Utah Public Health Laboratory |
| EPI_ISL_420820 | Utah Public Health Laboratory | Utah Public Health Laboratory |
| EPI_ISL_420821 | Utah Public Health Laboratory | Utah Public Health Laboratory |
| EPI_ISL_420822 | Utah Public Health Laboratory | Utah Public Health Laboratory |
| EPI_ISL_420823 | Utah Public Health Laboratory | Utah Public Health Laboratory |
| EPI_ISL_420824 | Utah Public Health Laboratory | Utah Public Health Laboratory |
| EPI_ISL_420825 | Utah Public Health Laboratory | Utah Public Health Laboratory |
| EPI_ISL_420838 | Viral Respiratory Lab, National Institute for Biomedical Research (INRB) | Pathogen Sequencing Lab, National Institute for Biomedical Research (INRB) |
| EPI_ISL_420839 | Viral Respiratory Lab, National Institute for Biomedical Research (INRB) | Pathogen Sequencing Lab, National Institute for Biomedical Research (INRB) |
| EPI_ISL_420840 | Viral Respiratory Lab, National Institute for Biomedical Research (INRB) | Pathogen Sequencing Lab, National Institute for Biomedical Research (INRB) |
| EPI_ISL_420841 | Viral Respiratory Lab, National Institute for Biomedical Research (INRB) | Pathogen Sequencing Lab, National Institute for Biomedical Research (INRB) |
| EPI_ISL_420845 | Viral Respiratory Lab, National Institute for Biomedical Research (INRB) | Pathogen Sequencing Lab, National Institute for Biomedical Research (INRB) |
| EPI_ISL_420847 | Viral Respiratory Lab, National Institute for Biomedical Research (INRB) | Pathogen Sequencing Lab, National Institute for Biomedical Research (INRB) |
| EPI_ISL_420849 | Viral Respiratory Lab, National Institute for Biomedical Research (INRB) | Pathogen Sequencing Lab, National Institute for Biomedical Research (INRB) |
| EPI_ISL_420851 | Viral Respiratory Lab, National Institute for Biomedical Research (INRB) | Pathogen Sequencing Lab, National Institute for Biomedical Research (INRB) |

| EPI_Accession_ID | Originating Laboratory | Submitting Laboratory |
|---|---|---|
| EPI_ISL_420852 | Viral Respiratory Lab, National Institute for Biomedical Research (INRB) | Pathogen Sequencing Lab, National Institute for Biomedical Research (INRB) |
| EPI_ISL_420853 | Viral Respiratory Lab, National Institute for Biomedical Research (INRB) | Pathogen Sequencing Lab, National Institute for Biomedical Research (INRB) |
| EPI_ISL_420854 | Viral Respiratory Lab, National Institute for Biomedical Research (INRB) | Pathogen Sequencing Lab, National Institute for Biomedical Research (INRB) |
| EPI_ISL_420855 | Geelong Centre for Emerging Infectious Diseases | Geelong Centre for Emerging Infectious Diseases |
| EPI_ISL_420876 | Geelong Centre for Emerging Infectious Diseases | Geelong Centre for Emerging Infectious Diseases |
| EPI_ISL_420877 | Geelong Centre for Emerging Infectious Diseases | Geelong Centre for Emerging Infectious Diseases |
| EPI_ISL_420878 | Mater Pathology | Public Health Virology Laboratory |
| EPI_ISL_420879 | Mater Pathology | Public Health Virology Laboratory |
| EPI_ISL_420889 | unknown | Takayuki Hishiki Kanagawa Prefectural Institute of Public Health |
| EPI_ISL_420890 | unknown | Takayuki Hishiki Kanagawa Prefectural Institute of Public Health |
| EPI_ISL_420898 | Max von Pettenkofer Institute, Virology, National Reference Center for Retroviruses, LMU Munich | Laboratory for Functional Genome Analysis, Dept. Genomics, Gene Center of the LMU Munich |
| EPI_ISL_420899 | Max von Pettenkofer Institute, Virology, National Reference Center for Retroviruses, LMU Munich | Laboratory for Functional Genome Analysis, Dept. Genomics, Gene Center of the LMU Munich |
| EPI_ISL_420900 | Max von Pettenkofer Institute, Virology, National Reference Center for Retroviruses, LMU Munich | Laboratory for Functional Genome Analysis, Dept. Genomics, Gene Center of the LMU Munich |
| EPI_ISL_420901 | Max von Pettenkofer Institute, Virology, National Reference Center for Retroviruses, LMU Munich | Laboratory for Functional Genome Analysis, Dept. Genomics, Gene Center of the LMU Munich |
| EPI_ISL_420902 | Max von Pettenkofer Institute, Virology, National Reference Center for Retroviruses, LMU Munich | Laboratory for Functional Genome Analysis, Dept. Genomics, Gene Center of the LMU Munich |
| EPI_ISL_420903 | Max von Pettenkofer Institute, Virology, National Reference Center for Retroviruses, LMU Munich | Laboratory for Functional Genome Analysis, Dept. Genomics, Gene Center of the LMU Munich |
| EPI_ISL_420904 | Max von Pettenkofer Institute, Virology, National Reference Center for Retroviruses, LMU Munich | Laboratory for Functional Genome Analysis, Dept. Genomics, Gene Center of the LMU Munich |
| EPI_ISL_420905 | Max von Pettenkofer Institute, Virology, National Reference Center for Retroviruses, LMU Munich | Laboratory for Functional Genome Analysis, Dept. Genomics, Gene Center of the LMU Munich |
| EPI_ISL_420906 | Max von Pettenkofer Institute, Virology, National Reference Center for Retroviruses, LMU Munich | Laboratory for Functional Genome Analysis, Dept. Genomics, Gene Center of the LMU Munich |

| EPI_Accession_ID | Originating Laboratory | Submitting Laboratory |
|---|---|---|
| EPI_ISL_420907 | Max von Pettenkofer Institute, Virology, National Reference Center for Retroviruses, LMU Munich | Laboratory for Functional Genome Analysis, Dept. Genomics, Gene Center of the LMU Munich |
| EPI_ISL_420908 | Max von Pettenkofer Institute, Virology, National Reference Center for Retroviruses, LMU Munich | Laboratory for Functional Genome Analysis, Dept. Genomics, Gene Center of the LMU Munich |
| EPI_ISL_420909 | Max von Pettenkofer Institute, Virology, National Reference Center for Retroviruses, LMU Munich | Laboratory for Functional Genome Analysis, Dept. Genomics, Gene Center of the LMU Munich |
| EPI_ISL_420910 | Respiratory Virus Unit, Microbiology Services Colindale, Public Health England | Respiratory Virus Unit, Microbiology Services Colindale, Public Health England |
| EPI_ISL_420911 | Max von Pettenkofer Institute, Virology, National Reference Center for Retroviruses, LMU Munich | Laboratory for Functional Genome Analysis, Dept. Genomics, Gene Center of the LMU Munich |
| EPI_ISL_420912 | Max von Pettenkofer Institute, Virology, National Reference Center for Retroviruses, LMU Munich | Laboratory for Functional Genome Analysis, Dept. Genomics, Gene Center of the LMU Munich |
| EPI_ISL_420913 | Wales Specialist Virology Centre | Public Health Wales Microbiology Cardiff |
| EPI_ISL_420914 | Wales Specialist Virology Centre | Public Health Wales Microbiology Cardiff |
| EPI_ISL_420915 | Wales Specialist Virology Centre | Public Health Wales Microbiology Cardiff |
| EPI_ISL_420916 | Wales Specialist Virology Centre | Public Health Wales Microbiology Cardiff |
| EPI_ISL_420917 | Wales Specialist Virology Centre | Public Health Wales Microbiology Cardiff |
| EPI_ISL_420918 | Wales Specialist Virology Centre | Public Health Wales Microbiology Cardiff |
| EPI_ISL_420919 | Wales Specialist Virology Centre | Public Health Wales Microbiology Cardiff |
| EPI_ISL_420920 | Wales Specialist Virology Centre | Public Health Wales Microbiology Cardiff |
| EPI_ISL_420921 | Wales Specialist Virology Centre | Public Health Wales Microbiology Cardiff |
| EPI_ISL_420922 | Wales Specialist Virology Centre | Public Health Wales Microbiology Cardiff |
| EPI_ISL_420923 | Wales Specialist Virology Centre | Public Health Wales Microbiology Cardiff |
| EPI_ISL_420924 | Wales Specialist Virology Centre | Public Health Wales Microbiology Cardiff |
| EPI_ISL_420925 | Wales Specialist Virology Centre | Public Health Wales Microbiology Cardiff |
| EPI_ISL_420926 | Wales Specialist Virology Centre | Public Health Wales Microbiology Cardiff |
| EPI_ISL_420927 | Wales Specialist Virology Centre | Public Health Wales Microbiology Cardiff |
| EPI_ISL_420928 | Wales Specialist Virology Centre | Public Health Wales Microbiology Cardiff |
| EPI_ISL_420929 | Wales Specialist Virology Centre | Public Health Wales Microbiology Cardiff |
| EPI_ISL_420931 | Wales Specialist Virology Centre | Public Health Wales Microbiology Cardiff |
| EPI_ISL_420932 | Wales Specialist Virology Centre | Public Health Wales Microbiology Cardiff |
| EPI_ISL_420933 | Wales Specialist Virology Centre | Public Health Wales Microbiology Cardiff |
| EPI_ISL_420934 | Wales Specialist Virology Centre | Public Health Wales Microbiology Cardiff |

| EPI_Accession_ID | Originating Laboratory | Submitting Laboratory |
|---|---|---|
| EPI_ISL_420935 | Wales Specialist Virology Centre | Public Health Wales Microbiology Cardiff |
| EPI_ISL_420936 | Wales Specialist Virology Centre | Public Health Wales Microbiology Cardiff |
| EPI_ISL_420937 | Wales Specialist Virology Centre | Public Health Wales Microbiology Cardiff |
| EPI_ISL_420938 | Wales Specialist Virology Centre | Public Health Wales Microbiology Cardiff |
| EPI_ISL_420939 | Wales Specialist Virology Centre | Public Health Wales Microbiology Cardiff |
| EPI_ISL_420940 | Wales Specialist Virology Centre | Public Health Wales Microbiology Cardiff |
| EPI_ISL_420941 | Wales Specialist Virology Centre | Public Health Wales Microbiology Cardiff |
| EPI_ISL_420942 | Wales Specialist Virology Centre | Public Health Wales Microbiology Cardiff |
| EPI_ISL_420943 | Wales Specialist Virology Centre | Public Health Wales Microbiology Cardiff |
| EPI_ISL_420944 | Wales Specialist Virology Centre | Public Health Wales Microbiology Cardiff |
| EPI_ISL_420945 | Wales Specialist Virology Centre | Public Health Wales Microbiology Cardiff |
| EPI_ISL_420946 | Wales Specialist Virology Centre | Public Health Wales Microbiology Cardiff |
| EPI_ISL_420947 | Wales Specialist Virology Centre | Public Health Wales Microbiology Cardiff |
| EPI_ISL_420948 | Wales Specialist Virology Centre | Public Health Wales Microbiology Cardiff |
| EPI_ISL_420949 | Wales Specialist Virology Centre | Public Health Wales Microbiology Cardiff |
| EPI_ISL_420950 | Wales Specialist Virology Centre | Public Health Wales Microbiology Cardiff |
| EPI_ISL_420951 | Wales Specialist Virology Centre | Public Health Wales Microbiology Cardiff |
| EPI_ISL_420952 | Wales Specialist Virology Centre | Public Health Wales Microbiology Cardiff |
| EPI_ISL_420953 | Wales Specialist Virology Centre | Public Health Wales Microbiology Cardiff |
| EPI_ISL_420954 | Wales Specialist Virology Centre | Public Health Wales Microbiology Cardiff |
| EPI_ISL_420955 | Wales Specialist Virology Centre | Public Health Wales Microbiology Cardiff |
| EPI_ISL_420956 | Wales Specialist Virology Centre | Public Health Wales Microbiology Cardiff |
| EPI_ISL_420957 | Wales Specialist Virology Centre | Public Health Wales Microbiology Cardiff |
| EPI_ISL_420958 | Wales Specialist Virology Centre | Public Health Wales Microbiology Cardiff |
| EPI_ISL_420959 | Wales Specialist Virology Centre | Public Health Wales Microbiology Cardiff |
| EPI_ISL_420960 | Wales Specialist Virology Centre | Public Health Wales Microbiology Cardiff |
| EPI_ISL_420961 | Wales Specialist Virology Centre | Public Health Wales Microbiology Cardiff |
| EPI_ISL_420962 | Wales Specialist Virology Centre | Public Health Wales Microbiology Cardiff |
| EPI_ISL_420963 | Wales Specialist Virology Centre | Public Health Wales Microbiology Cardiff |
| EPI_ISL_420964 | Wales Specialist Virology Centre | Public Health Wales Microbiology Cardiff |
| EPI_ISL_420965 | Wales Specialist Virology Centre | Public Health Wales Microbiology Cardiff |
| EPI_ISL_420966 | Wales Specialist Virology Centre | Public Health Wales Microbiology Cardiff |
| EPI_ISL_420967 | Wales Specialist Virology Centre | Public Health Wales Microbiology Cardiff |

| EPI_Accession_ID | Originating Laboratory | Submitting Laboratory |
|---|---|---|
| EPI_ISL_420968 | Wales Specialist Virology Centre | Public Health Wales Microbiology Cardiff |
| EPI_ISL_420969 | Wales Specialist Virology Centre | Public Health Wales Microbiology Cardiff |
| EPI_ISL_420970 | Wales Specialist Virology Centre | Public Health Wales Microbiology Cardiff |
| EPI_ISL_420971 | Wales Specialist Virology Centre | Public Health Wales Microbiology Cardiff |
| EPI_ISL_420972 | Wales Specialist Virology Centre | Public Health Wales Microbiology Cardiff |
| EPI_ISL_420973 | Wales Specialist Virology Centre | Public Health Wales Microbiology Cardiff |
| EPI_ISL_420974 | Wales Specialist Virology Centre | Public Health Wales Microbiology Cardiff |
| EPI_ISL_420975 | Wales Specialist Virology Centre | Public Health Wales Microbiology Cardiff |
| EPI_ISL_420976 | Wales Specialist Virology Centre | Public Health Wales Microbiology Cardiff |
| EPI_ISL_420977 | Wales Specialist Virology Centre | Public Health Wales Microbiology Cardiff |
| EPI_ISL_420978 | Wales Specialist Virology Centre | Public Health Wales Microbiology Cardiff |
| EPI_ISL_420979 | Wales Specialist Virology Centre | Public Health Wales Microbiology Cardiff |
| EPI_ISL_420980 | Wales Specialist Virology Centre | Public Health Wales Microbiology Cardiff |
| EPI_ISL_420981 | Wales Specialist Virology Centre | Public Health Wales Microbiology Cardiff |
| EPI_ISL_420982 | Wales Specialist Virology Centre | Public Health Wales Microbiology Cardiff |
| EPI_ISL_420983 | Wales Specialist Virology Centre | Public Health Wales Microbiology Cardiff |
| EPI_ISL_420984 | Wales Specialist Virology Centre | Public Health Wales Microbiology Cardiff |
| EPI_ISL_420985 | Wales Specialist Virology Centre | Public Health Wales Microbiology Cardiff |
| EPI_ISL_420986 | Wales Specialist Virology Centre | Public Health Wales Microbiology Cardiff |
| EPI_ISL_420987 | Wales Specialist Virology Centre | Public Health Wales Microbiology Cardiff |
| EPI_ISL_420988 | Wales Specialist Virology Centre | Public Health Wales Microbiology Cardiff |
| EPI_ISL_420989 | Wales Specialist Virology Centre | Public Health Wales Microbiology Cardiff |
| EPI_ISL_420990 | Wales Specialist Virology Centre | Public Health Wales Microbiology Cardiff |
| EPI_ISL_420991 | Wales Specialist Virology Centre | Public Health Wales Microbiology Cardiff |
| EPI_ISL_420992 | Wales Specialist Virology Centre | Public Health Wales Microbiology Cardiff |
| EPI_ISL_420993 | Wales Specialist Virology Centre | Public Health Wales Microbiology Cardiff |
| EPI_ISL_420994 | Wales Specialist Virology Centre | Public Health Wales Microbiology Cardiff |
| EPI_ISL_420995 | Wales Specialist Virology Centre | Public Health Wales Microbiology Cardiff |
| EPI_ISL_420996 | Wales Specialist Virology Centre | Public Health Wales Microbiology Cardiff |
| EPI_ISL_420997 | Wales Specialist Virology Centre | Public Health Wales Microbiology Cardiff |
| EPI_ISL_420998 | Wales Specialist Virology Centre | Public Health Wales Microbiology Cardiff |
| EPI_ISL_420999 | Wales Specialist Virology Centre | Public Health Wales Microbiology Cardiff |
| EPI_ISL_421000 | Wales Specialist Virology Centre | Public Health Wales Microbiology Cardiff |

| EPI_Accession_ID | Originating Laboratory | Submitting Laboratory |
|---|---|---|
| EPI_ISL_421001 | Wales Specialist Virology Centre | Public Health Wales Microbiology Cardiff |
| EPI_ISL_421002 | Wales Specialist Virology Centre | Public Health Wales Microbiology Cardiff |
| EPI_ISL_421003 | Wales Specialist Virology Centre | Public Health Wales Microbiology Cardiff |
| EPI_ISL_421004 | Wales Specialist Virology Centre | Public Health Wales Microbiology Cardiff |
| EPI_ISL_421005 | Wales Specialist Virology Centre | Public Health Wales Microbiology Cardiff |
| EPI_ISL_421006 | Wales Specialist Virology Centre | Public Health Wales Microbiology Cardiff |
| EPI_ISL_421007 | Wales Specialist Virology Centre | Public Health Wales Microbiology Cardiff |
| EPI_ISL_421008 | Wales Specialist Virology Centre | Public Health Wales Microbiology Cardiff |
| EPI_ISL_421009 | Wales Specialist Virology Centre | Public Health Wales Microbiology Cardiff |
| EPI_ISL_421010 | Wales Specialist Virology Centre | Public Health Wales Microbiology Cardiff |
| EPI_ISL_421011 | Wales Specialist Virology Centre | Public Health Wales Microbiology Cardiff |
| EPI_ISL_421171 | Hospital Universitario 12 de Octubre | Hospital Universitario 12 de Octubre |
| EPI_ISL_421172 | Hospital Universitario 12 de Octubre | Hospital Universitario 12 de Octubre |
| EPI_ISL_421173 | Hospital Universitario 12 de Octubre | Hospital Universitario 12 de Octubre |
| EPI_ISL_421174 | Hospital Universitario 12 de Octubre | Hospital Universitario 12 de Octubre |
| EPI_ISL_421175 | Hospital Universitario 12 de Octubre | Hospital Universitario 12 de Octubre |
| EPI_ISL_421176 | Hospital Universitario 12 de Octubre | Hospital Universitario 12 de Octubre |
| EPI_ISL_421177 | Hospital Universitario 12 de Octubre | Hospital Universitario 12 de Octubre |
| EPI_ISL_421178 | Hospital Universitario 12 de Octubre | Hospital Universitario 12 de Octubre |
| EPI_ISL_421179 | Hospital Universitario 12 de Octubre | Hospital Universitario 12 de Octubre |
| EPI_ISL_421180 | Hospital Universitario 12 de Octubre | Hospital Universitario 12 de Octubre |
| EPI_ISL_421182 | Department of Clinical Microbiology | GIGA Medical Genomics |
| EPI_ISL_421183 | Department of Clinical Microbiology | GIGA Medical Genomics |
| EPI_ISL_421184 | Department of Clinical Microbiology | GIGA Medical Genomics |
| EPI_ISL_421185 | Department of Clinical Microbiology | GIGA Medical Genomics |
| EPI_ISL_421186 | Department of Clinical Microbiology | GIGA Medical Genomics |
| EPI_ISL_421187 | Department of Clinical Microbiology | GIGA Medical Genomics |
| EPI_ISL_421188 | Department of Clinical Microbiology | GIGA Medical Genomics |
| EPI_ISL_421189 | Department of Clinical Microbiology | GIGA Medical Genomics |
| EPI_ISL_421190 | Department of Clinical Microbiology | GIGA Medical Genomics |
| EPI_ISL_421191 | Department of Clinical Microbiology | GIGA Medical Genomics |
| EPI_ISL_421192 | Department of Clinical Microbiology | GIGA Medical Genomics |
| EPI_ISL_421193 | Department of Clinical Microbiology | GIGA Medical Genomics |

| EPI_Accession_ID | Originating Laboratory | Submitting Laboratory |
|---|---|---|
| EPI_ISL_421194 | Department of Clinical Microbiology | GIGA Medical Genomics |
| EPI_ISL_421195 | Department of Clinical Microbiology | GIGA Medical Genomics |
| EPI_ISL_421196 | Department of Clinical Microbiology | GIGA Medical Genomics |
| EPI_ISL_421197 | Department of Clinical Microbiology | GIGA Medical Genomics |
| EPI_ISL_421198 | Department of Clinical Microbiology | GIGA Medical Genomics |
| EPI_ISL_421199 | Department of Clinical Microbiology | GIGA Medical Genomics |
| EPI_ISL_421200 | Department of Clinical Microbiology | GIGA Medical Genomics |
| EPI_ISL_421201 | Department of Clinical Microbiology | GIGA Medical Genomics |
| EPI_ISL_421202 | Department of Clinical Microbiology | GIGA Medical Genomics |
| EPI_ISL_421203 | Department of Clinical Microbiology | GIGA Medical Genomics |
| EPI_ISL_421204 | Department of Clinical Microbiology | GIGA Medical Genomics |
| EPI_ISL_421205 | Department of Clinical Microbiology | GIGA Medical Genomics |
| EPI_ISL_421206 | Department of Clinical Microbiology | GIGA Medical Genomics |
| EPI_ISL_421207 | Department of Clinical Microbiology | GIGA Medical Genomics |
| EPI_ISL_421208 | Department of Clinical Microbiology | GIGA Medical Genomics |
| EPI_ISL_421209 | Department of Clinical Microbiology | GIGA Medical Genomics |
| EPI_ISL_421210 | Department of Clinical Microbiology | GIGA Medical Genomics |
| EPI_ISL_421211 | Department of Clinical Microbiology | GIGA Medical Genomics |
| EPI_ISL_421212 | Department of Clinical Microbiology | GIGA Medical Genomics |
| EPI_ISL_421213 | Department of Clinical Microbiology | GIGA Medical Genomics |
| EPI_ISL_421214 | Department of Clinical Microbiology | GIGA Medical Genomics |
| EPI_ISL_421221 | Hangzhou Center for Diseases Control and Prevention | Hangzhou Center for Diseases Control and Prevention |
| EPI_ISL_421222 | Hangzhou Center for Diseases Control and Prevention | Hangzhou Center for Diseases Control and Prevention |
| EPI_ISL_421223 | Hangzhou Center for Diseases Control and Prevention | Hangzhou Center for Diseases Control and Prevention |
| EPI_ISL_421224 | Hangzhou Center for Diseases Control and Prevention | Hangzhou Center for Diseases Control and Prevention |
| EPI_ISL_421225 | Hangzhou Center for Diseases Control and Prevention | Hangzhou Center for Diseases Control and Prevention |
| EPI_ISL_421226 | Hangzhou Center for Diseases Control and Prevention | Hangzhou Center for Diseases Control and Prevention |
| EPI_ISL_421227 | Hangzhou Center for Diseases Control and Prevention | Hangzhou Center for Diseases Control and Prevention |
| EPI_ISL_421228 | Hangzhou Center for Diseases Control and Prevention | Hangzhou Center for Diseases Control and Prevention |
| EPI_ISL_421229 | Hangzhou Center for Diseases Control and Prevention | Hangzhou Center for Diseases Control and Prevention |
| EPI_ISL_421230 | Hangzhou Center for Diseases Control and Prevention | Hangzhou Center for Diseases Control and Prevention |
| EPI_ISL_421231 | Hangzhou Center for Diseases Control and Prevention | Hangzhou Center for Diseases Control and Prevention |
| EPI_ISL_421232 | Hangzhou Center for Diseases Control and Prevention | Hangzhou Center for Diseases Control and Prevention |

| EPI_Accession_ID | Originating Laboratory | Submitting Laboratory |
|---|---|---|
| EPI_ISL_421233 | Hangzhou Center for Diseases Control and Prevention | Hangzhou Center for Diseases Control and Prevention |
| EPI_ISL_421234 | Hangzhou Center for Diseases Control and Prevention | Hangzhou Center for Diseases Control and Prevention |
| EPI_ISL_421235 | Hangzhou Center for Diseases Control and Prevention | Hangzhou Center for Diseases Control and Prevention |
| EPI_ISL_421236 | Hangzhou Center for Diseases Control and Prevention | Hangzhou Center for Diseases Control and Prevention |
| EPI_ISL_421237 | Jiangxi Province Center for Disease Control and Prevention | Jiangxi Province Center for Disease Control and Prevention |
| EPI_ISL_421238 | Jiangxi Province Center for Disease Control and Prevention | Jiangxi Province Center for Disease Control and Prevention |
| EPI_ISL_421239 | Jiangxi Province Center for Disease Control and Prevention | Jiangxi Province Center for Disease Control and Prevention |
| EPI_ISL_421240 | Jiangxi Province Center for Disease Control and Prevention | Jiangxi Province Center for Disease Control and Prevention |
| EPI_ISL_421241 | Jiangxi Province Center for Disease Control and Prevention | Jiangxi Province Center for Disease Control and Prevention |
| EPI_ISL_421242 | Jiangxi Province Center for Disease Control and Prevention | Jiangxi Province Center for Disease Control and Prevention |
| EPI_ISL_421243 | Jiangxi Province Center for Disease Control and Prevention | Jiangxi Province Center for Disease Control and Prevention |
| EPI_ISL_421244 | Jiangxi Province Center for Disease Control and Prevention | Jiangxi Province Center for Disease Control and Prevention |
| EPI_ISL_421245 | Jiangxi Province Center for Disease Control and Prevention | Jiangxi Province Center for Disease Control and Prevention |
| EPI_ISL_421246 | Jiangxi Province Center for Disease Control and Prevention | Jiangxi Province Center for Disease Control and Prevention |
| EPI_ISL_421247 | Jiangxi Province Center for Disease Control and Prevention | Jiangxi Province Center for Disease Control and Prevention |
| EPI_ISL_421248 | Jiangxi Province Center for Disease Control and Prevention | Jiangxi Province Center for Disease Control and Prevention |
| EPI_ISL_421249 | Jiangxi Province Center for Disease Control and Prevention | Jiangxi Province Center for Disease Control and Prevention |
| EPI_ISL_421250 | Jiangxi Province Center for Disease Control and Prevention | Jiangxi Province Center for Disease Control and Prevention |

| EPI_Accession_ID | Originating Laboratory | Submitting Laboratory |
|---|---|---|
| EPI_ISL_421251 | Jiangxi Province Center for Disease Control and Prevention | Jiangxi Province Center for Disease Control and Prevention |
| EPI_ISL_421252 | Jiangxi Province Center for Disease Control and Prevention | Jiangxi Province Center for Disease Control and Prevention |
| EPI_ISL_421253 | Jiangxi Province Center for Disease Control and Prevention | Jiangxi Province Center for Disease Control and Prevention |
| EPI_ISL_421254 | Jiangxi Province Center for Disease Control and Prevention | Jiangxi Province Center for Disease Control and Prevention |
| EPI_ISL_421256 | Jiangxi Province Center for Disease Control and Prevention | Jiangxi Province Center for Disease Control and Prevention |
| EPI_ISL_421257 | Jiangxi Province Center for Disease Control and Prevention | Jiangxi Province Center for Disease Control and Prevention |
| EPI_ISL_421258 | Jiangxi Province Center for Disease Control and Prevention | Jiangxi Province Center for Disease Control and Prevention |
| EPI_ISL_421259 | Jiangxi Province Center for Disease Control and Prevention | Jiangxi Province Center for Disease Control and Prevention |
| EPI_ISL_421260 | Jiangxi Province Center for Disease Control and Prevention | Jiangxi Province Center for Disease Control and Prevention |
| EPI_ISL_421261 | Jiangxi Province Center for Disease Control and Prevention | Jiangxi Province Center for Disease Control and Prevention |
| EPI_ISL_421262 | Jiangxi Province Center for Disease Control and Prevention | Jiangxi Province Center for Disease Control and Prevention |
| EPI_ISL_421272 | Wyoming Public Health Laboratory | Center for Global Health, University of New Mexico Health Sciences Center |
| EPI_ISL_421275 | Russian State Collection of Viruses | Pathogenic Microorganisms Variability Laboratory |
| EPI_ISL_421279 | Clinical Diagnostics Laboratory, Diagnostic & Experimental Pathology, Lilly Research Laboratories | Clinical Diagnostics Laboratory, Diagnostic & Experimental Pathology, Lilly Research Laboratories |
| EPI_ISL_421281 | Clinical Diagnostics Laboratory, Diagnostic & Experimental Pathology, Lilly Research Laboratories | Clinical Diagnostics Laboratory, Diagnostic & Experimental Pathology, Lilly Research Laboratories |
| EPI_ISL_421283 | University of Wisconsin-Madison AIDS Vaccine Research Laboratories | University of Wisconsin-Madison AIDS Vaccine Research Laboratories |
| EPI_ISL_421284 | University of Wisconsin-Madison AIDS Vaccine Research Laboratories | University of Wisconsin-Madison AIDS Vaccine Research Laboratories |

| EPI_Accession_ID | Originating Laboratory | Submitting Laboratory |
|---|---|---|
| EPI_ISL_421285 | University of Wisconsin-Madison AIDS Vaccine Research Laboratories | University of Wisconsin-Madison AIDS Vaccine Research Laboratories |
| EPI_ISL_421286 | University of Wisconsin-Madison AIDS Vaccine Research Laboratories | University of Wisconsin-Madison AIDS Vaccine Research Laboratories |
| EPI_ISL_421287 | University of Wisconsin-Madison AIDS Vaccine Research Laboratories | University of Wisconsin-Madison AIDS Vaccine Research Laboratories |
| EPI_ISL_421288 | University of Wisconsin-Madison AIDS Vaccine Research Laboratories | University of Wisconsin-Madison AIDS Vaccine Research Laboratories |
| EPI_ISL_421289 | University of Wisconsin-Madison AIDS Vaccine Research Laboratories | University of Wisconsin-Madison AIDS Vaccine Research Laboratories |
| EPI_ISL_421290 | University of Wisconsin-Madison AIDS Vaccine Research Laboratories | University of Wisconsin-Madison AIDS Vaccine Research Laboratories |
| EPI_ISL_421291 | University of Wisconsin-Madison AIDS Vaccine Research Laboratories | University of Wisconsin-Madison AIDS Vaccine Research Laboratories |
| EPI_ISL_421292 | University of Wisconsin-Madison AIDS Vaccine Research Laboratories | University of Wisconsin-Madison AIDS Vaccine Research Laboratories |
| EPI_ISL_421293 | University of Wisconsin-Madison AIDS Vaccine Research Laboratories | University of Wisconsin-Madison AIDS Vaccine Research Laboratories |
| EPI_ISL_421294 | University of Wisconsin-Madison AIDS Vaccine Research Laboratories | University of Wisconsin-Madison AIDS Vaccine Research Laboratories |
| EPI_ISL_421295 | University of Wisconsin-Madison AIDS Vaccine Research Laboratories | University of Wisconsin-Madison AIDS Vaccine Research Laboratories |
| EPI_ISL_421296 | University of Wisconsin-Madison AIDS Vaccine Research Laboratories | University of Wisconsin-Madison AIDS Vaccine Research Laboratories |
| EPI_ISL_421297 | University of Wisconsin-Madison AIDS Vaccine Research Laboratories | University of Wisconsin-Madison AIDS Vaccine Research Laboratories |
| EPI_ISL_421298 | University of Wisconsin-Madison AIDS Vaccine Research Laboratories | University of Wisconsin-Madison AIDS Vaccine Research Laboratories |
| EPI_ISL_421299 | University of Wisconsin-Madison AIDS Vaccine Research Laboratories | University of Wisconsin-Madison AIDS Vaccine Research Laboratories |
| EPI_ISL_421300 | University of Wisconsin-Madison AIDS Vaccine Research Laboratories | University of Wisconsin-Madison AIDS Vaccine Research Laboratories |

| EPI_Accession_ID | Originating Laboratory | Submitting Laboratory |
|---|---|---|
| EPI_ISL_421301 | University of Wisconsin-Madison AIDS Vaccine Research Laboratories | University of Wisconsin-Madison AIDS Vaccine Research Laboratories |
| EPI_ISL_421302 | University of Wisconsin-Madison AIDS Vaccine Research Laboratories | University of Wisconsin-Madison AIDS Vaccine Research Laboratories |
| EPI_ISL_421303 | University of Wisconsin-Madison AIDS Vaccine Research Laboratories | University of Wisconsin-Madison AIDS Vaccine Research Laboratories |
| EPI_ISL_421304 | University of Wisconsin-Madison AIDS Vaccine Research Laboratories | University of Wisconsin-Madison AIDS Vaccine Research Laboratories |
| EPI_ISL_421305 | University of Wisconsin-Madison AIDS Vaccine Research Laboratories | University of Wisconsin-Madison AIDS Vaccine Research Laboratories |
| EPI_ISL_421306 | University of Wisconsin-Madison AIDS Vaccine Research Laboratories | University of Wisconsin-Madison AIDS Vaccine Research Laboratories |
| EPI_ISL_421307 | University of Wisconsin-Madison AIDS Vaccine Research Laboratories | University of Wisconsin-Madison AIDS Vaccine Research Laboratories |
| EPI_ISL_421308 | University of Wisconsin-Madison AIDS Vaccine Research Laboratories | University of Wisconsin-Madison AIDS Vaccine Research Laboratories |
| EPI_ISL_421309 | University of Wisconsin-Madison AIDS Vaccine Research Laboratories | University of Wisconsin-Madison AIDS Vaccine Research Laboratories |
| EPI_ISL_421310 | University of Wisconsin-Madison AIDS Vaccine Research Laboratories | University of Wisconsin-Madison AIDS Vaccine Research Laboratories |
| EPI_ISL_421311 | University of Wisconsin-Madison AIDS Vaccine Research Laboratories | University of Wisconsin-Madison AIDS Vaccine Research Laboratories |
| EPI_ISL_421312 | University of Wisconsin-Madison AIDS Vaccine Research Laboratories | University of Wisconsin-Madison AIDS Vaccine Research Laboratories |
| EPI_ISL_421313 | University of Wisconsin-Madison AIDS Vaccine Research Laboratories | University of Wisconsin-Madison AIDS Vaccine Research Laboratories |
| EPI_ISL_421314 | University of Wisconsin-Madison AIDS Vaccine Research Laboratories | University of Wisconsin-Madison AIDS Vaccine Research Laboratories |
| EPI_ISL_421315 | University of Wisconsin-Madison AIDS Vaccine Research Laboratories | University of Wisconsin-Madison AIDS Vaccine Research Laboratories |
| EPI_ISL_421316 | University of Wisconsin-Madison AIDS Vaccine Research Laboratories | University of Wisconsin-Madison AIDS Vaccine Research Laboratories |

| EPI_Accession_ID | Originating Laboratory | Submitting Laboratory |
|---|---|---|
| EPI_ISL_421317 | University of Wisconsin-Madison AIDS Vaccine Research Laboratories | University of Wisconsin-Madison AIDS Vaccine Research Laboratories |
| EPI_ISL_421318 | University of Wisconsin-Madison AIDS Vaccine Research Laboratories | University of Wisconsin-Madison AIDS Vaccine Research Laboratories |
| EPI_ISL_421319 | University of Wisconsin-Madison AIDS Vaccine Research Laboratories | University of Wisconsin-Madison AIDS Vaccine Research Laboratories |
| EPI_ISL_421320 | University of Wisconsin-Madison AIDS Vaccine Research Laboratories | University of Wisconsin-Madison AIDS Vaccine Research Laboratories |
| EPI_ISL_421321 | University of Wisconsin-Madison AIDS Vaccine Research Laboratories | University of Wisconsin-Madison AIDS Vaccine Research Laboratories |
| EPI_ISL_421322 | University of Wisconsin-Madison AIDS Vaccine Research Laboratories | University of Wisconsin-Madison AIDS Vaccine Research Laboratories |
| EPI_ISL_421323 | University of Wisconsin-Madison AIDS Vaccine Research Laboratories | University of Wisconsin-Madison AIDS Vaccine Research Laboratories |
| EPI_ISL_421324 | University of Wisconsin-Madison AIDS Vaccine Research Laboratories | University of Wisconsin-Madison AIDS Vaccine Research Laboratories |
| EPI_ISL_421325 | University of Wisconsin-Madison AIDS Vaccine Research Laboratories | University of Wisconsin-Madison AIDS Vaccine Research Laboratories |
| EPI_ISL_421326 | University of Wisconsin-Madison AIDS Vaccine Research Laboratories | University of Wisconsin-Madison AIDS Vaccine Research Laboratories |
| EPI_ISL_421327 | University of Wisconsin-Madison AIDS Vaccine Research Laboratories | University of Wisconsin-Madison AIDS Vaccine Research Laboratories |
| EPI_ISL_421328 | University of Wisconsin-Madison AIDS Vaccine Research Laboratories | University of Wisconsin-Madison AIDS Vaccine Research Laboratories |
| EPI_ISL_421329 | University of Wisconsin-Madison AIDS Vaccine Research Laboratories | University of Wisconsin-Madison AIDS Vaccine Research Laboratories |
| EPI_ISL_421330 | University of Wisconsin-Madison AIDS Vaccine Research Laboratories | University of Wisconsin-Madison AIDS Vaccine Research Laboratories |
| EPI_ISL_421331 | University of Wisconsin-Madison AIDS Vaccine Research Laboratories | University of Wisconsin-Madison AIDS Vaccine Research Laboratories |
| EPI_ISL_421332 | University of Wisconsin-Madison AIDS Vaccine Research Laboratories | University of Wisconsin-Madison AIDS Vaccine Research Laboratories |

| EPI_Accession_ID | Originating Laboratory | Submitting Laboratory |
|---|---|---|
| EPI_ISL_421333 | University of Wisconsin-Madison AIDS Vaccine Research Laboratories | University of Wisconsin-Madison AIDS Vaccine Research Laboratories |
| EPI_ISL_421334 | University of Wisconsin-Madison AIDS Vaccine Research Laboratories | University of Wisconsin-Madison AIDS Vaccine Research Laboratories |
| EPI_ISL_421335 | University of Wisconsin-Madison AIDS Vaccine Research Laboratories | University of Wisconsin-Madison AIDS Vaccine Research Laboratories |
| EPI_ISL_421336 | University of Wisconsin-Madison AIDS Vaccine Research Laboratories | University of Wisconsin-Madison AIDS Vaccine Research Laboratories |
| EPI_ISL_421338 | University of Wisconsin-Madison AIDS Vaccine Research Laboratories | University of Wisconsin-Madison AIDS Vaccine Research Laboratories |
| EPI_ISL_421339 | University of Wisconsin-Madison AIDS Vaccine Research Laboratories | University of Wisconsin-Madison AIDS Vaccine Research Laboratories |
| EPI_ISL_421340 | University of Wisconsin-Madison AIDS Vaccine Research Laboratories | University of Wisconsin-Madison AIDS Vaccine Research Laboratories |
| EPI_ISL_421341 | University of Wisconsin-Madison AIDS Vaccine Research Laboratories | University of Wisconsin-Madison AIDS Vaccine Research Laboratories |
| EPI_ISL_421342 | University of Wisconsin-Madison AIDS Vaccine Research Laboratories | University of Wisconsin-Madison AIDS Vaccine Research Laboratories |
| EPI_ISL_421343 | University of Wisconsin-Madison AIDS Vaccine Research Laboratories | University of Wisconsin-Madison AIDS Vaccine Research Laboratories |
| EPI_ISL_421346 | Wyoming Public Health Laboratory | Center for Global Health, University of New Mexico Health Sciences Center |
| EPI_ISL_421347 | Wyoming Public Health Laboratory | Center for Global Health, University of New Mexico Health Sciences Center |
| EPI_ISL_421348 | MSHS Clinical Microbiology Laboratories | MSHS Pathogen Surveillance Program |
| EPI_ISL_421349 | MSHS Clinical Microbiology Laboratories | MSHS Pathogen Surveillance Program |
| EPI_ISL_421350 | MSHS Clinical Microbiology Laboratories | MSHS Pathogen Surveillance Program |
| EPI_ISL_421351 | MSHS Clinical Microbiology Laboratories | MSHS Pathogen Surveillance Program |
| EPI_ISL_421352 | MSHS Clinical Microbiology Laboratories | MSHS Pathogen Surveillance Program |
| EPI_ISL_421353 | MSHS Clinical Microbiology Laboratories | MSHS Pathogen Surveillance Program |
| EPI_ISL_421354 | MSHS Clinical Microbiology Laboratories | MSHS Pathogen Surveillance Program |
| EPI_ISL_421355 | MSHS Clinical Microbiology Laboratories | MSHS Pathogen Surveillance Program |
| EPI_ISL_421356 | MSHS Clinical Microbiology Laboratories | MSHS Pathogen Surveillance Program |

| EPI_Accession_ID | Originating Laboratory | Submitting Laboratory |
|---|---|---|
| EPI_ISL_421357 | MSHS Clinical Microbiology Laboratories | MSHS Pathogen Surveillance Program |
| EPI_ISL_421358 | MSHS Clinical Microbiology Laboratories | MSHS Pathogen Surveillance Program |
| EPI_ISL_421359 | MSHS Clinical Microbiology Laboratories | MSHS Pathogen Surveillance Program |
| EPI_ISL_421360 | MSHS Clinical Microbiology Laboratories | MSHS Pathogen Surveillance Program |
| EPI_ISL_421361 | MSHS Clinical Microbiology Laboratories | MSHS Pathogen Surveillance Program |
| EPI_ISL_421362 | MSHS Clinical Microbiology Laboratories | MSHS Pathogen Surveillance Program |
| EPI_ISL_421363 | MSHS Clinical Microbiology Laboratories | MSHS Pathogen Surveillance Program |
| EPI_ISL_421364 | MSHS Clinical Microbiology Laboratories | MSHS Pathogen Surveillance Program |
| EPI_ISL_421365 | MSHS Clinical Microbiology Laboratories | MSHS Pathogen Surveillance Program |
| EPI_ISL_421366 | MSHS Clinical Microbiology Laboratories | MSHS Pathogen Surveillance Program |
| EPI_ISL_421367 | MSHS Clinical Microbiology Laboratories | MSHS Pathogen Surveillance Program |
| EPI_ISL_421368 | MSHS Clinical Microbiology Laboratories | MSHS Pathogen Surveillance Program |
| EPI_ISL_421369 | MSHS Clinical Microbiology Laboratories | MSHS Pathogen Surveillance Program |
| EPI_ISL_421370 | MSHS Clinical Microbiology Laboratories | MSHS Pathogen Surveillance Program |
| EPI_ISL_421371 | MSHS Clinical Microbiology Laboratories | MSHS Pathogen Surveillance Program |
| EPI_ISL_421372 | MSHS Clinical Microbiology Laboratories | MSHS Pathogen Surveillance Program |
| EPI_ISL_421373 | MSHS Clinical Microbiology Laboratories | MSHS Pathogen Surveillance Program |
| EPI_ISL_421374 | MSHS Clinical Microbiology Laboratories | MSHS Pathogen Surveillance Program |
| EPI_ISL_421375 | MSHS Clinical Microbiology Laboratories | MSHS Pathogen Surveillance Program |
| EPI_ISL_421376 | MSHS Clinical Microbiology Laboratories | MSHS Pathogen Surveillance Program |
| EPI_ISL_421377 | MSHS Clinical Microbiology Laboratories | MSHS Pathogen Surveillance Program |
| EPI_ISL_421378 | MSHS Clinical Microbiology Laboratories | MSHS Pathogen Surveillance Program |
| EPI_ISL_421379 | MSHS Clinical Microbiology Laboratories | MSHS Pathogen Surveillance Program |
| EPI_ISL_421380 | MSHS Clinical Microbiology Laboratories | MSHS Pathogen Surveillance Program |
| EPI_ISL_421381 | MSHS Clinical Microbiology Laboratories | MSHS Pathogen Surveillance Program |
| EPI_ISL_421382 | MSHS Clinical Microbiology Laboratories | MSHS Pathogen Surveillance Program |
| EPI_ISL_421383 | MSHS Clinical Microbiology Laboratories | MSHS Pathogen Surveillance Program |
| EPI_ISL_421384 | MSHS Clinical Microbiology Laboratories | MSHS Pathogen Surveillance Program |
| EPI_ISL_421385 | MSHS Clinical Microbiology Laboratories | MSHS Pathogen Surveillance Program |
| EPI_ISL_421386 | MSHS Clinical Microbiology Laboratories | MSHS Pathogen Surveillance Program |
| EPI_ISL_421387 | MSHS Clinical Microbiology Laboratories | MSHS Pathogen Surveillance Program |
| EPI_ISL_421388 | MSHS Clinical Microbiology Laboratories | MSHS Pathogen Surveillance Program |
| EPI_ISL_421389 | MSHS Clinical Microbiology Laboratories | MSHS Pathogen Surveillance Program |

| EPI_Accession_ID | Originating Laboratory | Submitting Laboratory |
|---|---|---|
| EPI_ISL_421390 | MSHS Clinical Microbiology Laboratories | MSHS Pathogen Surveillance Program |
| EPI_ISL_421391 | MSHS Clinical Microbiology Laboratories | MSHS Pathogen Surveillance Program |
| EPI_ISL_421392 | MSHS Clinical Microbiology Laboratories | MSHS Pathogen Surveillance Program |
| EPI_ISL_421393 | MSHS Clinical Microbiology Laboratories | MSHS Pathogen Surveillance Program |
| EPI_ISL_421394 | MSHS Clinical Microbiology Laboratories | MSHS Pathogen Surveillance Program |
| EPI_ISL_421395 | MSHS Clinical Microbiology Laboratories | MSHS Pathogen Surveillance Program |
| EPI_ISL_421396 | MSHS Clinical Microbiology Laboratories | MSHS Pathogen Surveillance Program |
| EPI_ISL_421397 | MSHS Clinical Microbiology Laboratories | MSHS Pathogen Surveillance Program |
| EPI_ISL_421398 | MSHS Clinical Microbiology Laboratories | MSHS Pathogen Surveillance Program |
| EPI_ISL_421399 | MSHS Clinical Microbiology Laboratories | MSHS Pathogen Surveillance Program |
| EPI_ISL_421400 | MSHS Clinical Microbiology Laboratories | MSHS Pathogen Surveillance Program |
| EPI_ISL_421401 | MSHS Clinical Microbiology Laboratories | MSHS Pathogen Surveillance Program |
| EPI_ISL_421402 | MSHS Clinical Microbiology Laboratories | MSHS Pathogen Surveillance Program |
| EPI_ISL_421403 | MSHS Clinical Microbiology Laboratories | MSHS Pathogen Surveillance Program |
| EPI_ISL_421404 | MSHS Clinical Microbiology Laboratories | MSHS Pathogen Surveillance Program |
| EPI_ISL_421405 | MSHS Clinical Microbiology Laboratories | MSHS Pathogen Surveillance Program |
| EPI_ISL_421406 | MSHS Clinical Microbiology Laboratories | MSHS Pathogen Surveillance Program |
| EPI_ISL_421407 | MSHS Clinical Microbiology Laboratories | MSHS Pathogen Surveillance Program |
| EPI_ISL_421408 | MSHS Clinical Microbiology Laboratories | MSHS Pathogen Surveillance Program |
| EPI_ISL_421409 | MSHS Clinical Microbiology Laboratories | MSHS Pathogen Surveillance Program |
| EPI_ISL_421410 | MSHS Clinical Microbiology Laboratories | MSHS Pathogen Surveillance Program |
| EPI_ISL_421411 | MSHS Clinical Microbiology Laboratories | MSHS Pathogen Surveillance Program |
| EPI_ISL_421412 | MSHS Clinical Microbiology Laboratories | MSHS Pathogen Surveillance Program |
| EPI_ISL_421413 | MSHS Clinical Microbiology Laboratories | MSHS Pathogen Surveillance Program |
| EPI_ISL_421414 | MSHS Clinical Microbiology Laboratories | MSHS Pathogen Surveillance Program |
| EPI_ISL_421415 | MSHS Clinical Microbiology Laboratories | MSHS Pathogen Surveillance Program |
| EPI_ISL_421416 | MSHS Clinical Microbiology Laboratories | MSHS Pathogen Surveillance Program |
| EPI_ISL_421417 | MSHS Clinical Microbiology Laboratories | MSHS Pathogen Surveillance Program |
| EPI_ISL_421418 | MSHS Clinical Microbiology Laboratories | MSHS Pathogen Surveillance Program |
| EPI_ISL_421419 | MSHS Clinical Microbiology Laboratories | MSHS Pathogen Surveillance Program |
| EPI_ISL_421420 | MSHS Clinical Microbiology Laboratories | MSHS Pathogen Surveillance Program |
| EPI_ISL_421421 | MSHS Clinical Microbiology Laboratories | MSHS Pathogen Surveillance Program |
| EPI_ISL_421422 | MSHS Clinical Microbiology Laboratories | MSHS Pathogen Surveillance Program |

| EPI_Accession_ID | Originating Laboratory | Submitting Laboratory |
|---|---|---|
| EPI_ISL_421423 | MSHS Clinical Microbiology Laboratories | MSHS Pathogen Surveillance Program |
| EPI_ISL_421424 | MSHS Clinical Microbiology Laboratories | MSHS Pathogen Surveillance Program |
| EPI_ISL_421425 | MSHS Clinical Microbiology Laboratories | MSHS Pathogen Surveillance Program |
| EPI_ISL_421426 | MSHS Clinical Microbiology Laboratories | MSHS Pathogen Surveillance Program |
| EPI_ISL_421427 | MSHS Clinical Microbiology Laboratories | MSHS Pathogen Surveillance Program |
| EPI_ISL_421428 | MSHS Clinical Microbiology Laboratories | MSHS Pathogen Surveillance Program |
| EPI_ISL_421429 | MSHS Clinical Microbiology Laboratories | MSHS Pathogen Surveillance Program |
| EPI_ISL_421430 | MSHS Clinical Microbiology Laboratories | MSHS Pathogen Surveillance Program |
| EPI_ISL_421431 | MSHS Clinical Microbiology Laboratories | MSHS Pathogen Surveillance Program |
| EPI_ISL_421432 | MSHS Clinical Microbiology Laboratories | MSHS Pathogen Surveillance Program |
| EPI_ISL_421433 | MSHS Clinical Microbiology Laboratories | MSHS Pathogen Surveillance Program |
| EPI_ISL_421434 | MSHS Clinical Microbiology Laboratories | MSHS Pathogen Surveillance Program |
| EPI_ISL_421435 | MSHS Clinical Microbiology Laboratories | MSHS Pathogen Surveillance Program |
| EPI_ISL_421446 | H Guimaraes | Instituto Nacional de Saude (INSA) |
| EPI_ISL_421447 | H Guimaraes | Instituto Nacional de Saude (INSA) |
| EPI_ISL_421448 | H Guimaraes | Instituto Nacional de Saude (INSA) |
| EPI_ISL_421449 | H Dr. Nelio Mendonca - Funchal | Instituto Nacional de Saude (INSA) |
| EPI_ISL_421450 | Instituto Nacional de Saude (INSA) | Instituto Nacional de Saude (INSA) |
| EPI_ISL_421451 | Instituto Nacional de Saude (INSA) | Instituto Nacional de Saude (INSA) |
| EPI_ISL_421452 | Instituto Nacional de Saude (INSA) | Instituto Nacional de Saude (INSA) |
| EPI_ISL_421453 | CHTMAD | Instituto Nacional de Saude (INSA) |
| EPI_ISL_421454 | H Beatriz Angelo | Instituto Nacional de Saude (INSA) |
| EPI_ISL_421455 | CH Barreiro Montijo | Instituto Nacional de Saude (INSA) |
| EPI_ISL_421456 | Instituto Nacional de Saude (INSA) | Instituto Nacional de Saude (INSA) |
| EPI_ISL_421457 | H Dr Nelio Mendonca - Funchal | Instituto Nacional de Saude (INSA) |
| EPI_ISL_421458 | H Beatriz Angelo | Instituto Nacional de Saude (INSA) |
| EPI_ISL_421459 | H Dr. Nelio Mendonca - Funchal | Instituto Nacional de Saude (INSA) |
| EPI_ISL_421460 | H Dr. Nelio Mendonca - Funchal | Instituto Nacional de Saude (INSA) |
| EPI_ISL_421461 | H Dr. Nelio Mendonca - Funchal | Instituto Nacional de Saude (INSA) |
| EPI_ISL_421462 | H Santarem | Instituto Nacional de Saude (INSA) |
| EPI_ISL_421463 | HSE Ilha Terceira - Angra do Heroismo | Instituto Nacional de Saude (INSA) |
| EPI_ISL_421464 | CHTMAD | Instituto Nacional de Saude (INSA) |
| EPI_ISL_421465 | CHTMAD | Instituto Nacional de Saude (INSA) |

| EPI_Accession_ID | Originating Laboratory | Submitting Laboratory |
|---|---|---|
| EPI_ISL_421466 | H Evora | Instituto Nacional de Saude (INSA) |
| EPI_ISL_421467 | H Evora | Instituto Nacional de Saude (INSA) |
| EPI_ISL_421468 | H Santarem | Instituto Nacional de Saude (INSA) |
| EPI_ISL_421469 | H Santarem | Instituto Nacional de Saude (INSA) |
| EPI_ISL_421470 | H Santarem | Instituto Nacional de Saude (INSA) |
| EPI_ISL_421471 | H Santarem | Instituto Nacional de Saude (INSA) |
| EPI_ISL_421472 | Instituto Nacional de Saude (INSA) | Instituto Nacional de Saude (INSA) |
| EPI_ISL_421473 | Instituto Nacional de Saude (INSA) | Instituto Nacional de Saude (INSA) |
| EPI_ISL_421474 | Instituto Nacional de Saude (INSA) | Instituto Nacional de Saude (INSA) |
| EPI_ISL_421475 | Instituto Nacional de Saude (INSA) | Instituto Nacional de Saude (INSA) |
| EPI_ISL_421476 | Instituto Nacional de Saude (INSA) | Instituto Nacional de Saude (INSA) |
| EPI_ISL_421477 | Instituto Nacional de Saude (INSA) | Instituto Nacional de Saude (INSA) |
| EPI_ISL_421478 | Instituto Nacional de Saude (INSA) | Instituto Nacional de Saude (INSA) |
| EPI_ISL_421479 | CH Barreiro Montijo | Instituto Nacional de Saude (INSA) |
| EPI_ISL_421480 | CH Barreiro Montijo | Instituto Nacional de Saude (INSA) |
| EPI_ISL_421481 | H Beatriz Angelo | Instituto Nacional de Saude (INSA) |
| EPI_ISL_421482 | CH VN Gaia - Espinho | Instituto Nacional de Saude (INSA) |
| EPI_ISL_421483 | CH VN Gaia - Espinho | Instituto Nacional de Saude (INSA) |
| EPI_ISL_421484 | CH VN Gaia - Espinho | Instituto Nacional de Saude (INSA) |
| EPI_ISL_421485 | CH Barreiro Montijo | Instituto Nacional de Saude (INSA) |
| EPI_ISL_421486 | CH Barreiro Montijo | Instituto Nacional de Saude (INSA) |
| EPI_ISL_421487 | H Beatriz Angelo | Instituto Nacional de Saude (INSA) |
| EPI_ISL_421488 | H Santarem | Instituto Nacional de Saude (INSA) |
| EPI_ISL_421489 | HSE Ilha Terceira - Angra do Heroismo | Instituto Nacional de Saude (INSA) |
| EPI_ISL_421490 | H Santarem | Instituto Nacional de Saude (INSA) |
| EPI_ISL_421491 | H Beatriz Angelo | Instituto Nacional de Saude (INSA) |
| EPI_ISL_421492 | H Santarem | Instituto Nacional de Saude (INSA) |
| EPI_ISL_421493 | HSE Ilha Terceira - Angra do Heroismo | Instituto Nacional de Saude (INSA) |
| EPI_ISL_421494 | HSE Ilha Terceira - Angra do Heroismo | Instituto Nacional de Saude (INSA) |
| EPI_ISL_421495 | H Santarem | Instituto Nacional de Saude (INSA) |
| EPI_ISL_421496 | Instituto Nacional de Saude (INSA) | Instituto Nacional de Saude (INSA) |
| EPI_ISL_421497 | Instituto Nacional de Saude (INSA) | Instituto Nacional de Saude (INSA) |
| EPI_ISL_421498 | Instituto Nacional de Saude (INSA) | Instituto Nacional de Saude (INSA) |

| EPI_Accession_ID | Originating Laboratory | Submitting Laboratory |
|---|---|---|
| EPI_ISL_421499 | Instituto Nacional de Saude (INSA) | Instituto Nacional de Saude (INSA) |
| EPI_ISL_421500 | CH CompiÃ¨gne Laboratoire de Biologie | National Reference Center for Viruses of Respiratory Infections, Institut Pasteur, Paris |
| EPI_ISL_421501 | Service de Biologie MÃ©dicale - BP 125 | National Reference Center for Viruses of Respiratory Infections, Institut Pasteur, Paris |
| EPI_ISL_421502 | Parc des Dames | National Reference Center for Viruses of Respiratory Infections, Institut Pasteur, Paris |
| EPI_ISL_421503 | Parc des Dames | National Reference Center for Viruses of Respiratory Infections, Institut Pasteur, Paris |
| EPI_ISL_421504 | Service de Biologie MÃ©dicale - BP 125 | National Reference Center for Viruses of Respiratory Infections, Institut Pasteur, Paris |
| EPI_ISL_421505 | Service de Biologie MÃ©dicale - BP 125 | National Reference Center for Viruses of Respiratory Infections, Institut Pasteur, Paris |
| EPI_ISL_421506 | Service de Biologie MÃ©dicale - BP 125 | National Reference Center for Viruses of Respiratory Infections, Institut Pasteur, Paris |
| EPI_ISL_421507 | Le ChÃ¢teau de Seine-Port | National Reference Center for Viruses of Respiratory Infections, Institut Pasteur, Paris |
| EPI_ISL_421508 | Le ChÃ¢teau de Seine-Port | National Reference Center for Viruses of Respiratory Infections, Institut Pasteur, Paris |
| EPI_ISL_421509 | CH CompiÃ¨gne Laboratoire de Biologie | National Reference Center for Viruses of Respiratory Infections, Institut Pasteur, Paris |
| EPI_ISL_421510 | CH CompiÃ¨gne Laboratoire de Biologie | National Reference Center for Viruses of Respiratory Infections, Institut Pasteur, Paris |
| EPI_ISL_421511 | CH CompiÃ¨gne Laboratoire de Biologie | National Reference Center for Viruses of Respiratory Infections, Institut Pasteur, Paris |
| EPI_ISL_421512 | Service de Biologie MÃ©dicale - BP 125 | National Reference Center for Viruses of Respiratory Infections, Institut Pasteur, Paris |
| EPI_ISL_421513 | Service de Biologie clinique | National Reference Center for Viruses of Respiratory Infections, Institut Pasteur, Paris |
| EPI_ISL_421514 | Sentinelles network | National Reference Center for Viruses of Respiratory Infections, Institut Pasteur, Paris |
| EPI_ISL_421515 | Servicio de Microbiologia. Hospital Clinico Universitario de Valencia | Sequencing and Bioinformatics Service and Molecular Epidemiology Research Group. FISABIO-Public Health |

| EPI_Accession_ID | Originating Laboratory | Submitting Laboratory |
|---|---|---|
| EPI_ISL_421516 | Servicio de Microbiologia. Hospital Clinico Universitario de Valencia | Sequencing and Bioinformatics Service and Molecular Epidemiology Research Group. FISABIO-Public Health |
| EPI_ISL_421517 | Servicio de Microbiologia. Hospital Clinico Universitario de Valencia | Sequencing and Bioinformatics Service and Molecular Epidemiology Research Group. FISABIO-Public Health |
| EPI_ISL_421518 | Servicio de Microbiologia. Hospital Clinico Universitario de Valencia | Sequencing and Bioinformatics Service and Molecular Epidemiology Research Group. FISABIO-Public Health |
| EPI_ISL_421519 | Servicio de Microbiologia. Hospital Clinico Universitario de Valencia | Sequencing and Bioinformatics Service and Molecular Epidemiology Research Group. FISABIO-Public Health |
| EPI_ISL_421520 | Servicio de Microbiologia. Hospital Clinico Universitario de Valencia | Sequencing and Bioinformatics Service and Molecular Epidemiology Research Group. FISABIO-Public Health |
| EPI_ISL_421543 | Wyoming Public Health Laboratory | Center for Global Health, University of New Mexico Health Sciences Center |
| EPI_ISL_421544 | Wyoming Public Health Laboratory | Center for Global Health, University of New Mexico Health Sciences Center |
| EPI_ISL_421545 | Wyoming Public Health Laboratory | Center for Global Health, University of New Mexico Health Sciences Center |
| EPI_ISL_421547 | Wyoming Public Health Laboratory | Center for Global Health, University of New Mexico Health Sciences Center |
| EPI_ISL_421548 | Wyoming Public Health Laboratory | Center for Global Health, University of New Mexico Health Sciences Center |
| EPI_ISL_421549 | Wyoming Public Health Laboratory | Center for Global Health, University of New Mexico Health Sciences Center |
| EPI_ISL_421550 | Wyoming Public Health Laboratory | Center for Global Health, University of New Mexico Health Sciences Center |
| EPI_ISL_421551 | Wyoming Public Health Laboratory | Center for Global Health, University of New Mexico Health Sciences Center |
| EPI_ISL_421552 | Wyoming Public Health Laboratory | Center for Global Health, University of New Mexico Health Sciences Center |
| EPI_ISL_421553 | Wyoming Public Health Laboratory | Center for Global Health, University of New Mexico Health Sciences Center |
| EPI_ISL_421560 | Utah Public Health Laboratory | Utah Public Health Laboratory |
| EPI_ISL_421561 | Utah Public Health Laboratory | Utah Public Health Laboratory |
| EPI_ISL_421562 | Utah Public Health Laboratory | Utah Public Health Laboratory |

| EPI_Accession_ID | Originating Laboratory | Submitting Laboratory |
|---|---|---|
| EPI_ISL_421563 | Utah Public Health Laboratory | Utah Public Health Laboratory |
| EPI_ISL_421572 | Molecular Diagnostic Services and FLowpath | KRISP, KZN Research Innovation and Sequencing Platform |
| EPI_ISL_421573 | Molecular Diagnostic Services | KRISP, KZN Research Innovation and Sequencing Platform |
| EPI_ISL_421574 | Molecular Diagnostic Services | KRISP, KZN Research Innovation and Sequencing Platform |
| EPI_ISL_421575 | Molecular Diagnostic Services | KRISP, KZN Research Innovation and Sequencing Platform |
| EPI_ISL_421576 | Molecular Diagnostic Services | KRISP, KZN Research Innovation and Sequencing Platform |
| EPI_ISL_421577 | NYU Langone Health | Departments of Pathology and Medicine, New York University School of Medicine |
| EPI_ISL_421578 | NYU Langone Health | Departments of Pathology and Medicine, New York University School of Medicine |
| EPI_ISL_421579 | NYU Langone Health | Departments of Pathology and Medicine, New York University School of Medicine |
| EPI_ISL_421580 | NYU Langone Health | Departments of Pathology and Medicine, New York University School of Medicine |
| EPI_ISL_421581 | NYU Langone Health | Departments of Pathology and Medicine, New York University School of Medicine |
| EPI_ISL_421582 | NYU Langone Health | Departments of Pathology and Medicine, New York University School of Medicine |
| EPI_ISL_421583 | NYU Langone Health | Departments of Pathology and Medicine, New York University School of Medicine |
| EPI_ISL_421584 | NYU Langone Health | Departments of Pathology and Medicine, New York University School of Medicine |
| EPI_ISL_421585 | NYU Langone Health | Departments of Pathology and Medicine, New York University School of Medicine |
| EPI_ISL_421586 | NYU Langone Health | Departments of Pathology and Medicine, New York University School of Medicine |
| EPI_ISL_421587 | NYU Langone Health | Departments of Pathology and Medicine, New York University School of Medicine |

| EPI_Accession_ID | Originating Laboratory | Submitting Laboratory |
|---|---|---|
| EPI_ISL_421588 | NYU Langone Health | Departments of Pathology and Medicine, New York University School of Medicine |
| EPI_ISL_421589 | NYU Langone Health | Departments of Pathology and Medicine, New York University School of Medicine |
| EPI_ISL_421590 | NYU Langone Health | Departments of Pathology and Medicine, New York University School of Medicine |
| EPI_ISL_421591 | NYU Langone Health | Departments of Pathology and Medicine, New York University School of Medicine |
| EPI_ISL_421592 | NYU Langone Health | Departments of Pathology and Medicine, New York University School of Medicine |
| EPI_ISL_421593 | MSHS Clinical Microbiology Laboratories | MSHS Pathogen Surveillance Program |
| EPI_ISL_421594 | MSHS Clinical Microbiology Laboratories | MSHS Pathogen Surveillance Program |
| EPI_ISL_421595 | MSHS Clinical Microbiology Laboratories | MSHS Pathogen Surveillance Program |
| EPI_ISL_421596 | MSHS Clinical Microbiology Laboratories | MSHS Pathogen Surveillance Program |
| EPI_ISL_421597 | MSHS Clinical Microbiology Laboratories | MSHS Pathogen Surveillance Program |
| EPI_ISL_421598 | MSHS Clinical Microbiology Laboratories | MSHS Pathogen Surveillance Program |
| EPI_ISL_421599 | MSHS Clinical Microbiology Laboratories | MSHS Pathogen Surveillance Program |
| EPI_ISL_421600 | MSHS Clinical Microbiology Laboratories | MSHS Pathogen Surveillance Program |
| EPI_ISL_421601 | MSHS Clinical Microbiology Laboratories | MSHS Pathogen Surveillance Program |
| EPI_ISL_421602 | MSHS Clinical Microbiology Laboratories | MSHS Pathogen Surveillance Program |
| EPI_ISL_421603 | MSHS Clinical Microbiology Laboratories | MSHS Pathogen Surveillance Program |
| EPI_ISL_421604 | MSHS Clinical Microbiology Laboratories | MSHS Pathogen Surveillance Program |
| EPI_ISL_421605 | MSHS Clinical Microbiology Laboratories | MSHS Pathogen Surveillance Program |
| EPI_ISL_421606 | MSHS Clinical Microbiology Laboratories | MSHS Pathogen Surveillance Program |
| EPI_ISL_421607 | MSHS Clinical Microbiology Laboratories | MSHS Pathogen Surveillance Program |
| EPI_ISL_421608 | MSHS Clinical Microbiology Laboratories | MSHS Pathogen Surveillance Program |
| EPI_ISL_421609 | MSHS Clinical Microbiology Laboratories | MSHS Pathogen Surveillance Program |
| EPI_ISL_421610 | MSHS Clinical Microbiology Laboratories | MSHS Pathogen Surveillance Program |
| EPI_ISL_421611 | MSHS Clinical Microbiology Laboratories | MSHS Pathogen Surveillance Program |
| EPI_ISL_421612 | MSHS Clinical Microbiology Laboratories | MSHS Pathogen Surveillance Program |
| EPI_ISL_421613 | MSHS Clinical Microbiology Laboratories | MSHS Pathogen Surveillance Program |
| EPI_ISL_421614 | MSHS Clinical Microbiology Laboratories | MSHS Pathogen Surveillance Program |
| EPI_ISL_421615 | MSHS Clinical Microbiology Laboratories | MSHS Pathogen Surveillance Program |

| EPI_Accession_ID | Originating Laboratory | Submitting Laboratory |
|---|---|---|
| EPI_ISL_421616 | MSHS Clinical Microbiology Laboratories | MSHS Pathogen Surveillance Program |
| EPI_ISL_421617 | MSHS Clinical Microbiology Laboratories | MSHS Pathogen Surveillance Program |
| EPI_ISL_421618 | MSHS Clinical Microbiology Laboratories | MSHS Pathogen Surveillance Program |
| EPI_ISL_421619 | MSHS Clinical Microbiology Laboratories | MSHS Pathogen Surveillance Program |
| EPI_ISL_421620 | MSHS Clinical Microbiology Laboratories | MSHS Pathogen Surveillance Program |
| EPI_ISL_421621 | MSHS Clinical Microbiology Laboratories | MSHS Pathogen Surveillance Program |
| EPI_ISL_421622 | MSHS Clinical Microbiology Laboratories | MSHS Pathogen Surveillance Program |
| EPI_ISL_421623 | MSHS Clinical Microbiology Laboratories | MSHS Pathogen Surveillance Program |
| EPI_ISL_421624 | MSHS Clinical Microbiology Laboratories | MSHS Pathogen Surveillance Program |
| EPI_ISL_421625 | MSHS Clinical Microbiology Laboratories | MSHS Pathogen Surveillance Program |
| EPI_ISL_421626 | MSHS Clinical Microbiology Laboratories | MSHS Pathogen Surveillance Program |
| EPI_ISL_421627 | MSHS Clinical Microbiology Laboratories | MSHS Pathogen Surveillance Program |
| EPI_ISL_421628 | MSHS Clinical Microbiology Laboratories | MSHS Pathogen Surveillance Program |
| EPI_ISL_421629 | MSHS Clinical Microbiology Laboratories | MSHS Pathogen Surveillance Program |
| EPI_ISL_421630 | MSHS Clinical Microbiology Laboratories | MSHS Pathogen Surveillance Program |
| EPI_ISL_421631 | MSHS Clinical Microbiology Laboratories | MSHS Pathogen Surveillance Program |
| EPI_ISL_421632 | MSHS Clinical Microbiology Laboratories | MSHS Pathogen Surveillance Program |
| EPI_ISL_421633 | MSHS Clinical Microbiology Laboratories | MSHS Pathogen Surveillance Program |
| EPI_ISL_421634 | MSHS Clinical Microbiology Laboratories | MSHS Pathogen Surveillance Program |
| EPI_ISL_421635 | MSHS Clinical Microbiology Laboratories | MSHS Pathogen Surveillance Program |
| EPI_ISL_421636 | Pathology North | Public Health Virology Laboratory |
| EPI_ISL_421641 | Centers for Disease Control, R.O.C. (Taiwan) | Centers for Disease Control, R.O.C. (Taiwan) |
| EPI_ISL_421651 | Centers for Disease Control, R.O.C. (Taiwan) | Centers for Disease Control, R.O.C. (Taiwan) |
| EPI_ISL_421652 | Dasman Diabetes Institute | Dasman Diabetes Institute |
| EPI_ISL_421653 | E. Gulbja Laboratorija | Latvian Biomedical Research and Study Centre |
| EPI_ISL_421654 | E. Gulbja Laboratorija | Latvian Biomedical Research and Study Centre |
| EPI_ISL_421655 | E. Gulbja Laboratorija | Latvian Biomedical Research and Study Centre |
| EPI_ISL_421656 | E. Gulbja Laboratorija | Latvian Biomedical Research and Study Centre |
| EPI_ISL_421660 | The Ohio State University | The Ohio State University-James Molecular Lab at Polaris |
| EPI_ISL_421662 | National Influenza Center, Indian Council of Medical Research - National Institute of Virology | Indian Council of Medical Research-National Institute of Virology, Microbial Containment Complex |

| EPI_Accession_ID | Originating Laboratory | Submitting Laboratory |
|---|---|---|
| EPI_ISL_421663 | National Influenza Center, Indian Council of Medical Research - National Institute of Virology | Indian Council of Medical Research-National Institute of Virology, Microbial Containment Complex |
| EPI_ISL_421664 | National Influenza Center, Indian Council of Medical Research - National Institute of Virology | Indian Council of Medical Research-National Institute of Virology, Microbial Containment Complex |
| EPI_ISL_421665 | National Influenza Center, Indian Council of Medical Research - National Institute of Virology | Indian Council of Medical Research-National Institute of Virology, Microbial Containment Complex |
| EPI_ISL_421666 | National Influenza Center, Indian Council of Medical Research - National Institute of Virology | Indian Council of Medical Research-National Institute of Virology, Microbial Containment Complex |
| EPI_ISL_421667 | National Influenza Center, Indian Council of Medical Research - National Institute of Virology | Indian Council of Medical Research-National Institute of Virology, Microbial Containment Complex |
| EPI_ISL_421668 | National Influenza Center, Indian Council of Medical Research - National Institute of Virology | Indian Council of Medical Research-National Institute of Virology, Microbial Containment Complex |
| EPI_ISL_421669 | National Influenza Center, Indian Council of Medical Research - National Institute of Virology | Indian Council of Medical Research-National Institute of Virology, Microbial Containment Complex |
| EPI_ISL_421670 | National Influenza Center, Indian Council of Medical Research - National Institute of Virology | Indian Council of Medical Research-National Institute of Virology, Microbial Containment Complex |
| EPI_ISL_421671 | National Influenza Center, Indian Council of Medical Research - National Institute of Virology | Indian Council of Medical Research-National Institute of Virology, Microbial Containment Complex |
| EPI_ISL_421672 | National Influenza Center, Indian Council of Medical Research - National Institute of Virology | Indian Council of Medical Research-National Institute of Virology, Microbial Containment Complex |
| EPI_ISL_421675 | The Ohio State University Wexner Medical Center | The Ohio State University James Molecular lab |
| EPI_ISL_421683 | Minnesota Department of Health, Public Health Laboratory | Minnesota Department of Health, Public Health Laboratory |
| EPI_ISL_421684 | Minnesota Department of Health, Public Health Laboratory | Minnesota Department of Health, Public Health Laboratory |
| EPI_ISL_421685 | Minnesota Department of Health, Public Health Laboratory | Minnesota Department of Health, Public Health Laboratory |
| EPI_ISL_421686 | Minnesota Department of Health, Public Health Laboratory | Minnesota Department of Health, Public Health Laboratory |
| EPI_ISL_421687 | Minnesota Department of Health, Public Health Laboratory | Minnesota Department of Health, Public Health Laboratory |
| EPI_ISL_421688 | Minnesota Department of Health, Public Health Laboratory | Minnesota Department of Health, Public Health Laboratory |

| EPI_Accession_ID | Originating Laboratory | Submitting Laboratory |
|---|---|---|
| EPI_ISL_421689 | Minnesota Department of Health, Public Health Laboratory | Minnesota Department of Health, Public Health Laboratory |
| EPI_ISL_421690 | Minnesota Department of Health, Public Health Laboratory | Minnesota Department of Health, Public Health Laboratory |
| EPI_ISL_421691 | Minnesota Department of Health, Public Health Laboratory | Minnesota Department of Health, Public Health Laboratory |
| EPI_ISL_421692 | Minnesota Department of Health, Public Health Laboratory | Minnesota Department of Health, Public Health Laboratory |
| EPI_ISL_421704 | NYU Langone Health | Departments of Pathology and Medicine, New York University School of Medicine |
| EPI_ISL_421705 | NYU Langone Health | Departments of Pathology and Medicine, New York University School of Medicine |
| EPI_ISL_421706 | NYU Langone Health | Departments of Pathology and Medicine, New York University School of Medicine |
| EPI_ISL_421707 | NYU Langone Health | Departments of Pathology and Medicine, New York University School of Medicine |
| EPI_ISL_421708 | NYU Langone Health | Departments of Pathology and Medicine, New York University School of Medicine |
| EPI_ISL_421709 | NYU Langone Health | Departments of Pathology and Medicine, New York University School of Medicine |
| EPI_ISL_421710 | NYU Langone Health | Departments of Pathology and Medicine, New York University School of Medicine |
| EPI_ISL_421711 | NYU Langone Health | Departments of Pathology and Medicine, New York University School of Medicine |
| EPI_ISL_421712 | NYU Langone Health | Departments of Pathology and Medicine, New York University School of Medicine |
| EPI_ISL_421713 | NYU Langone Health | Departments of Pathology and Medicine, New York University School of Medicine |
| EPI_ISL_421714 | NYU Langone Health | Departments of Pathology and Medicine, New York University School of Medicine |
| EPI_ISL_421715 | NYU Langone Health | Departments of Pathology and Medicine, New York University School of Medicine |

| EPI_Accession_ID | Originating Laboratory | Submitting Laboratory |
|---|---|---|
| EPI_ISL_421716 | NYU Langone Health | Departments of Pathology and Medicine, New York University School of Medicine |
| EPI_ISL_421717 | NYU Langone Health | Departments of Pathology and Medicine, New York University School of Medicine |
| EPI_ISL_421718 | NYU Langone Health | Departments of Pathology and Medicine, New York University School of Medicine |
| EPI_ISL_421719 | NYU Langone Health | Departments of Pathology and Medicine, New York University School of Medicine |
| EPI_ISL_421720 | NYU Langone Health | Departments of Pathology and Medicine, New York University School of Medicine |
| EPI_ISL_421721 | NYU Langone Health | Departments of Pathology and Medicine, New York University School of Medicine |
| EPI_ISL_421722 | NYU Langone Health | Departments of Pathology and Medicine, New York University School of Medicine |
| EPI_ISL_421723 | NYU Langone Health | Departments of Pathology and Medicine, New York University School of Medicine |
| EPI_ISL_421724 | NYU Langone Health | Departments of Pathology and Medicine, New York University School of Medicine |
| EPI_ISL_421725 | NYU Langone Health | Departments of Pathology and Medicine, New York University School of Medicine |
| EPI_ISL_421726 | NYU Langone Health | Departments of Pathology and Medicine, New York University School of Medicine |
| EPI_ISL_421727 | NYU Langone Health | Departments of Pathology and Medicine, New York University School of Medicine |
| EPI_ISL_421728 | NYU Langone Health | Departments of Pathology and Medicine, New York University School of Medicine |
| EPI_ISL_421729 | NYU Langone Health | Departments of Pathology and Medicine, New York University School of Medicine |
| EPI_ISL_421730 | NYU Langone Health | Departments of Pathology and Medicine, New York University School of Medicine |
| EPI_ISL_421731 | NYU Langone Health | Departments of Pathology and Medicine, New York University School of Medicine |

| EPI_Accession_ID | Originating Laboratory | Submitting Laboratory |
|---|---|---|
| EPI_ISL_421732 | NYU Langone Health | Departments of Pathology and Medicine, New York University School of Medicine |
| EPI_ISL_421733 | NYU Langone Health | Departments of Pathology and Medicine, New York University School of Medicine |
| EPI_ISL_421734 | Laboratoire National de Sante, Microbiology, Virology | Laboratoire National de Sante, Microbiology, Epidemiology and Microbial Genomics |
| EPI_ISL_421735 | Laboratoire National de Sante, Microbiology, Virology | Laboratoire National de Sante, Microbiology, Epidemiology and Microbial Genomics |
| EPI_ISL_421736 | Laboratoire National de Sante, Microbiology, Virology | Laboratoire National de Sante, Microbiology, Epidemiology and Microbial Genomics |
| EPI_ISL_421737 | Laboratoire National de Sante, Microbiology, Virology | Laboratoire National de Sante, Microbiology, Epidemiology and Microbial Genomics |
| EPI_ISL_421738 | Laboratoire National de Sante, Microbiology, Virology | Laboratoire National de Sante, Microbiology, Epidemiology and Microbial Genomics |
| EPI_ISL_421739 | Laboratoire National de Sante, Microbiology, Virology | Laboratoire National de Sante, Microbiology, Epidemiology and Microbial Genomics |
| EPI_ISL_421740 | Laboratoire National de Sante, Microbiology, Virology | Laboratoire National de Sante, Microbiology, Epidemiology and Microbial Genomics |
| EPI_ISL_421741 | Laboratoire National de Sante, Microbiology, Virology | Laboratoire National de Sante, Microbiology, Epidemiology and Microbial Genomics |
| EPI_ISL_421742 | Laboratoire National de Sante, Microbiology, Virology | Laboratoire National de Sante, Microbiology, Epidemiology and Microbial Genomics |
| EPI_ISL_421743 | Laboratoire National de Sante, Microbiology, Virology | Laboratoire National de Sante, Microbiology, Epidemiology and Microbial Genomics |
| EPI_ISL_421744 | Laboratoire National de Sante, Microbiology, Virology | Laboratoire National de Sante, Microbiology, Epidemiology and Microbial Genomics |
| EPI_ISL_421745 | Laboratoire National de Sante, Microbiology, Virology | Laboratoire National de Sante, Microbiology, Epidemiology and Microbial Genomics |
| EPI_ISL_421746 | Laboratoire National de Sante, Microbiology, Virology | Laboratoire National de Sante, Microbiology, Epidemiology and Microbial Genomics |
| EPI_ISL_421747 | Laboratoire National de Sante, Microbiology, Virology | Laboratoire National de Sante, Microbiology, Epidemiology and Microbial Genomics |

| EPI_Accession_ID | Originating Laboratory | Submitting Laboratory |
| --- | --- | --- |
| EPI_ISL_421748 | Laboratoire National de Sante, Microbiology, Virology | Laboratoire National de Sante, Microbiology, Epidemiology and Microbial Genomics |
| EPI_ISL_421749 | Laboratoire National de Sante, Microbiology, Virology | Laboratoire National de Sante, Microbiology, Epidemiology and Microbial Genomics |
| EPI_ISL_421750 | Laboratoire National de Sante, Microbiology, Virology | Laboratoire National de Sante, Microbiology, Epidemiology and Microbial Genomics |
| EPI_ISL_421751 | Laboratoire National de Sante, Microbiology, Virology | Laboratoire National de Sante, Microbiology, Epidemiology and Microbial Genomics |
| EPI_ISL_421752 | Laboratoire National de Sante, Microbiology, Virology | Laboratoire National de Sante, Microbiology, Epidemiology and Microbial Genomics |
| EPI_ISL_421753 | Laboratoire National de Sante, Microbiology, Virology | Laboratoire National de Sante, Microbiology, Epidemiology and Microbial Genomics |
| EPI_ISL_421754 | Laboratoire National de Sante, Microbiology, Virology | Laboratoire National de Sante, Microbiology, Epidemiology and Microbial Genomics |
| EPI_ISL_421755 | Laboratoire National de Sante, Microbiology, Virology | Laboratoire National de Sante, Microbiology, Epidemiology and Microbial Genomics |
| EPI_ISL_421756 | Laboratoire National de Sante, Microbiology, Virology | Laboratoire National de Sante, Microbiology, Epidemiology and Microbial Genomics |
| EPI_ISL_421757 | Laboratoire National de Sante, Microbiology, Virology | Laboratoire National de Sante, Microbiology, Epidemiology and Microbial Genomics |
| EPI_ISL_421758 | Laboratoire National de Sante, Microbiology, Virology | Laboratoire National de Sante, Microbiology, Epidemiology and Microbial Genomics |
| EPI_ISL_421759 | Laboratoire National de Sante, Microbiology, Virology | Laboratoire National de Sante, Microbiology, Epidemiology and Microbial Genomics |
| EPI_ISL_421760 | Laboratoire National de Sante, Microbiology, Virology | Laboratoire National de Sante, Microbiology, Epidemiology and Microbial Genomics |
| EPI_ISL_421761 | Laboratoire National de Sante, Microbiology, Virology | Laboratoire National de Sante, Microbiology, Epidemiology and Microbial Genomics |
| EPI_ISL_421762 | Laboratoire National de Sante, Microbiology, Virology | Laboratoire National de Sante, Microbiology, Epidemiology and Microbial Genomics |
| EPI_ISL_421763 | Laboratoire National de Sante, Microbiology, Virology | Laboratoire National de Sante, Microbiology, Epidemiology and Microbial Genomics |

| EPI_Accession_ID | Originating Laboratory | Submitting Laboratory |
|---|---|---|
| EPI_ISL_421768 | Respiratory Virus Unit, Microbiology Services Colindale, Public Health England | Respiratory Virus Unit, Microbiology Services Colindale, Public Health England |
| EPI_ISL_421769 | Respiratory Virus Unit, Microbiology Services Colindale, Public Health England | Respiratory Virus Unit, Microbiology Services Colindale, Public Health England |
| EPI_ISL_421770 | Respiratory Virus Unit, Microbiology Services Colindale, Public Health England | Respiratory Virus Unit, Microbiology Services Colindale, Public Health England |
| EPI_ISL_421771 | Respiratory Virus Unit, Microbiology Services Colindale, Public Health England | Respiratory Virus Unit, Microbiology Services Colindale, Public Health England |
| EPI_ISL_421772 | Respiratory Virus Unit, Microbiology Services Colindale, Public Health England | Respiratory Virus Unit, Microbiology Services Colindale, Public Health England |
| EPI_ISL_421773 | Respiratory Virus Unit, Microbiology Services Colindale, Public Health England | Respiratory Virus Unit, Microbiology Services Colindale, Public Health England |
| EPI_ISL_421774 | Respiratory Virus Unit, Microbiology Services Colindale, Public Health England | Respiratory Virus Unit, Microbiology Services Colindale, Public Health England |
| EPI_ISL_421775 | Respiratory Virus Unit, Microbiology Services Colindale, Public Health England | Respiratory Virus Unit, Microbiology Services Colindale, Public Health England |
| EPI_ISL_421776 | Respiratory Virus Unit, Microbiology Services Colindale, Public Health England | Respiratory Virus Unit, Microbiology Services Colindale, Public Health England |
| EPI_ISL_421777 | Respiratory Virus Unit, Microbiology Services Colindale, Public Health England | Respiratory Virus Unit, Microbiology Services Colindale, Public Health England |
| EPI_ISL_421778 | Respiratory Virus Unit, Microbiology Services Colindale, Public Health England | Respiratory Virus Unit, Microbiology Services Colindale, Public Health England |
| EPI_ISL_421779 | Respiratory Virus Unit, Microbiology Services Colindale, Public Health England | Respiratory Virus Unit, Microbiology Services Colindale, Public Health England |
| EPI_ISL_421780 | Respiratory Virus Unit, Microbiology Services Colindale, Public Health England | Respiratory Virus Unit, Microbiology Services Colindale, Public Health England |
| EPI_ISL_421781 | Respiratory Virus Unit, Microbiology Services Colindale, Public Health England | Respiratory Virus Unit, Microbiology Services Colindale, Public Health England |
| EPI_ISL_421782 | Respiratory Virus Unit, Microbiology Services Colindale, Public Health England | Respiratory Virus Unit, Microbiology Services Colindale, Public Health England |
| EPI_ISL_421783 | Respiratory Virus Unit, Microbiology Services Colindale, Public Health England | Respiratory Virus Unit, Microbiology Services Colindale, Public Health England |

| EPI_Accession_ID | Originating Laboratory | Submitting Laboratory |
|---|---|---|
| EPI_ISL_421784 | Respiratory Virus Unit, Microbiology Services Colindale, Public Health England | Respiratory Virus Unit, Microbiology Services Colindale, Public Health England |
| EPI_ISL_421785 | Respiratory Virus Unit, Microbiology Services Colindale, Public Health England | Respiratory Virus Unit, Microbiology Services Colindale, Public Health England |
| EPI_ISL_421786 | Respiratory Virus Unit, Microbiology Services Colindale, Public Health England | Respiratory Virus Unit, Microbiology Services Colindale, Public Health England |
| EPI_ISL_421787 | Respiratory Virus Unit, Microbiology Services Colindale, Public Health England | Respiratory Virus Unit, Microbiology Services Colindale, Public Health England |
| EPI_ISL_421788 | Respiratory Virus Unit, Microbiology Services Colindale, Public Health England | Respiratory Virus Unit, Microbiology Services Colindale, Public Health England |
| EPI_ISL_421789 | Respiratory Virus Unit, Microbiology Services Colindale, Public Health England | Respiratory Virus Unit, Microbiology Services Colindale, Public Health England |
| EPI_ISL_421790 | Respiratory Virus Unit, Microbiology Services Colindale, Public Health England | Respiratory Virus Unit, Microbiology Services Colindale, Public Health England |
| EPI_ISL_421791 | Respiratory Virus Unit, Microbiology Services Colindale, Public Health England | Respiratory Virus Unit, Microbiology Services Colindale, Public Health England |
| EPI_ISL_421792 | Respiratory Virus Unit, Microbiology Services Colindale, Public Health England | Respiratory Virus Unit, Microbiology Services Colindale, Public Health England |
| EPI_ISL_421793 | Respiratory Virus Unit, Microbiology Services Colindale, Public Health England | Respiratory Virus Unit, Microbiology Services Colindale, Public Health England |
| EPI_ISL_421794 | Respiratory Virus Unit, Microbiology Services Colindale, Public Health England | Respiratory Virus Unit, Microbiology Services Colindale, Public Health England |
| EPI_ISL_421795 | Respiratory Virus Unit, Microbiology Services Colindale, Public Health England | Respiratory Virus Unit, Microbiology Services Colindale, Public Health England |
| EPI_ISL_421796 | Respiratory Virus Unit, Microbiology Services Colindale, Public Health England | Respiratory Virus Unit, Microbiology Services Colindale, Public Health England |
| EPI_ISL_421797 | Respiratory Virus Unit, Microbiology Services Colindale, Public Health England | Respiratory Virus Unit, Microbiology Services Colindale, Public Health England |
| EPI_ISL_421798 | Respiratory Virus Unit, Microbiology Services Colindale, Public Health England | Respiratory Virus Unit, Microbiology Services Colindale, Public Health England |
| EPI_ISL_421799 | Respiratory Virus Unit, Microbiology Services Colindale, Public Health England | Respiratory Virus Unit, Microbiology Services Colindale, Public Health England |

| EPI_Accession_ID | Originating Laboratory | Submitting Laboratory |
|---|---|---|
| EPI_ISL_421800 | Respiratory Virus Unit, Microbiology Services Colindale, Public Health England | Respiratory Virus Unit, Microbiology Services Colindale, Public Health England |
| EPI_ISL_421801 | Respiratory Virus Unit, Microbiology Services Colindale, Public Health England | Respiratory Virus Unit, Microbiology Services Colindale, Public Health England |
| EPI_ISL_421802 | Respiratory Virus Unit, Microbiology Services Colindale, Public Health England | Respiratory Virus Unit, Microbiology Services Colindale, Public Health England |
| EPI_ISL_421803 | Respiratory Virus Unit, Microbiology Services Colindale, Public Health England | Respiratory Virus Unit, Microbiology Services Colindale, Public Health England |
| EPI_ISL_421804 | Respiratory Virus Unit, Microbiology Services Colindale, Public Health England | Respiratory Virus Unit, Microbiology Services Colindale, Public Health England |
| EPI_ISL_421805 | Respiratory Virus Unit, Microbiology Services Colindale, Public Health England | Respiratory Virus Unit, Microbiology Services Colindale, Public Health England |
| EPI_ISL_421806 | Respiratory Virus Unit, Microbiology Services Colindale, Public Health England | Respiratory Virus Unit, Microbiology Services Colindale, Public Health England |
| EPI_ISL_421807 | Respiratory Virus Unit, Microbiology Services Colindale, Public Health England | Respiratory Virus Unit, Microbiology Services Colindale, Public Health England |
| EPI_ISL_421808 | Respiratory Virus Unit, Microbiology Services Colindale, Public Health England | Respiratory Virus Unit, Microbiology Services Colindale, Public Health England |
| EPI_ISL_421809 | Respiratory Virus Unit, Microbiology Services Colindale, Public Health England | Respiratory Virus Unit, Microbiology Services Colindale, Public Health England |
| EPI_ISL_421810 | Respiratory Virus Unit, Microbiology Services Colindale, Public Health England | Respiratory Virus Unit, Microbiology Services Colindale, Public Health England |
| EPI_ISL_421811 | Respiratory Virus Unit, Microbiology Services Colindale, Public Health England | Respiratory Virus Unit, Microbiology Services Colindale, Public Health England |
| EPI_ISL_421812 | Respiratory Virus Unit, Microbiology Services Colindale, Public Health England | Respiratory Virus Unit, Microbiology Services Colindale, Public Health England |
| EPI_ISL_421813 | Respiratory Virus Unit, Microbiology Services Colindale, Public Health England | Respiratory Virus Unit, Microbiology Services Colindale, Public Health England |
| EPI_ISL_421814 | Respiratory Virus Unit, Microbiology Services Colindale, Public Health England | Respiratory Virus Unit, Microbiology Services Colindale, Public Health England |
| EPI_ISL_421815 | Respiratory Virus Unit, Microbiology Services Colindale, Public Health England | Respiratory Virus Unit, Microbiology Services Colindale, Public Health England |

| EPI_Accession_ID | Originating Laboratory | Submitting Laboratory |
|---|---|---|
| EPI_ISL_421816 | Respiratory Virus Unit, Microbiology Services Colindale, Public Health England | Respiratory Virus Unit, Microbiology Services Colindale, Public Health England |
| EPI_ISL_421817 | Respiratory Virus Unit, Microbiology Services Colindale, Public Health England | Respiratory Virus Unit, Microbiology Services Colindale, Public Health England |
| EPI_ISL_421818 | Respiratory Virus Unit, Microbiology Services Colindale, Public Health England | Respiratory Virus Unit, Microbiology Services Colindale, Public Health England |
| EPI_ISL_421819 | Respiratory Virus Unit, Microbiology Services Colindale, Public Health England | Respiratory Virus Unit, Microbiology Services Colindale, Public Health England |
| EPI_ISL_421820 | Respiratory Virus Unit, Microbiology Services Colindale, Public Health England | Respiratory Virus Unit, Microbiology Services Colindale, Public Health England |
| EPI_ISL_421821 | Respiratory Virus Unit, Microbiology Services Colindale, Public Health England | Respiratory Virus Unit, Microbiology Services Colindale, Public Health England |
| EPI_ISL_421822 | Respiratory Virus Unit, Microbiology Services Colindale, Public Health England | Respiratory Virus Unit, Microbiology Services Colindale, Public Health England |
| EPI_ISL_421823 | Respiratory Virus Unit, Microbiology Services Colindale, Public Health England | Respiratory Virus Unit, Microbiology Services Colindale, Public Health England |
| EPI_ISL_421824 | Respiratory Virus Unit, Microbiology Services Colindale, Public Health England | Respiratory Virus Unit, Microbiology Services Colindale, Public Health England |
| EPI_ISL_421825 | Respiratory Virus Unit, Microbiology Services Colindale, Public Health England | Respiratory Virus Unit, Microbiology Services Colindale, Public Health England |
| EPI_ISL_421826 | Respiratory Virus Unit, Microbiology Services Colindale, Public Health England | Respiratory Virus Unit, Microbiology Services Colindale, Public Health England |
| EPI_ISL_421827 | Respiratory Virus Unit, Microbiology Services Colindale, Public Health England | Respiratory Virus Unit, Microbiology Services Colindale, Public Health England |
| EPI_ISL_421828 | Respiratory Virus Unit, Microbiology Services Colindale, Public Health England | Respiratory Virus Unit, Microbiology Services Colindale, Public Health England |
| EPI_ISL_421829 | Respiratory Virus Unit, Microbiology Services Colindale, Public Health England | Respiratory Virus Unit, Microbiology Services Colindale, Public Health England |
| EPI_ISL_421830 | Respiratory Virus Unit, Microbiology Services Colindale, Public Health England | Respiratory Virus Unit, Microbiology Services Colindale, Public Health England |
| EPI_ISL_421831 | Respiratory Virus Unit, Microbiology Services Colindale, Public Health England | Respiratory Virus Unit, Microbiology Services Colindale, Public Health England |

| EPI_Accession_ID | Originating Laboratory | Submitting Laboratory |
|---|---|---|
| EPI_ISL_421832 | Respiratory Virus Unit, Microbiology Services Colindale, Public Health England | Respiratory Virus Unit, Microbiology Services Colindale, Public Health England |
| EPI_ISL_421833 | Respiratory Virus Unit, Microbiology Services Colindale, Public Health England | Respiratory Virus Unit, Microbiology Services Colindale, Public Health England |
| EPI_ISL_421834 | Respiratory Virus Unit, Microbiology Services Colindale, Public Health England | Respiratory Virus Unit, Microbiology Services Colindale, Public Health England |
| EPI_ISL_421835 | Respiratory Virus Unit, Microbiology Services Colindale, Public Health England | Respiratory Virus Unit, Microbiology Services Colindale, Public Health England |
| EPI_ISL_421836 | Respiratory Virus Unit, Microbiology Services Colindale, Public Health England | Respiratory Virus Unit, Microbiology Services Colindale, Public Health England |
| EPI_ISL_421837 | Respiratory Virus Unit, Microbiology Services Colindale, Public Health England | Respiratory Virus Unit, Microbiology Services Colindale, Public Health England |
| EPI_ISL_421838 | Respiratory Virus Unit, Microbiology Services Colindale, Public Health England | Respiratory Virus Unit, Microbiology Services Colindale, Public Health England |
| EPI_ISL_421839 | Respiratory Virus Unit, Microbiology Services Colindale, Public Health England | Respiratory Virus Unit, Microbiology Services Colindale, Public Health England |
| EPI_ISL_421840 | Respiratory Virus Unit, Microbiology Services Colindale, Public Health England | Respiratory Virus Unit, Microbiology Services Colindale, Public Health England |
| EPI_ISL_421841 | Respiratory Virus Unit, Microbiology Services Colindale, Public Health England | Respiratory Virus Unit, Microbiology Services Colindale, Public Health England |
| EPI_ISL_421842 | Respiratory Virus Unit, Microbiology Services Colindale, Public Health England | Respiratory Virus Unit, Microbiology Services Colindale, Public Health England |
| EPI_ISL_421843 | Respiratory Virus Unit, Microbiology Services Colindale, Public Health England | Respiratory Virus Unit, Microbiology Services Colindale, Public Health England |
| EPI_ISL_421844 | Respiratory Virus Unit, Microbiology Services Colindale, Public Health England | Respiratory Virus Unit, Microbiology Services Colindale, Public Health England |
| EPI_ISL_421845 | Respiratory Virus Unit, Microbiology Services Colindale, Public Health England | Respiratory Virus Unit, Microbiology Services Colindale, Public Health England |
| EPI_ISL_421846 | Respiratory Virus Unit, Microbiology Services Colindale, Public Health England | Respiratory Virus Unit, Microbiology Services Colindale, Public Health England |
| EPI_ISL_421847 | Respiratory Virus Unit, Microbiology Services Colindale, Public Health England | Respiratory Virus Unit, Microbiology Services Colindale, Public Health England |

| EPI_Accession_ID | Originating Laboratory | Submitting Laboratory |
|---|---|---|
| EPI_ISL_421848 | Respiratory Virus Unit, Microbiology Services Colindale, Public Health England | Respiratory Virus Unit, Microbiology Services Colindale, Public Health England |
| EPI_ISL_421849 | Respiratory Virus Unit, Microbiology Services Colindale, Public Health England | Respiratory Virus Unit, Microbiology Services Colindale, Public Health England |
| EPI_ISL_421850 | Respiratory Virus Unit, Microbiology Services Colindale, Public Health England | Respiratory Virus Unit, Microbiology Services Colindale, Public Health England |
| EPI_ISL_421851 | Respiratory Virus Unit, Microbiology Services Colindale, Public Health England | Respiratory Virus Unit, Microbiology Services Colindale, Public Health England |
| EPI_ISL_421852 | Respiratory Virus Unit, Microbiology Services Colindale, Public Health England | Respiratory Virus Unit, Microbiology Services Colindale, Public Health England |
| EPI_ISL_421853 | Respiratory Virus Unit, Microbiology Services Colindale, Public Health England | Respiratory Virus Unit, Microbiology Services Colindale, Public Health England |
| EPI_ISL_421854 | Respiratory Virus Unit, Microbiology Services Colindale, Public Health England | Respiratory Virus Unit, Microbiology Services Colindale, Public Health England |
| EPI_ISL_421855 | Respiratory Virus Unit, Microbiology Services Colindale, Public Health England | Respiratory Virus Unit, Microbiology Services Colindale, Public Health England |
| EPI_ISL_421856 | Respiratory Virus Unit, Microbiology Services Colindale, Public Health England | Respiratory Virus Unit, Microbiology Services Colindale, Public Health England |
| EPI_ISL_421857 | Respiratory Virus Unit, Microbiology Services Colindale, Public Health England | Respiratory Virus Unit, Microbiology Services Colindale, Public Health England |
| EPI_ISL_421858 | Respiratory Virus Unit, Microbiology Services Colindale, Public Health England | Respiratory Virus Unit, Microbiology Services Colindale, Public Health England |
| EPI_ISL_421859 | Respiratory Virus Unit, Microbiology Services Colindale, Public Health England | Respiratory Virus Unit, Microbiology Services Colindale, Public Health England |
| EPI_ISL_421860 | Respiratory Virus Unit, Microbiology Services Colindale, Public Health England | Respiratory Virus Unit, Microbiology Services Colindale, Public Health England |
| EPI_ISL_421861 | Respiratory Virus Unit, Microbiology Services Colindale, Public Health England | Respiratory Virus Unit, Microbiology Services Colindale, Public Health England |
| EPI_ISL_421862 | Respiratory Virus Unit, Microbiology Services Colindale, Public Health England | Respiratory Virus Unit, Microbiology Services Colindale, Public Health England |
| EPI_ISL_421863 | Respiratory Virus Unit, Microbiology Services Colindale, Public Health England | Respiratory Virus Unit, Microbiology Services Colindale, Public Health England |

| EPI_Accession_ID | Originating Laboratory | Submitting Laboratory |
|---|---|---|
| EPI_ISL_421864 | Respiratory Virus Unit, Microbiology Services Colindale, Public Health England | Respiratory Virus Unit, Microbiology Services Colindale, Public Health England |
| EPI_ISL_421865 | Respiratory Virus Unit, Microbiology Services Colindale, Public Health England | Respiratory Virus Unit, Microbiology Services Colindale, Public Health England |
| EPI_ISL_421866 | Respiratory Virus Unit, Microbiology Services Colindale, Public Health England | Respiratory Virus Unit, Microbiology Services Colindale, Public Health England |
| EPI_ISL_421867 | Respiratory Virus Unit, Microbiology Services Colindale, Public Health England | Respiratory Virus Unit, Microbiology Services Colindale, Public Health England |
| EPI_ISL_421868 | Respiratory Virus Unit, Microbiology Services Colindale, Public Health England | Respiratory Virus Unit, Microbiology Services Colindale, Public Health England |
| EPI_ISL_421869 | Respiratory Virus Unit, Microbiology Services Colindale, Public Health England | Respiratory Virus Unit, Microbiology Services Colindale, Public Health England |
| EPI_ISL_421870 | Respiratory Virus Unit, Microbiology Services Colindale, Public Health England | Respiratory Virus Unit, Microbiology Services Colindale, Public Health England |
| EPI_ISL_421871 | Respiratory Virus Unit, Microbiology Services Colindale, Public Health England | Respiratory Virus Unit, Microbiology Services Colindale, Public Health England |
| EPI_ISL_421872 | Respiratory Virus Unit, Microbiology Services Colindale, Public Health England | Respiratory Virus Unit, Microbiology Services Colindale, Public Health England |
| EPI_ISL_421873 | Respiratory Virus Unit, Microbiology Services Colindale, Public Health England | Respiratory Virus Unit, Microbiology Services Colindale, Public Health England |
| EPI_ISL_421874 | Respiratory Virus Unit, Microbiology Services Colindale, Public Health England | Respiratory Virus Unit, Microbiology Services Colindale, Public Health England |
| EPI_ISL_421875 | Respiratory Virus Unit, Microbiology Services Colindale, Public Health England | Respiratory Virus Unit, Microbiology Services Colindale, Public Health England |
| EPI_ISL_421876 | Respiratory Virus Unit, Microbiology Services Colindale, Public Health England | Respiratory Virus Unit, Microbiology Services Colindale, Public Health England |
| EPI_ISL_421877 | Respiratory Virus Unit, Microbiology Services Colindale, Public Health England | Respiratory Virus Unit, Microbiology Services Colindale, Public Health England |
| EPI_ISL_421878 | Respiratory Virus Unit, Microbiology Services Colindale, Public Health England | Respiratory Virus Unit, Microbiology Services Colindale, Public Health England |
| EPI_ISL_421879 | Respiratory Virus Unit, Microbiology Services Colindale, Public Health England | Respiratory Virus Unit, Microbiology Services Colindale, Public Health England |

| EPI_Accession_ID | Originating Laboratory | Submitting Laboratory |
|---|---|---|
| EPI_ISL_421880 | Respiratory Virus Unit, Microbiology Services Colindale, Public Health England | Respiratory Virus Unit, Microbiology Services Colindale, Public Health England |
| EPI_ISL_421881 | Respiratory Virus Unit, Microbiology Services Colindale, Public Health England | Respiratory Virus Unit, Microbiology Services Colindale, Public Health England |
| EPI_ISL_421882 | Respiratory Virus Unit, Microbiology Services Colindale, Public Health England | Respiratory Virus Unit, Microbiology Services Colindale, Public Health England |
| EPI_ISL_421883 | Respiratory Virus Unit, Microbiology Services Colindale, Public Health England | Respiratory Virus Unit, Microbiology Services Colindale, Public Health England |
| EPI_ISL_421884 | Respiratory Virus Unit, Microbiology Services Colindale, Public Health England | Respiratory Virus Unit, Microbiology Services Colindale, Public Health England |
| EPI_ISL_421885 | Respiratory Virus Unit, Microbiology Services Colindale, Public Health England | Respiratory Virus Unit, Microbiology Services Colindale, Public Health England |
| EPI_ISL_421886 | Respiratory Virus Unit, Microbiology Services Colindale, Public Health England | Respiratory Virus Unit, Microbiology Services Colindale, Public Health England |
| EPI_ISL_421887 | Respiratory Virus Unit, Microbiology Services Colindale, Public Health England | Respiratory Virus Unit, Microbiology Services Colindale, Public Health England |
| EPI_ISL_421888 | Respiratory Virus Unit, Microbiology Services Colindale, Public Health England | Respiratory Virus Unit, Microbiology Services Colindale, Public Health England |
| EPI_ISL_421889 | Respiratory Virus Unit, Microbiology Services Colindale, Public Health England | Respiratory Virus Unit, Microbiology Services Colindale, Public Health England |
| EPI_ISL_421890 | Respiratory Virus Unit, Microbiology Services Colindale, Public Health England | Respiratory Virus Unit, Microbiology Services Colindale, Public Health England |
| EPI_ISL_421891 | Respiratory Virus Unit, Microbiology Services Colindale, Public Health England | Respiratory Virus Unit, Microbiology Services Colindale, Public Health England |
| EPI_ISL_421892 | Respiratory Virus Unit, Microbiology Services Colindale, Public Health England | Respiratory Virus Unit, Microbiology Services Colindale, Public Health England |
| EPI_ISL_421893 | Respiratory Virus Unit, Microbiology Services Colindale, Public Health England | Respiratory Virus Unit, Microbiology Services Colindale, Public Health England |
| EPI_ISL_421894 | Respiratory Virus Unit, Microbiology Services Colindale, Public Health England | Respiratory Virus Unit, Microbiology Services Colindale, Public Health England |
| EPI_ISL_421895 | Respiratory Virus Unit, Microbiology Services Colindale, Public Health England | Respiratory Virus Unit, Microbiology Services Colindale, Public Health England |

| EPI_Accession_ID | Originating Laboratory | Submitting Laboratory |
|---|---|---|
| EPI_ISL_421896 | Respiratory Virus Unit, Microbiology Services Colindale, Public Health England | Respiratory Virus Unit, Microbiology Services Colindale, Public Health England |
| EPI_ISL_421897 | Respiratory Virus Unit, Microbiology Services Colindale, Public Health England | Respiratory Virus Unit, Microbiology Services Colindale, Public Health England |
| EPI_ISL_421898 | Respiratory Virus Unit, Microbiology Services Colindale, Public Health England | Respiratory Virus Unit, Microbiology Services Colindale, Public Health England |
| EPI_ISL_421899 | Respiratory Virus Unit, Microbiology Services Colindale, Public Health England | Respiratory Virus Unit, Microbiology Services Colindale, Public Health England |
| EPI_ISL_421900 | Respiratory Virus Unit, Microbiology Services Colindale, Public Health England | Respiratory Virus Unit, Microbiology Services Colindale, Public Health England |
| EPI_ISL_421901 | Respiratory Virus Unit, Microbiology Services Colindale, Public Health England | Respiratory Virus Unit, Microbiology Services Colindale, Public Health England |
| EPI_ISL_421902 | Respiratory Virus Unit, Microbiology Services Colindale, Public Health England | Respiratory Virus Unit, Microbiology Services Colindale, Public Health England |
| EPI_ISL_421903 | Respiratory Virus Unit, Microbiology Services Colindale, Public Health England | Respiratory Virus Unit, Microbiology Services Colindale, Public Health England |
| EPI_ISL_421904 | Respiratory Virus Unit, Microbiology Services Colindale, Public Health England | Respiratory Virus Unit, Microbiology Services Colindale, Public Health England |
| EPI_ISL_421905 | Respiratory Virus Unit, Microbiology Services Colindale, Public Health England | Respiratory Virus Unit, Microbiology Services Colindale, Public Health England |
| EPI_ISL_421906 | Respiratory Virus Unit, Microbiology Services Colindale, Public Health England | Respiratory Virus Unit, Microbiology Services Colindale, Public Health England |
| EPI_ISL_421907 | Respiratory Virus Unit, Microbiology Services Colindale, Public Health England | Respiratory Virus Unit, Microbiology Services Colindale, Public Health England |
| EPI_ISL_421908 | Respiratory Virus Unit, Microbiology Services Colindale, Public Health England | Respiratory Virus Unit, Microbiology Services Colindale, Public Health England |
| EPI_ISL_421909 | Respiratory Virus Unit, Microbiology Services Colindale, Public Health England | Respiratory Virus Unit, Microbiology Services Colindale, Public Health England |
| EPI_ISL_421910 | Respiratory Virus Unit, Microbiology Services Colindale, Public Health England | Respiratory Virus Unit, Microbiology Services Colindale, Public Health England |
| EPI_ISL_421911 | Respiratory Virus Unit, Microbiology Services Colindale, Public Health England | Respiratory Virus Unit, Microbiology Services Colindale, Public Health England |

| EPI_Accession_ID | Originating Laboratory | Submitting Laboratory |
|---|---|---|
| EPI_ISL_421912 | Respiratory Virus Unit, Microbiology Services Colindale, Public Health England | Respiratory Virus Unit, Microbiology Services Colindale, Public Health England |
| EPI_ISL_421913 | Respiratory Virus Unit, Microbiology Services Colindale, Public Health England | Respiratory Virus Unit, Microbiology Services Colindale, Public Health England |
| EPI_ISL_421914 | Respiratory Virus Unit, Microbiology Services Colindale, Public Health England | Respiratory Virus Unit, Microbiology Services Colindale, Public Health England |
| EPI_ISL_421915 | Respiratory Virus Unit, Microbiology Services Colindale, Public Health England | Respiratory Virus Unit, Microbiology Services Colindale, Public Health England |
| EPI_ISL_421916 | Respiratory Virus Unit, Microbiology Services Colindale, Public Health England | Respiratory Virus Unit, Microbiology Services Colindale, Public Health England |
| EPI_ISL_421917 | Respiratory Virus Unit, Microbiology Services Colindale, Public Health England | Respiratory Virus Unit, Microbiology Services Colindale, Public Health England |
| EPI_ISL_421918 | Respiratory Virus Unit, Microbiology Services Colindale, Public Health England | Respiratory Virus Unit, Microbiology Services Colindale, Public Health England |
| EPI_ISL_421919 | Respiratory Virus Unit, Microbiology Services Colindale, Public Health England | Respiratory Virus Unit, Microbiology Services Colindale, Public Health England |
| EPI_ISL_421920 | Respiratory Virus Unit, Microbiology Services Colindale, Public Health England | Respiratory Virus Unit, Microbiology Services Colindale, Public Health England |
| EPI_ISL_421921 | Respiratory Virus Unit, Microbiology Services Colindale, Public Health England | Respiratory Virus Unit, Microbiology Services Colindale, Public Health England |
| EPI_ISL_421922 | Respiratory Virus Unit, Microbiology Services Colindale, Public Health England | Respiratory Virus Unit, Microbiology Services Colindale, Public Health England |
| EPI_ISL_421923 | Respiratory Virus Unit, Microbiology Services Colindale, Public Health England | Respiratory Virus Unit, Microbiology Services Colindale, Public Health England |
| EPI_ISL_421924 | Respiratory Virus Unit, Microbiology Services Colindale, Public Health England | Respiratory Virus Unit, Microbiology Services Colindale, Public Health England |
| EPI_ISL_421925 | Respiratory Virus Unit, Microbiology Services Colindale, Public Health England | Respiratory Virus Unit, Microbiology Services Colindale, Public Health England |
| EPI_ISL_421926 | Respiratory Virus Unit, Microbiology Services Colindale, Public Health England | Respiratory Virus Unit, Microbiology Services Colindale, Public Health England |
| EPI_ISL_421927 | Respiratory Virus Unit, Microbiology Services Colindale, Public Health England | Respiratory Virus Unit, Microbiology Services Colindale, Public Health England |

| EPI_Accession_ID | Originating Laboratory | Submitting Laboratory |
|---|---|---|
| EPI_ISL_421928 | Respiratory Virus Unit, Microbiology Services Colindale, Public Health England | Respiratory Virus Unit, Microbiology Services Colindale, Public Health England |
| EPI_ISL_421929 | Respiratory Virus Unit, Microbiology Services Colindale, Public Health England | Respiratory Virus Unit, Microbiology Services Colindale, Public Health England |
| EPI_ISL_421930 | Respiratory Virus Unit, Microbiology Services Colindale, Public Health England | Respiratory Virus Unit, Microbiology Services Colindale, Public Health England |
| EPI_ISL_421931 | Respiratory Virus Unit, Microbiology Services Colindale, Public Health England | Respiratory Virus Unit, Microbiology Services Colindale, Public Health England |
| EPI_ISL_421932 | Respiratory Virus Unit, Microbiology Services Colindale, Public Health England | Respiratory Virus Unit, Microbiology Services Colindale, Public Health England |
| EPI_ISL_421933 | Respiratory Virus Unit, Microbiology Services Colindale, Public Health England | Respiratory Virus Unit, Microbiology Services Colindale, Public Health England |
| EPI_ISL_421934 | Respiratory Virus Unit, Microbiology Services Colindale, Public Health England | Respiratory Virus Unit, Microbiology Services Colindale, Public Health England |
| EPI_ISL_421935 | Respiratory Virus Unit, Microbiology Services Colindale, Public Health England | Respiratory Virus Unit, Microbiology Services Colindale, Public Health England |
| EPI_ISL_421936 | Respiratory Virus Unit, Microbiology Services Colindale, Public Health England | Respiratory Virus Unit, Microbiology Services Colindale, Public Health England |
| EPI_ISL_421937 | Respiratory Virus Unit, Microbiology Services Colindale, Public Health England | Respiratory Virus Unit, Microbiology Services Colindale, Public Health England |
| EPI_ISL_421938 | Respiratory Virus Unit, Microbiology Services Colindale, Public Health England | Respiratory Virus Unit, Microbiology Services Colindale, Public Health England |
| EPI_ISL_421939 | Respiratory Virus Unit, Microbiology Services Colindale, Public Health England | Respiratory Virus Unit, Microbiology Services Colindale, Public Health England |
| EPI_ISL_421940 | Respiratory Virus Unit, Microbiology Services Colindale, Public Health England | Respiratory Virus Unit, Microbiology Services Colindale, Public Health England |
| EPI_ISL_421941 | Respiratory Virus Unit, Microbiology Services Colindale, Public Health England | Respiratory Virus Unit, Microbiology Services Colindale, Public Health England |
| EPI_ISL_421942 | Respiratory Virus Unit, Microbiology Services Colindale, Public Health England | Respiratory Virus Unit, Microbiology Services Colindale, Public Health England |
| EPI_ISL_421943 | Respiratory Virus Unit, Microbiology Services Colindale, Public Health England | Respiratory Virus Unit, Microbiology Services Colindale, Public Health England |

| EPI_Accession_ID | Originating Laboratory | Submitting Laboratory |
|---|---|---|
| EPI_ISL_421944 | Respiratory Virus Unit, Microbiology Services Colindale, Public Health England | Respiratory Virus Unit, Microbiology Services Colindale, Public Health England |
| EPI_ISL_421945 | Respiratory Virus Unit, Microbiology Services Colindale, Public Health England | Respiratory Virus Unit, Microbiology Services Colindale, Public Health England |
| EPI_ISL_421946 | Respiratory Virus Unit, Microbiology Services Colindale, Public Health England | Respiratory Virus Unit, Microbiology Services Colindale, Public Health England |
| EPI_ISL_421947 | Respiratory Virus Unit, Microbiology Services Colindale, Public Health England | Respiratory Virus Unit, Microbiology Services Colindale, Public Health England |
| EPI_ISL_421948 | Respiratory Virus Unit, Microbiology Services Colindale, Public Health England | Respiratory Virus Unit, Microbiology Services Colindale, Public Health England |
| EPI_ISL_421949 | Respiratory Virus Unit, Microbiology Services Colindale, Public Health England | Respiratory Virus Unit, Microbiology Services Colindale, Public Health England |
| EPI_ISL_421951 | Respiratory Virus Unit, Microbiology Services Colindale, Public Health England | Respiratory Virus Unit, Microbiology Services Colindale, Public Health England |
| EPI_ISL_421952 | Respiratory Virus Unit, Microbiology Services Colindale, Public Health England | Respiratory Virus Unit, Microbiology Services Colindale, Public Health England |
| EPI_ISL_421953 | Respiratory Virus Unit, Microbiology Services Colindale, Public Health England | Respiratory Virus Unit, Microbiology Services Colindale, Public Health England |
| EPI_ISL_421954 | Respiratory Virus Unit, Microbiology Services Colindale, Public Health England | Respiratory Virus Unit, Microbiology Services Colindale, Public Health England |
| EPI_ISL_421955 | Respiratory Virus Unit, Microbiology Services Colindale, Public Health England | Respiratory Virus Unit, Microbiology Services Colindale, Public Health England |
| EPI_ISL_421956 | Respiratory Virus Unit, Microbiology Services Colindale, Public Health England | Respiratory Virus Unit, Microbiology Services Colindale, Public Health England |
| EPI_ISL_421957 | Respiratory Virus Unit, Microbiology Services Colindale, Public Health England | Respiratory Virus Unit, Microbiology Services Colindale, Public Health England |
| EPI_ISL_421958 | Respiratory Virus Unit, Microbiology Services Colindale, Public Health England | Respiratory Virus Unit, Microbiology Services Colindale, Public Health England |
| EPI_ISL_421959 | Respiratory Virus Unit, Microbiology Services Colindale, Public Health England | Respiratory Virus Unit, Microbiology Services Colindale, Public Health England |
| EPI_ISL_421960 | Respiratory Virus Unit, Microbiology Services Colindale, Public Health England | Respiratory Virus Unit, Microbiology Services Colindale, Public Health England |

| EPI_Accession_ID | Originating Laboratory | Submitting Laboratory |
|---|---|---|
| EPI_ISL_421961 | Respiratory Virus Unit, Microbiology Services Colindale, Public Health England | Respiratory Virus Unit, Microbiology Services Colindale, Public Health England |
| EPI_ISL_421962 | Respiratory Virus Unit, Microbiology Services Colindale, Public Health England | Respiratory Virus Unit, Microbiology Services Colindale, Public Health England |
| EPI_ISL_421963 | Respiratory Virus Unit, Microbiology Services Colindale, Public Health England | Respiratory Virus Unit, Microbiology Services Colindale, Public Health England |
| EPI_ISL_421964 | Respiratory Virus Unit, Microbiology Services Colindale, Public Health England | Respiratory Virus Unit, Microbiology Services Colindale, Public Health England |
| EPI_ISL_421965 | Respiratory Virus Unit, Microbiology Services Colindale, Public Health England | Respiratory Virus Unit, Microbiology Services Colindale, Public Health England |
| EPI_ISL_421966 | Respiratory Virus Unit, Microbiology Services Colindale, Public Health England | Respiratory Virus Unit, Microbiology Services Colindale, Public Health England |
| EPI_ISL_421967 | Respiratory Virus Unit, Microbiology Services Colindale, Public Health England | Respiratory Virus Unit, Microbiology Services Colindale, Public Health England |
| EPI_ISL_421968 | Respiratory Virus Unit, Microbiology Services Colindale, Public Health England | Respiratory Virus Unit, Microbiology Services Colindale, Public Health England |
| EPI_ISL_421969 | Respiratory Virus Unit, Microbiology Services Colindale, Public Health England | Respiratory Virus Unit, Microbiology Services Colindale, Public Health England |
| EPI_ISL_421970 | Respiratory Virus Unit, Microbiology Services Colindale, Public Health England | Respiratory Virus Unit, Microbiology Services Colindale, Public Health England |
| EPI_ISL_421971 | Respiratory Virus Unit, Microbiology Services Colindale, Public Health England | Respiratory Virus Unit, Microbiology Services Colindale, Public Health England |
| EPI_ISL_421972 | Respiratory Virus Unit, Microbiology Services Colindale, Public Health England | Respiratory Virus Unit, Microbiology Services Colindale, Public Health England |
| EPI_ISL_421973 | Respiratory Virus Unit, Microbiology Services Colindale, Public Health England | Respiratory Virus Unit, Microbiology Services Colindale, Public Health England |
| EPI_ISL_421974 | Respiratory Virus Unit, Microbiology Services Colindale, Public Health England | Respiratory Virus Unit, Microbiology Services Colindale, Public Health England |
| EPI_ISL_421975 | Respiratory Virus Unit, Microbiology Services Colindale, Public Health England | Respiratory Virus Unit, Microbiology Services Colindale, Public Health England |
| EPI_ISL_421976 | Respiratory Virus Unit, Microbiology Services Colindale, Public Health England | Respiratory Virus Unit, Microbiology Services Colindale, Public Health England |

| EPI_Accession_ID | Originating Laboratory | Submitting Laboratory |
|---|---|---|
| EPI_ISL_421977 | Respiratory Virus Unit, Microbiology Services Colindale, Public Health England | Respiratory Virus Unit, Microbiology Services Colindale, Public Health England |
| EPI_ISL_421978 | Respiratory Virus Unit, Microbiology Services Colindale, Public Health England | Respiratory Virus Unit, Microbiology Services Colindale, Public Health England |
| EPI_ISL_421979 | Respiratory Virus Unit, Microbiology Services Colindale, Public Health England | Respiratory Virus Unit, Microbiology Services Colindale, Public Health England |
| EPI_ISL_421980 | Respiratory Virus Unit, Microbiology Services Colindale, Public Health England | Respiratory Virus Unit, Microbiology Services Colindale, Public Health England |
| EPI_ISL_421981 | Respiratory Virus Unit, Microbiology Services Colindale, Public Health England | Respiratory Virus Unit, Microbiology Services Colindale, Public Health England |
| EPI_ISL_421982 | Respiratory Virus Unit, Microbiology Services Colindale, Public Health England | Respiratory Virus Unit, Microbiology Services Colindale, Public Health England |
| EPI_ISL_421983 | Respiratory Virus Unit, Microbiology Services Colindale, Public Health England | Respiratory Virus Unit, Microbiology Services Colindale, Public Health England |
| EPI_ISL_421984 | Respiratory Virus Unit, Microbiology Services Colindale, Public Health England | Respiratory Virus Unit, Microbiology Services Colindale, Public Health England |
| EPI_ISL_421985 | Respiratory Virus Unit, Microbiology Services Colindale, Public Health England | Respiratory Virus Unit, Microbiology Services Colindale, Public Health England |
| EPI_ISL_421986 | Respiratory Virus Unit, Microbiology Services Colindale, Public Health England | Respiratory Virus Unit, Microbiology Services Colindale, Public Health England |
| EPI_ISL_421987 | Respiratory Virus Unit, Microbiology Services Colindale, Public Health England | Respiratory Virus Unit, Microbiology Services Colindale, Public Health England |
| EPI_ISL_421988 | Respiratory Virus Unit, Microbiology Services Colindale, Public Health England | Respiratory Virus Unit, Microbiology Services Colindale, Public Health England |
| EPI_ISL_421989 | Respiratory Virus Unit, Microbiology Services Colindale, Public Health England | Respiratory Virus Unit, Microbiology Services Colindale, Public Health England |
| EPI_ISL_421990 | Respiratory Virus Unit, Microbiology Services Colindale, Public Health England | Respiratory Virus Unit, Microbiology Services Colindale, Public Health England |
| EPI_ISL_421991 | Respiratory Virus Unit, Microbiology Services Colindale, Public Health England | Respiratory Virus Unit, Microbiology Services Colindale, Public Health England |
| EPI_ISL_421992 | Respiratory Virus Unit, Microbiology Services Colindale, Public Health England | Respiratory Virus Unit, Microbiology Services Colindale, Public Health England |

| EPI_Accession_ID | Originating Laboratory | Submitting Laboratory |
|---|---|---|
| EPI_ISL_421993 | Respiratory Virus Unit, Microbiology Services Colindale, Public Health England | Respiratory Virus Unit, Microbiology Services Colindale, Public Health England |
| EPI_ISL_421994 | Respiratory Virus Unit, Microbiology Services Colindale, Public Health England | Respiratory Virus Unit, Microbiology Services Colindale, Public Health England |
| EPI_ISL_421995 | Respiratory Virus Unit, Microbiology Services Colindale, Public Health England | Respiratory Virus Unit, Microbiology Services Colindale, Public Health England |
| EPI_ISL_421996 | Respiratory Virus Unit, Microbiology Services Colindale, Public Health England | Respiratory Virus Unit, Microbiology Services Colindale, Public Health England |
| EPI_ISL_421997 | Respiratory Virus Unit, Microbiology Services Colindale, Public Health England | Respiratory Virus Unit, Microbiology Services Colindale, Public Health England |
| EPI_ISL_421998 | Respiratory Virus Unit, Microbiology Services Colindale, Public Health England | Respiratory Virus Unit, Microbiology Services Colindale, Public Health England |
| EPI_ISL_421999 | Respiratory Virus Unit, Microbiology Services Colindale, Public Health England | Respiratory Virus Unit, Microbiology Services Colindale, Public Health England |
| EPI_ISL_422000 | Respiratory Virus Unit, Microbiology Services Colindale, Public Health England | Respiratory Virus Unit, Microbiology Services Colindale, Public Health England |
| EPI_ISL_422001 | Respiratory Virus Unit, Microbiology Services Colindale, Public Health England | Respiratory Virus Unit, Microbiology Services Colindale, Public Health England |
| EPI_ISL_422002 | Respiratory Virus Unit, Microbiology Services Colindale, Public Health England | Respiratory Virus Unit, Microbiology Services Colindale, Public Health England |
| EPI_ISL_422003 | Respiratory Virus Unit, Microbiology Services Colindale, Public Health England | Respiratory Virus Unit, Microbiology Services Colindale, Public Health England |
| EPI_ISL_422004 | Respiratory Virus Unit, Microbiology Services Colindale, Public Health England | Respiratory Virus Unit, Microbiology Services Colindale, Public Health England |
| EPI_ISL_422005 | Respiratory Virus Unit, Microbiology Services Colindale, Public Health England | Respiratory Virus Unit, Microbiology Services Colindale, Public Health England |
| EPI_ISL_422006 | Respiratory Virus Unit, Microbiology Services Colindale, Public Health England | Respiratory Virus Unit, Microbiology Services Colindale, Public Health England |
| EPI_ISL_422007 | Respiratory Virus Unit, Microbiology Services Colindale, Public Health England | Respiratory Virus Unit, Microbiology Services Colindale, Public Health England |
| EPI_ISL_422008 | Respiratory Virus Unit, Microbiology Services Colindale, Public Health England | Respiratory Virus Unit, Microbiology Services Colindale, Public Health England |

| EPI_Accession_ID | Originating Laboratory | Submitting Laboratory |
|---|---|---|
| EPI_ISL_422009 | Respiratory Virus Unit, Microbiology Services Colindale, Public Health England | Respiratory Virus Unit, Microbiology Services Colindale, Public Health England |
| EPI_ISL_422010 | Respiratory Virus Unit, Microbiology Services Colindale, Public Health England | Respiratory Virus Unit, Microbiology Services Colindale, Public Health England |
| EPI_ISL_422011 | Respiratory Virus Unit, Microbiology Services Colindale, Public Health England | Respiratory Virus Unit, Microbiology Services Colindale, Public Health England |
| EPI_ISL_422012 | Respiratory Virus Unit, Microbiology Services Colindale, Public Health England | Respiratory Virus Unit, Microbiology Services Colindale, Public Health England |
| EPI_ISL_422016 | Wales Specialist Virology Centre | Public Health Wales Microbiology Cardiff |
| EPI_ISL_422017 | Wales Specialist Virology Centre | Public Health Wales Microbiology Cardiff |
| EPI_ISL_422018 | Wales Specialist Virology Centre | Public Health Wales Microbiology Cardiff |
| EPI_ISL_422019 | Wales Specialist Virology Centre | Public Health Wales Microbiology Cardiff |
| EPI_ISL_422020 | Wales Specialist Virology Centre | Public Health Wales Microbiology Cardiff |
| EPI_ISL_422021 | Wales Specialist Virology Centre | Public Health Wales Microbiology Cardiff |
| EPI_ISL_422022 | Wales Specialist Virology Centre | Public Health Wales Microbiology Cardiff |
| EPI_ISL_422023 | Wales Specialist Virology Centre | Public Health Wales Microbiology Cardiff |
| EPI_ISL_422024 | Wales Specialist Virology Centre | Public Health Wales Microbiology Cardiff |
| EPI_ISL_422025 | Wales Specialist Virology Centre | Public Health Wales Microbiology Cardiff |
| EPI_ISL_422026 | Wales Specialist Virology Centre | Public Health Wales Microbiology Cardiff |
| EPI_ISL_422027 | Wales Specialist Virology Centre | Public Health Wales Microbiology Cardiff |
| EPI_ISL_422028 | Wales Specialist Virology Centre | Public Health Wales Microbiology Cardiff |
| EPI_ISL_422029 | Wales Specialist Virology Centre | Public Health Wales Microbiology Cardiff |
| EPI_ISL_422030 | Wales Specialist Virology Centre | Public Health Wales Microbiology Cardiff |
| EPI_ISL_422031 | Wales Specialist Virology Centre | Public Health Wales Microbiology Cardiff |
| EPI_ISL_422032 | Wales Specialist Virology Centre | Public Health Wales Microbiology Cardiff |
| EPI_ISL_422033 | Wales Specialist Virology Centre | Public Health Wales Microbiology Cardiff |
| EPI_ISL_422034 | Wales Specialist Virology Centre | Public Health Wales Microbiology Cardiff |
| EPI_ISL_422035 | Wales Specialist Virology Centre | Public Health Wales Microbiology Cardiff |
| EPI_ISL_422036 | Wales Specialist Virology Centre | Public Health Wales Microbiology Cardiff |
| EPI_ISL_422037 | Wales Specialist Virology Centre | Public Health Wales Microbiology Cardiff |
| EPI_ISL_422038 | Wales Specialist Virology Centre | Public Health Wales Microbiology Cardiff |
| EPI_ISL_422039 | Wales Specialist Virology Centre | Public Health Wales Microbiology Cardiff |
| EPI_ISL_422040 | Wales Specialist Virology Centre | Public Health Wales Microbiology Cardiff |

| EPI_Accession_ID | Originating Laboratory | Submitting Laboratory |
|---|---|---|
| EPI_ISL_422041 | Wales Specialist Virology Centre | Public Health Wales Microbiology Cardiff |
| EPI_ISL_422042 | Wales Specialist Virology Centre | Public Health Wales Microbiology Cardiff |
| EPI_ISL_422043 | Wales Specialist Virology Centre | Public Health Wales Microbiology Cardiff |
| EPI_ISL_422044 | Wales Specialist Virology Centre | Public Health Wales Microbiology Cardiff |
| EPI_ISL_422045 | Wales Specialist Virology Centre | Public Health Wales Microbiology Cardiff |
| EPI_ISL_422046 | Wales Specialist Virology Centre | Public Health Wales Microbiology Cardiff |
| EPI_ISL_422047 | Wales Specialist Virology Centre | Public Health Wales Microbiology Cardiff |
| EPI_ISL_422048 | Wales Specialist Virology Centre | Public Health Wales Microbiology Cardiff |
| EPI_ISL_422049 | Wales Specialist Virology Centre | Public Health Wales Microbiology Cardiff |
| EPI_ISL_422050 | Wales Specialist Virology Centre | Public Health Wales Microbiology Cardiff |
| EPI_ISL_422051 | Wales Specialist Virology Centre | Public Health Wales Microbiology Cardiff |
| EPI_ISL_422052 | Wales Specialist Virology Centre | Public Health Wales Microbiology Cardiff |
| EPI_ISL_422053 | Wales Specialist Virology Centre | Public Health Wales Microbiology Cardiff |
| EPI_ISL_422054 | Wales Specialist Virology Centre | Public Health Wales Microbiology Cardiff |
| EPI_ISL_422055 | Wales Specialist Virology Centre | Public Health Wales Microbiology Cardiff |
| EPI_ISL_422056 | Wales Specialist Virology Centre | Public Health Wales Microbiology Cardiff |
| EPI_ISL_422057 | Wales Specialist Virology Centre | Public Health Wales Microbiology Cardiff |
| EPI_ISL_422058 | Wales Specialist Virology Centre | Public Health Wales Microbiology Cardiff |
| EPI_ISL_422059 | Wales Specialist Virology Centre | Public Health Wales Microbiology Cardiff |
| EPI_ISL_422060 | Wales Specialist Virology Centre | Public Health Wales Microbiology Cardiff |
| EPI_ISL_422061 | Wales Specialist Virology Centre | Public Health Wales Microbiology Cardiff |
| EPI_ISL_422062 | Wales Specialist Virology Centre | Public Health Wales Microbiology Cardiff |
| EPI_ISL_422063 | Wales Specialist Virology Centre | Public Health Wales Microbiology Cardiff |
| EPI_ISL_422064 | Wales Specialist Virology Centre | Public Health Wales Microbiology Cardiff |
| EPI_ISL_422065 | Wales Specialist Virology Centre | Public Health Wales Microbiology Cardiff |
| EPI_ISL_422066 | Wales Specialist Virology Centre | Public Health Wales Microbiology Cardiff |
| EPI_ISL_422067 | Wales Specialist Virology Centre | Public Health Wales Microbiology Cardiff |
| EPI_ISL_422068 | Wales Specialist Virology Centre | Public Health Wales Microbiology Cardiff |
| EPI_ISL_422069 | Wales Specialist Virology Centre | Public Health Wales Microbiology Cardiff |
| EPI_ISL_422070 | Wales Specialist Virology Centre | Public Health Wales Microbiology Cardiff |
| EPI_ISL_422071 | Wales Specialist Virology Centre | Public Health Wales Microbiology Cardiff |
| EPI_ISL_422072 | Wales Specialist Virology Centre | Public Health Wales Microbiology Cardiff |
| EPI_ISL_422073 | Wales Specialist Virology Centre | Public Health Wales Microbiology Cardiff |

| EPI_Accession_ID | Originating Laboratory | Submitting Laboratory |
|---|---|---|
| EPI_ISL_422074 | Wales Specialist Virology Centre | Public Health Wales Microbiology Cardiff |
| EPI_ISL_422075 | Wales Specialist Virology Centre | Public Health Wales Microbiology Cardiff |
| EPI_ISL_422076 | Wales Specialist Virology Centre | Public Health Wales Microbiology Cardiff |
| EPI_ISL_422077 | Wales Specialist Virology Centre | Public Health Wales Microbiology Cardiff |
| EPI_ISL_422078 | Wales Specialist Virology Centre | Public Health Wales Microbiology Cardiff |
| EPI_ISL_422079 | Wales Specialist Virology Centre | Public Health Wales Microbiology Cardiff |
| EPI_ISL_422080 | Wales Specialist Virology Centre | Public Health Wales Microbiology Cardiff |
| EPI_ISL_422081 | Wales Specialist Virology Centre | Public Health Wales Microbiology Cardiff |
| EPI_ISL_422082 | Wales Specialist Virology Centre | Public Health Wales Microbiology Cardiff |
| EPI_ISL_422083 | Wales Specialist Virology Centre | Public Health Wales Microbiology Cardiff |
| EPI_ISL_422084 | Wales Specialist Virology Centre | Public Health Wales Microbiology Cardiff |
| EPI_ISL_422085 | Wales Specialist Virology Centre | Public Health Wales Microbiology Cardiff |
| EPI_ISL_422086 | Wales Specialist Virology Centre | Public Health Wales Microbiology Cardiff |
| EPI_ISL_422087 | Wales Specialist Virology Centre | Public Health Wales Microbiology Cardiff |
| EPI_ISL_422088 | Wales Specialist Virology Centre | Public Health Wales Microbiology Cardiff |
| EPI_ISL_422089 | Wales Specialist Virology Centre | Public Health Wales Microbiology Cardiff |
| EPI_ISL_422090 | Wales Specialist Virology Centre | Public Health Wales Microbiology Cardiff |
| EPI_ISL_422091 | Wales Specialist Virology Centre | Public Health Wales Microbiology Cardiff |
| EPI_ISL_422092 | Wales Specialist Virology Centre | Public Health Wales Microbiology Cardiff |
| EPI_ISL_422093 | Wales Specialist Virology Centre | Public Health Wales Microbiology Cardiff |
| EPI_ISL_422094 | Wales Specialist Virology Centre | Public Health Wales Microbiology Cardiff |
| EPI_ISL_422095 | Wales Specialist Virology Centre | Public Health Wales Microbiology Cardiff |
| EPI_ISL_422096 | Wales Specialist Virology Centre | Public Health Wales Microbiology Cardiff |
| EPI_ISL_422097 | Wales Specialist Virology Centre | Public Health Wales Microbiology Cardiff |
| EPI_ISL_422098 | Wales Specialist Virology Centre | Public Health Wales Microbiology Cardiff |
| EPI_ISL_422099 | Wales Specialist Virology Centre | Public Health Wales Microbiology Cardiff |
| EPI_ISL_422100 | Wales Specialist Virology Centre | Public Health Wales Microbiology Cardiff |
| EPI_ISL_422102 | Wales Specialist Virology Centre | Public Health Wales Microbiology Cardiff |
| EPI_ISL_422103 | Wales Specialist Virology Centre | Public Health Wales Microbiology Cardiff |
| EPI_ISL_422104 | Wales Specialist Virology Centre | Public Health Wales Microbiology Cardiff |
| EPI_ISL_422105 | Wales Specialist Virology Centre | Public Health Wales Microbiology Cardiff |
| EPI_ISL_422106 | Wales Specialist Virology Centre | Public Health Wales Microbiology Cardiff |
| EPI_ISL_422107 | Wales Specialist Virology Centre | Public Health Wales Microbiology Cardiff |

| EPI_Accession_ID | Originating Laboratory | Submitting Laboratory |
|---|---|---|
| EPI_ISL_422108 | Wales Specialist Virology Centre | Public Health Wales Microbiology Cardiff |
| EPI_ISL_422109 | Wales Specialist Virology Centre | Public Health Wales Microbiology Cardiff |
| EPI_ISL_422110 | Wales Specialist Virology Centre | Public Health Wales Microbiology Cardiff |
| EPI_ISL_422111 | Wales Specialist Virology Centre | Public Health Wales Microbiology Cardiff |
| EPI_ISL_422112 | Wales Specialist Virology Centre | Public Health Wales Microbiology Cardiff |
| EPI_ISL_422113 | Wales Specialist Virology Centre | Public Health Wales Microbiology Cardiff |
| EPI_ISL_422114 | Wales Specialist Virology Centre | Public Health Wales Microbiology Cardiff |
| EPI_ISL_422115 | Wales Specialist Virology Centre | Public Health Wales Microbiology Cardiff |
| EPI_ISL_422116 | Wales Specialist Virology Centre | Public Health Wales Microbiology Cardiff |
| EPI_ISL_422117 | Wales Specialist Virology Centre | Public Health Wales Microbiology Cardiff |
| EPI_ISL_422118 | Wales Specialist Virology Centre | Public Health Wales Microbiology Cardiff |
| EPI_ISL_422119 | Wales Specialist Virology Centre | Public Health Wales Microbiology Cardiff |
| EPI_ISL_422120 | Wales Specialist Virology Centre | Public Health Wales Microbiology Cardiff |
| EPI_ISL_422121 | Wales Specialist Virology Centre | Public Health Wales Microbiology Cardiff |
| EPI_ISL_422122 | Wales Specialist Virology Centre | Public Health Wales Microbiology Cardiff |
| EPI_ISL_422123 | Wales Specialist Virology Centre | Public Health Wales Microbiology Cardiff |
| EPI_ISL_422124 | Wales Specialist Virology Centre | Public Health Wales Microbiology Cardiff |
| EPI_ISL_422125 | Wales Specialist Virology Centre | Public Health Wales Microbiology Cardiff |
| EPI_ISL_422126 | Wales Specialist Virology Centre | Public Health Wales Microbiology Cardiff |
| EPI_ISL_422127 | Wales Specialist Virology Centre | Public Health Wales Microbiology Cardiff |
| EPI_ISL_422128 | Wales Specialist Virology Centre | Public Health Wales Microbiology Cardiff |
| EPI_ISL_422129 | Wales Specialist Virology Centre | Public Health Wales Microbiology Cardiff |
| EPI_ISL_422130 | Wales Specialist Virology Centre | Public Health Wales Microbiology Cardiff |
| EPI_ISL_422131 | Wales Specialist Virology Centre | Public Health Wales Microbiology Cardiff |
| EPI_ISL_422132 | Wales Specialist Virology Centre | Public Health Wales Microbiology Cardiff |
| EPI_ISL_422133 | Wales Specialist Virology Centre | Public Health Wales Microbiology Cardiff |
| EPI_ISL_422134 | Wales Specialist Virology Centre | Public Health Wales Microbiology Cardiff |
| EPI_ISL_422135 | Wales Specialist Virology Centre | Public Health Wales Microbiology Cardiff |
| EPI_ISL_422136 | Wales Specialist Virology Centre | Public Health Wales Microbiology Cardiff |
| EPI_ISL_422137 | Wales Specialist Virology Centre | Public Health Wales Microbiology Cardiff |
| EPI_ISL_422138 | Wales Specialist Virology Centre | Public Health Wales Microbiology Cardiff |
| EPI_ISL_422140 | Wales Specialist Virology Centre | Public Health Wales Microbiology Cardiff |
| EPI_ISL_422141 | Wales Specialist Virology Centre | Public Health Wales Microbiology Cardiff |

| EPI_Accession_ID | Originating Laboratory | Submitting Laboratory |
|---|---|---|
| EPI_ISL_422142 | Wales Specialist Virology Centre | Public Health Wales Microbiology Cardiff |
| EPI_ISL_422143 | Wales Specialist Virology Centre | Public Health Wales Microbiology Cardiff |
| EPI_ISL_422144 | Wales Specialist Virology Centre | Public Health Wales Microbiology Cardiff |
| EPI_ISL_422145 | Wales Specialist Virology Centre | Public Health Wales Microbiology Cardiff |
| EPI_ISL_422146 | Wales Specialist Virology Centre | Public Health Wales Microbiology Cardiff |
| EPI_ISL_422147 | Wales Specialist Virology Centre | Public Health Wales Microbiology Cardiff |
| EPI_ISL_422148 | Wales Specialist Virology Centre | Public Health Wales Microbiology Cardiff |
| EPI_ISL_422149 | Wales Specialist Virology Centre | Public Health Wales Microbiology Cardiff |
| EPI_ISL_422150 | Wales Specialist Virology Centre | Public Health Wales Microbiology Cardiff |
| EPI_ISL_422151 | Wales Specialist Virology Centre | Public Health Wales Microbiology Cardiff |
| EPI_ISL_422152 | Wales Specialist Virology Centre | Public Health Wales Microbiology Cardiff |
| EPI_ISL_422153 | Wales Specialist Virology Centre | Public Health Wales Microbiology Cardiff |
| EPI_ISL_422154 | Wales Specialist Virology Centre | Public Health Wales Microbiology Cardiff |
| EPI_ISL_422155 | Wales Specialist Virology Centre | Public Health Wales Microbiology Cardiff |
| EPI_ISL_422156 | Wales Specialist Virology Centre | Public Health Wales Microbiology Cardiff |
| EPI_ISL_422157 | Wales Specialist Virology Centre | Public Health Wales Microbiology Cardiff |
| EPI_ISL_422158 | Wales Specialist Virology Centre | Public Health Wales Microbiology Cardiff |
| EPI_ISL_422159 | Wales Specialist Virology Centre | Public Health Wales Microbiology Cardiff |
| EPI_ISL_422160 | Wales Specialist Virology Centre | Public Health Wales Microbiology Cardiff |
| EPI_ISL_422161 | Wales Specialist Virology Centre | Public Health Wales Microbiology Cardiff |
| EPI_ISL_422162 | Wales Specialist Virology Centre | Public Health Wales Microbiology Cardiff |
| EPI_ISL_422163 | Wales Specialist Virology Centre | Public Health Wales Microbiology Cardiff |
| EPI_ISL_422164 | Wales Specialist Virology Centre | Public Health Wales Microbiology Cardiff |
| EPI_ISL_422165 | Wales Specialist Virology Centre | Public Health Wales Microbiology Cardiff |
| EPI_ISL_422166 | Wales Specialist Virology Centre | Public Health Wales Microbiology Cardiff |
| EPI_ISL_422167 | Wales Specialist Virology Centre | Public Health Wales Microbiology Cardiff |
| EPI_ISL_422168 | Wales Specialist Virology Centre | Public Health Wales Microbiology Cardiff |
| EPI_ISL_422169 | Wales Specialist Virology Centre | Public Health Wales Microbiology Cardiff |
| EPI_ISL_422170 | Wales Specialist Virology Centre | Public Health Wales Microbiology Cardiff |
| EPI_ISL_422171 | Wales Specialist Virology Centre | Public Health Wales Microbiology Cardiff |
| EPI_ISL_422172 | Wales Specialist Virology Centre | Public Health Wales Microbiology Cardiff |
| EPI_ISL_422173 | Wales Specialist Virology Centre | Public Health Wales Microbiology Cardiff |
| EPI_ISL_422174 | Wales Specialist Virology Centre | Public Health Wales Microbiology Cardiff |

| EPI_Accession_ID | Originating Laboratory | Submitting Laboratory |
|---|---|---|
| EPI_ISL_422175 | Wales Specialist Virology Centre | Public Health Wales Microbiology Cardiff |
| EPI_ISL_422176 | Wales Specialist Virology Centre | Public Health Wales Microbiology Cardiff |
| EPI_ISL_422177 | Wales Specialist Virology Centre | Public Health Wales Microbiology Cardiff |
| EPI_ISL_422178 | Wales Specialist Virology Centre | Public Health Wales Microbiology Cardiff |
| EPI_ISL_422179 | Wales Specialist Virology Centre | Public Health Wales Microbiology Cardiff |
| EPI_ISL_422180 | Wales Specialist Virology Centre | Public Health Wales Microbiology Cardiff |
| EPI_ISL_422181 | Wales Specialist Virology Centre | Public Health Wales Microbiology Cardiff |
| EPI_ISL_422182 | Wales Specialist Virology Centre | Public Health Wales Microbiology Cardiff |
| EPI_ISL_422183 | Wales Specialist Virology Centre | Public Health Wales Microbiology Cardiff |
| EPI_ISL_422184 | Wales Specialist Virology Centre | Public Health Wales Microbiology Cardiff |
| EPI_ISL_422185 | Wales Specialist Virology Centre | Public Health Wales Microbiology Cardiff |
| EPI_ISL_422186 | Wales Specialist Virology Centre | Public Health Wales Microbiology Cardiff |
| EPI_ISL_422187 | Wales Specialist Virology Centre | Public Health Wales Microbiology Cardiff |
| EPI_ISL_422188 | Wales Specialist Virology Centre | Public Health Wales Microbiology Cardiff |
| EPI_ISL_422189 | Wales Specialist Virology Centre | Public Health Wales Microbiology Cardiff |
| EPI_ISL_422190 | Wales Specialist Virology Centre | Public Health Wales Microbiology Cardiff |
| EPI_ISL_422191 | Wales Specialist Virology Centre | Public Health Wales Microbiology Cardiff |
| EPI_ISL_422192 | Wales Specialist Virology Centre | Public Health Wales Microbiology Cardiff |
| EPI_ISL_422193 | Wales Specialist Virology Centre | Public Health Wales Microbiology Cardiff |
| EPI_ISL_422194 | Wales Specialist Virology Centre | Public Health Wales Microbiology Cardiff |
| EPI_ISL_422195 | Wales Specialist Virology Centre | Public Health Wales Microbiology Cardiff |
| EPI_ISL_422196 | Wales Specialist Virology Centre | Public Health Wales Microbiology Cardiff |
| EPI_ISL_422197 | Wales Specialist Virology Centre | Public Health Wales Microbiology Cardiff |
| EPI_ISL_422198 | Wales Specialist Virology Centre | Public Health Wales Microbiology Cardiff |
| EPI_ISL_422199 | Wales Specialist Virology Centre | Public Health Wales Microbiology Cardiff |
| EPI_ISL_422200 | Wales Specialist Virology Centre | Public Health Wales Microbiology Cardiff |
| EPI_ISL_422201 | Wales Specialist Virology Centre | Public Health Wales Microbiology Cardiff |
| EPI_ISL_422202 | Wales Specialist Virology Centre | Public Health Wales Microbiology Cardiff |
| EPI_ISL_422203 | Wales Specialist Virology Centre | Public Health Wales Microbiology Cardiff |
| EPI_ISL_422204 | Wales Specialist Virology Centre | Public Health Wales Microbiology Cardiff |
| EPI_ISL_422205 | Wales Specialist Virology Centre | Public Health Wales Microbiology Cardiff |
| EPI_ISL_422206 | Wales Specialist Virology Centre | Public Health Wales Microbiology Cardiff |
| EPI_ISL_422207 | Wales Specialist Virology Centre | Public Health Wales Microbiology Cardiff |

| EPI_Accession_ID | Originating Laboratory | Submitting Laboratory |
|---|---|---|
| EPI_ISL_422208 | Wales Specialist Virology Centre | Public Health Wales Microbiology Cardiff |
| EPI_ISL_422209 | Wales Specialist Virology Centre | Public Health Wales Microbiology Cardiff |
| EPI_ISL_422210 | Wales Specialist Virology Centre | Public Health Wales Microbiology Cardiff |
| EPI_ISL_422211 | Wales Specialist Virology Centre | Public Health Wales Microbiology Cardiff |
| EPI_ISL_422212 | Wales Specialist Virology Centre | Public Health Wales Microbiology Cardiff |
| EPI_ISL_422213 | Wales Specialist Virology Centre | Public Health Wales Microbiology Cardiff |
| EPI_ISL_422214 | Wales Specialist Virology Centre | Public Health Wales Microbiology Cardiff |
| EPI_ISL_422215 | Wales Specialist Virology Centre | Public Health Wales Microbiology Cardiff |
| EPI_ISL_422216 | Wales Specialist Virology Centre | Public Health Wales Microbiology Cardiff |
| EPI_ISL_422217 | Wales Specialist Virology Centre | Public Health Wales Microbiology Cardiff |
| EPI_ISL_422218 | Wales Specialist Virology Centre | Public Health Wales Microbiology Cardiff |
| EPI_ISL_422219 | Wales Specialist Virology Centre | Public Health Wales Microbiology Cardiff |
| EPI_ISL_422220 | Wales Specialist Virology Centre | Public Health Wales Microbiology Cardiff |
| EPI_ISL_422221 | Wales Specialist Virology Centre | Public Health Wales Microbiology Cardiff |
| EPI_ISL_422222 | Wales Specialist Virology Centre | Public Health Wales Microbiology Cardiff |
| EPI_ISL_422223 | Wales Specialist Virology Centre | Public Health Wales Microbiology Cardiff |
| EPI_ISL_422224 | Wales Specialist Virology Centre | Public Health Wales Microbiology Cardiff |
| EPI_ISL_422225 | Wales Specialist Virology Centre | Public Health Wales Microbiology Cardiff |
| EPI_ISL_422226 | Wales Specialist Virology Centre | Public Health Wales Microbiology Cardiff |
| EPI_ISL_422227 | Wales Specialist Virology Centre | Public Health Wales Microbiology Cardiff |
| EPI_ISL_422228 | Wales Specialist Virology Centre | Public Health Wales Microbiology Cardiff |
| EPI_ISL_422229 | Wales Specialist Virology Centre | Public Health Wales Microbiology Cardiff |
| EPI_ISL_422230 | Wales Specialist Virology Centre | Public Health Wales Microbiology Cardiff |
| EPI_ISL_422231 | Wales Specialist Virology Centre | Public Health Wales Microbiology Cardiff |
| EPI_ISL_422232 | Wales Specialist Virology Centre | Public Health Wales Microbiology Cardiff |
| EPI_ISL_422233 | Wales Specialist Virology Centre | Public Health Wales Microbiology Cardiff |
| EPI_ISL_422234 | Wales Specialist Virology Centre | Public Health Wales Microbiology Cardiff |
| EPI_ISL_422235 | Wales Specialist Virology Centre | Public Health Wales Microbiology Cardiff |
| EPI_ISL_422236 | Wales Specialist Virology Centre | Public Health Wales Microbiology Cardiff |
| EPI_ISL_422237 | Wales Specialist Virology Centre | Public Health Wales Microbiology Cardiff |
| EPI_ISL_422238 | Wales Specialist Virology Centre | Public Health Wales Microbiology Cardiff |
| EPI_ISL_422239 | Wales Specialist Virology Centre | Public Health Wales Microbiology Cardiff |
| EPI_ISL_422240 | Wales Specialist Virology Centre | Public Health Wales Microbiology Cardiff |

| EPI_Accession_ID | Originating Laboratory | Submitting Laboratory |
|---|---|---|
| EPI_ISL_422241 | Wales Specialist Virology Centre | Public Health Wales Microbiology Cardiff |
| EPI_ISL_422242 | Wales Specialist Virology Centre | Public Health Wales Microbiology Cardiff |
| EPI_ISL_422243 | Wales Specialist Virology Centre | Public Health Wales Microbiology Cardiff |
| EPI_ISL_422244 | Wales Specialist Virology Centre | Public Health Wales Microbiology Cardiff |
| EPI_ISL_422245 | Wales Specialist Virology Centre | Public Health Wales Microbiology Cardiff |
| EPI_ISL_422246 | Wales Specialist Virology Centre | Public Health Wales Microbiology Cardiff |
| EPI_ISL_422247 | Wales Specialist Virology Centre | Public Health Wales Microbiology Cardiff |
| EPI_ISL_422248 | Wales Specialist Virology Centre | Public Health Wales Microbiology Cardiff |
| EPI_ISL_422249 | Wales Specialist Virology Centre | Public Health Wales Microbiology Cardiff |
| EPI_ISL_422250 | Wales Specialist Virology Centre | Public Health Wales Microbiology Cardiff |
| EPI_ISL_422251 | Wales Specialist Virology Centre | Public Health Wales Microbiology Cardiff |
| EPI_ISL_422252 | Wales Specialist Virology Centre | Public Health Wales Microbiology Cardiff |
| EPI_ISL_422253 | Wales Specialist Virology Centre | Public Health Wales Microbiology Cardiff |
| EPI_ISL_422254 | Wales Specialist Virology Centre | Public Health Wales Microbiology Cardiff |
| EPI_ISL_422255 | Wales Specialist Virology Centre | Public Health Wales Microbiology Cardiff |
| EPI_ISL_422256 | Wales Specialist Virology Centre | Public Health Wales Microbiology Cardiff |
| EPI_ISL_422257 | Wales Specialist Virology Centre | Public Health Wales Microbiology Cardiff |
| EPI_ISL_422258 | Wales Specialist Virology Centre | Public Health Wales Microbiology Cardiff |
| EPI_ISL_422259 | Wales Specialist Virology Centre | Public Health Wales Microbiology Cardiff |
| EPI_ISL_422260 | Wales Specialist Virology Centre | Public Health Wales Microbiology Cardiff |
| EPI_ISL_422261 | Wales Specialist Virology Centre | Public Health Wales Microbiology Cardiff |
| EPI_ISL_422262 | Wales Specialist Virology Centre | Public Health Wales Microbiology Cardiff |
| EPI_ISL_422263 | Wales Specialist Virology Centre | Public Health Wales Microbiology Cardiff |
| EPI_ISL_422264 | Wales Specialist Virology Centre | Public Health Wales Microbiology Cardiff |
| EPI_ISL_422265 | Wales Specialist Virology Centre | Public Health Wales Microbiology Cardiff |
| EPI_ISL_422266 | Wales Specialist Virology Centre | Public Health Wales Microbiology Cardiff |
| EPI_ISL_422267 | Wales Specialist Virology Centre | Public Health Wales Microbiology Cardiff |
| EPI_ISL_422268 | Wales Specialist Virology Centre | Public Health Wales Microbiology Cardiff |
| EPI_ISL_422269 | Wales Specialist Virology Centre | Public Health Wales Microbiology Cardiff |
| EPI_ISL_422270 | Wales Specialist Virology Centre | Public Health Wales Microbiology Cardiff |
| EPI_ISL_422271 | Wales Specialist Virology Centre | Public Health Wales Microbiology Cardiff |
| EPI_ISL_422272 | Wales Specialist Virology Centre | Public Health Wales Microbiology Cardiff |
| EPI_ISL_422273 | Wales Specialist Virology Centre | Public Health Wales Microbiology Cardiff |

| EPI_Accession_ID | Originating Laboratory | Submitting Laboratory |
| --- | --- | --- |
| EPI_ISL_422274 | Wales Specialist Virology Centre | Public Health Wales Microbiology Cardiff |
| EPI_ISL_422275 | Wales Specialist Virology Centre | Public Health Wales Microbiology Cardiff |
| EPI_ISL_422276 | Wales Specialist Virology Centre | Public Health Wales Microbiology Cardiff |
| EPI_ISL_422277 | Wales Specialist Virology Centre | Public Health Wales Microbiology Cardiff |
| EPI_ISL_422278 | Wales Specialist Virology Centre | Public Health Wales Microbiology Cardiff |
| EPI_ISL_422279 | Wales Specialist Virology Centre | Public Health Wales Microbiology Cardiff |
| EPI_ISL_422280 | Wales Specialist Virology Centre | Public Health Wales Microbiology Cardiff |
| EPI_ISL_422281 | Wales Specialist Virology Centre | Public Health Wales Microbiology Cardiff |
| EPI_ISL_422282 | Wales Specialist Virology Centre | Public Health Wales Microbiology Cardiff |
| EPI_ISL_422283 | Wales Specialist Virology Centre | Public Health Wales Microbiology Cardiff |
| EPI_ISL_422284 | Wales Specialist Virology Centre | Public Health Wales Microbiology Cardiff |
| EPI_ISL_422285 | Wales Specialist Virology Centre | Public Health Wales Microbiology Cardiff |
| EPI_ISL_422286 | Wales Specialist Virology Centre | Public Health Wales Microbiology Cardiff |
| EPI_ISL_422287 | Wales Specialist Virology Centre | Public Health Wales Microbiology Cardiff |
| EPI_ISL_422288 | Wales Specialist Virology Centre | Public Health Wales Microbiology Cardiff |
| EPI_ISL_422289 | Wales Specialist Virology Centre | Public Health Wales Microbiology Cardiff |
| EPI_ISL_422290 | Wales Specialist Virology Centre | Public Health Wales Microbiology Cardiff |
| EPI_ISL_422291 | Wales Specialist Virology Centre | Public Health Wales Microbiology Cardiff |
| EPI_ISL_422292 | Wales Specialist Virology Centre | Public Health Wales Microbiology Cardiff |
| EPI_ISL_422293 | Wales Specialist Virology Centre | Public Health Wales Microbiology Cardiff |
| EPI_ISL_422294 | Wales Specialist Virology Centre | Public Health Wales Microbiology Cardiff |
| EPI_ISL_422295 | Wales Specialist Virology Centre | Public Health Wales Microbiology Cardiff |
| EPI_ISL_422296 | Wales Specialist Virology Centre | Public Health Wales Microbiology Cardiff |
| EPI_ISL_422297 | Wales Specialist Virology Centre | Public Health Wales Microbiology Cardiff |
| EPI_ISL_422298 | Wales Specialist Virology Centre | Public Health Wales Microbiology Cardiff |
| EPI_ISL_422299 | Wales Specialist Virology Centre | Public Health Wales Microbiology Cardiff |
| EPI_ISL_422300 | Wales Specialist Virology Centre | Public Health Wales Microbiology Cardiff |
| EPI_ISL_422301 | Wales Specialist Virology Centre | Public Health Wales Microbiology Cardiff |
| EPI_ISL_422302 | Wales Specialist Virology Centre | Public Health Wales Microbiology Cardiff |
| EPI_ISL_422303 | Wales Specialist Virology Centre | Public Health Wales Microbiology Cardiff |
| EPI_ISL_422304 | Wales Specialist Virology Centre | Public Health Wales Microbiology Cardiff |
| EPI_ISL_422305 | Wales Specialist Virology Centre | Public Health Wales Microbiology Cardiff |
| EPI_ISL_422306 | Wales Specialist Virology Centre | Public Health Wales Microbiology Cardiff |

| EPI_Accession_ID | Originating Laboratory | Submitting Laboratory |
|---|---|---|
| EPI_ISL_422307 | Wales Specialist Virology Centre | Public Health Wales Microbiology Cardiff |
| EPI_ISL_422308 | Wales Specialist Virology Centre | Public Health Wales Microbiology Cardiff |
| EPI_ISL_422309 | Wales Specialist Virology Centre | Public Health Wales Microbiology Cardiff |
| EPI_ISL_422310 | Wales Specialist Virology Centre | Public Health Wales Microbiology Cardiff |
| EPI_ISL_422311 | Wales Specialist Virology Centre | Public Health Wales Microbiology Cardiff |
| EPI_ISL_422312 | Wales Specialist Virology Centre | Public Health Wales Microbiology Cardiff |
| EPI_ISL_422313 | Wales Specialist Virology Centre | Public Health Wales Microbiology Cardiff |
| EPI_ISL_422314 | Wales Specialist Virology Centre | Public Health Wales Microbiology Cardiff |
| EPI_ISL_422315 | Wales Specialist Virology Centre | Public Health Wales Microbiology Cardiff |
| EPI_ISL_422316 | Wales Specialist Virology Centre | Public Health Wales Microbiology Cardiff |
| EPI_ISL_422317 | Wales Specialist Virology Centre | Public Health Wales Microbiology Cardiff |
| EPI_ISL_422318 | Wales Specialist Virology Centre | Public Health Wales Microbiology Cardiff |
| EPI_ISL_422319 | Wales Specialist Virology Centre | Public Health Wales Microbiology Cardiff |
| EPI_ISL_422320 | Wales Specialist Virology Centre | Public Health Wales Microbiology Cardiff |
| EPI_ISL_422321 | Wales Specialist Virology Centre | Public Health Wales Microbiology Cardiff |
| EPI_ISL_422322 | Wales Specialist Virology Centre | Public Health Wales Microbiology Cardiff |
| EPI_ISL_422323 | Wales Specialist Virology Centre | Public Health Wales Microbiology Cardiff |
| EPI_ISL_422324 | Wales Specialist Virology Centre | Public Health Wales Microbiology Cardiff |
| EPI_ISL_422325 | Wales Specialist Virology Centre | Public Health Wales Microbiology Cardiff |
| EPI_ISL_422326 | Wales Specialist Virology Centre | Public Health Wales Microbiology Cardiff |
| EPI_ISL_422327 | Wales Specialist Virology Centre | Public Health Wales Microbiology Cardiff |
| EPI_ISL_422328 | Wales Specialist Virology Centre | Public Health Wales Microbiology Cardiff |
| EPI_ISL_422329 | Wales Specialist Virology Centre | Public Health Wales Microbiology Cardiff |
| EPI_ISL_422330 | Wales Specialist Virology Centre | Public Health Wales Microbiology Cardiff |
| EPI_ISL_422331 | Wales Specialist Virology Centre | Public Health Wales Microbiology Cardiff |
| EPI_ISL_422332 | Wales Specialist Virology Centre | Public Health Wales Microbiology Cardiff |
| EPI_ISL_422333 | Wales Specialist Virology Centre | Public Health Wales Microbiology Cardiff |
| EPI_ISL_422334 | Wales Specialist Virology Centre | Public Health Wales Microbiology Cardiff |
| EPI_ISL_422335 | Wales Specialist Virology Centre | Public Health Wales Microbiology Cardiff |
| EPI_ISL_422336 | Wales Specialist Virology Centre | Public Health Wales Microbiology Cardiff |
| EPI_ISL_422337 | Wales Specialist Virology Centre | Public Health Wales Microbiology Cardiff |
| EPI_ISL_422338 | Wales Specialist Virology Centre | Public Health Wales Microbiology Cardiff |
| EPI_ISL_422339 | Wales Specialist Virology Centre | Public Health Wales Microbiology Cardiff |

| EPI_Accession_ID | Originating Laboratory | Submitting Laboratory |
|---|---|---|
| EPI_ISL_422340 | Wales Specialist Virology Centre | Public Health Wales Microbiology Cardiff |
| EPI_ISL_422341 | Wales Specialist Virology Centre | Public Health Wales Microbiology Cardiff |
| EPI_ISL_422342 | Wales Specialist Virology Centre | Public Health Wales Microbiology Cardiff |
| EPI_ISL_422343 | Wales Specialist Virology Centre | Public Health Wales Microbiology Cardiff |
| EPI_ISL_422344 | Wales Specialist Virology Centre | Public Health Wales Microbiology Cardiff |
| EPI_ISL_422345 | Wales Specialist Virology Centre | Public Health Wales Microbiology Cardiff |
| EPI_ISL_422346 | Wales Specialist Virology Centre | Public Health Wales Microbiology Cardiff |
| EPI_ISL_422347 | Wales Specialist Virology Centre | Public Health Wales Microbiology Cardiff |
| EPI_ISL_422348 | Wales Specialist Virology Centre | Public Health Wales Microbiology Cardiff |
| EPI_ISL_422349 | Wales Specialist Virology Centre | Public Health Wales Microbiology Cardiff |
| EPI_ISL_422350 | Wales Specialist Virology Centre | Public Health Wales Microbiology Cardiff |
| EPI_ISL_422351 | Wales Specialist Virology Centre | Public Health Wales Microbiology Cardiff |
| EPI_ISL_422352 | Wales Specialist Virology Centre | Public Health Wales Microbiology Cardiff |
| EPI_ISL_422353 | Wales Specialist Virology Centre | Public Health Wales Microbiology Cardiff |
| EPI_ISL_422354 | Wales Specialist Virology Centre | Public Health Wales Microbiology Cardiff |
| EPI_ISL_422355 | Wales Specialist Virology Centre | Public Health Wales Microbiology Cardiff |
| EPI_ISL_422356 | Wales Specialist Virology Centre | Public Health Wales Microbiology Cardiff |
| EPI_ISL_422357 | Wales Specialist Virology Centre | Public Health Wales Microbiology Cardiff |
| EPI_ISL_422358 | Wales Specialist Virology Centre | Public Health Wales Microbiology Cardiff |
| EPI_ISL_422359 | Wales Specialist Virology Centre | Public Health Wales Microbiology Cardiff |
| EPI_ISL_422360 | Wales Specialist Virology Centre | Public Health Wales Microbiology Cardiff |
| EPI_ISL_422361 | Wales Specialist Virology Centre | Public Health Wales Microbiology Cardiff |
| EPI_ISL_422362 | Wales Specialist Virology Centre | Public Health Wales Microbiology Cardiff |
| EPI_ISL_422363 | Wales Specialist Virology Centre | Public Health Wales Microbiology Cardiff |
| EPI_ISL_422364 | Wales Specialist Virology Centre | Public Health Wales Microbiology Cardiff |
| EPI_ISL_422365 | Wales Specialist Virology Centre | Public Health Wales Microbiology Cardiff |
| EPI_ISL_422366 | Wales Specialist Virology Centre | Public Health Wales Microbiology Cardiff |
| EPI_ISL_422368 | Wales Specialist Virology Centre | Public Health Wales Microbiology Cardiff |
| EPI_ISL_422369 | Wales Specialist Virology Centre | Public Health Wales Microbiology Cardiff |
| EPI_ISL_422370 | Wales Specialist Virology Centre | Public Health Wales Microbiology Cardiff |
| EPI_ISL_422371 | Wales Specialist Virology Centre | Public Health Wales Microbiology Cardiff |
| EPI_ISL_422372 | Wales Specialist Virology Centre | Public Health Wales Microbiology Cardiff |
| EPI_ISL_422373 | Wales Specialist Virology Centre | Public Health Wales Microbiology Cardiff |

| EPI_Accession_ID | Originating Laboratory | Submitting Laboratory |
|---|---|---|
| EPI_ISL_422374 | Wales Specialist Virology Centre | Public Health Wales Microbiology Cardiff |
| EPI_ISL_422375 | Wales Specialist Virology Centre | Public Health Wales Microbiology Cardiff |
| EPI_ISL_422376 | Wales Specialist Virology Centre | Public Health Wales Microbiology Cardiff |
| EPI_ISL_422377 | Wales Specialist Virology Centre | Public Health Wales Microbiology Cardiff |
| EPI_ISL_422384 | NMIMR, Department of Virology | WACCBIP, University of Ghana |
| EPI_ISL_422387 | NMIMR, Department of Virology | WACCBIP, University of Ghana |
| EPI_ISL_422390 | NMIMR, Department of Virology | WACCBIP, University of Ghana |
| EPI_ISL_422394 | NMIMR, Department of Virology | WACCBIP, University of Ghana |
| EPI_ISL_422397 | NMIMR, Department of Virology | WACCBIP, University of Ghana |
| EPI_ISL_422398 | NMIMR, Department of Virology | WACCBIP, University of Ghana |
| EPI_ISL_422399 | NMIMR, Department of Virology | WACCBIP, University of Ghana |
| EPI_ISL_422400 | NMIMR, Department of Virology | WACCBIP, University of Ghana |
| EPI_ISL_422401 | NMIMR, Department of Virology | WACCBIP, University of Ghana |
| EPI_ISL_422402 | NMIMR, Department of Virology | WACCBIP, University of Ghana |
| EPI_ISL_422403 | NMIMR, Department of Virology | WACCBIP, University of Ghana |
| EPI_ISL_422404 | NMIMR, Department of Virology | WACCBIP, University of Ghana |
| EPI_ISL_422405 | NMIMR, Department of Virology | WACCBIP, University of Ghana |
| EPI_ISL_422406 | NMIMR, Department of Virology | WACCBIP, University of Ghana |
| EPI_ISL_422407 | Department of Laboratory Medicine, National Taiwan University Hospital | Microbial Genomics Core Lab, National Taiwan University Centers of Genomic and Precision Medicine |
| EPI_ISL_422408 | Department of Laboratory Medicine, National Taiwan University Hospital | Microbial Genomics Core Lab, National Taiwan University Centers of Genomic and Precision Medicine |
| EPI_ISL_422409 | Department of Laboratory Medicine, National Taiwan University Hospital | Microbial Genomics Core Lab, National Taiwan University Centers of Genomic and Precision Medicine |
| EPI_ISL_422410 | Department of Laboratory Medicine, National Taiwan University Hospital | Microbial Genomics Core Lab, National Taiwan University Centers of Genomic and Precision Medicine |
| EPI_ISL_422411 | Department of Laboratory Medicine, National Taiwan University Hospital | Microbial Genomics Core Lab, National Taiwan University Centers of Genomic and Precision Medicine |
| EPI_ISL_422412 | Department of Laboratory Medicine, National Taiwan University Hospital | Microbial Genomics Core Lab, National Taiwan University Centers of Genomic and Precision Medicine |
| EPI_ISL_422413 | Department of Laboratory Medicine, National Taiwan University Hospital | Microbial Genomics Core Lab, National Taiwan University Centers of Genomic and Precision Medicine |

| EPI_Accession_ID | Originating Laboratory | Submitting Laboratory |
|---|---|---|
| EPI_ISL_422414 | Department of Laboratory Medicine, National Taiwan University Hospital | Microbial Genomics Core Lab, National Taiwan University Centers of Genomic and Precision Medicine |
| EPI_ISL_422415 | Department of Laboratory Medicine, National Taiwan University Hospital | Microbial Genomics Core Lab, National Taiwan University Centers of Genomic and Precision Medicine |
| EPI_ISL_422416 | Department of Laboratory Medicine, National Taiwan University Hospital | Microbial Genomics Core Lab, National Taiwan University Centers of Genomic and Precision Medicine |
| EPI_ISL_422417 | Department of Laboratory Medicine, National Taiwan University Hospital | Microbial Genomics Core Lab, National Taiwan University Centers of Genomic and Precision Medicine |
| EPI_ISL_422418 | Department of Laboratory Medicine, National Taiwan University Hospital | Microbial Genomics Core Lab, National Taiwan University Centers of Genomic and Precision Medicine |
| EPI_ISL_422419 | Department of Laboratory Medicine, National Taiwan University Hospital | Microbial Genomics Core Lab, National Taiwan University Centers of Genomic and Precision Medicine |
| EPI_ISL_422420 | Department of Laboratory Medicine, National Taiwan University Hospital | Microbial Genomics Core Lab, National Taiwan University Centers of Genomic and Precision Medicine |
| EPI_ISL_422421 | Department of Laboratory Medicine, National Taiwan University Hospital | Microbial Genomics Core Lab, National Taiwan University Centers of Genomic and Precision Medicine |
| EPI_ISL_422422 | Department of Laboratory Medicine, National Taiwan University Hospital | Microbial Genomics Core Lab, National Taiwan University Centers of Genomic and Precision Medicine |
| EPI_ISL_422424 | Jaber Al Ahmad Al Sabah Hospital | Dasman diabetes Institute |
| EPI_ISL_422425 | Zhejiang Provincial Center for Disease Control and Prevention | Zhejiang Provincial Center for Disease Control and Prevention |
| EPI_ISL_422426 | JABER AL AHMAD AL SABAH HOSPITAL â€“ KUWAIT CITY | Dasman Diabetes Institute |
| EPI_ISL_422427 | JABER AL AHMAD AL SABAH HOSPITAL â€“ KUWAIT CITY | Dasman Diabetes Institute |
| EPI_ISL_422428 | National Public Health Laboratory, National Centre for Infectious Diseases | National Public Health Laboratory, National Centre for Infectious Diseases |
| EPI_ISL_422429 | National Public Health Laboratory, National Centre for Infectious Diseases | National Public Health Laboratory, National Centre for Infectious Diseases |
| EPI_ISL_422430 | National Public Health Laboratory, National Centre for Infectious Diseases | National Public Health Laboratory, National Centre for Infectious Diseases |
| EPI_ISL_422431 | National Public Health Laboratory, National Centre for Infectious Diseases | National Public Health Laboratory, National Centre for Infectious Diseases |
| EPI_ISL_422432 | National Public Health Laboratory, National Centre for Infectious Diseases | National Public Health Laboratory, National Centre for Infectious Diseases |

| EPI_Accession_ID | Originating Laboratory | Submitting Laboratory |
|---|---|---|
| EPI_ISL_422433 | National Public Health Laboratory, National Centre for Infectious Diseases | National Public Health Laboratory, National Centre for Infectious Diseases |
| EPI_ISL_422434 | National Public Health Laboratory, National Centre for Infectious Diseases | National Public Health Laboratory, National Centre for Infectious Diseases |
| EPI_ISL_422435 | National Public Health Laboratory, National Centre for Infectious Diseases | National Public Health Laboratory, National Centre for Infectious Diseases |
| EPI_ISL_422436 | Respiratory Virus Unit, Microbiology Services Colindale, Public Health England | Respiratory Virus Unit, Microbiology Services Colindale, Public Health England |
| EPI_ISL_422437 | ULSS9 Distretto di Bussolengo | Istituto Zooprofilattico Sperimentale delle Venezie |
| EPI_ISL_422438 | ULSS9 Distretto di Bussolengo | Istituto Zooprofilattico Sperimentale delle Venezie |
| EPI_ISL_422453 | Gundersen Molecular Diagnostics Laboratory | Kabara Cancer Research Institute |
| EPI_ISL_422459 | Gundersen Molecular Diagnostics Laboratory | Kabara Cancer Research Institute |
| EPI_ISL_422461 | Gundersen Molecular Diagnostics Laboratory | Kabara Cancer Research Institute |
| EPI_ISL_422462 | Gundersen Molecular Diagnostics Laboratory | Kabara Cancer Research Institute |
| EPI_ISL_422463 | Gundersen Molecular Diagnostics Laboratory | Kabara Cancer Research Institute |
| EPI_ISL_422465 | Gundersen Molecular Diagnostics Laboratory | Kabara Cancer Research Institute |
| EPI_ISL_422488 | MSHS Clinical Microbiology Laboratories | MSHS Pathogen Surveillance Program |
| EPI_ISL_422489 | MSHS Clinical Microbiology Laboratories | MSHS Pathogen Surveillance Program |
| EPI_ISL_422490 | MSHS Clinical Microbiology Laboratories | MSHS Pathogen Surveillance Program |
| EPI_ISL_422491 | MSHS Clinical Microbiology Laboratories | MSHS Pathogen Surveillance Program |
| EPI_ISL_422492 | MSHS Clinical Microbiology Laboratories | MSHS Pathogen Surveillance Program |
| EPI_ISL_422493 | MSHS Clinical Microbiology Laboratories | MSHS Pathogen Surveillance Program |
| EPI_ISL_422494 | MSHS Clinical Microbiology Laboratories | MSHS Pathogen Surveillance Program |
| EPI_ISL_422495 | MSHS Clinical Microbiology Laboratories | MSHS Pathogen Surveillance Program |
| EPI_ISL_422496 | MSHS Clinical Microbiology Laboratories | MSHS Pathogen Surveillance Program |
| EPI_ISL_422497 | MSHS Clinical Microbiology Laboratories | MSHS Pathogen Surveillance Program |
| EPI_ISL_422498 | MSHS Clinical Microbiology Laboratories | MSHS Pathogen Surveillance Program |
| EPI_ISL_422499 | MSHS Clinical Microbiology Laboratories | MSHS Pathogen Surveillance Program |
| EPI_ISL_422500 | MSHS Clinical Microbiology Laboratories | MSHS Pathogen Surveillance Program |
| EPI_ISL_422501 | MSHS Clinical Microbiology Laboratories | MSHS Pathogen Surveillance Program |
| EPI_ISL_422502 | MSHS Clinical Microbiology Laboratories | MSHS Pathogen Surveillance Program |
| EPI_ISL_422503 | MSHS Clinical Microbiology Laboratories | MSHS Pathogen Surveillance Program |
| EPI_ISL_422504 | MSHS Clinical Microbiology Laboratories | MSHS Pathogen Surveillance Program |

| EPI_Accession_ID | Originating Laboratory | Submitting Laboratory |
|---|---|---|
| EPI_ISL_422505 | MSHS Clinical Microbiology Laboratories | MSHS Pathogen Surveillance Program |
| EPI_ISL_422506 | MSHS Clinical Microbiology Laboratories | MSHS Pathogen Surveillance Program |
| EPI_ISL_422507 | MSHS Clinical Microbiology Laboratories | MSHS Pathogen Surveillance Program |
| EPI_ISL_422508 | MSHS Clinical Microbiology Laboratories | MSHS Pathogen Surveillance Program |
| EPI_ISL_422509 | MSHS Clinical Microbiology Laboratories | MSHS Pathogen Surveillance Program |
| EPI_ISL_422510 | MSHS Clinical Microbiology Laboratories | MSHS Pathogen Surveillance Program |
| EPI_ISL_422511 | MSHS Clinical Microbiology Laboratories | MSHS Pathogen Surveillance Program |
| EPI_ISL_422512 | MSHS Clinical Microbiology Laboratories | MSHS Pathogen Surveillance Program |
| EPI_ISL_422513 | MSHS Clinical Microbiology Laboratories | MSHS Pathogen Surveillance Program |
| EPI_ISL_422514 | MSHS Clinical Microbiology Laboratories | MSHS Pathogen Surveillance Program |
| EPI_ISL_422515 | MSHS Clinical Microbiology Laboratories | MSHS Pathogen Surveillance Program |
| EPI_ISL_422516 | MSHS Clinical Microbiology Laboratories | MSHS Pathogen Surveillance Program |
| EPI_ISL_422517 | MSHS Clinical Microbiology Laboratories | MSHS Pathogen Surveillance Program |
| EPI_ISL_422518 | MSHS Clinical Microbiology Laboratories | MSHS Pathogen Surveillance Program |
| EPI_ISL_422519 | MSHS Clinical Microbiology Laboratories | MSHS Pathogen Surveillance Program |
| EPI_ISL_422520 | MSHS Clinical Microbiology Laboratories | MSHS Pathogen Surveillance Program |
| EPI_ISL_422521 | MSHS Clinical Microbiology Laboratories | MSHS Pathogen Surveillance Program |
| EPI_ISL_422522 | MSHS Clinical Microbiology Laboratories | MSHS Pathogen Surveillance Program |
| EPI_ISL_422523 | MSHS Clinical Microbiology Laboratories | MSHS Pathogen Surveillance Program |
| EPI_ISL_422524 | MSHS Clinical Microbiology Laboratories | MSHS Pathogen Surveillance Program |
| EPI_ISL_422525 | MSHS Clinical Microbiology Laboratories | MSHS Pathogen Surveillance Program |
| EPI_ISL_422526 | MSHS Clinical Microbiology Laboratories | MSHS Pathogen Surveillance Program |
| EPI_ISL_422527 | MSHS Clinical Microbiology Laboratories | MSHS Pathogen Surveillance Program |
| EPI_ISL_422528 | MSHS Clinical Microbiology Laboratories | MSHS Pathogen Surveillance Program |
| EPI_ISL_422529 | MSHS Clinical Microbiology Laboratories | MSHS Pathogen Surveillance Program |
| EPI_ISL_422530 | MSHS Clinical Microbiology Laboratories | MSHS Pathogen Surveillance Program |
| EPI_ISL_422531 | MSHS Clinical Microbiology Laboratories | MSHS Pathogen Surveillance Program |
| EPI_ISL_422532 | MSHS Clinical Microbiology Laboratories | MSHS Pathogen Surveillance Program |
| EPI_ISL_422533 | MSHS Clinical Microbiology Laboratories | MSHS Pathogen Surveillance Program |
| EPI_ISL_422534 | MSHS Clinical Microbiology Laboratories | MSHS Pathogen Surveillance Program |
| EPI_ISL_422535 | MSHS Clinical Microbiology Laboratories | MSHS Pathogen Surveillance Program |
| EPI_ISL_422536 | MSHS Clinical Microbiology Laboratories | MSHS Pathogen Surveillance Program |
| EPI_ISL_422537 | MSHS Clinical Microbiology Laboratories | MSHS Pathogen Surveillance Program |

| EPI_Accession_ID | Originating Laboratory | Submitting Laboratory |
|---|---|---|
| EPI_ISL_422538 | MSHS Clinical Microbiology Laboratories | MSHS Pathogen Surveillance Program |
| EPI_ISL_422539 | MSHS Clinical Microbiology Laboratories | MSHS Pathogen Surveillance Program |
| EPI_ISL_422540 | MSHS Clinical Microbiology Laboratories | MSHS Pathogen Surveillance Program |
| EPI_ISL_422541 | MSHS Clinical Microbiology Laboratories | MSHS Pathogen Surveillance Program |
| EPI_ISL_422542 | MSHS Clinical Microbiology Laboratories | MSHS Pathogen Surveillance Program |
| EPI_ISL_422543 | MSHS Clinical Microbiology Laboratories | MSHS Pathogen Surveillance Program |
| EPI_ISL_422544 | MSHS Clinical Microbiology Laboratories | MSHS Pathogen Surveillance Program |
| EPI_ISL_422545 | MSHS Clinical Microbiology Laboratories | MSHS Pathogen Surveillance Program |
| EPI_ISL_422546 | MSHS Clinical Microbiology Laboratories | MSHS Pathogen Surveillance Program |
| EPI_ISL_422547 | MSHS Clinical Microbiology Laboratories | MSHS Pathogen Surveillance Program |
| EPI_ISL_422548 | MSHS Clinical Microbiology Laboratories | MSHS Pathogen Surveillance Program |
| EPI_ISL_422549 | MSHS Clinical Microbiology Laboratories | MSHS Pathogen Surveillance Program |
| EPI_ISL_422550 | MSHS Clinical Microbiology Laboratories | MSHS Pathogen Surveillance Program |
| EPI_ISL_422551 | MSHS Clinical Microbiology Laboratories | MSHS Pathogen Surveillance Program |
| EPI_ISL_422552 | MSHS Clinical Microbiology Laboratories | MSHS Pathogen Surveillance Program |
| EPI_ISL_422553 | MSHS Clinical Microbiology Laboratories | MSHS Pathogen Surveillance Program |
| EPI_ISL_422554 | MSHS Clinical Microbiology Laboratories | MSHS Pathogen Surveillance Program |
| EPI_ISL_422555 | MSHS Clinical Microbiology Laboratories | MSHS Pathogen Surveillance Program |
| EPI_ISL_422556 | MSHS Clinical Microbiology Laboratories | MSHS Pathogen Surveillance Program |
| EPI_ISL_422557 | MSHS Clinical Microbiology Laboratories | MSHS Pathogen Surveillance Program |
| EPI_ISL_422558 | MSHS Clinical Microbiology Laboratories | MSHS Pathogen Surveillance Program |
| EPI_ISL_422559 | MSHS Clinical Microbiology Laboratories | MSHS Pathogen Surveillance Program |
| EPI_ISL_422560 | MSHS Clinical Microbiology Laboratories | MSHS Pathogen Surveillance Program |
| EPI_ISL_422561 | MSHS Clinical Microbiology Laboratories | MSHS Pathogen Surveillance Program |
| EPI_ISL_422562 | MSHS Clinical Microbiology Laboratories | MSHS Pathogen Surveillance Program |
| EPI_ISL_422563 | Institute of Microbiology Universidad San Francisco de Quito | Institute of Microbiology Universidad San Francisco de Quito |
| EPI_ISL_422566 | Dutch COVID-19 response team | Erasmus Medical Center |
| EPI_ISL_422567 | Dutch COVID-19 response team | Erasmus Medical Center |
| EPI_ISL_422568 | Dutch COVID-19 response team | Erasmus Medical Center |
| EPI_ISL_422569 | Dutch COVID-19 response team | Erasmus Medical Center |
| EPI_ISL_422570 | Dutch COVID-19 response team | Erasmus Medical Center |
| EPI_ISL_422571 | Dutch COVID-19 response team | Erasmus Medical Center |

| EPI_Accession_ID | Originating Laboratory | Submitting Laboratory |
|---|---|---|
| EPI_ISL_422572 | Dutch COVID-19 response team | Erasmus Medical Center |
| EPI_ISL_422573 | Dutch COVID-19 response team | Erasmus Medical Center |
| EPI_ISL_422574 | Dutch COVID-19 response team | Erasmus Medical Center |
| EPI_ISL_422575 | Dutch COVID-19 response team | Erasmus Medical Center |
| EPI_ISL_422576 | Dutch COVID-19 response team | Erasmus Medical Center |
| EPI_ISL_422577 | Dutch COVID-19 response team | Erasmus Medical Center |
| EPI_ISL_422578 | Dutch COVID-19 response team | Erasmus Medical Center |
| EPI_ISL_422579 | Dutch COVID-19 response team | Erasmus Medical Center |
| EPI_ISL_422580 | Dutch COVID-19 response team | Erasmus Medical Center |
| EPI_ISL_422581 | Dutch COVID-19 response team | Erasmus Medical Center |
| EPI_ISL_422582 | Dutch COVID-19 response team | Erasmus Medical Center |
| EPI_ISL_422583 | Dutch COVID-19 response team | Erasmus Medical Center |
| EPI_ISL_422584 | Dutch COVID-19 response team | Erasmus Medical Center |
| EPI_ISL_422585 | Dutch COVID-19 response team | Erasmus Medical Center |
| EPI_ISL_422586 | Dutch COVID-19 response team | Erasmus Medical Center |
| EPI_ISL_422587 | Dutch COVID-19 response team | Erasmus Medical Center |
| EPI_ISL_422588 | Dutch COVID-19 response team | Erasmus Medical Center |
| EPI_ISL_422589 | Dutch COVID-19 response team | Erasmus Medical Center |
| EPI_ISL_422590 | Dutch COVID-19 response team | Erasmus Medical Center |
| EPI_ISL_422591 | Dutch COVID-19 response team | Erasmus Medical Center |
| EPI_ISL_422592 | Dutch COVID-19 response team | Erasmus Medical Center |
| EPI_ISL_422593 | Dutch COVID-19 response team | Erasmus Medical Center |
| EPI_ISL_422594 | Dutch COVID-19 response team | Erasmus Medical Center |
| EPI_ISL_422595 | Dutch COVID-19 response team | Erasmus Medical Center |
| EPI_ISL_422596 | Dutch COVID-19 response team | Erasmus Medical Center |
| EPI_ISL_422597 | Dutch COVID-19 response team | Erasmus Medical Center |
| EPI_ISL_422598 | Dutch COVID-19 response team | Erasmus Medical Center |
| EPI_ISL_422599 | Dutch COVID-19 response team | Erasmus Medical Center |
| EPI_ISL_422600 | Dutch COVID-19 response team | Erasmus Medical Center |
| EPI_ISL_422601 | Dutch COVID-19 response team | Erasmus Medical Center |
| EPI_ISL_422602 | Dutch COVID-19 response team | Erasmus Medical Center |
| EPI_ISL_422603 | Dutch COVID-19 response team | Erasmus Medical Center |
| EPI_ISL_422604 | Dutch COVID-19 response team | Erasmus Medical Center |

| EPI_Accession_ID | Originating Laboratory | Submitting Laboratory |
|---|---|---|
| EPI_ISL_422605 | Dutch COVID-19 response team | Erasmus Medical Center |
| EPI_ISL_422606 | Dutch COVID-19 response team | Erasmus Medical Center |
| EPI_ISL_422607 | Dutch COVID-19 response team | Erasmus Medical Center |
| EPI_ISL_422608 | Dutch COVID-19 response team | Erasmus Medical Center |
| EPI_ISL_422609 | Dutch COVID-19 response team | Erasmus Medical Center |
| EPI_ISL_422610 | Dutch COVID-19 response team | Erasmus Medical Center |
| EPI_ISL_422611 | Dutch COVID-19 response team | Erasmus Medical Center |
| EPI_ISL_422612 | Dutch COVID-19 response team | Erasmus Medical Center |
| EPI_ISL_422613 | Dutch COVID-19 response team | Erasmus Medical Center |
| EPI_ISL_422614 | Dutch COVID-19 response team | Erasmus Medical Center |
| EPI_ISL_422615 | Dutch COVID-19 response team | Erasmus Medical Center |
| EPI_ISL_422616 | Dutch COVID-19 response team | Erasmus Medical Center |
| EPI_ISL_422617 | Dutch COVID-19 response team | Erasmus Medical Center |
| EPI_ISL_422618 | Dutch COVID-19 response team | Erasmus Medical Center |
| EPI_ISL_422619 | Dutch COVID-19 response team | Erasmus Medical Center |
| EPI_ISL_422620 | Dutch COVID-19 response team | Erasmus Medical Center |
| EPI_ISL_422621 | Dutch COVID-19 response team | Erasmus Medical Center |
| EPI_ISL_422622 | Dutch COVID-19 response team | Erasmus Medical Center |
| EPI_ISL_422623 | Dutch COVID-19 response team | Erasmus Medical Center |
| EPI_ISL_422624 | Dutch COVID-19 response team | Erasmus Medical Center |
| EPI_ISL_422625 | Dutch COVID-19 response team | Erasmus Medical Center |
| EPI_ISL_422626 | Dutch COVID-19 response team | Erasmus Medical Center |
| EPI_ISL_422627 | Dutch COVID-19 response team | Erasmus Medical Center |
| EPI_ISL_422628 | Dutch COVID-19 response team | Erasmus Medical Center |
| EPI_ISL_422629 | Dutch COVID-19 response team | Erasmus Medical Center |
| EPI_ISL_422630 | Dutch COVID-19 response team | Erasmus Medical Center |
| EPI_ISL_422631 | Dutch COVID-19 response team | Erasmus Medical Center |
| EPI_ISL_422632 | Dutch COVID-19 response team | Erasmus Medical Center |
| EPI_ISL_422633 | Dutch COVID-19 response team | Erasmus Medical Center |
| EPI_ISL_422634 | Dutch COVID-19 response team | Erasmus Medical Center |
| EPI_ISL_422635 | Dutch COVID-19 response team | Erasmus Medical Center |
| EPI_ISL_422636 | The National Institute of Public Health Center for Epidemiology and Microbiology | State Veterinary Institute Prague |

| EPI_Accession_ID | Originating Laboratory | Submitting Laboratory |
|---|---|---|
| EPI_ISL_422637 | Dutch COVID-19 response team | Erasmus Medical Center |
| EPI_ISL_422638 | Dutch COVID-19 response team | Erasmus Medical Center |
| EPI_ISL_422639 | Dutch COVID-19 response team | Erasmus Medical Center |
| EPI_ISL_422640 | Dutch COVID-19 response team | Erasmus Medical Center |
| EPI_ISL_422641 | Dutch COVID-19 response team | Erasmus Medical Center |
| EPI_ISL_422642 | Dutch COVID-19 response team | Erasmus Medical Center |
| EPI_ISL_422643 | Dutch COVID-19 response team | Erasmus Medical Center |
| EPI_ISL_422644 | Dutch COVID-19 response team | Erasmus Medical Center |
| EPI_ISL_422645 | Dutch COVID-19 response team | Erasmus Medical Center |
| EPI_ISL_422646 | Dutch COVID-19 response team | Erasmus Medical Center |
| EPI_ISL_422647 | Dutch COVID-19 response team | Erasmus Medical Center |
| EPI_ISL_422648 | Dutch COVID-19 response team | Erasmus Medical Center |
| EPI_ISL_422649 | Dutch COVID-19 response team | Erasmus Medical Center |
| EPI_ISL_422650 | Dutch COVID-19 response team | Erasmus Medical Center |
| EPI_ISL_422651 | Dutch COVID-19 response team | Erasmus Medical Center |
| EPI_ISL_422652 | Dutch COVID-19 response team | Erasmus Medical Center |
| EPI_ISL_422653 | Dutch COVID-19 response team | Erasmus Medical Center |
| EPI_ISL_422654 | Dutch COVID-19 response team | Erasmus Medical Center |
| EPI_ISL_422655 | Dutch COVID-19 response team | Erasmus Medical Center |
| EPI_ISL_422656 | Dutch COVID-19 response team | Erasmus Medical Center |
| EPI_ISL_422657 | Dutch COVID-19 response team | Erasmus Medical Center |
| EPI_ISL_422658 | Dutch COVID-19 response team | Erasmus Medical Center |
| EPI_ISL_422659 | Dutch COVID-19 response team | Erasmus Medical Center |
| EPI_ISL_422660 | Dutch COVID-19 response team | Erasmus Medical Center |
| EPI_ISL_422661 | Dutch COVID-19 response team | Erasmus Medical Center |
| EPI_ISL_422662 | Dutch COVID-19 response team | Erasmus Medical Center |
| EPI_ISL_422663 | Dutch COVID-19 response team | Erasmus Medical Center |
| EPI_ISL_422664 | Dutch COVID-19 response team | Erasmus Medical Center |
| EPI_ISL_422665 | Dutch COVID-19 response team | Erasmus Medical Center |
| EPI_ISL_422666 | Dutch COVID-19 response team | Erasmus Medical Center |
| EPI_ISL_422667 | Dutch COVID-19 response team | Erasmus Medical Center |
| EPI_ISL_422668 | Dutch COVID-19 response team | Erasmus Medical Center |
| EPI_ISL_422669 | Dutch COVID-19 response team | Erasmus Medical Center |

| EPI_Accession_ID | Originating Laboratory | Submitting Laboratory |
|---|---|---|
| EPI_ISL_422670 | Dutch COVID-19 response team | Erasmus Medical Center |
| EPI_ISL_422671 | Dutch COVID-19 response team | Erasmus Medical Center |
| EPI_ISL_422672 | Dutch COVID-19 response team | Erasmus Medical Center |
| EPI_ISL_422673 | Dutch COVID-19 response team | Erasmus Medical Center |
| EPI_ISL_422674 | Dutch COVID-19 response team | Erasmus Medical Center |
| EPI_ISL_422675 | Dutch COVID-19 response team | Erasmus Medical Center |
| EPI_ISL_422676 | Dutch COVID-19 response team | Erasmus Medical Center |
| EPI_ISL_422677 | Dutch COVID-19 response team | Erasmus Medical Center |
| EPI_ISL_422678 | Dutch COVID-19 response team | Erasmus Medical Center |
| EPI_ISL_422679 | Dutch COVID-19 response team | Erasmus Medical Center |
| EPI_ISL_422680 | Dutch COVID-19 response team | Erasmus Medical Center |
| EPI_ISL_422681 | Dutch COVID-19 response team | Erasmus Medical Center |
| EPI_ISL_422682 | Dutch COVID-19 response team | Erasmus Medical Center |
| EPI_ISL_422683 | Dutch COVID-19 response team | Erasmus Medical Center |
| EPI_ISL_422684 | Dutch COVID-19 response team | Erasmus Medical Center |
| EPI_ISL_422685 | Dutch COVID-19 response team | Erasmus Medical Center |
| EPI_ISL_422686 | Dutch COVID-19 response team | Erasmus Medical Center |
| EPI_ISL_422687 | Dutch COVID-19 response team | Erasmus Medical Center |
| EPI_ISL_422688 | Dutch COVID-19 response team | Erasmus Medical Center |
| EPI_ISL_422689 | Dutch COVID-19 response team | Erasmus Medical Center |
| EPI_ISL_422690 | Dutch COVID-19 response team | Erasmus Medical Center |
| EPI_ISL_422691 | Dutch COVID-19 response team | Erasmus Medical Center |
| EPI_ISL_422692 | Dutch COVID-19 response team | Erasmus Medical Center |
| EPI_ISL_422693 | Dutch COVID-19 response team | Erasmus Medical Center |
| EPI_ISL_422694 | Dutch COVID-19 response team | Erasmus Medical Center |
| EPI_ISL_422695 | Dutch COVID-19 response team | Erasmus Medical Center |
| EPI_ISL_422696 | Dutch COVID-19 response team | Erasmus Medical Center |
| EPI_ISL_422697 | Dutch COVID-19 response team | Erasmus Medical Center |
| EPI_ISL_422698 | Dutch COVID-19 response team | Erasmus Medical Center |
| EPI_ISL_422699 | Dutch COVID-19 response team | Erasmus Medical Center |
| EPI_ISL_422700 | Dutch COVID-19 response team | Erasmus Medical Center |
| EPI_ISL_422701 | Dutch COVID-19 response team | Erasmus Medical Center |
| EPI_ISL_422702 | Dutch COVID-19 response team | Erasmus Medical Center |

| EPI_Accession_ID | Originating Laboratory | Submitting Laboratory |
|---|---|---|
| EPI_ISL_422703 | Dutch COVID-19 response team | Erasmus Medical Center |
| EPI_ISL_422704 | Dutch COVID-19 response team | Erasmus Medical Center |
| EPI_ISL_422705 | Dutch COVID-19 response team | Erasmus Medical Center |
| EPI_ISL_422706 | Dutch COVID-19 response team | Erasmus Medical Center |
| EPI_ISL_422707 | Dutch COVID-19 response team | Erasmus Medical Center |
| EPI_ISL_422708 | Dutch COVID-19 response team | Erasmus Medical Center |
| EPI_ISL_422709 | Dutch COVID-19 response team | Erasmus Medical Center |
| EPI_ISL_422710 | Dutch COVID-19 response team | Erasmus Medical Center |
| EPI_ISL_422711 | Dutch COVID-19 response team | Erasmus Medical Center |
| EPI_ISL_422712 | Dutch COVID-19 response team | Erasmus Medical Center |
| EPI_ISL_422713 | Dutch COVID-19 response team | Erasmus Medical Center |
| EPI_ISL_422714 | Dutch COVID-19 response team | Erasmus Medical Center |
| EPI_ISL_422715 | Dutch COVID-19 response team | Erasmus Medical Center |
| EPI_ISL_422716 | Dutch COVID-19 response team | Erasmus Medical Center |
| EPI_ISL_422717 | Dutch COVID-19 response team | Erasmus Medical Center |
| EPI_ISL_422718 | Dutch COVID-19 response team | Erasmus Medical Center |
| EPI_ISL_422719 | Dutch COVID-19 response team | Erasmus Medical Center |
| EPI_ISL_422720 | Dutch COVID-19 response team | Erasmus Medical Center |
| EPI_ISL_422721 | Dutch COVID-19 response team | Erasmus Medical Center |
| EPI_ISL_422722 | Dutch COVID-19 response team | Erasmus Medical Center |
| EPI_ISL_422723 | Dutch COVID-19 response team | Erasmus Medical Center |
| EPI_ISL_422724 | Dutch COVID-19 response team | Erasmus Medical Center |
| EPI_ISL_422725 | Dutch COVID-19 response team | Erasmus Medical Center |
| EPI_ISL_422726 | Dutch COVID-19 response team | Erasmus Medical Center |
| EPI_ISL_422727 | Dutch COVID-19 response team | Erasmus Medical Center |
| EPI_ISL_422728 | Dutch COVID-19 response team | Erasmus Medical Center |
| EPI_ISL_422729 | Dutch COVID-19 response team | Erasmus Medical Center |
| EPI_ISL_422730 | Dutch COVID-19 response team | Erasmus Medical Center |
| EPI_ISL_422731 | Dutch COVID-19 response team | Erasmus Medical Center |
| EPI_ISL_422732 | Dutch COVID-19 response team | Erasmus Medical Center |
| EPI_ISL_422733 | Dutch COVID-19 response team | Erasmus Medical Center |
| EPI_ISL_422734 | Dutch COVID-19 response team | Erasmus Medical Center |
| EPI_ISL_422735 | Dutch COVID-19 response team | Erasmus Medical Center |

| EPI_Accession_ID | Originating Laboratory | Submitting Laboratory |
|---|---|---|
| EPI_ISL_422736 | Dutch COVID-19 response team | Erasmus Medical Center |
| EPI_ISL_422737 | Dutch COVID-19 response team | Erasmus Medical Center |
| EPI_ISL_422738 | Dutch COVID-19 response team | Erasmus Medical Center |
| EPI_ISL_422739 | Dutch COVID-19 response team | Erasmus Medical Center |
| EPI_ISL_422740 | Dutch COVID-19 response team | Erasmus Medical Center |
| EPI_ISL_422741 | Dutch COVID-19 response team | Erasmus Medical Center |
| EPI_ISL_422742 | Dutch COVID-19 response team | Erasmus Medical Center |
| EPI_ISL_422743 | Dutch COVID-19 response team | Erasmus Medical Center |
| EPI_ISL_422744 | Dutch COVID-19 response team | Erasmus Medical Center |
| EPI_ISL_422745 | Dutch COVID-19 response team | Erasmus Medical Center |
| EPI_ISL_422746 | Dutch COVID-19 response team | Erasmus Medical Center |
| EPI_ISL_422747 | Dutch COVID-19 response team | Erasmus Medical Center |
| EPI_ISL_422748 | Dutch COVID-19 response team | Erasmus Medical Center |
| EPI_ISL_422749 | Dutch COVID-19 response team | Erasmus Medical Center |
| EPI_ISL_422750 | Dutch COVID-19 response team | Erasmus Medical Center |
| EPI_ISL_422751 | Dutch COVID-19 response team | Erasmus Medical Center |
| EPI_ISL_422752 | Dutch COVID-19 response team | Erasmus Medical Center |
| EPI_ISL_422753 | Dutch COVID-19 response team | Erasmus Medical Center |
| EPI_ISL_422754 | Dutch COVID-19 response team | Erasmus Medical Center |
| EPI_ISL_422755 | Dutch COVID-19 response team | Erasmus Medical Center |
| EPI_ISL_422756 | Dutch COVID-19 response team | Erasmus Medical Center |
| EPI_ISL_422757 | Dutch COVID-19 response team | Erasmus Medical Center |
| EPI_ISL_422758 | Dutch COVID-19 response team | Erasmus Medical Center |
| EPI_ISL_422759 | Dutch COVID-19 response team | Erasmus Medical Center |
| EPI_ISL_422760 | Dutch COVID-19 response team | Erasmus Medical Center |
| EPI_ISL_422761 | Dutch COVID-19 response team | Erasmus Medical Center |
| EPI_ISL_422762 | Dutch COVID-19 response team | Erasmus Medical Center |
| EPI_ISL_422763 | Dutch COVID-19 response team | Erasmus Medical Center |
| EPI_ISL_422764 | Dutch COVID-19 response team | Erasmus Medical Center |
| EPI_ISL_422765 | Dutch COVID-19 response team | Erasmus Medical Center |
| EPI_ISL_422766 | Dutch COVID-19 response team | Erasmus Medical Center |
| EPI_ISL_422767 | Dutch COVID-19 response team | Erasmus Medical Center |
| EPI_ISL_422768 | Dutch COVID-19 response team | Erasmus Medical Center |

| EPI_Accession_ID | Originating Laboratory | Submitting Laboratory |
|---|---|---|
| EPI_ISL_422769 | Dutch COVID-19 response team | Erasmus Medical Center |
| EPI_ISL_422770 | Dutch COVID-19 response team | Erasmus Medical Center |
| EPI_ISL_422771 | Dutch COVID-19 response team | Erasmus Medical Center |
| EPI_ISL_422772 | Dutch COVID-19 response team | Erasmus Medical Center |
| EPI_ISL_422773 | Dutch COVID-19 response team | Erasmus Medical Center |
| EPI_ISL_422774 | Dutch COVID-19 response team | Erasmus Medical Center |
| EPI_ISL_422775 | Dutch COVID-19 response team | Erasmus Medical Center |
| EPI_ISL_422776 | Dutch COVID-19 response team | Erasmus Medical Center |
| EPI_ISL_422777 | Dutch COVID-19 response team | Erasmus Medical Center |
| EPI_ISL_422778 | Dutch COVID-19 response team | Erasmus Medical Center |
| EPI_ISL_422779 | Dutch COVID-19 response team | Erasmus Medical Center |
| EPI_ISL_422780 | Dutch COVID-19 response team | Erasmus Medical Center |
| EPI_ISL_422781 | Dutch COVID-19 response team | Erasmus Medical Center |
| EPI_ISL_422782 | Dutch COVID-19 response team | Erasmus Medical Center |
| EPI_ISL_422783 | Dutch COVID-19 response team | Erasmus Medical Center |
| EPI_ISL_422784 | Dutch COVID-19 response team | Erasmus Medical Center |
| EPI_ISL_422785 | Dutch COVID-19 response team | Erasmus Medical Center |
| EPI_ISL_422786 | Dutch COVID-19 response team | Erasmus Medical Center |
| EPI_ISL_422787 | Dutch COVID-19 response team | Erasmus Medical Center |
| EPI_ISL_422788 | Dutch COVID-19 response team | Erasmus Medical Center |
| EPI_ISL_422789 | Dutch COVID-19 response team | Erasmus Medical Center |
| EPI_ISL_422790 | Dutch COVID-19 response team | Erasmus Medical Center |
| EPI_ISL_422791 | Dutch COVID-19 response team | Erasmus Medical Center |
| EPI_ISL_422792 | Dutch COVID-19 response team | Erasmus Medical Center |
| EPI_ISL_422793 | Dutch COVID-19 response team | Erasmus Medical Center |
| EPI_ISL_422794 | Dutch COVID-19 response team | Erasmus Medical Center |
| EPI_ISL_422795 | Dutch COVID-19 response team | Erasmus Medical Center |
| EPI_ISL_422796 | Dutch COVID-19 response team | Erasmus Medical Center |
| EPI_ISL_422797 | Dutch COVID-19 response team | Erasmus Medical Center |
| EPI_ISL_422798 | Dutch COVID-19 response team | Erasmus Medical Center |
| EPI_ISL_422799 | Dutch COVID-19 response team | Erasmus Medical Center |
| EPI_ISL_422800 | Dutch COVID-19 response team | Erasmus Medical Center |
| EPI_ISL_422801 | Dutch COVID-19 response team | Erasmus Medical Center |

| EPI_Accession_ID | Originating Laboratory | Submitting Laboratory |
|---|---|---|
| EPI_ISL_422802 | Dutch COVID-19 response team | Erasmus Medical Center |
| EPI_ISL_422803 | Dutch COVID-19 response team | Erasmus Medical Center |
| EPI_ISL_422804 | Dutch COVID-19 response team | Erasmus Medical Center |
| EPI_ISL_422805 | Dutch COVID-19 response team | Erasmus Medical Center |
| EPI_ISL_422806 | Dutch COVID-19 response team | Erasmus Medical Center |
| EPI_ISL_422807 | Dutch COVID-19 response team | Erasmus Medical Center |
| EPI_ISL_422808 | Dutch COVID-19 response team | Erasmus Medical Center |
| EPI_ISL_422809 | Dutch COVID-19 response team | Erasmus Medical Center |
| EPI_ISL_422810 | Dutch COVID-19 response team | Erasmus Medical Center |
| EPI_ISL_422811 | Dutch COVID-19 response team | Erasmus Medical Center |
| EPI_ISL_422812 | Dutch COVID-19 response team | Erasmus Medical Center |
| EPI_ISL_422813 | Dutch COVID-19 response team | Erasmus Medical Center |
| EPI_ISL_422814 | Dutch COVID-19 response team | Erasmus Medical Center |
| EPI_ISL_422815 | Dutch COVID-19 response team | Erasmus Medical Center |
| EPI_ISL_422816 | Dutch COVID-19 response team | Erasmus Medical Center |
| EPI_ISL_422817 | Dutch COVID-19 response team | Erasmus Medical Center |
| EPI_ISL_422818 | Dutch COVID-19 response team | Erasmus Medical Center |
| EPI_ISL_422819 | Dutch COVID-19 response team | Erasmus Medical Center |
| EPI_ISL_422820 | Dutch COVID-19 response team | Erasmus Medical Center |
| EPI_ISL_422821 | Dutch COVID-19 response team | Erasmus Medical Center |
| EPI_ISL_422822 | Dutch COVID-19 response team | Erasmus Medical Center |
| EPI_ISL_422823 | Dutch COVID-19 response team | Erasmus Medical Center |
| EPI_ISL_422824 | Dutch COVID-19 response team | Erasmus Medical Center |
| EPI_ISL_422825 | Dutch COVID-19 response team | Erasmus Medical Center |
| EPI_ISL_422826 | Dutch COVID-19 response team | Erasmus Medical Center |
| EPI_ISL_422827 | Dutch COVID-19 response team | Erasmus Medical Center |
| EPI_ISL_422828 | Dutch COVID-19 response team | Erasmus Medical Center |
| EPI_ISL_422829 | Dutch COVID-19 response team | Erasmus Medical Center |
| EPI_ISL_422830 | Dutch COVID-19 response team | Erasmus Medical Center |
| EPI_ISL_422831 | Dutch COVID-19 response team | Erasmus Medical Center |
| EPI_ISL_422832 | Dutch COVID-19 response team | Erasmus Medical Center |
| EPI_ISL_422833 | Dutch COVID-19 response team | Erasmus Medical Center |
| EPI_ISL_422834 | Dutch COVID-19 response team | Erasmus Medical Center |

| EPI_Accession_ID | Originating Laboratory | Submitting Laboratory |
|---|---|---|
| EPI_ISL_422835 | Dutch COVID-19 response team | Erasmus Medical Center |
| EPI_ISL_422836 | Dutch COVID-19 response team | Erasmus Medical Center |
| EPI_ISL_422837 | Dutch COVID-19 response team | Erasmus Medical Center |
| EPI_ISL_422838 | Dutch COVID-19 response team | Erasmus Medical Center |
| EPI_ISL_422839 | Dutch COVID-19 response team | Erasmus Medical Center |
| EPI_ISL_422840 | Dutch COVID-19 response team | Erasmus Medical Center |
| EPI_ISL_422841 | Dutch COVID-19 response team | Erasmus Medical Center |
| EPI_ISL_422842 | Dutch COVID-19 response team | Erasmus Medical Center |
| EPI_ISL_422843 | Dutch COVID-19 response team | Erasmus Medical Center |
| EPI_ISL_422844 | Dutch COVID-19 response team | Erasmus Medical Center |
| EPI_ISL_422845 | Dutch COVID-19 response team | Erasmus Medical Center |
| EPI_ISL_422846 | Dutch COVID-19 response team | Erasmus Medical Center |
| EPI_ISL_422847 | Dutch COVID-19 response team | Erasmus Medical Center |
| EPI_ISL_422848 | Dutch COVID-19 response team | Erasmus Medical Center |
| EPI_ISL_422849 | Dutch COVID-19 response team | Erasmus Medical Center |
| EPI_ISL_422850 | Dutch COVID-19 response team | Erasmus Medical Center |
| EPI_ISL_422851 | Dutch COVID-19 response team | Erasmus Medical Center |
| EPI_ISL_422852 | Dutch COVID-19 response team | Erasmus Medical Center |
| EPI_ISL_422853 | Dutch COVID-19 response team | Erasmus Medical Center |
| EPI_ISL_422854 | Dutch COVID-19 response team | Erasmus Medical Center |
| EPI_ISL_422855 | Dutch COVID-19 response team | Erasmus Medical Center |
| EPI_ISL_422856 | Dutch COVID-19 response team | Erasmus Medical Center |
| EPI_ISL_422857 | Dutch COVID-19 response team | Erasmus Medical Center |
| EPI_ISL_422858 | Dutch COVID-19 response team | Erasmus Medical Center |
| EPI_ISL_422859 | Dutch COVID-19 response team | Erasmus Medical Center |
| EPI_ISL_422860 | Dutch COVID-19 response team | Erasmus Medical Center |
| EPI_ISL_422861 | Dutch COVID-19 response team | Erasmus Medical Center |
| EPI_ISL_422862 | Dutch COVID-19 response team | Erasmus Medical Center |
| EPI_ISL_422863 | Dutch COVID-19 response team | Erasmus Medical Center |
| EPI_ISL_422864 | Dutch COVID-19 response team | Erasmus Medical Center |
| EPI_ISL_422865 | Dutch COVID-19 response team | Erasmus Medical Center |
| EPI_ISL_422866 | Dutch COVID-19 response team | Erasmus Medical Center |
| EPI_ISL_422867 | Dutch COVID-19 response team | Erasmus Medical Center |

| EPI_Accession_ID | Originating Laboratory | Submitting Laboratory |
|---|---|---|
| EPI_ISL_422868 | Dutch COVID-19 response team | Erasmus Medical Center |
| EPI_ISL_422869 | Dutch COVID-19 response team | Erasmus Medical Center |
| EPI_ISL_422870 | Dutch COVID-19 response team | Erasmus Medical Center |
| EPI_ISL_422871 | Dutch COVID-19 response team | Erasmus Medical Center |
| EPI_ISL_422872 | Dutch COVID-19 response team | Erasmus Medical Center |
| EPI_ISL_422873 | Dutch COVID-19 response team | Erasmus Medical Center |
| EPI_ISL_422874 | Dutch COVID-19 response team | Erasmus Medical Center |
| EPI_ISL_422875 | Dutch COVID-19 response team | Erasmus Medical Center |
| EPI_ISL_422876 | Dutch COVID-19 response team | Erasmus Medical Center |
| EPI_ISL_422877 | Dutch COVID-19 response team | Erasmus Medical Center |
| EPI_ISL_422878 | Dutch COVID-19 response team | Erasmus Medical Center |
| EPI_ISL_422879 | Dutch COVID-19 response team | Erasmus Medical Center |
| EPI_ISL_422880 | Dutch COVID-19 response team | Erasmus Medical Center |
| EPI_ISL_422881 | Dutch COVID-19 response team | Erasmus Medical Center |
| EPI_ISL_422885 | Dutch COVID-19 response team | Erasmus Medical Center |
| EPI_ISL_422886 | Dutch COVID-19 response team | Erasmus Medical Center |
| EPI_ISL_422887 | Dutch COVID-19 response team | Erasmus Medical Center |
| EPI_ISL_422888 | Dutch COVID-19 response team | Erasmus Medical Center |
| EPI_ISL_422889 | Dutch COVID-19 response team | Erasmus Medical Center |
| EPI_ISL_422890 | Dutch COVID-19 response team | Erasmus Medical Center |
| EPI_ISL_422891 | Dutch COVID-19 response team | Erasmus Medical Center |
| EPI_ISL_422892 | Dutch COVID-19 response team | Erasmus Medical Center |
| EPI_ISL_422893 | Dutch COVID-19 response team | Erasmus Medical Center |
| EPI_ISL_422894 | Dutch COVID-19 response team | Erasmus Medical Center |
| EPI_ISL_422895 | Dutch COVID-19 response team | Erasmus Medical Center |
| EPI_ISL_422896 | Dutch COVID-19 response team | Erasmus Medical Center |
| EPI_ISL_422897 | Dutch COVID-19 response team | Erasmus Medical Center |
| EPI_ISL_422898 | Dutch COVID-19 response team | Erasmus Medical Center |
| EPI_ISL_422899 | Dutch COVID-19 response team | Erasmus Medical Center |
| EPI_ISL_422901 | Dutch COVID-19 response team | Erasmus Medical Center |
| EPI_ISL_422902 | Dutch COVID-19 response team | Erasmus Medical Center |
| EPI_ISL_422903 | Dutch COVID-19 response team | Erasmus Medical Center |
| EPI_ISL_422904 | Dutch COVID-19 response team | Erasmus Medical Center |

| EPI_Accession_ID | Originating Laboratory | Submitting Laboratory |
|---|---|---|
| EPI_ISL_422905 | Dutch COVID-19 response team | Erasmus Medical Center |
| EPI_ISL_422906 | Dutch COVID-19 response team | Erasmus Medical Center |
| EPI_ISL_422907 | Dutch COVID-19 response team | Erasmus Medical Center |
| EPI_ISL_422908 | Dutch COVID-19 response team | Erasmus Medical Center |
| EPI_ISL_422909 | Dutch COVID-19 response team | Erasmus Medical Center |
| EPI_ISL_422910 | Dutch COVID-19 response team | Erasmus Medical Center |
| EPI_ISL_422911 | Dutch COVID-19 response team | Erasmus Medical Center |
| EPI_ISL_422912 | Dutch COVID-19 response team | Erasmus Medical Center |
| EPI_ISL_422913 | Dutch COVID-19 response team | Erasmus Medical Center |
| EPI_ISL_422914 | Dutch COVID-19 response team | Erasmus Medical Center |
| EPI_ISL_422915 | Dutch COVID-19 response team | Erasmus Medical Center |
| EPI_ISL_422916 | Dutch COVID-19 response team | Erasmus Medical Center |
| EPI_ISL_422917 | Dutch COVID-19 response team | Erasmus Medical Center |
| EPI_ISL_422918 | Dutch COVID-19 response team | Erasmus Medical Center |
| EPI_ISL_422919 | Dutch COVID-19 response team | Erasmus Medical Center |
| EPI_ISL_422920 | Dutch COVID-19 response team | Erasmus Medical Center |
| EPI_ISL_422921 | Dutch COVID-19 response team | Erasmus Medical Center |
| EPI_ISL_422922 | Dutch COVID-19 response team | Erasmus Medical Center |
| EPI_ISL_422923 | Dutch COVID-19 response team | Erasmus Medical Center |
| EPI_ISL_422924 | Dutch COVID-19 response team | Erasmus Medical Center |
| EPI_ISL_422925 | Dutch COVID-19 response team | Erasmus Medical Center |
| EPI_ISL_422926 | Dutch COVID-19 response team | Erasmus Medical Center |
| EPI_ISL_422927 | Dutch COVID-19 response team | Erasmus Medical Center |
| EPI_ISL_422928 | Dutch COVID-19 response team | Erasmus Medical Center |
| EPI_ISL_422929 | Dutch COVID-19 response team | Erasmus Medical Center |
| EPI_ISL_422930 | Dutch COVID-19 response team | Erasmus Medical Center |
| EPI_ISL_422931 | Dutch COVID-19 response team | Erasmus Medical Center |
| EPI_ISL_422932 | Dutch COVID-19 response team | Erasmus Medical Center |
| EPI_ISL_422933 | Dutch COVID-19 response team | Erasmus Medical Center |
| EPI_ISL_422934 | Dutch COVID-19 response team | Erasmus Medical Center |
| EPI_ISL_422935 | Dutch COVID-19 response team | Erasmus Medical Center |
| EPI_ISL_422936 | Dutch COVID-19 response team | Erasmus Medical Center |
| EPI_ISL_422937 | Dutch COVID-19 response team | Erasmus Medical Center |

| EPI_Accession_ID | Originating Laboratory | Submitting Laboratory |
|---|---|---|
| EPI_ISL_422938 | Dutch COVID-19 response team | Erasmus Medical Center |
| EPI_ISL_422939 | Dutch COVID-19 response team | Erasmus Medical Center |
| EPI_ISL_422940 | Dutch COVID-19 response team | Erasmus Medical Center |
| EPI_ISL_422941 | Dutch COVID-19 response team | Erasmus Medical Center |
| EPI_ISL_422942 | Dutch COVID-19 response team | Erasmus Medical Center |
| EPI_ISL_422943 | Dutch COVID-19 response team | Erasmus Medical Center |
| EPI_ISL_422944 | Dutch COVID-19 response team | Erasmus Medical Center |
| EPI_ISL_422945 | Dutch COVID-19 response team | Erasmus Medical Center |
| EPI_ISL_422946 | Dutch COVID-19 response team | Erasmus Medical Center |
| EPI_ISL_422947 | Dutch COVID-19 response team | Erasmus Medical Center |
| EPI_ISL_422948 | Dutch COVID-19 response team | Erasmus Medical Center |
| EPI_ISL_422949 | Dutch COVID-19 response team | Erasmus Medical Center |
| EPI_ISL_422950 | Dutch COVID-19 response team | Erasmus Medical Center |
| EPI_ISL_422951 | Dutch COVID-19 response team | Erasmus Medical Center |
| EPI_ISL_422952 | Dutch COVID-19 response team | Erasmus Medical Center |
| EPI_ISL_422953 | Dutch COVID-19 response team | Erasmus Medical Center |
| EPI_ISL_422954 | Dutch COVID-19 response team | Erasmus Medical Center |
| EPI_ISL_422955 | Dutch COVID-19 response team | Erasmus Medical Center |
| EPI_ISL_422956 | Dutch COVID-19 response team | Erasmus Medical Center |
| EPI_ISL_422957 | Dutch COVID-19 response team | Erasmus Medical Center |
| EPI_ISL_422958 | Dutch COVID-19 response team | Erasmus Medical Center |
| EPI_ISL_422959 | Dutch COVID-19 response team | Erasmus Medical Center |
| EPI_ISL_422960 | Dutch COVID-19 response team | Erasmus Medical Center |
| EPI_ISL_422961 | UW Virology Lab | UW Virology Lab |
| EPI_ISL_422962 | UW Virology Lab | UW Virology Lab |
| EPI_ISL_422963 | UW Virology Lab | UW Virology Lab |
| EPI_ISL_422964 | UW Virology Lab | UW Virology Lab |
| EPI_ISL_422965 | UW Virology Lab | UW Virology Lab |
| EPI_ISL_422966 | UW Virology Lab | UW Virology Lab |
| EPI_ISL_422967 | UW Virology Lab | UW Virology Lab |
| EPI_ISL_422969 | UW Virology Lab | UW Virology Lab |
| EPI_ISL_422970 | UW Virology Lab | UW Virology Lab |
| EPI_ISL_422971 | UW Virology Lab | UW Virology Lab |

| EPI_Accession_ID | Originating Laboratory | Submitting Laboratory |
|---|---|---|
| EPI_ISL_422972 | UW Virology Lab | UW Virology Lab |
| EPI_ISL_422973 | UW Virology Lab | UW Virology Lab |
| EPI_ISL_422974 | UW Virology Lab | UW Virology Lab |
| EPI_ISL_422975 | UW Virology Lab | UW Virology Lab |
| EPI_ISL_422976 | UW Virology Lab | UW Virology Lab |
| EPI_ISL_422977 | UW Virology Lab | UW Virology Lab |
| EPI_ISL_422978 | UW Virology Lab | UW Virology Lab |
| EPI_ISL_422979 | UW Virology Lab | UW Virology Lab |
| EPI_ISL_422980 | UW Virology Lab | UW Virology Lab |
| EPI_ISL_422981 | UW Virology Lab | UW Virology Lab |
| EPI_ISL_422982 | UW Virology Lab | UW Virology Lab |
| EPI_ISL_422983 | UW Virology Lab | UW Virology Lab |
| EPI_ISL_422984 | UW Virology Lab | UW Virology Lab |
| EPI_ISL_422985 | UW Virology Lab | UW Virology Lab |
| EPI_ISL_422986 | UW Virology Lab | UW Virology Lab |
| EPI_ISL_422987 | UW Virology Lab | UW Virology Lab |
| EPI_ISL_422988 | UW Virology Lab | UW Virology Lab |
| EPI_ISL_422989 | UW Virology Lab | UW Virology Lab |
| EPI_ISL_422990 | UW Virology Lab | UW Virology Lab |
| EPI_ISL_422991 | UW Virology Lab | UW Virology Lab |
| EPI_ISL_422992 | UW Virology Lab | UW Virology Lab |
| EPI_ISL_422993 | UW Virology Lab | UW Virology Lab |
| EPI_ISL_422994 | UW Virology Lab | UW Virology Lab |
| EPI_ISL_422995 | UW Virology Lab | UW Virology Lab |
| EPI_ISL_422996 | UW Virology Lab | UW Virology Lab |
| EPI_ISL_422997 | UW Virology Lab | UW Virology Lab |
| EPI_ISL_422998 | UW Virology Lab | UW Virology Lab |
| EPI_ISL_422999 | UW Virology Lab | UW Virology Lab |
| EPI_ISL_423000 | UW Virology Lab | UW Virology Lab |
| EPI_ISL_423001 | UW Virology Lab | UW Virology Lab |
| EPI_ISL_423002 | UW Virology Lab | UW Virology Lab |
| EPI_ISL_423003 | UW Virology Lab | UW Virology Lab |
| EPI_ISL_423004 | UW Virology Lab | UW Virology Lab |

| EPI_Accession_ID | Originating Laboratory | Submitting Laboratory |
|---|---|---|
| EPI_ISL_423005 | UW Virology Lab | UW Virology Lab |
| EPI_ISL_423006 | UW Virology Lab | UW Virology Lab |
| EPI_ISL_423007 | UW Virology Lab | UW Virology Lab |
| EPI_ISL_423008 | UW Virology Lab | UW Virology Lab |
| EPI_ISL_423009 | UW Virology Lab | UW Virology Lab |
| EPI_ISL_423010 | UW Virology Lab | UW Virology Lab |
| EPI_ISL_423011 | UW Virology Lab | UW Virology Lab |
| EPI_ISL_423013 | UW Virology Lab | UW Virology Lab |
| EPI_ISL_423014 | UW Virology Lab | UW Virology Lab |
| EPI_ISL_423015 | UW Virology Lab | UW Virology Lab |
| EPI_ISL_423016 | UW Virology Lab | UW Virology Lab |
| EPI_ISL_423017 | UW Virology Lab | UW Virology Lab |
| EPI_ISL_423018 | UW Virology Lab | UW Virology Lab |
| EPI_ISL_423019 | UW Virology Lab | UW Virology Lab |
| EPI_ISL_423020 | UW Virology Lab | UW Virology Lab |
| EPI_ISL_423021 | UW Virology Lab | UW Virology Lab |
| EPI_ISL_423022 | UW Virology Lab | UW Virology Lab |
| EPI_ISL_423024 | UW Virology Lab | UW Virology Lab |
| EPI_ISL_423025 | UW Virology Lab | UW Virology Lab |
| EPI_ISL_423026 | UW Virology Lab | UW Virology Lab |
| EPI_ISL_423027 | UW Virology Lab | UW Virology Lab |
| EPI_ISL_423028 | UW Virology Lab | UW Virology Lab |
| EPI_ISL_423029 | UW Virology Lab | UW Virology Lab |
| EPI_ISL_423030 | UW Virology Lab | UW Virology Lab |
| EPI_ISL_423031 | UW Virology Lab | UW Virology Lab |
| EPI_ISL_423032 | UW Virology Lab | UW Virology Lab |
| EPI_ISL_423033 | UW Virology Lab | UW Virology Lab |
| EPI_ISL_423034 | Dutch COVID-19 response team | Erasmus Medical Center |
| EPI_ISL_423039 | Ramathibodi Hospital | COVID-19 Network Investigations (CONI) Alliance |
| EPI_ISL_423040 | Ramathibodi Hospital | COVID-19 Network Investigations (CONI) Alliance |
| EPI_ISL_423041 | Ramathibodi Hospital | COVID-19 Network Investigations (CONI) Alliance |
| EPI_ISL_423042 | Ramathibodi Hospital | COVID-19 Network Investigations (CONI) Alliance |
| EPI_ISL_423043 | Ramathibodi Hospital | COVID-19 Network Investigations (CONI) Alliance |

| EPI_Accession_ID | Originating Laboratory | Submitting Laboratory |
|---|---|---|
| EPI_ISL_423044 | Respiratory Virus Unit, Microbiology Services Colindale, Public Health England | Respiratory Virus Unit, Microbiology Services Colindale, Public Health England |
| EPI_ISL_423045 | Respiratory Virus Unit, Microbiology Services Colindale, Public Health England | Respiratory Virus Unit, Microbiology Services Colindale, Public Health England |
| EPI_ISL_423046 | Respiratory Virus Unit, Microbiology Services Colindale, Public Health England | Respiratory Virus Unit, Microbiology Services Colindale, Public Health England |
| EPI_ISL_423047 | Respiratory Virus Unit, Microbiology Services Colindale, Public Health England | Respiratory Virus Unit, Microbiology Services Colindale, Public Health England |
| EPI_ISL_423048 | Respiratory Virus Unit, Microbiology Services Colindale, Public Health England | Respiratory Virus Unit, Microbiology Services Colindale, Public Health England |
| EPI_ISL_423049 | Respiratory Virus Unit, Microbiology Services Colindale, Public Health England | Respiratory Virus Unit, Microbiology Services Colindale, Public Health England |
| EPI_ISL_423050 | Respiratory Virus Unit, Microbiology Services Colindale, Public Health England | Respiratory Virus Unit, Microbiology Services Colindale, Public Health England |
| EPI_ISL_423051 | Respiratory Virus Unit, Microbiology Services Colindale, Public Health England | Respiratory Virus Unit, Microbiology Services Colindale, Public Health England |
| EPI_ISL_423052 | Respiratory Virus Unit, Microbiology Services Colindale, Public Health England | Respiratory Virus Unit, Microbiology Services Colindale, Public Health England |
| EPI_ISL_423053 | Respiratory Virus Unit, Microbiology Services Colindale, Public Health England | Respiratory Virus Unit, Microbiology Services Colindale, Public Health England |
| EPI_ISL_423054 | Respiratory Virus Unit, Microbiology Services Colindale, Public Health England | Respiratory Virus Unit, Microbiology Services Colindale, Public Health England |
| EPI_ISL_423055 | Respiratory Virus Unit, Microbiology Services Colindale, Public Health England | Respiratory Virus Unit, Microbiology Services Colindale, Public Health England |
| EPI_ISL_423056 | Respiratory Virus Unit, Microbiology Services Colindale, Public Health England | Respiratory Virus Unit, Microbiology Services Colindale, Public Health England |
| EPI_ISL_423057 | Respiratory Virus Unit, Microbiology Services Colindale, Public Health England | Respiratory Virus Unit, Microbiology Services Colindale, Public Health England |
| EPI_ISL_423058 | Respiratory Virus Unit, Microbiology Services Colindale, Public Health England | Respiratory Virus Unit, Microbiology Services Colindale, Public Health England |
| EPI_ISL_423059 | Respiratory Virus Unit, Microbiology Services Colindale, Public Health England | Respiratory Virus Unit, Microbiology Services Colindale, Public Health England |

| EPI_Accession_ID | Originating Laboratory | Submitting Laboratory |
|---|---|---|
| EPI_ISL_423060 | Respiratory Virus Unit, Microbiology Services Colindale, Public Health England | Respiratory Virus Unit, Microbiology Services Colindale, Public Health England |
| EPI_ISL_423061 | Respiratory Virus Unit, Microbiology Services Colindale, Public Health England | Respiratory Virus Unit, Microbiology Services Colindale, Public Health England |
| EPI_ISL_423062 | Respiratory Virus Unit, Microbiology Services Colindale, Public Health England | Respiratory Virus Unit, Microbiology Services Colindale, Public Health England |
| EPI_ISL_423063 | Respiratory Virus Unit, Microbiology Services Colindale, Public Health England | Respiratory Virus Unit, Microbiology Services Colindale, Public Health England |
| EPI_ISL_423064 | Respiratory Virus Unit, Microbiology Services Colindale, Public Health England | Respiratory Virus Unit, Microbiology Services Colindale, Public Health England |
| EPI_ISL_423065 | Respiratory Virus Unit, Microbiology Services Colindale, Public Health England | Respiratory Virus Unit, Microbiology Services Colindale, Public Health England |
| EPI_ISL_423066 | Respiratory Virus Unit, Microbiology Services Colindale, Public Health England | Respiratory Virus Unit, Microbiology Services Colindale, Public Health England |
| EPI_ISL_423067 | Respiratory Virus Unit, Microbiology Services Colindale, Public Health England | Respiratory Virus Unit, Microbiology Services Colindale, Public Health England |
| EPI_ISL_423068 | Respiratory Virus Unit, Microbiology Services Colindale, Public Health England | Respiratory Virus Unit, Microbiology Services Colindale, Public Health England |
| EPI_ISL_423069 | Respiratory Virus Unit, Microbiology Services Colindale, Public Health England | Respiratory Virus Unit, Microbiology Services Colindale, Public Health England |
| EPI_ISL_423070 | Respiratory Virus Unit, Microbiology Services Colindale, Public Health England | Respiratory Virus Unit, Microbiology Services Colindale, Public Health England |
| EPI_ISL_423071 | Respiratory Virus Unit, Microbiology Services Colindale, Public Health England | Respiratory Virus Unit, Microbiology Services Colindale, Public Health England |
| EPI_ISL_423072 | Respiratory Virus Unit, Microbiology Services Colindale, Public Health England | Respiratory Virus Unit, Microbiology Services Colindale, Public Health England |
| EPI_ISL_423073 | Respiratory Virus Unit, Microbiology Services Colindale, Public Health England | Respiratory Virus Unit, Microbiology Services Colindale, Public Health England |
| EPI_ISL_423074 | Respiratory Virus Unit, Microbiology Services Colindale, Public Health England | Respiratory Virus Unit, Microbiology Services Colindale, Public Health England |
| EPI_ISL_423075 | Respiratory Virus Unit, Microbiology Services Colindale, Public Health England | Respiratory Virus Unit, Microbiology Services Colindale, Public Health England |

| EPI_Accession_ID | Originating Laboratory | Submitting Laboratory |
|---|---|---|
| EPI_ISL_423076 | Respiratory Virus Unit, Microbiology Services Colindale, Public Health England | Respiratory Virus Unit, Microbiology Services Colindale, Public Health England |
| EPI_ISL_423077 | Respiratory Virus Unit, Microbiology Services Colindale, Public Health England | Respiratory Virus Unit, Microbiology Services Colindale, Public Health England |
| EPI_ISL_423078 | Respiratory Virus Unit, Microbiology Services Colindale, Public Health England | Respiratory Virus Unit, Microbiology Services Colindale, Public Health England |
| EPI_ISL_423079 | Respiratory Virus Unit, Microbiology Services Colindale, Public Health England | Respiratory Virus Unit, Microbiology Services Colindale, Public Health England |
| EPI_ISL_423080 | Respiratory Virus Unit, Microbiology Services Colindale, Public Health England | Respiratory Virus Unit, Microbiology Services Colindale, Public Health England |
| EPI_ISL_423081 | Respiratory Virus Unit, Microbiology Services Colindale, Public Health England | Respiratory Virus Unit, Microbiology Services Colindale, Public Health England |
| EPI_ISL_423082 | Respiratory Virus Unit, Microbiology Services Colindale, Public Health England | Respiratory Virus Unit, Microbiology Services Colindale, Public Health England |
| EPI_ISL_423083 | Respiratory Virus Unit, Microbiology Services Colindale, Public Health England | Respiratory Virus Unit, Microbiology Services Colindale, Public Health England |
| EPI_ISL_423084 | Respiratory Virus Unit, Microbiology Services Colindale, Public Health England | Respiratory Virus Unit, Microbiology Services Colindale, Public Health England |
| EPI_ISL_423085 | Respiratory Virus Unit, Microbiology Services Colindale, Public Health England | Respiratory Virus Unit, Microbiology Services Colindale, Public Health England |
| EPI_ISL_423086 | Respiratory Virus Unit, Microbiology Services Colindale, Public Health England | Respiratory Virus Unit, Microbiology Services Colindale, Public Health England |
| EPI_ISL_423087 | Respiratory Virus Unit, Microbiology Services Colindale, Public Health England | Respiratory Virus Unit, Microbiology Services Colindale, Public Health England |
| EPI_ISL_423088 | Respiratory Virus Unit, Microbiology Services Colindale, Public Health England | Respiratory Virus Unit, Microbiology Services Colindale, Public Health England |
| EPI_ISL_423089 | Respiratory Virus Unit, Microbiology Services Colindale, Public Health England | Respiratory Virus Unit, Microbiology Services Colindale, Public Health England |
| EPI_ISL_423090 | Respiratory Virus Unit, Microbiology Services Colindale, Public Health England | Respiratory Virus Unit, Microbiology Services Colindale, Public Health England |
| EPI_ISL_423091 | Respiratory Virus Unit, Microbiology Services Colindale, Public Health England | Respiratory Virus Unit, Microbiology Services Colindale, Public Health England |

| EPI_Accession_ID | Originating Laboratory | Submitting Laboratory |
|---|---|---|
| EPI_ISL_423092 | Respiratory Virus Unit, Microbiology Services Colindale, Public Health England | Respiratory Virus Unit, Microbiology Services Colindale, Public Health England |
| EPI_ISL_423093 | Respiratory Virus Unit, Microbiology Services Colindale, Public Health England | Respiratory Virus Unit, Microbiology Services Colindale, Public Health England |
| EPI_ISL_423094 | Respiratory Virus Unit, Microbiology Services Colindale, Public Health England | Respiratory Virus Unit, Microbiology Services Colindale, Public Health England |
| EPI_ISL_423095 | Respiratory Virus Unit, Microbiology Services Colindale, Public Health England | Respiratory Virus Unit, Microbiology Services Colindale, Public Health England |
| EPI_ISL_423096 | Respiratory Virus Unit, Microbiology Services Colindale, Public Health England | Respiratory Virus Unit, Microbiology Services Colindale, Public Health England |
| EPI_ISL_423097 | Respiratory Virus Unit, Microbiology Services Colindale, Public Health England | Respiratory Virus Unit, Microbiology Services Colindale, Public Health England |
| EPI_ISL_423098 | Respiratory Virus Unit, Microbiology Services Colindale, Public Health England | Respiratory Virus Unit, Microbiology Services Colindale, Public Health England |
| EPI_ISL_423099 | Respiratory Virus Unit, Microbiology Services Colindale, Public Health England | Respiratory Virus Unit, Microbiology Services Colindale, Public Health England |
| EPI_ISL_423100 | Respiratory Virus Unit, Microbiology Services Colindale, Public Health England | Respiratory Virus Unit, Microbiology Services Colindale, Public Health England |
| EPI_ISL_423101 | Respiratory Virus Unit, Microbiology Services Colindale, Public Health England | Respiratory Virus Unit, Microbiology Services Colindale, Public Health England |
| EPI_ISL_423102 | Respiratory Virus Unit, Microbiology Services Colindale, Public Health England | Respiratory Virus Unit, Microbiology Services Colindale, Public Health England |
| EPI_ISL_423103 | Respiratory Virus Unit, Microbiology Services Colindale, Public Health England | Respiratory Virus Unit, Microbiology Services Colindale, Public Health England |
| EPI_ISL_423104 | Respiratory Virus Unit, Microbiology Services Colindale, Public Health England | Respiratory Virus Unit, Microbiology Services Colindale, Public Health England |
| EPI_ISL_423105 | Respiratory Virus Unit, Microbiology Services Colindale, Public Health England | Respiratory Virus Unit, Microbiology Services Colindale, Public Health England |
| EPI_ISL_423106 | Respiratory Virus Unit, Microbiology Services Colindale, Public Health England | Respiratory Virus Unit, Microbiology Services Colindale, Public Health England |
| EPI_ISL_423107 | Respiratory Virus Unit, Microbiology Services Colindale, Public Health England | Respiratory Virus Unit, Microbiology Services Colindale, Public Health England |

| EPI_Accession_ID | Originating Laboratory | Submitting Laboratory |
|---|---|---|
| EPI_ISL_423108 | Respiratory Virus Unit, Microbiology Services Colindale, Public Health England | Respiratory Virus Unit, Microbiology Services Colindale, Public Health England |
| EPI_ISL_423109 | Respiratory Virus Unit, Microbiology Services Colindale, Public Health England | Respiratory Virus Unit, Microbiology Services Colindale, Public Health England |
| EPI_ISL_423110 | Respiratory Virus Unit, Microbiology Services Colindale, Public Health England | Respiratory Virus Unit, Microbiology Services Colindale, Public Health England |
| EPI_ISL_423111 | Respiratory Virus Unit, Microbiology Services Colindale, Public Health England | Respiratory Virus Unit, Microbiology Services Colindale, Public Health England |
| EPI_ISL_423112 | Respiratory Virus Unit, Microbiology Services Colindale, Public Health England | Respiratory Virus Unit, Microbiology Services Colindale, Public Health England |
| EPI_ISL_423113 | Respiratory Virus Unit, Microbiology Services Colindale, Public Health England | Respiratory Virus Unit, Microbiology Services Colindale, Public Health England |
| EPI_ISL_423114 | Respiratory Virus Unit, Microbiology Services Colindale, Public Health England | Respiratory Virus Unit, Microbiology Services Colindale, Public Health England |
| EPI_ISL_423115 | Respiratory Virus Unit, Microbiology Services Colindale, Public Health England | Respiratory Virus Unit, Microbiology Services Colindale, Public Health England |
| EPI_ISL_423116 | Respiratory Virus Unit, Microbiology Services Colindale, Public Health England | Respiratory Virus Unit, Microbiology Services Colindale, Public Health England |
| EPI_ISL_423117 | Respiratory Virus Unit, Microbiology Services Colindale, Public Health England | Respiratory Virus Unit, Microbiology Services Colindale, Public Health England |
| EPI_ISL_423118 | Respiratory Virus Unit, Microbiology Services Colindale, Public Health England | Respiratory Virus Unit, Microbiology Services Colindale, Public Health England |
| EPI_ISL_423119 | Respiratory Virus Unit, Microbiology Services Colindale, Public Health England | Respiratory Virus Unit, Microbiology Services Colindale, Public Health England |
| EPI_ISL_423120 | Respiratory Virus Unit, Microbiology Services Colindale, Public Health England | Respiratory Virus Unit, Microbiology Services Colindale, Public Health England |
| EPI_ISL_423121 | Respiratory Virus Unit, Microbiology Services Colindale, Public Health England | Respiratory Virus Unit, Microbiology Services Colindale, Public Health England |
| EPI_ISL_423122 | Respiratory Virus Unit, Microbiology Services Colindale, Public Health England | Respiratory Virus Unit, Microbiology Services Colindale, Public Health England |
| EPI_ISL_423123 | Respiratory Virus Unit, Microbiology Services Colindale, Public Health England | Respiratory Virus Unit, Microbiology Services Colindale, Public Health England |

| EPI_Accession_ID | Originating Laboratory | Submitting Laboratory |
|---|---|---|
| EPI_ISL_423124 | Respiratory Virus Unit, Microbiology Services Colindale, Public Health England | Respiratory Virus Unit, Microbiology Services Colindale, Public Health England |
| EPI_ISL_423125 | Respiratory Virus Unit, Microbiology Services Colindale, Public Health England | Respiratory Virus Unit, Microbiology Services Colindale, Public Health England |
| EPI_ISL_423126 | Respiratory Virus Unit, Microbiology Services Colindale, Public Health England | Respiratory Virus Unit, Microbiology Services Colindale, Public Health England |
| EPI_ISL_423127 | Respiratory Virus Unit, Microbiology Services Colindale, Public Health England | Respiratory Virus Unit, Microbiology Services Colindale, Public Health England |
| EPI_ISL_423128 | Respiratory Virus Unit, Microbiology Services Colindale, Public Health England | Respiratory Virus Unit, Microbiology Services Colindale, Public Health England |
| EPI_ISL_423129 | Respiratory Virus Unit, Microbiology Services Colindale, Public Health England | Respiratory Virus Unit, Microbiology Services Colindale, Public Health England |
| EPI_ISL_423130 | Respiratory Virus Unit, Microbiology Services Colindale, Public Health England | Respiratory Virus Unit, Microbiology Services Colindale, Public Health England |
| EPI_ISL_423131 | Respiratory Virus Unit, Microbiology Services Colindale, Public Health England | Respiratory Virus Unit, Microbiology Services Colindale, Public Health England |
| EPI_ISL_423132 | Respiratory Virus Unit, Microbiology Services Colindale, Public Health England | Respiratory Virus Unit, Microbiology Services Colindale, Public Health England |
| EPI_ISL_423133 | Respiratory Virus Unit, Microbiology Services Colindale, Public Health England | Respiratory Virus Unit, Microbiology Services Colindale, Public Health England |
| EPI_ISL_423134 | Respiratory Virus Unit, Microbiology Services Colindale, Public Health England | Respiratory Virus Unit, Microbiology Services Colindale, Public Health England |
| EPI_ISL_423135 | Respiratory Virus Unit, Microbiology Services Colindale, Public Health England | Respiratory Virus Unit, Microbiology Services Colindale, Public Health England |
| EPI_ISL_423136 | Respiratory Virus Unit, Microbiology Services Colindale, Public Health England | Respiratory Virus Unit, Microbiology Services Colindale, Public Health England |
| EPI_ISL_423137 | Respiratory Virus Unit, Microbiology Services Colindale, Public Health England | Respiratory Virus Unit, Microbiology Services Colindale, Public Health England |
| EPI_ISL_423138 | Respiratory Virus Unit, Microbiology Services Colindale, Public Health England | Respiratory Virus Unit, Microbiology Services Colindale, Public Health England |
| EPI_ISL_423139 | Respiratory Virus Unit, Microbiology Services Colindale, Public Health England | Respiratory Virus Unit, Microbiology Services Colindale, Public Health England |

| EPI_Accession_ID | Originating Laboratory | Submitting Laboratory |
|---|---|---|
| EPI_ISL_423140 | Respiratory Virus Unit, Microbiology Services Colindale, Public Health England | Respiratory Virus Unit, Microbiology Services Colindale, Public Health England |
| EPI_ISL_423141 | Respiratory Virus Unit, Microbiology Services Colindale, Public Health England | Respiratory Virus Unit, Microbiology Services Colindale, Public Health England |
| EPI_ISL_423142 | Respiratory Virus Unit, Microbiology Services Colindale, Public Health England | Respiratory Virus Unit, Microbiology Services Colindale, Public Health England |
| EPI_ISL_423143 | Respiratory Virus Unit, Microbiology Services Colindale, Public Health England | Respiratory Virus Unit, Microbiology Services Colindale, Public Health England |
| EPI_ISL_423144 | Respiratory Virus Unit, Microbiology Services Colindale, Public Health England | Respiratory Virus Unit, Microbiology Services Colindale, Public Health England |
| EPI_ISL_423145 | Respiratory Virus Unit, Microbiology Services Colindale, Public Health England | Respiratory Virus Unit, Microbiology Services Colindale, Public Health England |
| EPI_ISL_423147 | Respiratory Virus Unit, Microbiology Services Colindale, Public Health England | Respiratory Virus Unit, Microbiology Services Colindale, Public Health England |
| EPI_ISL_423148 | Respiratory Virus Unit, Microbiology Services Colindale, Public Health England | Respiratory Virus Unit, Microbiology Services Colindale, Public Health England |
| EPI_ISL_423149 | Respiratory Virus Unit, Microbiology Services Colindale, Public Health England | Respiratory Virus Unit, Microbiology Services Colindale, Public Health England |
| EPI_ISL_423150 | Respiratory Virus Unit, Microbiology Services Colindale, Public Health England | Respiratory Virus Unit, Microbiology Services Colindale, Public Health England |
| EPI_ISL_423151 | Respiratory Virus Unit, Microbiology Services Colindale, Public Health England | Respiratory Virus Unit, Microbiology Services Colindale, Public Health England |
| EPI_ISL_423152 | Respiratory Virus Unit, Microbiology Services Colindale, Public Health England | Respiratory Virus Unit, Microbiology Services Colindale, Public Health England |
| EPI_ISL_423153 | Respiratory Virus Unit, Microbiology Services Colindale, Public Health England | Respiratory Virus Unit, Microbiology Services Colindale, Public Health England |
| EPI_ISL_423154 | Respiratory Virus Unit, Microbiology Services Colindale, Public Health England | Respiratory Virus Unit, Microbiology Services Colindale, Public Health England |
| EPI_ISL_423155 | Respiratory Virus Unit, Microbiology Services Colindale, Public Health England | Respiratory Virus Unit, Microbiology Services Colindale, Public Health England |
| EPI_ISL_423156 | Respiratory Virus Unit, Microbiology Services Colindale, Public Health England | Respiratory Virus Unit, Microbiology Services Colindale, Public Health England |

| EPI_Accession_ID | Originating Laboratory | Submitting Laboratory |
|---|---|---|
| EPI_ISL_423157 | Respiratory Virus Unit, Microbiology Services Colindale, Public Health England | Respiratory Virus Unit, Microbiology Services Colindale, Public Health England |
| EPI_ISL_423158 | Respiratory Virus Unit, Microbiology Services Colindale, Public Health England | Respiratory Virus Unit, Microbiology Services Colindale, Public Health England |
| EPI_ISL_423159 | Respiratory Virus Unit, Microbiology Services Colindale, Public Health England | Respiratory Virus Unit, Microbiology Services Colindale, Public Health England |
| EPI_ISL_423160 | Respiratory Virus Unit, Microbiology Services Colindale, Public Health England | Respiratory Virus Unit, Microbiology Services Colindale, Public Health England |
| EPI_ISL_423161 | Respiratory Virus Unit, Microbiology Services Colindale, Public Health England | Respiratory Virus Unit, Microbiology Services Colindale, Public Health England |
| EPI_ISL_423162 | Respiratory Virus Unit, Microbiology Services Colindale, Public Health England | Respiratory Virus Unit, Microbiology Services Colindale, Public Health England |
| EPI_ISL_423163 | Respiratory Virus Unit, Microbiology Services Colindale, Public Health England | Respiratory Virus Unit, Microbiology Services Colindale, Public Health England |
| EPI_ISL_423164 | Respiratory Virus Unit, Microbiology Services Colindale, Public Health England | Respiratory Virus Unit, Microbiology Services Colindale, Public Health England |
| EPI_ISL_423165 | Respiratory Virus Unit, Microbiology Services Colindale, Public Health England | Respiratory Virus Unit, Microbiology Services Colindale, Public Health England |
| EPI_ISL_423166 | Respiratory Virus Unit, Microbiology Services Colindale, Public Health England | Respiratory Virus Unit, Microbiology Services Colindale, Public Health England |
| EPI_ISL_423167 | Respiratory Virus Unit, Microbiology Services Colindale, Public Health England | Respiratory Virus Unit, Microbiology Services Colindale, Public Health England |
| EPI_ISL_423168 | Respiratory Virus Unit, Microbiology Services Colindale, Public Health England | Respiratory Virus Unit, Microbiology Services Colindale, Public Health England |
| EPI_ISL_423169 | Respiratory Virus Unit, Microbiology Services Colindale, Public Health England | Respiratory Virus Unit, Microbiology Services Colindale, Public Health England |
| EPI_ISL_423170 | Respiratory Virus Unit, Microbiology Services Colindale, Public Health England | Respiratory Virus Unit, Microbiology Services Colindale, Public Health England |
| EPI_ISL_423172 | Respiratory Virus Unit, Microbiology Services Colindale, Public Health England | Respiratory Virus Unit, Microbiology Services Colindale, Public Health England |
| EPI_ISL_423173 | Respiratory Virus Unit, Microbiology Services Colindale, Public Health England | Respiratory Virus Unit, Microbiology Services Colindale, Public Health England |

| EPI_Accession_ID | Originating Laboratory | Submitting Laboratory |
|---|---|---|
| EPI_ISL_423174 | Respiratory Virus Unit, Microbiology Services Colindale, Public Health England | Respiratory Virus Unit, Microbiology Services Colindale, Public Health England |
| EPI_ISL_423175 | Respiratory Virus Unit, Microbiology Services Colindale, Public Health England | Respiratory Virus Unit, Microbiology Services Colindale, Public Health England |
| EPI_ISL_423176 | Respiratory Virus Unit, Microbiology Services Colindale, Public Health England | Respiratory Virus Unit, Microbiology Services Colindale, Public Health England |
| EPI_ISL_423177 | Respiratory Virus Unit, Microbiology Services Colindale, Public Health England | Respiratory Virus Unit, Microbiology Services Colindale, Public Health England |
| EPI_ISL_423178 | Respiratory Virus Unit, Microbiology Services Colindale, Public Health England | Respiratory Virus Unit, Microbiology Services Colindale, Public Health England |
| EPI_ISL_423179 | Respiratory Virus Unit, Microbiology Services Colindale, Public Health England | Respiratory Virus Unit, Microbiology Services Colindale, Public Health England |
| EPI_ISL_423180 | Respiratory Virus Unit, Microbiology Services Colindale, Public Health England | Respiratory Virus Unit, Microbiology Services Colindale, Public Health England |
| EPI_ISL_423181 | Respiratory Virus Unit, Microbiology Services Colindale, Public Health England | Respiratory Virus Unit, Microbiology Services Colindale, Public Health England |
| EPI_ISL_423182 | Respiratory Virus Unit, Microbiology Services Colindale, Public Health England | Respiratory Virus Unit, Microbiology Services Colindale, Public Health England |
| EPI_ISL_423183 | Respiratory Virus Unit, Microbiology Services Colindale, Public Health England | Respiratory Virus Unit, Microbiology Services Colindale, Public Health England |
| EPI_ISL_423184 | Respiratory Virus Unit, Microbiology Services Colindale, Public Health England | Respiratory Virus Unit, Microbiology Services Colindale, Public Health England |
| EPI_ISL_423185 | Respiratory Virus Unit, Microbiology Services Colindale, Public Health England | Respiratory Virus Unit, Microbiology Services Colindale, Public Health England |
| EPI_ISL_423186 | Respiratory Virus Unit, Microbiology Services Colindale, Public Health England | Respiratory Virus Unit, Microbiology Services Colindale, Public Health England |
| EPI_ISL_423187 | Respiratory Virus Unit, Microbiology Services Colindale, Public Health England | Respiratory Virus Unit, Microbiology Services Colindale, Public Health England |
| EPI_ISL_423188 | Respiratory Virus Unit, Microbiology Services Colindale, Public Health England | Respiratory Virus Unit, Microbiology Services Colindale, Public Health England |
| EPI_ISL_423189 | Respiratory Virus Unit, Microbiology Services Colindale, Public Health England | Respiratory Virus Unit, Microbiology Services Colindale, Public Health England |

| EPI_Accession_ID | Originating Laboratory | Submitting Laboratory |
|---|---|---|
| EPI_ISL_423190 | Respiratory Virus Unit, Microbiology Services Colindale, Public Health England | Respiratory Virus Unit, Microbiology Services Colindale, Public Health England |
| EPI_ISL_423191 | Respiratory Virus Unit, Microbiology Services Colindale, Public Health England | Respiratory Virus Unit, Microbiology Services Colindale, Public Health England |
| EPI_ISL_423192 | Respiratory Virus Unit, Microbiology Services Colindale, Public Health England | Respiratory Virus Unit, Microbiology Services Colindale, Public Health England |
| EPI_ISL_423193 | Respiratory Virus Unit, Microbiology Services Colindale, Public Health England | Respiratory Virus Unit, Microbiology Services Colindale, Public Health England |
| EPI_ISL_423194 | Respiratory Virus Unit, Microbiology Services Colindale, Public Health England | Respiratory Virus Unit, Microbiology Services Colindale, Public Health England |
| EPI_ISL_423195 | Respiratory Virus Unit, Microbiology Services Colindale, Public Health England | Respiratory Virus Unit, Microbiology Services Colindale, Public Health England |
| EPI_ISL_423196 | Respiratory Virus Unit, Microbiology Services Colindale, Public Health England | Respiratory Virus Unit, Microbiology Services Colindale, Public Health England |
| EPI_ISL_423197 | Respiratory Virus Unit, Microbiology Services Colindale, Public Health England | Respiratory Virus Unit, Microbiology Services Colindale, Public Health England |
| EPI_ISL_423198 | Respiratory Virus Unit, Microbiology Services Colindale, Public Health England | Respiratory Virus Unit, Microbiology Services Colindale, Public Health England |
| EPI_ISL_423199 | Respiratory Virus Unit, Microbiology Services Colindale, Public Health England | Respiratory Virus Unit, Microbiology Services Colindale, Public Health England |
| EPI_ISL_423200 | Respiratory Virus Unit, Microbiology Services Colindale, Public Health England | Respiratory Virus Unit, Microbiology Services Colindale, Public Health England |
| EPI_ISL_423201 | Respiratory Virus Unit, Microbiology Services Colindale, Public Health England | Respiratory Virus Unit, Microbiology Services Colindale, Public Health England |
| EPI_ISL_423202 | Respiratory Virus Unit, Microbiology Services Colindale, Public Health England | Respiratory Virus Unit, Microbiology Services Colindale, Public Health England |
| EPI_ISL_423203 | Respiratory Virus Unit, Microbiology Services Colindale, Public Health England | Respiratory Virus Unit, Microbiology Services Colindale, Public Health England |
| EPI_ISL_423204 | Respiratory Virus Unit, Microbiology Services Colindale, Public Health England | Respiratory Virus Unit, Microbiology Services Colindale, Public Health England |
| EPI_ISL_423205 | Respiratory Virus Unit, Microbiology Services Colindale, Public Health England | Respiratory Virus Unit, Microbiology Services Colindale, Public Health England |

| EPI_Accession_ID | Originating Laboratory | Submitting Laboratory |
|---|---|---|
| EPI_ISL_423206 | Respiratory Virus Unit, Microbiology Services Colindale, Public Health England | Respiratory Virus Unit, Microbiology Services Colindale, Public Health England |
| EPI_ISL_423207 | Respiratory Virus Unit, Microbiology Services Colindale, Public Health England | Respiratory Virus Unit, Microbiology Services Colindale, Public Health England |
| EPI_ISL_423208 | Respiratory Virus Unit, Microbiology Services Colindale, Public Health England | Respiratory Virus Unit, Microbiology Services Colindale, Public Health England |
| EPI_ISL_423209 | Respiratory Virus Unit, Microbiology Services Colindale, Public Health England | Respiratory Virus Unit, Microbiology Services Colindale, Public Health England |
| EPI_ISL_423210 | Respiratory Virus Unit, Microbiology Services Colindale, Public Health England | Respiratory Virus Unit, Microbiology Services Colindale, Public Health England |
| EPI_ISL_423211 | Respiratory Virus Unit, Microbiology Services Colindale, Public Health England | Respiratory Virus Unit, Microbiology Services Colindale, Public Health England |
| EPI_ISL_423212 | Respiratory Virus Unit, Microbiology Services Colindale, Public Health England | Respiratory Virus Unit, Microbiology Services Colindale, Public Health England |
| EPI_ISL_423213 | Respiratory Virus Unit, Microbiology Services Colindale, Public Health England | Respiratory Virus Unit, Microbiology Services Colindale, Public Health England |
| EPI_ISL_423214 | Respiratory Virus Unit, Microbiology Services Colindale, Public Health England | Respiratory Virus Unit, Microbiology Services Colindale, Public Health England |
| EPI_ISL_423215 | Respiratory Virus Unit, Microbiology Services Colindale, Public Health England | Respiratory Virus Unit, Microbiology Services Colindale, Public Health England |
| EPI_ISL_423216 | Respiratory Virus Unit, Microbiology Services Colindale, Public Health England | Respiratory Virus Unit, Microbiology Services Colindale, Public Health England |
| EPI_ISL_423217 | Respiratory Virus Unit, Microbiology Services Colindale, Public Health England | Respiratory Virus Unit, Microbiology Services Colindale, Public Health England |
| EPI_ISL_423218 | Respiratory Virus Unit, Microbiology Services Colindale, Public Health England | Respiratory Virus Unit, Microbiology Services Colindale, Public Health England |
| EPI_ISL_423219 | Respiratory Virus Unit, Microbiology Services Colindale, Public Health England | Respiratory Virus Unit, Microbiology Services Colindale, Public Health England |
| EPI_ISL_423220 | Respiratory Virus Unit, Microbiology Services Colindale, Public Health England | Respiratory Virus Unit, Microbiology Services Colindale, Public Health England |
| EPI_ISL_423221 | Respiratory Virus Unit, Microbiology Services Colindale, Public Health England | Respiratory Virus Unit, Microbiology Services Colindale, Public Health England |

| EPI_Accession_ID | Originating Laboratory | Submitting Laboratory |
|---|---|---|
| EPI_ISL_423222 | Respiratory Virus Unit, Microbiology Services Colindale, Public Health England | Respiratory Virus Unit, Microbiology Services Colindale, Public Health England |
| EPI_ISL_423223 | Respiratory Virus Unit, Microbiology Services Colindale, Public Health England | Respiratory Virus Unit, Microbiology Services Colindale, Public Health England |
| EPI_ISL_423224 | Respiratory Virus Unit, Microbiology Services Colindale, Public Health England | Respiratory Virus Unit, Microbiology Services Colindale, Public Health England |
| EPI_ISL_423225 | Respiratory Virus Unit, Microbiology Services Colindale, Public Health England | Respiratory Virus Unit, Microbiology Services Colindale, Public Health England |
| EPI_ISL_423226 | Respiratory Virus Unit, Microbiology Services Colindale, Public Health England | Respiratory Virus Unit, Microbiology Services Colindale, Public Health England |
| EPI_ISL_423227 | Respiratory Virus Unit, Microbiology Services Colindale, Public Health England | Respiratory Virus Unit, Microbiology Services Colindale, Public Health England |
| EPI_ISL_423228 | Respiratory Virus Unit, Microbiology Services Colindale, Public Health England | Respiratory Virus Unit, Microbiology Services Colindale, Public Health England |
| EPI_ISL_423229 | Respiratory Virus Unit, Microbiology Services Colindale, Public Health England | Respiratory Virus Unit, Microbiology Services Colindale, Public Health England |
| EPI_ISL_423230 | Respiratory Virus Unit, Microbiology Services Colindale, Public Health England | Respiratory Virus Unit, Microbiology Services Colindale, Public Health England |
| EPI_ISL_423231 | Respiratory Virus Unit, Microbiology Services Colindale, Public Health England | Respiratory Virus Unit, Microbiology Services Colindale, Public Health England |
| EPI_ISL_423232 | Respiratory Virus Unit, Microbiology Services Colindale, Public Health England | Respiratory Virus Unit, Microbiology Services Colindale, Public Health England |
| EPI_ISL_423233 | Respiratory Virus Unit, Microbiology Services Colindale, Public Health England | Respiratory Virus Unit, Microbiology Services Colindale, Public Health England |
| EPI_ISL_423234 | Respiratory Virus Unit, Microbiology Services Colindale, Public Health England | Respiratory Virus Unit, Microbiology Services Colindale, Public Health England |
| EPI_ISL_423235 | Respiratory Virus Unit, Microbiology Services Colindale, Public Health England | Respiratory Virus Unit, Microbiology Services Colindale, Public Health England |
| EPI_ISL_423236 | Respiratory Virus Unit, Microbiology Services Colindale, Public Health England | Respiratory Virus Unit, Microbiology Services Colindale, Public Health England |
| EPI_ISL_423237 | Respiratory Virus Unit, Microbiology Services Colindale, Public Health England | Respiratory Virus Unit, Microbiology Services Colindale, Public Health England |

| EPI_Accession_ID | Originating Laboratory | Submitting Laboratory |
| --- | --- | --- |
| EPI_ISL_423238 | Respiratory Virus Unit, Microbiology Services Colindale, Public Health England | Respiratory Virus Unit, Microbiology Services Colindale, Public Health England |
| EPI_ISL_423239 | Respiratory Virus Unit, Microbiology Services Colindale, Public Health England | Respiratory Virus Unit, Microbiology Services Colindale, Public Health England |
| EPI_ISL_423240 | Respiratory Virus Unit, Microbiology Services Colindale, Public Health England | Respiratory Virus Unit, Microbiology Services Colindale, Public Health England |
| EPI_ISL_423241 | Respiratory Virus Unit, Microbiology Services Colindale, Public Health England | Respiratory Virus Unit, Microbiology Services Colindale, Public Health England |
| EPI_ISL_423242 | Respiratory Virus Unit, Microbiology Services Colindale, Public Health England | Respiratory Virus Unit, Microbiology Services Colindale, Public Health England |
| EPI_ISL_423243 | Respiratory Virus Unit, Microbiology Services Colindale, Public Health England | Respiratory Virus Unit, Microbiology Services Colindale, Public Health England |
| EPI_ISL_423244 | Respiratory Virus Unit, Microbiology Services Colindale, Public Health England | Respiratory Virus Unit, Microbiology Services Colindale, Public Health England |
| EPI_ISL_423245 | Respiratory Virus Unit, Microbiology Services Colindale, Public Health England | Respiratory Virus Unit, Microbiology Services Colindale, Public Health England |
| EPI_ISL_423246 | Respiratory Virus Unit, Microbiology Services Colindale, Public Health England | Respiratory Virus Unit, Microbiology Services Colindale, Public Health England |
| EPI_ISL_423247 | Respiratory Virus Unit, Microbiology Services Colindale, Public Health England | Respiratory Virus Unit, Microbiology Services Colindale, Public Health England |
| EPI_ISL_423248 | Respiratory Virus Unit, Microbiology Services Colindale, Public Health England | Respiratory Virus Unit, Microbiology Services Colindale, Public Health England |
| EPI_ISL_423249 | Respiratory Virus Unit, Microbiology Services Colindale, Public Health England | Respiratory Virus Unit, Microbiology Services Colindale, Public Health England |
| EPI_ISL_423250 | Respiratory Virus Unit, Microbiology Services Colindale, Public Health England | Respiratory Virus Unit, Microbiology Services Colindale, Public Health England |
| EPI_ISL_423251 | Respiratory Virus Unit, Microbiology Services Colindale, Public Health England | Respiratory Virus Unit, Microbiology Services Colindale, Public Health England |
| EPI_ISL_423252 | Respiratory Virus Unit, Microbiology Services Colindale, Public Health England | Respiratory Virus Unit, Microbiology Services Colindale, Public Health England |
| EPI_ISL_423253 | Respiratory Virus Unit, Microbiology Services Colindale, Public Health England | Respiratory Virus Unit, Microbiology Services Colindale, Public Health England |

| EPI_Accession_ID | Originating Laboratory | Submitting Laboratory |
|---|---|---|
| EPI_ISL_423254 | Respiratory Virus Unit, Microbiology Services Colindale, Public Health England | Respiratory Virus Unit, Microbiology Services Colindale, Public Health England |
| EPI_ISL_423255 | Respiratory Virus Unit, Microbiology Services Colindale, Public Health England | Respiratory Virus Unit, Microbiology Services Colindale, Public Health England |
| EPI_ISL_423256 | Respiratory Virus Unit, Microbiology Services Colindale, Public Health England | Respiratory Virus Unit, Microbiology Services Colindale, Public Health England |
| EPI_ISL_423258 | Respiratory Virus Unit, Microbiology Services Colindale, Public Health England | Respiratory Virus Unit, Microbiology Services Colindale, Public Health England |
| EPI_ISL_423259 | Respiratory Virus Unit, Microbiology Services Colindale, Public Health England | Respiratory Virus Unit, Microbiology Services Colindale, Public Health England |
| EPI_ISL_423260 | Respiratory Virus Unit, Microbiology Services Colindale, Public Health England | Respiratory Virus Unit, Microbiology Services Colindale, Public Health England |
| EPI_ISL_423261 | Respiratory Virus Unit, Microbiology Services Colindale, Public Health England | Respiratory Virus Unit, Microbiology Services Colindale, Public Health England |
| EPI_ISL_423262 | Respiratory Virus Unit, Microbiology Services Colindale, Public Health England | Respiratory Virus Unit, Microbiology Services Colindale, Public Health England |
| EPI_ISL_423263 | Respiratory Virus Unit, Microbiology Services Colindale, Public Health England | Respiratory Virus Unit, Microbiology Services Colindale, Public Health England |
| EPI_ISL_423264 | Respiratory Virus Unit, Microbiology Services Colindale, Public Health England | Respiratory Virus Unit, Microbiology Services Colindale, Public Health England |
| EPI_ISL_423265 | Respiratory Virus Unit, Microbiology Services Colindale, Public Health England | Respiratory Virus Unit, Microbiology Services Colindale, Public Health England |
| EPI_ISL_423266 | Respiratory Virus Unit, Microbiology Services Colindale, Public Health England | Respiratory Virus Unit, Microbiology Services Colindale, Public Health England |
| EPI_ISL_423267 | Respiratory Virus Unit, Microbiology Services Colindale, Public Health England | Respiratory Virus Unit, Microbiology Services Colindale, Public Health England |
| EPI_ISL_423268 | Respiratory Virus Unit, Microbiology Services Colindale, Public Health England | Respiratory Virus Unit, Microbiology Services Colindale, Public Health England |
| EPI_ISL_423269 | Respiratory Virus Unit, Microbiology Services Colindale, Public Health England | Respiratory Virus Unit, Microbiology Services Colindale, Public Health England |
| EPI_ISL_423270 | Respiratory Virus Unit, Microbiology Services Colindale, Public Health England | Respiratory Virus Unit, Microbiology Services Colindale, Public Health England |

| EPI_Accession_ID | Originating Laboratory | Submitting Laboratory |
|---|---|---|
| EPI_ISL_423271 | Respiratory Virus Unit, Microbiology Services Colindale, Public Health England | Respiratory Virus Unit, Microbiology Services Colindale, Public Health England |
| EPI_ISL_423272 | Respiratory Virus Unit, Microbiology Services Colindale, Public Health England | Respiratory Virus Unit, Microbiology Services Colindale, Public Health England |
| EPI_ISL_423273 | Respiratory Virus Unit, Microbiology Services Colindale, Public Health England | Respiratory Virus Unit, Microbiology Services Colindale, Public Health England |
| EPI_ISL_423274 | Respiratory Virus Unit, Microbiology Services Colindale, Public Health England | Respiratory Virus Unit, Microbiology Services Colindale, Public Health England |
| EPI_ISL_423275 | Respiratory Virus Unit, Microbiology Services Colindale, Public Health England | Respiratory Virus Unit, Microbiology Services Colindale, Public Health England |
| EPI_ISL_423276 | Respiratory Virus Unit, Microbiology Services Colindale, Public Health England | Respiratory Virus Unit, Microbiology Services Colindale, Public Health England |
| EPI_ISL_423277 | Respiratory Virus Unit, Microbiology Services Colindale, Public Health England | Respiratory Virus Unit, Microbiology Services Colindale, Public Health England |
| EPI_ISL_423278 | Respiratory Virus Unit, Microbiology Services Colindale, Public Health England | Respiratory Virus Unit, Microbiology Services Colindale, Public Health England |
| EPI_ISL_423279 | Respiratory Virus Unit, Microbiology Services Colindale, Public Health England | Respiratory Virus Unit, Microbiology Services Colindale, Public Health England |
| EPI_ISL_423280 | Respiratory Virus Unit, Microbiology Services Colindale, Public Health England | Respiratory Virus Unit, Microbiology Services Colindale, Public Health England |
| EPI_ISL_423281 | Respiratory Virus Unit, Microbiology Services Colindale, Public Health England | Respiratory Virus Unit, Microbiology Services Colindale, Public Health England |
| EPI_ISL_423282 | Respiratory Virus Unit, Microbiology Services Colindale, Public Health England | Respiratory Virus Unit, Microbiology Services Colindale, Public Health England |
| EPI_ISL_423283 | Respiratory Virus Unit, Microbiology Services Colindale, Public Health England | Respiratory Virus Unit, Microbiology Services Colindale, Public Health England |
| EPI_ISL_423284 | Respiratory Virus Unit, Microbiology Services Colindale, Public Health England | Respiratory Virus Unit, Microbiology Services Colindale, Public Health England |
| EPI_ISL_423285 | Respiratory Virus Unit, Microbiology Services Colindale, Public Health England | Respiratory Virus Unit, Microbiology Services Colindale, Public Health England |
| EPI_ISL_423286 | Respiratory Virus Unit, Microbiology Services Colindale, Public Health England | Respiratory Virus Unit, Microbiology Services Colindale, Public Health England |

| EPI_Accession_ID | Originating Laboratory | Submitting Laboratory |
|---|---|---|
| EPI_ISL_423287 | Respiratory Virus Unit, Microbiology Services Colindale, Public Health England | Respiratory Virus Unit, Microbiology Services Colindale, Public Health England |
| EPI_ISL_423288 | Respiratory Virus Unit, Microbiology Services Colindale, Public Health England | Respiratory Virus Unit, Microbiology Services Colindale, Public Health England |
| EPI_ISL_423289 | Respiratory Virus Unit, Microbiology Services Colindale, Public Health England | Respiratory Virus Unit, Microbiology Services Colindale, Public Health England |
| EPI_ISL_423290 | Respiratory Virus Unit, Microbiology Services Colindale, Public Health England | Respiratory Virus Unit, Microbiology Services Colindale, Public Health England |
| EPI_ISL_423291 | Respiratory Virus Unit, Microbiology Services Colindale, Public Health England | Respiratory Virus Unit, Microbiology Services Colindale, Public Health England |
| EPI_ISL_423292 | Respiratory Virus Unit, Microbiology Services Colindale, Public Health England | Respiratory Virus Unit, Microbiology Services Colindale, Public Health England |
| EPI_ISL_423293 | Respiratory Virus Unit, Microbiology Services Colindale, Public Health England | Respiratory Virus Unit, Microbiology Services Colindale, Public Health England |
| EPI_ISL_423294 | Respiratory Virus Unit, Microbiology Services Colindale, Public Health England | Respiratory Virus Unit, Microbiology Services Colindale, Public Health England |
| EPI_ISL_423295 | Respiratory Virus Unit, Microbiology Services Colindale, Public Health England | Respiratory Virus Unit, Microbiology Services Colindale, Public Health England |
| EPI_ISL_423296 | Respiratory Virus Unit, Microbiology Services Colindale, Public Health England | Respiratory Virus Unit, Microbiology Services Colindale, Public Health England |
| EPI_ISL_423297 | Respiratory Virus Unit, Microbiology Services Colindale, Public Health England | Respiratory Virus Unit, Microbiology Services Colindale, Public Health England |
| EPI_ISL_423298 | Respiratory Virus Unit, Microbiology Services Colindale, Public Health England | Respiratory Virus Unit, Microbiology Services Colindale, Public Health England |
| EPI_ISL_423299 | Respiratory Virus Unit, Microbiology Services Colindale, Public Health England | Respiratory Virus Unit, Microbiology Services Colindale, Public Health England |
| EPI_ISL_423300 | Respiratory Virus Unit, Microbiology Services Colindale, Public Health England | Respiratory Virus Unit, Microbiology Services Colindale, Public Health England |
| EPI_ISL_423301 | Respiratory Virus Unit, Microbiology Services Colindale, Public Health England | Respiratory Virus Unit, Microbiology Services Colindale, Public Health England |
| EPI_ISL_423302 | Respiratory Virus Unit, Microbiology Services Colindale, Public Health England | Respiratory Virus Unit, Microbiology Services Colindale, Public Health England |

| EPI_Accession_ID | Originating Laboratory | Submitting Laboratory |
|---|---|---|
| EPI_ISL_423303 | Respiratory Virus Unit, Microbiology Services Colindale, Public Health England | Respiratory Virus Unit, Microbiology Services Colindale, Public Health England |
| EPI_ISL_423304 | Respiratory Virus Unit, Microbiology Services Colindale, Public Health England | Respiratory Virus Unit, Microbiology Services Colindale, Public Health England |
| EPI_ISL_423305 | Respiratory Virus Unit, Microbiology Services Colindale, Public Health England | Respiratory Virus Unit, Microbiology Services Colindale, Public Health England |
| EPI_ISL_423306 | Respiratory Virus Unit, Microbiology Services Colindale, Public Health England | Respiratory Virus Unit, Microbiology Services Colindale, Public Health England |
| EPI_ISL_423307 | Respiratory Virus Unit, Microbiology Services Colindale, Public Health England | Respiratory Virus Unit, Microbiology Services Colindale, Public Health England |
| EPI_ISL_423308 | Respiratory Virus Unit, Microbiology Services Colindale, Public Health England | Respiratory Virus Unit, Microbiology Services Colindale, Public Health England |
| EPI_ISL_423309 | Respiratory Virus Unit, Microbiology Services Colindale, Public Health England | Respiratory Virus Unit, Microbiology Services Colindale, Public Health England |
| EPI_ISL_423310 | Respiratory Virus Unit, Microbiology Services Colindale, Public Health England | Respiratory Virus Unit, Microbiology Services Colindale, Public Health England |
| EPI_ISL_423311 | Respiratory Virus Unit, Microbiology Services Colindale, Public Health England | Respiratory Virus Unit, Microbiology Services Colindale, Public Health England |
| EPI_ISL_423312 | Respiratory Virus Unit, Microbiology Services Colindale, Public Health England | Respiratory Virus Unit, Microbiology Services Colindale, Public Health England |
| EPI_ISL_423313 | Respiratory Virus Unit, Microbiology Services Colindale, Public Health England | Respiratory Virus Unit, Microbiology Services Colindale, Public Health England |
| EPI_ISL_423314 | Respiratory Virus Unit, Microbiology Services Colindale, Public Health England | Respiratory Virus Unit, Microbiology Services Colindale, Public Health England |
| EPI_ISL_423315 | Respiratory Virus Unit, Microbiology Services Colindale, Public Health England | Respiratory Virus Unit, Microbiology Services Colindale, Public Health England |
| EPI_ISL_423316 | Respiratory Virus Unit, Microbiology Services Colindale, Public Health England | Respiratory Virus Unit, Microbiology Services Colindale, Public Health England |
| EPI_ISL_423317 | Respiratory Virus Unit, Microbiology Services Colindale, Public Health England | Respiratory Virus Unit, Microbiology Services Colindale, Public Health England |
| EPI_ISL_423318 | Respiratory Virus Unit, Microbiology Services Colindale, Public Health England | Respiratory Virus Unit, Microbiology Services Colindale, Public Health England |

| EPI_Accession_ID | Originating Laboratory | Submitting Laboratory |
|---|---|---|
| EPI_ISL_423319 | Respiratory Virus Unit, Microbiology Services Colindale, Public Health England | Respiratory Virus Unit, Microbiology Services Colindale, Public Health England |
| EPI_ISL_423320 | Respiratory Virus Unit, Microbiology Services Colindale, Public Health England | Respiratory Virus Unit, Microbiology Services Colindale, Public Health England |
| EPI_ISL_423321 | Respiratory Virus Unit, Microbiology Services Colindale, Public Health England | Respiratory Virus Unit, Microbiology Services Colindale, Public Health England |
| EPI_ISL_423322 | Respiratory Virus Unit, Microbiology Services Colindale, Public Health England | Respiratory Virus Unit, Microbiology Services Colindale, Public Health England |
| EPI_ISL_423323 | Respiratory Virus Unit, Microbiology Services Colindale, Public Health England | Respiratory Virus Unit, Microbiology Services Colindale, Public Health England |
| EPI_ISL_423324 | Respiratory Virus Unit, Microbiology Services Colindale, Public Health England | Respiratory Virus Unit, Microbiology Services Colindale, Public Health England |
| EPI_ISL_423325 | Respiratory Virus Unit, Microbiology Services Colindale, Public Health England | Respiratory Virus Unit, Microbiology Services Colindale, Public Health England |
| EPI_ISL_423326 | Respiratory Virus Unit, Microbiology Services Colindale, Public Health England | Respiratory Virus Unit, Microbiology Services Colindale, Public Health England |
| EPI_ISL_423327 | Respiratory Virus Unit, Microbiology Services Colindale, Public Health England | Respiratory Virus Unit, Microbiology Services Colindale, Public Health England |
| EPI_ISL_423328 | Respiratory Virus Unit, Microbiology Services Colindale, Public Health England | Respiratory Virus Unit, Microbiology Services Colindale, Public Health England |
| EPI_ISL_423329 | Respiratory Virus Unit, Microbiology Services Colindale, Public Health England | Respiratory Virus Unit, Microbiology Services Colindale, Public Health England |
| EPI_ISL_423330 | Respiratory Virus Unit, Microbiology Services Colindale, Public Health England | Respiratory Virus Unit, Microbiology Services Colindale, Public Health England |
| EPI_ISL_423331 | Respiratory Virus Unit, Microbiology Services Colindale, Public Health England | Respiratory Virus Unit, Microbiology Services Colindale, Public Health England |
| EPI_ISL_423332 | Respiratory Virus Unit, Microbiology Services Colindale, Public Health England | Respiratory Virus Unit, Microbiology Services Colindale, Public Health England |
| EPI_ISL_423333 | Respiratory Virus Unit, Microbiology Services Colindale, Public Health England | Respiratory Virus Unit, Microbiology Services Colindale, Public Health England |
| EPI_ISL_423334 | Respiratory Virus Unit, Microbiology Services Colindale, Public Health England | Respiratory Virus Unit, Microbiology Services Colindale, Public Health England |

| EPI_Accession_ID | Originating Laboratory | Submitting Laboratory |
|---|---|---|
| EPI_ISL_423335 | Respiratory Virus Unit, Microbiology Services Colindale, Public Health England | Respiratory Virus Unit, Microbiology Services Colindale, Public Health England |
| EPI_ISL_423336 | Respiratory Virus Unit, Microbiology Services Colindale, Public Health England | Respiratory Virus Unit, Microbiology Services Colindale, Public Health England |
| EPI_ISL_423337 | Respiratory Virus Unit, Microbiology Services Colindale, Public Health England | Respiratory Virus Unit, Microbiology Services Colindale, Public Health England |
| EPI_ISL_423338 | Respiratory Virus Unit, Microbiology Services Colindale, Public Health England | Respiratory Virus Unit, Microbiology Services Colindale, Public Health England |
| EPI_ISL_423339 | Respiratory Virus Unit, Microbiology Services Colindale, Public Health England | Respiratory Virus Unit, Microbiology Services Colindale, Public Health England |
| EPI_ISL_423340 | Respiratory Virus Unit, Microbiology Services Colindale, Public Health England | Respiratory Virus Unit, Microbiology Services Colindale, Public Health England |
| EPI_ISL_423341 | Respiratory Virus Unit, Microbiology Services Colindale, Public Health England | Respiratory Virus Unit, Microbiology Services Colindale, Public Health England |
| EPI_ISL_423342 | Respiratory Virus Unit, Microbiology Services Colindale, Public Health England | Respiratory Virus Unit, Microbiology Services Colindale, Public Health England |
| EPI_ISL_423343 | Respiratory Virus Unit, Microbiology Services Colindale, Public Health England | Respiratory Virus Unit, Microbiology Services Colindale, Public Health England |
| EPI_ISL_423344 | Respiratory Virus Unit, Microbiology Services Colindale, Public Health England | Respiratory Virus Unit, Microbiology Services Colindale, Public Health England |
| EPI_ISL_423345 | Respiratory Virus Unit, Microbiology Services Colindale, Public Health England | Respiratory Virus Unit, Microbiology Services Colindale, Public Health England |
| EPI_ISL_423346 | Respiratory Virus Unit, Microbiology Services Colindale, Public Health England | Respiratory Virus Unit, Microbiology Services Colindale, Public Health England |
| EPI_ISL_423347 | Respiratory Virus Unit, Microbiology Services Colindale, Public Health England | Respiratory Virus Unit, Microbiology Services Colindale, Public Health England |
| EPI_ISL_423348 | Respiratory Virus Unit, Microbiology Services Colindale, Public Health England | Respiratory Virus Unit, Microbiology Services Colindale, Public Health England |
| EPI_ISL_423349 | Respiratory Virus Unit, Microbiology Services Colindale, Public Health England | Respiratory Virus Unit, Microbiology Services Colindale, Public Health England |
| EPI_ISL_423350 | Respiratory Virus Unit, Microbiology Services Colindale, Public Health England | Respiratory Virus Unit, Microbiology Services Colindale, Public Health England |

| EPI_Accession_ID | Originating Laboratory | Submitting Laboratory |
|---|---|---|
| EPI_ISL_423351 | Respiratory Virus Unit, Microbiology Services Colindale, Public Health England | Respiratory Virus Unit, Microbiology Services Colindale, Public Health England |
| EPI_ISL_423352 | Respiratory Virus Unit, Microbiology Services Colindale, Public Health England | Respiratory Virus Unit, Microbiology Services Colindale, Public Health England |
| EPI_ISL_423353 | Respiratory Virus Unit, Microbiology Services Colindale, Public Health England | Respiratory Virus Unit, Microbiology Services Colindale, Public Health England |
| EPI_ISL_423354 | Respiratory Virus Unit, Microbiology Services Colindale, Public Health England | Respiratory Virus Unit, Microbiology Services Colindale, Public Health England |
| EPI_ISL_423355 | Respiratory Virus Unit, Microbiology Services Colindale, Public Health England | Respiratory Virus Unit, Microbiology Services Colindale, Public Health England |
| EPI_ISL_423356 | Respiratory Virus Unit, Microbiology Services Colindale, Public Health England | Respiratory Virus Unit, Microbiology Services Colindale, Public Health England |
| EPI_ISL_423357 | Respiratory Virus Unit, Microbiology Services Colindale, Public Health England | Respiratory Virus Unit, Microbiology Services Colindale, Public Health England |
| EPI_ISL_423358 | Respiratory Virus Unit, Microbiology Services Colindale, Public Health England | Respiratory Virus Unit, Microbiology Services Colindale, Public Health England |
| EPI_ISL_423359 | Respiratory Virus Unit, Microbiology Services Colindale, Public Health England | Respiratory Virus Unit, Microbiology Services Colindale, Public Health England |
| EPI_ISL_423360 | Respiratory Virus Unit, Microbiology Services Colindale, Public Health England | Respiratory Virus Unit, Microbiology Services Colindale, Public Health England |
| EPI_ISL_423361 | Respiratory Virus Unit, Microbiology Services Colindale, Public Health England | Respiratory Virus Unit, Microbiology Services Colindale, Public Health England |
| EPI_ISL_423362 | Respiratory Virus Unit, Microbiology Services Colindale, Public Health England | Respiratory Virus Unit, Microbiology Services Colindale, Public Health England |
| EPI_ISL_423363 | Respiratory Virus Unit, Microbiology Services Colindale, Public Health England | Respiratory Virus Unit, Microbiology Services Colindale, Public Health England |
| EPI_ISL_423364 | Respiratory Virus Unit, Microbiology Services Colindale, Public Health England | Respiratory Virus Unit, Microbiology Services Colindale, Public Health England |
| EPI_ISL_423365 | Respiratory Virus Unit, Microbiology Services Colindale, Public Health England | Respiratory Virus Unit, Microbiology Services Colindale, Public Health England |
| EPI_ISL_423366 | Respiratory Virus Unit, Microbiology Services Colindale, Public Health England | Respiratory Virus Unit, Microbiology Services Colindale, Public Health England |

| EPI_Accession_ID | Originating Laboratory | Submitting Laboratory |
|---|---|---|
| EPI_ISL_423367 | Respiratory Virus Unit, Microbiology Services Colindale, Public Health England | Respiratory Virus Unit, Microbiology Services Colindale, Public Health England |
| EPI_ISL_423368 | Respiratory Virus Unit, Microbiology Services Colindale, Public Health England | Respiratory Virus Unit, Microbiology Services Colindale, Public Health England |
| EPI_ISL_423369 | Respiratory Virus Unit, Microbiology Services Colindale, Public Health England | Respiratory Virus Unit, Microbiology Services Colindale, Public Health England |
| EPI_ISL_423370 | Respiratory Virus Unit, Microbiology Services Colindale, Public Health England | Respiratory Virus Unit, Microbiology Services Colindale, Public Health England |
| EPI_ISL_423371 | Respiratory Virus Unit, Microbiology Services Colindale, Public Health England | Respiratory Virus Unit, Microbiology Services Colindale, Public Health England |
| EPI_ISL_423372 | Respiratory Virus Unit, Microbiology Services Colindale, Public Health England | Respiratory Virus Unit, Microbiology Services Colindale, Public Health England |
| EPI_ISL_423373 | Respiratory Virus Unit, Microbiology Services Colindale, Public Health England | Respiratory Virus Unit, Microbiology Services Colindale, Public Health England |
| EPI_ISL_423374 | Respiratory Virus Unit, Microbiology Services Colindale, Public Health England | Respiratory Virus Unit, Microbiology Services Colindale, Public Health England |
| EPI_ISL_423375 | Respiratory Virus Unit, Microbiology Services Colindale, Public Health England | Respiratory Virus Unit, Microbiology Services Colindale, Public Health England |
| EPI_ISL_423376 | Respiratory Virus Unit, Microbiology Services Colindale, Public Health England | Respiratory Virus Unit, Microbiology Services Colindale, Public Health England |
| EPI_ISL_423377 | Respiratory Virus Unit, Microbiology Services Colindale, Public Health England | Respiratory Virus Unit, Microbiology Services Colindale, Public Health England |
| EPI_ISL_423378 | Respiratory Virus Unit, Microbiology Services Colindale, Public Health England | Respiratory Virus Unit, Microbiology Services Colindale, Public Health England |
| EPI_ISL_423379 | Respiratory Virus Unit, Microbiology Services Colindale, Public Health England | Respiratory Virus Unit, Microbiology Services Colindale, Public Health England |
| EPI_ISL_423380 | Respiratory Virus Unit, Microbiology Services Colindale, Public Health England | Respiratory Virus Unit, Microbiology Services Colindale, Public Health England |
| EPI_ISL_423381 | Respiratory Virus Unit, Microbiology Services Colindale, Public Health England | Respiratory Virus Unit, Microbiology Services Colindale, Public Health England |
| EPI_ISL_423382 | Respiratory Virus Unit, Microbiology Services Colindale, Public Health England | Respiratory Virus Unit, Microbiology Services Colindale, Public Health England |

| EPI_Accession_ID | Originating Laboratory | Submitting Laboratory |
|---|---|---|
| EPI_ISL_423383 | Respiratory Virus Unit, Microbiology Services Colindale, Public Health England | Respiratory Virus Unit, Microbiology Services Colindale, Public Health England |
| EPI_ISL_423384 | Respiratory Virus Unit, Microbiology Services Colindale, Public Health England | Respiratory Virus Unit, Microbiology Services Colindale, Public Health England |
| EPI_ISL_423385 | Respiratory Virus Unit, Microbiology Services Colindale, Public Health England | Respiratory Virus Unit, Microbiology Services Colindale, Public Health England |
| EPI_ISL_423386 | Respiratory Virus Unit, Microbiology Services Colindale, Public Health England | Respiratory Virus Unit, Microbiology Services Colindale, Public Health England |
| EPI_ISL_423387 | Respiratory Virus Unit, Microbiology Services Colindale, Public Health England | Respiratory Virus Unit, Microbiology Services Colindale, Public Health England |
| EPI_ISL_423388 | Respiratory Virus Unit, Microbiology Services Colindale, Public Health England | Respiratory Virus Unit, Microbiology Services Colindale, Public Health England |
| EPI_ISL_423389 | Respiratory Virus Unit, Microbiology Services Colindale, Public Health England | Respiratory Virus Unit, Microbiology Services Colindale, Public Health England |
| EPI_ISL_423390 | Respiratory Virus Unit, Microbiology Services Colindale, Public Health England | Respiratory Virus Unit, Microbiology Services Colindale, Public Health England |
| EPI_ISL_423391 | Respiratory Virus Unit, Microbiology Services Colindale, Public Health England | Respiratory Virus Unit, Microbiology Services Colindale, Public Health England |
| EPI_ISL_423392 | Respiratory Virus Unit, Microbiology Services Colindale, Public Health England | Respiratory Virus Unit, Microbiology Services Colindale, Public Health England |
| EPI_ISL_423393 | Respiratory Virus Unit, Microbiology Services Colindale, Public Health England | Respiratory Virus Unit, Microbiology Services Colindale, Public Health England |
| EPI_ISL_423394 | Respiratory Virus Unit, Microbiology Services Colindale, Public Health England | Respiratory Virus Unit, Microbiology Services Colindale, Public Health England |
| EPI_ISL_423395 | Respiratory Virus Unit, Microbiology Services Colindale, Public Health England | Respiratory Virus Unit, Microbiology Services Colindale, Public Health England |
| EPI_ISL_423396 | Respiratory Virus Unit, Microbiology Services Colindale, Public Health England | Respiratory Virus Unit, Microbiology Services Colindale, Public Health England |
| EPI_ISL_423397 | Respiratory Virus Unit, Microbiology Services Colindale, Public Health England | Respiratory Virus Unit, Microbiology Services Colindale, Public Health England |
| EPI_ISL_423398 | Respiratory Virus Unit, Microbiology Services Colindale, Public Health England | Respiratory Virus Unit, Microbiology Services Colindale, Public Health England |

| EPI_Accession_ID | Originating Laboratory | Submitting Laboratory |
|---|---|---|
| EPI_ISL_423399 | Respiratory Virus Unit, Microbiology Services Colindale, Public Health England | Respiratory Virus Unit, Microbiology Services Colindale, Public Health England |
| EPI_ISL_423400 | Respiratory Virus Unit, Microbiology Services Colindale, Public Health England | Respiratory Virus Unit, Microbiology Services Colindale, Public Health England |
| EPI_ISL_423401 | Respiratory Virus Unit, Microbiology Services Colindale, Public Health England | Respiratory Virus Unit, Microbiology Services Colindale, Public Health England |
| EPI_ISL_423402 | Respiratory Virus Unit, Microbiology Services Colindale, Public Health England | Respiratory Virus Unit, Microbiology Services Colindale, Public Health England |
| EPI_ISL_423403 | Respiratory Virus Unit, Microbiology Services Colindale, Public Health England | Respiratory Virus Unit, Microbiology Services Colindale, Public Health England |
| EPI_ISL_423404 | Respiratory Virus Unit, Microbiology Services Colindale, Public Health England | Respiratory Virus Unit, Microbiology Services Colindale, Public Health England |
| EPI_ISL_423405 | Respiratory Virus Unit, Microbiology Services Colindale, Public Health England | Respiratory Virus Unit, Microbiology Services Colindale, Public Health England |
| EPI_ISL_423406 | Respiratory Virus Unit, Microbiology Services Colindale, Public Health England | Respiratory Virus Unit, Microbiology Services Colindale, Public Health England |
| EPI_ISL_423407 | Respiratory Virus Unit, Microbiology Services Colindale, Public Health England | Respiratory Virus Unit, Microbiology Services Colindale, Public Health England |
| EPI_ISL_423408 | Respiratory Virus Unit, Microbiology Services Colindale, Public Health England | Respiratory Virus Unit, Microbiology Services Colindale, Public Health England |
| EPI_ISL_423409 | Respiratory Virus Unit, Microbiology Services Colindale, Public Health England | Respiratory Virus Unit, Microbiology Services Colindale, Public Health England |
| EPI_ISL_423410 | Respiratory Virus Unit, Microbiology Services Colindale, Public Health England | Respiratory Virus Unit, Microbiology Services Colindale, Public Health England |
| EPI_ISL_423411 | Respiratory Virus Unit, Microbiology Services Colindale, Public Health England | Respiratory Virus Unit, Microbiology Services Colindale, Public Health England |
| EPI_ISL_423412 | Respiratory Virus Unit, Microbiology Services Colindale, Public Health England | Respiratory Virus Unit, Microbiology Services Colindale, Public Health England |
| EPI_ISL_423413 | Respiratory Virus Unit, Microbiology Services Colindale, Public Health England | Respiratory Virus Unit, Microbiology Services Colindale, Public Health England |
| EPI_ISL_423414 | Respiratory Virus Unit, Microbiology Services Colindale, Public Health England | Respiratory Virus Unit, Microbiology Services Colindale, Public Health England |

| EPI_Accession_ID | Originating Laboratory | Submitting Laboratory |
|---|---|---|
| EPI_ISL_423415 | Respiratory Virus Unit, Microbiology Services Colindale, Public Health England | Respiratory Virus Unit, Microbiology Services Colindale, Public Health England |
| EPI_ISL_423416 | Respiratory Virus Unit, Microbiology Services Colindale, Public Health England | Respiratory Virus Unit, Microbiology Services Colindale, Public Health England |
| EPI_ISL_423417 | Respiratory Virus Unit, Microbiology Services Colindale, Public Health England | Respiratory Virus Unit, Microbiology Services Colindale, Public Health England |
| EPI_ISL_423418 | Respiratory Virus Unit, Microbiology Services Colindale, Public Health England | Respiratory Virus Unit, Microbiology Services Colindale, Public Health England |
| EPI_ISL_423419 | Respiratory Virus Unit, Microbiology Services Colindale, Public Health England | Respiratory Virus Unit, Microbiology Services Colindale, Public Health England |
| EPI_ISL_423420 | Respiratory Virus Unit, Microbiology Services Colindale, Public Health England | Respiratory Virus Unit, Microbiology Services Colindale, Public Health England |
| EPI_ISL_423421 | Respiratory Virus Unit, Microbiology Services Colindale, Public Health England | Respiratory Virus Unit, Microbiology Services Colindale, Public Health England |
| EPI_ISL_423422 | Respiratory Virus Unit, Microbiology Services Colindale, Public Health England | Respiratory Virus Unit, Microbiology Services Colindale, Public Health England |
| EPI_ISL_423423 | Respiratory Virus Unit, Microbiology Services Colindale, Public Health England | Respiratory Virus Unit, Microbiology Services Colindale, Public Health England |
| EPI_ISL_423424 | Respiratory Virus Unit, Microbiology Services Colindale, Public Health England | Respiratory Virus Unit, Microbiology Services Colindale, Public Health England |
| EPI_ISL_423425 | Respiratory Virus Unit, Microbiology Services Colindale, Public Health England | Respiratory Virus Unit, Microbiology Services Colindale, Public Health England |
| EPI_ISL_423426 | Respiratory Virus Unit, Microbiology Services Colindale, Public Health England | Respiratory Virus Unit, Microbiology Services Colindale, Public Health England |
| EPI_ISL_423427 | Respiratory Virus Unit, Microbiology Services Colindale, Public Health England | Respiratory Virus Unit, Microbiology Services Colindale, Public Health England |
| EPI_ISL_423428 | Respiratory Virus Unit, Microbiology Services Colindale, Public Health England | Respiratory Virus Unit, Microbiology Services Colindale, Public Health England |
| EPI_ISL_423429 | Respiratory Virus Unit, Microbiology Services Colindale, Public Health England | Respiratory Virus Unit, Microbiology Services Colindale, Public Health England |
| EPI_ISL_423430 | Respiratory Virus Unit, Microbiology Services Colindale, Public Health England | Respiratory Virus Unit, Microbiology Services Colindale, Public Health England |

| EPI_Accession_ID | Originating Laboratory | Submitting Laboratory |
|---|---|---|
| EPI_ISL_423431 | Respiratory Virus Unit, Microbiology Services Colindale, Public Health England | Respiratory Virus Unit, Microbiology Services Colindale, Public Health England |
| EPI_ISL_423432 | Respiratory Virus Unit, Microbiology Services Colindale, Public Health England | Respiratory Virus Unit, Microbiology Services Colindale, Public Health England |
| EPI_ISL_423433 | Respiratory Virus Unit, Microbiology Services Colindale, Public Health England | Respiratory Virus Unit, Microbiology Services Colindale, Public Health England |
| EPI_ISL_423434 | Respiratory Virus Unit, Microbiology Services Colindale, Public Health England | Respiratory Virus Unit, Microbiology Services Colindale, Public Health England |
| EPI_ISL_423435 | Respiratory Virus Unit, Microbiology Services Colindale, Public Health England | Respiratory Virus Unit, Microbiology Services Colindale, Public Health England |
| EPI_ISL_423436 | Respiratory Virus Unit, Microbiology Services Colindale, Public Health England | Respiratory Virus Unit, Microbiology Services Colindale, Public Health England |
| EPI_ISL_423437 | Respiratory Virus Unit, Microbiology Services Colindale, Public Health England | Respiratory Virus Unit, Microbiology Services Colindale, Public Health England |
| EPI_ISL_423438 | Respiratory Virus Unit, Microbiology Services Colindale, Public Health England | Respiratory Virus Unit, Microbiology Services Colindale, Public Health England |
| EPI_ISL_423439 | Respiratory Virus Unit, Microbiology Services Colindale, Public Health England | Respiratory Virus Unit, Microbiology Services Colindale, Public Health England |
| EPI_ISL_423440 | Respiratory Virus Unit, Microbiology Services Colindale, Public Health England | Respiratory Virus Unit, Microbiology Services Colindale, Public Health England |
| EPI_ISL_423441 | Respiratory Virus Unit, Microbiology Services Colindale, Public Health England | Respiratory Virus Unit, Microbiology Services Colindale, Public Health England |
| EPI_ISL_423442 | Respiratory Virus Unit, Microbiology Services Colindale, Public Health England | Respiratory Virus Unit, Microbiology Services Colindale, Public Health England |
| EPI_ISL_423443 | Respiratory Virus Unit, Microbiology Services Colindale, Public Health England | Respiratory Virus Unit, Microbiology Services Colindale, Public Health England |
| EPI_ISL_423444 | Respiratory Virus Unit, Microbiology Services Colindale, Public Health England | Respiratory Virus Unit, Microbiology Services Colindale, Public Health England |
| EPI_ISL_423445 | Respiratory Virus Unit, Microbiology Services Colindale, Public Health England | Respiratory Virus Unit, Microbiology Services Colindale, Public Health England |
| EPI_ISL_423446 | Respiratory Virus Unit, Microbiology Services Colindale, Public Health England | Respiratory Virus Unit, Microbiology Services Colindale, Public Health England |

| EPI_Accession_ID | Originating Laboratory | Submitting Laboratory |
|---|---|---|
| EPI_ISL_423447 | Respiratory Virus Unit, Microbiology Services Colindale, Public Health England | Respiratory Virus Unit, Microbiology Services Colindale, Public Health England |
| EPI_ISL_423448 | Respiratory Virus Unit, Microbiology Services Colindale, Public Health England | Respiratory Virus Unit, Microbiology Services Colindale, Public Health England |
| EPI_ISL_423449 | Respiratory Virus Unit, Microbiology Services Colindale, Public Health England | Respiratory Virus Unit, Microbiology Services Colindale, Public Health England |
| EPI_ISL_423450 | Respiratory Virus Unit, Microbiology Services Colindale, Public Health England | Respiratory Virus Unit, Microbiology Services Colindale, Public Health England |
| EPI_ISL_423451 | Respiratory Virus Unit, Microbiology Services Colindale, Public Health England | Respiratory Virus Unit, Microbiology Services Colindale, Public Health England |
| EPI_ISL_423452 | Respiratory Virus Unit, Microbiology Services Colindale, Public Health England | Respiratory Virus Unit, Microbiology Services Colindale, Public Health England |
| EPI_ISL_423453 | Respiratory Virus Unit, Microbiology Services Colindale, Public Health England | Respiratory Virus Unit, Microbiology Services Colindale, Public Health England |
| EPI_ISL_423454 | Respiratory Virus Unit, Microbiology Services Colindale, Public Health England | Respiratory Virus Unit, Microbiology Services Colindale, Public Health England |
| EPI_ISL_423455 | Respiratory Virus Unit, Microbiology Services Colindale, Public Health England | Respiratory Virus Unit, Microbiology Services Colindale, Public Health England |
| EPI_ISL_423456 | Respiratory Virus Unit, Microbiology Services Colindale, Public Health England | Respiratory Virus Unit, Microbiology Services Colindale, Public Health England |
| EPI_ISL_423457 | Respiratory Virus Unit, Microbiology Services Colindale, Public Health England | Respiratory Virus Unit, Microbiology Services Colindale, Public Health England |
| EPI_ISL_423458 | Respiratory Virus Unit, Microbiology Services Colindale, Public Health England | Respiratory Virus Unit, Microbiology Services Colindale, Public Health England |
| EPI_ISL_423459 | Respiratory Virus Unit, Microbiology Services Colindale, Public Health England | Respiratory Virus Unit, Microbiology Services Colindale, Public Health England |
| EPI_ISL_423460 | Respiratory Virus Unit, Microbiology Services Colindale, Public Health England | Respiratory Virus Unit, Microbiology Services Colindale, Public Health England |
| EPI_ISL_423461 | Respiratory Virus Unit, Microbiology Services Colindale, Public Health England | Respiratory Virus Unit, Microbiology Services Colindale, Public Health England |
| EPI_ISL_423462 | Respiratory Virus Unit, Microbiology Services Colindale, Public Health England | Respiratory Virus Unit, Microbiology Services Colindale, Public Health England |

| EPI_Accession_ID | Originating Laboratory | Submitting Laboratory |
|---|---|---|
| EPI_ISL_423463 | Respiratory Virus Unit, Microbiology Services Colindale, Public Health England | Respiratory Virus Unit, Microbiology Services Colindale, Public Health England |
| EPI_ISL_423465 | Respiratory Virus Unit, Microbiology Services Colindale, Public Health England | Respiratory Virus Unit, Microbiology Services Colindale, Public Health England |
| EPI_ISL_423466 | Respiratory Virus Unit, Microbiology Services Colindale, Public Health England | Respiratory Virus Unit, Microbiology Services Colindale, Public Health England |
| EPI_ISL_423467 | Respiratory Virus Unit, Microbiology Services Colindale, Public Health England | Respiratory Virus Unit, Microbiology Services Colindale, Public Health England |
| EPI_ISL_423468 | Respiratory Virus Unit, Microbiology Services Colindale, Public Health England | Respiratory Virus Unit, Microbiology Services Colindale, Public Health England |
| EPI_ISL_423469 | Respiratory Virus Unit, Microbiology Services Colindale, Public Health England | Respiratory Virus Unit, Microbiology Services Colindale, Public Health England |
| EPI_ISL_423470 | Respiratory Virus Unit, Microbiology Services Colindale, Public Health England | Respiratory Virus Unit, Microbiology Services Colindale, Public Health England |
| EPI_ISL_423471 | Respiratory Virus Unit, Microbiology Services Colindale, Public Health England | Respiratory Virus Unit, Microbiology Services Colindale, Public Health England |
| EPI_ISL_423472 | Respiratory Virus Unit, Microbiology Services Colindale, Public Health England | Respiratory Virus Unit, Microbiology Services Colindale, Public Health England |
| EPI_ISL_423473 | Respiratory Virus Unit, Microbiology Services Colindale, Public Health England | Respiratory Virus Unit, Microbiology Services Colindale, Public Health England |
| EPI_ISL_423474 | Respiratory Virus Unit, Microbiology Services Colindale, Public Health England | Respiratory Virus Unit, Microbiology Services Colindale, Public Health England |
| EPI_ISL_423475 | Respiratory Virus Unit, Microbiology Services Colindale, Public Health England | Respiratory Virus Unit, Microbiology Services Colindale, Public Health England |
| EPI_ISL_423476 | Respiratory Virus Unit, Microbiology Services Colindale, Public Health England | Respiratory Virus Unit, Microbiology Services Colindale, Public Health England |
| EPI_ISL_423477 | Respiratory Virus Unit, Microbiology Services Colindale, Public Health England | Respiratory Virus Unit, Microbiology Services Colindale, Public Health England |
| EPI_ISL_423478 | Respiratory Virus Unit, Microbiology Services Colindale, Public Health England | Respiratory Virus Unit, Microbiology Services Colindale, Public Health England |
| EPI_ISL_423479 | Respiratory Virus Unit, Microbiology Services Colindale, Public Health England | Respiratory Virus Unit, Microbiology Services Colindale, Public Health England |

| EPI_Accession_ID | Originating Laboratory | Submitting Laboratory |
|---|---|---|
| EPI_ISL_423480 | Respiratory Virus Unit, Microbiology Services Colindale, Public Health England | Respiratory Virus Unit, Microbiology Services Colindale, Public Health England |
| EPI_ISL_423481 | Respiratory Virus Unit, Microbiology Services Colindale, Public Health England | Respiratory Virus Unit, Microbiology Services Colindale, Public Health England |
| EPI_ISL_423482 | Respiratory Virus Unit, Microbiology Services Colindale, Public Health England | Respiratory Virus Unit, Microbiology Services Colindale, Public Health England |
| EPI_ISL_423483 | Respiratory Virus Unit, Microbiology Services Colindale, Public Health England | Respiratory Virus Unit, Microbiology Services Colindale, Public Health England |
| EPI_ISL_423484 | Respiratory Virus Unit, Microbiology Services Colindale, Public Health England | Respiratory Virus Unit, Microbiology Services Colindale, Public Health England |
| EPI_ISL_423485 | Respiratory Virus Unit, Microbiology Services Colindale, Public Health England | Respiratory Virus Unit, Microbiology Services Colindale, Public Health England |
| EPI_ISL_423486 | Respiratory Virus Unit, Microbiology Services Colindale, Public Health England | Respiratory Virus Unit, Microbiology Services Colindale, Public Health England |
| EPI_ISL_423487 | Respiratory Virus Unit, Microbiology Services Colindale, Public Health England | Respiratory Virus Unit, Microbiology Services Colindale, Public Health England |
| EPI_ISL_423488 | Respiratory Virus Unit, Microbiology Services Colindale, Public Health England | Respiratory Virus Unit, Microbiology Services Colindale, Public Health England |
| EPI_ISL_423489 | Respiratory Virus Unit, Microbiology Services Colindale, Public Health England | Respiratory Virus Unit, Microbiology Services Colindale, Public Health England |
| EPI_ISL_423490 | Respiratory Virus Unit, Microbiology Services Colindale, Public Health England | Respiratory Virus Unit, Microbiology Services Colindale, Public Health England |
| EPI_ISL_423491 | Respiratory Virus Unit, Microbiology Services Colindale, Public Health England | Respiratory Virus Unit, Microbiology Services Colindale, Public Health England |
| EPI_ISL_423492 | Respiratory Virus Unit, Microbiology Services Colindale, Public Health England | Respiratory Virus Unit, Microbiology Services Colindale, Public Health England |
| EPI_ISL_423493 | Respiratory Virus Unit, Microbiology Services Colindale, Public Health England | Respiratory Virus Unit, Microbiology Services Colindale, Public Health England |
| EPI_ISL_423494 | Respiratory Virus Unit, Microbiology Services Colindale, Public Health England | Respiratory Virus Unit, Microbiology Services Colindale, Public Health England |
| EPI_ISL_423495 | Respiratory Virus Unit, Microbiology Services Colindale, Public Health England | Respiratory Virus Unit, Microbiology Services Colindale, Public Health England |

| EPI_Accession_ID | Originating Laboratory | Submitting Laboratory |
|---|---|---|
| EPI_ISL_423496 | Respiratory Virus Unit, Microbiology Services Colindale, Public Health England | Respiratory Virus Unit, Microbiology Services Colindale, Public Health England |
| EPI_ISL_423497 | Respiratory Virus Unit, Microbiology Services Colindale, Public Health England | Respiratory Virus Unit, Microbiology Services Colindale, Public Health England |
| EPI_ISL_423498 | Respiratory Virus Unit, Microbiology Services Colindale, Public Health England | Respiratory Virus Unit, Microbiology Services Colindale, Public Health England |
| EPI_ISL_423499 | Respiratory Virus Unit, Microbiology Services Colindale, Public Health England | Respiratory Virus Unit, Microbiology Services Colindale, Public Health England |
| EPI_ISL_423500 | Respiratory Virus Unit, Microbiology Services Colindale, Public Health England | Respiratory Virus Unit, Microbiology Services Colindale, Public Health England |
| EPI_ISL_423501 | Respiratory Virus Unit, Microbiology Services Colindale, Public Health England | Respiratory Virus Unit, Microbiology Services Colindale, Public Health England |
| EPI_ISL_423502 | Respiratory Virus Unit, Microbiology Services Colindale, Public Health England | Respiratory Virus Unit, Microbiology Services Colindale, Public Health England |
| EPI_ISL_423503 | Respiratory Virus Unit, Microbiology Services Colindale, Public Health England | Respiratory Virus Unit, Microbiology Services Colindale, Public Health England |
| EPI_ISL_423504 | Respiratory Virus Unit, Microbiology Services Colindale, Public Health England | Respiratory Virus Unit, Microbiology Services Colindale, Public Health England |
| EPI_ISL_423505 | Respiratory Virus Unit, Microbiology Services Colindale, Public Health England | Respiratory Virus Unit, Microbiology Services Colindale, Public Health England |
| EPI_ISL_423506 | Respiratory Virus Unit, Microbiology Services Colindale, Public Health England | Respiratory Virus Unit, Microbiology Services Colindale, Public Health England |
| EPI_ISL_423507 | Respiratory Virus Unit, Microbiology Services Colindale, Public Health England | Respiratory Virus Unit, Microbiology Services Colindale, Public Health England |
| EPI_ISL_423508 | Respiratory Virus Unit, Microbiology Services Colindale, Public Health England | Respiratory Virus Unit, Microbiology Services Colindale, Public Health England |
| EPI_ISL_423509 | Respiratory Virus Unit, Microbiology Services Colindale, Public Health England | Respiratory Virus Unit, Microbiology Services Colindale, Public Health England |
| EPI_ISL_423510 | Respiratory Virus Unit, Microbiology Services Colindale, Public Health England | Respiratory Virus Unit, Microbiology Services Colindale, Public Health England |
| EPI_ISL_423511 | Respiratory Virus Unit, Microbiology Services Colindale, Public Health England | Respiratory Virus Unit, Microbiology Services Colindale, Public Health England |

| EPI_Accession_ID | Originating Laboratory | Submitting Laboratory |
| --- | --- | --- |
| EPI_ISL_423512 | Respiratory Virus Unit, Microbiology Services Colindale, Public Health England | Respiratory Virus Unit, Microbiology Services Colindale, Public Health England |
| EPI_ISL_423513 | Respiratory Virus Unit, Microbiology Services Colindale, Public Health England | Respiratory Virus Unit, Microbiology Services Colindale, Public Health England |
| EPI_ISL_423514 | Respiratory Virus Unit, Microbiology Services Colindale, Public Health England | Respiratory Virus Unit, Microbiology Services Colindale, Public Health England |
| EPI_ISL_423515 | Respiratory Virus Unit, Microbiology Services Colindale, Public Health England | Respiratory Virus Unit, Microbiology Services Colindale, Public Health England |
| EPI_ISL_423516 | Respiratory Virus Unit, Microbiology Services Colindale, Public Health England | Respiratory Virus Unit, Microbiology Services Colindale, Public Health England |
| EPI_ISL_423517 | Respiratory Virus Unit, Microbiology Services Colindale, Public Health England | Respiratory Virus Unit, Microbiology Services Colindale, Public Health England |
| EPI_ISL_423518 | Respiratory Virus Unit, Microbiology Services Colindale, Public Health England | Respiratory Virus Unit, Microbiology Services Colindale, Public Health England |
| EPI_ISL_423519 | Respiratory Virus Unit, Microbiology Services Colindale, Public Health England | Respiratory Virus Unit, Microbiology Services Colindale, Public Health England |
| EPI_ISL_423520 | Respiratory Virus Unit, Microbiology Services Colindale, Public Health England | Respiratory Virus Unit, Microbiology Services Colindale, Public Health England |
| EPI_ISL_423521 | Respiratory Virus Unit, Microbiology Services Colindale, Public Health England | Respiratory Virus Unit, Microbiology Services Colindale, Public Health England |
| EPI_ISL_423522 | Respiratory Virus Unit, Microbiology Services Colindale, Public Health England | Respiratory Virus Unit, Microbiology Services Colindale, Public Health England |
| EPI_ISL_423523 | Respiratory Virus Unit, Microbiology Services Colindale, Public Health England | Respiratory Virus Unit, Microbiology Services Colindale, Public Health England |
| EPI_ISL_423524 | Respiratory Virus Unit, Microbiology Services Colindale, Public Health England | Respiratory Virus Unit, Microbiology Services Colindale, Public Health England |
| EPI_ISL_423526 | Respiratory Virus Unit, Microbiology Services Colindale, Public Health England | Respiratory Virus Unit, Microbiology Services Colindale, Public Health England |
| EPI_ISL_423527 | Respiratory Virus Unit, Microbiology Services Colindale, Public Health England | Respiratory Virus Unit, Microbiology Services Colindale, Public Health England |
| EPI_ISL_423528 | Respiratory Virus Unit, Microbiology Services Colindale, Public Health England | Respiratory Virus Unit, Microbiology Services Colindale, Public Health England |

| EPI_Accession_ID | Originating Laboratory | Submitting Laboratory |
|---|---|---|
| EPI_ISL_423529 | Respiratory Virus Unit, Microbiology Services Colindale, Public Health England | Respiratory Virus Unit, Microbiology Services Colindale, Public Health England |
| EPI_ISL_423531 | Respiratory Virus Unit, Microbiology Services Colindale, Public Health England | Respiratory Virus Unit, Microbiology Services Colindale, Public Health England |
| EPI_ISL_423532 | Respiratory Virus Unit, Microbiology Services Colindale, Public Health England | Respiratory Virus Unit, Microbiology Services Colindale, Public Health England |
| EPI_ISL_423533 | Respiratory Virus Unit, Microbiology Services Colindale, Public Health England | Respiratory Virus Unit, Microbiology Services Colindale, Public Health England |
| EPI_ISL_423534 | Respiratory Virus Unit, Microbiology Services Colindale, Public Health England | Respiratory Virus Unit, Microbiology Services Colindale, Public Health England |
| EPI_ISL_423535 | Respiratory Virus Unit, Microbiology Services Colindale, Public Health England | Respiratory Virus Unit, Microbiology Services Colindale, Public Health England |
| EPI_ISL_423536 | Respiratory Virus Unit, Microbiology Services Colindale, Public Health England | Respiratory Virus Unit, Microbiology Services Colindale, Public Health England |
| EPI_ISL_423537 | Respiratory Virus Unit, Microbiology Services Colindale, Public Health England | Respiratory Virus Unit, Microbiology Services Colindale, Public Health England |
| EPI_ISL_423538 | Respiratory Virus Unit, Microbiology Services Colindale, Public Health England | Respiratory Virus Unit, Microbiology Services Colindale, Public Health England |
| EPI_ISL_423539 | Respiratory Virus Unit, Microbiology Services Colindale, Public Health England | Respiratory Virus Unit, Microbiology Services Colindale, Public Health England |
| EPI_ISL_423541 | Respiratory Virus Unit, Microbiology Services Colindale, Public Health England | Respiratory Virus Unit, Microbiology Services Colindale, Public Health England |
| EPI_ISL_423543 | Respiratory Virus Unit, Microbiology Services Colindale, Public Health England | Respiratory Virus Unit, Microbiology Services Colindale, Public Health England |
| EPI_ISL_423544 | Respiratory Virus Unit, Microbiology Services Colindale, Public Health England | Respiratory Virus Unit, Microbiology Services Colindale, Public Health England |
| EPI_ISL_423545 | Respiratory Virus Unit, Microbiology Services Colindale, Public Health England | Respiratory Virus Unit, Microbiology Services Colindale, Public Health England |
| EPI_ISL_423546 | Respiratory Virus Unit, Microbiology Services Colindale, Public Health England | Respiratory Virus Unit, Microbiology Services Colindale, Public Health England |
| EPI_ISL_423548 | Respiratory Virus Unit, Microbiology Services Colindale, Public Health England | Respiratory Virus Unit, Microbiology Services Colindale, Public Health England |

| EPI_Accession_ID | Originating Laboratory | Submitting Laboratory |
|---|---|---|
| EPI_ISL_423549 | Respiratory Virus Unit, Microbiology Services Colindale, Public Health England | Respiratory Virus Unit, Microbiology Services Colindale, Public Health England |
| EPI_ISL_423550 | Respiratory Virus Unit, Microbiology Services Colindale, Public Health England | Respiratory Virus Unit, Microbiology Services Colindale, Public Health England |
| EPI_ISL_423551 | Respiratory Virus Unit, Microbiology Services Colindale, Public Health England | Respiratory Virus Unit, Microbiology Services Colindale, Public Health England |
| EPI_ISL_423552 | Respiratory Virus Unit, Microbiology Services Colindale, Public Health England | Respiratory Virus Unit, Microbiology Services Colindale, Public Health England |
| EPI_ISL_423553 | Respiratory Virus Unit, Microbiology Services Colindale, Public Health England | Respiratory Virus Unit, Microbiology Services Colindale, Public Health England |
| EPI_ISL_423554 | Respiratory Virus Unit, Microbiology Services Colindale, Public Health England | Respiratory Virus Unit, Microbiology Services Colindale, Public Health England |
| EPI_ISL_423555 | Respiratory Virus Unit, Microbiology Services Colindale, Public Health England | Respiratory Virus Unit, Microbiology Services Colindale, Public Health England |
| EPI_ISL_423556 | Respiratory Virus Unit, Microbiology Services Colindale, Public Health England | Respiratory Virus Unit, Microbiology Services Colindale, Public Health England |
| EPI_ISL_423557 | Respiratory Virus Unit, Microbiology Services Colindale, Public Health England | Respiratory Virus Unit, Microbiology Services Colindale, Public Health England |
| EPI_ISL_423558 | Respiratory Virus Unit, Microbiology Services Colindale, Public Health England | Respiratory Virus Unit, Microbiology Services Colindale, Public Health England |
| EPI_ISL_423559 | Respiratory Virus Unit, Microbiology Services Colindale, Public Health England | Respiratory Virus Unit, Microbiology Services Colindale, Public Health England |
| EPI_ISL_423560 | Respiratory Virus Unit, Microbiology Services Colindale, Public Health England | Respiratory Virus Unit, Microbiology Services Colindale, Public Health England |
| EPI_ISL_423561 | Respiratory Virus Unit, Microbiology Services Colindale, Public Health England | Respiratory Virus Unit, Microbiology Services Colindale, Public Health England |
| EPI_ISL_423562 | Respiratory Virus Unit, Microbiology Services Colindale, Public Health England | Respiratory Virus Unit, Microbiology Services Colindale, Public Health England |
| EPI_ISL_423563 | Respiratory Virus Unit, Microbiology Services Colindale, Public Health England | Respiratory Virus Unit, Microbiology Services Colindale, Public Health England |
| EPI_ISL_423564 | Respiratory Virus Unit, Microbiology Services Colindale, Public Health England | Respiratory Virus Unit, Microbiology Services Colindale, Public Health England |

| EPI_Accession_ID | Originating Laboratory | Submitting Laboratory |
|---|---|---|
| EPI_ISL_423565 | Respiratory Virus Unit, Microbiology Services Colindale, Public Health England | Respiratory Virus Unit, Microbiology Services Colindale, Public Health England |
| EPI_ISL_423566 | Respiratory Virus Unit, Microbiology Services Colindale, Public Health England | Respiratory Virus Unit, Microbiology Services Colindale, Public Health England |
| EPI_ISL_423567 | Respiratory Virus Unit, Microbiology Services Colindale, Public Health England | Respiratory Virus Unit, Microbiology Services Colindale, Public Health England |
| EPI_ISL_423569 | Respiratory Virus Unit, Microbiology Services Colindale, Public Health England | Respiratory Virus Unit, Microbiology Services Colindale, Public Health England |
| EPI_ISL_423570 | Respiratory Virus Unit, Microbiology Services Colindale, Public Health England | Respiratory Virus Unit, Microbiology Services Colindale, Public Health England |
| EPI_ISL_423571 | Respiratory Virus Unit, Microbiology Services Colindale, Public Health England | Respiratory Virus Unit, Microbiology Services Colindale, Public Health England |
| EPI_ISL_423572 | Respiratory Virus Unit, Microbiology Services Colindale, Public Health England | Respiratory Virus Unit, Microbiology Services Colindale, Public Health England |
| EPI_ISL_423573 | Respiratory Virus Unit, Microbiology Services Colindale, Public Health England | Respiratory Virus Unit, Microbiology Services Colindale, Public Health England |
| EPI_ISL_423574 | Respiratory Virus Unit, Microbiology Services Colindale, Public Health England | Respiratory Virus Unit, Microbiology Services Colindale, Public Health England |
| EPI_ISL_423575 | Respiratory Virus Unit, Microbiology Services Colindale, Public Health England | Respiratory Virus Unit, Microbiology Services Colindale, Public Health England |
| EPI_ISL_423576 | Respiratory Virus Unit, Microbiology Services Colindale, Public Health England | Respiratory Virus Unit, Microbiology Services Colindale, Public Health England |
| EPI_ISL_423577 | Respiratory Virus Unit, Microbiology Services Colindale, Public Health England | Respiratory Virus Unit, Microbiology Services Colindale, Public Health England |
| EPI_ISL_423579 | Respiratory Virus Unit, Microbiology Services Colindale, Public Health England | Respiratory Virus Unit, Microbiology Services Colindale, Public Health England |
| EPI_ISL_423581 | Respiratory Virus Unit, Microbiology Services Colindale, Public Health England | Respiratory Virus Unit, Microbiology Services Colindale, Public Health England |
| EPI_ISL_423582 | Respiratory Virus Unit, Microbiology Services Colindale, Public Health England | Respiratory Virus Unit, Microbiology Services Colindale, Public Health England |
| EPI_ISL_423583 | Respiratory Virus Unit, Microbiology Services Colindale, Public Health England | Respiratory Virus Unit, Microbiology Services Colindale, Public Health England |

| EPI_Accession_ID | Originating Laboratory | Submitting Laboratory |
|---|---|---|
| EPI_ISL_423584 | Respiratory Virus Unit, Microbiology Services Colindale, Public Health England | Respiratory Virus Unit, Microbiology Services Colindale, Public Health England |
| EPI_ISL_423585 | Respiratory Virus Unit, Microbiology Services Colindale, Public Health England | Respiratory Virus Unit, Microbiology Services Colindale, Public Health England |
| EPI_ISL_423586 | Respiratory Virus Unit, Microbiology Services Colindale, Public Health England | Respiratory Virus Unit, Microbiology Services Colindale, Public Health England |
| EPI_ISL_423587 | Respiratory Virus Unit, Microbiology Services Colindale, Public Health England | Respiratory Virus Unit, Microbiology Services Colindale, Public Health England |
| EPI_ISL_423588 | Respiratory Virus Unit, Microbiology Services Colindale, Public Health England | Respiratory Virus Unit, Microbiology Services Colindale, Public Health England |
| EPI_ISL_423589 | Respiratory Virus Unit, Microbiology Services Colindale, Public Health England | Respiratory Virus Unit, Microbiology Services Colindale, Public Health England |
| EPI_ISL_423590 | Respiratory Virus Unit, Microbiology Services Colindale, Public Health England | Respiratory Virus Unit, Microbiology Services Colindale, Public Health England |
| EPI_ISL_423591 | Respiratory Virus Unit, Microbiology Services Colindale, Public Health England | Respiratory Virus Unit, Microbiology Services Colindale, Public Health England |
| EPI_ISL_423592 | Respiratory Virus Unit, Microbiology Services Colindale, Public Health England | Respiratory Virus Unit, Microbiology Services Colindale, Public Health England |
| EPI_ISL_423593 | Respiratory Virus Unit, Microbiology Services Colindale, Public Health England | Respiratory Virus Unit, Microbiology Services Colindale, Public Health England |
| EPI_ISL_423594 | Respiratory Virus Unit, Microbiology Services Colindale, Public Health England | Respiratory Virus Unit, Microbiology Services Colindale, Public Health England |
| EPI_ISL_423595 | Respiratory Virus Unit, Microbiology Services Colindale, Public Health England | Respiratory Virus Unit, Microbiology Services Colindale, Public Health England |
| EPI_ISL_423596 | Respiratory Virus Unit, Microbiology Services Colindale, Public Health England | Respiratory Virus Unit, Microbiology Services Colindale, Public Health England |
| EPI_ISL_423597 | Respiratory Virus Unit, Microbiology Services Colindale, Public Health England | Respiratory Virus Unit, Microbiology Services Colindale, Public Health England |
| EPI_ISL_423598 | Respiratory Virus Unit, Microbiology Services Colindale, Public Health England | Respiratory Virus Unit, Microbiology Services Colindale, Public Health England |
| EPI_ISL_423599 | Respiratory Virus Unit, Microbiology Services Colindale, Public Health England | Respiratory Virus Unit, Microbiology Services Colindale, Public Health England |

| EPI_Accession_ID | Originating Laboratory | Submitting Laboratory |
| --- | --- | --- |
| EPI_ISL_423600 | Respiratory Virus Unit, Microbiology Services Colindale, Public Health England | Respiratory Virus Unit, Microbiology Services Colindale, Public Health England |
| EPI_ISL_423601 | Respiratory Virus Unit, Microbiology Services Colindale, Public Health England | Respiratory Virus Unit, Microbiology Services Colindale, Public Health England |
| EPI_ISL_423602 | Respiratory Virus Unit, Microbiology Services Colindale, Public Health England | Respiratory Virus Unit, Microbiology Services Colindale, Public Health England |
| EPI_ISL_423603 | Respiratory Virus Unit, Microbiology Services Colindale, Public Health England | Respiratory Virus Unit, Microbiology Services Colindale, Public Health England |
| EPI_ISL_423604 | Respiratory Virus Unit, Microbiology Services Colindale, Public Health England | Respiratory Virus Unit, Microbiology Services Colindale, Public Health England |
| EPI_ISL_423605 | Respiratory Virus Unit, Microbiology Services Colindale, Public Health England | Respiratory Virus Unit, Microbiology Services Colindale, Public Health England |
| EPI_ISL_423606 | Respiratory Virus Unit, Microbiology Services Colindale, Public Health England | Respiratory Virus Unit, Microbiology Services Colindale, Public Health England |
| EPI_ISL_423607 | Respiratory Virus Unit, Microbiology Services Colindale, Public Health England | Respiratory Virus Unit, Microbiology Services Colindale, Public Health England |
| EPI_ISL_423608 | Respiratory Virus Unit, Microbiology Services Colindale, Public Health England | Respiratory Virus Unit, Microbiology Services Colindale, Public Health England |
| EPI_ISL_423609 | Respiratory Virus Unit, Microbiology Services Colindale, Public Health England | Respiratory Virus Unit, Microbiology Services Colindale, Public Health England |
| EPI_ISL_423611 | Respiratory Virus Unit, Microbiology Services Colindale, Public Health England | Respiratory Virus Unit, Microbiology Services Colindale, Public Health England |
| EPI_ISL_423612 | Respiratory Virus Unit, Microbiology Services Colindale, Public Health England | Respiratory Virus Unit, Microbiology Services Colindale, Public Health England |
| EPI_ISL_423613 | Respiratory Virus Unit, Microbiology Services Colindale, Public Health England | Respiratory Virus Unit, Microbiology Services Colindale, Public Health England |
| EPI_ISL_423614 | Respiratory Virus Unit, Microbiology Services Colindale, Public Health England | Respiratory Virus Unit, Microbiology Services Colindale, Public Health England |
| EPI_ISL_423615 | Respiratory Virus Unit, Microbiology Services Colindale, Public Health England | Respiratory Virus Unit, Microbiology Services Colindale, Public Health England |
| EPI_ISL_423616 | Respiratory Virus Unit, Microbiology Services Colindale, Public Health England | Respiratory Virus Unit, Microbiology Services Colindale, Public Health England |

| EPI_Accession_ID | Originating Laboratory | Submitting Laboratory |
|---|---|---|
| EPI_ISL_423617 | Respiratory Virus Unit, Microbiology Services Colindale, Public Health England | Respiratory Virus Unit, Microbiology Services Colindale, Public Health England |
| EPI_ISL_423618 | Respiratory Virus Unit, Microbiology Services Colindale, Public Health England | Respiratory Virus Unit, Microbiology Services Colindale, Public Health England |
| EPI_ISL_423619 | Respiratory Virus Unit, Microbiology Services Colindale, Public Health England | Respiratory Virus Unit, Microbiology Services Colindale, Public Health England |
| EPI_ISL_423620 | Respiratory Virus Unit, Microbiology Services Colindale, Public Health England | Respiratory Virus Unit, Microbiology Services Colindale, Public Health England |
| EPI_ISL_423621 | Respiratory Virus Unit, Microbiology Services Colindale, Public Health England | Respiratory Virus Unit, Microbiology Services Colindale, Public Health England |
| EPI_ISL_423622 | Respiratory Virus Unit, Microbiology Services Colindale, Public Health England | Respiratory Virus Unit, Microbiology Services Colindale, Public Health England |
| EPI_ISL_423623 | Respiratory Virus Unit, Microbiology Services Colindale, Public Health England | Respiratory Virus Unit, Microbiology Services Colindale, Public Health England |
| EPI_ISL_423624 | Respiratory Virus Unit, Microbiology Services Colindale, Public Health England | Respiratory Virus Unit, Microbiology Services Colindale, Public Health England |
| EPI_ISL_423625 | Respiratory Virus Unit, Microbiology Services Colindale, Public Health England | Respiratory Virus Unit, Microbiology Services Colindale, Public Health England |
| EPI_ISL_423626 | Respiratory Virus Unit, Microbiology Services Colindale, Public Health England | Respiratory Virus Unit, Microbiology Services Colindale, Public Health England |
| EPI_ISL_423627 | Respiratory Virus Unit, Microbiology Services Colindale, Public Health England | Respiratory Virus Unit, Microbiology Services Colindale, Public Health England |
| EPI_ISL_423628 | Respiratory Virus Unit, Microbiology Services Colindale, Public Health England | Respiratory Virus Unit, Microbiology Services Colindale, Public Health England |
| EPI_ISL_423629 | Respiratory Virus Unit, Microbiology Services Colindale, Public Health England | Respiratory Virus Unit, Microbiology Services Colindale, Public Health England |
| EPI_ISL_423630 | Respiratory Virus Unit, Microbiology Services Colindale, Public Health England | Respiratory Virus Unit, Microbiology Services Colindale, Public Health England |
| EPI_ISL_423631 | Respiratory Virus Unit, Microbiology Services Colindale, Public Health England | Respiratory Virus Unit, Microbiology Services Colindale, Public Health England |
| EPI_ISL_423632 | Respiratory Virus Unit, Microbiology Services Colindale, Public Health England | Respiratory Virus Unit, Microbiology Services Colindale, Public Health England |

| EPI_Accession_ID | Originating Laboratory | Submitting Laboratory |
|---|---|---|
| EPI_ISL_423634 | Respiratory Virus Unit, Microbiology Services Colindale, Public Health England | Respiratory Virus Unit, Microbiology Services Colindale, Public Health England |
| EPI_ISL_423635 | Respiratory Virus Unit, Microbiology Services Colindale, Public Health England | Respiratory Virus Unit, Microbiology Services Colindale, Public Health England |
| EPI_ISL_423636 | Respiratory Virus Unit, Microbiology Services Colindale, Public Health England | Respiratory Virus Unit, Microbiology Services Colindale, Public Health England |
| EPI_ISL_423637 | Respiratory Virus Unit, Microbiology Services Colindale, Public Health England | Respiratory Virus Unit, Microbiology Services Colindale, Public Health England |
| EPI_ISL_423638 | Respiratory Virus Unit, Microbiology Services Colindale, Public Health England | Respiratory Virus Unit, Microbiology Services Colindale, Public Health England |
| EPI_ISL_423639 | Respiratory Virus Unit, Microbiology Services Colindale, Public Health England | Respiratory Virus Unit, Microbiology Services Colindale, Public Health England |
| EPI_ISL_423641 | Respiratory Virus Unit, Microbiology Services Colindale, Public Health England | Respiratory Virus Unit, Microbiology Services Colindale, Public Health England |
| EPI_ISL_423643 | Respiratory Virus Unit, Microbiology Services Colindale, Public Health England | Respiratory Virus Unit, Microbiology Services Colindale, Public Health England |
| EPI_ISL_423644 | Respiratory Virus Unit, Microbiology Services Colindale, Public Health England | Respiratory Virus Unit, Microbiology Services Colindale, Public Health England |
| EPI_ISL_423645 | Respiratory Virus Unit, Microbiology Services Colindale, Public Health England | Respiratory Virus Unit, Microbiology Services Colindale, Public Health England |
| EPI_ISL_423646 | Respiratory Virus Unit, Microbiology Services Colindale, Public Health England | Respiratory Virus Unit, Microbiology Services Colindale, Public Health England |
| EPI_ISL_423647 | Respiratory Virus Unit, Microbiology Services Colindale, Public Health England | Respiratory Virus Unit, Microbiology Services Colindale, Public Health England |
| EPI_ISL_423648 | Respiratory Virus Unit, Microbiology Services Colindale, Public Health England | Respiratory Virus Unit, Microbiology Services Colindale, Public Health England |
| EPI_ISL_423649 | Respiratory Virus Unit, Microbiology Services Colindale, Public Health England | Respiratory Virus Unit, Microbiology Services Colindale, Public Health England |
| EPI_ISL_423650 | Respiratory Virus Unit, Microbiology Services Colindale, Public Health England | Respiratory Virus Unit, Microbiology Services Colindale, Public Health England |
| EPI_ISL_423651 | Respiratory Virus Unit, Microbiology Services Colindale, Public Health England | Respiratory Virus Unit, Microbiology Services Colindale, Public Health England |

| EPI_Accession_ID | Originating Laboratory | Submitting Laboratory |
|---|---|---|
| EPI_ISL_423652 | Respiratory Virus Unit, Microbiology Services Colindale, Public Health England | Respiratory Virus Unit, Microbiology Services Colindale, Public Health England |
| EPI_ISL_423653 | Respiratory Virus Unit, Microbiology Services Colindale, Public Health England | Respiratory Virus Unit, Microbiology Services Colindale, Public Health England |
| EPI_ISL_423654 | Respiratory Virus Unit, Microbiology Services Colindale, Public Health England | Respiratory Virus Unit, Microbiology Services Colindale, Public Health England |
| EPI_ISL_423655 | Respiratory Virus Unit, Microbiology Services Colindale, Public Health England | Respiratory Virus Unit, Microbiology Services Colindale, Public Health England |
| EPI_ISL_423656 | Respiratory Virus Unit, Microbiology Services Colindale, Public Health England | Respiratory Virus Unit, Microbiology Services Colindale, Public Health England |
| EPI_ISL_423657 | Respiratory Virus Unit, Microbiology Services Colindale, Public Health England | Respiratory Virus Unit, Microbiology Services Colindale, Public Health England |
| EPI_ISL_423658 | Respiratory Virus Unit, Microbiology Services Colindale, Public Health England | Respiratory Virus Unit, Microbiology Services Colindale, Public Health England |
| EPI_ISL_423659 | Respiratory Virus Unit, Microbiology Services Colindale, Public Health England | Respiratory Virus Unit, Microbiology Services Colindale, Public Health England |
| EPI_ISL_423660 | Respiratory Virus Unit, Microbiology Services Colindale, Public Health England | Respiratory Virus Unit, Microbiology Services Colindale, Public Health England |
| EPI_ISL_423661 | Respiratory Virus Unit, Microbiology Services Colindale, Public Health England | Respiratory Virus Unit, Microbiology Services Colindale, Public Health England |
| EPI_ISL_423662 | Respiratory Virus Unit, Microbiology Services Colindale, Public Health England | Respiratory Virus Unit, Microbiology Services Colindale, Public Health England |
| EPI_ISL_423663 | Respiratory Virus Unit, Microbiology Services Colindale, Public Health England | Respiratory Virus Unit, Microbiology Services Colindale, Public Health England |
| EPI_ISL_423664 | Respiratory Virus Unit, Microbiology Services Colindale, Public Health England | Respiratory Virus Unit, Microbiology Services Colindale, Public Health England |
| EPI_ISL_423665 | Respiratory Virus Unit, Microbiology Services Colindale, Public Health England | Respiratory Virus Unit, Microbiology Services Colindale, Public Health England |
| EPI_ISL_423666 | Respiratory Virus Unit, Microbiology Services Colindale, Public Health England | Respiratory Virus Unit, Microbiology Services Colindale, Public Health England |
| EPI_ISL_423667 | Respiratory Virus Unit, Microbiology Services Colindale, Public Health England | Respiratory Virus Unit, Microbiology Services Colindale, Public Health England |

| EPI_Accession_ID | Originating Laboratory | Submitting Laboratory |
|---|---|---|
| EPI_ISL_423668 | Respiratory Virus Unit, Microbiology Services Colindale, Public Health England | Respiratory Virus Unit, Microbiology Services Colindale, Public Health England |
| EPI_ISL_423669 | Respiratory Virus Unit, Microbiology Services Colindale, Public Health England | Respiratory Virus Unit, Microbiology Services Colindale, Public Health England |
| EPI_ISL_423670 | Respiratory Virus Unit, Microbiology Services Colindale, Public Health England | Respiratory Virus Unit, Microbiology Services Colindale, Public Health England |
| EPI_ISL_423671 | Respiratory Virus Unit, Microbiology Services Colindale, Public Health England | Respiratory Virus Unit, Microbiology Services Colindale, Public Health England |
| EPI_ISL_423672 | Respiratory Virus Unit, Microbiology Services Colindale, Public Health England | Respiratory Virus Unit, Microbiology Services Colindale, Public Health England |
| EPI_ISL_423673 | Respiratory Virus Unit, Microbiology Services Colindale, Public Health England | Respiratory Virus Unit, Microbiology Services Colindale, Public Health England |
| EPI_ISL_423674 | Respiratory Virus Unit, Microbiology Services Colindale, Public Health England | Respiratory Virus Unit, Microbiology Services Colindale, Public Health England |
| EPI_ISL_423675 | Respiratory Virus Unit, Microbiology Services Colindale, Public Health England | Respiratory Virus Unit, Microbiology Services Colindale, Public Health England |
| EPI_ISL_423677 | Respiratory Virus Unit, Microbiology Services Colindale, Public Health England | Respiratory Virus Unit, Microbiology Services Colindale, Public Health England |
| EPI_ISL_423678 | Respiratory Virus Unit, Microbiology Services Colindale, Public Health England | Respiratory Virus Unit, Microbiology Services Colindale, Public Health England |
| EPI_ISL_423679 | Respiratory Virus Unit, Microbiology Services Colindale, Public Health England | Respiratory Virus Unit, Microbiology Services Colindale, Public Health England |
| EPI_ISL_423680 | Respiratory Virus Unit, Microbiology Services Colindale, Public Health England | Respiratory Virus Unit, Microbiology Services Colindale, Public Health England |
| EPI_ISL_423681 | Respiratory Virus Unit, Microbiology Services Colindale, Public Health England | Respiratory Virus Unit, Microbiology Services Colindale, Public Health England |
| EPI_ISL_423682 | Respiratory Virus Unit, Microbiology Services Colindale, Public Health England | Respiratory Virus Unit, Microbiology Services Colindale, Public Health England |
| EPI_ISL_423683 | Respiratory Virus Unit, Microbiology Services Colindale, Public Health England | Respiratory Virus Unit, Microbiology Services Colindale, Public Health England |
| EPI_ISL_423684 | Respiratory Virus Unit, Microbiology Services Colindale, Public Health England | Respiratory Virus Unit, Microbiology Services Colindale, Public Health England |

| EPI_Accession_ID | Originating Laboratory | Submitting Laboratory |
|---|---|---|
| EPI_ISL_423685 | Respiratory Virus Unit, Microbiology Services Colindale, Public Health England | Respiratory Virus Unit, Microbiology Services Colindale, Public Health England |
| EPI_ISL_423686 | Respiratory Virus Unit, Microbiology Services Colindale, Public Health England | Respiratory Virus Unit, Microbiology Services Colindale, Public Health England |
| EPI_ISL_423687 | Respiratory Virus Unit, Microbiology Services Colindale, Public Health England | Respiratory Virus Unit, Microbiology Services Colindale, Public Health England |
| EPI_ISL_423688 | Respiratory Virus Unit, Microbiology Services Colindale, Public Health England | Respiratory Virus Unit, Microbiology Services Colindale, Public Health England |
| EPI_ISL_423689 | Respiratory Virus Unit, Microbiology Services Colindale, Public Health England | Respiratory Virus Unit, Microbiology Services Colindale, Public Health England |
| EPI_ISL_423690 | Respiratory Virus Unit, Microbiology Services Colindale, Public Health England | Respiratory Virus Unit, Microbiology Services Colindale, Public Health England |
| EPI_ISL_423691 | Respiratory Virus Unit, Microbiology Services Colindale, Public Health England | Respiratory Virus Unit, Microbiology Services Colindale, Public Health England |
| EPI_ISL_423692 | Respiratory Virus Unit, Microbiology Services Colindale, Public Health England | Respiratory Virus Unit, Microbiology Services Colindale, Public Health England |
| EPI_ISL_423693 | Respiratory Virus Unit, Microbiology Services Colindale, Public Health England | Respiratory Virus Unit, Microbiology Services Colindale, Public Health England |
| EPI_ISL_423694 | Respiratory Virus Unit, Microbiology Services Colindale, Public Health England | Respiratory Virus Unit, Microbiology Services Colindale, Public Health England |
| EPI_ISL_423695 | Respiratory Virus Unit, Microbiology Services Colindale, Public Health England | Respiratory Virus Unit, Microbiology Services Colindale, Public Health England |
| EPI_ISL_423696 | Respiratory Virus Unit, Microbiology Services Colindale, Public Health England | Respiratory Virus Unit, Microbiology Services Colindale, Public Health England |
| EPI_ISL_423697 | Respiratory Virus Unit, Microbiology Services Colindale, Public Health England | Respiratory Virus Unit, Microbiology Services Colindale, Public Health England |
| EPI_ISL_423698 | Respiratory Virus Unit, Microbiology Services Colindale, Public Health England | Respiratory Virus Unit, Microbiology Services Colindale, Public Health England |
| EPI_ISL_423699 | Respiratory Virus Unit, Microbiology Services Colindale, Public Health England | Respiratory Virus Unit, Microbiology Services Colindale, Public Health England |
| EPI_ISL_423700 | Respiratory Virus Unit, Microbiology Services Colindale, Public Health England | Respiratory Virus Unit, Microbiology Services Colindale, Public Health England |

| EPI_Accession_ID | Originating Laboratory | Submitting Laboratory |
|---|---|---|
| EPI_ISL_423701 | Respiratory Virus Unit, Microbiology Services Colindale, Public Health England | Respiratory Virus Unit, Microbiology Services Colindale, Public Health England |
| EPI_ISL_423702 | Respiratory Virus Unit, Microbiology Services Colindale, Public Health England | Respiratory Virus Unit, Microbiology Services Colindale, Public Health England |
| EPI_ISL_423703 | Respiratory Virus Unit, Microbiology Services Colindale, Public Health England | Respiratory Virus Unit, Microbiology Services Colindale, Public Health England |
| EPI_ISL_423704 | Respiratory Virus Unit, Microbiology Services Colindale, Public Health England | Respiratory Virus Unit, Microbiology Services Colindale, Public Health England |
| EPI_ISL_423705 | Respiratory Virus Unit, Microbiology Services Colindale, Public Health England | Respiratory Virus Unit, Microbiology Services Colindale, Public Health England |
| EPI_ISL_423706 | Respiratory Virus Unit, Microbiology Services Colindale, Public Health England | Respiratory Virus Unit, Microbiology Services Colindale, Public Health England |
| EPI_ISL_423708 | Respiratory Virus Unit, Microbiology Services Colindale, Public Health England | Respiratory Virus Unit, Microbiology Services Colindale, Public Health England |
| EPI_ISL_423709 | Respiratory Virus Unit, Microbiology Services Colindale, Public Health England | Respiratory Virus Unit, Microbiology Services Colindale, Public Health England |
| EPI_ISL_423710 | Respiratory Virus Unit, Microbiology Services Colindale, Public Health England | Respiratory Virus Unit, Microbiology Services Colindale, Public Health England |
| EPI_ISL_423711 | Respiratory Virus Unit, Microbiology Services Colindale, Public Health England | Respiratory Virus Unit, Microbiology Services Colindale, Public Health England |
| EPI_ISL_423712 | Respiratory Virus Unit, Microbiology Services Colindale, Public Health England | Respiratory Virus Unit, Microbiology Services Colindale, Public Health England |
| EPI_ISL_423713 | Respiratory Virus Unit, Microbiology Services Colindale, Public Health England | Respiratory Virus Unit, Microbiology Services Colindale, Public Health England |
| EPI_ISL_423714 | Respiratory Virus Unit, Microbiology Services Colindale, Public Health England | Respiratory Virus Unit, Microbiology Services Colindale, Public Health England |
| EPI_ISL_423715 | Respiratory Virus Unit, Microbiology Services Colindale, Public Health England | Respiratory Virus Unit, Microbiology Services Colindale, Public Health England |
| EPI_ISL_423716 | Respiratory Virus Unit, Microbiology Services Colindale, Public Health England | Respiratory Virus Unit, Microbiology Services Colindale, Public Health England |
| EPI_ISL_423717 | Respiratory Virus Unit, Microbiology Services Colindale, Public Health England | Respiratory Virus Unit, Microbiology Services Colindale, Public Health England |

| EPI_Accession_ID | Originating Laboratory | Submitting Laboratory |
|---|---|---|
| EPI_ISL_423718 | Respiratory Virus Unit, Microbiology Services Colindale, Public Health England | Respiratory Virus Unit, Microbiology Services Colindale, Public Health England |
| EPI_ISL_423719 | Respiratory Virus Unit, Microbiology Services Colindale, Public Health England | Respiratory Virus Unit, Microbiology Services Colindale, Public Health England |
| EPI_ISL_423720 | Respiratory Virus Unit, Microbiology Services Colindale, Public Health England | Respiratory Virus Unit, Microbiology Services Colindale, Public Health England |
| EPI_ISL_423721 | Respiratory Virus Unit, Microbiology Services Colindale, Public Health England | Respiratory Virus Unit, Microbiology Services Colindale, Public Health England |
| EPI_ISL_423722 | Respiratory Virus Unit, Microbiology Services Colindale, Public Health England | Respiratory Virus Unit, Microbiology Services Colindale, Public Health England |
| EPI_ISL_423723 | Respiratory Virus Unit, Microbiology Services Colindale, Public Health England | Respiratory Virus Unit, Microbiology Services Colindale, Public Health England |
| EPI_ISL_423724 | Respiratory Virus Unit, Microbiology Services Colindale, Public Health England | Respiratory Virus Unit, Microbiology Services Colindale, Public Health England |
| EPI_ISL_423725 | Respiratory Virus Unit, Microbiology Services Colindale, Public Health England | Respiratory Virus Unit, Microbiology Services Colindale, Public Health England |
| EPI_ISL_423726 | Respiratory Virus Unit, Microbiology Services Colindale, Public Health England | Respiratory Virus Unit, Microbiology Services Colindale, Public Health England |
| EPI_ISL_423727 | Respiratory Virus Unit, Microbiology Services Colindale, Public Health England | Respiratory Virus Unit, Microbiology Services Colindale, Public Health England |
| EPI_ISL_423728 | Respiratory Virus Unit, Microbiology Services Colindale, Public Health England | Respiratory Virus Unit, Microbiology Services Colindale, Public Health England |
| EPI_ISL_423729 | Respiratory Virus Unit, Microbiology Services Colindale, Public Health England | Respiratory Virus Unit, Microbiology Services Colindale, Public Health England |
| EPI_ISL_423730 | Respiratory Virus Unit, Microbiology Services Colindale, Public Health England | Respiratory Virus Unit, Microbiology Services Colindale, Public Health England |
| EPI_ISL_423731 | Respiratory Virus Unit, Microbiology Services Colindale, Public Health England | Respiratory Virus Unit, Microbiology Services Colindale, Public Health England |
| EPI_ISL_423732 | Respiratory Virus Unit, Microbiology Services Colindale, Public Health England | Respiratory Virus Unit, Microbiology Services Colindale, Public Health England |
| EPI_ISL_423733 | Respiratory Virus Unit, Microbiology Services Colindale, Public Health England | Respiratory Virus Unit, Microbiology Services Colindale, Public Health England |

| EPI_Accession_ID | Originating Laboratory | Submitting Laboratory |
|---|---|---|
| EPI_ISL_423734 | Respiratory Virus Unit, Microbiology Services Colindale, Public Health England | Respiratory Virus Unit, Microbiology Services Colindale, Public Health England |
| EPI_ISL_423735 | Respiratory Virus Unit, Microbiology Services Colindale, Public Health England | Respiratory Virus Unit, Microbiology Services Colindale, Public Health England |
| EPI_ISL_423736 | Respiratory Virus Unit, Microbiology Services Colindale, Public Health England | Respiratory Virus Unit, Microbiology Services Colindale, Public Health England |
| EPI_ISL_423737 | Respiratory Virus Unit, Microbiology Services Colindale, Public Health England | Respiratory Virus Unit, Microbiology Services Colindale, Public Health England |
| EPI_ISL_423738 | Respiratory Virus Unit, Microbiology Services Colindale, Public Health England | Respiratory Virus Unit, Microbiology Services Colindale, Public Health England |
| EPI_ISL_423739 | Respiratory Virus Unit, Microbiology Services Colindale, Public Health England | Respiratory Virus Unit, Microbiology Services Colindale, Public Health England |
| EPI_ISL_423740 | Respiratory Virus Unit, Microbiology Services Colindale, Public Health England | Respiratory Virus Unit, Microbiology Services Colindale, Public Health England |
| EPI_ISL_423741 | Respiratory Virus Unit, Microbiology Services Colindale, Public Health England | Respiratory Virus Unit, Microbiology Services Colindale, Public Health England |
| EPI_ISL_423742 | Respiratory Virus Unit, Microbiology Services Colindale, Public Health England | Respiratory Virus Unit, Microbiology Services Colindale, Public Health England |
| EPI_ISL_423743 | Respiratory Virus Unit, Microbiology Services Colindale, Public Health England | Respiratory Virus Unit, Microbiology Services Colindale, Public Health England |
| EPI_ISL_423744 | Respiratory Virus Unit, Microbiology Services Colindale, Public Health England | Respiratory Virus Unit, Microbiology Services Colindale, Public Health England |
| EPI_ISL_423745 | Respiratory Virus Unit, Microbiology Services Colindale, Public Health England | Respiratory Virus Unit, Microbiology Services Colindale, Public Health England |
| EPI_ISL_423746 | Respiratory Virus Unit, Microbiology Services Colindale, Public Health England | Respiratory Virus Unit, Microbiology Services Colindale, Public Health England |
| EPI_ISL_423747 | Respiratory Virus Unit, Microbiology Services Colindale, Public Health England | Respiratory Virus Unit, Microbiology Services Colindale, Public Health England |
| EPI_ISL_423748 | Respiratory Virus Unit, Microbiology Services Colindale, Public Health England | Respiratory Virus Unit, Microbiology Services Colindale, Public Health England |
| EPI_ISL_423749 | Respiratory Virus Unit, Microbiology Services Colindale, Public Health England | Respiratory Virus Unit, Microbiology Services Colindale, Public Health England |

| EPI_Accession_ID | Originating Laboratory | Submitting Laboratory |
|---|---|---|
| EPI_ISL_423750 | Respiratory Virus Unit, Microbiology Services Colindale, Public Health England | Respiratory Virus Unit, Microbiology Services Colindale, Public Health England |
| EPI_ISL_423751 | Respiratory Virus Unit, Microbiology Services Colindale, Public Health England | Respiratory Virus Unit, Microbiology Services Colindale, Public Health England |
| EPI_ISL_423752 | Respiratory Virus Unit, Microbiology Services Colindale, Public Health England | Respiratory Virus Unit, Microbiology Services Colindale, Public Health England |
| EPI_ISL_423753 | Respiratory Virus Unit, Microbiology Services Colindale, Public Health England | Respiratory Virus Unit, Microbiology Services Colindale, Public Health England |
| EPI_ISL_423754 | Respiratory Virus Unit, Microbiology Services Colindale, Public Health England | Respiratory Virus Unit, Microbiology Services Colindale, Public Health England |
| EPI_ISL_423755 | Respiratory Virus Unit, Microbiology Services Colindale, Public Health England | Respiratory Virus Unit, Microbiology Services Colindale, Public Health England |
| EPI_ISL_423756 | Respiratory Virus Unit, Microbiology Services Colindale, Public Health England | Respiratory Virus Unit, Microbiology Services Colindale, Public Health England |
| EPI_ISL_423757 | Respiratory Virus Unit, Microbiology Services Colindale, Public Health England | Respiratory Virus Unit, Microbiology Services Colindale, Public Health England |
| EPI_ISL_423758 | Respiratory Virus Unit, Microbiology Services Colindale, Public Health England | Respiratory Virus Unit, Microbiology Services Colindale, Public Health England |
| EPI_ISL_423759 | Respiratory Virus Unit, Microbiology Services Colindale, Public Health England | Respiratory Virus Unit, Microbiology Services Colindale, Public Health England |
| EPI_ISL_423760 | Respiratory Virus Unit, Microbiology Services Colindale, Public Health England | Respiratory Virus Unit, Microbiology Services Colindale, Public Health England |
| EPI_ISL_423761 | Respiratory Virus Unit, Microbiology Services Colindale, Public Health England | Respiratory Virus Unit, Microbiology Services Colindale, Public Health England |
| EPI_ISL_423762 | Respiratory Virus Unit, Microbiology Services Colindale, Public Health England | Respiratory Virus Unit, Microbiology Services Colindale, Public Health England |
| EPI_ISL_423763 | Respiratory Virus Unit, Microbiology Services Colindale, Public Health England | Respiratory Virus Unit, Microbiology Services Colindale, Public Health England |
| EPI_ISL_423764 | Respiratory Virus Unit, Microbiology Services Colindale, Public Health England | Respiratory Virus Unit, Microbiology Services Colindale, Public Health England |
| EPI_ISL_423765 | Respiratory Virus Unit, Microbiology Services Colindale, Public Health England | Respiratory Virus Unit, Microbiology Services Colindale, Public Health England |

| EPI_Accession_ID | Originating Laboratory | Submitting Laboratory |
|---|---|---|
| EPI_ISL_423766 | Respiratory Virus Unit, Microbiology Services Colindale, Public Health England | Respiratory Virus Unit, Microbiology Services Colindale, Public Health England |
| EPI_ISL_423767 | Respiratory Virus Unit, Microbiology Services Colindale, Public Health England | Respiratory Virus Unit, Microbiology Services Colindale, Public Health England |
| EPI_ISL_423768 | Respiratory Virus Unit, Microbiology Services Colindale, Public Health England | Respiratory Virus Unit, Microbiology Services Colindale, Public Health England |
| EPI_ISL_423769 | Respiratory Virus Unit, Microbiology Services Colindale, Public Health England | Respiratory Virus Unit, Microbiology Services Colindale, Public Health England |
| EPI_ISL_423770 | Respiratory Virus Unit, Microbiology Services Colindale, Public Health England | Respiratory Virus Unit, Microbiology Services Colindale, Public Health England |
| EPI_ISL_423771 | Respiratory Virus Unit, Microbiology Services Colindale, Public Health England | Respiratory Virus Unit, Microbiology Services Colindale, Public Health England |
| EPI_ISL_423772 | Respiratory Virus Unit, Microbiology Services Colindale, Public Health England | Respiratory Virus Unit, Microbiology Services Colindale, Public Health England |
| EPI_ISL_423773 | Respiratory Virus Unit, Microbiology Services Colindale, Public Health England | Respiratory Virus Unit, Microbiology Services Colindale, Public Health England |
| EPI_ISL_423774 | Respiratory Virus Unit, Microbiology Services Colindale, Public Health England | Respiratory Virus Unit, Microbiology Services Colindale, Public Health England |
| EPI_ISL_423775 | Respiratory Virus Unit, Microbiology Services Colindale, Public Health England | Respiratory Virus Unit, Microbiology Services Colindale, Public Health England |
| EPI_ISL_423776 | Respiratory Virus Unit, Microbiology Services Colindale, Public Health England | Respiratory Virus Unit, Microbiology Services Colindale, Public Health England |
| EPI_ISL_423777 | Respiratory Virus Unit, Microbiology Services Colindale, Public Health England | Respiratory Virus Unit, Microbiology Services Colindale, Public Health England |
| EPI_ISL_423778 | Respiratory Virus Unit, Microbiology Services Colindale, Public Health England | Respiratory Virus Unit, Microbiology Services Colindale, Public Health England |
| EPI_ISL_423779 | Respiratory Virus Unit, Microbiology Services Colindale, Public Health England | Respiratory Virus Unit, Microbiology Services Colindale, Public Health England |
| EPI_ISL_423780 | Respiratory Virus Unit, Microbiology Services Colindale, Public Health England | Respiratory Virus Unit, Microbiology Services Colindale, Public Health England |
| EPI_ISL_423781 | Respiratory Virus Unit, Microbiology Services Colindale, Public Health England | Respiratory Virus Unit, Microbiology Services Colindale, Public Health England |

| EPI_Accession_ID | Originating Laboratory | Submitting Laboratory |
|---|---|---|
| EPI_ISL_423782 | Respiratory Virus Unit, Microbiology Services Colindale, Public Health England | Respiratory Virus Unit, Microbiology Services Colindale, Public Health England |
| EPI_ISL_423783 | Respiratory Virus Unit, Microbiology Services Colindale, Public Health England | Respiratory Virus Unit, Microbiology Services Colindale, Public Health England |
| EPI_ISL_423784 | Respiratory Virus Unit, Microbiology Services Colindale, Public Health England | Respiratory Virus Unit, Microbiology Services Colindale, Public Health England |
| EPI_ISL_423785 | Respiratory Virus Unit, Microbiology Services Colindale, Public Health England | Respiratory Virus Unit, Microbiology Services Colindale, Public Health England |
| EPI_ISL_423786 | Respiratory Virus Unit, Microbiology Services Colindale, Public Health England | Respiratory Virus Unit, Microbiology Services Colindale, Public Health England |
| EPI_ISL_423787 | Respiratory Virus Unit, Microbiology Services Colindale, Public Health England | Respiratory Virus Unit, Microbiology Services Colindale, Public Health England |
| EPI_ISL_423788 | Respiratory Virus Unit, Microbiology Services Colindale, Public Health England | Respiratory Virus Unit, Microbiology Services Colindale, Public Health England |
| EPI_ISL_423789 | Respiratory Virus Unit, Microbiology Services Colindale, Public Health England | Respiratory Virus Unit, Microbiology Services Colindale, Public Health England |
| EPI_ISL_423790 | Respiratory Virus Unit, Microbiology Services Colindale, Public Health England | Respiratory Virus Unit, Microbiology Services Colindale, Public Health England |
| EPI_ISL_423791 | Respiratory Virus Unit, Microbiology Services Colindale, Public Health England | Respiratory Virus Unit, Microbiology Services Colindale, Public Health England |
| EPI_ISL_423792 | Respiratory Virus Unit, Microbiology Services Colindale, Public Health England | Respiratory Virus Unit, Microbiology Services Colindale, Public Health England |
| EPI_ISL_423793 | Respiratory Virus Unit, Microbiology Services Colindale, Public Health England | Respiratory Virus Unit, Microbiology Services Colindale, Public Health England |
| EPI_ISL_423794 | Respiratory Virus Unit, Microbiology Services Colindale, Public Health England | Respiratory Virus Unit, Microbiology Services Colindale, Public Health England |
| EPI_ISL_423795 | Respiratory Virus Unit, Microbiology Services Colindale, Public Health England | Respiratory Virus Unit, Microbiology Services Colindale, Public Health England |
| EPI_ISL_423796 | Respiratory Virus Unit, Microbiology Services Colindale, Public Health England | Respiratory Virus Unit, Microbiology Services Colindale, Public Health England |
| EPI_ISL_423797 | Respiratory Virus Unit, Microbiology Services Colindale, Public Health England | Respiratory Virus Unit, Microbiology Services Colindale, Public Health England |

| EPI_Accession_ID | Originating Laboratory | Submitting Laboratory |
|---|---|---|
| EPI_ISL_423798 | Respiratory Virus Unit, Microbiology Services Colindale, Public Health England | Respiratory Virus Unit, Microbiology Services Colindale, Public Health England |
| EPI_ISL_423799 | Respiratory Virus Unit, Microbiology Services Colindale, Public Health England | Respiratory Virus Unit, Microbiology Services Colindale, Public Health England |
| EPI_ISL_423800 | Respiratory Virus Unit, Microbiology Services Colindale, Public Health England | Respiratory Virus Unit, Microbiology Services Colindale, Public Health England |
| EPI_ISL_423801 | Respiratory Virus Unit, Microbiology Services Colindale, Public Health England | Respiratory Virus Unit, Microbiology Services Colindale, Public Health England |
| EPI_ISL_423802 | Respiratory Virus Unit, Microbiology Services Colindale, Public Health England | Respiratory Virus Unit, Microbiology Services Colindale, Public Health England |
| EPI_ISL_423803 | Respiratory Virus Unit, Microbiology Services Colindale, Public Health England | Respiratory Virus Unit, Microbiology Services Colindale, Public Health England |
| EPI_ISL_423804 | Respiratory Virus Unit, Microbiology Services Colindale, Public Health England | Respiratory Virus Unit, Microbiology Services Colindale, Public Health England |
| EPI_ISL_423805 | Respiratory Virus Unit, Microbiology Services Colindale, Public Health England | Respiratory Virus Unit, Microbiology Services Colindale, Public Health England |
| EPI_ISL_423806 | Respiratory Virus Unit, Microbiology Services Colindale, Public Health England | Respiratory Virus Unit, Microbiology Services Colindale, Public Health England |
| EPI_ISL_423807 | Respiratory Virus Unit, Microbiology Services Colindale, Public Health England | Respiratory Virus Unit, Microbiology Services Colindale, Public Health England |
| EPI_ISL_423808 | Respiratory Virus Unit, Microbiology Services Colindale, Public Health England | Respiratory Virus Unit, Microbiology Services Colindale, Public Health England |
| EPI_ISL_423809 | Respiratory Virus Unit, Microbiology Services Colindale, Public Health England | Respiratory Virus Unit, Microbiology Services Colindale, Public Health England |
| EPI_ISL_423810 | Respiratory Virus Unit, Microbiology Services Colindale, Public Health England | Respiratory Virus Unit, Microbiology Services Colindale, Public Health England |
| EPI_ISL_423811 | Respiratory Virus Unit, Microbiology Services Colindale, Public Health England | Respiratory Virus Unit, Microbiology Services Colindale, Public Health England |
| EPI_ISL_423812 | Respiratory Virus Unit, Microbiology Services Colindale, Public Health England | Respiratory Virus Unit, Microbiology Services Colindale, Public Health England |
| EPI_ISL_423813 | Respiratory Virus Unit, Microbiology Services Colindale, Public Health England | Respiratory Virus Unit, Microbiology Services Colindale, Public Health England |

| EPI_Accession_ID | Originating Laboratory | Submitting Laboratory |
|---|---|---|
| EPI_ISL_423814 | Respiratory Virus Unit, Microbiology Services Colindale, Public Health England | Respiratory Virus Unit, Microbiology Services Colindale, Public Health England |
| EPI_ISL_423816 | Respiratory Virus Unit, Microbiology Services Colindale, Public Health England | Respiratory Virus Unit, Microbiology Services Colindale, Public Health England |
| EPI_ISL_423817 | Respiratory Virus Unit, Microbiology Services Colindale, Public Health England | Respiratory Virus Unit, Microbiology Services Colindale, Public Health England |
| EPI_ISL_423818 | Respiratory Virus Unit, Microbiology Services Colindale, Public Health England | Respiratory Virus Unit, Microbiology Services Colindale, Public Health England |
| EPI_ISL_423819 | Respiratory Virus Unit, Microbiology Services Colindale, Public Health England | Respiratory Virus Unit, Microbiology Services Colindale, Public Health England |
| EPI_ISL_423820 | Respiratory Virus Unit, Microbiology Services Colindale, Public Health England | Respiratory Virus Unit, Microbiology Services Colindale, Public Health England |
| EPI_ISL_423822 | Respiratory Virus Unit, Microbiology Services Colindale, Public Health England | Respiratory Virus Unit, Microbiology Services Colindale, Public Health England |
| EPI_ISL_423823 | Respiratory Virus Unit, Microbiology Services Colindale, Public Health England | Respiratory Virus Unit, Microbiology Services Colindale, Public Health England |
| EPI_ISL_423824 | Respiratory Virus Unit, Microbiology Services Colindale, Public Health England | Respiratory Virus Unit, Microbiology Services Colindale, Public Health England |
| EPI_ISL_423825 | Respiratory Virus Unit, Microbiology Services Colindale, Public Health England | Respiratory Virus Unit, Microbiology Services Colindale, Public Health England |
| EPI_ISL_423826 | Respiratory Virus Unit, Microbiology Services Colindale, Public Health England | Respiratory Virus Unit, Microbiology Services Colindale, Public Health England |
| EPI_ISL_423827 | Respiratory Virus Unit, Microbiology Services Colindale, Public Health England | Respiratory Virus Unit, Microbiology Services Colindale, Public Health England |
| EPI_ISL_423828 | Respiratory Virus Unit, Microbiology Services Colindale, Public Health England | Respiratory Virus Unit, Microbiology Services Colindale, Public Health England |
| EPI_ISL_423829 | Respiratory Virus Unit, Microbiology Services Colindale, Public Health England | Respiratory Virus Unit, Microbiology Services Colindale, Public Health England |
| EPI_ISL_423830 | Respiratory Virus Unit, Microbiology Services Colindale, Public Health England | Respiratory Virus Unit, Microbiology Services Colindale, Public Health England |
| EPI_ISL_423831 | Respiratory Virus Unit, Microbiology Services Colindale, Public Health England | Respiratory Virus Unit, Microbiology Services Colindale, Public Health England |

| EPI_Accession_ID | Originating Laboratory | Submitting Laboratory |
|---|---|---|
| EPI_ISL_423832 | Respiratory Virus Unit, Microbiology Services Colindale, Public Health England | Respiratory Virus Unit, Microbiology Services Colindale, Public Health England |
| EPI_ISL_423833 | Respiratory Virus Unit, Microbiology Services Colindale, Public Health England | Respiratory Virus Unit, Microbiology Services Colindale, Public Health England |
| EPI_ISL_423834 | Respiratory Virus Unit, Microbiology Services Colindale, Public Health England | Respiratory Virus Unit, Microbiology Services Colindale, Public Health England |
| EPI_ISL_423835 | Respiratory Virus Unit, Microbiology Services Colindale, Public Health England | Respiratory Virus Unit, Microbiology Services Colindale, Public Health England |
| EPI_ISL_423836 | Respiratory Virus Unit, Microbiology Services Colindale, Public Health England | Respiratory Virus Unit, Microbiology Services Colindale, Public Health England |
| EPI_ISL_423837 | Respiratory Virus Unit, Microbiology Services Colindale, Public Health England | Respiratory Virus Unit, Microbiology Services Colindale, Public Health England |
| EPI_ISL_423838 | Respiratory Virus Unit, Microbiology Services Colindale, Public Health England | Respiratory Virus Unit, Microbiology Services Colindale, Public Health England |
| EPI_ISL_423839 | Respiratory Virus Unit, Microbiology Services Colindale, Public Health England | Respiratory Virus Unit, Microbiology Services Colindale, Public Health England |
| EPI_ISL_423840 | Respiratory Virus Unit, Microbiology Services Colindale, Public Health England | Respiratory Virus Unit, Microbiology Services Colindale, Public Health England |
| EPI_ISL_423841 | Respiratory Virus Unit, Microbiology Services Colindale, Public Health England | Respiratory Virus Unit, Microbiology Services Colindale, Public Health England |
| EPI_ISL_423842 | Respiratory Virus Unit, Microbiology Services Colindale, Public Health England | Respiratory Virus Unit, Microbiology Services Colindale, Public Health England |
| EPI_ISL_423843 | Respiratory Virus Unit, Microbiology Services Colindale, Public Health England | Respiratory Virus Unit, Microbiology Services Colindale, Public Health England |
| EPI_ISL_423844 | Respiratory Virus Unit, Microbiology Services Colindale, Public Health England | Respiratory Virus Unit, Microbiology Services Colindale, Public Health England |
| EPI_ISL_423845 | Respiratory Virus Unit, Microbiology Services Colindale, Public Health England | Respiratory Virus Unit, Microbiology Services Colindale, Public Health England |
| EPI_ISL_423846 | Respiratory Virus Unit, Microbiology Services Colindale, Public Health England | Respiratory Virus Unit, Microbiology Services Colindale, Public Health England |
| EPI_ISL_423847 | Respiratory Virus Unit, Microbiology Services Colindale, Public Health England | Respiratory Virus Unit, Microbiology Services Colindale, Public Health England |

| EPI_Accession_ID | Originating Laboratory | Submitting Laboratory |
|---|---|---|
| EPI_ISL_423848 | Respiratory Virus Unit, Microbiology Services Colindale, Public Health England | Respiratory Virus Unit, Microbiology Services Colindale, Public Health England |
| EPI_ISL_423849 | Respiratory Virus Unit, Microbiology Services Colindale, Public Health England | Respiratory Virus Unit, Microbiology Services Colindale, Public Health England |
| EPI_ISL_423850 | Respiratory Virus Unit, Microbiology Services Colindale, Public Health England | Respiratory Virus Unit, Microbiology Services Colindale, Public Health England |
| EPI_ISL_423851 | Respiratory Virus Unit, Microbiology Services Colindale, Public Health England | Respiratory Virus Unit, Microbiology Services Colindale, Public Health England |
| EPI_ISL_423852 | Respiratory Virus Unit, Microbiology Services Colindale, Public Health England | Respiratory Virus Unit, Microbiology Services Colindale, Public Health England |
| EPI_ISL_423853 | Respiratory Virus Unit, Microbiology Services Colindale, Public Health England | Respiratory Virus Unit, Microbiology Services Colindale, Public Health England |
| EPI_ISL_423854 | Respiratory Virus Unit, Microbiology Services Colindale, Public Health England | Respiratory Virus Unit, Microbiology Services Colindale, Public Health England |
| EPI_ISL_423855 | Respiratory Virus Unit, Microbiology Services Colindale, Public Health England | Respiratory Virus Unit, Microbiology Services Colindale, Public Health England |
| EPI_ISL_423856 | Respiratory Virus Unit, Microbiology Services Colindale, Public Health England | Respiratory Virus Unit, Microbiology Services Colindale, Public Health England |
| EPI_ISL_423857 | Respiratory Virus Unit, Microbiology Services Colindale, Public Health England | Respiratory Virus Unit, Microbiology Services Colindale, Public Health England |
| EPI_ISL_423858 | Respiratory Virus Unit, Microbiology Services Colindale, Public Health England | Respiratory Virus Unit, Microbiology Services Colindale, Public Health England |
| EPI_ISL_423859 | Respiratory Virus Unit, Microbiology Services Colindale, Public Health England | Respiratory Virus Unit, Microbiology Services Colindale, Public Health England |
| EPI_ISL_423860 | Respiratory Virus Unit, Microbiology Services Colindale, Public Health England | Respiratory Virus Unit, Microbiology Services Colindale, Public Health England |
| EPI_ISL_423861 | Respiratory Virus Unit, Microbiology Services Colindale, Public Health England | Respiratory Virus Unit, Microbiology Services Colindale, Public Health England |
| EPI_ISL_423862 | Respiratory Virus Unit, Microbiology Services Colindale, Public Health England | Respiratory Virus Unit, Microbiology Services Colindale, Public Health England |
| EPI_ISL_423863 | Respiratory Virus Unit, Microbiology Services Colindale, Public Health England | Respiratory Virus Unit, Microbiology Services Colindale, Public Health England |

| EPI_Accession_ID | Originating Laboratory | Submitting Laboratory |
|---|---|---|
| EPI_ISL_423864 | Respiratory Virus Unit, Microbiology Services Colindale, Public Health England | Respiratory Virus Unit, Microbiology Services Colindale, Public Health England |
| EPI_ISL_423865 | Respiratory Virus Unit, Microbiology Services Colindale, Public Health England | Respiratory Virus Unit, Microbiology Services Colindale, Public Health England |
| EPI_ISL_423866 | Respiratory Virus Unit, Microbiology Services Colindale, Public Health England | Respiratory Virus Unit, Microbiology Services Colindale, Public Health England |
| EPI_ISL_423867 | Respiratory Virus Unit, Microbiology Services Colindale, Public Health England | Respiratory Virus Unit, Microbiology Services Colindale, Public Health England |
| EPI_ISL_423868 | Respiratory Virus Unit, Microbiology Services Colindale, Public Health England | Respiratory Virus Unit, Microbiology Services Colindale, Public Health England |
| EPI_ISL_423869 | Respiratory Virus Unit, Microbiology Services Colindale, Public Health England | Respiratory Virus Unit, Microbiology Services Colindale, Public Health England |
| EPI_ISL_423870 | Respiratory Virus Unit, Microbiology Services Colindale, Public Health England | Respiratory Virus Unit, Microbiology Services Colindale, Public Health England |
| EPI_ISL_423871 | Respiratory Virus Unit, Microbiology Services Colindale, Public Health England | Respiratory Virus Unit, Microbiology Services Colindale, Public Health England |
| EPI_ISL_423872 | Respiratory Virus Unit, Microbiology Services Colindale, Public Health England | Respiratory Virus Unit, Microbiology Services Colindale, Public Health England |
| EPI_ISL_423873 | Respiratory Virus Unit, Microbiology Services Colindale, Public Health England | Respiratory Virus Unit, Microbiology Services Colindale, Public Health England |
| EPI_ISL_423874 | Respiratory Virus Unit, Microbiology Services Colindale, Public Health England | Respiratory Virus Unit, Microbiology Services Colindale, Public Health England |
| EPI_ISL_423875 | Respiratory Virus Unit, Microbiology Services Colindale, Public Health England | Respiratory Virus Unit, Microbiology Services Colindale, Public Health England |
| EPI_ISL_423876 | Respiratory Virus Unit, Microbiology Services Colindale, Public Health England | Respiratory Virus Unit, Microbiology Services Colindale, Public Health England |
| EPI_ISL_423877 | Respiratory Virus Unit, Microbiology Services Colindale, Public Health England | Respiratory Virus Unit, Microbiology Services Colindale, Public Health England |
| EPI_ISL_423878 | Respiratory Virus Unit, Microbiology Services Colindale, Public Health England | Respiratory Virus Unit, Microbiology Services Colindale, Public Health England |
| EPI_ISL_423879 | Respiratory Virus Unit, Microbiology Services Colindale, Public Health England | Respiratory Virus Unit, Microbiology Services Colindale, Public Health England |

| EPI_Accession_ID | Originating Laboratory | Submitting Laboratory |
|---|---|---|
| EPI_ISL_423880 | Respiratory Virus Unit, Microbiology Services Colindale, Public Health England | Respiratory Virus Unit, Microbiology Services Colindale, Public Health England |
| EPI_ISL_423881 | Respiratory Virus Unit, Microbiology Services Colindale, Public Health England | Respiratory Virus Unit, Microbiology Services Colindale, Public Health England |
| EPI_ISL_423882 | Respiratory Virus Unit, Microbiology Services Colindale, Public Health England | Respiratory Virus Unit, Microbiology Services Colindale, Public Health England |
| EPI_ISL_423883 | Respiratory Virus Unit, Microbiology Services Colindale, Public Health England | Respiratory Virus Unit, Microbiology Services Colindale, Public Health England |
| EPI_ISL_423884 | Respiratory Virus Unit, Microbiology Services Colindale, Public Health England | Respiratory Virus Unit, Microbiology Services Colindale, Public Health England |
| EPI_ISL_423885 | Respiratory Virus Unit, Microbiology Services Colindale, Public Health England | Respiratory Virus Unit, Microbiology Services Colindale, Public Health England |
| EPI_ISL_423886 | Respiratory Virus Unit, Microbiology Services Colindale, Public Health England | Respiratory Virus Unit, Microbiology Services Colindale, Public Health England |
| EPI_ISL_423887 | Respiratory Virus Unit, Microbiology Services Colindale, Public Health England | Respiratory Virus Unit, Microbiology Services Colindale, Public Health England |
| EPI_ISL_423888 | Respiratory Virus Unit, Microbiology Services Colindale, Public Health England | Respiratory Virus Unit, Microbiology Services Colindale, Public Health England |
| EPI_ISL_423889 | Respiratory Virus Unit, Microbiology Services Colindale, Public Health England | Respiratory Virus Unit, Microbiology Services Colindale, Public Health England |
| EPI_ISL_423890 | Respiratory Virus Unit, Microbiology Services Colindale, Public Health England | Respiratory Virus Unit, Microbiology Services Colindale, Public Health England |
| EPI_ISL_423891 | Respiratory Virus Unit, Microbiology Services Colindale, Public Health England | Respiratory Virus Unit, Microbiology Services Colindale, Public Health England |
| EPI_ISL_423892 | Respiratory Virus Unit, Microbiology Services Colindale, Public Health England | Respiratory Virus Unit, Microbiology Services Colindale, Public Health England |
| EPI_ISL_423893 | Respiratory Virus Unit, Microbiology Services Colindale, Public Health England | Respiratory Virus Unit, Microbiology Services Colindale, Public Health England |
| EPI_ISL_423894 | Respiratory Virus Unit, Microbiology Services Colindale, Public Health England | Respiratory Virus Unit, Microbiology Services Colindale, Public Health England |
| EPI_ISL_423895 | Respiratory Virus Unit, Microbiology Services Colindale, Public Health England | Respiratory Virus Unit, Microbiology Services Colindale, Public Health England |

| EPI_Accession_ID | Originating Laboratory | Submitting Laboratory |
|---|---|---|
| EPI_ISL_423896 | Respiratory Virus Unit, Microbiology Services Colindale, Public Health England | Respiratory Virus Unit, Microbiology Services Colindale, Public Health England |
| EPI_ISL_423897 | Respiratory Virus Unit, Microbiology Services Colindale, Public Health England | Respiratory Virus Unit, Microbiology Services Colindale, Public Health England |
| EPI_ISL_423898 | Respiratory Virus Unit, Microbiology Services Colindale, Public Health England | Respiratory Virus Unit, Microbiology Services Colindale, Public Health England |
| EPI_ISL_423900 | Respiratory Virus Unit, Microbiology Services Colindale, Public Health England | Respiratory Virus Unit, Microbiology Services Colindale, Public Health England |
| EPI_ISL_423901 | Respiratory Virus Unit, Microbiology Services Colindale, Public Health England | Respiratory Virus Unit, Microbiology Services Colindale, Public Health England |
| EPI_ISL_423902 | Respiratory Virus Unit, Microbiology Services Colindale, Public Health England | Respiratory Virus Unit, Microbiology Services Colindale, Public Health England |
| EPI_ISL_423903 | Respiratory Virus Unit, Microbiology Services Colindale, Public Health England | Respiratory Virus Unit, Microbiology Services Colindale, Public Health England |
| EPI_ISL_423904 | Respiratory Virus Unit, Microbiology Services Colindale, Public Health England | Respiratory Virus Unit, Microbiology Services Colindale, Public Health England |
| EPI_ISL_423905 | Respiratory Virus Unit, Microbiology Services Colindale, Public Health England | Respiratory Virus Unit, Microbiology Services Colindale, Public Health England |
| EPI_ISL_423906 | Respiratory Virus Unit, Microbiology Services Colindale, Public Health England | Respiratory Virus Unit, Microbiology Services Colindale, Public Health England |
| EPI_ISL_423907 | Respiratory Virus Unit, Microbiology Services Colindale, Public Health England | Respiratory Virus Unit, Microbiology Services Colindale, Public Health England |
| EPI_ISL_423908 | Respiratory Virus Unit, Microbiology Services Colindale, Public Health England | Respiratory Virus Unit, Microbiology Services Colindale, Public Health England |
| EPI_ISL_423909 | Respiratory Virus Unit, Microbiology Services Colindale, Public Health England | Respiratory Virus Unit, Microbiology Services Colindale, Public Health England |
| EPI_ISL_423910 | Respiratory Virus Unit, Microbiology Services Colindale, Public Health England | Respiratory Virus Unit, Microbiology Services Colindale, Public Health England |
| EPI_ISL_423911 | Respiratory Virus Unit, Microbiology Services Colindale, Public Health England | Respiratory Virus Unit, Microbiology Services Colindale, Public Health England |
| EPI_ISL_423912 | Respiratory Virus Unit, Microbiology Services Colindale, Public Health England | Respiratory Virus Unit, Microbiology Services Colindale, Public Health England |

| EPI_Accession_ID | Originating Laboratory | Submitting Laboratory |
|---|---|---|
| EPI_ISL_423913 | Respiratory Virus Unit, Microbiology Services Colindale, Public Health England | Respiratory Virus Unit, Microbiology Services Colindale, Public Health England |
| EPI_ISL_423914 | Respiratory Virus Unit, Microbiology Services Colindale, Public Health England | Respiratory Virus Unit, Microbiology Services Colindale, Public Health England |
| EPI_ISL_423915 | Respiratory Virus Unit, Microbiology Services Colindale, Public Health England | Respiratory Virus Unit, Microbiology Services Colindale, Public Health England |
| EPI_ISL_423916 | Respiratory Virus Unit, Microbiology Services Colindale, Public Health England | Respiratory Virus Unit, Microbiology Services Colindale, Public Health England |
| EPI_ISL_423917 | Respiratory Virus Unit, Microbiology Services Colindale, Public Health England | Respiratory Virus Unit, Microbiology Services Colindale, Public Health England |
| EPI_ISL_423918 | Respiratory Virus Unit, Microbiology Services Colindale, Public Health England | Respiratory Virus Unit, Microbiology Services Colindale, Public Health England |
| EPI_ISL_423919 | Respiratory Virus Unit, Microbiology Services Colindale, Public Health England | Respiratory Virus Unit, Microbiology Services Colindale, Public Health England |
| EPI_ISL_423920 | Respiratory Virus Unit, Microbiology Services Colindale, Public Health England | Respiratory Virus Unit, Microbiology Services Colindale, Public Health England |
| EPI_ISL_423921 | Respiratory Virus Unit, Microbiology Services Colindale, Public Health England | Respiratory Virus Unit, Microbiology Services Colindale, Public Health England |
| EPI_ISL_423922 | Respiratory Virus Unit, Microbiology Services Colindale, Public Health England | Respiratory Virus Unit, Microbiology Services Colindale, Public Health England |
| EPI_ISL_423923 | Respiratory Virus Unit, Microbiology Services Colindale, Public Health England | Respiratory Virus Unit, Microbiology Services Colindale, Public Health England |
| EPI_ISL_423924 | Respiratory Virus Unit, Microbiology Services Colindale, Public Health England | Respiratory Virus Unit, Microbiology Services Colindale, Public Health England |
| EPI_ISL_423925 | Respiratory Virus Unit, Microbiology Services Colindale, Public Health England | Respiratory Virus Unit, Microbiology Services Colindale, Public Health England |
| EPI_ISL_423926 | Respiratory Virus Unit, Microbiology Services Colindale, Public Health England | Respiratory Virus Unit, Microbiology Services Colindale, Public Health England |
| EPI_ISL_423927 | Respiratory Virus Unit, Microbiology Services Colindale, Public Health England | Respiratory Virus Unit, Microbiology Services Colindale, Public Health England |
| EPI_ISL_423928 | Respiratory Virus Unit, Microbiology Services Colindale, Public Health England | Respiratory Virus Unit, Microbiology Services Colindale, Public Health England |

| EPI_Accession_ID | Originating Laboratory | Submitting Laboratory |
| --- | --- | --- |
| EPI_ISL_423929 | Respiratory Virus Unit, Microbiology Services Colindale, Public Health England | Respiratory Virus Unit, Microbiology Services Colindale, Public Health England |
| EPI_ISL_423930 | Respiratory Virus Unit, Microbiology Services Colindale, Public Health England | Respiratory Virus Unit, Microbiology Services Colindale, Public Health England |
| EPI_ISL_423931 | Respiratory Virus Unit, Microbiology Services Colindale, Public Health England | Respiratory Virus Unit, Microbiology Services Colindale, Public Health England |
| EPI_ISL_423932 | Respiratory Virus Unit, Microbiology Services Colindale, Public Health England | Respiratory Virus Unit, Microbiology Services Colindale, Public Health England |
| EPI_ISL_423933 | Respiratory Virus Unit, Microbiology Services Colindale, Public Health England | Respiratory Virus Unit, Microbiology Services Colindale, Public Health England |
| EPI_ISL_423934 | Respiratory Virus Unit, Microbiology Services Colindale, Public Health England | Respiratory Virus Unit, Microbiology Services Colindale, Public Health England |
| EPI_ISL_423935 | Respiratory Virus Unit, Microbiology Services Colindale, Public Health England | Respiratory Virus Unit, Microbiology Services Colindale, Public Health England |
| EPI_ISL_423936 | Respiratory Virus Unit, Microbiology Services Colindale, Public Health England | Respiratory Virus Unit, Microbiology Services Colindale, Public Health England |
| EPI_ISL_423937 | Respiratory Virus Unit, Microbiology Services Colindale, Public Health England | Respiratory Virus Unit, Microbiology Services Colindale, Public Health England |
| EPI_ISL_423938 | Respiratory Virus Unit, Microbiology Services Colindale, Public Health England | Respiratory Virus Unit, Microbiology Services Colindale, Public Health England |
| EPI_ISL_423939 | Respiratory Virus Unit, Microbiology Services Colindale, Public Health England | Respiratory Virus Unit, Microbiology Services Colindale, Public Health England |
| EPI_ISL_423940 | Respiratory Virus Unit, Microbiology Services Colindale, Public Health England | Respiratory Virus Unit, Microbiology Services Colindale, Public Health England |
| EPI_ISL_423941 | Respiratory Virus Unit, Microbiology Services Colindale, Public Health England | Respiratory Virus Unit, Microbiology Services Colindale, Public Health England |
| EPI_ISL_423942 | Respiratory Virus Unit, Microbiology Services Colindale, Public Health England | Respiratory Virus Unit, Microbiology Services Colindale, Public Health England |
| EPI_ISL_423943 | Respiratory Virus Unit, Microbiology Services Colindale, Public Health England | Respiratory Virus Unit, Microbiology Services Colindale, Public Health England |
| EPI_ISL_423944 | Respiratory Virus Unit, Microbiology Services Colindale, Public Health England | Respiratory Virus Unit, Microbiology Services Colindale, Public Health England |

| EPI_Accession_ID | Originating Laboratory | Submitting Laboratory |
|---|---|---|
| EPI_ISL_423945 | Respiratory Virus Unit, Microbiology Services Colindale, Public Health England | Respiratory Virus Unit, Microbiology Services Colindale, Public Health England |
| EPI_ISL_423946 | Respiratory Virus Unit, Microbiology Services Colindale, Public Health England | Respiratory Virus Unit, Microbiology Services Colindale, Public Health England |
| EPI_ISL_423947 | Respiratory Virus Unit, Microbiology Services Colindale, Public Health England | Respiratory Virus Unit, Microbiology Services Colindale, Public Health England |
| EPI_ISL_423948 | Respiratory Virus Unit, Microbiology Services Colindale, Public Health England | Respiratory Virus Unit, Microbiology Services Colindale, Public Health England |
| EPI_ISL_423949 | Respiratory Virus Unit, Microbiology Services Colindale, Public Health England | Respiratory Virus Unit, Microbiology Services Colindale, Public Health England |
| EPI_ISL_423950 | Respiratory Virus Unit, Microbiology Services Colindale, Public Health England | Respiratory Virus Unit, Microbiology Services Colindale, Public Health England |
| EPI_ISL_423951 | Respiratory Virus Unit, Microbiology Services Colindale, Public Health England | Respiratory Virus Unit, Microbiology Services Colindale, Public Health England |
| EPI_ISL_423952 | Respiratory Virus Unit, Microbiology Services Colindale, Public Health England | Respiratory Virus Unit, Microbiology Services Colindale, Public Health England |
| EPI_ISL_423953 | Respiratory Virus Unit, Microbiology Services Colindale, Public Health England | Respiratory Virus Unit, Microbiology Services Colindale, Public Health England |
| EPI_ISL_423954 | Respiratory Virus Unit, Microbiology Services Colindale, Public Health England | Respiratory Virus Unit, Microbiology Services Colindale, Public Health England |
| EPI_ISL_423955 | Respiratory Virus Unit, Microbiology Services Colindale, Public Health England | Respiratory Virus Unit, Microbiology Services Colindale, Public Health England |
| EPI_ISL_423956 | Respiratory Virus Unit, Microbiology Services Colindale, Public Health England | Respiratory Virus Unit, Microbiology Services Colindale, Public Health England |
| EPI_ISL_423957 | Respiratory Virus Unit, Microbiology Services Colindale, Public Health England | Respiratory Virus Unit, Microbiology Services Colindale, Public Health England |
| EPI_ISL_423958 | Respiratory Virus Unit, Microbiology Services Colindale, Public Health England | Respiratory Virus Unit, Microbiology Services Colindale, Public Health England |
| EPI_ISL_423959 | Respiratory Virus Unit, Microbiology Services Colindale, Public Health England | Respiratory Virus Unit, Microbiology Services Colindale, Public Health England |
| EPI_ISL_423960 | Respiratory Virus Unit, Microbiology Services Colindale, Public Health England | Respiratory Virus Unit, Microbiology Services Colindale, Public Health England |

| EPI_Accession_ID | Originating Laboratory | Submitting Laboratory |
|---|---|---|
| EPI_ISL_423961 | Respiratory Virus Unit, Microbiology Services Colindale, Public Health England | Respiratory Virus Unit, Microbiology Services Colindale, Public Health England |
| EPI_ISL_423962 | Respiratory Virus Unit, Microbiology Services Colindale, Public Health England | Respiratory Virus Unit, Microbiology Services Colindale, Public Health England |
| EPI_ISL_423963 | Respiratory Virus Unit, Microbiology Services Colindale, Public Health England | Respiratory Virus Unit, Microbiology Services Colindale, Public Health England |
| EPI_ISL_423964 | Respiratory Virus Unit, Microbiology Services Colindale, Public Health England | Respiratory Virus Unit, Microbiology Services Colindale, Public Health England |
| EPI_ISL_423965 | Respiratory Virus Unit, Microbiology Services Colindale, Public Health England | Respiratory Virus Unit, Microbiology Services Colindale, Public Health England |
| EPI_ISL_423966 | Respiratory Virus Unit, Microbiology Services Colindale, Public Health England | Respiratory Virus Unit, Microbiology Services Colindale, Public Health England |
| EPI_ISL_423967 | Respiratory Virus Unit, Microbiology Services Colindale, Public Health England | Respiratory Virus Unit, Microbiology Services Colindale, Public Health England |
| EPI_ISL_423968 | Respiratory Virus Unit, Microbiology Services Colindale, Public Health England | Respiratory Virus Unit, Microbiology Services Colindale, Public Health England |
| EPI_ISL_423969 | Respiratory Virus Unit, Microbiology Services Colindale, Public Health England | Respiratory Virus Unit, Microbiology Services Colindale, Public Health England |
| EPI_ISL_423970 | Respiratory Virus Unit, Microbiology Services Colindale, Public Health England | Respiratory Virus Unit, Microbiology Services Colindale, Public Health England |
| EPI_ISL_423971 | Respiratory Virus Unit, Microbiology Services Colindale, Public Health England | Respiratory Virus Unit, Microbiology Services Colindale, Public Health England |
| EPI_ISL_423972 | Respiratory Virus Unit, Microbiology Services Colindale, Public Health England | Respiratory Virus Unit, Microbiology Services Colindale, Public Health England |
| EPI_ISL_423973 | Respiratory Virus Unit, Microbiology Services Colindale, Public Health England | Respiratory Virus Unit, Microbiology Services Colindale, Public Health England |
| EPI_ISL_423974 | Respiratory Virus Unit, Microbiology Services Colindale, Public Health England | Respiratory Virus Unit, Microbiology Services Colindale, Public Health England |
| EPI_ISL_423975 | Respiratory Virus Unit, Microbiology Services Colindale, Public Health England | Respiratory Virus Unit, Microbiology Services Colindale, Public Health England |
| EPI_ISL_423976 | Respiratory Virus Unit, Microbiology Services Colindale, Public Health England | Respiratory Virus Unit, Microbiology Services Colindale, Public Health England |

| EPI_Accession_ID | Originating Laboratory | Submitting Laboratory |
| --- | --- | --- |
| EPI_ISL_423977 | Respiratory Virus Unit, Microbiology Services Colindale, Public Health England | Respiratory Virus Unit, Microbiology Services Colindale, Public Health England |
| EPI_ISL_423978 | Respiratory Virus Unit, Microbiology Services Colindale, Public Health England | Respiratory Virus Unit, Microbiology Services Colindale, Public Health England |
| EPI_ISL_423979 | Respiratory Virus Unit, Microbiology Services Colindale, Public Health England | Respiratory Virus Unit, Microbiology Services Colindale, Public Health England |
| EPI_ISL_423980 | Respiratory Virus Unit, Microbiology Services Colindale, Public Health England | Respiratory Virus Unit, Microbiology Services Colindale, Public Health England |
| EPI_ISL_423981 | Respiratory Virus Unit, Microbiology Services Colindale, Public Health England | Respiratory Virus Unit, Microbiology Services Colindale, Public Health England |
| EPI_ISL_423982 | Respiratory Virus Unit, Microbiology Services Colindale, Public Health England | Respiratory Virus Unit, Microbiology Services Colindale, Public Health England |
| EPI_ISL_423983 | Respiratory Virus Unit, Microbiology Services Colindale, Public Health England | Respiratory Virus Unit, Microbiology Services Colindale, Public Health England |
| EPI_ISL_423984 | Respiratory Virus Unit, Microbiology Services Colindale, Public Health England | Respiratory Virus Unit, Microbiology Services Colindale, Public Health England |
| EPI_ISL_423985 | Respiratory Virus Unit, Microbiology Services Colindale, Public Health England | Respiratory Virus Unit, Microbiology Services Colindale, Public Health England |
| EPI_ISL_423986 | Respiratory Virus Unit, Microbiology Services Colindale, Public Health England | Respiratory Virus Unit, Microbiology Services Colindale, Public Health England |
| EPI_ISL_423987 | Respiratory Virus Unit, Microbiology Services Colindale, Public Health England | Respiratory Virus Unit, Microbiology Services Colindale, Public Health England |
| EPI_ISL_423988 | Respiratory Virus Unit, Microbiology Services Colindale, Public Health England | Respiratory Virus Unit, Microbiology Services Colindale, Public Health England |
| EPI_ISL_423989 | Respiratory Virus Unit, Microbiology Services Colindale, Public Health England | Respiratory Virus Unit, Microbiology Services Colindale, Public Health England |
| EPI_ISL_423990 | Respiratory Virus Unit, Microbiology Services Colindale, Public Health England | Respiratory Virus Unit, Microbiology Services Colindale, Public Health England |
| EPI_ISL_423991 | Respiratory Virus Unit, Microbiology Services Colindale, Public Health England | Respiratory Virus Unit, Microbiology Services Colindale, Public Health England |
| EPI_ISL_423992 | Respiratory Virus Unit, Microbiology Services Colindale, Public Health England | Respiratory Virus Unit, Microbiology Services Colindale, Public Health England |

| EPI_Accession_ID | Originating Laboratory | Submitting Laboratory |
|---|---|---|
| EPI_ISL_423993 | Respiratory Virus Unit, Microbiology Services Colindale, Public Health England | Respiratory Virus Unit, Microbiology Services Colindale, Public Health England |
| EPI_ISL_423994 | Respiratory Virus Unit, Microbiology Services Colindale, Public Health England | Respiratory Virus Unit, Microbiology Services Colindale, Public Health England |
| EPI_ISL_423995 | Respiratory Virus Unit, Microbiology Services Colindale, Public Health England | Respiratory Virus Unit, Microbiology Services Colindale, Public Health England |
| EPI_ISL_423996 | Respiratory Virus Unit, Microbiology Services Colindale, Public Health England | Respiratory Virus Unit, Microbiology Services Colindale, Public Health England |
| EPI_ISL_423997 | Respiratory Virus Unit, Microbiology Services Colindale, Public Health England | Respiratory Virus Unit, Microbiology Services Colindale, Public Health England |
| EPI_ISL_423998 | Respiratory Virus Unit, Microbiology Services Colindale, Public Health England | Respiratory Virus Unit, Microbiology Services Colindale, Public Health England |
| EPI_ISL_423999 | Respiratory Virus Unit, Microbiology Services Colindale, Public Health England | Respiratory Virus Unit, Microbiology Services Colindale, Public Health England |
| EPI_ISL_424000 | Respiratory Virus Unit, Microbiology Services Colindale, Public Health England | Respiratory Virus Unit, Microbiology Services Colindale, Public Health England |
| EPI_ISL_424001 | Respiratory Virus Unit, Microbiology Services Colindale, Public Health England | Respiratory Virus Unit, Microbiology Services Colindale, Public Health England |
| EPI_ISL_424002 | Respiratory Virus Unit, Microbiology Services Colindale, Public Health England | Respiratory Virus Unit, Microbiology Services Colindale, Public Health England |
| EPI_ISL_424003 | Respiratory Virus Unit, Microbiology Services Colindale, Public Health England | Respiratory Virus Unit, Microbiology Services Colindale, Public Health England |
| EPI_ISL_424004 | Respiratory Virus Unit, Microbiology Services Colindale, Public Health England | Respiratory Virus Unit, Microbiology Services Colindale, Public Health England |
| EPI_ISL_424005 | Respiratory Virus Unit, Microbiology Services Colindale, Public Health England | Respiratory Virus Unit, Microbiology Services Colindale, Public Health England |
| EPI_ISL_424006 | Respiratory Virus Unit, Microbiology Services Colindale, Public Health England | Respiratory Virus Unit, Microbiology Services Colindale, Public Health England |
| EPI_ISL_424007 | Respiratory Virus Unit, Microbiology Services Colindale, Public Health England | Respiratory Virus Unit, Microbiology Services Colindale, Public Health England |
| EPI_ISL_424008 | Respiratory Virus Unit, Microbiology Services Colindale, Public Health England | Respiratory Virus Unit, Microbiology Services Colindale, Public Health England |

| EPI_Accession_ID | Originating Laboratory | Submitting Laboratory |
|---|---|---|
| EPI_ISL_424009 | Respiratory Virus Unit, Microbiology Services Colindale, Public Health England | Respiratory Virus Unit, Microbiology Services Colindale, Public Health England |
| EPI_ISL_424010 | Respiratory Virus Unit, Microbiology Services Colindale, Public Health England | Respiratory Virus Unit, Microbiology Services Colindale, Public Health England |
| EPI_ISL_424011 | Respiratory Virus Unit, Microbiology Services Colindale, Public Health England | Respiratory Virus Unit, Microbiology Services Colindale, Public Health England |
| EPI_ISL_424012 | Respiratory Virus Unit, Microbiology Services Colindale, Public Health England | Respiratory Virus Unit, Microbiology Services Colindale, Public Health England |
| EPI_ISL_424013 | Respiratory Virus Unit, Microbiology Services Colindale, Public Health England | Respiratory Virus Unit, Microbiology Services Colindale, Public Health England |
| EPI_ISL_424014 | Respiratory Virus Unit, Microbiology Services Colindale, Public Health England | Respiratory Virus Unit, Microbiology Services Colindale, Public Health England |
| EPI_ISL_424015 | Respiratory Virus Unit, Microbiology Services Colindale, Public Health England | Respiratory Virus Unit, Microbiology Services Colindale, Public Health England |
| EPI_ISL_424016 | Respiratory Virus Unit, Microbiology Services Colindale, Public Health England | Respiratory Virus Unit, Microbiology Services Colindale, Public Health England |
| EPI_ISL_424017 | Respiratory Virus Unit, Microbiology Services Colindale, Public Health England | Respiratory Virus Unit, Microbiology Services Colindale, Public Health England |
| EPI_ISL_424018 | Respiratory Virus Unit, Microbiology Services Colindale, Public Health England | Respiratory Virus Unit, Microbiology Services Colindale, Public Health England |
| EPI_ISL_424019 | Respiratory Virus Unit, Microbiology Services Colindale, Public Health England | Respiratory Virus Unit, Microbiology Services Colindale, Public Health England |
| EPI_ISL_424020 | Respiratory Virus Unit, Microbiology Services Colindale, Public Health England | Respiratory Virus Unit, Microbiology Services Colindale, Public Health England |
| EPI_ISL_424021 | Respiratory Virus Unit, Microbiology Services Colindale, Public Health England | Respiratory Virus Unit, Microbiology Services Colindale, Public Health England |
| EPI_ISL_424022 | Respiratory Virus Unit, Microbiology Services Colindale, Public Health England | Respiratory Virus Unit, Microbiology Services Colindale, Public Health England |
| EPI_ISL_424023 | Respiratory Virus Unit, Microbiology Services Colindale, Public Health England | Respiratory Virus Unit, Microbiology Services Colindale, Public Health England |
| EPI_ISL_424024 | Respiratory Virus Unit, Microbiology Services Colindale, Public Health England | Respiratory Virus Unit, Microbiology Services Colindale, Public Health England |

| EPI_Accession_ID | Originating Laboratory | Submitting Laboratory |
|---|---|---|
| EPI_ISL_424025 | Respiratory Virus Unit, Microbiology Services Colindale, Public Health England | Respiratory Virus Unit, Microbiology Services Colindale, Public Health England |
| EPI_ISL_424026 | Respiratory Virus Unit, Microbiology Services Colindale, Public Health England | Respiratory Virus Unit, Microbiology Services Colindale, Public Health England |
| EPI_ISL_424027 | Respiratory Virus Unit, Microbiology Services Colindale, Public Health England | Respiratory Virus Unit, Microbiology Services Colindale, Public Health England |
| EPI_ISL_424028 | Respiratory Virus Unit, Microbiology Services Colindale, Public Health England | Respiratory Virus Unit, Microbiology Services Colindale, Public Health England |
| EPI_ISL_424029 | Respiratory Virus Unit, Microbiology Services Colindale, Public Health England | Respiratory Virus Unit, Microbiology Services Colindale, Public Health England |
| EPI_ISL_424030 | Respiratory Virus Unit, Microbiology Services Colindale, Public Health England | Respiratory Virus Unit, Microbiology Services Colindale, Public Health England |
| EPI_ISL_424031 | Respiratory Virus Unit, Microbiology Services Colindale, Public Health England | Respiratory Virus Unit, Microbiology Services Colindale, Public Health England |
| EPI_ISL_424032 | Respiratory Virus Unit, Microbiology Services Colindale, Public Health England | Respiratory Virus Unit, Microbiology Services Colindale, Public Health England |
| EPI_ISL_424033 | Respiratory Virus Unit, Microbiology Services Colindale, Public Health England | Respiratory Virus Unit, Microbiology Services Colindale, Public Health England |
| EPI_ISL_424034 | Respiratory Virus Unit, Microbiology Services Colindale, Public Health England | Respiratory Virus Unit, Microbiology Services Colindale, Public Health England |
| EPI_ISL_424035 | Respiratory Virus Unit, Microbiology Services Colindale, Public Health England | Respiratory Virus Unit, Microbiology Services Colindale, Public Health England |
| EPI_ISL_424036 | Respiratory Virus Unit, Microbiology Services Colindale, Public Health England | Respiratory Virus Unit, Microbiology Services Colindale, Public Health England |
| EPI_ISL_424037 | Respiratory Virus Unit, Microbiology Services Colindale, Public Health England | Respiratory Virus Unit, Microbiology Services Colindale, Public Health England |
| EPI_ISL_424038 | Respiratory Virus Unit, Microbiology Services Colindale, Public Health England | Respiratory Virus Unit, Microbiology Services Colindale, Public Health England |
| EPI_ISL_424039 | Respiratory Virus Unit, Microbiology Services Colindale, Public Health England | Respiratory Virus Unit, Microbiology Services Colindale, Public Health England |
| EPI_ISL_424040 | Respiratory Virus Unit, Microbiology Services Colindale, Public Health England | Respiratory Virus Unit, Microbiology Services Colindale, Public Health England |

| EPI_Accession_ID | Originating Laboratory | Submitting Laboratory |
|---|---|---|
| EPI_ISL_424041 | Respiratory Virus Unit, Microbiology Services Colindale, Public Health England | Respiratory Virus Unit, Microbiology Services Colindale, Public Health England |
| EPI_ISL_424042 | Respiratory Virus Unit, Microbiology Services Colindale, Public Health England | Respiratory Virus Unit, Microbiology Services Colindale, Public Health England |
| EPI_ISL_424043 | Respiratory Virus Unit, Microbiology Services Colindale, Public Health England | Respiratory Virus Unit, Microbiology Services Colindale, Public Health England |
| EPI_ISL_424044 | Respiratory Virus Unit, Microbiology Services Colindale, Public Health England | Respiratory Virus Unit, Microbiology Services Colindale, Public Health England |
| EPI_ISL_424045 | Respiratory Virus Unit, Microbiology Services Colindale, Public Health England | Respiratory Virus Unit, Microbiology Services Colindale, Public Health England |
| EPI_ISL_424046 | Respiratory Virus Unit, Microbiology Services Colindale, Public Health England | Respiratory Virus Unit, Microbiology Services Colindale, Public Health England |
| EPI_ISL_424047 | Respiratory Virus Unit, Microbiology Services Colindale, Public Health England | Respiratory Virus Unit, Microbiology Services Colindale, Public Health England |
| EPI_ISL_424048 | Respiratory Virus Unit, Microbiology Services Colindale, Public Health England | Respiratory Virus Unit, Microbiology Services Colindale, Public Health England |
| EPI_ISL_424049 | Respiratory Virus Unit, Microbiology Services Colindale, Public Health England | Respiratory Virus Unit, Microbiology Services Colindale, Public Health England |
| EPI_ISL_424050 | Respiratory Virus Unit, Microbiology Services Colindale, Public Health England | Respiratory Virus Unit, Microbiology Services Colindale, Public Health England |
| EPI_ISL_424052 | Respiratory Virus Unit, Microbiology Services Colindale, Public Health England | Respiratory Virus Unit, Microbiology Services Colindale, Public Health England |
| EPI_ISL_424053 | Respiratory Virus Unit, Microbiology Services Colindale, Public Health England | Respiratory Virus Unit, Microbiology Services Colindale, Public Health England |
| EPI_ISL_424054 | Respiratory Virus Unit, Microbiology Services Colindale, Public Health England | Respiratory Virus Unit, Microbiology Services Colindale, Public Health England |
| EPI_ISL_424055 | Respiratory Virus Unit, Microbiology Services Colindale, Public Health England | Respiratory Virus Unit, Microbiology Services Colindale, Public Health England |
| EPI_ISL_424056 | Respiratory Virus Unit, Microbiology Services Colindale, Public Health England | Respiratory Virus Unit, Microbiology Services Colindale, Public Health England |
| EPI_ISL_424057 | Respiratory Virus Unit, Microbiology Services Colindale, Public Health England | Respiratory Virus Unit, Microbiology Services Colindale, Public Health England |

| EPI_Accession_ID | Originating Laboratory | Submitting Laboratory |
|---|---|---|
| EPI_ISL_424058 | Respiratory Virus Unit, Microbiology Services Colindale, Public Health England | Respiratory Virus Unit, Microbiology Services Colindale, Public Health England |
| EPI_ISL_424059 | Respiratory Virus Unit, Microbiology Services Colindale, Public Health England | Respiratory Virus Unit, Microbiology Services Colindale, Public Health England |
| EPI_ISL_424060 | Respiratory Virus Unit, Microbiology Services Colindale, Public Health England | Respiratory Virus Unit, Microbiology Services Colindale, Public Health England |
| EPI_ISL_424061 | Respiratory Virus Unit, Microbiology Services Colindale, Public Health England | Respiratory Virus Unit, Microbiology Services Colindale, Public Health England |
| EPI_ISL_424062 | Respiratory Virus Unit, Microbiology Services Colindale, Public Health England | Respiratory Virus Unit, Microbiology Services Colindale, Public Health England |
| EPI_ISL_424063 | Respiratory Virus Unit, Microbiology Services Colindale, Public Health England | Respiratory Virus Unit, Microbiology Services Colindale, Public Health England |
| EPI_ISL_424064 | Respiratory Virus Unit, Microbiology Services Colindale, Public Health England | Respiratory Virus Unit, Microbiology Services Colindale, Public Health England |
| EPI_ISL_424065 | Respiratory Virus Unit, Microbiology Services Colindale, Public Health England | Respiratory Virus Unit, Microbiology Services Colindale, Public Health England |
| EPI_ISL_424066 | Respiratory Virus Unit, Microbiology Services Colindale, Public Health England | Respiratory Virus Unit, Microbiology Services Colindale, Public Health England |
| EPI_ISL_424067 | Respiratory Virus Unit, Microbiology Services Colindale, Public Health England | Respiratory Virus Unit, Microbiology Services Colindale, Public Health England |
| EPI_ISL_424068 | Respiratory Virus Unit, Microbiology Services Colindale, Public Health England | Respiratory Virus Unit, Microbiology Services Colindale, Public Health England |
| EPI_ISL_424069 | Respiratory Virus Unit, Microbiology Services Colindale, Public Health England | Respiratory Virus Unit, Microbiology Services Colindale, Public Health England |
| EPI_ISL_424070 | Respiratory Virus Unit, Microbiology Services Colindale, Public Health England | Respiratory Virus Unit, Microbiology Services Colindale, Public Health England |
| EPI_ISL_424071 | Respiratory Virus Unit, Microbiology Services Colindale, Public Health England | Respiratory Virus Unit, Microbiology Services Colindale, Public Health England |
| EPI_ISL_424072 | Respiratory Virus Unit, Microbiology Services Colindale, Public Health England | Respiratory Virus Unit, Microbiology Services Colindale, Public Health England |
| EPI_ISL_424073 | Respiratory Virus Unit, Microbiology Services Colindale, Public Health England | Respiratory Virus Unit, Microbiology Services Colindale, Public Health England |

| EPI_Accession_ID | Originating Laboratory | Submitting Laboratory |
|---|---|---|
| EPI_ISL_424074 | Respiratory Virus Unit, Microbiology Services Colindale, Public Health England | Respiratory Virus Unit, Microbiology Services Colindale, Public Health England |
| EPI_ISL_424075 | Respiratory Virus Unit, Microbiology Services Colindale, Public Health England | Respiratory Virus Unit, Microbiology Services Colindale, Public Health England |
| EPI_ISL_424076 | Respiratory Virus Unit, Microbiology Services Colindale, Public Health England | Respiratory Virus Unit, Microbiology Services Colindale, Public Health England |
| EPI_ISL_424077 | Respiratory Virus Unit, Microbiology Services Colindale, Public Health England | Respiratory Virus Unit, Microbiology Services Colindale, Public Health England |
| EPI_ISL_424078 | Respiratory Virus Unit, Microbiology Services Colindale, Public Health England | Respiratory Virus Unit, Microbiology Services Colindale, Public Health England |
| EPI_ISL_424079 | Respiratory Virus Unit, Microbiology Services Colindale, Public Health England | Respiratory Virus Unit, Microbiology Services Colindale, Public Health England |
| EPI_ISL_424080 | Respiratory Virus Unit, Microbiology Services Colindale, Public Health England | Respiratory Virus Unit, Microbiology Services Colindale, Public Health England |
| EPI_ISL_424081 | Respiratory Virus Unit, Microbiology Services Colindale, Public Health England | Respiratory Virus Unit, Microbiology Services Colindale, Public Health England |
| EPI_ISL_424082 | Respiratory Virus Unit, Microbiology Services Colindale, Public Health England | Respiratory Virus Unit, Microbiology Services Colindale, Public Health England |
| EPI_ISL_424083 | Respiratory Virus Unit, Microbiology Services Colindale, Public Health England | Respiratory Virus Unit, Microbiology Services Colindale, Public Health England |
| EPI_ISL_424084 | Respiratory Virus Unit, Microbiology Services Colindale, Public Health England | Respiratory Virus Unit, Microbiology Services Colindale, Public Health England |
| EPI_ISL_424085 | Respiratory Virus Unit, Microbiology Services Colindale, Public Health England | Respiratory Virus Unit, Microbiology Services Colindale, Public Health England |
| EPI_ISL_424086 | Respiratory Virus Unit, Microbiology Services Colindale, Public Health England | Respiratory Virus Unit, Microbiology Services Colindale, Public Health England |
| EPI_ISL_424087 | Respiratory Virus Unit, Microbiology Services Colindale, Public Health England | Respiratory Virus Unit, Microbiology Services Colindale, Public Health England |
| EPI_ISL_424088 | Respiratory Virus Unit, Microbiology Services Colindale, Public Health England | Respiratory Virus Unit, Microbiology Services Colindale, Public Health England |
| EPI_ISL_424089 | Respiratory Virus Unit, Microbiology Services Colindale, Public Health England | Respiratory Virus Unit, Microbiology Services Colindale, Public Health England |

| EPI_Accession_ID | Originating Laboratory | Submitting Laboratory |
|---|---|---|
| EPI_ISL_424090 | Respiratory Virus Unit, Microbiology Services Colindale, Public Health England | Respiratory Virus Unit, Microbiology Services Colindale, Public Health England |
| EPI_ISL_424091 | Respiratory Virus Unit, Microbiology Services Colindale, Public Health England | Respiratory Virus Unit, Microbiology Services Colindale, Public Health England |
| EPI_ISL_424092 | Respiratory Virus Unit, Microbiology Services Colindale, Public Health England | Respiratory Virus Unit, Microbiology Services Colindale, Public Health England |
| EPI_ISL_424093 | Respiratory Virus Unit, Microbiology Services Colindale, Public Health England | Respiratory Virus Unit, Microbiology Services Colindale, Public Health England |
| EPI_ISL_424094 | Respiratory Virus Unit, Microbiology Services Colindale, Public Health England | Respiratory Virus Unit, Microbiology Services Colindale, Public Health England |
| EPI_ISL_424095 | Respiratory Virus Unit, Microbiology Services Colindale, Public Health England | Respiratory Virus Unit, Microbiology Services Colindale, Public Health England |
| EPI_ISL_424096 | Respiratory Virus Unit, Microbiology Services Colindale, Public Health England | Respiratory Virus Unit, Microbiology Services Colindale, Public Health England |
| EPI_ISL_424097 | Respiratory Virus Unit, Microbiology Services Colindale, Public Health England | Respiratory Virus Unit, Microbiology Services Colindale, Public Health England |
| EPI_ISL_424098 | Respiratory Virus Unit, Microbiology Services Colindale, Public Health England | Respiratory Virus Unit, Microbiology Services Colindale, Public Health England |
| EPI_ISL_424099 | Respiratory Virus Unit, Microbiology Services Colindale, Public Health England | Respiratory Virus Unit, Microbiology Services Colindale, Public Health England |
| EPI_ISL_424100 | Respiratory Virus Unit, Microbiology Services Colindale, Public Health England | Respiratory Virus Unit, Microbiology Services Colindale, Public Health England |
| EPI_ISL_424101 | Respiratory Virus Unit, Microbiology Services Colindale, Public Health England | Respiratory Virus Unit, Microbiology Services Colindale, Public Health England |
| EPI_ISL_424102 | Respiratory Virus Unit, Microbiology Services Colindale, Public Health England | Respiratory Virus Unit, Microbiology Services Colindale, Public Health England |
| EPI_ISL_424103 | Respiratory Virus Unit, Microbiology Services Colindale, Public Health England | Respiratory Virus Unit, Microbiology Services Colindale, Public Health England |
| EPI_ISL_424104 | Respiratory Virus Unit, Microbiology Services Colindale, Public Health England | Respiratory Virus Unit, Microbiology Services Colindale, Public Health England |
| EPI_ISL_424105 | Respiratory Virus Unit, Microbiology Services Colindale, Public Health England | Respiratory Virus Unit, Microbiology Services Colindale, Public Health England |

| EPI_Accession_ID | Originating Laboratory | Submitting Laboratory |
|---|---|---|
| EPI_ISL_424106 | Respiratory Virus Unit, Microbiology Services Colindale, Public Health England | Respiratory Virus Unit, Microbiology Services Colindale, Public Health England |
| EPI_ISL_424107 | Respiratory Virus Unit, Microbiology Services Colindale, Public Health England | Respiratory Virus Unit, Microbiology Services Colindale, Public Health England |
| EPI_ISL_424108 | Respiratory Virus Unit, Microbiology Services Colindale, Public Health England | Respiratory Virus Unit, Microbiology Services Colindale, Public Health England |
| EPI_ISL_424109 | Respiratory Virus Unit, Microbiology Services Colindale, Public Health England | Respiratory Virus Unit, Microbiology Services Colindale, Public Health England |
| EPI_ISL_424110 | Respiratory Virus Unit, Microbiology Services Colindale, Public Health England | Respiratory Virus Unit, Microbiology Services Colindale, Public Health England |
| EPI_ISL_424111 | Respiratory Virus Unit, Microbiology Services Colindale, Public Health England | Respiratory Virus Unit, Microbiology Services Colindale, Public Health England |
| EPI_ISL_424112 | Respiratory Virus Unit, Microbiology Services Colindale, Public Health England | Respiratory Virus Unit, Microbiology Services Colindale, Public Health England |
| EPI_ISL_424113 | Respiratory Virus Unit, Microbiology Services Colindale, Public Health England | Respiratory Virus Unit, Microbiology Services Colindale, Public Health England |
| EPI_ISL_424114 | Respiratory Virus Unit, Microbiology Services Colindale, Public Health England | Respiratory Virus Unit, Microbiology Services Colindale, Public Health England |
| EPI_ISL_424115 | Respiratory Virus Unit, Microbiology Services Colindale, Public Health England | Respiratory Virus Unit, Microbiology Services Colindale, Public Health England |
| EPI_ISL_424116 | Respiratory Virus Unit, Microbiology Services Colindale, Public Health England | Respiratory Virus Unit, Microbiology Services Colindale, Public Health England |
| EPI_ISL_424117 | Respiratory Virus Unit, Microbiology Services Colindale, Public Health England | Respiratory Virus Unit, Microbiology Services Colindale, Public Health England |
| EPI_ISL_424118 | Respiratory Virus Unit, Microbiology Services Colindale, Public Health England | Respiratory Virus Unit, Microbiology Services Colindale, Public Health England |
| EPI_ISL_424119 | Respiratory Virus Unit, Microbiology Services Colindale, Public Health England | Respiratory Virus Unit, Microbiology Services Colindale, Public Health England |
| EPI_ISL_424120 | Respiratory Virus Unit, Microbiology Services Colindale, Public Health England | Respiratory Virus Unit, Microbiology Services Colindale, Public Health England |
| EPI_ISL_424121 | Respiratory Virus Unit, Microbiology Services Colindale, Public Health England | Respiratory Virus Unit, Microbiology Services Colindale, Public Health England |

| EPI_Accession_ID | Originating Laboratory | Submitting Laboratory |
|---|---|---|
| EPI_ISL_424122 | Respiratory Virus Unit, Microbiology Services Colindale, Public Health England | Respiratory Virus Unit, Microbiology Services Colindale, Public Health England |
| EPI_ISL_424123 | Respiratory Virus Unit, Microbiology Services Colindale, Public Health England | Respiratory Virus Unit, Microbiology Services Colindale, Public Health England |
| EPI_ISL_424124 | Respiratory Virus Unit, Microbiology Services Colindale, Public Health England | Respiratory Virus Unit, Microbiology Services Colindale, Public Health England |
| EPI_ISL_424125 | Respiratory Virus Unit, Microbiology Services Colindale, Public Health England | Respiratory Virus Unit, Microbiology Services Colindale, Public Health England |
| EPI_ISL_424126 | Respiratory Virus Unit, Microbiology Services Colindale, Public Health England | Respiratory Virus Unit, Microbiology Services Colindale, Public Health England |
| EPI_ISL_424127 | Respiratory Virus Unit, Microbiology Services Colindale, Public Health England | Respiratory Virus Unit, Microbiology Services Colindale, Public Health England |
| EPI_ISL_424128 | Respiratory Virus Unit, Microbiology Services Colindale, Public Health England | Respiratory Virus Unit, Microbiology Services Colindale, Public Health England |
| EPI_ISL_424129 | Respiratory Virus Unit, Microbiology Services Colindale, Public Health England | Respiratory Virus Unit, Microbiology Services Colindale, Public Health England |
| EPI_ISL_424130 | Respiratory Virus Unit, Microbiology Services Colindale, Public Health England | Respiratory Virus Unit, Microbiology Services Colindale, Public Health England |
| EPI_ISL_424131 | Respiratory Virus Unit, Microbiology Services Colindale, Public Health England | Respiratory Virus Unit, Microbiology Services Colindale, Public Health England |
| EPI_ISL_424132 | Respiratory Virus Unit, Microbiology Services Colindale, Public Health England | Respiratory Virus Unit, Microbiology Services Colindale, Public Health England |
| EPI_ISL_424133 | Respiratory Virus Unit, Microbiology Services Colindale, Public Health England | Respiratory Virus Unit, Microbiology Services Colindale, Public Health England |
| EPI_ISL_424134 | Respiratory Virus Unit, Microbiology Services Colindale, Public Health England | Respiratory Virus Unit, Microbiology Services Colindale, Public Health England |
| EPI_ISL_424135 | Respiratory Virus Unit, Microbiology Services Colindale, Public Health England | Respiratory Virus Unit, Microbiology Services Colindale, Public Health England |
| EPI_ISL_424136 | Respiratory Virus Unit, Microbiology Services Colindale, Public Health England | Respiratory Virus Unit, Microbiology Services Colindale, Public Health England |
| EPI_ISL_424138 | Respiratory Virus Unit, Microbiology Services Colindale, Public Health England | Respiratory Virus Unit, Microbiology Services Colindale, Public Health England |

| EPI_Accession_ID | Originating Laboratory | Submitting Laboratory |
|---|---|---|
| EPI_ISL_424139 | Respiratory Virus Unit, Microbiology Services Colindale, Public Health England | Respiratory Virus Unit, Microbiology Services Colindale, Public Health England |
| EPI_ISL_424140 | Respiratory Virus Unit, Microbiology Services Colindale, Public Health England | Respiratory Virus Unit, Microbiology Services Colindale, Public Health England |
| EPI_ISL_424141 | Respiratory Virus Unit, Microbiology Services Colindale, Public Health England | Respiratory Virus Unit, Microbiology Services Colindale, Public Health England |
| EPI_ISL_424142 | Respiratory Virus Unit, Microbiology Services Colindale, Public Health England | Respiratory Virus Unit, Microbiology Services Colindale, Public Health England |
| EPI_ISL_424143 | Respiratory Virus Unit, Microbiology Services Colindale, Public Health England | Respiratory Virus Unit, Microbiology Services Colindale, Public Health England |
| EPI_ISL_424144 | Respiratory Virus Unit, Microbiology Services Colindale, Public Health England | Respiratory Virus Unit, Microbiology Services Colindale, Public Health England |
| EPI_ISL_424145 | Respiratory Virus Unit, Microbiology Services Colindale, Public Health England | Respiratory Virus Unit, Microbiology Services Colindale, Public Health England |
| EPI_ISL_424146 | Respiratory Virus Unit, Microbiology Services Colindale, Public Health England | Respiratory Virus Unit, Microbiology Services Colindale, Public Health England |
| EPI_ISL_424147 | Respiratory Virus Unit, Microbiology Services Colindale, Public Health England | Respiratory Virus Unit, Microbiology Services Colindale, Public Health England |
| EPI_ISL_424148 | Respiratory Virus Unit, Microbiology Services Colindale, Public Health England | Respiratory Virus Unit, Microbiology Services Colindale, Public Health England |
| EPI_ISL_424149 | Respiratory Virus Unit, Microbiology Services Colindale, Public Health England | Respiratory Virus Unit, Microbiology Services Colindale, Public Health England |
| EPI_ISL_424150 | Respiratory Virus Unit, Microbiology Services Colindale, Public Health England | Respiratory Virus Unit, Microbiology Services Colindale, Public Health England |
| EPI_ISL_424151 | Respiratory Virus Unit, Microbiology Services Colindale, Public Health England | Respiratory Virus Unit, Microbiology Services Colindale, Public Health England |
| EPI_ISL_424152 | Respiratory Virus Unit, Microbiology Services Colindale, Public Health England | Respiratory Virus Unit, Microbiology Services Colindale, Public Health England |
| EPI_ISL_424153 | Respiratory Virus Unit, Microbiology Services Colindale, Public Health England | Respiratory Virus Unit, Microbiology Services Colindale, Public Health England |
| EPI_ISL_424154 | Respiratory Virus Unit, Microbiology Services Colindale, Public Health England | Respiratory Virus Unit, Microbiology Services Colindale, Public Health England |

| EPI_Accession_ID | Originating Laboratory | Submitting Laboratory |
|---|---|---|
| EPI_ISL_424155 | Respiratory Virus Unit, Microbiology Services Colindale, Public Health England | Respiratory Virus Unit, Microbiology Services Colindale, Public Health England |
| EPI_ISL_424156 | Respiratory Virus Unit, Microbiology Services Colindale, Public Health England | Respiratory Virus Unit, Microbiology Services Colindale, Public Health England |
| EPI_ISL_424157 | Respiratory Virus Unit, Microbiology Services Colindale, Public Health England | Respiratory Virus Unit, Microbiology Services Colindale, Public Health England |
| EPI_ISL_424158 | Respiratory Virus Unit, Microbiology Services Colindale, Public Health England | Respiratory Virus Unit, Microbiology Services Colindale, Public Health England |
| EPI_ISL_424159 | Respiratory Virus Unit, Microbiology Services Colindale, Public Health England | Respiratory Virus Unit, Microbiology Services Colindale, Public Health England |
| EPI_ISL_424160 | Respiratory Virus Unit, Microbiology Services Colindale, Public Health England | Respiratory Virus Unit, Microbiology Services Colindale, Public Health England |
| EPI_ISL_424161 | Respiratory Virus Unit, Microbiology Services Colindale, Public Health England | Respiratory Virus Unit, Microbiology Services Colindale, Public Health England |
| EPI_ISL_424162 | Respiratory Virus Unit, Microbiology Services Colindale, Public Health England | Respiratory Virus Unit, Microbiology Services Colindale, Public Health England |
| EPI_ISL_424163 | Respiratory Virus Unit, Microbiology Services Colindale, Public Health England | Respiratory Virus Unit, Microbiology Services Colindale, Public Health England |
| EPI_ISL_424164 | Respiratory Virus Unit, Microbiology Services Colindale, Public Health England | Respiratory Virus Unit, Microbiology Services Colindale, Public Health England |
| EPI_ISL_424165 | Respiratory Virus Unit, Microbiology Services Colindale, Public Health England | Respiratory Virus Unit, Microbiology Services Colindale, Public Health England |
| EPI_ISL_424166 | UW Virology Lab | UW Virology Lab |
| EPI_ISL_424167 | UW Virology Lab | UW Virology Lab |
| EPI_ISL_424168 | UW Virology Lab | UW Virology Lab |
| EPI_ISL_424169 | UW Virology Lab | UW Virology Lab |
| EPI_ISL_424170 | UW Virology Lab | UW Virology Lab |
| EPI_ISL_424172 | UW Virology Lab | UW Virology Lab |
| EPI_ISL_424173 | UW Virology Lab | UW Virology Lab |
| EPI_ISL_424174 | UW Virology Lab | UW Virology Lab |
| EPI_ISL_424175 | UW Virology Lab | UW Virology Lab |
| EPI_ISL_424176 | UW Virology Lab | UW Virology Lab |
| EPI_ISL_424177 | UW Virology Lab | UW Virology Lab |

| EPI_Accession_ID | Originating Laboratory | Submitting Laboratory |
|---|---|---|
| EPI_ISL_424178 | UW Virology Lab | UW Virology Lab |
| EPI_ISL_424179 | UW Virology Lab | UW Virology Lab |
| EPI_ISL_424180 | UW Virology Lab | UW Virology Lab |
| EPI_ISL_424181 | UW Virology Lab | UW Virology Lab |
| EPI_ISL_424182 | UW Virology Lab | UW Virology Lab |
| EPI_ISL_424183 | UW Virology Lab | UW Virology Lab |
| EPI_ISL_424184 | UW Virology Lab | UW Virology Lab |
| EPI_ISL_424186 | UW Virology Lab | UW Virology Lab |
| EPI_ISL_424187 | UW Virology Lab | UW Virology Lab |
| EPI_ISL_424188 | UW Virology Lab | UW Virology Lab |
| EPI_ISL_424190 | UW Virology Lab | UW Virology Lab |
| EPI_ISL_424191 | UW Virology Lab | UW Virology Lab |
| EPI_ISL_424192 | UW Virology Lab | UW Virology Lab |
| EPI_ISL_424193 | UW Virology Lab | UW Virology Lab |
| EPI_ISL_424194 | UW Virology Lab | UW Virology Lab |
| EPI_ISL_424195 | UW Virology Lab | UW Virology Lab |
| EPI_ISL_424196 | UW Virology Lab | UW Virology Lab |
| EPI_ISL_424197 | UW Virology Lab | UW Virology Lab |
| EPI_ISL_424198 | UW Virology Lab | UW Virology Lab |
| EPI_ISL_424199 | UW Virology Lab | UW Virology Lab |
| EPI_ISL_424200 | UW Virology Lab | UW Virology Lab |
| EPI_ISL_424201 | UW Virology Lab | UW Virology Lab |
| EPI_ISL_424202 | UW Virology Lab | UW Virology Lab |
| EPI_ISL_424203 | UW Virology Lab | UW Virology Lab |
| EPI_ISL_424204 | UW Virology Lab | UW Virology Lab |
| EPI_ISL_424205 | UW Virology Lab | UW Virology Lab |
| EPI_ISL_424206 | UW Virology Lab | UW Virology Lab |
| EPI_ISL_424207 | UW Virology Lab | UW Virology Lab |
| EPI_ISL_424208 | UW Virology Lab | UW Virology Lab |
| EPI_ISL_424209 | UW Virology Lab | UW Virology Lab |
| EPI_ISL_424210 | UW Virology Lab | UW Virology Lab |
| EPI_ISL_424211 | UW Virology Lab | UW Virology Lab |
| EPI_ISL_424212 | UW Virology Lab | UW Virology Lab |

| EPI_Accession_ID | Originating Laboratory | Submitting Laboratory |
|---|---|---|
| EPI_ISL_424213 | UW Virology Lab | UW Virology Lab |
| EPI_ISL_424214 | UW Virology Lab | UW Virology Lab |
| EPI_ISL_424215 | UW Virology Lab | UW Virology Lab |
| EPI_ISL_424216 | UW Virology Lab | UW Virology Lab |
| EPI_ISL_424217 | UW Virology Lab | UW Virology Lab |
| EPI_ISL_424218 | UW Virology Lab | UW Virology Lab |
| EPI_ISL_424219 | UW Virology Lab | UW Virology Lab |
| EPI_ISL_424220 | UW Virology Lab | UW Virology Lab |
| EPI_ISL_424221 | UW Virology Lab | UW Virology Lab |
| EPI_ISL_424222 | UW Virology Lab | UW Virology Lab |
| EPI_ISL_424223 | UW Virology Lab | UW Virology Lab |
| EPI_ISL_424224 | UW Virology Lab | UW Virology Lab |
| EPI_ISL_424225 | UW Virology Lab | UW Virology Lab |
| EPI_ISL_424226 | UW Virology Lab | UW Virology Lab |
| EPI_ISL_424227 | UW Virology Lab | UW Virology Lab |
| EPI_ISL_424228 | UW Virology Lab | UW Virology Lab |
| EPI_ISL_424229 | UW Virology Lab | UW Virology Lab |
| EPI_ISL_424230 | UW Virology Lab | UW Virology Lab |
| EPI_ISL_424231 | UW Virology Lab | UW Virology Lab |
| EPI_ISL_424232 | UW Virology Lab | UW Virology Lab |
| EPI_ISL_424233 | UW Virology Lab | UW Virology Lab |
| EPI_ISL_424234 | UW Virology Lab | UW Virology Lab |
| EPI_ISL_424235 | UW Virology Lab | UW Virology Lab |
| EPI_ISL_424236 | UW Virology Lab | UW Virology Lab |
| EPI_ISL_424237 | UW Virology Lab | UW Virology Lab |
| EPI_ISL_424238 | UW Virology Lab | UW Virology Lab |
| EPI_ISL_424239 | UW Virology Lab | UW Virology Lab |
| EPI_ISL_424240 | UW Virology Lab | UW Virology Lab |
| EPI_ISL_424241 | UW Virology Lab | UW Virology Lab |
| EPI_ISL_424242 | UW Virology Lab | UW Virology Lab |
| EPI_ISL_424243 | UW Virology Lab | UW Virology Lab |
| EPI_ISL_424244 | UW Virology Lab | UW Virology Lab |
| EPI_ISL_424245 | UW Virology Lab | UW Virology Lab |

| EPI_Accession_ID | Originating Laboratory | Submitting Laboratory |
|---|---|---|
| EPI_ISL_424246 | UW Virology Lab | UW Virology Lab |
| EPI_ISL_424247 | UW Virology Lab | UW Virology Lab |
| EPI_ISL_424249 | UW Virology Lab | UW Virology Lab |
| EPI_ISL_424250 | UW Virology Lab | UW Virology Lab |
| EPI_ISL_424251 | UW Virology Lab | UW Virology Lab |
| EPI_ISL_424252 | UW Virology Lab | UW Virology Lab |
| EPI_ISL_424253 | UW Virology Lab | UW Virology Lab |
| EPI_ISL_424254 | UW Virology Lab | UW Virology Lab |
| EPI_ISL_424255 | UW Virology Lab | UW Virology Lab |
| EPI_ISL_424256 | UW Virology Lab | UW Virology Lab |
| EPI_ISL_424257 | UW Virology Lab | UW Virology Lab |
| EPI_ISL_424258 | UW Virology Lab | UW Virology Lab |
| EPI_ISL_424259 | UW Virology Lab | UW Virology Lab |
| EPI_ISL_424260 | UW Virology Lab | UW Virology Lab |
| EPI_ISL_424261 | UW Virology Lab | UW Virology Lab |
| EPI_ISL_424262 | UW Virology Lab | UW Virology Lab |
| EPI_ISL_424263 | UW Virology Lab | UW Virology Lab |
| EPI_ISL_424264 | UW Virology Lab | UW Virology Lab |
| EPI_ISL_424265 | UW Virology Lab | UW Virology Lab |
| EPI_ISL_424266 | UW Virology Lab | UW Virology Lab |
| EPI_ISL_424267 | UW Virology Lab | UW Virology Lab |
| EPI_ISL_424268 | UW Virology Lab | UW Virology Lab |
| EPI_ISL_424269 | UW Virology Lab | UW Virology Lab |
| EPI_ISL_424270 | UW Virology Lab | UW Virology Lab |
| EPI_ISL_424271 | UW Virology Lab | UW Virology Lab |
| EPI_ISL_424272 | UW Virology Lab | UW Virology Lab |
| EPI_ISL_424273 | UW Virology Lab | UW Virology Lab |
| EPI_ISL_424274 | UW Virology Lab | UW Virology Lab |
| EPI_ISL_424275 | UW Virology Lab | UW Virology Lab |
| EPI_ISL_424276 | UW Virology Lab | UW Virology Lab |
| EPI_ISL_424277 | UW Virology Lab | UW Virology Lab |
| EPI_ISL_424278 | UW Virology Lab | UW Virology Lab |
| EPI_ISL_424279 | UW Virology Lab | UW Virology Lab |

| EPI_Accession_ID | Originating Laboratory | Submitting Laboratory |
|---|---|---|
| EPI_ISL_424280 | UW Virology Lab | UW Virology Lab |
| EPI_ISL_424281 | UW Virology Lab | UW Virology Lab |
| EPI_ISL_424282 | UW Virology Lab | UW Virology Lab |
| EPI_ISL_424283 | UW Virology Lab | UW Virology Lab |
| EPI_ISL_424284 | UW Virology Lab | UW Virology Lab |
| EPI_ISL_424285 | UW Virology Lab | UW Virology Lab |
| EPI_ISL_424286 | UW Virology Lab | UW Virology Lab |
| EPI_ISL_424287 | UW Virology Lab | UW Virology Lab |
| EPI_ISL_424288 | UW Virology Lab | UW Virology Lab |
| EPI_ISL_424289 | UW Virology Lab | UW Virology Lab |
| EPI_ISL_424290 | UW Virology Lab | UW Virology Lab |
| EPI_ISL_424291 | UW Virology Lab | UW Virology Lab |
| EPI_ISL_424292 | UW Virology Lab | UW Virology Lab |
| EPI_ISL_424293 | UW Virology Lab | UW Virology Lab |
| EPI_ISL_424294 | UW Virology Lab | UW Virology Lab |
| EPI_ISL_424295 | UW Virology Lab | UW Virology Lab |
| EPI_ISL_424296 | UW Virology Lab | UW Virology Lab |
| EPI_ISL_424297 | UW Virology Lab | UW Virology Lab |
| EPI_ISL_424298 | UW Virology Lab | UW Virology Lab |
| EPI_ISL_424299 | UW Virology Lab | UW Virology Lab |
| EPI_ISL_424300 | UW Virology Lab | UW Virology Lab |
| EPI_ISL_424301 | UW Virology Lab | UW Virology Lab |
| EPI_ISL_424302 | UW Virology Lab | UW Virology Lab |
| EPI_ISL_424303 | UW Virology Lab | UW Virology Lab |
| EPI_ISL_424304 | UW Virology Lab | UW Virology Lab |
| EPI_ISL_424305 | UW Virology Lab | UW Virology Lab |
| EPI_ISL_424306 | UW Virology Lab | UW Virology Lab |
| EPI_ISL_424307 | UW Virology Lab | UW Virology Lab |
| EPI_ISL_424308 | UW Virology Lab | UW Virology Lab |
| EPI_ISL_424309 | UW Virology Lab | UW Virology Lab |
| EPI_ISL_424310 | UW Virology Lab | UW Virology Lab |
| EPI_ISL_424311 | UW Virology Lab | UW Virology Lab |
| EPI_ISL_424312 | UW Virology Lab | UW Virology Lab |

| EPI_Accession_ID | Originating Laboratory | Submitting Laboratory |
|---|---|---|
| EPI_ISL_424313 | UW Virology Lab | UW Virology Lab |
| EPI_ISL_424314 | UW Virology Lab | UW Virology Lab |
| EPI_ISL_424315 | UW Virology Lab | UW Virology Lab |
| EPI_ISL_424316 | UW Virology Lab | UW Virology Lab |
| EPI_ISL_424317 | UW Virology Lab | UW Virology Lab |
| EPI_ISL_424318 | UW Virology Lab | UW Virology Lab |
| EPI_ISL_424319 | UW Virology Lab | UW Virology Lab |
| EPI_ISL_424320 | UW Virology Lab | UW Virology Lab |
| EPI_ISL_424321 | UW Virology Lab | UW Virology Lab |
| EPI_ISL_424322 | UW Virology Lab | UW Virology Lab |
| EPI_ISL_424323 | UW Virology Lab | UW Virology Lab |
| EPI_ISL_424324 | UW Virology Lab | UW Virology Lab |
| EPI_ISL_424325 | UW Virology Lab | UW Virology Lab |
| EPI_ISL_424326 | UW Virology Lab | UW Virology Lab |
| EPI_ISL_424327 | UW Virology Lab | UW Virology Lab |
| EPI_ISL_424328 | UW Virology Lab | UW Virology Lab |
| EPI_ISL_424329 | UW Virology Lab | UW Virology Lab |
| EPI_ISL_424330 | UW Virology Lab | UW Virology Lab |
| EPI_ISL_424331 | UW Virology Lab | UW Virology Lab |
| EPI_ISL_424332 | UW Virology Lab | UW Virology Lab |
| EPI_ISL_424333 | UW Virology Lab | UW Virology Lab |
| EPI_ISL_424334 | UW Virology Lab | UW Virology Lab |
| EPI_ISL_424335 | UW Virology Lab | UW Virology Lab |
| EPI_ISL_424336 | UW Virology Lab | UW Virology Lab |
| EPI_ISL_424337 | UW Virology Lab | UW Virology Lab |
| EPI_ISL_424338 | UW Virology Lab | UW Virology Lab |
| EPI_ISL_424339 | UW Virology Lab | UW Virology Lab |
| EPI_ISL_424340 | UW Virology Lab | UW Virology Lab |
| EPI_ISL_424341 | UW Virology Lab | UW Virology Lab |
| EPI_ISL_424342 | INMI Lazzaro Spallanzani IRCCS | Laboratory of Virology, INMI Lazzaro Spallanzani IRCCS |
| EPI_ISL_424343 | INMI Lazzaro Spallanzani IRCCS | Laboratory of Virology, INMI Lazzaro Spallanzani IRCCS |
| EPI_ISL_424344 | INMI Lazzaro Spallanzani IRCCS | Laboratory of Virology, INMI Lazzaro Spallanzani IRCCS |
| EPI_ISL_424345 | Instituto Nacional de Enfermedades Respiratorias | Instituto Nacional de Enfermedades Respiratorias |

| EPI_Accession_ID | Originating Laboratory | Submitting Laboratory |
|---|---|---|
| EPI_ISL_424346 | Alaska State Virology Laboratory | Alaska State Virology Laboratory |
| EPI_ISL_424347 | Alaska State Virology Laboratory | Alaska State Virology Laboratory |
| EPI_ISL_424348 | Instituto Nacional de Enfermedades Respiratorias | Instituto Nacional de Enfermedades Respiratorias |
| EPI_ISL_424349 | unknown | Human Genetic Research Center |
| EPI_ISL_424352 | unknown | Clinical Laboratory |
| EPI_ISL_424355 | unknown | Beijing Institute of Microbiology and Epidemiology |
| EPI_ISL_424356 | unknown | Beijing Institute of Microbiology and Epidemiology |
| EPI_ISL_424357 | unknown | Beijing Institute of Microbiology and Epidemiology |
| EPI_ISL_424358 | unknown | Beijing Institute of Microbiology and Epidemiology |
| EPI_ISL_424359 | unknown | Beijing Institute of Microbiology and Epidemiology |
| EPI_ISL_424360 | unknown | Beijing Institute of Microbiology and Epidemiology |
| EPI_ISL_424361 | National Influenza Center, Indian Council of Medical Research - National Institute of Virology | Indian Council of Medical Research-National Institute of Virology, Microbial Containment Complex |
| EPI_ISL_424362 | National Influenza Center, Indian Council of Medical Research - National Institute of Virology | Indian Council of Medical Research-National Institute of Virology, Microbial Containment Complex |
| EPI_ISL_424363 | National Influenza Center, Indian Council of Medical Research - National Institute of Virology | Indian Council of Medical Research-National Institute of Virology, Microbial Containment Complex |
| EPI_ISL_424364 | National Influenza Center, Indian Council of Medical Research - National Institute of Virology | Indian Council of Medical Research-National Institute of Virology, Microbial Containment Complex |
| EPI_ISL_424365 | National Influenza Center, Indian Council of Medical Research - National Institute of Virology | Indian Council of Medical Research-National Institute of Virology, Microbial Containment Complex |
| EPI_ISL_424366 | Vaccine Research, Development and Application Center, Erciyes University | Gen Era Diagnostics Inc. |
| EPI_ISL_424377 | deCODE genetics | deCODE genetics |
| EPI_ISL_424378 | deCODE genetics | deCODE genetics |
| EPI_ISL_424379 | The National University Hospital of Iceland | deCODE genetics |
| EPI_ISL_424380 | The National University Hospital of Iceland | deCODE genetics |
| EPI_ISL_424381 | The National University Hospital of Iceland | deCODE genetics |
| EPI_ISL_424382 | The National University Hospital of Iceland | deCODE genetics |
| EPI_ISL_424383 | The National University Hospital of Iceland | deCODE genetics |
| EPI_ISL_424384 | The National University Hospital of Iceland | deCODE genetics |
| EPI_ISL_424385 | The National University Hospital of Iceland | deCODE genetics |
| EPI_ISL_424386 | The National University Hospital of Iceland | deCODE genetics |

| EPI_Accession_ID | Originating Laboratory | Submitting Laboratory |
|---|---|---|
| EPI_ISL_424387 | The National University Hospital of Iceland | deCODE genetics |
| EPI_ISL_424388 | The National University Hospital of Iceland | deCODE genetics |
| EPI_ISL_424389 | The National University Hospital of Iceland | deCODE genetics |
| EPI_ISL_424390 | The National University Hospital of Iceland | deCODE genetics |
| EPI_ISL_424391 | The National University Hospital of Iceland | deCODE genetics |
| EPI_ISL_424392 | The National University Hospital of Iceland | deCODE genetics |
| EPI_ISL_424393 | The National University Hospital of Iceland | deCODE genetics |
| EPI_ISL_424394 | The National University Hospital of Iceland | deCODE genetics |
| EPI_ISL_424395 | The National University Hospital of Iceland | deCODE genetics |
| EPI_ISL_424396 | The National University Hospital of Iceland | deCODE genetics |
| EPI_ISL_424397 | The National University Hospital of Iceland | deCODE genetics |
| EPI_ISL_424398 | The National University Hospital of Iceland | deCODE genetics |
| EPI_ISL_424399 | The National University Hospital of Iceland | deCODE genetics |
| EPI_ISL_424400 | The National University Hospital of Iceland | deCODE genetics |
| EPI_ISL_424401 | The National University Hospital of Iceland | deCODE genetics |
| EPI_ISL_424402 | The National University Hospital of Iceland | deCODE genetics |
| EPI_ISL_424403 | The National University Hospital of Iceland | deCODE genetics |
| EPI_ISL_424404 | The National University Hospital of Iceland | deCODE genetics |
| EPI_ISL_424405 | The National University Hospital of Iceland | deCODE genetics |
| EPI_ISL_424406 | The National University Hospital of Iceland | deCODE genetics |
| EPI_ISL_424407 | The National University Hospital of Iceland | deCODE genetics |
| EPI_ISL_424409 | The National University Hospital of Iceland | deCODE genetics |
| EPI_ISL_424410 | The National University Hospital of Iceland | deCODE genetics |
| EPI_ISL_424411 | The National University Hospital of Iceland | deCODE genetics |
| EPI_ISL_424412 | The National University Hospital of Iceland | deCODE genetics |
| EPI_ISL_424413 | The National University Hospital of Iceland | deCODE genetics |
| EPI_ISL_424414 | The National University Hospital of Iceland | deCODE genetics |
| EPI_ISL_424415 | The National University Hospital of Iceland | deCODE genetics |
| EPI_ISL_424416 | The National University Hospital of Iceland | deCODE genetics |
| EPI_ISL_424417 | The National University Hospital of Iceland | deCODE genetics |
| EPI_ISL_424418 | The National University Hospital of Iceland | deCODE genetics |
| EPI_ISL_424419 | The National University Hospital of Iceland | deCODE genetics |
| EPI_ISL_424420 | The National University Hospital of Iceland | deCODE genetics |

| EPI_Accession_ID | Originating Laboratory | Submitting Laboratory |
|---|---|---|
| EPI_ISL_424421 | The National University Hospital of Iceland | deCODE genetics |
| EPI_ISL_424422 | The National University Hospital of Iceland | deCODE genetics |
| EPI_ISL_424423 | The National University Hospital of Iceland | deCODE genetics |
| EPI_ISL_424424 | The National University Hospital of Iceland | deCODE genetics |
| EPI_ISL_424425 | The National University Hospital of Iceland | deCODE genetics |
| EPI_ISL_424426 | The National University Hospital of Iceland | deCODE genetics |
| EPI_ISL_424427 | The National University Hospital of Iceland | deCODE genetics |
| EPI_ISL_424428 | The National University Hospital of Iceland | deCODE genetics |
| EPI_ISL_424429 | The National University Hospital of Iceland | deCODE genetics |
| EPI_ISL_424430 | The National University Hospital of Iceland | deCODE genetics |
| EPI_ISL_424431 | The National University Hospital of Iceland | deCODE genetics |
| EPI_ISL_424432 | The National University Hospital of Iceland | deCODE genetics |
| EPI_ISL_424433 | The National University Hospital of Iceland | deCODE genetics |
| EPI_ISL_424434 | The National University Hospital of Iceland | deCODE genetics |
| EPI_ISL_424435 | The National University Hospital of Iceland | deCODE genetics |
| EPI_ISL_424436 | The National University Hospital of Iceland | deCODE genetics |
| EPI_ISL_424437 | The National University Hospital of Iceland | deCODE genetics |
| EPI_ISL_424438 | The National University Hospital of Iceland | deCODE genetics |
| EPI_ISL_424439 | The National University Hospital of Iceland | deCODE genetics |
| EPI_ISL_424440 | The National University Hospital of Iceland | deCODE genetics |
| EPI_ISL_424441 | The National University Hospital of Iceland | deCODE genetics |
| EPI_ISL_424442 | The National University Hospital of Iceland | deCODE genetics |
| EPI_ISL_424443 | The National University Hospital of Iceland | deCODE genetics |
| EPI_ISL_424444 | The National University Hospital of Iceland | deCODE genetics |
| EPI_ISL_424445 | The National University Hospital of Iceland | deCODE genetics |
| EPI_ISL_424446 | The National University Hospital of Iceland | deCODE genetics |
| EPI_ISL_424447 | The National University Hospital of Iceland | deCODE genetics |
| EPI_ISL_424448 | The National University Hospital of Iceland | deCODE genetics |
| EPI_ISL_424449 | The National University Hospital of Iceland | deCODE genetics |
| EPI_ISL_424450 | The National University Hospital of Iceland | deCODE genetics |
| EPI_ISL_424451 | The National University Hospital of Iceland | deCODE genetics |
| EPI_ISL_424452 | The National University Hospital of Iceland | deCODE genetics |
| EPI_ISL_424453 | The National University Hospital of Iceland | deCODE genetics |

| EPI_Accession_ID | Originating Laboratory | Submitting Laboratory |
|---|---|---|
| EPI_ISL_424454 | The National University Hospital of Iceland | deCODE genetics |
| EPI_ISL_424455 | The National University Hospital of Iceland | deCODE genetics |
| EPI_ISL_424456 | The National University Hospital of Iceland | deCODE genetics |
| EPI_ISL_424457 | The National University Hospital of Iceland | deCODE genetics |
| EPI_ISL_424458 | The National University Hospital of Iceland | deCODE genetics |
| EPI_ISL_424459 | The National University Hospital of Iceland | deCODE genetics |
| EPI_ISL_424460 | The National University Hospital of Iceland | deCODE genetics |
| EPI_ISL_424461 | The National University Hospital of Iceland | deCODE genetics |
| EPI_ISL_424462 | The National University Hospital of Iceland | deCODE genetics |
| EPI_ISL_424463 | The National University Hospital of Iceland | deCODE genetics |
| EPI_ISL_424464 | The National University Hospital of Iceland | deCODE genetics |
| EPI_ISL_424465 | The National University Hospital of Iceland | deCODE genetics |
| EPI_ISL_424466 | The National University Hospital of Iceland | deCODE genetics |
| EPI_ISL_424467 | The National University Hospital of Iceland | deCODE genetics |
| EPI_ISL_424468 | The National University Hospital of Iceland | deCODE genetics |
| EPI_ISL_424469 | The National University Hospital of Iceland | deCODE genetics |
| EPI_ISL_424470 | The National University Hospital of Iceland | deCODE genetics |
| EPI_ISL_424471 | The National University Hospital of Iceland | deCODE genetics |
| EPI_ISL_424472 | The National University Hospital of Iceland | deCODE genetics |
| EPI_ISL_424473 | The National University Hospital of Iceland | deCODE genetics |
| EPI_ISL_424474 | The National University Hospital of Iceland | deCODE genetics |
| EPI_ISL_424475 | The National University Hospital of Iceland | deCODE genetics |
| EPI_ISL_424476 | The National University Hospital of Iceland | deCODE genetics |
| EPI_ISL_424477 | The National University Hospital of Iceland | deCODE genetics |
| EPI_ISL_424478 | The National University Hospital of Iceland | deCODE genetics |
| EPI_ISL_424479 | The National University Hospital of Iceland | deCODE genetics |
| EPI_ISL_424480 | The National University Hospital of Iceland | deCODE genetics |
| EPI_ISL_424481 | The National University Hospital of Iceland | deCODE genetics |
| EPI_ISL_424482 | The National University Hospital of Iceland | deCODE genetics |
| EPI_ISL_424483 | The National University Hospital of Iceland | deCODE genetics |
| EPI_ISL_424484 | The National University Hospital of Iceland | deCODE genetics |
| EPI_ISL_424485 | The National University Hospital of Iceland | deCODE genetics |
| EPI_ISL_424486 | The National University Hospital of Iceland | deCODE genetics |

| EPI_Accession_ID | Originating Laboratory | Submitting Laboratory |
| --- | --- | --- |
| EPI_ISL_424487 | The National University Hospital of Iceland | deCODE genetics |
| EPI_ISL_424488 | The National University Hospital of Iceland | deCODE genetics |
| EPI_ISL_424489 | The National University Hospital of Iceland | deCODE genetics |
| EPI_ISL_424490 | The National University Hospital of Iceland | deCODE genetics |
| EPI_ISL_424491 | The National University Hospital of Iceland | deCODE genetics |
| EPI_ISL_424492 | The National University Hospital of Iceland | deCODE genetics |
| EPI_ISL_424493 | The National University Hospital of Iceland | deCODE genetics |
| EPI_ISL_424494 | The National University Hospital of Iceland | deCODE genetics |
| EPI_ISL_424495 | The National University Hospital of Iceland | deCODE genetics |
| EPI_ISL_424496 | The National University Hospital of Iceland | deCODE genetics |
| EPI_ISL_424497 | The National University Hospital of Iceland | deCODE genetics |
| EPI_ISL_424498 | The National University Hospital of Iceland | deCODE genetics |
| EPI_ISL_424499 | The National University Hospital of Iceland | deCODE genetics |
| EPI_ISL_424500 | The National University Hospital of Iceland | deCODE genetics |
| EPI_ISL_424501 | The National University Hospital of Iceland | deCODE genetics |
| EPI_ISL_424502 | The National University Hospital of Iceland | deCODE genetics |
| EPI_ISL_424503 | The National University Hospital of Iceland | deCODE genetics |
| EPI_ISL_424505 | The National University Hospital of Iceland | deCODE genetics |
| EPI_ISL_424506 | The National University Hospital of Iceland | deCODE genetics |
| EPI_ISL_424507 | The National University Hospital of Iceland | deCODE genetics |
| EPI_ISL_424508 | The National University Hospital of Iceland | deCODE genetics |
| EPI_ISL_424509 | The National University Hospital of Iceland | deCODE genetics |
| EPI_ISL_424510 | The National University Hospital of Iceland | deCODE genetics |
| EPI_ISL_424511 | The National University Hospital of Iceland | deCODE genetics |
| EPI_ISL_424512 | The National University Hospital of Iceland | deCODE genetics |
| EPI_ISL_424513 | The National University Hospital of Iceland | deCODE genetics |
| EPI_ISL_424514 | The National University Hospital of Iceland | deCODE genetics |
| EPI_ISL_424515 | The National University Hospital of Iceland | deCODE genetics |
| EPI_ISL_424516 | The National University Hospital of Iceland | deCODE genetics |
| EPI_ISL_424517 | The National University Hospital of Iceland | deCODE genetics |
| EPI_ISL_424518 | The National University Hospital of Iceland | deCODE genetics |
| EPI_ISL_424519 | The National University Hospital of Iceland | deCODE genetics |
| EPI_ISL_424520 | The National University Hospital of Iceland | deCODE genetics |

| EPI_Accession_ID | Originating Laboratory | Submitting Laboratory |
|---|---|---|
| EPI_ISL_424521 | The National University Hospital of Iceland | deCODE genetics |
| EPI_ISL_424522 | The National University Hospital of Iceland | deCODE genetics |
| EPI_ISL_424523 | The National University Hospital of Iceland | deCODE genetics |
| EPI_ISL_424524 | The National University Hospital of Iceland | deCODE genetics |
| EPI_ISL_424525 | The National University Hospital of Iceland | deCODE genetics |
| EPI_ISL_424526 | The National University Hospital of Iceland | deCODE genetics |
| EPI_ISL_424527 | The National University Hospital of Iceland | deCODE genetics |
| EPI_ISL_424528 | The National University Hospital of Iceland | deCODE genetics |
| EPI_ISL_424529 | The National University Hospital of Iceland | deCODE genetics |
| EPI_ISL_424530 | The National University Hospital of Iceland | deCODE genetics |
| EPI_ISL_424531 | The National University Hospital of Iceland | deCODE genetics |
| EPI_ISL_424532 | The National University Hospital of Iceland | deCODE genetics |
| EPI_ISL_424533 | deCODE genetics | deCODE genetics |
| EPI_ISL_424535 | deCODE genetics | deCODE genetics |
| EPI_ISL_424536 | deCODE genetics | deCODE genetics |
| EPI_ISL_424538 | deCODE genetics | deCODE genetics |
| EPI_ISL_424539 | deCODE genetics | deCODE genetics |
| EPI_ISL_424543 | deCODE genetics | deCODE genetics |
| EPI_ISL_424544 | deCODE genetics | deCODE genetics |
| EPI_ISL_424545 | deCODE genetics | deCODE genetics |
| EPI_ISL_424546 | deCODE genetics | deCODE genetics |
| EPI_ISL_424551 | deCODE genetics | deCODE genetics |
| EPI_ISL_424552 | The National University Hospital of Iceland | deCODE genetics |
| EPI_ISL_424554 | The National University Hospital of Iceland | deCODE genetics |
| EPI_ISL_424555 | The National University Hospital of Iceland | deCODE genetics |
| EPI_ISL_424556 | The National University Hospital of Iceland | deCODE genetics |
| EPI_ISL_424557 | The National University Hospital of Iceland | deCODE genetics |
| EPI_ISL_424558 | The National University Hospital of Iceland | deCODE genetics |
| EPI_ISL_424559 | The National University Hospital of Iceland | deCODE genetics |
| EPI_ISL_424560 | deCODE genetics | deCODE genetics |
| EPI_ISL_424561 | deCODE genetics | deCODE genetics |
| EPI_ISL_424562 | deCODE genetics | deCODE genetics |
| EPI_ISL_424563 | The National University Hospital of Iceland | deCODE genetics |

| EPI_Accession_ID | Originating Laboratory | Submitting Laboratory |
|---|---|---|
| EPI_ISL_424564 | The National University Hospital of Iceland | deCODE genetics |
| EPI_ISL_424566 | deCODE genetics | deCODE genetics |
| EPI_ISL_424567 | deCODE genetics | deCODE genetics |
| EPI_ISL_424568 | deCODE genetics | deCODE genetics |
| EPI_ISL_424570 | deCODE genetics | deCODE genetics |
| EPI_ISL_424571 | The National University Hospital of Iceland | deCODE genetics |
| EPI_ISL_424572 | The National University Hospital of Iceland | deCODE genetics |
| EPI_ISL_424573 | The National University Hospital of Iceland | deCODE genetics |
| EPI_ISL_424574 | The National University Hospital of Iceland | deCODE genetics |
| EPI_ISL_424575 | The National University Hospital of Iceland | deCODE genetics |
| EPI_ISL_424576 | The National University Hospital of Iceland | deCODE genetics |
| EPI_ISL_424577 | The National University Hospital of Iceland | deCODE genetics |
| EPI_ISL_424578 | The National University Hospital of Iceland | deCODE genetics |
| EPI_ISL_424579 | The National University Hospital of Iceland | deCODE genetics |
| EPI_ISL_424580 | The National University Hospital of Iceland | deCODE genetics |
| EPI_ISL_424581 | The National University Hospital of Iceland | deCODE genetics |
| EPI_ISL_424582 | The National University Hospital of Iceland | deCODE genetics |
| EPI_ISL_424583 | The National University Hospital of Iceland | deCODE genetics |
| EPI_ISL_424584 | The National University Hospital of Iceland | deCODE genetics |
| EPI_ISL_424585 | The National University Hospital of Iceland | deCODE genetics |
| EPI_ISL_424586 | The National University Hospital of Iceland | deCODE genetics |
| EPI_ISL_424588 | The National University Hospital of Iceland | deCODE genetics |
| EPI_ISL_424589 | The National University Hospital of Iceland | deCODE genetics |
| EPI_ISL_424590 | The National University Hospital of Iceland | deCODE genetics |
| EPI_ISL_424591 | The National University Hospital of Iceland | deCODE genetics |
| EPI_ISL_424592 | The National University Hospital of Iceland | deCODE genetics |
| EPI_ISL_424593 | The National University Hospital of Iceland | deCODE genetics |
| EPI_ISL_424595 | The National University Hospital of Iceland | deCODE genetics |
| EPI_ISL_424596 | The National University Hospital of Iceland | deCODE genetics |
| EPI_ISL_424597 | The National University Hospital of Iceland | deCODE genetics |
| EPI_ISL_424598 | The National University Hospital of Iceland | deCODE genetics |
| EPI_ISL_424599 | The National University Hospital of Iceland | deCODE genetics |
| EPI_ISL_424600 | The National University Hospital of Iceland | deCODE genetics |

| EPI_Accession_ID | Originating Laboratory | Submitting Laboratory |
|---|---|---|
| EPI_ISL_424601 | The National University Hospital of Iceland | deCODE genetics |
| EPI_ISL_424602 | The National University Hospital of Iceland | deCODE genetics |
| EPI_ISL_424603 | The National University Hospital of Iceland | deCODE genetics |
| EPI_ISL_424604 | The National University Hospital of Iceland | deCODE genetics |
| EPI_ISL_424605 | The National University Hospital of Iceland | deCODE genetics |
| EPI_ISL_424606 | The National University Hospital of Iceland | deCODE genetics |
| EPI_ISL_424608 | deCODE genetics | deCODE genetics |
| EPI_ISL_424609 | deCODE genetics | deCODE genetics |
| EPI_ISL_424610 | The National University Hospital of Iceland | deCODE genetics |
| EPI_ISL_424612 | The National University Hospital of Iceland | deCODE genetics |
| EPI_ISL_424613 | The National University Hospital of Iceland | deCODE genetics |
| EPI_ISL_424614 | The National University Hospital of Iceland | deCODE genetics |
| EPI_ISL_424615 | The National University Hospital of Iceland | deCODE genetics |
| EPI_ISL_424616 | The National University Hospital of Iceland | deCODE genetics |
| EPI_ISL_424617 | The National University Hospital of Iceland | deCODE genetics |
| EPI_ISL_424618 | The National University Hospital of Iceland | deCODE genetics |
| EPI_ISL_424619 | The National University Hospital of Iceland | deCODE genetics |
| EPI_ISL_424620 | The National University Hospital of Iceland | deCODE genetics |
| EPI_ISL_424621 | The National University Hospital of Iceland | deCODE genetics |
| EPI_ISL_424622 | The National University Hospital of Iceland | deCODE genetics |
| EPI_ISL_424623 | The National University Hospital of Iceland | deCODE genetics |
| EPI_ISL_424624 | The National University Hospital of Iceland | deCODE genetics |
| EPI_ISL_424626 | Instituto Nacional de Enfermedades Respiratorias | Instituto Nacional de Enfermedades Respiratorias |
| EPI_ISL_424627 | Instituto Nacional de Enfermedades Respiratorias | Instituto Nacional de Enfermedades Respiratorias |
| EPI_ISL_424628 | Department of Clinical Microbiology | GIGA Medical Genomics |
| EPI_ISL_424629 | Department of Clinical Microbiology | GIGA Medical Genomics |
| EPI_ISL_424630 | Department of Clinical Microbiology | GIGA Medical Genomics |
| EPI_ISL_424631 | Department of Clinical Microbiology | GIGA Medical Genomics |
| EPI_ISL_424632 | Department of Clinical Microbiology | GIGA Medical Genomics |
| EPI_ISL_424633 | Department of Clinical Microbiology | GIGA Medical Genomics |
| EPI_ISL_424634 | Department of Clinical Microbiology | GIGA Medical Genomics |
| EPI_ISL_424635 | Department of Clinical Microbiology | GIGA Medical Genomics |
| EPI_ISL_424636 | Department of Clinical Microbiology | GIGA Medical Genomics |

| EPI_Accession_ID | Originating Laboratory | Submitting Laboratory |
|---|---|---|
| EPI_ISL_424637 | Department of Clinical Microbiology | GIGA Medical Genomics |
| EPI_ISL_424638 | Department of Clinical Microbiology | GIGA Medical Genomics |
| EPI_ISL_424639 | Department of Clinical Microbiology | GIGA Medical Genomics |
| EPI_ISL_424640 | Department of Clinical Microbiology | GIGA Medical Genomics |
| EPI_ISL_424641 | Department of Clinical Microbiology | GIGA Medical Genomics |
| EPI_ISL_424642 | Department of Clinical Microbiology | GIGA Medical Genomics |
| EPI_ISL_424643 | Department of Clinical Microbiology | GIGA Medical Genomics |
| EPI_ISL_424644 | Department of Clinical Microbiology | GIGA Medical Genomics |
| EPI_ISL_424645 | Department of Clinical Microbiology | GIGA Medical Genomics |
| EPI_ISL_424646 | Department of Clinical Microbiology | GIGA Medical Genomics |
| EPI_ISL_424647 | Department of Clinical Microbiology | GIGA Medical Genomics |
| EPI_ISL_424648 | Department of Clinical Microbiology | GIGA Medical Genomics |
| EPI_ISL_424649 | Department of Clinical Microbiology | GIGA Medical Genomics |
| EPI_ISL_424650 | Department of Clinical Microbiology | GIGA Medical Genomics |
| EPI_ISL_424651 | Department of Clinical Microbiology | GIGA Medical Genomics |
| EPI_ISL_424652 | Department of Clinical Microbiology | GIGA Medical Genomics |
| EPI_ISL_424653 | Department of Clinical Microbiology | GIGA Medical Genomics |
| EPI_ISL_424654 | Department of Clinical Microbiology | GIGA Medical Genomics |
| EPI_ISL_424655 | Department of Clinical Microbiology | GIGA Medical Genomics |
| EPI_ISL_424656 | Department of Clinical Microbiology | GIGA Medical Genomics |
| EPI_ISL_424657 | Department of Clinical Microbiology | GIGA Medical Genomics |
| EPI_ISL_424658 | Department of Clinical Microbiology | GIGA Medical Genomics |
| EPI_ISL_424659 | Department of Clinical Microbiology | GIGA Medical Genomics |
| EPI_ISL_424660 | Department of Clinical Microbiology | GIGA Medical Genomics |
| EPI_ISL_424661 | Department of Clinical Microbiology | GIGA Medical Genomics |
| EPI_ISL_424662 | Department of Clinical Microbiology | GIGA Medical Genomics |
| EPI_ISL_424663 | Department of Clinical Microbiology | GIGA Medical Genomics |
| EPI_ISL_424664 | Department of Clinical Microbiology | GIGA Medical Genomics |
| EPI_ISL_424665 | Department of Clinical Microbiology | GIGA Medical Genomics |
| EPI_ISL_424667 | Laboratorio Estatal de Salud Publica del Estado de MÃ©xico | Instituto de DiagnÃ³stico y Referencia EpidemiolÃ³gicos |
| EPI_ISL_424668 | Arizona State University Health Services | Arizona State University |
| EPI_ISL_424669 | Arizona State University Health Services | Arizona State University |

| EPI_Accession_ID | Originating Laboratory | Submitting Laboratory |
|---|---|---|
| EPI_ISL_424670 | Laboratorio Estatal de Salud Publica del Estado de Queretaro | Instituto de DiagnÃ³stico y Referencia Epidemiologicos |
| EPI_ISL_424671 | Arizona State University Health Services | Arizona State University |
| EPI_ISL_424672 | Laboratorio Estatal de Salud Publica del Estado de Puebla | Instituto de Diagnostico y Referencia Epidemiologicos |
| EPI_ISL_424673 | Instituto de Diagnostico y Referencia Epidemiologicos | Instituto de Diagnostico y Referencia Epidemiologicos |
| EPI_ISL_424703 | Klinisk mikrobiologi, Region VÃ¤sterbotten | Unit for Biological Agents, Department for CBRN Defence and Security, Swedish Defence Research Agency |
| EPI_ISL_424731 | Hospital General Regional No.66, Ciudad JuÃ¡rez, Chihuahua. | Laboratorio Central de EpidemiologÃ-a-DLVIE / Laboratorio de SecuenciaciÃ³n-Centro de Instrumentos. Instituto Mexicano del Seguro Social |
| EPI_ISL_424841 | SC Dept of Health and Env. Control-Bureau of Laboratories | Pathogen Discovery, Respiratory Viruses Branch, Division of Viral Diseases, Centers for Disease Control and Prevention |
| EPI_ISL_424842 | SC Dept of Health and Env. Control-Bureau of Laboratories | Pathogen Discovery, Respiratory Viruses Branch, Division of Viral Diseases, Centers for Disease Control and Prevention |
| EPI_ISL_424843 | MA State Public Health Laboratory | Pathogen Discovery, Respiratory Viruses Branch, Division of Viral Diseases, Centers for Disease Control and Prevention |
| EPI_ISL_424844 | MA State Public Health Laboratory | Pathogen Discovery, Respiratory Viruses Branch, Division of Viral Diseases, Centers for Disease Control and Prevention |
| EPI_ISL_424845 | MA State Public Health Laboratory | Pathogen Discovery, Respiratory Viruses Branch, Division of Viral Diseases, Centers for Disease Control and Prevention |
| EPI_ISL_424846 | MA State Public Health Laboratory | Pathogen Discovery, Respiratory Viruses Branch, Division of Viral Diseases, Centers for Disease Control and Prevention |
| EPI_ISL_424847 | MA State Public Health Laboratory | Pathogen Discovery, Respiratory Viruses Branch, Division of Viral Diseases, Centers for Disease Control and Prevention |

| EPI_Accession_ID | Originating Laboratory | Submitting Laboratory |
|---|---|---|
| EPI_ISL_424848 | AZ SPHL, Arizona Department of Health Services | Pathogen Discovery, Respiratory Viruses Branch, Division of Viral Diseases, Centers for Disease Control and Prevention |
| EPI_ISL_424849 | AZ SPHL, Arizona Department of Health Services | Pathogen Discovery, Respiratory Viruses Branch, Division of Viral Diseases, Centers for Disease Control and Prevention |
| EPI_ISL_424850 | IL Department of Public Health Chicago Laboratory | Pathogen Discovery, Respiratory Viruses Branch, Division of Viral Diseases, Centers for Disease Control and Prevention |
| EPI_ISL_424851 | IL Department of Public Health Chicago Laboratory | Pathogen Discovery, Respiratory Viruses Branch, Division of Viral Diseases, Centers for Disease Control and Prevention |
| EPI_ISL_424852 | DC Public Health Lab/ Dept. of Forensic Sciences | Pathogen Discovery, Respiratory Viruses Branch, Division of Viral Diseases, Centers for Disease Control and Prevention |
| EPI_ISL_424853 | FL Bureau of Public Health Laboratories-Miami | Pathogen Discovery, Respiratory Viruses Branch, Division of Viral Diseases, Centers for Disease Control and Prevention |
| EPI_ISL_424854 | FL Bureau of Public Health Laboratories-Miami | Pathogen Discovery, Respiratory Viruses Branch, Division of Viral Diseases, Centers for Disease Control and Prevention |
| EPI_ISL_424855 | FL Bureau of Public Health Laboratories-Tampa | Pathogen Discovery, Respiratory Viruses Branch, Division of Viral Diseases, Centers for Disease Control and Prevention |
| EPI_ISL_424856 | FL Bur. of Public Health Laboratories-Jacksonville | Pathogen Discovery, Respiratory Viruses Branch, Division of Viral Diseases, Centers for Disease Control and Prevention |
| EPI_ISL_424857 | FL Bur. of Public Health Laboratories-Jacksonville | Pathogen Discovery, Respiratory Viruses Branch, Division of Viral Diseases, Centers for Disease Control and Prevention |
| EPI_ISL_424858 | GA Department of Public Health Laboratory | Pathogen Discovery, Respiratory Viruses Branch, Division of Viral Diseases, Centers for Disease Control and Prevention |

| EPI_Accession_ID | Originating Laboratory | Submitting Laboratory |
|---|---|---|
| EPI_ISL_424859 | GA Department of Public Health Laboratory | Pathogen Discovery, Respiratory Viruses Branch, Division of Viral Diseases, Centers for Disease Control and Prevention |
| EPI_ISL_424861 | GA Department of Public Health Laboratory | Pathogen Discovery, Respiratory Viruses Branch, Division of Viral Diseases, Centers for Disease Control and Prevention |
| EPI_ISL_424864 | GA Department of Public Health Laboratory | Pathogen Discovery, Respiratory Viruses Branch, Division of Viral Diseases, Centers for Disease Control and Prevention |
| EPI_ISL_424865 | IA State Hygienic Laboratory | Pathogen Discovery, Respiratory Viruses Branch, Division of Viral Diseases, Centers for Disease Control and Prevention |
| EPI_ISL_424866 | IN State Department of Health Laboratory Services | Pathogen Discovery, Respiratory Viruses Branch, Division of Viral Diseases, Centers for Disease Control and Prevention |
| EPI_ISL_424867 | KS Health and Environmental Laboratories | Pathogen Discovery, Respiratory Viruses Branch, Division of Viral Diseases, Centers for Disease Control and Prevention |
| EPI_ISL_424868 | LA Office of Public Health Laboratories | Pathogen Discovery, Respiratory Viruses Branch, Division of Viral Diseases, Centers for Disease Control and Prevention |
| EPI_ISL_424869 | MD DOH Laboratories Administration | Pathogen Discovery, Respiratory Viruses Branch, Division of Viral Diseases, Centers for Disease Control and Prevention |
| EPI_ISL_424870 | MO State Public Health Laboratory | Pathogen Discovery, Respiratory Viruses Branch, Division of Viral Diseases, Centers for Disease Control and Prevention |
| EPI_ISL_424871 | NC State Laboratory of Public Health | Pathogen Discovery, Respiratory Viruses Branch, Division of Viral Diseases, Centers for Disease Control and Prevention |
| EPI_ISL_424872 | NC State Laboratory of Public Health | Pathogen Discovery, Respiratory Viruses Branch, Division of Viral Diseases, Centers for Disease Control and Prevention |

| EPI_Accession_ID | Originating Laboratory | Submitting Laboratory |
|---|---|---|
| EPI_ISL_424873 | NC State Laboratory of Public Health | Pathogen Discovery, Respiratory Viruses Branch, Division of Viral Diseases, Centers for Disease Control and Prevention |
| EPI_ISL_424874 | NE Public Health Laboratory | Pathogen Discovery, Respiratory Viruses Branch, Division of Viral Diseases, Centers for Disease Control and Prevention |
| EPI_ISL_424875 | NE Public Health Laboratory | Pathogen Discovery, Respiratory Viruses Branch, Division of Viral Diseases, Centers for Disease Control and Prevention |
| EPI_ISL_424876 | NH Dept. of Health and Human Services Public Health Labs | Pathogen Discovery, Respiratory Viruses Branch, Division of Viral Diseases, Centers for Disease Control and Prevention |
| EPI_ISL_424877 | NH Dept. of Health and Human Services Public Health Labs | Pathogen Discovery, Respiratory Viruses Branch, Division of Viral Diseases, Centers for Disease Control and Prevention |
| EPI_ISL_424878 | NJ Public Health and Environmental Laboratories | Pathogen Discovery, Respiratory Viruses Branch, Division of Viral Diseases, Centers for Disease Control and Prevention |
| EPI_ISL_424879 | NV-Southern Nevada Public Health Laboratory | Pathogen Discovery, Respiratory Viruses Branch, Division of Viral Diseases, Centers for Disease Control and Prevention |
| EPI_ISL_424880 | OH Department of Health Laboratory | Pathogen Discovery, Respiratory Viruses Branch, Division of Viral Diseases, Centers for Disease Control and Prevention |
| EPI_ISL_424881 | PA Department of Health, Bureau of Laboratories | Pathogen Discovery, Respiratory Viruses Branch, Division of Viral Diseases, Centers for Disease Control and Prevention |
| EPI_ISL_424882 | PA Department of Health, Bureau of Laboratories | Pathogen Discovery, Respiratory Viruses Branch, Division of Viral Diseases, Centers for Disease Control and Prevention |
| EPI_ISL_424883 | PA Department of Health, Bureau of Laboratories | Pathogen Discovery, Respiratory Viruses Branch, Division of Viral Diseases, Centers for Disease Control and Prevention |

| EPI_Accession_ID | Originating Laboratory | Submitting Laboratory |
|---|---|---|
| EPI_ISL_424884 | PA Department of Health, Bureau of Laboratories | Pathogen Discovery, Respiratory Viruses Branch, Division of Viral Diseases, Centers for Disease Control and Prevention |
| EPI_ISL_424885 | PA Department of Health, Bureau of Laboratories | Pathogen Discovery, Respiratory Viruses Branch, Division of Viral Diseases, Centers for Disease Control and Prevention |
| EPI_ISL_424886 | PA Department of Health, Bureau of Laboratories | Pathogen Discovery, Respiratory Viruses Branch, Division of Viral Diseases, Centers for Disease Control and Prevention |
| EPI_ISL_424887 | RI State Health Laboratories | Pathogen Discovery, Respiratory Viruses Branch, Division of Viral Diseases, Centers for Disease Control and Prevention |
| EPI_ISL_424888 | SC Dept of Health and Env. Control-Bureau of Laboratories | Pathogen Discovery, Respiratory Viruses Branch, Division of Viral Diseases, Centers for Disease Control and Prevention |
| EPI_ISL_424889 | UT-Unified State Labs: Public Health Utah DOH | Pathogen Discovery, Respiratory Viruses Branch, Division of Viral Diseases, Centers for Disease Control and Prevention |
| EPI_ISL_424890 | UT-Unified State Labs: Public Health Utah DOH | Pathogen Discovery, Respiratory Viruses Branch, Division of Viral Diseases, Centers for Disease Control and Prevention |
| EPI_ISL_424891 | VA-Division of Consolidated Laboratory Services | Pathogen Discovery, Respiratory Viruses Branch, Division of Viral Diseases, Centers for Disease Control and Prevention |
| EPI_ISL_424892 | VA-Division of Consolidated Laboratory Services | Pathogen Discovery, Respiratory Viruses Branch, Division of Viral Diseases, Centers for Disease Control and Prevention |
| EPI_ISL_424893 | VA-Division of Consolidated Laboratory Services | Pathogen Discovery, Respiratory Viruses Branch, Division of Viral Diseases, Centers for Disease Control and Prevention |
| EPI_ISL_424894 | VA-Division of Consolidated Laboratory Services | Pathogen Discovery, Respiratory Viruses Branch, Division of Viral Diseases, Centers for Disease Control and Prevention |

| EPI_Accession_ID | Originating Laboratory | Submitting Laboratory |
|---|---|---|
| EPI_ISL_424895 | IA State Hygienic Laboratory | Pathogen Discovery, Respiratory Viruses Branch, Division of Viral Diseases, Centers for Disease Control and Prevention |
| EPI_ISL_424896 | IA State Hygienic Laboratory | Pathogen Discovery, Respiratory Viruses Branch, Division of Viral Diseases, Centers for Disease Control and Prevention |
| EPI_ISL_424897 | IA State Hygienic Laboratory | Pathogen Discovery, Respiratory Viruses Branch, Division of Viral Diseases, Centers for Disease Control and Prevention |
| EPI_ISL_424898 | IA State Hygienic Laboratory | Pathogen Discovery, Respiratory Viruses Branch, Division of Viral Diseases, Centers for Disease Control and Prevention |
| EPI_ISL_424899 | IA State Hygienic Laboratory | Pathogen Discovery, Respiratory Viruses Branch, Division of Viral Diseases, Centers for Disease Control and Prevention |
| EPI_ISL_424900 | IA State Hygienic Laboratory | Pathogen Discovery, Respiratory Viruses Branch, Division of Viral Diseases, Centers for Disease Control and Prevention |
| EPI_ISL_424901 | NJ Public Health and Environmental Laboratories | Pathogen Discovery, Respiratory Viruses Branch, Division of Viral Diseases, Centers for Disease Control and Prevention |
| EPI_ISL_424902 | SC Dept of Health and Env. Control-Bureau of Laboratories | Pathogen Discovery, Respiratory Viruses Branch, Division of Viral Diseases, Centers for Disease Control and Prevention |
| EPI_ISL_424903 | SC Dept of Health and Env. Control-Bureau of Laboratories | Pathogen Discovery, Respiratory Viruses Branch, Division of Viral Diseases, Centers for Disease Control and Prevention |
| EPI_ISL_424904 | SC Dept of Health and Env. Control-Bureau of Laboratories | Pathogen Discovery, Respiratory Viruses Branch, Division of Viral Diseases, Centers for Disease Control and Prevention |
| EPI_ISL_424905 | SC Dept of Health and Env. Control-Bureau of Laboratories | Pathogen Discovery, Respiratory Viruses Branch, Division of Viral Diseases, Centers for Disease Control and Prevention |

| EPI_Accession_ID | Originating Laboratory | Submitting Laboratory |
|---|---|---|
| EPI_ISL_424906 | NYC Department of Health and Mental Hygiene | Pathogen Discovery, Respiratory Viruses Branch, Division of Viral Diseases, Centers for Disease Control and Prevention |
| EPI_ISL_424907 | VA-Division of Consolidated Laboratory Services | Pathogen Discovery, Respiratory Viruses Branch, Division of Viral Diseases, Centers for Disease Control and Prevention |
| EPI_ISL_424908 | MA State Public Health Laboratory | Pathogen Discovery, Respiratory Viruses Branch, Division of Viral Diseases, Centers for Disease Control and Prevention |
| EPI_ISL_424909 | MA State Public Health Laboratory | Pathogen Discovery, Respiratory Viruses Branch, Division of Viral Diseases, Centers for Disease Control and Prevention |
| EPI_ISL_424910 | MA State Public Health Laboratory | Pathogen Discovery, Respiratory Viruses Branch, Division of Viral Diseases, Centers for Disease Control and Prevention |
| EPI_ISL_424911 | MA State Public Health Laboratory | Pathogen Discovery, Respiratory Viruses Branch, Division of Viral Diseases, Centers for Disease Control and Prevention |
| EPI_ISL_424912 | MA State Public Health Laboratory | Pathogen Discovery, Respiratory Viruses Branch, Division of Viral Diseases, Centers for Disease Control and Prevention |
| EPI_ISL_424913 | MA State Public Health Laboratory | Pathogen Discovery, Respiratory Viruses Branch, Division of Viral Diseases, Centers for Disease Control and Prevention |
| EPI_ISL_424914 | MA State Public Health Laboratory | Pathogen Discovery, Respiratory Viruses Branch, Division of Viral Diseases, Centers for Disease Control and Prevention |
| EPI_ISL_424915 | MA State Public Health Laboratory | Pathogen Discovery, Respiratory Viruses Branch, Division of Viral Diseases, Centers for Disease Control and Prevention |
| EPI_ISL_424916 | MA State Public Health Laboratory | Pathogen Discovery, Respiratory Viruses Branch, Division of Viral Diseases, Centers for Disease Control and Prevention |

| EPI_Accession_ID | Originating Laboratory | Submitting Laboratory |
|---|---|---|
| EPI_ISL_424917 | MA State Public Health Laboratory | Pathogen Discovery, Respiratory Viruses Branch, Division of Viral Diseases, Centers for Disease Control and Prevention |
| EPI_ISL_424918 | MA State Public Health Laboratory | Pathogen Discovery, Respiratory Viruses Branch, Division of Viral Diseases, Centers for Disease Control and Prevention |
| EPI_ISL_424919 | MA State Public Health Laboratory | Pathogen Discovery, Respiratory Viruses Branch, Division of Viral Diseases, Centers for Disease Control and Prevention |
| EPI_ISL_424920 | MA State Public Health Laboratory | Pathogen Discovery, Respiratory Viruses Branch, Division of Viral Diseases, Centers for Disease Control and Prevention |
| EPI_ISL_424929 | NYU Langone Health | Departments of Pathology and Medicine, New York University School of Medicine |
| EPI_ISL_424930 | NYU Langone Health | Departments of Pathology and Medicine, New York University School of Medicine |
| EPI_ISL_424931 | NYU Langone Health | Departments of Pathology and Medicine, New York University School of Medicine |
| EPI_ISL_424932 | NYU Langone Health | Departments of Pathology and Medicine, New York University School of Medicine |
| EPI_ISL_424933 | NYU Langone Health | Departments of Pathology and Medicine, New York University School of Medicine |
| EPI_ISL_424934 | NYU Langone Health | Departments of Pathology and Medicine, New York University School of Medicine |
| EPI_ISL_424935 | NYU Langone Health | Departments of Pathology and Medicine, New York University School of Medicine |
| EPI_ISL_424936 | NYU Langone Health | Departments of Pathology and Medicine, New York University School of Medicine |
| EPI_ISL_424937 | NYU Langone Health | Departments of Pathology and Medicine, New York University School of Medicine |
| EPI_ISL_424938 | NYU Langone Health | Departments of Pathology and Medicine, New York University School of Medicine |

| EPI_Accession_ID | Originating Laboratory | Submitting Laboratory |
|---|---|---|
| EPI_ISL_424939 | NYU Langone Health | Departments of Pathology and Medicine, New York University School of Medicine |
| EPI_ISL_424940 | NYU Langone Health | Departments of Pathology and Medicine, New York University School of Medicine |
| EPI_ISL_424941 | NYU Langone Health | Departments of Pathology and Medicine, New York University School of Medicine |
| EPI_ISL_424942 | NYU Langone Health | Departments of Pathology and Medicine, New York University School of Medicine |
| EPI_ISL_424943 | NYU Langone Health | Departments of Pathology and Medicine, New York University School of Medicine |
| EPI_ISL_424944 | NYU Langone Health | Departments of Pathology and Medicine, New York University School of Medicine |
| EPI_ISL_424945 | NYU Langone Health | Departments of Pathology and Medicine, New York University School of Medicine |
| EPI_ISL_424946 | NYU Langone Health | Departments of Pathology and Medicine, New York University School of Medicine |
| EPI_ISL_424947 | NYU Langone Health | Departments of Pathology and Medicine, New York University School of Medicine |
| EPI_ISL_424948 | NYU Langone Health | Departments of Pathology and Medicine, New York University School of Medicine |
| EPI_ISL_424949 | NYU Langone Health | Departments of Pathology and Medicine, New York University School of Medicine |
| EPI_ISL_424950 | NYU Langone Health | Departments of Pathology and Medicine, New York University School of Medicine |
| EPI_ISL_424951 | NYU Langone Health | Departments of Pathology and Medicine, New York University School of Medicine |
| EPI_ISL_424952 | NYU Langone Health | Departments of Pathology and Medicine, New York University School of Medicine |
| EPI_ISL_424953 | NYU Langone Health | Departments of Pathology and Medicine, New York University School of Medicine |
| EPI_ISL_424954 | NYU Langone Health | Departments of Pathology and Medicine, New York University School of Medicine |

| EPI_Accession_ID | Originating Laboratory | Submitting Laboratory |
|---|---|---|
| EPI_ISL_424955 | NYU Langone Health | Departments of Pathology and Medicine, New York University School of Medicine |
| EPI_ISL_424956 | NYU Langone Health | Departments of Pathology and Medicine, New York University School of Medicine |
| EPI_ISL_424957 | NYU Langone Health | Departments of Pathology and Medicine, New York University School of Medicine |
| EPI_ISL_424958 | NYU Langone Health | Departments of Pathology and Medicine, New York University School of Medicine |
| EPI_ISL_424959 | NYU Langone Health | Departments of Pathology and Medicine, New York University School of Medicine |
| EPI_ISL_424960 | NYU Langone Health | Departments of Pathology and Medicine, New York University School of Medicine |
| EPI_ISL_424961 | NYU Langone Health | Departments of Pathology and Medicine, New York University School of Medicine |
| EPI_ISL_424962 | NYU Langone Health | Departments of Pathology and Medicine, New York University School of Medicine |
| EPI_ISL_424963 | NYU Langone Health | Departments of Pathology and Medicine, New York University School of Medicine |
| EPI_ISL_424964 | NYU Langone Health | Departments of Pathology and Medicine, New York University School of Medicine |
| EPI_ISL_424965 | NYU Langone Health | Departments of Pathology and Medicine, New York University School of Medicine |
| EPI_ISL_424966 | NYU Langone Health | Departments of Pathology and Medicine, New York University School of Medicine |
| EPI_ISL_424967 | NYU Langone Health | Departments of Pathology and Medicine, New York University School of Medicine |
| EPI_ISL_424968 | NYU Langone Health | Departments of Pathology and Medicine, New York University School of Medicine |
| EPI_ISL_424969 | Laboratory Medicine | Department of Laboratory Medicine, Lin-Kou Chang Gung Memorial Hospital, Taoyuan, Taiwan |
| EPI_ISL_424970 | Laboratory Medicine | Department of Laboratory Medicine, Lin-Kou Chang Gung Memorial Hospital, Taoyuan, Taiwan |

| EPI_Accession_ID | Originating Laboratory | Submitting Laboratory |
|---|---|---|
| EPI_ISL_424971 | Laboratory Medicine | Department of Laboratory Medicine, Lin-Kou Chang Gung Memorial Hospital, Taoyuan, Taiwan |
| EPI_ISL_424972 | Laboratory Medicine | Department of Laboratory Medicine, Lin-Kou Chang Gung Memorial Hospital, Taoyuan, Taiwan |
| EPI_ISL_424973 | Laboratory Medicine | Department of Laboratory Medicine, Lin-Kou Chang Gung Memorial Hospital, Taoyuan, Taiwan |
| EPI_ISL_424974 | Laboratory Medicine | Department of Laboratory Medicine, Lin-Kou Chang Gung Memorial Hospital, Taoyuan, Taiwan |
| EPI_ISL_424975 | Laboratory Medicine | Department of Laboratory Medicine, Lin-Kou Chang Gung Memorial Hospital, Taoyuan, Taiwan |
| EPI_ISL_424978 | Laboratory Medicine | Department of Laboratory Medicine, Lin-Kou Chang Gung Memorial Hospital, Taoyuan, Taiwan |
| EPI_ISL_424981 | unknown | Bureau of Laboratories |
| EPI_ISL_424982 | unknown | Bureau of Laboratories |
| EPI_ISL_424984 | unknown | Bureau of Laboratories |
| EPI_ISL_424985 | unknown | Bureau of Laboratories |
| EPI_ISL_424986 | unknown | Bureau of Laboratories |
| EPI_ISL_424988 | unknown | Bureau of Laboratories |
| EPI_ISL_424989 | unknown | Bureau of Laboratories |
| EPI_ISL_424990 | unknown | Bureau of Laboratories |
| EPI_ISL_424992 | unknown | Bureau of Laboratories |
| EPI_ISL_425023 | Respiratory Virus Unit, Microbiology Services Colindale, Public Health England | Respiratory Virus Unit, Microbiology Services Colindale, Public Health England |
| EPI_ISL_425024 | Respiratory Virus Unit, Microbiology Services Colindale, Public Health England | Respiratory Virus Unit, Microbiology Services Colindale, Public Health England |
| EPI_ISL_425048 | Lab voor klinische biologie | Onderzoeksgroep Virologie |
| EPI_ISL_425050 | Lab voor klinische biologie | Onderzoeksgroep Virologie |
| EPI_ISL_425051 | Lab voor klinische biologie | Onderzoeksgroep Virologie |
| EPI_ISL_425052 | Lab voor klinische biologie | Onderzoeksgroep Virologie |
| EPI_ISL_425053 | Lab voor klinische biologie | Onderzoeksgroep Virologie |
| EPI_ISL_425054 | Lab voor klinische biologie | Onderzoeksgroep Virologie |
| EPI_ISL_425055 | Lab voor klinische biologie | Onderzoeksgroep Virologie |
| EPI_ISL_425056 | Lab voor klinische biologie | Onderzoeksgroep Virologie |

| EPI_Accession_ID | Originating Laboratory | Submitting Laboratory |
|---|---|---|
| EPI_ISL_425057 | Lab voor klinische biologie | Onderzoeksgroep Virologie |
| EPI_ISL_425058 | Lab voor klinische biologie | Onderzoeksgroep Virologie |
| EPI_ISL_425059 | Lab voor klinische biologie | Onderzoeksgroep Virologie |
| EPI_ISL_425060 | Lab voor klinische biologie | Onderzoeksgroep Virologie |
| EPI_ISL_425061 | Lab voor klinische biologie | Onderzoeksgroep Virologie |
| EPI_ISL_425062 | Lab voor klinische biologie | Onderzoeksgroep Virologie |
| EPI_ISL_425063 | Lab voor klinische biologie | Onderzoeksgroep Virologie |
| EPI_ISL_425064 | Lab voor klinische biologie | Onderzoeksgroep Virologie |
| EPI_ISL_425117 | Division of Viral Diseases, Center for Laboratory Control of Infectious Diseases, Korea Centers for Diseases Control and Prevention | Division of Viral Diseases, Center for Laboratory Control of Infectious Diseases, Korea Centers for Diseases Control and Prevention |
| EPI_ISL_425118 | Division of Viral Diseases, Center for Laboratory Control of Infectious Diseases, Korea Centers for Diseases Control and Prevention | Division of Viral Diseases, Center for Laboratory Control of Infectious Diseases, Korea Centers for Diseases Control and Prevention |
| EPI_ISL_425120 | Center of Medical Microbiology, Virology, and Hospital Hygiene, University of Duesseldorf | Center of Medical Microbiology, Virology, and Hospital Hygiene, University of Duesseldorf |
| EPI_ISL_425121 | Center of Medical Microbiology, Virology, and Hospital Hygiene, University of Duesseldorf | Center of Medical Microbiology, Virology, and Hospital Hygiene, University of Duesseldorf |
| EPI_ISL_425122 | Center of Medical Microbiology, Virology, and Hospital Hygiene, University of Duesseldorf | Center of Medical Microbiology, Virology, and Hospital Hygiene, University of Duesseldorf |
| EPI_ISL_425123 | Center of Medical Microbiology, Virology, and Hospital Hygiene, University of Duesseldorf | Center of Medical Microbiology, Virology, and Hospital Hygiene, University of Duesseldorf |
| EPI_ISL_425124 | Center of Medical Microbiology, Virology, and Hospital Hygiene, University of Duesseldorf | Center of Medical Microbiology, Virology, and Hospital Hygiene, University of Duesseldorf |
| EPI_ISL_425125 | Center of Medical Microbiology, Virology, and Hospital Hygiene, University of Duesseldorf | Center of Medical Microbiology, Virology, and Hospital Hygiene, University of Duesseldorf |
| EPI_ISL_425126 | Center of Medical Microbiology, Virology, and Hospital Hygiene, University of Duesseldorf | Center of Medical Microbiology, Virology, and Hospital Hygiene, University of Duesseldorf |
| EPI_ISL_425127 | Center of Medical Microbiology, Virology, and Hospital Hygiene, University of Duesseldorf | Center of Medical Microbiology, Virology, and Hospital Hygiene, University of Duesseldorf |
| EPI_ISL_425128 | Center of Medical Microbiology, Virology, and Hospital Hygiene, University of Duesseldorf | Center of Medical Microbiology, Virology, and Hospital Hygiene, University of Duesseldorf |

| EPI_Accession_ID | Originating Laboratory | Submitting Laboratory |
|---|---|---|
| EPI_ISL_425129 | Center of Medical Microbiology, Virology, and Hospital Hygiene, University of Duesseldorf | Center of Medical Microbiology, Virology, and Hospital Hygiene, University of Duesseldorf |
| EPI_ISL_425130 | Center of Medical Microbiology, Virology, and Hospital Hygiene, University of Duesseldorf | Center of Medical Microbiology, Virology, and Hospital Hygiene, University of Duesseldorf |
| EPI_ISL_425131 | Center of Medical Microbiology, Virology, and Hospital Hygiene, University of Duesseldorf | Center of Medical Microbiology, Virology, and Hospital Hygiene, University of Duesseldorf |
| EPI_ISL_425132 | Center of Medical Microbiology, Virology, and Hospital Hygiene, University of Duesseldorf | Center of Medical Microbiology, Virology, and Hospital Hygiene, University of Duesseldorf |
| EPI_ISL_425133 | Center of Medical Microbiology, Virology, and Hospital Hygiene, University of Duesseldorf | Center of Medical Microbiology, Virology, and Hospital Hygiene, University of Duesseldorf |
| EPI_ISL_425134 | Center of Medical Microbiology, Virology, and Hospital Hygiene, University of Duesseldorf | Center of Medical Microbiology, Virology, and Hospital Hygiene, University of Duesseldorf |
| EPI_ISL_425135 | Center of Medical Microbiology, Virology, and Hospital Hygiene, University of Duesseldorf | Center of Medical Microbiology, Virology, and Hospital Hygiene, University of Duesseldorf |
| EPI_ISL_425136 | Center of Medical Microbiology, Virology, and Hospital Hygiene, University of Duesseldorf | Center of Medical Microbiology, Virology, and Hospital Hygiene, University of Duesseldorf |
| EPI_ISL_425137 | Center of Medical Microbiology, Virology, and Hospital Hygiene, University of Duesseldorf | Center of Medical Microbiology, Virology, and Hospital Hygiene, University of Duesseldorf |
| EPI_ISL_425138 | Center of Medical Microbiology, Virology, and Hospital Hygiene, University of Duesseldorf | Center of Medical Microbiology, Virology, and Hospital Hygiene, University of Duesseldorf |
| EPI_ISL_425139 | Center of Medical Microbiology, Virology, and Hospital Hygiene, University of Duesseldorf | Center of Medical Microbiology, Virology, and Hospital Hygiene, University of Duesseldorf |
| EPI_ISL_425140 | Center of Medical Microbiology, Virology, and Hospital Hygiene, University of Duesseldorf | Center of Medical Microbiology, Virology, and Hospital Hygiene, University of Duesseldorf |
| EPI_ISL_425142 | University of Wisconsin-Madison AIDS Vaccine Research Laboratories | University of Wisconsin-Madison AIDS Vaccine Research Laboratories |
| EPI_ISL_425143 | University of Wisconsin-Madison AIDS Vaccine Research Laboratories | University of Wisconsin-Madison AIDS Vaccine Research Laboratories |
| EPI_ISL_425144 | University of Wisconsin-Madison AIDS Vaccine Research Laboratories | University of Wisconsin-Madison AIDS Vaccine Research Laboratories |
| EPI_ISL_425145 | University of Wisconsin-Madison AIDS Vaccine Research Laboratories | University of Wisconsin-Madison AIDS Vaccine Research Laboratories |

| EPI_Accession_ID | Originating Laboratory | Submitting Laboratory |
|---|---|---|
| EPI_ISL_425146 | University of Wisconsin-Madison AIDS Vaccine Research Laboratories | University of Wisconsin-Madison AIDS Vaccine Research Laboratories |
| EPI_ISL_425147 | University of Wisconsin-Madison AIDS Vaccine Research Laboratories | University of Wisconsin-Madison AIDS Vaccine Research Laboratories |
| EPI_ISL_425148 | University of Wisconsin-Madison AIDS Vaccine Research Laboratories | University of Wisconsin-Madison AIDS Vaccine Research Laboratories |
| EPI_ISL_425149 | University of Wisconsin-Madison AIDS Vaccine Research Laboratories | University of Wisconsin-Madison AIDS Vaccine Research Laboratories |
| EPI_ISL_425150 | University of Wisconsin-Madison AIDS Vaccine Research Laboratories | University of Wisconsin-Madison AIDS Vaccine Research Laboratories |
| EPI_ISL_425151 | University of Wisconsin-Madison AIDS Vaccine Research Laboratories | University of Wisconsin-Madison AIDS Vaccine Research Laboratories |
| EPI_ISL_425152 | University of Wisconsin-Madison AIDS Vaccine Research Laboratories | University of Wisconsin-Madison AIDS Vaccine Research Laboratories |
| EPI_ISL_425153 | University of Wisconsin-Madison AIDS Vaccine Research Laboratories | University of Wisconsin-Madison AIDS Vaccine Research Laboratories |
| EPI_ISL_425154 | University of Wisconsin-Madison AIDS Vaccine Research Laboratories | University of Wisconsin-Madison AIDS Vaccine Research Laboratories |
| EPI_ISL_425155 | University of Wisconsin-Madison AIDS Vaccine Research Laboratories | University of Wisconsin-Madison AIDS Vaccine Research Laboratories |
| EPI_ISL_425156 | University of Wisconsin-Madison AIDS Vaccine Research Laboratories | University of Wisconsin-Madison AIDS Vaccine Research Laboratories |
| EPI_ISL_425157 | University of Wisconsin-Madison AIDS Vaccine Research Laboratories | University of Wisconsin-Madison AIDS Vaccine Research Laboratories |
| EPI_ISL_425158 | University of Wisconsin-Madison AIDS Vaccine Research Laboratories | University of Wisconsin-Madison AIDS Vaccine Research Laboratories |
| EPI_ISL_425159 | University of Wisconsin-Madison AIDS Vaccine Research Laboratories | University of Wisconsin-Madison AIDS Vaccine Research Laboratories |
| EPI_ISL_425160 | University of Wisconsin-Madison AIDS Vaccine Research Laboratories | University of Wisconsin-Madison AIDS Vaccine Research Laboratories |
| EPI_ISL_425161 | University of Wisconsin-Madison AIDS Vaccine Research Laboratories | University of Wisconsin-Madison AIDS Vaccine Research Laboratories |

| EPI_Accession_ID | Originating Laboratory | Submitting Laboratory |
|---|---|---|
| EPI_ISL_425162 | University of Wisconsin-Madison AIDS Vaccine Research Laboratories | University of Wisconsin-Madison AIDS Vaccine Research Laboratories |
| EPI_ISL_425163 | University of Wisconsin-Madison AIDS Vaccine Research Laboratories | University of Wisconsin-Madison AIDS Vaccine Research Laboratories |
| EPI_ISL_425164 | University of Wisconsin-Madison AIDS Vaccine Research Laboratories | University of Wisconsin-Madison AIDS Vaccine Research Laboratories |
| EPI_ISL_425165 | University of Wisconsin-Madison AIDS Vaccine Research Laboratories | University of Wisconsin-Madison AIDS Vaccine Research Laboratories |
| EPI_ISL_425166 | University of Wisconsin-Madison AIDS Vaccine Research Laboratories | University of Wisconsin-Madison AIDS Vaccine Research Laboratories |
| EPI_ISL_425167 | University of Wisconsin-Madison AIDS Vaccine Research Laboratories | University of Wisconsin-Madison AIDS Vaccine Research Laboratories |
| EPI_ISL_425168 | University of Wisconsin-Madison AIDS Vaccine Research Laboratories | University of Wisconsin-Madison AIDS Vaccine Research Laboratories |
| EPI_ISL_425169 | University of Wisconsin-Madison AIDS Vaccine Research Laboratories | University of Wisconsin-Madison AIDS Vaccine Research Laboratories |
| EPI_ISL_425170 | University of Wisconsin-Madison AIDS Vaccine Research Laboratories | University of Wisconsin-Madison AIDS Vaccine Research Laboratories |
| EPI_ISL_425171 | University of Wisconsin-Madison AIDS Vaccine Research Laboratories | University of Wisconsin-Madison AIDS Vaccine Research Laboratories |
| EPI_ISL_425172 | University of Wisconsin-Madison AIDS Vaccine Research Laboratories | University of Wisconsin-Madison AIDS Vaccine Research Laboratories |
| EPI_ISL_425173 | University of Wisconsin-Madison AIDS Vaccine Research Laboratories | University of Wisconsin-Madison AIDS Vaccine Research Laboratories |
| EPI_ISL_425174 | University of Wisconsin-Madison AIDS Vaccine Research Laboratories | University of Wisconsin-Madison AIDS Vaccine Research Laboratories |
| EPI_ISL_425175 | University of Wisconsin-Madison AIDS Vaccine Research Laboratories | University of Wisconsin-Madison AIDS Vaccine Research Laboratories |
| EPI_ISL_425177 | Public Health Ontario | Public Health Agency of Canada - National Microbiology Laboratory |
| EPI_ISL_425178 | Servicio de MicrobiologÃ-a. Consorcio Hospital General Universitario de Valencia | Sequencing and Bioinformatics Service and Molecular Epidemiology Research Group. FISABIO-Public Health |

| EPI_Accession_ID | Originating Laboratory | Submitting Laboratory |
|---|---|---|
| EPI_ISL_425179 | Servicio de MicrobiologÃ-a. Consorcio Hospital General Universitario de Valencia | Sequencing and Bioinformatics Service and Molecular Epidemiology Research Group. FISABIO-Public Health |
| EPI_ISL_425180 | Servicio de MicrobiologÃ-a. Consorcio Hospital General Universitario de Valencia | Sequencing and Bioinformatics Service and Molecular Epidemiology Research Group. FISABIO-Public Health |
| EPI_ISL_425181 | Servicio de MicrobiologÃ-a. Consorcio Hospital General Universitario de Valencia | Sequencing and Bioinformatics Service and Molecular Epidemiology Research Group. FISABIO-Public Health |
| EPI_ISL_425182 | Servicio de MicrobiologÃ-a. Consorcio Hospital General Universitario de Valencia | Sequencing and Bioinformatics Service and Molecular Epidemiology Research Group. FISABIO-Public Health |
| EPI_ISL_425183 | Servicio de MicrobiologÃ-a. Consorcio Hospital General Universitario de Valencia | Sequencing and Bioinformatics Service and Molecular Epidemiology Research Group. FISABIO-Public Health |
| EPI_ISL_425184 | Servicio de MicrobiologÃ-a. Consorcio Hospital General Universitario de Valencia | Sequencing and Bioinformatics Service and Molecular Epidemiology Research Group. FISABIO-Public Health |
| EPI_ISL_425185 | Servicio de MicrobiologÃ-a. Consorcio Hospital General Universitario de Valencia | Sequencing and Bioinformatics Service and Molecular Epidemiology Research Group. FISABIO-Public Health |
| EPI_ISL_425186 | Servicio de MicrobiologÃ-a. Consorcio Hospital General Universitario de Valencia | Sequencing and Bioinformatics Service and Molecular Epidemiology Research Group. FISABIO-Public Health |
| EPI_ISL_425187 | Servicio de MicrobiologÃ-a. Consorcio Hospital General Universitario de Valencia | Sequencing and Bioinformatics Service and Molecular Epidemiology Research Group. FISABIO-Public Health |
| EPI_ISL_425188 | Servicio de MicrobiologÃ-a. Consorcio Hospital General Universitario de Valencia | Sequencing and Bioinformatics Service and Molecular Epidemiology Research Group. FISABIO-Public Health |
| EPI_ISL_425189 | Servicio de MicrobiologÃ-a. Consorcio Hospital General Universitario de Valencia | Sequencing and Bioinformatics Service and Molecular Epidemiology Research Group. FISABIO-Public Health |
| EPI_ISL_425190 | Servicio de MicrobiologÃ-a. Consorcio Hospital General Universitario de Valencia | Sequencing and Bioinformatics Service and Molecular Epidemiology Research Group. FISABIO-Public Health |
| EPI_ISL_425191 | Servicio de MicrobiologÃ-a. Consorcio Hospital General Universitario de Valencia | Sequencing and Bioinformatics Service and Molecular Epidemiology Research Group. FISABIO-Public Health |
| EPI_ISL_425192 | Servicio de MicrobiologÃ-a. Consorcio Hospital General Universitario de Valencia | Sequencing and Bioinformatics Service and Molecular Epidemiology Research Group. FISABIO-Public Health |
| EPI_ISL_425193 | Servicio de MicrobiologÃ-a. Consorcio Hospital General Universitario de Valencia | Sequencing and Bioinformatics Service and Molecular Epidemiology Research Group. FISABIO-Public Health |
| EPI_ISL_425194 | Servicio de MicrobiologÃ-a. Consorcio Hospital General Universitario de Valencia | Sequencing and Bioinformatics Service and Molecular Epidemiology Research Group. FISABIO-Public Health |

| EPI_Accession_ID | Originating Laboratory | Submitting Laboratory |
| --- | --- | --- |
| EPI_ISL_425195 | Servicio de MicrobiologÃ-a. Consorcio Hospital General Universitario de Valencia | Sequencing and Bioinformatics Service and Molecular Epidemiology Research Group. FISABIO-Public Health |
| EPI_ISL_425196 | Servicio de MicrobiologÃ-a. Consorcio Hospital General Universitario de Valencia | Sequencing and Bioinformatics Service and Molecular Epidemiology Research Group. FISABIO-Public Health |
| EPI_ISL_425197 | Servicio de MicrobiologÃ-a. Consorcio Hospital General Universitario de Valencia | Sequencing and Bioinformatics Service and Molecular Epidemiology Research Group. FISABIO-Public Health |
| EPI_ISL_425198 | Servicio de MicrobiologÃ-a. Consorcio Hospital General Universitario de Valencia | Sequencing and Bioinformatics Service and Molecular Epidemiology Research Group. FISABIO-Public Health |
| EPI_ISL_425199 | Servicio de Microbiologia. Hospital Clinico Universitario de Valencia | Sequencing and Bioinformatics Service and Molecular Epidemiology Research Group. FISABIO-Public Health |
| EPI_ISL_425200 | Servicio de Microbiologia. Hospital Clinico Universitario de Valencia | Sequencing and Bioinformatics Service and Molecular Epidemiology Research Group. FISABIO-Public Health |
| EPI_ISL_425201 | Servicio de Microbiologia. Hospital Clinico Universitario de Valencia | Sequencing and Bioinformatics Service and Molecular Epidemiology Research Group. FISABIO-Public Health |
| EPI_ISL_425202 | Servicio de Microbiologia. Hospital Clinico Universitario de Valencia | Sequencing and Bioinformatics Service and Molecular Epidemiology Research Group. FISABIO-Public Health |
| EPI_ISL_425203 | Servicio de Microbiologia. Hospital Clinico Universitario de Valencia | Sequencing and Bioinformatics Service and Molecular Epidemiology Research Group. FISABIO-Public Health |
| EPI_ISL_425204 | Servicio de Microbiologia. Hospital Clinico Universitario de Valencia | Sequencing and Bioinformatics Service and Molecular Epidemiology Research Group. FISABIO-Public Health |
| EPI_ISL_425205 | Servicio de Microbiologia. Hospital Clinico Universitario de Valencia | Sequencing and Bioinformatics Service and Molecular Epidemiology Research Group. FISABIO-Public Health |
| EPI_ISL_425206 | Servicio de Microbiologia. Hospital Clinico Universitario de Valencia | Sequencing and Bioinformatics Service and Molecular Epidemiology Research Group. FISABIO-Public Health |
| EPI_ISL_425207 | Servicio de Microbiologia. Hospital Clinico Universitario de Valencia | Sequencing and Bioinformatics Service and Molecular Epidemiology Research Group. FISABIO-Public Health |
| EPI_ISL_425208 | Servicio de Microbiologia. Hospital Clinico Universitario de Valencia | Sequencing and Bioinformatics Service and Molecular Epidemiology Research Group. FISABIO-Public Health |
| EPI_ISL_425209 | Servicio de Microbiologia. Hospital Clinico Universitario de Valencia | Sequencing and Bioinformatics Service and Molecular Epidemiology Research Group. FISABIO-Public Health |
| EPI_ISL_425210 | Servicio de Microbiologia. Hospital Clinico Universitario de Valencia | Sequencing and Bioinformatics Service and Molecular Epidemiology Research Group. FISABIO-Public Health |

| EPI_Accession_ID | Originating Laboratory | Submitting Laboratory |
|---|---|---|
| EPI_ISL_425211 | Servicio de Microbiologia. Hospital Clinico Universitario de Valencia | Sequencing and Bioinformatics Service and Molecular Epidemiology Research Group. FISABIO-Public Health |
| EPI_ISL_425212 | Servicio de MicrobiologÃ-a. Consorcio Hospital General Universitario de Valencia | Sequencing and Bioinformatics Service and Molecular Epidemiology Research Group. FISABIO-Public Health |
| EPI_ISL_425213 | Servicio de MicrobiologÃ-a. Consorcio Hospital General Universitario de Valencia | Sequencing and Bioinformatics Service and Molecular Epidemiology Research Group. FISABIO-Public Health |
| EPI_ISL_425214 | Servicio de MicrobiologÃ-a. Consorcio Hospital General Universitario de Valencia | Sequencing and Bioinformatics Service and Molecular Epidemiology Research Group. FISABIO-Public Health |
| EPI_ISL_425215 | Servicio de MicrobiologÃ-a. Consorcio Hospital General Universitario de Valencia | Sequencing and Bioinformatics Service and Molecular Epidemiology Research Group. FISABIO-Public Health |
| EPI_ISL_425216 | Servicio de MicrobiologÃ-a. Consorcio Hospital General Universitario de Valencia | Sequencing and Bioinformatics Service and Molecular Epidemiology Research Group. FISABIO-Public Health |
| EPI_ISL_425217 | Servicio de MicrobiologÃ-a. Consorcio Hospital General Universitario de Valencia | Sequencing and Bioinformatics Service and Molecular Epidemiology Research Group. FISABIO-Public Health |
| EPI_ISL_425218 | Servicio de MicrobiologÃ-a. Consorcio Hospital General Universitario de Valencia | Sequencing and Bioinformatics Service and Molecular Epidemiology Research Group. FISABIO-Public Health |
| EPI_ISL_425219 | Servicio de MicrobiologÃ-a. Consorcio Hospital General Universitario de Valencia | Sequencing and Bioinformatics Service and Molecular Epidemiology Research Group. FISABIO-Public Health |
| EPI_ISL_425220 | Servicio de MicrobiologÃ-a. Consorcio Hospital General Universitario de Valencia | Sequencing and Bioinformatics Service and Molecular Epidemiology Research Group. FISABIO-Public Health |
| EPI_ISL_425221 | Servicio de MicrobiologÃ-a. Consorcio Hospital General Universitario de Valencia | Sequencing and Bioinformatics Service and Molecular Epidemiology Research Group. FISABIO-Public Health |
| EPI_ISL_425222 | Servicio de MicrobiologÃ-a. Consorcio Hospital General Universitario de Valencia | Sequencing and Bioinformatics Service and Molecular Epidemiology Research Group. FISABIO-Public Health |
| EPI_ISL_425223 | Servicio de MicrobiologÃ-a. Consorcio Hospital General Universitario de Valencia | Sequencing and Bioinformatics Service and Molecular Epidemiology Research Group. FISABIO-Public Health |
| EPI_ISL_425224 | Wadsworth Center, New York State Department of Health | Wadsworth Center, New York State Department of Health |
| EPI_ISL_425228 | Laboratory of Molecular Genetics, 2nd Faculty of Medicine, Charles University in Prague, Prague, Czech Republic | Laboratory of Molecular Genetics, 2nd Faculty of Medicine, Charles University in Prague, Prague, Czech Republic |

| EPI_Accession_ID | Originating Laboratory | Submitting Laboratory |
| --- | --- | --- |
| EPI_ISL_425229 | Laboratory of Molecular Genetics, 2nd Faculty of Medicine, Charles University in Prague, Prague, Czech Republic | Laboratory of Molecular Genetics, 2nd Faculty of Medicine, Charles University in Prague, Prague, Czech Republic |
| EPI_ISL_425231 | Department of Pathology, University of Cambridge | COVID-19 Genomics UK (COG-UK) Consortium |
| EPI_ISL_425232 | Department of Pathology, University of Cambridge | COVID-19 Genomics UK (COG-UK) Consortium |
| EPI_ISL_425233 | Department of Pathology, University of Cambridge | COVID-19 Genomics UK (COG-UK) Consortium |
| EPI_ISL_425234 | Department of Pathology, University of Cambridge | COVID-19 Genomics UK (COG-UK) Consortium |
| EPI_ISL_425235 | Department of Pathology, University of Cambridge | COVID-19 Genomics UK (COG-UK) Consortium |
| EPI_ISL_425236 | Department of Pathology, University of Cambridge | COVID-19 Genomics UK (COG-UK) Consortium |
| EPI_ISL_425237 | Department of Pathology, University of Cambridge | COVID-19 Genomics UK (COG-UK) Consortium |
| EPI_ISL_425238 | Department of Pathology, University of Cambridge | COVID-19 Genomics UK (COG-UK) Consortium |
| EPI_ISL_425239 | Department of Pathology, University of Cambridge | COVID-19 Genomics UK (COG-UK) Consortium |
| EPI_ISL_425240 | Department of Pathology, University of Cambridge | COVID-19 Genomics UK (COG-UK) Consortium |
| EPI_ISL_425241 | Department of Pathology, University of Cambridge | COVID-19 Genomics UK (COG-UK) Consortium |
| EPI_ISL_425242 | Department of Pathology, University of Cambridge | COVID-19 Genomics UK (COG-UK) Consortium |
| EPI_ISL_425243 | Department of Pathology, University of Cambridge | COVID-19 Genomics UK (COG-UK) Consortium |
| EPI_ISL_425244 | Department of Pathology, University of Cambridge | COVID-19 Genomics UK (COG-UK) Consortium |
| EPI_ISL_425245 | Department of Pathology, University of Cambridge | COVID-19 Genomics UK (COG-UK) Consortium |
| EPI_ISL_425246 | Department of Pathology, University of Cambridge | COVID-19 Genomics UK (COG-UK) Consortium |
| EPI_ISL_425247 | Department of Pathology, University of Cambridge | COVID-19 Genomics UK (COG-UK) Consortium |
| EPI_ISL_425248 | Department of Pathology, University of Cambridge | COVID-19 Genomics UK (COG-UK) Consortium |
| EPI_ISL_425249 | Department of Pathology, University of Cambridge | COVID-19 Genomics UK (COG-UK) Consortium |
| EPI_ISL_425250 | Department of Pathology, University of Cambridge | COVID-19 Genomics UK (COG-UK) Consortium |
| EPI_ISL_425251 | Department of Pathology, University of Cambridge | COVID-19 Genomics UK (COG-UK) Consortium |
| EPI_ISL_425252 | Department of Pathology, University of Cambridge | COVID-19 Genomics UK (COG-UK) Consortium |
| EPI_ISL_425253 | Department of Pathology, University of Cambridge | COVID-19 Genomics UK (COG-UK) Consortium |
| EPI_ISL_425254 | Department of Pathology, University of Cambridge | COVID-19 Genomics UK (COG-UK) Consortium |
| EPI_ISL_425255 | Department of Pathology, University of Cambridge | COVID-19 Genomics UK (COG-UK) Consortium |
| EPI_ISL_425256 | Department of Pathology, University of Cambridge | COVID-19 Genomics UK (COG-UK) Consortium |
| EPI_ISL_425257 | Department of Pathology, University of Cambridge | COVID-19 Genomics UK (COG-UK) Consortium |
| EPI_ISL_425260 | Department of Pathology, University of Cambridge | COVID-19 Genomics UK (COG-UK) Consortium |
| EPI_ISL_425262 | Department of Pathology, University of Cambridge | COVID-19 Genomics UK (COG-UK) Consortium |
| EPI_ISL_425263 | Department of Pathology, University of Cambridge | COVID-19 Genomics UK (COG-UK) Consortium |

| EPI_Accession_ID | Originating Laboratory | Submitting Laboratory |
|---|---|---|
| EPI_ISL_425264 | Department of Pathology, University of Cambridge | COVID-19 Genomics UK (COG-UK) Consortium |
| EPI_ISL_425265 | Department of Pathology, University of Cambridge | COVID-19 Genomics UK (COG-UK) Consortium |
| EPI_ISL_425266 | Department of Pathology, University of Cambridge | COVID-19 Genomics UK (COG-UK) Consortium |
| EPI_ISL_425267 | Department of Pathology, University of Cambridge | COVID-19 Genomics UK (COG-UK) Consortium |
| EPI_ISL_425268 | Department of Pathology, University of Cambridge | COVID-19 Genomics UK (COG-UK) Consortium |
| EPI_ISL_425269 | Department of Pathology, University of Cambridge | COVID-19 Genomics UK (COG-UK) Consortium |
| EPI_ISL_425270 | Department of Pathology, University of Cambridge | COVID-19 Genomics UK (COG-UK) Consortium |
| EPI_ISL_425271 | Department of Pathology, University of Cambridge | COVID-19 Genomics UK (COG-UK) Consortium |
| EPI_ISL_425272 | Department of Pathology, University of Cambridge | COVID-19 Genomics UK (COG-UK) Consortium |
| EPI_ISL_425273 | Department of Pathology, University of Cambridge | COVID-19 Genomics UK (COG-UK) Consortium |
| EPI_ISL_425275 | Department of Pathology, University of Cambridge | COVID-19 Genomics UK (COG-UK) Consortium |
| EPI_ISL_425276 | Department of Pathology, University of Cambridge | COVID-19 Genomics UK (COG-UK) Consortium |
| EPI_ISL_425277 | Department of Pathology, University of Cambridge | COVID-19 Genomics UK (COG-UK) Consortium |
| EPI_ISL_425278 | Department of Pathology, University of Cambridge | COVID-19 Genomics UK (COG-UK) Consortium |
| EPI_ISL_425279 | Department of Pathology, University of Cambridge | COVID-19 Genomics UK (COG-UK) Consortium |
| EPI_ISL_425281 | Department of Pathology, University of Cambridge | COVID-19 Genomics UK (COG-UK) Consortium |
| EPI_ISL_425282 | Department of Pathology, University of Cambridge | COVID-19 Genomics UK (COG-UK) Consortium |
| EPI_ISL_425283 | Department of Pathology, University of Cambridge | COVID-19 Genomics UK (COG-UK) Consortium |
| EPI_ISL_425284 | Department of Pathology, University of Cambridge | COVID-19 Genomics UK (COG-UK) Consortium |
| EPI_ISL_425285 | Department of Pathology, University of Cambridge | COVID-19 Genomics UK (COG-UK) Consortium |
| EPI_ISL_425286 | Department of Pathology, University of Cambridge | COVID-19 Genomics UK (COG-UK) Consortium |
| EPI_ISL_425287 | Department of Pathology, University of Cambridge | COVID-19 Genomics UK (COG-UK) Consortium |
| EPI_ISL_425288 | Department of Pathology, University of Cambridge | COVID-19 Genomics UK (COG-UK) Consortium |
| EPI_ISL_425289 | Department of Pathology, University of Cambridge | COVID-19 Genomics UK (COG-UK) Consortium |
| EPI_ISL_425290 | Department of Pathology, University of Cambridge | COVID-19 Genomics UK (COG-UK) Consortium |
| EPI_ISL_425291 | Department of Pathology, University of Cambridge | COVID-19 Genomics UK (COG-UK) Consortium |
| EPI_ISL_425292 | Department of Pathology, University of Cambridge | COVID-19 Genomics UK (COG-UK) Consortium |
| EPI_ISL_425293 | Department of Pathology, University of Cambridge | COVID-19 Genomics UK (COG-UK) Consortium |
| EPI_ISL_425294 | Department of Pathology, University of Cambridge | COVID-19 Genomics UK (COG-UK) Consortium |
| EPI_ISL_425295 | Department of Pathology, University of Cambridge | COVID-19 Genomics UK (COG-UK) Consortium |
| EPI_ISL_425296 | Department of Pathology, University of Cambridge | COVID-19 Genomics UK (COG-UK) Consortium |
| EPI_ISL_425297 | Department of Pathology, University of Cambridge | COVID-19 Genomics UK (COG-UK) Consortium |
| EPI_ISL_425298 | Department of Pathology, University of Cambridge | COVID-19 Genomics UK (COG-UK) Consortium |

| EPI_Accession_ID | Originating Laboratory | Submitting Laboratory |
|---|---|---|
| EPI_ISL_425299 | Department of Pathology, University of Cambridge | COVID-19 Genomics UK (COG-UK) Consortium |
| EPI_ISL_425300 | Department of Pathology, University of Cambridge | COVID-19 Genomics UK (COG-UK) Consortium |
| EPI_ISL_425301 | Department of Pathology, University of Cambridge | COVID-19 Genomics UK (COG-UK) Consortium |
| EPI_ISL_425302 | Department of Pathology, University of Cambridge | COVID-19 Genomics UK (COG-UK) Consortium |
| EPI_ISL_425303 | Department of Pathology, University of Cambridge | COVID-19 Genomics UK (COG-UK) Consortium |
| EPI_ISL_425304 | Department of Pathology, University of Cambridge | COVID-19 Genomics UK (COG-UK) Consortium |
| EPI_ISL_425305 | Department of Pathology, University of Cambridge | COVID-19 Genomics UK (COG-UK) Consortium |
| EPI_ISL_425306 | Department of Pathology, University of Cambridge | COVID-19 Genomics UK (COG-UK) Consortium |
| EPI_ISL_425307 | Department of Pathology, University of Cambridge | COVID-19 Genomics UK (COG-UK) Consortium |
| EPI_ISL_425308 | Department of Pathology, University of Cambridge | COVID-19 Genomics UK (COG-UK) Consortium |
| EPI_ISL_425309 | Department of Pathology, University of Cambridge | COVID-19 Genomics UK (COG-UK) Consortium |
| EPI_ISL_425310 | Department of Pathology, University of Cambridge | COVID-19 Genomics UK (COG-UK) Consortium |
| EPI_ISL_425311 | Department of Pathology, University of Cambridge | COVID-19 Genomics UK (COG-UK) Consortium |
| EPI_ISL_425312 | Department of Pathology, University of Cambridge | COVID-19 Genomics UK (COG-UK) Consortium |
| EPI_ISL_425313 | Department of Pathology, University of Cambridge | COVID-19 Genomics UK (COG-UK) Consortium |
| EPI_ISL_425314 | Department of Pathology, University of Cambridge | COVID-19 Genomics UK (COG-UK) Consortium |
| EPI_ISL_425315 | Department of Pathology, University of Cambridge | COVID-19 Genomics UK (COG-UK) Consortium |
| EPI_ISL_425316 | Department of Pathology, University of Cambridge | COVID-19 Genomics UK (COG-UK) Consortium |
| EPI_ISL_425317 | Department of Pathology, University of Cambridge | COVID-19 Genomics UK (COG-UK) Consortium |
| EPI_ISL_425318 | Department of Pathology, University of Cambridge | COVID-19 Genomics UK (COG-UK) Consortium |
| EPI_ISL_425319 | Department of Pathology, University of Cambridge | COVID-19 Genomics UK (COG-UK) Consortium |
| EPI_ISL_425320 | Department of Pathology, University of Cambridge | COVID-19 Genomics UK (COG-UK) Consortium |
| EPI_ISL_425321 | Department of Pathology, University of Cambridge | COVID-19 Genomics UK (COG-UK) Consortium |
| EPI_ISL_425322 | Department of Pathology, University of Cambridge | COVID-19 Genomics UK (COG-UK) Consortium |
| EPI_ISL_425323 | Department of Pathology, University of Cambridge | COVID-19 Genomics UK (COG-UK) Consortium |
| EPI_ISL_425324 | Department of Pathology, University of Cambridge | COVID-19 Genomics UK (COG-UK) Consortium |
| EPI_ISL_425325 | Department of Pathology, University of Cambridge | COVID-19 Genomics UK (COG-UK) Consortium |
| EPI_ISL_425326 | Department of Pathology, University of Cambridge | COVID-19 Genomics UK (COG-UK) Consortium |
| EPI_ISL_425327 | Department of Pathology, University of Cambridge | COVID-19 Genomics UK (COG-UK) Consortium |
| EPI_ISL_425328 | Department of Pathology, University of Cambridge | COVID-19 Genomics UK (COG-UK) Consortium |
| EPI_ISL_425329 | Department of Pathology, University of Cambridge | COVID-19 Genomics UK (COG-UK) Consortium |
| EPI_ISL_425330 | Department of Pathology, University of Cambridge | COVID-19 Genomics UK (COG-UK) Consortium |
| EPI_ISL_425331 | Department of Pathology, University of Cambridge | COVID-19 Genomics UK (COG-UK) Consortium |

| EPI_Accession_ID | Originating Laboratory | Submitting Laboratory |
|---|---|---|
| EPI_ISL_425332 | Department of Pathology, University of Cambridge | COVID-19 Genomics UK (COG-UK) Consortium |
| EPI_ISL_425333 | Department of Pathology, University of Cambridge | COVID-19 Genomics UK (COG-UK) Consortium |
| EPI_ISL_425334 | Department of Pathology, University of Cambridge | COVID-19 Genomics UK (COG-UK) Consortium |
| EPI_ISL_425335 | Department of Pathology, University of Cambridge | COVID-19 Genomics UK (COG-UK) Consortium |
| EPI_ISL_425336 | Department of Pathology, University of Cambridge | COVID-19 Genomics UK (COG-UK) Consortium |
| EPI_ISL_425337 | Department of Pathology, University of Cambridge | COVID-19 Genomics UK (COG-UK) Consortium |
| EPI_ISL_425338 | Department of Pathology, University of Cambridge | COVID-19 Genomics UK (COG-UK) Consortium |
| EPI_ISL_425339 | Department of Pathology, University of Cambridge | COVID-19 Genomics UK (COG-UK) Consortium |
| EPI_ISL_425340 | Department of Pathology, University of Cambridge | COVID-19 Genomics UK (COG-UK) Consortium |
| EPI_ISL_425341 | Department of Pathology, University of Cambridge | COVID-19 Genomics UK (COG-UK) Consortium |
| EPI_ISL_425342 | Department of Pathology, University of Cambridge | COVID-19 Genomics UK (COG-UK) Consortium |
| EPI_ISL_425343 | Department of Pathology, University of Cambridge | COVID-19 Genomics UK (COG-UK) Consortium |
| EPI_ISL_425344 | Department of Pathology, University of Cambridge | COVID-19 Genomics UK (COG-UK) Consortium |
| EPI_ISL_425345 | Department of Pathology, University of Cambridge | COVID-19 Genomics UK (COG-UK) Consortium |
| EPI_ISL_425347 | Department of Pathology, University of Cambridge | COVID-19 Genomics UK (COG-UK) Consortium |
| EPI_ISL_425348 | Department of Pathology, University of Cambridge | COVID-19 Genomics UK (COG-UK) Consortium |
| EPI_ISL_425349 | Department of Pathology, University of Cambridge | COVID-19 Genomics UK (COG-UK) Consortium |
| EPI_ISL_425350 | Department of Pathology, University of Cambridge | COVID-19 Genomics UK (COG-UK) Consortium |
| EPI_ISL_425351 | Department of Pathology, University of Cambridge | COVID-19 Genomics UK (COG-UK) Consortium |
| EPI_ISL_425352 | Department of Pathology, University of Cambridge | COVID-19 Genomics UK (COG-UK) Consortium |
| EPI_ISL_425353 | Department of Pathology, University of Cambridge | COVID-19 Genomics UK (COG-UK) Consortium |
| EPI_ISL_425354 | Department of Pathology, University of Cambridge | COVID-19 Genomics UK (COG-UK) Consortium |
| EPI_ISL_425355 | Department of Pathology, University of Cambridge | COVID-19 Genomics UK (COG-UK) Consortium |
| EPI_ISL_425356 | Department of Pathology, University of Cambridge | COVID-19 Genomics UK (COG-UK) Consortium |
| EPI_ISL_425357 | Department of Pathology, University of Cambridge | COVID-19 Genomics UK (COG-UK) Consortium |
| EPI_ISL_425358 | Department of Pathology, University of Cambridge | COVID-19 Genomics UK (COG-UK) Consortium |
| EPI_ISL_425359 | Department of Pathology, University of Cambridge | COVID-19 Genomics UK (COG-UK) Consortium |
| EPI_ISL_425360 | Department of Pathology, University of Cambridge | COVID-19 Genomics UK (COG-UK) Consortium |
| EPI_ISL_425361 | Department of Pathology, University of Cambridge | COVID-19 Genomics UK (COG-UK) Consortium |
| EPI_ISL_425362 | Department of Pathology, University of Cambridge | COVID-19 Genomics UK (COG-UK) Consortium |
| EPI_ISL_425363 | Department of Pathology, University of Cambridge | COVID-19 Genomics UK (COG-UK) Consortium |
| EPI_ISL_425364 | Department of Pathology, University of Cambridge | COVID-19 Genomics UK (COG-UK) Consortium |
| EPI_ISL_425365 | Department of Pathology, University of Cambridge | COVID-19 Genomics UK (COG-UK) Consortium |

| EPI_Accession_ID | Originating Laboratory | Submitting Laboratory |
|---|---|---|
| EPI_ISL_425366 | Department of Pathology, University of Cambridge | COVID-19 Genomics UK (COG-UK) Consortium |
| EPI_ISL_425367 | Department of Pathology, University of Cambridge | COVID-19 Genomics UK (COG-UK) Consortium |
| EPI_ISL_425368 | Department of Pathology, University of Cambridge | COVID-19 Genomics UK (COG-UK) Consortium |
| EPI_ISL_425369 | Department of Pathology, University of Cambridge | COVID-19 Genomics UK (COG-UK) Consortium |
| EPI_ISL_425370 | Department of Pathology, University of Cambridge | COVID-19 Genomics UK (COG-UK) Consortium |
| EPI_ISL_425371 | Department of Pathology, University of Cambridge | COVID-19 Genomics UK (COG-UK) Consortium |
| EPI_ISL_425373 | Department of Pathology, University of Cambridge | COVID-19 Genomics UK (COG-UK) Consortium |
| EPI_ISL_425374 | Department of Pathology, University of Cambridge | COVID-19 Genomics UK (COG-UK) Consortium |
| EPI_ISL_425375 | Department of Pathology, University of Cambridge | COVID-19 Genomics UK (COG-UK) Consortium |
| EPI_ISL_425376 | Department of Pathology, University of Cambridge | COVID-19 Genomics UK (COG-UK) Consortium |
| EPI_ISL_425377 | Department of Pathology, University of Cambridge | COVID-19 Genomics UK (COG-UK) Consortium |
| EPI_ISL_425378 | Department of Pathology, University of Cambridge | COVID-19 Genomics UK (COG-UK) Consortium |
| EPI_ISL_425379 | Department of Pathology, University of Cambridge | COVID-19 Genomics UK (COG-UK) Consortium |
| EPI_ISL_425380 | Department of Pathology, University of Cambridge | COVID-19 Genomics UK (COG-UK) Consortium |
| EPI_ISL_425381 | Department of Pathology, University of Cambridge | COVID-19 Genomics UK (COG-UK) Consortium |
| EPI_ISL_425382 | Department of Pathology, University of Cambridge | COVID-19 Genomics UK (COG-UK) Consortium |
| EPI_ISL_425383 | Department of Pathology, University of Cambridge | COVID-19 Genomics UK (COG-UK) Consortium |
| EPI_ISL_425384 | Department of Pathology, University of Cambridge | COVID-19 Genomics UK (COG-UK) Consortium |
| EPI_ISL_425385 | Department of Pathology, University of Cambridge | COVID-19 Genomics UK (COG-UK) Consortium |
| EPI_ISL_425387 | Department of Pathology, University of Cambridge | COVID-19 Genomics UK (COG-UK) Consortium |
| EPI_ISL_425388 | Department of Pathology, University of Cambridge | COVID-19 Genomics UK (COG-UK) Consortium |
| EPI_ISL_425389 | Department of Pathology, University of Cambridge | COVID-19 Genomics UK (COG-UK) Consortium |
| EPI_ISL_425390 | Department of Pathology, University of Cambridge | COVID-19 Genomics UK (COG-UK) Consortium |
| EPI_ISL_425391 | Department of Pathology, University of Cambridge | COVID-19 Genomics UK (COG-UK) Consortium |
| EPI_ISL_425392 | Department of Pathology, University of Cambridge | COVID-19 Genomics UK (COG-UK) Consortium |
| EPI_ISL_425393 | Department of Pathology, University of Cambridge | COVID-19 Genomics UK (COG-UK) Consortium |
| EPI_ISL_425394 | Department of Pathology, University of Cambridge | COVID-19 Genomics UK (COG-UK) Consortium |
| EPI_ISL_425395 | Department of Pathology, University of Cambridge | COVID-19 Genomics UK (COG-UK) Consortium |
| EPI_ISL_425396 | Department of Pathology, University of Cambridge | COVID-19 Genomics UK (COG-UK) Consortium |
| EPI_ISL_425397 | Department of Pathology, University of Cambridge | COVID-19 Genomics UK (COG-UK) Consortium |
| EPI_ISL_425398 | Department of Pathology, University of Cambridge | COVID-19 Genomics UK (COG-UK) Consortium |
| EPI_ISL_425399 | Department of Pathology, University of Cambridge | COVID-19 Genomics UK (COG-UK) Consortium |
| EPI_ISL_425400 | Department of Pathology, University of Cambridge | COVID-19 Genomics UK (COG-UK) Consortium |

| EPI_Accession_ID | Originating Laboratory | Submitting Laboratory |
|---|---|---|
| EPI_ISL_425401 | Department of Pathology, University of Cambridge | COVID-19 Genomics UK (COG-UK) Consortium |
| EPI_ISL_425402 | Department of Pathology, University of Cambridge | COVID-19 Genomics UK (COG-UK) Consortium |
| EPI_ISL_425403 | Department of Pathology, University of Cambridge | COVID-19 Genomics UK (COG-UK) Consortium |
| EPI_ISL_425404 | Department of Pathology, University of Cambridge | COVID-19 Genomics UK (COG-UK) Consortium |
| EPI_ISL_425406 | Department of Pathology, University of Cambridge | COVID-19 Genomics UK (COG-UK) Consortium |
| EPI_ISL_425407 | Department of Pathology, University of Cambridge | COVID-19 Genomics UK (COG-UK) Consortium |
| EPI_ISL_425408 | Department of Pathology, University of Cambridge | COVID-19 Genomics UK (COG-UK) Consortium |
| EPI_ISL_425409 | Department of Pathology, University of Cambridge | COVID-19 Genomics UK (COG-UK) Consortium |
| EPI_ISL_425411 | Department of Pathology, University of Cambridge | COVID-19 Genomics UK (COG-UK) Consortium |
| EPI_ISL_425412 | Department of Pathology, University of Cambridge | COVID-19 Genomics UK (COG-UK) Consortium |
| EPI_ISL_425413 | Department of Pathology, University of Cambridge | COVID-19 Genomics UK (COG-UK) Consortium |
| EPI_ISL_425414 | Department of Pathology, University of Cambridge | COVID-19 Genomics UK (COG-UK) Consortium |
| EPI_ISL_425415 | Department of Pathology, University of Cambridge | COVID-19 Genomics UK (COG-UK) Consortium |
| EPI_ISL_425416 | Department of Pathology, University of Cambridge | COVID-19 Genomics UK (COG-UK) Consortium |
| EPI_ISL_425417 | Department of Pathology, University of Cambridge | COVID-19 Genomics UK (COG-UK) Consortium |
| EPI_ISL_425418 | Department of Pathology, University of Cambridge | COVID-19 Genomics UK (COG-UK) Consortium |
| EPI_ISL_425420 | Department of Pathology, University of Cambridge | COVID-19 Genomics UK (COG-UK) Consortium |
| EPI_ISL_425421 | Department of Pathology, University of Cambridge | COVID-19 Genomics UK (COG-UK) Consortium |
| EPI_ISL_425423 | Department of Pathology, University of Cambridge | COVID-19 Genomics UK (COG-UK) Consortium |
| EPI_ISL_425424 | Department of Pathology, University of Cambridge | COVID-19 Genomics UK (COG-UK) Consortium |
| EPI_ISL_425425 | Department of Pathology, University of Cambridge | COVID-19 Genomics UK (COG-UK) Consortium |
| EPI_ISL_425426 | Department of Pathology, University of Cambridge | COVID-19 Genomics UK (COG-UK) Consortium |
| EPI_ISL_425427 | Department of Pathology, University of Cambridge | COVID-19 Genomics UK (COG-UK) Consortium |
| EPI_ISL_425428 | Department of Pathology, University of Cambridge | COVID-19 Genomics UK (COG-UK) Consortium |
| EPI_ISL_425429 | Department of Pathology, University of Cambridge | COVID-19 Genomics UK (COG-UK) Consortium |
| EPI_ISL_425430 | Department of Pathology, University of Cambridge | COVID-19 Genomics UK (COG-UK) Consortium |
| EPI_ISL_425431 | Department of Pathology, University of Cambridge | COVID-19 Genomics UK (COG-UK) Consortium |
| EPI_ISL_425432 | Department of Pathology, University of Cambridge | COVID-19 Genomics UK (COG-UK) Consortium |
| EPI_ISL_425433 | Department of Pathology, University of Cambridge | COVID-19 Genomics UK (COG-UK) Consortium |
| EPI_ISL_425434 | Department of Pathology, University of Cambridge | COVID-19 Genomics UK (COG-UK) Consortium |
| EPI_ISL_425435 | Department of Pathology, University of Cambridge | COVID-19 Genomics UK (COG-UK) Consortium |
| EPI_ISL_425436 | Department of Pathology, University of Cambridge | COVID-19 Genomics UK (COG-UK) Consortium |
| EPI_ISL_425437 | Department of Pathology, University of Cambridge | COVID-19 Genomics UK (COG-UK) Consortium |

| EPI_Accession_ID | Originating Laboratory | Submitting Laboratory |
|---|---|---|
| EPI_ISL_425438 | Department of Pathology, University of Cambridge | COVID-19 Genomics UK (COG-UK) Consortium |
| EPI_ISL_425441 | Department of Pathology, University of Cambridge | COVID-19 Genomics UK (COG-UK) Consortium |
| EPI_ISL_425442 | Department of Pathology, University of Cambridge | COVID-19 Genomics UK (COG-UK) Consortium |
| EPI_ISL_425443 | Department of Pathology, University of Cambridge | COVID-19 Genomics UK (COG-UK) Consortium |
| EPI_ISL_425444 | Department of Pathology, University of Cambridge | COVID-19 Genomics UK (COG-UK) Consortium |
| EPI_ISL_425445 | Department of Pathology, University of Cambridge | COVID-19 Genomics UK (COG-UK) Consortium |
| EPI_ISL_425446 | Department of Pathology, University of Cambridge | COVID-19 Genomics UK (COG-UK) Consortium |
| EPI_ISL_425447 | Department of Pathology, University of Cambridge | COVID-19 Genomics UK (COG-UK) Consortium |
| EPI_ISL_425448 | Department of Pathology, University of Cambridge | COVID-19 Genomics UK (COG-UK) Consortium |
| EPI_ISL_425449 | Department of Pathology, University of Cambridge | COVID-19 Genomics UK (COG-UK) Consortium |
| EPI_ISL_425450 | Department of Pathology, University of Cambridge | COVID-19 Genomics UK (COG-UK) Consortium |
| EPI_ISL_425451 | Department of Pathology, University of Cambridge | COVID-19 Genomics UK (COG-UK) Consortium |
| EPI_ISL_425452 | Department of Pathology, University of Cambridge | COVID-19 Genomics UK (COG-UK) Consortium |
| EPI_ISL_425453 | Department of Pathology, University of Cambridge | COVID-19 Genomics UK (COG-UK) Consortium |
| EPI_ISL_425454 | Department of Pathology, University of Cambridge | COVID-19 Genomics UK (COG-UK) Consortium |
| EPI_ISL_425455 | Department of Pathology, University of Cambridge | COVID-19 Genomics UK (COG-UK) Consortium |
| EPI_ISL_425456 | Department of Pathology, University of Cambridge | COVID-19 Genomics UK (COG-UK) Consortium |
| EPI_ISL_425457 | Department of Pathology, University of Cambridge | COVID-19 Genomics UK (COG-UK) Consortium |
| EPI_ISL_425458 | Department of Pathology, University of Cambridge | COVID-19 Genomics UK (COG-UK) Consortium |
| EPI_ISL_425459 | Department of Pathology, University of Cambridge | COVID-19 Genomics UK (COG-UK) Consortium |
| EPI_ISL_425460 | Department of Pathology, University of Cambridge | COVID-19 Genomics UK (COG-UK) Consortium |
| EPI_ISL_425461 | Department of Pathology, University of Cambridge | COVID-19 Genomics UK (COG-UK) Consortium |
| EPI_ISL_425462 | Queens Medical Centre, Clinical Microbiology Department / DeepSeq Nottingham | COVID-19 Genomics UK (COG-UK) Consortium |
| EPI_ISL_425463 | Queens Medical Centre, Clinical Microbiology Department / DeepSeq Nottingham | COVID-19 Genomics UK (COG-UK) Consortium |
| EPI_ISL_425464 | Queens Medical Centre, Clinical Microbiology Department / DeepSeq Nottingham | COVID-19 Genomics UK (COG-UK) Consortium |
| EPI_ISL_425465 | Queens Medical Centre, Clinical Microbiology Department / DeepSeq Nottingham | COVID-19 Genomics UK (COG-UK) Consortium |
| EPI_ISL_425466 | Queens Medical Centre, Clinical Microbiology Department / DeepSeq Nottingham | COVID-19 Genomics UK (COG-UK) Consortium |

| EPI_Accession_ID | Originating Laboratory | Submitting Laboratory |
|---|---|---|
| EPI_ISL_425467 | Queens Medical Centre, Clinical Microbiology Department / DeepSeq Nottingham | COVID-19 Genomics UK (COG-UK) Consortium |
| EPI_ISL_425468 | Queens Medical Centre, Clinical Microbiology Department / DeepSeq Nottingham | COVID-19 Genomics UK (COG-UK) Consortium |
| EPI_ISL_425469 | Queens Medical Centre, Clinical Microbiology Department / DeepSeq Nottingham | COVID-19 Genomics UK (COG-UK) Consortium |
| EPI_ISL_425470 | Queens Medical Centre, Clinical Microbiology Department / DeepSeq Nottingham | COVID-19 Genomics UK (COG-UK) Consortium |
| EPI_ISL_425471 | Queens Medical Centre, Clinical Microbiology Department / DeepSeq Nottingham | COVID-19 Genomics UK (COG-UK) Consortium |
| EPI_ISL_425472 | Queens Medical Centre, Clinical Microbiology Department / DeepSeq Nottingham | COVID-19 Genomics UK (COG-UK) Consortium |
| EPI_ISL_425473 | Queens Medical Centre, Clinical Microbiology Department / DeepSeq Nottingham | COVID-19 Genomics UK (COG-UK) Consortium |
| EPI_ISL_425474 | Queens Medical Centre, Clinical Microbiology Department / DeepSeq Nottingham | COVID-19 Genomics UK (COG-UK) Consortium |
| EPI_ISL_425475 | Queens Medical Centre, Clinical Microbiology Department / DeepSeq Nottingham | COVID-19 Genomics UK (COG-UK) Consortium |
| EPI_ISL_425476 | Queens Medical Centre, Clinical Microbiology Department / DeepSeq Nottingham | COVID-19 Genomics UK (COG-UK) Consortium |
| EPI_ISL_425477 | Queens Medical Centre, Clinical Microbiology Department / DeepSeq Nottingham | COVID-19 Genomics UK (COG-UK) Consortium |
| EPI_ISL_425478 | Queens Medical Centre, Clinical Microbiology Department / DeepSeq Nottingham | COVID-19 Genomics UK (COG-UK) Consortium |
| EPI_ISL_425479 | Queens Medical Centre, Clinical Microbiology Department / DeepSeq Nottingham | COVID-19 Genomics UK (COG-UK) Consortium |
| EPI_ISL_425480 | Queens Medical Centre, Clinical Microbiology Department / DeepSeq Nottingham | COVID-19 Genomics UK (COG-UK) Consortium |
| EPI_ISL_425481 | Queens Medical Centre, Clinical Microbiology Department / DeepSeq Nottingham | COVID-19 Genomics UK (COG-UK) Consortium |
| EPI_ISL_425482 | Queens Medical Centre, Clinical Microbiology Department / DeepSeq Nottingham | COVID-19 Genomics UK (COG-UK) Consortium |

| EPI_Accession_ID | Originating Laboratory | Submitting Laboratory |
| --- | --- | --- |
| EPI_ISL_425483 | Queens Medical Centre, Clinical Microbiology Department / DeepSeq Nottingham | COVID-19 Genomics UK (COG-UK) Consortium |
| EPI_ISL_425484 | Queens Medical Centre, Clinical Microbiology Department / DeepSeq Nottingham | COVID-19 Genomics UK (COG-UK) Consortium |
| EPI_ISL_425485 | Queens Medical Centre, Clinical Microbiology Department / DeepSeq Nottingham | COVID-19 Genomics UK (COG-UK) Consortium |
| EPI_ISL_425486 | Queens Medical Centre, Clinical Microbiology Department / DeepSeq Nottingham | COVID-19 Genomics UK (COG-UK) Consortium |
| EPI_ISL_425487 | Queens Medical Centre, Clinical Microbiology Department / DeepSeq Nottingham | COVID-19 Genomics UK (COG-UK) Consortium |
| EPI_ISL_425488 | Queens Medical Centre, Clinical Microbiology Department / DeepSeq Nottingham | COVID-19 Genomics UK (COG-UK) Consortium |
| EPI_ISL_425489 | Queens Medical Centre, Clinical Microbiology Department / DeepSeq Nottingham | COVID-19 Genomics UK (COG-UK) Consortium |
| EPI_ISL_425490 | Queens Medical Centre, Clinical Microbiology Department / DeepSeq Nottingham | COVID-19 Genomics UK (COG-UK) Consortium |
| EPI_ISL_425491 | Queens Medical Centre, Clinical Microbiology Department / DeepSeq Nottingham | COVID-19 Genomics UK (COG-UK) Consortium |
| EPI_ISL_425492 | Queens Medical Centre, Clinical Microbiology Department / DeepSeq Nottingham | COVID-19 Genomics UK (COG-UK) Consortium |
| EPI_ISL_425493 | Queens Medical Centre, Clinical Microbiology Department / DeepSeq Nottingham | COVID-19 Genomics UK (COG-UK) Consortium |
| EPI_ISL_425494 | Queens Medical Centre, Clinical Microbiology Department / DeepSeq Nottingham | COVID-19 Genomics UK (COG-UK) Consortium |
| EPI_ISL_425495 | Queens Medical Centre, Clinical Microbiology Department / DeepSeq Nottingham | COVID-19 Genomics UK (COG-UK) Consortium |
| EPI_ISL_425496 | Queens Medical Centre, Clinical Microbiology Department / DeepSeq Nottingham | COVID-19 Genomics UK (COG-UK) Consortium |
| EPI_ISL_425497 | Queens Medical Centre, Clinical Microbiology Department / DeepSeq Nottingham | COVID-19 Genomics UK (COG-UK) Consortium |
| EPI_ISL_425498 | Queens Medical Centre, Clinical Microbiology Department / DeepSeq Nottingham | COVID-19 Genomics UK (COG-UK) Consortium |

| EPI_Accession_ID | Originating Laboratory | Submitting Laboratory |
|---|---|---|
| EPI_ISL_425499 | Queens Medical Centre, Clinical Microbiology Department / DeepSeq Nottingham | COVID-19 Genomics UK (COG-UK) Consortium |
| EPI_ISL_425500 | Queens Medical Centre, Clinical Microbiology Department / DeepSeq Nottingham | COVID-19 Genomics UK (COG-UK) Consortium |
| EPI_ISL_425501 | Queens Medical Centre, Clinical Microbiology Department / DeepSeq Nottingham | COVID-19 Genomics UK (COG-UK) Consortium |
| EPI_ISL_425502 | Queens Medical Centre, Clinical Microbiology Department / DeepSeq Nottingham | COVID-19 Genomics UK (COG-UK) Consortium |
| EPI_ISL_425503 | Queens Medical Centre, Clinical Microbiology Department / DeepSeq Nottingham | COVID-19 Genomics UK (COG-UK) Consortium |
| EPI_ISL_425504 | Queens Medical Centre, Clinical Microbiology Department / DeepSeq Nottingham | COVID-19 Genomics UK (COG-UK) Consortium |
| EPI_ISL_425505 | Queens Medical Centre, Clinical Microbiology Department / DeepSeq Nottingham | COVID-19 Genomics UK (COG-UK) Consortium |
| EPI_ISL_425506 | Queens Medical Centre, Clinical Microbiology Department / DeepSeq Nottingham | COVID-19 Genomics UK (COG-UK) Consortium |
| EPI_ISL_425507 | Queens Medical Centre, Clinical Microbiology Department / DeepSeq Nottingham | COVID-19 Genomics UK (COG-UK) Consortium |
| EPI_ISL_425508 | Queens Medical Centre, Clinical Microbiology Department / DeepSeq Nottingham | COVID-19 Genomics UK (COG-UK) Consortium |
| EPI_ISL_425509 | Queens Medical Centre, Clinical Microbiology Department / DeepSeq Nottingham | COVID-19 Genomics UK (COG-UK) Consortium |
| EPI_ISL_425510 | Queens Medical Centre, Clinical Microbiology Department / DeepSeq Nottingham | COVID-19 Genomics UK (COG-UK) Consortium |
| EPI_ISL_425511 | Queens Medical Centre, Clinical Microbiology Department / DeepSeq Nottingham | COVID-19 Genomics UK (COG-UK) Consortium |
| EPI_ISL_425512 | Queens Medical Centre, Clinical Microbiology Department / DeepSeq Nottingham | COVID-19 Genomics UK (COG-UK) Consortium |
| EPI_ISL_425513 | Queens Medical Centre, Clinical Microbiology Department / DeepSeq Nottingham | COVID-19 Genomics UK (COG-UK) Consortium |
| EPI_ISL_425514 | Queens Medical Centre, Clinical Microbiology Department / DeepSeq Nottingham | COVID-19 Genomics UK (COG-UK) Consortium |

| EPI_Accession_ID | Originating Laboratory | Submitting Laboratory |
|---|---|---|
| EPI_ISL_425515 | Queens Medical Centre, Clinical Microbiology Department / DeepSeq Nottingham | COVID-19 Genomics UK (COG-UK) Consortium |
| EPI_ISL_425516 | Queens Medical Centre, Clinical Microbiology Department / DeepSeq Nottingham | COVID-19 Genomics UK (COG-UK) Consortium |
| EPI_ISL_425517 | Queens Medical Centre, Clinical Microbiology Department / DeepSeq Nottingham | COVID-19 Genomics UK (COG-UK) Consortium |
| EPI_ISL_425518 | Queens Medical Centre, Clinical Microbiology Department / DeepSeq Nottingham | COVID-19 Genomics UK (COG-UK) Consortium |
| EPI_ISL_425519 | Queens Medical Centre, Clinical Microbiology Department / DeepSeq Nottingham | COVID-19 Genomics UK (COG-UK) Consortium |
| EPI_ISL_425520 | Queens Medical Centre, Clinical Microbiology Department / DeepSeq Nottingham | COVID-19 Genomics UK (COG-UK) Consortium |
| EPI_ISL_425521 | Queens Medical Centre, Clinical Microbiology Department / DeepSeq Nottingham | COVID-19 Genomics UK (COG-UK) Consortium |
| EPI_ISL_425522 | Queens Medical Centre, Clinical Microbiology Department / DeepSeq Nottingham | COVID-19 Genomics UK (COG-UK) Consortium |
| EPI_ISL_425523 | Queens Medical Centre, Clinical Microbiology Department / DeepSeq Nottingham | COVID-19 Genomics UK (COG-UK) Consortium |
| EPI_ISL_425524 | Queens Medical Centre, Clinical Microbiology Department / DeepSeq Nottingham | COVID-19 Genomics UK (COG-UK) Consortium |
| EPI_ISL_425525 | Queens Medical Centre, Clinical Microbiology Department / DeepSeq Nottingham | COVID-19 Genomics UK (COG-UK) Consortium |
| EPI_ISL_425526 | Queens Medical Centre, Clinical Microbiology Department / DeepSeq Nottingham | COVID-19 Genomics UK (COG-UK) Consortium |
| EPI_ISL_425527 | Queens Medical Centre, Clinical Microbiology Department / DeepSeq Nottingham | COVID-19 Genomics UK (COG-UK) Consortium |
| EPI_ISL_425528 | Queens Medical Centre, Clinical Microbiology Department / DeepSeq Nottingham | COVID-19 Genomics UK (COG-UK) Consortium |
| EPI_ISL_425529 | Queens Medical Centre, Clinical Microbiology Department / DeepSeq Nottingham | COVID-19 Genomics UK (COG-UK) Consortium |
| EPI_ISL_425530 | Queens Medical Centre, Clinical Microbiology Department / DeepSeq Nottingham | COVID-19 Genomics UK (COG-UK) Consortium |

| EPI_Accession_ID | Originating Laboratory | Submitting Laboratory |
|---|---|---|
| EPI_ISL_425531 | Queens Medical Centre, Clinical Microbiology Department / DeepSeq Nottingham | COVID-19 Genomics UK (COG-UK) Consortium |
| EPI_ISL_425532 | Queens Medical Centre, Clinical Microbiology Department / DeepSeq Nottingham | COVID-19 Genomics UK (COG-UK) Consortium |
| EPI_ISL_425533 | Queens Medical Centre, Clinical Microbiology Department / DeepSeq Nottingham | COVID-19 Genomics UK (COG-UK) Consortium |
| EPI_ISL_425534 | Queens Medical Centre, Clinical Microbiology Department / DeepSeq Nottingham | COVID-19 Genomics UK (COG-UK) Consortium |
| EPI_ISL_425535 | Queens Medical Centre, Clinical Microbiology Department / DeepSeq Nottingham | COVID-19 Genomics UK (COG-UK) Consortium |
| EPI_ISL_425536 | Queens Medical Centre, Clinical Microbiology Department / DeepSeq Nottingham | COVID-19 Genomics UK (COG-UK) Consortium |
| EPI_ISL_425537 | Queens Medical Centre, Clinical Microbiology Department / DeepSeq Nottingham | COVID-19 Genomics UK (COG-UK) Consortium |
| EPI_ISL_425538 | Queens Medical Centre, Clinical Microbiology Department / DeepSeq Nottingham | COVID-19 Genomics UK (COG-UK) Consortium |
| EPI_ISL_425539 | Queens Medical Centre, Clinical Microbiology Department / DeepSeq Nottingham | COVID-19 Genomics UK (COG-UK) Consortium |
| EPI_ISL_425540 | Queens Medical Centre, Clinical Microbiology Department / DeepSeq Nottingham | COVID-19 Genomics UK (COG-UK) Consortium |
| EPI_ISL_425541 | Queens Medical Centre, Clinical Microbiology Department / DeepSeq Nottingham | COVID-19 Genomics UK (COG-UK) Consortium |
| EPI_ISL_425542 | Queens Medical Centre, Clinical Microbiology Department / DeepSeq Nottingham | COVID-19 Genomics UK (COG-UK) Consortium |
| EPI_ISL_425543 | Queens Medical Centre, Clinical Microbiology Department / DeepSeq Nottingham | COVID-19 Genomics UK (COG-UK) Consortium |
| EPI_ISL_425544 | Queens Medical Centre, Clinical Microbiology Department / DeepSeq Nottingham | COVID-19 Genomics UK (COG-UK) Consortium |
| EPI_ISL_425545 | Queens Medical Centre, Clinical Microbiology Department / DeepSeq Nottingham | COVID-19 Genomics UK (COG-UK) Consortium |
| EPI_ISL_425546 | Queens Medical Centre, Clinical Microbiology Department / DeepSeq Nottingham | COVID-19 Genomics UK (COG-UK) Consortium |

| EPI_Accession_ID | Originating Laboratory | Submitting Laboratory |
| --- | --- | --- |
| EPI_ISL_425547 | Queens Medical Centre, Clinical Microbiology Department / DeepSeq Nottingham | COVID-19 Genomics UK (COG-UK) Consortium |
| EPI_ISL_425548 | Queens Medical Centre, Clinical Microbiology Department / DeepSeq Nottingham | COVID-19 Genomics UK (COG-UK) Consortium |
| EPI_ISL_425549 | Queens Medical Centre, Clinical Microbiology Department / DeepSeq Nottingham | COVID-19 Genomics UK (COG-UK) Consortium |
| EPI_ISL_425550 | Queens Medical Centre, Clinical Microbiology Department / DeepSeq Nottingham | COVID-19 Genomics UK (COG-UK) Consortium |
| EPI_ISL_425551 | Queens Medical Centre, Clinical Microbiology Department / DeepSeq Nottingham | COVID-19 Genomics UK (COG-UK) Consortium |
| EPI_ISL_425552 | Queens Medical Centre, Clinical Microbiology Department / DeepSeq Nottingham | COVID-19 Genomics UK (COG-UK) Consortium |
| EPI_ISL_425553 | Queens Medical Centre, Clinical Microbiology Department / DeepSeq Nottingham | COVID-19 Genomics UK (COG-UK) Consortium |
| EPI_ISL_425554 | Queens Medical Centre, Clinical Microbiology Department / DeepSeq Nottingham | COVID-19 Genomics UK (COG-UK) Consortium |
| EPI_ISL_425555 | Queens Medical Centre, Clinical Microbiology Department / DeepSeq Nottingham | COVID-19 Genomics UK (COG-UK) Consortium |
| EPI_ISL_425556 | Queens Medical Centre, Clinical Microbiology Department / DeepSeq Nottingham | COVID-19 Genomics UK (COG-UK) Consortium |
| EPI_ISL_425557 | Queens Medical Centre, Clinical Microbiology Department / DeepSeq Nottingham | COVID-19 Genomics UK (COG-UK) Consortium |
| EPI_ISL_425558 | Queens Medical Centre, Clinical Microbiology Department / DeepSeq Nottingham | COVID-19 Genomics UK (COG-UK) Consortium |
| EPI_ISL_425559 | Queens Medical Centre, Clinical Microbiology Department / DeepSeq Nottingham | COVID-19 Genomics UK (COG-UK) Consortium |
| EPI_ISL_425560 | Queens Medical Centre, Clinical Microbiology Department / DeepSeq Nottingham | COVID-19 Genomics UK (COG-UK) Consortium |
| EPI_ISL_425561 | Queens Medical Centre, Clinical Microbiology Department / DeepSeq Nottingham | COVID-19 Genomics UK (COG-UK) Consortium |
| EPI_ISL_425562 | Queens Medical Centre, Clinical Microbiology Department / DeepSeq Nottingham | COVID-19 Genomics UK (COG-UK) Consortium |

| EPI_Accession_ID | Originating Laboratory | Submitting Laboratory |
|---|---|---|
| EPI_ISL_425563 | Queens Medical Centre, Clinical Microbiology Department / DeepSeq Nottingham | COVID-19 Genomics UK (COG-UK) Consortium |
| EPI_ISL_425564 | Queens Medical Centre, Clinical Microbiology Department / DeepSeq Nottingham | COVID-19 Genomics UK (COG-UK) Consortium |
| EPI_ISL_425565 | Queens Medical Centre, Clinical Microbiology Department / DeepSeq Nottingham | COVID-19 Genomics UK (COG-UK) Consortium |
| EPI_ISL_425566 | Queens Medical Centre, Clinical Microbiology Department / DeepSeq Nottingham | COVID-19 Genomics UK (COG-UK) Consortium |
| EPI_ISL_425567 | Queens Medical Centre, Clinical Microbiology Department / DeepSeq Nottingham | COVID-19 Genomics UK (COG-UK) Consortium |
| EPI_ISL_425568 | Queens Medical Centre, Clinical Microbiology Department / DeepSeq Nottingham | COVID-19 Genomics UK (COG-UK) Consortium |
| EPI_ISL_425569 | Queens Medical Centre, Clinical Microbiology Department / DeepSeq Nottingham | COVID-19 Genomics UK (COG-UK) Consortium |
| EPI_ISL_425570 | Queens Medical Centre, Clinical Microbiology Department / DeepSeq Nottingham | COVID-19 Genomics UK (COG-UK) Consortium |
| EPI_ISL_425571 | Queens Medical Centre, Clinical Microbiology Department / DeepSeq Nottingham | COVID-19 Genomics UK (COG-UK) Consortium |
| EPI_ISL_425572 | Queens Medical Centre, Clinical Microbiology Department / DeepSeq Nottingham | COVID-19 Genomics UK (COG-UK) Consortium |
| EPI_ISL_425573 | Queens Medical Centre, Clinical Microbiology Department / DeepSeq Nottingham | COVID-19 Genomics UK (COG-UK) Consortium |
| EPI_ISL_425574 | Queens Medical Centre, Clinical Microbiology Department / DeepSeq Nottingham | COVID-19 Genomics UK (COG-UK) Consortium |
| EPI_ISL_425575 | Queens Medical Centre, Clinical Microbiology Department / DeepSeq Nottingham | COVID-19 Genomics UK (COG-UK) Consortium |
| EPI_ISL_425576 | Queens Medical Centre, Clinical Microbiology Department / DeepSeq Nottingham | COVID-19 Genomics UK (COG-UK) Consortium |
| EPI_ISL_425577 | Queens Medical Centre, Clinical Microbiology Department / DeepSeq Nottingham | COVID-19 Genomics UK (COG-UK) Consortium |
| EPI_ISL_425579 | Queens Medical Centre, Clinical Microbiology Department / DeepSeq Nottingham | COVID-19 Genomics UK (COG-UK) Consortium |

| EPI_Accession_ID | Originating Laboratory | Submitting Laboratory |
|---|---|---|
| EPI_ISL_425580 | Queens Medical Centre, Clinical Microbiology Department / DeepSeq Nottingham | COVID-19 Genomics UK (COG-UK) Consortium |
| EPI_ISL_425581 | Queens Medical Centre, Clinical Microbiology Department / DeepSeq Nottingham | COVID-19 Genomics UK (COG-UK) Consortium |
| EPI_ISL_425582 | Queens Medical Centre, Clinical Microbiology Department / DeepSeq Nottingham | COVID-19 Genomics UK (COG-UK) Consortium |
| EPI_ISL_425583 | Queens Medical Centre, Clinical Microbiology Department / DeepSeq Nottingham | COVID-19 Genomics UK (COG-UK) Consortium |
| EPI_ISL_425584 | Queens Medical Centre, Clinical Microbiology Department / DeepSeq Nottingham | COVID-19 Genomics UK (COG-UK) Consortium |
| EPI_ISL_425585 | Queens Medical Centre, Clinical Microbiology Department / DeepSeq Nottingham | COVID-19 Genomics UK (COG-UK) Consortium |
| EPI_ISL_425586 | Queens Medical Centre, Clinical Microbiology Department / DeepSeq Nottingham | COVID-19 Genomics UK (COG-UK) Consortium |
| EPI_ISL_425587 | Queens Medical Centre, Clinical Microbiology Department / DeepSeq Nottingham | COVID-19 Genomics UK (COG-UK) Consortium |
| EPI_ISL_425588 | Queens Medical Centre, Clinical Microbiology Department / DeepSeq Nottingham | COVID-19 Genomics UK (COG-UK) Consortium |
| EPI_ISL_425589 | Queens Medical Centre, Clinical Microbiology Department / DeepSeq Nottingham | COVID-19 Genomics UK (COG-UK) Consortium |
| EPI_ISL_425590 | Queens Medical Centre, Clinical Microbiology Department / DeepSeq Nottingham | COVID-19 Genomics UK (COG-UK) Consortium |
| EPI_ISL_425591 | Queens Medical Centre, Clinical Microbiology Department / DeepSeq Nottingham | COVID-19 Genomics UK (COG-UK) Consortium |
| EPI_ISL_425592 | Queens Medical Centre, Clinical Microbiology Department / DeepSeq Nottingham | COVID-19 Genomics UK (COG-UK) Consortium |
| EPI_ISL_425593 | Queens Medical Centre, Clinical Microbiology Department / DeepSeq Nottingham | COVID-19 Genomics UK (COG-UK) Consortium |
| EPI_ISL_425594 | Queens Medical Centre, Clinical Microbiology Department / DeepSeq Nottingham | COVID-19 Genomics UK (COG-UK) Consortium |
| EPI_ISL_425595 | Queens Medical Centre, Clinical Microbiology Department / DeepSeq Nottingham | COVID-19 Genomics UK (COG-UK) Consortium |

| EPI_Accession_ID | Originating Laboratory | Submitting Laboratory |
|---|---|---|
| EPI_ISL_425596 | Queens Medical Centre, Clinical Microbiology Department / DeepSeq Nottingham | COVID-19 Genomics UK (COG-UK) Consortium |
| EPI_ISL_425597 | Queens Medical Centre, Clinical Microbiology Department / DeepSeq Nottingham | COVID-19 Genomics UK (COG-UK) Consortium |
| EPI_ISL_425598 | Queens Medical Centre, Clinical Microbiology Department / DeepSeq Nottingham | COVID-19 Genomics UK (COG-UK) Consortium |
| EPI_ISL_425599 | Queens Medical Centre, Clinical Microbiology Department / DeepSeq Nottingham | COVID-19 Genomics UK (COG-UK) Consortium |
| EPI_ISL_425600 | Queens Medical Centre, Clinical Microbiology Department / DeepSeq Nottingham | COVID-19 Genomics UK (COG-UK) Consortium |
| EPI_ISL_425601 | Queens Medical Centre, Clinical Microbiology Department / DeepSeq Nottingham | COVID-19 Genomics UK (COG-UK) Consortium |
| EPI_ISL_425602 | Queens Medical Centre, Clinical Microbiology Department / DeepSeq Nottingham | COVID-19 Genomics UK (COG-UK) Consortium |
| EPI_ISL_425603 | Queens Medical Centre, Clinical Microbiology Department / DeepSeq Nottingham | COVID-19 Genomics UK (COG-UK) Consortium |
| EPI_ISL_425604 | Queens Medical Centre, Clinical Microbiology Department / DeepSeq Nottingham | COVID-19 Genomics UK (COG-UK) Consortium |
| EPI_ISL_425605 | Queens Medical Centre, Clinical Microbiology Department / DeepSeq Nottingham | COVID-19 Genomics UK (COG-UK) Consortium |
| EPI_ISL_425606 | Queens Medical Centre, Clinical Microbiology Department / DeepSeq Nottingham | COVID-19 Genomics UK (COG-UK) Consortium |
| EPI_ISL_425607 | Queens Medical Centre, Clinical Microbiology Department / DeepSeq Nottingham | COVID-19 Genomics UK (COG-UK) Consortium |
| EPI_ISL_425608 | Queens Medical Centre, Clinical Microbiology Department / DeepSeq Nottingham | COVID-19 Genomics UK (COG-UK) Consortium |
| EPI_ISL_425609 | Queens Medical Centre, Clinical Microbiology Department / DeepSeq Nottingham | COVID-19 Genomics UK (COG-UK) Consortium |
| EPI_ISL_425610 | Queens Medical Centre, Clinical Microbiology Department / DeepSeq Nottingham | COVID-19 Genomics UK (COG-UK) Consortium |
| EPI_ISL_425611 | Queens Medical Centre, Clinical Microbiology Department / DeepSeq Nottingham | COVID-19 Genomics UK (COG-UK) Consortium |

| EPI_Accession_ID | Originating Laboratory | Submitting Laboratory |
|---|---|---|
| EPI_ISL_425612 | Queens Medical Centre, Clinical Microbiology Department / DeepSeq Nottingham | COVID-19 Genomics UK (COG-UK) Consortium |
| EPI_ISL_425613 | Queens Medical Centre, Clinical Microbiology Department / DeepSeq Nottingham | COVID-19 Genomics UK (COG-UK) Consortium |
| EPI_ISL_425614 | Queens Medical Centre, Clinical Microbiology Department / DeepSeq Nottingham | COVID-19 Genomics UK (COG-UK) Consortium |
| EPI_ISL_425615 | Queens Medical Centre, Clinical Microbiology Department / DeepSeq Nottingham | COVID-19 Genomics UK (COG-UK) Consortium |
| EPI_ISL_425616 | Queens Medical Centre, Clinical Microbiology Department / DeepSeq Nottingham | COVID-19 Genomics UK (COG-UK) Consortium |
| EPI_ISL_425617 | Queens Medical Centre, Clinical Microbiology Department / DeepSeq Nottingham | COVID-19 Genomics UK (COG-UK) Consortium |
| EPI_ISL_425618 | Queens Medical Centre, Clinical Microbiology Department / DeepSeq Nottingham | COVID-19 Genomics UK (COG-UK) Consortium |
| EPI_ISL_425619 | Queens Medical Centre, Clinical Microbiology Department / DeepSeq Nottingham | COVID-19 Genomics UK (COG-UK) Consortium |
| EPI_ISL_425620 | Queens Medical Centre, Clinical Microbiology Department / DeepSeq Nottingham | COVID-19 Genomics UK (COG-UK) Consortium |
| EPI_ISL_425621 | Queens Medical Centre, Clinical Microbiology Department / DeepSeq Nottingham | COVID-19 Genomics UK (COG-UK) Consortium |
| EPI_ISL_425622 | Queens Medical Centre, Clinical Microbiology Department / DeepSeq Nottingham | COVID-19 Genomics UK (COG-UK) Consortium |
| EPI_ISL_425623 | Queens Medical Centre, Clinical Microbiology Department / DeepSeq Nottingham | COVID-19 Genomics UK (COG-UK) Consortium |
| EPI_ISL_425624 | Queens Medical Centre, Clinical Microbiology Department / DeepSeq Nottingham | COVID-19 Genomics UK (COG-UK) Consortium |
| EPI_ISL_425625 | Queens Medical Centre, Clinical Microbiology Department / DeepSeq Nottingham | COVID-19 Genomics UK (COG-UK) Consortium |
| EPI_ISL_425626 | Queens Medical Centre, Clinical Microbiology Department / DeepSeq Nottingham | COVID-19 Genomics UK (COG-UK) Consortium |
| EPI_ISL_425627 | Queens Medical Centre, Clinical Microbiology Department / DeepSeq Nottingham | COVID-19 Genomics UK (COG-UK) Consortium |

| EPI_Accession_ID | Originating Laboratory | Submitting Laboratory |
|---|---|---|
| EPI_ISL_425628 | Queens Medical Centre, Clinical Microbiology Department / DeepSeq Nottingham | COVID-19 Genomics UK (COG-UK) Consortium |
| EPI_ISL_425629 | Queens Medical Centre, Clinical Microbiology Department / DeepSeq Nottingham | COVID-19 Genomics UK (COG-UK) Consortium |
| EPI_ISL_425630 | Queens Medical Centre, Clinical Microbiology Department / DeepSeq Nottingham | COVID-19 Genomics UK (COG-UK) Consortium |
| EPI_ISL_425631 | Queens Medical Centre, Clinical Microbiology Department / DeepSeq Nottingham | COVID-19 Genomics UK (COG-UK) Consortium |
| EPI_ISL_425632 | Queens Medical Centre, Clinical Microbiology Department / DeepSeq Nottingham | COVID-19 Genomics UK (COG-UK) Consortium |
| EPI_ISL_425633 | Queens Medical Centre, Clinical Microbiology Department / DeepSeq Nottingham | COVID-19 Genomics UK (COG-UK) Consortium |
| EPI_ISL_425634 | Queens Medical Centre, Clinical Microbiology Department / DeepSeq Nottingham | COVID-19 Genomics UK (COG-UK) Consortium |
| EPI_ISL_425635 | Queens Medical Centre, Clinical Microbiology Department / DeepSeq Nottingham | COVID-19 Genomics UK (COG-UK) Consortium |
| EPI_ISL_425636 | Queens Medical Centre, Clinical Microbiology Department / DeepSeq Nottingham | COVID-19 Genomics UK (COG-UK) Consortium |
| EPI_ISL_425637 | Queens Medical Centre, Clinical Microbiology Department / DeepSeq Nottingham | COVID-19 Genomics UK (COG-UK) Consortium |
| EPI_ISL_425638 | Queens Medical Centre, Clinical Microbiology Department / DeepSeq Nottingham | COVID-19 Genomics UK (COG-UK) Consortium |
| EPI_ISL_425639 | Queens Medical Centre, Clinical Microbiology Department / DeepSeq Nottingham | COVID-19 Genomics UK (COG-UK) Consortium |
| EPI_ISL_425640 | Queens Medical Centre, Clinical Microbiology Department / DeepSeq Nottingham | COVID-19 Genomics UK (COG-UK) Consortium |
| EPI_ISL_425641 | Queens Medical Centre, Clinical Microbiology Department / DeepSeq Nottingham | COVID-19 Genomics UK (COG-UK) Consortium |
| EPI_ISL_425642 | Queens Medical Centre, Clinical Microbiology Department / DeepSeq Nottingham | COVID-19 Genomics UK (COG-UK) Consortium |
| EPI_ISL_425643 | Queens Medical Centre, Clinical Microbiology Department / DeepSeq Nottingham | COVID-19 Genomics UK (COG-UK) Consortium |

| EPI_Accession_ID | Originating Laboratory | Submitting Laboratory |
|---|---|---|
| EPI_ISL_425644 | Queens Medical Centre, Clinical Microbiology Department / DeepSeq Nottingham | COVID-19 Genomics UK (COG-UK) Consortium |
| EPI_ISL_425645 | Queens Medical Centre, Clinical Microbiology Department / DeepSeq Nottingham | COVID-19 Genomics UK (COG-UK) Consortium |
| EPI_ISL_425646 | Queens Medical Centre, Clinical Microbiology Department / DeepSeq Nottingham | COVID-19 Genomics UK (COG-UK) Consortium |
| EPI_ISL_425647 | West of Scotland Specialist Virology Centre, NHSGGC / MRC-University of Glasgow Centre for Virus Research | COVID-19 Genomics UK (COG-UK) Consortium |
| EPI_ISL_425648 | West of Scotland Specialist Virology Centre, NHSGGC / MRC-University of Glasgow Centre for Virus Research | COVID-19 Genomics UK (COG-UK) Consortium |
| EPI_ISL_425649 | West of Scotland Specialist Virology Centre, NHSGGC / MRC-University of Glasgow Centre for Virus Research | COVID-19 Genomics UK (COG-UK) Consortium |
| EPI_ISL_425650 | West of Scotland Specialist Virology Centre, NHSGGC / MRC-University of Glasgow Centre for Virus Research | COVID-19 Genomics UK (COG-UK) Consortium |
| EPI_ISL_425651 | West of Scotland Specialist Virology Centre, NHSGGC / MRC-University of Glasgow Centre for Virus Research | COVID-19 Genomics UK (COG-UK) Consortium |
| EPI_ISL_425652 | West of Scotland Specialist Virology Centre, NHSGGC / MRC-University of Glasgow Centre for Virus Research | COVID-19 Genomics UK (COG-UK) Consortium |
| EPI_ISL_425653 | West of Scotland Specialist Virology Centre, NHSGGC / MRC-University of Glasgow Centre for Virus Research | COVID-19 Genomics UK (COG-UK) Consortium |
| EPI_ISL_425654 | West of Scotland Specialist Virology Centre, NHSGGC / MRC-University of Glasgow Centre for Virus Research | COVID-19 Genomics UK (COG-UK) Consortium |
| EPI_ISL_425655 | West of Scotland Specialist Virology Centre, NHSGGC / MRC-University of Glasgow Centre for Virus Research | COVID-19 Genomics UK (COG-UK) Consortium |
| EPI_ISL_425656 | West of Scotland Specialist Virology Centre, NHSGGC / MRC-University of Glasgow Centre for Virus Research | COVID-19 Genomics UK (COG-UK) Consortium |
| EPI_ISL_425657 | West of Scotland Specialist Virology Centre, NHSGGC / MRC-University of Glasgow Centre for Virus Research | COVID-19 Genomics UK (COG-UK) Consortium |
| EPI_ISL_425658 | West of Scotland Specialist Virology Centre, NHSGGC / MRC-University of Glasgow Centre for Virus Research | COVID-19 Genomics UK (COG-UK) Consortium |
| EPI_ISL_425659 | West of Scotland Specialist Virology Centre, NHSGGC / MRC-University of Glasgow Centre for Virus Research | COVID-19 Genomics UK (COG-UK) Consortium |

| EPI_Accession_ID | Originating Laboratory | Submitting Laboratory |
|---|---|---|
| EPI_ISL_425660 | West of Scotland Specialist Virology Centre, NHSGGC / MRC-University of Glasgow Centre for Virus Research | COVID-19 Genomics UK (COG-UK) Consortium |
| EPI_ISL_425661 | West of Scotland Specialist Virology Centre, NHSGGC / MRC-University of Glasgow Centre for Virus Research | COVID-19 Genomics UK (COG-UK) Consortium |
| EPI_ISL_425662 | West of Scotland Specialist Virology Centre, NHSGGC / MRC-University of Glasgow Centre for Virus Research | COVID-19 Genomics UK (COG-UK) Consortium |
| EPI_ISL_425663 | West of Scotland Specialist Virology Centre, NHSGGC / MRC-University of Glasgow Centre for Virus Research | COVID-19 Genomics UK (COG-UK) Consortium |
| EPI_ISL_425664 | West of Scotland Specialist Virology Centre, NHSGGC / MRC-University of Glasgow Centre for Virus Research | COVID-19 Genomics UK (COG-UK) Consortium |
| EPI_ISL_425665 | West of Scotland Specialist Virology Centre, NHSGGC / MRC-University of Glasgow Centre for Virus Research | COVID-19 Genomics UK (COG-UK) Consortium |
| EPI_ISL_425666 | West of Scotland Specialist Virology Centre, NHSGGC / MRC-University of Glasgow Centre for Virus Research | COVID-19 Genomics UK (COG-UK) Consortium |
| EPI_ISL_425667 | West of Scotland Specialist Virology Centre, NHSGGC / MRC-University of Glasgow Centre for Virus Research | COVID-19 Genomics UK (COG-UK) Consortium |
| EPI_ISL_425668 | West of Scotland Specialist Virology Centre, NHSGGC / MRC-University of Glasgow Centre for Virus Research | COVID-19 Genomics UK (COG-UK) Consortium |
| EPI_ISL_425669 | West of Scotland Specialist Virology Centre, NHSGGC / MRC-University of Glasgow Centre for Virus Research | COVID-19 Genomics UK (COG-UK) Consortium |
| EPI_ISL_425670 | West of Scotland Specialist Virology Centre, NHSGGC / MRC-University of Glasgow Centre for Virus Research | COVID-19 Genomics UK (COG-UK) Consortium |
| EPI_ISL_425671 | West of Scotland Specialist Virology Centre, NHSGGC / MRC-University of Glasgow Centre for Virus Research | COVID-19 Genomics UK (COG-UK) Consortium |
| EPI_ISL_425672 | West of Scotland Specialist Virology Centre, NHSGGC / MRC-University of Glasgow Centre for Virus Research | COVID-19 Genomics UK (COG-UK) Consortium |
| EPI_ISL_425673 | West of Scotland Specialist Virology Centre, NHSGGC / MRC-University of Glasgow Centre for Virus Research | COVID-19 Genomics UK (COG-UK) Consortium |
| EPI_ISL_425674 | West of Scotland Specialist Virology Centre, NHSGGC / MRC-University of Glasgow Centre for Virus Research | COVID-19 Genomics UK (COG-UK) Consortium |
| EPI_ISL_425675 | West of Scotland Specialist Virology Centre, NHSGGC / MRC-University of Glasgow Centre for Virus Research | COVID-19 Genomics UK (COG-UK) Consortium |

| EPI_Accession_ID | Originating Laboratory | Submitting Laboratory |
|---|---|---|
| EPI_ISL_425676 | West of Scotland Specialist Virology Centre, NHSGGC / MRC-University of Glasgow Centre for Virus Research | COVID-19 Genomics UK (COG-UK) Consortium |
| EPI_ISL_425677 | West of Scotland Specialist Virology Centre, NHSGGC / MRC-University of Glasgow Centre for Virus Research | COVID-19 Genomics UK (COG-UK) Consortium |
| EPI_ISL_425678 | West of Scotland Specialist Virology Centre, NHSGGC / MRC-University of Glasgow Centre for Virus Research | COVID-19 Genomics UK (COG-UK) Consortium |
| EPI_ISL_425679 | West of Scotland Specialist Virology Centre, NHSGGC / MRC-University of Glasgow Centre for Virus Research | COVID-19 Genomics UK (COG-UK) Consortium |
| EPI_ISL_425680 | West of Scotland Specialist Virology Centre, NHSGGC / MRC-University of Glasgow Centre for Virus Research | COVID-19 Genomics UK (COG-UK) Consortium |
| EPI_ISL_425681 | West of Scotland Specialist Virology Centre, NHSGGC / MRC-University of Glasgow Centre for Virus Research | COVID-19 Genomics UK (COG-UK) Consortium |
| EPI_ISL_425682 | West of Scotland Specialist Virology Centre, NHSGGC / MRC-University of Glasgow Centre for Virus Research | COVID-19 Genomics UK (COG-UK) Consortium |
| EPI_ISL_425683 | West of Scotland Specialist Virology Centre, NHSGGC / MRC-University of Glasgow Centre for Virus Research | COVID-19 Genomics UK (COG-UK) Consortium |
| EPI_ISL_425684 | West of Scotland Specialist Virology Centre, NHSGGC / MRC-University of Glasgow Centre for Virus Research | COVID-19 Genomics UK (COG-UK) Consortium |
| EPI_ISL_425685 | West of Scotland Specialist Virology Centre, NHSGGC / MRC-University of Glasgow Centre for Virus Research | COVID-19 Genomics UK (COG-UK) Consortium |
| EPI_ISL_425686 | West of Scotland Specialist Virology Centre, NHSGGC / MRC-University of Glasgow Centre for Virus Research | COVID-19 Genomics UK (COG-UK) Consortium |
| EPI_ISL_425687 | West of Scotland Specialist Virology Centre, NHSGGC / MRC-University of Glasgow Centre for Virus Research | COVID-19 Genomics UK (COG-UK) Consortium |
| EPI_ISL_425688 | West of Scotland Specialist Virology Centre, NHSGGC / MRC-University of Glasgow Centre for Virus Research | COVID-19 Genomics UK (COG-UK) Consortium |
| EPI_ISL_425689 | West of Scotland Specialist Virology Centre, NHSGGC / MRC-University of Glasgow Centre for Virus Research | COVID-19 Genomics UK (COG-UK) Consortium |
| EPI_ISL_425690 | West of Scotland Specialist Virology Centre, NHSGGC / MRC-University of Glasgow Centre for Virus Research | COVID-19 Genomics UK (COG-UK) Consortium |
| EPI_ISL_425691 | West of Scotland Specialist Virology Centre, NHSGGC / MRC-University of Glasgow Centre for Virus Research | COVID-19 Genomics UK (COG-UK) Consortium |

| EPI_Accession_ID | Originating Laboratory | Submitting Laboratory |
|---|---|---|
| EPI_ISL_425692 | West of Scotland Specialist Virology Centre, NHSGGC / MRC-University of Glasgow Centre for Virus Research | COVID-19 Genomics UK (COG-UK) Consortium |
| EPI_ISL_425693 | West of Scotland Specialist Virology Centre, NHSGGC / MRC-University of Glasgow Centre for Virus Research | COVID-19 Genomics UK (COG-UK) Consortium |
| EPI_ISL_425694 | West of Scotland Specialist Virology Centre, NHSGGC / MRC-University of Glasgow Centre for Virus Research | COVID-19 Genomics UK (COG-UK) Consortium |
| EPI_ISL_425695 | West of Scotland Specialist Virology Centre, NHSGGC / MRC-University of Glasgow Centre for Virus Research | COVID-19 Genomics UK (COG-UK) Consortium |
| EPI_ISL_425696 | West of Scotland Specialist Virology Centre, NHSGGC / MRC-University of Glasgow Centre for Virus Research | COVID-19 Genomics UK (COG-UK) Consortium |
| EPI_ISL_425697 | West of Scotland Specialist Virology Centre, NHSGGC / MRC-University of Glasgow Centre for Virus Research | COVID-19 Genomics UK (COG-UK) Consortium |
| EPI_ISL_425698 | West of Scotland Specialist Virology Centre, NHSGGC / MRC-University of Glasgow Centre for Virus Research | COVID-19 Genomics UK (COG-UK) Consortium |
| EPI_ISL_425699 | West of Scotland Specialist Virology Centre, NHSGGC / MRC-University of Glasgow Centre for Virus Research | COVID-19 Genomics UK (COG-UK) Consortium |
| EPI_ISL_425700 | West of Scotland Specialist Virology Centre, NHSGGC / MRC-University of Glasgow Centre for Virus Research | COVID-19 Genomics UK (COG-UK) Consortium |
| EPI_ISL_425701 | West of Scotland Specialist Virology Centre, NHSGGC / MRC-University of Glasgow Centre for Virus Research | COVID-19 Genomics UK (COG-UK) Consortium |
| EPI_ISL_425702 | West of Scotland Specialist Virology Centre, NHSGGC / MRC-University of Glasgow Centre for Virus Research | COVID-19 Genomics UK (COG-UK) Consortium |
| EPI_ISL_425703 | West of Scotland Specialist Virology Centre, NHSGGC / MRC-University of Glasgow Centre for Virus Research | COVID-19 Genomics UK (COG-UK) Consortium |
| EPI_ISL_425704 | West of Scotland Specialist Virology Centre, NHSGGC / MRC-University of Glasgow Centre for Virus Research | COVID-19 Genomics UK (COG-UK) Consortium |
| EPI_ISL_425705 | West of Scotland Specialist Virology Centre, NHSGGC / MRC-University of Glasgow Centre for Virus Research | COVID-19 Genomics UK (COG-UK) Consortium |
| EPI_ISL_425706 | West of Scotland Specialist Virology Centre, NHSGGC / MRC-University of Glasgow Centre for Virus Research | COVID-19 Genomics UK (COG-UK) Consortium |
| EPI_ISL_425707 | West of Scotland Specialist Virology Centre, NHSGGC / MRC-University of Glasgow Centre for Virus Research | COVID-19 Genomics UK (COG-UK) Consortium |

| EPI_Accession_ID | Originating Laboratory | Submitting Laboratory |
|---|---|---|
| EPI_ISL_425708 | West of Scotland Specialist Virology Centre, NHSGGC / MRC-University of Glasgow Centre for Virus Research | COVID-19 Genomics UK (COG-UK) Consortium |
| EPI_ISL_425709 | West of Scotland Specialist Virology Centre, NHSGGC / MRC-University of Glasgow Centre for Virus Research | COVID-19 Genomics UK (COG-UK) Consortium |
| EPI_ISL_425710 | West of Scotland Specialist Virology Centre, NHSGGC / MRC-University of Glasgow Centre for Virus Research | COVID-19 Genomics UK (COG-UK) Consortium |
| EPI_ISL_425711 | West of Scotland Specialist Virology Centre, NHSGGC / MRC-University of Glasgow Centre for Virus Research | COVID-19 Genomics UK (COG-UK) Consortium |
| EPI_ISL_425712 | West of Scotland Specialist Virology Centre, NHSGGC / MRC-University of Glasgow Centre for Virus Research | COVID-19 Genomics UK (COG-UK) Consortium |
| EPI_ISL_425713 | West of Scotland Specialist Virology Centre, NHSGGC / MRC-University of Glasgow Centre for Virus Research | COVID-19 Genomics UK (COG-UK) Consortium |
| EPI_ISL_425714 | West of Scotland Specialist Virology Centre, NHSGGC / MRC-University of Glasgow Centre for Virus Research | COVID-19 Genomics UK (COG-UK) Consortium |
| EPI_ISL_425715 | West of Scotland Specialist Virology Centre, NHSGGC / MRC-University of Glasgow Centre for Virus Research | COVID-19 Genomics UK (COG-UK) Consortium |
| EPI_ISL_425716 | West of Scotland Specialist Virology Centre, NHSGGC / MRC-University of Glasgow Centre for Virus Research | COVID-19 Genomics UK (COG-UK) Consortium |
| EPI_ISL_425717 | West of Scotland Specialist Virology Centre, NHSGGC / MRC-University of Glasgow Centre for Virus Research | COVID-19 Genomics UK (COG-UK) Consortium |
| EPI_ISL_425718 | West of Scotland Specialist Virology Centre, NHSGGC / MRC-University of Glasgow Centre for Virus Research | COVID-19 Genomics UK (COG-UK) Consortium |
| EPI_ISL_425719 | West of Scotland Specialist Virology Centre, NHSGGC / MRC-University of Glasgow Centre for Virus Research | COVID-19 Genomics UK (COG-UK) Consortium |
| EPI_ISL_425720 | West of Scotland Specialist Virology Centre, NHSGGC / MRC-University of Glasgow Centre for Virus Research | COVID-19 Genomics UK (COG-UK) Consortium |
| EPI_ISL_425721 | West of Scotland Specialist Virology Centre, NHSGGC / MRC-University of Glasgow Centre for Virus Research | COVID-19 Genomics UK (COG-UK) Consortium |
| EPI_ISL_425722 | West of Scotland Specialist Virology Centre, NHSGGC / MRC-University of Glasgow Centre for Virus Research | COVID-19 Genomics UK (COG-UK) Consortium |
| EPI_ISL_425723 | West of Scotland Specialist Virology Centre, NHSGGC / MRC-University of Glasgow Centre for Virus Research | COVID-19 Genomics UK (COG-UK) Consortium |

| EPI_Accession_ID | Originating Laboratory | Submitting Laboratory |
|---|---|---|
| EPI_ISL_425724 | West of Scotland Specialist Virology Centre, NHSGGC / MRC-University of Glasgow Centre for Virus Research | COVID-19 Genomics UK (COG-UK) Consortium |
| EPI_ISL_425725 | West of Scotland Specialist Virology Centre, NHSGGC / MRC-University of Glasgow Centre for Virus Research | COVID-19 Genomics UK (COG-UK) Consortium |
| EPI_ISL_425726 | West of Scotland Specialist Virology Centre, NHSGGC / MRC-University of Glasgow Centre for Virus Research | COVID-19 Genomics UK (COG-UK) Consortium |
| EPI_ISL_425727 | West of Scotland Specialist Virology Centre, NHSGGC / MRC-University of Glasgow Centre for Virus Research | COVID-19 Genomics UK (COG-UK) Consortium |
| EPI_ISL_425728 | West of Scotland Specialist Virology Centre, NHSGGC / MRC-University of Glasgow Centre for Virus Research | COVID-19 Genomics UK (COG-UK) Consortium |
| EPI_ISL_425729 | West of Scotland Specialist Virology Centre, NHSGGC / MRC-University of Glasgow Centre for Virus Research | COVID-19 Genomics UK (COG-UK) Consortium |
| EPI_ISL_425730 | West of Scotland Specialist Virology Centre, NHSGGC / MRC-University of Glasgow Centre for Virus Research | COVID-19 Genomics UK (COG-UK) Consortium |
| EPI_ISL_425731 | West of Scotland Specialist Virology Centre, NHSGGC / MRC-University of Glasgow Centre for Virus Research | COVID-19 Genomics UK (COG-UK) Consortium |
| EPI_ISL_425732 | West of Scotland Specialist Virology Centre, NHSGGC / MRC-University of Glasgow Centre for Virus Research | COVID-19 Genomics UK (COG-UK) Consortium |
| EPI_ISL_425733 | West of Scotland Specialist Virology Centre, NHSGGC / MRC-University of Glasgow Centre for Virus Research | COVID-19 Genomics UK (COG-UK) Consortium |
| EPI_ISL_425734 | West of Scotland Specialist Virology Centre, NHSGGC / MRC-University of Glasgow Centre for Virus Research | COVID-19 Genomics UK (COG-UK) Consortium |
| EPI_ISL_425735 | West of Scotland Specialist Virology Centre, NHSGGC / MRC-University of Glasgow Centre for Virus Research | COVID-19 Genomics UK (COG-UK) Consortium |
| EPI_ISL_425736 | West of Scotland Specialist Virology Centre, NHSGGC / MRC-University of Glasgow Centre for Virus Research | COVID-19 Genomics UK (COG-UK) Consortium |
| EPI_ISL_425737 | West of Scotland Specialist Virology Centre, NHSGGC / MRC-University of Glasgow Centre for Virus Research | COVID-19 Genomics UK (COG-UK) Consortium |
| EPI_ISL_425738 | West of Scotland Specialist Virology Centre, NHSGGC / MRC-University of Glasgow Centre for Virus Research | COVID-19 Genomics UK (COG-UK) Consortium |
| EPI_ISL_425739 | West of Scotland Specialist Virology Centre, NHSGGC / MRC-University of Glasgow Centre for Virus Research | COVID-19 Genomics UK (COG-UK) Consortium |

| EPI_Accession_ID | Originating Laboratory | Submitting Laboratory |
|---|---|---|
| EPI_ISL_425740 | West of Scotland Specialist Virology Centre, NHSGGC / MRC-University of Glasgow Centre for Virus Research | COVID-19 Genomics UK (COG-UK) Consortium |
| EPI_ISL_425741 | West of Scotland Specialist Virology Centre, NHSGGC / MRC-University of Glasgow Centre for Virus Research | COVID-19 Genomics UK (COG-UK) Consortium |
| EPI_ISL_425742 | West of Scotland Specialist Virology Centre, NHSGGC / MRC-University of Glasgow Centre for Virus Research | COVID-19 Genomics UK (COG-UK) Consortium |
| EPI_ISL_425743 | West of Scotland Specialist Virology Centre, NHSGGC / MRC-University of Glasgow Centre for Virus Research | COVID-19 Genomics UK (COG-UK) Consortium |
| EPI_ISL_425744 | West of Scotland Specialist Virology Centre, NHSGGC / MRC-University of Glasgow Centre for Virus Research | COVID-19 Genomics UK (COG-UK) Consortium |
| EPI_ISL_425745 | West of Scotland Specialist Virology Centre, NHSGGC / MRC-University of Glasgow Centre for Virus Research | COVID-19 Genomics UK (COG-UK) Consortium |
| EPI_ISL_425746 | West of Scotland Specialist Virology Centre, NHSGGC / MRC-University of Glasgow Centre for Virus Research | COVID-19 Genomics UK (COG-UK) Consortium |
| EPI_ISL_425747 | West of Scotland Specialist Virology Centre, NHSGGC / MRC-University of Glasgow Centre for Virus Research | COVID-19 Genomics UK (COG-UK) Consortium |
| EPI_ISL_425748 | West of Scotland Specialist Virology Centre, NHSGGC / MRC-University of Glasgow Centre for Virus Research | COVID-19 Genomics UK (COG-UK) Consortium |
| EPI_ISL_425749 | West of Scotland Specialist Virology Centre, NHSGGC / MRC-University of Glasgow Centre for Virus Research | COVID-19 Genomics UK (COG-UK) Consortium |
| EPI_ISL_425750 | West of Scotland Specialist Virology Centre, NHSGGC / MRC-University of Glasgow Centre for Virus Research | COVID-19 Genomics UK (COG-UK) Consortium |
| EPI_ISL_425751 | West of Scotland Specialist Virology Centre, NHSGGC / MRC-University of Glasgow Centre for Virus Research | COVID-19 Genomics UK (COG-UK) Consortium |
| EPI_ISL_425752 | West of Scotland Specialist Virology Centre, NHSGGC / MRC-University of Glasgow Centre for Virus Research | COVID-19 Genomics UK (COG-UK) Consortium |
| EPI_ISL_425753 | West of Scotland Specialist Virology Centre, NHSGGC / MRC-University of Glasgow Centre for Virus Research | COVID-19 Genomics UK (COG-UK) Consortium |
| EPI_ISL_425754 | West of Scotland Specialist Virology Centre, NHSGGC / MRC-University of Glasgow Centre for Virus Research | COVID-19 Genomics UK (COG-UK) Consortium |
| EPI_ISL_425755 | West of Scotland Specialist Virology Centre, NHSGGC / MRC-University of Glasgow Centre for Virus Research | COVID-19 Genomics UK (COG-UK) Consortium |

| EPI_Accession_ID | Originating Laboratory | Submitting Laboratory |
|---|---|---|
| EPI_ISL_425756 | West of Scotland Specialist Virology Centre, NHSGGC / MRC-University of Glasgow Centre for Virus Research | COVID-19 Genomics UK (COG-UK) Consortium |
| EPI_ISL_425757 | West of Scotland Specialist Virology Centre, NHSGGC / MRC-University of Glasgow Centre for Virus Research | COVID-19 Genomics UK (COG-UK) Consortium |
| EPI_ISL_425758 | West of Scotland Specialist Virology Centre, NHSGGC / MRC-University of Glasgow Centre for Virus Research | COVID-19 Genomics UK (COG-UK) Consortium |
| EPI_ISL_425759 | West of Scotland Specialist Virology Centre, NHSGGC / MRC-University of Glasgow Centre for Virus Research | COVID-19 Genomics UK (COG-UK) Consortium |
| EPI_ISL_425760 | West of Scotland Specialist Virology Centre, NHSGGC / MRC-University of Glasgow Centre for Virus Research | COVID-19 Genomics UK (COG-UK) Consortium |
| EPI_ISL_425761 | West of Scotland Specialist Virology Centre, NHSGGC / MRC-University of Glasgow Centre for Virus Research | COVID-19 Genomics UK (COG-UK) Consortium |
| EPI_ISL_425762 | West of Scotland Specialist Virology Centre, NHSGGC / MRC-University of Glasgow Centre for Virus Research | COVID-19 Genomics UK (COG-UK) Consortium |
| EPI_ISL_425763 | West of Scotland Specialist Virology Centre, NHSGGC / MRC-University of Glasgow Centre for Virus Research | COVID-19 Genomics UK (COG-UK) Consortium |
| EPI_ISL_425764 | West of Scotland Specialist Virology Centre, NHSGGC / MRC-University of Glasgow Centre for Virus Research | COVID-19 Genomics UK (COG-UK) Consortium |
| EPI_ISL_425765 | West of Scotland Specialist Virology Centre, NHSGGC / MRC-University of Glasgow Centre for Virus Research | COVID-19 Genomics UK (COG-UK) Consortium |
| EPI_ISL_425766 | West of Scotland Specialist Virology Centre, NHSGGC / MRC-University of Glasgow Centre for Virus Research | COVID-19 Genomics UK (COG-UK) Consortium |
| EPI_ISL_425767 | West of Scotland Specialist Virology Centre, NHSGGC / MRC-University of Glasgow Centre for Virus Research | COVID-19 Genomics UK (COG-UK) Consortium |
| EPI_ISL_425768 | West of Scotland Specialist Virology Centre, NHSGGC / MRC-University of Glasgow Centre for Virus Research | COVID-19 Genomics UK (COG-UK) Consortium |
| EPI_ISL_425769 | West of Scotland Specialist Virology Centre, NHSGGC / MRC-University of Glasgow Centre for Virus Research | COVID-19 Genomics UK (COG-UK) Consortium |
| EPI_ISL_425770 | West of Scotland Specialist Virology Centre, NHSGGC / MRC-University of Glasgow Centre for Virus Research | COVID-19 Genomics UK (COG-UK) Consortium |
| EPI_ISL_425771 | West of Scotland Specialist Virology Centre, NHSGGC / MRC-University of Glasgow Centre for Virus Research | COVID-19 Genomics UK (COG-UK) Consortium |

| EPI_Accession_ID | Originating Laboratory | Submitting Laboratory |
|---|---|---|
| EPI_ISL_425772 | West of Scotland Specialist Virology Centre, NHSGGC / MRC-University of Glasgow Centre for Virus Research | COVID-19 Genomics UK (COG-UK) Consortium |
| EPI_ISL_425773 | West of Scotland Specialist Virology Centre, NHSGGC / MRC-University of Glasgow Centre for Virus Research | COVID-19 Genomics UK (COG-UK) Consortium |
| EPI_ISL_425774 | West of Scotland Specialist Virology Centre, NHSGGC / MRC-University of Glasgow Centre for Virus Research | COVID-19 Genomics UK (COG-UK) Consortium |
| EPI_ISL_425775 | West of Scotland Specialist Virology Centre, NHSGGC / MRC-University of Glasgow Centre for Virus Research | COVID-19 Genomics UK (COG-UK) Consortium |
| EPI_ISL_425776 | West of Scotland Specialist Virology Centre, NHSGGC / MRC-University of Glasgow Centre for Virus Research | COVID-19 Genomics UK (COG-UK) Consortium |
| EPI_ISL_425777 | West of Scotland Specialist Virology Centre, NHSGGC / MRC-University of Glasgow Centre for Virus Research | COVID-19 Genomics UK (COG-UK) Consortium |
| EPI_ISL_425778 | West of Scotland Specialist Virology Centre, NHSGGC / MRC-University of Glasgow Centre for Virus Research | COVID-19 Genomics UK (COG-UK) Consortium |
| EPI_ISL_425779 | West of Scotland Specialist Virology Centre, NHSGGC / MRC-University of Glasgow Centre for Virus Research | COVID-19 Genomics UK (COG-UK) Consortium |
| EPI_ISL_425780 | West of Scotland Specialist Virology Centre, NHSGGC / MRC-University of Glasgow Centre for Virus Research | COVID-19 Genomics UK (COG-UK) Consortium |
| EPI_ISL_425781 | West of Scotland Specialist Virology Centre, NHSGGC / MRC-University of Glasgow Centre for Virus Research | COVID-19 Genomics UK (COG-UK) Consortium |
| EPI_ISL_425782 | West of Scotland Specialist Virology Centre, NHSGGC / MRC-University of Glasgow Centre for Virus Research | COVID-19 Genomics UK (COG-UK) Consortium |
| EPI_ISL_425783 | West of Scotland Specialist Virology Centre, NHSGGC / MRC-University of Glasgow Centre for Virus Research | COVID-19 Genomics UK (COG-UK) Consortium |
| EPI_ISL_425784 | West of Scotland Specialist Virology Centre, NHSGGC / MRC-University of Glasgow Centre for Virus Research | COVID-19 Genomics UK (COG-UK) Consortium |
| EPI_ISL_425785 | West of Scotland Specialist Virology Centre, NHSGGC / MRC-University of Glasgow Centre for Virus Research | COVID-19 Genomics UK (COG-UK) Consortium |
| EPI_ISL_425786 | West of Scotland Specialist Virology Centre, NHSGGC / MRC-University of Glasgow Centre for Virus Research | COVID-19 Genomics UK (COG-UK) Consortium |
| EPI_ISL_425787 | West of Scotland Specialist Virology Centre, NHSGGC / MRC-University of Glasgow Centre for Virus Research | COVID-19 Genomics UK (COG-UK) Consortium |

| EPI_Accession_ID | Originating Laboratory | Submitting Laboratory |
|---|---|---|
| EPI_ISL_425788 | West of Scotland Specialist Virology Centre, NHSGGC / MRC-University of Glasgow Centre for Virus Research | COVID-19 Genomics UK (COG-UK) Consortium |
| EPI_ISL_425789 | West of Scotland Specialist Virology Centre, NHSGGC / MRC-University of Glasgow Centre for Virus Research | COVID-19 Genomics UK (COG-UK) Consortium |
| EPI_ISL_425790 | West of Scotland Specialist Virology Centre, NHSGGC / MRC-University of Glasgow Centre for Virus Research | COVID-19 Genomics UK (COG-UK) Consortium |
| EPI_ISL_425791 | West of Scotland Specialist Virology Centre, NHSGGC / MRC-University of Glasgow Centre for Virus Research | COVID-19 Genomics UK (COG-UK) Consortium |
| EPI_ISL_425792 | West of Scotland Specialist Virology Centre, NHSGGC / MRC-University of Glasgow Centre for Virus Research | COVID-19 Genomics UK (COG-UK) Consortium |
| EPI_ISL_425793 | West of Scotland Specialist Virology Centre, NHSGGC / MRC-University of Glasgow Centre for Virus Research | COVID-19 Genomics UK (COG-UK) Consortium |
| EPI_ISL_425794 | West of Scotland Specialist Virology Centre, NHSGGC / MRC-University of Glasgow Centre for Virus Research | COVID-19 Genomics UK (COG-UK) Consortium |
| EPI_ISL_425795 | West of Scotland Specialist Virology Centre, NHSGGC / MRC-University of Glasgow Centre for Virus Research | COVID-19 Genomics UK (COG-UK) Consortium |
| EPI_ISL_425796 | West of Scotland Specialist Virology Centre, NHSGGC / MRC-University of Glasgow Centre for Virus Research | COVID-19 Genomics UK (COG-UK) Consortium |
| EPI_ISL_425797 | West of Scotland Specialist Virology Centre, NHSGGC / MRC-University of Glasgow Centre for Virus Research | COVID-19 Genomics UK (COG-UK) Consortium |
| EPI_ISL_425798 | West of Scotland Specialist Virology Centre, NHSGGC / MRC-University of Glasgow Centre for Virus Research | COVID-19 Genomics UK (COG-UK) Consortium |
| EPI_ISL_425799 | West of Scotland Specialist Virology Centre, NHSGGC / MRC-University of Glasgow Centre for Virus Research | COVID-19 Genomics UK (COG-UK) Consortium |
| EPI_ISL_425800 | West of Scotland Specialist Virology Centre, NHSGGC / MRC-University of Glasgow Centre for Virus Research | COVID-19 Genomics UK (COG-UK) Consortium |
| EPI_ISL_425801 | West of Scotland Specialist Virology Centre, NHSGGC / MRC-University of Glasgow Centre for Virus Research | COVID-19 Genomics UK (COG-UK) Consortium |
| EPI_ISL_425802 | West of Scotland Specialist Virology Centre, NHSGGC / MRC-University of Glasgow Centre for Virus Research | COVID-19 Genomics UK (COG-UK) Consortium |
| EPI_ISL_425803 | West of Scotland Specialist Virology Centre, NHSGGC / MRC-University of Glasgow Centre for Virus Research | COVID-19 Genomics UK (COG-UK) Consortium |

| EPI_Accession_ID | Originating Laboratory | Submitting Laboratory |
|---|---|---|
| EPI_ISL_425804 | West of Scotland Specialist Virology Centre, NHSGGC / MRC-University of Glasgow Centre for Virus Research | COVID-19 Genomics UK (COG-UK) Consortium |
| EPI_ISL_425805 | West of Scotland Specialist Virology Centre, NHSGGC / MRC-University of Glasgow Centre for Virus Research | COVID-19 Genomics UK (COG-UK) Consortium |
| EPI_ISL_425806 | West of Scotland Specialist Virology Centre, NHSGGC / MRC-University of Glasgow Centre for Virus Research | COVID-19 Genomics UK (COG-UK) Consortium |
| EPI_ISL_425807 | West of Scotland Specialist Virology Centre, NHSGGC / MRC-University of Glasgow Centre for Virus Research | COVID-19 Genomics UK (COG-UK) Consortium |
| EPI_ISL_425808 | West of Scotland Specialist Virology Centre, NHSGGC / MRC-University of Glasgow Centre for Virus Research | COVID-19 Genomics UK (COG-UK) Consortium |
| EPI_ISL_425809 | West of Scotland Specialist Virology Centre, NHSGGC / MRC-University of Glasgow Centre for Virus Research | COVID-19 Genomics UK (COG-UK) Consortium |
| EPI_ISL_425810 | West of Scotland Specialist Virology Centre, NHSGGC / MRC-University of Glasgow Centre for Virus Research | COVID-19 Genomics UK (COG-UK) Consortium |
| EPI_ISL_425811 | West of Scotland Specialist Virology Centre, NHSGGC / MRC-University of Glasgow Centre for Virus Research | COVID-19 Genomics UK (COG-UK) Consortium |
| EPI_ISL_425812 | West of Scotland Specialist Virology Centre, NHSGGC / MRC-University of Glasgow Centre for Virus Research | COVID-19 Genomics UK (COG-UK) Consortium |
| EPI_ISL_425813 | West of Scotland Specialist Virology Centre, NHSGGC / MRC-University of Glasgow Centre for Virus Research | COVID-19 Genomics UK (COG-UK) Consortium |
| EPI_ISL_425814 | West of Scotland Specialist Virology Centre, NHSGGC / MRC-University of Glasgow Centre for Virus Research | COVID-19 Genomics UK (COG-UK) Consortium |
| EPI_ISL_425815 | West of Scotland Specialist Virology Centre, NHSGGC / MRC-University of Glasgow Centre for Virus Research | COVID-19 Genomics UK (COG-UK) Consortium |
| EPI_ISL_425816 | West of Scotland Specialist Virology Centre, NHSGGC / MRC-University of Glasgow Centre for Virus Research | COVID-19 Genomics UK (COG-UK) Consortium |
| EPI_ISL_425817 | West of Scotland Specialist Virology Centre, NHSGGC / MRC-University of Glasgow Centre for Virus Research | COVID-19 Genomics UK (COG-UK) Consortium |
| EPI_ISL_425818 | West of Scotland Specialist Virology Centre, NHSGGC / MRC-University of Glasgow Centre for Virus Research | COVID-19 Genomics UK (COG-UK) Consortium |

| EPI_Accession_ID | Originating Laboratory | Submitting Laboratory |
|---|---|---|
| EPI_ISL_425819 | Virology Department, Royal Infirmary of Edinburgh, NHS Lothian / School of Biological Sciences, University of Edinburgh / Institute of Genetics and Molecular Medicine, University of Edinburgh | COVID-19 Genomics UK (COG-UK) Consortium |
| EPI_ISL_425820 | Virology Department, Royal Infirmary of Edinburgh, NHS Lothian / School of Biological Sciences, University of Edinburgh / Institute of Genetics and Molecular Medicine, University of Edinburgh | COVID-19 Genomics UK (COG-UK) Consortium |
| EPI_ISL_425821 | Virology Department, Royal Infirmary of Edinburgh, NHS Lothian / School of Biological Sciences, University of Edinburgh / Institute of Genetics and Molecular Medicine, University of Edinburgh | COVID-19 Genomics UK (COG-UK) Consortium |
| EPI_ISL_425822 | Virology Department, Royal Infirmary of Edinburgh, NHS Lothian / School of Biological Sciences, University of Edinburgh / Institute of Genetics and Molecular Medicine, University of Edinburgh | COVID-19 Genomics UK (COG-UK) Consortium |
| EPI_ISL_425823 | Virology Department, Royal Infirmary of Edinburgh, NHS Lothian / School of Biological Sciences, University of Edinburgh / Institute of Genetics and Molecular Medicine, University of Edinburgh | COVID-19 Genomics UK (COG-UK) Consortium |
| EPI_ISL_425824 | Virology Department, Royal Infirmary of Edinburgh, NHS Lothian / School of Biological Sciences, University of Edinburgh / Institute of Genetics and Molecular Medicine, University of Edinburgh | COVID-19 Genomics UK (COG-UK) Consortium |
| EPI_ISL_425825 | Virology Department, Royal Infirmary of Edinburgh, NHS Lothian / School of Biological Sciences, University of Edinburgh / Institute of Genetics and Molecular Medicine, University of Edinburgh | COVID-19 Genomics UK (COG-UK) Consortium |
| EPI_ISL_425826 | Virology Department, Royal Infirmary of Edinburgh, NHS Lothian / School of Biological Sciences, University of Edinburgh / Institute of Genetics and Molecular Medicine, University of Edinburgh | COVID-19 Genomics UK (COG-UK) Consortium |

| EPI_Accession_ID | Originating Laboratory | Submitting Laboratory |
|---|---|---|
| EPI_ISL_425827 | Virology Department, Royal Infirmary of Edinburgh, NHS Lothian / School of Biological Sciences, University of Edinburgh / Institute of Genetics and Molecular Medicine, University of Edinburgh | COVID-19 Genomics UK (COG-UK) Consortium |
| EPI_ISL_425828 | Virology Department, Royal Infirmary of Edinburgh, NHS Lothian / School of Biological Sciences, University of Edinburgh / Institute of Genetics and Molecular Medicine, University of Edinburgh | COVID-19 Genomics UK (COG-UK) Consortium |
| EPI_ISL_425829 | Virology Department, Royal Infirmary of Edinburgh, NHS Lothian / School of Biological Sciences, University of Edinburgh / Institute of Genetics and Molecular Medicine, University of Edinburgh | COVID-19 Genomics UK (COG-UK) Consortium |
| EPI_ISL_425830 | Virology Department, Royal Infirmary of Edinburgh, NHS Lothian / School of Biological Sciences, University of Edinburgh / Institute of Genetics and Molecular Medicine, University of Edinburgh | COVID-19 Genomics UK (COG-UK) Consortium |
| EPI_ISL_425831 | Virology Department, Royal Infirmary of Edinburgh, NHS Lothian / School of Biological Sciences, University of Edinburgh / Institute of Genetics and Molecular Medicine, University of Edinburgh | COVID-19 Genomics UK (COG-UK) Consortium |
| EPI_ISL_425832 | Virology Department, Royal Infirmary of Edinburgh, NHS Lothian / School of Biological Sciences, University of Edinburgh / Institute of Genetics and Molecular Medicine, University of Edinburgh | COVID-19 Genomics UK (COG-UK) Consortium |
| EPI_ISL_425833 | Virology Department, Royal Infirmary of Edinburgh, NHS Lothian / School of Biological Sciences, University of Edinburgh / Institute of Genetics and Molecular Medicine, University of Edinburgh | COVID-19 Genomics UK (COG-UK) Consortium |
| EPI_ISL_425834 | Virology Department, Royal Infirmary of Edinburgh, NHS Lothian / School of Biological Sciences, University of Edinburgh / Institute of Genetics and Molecular Medicine, University of Edinburgh | COVID-19 Genomics UK (COG-UK) Consortium |

| EPI_Accession_ID | Originating Laboratory | Submitting Laboratory |
|---|---|---|
| EPI_ISL_425835 | Virology Department, Royal Infirmary of Edinburgh, NHS Lothian / School of Biological Sciences, University of Edinburgh / Institute of Genetics and Molecular Medicine, University of Edinburgh | COVID-19 Genomics UK (COG-UK) Consortium |
| EPI_ISL_425836 | Virology Department, Royal Infirmary of Edinburgh, NHS Lothian / School of Biological Sciences, University of Edinburgh / Institute of Genetics and Molecular Medicine, University of Edinburgh | COVID-19 Genomics UK (COG-UK) Consortium |
| EPI_ISL_425837 | Virology Department, Royal Infirmary of Edinburgh, NHS Lothian / School of Biological Sciences, University of Edinburgh / Institute of Genetics and Molecular Medicine, University of Edinburgh | COVID-19 Genomics UK (COG-UK) Consortium |
| EPI_ISL_425838 | Virology Department, Royal Infirmary of Edinburgh, NHS Lothian / School of Biological Sciences, University of Edinburgh / Institute of Genetics and Molecular Medicine, University of Edinburgh | COVID-19 Genomics UK (COG-UK) Consortium |
| EPI_ISL_425839 | Virology Department, Royal Infirmary of Edinburgh, NHS Lothian / School of Biological Sciences, University of Edinburgh / Institute of Genetics and Molecular Medicine, University of Edinburgh | COVID-19 Genomics UK (COG-UK) Consortium |
| EPI_ISL_425840 | Virology Department, Royal Infirmary of Edinburgh, NHS Lothian / School of Biological Sciences, University of Edinburgh / Institute of Genetics and Molecular Medicine, University of Edinburgh | COVID-19 Genomics UK (COG-UK) Consortium |
| EPI_ISL_425841 | Virology Department, Royal Infirmary of Edinburgh, NHS Lothian / School of Biological Sciences, University of Edinburgh / Institute of Genetics and Molecular Medicine, University of Edinburgh | COVID-19 Genomics UK (COG-UK) Consortium |
| EPI_ISL_425842 | Virology Department, Royal Infirmary of Edinburgh, NHS Lothian / School of Biological Sciences, University of Edinburgh / Institute of Genetics and Molecular Medicine, University of Edinburgh | COVID-19 Genomics UK (COG-UK) Consortium |

| EPI_Accession_ID | Originating Laboratory | Submitting Laboratory |
|---|---|---|
| EPI_ISL_425843 | Virology Department, Royal Infirmary of Edinburgh, NHS Lothian / School of Biological Sciences, University of Edinburgh / Institute of Genetics and Molecular Medicine, University of Edinburgh | COVID-19 Genomics UK (COG-UK) Consortium |
| EPI_ISL_425844 | Virology Department, Royal Infirmary of Edinburgh, NHS Lothian / School of Biological Sciences, University of Edinburgh / Institute of Genetics and Molecular Medicine, University of Edinburgh | COVID-19 Genomics UK (COG-UK) Consortium |
| EPI_ISL_425845 | Virology Department, Royal Infirmary of Edinburgh, NHS Lothian / School of Biological Sciences, University of Edinburgh / Institute of Genetics and Molecular Medicine, University of Edinburgh | COVID-19 Genomics UK (COG-UK) Consortium |
| EPI_ISL_425846 | Virology Department, Royal Infirmary of Edinburgh, NHS Lothian / School of Biological Sciences, University of Edinburgh / Institute of Genetics and Molecular Medicine, University of Edinburgh | COVID-19 Genomics UK (COG-UK) Consortium |
| EPI_ISL_425847 | Virology Department, Royal Infirmary of Edinburgh, NHS Lothian / School of Biological Sciences, University of Edinburgh / Institute of Genetics and Molecular Medicine, University of Edinburgh | COVID-19 Genomics UK (COG-UK) Consortium |
| EPI_ISL_425848 | Virology Department, Royal Infirmary of Edinburgh, NHS Lothian / School of Biological Sciences, University of Edinburgh / Institute of Genetics and Molecular Medicine, University of Edinburgh | COVID-19 Genomics UK (COG-UK) Consortium |
| EPI_ISL_425849 | Virology Department, Royal Infirmary of Edinburgh, NHS Lothian / School of Biological Sciences, University of Edinburgh / Institute of Genetics and Molecular Medicine, University of Edinburgh | COVID-19 Genomics UK (COG-UK) Consortium |
| EPI_ISL_425850 | Virology Department, Royal Infirmary of Edinburgh, NHS Lothian / School of Biological Sciences, University of Edinburgh / Institute of Genetics and Molecular Medicine, University of Edinburgh | COVID-19 Genomics UK (COG-UK) Consortium |

| EPI_Accession_ID | Originating Laboratory | Submitting Laboratory |
|---|---|---|
| EPI_ISL_425851 | Virology Department, Royal Infirmary of Edinburgh, NHS Lothian / School of Biological Sciences, University of Edinburgh / Institute of Genetics and Molecular Medicine, University of Edinburgh | COVID-19 Genomics UK (COG-UK) Consortium |
| EPI_ISL_425852 | Virology Department, Royal Infirmary of Edinburgh, NHS Lothian / School of Biological Sciences, University of Edinburgh / Institute of Genetics and Molecular Medicine, University of Edinburgh | COVID-19 Genomics UK (COG-UK) Consortium |
| EPI_ISL_425853 | Virology Department, Royal Infirmary of Edinburgh, NHS Lothian / School of Biological Sciences, University of Edinburgh / Institute of Genetics and Molecular Medicine, University of Edinburgh | COVID-19 Genomics UK (COG-UK) Consortium |
| EPI_ISL_425854 | Virology Department, Royal Infirmary of Edinburgh, NHS Lothian / School of Biological Sciences, University of Edinburgh / Institute of Genetics and Molecular Medicine, University of Edinburgh | COVID-19 Genomics UK (COG-UK) Consortium |
| EPI_ISL_425855 | Virology Department, Royal Infirmary of Edinburgh, NHS Lothian / School of Biological Sciences, University of Edinburgh / Institute of Genetics and Molecular Medicine, University of Edinburgh | COVID-19 Genomics UK (COG-UK) Consortium |
| EPI_ISL_425856 | Virology Department, Royal Infirmary of Edinburgh, NHS Lothian / School of Biological Sciences, University of Edinburgh / Institute of Genetics and Molecular Medicine, University of Edinburgh | COVID-19 Genomics UK (COG-UK) Consortium |
| EPI_ISL_425857 | Virology Department, Royal Infirmary of Edinburgh, NHS Lothian / School of Biological Sciences, University of Edinburgh / Institute of Genetics and Molecular Medicine, University of Edinburgh | COVID-19 Genomics UK (COG-UK) Consortium |
| EPI_ISL_425858 | Virology Department, Royal Infirmary of Edinburgh, NHS Lothian / School of Biological Sciences, University of Edinburgh / Institute of Genetics and Molecular Medicine, University of Edinburgh | COVID-19 Genomics UK (COG-UK) Consortium |

| EPI_Accession_ID | Originating Laboratory | Submitting Laboratory |
|---|---|---|
| EPI_ISL_425859 | Virology Department, Royal Infirmary of Edinburgh, NHS Lothian / School of Biological Sciences, University of Edinburgh / Institute of Genetics and Molecular Medicine, University of Edinburgh | COVID-19 Genomics UK (COG-UK) Consortium |
| EPI_ISL_425860 | Virology Department, Royal Infirmary of Edinburgh, NHS Lothian / School of Biological Sciences, University of Edinburgh / Institute of Genetics and Molecular Medicine, University of Edinburgh | COVID-19 Genomics UK (COG-UK) Consortium |
| EPI_ISL_425861 | Virology Department, Royal Infirmary of Edinburgh, NHS Lothian / School of Biological Sciences, University of Edinburgh / Institute of Genetics and Molecular Medicine, University of Edinburgh | COVID-19 Genomics UK (COG-UK) Consortium |
| EPI_ISL_425862 | Virology Department, Royal Infirmary of Edinburgh, NHS Lothian / School of Biological Sciences, University of Edinburgh / Institute of Genetics and Molecular Medicine, University of Edinburgh | COVID-19 Genomics UK (COG-UK) Consortium |
| EPI_ISL_425863 | Virology Department, Royal Infirmary of Edinburgh, NHS Lothian / School of Biological Sciences, University of Edinburgh / Institute of Genetics and Molecular Medicine, University of Edinburgh | COVID-19 Genomics UK (COG-UK) Consortium |
| EPI_ISL_425864 | Virology Department, Royal Infirmary of Edinburgh, NHS Lothian / School of Biological Sciences, University of Edinburgh / Institute of Genetics and Molecular Medicine, University of Edinburgh | COVID-19 Genomics UK (COG-UK) Consortium |
| EPI_ISL_425865 | Virology Department, Royal Infirmary of Edinburgh, NHS Lothian / School of Biological Sciences, University of Edinburgh / Institute of Genetics and Molecular Medicine, University of Edinburgh | COVID-19 Genomics UK (COG-UK) Consortium |
| EPI_ISL_425866 | Virology Department, Royal Infirmary of Edinburgh, NHS Lothian / School of Biological Sciences, University of Edinburgh / Institute of Genetics and Molecular Medicine, University of Edinburgh | COVID-19 Genomics UK (COG-UK) Consortium |

| EPI_Accession_ID | Originating Laboratory | Submitting Laboratory |
|---|---|---|
| EPI_ISL_425867 | Virology Department, Royal Infirmary of Edinburgh, NHS Lothian / School of Biological Sciences, University of Edinburgh / Institute of Genetics and Molecular Medicine, University of Edinburgh | COVID-19 Genomics UK (COG-UK) Consortium |
| EPI_ISL_425868 | Virology Department, Royal Infirmary of Edinburgh, NHS Lothian / School of Biological Sciences, University of Edinburgh / Institute of Genetics and Molecular Medicine, University of Edinburgh | COVID-19 Genomics UK (COG-UK) Consortium |
| EPI_ISL_425869 | Virology Department, Royal Infirmary of Edinburgh, NHS Lothian / School of Biological Sciences, University of Edinburgh / Institute of Genetics and Molecular Medicine, University of Edinburgh | COVID-19 Genomics UK (COG-UK) Consortium |
| EPI_ISL_425870 | Virology Department, Royal Infirmary of Edinburgh, NHS Lothian / School of Biological Sciences, University of Edinburgh / Institute of Genetics and Molecular Medicine, University of Edinburgh | COVID-19 Genomics UK (COG-UK) Consortium |
| EPI_ISL_425871 | Virology Department, Royal Infirmary of Edinburgh, NHS Lothian / School of Biological Sciences, University of Edinburgh / Institute of Genetics and Molecular Medicine, University of Edinburgh | COVID-19 Genomics UK (COG-UK) Consortium |
| EPI_ISL_425872 | Virology Department, Royal Infirmary of Edinburgh, NHS Lothian / School of Biological Sciences, University of Edinburgh / Institute of Genetics and Molecular Medicine, University of Edinburgh | COVID-19 Genomics UK (COG-UK) Consortium |
| EPI_ISL_425873 | Virology Department, Royal Infirmary of Edinburgh, NHS Lothian / School of Biological Sciences, University of Edinburgh / Institute of Genetics and Molecular Medicine, University of Edinburgh | COVID-19 Genomics UK (COG-UK) Consortium |
| EPI_ISL_425874 | Virology Department, Royal Infirmary of Edinburgh, NHS Lothian / School of Biological Sciences, University of Edinburgh / Institute of Genetics and Molecular Medicine, University of Edinburgh | COVID-19 Genomics UK (COG-UK) Consortium |

| EPI_Accession_ID | Originating Laboratory | Submitting Laboratory |
|---|---|---|
| EPI_ISL_425875 | Virology Department, Royal Infirmary of Edinburgh, NHS Lothian / School of Biological Sciences, University of Edinburgh / Institute of Genetics and Molecular Medicine, University of Edinburgh | COVID-19 Genomics UK (COG-UK) Consortium |
| EPI_ISL_425876 | Virology Department, Royal Infirmary of Edinburgh, NHS Lothian / School of Biological Sciences, University of Edinburgh / Institute of Genetics and Molecular Medicine, University of Edinburgh | COVID-19 Genomics UK (COG-UK) Consortium |
| EPI_ISL_425877 | Virology Department, Royal Infirmary of Edinburgh, NHS Lothian / School of Biological Sciences, University of Edinburgh / Institute of Genetics and Molecular Medicine, University of Edinburgh | COVID-19 Genomics UK (COG-UK) Consortium |
| EPI_ISL_425878 | Virology Department, Royal Infirmary of Edinburgh, NHS Lothian / School of Biological Sciences, University of Edinburgh / Institute of Genetics and Molecular Medicine, University of Edinburgh | COVID-19 Genomics UK (COG-UK) Consortium |
| EPI_ISL_425879 | Virology Department, Royal Infirmary of Edinburgh, NHS Lothian / School of Biological Sciences, University of Edinburgh / Institute of Genetics and Molecular Medicine, University of Edinburgh | COVID-19 Genomics UK (COG-UK) Consortium |
| EPI_ISL_425880 | Virology Department, Royal Infirmary of Edinburgh, NHS Lothian / School of Biological Sciences, University of Edinburgh / Institute of Genetics and Molecular Medicine, University of Edinburgh | COVID-19 Genomics UK (COG-UK) Consortium |
| EPI_ISL_425881 | Virology Department, Royal Infirmary of Edinburgh, NHS Lothian / School of Biological Sciences, University of Edinburgh / Institute of Genetics and Molecular Medicine, University of Edinburgh | COVID-19 Genomics UK (COG-UK) Consortium |
| EPI_ISL_425882 | Virology Department, Royal Infirmary of Edinburgh, NHS Lothian / School of Biological Sciences, University of Edinburgh / Institute of Genetics and Molecular Medicine, University of Edinburgh | COVID-19 Genomics UK (COG-UK) Consortium |

| EPI_Accession_ID | Originating Laboratory | Submitting Laboratory |
| --- | --- | --- |
| EPI_ISL_425883 | Virology Department, Royal Infirmary of Edinburgh, NHS Lothian / School of Biological Sciences, University of Edinburgh / Institute of Genetics and Molecular Medicine, University of Edinburgh | COVID-19 Genomics UK (COG-UK) Consortium |
| EPI_ISL_425884 | Virology Department, Royal Infirmary of Edinburgh, NHS Lothian / School of Biological Sciences, University of Edinburgh / Institute of Genetics and Molecular Medicine, University of Edinburgh | COVID-19 Genomics UK (COG-UK) Consortium |
| EPI_ISL_425885 | Virology Department, Royal Infirmary of Edinburgh, NHS Lothian / School of Biological Sciences, University of Edinburgh / Institute of Genetics and Molecular Medicine, University of Edinburgh | COVID-19 Genomics UK (COG-UK) Consortium |
| EPI_ISL_425886 | Virology Department, Royal Infirmary of Edinburgh, NHS Lothian / School of Biological Sciences, University of Edinburgh / Institute of Genetics and Molecular Medicine, University of Edinburgh | COVID-19 Genomics UK (COG-UK) Consortium |
| EPI_ISL_425887 | Virology Department, Royal Infirmary of Edinburgh, NHS Lothian / School of Biological Sciences, University of Edinburgh / Institute of Genetics and Molecular Medicine, University of Edinburgh | COVID-19 Genomics UK (COG-UK) Consortium |
| EPI_ISL_425888 | Virology Department, Royal Infirmary of Edinburgh, NHS Lothian / School of Biological Sciences, University of Edinburgh / Institute of Genetics and Molecular Medicine, University of Edinburgh | COVID-19 Genomics UK (COG-UK) Consortium |
| EPI_ISL_425889 | Virology Department, Royal Infirmary of Edinburgh, NHS Lothian / School of Biological Sciences, University of Edinburgh / Institute of Genetics and Molecular Medicine, University of Edinburgh | COVID-19 Genomics UK (COG-UK) Consortium |
| EPI_ISL_425890 | Virology Department, Royal Infirmary of Edinburgh, NHS Lothian / School of Biological Sciences, University of Edinburgh / Institute of Genetics and Molecular Medicine, University of Edinburgh | COVID-19 Genomics UK (COG-UK) Consortium |

| EPI_Accession_ID | Originating Laboratory | Submitting Laboratory |
|---|---|---|
| EPI_ISL_425891 | Virology Department, Royal Infirmary of Edinburgh, NHS Lothian / School of Biological Sciences, University of Edinburgh / Institute of Genetics and Molecular Medicine, University of Edinburgh | COVID-19 Genomics UK (COG-UK) Consortium |
| EPI_ISL_425892 | Virology Department, Royal Infirmary of Edinburgh, NHS Lothian / School of Biological Sciences, University of Edinburgh / Institute of Genetics and Molecular Medicine, University of Edinburgh | COVID-19 Genomics UK (COG-UK) Consortium |
| EPI_ISL_425893 | Virology Department, Royal Infirmary of Edinburgh, NHS Lothian / School of Biological Sciences, University of Edinburgh / Institute of Genetics and Molecular Medicine, University of Edinburgh | COVID-19 Genomics UK (COG-UK) Consortium |
| EPI_ISL_425894 | Virology Department, Royal Infirmary of Edinburgh, NHS Lothian / School of Biological Sciences, University of Edinburgh / Institute of Genetics and Molecular Medicine, University of Edinburgh | COVID-19 Genomics UK (COG-UK) Consortium |
| EPI_ISL_425895 | Virology Department, Royal Infirmary of Edinburgh, NHS Lothian / School of Biological Sciences, University of Edinburgh / Institute of Genetics and Molecular Medicine, University of Edinburgh | COVID-19 Genomics UK (COG-UK) Consortium |
| EPI_ISL_425896 | Virology Department, Royal Infirmary of Edinburgh, NHS Lothian / School of Biological Sciences, University of Edinburgh / Institute of Genetics and Molecular Medicine, University of Edinburgh | COVID-19 Genomics UK (COG-UK) Consortium |
| EPI_ISL_425897 | Virology Department, Royal Infirmary of Edinburgh, NHS Lothian / School of Biological Sciences, University of Edinburgh / Institute of Genetics and Molecular Medicine, University of Edinburgh | COVID-19 Genomics UK (COG-UK) Consortium |
| EPI_ISL_425898 | Virology Department, Royal Infirmary of Edinburgh, NHS Lothian / School of Biological Sciences, University of Edinburgh / Institute of Genetics and Molecular Medicine, University of Edinburgh | COVID-19 Genomics UK (COG-UK) Consortium |

| EPI_Accession_ID | Originating Laboratory | Submitting Laboratory |
|---|---|---|
| EPI_ISL_425899 | Virology Department, Royal Infirmary of Edinburgh, NHS Lothian / School of Biological Sciences, University of Edinburgh / Institute of Genetics and Molecular Medicine, University of Edinburgh | COVID-19 Genomics UK (COG-UK) Consortium |
| EPI_ISL_425900 | Virology Department, Royal Infirmary of Edinburgh, NHS Lothian / School of Biological Sciences, University of Edinburgh / Institute of Genetics and Molecular Medicine, University of Edinburgh | COVID-19 Genomics UK (COG-UK) Consortium |
| EPI_ISL_425901 | Virology Department, Royal Infirmary of Edinburgh, NHS Lothian / School of Biological Sciences, University of Edinburgh / Institute of Genetics and Molecular Medicine, University of Edinburgh | COVID-19 Genomics UK (COG-UK) Consortium |
| EPI_ISL_425902 | Virology Department, Royal Infirmary of Edinburgh, NHS Lothian / School of Biological Sciences, University of Edinburgh / Institute of Genetics and Molecular Medicine, University of Edinburgh | COVID-19 Genomics UK (COG-UK) Consortium |
| EPI_ISL_425903 | Virology Department, Royal Infirmary of Edinburgh, NHS Lothian / School of Biological Sciences, University of Edinburgh / Institute of Genetics and Molecular Medicine, University of Edinburgh | COVID-19 Genomics UK (COG-UK) Consortium |
| EPI_ISL_425904 | Virology Department, Royal Infirmary of Edinburgh, NHS Lothian / School of Biological Sciences, University of Edinburgh / Institute of Genetics and Molecular Medicine, University of Edinburgh | COVID-19 Genomics UK (COG-UK) Consortium |
| EPI_ISL_425905 | Virology Department, Royal Infirmary of Edinburgh, NHS Lothian / School of Biological Sciences, University of Edinburgh / Institute of Genetics and Molecular Medicine, University of Edinburgh | COVID-19 Genomics UK (COG-UK) Consortium |
| EPI_ISL_425906 | Virology Department, Royal Infirmary of Edinburgh, NHS Lothian / School of Biological Sciences, University of Edinburgh / Institute of Genetics and Molecular Medicine, University of Edinburgh | COVID-19 Genomics UK (COG-UK) Consortium |

| EPI_Accession_ID | Originating Laboratory | Submitting Laboratory |
|---|---|---|
| EPI_ISL_425907 | Virology Department, Royal Infirmary of Edinburgh, NHS Lothian / School of Biological Sciences, University of Edinburgh / Institute of Genetics and Molecular Medicine, University of Edinburgh | COVID-19 Genomics UK (COG-UK) Consortium |
| EPI_ISL_425908 | Virology Department, Royal Infirmary of Edinburgh, NHS Lothian / School of Biological Sciences, University of Edinburgh / Institute of Genetics and Molecular Medicine, University of Edinburgh | COVID-19 Genomics UK (COG-UK) Consortium |
| EPI_ISL_425909 | Virology Department, Royal Infirmary of Edinburgh, NHS Lothian / School of Biological Sciences, University of Edinburgh / Institute of Genetics and Molecular Medicine, University of Edinburgh | COVID-19 Genomics UK (COG-UK) Consortium |
| EPI_ISL_425910 | Virology Department, Royal Infirmary of Edinburgh, NHS Lothian / School of Biological Sciences, University of Edinburgh / Institute of Genetics and Molecular Medicine, University of Edinburgh | COVID-19 Genomics UK (COG-UK) Consortium |
| EPI_ISL_425911 | Virology Department, Royal Infirmary of Edinburgh, NHS Lothian / School of Biological Sciences, University of Edinburgh / Institute of Genetics and Molecular Medicine, University of Edinburgh | COVID-19 Genomics UK (COG-UK) Consortium |
| EPI_ISL_425912 | Virology Department, Royal Infirmary of Edinburgh, NHS Lothian / School of Biological Sciences, University of Edinburgh / Institute of Genetics and Molecular Medicine, University of Edinburgh | COVID-19 Genomics UK (COG-UK) Consortium |
| EPI_ISL_425913 | Virology Department, Royal Infirmary of Edinburgh, NHS Lothian / School of Biological Sciences, University of Edinburgh / Institute of Genetics and Molecular Medicine, University of Edinburgh | COVID-19 Genomics UK (COG-UK) Consortium |
| EPI_ISL_425914 | Virology Department, Royal Infirmary of Edinburgh, NHS Lothian / School of Biological Sciences, University of Edinburgh / Institute of Genetics and Molecular Medicine, University of Edinburgh | COVID-19 Genomics UK (COG-UK) Consortium |

| EPI_Accession_ID | Originating Laboratory | Submitting Laboratory |
|---|---|---|
| EPI_ISL_425916 | Virology Department, Royal Infirmary of Edinburgh, NHS Lothian / School of Biological Sciences, University of Edinburgh / Institute of Genetics and Molecular Medicine, University of Edinburgh | COVID-19 Genomics UK (COG-UK) Consortium |
| EPI_ISL_425917 | Virology Department, Royal Infirmary of Edinburgh, NHS Lothian / School of Biological Sciences, University of Edinburgh / Institute of Genetics and Molecular Medicine, University of Edinburgh | COVID-19 Genomics UK (COG-UK) Consortium |
| EPI_ISL_425918 | Virology Department, Royal Infirmary of Edinburgh, NHS Lothian / School of Biological Sciences, University of Edinburgh / Institute of Genetics and Molecular Medicine, University of Edinburgh | COVID-19 Genomics UK (COG-UK) Consortium |
| EPI_ISL_425919 | Virology Department, Royal Infirmary of Edinburgh, NHS Lothian / School of Biological Sciences, University of Edinburgh / Institute of Genetics and Molecular Medicine, University of Edinburgh | COVID-19 Genomics UK (COG-UK) Consortium |
| EPI_ISL_425920 | Virology Department, Royal Infirmary of Edinburgh, NHS Lothian / School of Biological Sciences, University of Edinburgh / Institute of Genetics and Molecular Medicine, University of Edinburgh | COVID-19 Genomics UK (COG-UK) Consortium |
| EPI_ISL_425921 | Virology Department, Royal Infirmary of Edinburgh, NHS Lothian / School of Biological Sciences, University of Edinburgh / Institute of Genetics and Molecular Medicine, University of Edinburgh | COVID-19 Genomics UK (COG-UK) Consortium |
| EPI_ISL_425922 | Virology Department, Royal Infirmary of Edinburgh, NHS Lothian / School of Biological Sciences, University of Edinburgh / Institute of Genetics and Molecular Medicine, University of Edinburgh | COVID-19 Genomics UK (COG-UK) Consortium |
| EPI_ISL_425923 | Virology Department, Royal Infirmary of Edinburgh, NHS Lothian / School of Biological Sciences, University of Edinburgh / Institute of Genetics and Molecular Medicine, University of Edinburgh | COVID-19 Genomics UK (COG-UK) Consortium |

| EPI_Accession_ID | Originating Laboratory | Submitting Laboratory |
|---|---|---|
| EPI_ISL_425924 | Virology Department, Royal Infirmary of Edinburgh, NHS Lothian / School of Biological Sciences, University of Edinburgh / Institute of Genetics and Molecular Medicine, University of Edinburgh | COVID-19 Genomics UK (COG-UK) Consortium |
| EPI_ISL_425925 | Virology Department, Royal Infirmary of Edinburgh, NHS Lothian / School of Biological Sciences, University of Edinburgh / Institute of Genetics and Molecular Medicine, University of Edinburgh | COVID-19 Genomics UK (COG-UK) Consortium |
| EPI_ISL_425926 | Virology Department, Royal Infirmary of Edinburgh, NHS Lothian / School of Biological Sciences, University of Edinburgh / Institute of Genetics and Molecular Medicine, University of Edinburgh | COVID-19 Genomics UK (COG-UK) Consortium |
| EPI_ISL_425927 | Virology Department, Royal Infirmary of Edinburgh, NHS Lothian / School of Biological Sciences, University of Edinburgh / Institute of Genetics and Molecular Medicine, University of Edinburgh | COVID-19 Genomics UK (COG-UK) Consortium |
| EPI_ISL_425928 | Virology Department, Royal Infirmary of Edinburgh, NHS Lothian / School of Biological Sciences, University of Edinburgh / Institute of Genetics and Molecular Medicine, University of Edinburgh | COVID-19 Genomics UK (COG-UK) Consortium |
| EPI_ISL_425929 | Virology Department, Royal Infirmary of Edinburgh, NHS Lothian / School of Biological Sciences, University of Edinburgh / Institute of Genetics and Molecular Medicine, University of Edinburgh | COVID-19 Genomics UK (COG-UK) Consortium |
| EPI_ISL_425930 | Virology Department, Royal Infirmary of Edinburgh, NHS Lothian / School of Biological Sciences, University of Edinburgh / Institute of Genetics and Molecular Medicine, University of Edinburgh | COVID-19 Genomics UK (COG-UK) Consortium |
| EPI_ISL_425931 | Virology Department, Royal Infirmary of Edinburgh, NHS Lothian / School of Biological Sciences, University of Edinburgh / Institute of Genetics and Molecular Medicine, University of Edinburgh | COVID-19 Genomics UK (COG-UK) Consortium |

| EPI_Accession_ID | Originating Laboratory | Submitting Laboratory |
|---|---|---|
| EPI_ISL_425932 | Virology Department, Royal Infirmary of Edinburgh, NHS Lothian / School of Biological Sciences, University of Edinburgh / Institute of Genetics and Molecular Medicine, University of Edinburgh | COVID-19 Genomics UK (COG-UK) Consortium |
| EPI_ISL_425933 | Virology Department, Royal Infirmary of Edinburgh, NHS Lothian / School of Biological Sciences, University of Edinburgh / Institute of Genetics and Molecular Medicine, University of Edinburgh | COVID-19 Genomics UK (COG-UK) Consortium |
| EPI_ISL_425934 | Virology Department, Royal Infirmary of Edinburgh, NHS Lothian / School of Biological Sciences, University of Edinburgh / Institute of Genetics and Molecular Medicine, University of Edinburgh | COVID-19 Genomics UK (COG-UK) Consortium |
| EPI_ISL_425935 | Virology Department, Royal Infirmary of Edinburgh, NHS Lothian / School of Biological Sciences, University of Edinburgh / Institute of Genetics and Molecular Medicine, University of Edinburgh | COVID-19 Genomics UK (COG-UK) Consortium |
| EPI_ISL_425936 | Virology Department, Royal Infirmary of Edinburgh, NHS Lothian / School of Biological Sciences, University of Edinburgh / Institute of Genetics and Molecular Medicine, University of Edinburgh | COVID-19 Genomics UK (COG-UK) Consortium |
| EPI_ISL_425937 | Virology Department, Royal Infirmary of Edinburgh, NHS Lothian / School of Biological Sciences, University of Edinburgh / Institute of Genetics and Molecular Medicine, University of Edinburgh | COVID-19 Genomics UK (COG-UK) Consortium |
| EPI_ISL_425938 | Virology Department, Royal Infirmary of Edinburgh, NHS Lothian / School of Biological Sciences, University of Edinburgh / Institute of Genetics and Molecular Medicine, University of Edinburgh | COVID-19 Genomics UK (COG-UK) Consortium |
| EPI_ISL_425939 | Virology Department, Royal Infirmary of Edinburgh, NHS Lothian / School of Biological Sciences, University of Edinburgh / Institute of Genetics and Molecular Medicine, University of Edinburgh | COVID-19 Genomics UK (COG-UK) Consortium |

| EPI_Accession_ID | Originating Laboratory | Submitting Laboratory |
|---|---|---|
| EPI_ISL_425940 | Virology Department, Royal Infirmary of Edinburgh, NHS Lothian / School of Biological Sciences, University of Edinburgh / Institute of Genetics and Molecular Medicine, University of Edinburgh | COVID-19 Genomics UK (COG-UK) Consortium |
| EPI_ISL_425941 | Virology Department, Royal Infirmary of Edinburgh, NHS Lothian / School of Biological Sciences, University of Edinburgh / Institute of Genetics and Molecular Medicine, University of Edinburgh | COVID-19 Genomics UK (COG-UK) Consortium |
| EPI_ISL_425942 | Virology Department, Royal Infirmary of Edinburgh, NHS Lothian / School of Biological Sciences, University of Edinburgh / Institute of Genetics and Molecular Medicine, University of Edinburgh | COVID-19 Genomics UK (COG-UK) Consortium |
| EPI_ISL_425943 | Virology Department, Royal Infirmary of Edinburgh, NHS Lothian / School of Biological Sciences, University of Edinburgh / Institute of Genetics and Molecular Medicine, University of Edinburgh | COVID-19 Genomics UK (COG-UK) Consortium |
| EPI_ISL_425944 | Virology Department, Royal Infirmary of Edinburgh, NHS Lothian / School of Biological Sciences, University of Edinburgh / Institute of Genetics and Molecular Medicine, University of Edinburgh | COVID-19 Genomics UK (COG-UK) Consortium |
| EPI_ISL_425945 | Virology Department, Royal Infirmary of Edinburgh, NHS Lothian / School of Biological Sciences, University of Edinburgh / Institute of Genetics and Molecular Medicine, University of Edinburgh | COVID-19 Genomics UK (COG-UK) Consortium |
| EPI_ISL_425946 | Virology Department, Royal Infirmary of Edinburgh, NHS Lothian / School of Biological Sciences, University of Edinburgh / Institute of Genetics and Molecular Medicine, University of Edinburgh | COVID-19 Genomics UK (COG-UK) Consortium |
| EPI_ISL_425947 | Virology Department, Royal Infirmary of Edinburgh, NHS Lothian / School of Biological Sciences, University of Edinburgh / Institute of Genetics and Molecular Medicine, University of Edinburgh | COVID-19 Genomics UK (COG-UK) Consortium |

| EPI_Accession_ID | Originating Laboratory | Submitting Laboratory |
|---|---|---|
| EPI_ISL_425948 | Virology Department, Royal Infirmary of Edinburgh, NHS Lothian / School of Biological Sciences, University of Edinburgh / Institute of Genetics and Molecular Medicine, University of Edinburgh | COVID-19 Genomics UK (COG-UK) Consortium |
| EPI_ISL_425949 | Virology Department, Royal Infirmary of Edinburgh, NHS Lothian / School of Biological Sciences, University of Edinburgh / Institute of Genetics and Molecular Medicine, University of Edinburgh | COVID-19 Genomics UK (COG-UK) Consortium |
| EPI_ISL_425950 | Virology Department, Royal Infirmary of Edinburgh, NHS Lothian / School of Biological Sciences, University of Edinburgh / Institute of Genetics and Molecular Medicine, University of Edinburgh | COVID-19 Genomics UK (COG-UK) Consortium |
| EPI_ISL_425951 | Virology Department, Royal Infirmary of Edinburgh, NHS Lothian / School of Biological Sciences, University of Edinburgh / Institute of Genetics and Molecular Medicine, University of Edinburgh | COVID-19 Genomics UK (COG-UK) Consortium |
| EPI_ISL_425952 | Virology Department, Royal Infirmary of Edinburgh, NHS Lothian / School of Biological Sciences, University of Edinburgh / Institute of Genetics and Molecular Medicine, University of Edinburgh | COVID-19 Genomics UK (COG-UK) Consortium |
| EPI_ISL_425953 | Virology Department, Royal Infirmary of Edinburgh, NHS Lothian / School of Biological Sciences, University of Edinburgh / Institute of Genetics and Molecular Medicine, University of Edinburgh | COVID-19 Genomics UK (COG-UK) Consortium |
| EPI_ISL_425954 | Virology Department, Royal Infirmary of Edinburgh, NHS Lothian / School of Biological Sciences, University of Edinburgh / Institute of Genetics and Molecular Medicine, University of Edinburgh | COVID-19 Genomics UK (COG-UK) Consortium |
| EPI_ISL_425955 | Virology Department, Royal Infirmary of Edinburgh, NHS Lothian / School of Biological Sciences, University of Edinburgh / Institute of Genetics and Molecular Medicine, University of Edinburgh | COVID-19 Genomics UK (COG-UK) Consortium |

| EPI_Accession_ID | Originating Laboratory | Submitting Laboratory |
|---|---|---|
| EPI_ISL_425956 | Virology Department, Royal Infirmary of Edinburgh, NHS Lothian / School of Biological Sciences, University of Edinburgh / Institute of Genetics and Molecular Medicine, University of Edinburgh | COVID-19 Genomics UK (COG-UK) Consortium |
| EPI_ISL_425957 | Virology Department, Royal Infirmary of Edinburgh, NHS Lothian / School of Biological Sciences, University of Edinburgh / Institute of Genetics and Molecular Medicine, University of Edinburgh | COVID-19 Genomics UK (COG-UK) Consortium |
| EPI_ISL_425958 | Virology Department, Royal Infirmary of Edinburgh, NHS Lothian / School of Biological Sciences, University of Edinburgh / Institute of Genetics and Molecular Medicine, University of Edinburgh | COVID-19 Genomics UK (COG-UK) Consortium |
| EPI_ISL_425959 | Virology Department, Royal Infirmary of Edinburgh, NHS Lothian / School of Biological Sciences, University of Edinburgh / Institute of Genetics and Molecular Medicine, University of Edinburgh | COVID-19 Genomics UK (COG-UK) Consortium |
| EPI_ISL_425960 | Virology Department, Royal Infirmary of Edinburgh, NHS Lothian / School of Biological Sciences, University of Edinburgh / Institute of Genetics and Molecular Medicine, University of Edinburgh | COVID-19 Genomics UK (COG-UK) Consortium |
| EPI_ISL_425961 | Virology Department, Royal Infirmary of Edinburgh, NHS Lothian / School of Biological Sciences, University of Edinburgh / Institute of Genetics and Molecular Medicine, University of Edinburgh | COVID-19 Genomics UK (COG-UK) Consortium |
| EPI_ISL_425962 | Virology Department, Royal Infirmary of Edinburgh, NHS Lothian / School of Biological Sciences, University of Edinburgh / Institute of Genetics and Molecular Medicine, University of Edinburgh | COVID-19 Genomics UK (COG-UK) Consortium |
| EPI_ISL_425963 | Virology Department, Royal Infirmary of Edinburgh, NHS Lothian / School of Biological Sciences, University of Edinburgh / Institute of Genetics and Molecular Medicine, University of Edinburgh | COVID-19 Genomics UK (COG-UK) Consortium |

| EPI_Accession_ID | Originating Laboratory | Submitting Laboratory |
|---|---|---|
| EPI_ISL_425964 | Virology Department, Royal Infirmary of Edinburgh, NHS Lothian / School of Biological Sciences, University of Edinburgh / Institute of Genetics and Molecular Medicine, University of Edinburgh | COVID-19 Genomics UK (COG-UK) Consortium |
| EPI_ISL_425965 | Virology Department, Royal Infirmary of Edinburgh, NHS Lothian / School of Biological Sciences, University of Edinburgh / Institute of Genetics and Molecular Medicine, University of Edinburgh | COVID-19 Genomics UK (COG-UK) Consortium |
| EPI_ISL_425966 | Virology Department, Royal Infirmary of Edinburgh, NHS Lothian / School of Biological Sciences, University of Edinburgh / Institute of Genetics and Molecular Medicine, University of Edinburgh | COVID-19 Genomics UK (COG-UK) Consortium |
| EPI_ISL_425967 | Virology Department, Royal Infirmary of Edinburgh, NHS Lothian / School of Biological Sciences, University of Edinburgh / Institute of Genetics and Molecular Medicine, University of Edinburgh | COVID-19 Genomics UK (COG-UK) Consortium |
| EPI_ISL_425968 | Virology Department, Royal Infirmary of Edinburgh, NHS Lothian / School of Biological Sciences, University of Edinburgh / Institute of Genetics and Molecular Medicine, University of Edinburgh | COVID-19 Genomics UK (COG-UK) Consortium |
| EPI_ISL_425969 | Virology Department, Royal Infirmary of Edinburgh, NHS Lothian / School of Biological Sciences, University of Edinburgh / Institute of Genetics and Molecular Medicine, University of Edinburgh | COVID-19 Genomics UK (COG-UK) Consortium |
| EPI_ISL_425970 | Virology Department, Royal Infirmary of Edinburgh, NHS Lothian / School of Biological Sciences, University of Edinburgh / Institute of Genetics and Molecular Medicine, University of Edinburgh | COVID-19 Genomics UK (COG-UK) Consortium |
| EPI_ISL_425971 | Virology Department, Royal Infirmary of Edinburgh, NHS Lothian / School of Biological Sciences, University of Edinburgh / Institute of Genetics and Molecular Medicine, University of Edinburgh | COVID-19 Genomics UK (COG-UK) Consortium |

| EPI_Accession_ID | Originating Laboratory | Submitting Laboratory |
|---|---|---|
| EPI_ISL_425972 | Virology Department, Royal Infirmary of Edinburgh, NHS Lothian / School of Biological Sciences, University of Edinburgh / Institute of Genetics and Molecular Medicine, University of Edinburgh | COVID-19 Genomics UK (COG-UK) Consortium |
| EPI_ISL_425973 | Virology Department, Royal Infirmary of Edinburgh, NHS Lothian / School of Biological Sciences, University of Edinburgh / Institute of Genetics and Molecular Medicine, University of Edinburgh | COVID-19 Genomics UK (COG-UK) Consortium |
| EPI_ISL_425974 | Virology Department, Royal Infirmary of Edinburgh, NHS Lothian / School of Biological Sciences, University of Edinburgh / Institute of Genetics and Molecular Medicine, University of Edinburgh | COVID-19 Genomics UK (COG-UK) Consortium |
| EPI_ISL_425975 | Virology Department, Royal Infirmary of Edinburgh, NHS Lothian / School of Biological Sciences, University of Edinburgh / Institute of Genetics and Molecular Medicine, University of Edinburgh | COVID-19 Genomics UK (COG-UK) Consortium |
| EPI_ISL_425976 | Virology Department, Royal Infirmary of Edinburgh, NHS Lothian / School of Biological Sciences, University of Edinburgh / Institute of Genetics and Molecular Medicine, University of Edinburgh | COVID-19 Genomics UK (COG-UK) Consortium |
| EPI_ISL_425977 | Virology Department, Royal Infirmary of Edinburgh, NHS Lothian / School of Biological Sciences, University of Edinburgh / Institute of Genetics and Molecular Medicine, University of Edinburgh | COVID-19 Genomics UK (COG-UK) Consortium |
| EPI_ISL_425978 | Virology Department, Royal Infirmary of Edinburgh, NHS Lothian / School of Biological Sciences, University of Edinburgh / Institute of Genetics and Molecular Medicine, University of Edinburgh | COVID-19 Genomics UK (COG-UK) Consortium |
| EPI_ISL_425979 | Virology Department, Royal Infirmary of Edinburgh, NHS Lothian / School of Biological Sciences, University of Edinburgh / Institute of Genetics and Molecular Medicine, University of Edinburgh | COVID-19 Genomics UK (COG-UK) Consortium |

| EPI_Accession_ID | Originating Laboratory | Submitting Laboratory |
|---|---|---|
| EPI_ISL_425980 | Virology Department, Royal Infirmary of Edinburgh, NHS Lothian / School of Biological Sciences, University of Edinburgh / Institute of Genetics and Molecular Medicine, University of Edinburgh | COVID-19 Genomics UK (COG-UK) Consortium |
| EPI_ISL_425981 | Virology Department, Royal Infirmary of Edinburgh, NHS Lothian / School of Biological Sciences, University of Edinburgh / Institute of Genetics and Molecular Medicine, University of Edinburgh | COVID-19 Genomics UK (COG-UK) Consortium |
| EPI_ISL_425982 | Virology Department, Royal Infirmary of Edinburgh, NHS Lothian / School of Biological Sciences, University of Edinburgh / Institute of Genetics and Molecular Medicine, University of Edinburgh | COVID-19 Genomics UK (COG-UK) Consortium |
| EPI_ISL_425983 | Virology Department, Royal Infirmary of Edinburgh, NHS Lothian / School of Biological Sciences, University of Edinburgh / Institute of Genetics and Molecular Medicine, University of Edinburgh | COVID-19 Genomics UK (COG-UK) Consortium |
| EPI_ISL_425984 | Virology Department, Royal Infirmary of Edinburgh, NHS Lothian / School of Biological Sciences, University of Edinburgh / Institute of Genetics and Molecular Medicine, University of Edinburgh | COVID-19 Genomics UK (COG-UK) Consortium |
| EPI_ISL_425985 | Virology Department, Royal Infirmary of Edinburgh, NHS Lothian / School of Biological Sciences, University of Edinburgh / Institute of Genetics and Molecular Medicine, University of Edinburgh | COVID-19 Genomics UK (COG-UK) Consortium |
| EPI_ISL_425986 | Virology Department, Royal Infirmary of Edinburgh, NHS Lothian / School of Biological Sciences, University of Edinburgh / Institute of Genetics and Molecular Medicine, University of Edinburgh | COVID-19 Genomics UK (COG-UK) Consortium |
| EPI_ISL_425987 | Virology Department, Royal Infirmary of Edinburgh, NHS Lothian / School of Biological Sciences, University of Edinburgh / Institute of Genetics and Molecular Medicine, University of Edinburgh | COVID-19 Genomics UK (COG-UK) Consortium |

| EPI_Accession_ID | Originating Laboratory | Submitting Laboratory |
|---|---|---|
| EPI_ISL_425988 | Virology Department, Royal Infirmary of Edinburgh, NHS Lothian / School of Biological Sciences, University of Edinburgh / Institute of Genetics and Molecular Medicine, University of Edinburgh | COVID-19 Genomics UK (COG-UK) Consortium |
| EPI_ISL_425989 | Virology Department, Royal Infirmary of Edinburgh, NHS Lothian / School of Biological Sciences, University of Edinburgh / Institute of Genetics and Molecular Medicine, University of Edinburgh | COVID-19 Genomics UK (COG-UK) Consortium |
| EPI_ISL_425990 | Virology Department, Royal Infirmary of Edinburgh, NHS Lothian / School of Biological Sciences, University of Edinburgh / Institute of Genetics and Molecular Medicine, University of Edinburgh | COVID-19 Genomics UK (COG-UK) Consortium |
| EPI_ISL_425991 | Virology Department, Royal Infirmary of Edinburgh, NHS Lothian / School of Biological Sciences, University of Edinburgh / Institute of Genetics and Molecular Medicine, University of Edinburgh | COVID-19 Genomics UK (COG-UK) Consortium |
| EPI_ISL_425992 | Virology Department, Royal Infirmary of Edinburgh, NHS Lothian / School of Biological Sciences, University of Edinburgh / Institute of Genetics and Molecular Medicine, University of Edinburgh | COVID-19 Genomics UK (COG-UK) Consortium |
| EPI_ISL_425993 | Virology Department, Royal Infirmary of Edinburgh, NHS Lothian / School of Biological Sciences, University of Edinburgh / Institute of Genetics and Molecular Medicine, University of Edinburgh | COVID-19 Genomics UK (COG-UK) Consortium |
| EPI_ISL_425994 | Virology Department, Royal Infirmary of Edinburgh, NHS Lothian / School of Biological Sciences, University of Edinburgh / Institute of Genetics and Molecular Medicine, University of Edinburgh | COVID-19 Genomics UK (COG-UK) Consortium |
| EPI_ISL_425995 | Virology Department, Royal Infirmary of Edinburgh, NHS Lothian / School of Biological Sciences, University of Edinburgh / Institute of Genetics and Molecular Medicine, University of Edinburgh | COVID-19 Genomics UK (COG-UK) Consortium |

| EPI_Accession_ID | Originating Laboratory | Submitting Laboratory |
|---|---|---|
| EPI_ISL_425996 | Virology Department, Royal Infirmary of Edinburgh, NHS Lothian / School of Biological Sciences, University of Edinburgh / Institute of Genetics and Molecular Medicine, University of Edinburgh | COVID-19 Genomics UK (COG-UK) Consortium |
| EPI_ISL_425997 | Virology Department, Royal Infirmary of Edinburgh, NHS Lothian / School of Biological Sciences, University of Edinburgh / Institute of Genetics and Molecular Medicine, University of Edinburgh | COVID-19 Genomics UK (COG-UK) Consortium |
| EPI_ISL_425998 | Virology Department, Royal Infirmary of Edinburgh, NHS Lothian / School of Biological Sciences, University of Edinburgh / Institute of Genetics and Molecular Medicine, University of Edinburgh | COVID-19 Genomics UK (COG-UK) Consortium |
| EPI_ISL_425999 | Virology Department, Royal Infirmary of Edinburgh, NHS Lothian / School of Biological Sciences, University of Edinburgh / Institute of Genetics and Molecular Medicine, University of Edinburgh | COVID-19 Genomics UK (COG-UK) Consortium |
| EPI_ISL_426000 | Virology Department, Royal Infirmary of Edinburgh, NHS Lothian / School of Biological Sciences, University of Edinburgh / Institute of Genetics and Molecular Medicine, University of Edinburgh | COVID-19 Genomics UK (COG-UK) Consortium |
| EPI_ISL_426001 | Virology Department, Royal Infirmary of Edinburgh, NHS Lothian / School of Biological Sciences, University of Edinburgh / Institute of Genetics and Molecular Medicine, University of Edinburgh | COVID-19 Genomics UK (COG-UK) Consortium |
| EPI_ISL_426002 | Virology Department, Royal Infirmary of Edinburgh, NHS Lothian / School of Biological Sciences, University of Edinburgh / Institute of Genetics and Molecular Medicine, University of Edinburgh | COVID-19 Genomics UK (COG-UK) Consortium |
| EPI_ISL_426003 | Virology Department, Royal Infirmary of Edinburgh, NHS Lothian / School of Biological Sciences, University of Edinburgh / Institute of Genetics and Molecular Medicine, University of Edinburgh | COVID-19 Genomics UK (COG-UK) Consortium |

| EPI_Accession_ID | Originating Laboratory | Submitting Laboratory |
|---|---|---|
| EPI_ISL_426004 | Virology Department, Royal Infirmary of Edinburgh, NHS Lothian / School of Biological Sciences, University of Edinburgh / Institute of Genetics and Molecular Medicine, University of Edinburgh | COVID-19 Genomics UK (COG-UK) Consortium |
| EPI_ISL_426005 | Virology Department, Royal Infirmary of Edinburgh, NHS Lothian / School of Biological Sciences, University of Edinburgh / Institute of Genetics and Molecular Medicine, University of Edinburgh | COVID-19 Genomics UK (COG-UK) Consortium |
| EPI_ISL_426006 | Virology Department, Royal Infirmary of Edinburgh, NHS Lothian / School of Biological Sciences, University of Edinburgh / Institute of Genetics and Molecular Medicine, University of Edinburgh | COVID-19 Genomics UK (COG-UK) Consortium |
| EPI_ISL_426007 | Virology Department, Royal Infirmary of Edinburgh, NHS Lothian / School of Biological Sciences, University of Edinburgh / Institute of Genetics and Molecular Medicine, University of Edinburgh | COVID-19 Genomics UK (COG-UK) Consortium |
| EPI_ISL_426008 | Virology Department, Royal Infirmary of Edinburgh, NHS Lothian / School of Biological Sciences, University of Edinburgh / Institute of Genetics and Molecular Medicine, University of Edinburgh | COVID-19 Genomics UK (COG-UK) Consortium |
| EPI_ISL_426009 | Virology Department, Royal Infirmary of Edinburgh, NHS Lothian / School of Biological Sciences, University of Edinburgh / Institute of Genetics and Molecular Medicine, University of Edinburgh | COVID-19 Genomics UK (COG-UK) Consortium |
| EPI_ISL_426010 | Virology Department, Royal Infirmary of Edinburgh, NHS Lothian / School of Biological Sciences, University of Edinburgh / Institute of Genetics and Molecular Medicine, University of Edinburgh | COVID-19 Genomics UK (COG-UK) Consortium |
| EPI_ISL_426011 | Virology Department, Royal Infirmary of Edinburgh, NHS Lothian / School of Biological Sciences, University of Edinburgh / Institute of Genetics and Molecular Medicine, University of Edinburgh | COVID-19 Genomics UK (COG-UK) Consortium |

| EPI_Accession_ID | Originating Laboratory | Submitting Laboratory |
|---|---|---|
| EPI_ISL_426012 | Virology Department, Royal Infirmary of Edinburgh, NHS Lothian / School of Biological Sciences, University of Edinburgh / Institute of Genetics and Molecular Medicine, University of Edinburgh | COVID-19 Genomics UK (COG-UK) Consortium |
| EPI_ISL_426013 | Virology Department, Royal Infirmary of Edinburgh, NHS Lothian / School of Biological Sciences, University of Edinburgh / Institute of Genetics and Molecular Medicine, University of Edinburgh | COVID-19 Genomics UK (COG-UK) Consortium |
| EPI_ISL_426014 | Virology Department, Royal Infirmary of Edinburgh, NHS Lothian / School of Biological Sciences, University of Edinburgh / Institute of Genetics and Molecular Medicine, University of Edinburgh | COVID-19 Genomics UK (COG-UK) Consortium |
| EPI_ISL_426015 | Virology Department, Royal Infirmary of Edinburgh, NHS Lothian / School of Biological Sciences, University of Edinburgh / Institute of Genetics and Molecular Medicine, University of Edinburgh | COVID-19 Genomics UK (COG-UK) Consortium |
| EPI_ISL_426016 | Virology Department, Royal Infirmary of Edinburgh, NHS Lothian / School of Biological Sciences, University of Edinburgh / Institute of Genetics and Molecular Medicine, University of Edinburgh | COVID-19 Genomics UK (COG-UK) Consortium |
| EPI_ISL_426017 | Virology Department, Royal Infirmary of Edinburgh, NHS Lothian / School of Biological Sciences, University of Edinburgh / Institute of Genetics and Molecular Medicine, University of Edinburgh | COVID-19 Genomics UK (COG-UK) Consortium |
| EPI_ISL_426018 | Virology Department, Royal Infirmary of Edinburgh, NHS Lothian / School of Biological Sciences, University of Edinburgh / Institute of Genetics and Molecular Medicine, University of Edinburgh | COVID-19 Genomics UK (COG-UK) Consortium |
| EPI_ISL_426019 | Virology Department, Royal Infirmary of Edinburgh, NHS Lothian / School of Biological Sciences, University of Edinburgh / Institute of Genetics and Molecular Medicine, University of Edinburgh | COVID-19 Genomics UK (COG-UK) Consortium |

| EPI_Accession_ID | Originating Laboratory | Submitting Laboratory |
|---|---|---|
| EPI_ISL_426020 | Virology Department, Royal Infirmary of Edinburgh, NHS Lothian / School of Biological Sciences, University of Edinburgh / Institute of Genetics and Molecular Medicine, University of Edinburgh | COVID-19 Genomics UK (COG-UK) Consortium |
| EPI_ISL_426021 | Virology Department, Royal Infirmary of Edinburgh, NHS Lothian / School of Biological Sciences, University of Edinburgh / Institute of Genetics and Molecular Medicine, University of Edinburgh | COVID-19 Genomics UK (COG-UK) Consortium |
| EPI_ISL_426022 | Virology Department, Royal Infirmary of Edinburgh, NHS Lothian / School of Biological Sciences, University of Edinburgh / Institute of Genetics and Molecular Medicine, University of Edinburgh | COVID-19 Genomics UK (COG-UK) Consortium |
| EPI_ISL_426023 | Virology Department, Royal Infirmary of Edinburgh, NHS Lothian / School of Biological Sciences, University of Edinburgh / Institute of Genetics and Molecular Medicine, University of Edinburgh | COVID-19 Genomics UK (COG-UK) Consortium |
| EPI_ISL_426024 | Virology Department, Royal Infirmary of Edinburgh, NHS Lothian / School of Biological Sciences, University of Edinburgh / Institute of Genetics and Molecular Medicine, University of Edinburgh | COVID-19 Genomics UK (COG-UK) Consortium |
| EPI_ISL_426025 | Wadsworth Center, New York State Department.of Health | Wadsworth Center, New York State Department.of Health |
| EPI_ISL_426026 | Wadsworth Center, New York State Department.of Health | Wadsworth Center, New York State Department.of Health |
| EPI_ISL_426027 | Wadsworth Center, New York State Department.of Health | Wadsworth Center, New York State Department.of Health |
| EPI_ISL_426028 | Wadsworth Center, New York State Department.of Health | Wadsworth Center, New York State Department.of Health |
| EPI_ISL_426029 | Wadsworth Center, New York State Department.of Health | Wadsworth Center, New York State Department.of Health |
| EPI_ISL_426030 | Wadsworth Center, New York State Department.of Health | Wadsworth Center, New York State Department.of Health |

| EPI_Accession_ID | Originating Laboratory | Submitting Laboratory |
|---|---|---|
| EPI_ISL_426031 | Wadsworth Center, New York State Department.of Health | Wadsworth Center, New York State Department.of Health |
| EPI_ISL_426032 | Wadsworth Center, New York State Department.of Health | Wadsworth Center, New York State Department.of Health |
| EPI_ISL_426033 | Wadsworth Center, New York State Department.of Health | Wadsworth Center, New York State Department.of Health |
| EPI_ISL_426034 | Wadsworth Center, New York State Department.of Health | Wadsworth Center, New York State Department.of Health |
| EPI_ISL_426035 | Wadsworth Center, New York State Department.of Health | Wadsworth Center, New York State Department.of Health |
| EPI_ISL_426036 | Wadsworth Center, New York State Department.of Health | Wadsworth Center, New York State Department.of Health |
| EPI_ISL_426037 | Wadsworth Center, New York State Department.of Health | Wadsworth Center, New York State Department.of Health |
| EPI_ISL_426038 | Wadsworth Center, New York State Department.of Health | Wadsworth Center, New York State Department.of Health |
| EPI_ISL_426039 | Wadsworth Center, New York State Department.of Health | Wadsworth Center, New York State Department.of Health |
| EPI_ISL_426040 | Wadsworth Center, New York State Department.of Health | Wadsworth Center, New York State Department.of Health |
| EPI_ISL_426041 | Wadsworth Center, New York State Department.of Health | Wadsworth Center, New York State Department.of Health |
| EPI_ISL_426042 | Wadsworth Center, New York State Department.of Health | Wadsworth Center, New York State Department.of Health |
| EPI_ISL_426043 | Wadsworth Center, New York State Department.of Health | Wadsworth Center, New York State Department.of Health |
| EPI_ISL_426044 | Wadsworth Center, New York State Department.of Health | Wadsworth Center, New York State Department.of Health |
| EPI_ISL_426045 | Wadsworth Center, New York State Department.of Health | Wadsworth Center, New York State Department.of Health |
| EPI_ISL_426046 | Wadsworth Center, New York State Department.of Health | Wadsworth Center, New York State Department.of Health |

| EPI_Accession_ID | Originating Laboratory | Submitting Laboratory |
|---|---|---|
| EPI_ISL_426047 | Wadsworth Center, New York State Department.of Health | Wadsworth Center, New York State Department.of Health |
| EPI_ISL_426048 | Wadsworth Center, New York State Department.of Health | Wadsworth Center, New York State Department.of Health |
| EPI_ISL_426049 | Wadsworth Center, New York State Department.of Health | Wadsworth Center, New York State Department.of Health |
| EPI_ISL_426050 | Wadsworth Center, New York State Department.of Health | Wadsworth Center, New York State Department.of Health |
| EPI_ISL_426051 | Laboratory of Molecular Genetics, 2nd Faculty of Medicine, Charles University in Prague, Prague, Czech Republic | Laboratory of Molecular Genetics, 2nd Faculty of Medicine, Charles University in Prague, Prague, Czech Republic |
| EPI_ISL_426052 | UW Virology Lab | UW Virology Lab |
| EPI_ISL_426053 | UW Virology Lab | UW Virology Lab |
| EPI_ISL_426054 | UW Virology Lab | UW Virology Lab |
| EPI_ISL_426055 | UW Virology Lab | UW Virology Lab |
| EPI_ISL_426056 | UW Virology Lab | UW Virology Lab |
| EPI_ISL_426057 | UW Virology Lab | UW Virology Lab |
| EPI_ISL_426058 | UW Virology Lab | UW Virology Lab |
| EPI_ISL_426059 | UW Virology Lab | UW Virology Lab |
| EPI_ISL_426060 | UW Virology Lab | UW Virology Lab |
| EPI_ISL_426061 | UW Virology Lab | UW Virology Lab |
| EPI_ISL_426062 | UW Virology Lab | UW Virology Lab |
| EPI_ISL_426063 | UW Virology Lab | UW Virology Lab |
| EPI_ISL_426064 | UW Virology Lab | UW Virology Lab |
| EPI_ISL_426065 | UW Virology Lab | UW Virology Lab |
| EPI_ISL_426066 | UW Virology Lab | UW Virology Lab |
| EPI_ISL_426067 | UW Virology Lab | UW Virology Lab |
| EPI_ISL_426068 | UW Virology Lab | UW Virology Lab |
| EPI_ISL_426069 | UW Virology Lab | UW Virology Lab |
| EPI_ISL_426070 | UW Virology Lab | UW Virology Lab |
| EPI_ISL_426071 | UW Virology Lab | UW Virology Lab |
| EPI_ISL_426072 | UW Virology Lab | UW Virology Lab |
| EPI_ISL_426073 | UW Virology Lab | UW Virology Lab |

| EPI_Accession_ID | Originating Laboratory | Submitting Laboratory |
|---|---|---|
| EPI_ISL_426074 | UW Virology Lab | UW Virology Lab |
| EPI_ISL_426075 | UW Virology Lab | UW Virology Lab |
| EPI_ISL_426076 | UW Virology Lab | UW Virology Lab |
| EPI_ISL_426077 | UW Virology Lab | UW Virology Lab |
| EPI_ISL_426078 | UW Virology Lab | UW Virology Lab |
| EPI_ISL_426079 | UW Virology Lab | UW Virology Lab |
| EPI_ISL_426080 | UW Virology Lab | UW Virology Lab |
| EPI_ISL_426081 | UW Virology Lab | UW Virology Lab |
| EPI_ISL_426082 | UW Virology Lab | UW Virology Lab |
| EPI_ISL_426083 | UW Virology Lab | UW Virology Lab |
| EPI_ISL_426084 | UW Virology Lab | UW Virology Lab |
| EPI_ISL_426085 | UW Virology Lab | UW Virology Lab |
| EPI_ISL_426086 | UW Virology Lab | UW Virology Lab |
| EPI_ISL_426087 | UW Virology Lab | UW Virology Lab |
| EPI_ISL_426089 | UW Virology Lab | UW Virology Lab |
| EPI_ISL_426090 | UW Virology Lab | UW Virology Lab |
| EPI_ISL_426091 | UW Virology Lab | UW Virology Lab |
| EPI_ISL_426092 | UW Virology Lab | UW Virology Lab |
| EPI_ISL_426093 | UW Virology Lab | UW Virology Lab |
| EPI_ISL_426094 | UW Virology Lab | UW Virology Lab |
| EPI_ISL_426095 | UW Virology Lab | UW Virology Lab |
| EPI_ISL_426096 | UW Virology Lab | UW Virology Lab |
| EPI_ISL_426097 | UW Virology Lab | UW Virology Lab |
| EPI_ISL_426098 | UW Virology Lab | UW Virology Lab |
| EPI_ISL_426099 | UW Virology Lab | UW Virology Lab |
| EPI_ISL_426100 | UW Virology Lab | UW Virology Lab |
| EPI_ISL_426101 | UW Virology Lab | UW Virology Lab |
| EPI_ISL_426102 | UW Virology Lab | UW Virology Lab |
| EPI_ISL_426103 | UW Virology Lab | UW Virology Lab |
| EPI_ISL_426104 | UW Virology Lab | UW Virology Lab |
| EPI_ISL_426105 | UW Virology Lab | UW Virology Lab |
| EPI_ISL_426106 | UW Virology Lab | UW Virology Lab |
| EPI_ISL_426107 | UW Virology Lab | UW Virology Lab |

| EPI_Accession_ID | Originating Laboratory | Submitting Laboratory |
|---|---|---|
| EPI_ISL_426108 | UW Virology Lab | UW Virology Lab |
| EPI_ISL_426109 | UW Virology Lab | UW Virology Lab |
| EPI_ISL_426110 | UW Virology Lab | UW Virology Lab |
| EPI_ISL_426111 | UW Virology Lab | UW Virology Lab |
| EPI_ISL_426112 | UW Virology Lab | UW Virology Lab |
| EPI_ISL_426113 | UW Virology Lab | UW Virology Lab |
| EPI_ISL_426114 | UW Virology Lab | UW Virology Lab |
| EPI_ISL_426115 | UW Virology Lab | UW Virology Lab |
| EPI_ISL_426116 | UW Virology Lab | UW Virology Lab |
| EPI_ISL_426117 | UW Virology Lab | UW Virology Lab |
| EPI_ISL_426118 | UW Virology Lab | UW Virology Lab |
| EPI_ISL_426119 | UW Virology Lab | UW Virology Lab |
| EPI_ISL_426120 | UW Virology Lab | UW Virology Lab |
| EPI_ISL_426121 | UW Virology Lab | UW Virology Lab |
| EPI_ISL_426122 | UW Virology Lab | UW Virology Lab |
| EPI_ISL_426123 | UW Virology Lab | UW Virology Lab |
| EPI_ISL_426124 | UW Virology Lab | UW Virology Lab |
| EPI_ISL_426125 | UW Virology Lab | UW Virology Lab |
| EPI_ISL_426126 | UW Virology Lab | UW Virology Lab |
| EPI_ISL_426127 | UW Virology Lab | UW Virology Lab |
| EPI_ISL_426128 | UW Virology Lab | UW Virology Lab |
| EPI_ISL_426129 | UW Virology Lab | UW Virology Lab |
| EPI_ISL_426130 | UW Virology Lab | UW Virology Lab |
| EPI_ISL_426131 | UW Virology Lab | UW Virology Lab |
| EPI_ISL_426132 | UW Virology Lab | UW Virology Lab |
| EPI_ISL_426133 | UW Virology Lab | UW Virology Lab |
| EPI_ISL_426134 | UW Virology Lab | UW Virology Lab |
| EPI_ISL_426135 | UW Virology Lab | UW Virology Lab |
| EPI_ISL_426136 | UW Virology Lab | UW Virology Lab |
| EPI_ISL_426137 | UW Virology Lab | UW Virology Lab |
| EPI_ISL_426159 | Gundersen Molecular Diagnostics Laboratory | Kabara Cancer Research Institute |
| EPI_ISL_426160 | Gundersen Molecular Diagnostics Laboratory | Kabara Cancer Research Institute |
| EPI_ISL_426161 | Gundersen Molecular Diagnostics Laboratory | Kabara Cancer Research Institute |

| EPI_Accession_ID | Originating Laboratory | Submitting Laboratory |
|---|---|---|
| EPI_ISL_426163 | Division of Viral Diseases, Center for Laboratory Control of Infectious Diseases, Korea Centers for Diseases Control and Prevention | Division of Viral Diseases, Center for Laboratory Control of Infectious Diseases, Korea Centers for Diseases Control and Prevention |
| EPI_ISL_426164 | Division of Viral Diseases, Center for Laboratory Control of Infectious Diseases, Korea Centers for Diseases Control and Prevention | Division of Viral Diseases, Center for Laboratory Control of Infectious Diseases, Korea Centers for Diseases Control and Prevention |
| EPI_ISL_426166 | Division of Viral Diseases, Center for Laboratory Control of Infectious Diseases, Korea Centers for Diseases Control and Prevention | Division of Viral Diseases, Center for Laboratory Control of Infectious Diseases, Korea Centers for Diseases Control and Prevention |
| EPI_ISL_426168 | Division of Viral Diseases, Center for Laboratory Control of Infectious Diseases, Korea Centers for Diseases Control and Prevention | Division of Viral Diseases, Center for Laboratory Control of Infectious Diseases, Korea Centers for Diseases Control and Prevention |
| EPI_ISL_426169 | Division of Viral Diseases, Center for Laboratory Control of Infectious Diseases, Korea Centers for Diseases Control and Prevention | Division of Viral Diseases, Center for Laboratory Control of Infectious Diseases, Korea Centers for Diseases Control and Prevention |
| EPI_ISL_426171 | Division of Viral Diseases, Center for Laboratory Control of Infectious Diseases, Korea Centers for Diseases Control and Prevention | Division of Viral Diseases, Center for Laboratory Control of Infectious Diseases, Korea Centers for Diseases Control and Prevention |
| EPI_ISL_426173 | Division of Viral Diseases, Center for Laboratory Control of Infectious Diseases, Korea Centers for Diseases Control and Prevention | Division of Viral Diseases, Center for Laboratory Control of Infectious Diseases, Korea Centers for Diseases Control and Prevention |
| EPI_ISL_426179 | National Influenza Center, Indian Council of Medical Research - National Institute of Virology | Indian Council of Medical Research-National Institute of Virology, Microbial Containment Complex |
| EPI_ISL_426180 | Division of Viral Diseases, Center for Laboratory Control of Infectious Diseases, Korea Centers for Diseases Control and Prevention | Division of Viral Diseases, Center for Laboratory Control of Infectious Diseases, Korea Centers for Diseases Control and Prevention |
| EPI_ISL_426181 | Division of Viral Diseases, Center for Laboratory Control of Infectious Diseases, Korea Centers for Diseases Control and Prevention | Division of Viral Diseases, Center for Laboratory Control of Infectious Diseases, Korea Centers for Diseases Control and Prevention |
| EPI_ISL_426182 | Division of Viral Diseases, Center for Laboratory Control of Infectious Diseases, Korea Centers for Diseases Control and Prevention | Division of Viral Diseases, Center for Laboratory Control of Infectious Diseases, Korea Centers for Diseases Control and Prevention |

| EPI_Accession_ID | Originating Laboratory | Submitting Laboratory |
|---|---|---|
| EPI_ISL_426183 | Division of Viral Diseases, Center for Laboratory Control of Infectious Diseases, Korea Centers for Diseases Control and Prevention | Division of Viral Diseases, Center for Laboratory Control of Infectious Diseases, Korea Centers for Diseases Control and Prevention |
| EPI_ISL_426187 | Division of Viral Diseases, Center for Laboratory Control of Infectious Diseases, Korea Centers for Diseases Control and Prevention | Division of Viral Diseases, Center for Laboratory Control of Infectious Diseases, Korea Centers for Diseases Control and Prevention |
| EPI_ISL_426285 | E. Gulbja Laboratorija | Latvian Biomedical Research and Study Centre |
| EPI_ISL_426286 | E. Gulbja Laboratorija | Latvian Biomedical Research and Study Centre |
| EPI_ISL_426287 | E. Gulbja Laboratorija | Latvian Biomedical Research and Study Centre |
| EPI_ISL_426288 | E. Gulbja Laboratorija | Latvian Biomedical Research and Study Centre |
| EPI_ISL_426289 | E. Gulbja Laboratorija | Latvian Biomedical Research and Study Centre |
| EPI_ISL_426290 | Wadsworth Center, New York State Department of Health | Wadsworth Center, New York State Department.of Health |
| EPI_ISL_426291 | Wadsworth Center, New York State Department.of Health | Wadsworth Center, New York State Department.of Health |
| EPI_ISL_426292 | Wadsworth Center, New York State Department.of Health | Wadsworth Center, New York State Department.of Health |
| EPI_ISL_426293 | Wadsworth Center, New York State Department.of Health | Wadsworth Center, New York State Department.of Health |
| EPI_ISL_426294 | Wadsworth Center, New York State Department.of Health | Wadsworth Center, New York State Department.of Health |
| EPI_ISL_426295 | Wadsworth Center, New York State Department.of Health | Wadsworth Center, New York State Department.of Health |
| EPI_ISL_426296 | Wadsworth Center, New York State Department.of Health | Wadsworth Center, New York State Department.of Health |
| EPI_ISL_426297 | Wadsworth Center, New York State Department.of Health | Wadsworth Center, New York State Department.of Health |
| EPI_ISL_426298 | Wadsworth Center, New York State Department.of Health | Wadsworth Center, New York State Department.of Health |
| EPI_ISL_426299 | Wadsworth Center, New York State Department.of Health | Wadsworth Center, New York State Department.of Health |
| EPI_ISL_426300 | Wadsworth Center, New York State Department.of Health | Wadsworth Center, New York State Department.of Health |

| EPI_Accession_ID | Originating Laboratory | Submitting Laboratory |
| --- | --- | --- |
| EPI_ISL_426301 | Wadsworth Center, New York State Department.of Health | Wadsworth Center, New York State Department.of Health |
| EPI_ISL_426302 | Wadsworth Center, New York State Department.of Health | Wadsworth Center, New York State Department.of Health |
| EPI_ISL_426303 | Wadsworth Center, New York State Department.of Health | Wadsworth Center, New York State Department.of Health |
| EPI_ISL_426304 | Wadsworth Center, New York State Department.of Health | Wadsworth Center, New York State Department.of Health |
| EPI_ISL_426305 | Wadsworth Center, New York State Department.of Health | Wadsworth Center, New York State Department.of Health |
| EPI_ISL_426306 | Wadsworth Center, New York State Department.of Health | Wadsworth Center, New York State Department.of Health |
| EPI_ISL_426307 | Wadsworth Center, New York State Department.of Health | Wadsworth Center, New York State Department.of Health |
| EPI_ISL_426308 | Wadsworth Center, New York State Department.of Health | Wadsworth Center, New York State Department.of Health |
| EPI_ISL_426309 | Wadsworth Center, New York State Department.of Health | Wadsworth Center, New York State Department.of Health |
| EPI_ISL_426310 | Wadsworth Center, New York State Department.of Health | Wadsworth Center, New York State Department.of Health |
| EPI_ISL_426311 | Wadsworth Center, New York State Department.of Health | Wadsworth Center, New York State Department.of Health |
| EPI_ISL_426312 | Wadsworth Center, New York State Department.of Health | Wadsworth Center, New York State Department.of Health |
| EPI_ISL_426313 | Wadsworth Center, New York State Department.of Health | Wadsworth Center, New York State Department.of Health |
| EPI_ISL_426314 | Wadsworth Center, New York State Department.of Health | Wadsworth Center, New York State Department.of Health |
| EPI_ISL_426315 | Wadsworth Center, New York State Department.of Health | Wadsworth Center, New York State Department.of Health |
| EPI_ISL_426316 | Wadsworth Center, New York State Department.of Health | Wadsworth Center, New York State Department.of Health |

| EPI_Accession_ID | Originating Laboratory | Submitting Laboratory |
|---|---|---|
| EPI_ISL_426317 | Wadsworth Center, New York State Department.of Health | Wadsworth Center, New York State Department.of Health |
| EPI_ISL_426318 | Wadsworth Center, New York State Department.of Health | Wadsworth Center, New York State Department.of Health |
| EPI_ISL_426319 | Wadsworth Center, New York State Department.of Health | Wadsworth Center, New York State Department.of Health |
| EPI_ISL_426320 | Wadsworth Center, New York State Department.of Health | Wadsworth Center, New York State Department.of Health |
| EPI_ISL_426321 | Wadsworth Center, New York State Department.of Health | Wadsworth Center, New York State Department.of Health |
| EPI_ISL_426322 | Wadsworth Center, New York State Department.of Health | Wadsworth Center, New York State Department.of Health |
| EPI_ISL_426323 | Wadsworth Center, New York State Department.of Health | Wadsworth Center, New York State Department.of Health |
| EPI_ISL_426324 | Wadsworth Center, New York State Department.of Health | Wadsworth Center, New York State Department.of Health |
| EPI_ISL_426325 | Wadsworth Center, New York State Department.of Health | Wadsworth Center, New York State Department.of Health |
| EPI_ISL_426326 | Wadsworth Center, New York State Department.of Health | Wadsworth Center, New York State Department.of Health |
| EPI_ISL_426327 | Wadsworth Center, New York State Department.of Health | Wadsworth Center, New York State Department.of Health |
| EPI_ISL_426328 | Wadsworth Center, New York State Department.of Health | Wadsworth Center, New York State Department.of Health |
| EPI_ISL_426356 | Laboratory of Molecular Genetics, 2nd Faculty of Medicine, Charles University in Prague, Prague, Czech Republic | Laboratory of Molecular Genetics, 2nd Faculty of Medicine, Charles University in Prague, Prague, Czech Republic |
| EPI_ISL_426357 | Laboratory of Molecular Genetics, 2nd Faculty of Medicine, Charles University in Prague, Prague, Czech Republic | Laboratory of Molecular Genetics, 2nd Faculty of Medicine, Charles University in Prague, Prague, Czech Republic |
| EPI_ISL_426358 | Laboratory of Molecular Genetics, 2nd Faculty of Medicine, Charles University in Prague, Prague, Czech Republic | Laboratory of Molecular Genetics, 2nd Faculty of Medicine, Charles University in Prague, Prague, Czech Republic |

| EPI_Accession_ID | Originating Laboratory | Submitting Laboratory |
|---|---|---|
| EPI_ISL_426359 | Laboratory of Molecular Genetics, 2nd Faculty of Medicine, Charles University in Prague, Prague, Czech Republic | Laboratory of Molecular Genetics, 2nd Faculty of Medicine, Charles University in Prague, Prague, Czech Republic |
| EPI_ISL_426360 | Laboratory of Molecular Genetics, 2nd Faculty of Medicine, Charles University in Prague, Prague, Czech Republic | Laboratory of Molecular Genetics, 2nd Faculty of Medicine, Charles University in Prague, Prague, Czech Republic |
| EPI_ISL_426361 | Instituto Nacional de Ciencias Medicas y Nutricion Salvador Zubiran | Instituto Nacional de Ciencias Medicas y Nutricion Salvador Zubiran |
| EPI_ISL_426362 | Instituto Nacional de Ciencias Medicas y Nutricion Salvador Zubiran | Instituto Nacional de Ciencias Medicas y Nutricion Salvador Zubiran |
| EPI_ISL_426363 | Instituto Nacional de Ciencias Medicas y Nutricion Salvador Zubiran | Instituto Nacional de Ciencias Medicas y Nutricion Salvador Zubiran |
| EPI_ISL_426364 | Instituto Nacional de Ciencias Medicas y Nutricion Salvador Zubiran | Instituto Nacional de Ciencias Medicas y Nutricion |
| EPI_ISL_426365 | Instituto Nacional de Ciencias Medicas y Nutricion Salvador Zubiran | Instituto Nacional de Ciencias Medicas y Nutricion Salvador Zubiran |
| EPI_ISL_426379 | The National Laboratory of Health, Environment and Food, Maribor, Slovenia | The National Laboratory of Health, Environment and Food, Maribor, Slovenia |
| EPI_ISL_426414 | Sir M P Shah Government Medical College | Gujarat Biotechnology Research Centre |
| EPI_ISL_426415 | Sir M P Shah Government Medical College, Jamnagar | Gujarat Biotechnology Research Centre, Gandhinagar |
| EPI_ISL_426416 | CT-Dr. Katherine A. Kelley State Public Health Lab | Pathogen Discovery, Respiratory Viruses Branch, Division of Viral Diseases, Centers for Disease Control and Prevention |
| EPI_ISL_426417 | GA Department of Public Health Laboratory | Pathogen Discovery, Respiratory Viruses Branch, Division of Viral Diseases, Centers for Disease Control and Prevention |
| EPI_ISL_426420 | HI Dept. of Health, State Laboratories Division | Pathogen Discovery, Respiratory Viruses Branch, Division of Viral Diseases, Centers for Disease Control and Prevention |
| EPI_ISL_426421 | HI Dept. of Health, State Laboratories Division | Pathogen Discovery, Respiratory Viruses Branch, Division of Viral Diseases, Centers for Disease Control and Prevention |

| EPI_Accession_ID | Originating Laboratory | Submitting Laboratory |
|---|---|---|
| EPI_ISL_426422 | IN State Department of Health Laboratory Services | Pathogen Discovery, Respiratory Viruses Branch, Division of Viral Diseases, Centers for Disease Control and Prevention |
| EPI_ISL_426423 | IN State Department of Health Laboratory Services | Pathogen Discovery, Respiratory Viruses Branch, Division of Viral Diseases, Centers for Disease Control and Prevention |
| EPI_ISL_426424 | IN State Department of Health Laboratory Services | Pathogen Discovery, Respiratory Viruses Branch, Division of Viral Diseases, Centers for Disease Control and Prevention |
| EPI_ISL_426425 | MD DOH Laboratories Administration | Pathogen Discovery, Respiratory Viruses Branch, Division of Viral Diseases, Centers for Disease Control and Prevention |
| EPI_ISL_426426 | MN PHL Division, Minnesota Department of Health | Pathogen Discovery, Respiratory Viruses Branch, Division of Viral Diseases, Centers for Disease Control and Prevention |
| EPI_ISL_426427 | MN PHL Division, Minnesota Department of Health | Pathogen Discovery, Respiratory Viruses Branch, Division of Viral Diseases, Centers for Disease Control and Prevention |
| EPI_ISL_426428 | NC State Laboratory of Public Health | Pathogen Discovery, Respiratory Viruses Branch, Division of Viral Diseases, Centers for Disease Control and Prevention |
| EPI_ISL_426429 | NV State Public Health Laboratory | Pathogen Discovery, Respiratory Viruses Branch, Division of Viral Diseases, Centers for Disease Control and Prevention |
| EPI_ISL_426430 | OH Department of Health Laboratory | Pathogen Discovery, Respiratory Viruses Branch, Division of Viral Diseases, Centers for Disease Control and Prevention |
| EPI_ISL_426431 | OH Department of Health Laboratory | Pathogen Discovery, Respiratory Viruses Branch, Division of Viral Diseases, Centers for Disease Control and Prevention |
| EPI_ISL_426432 | PA Department of Health, Bureau of Laboratories | Pathogen Discovery, Respiratory Viruses Branch, Division of Viral Diseases, Centers for Disease Control and Prevention |

| EPI_Accession_ID | Originating Laboratory | Submitting Laboratory |
|---|---|---|
| EPI_ISL_426433 | PA Department of Health, Bureau of Laboratories | Pathogen Discovery, Respiratory Viruses Branch, Division of Viral Diseases, Centers for Disease Control and Prevention |
| EPI_ISL_426434 | PA Department of Health, Bureau of Laboratories | Pathogen Discovery, Respiratory Viruses Branch, Division of Viral Diseases, Centers for Disease Control and Prevention |
| EPI_ISL_426435 | RI State Health Laboratories | Pathogen Discovery, Respiratory Viruses Branch, Division of Viral Diseases, Centers for Disease Control and Prevention |
| EPI_ISL_426436 | WA State Department of Health | Pathogen Discovery, Respiratory Viruses Branch, Division of Viral Diseases, Centers for Disease Control and Prevention |
| EPI_ISL_426437 | WA State Department of Health | Pathogen Discovery, Respiratory Viruses Branch, Division of Viral Diseases, Centers for Disease Control and Prevention |
| EPI_ISL_426438 | WA State Department of Health | Pathogen Discovery, Respiratory Viruses Branch, Division of Viral Diseases, Centers for Disease Control and Prevention |
| EPI_ISL_426439 | WA State Department of Health | Pathogen Discovery, Respiratory Viruses Branch, Division of Viral Diseases, Centers for Disease Control and Prevention |
| EPI_ISL_426440 | WA State Department of Health | Pathogen Discovery, Respiratory Viruses Branch, Division of Viral Diseases, Centers for Disease Control and Prevention |
| EPI_ISL_426441 | WA State Department of Health | Pathogen Discovery, Respiratory Viruses Branch, Division of Viral Diseases, Centers for Disease Control and Prevention |
| EPI_ISL_426442 | WA State Department of Health | Pathogen Discovery, Respiratory Viruses Branch, Division of Viral Diseases, Centers for Disease Control and Prevention |
| EPI_ISL_426443 | WA State Department of Health | Pathogen Discovery, Respiratory Viruses Branch, Division of Viral Diseases, Centers for Disease Control and Prevention |

| EPI_Accession_ID | Originating Laboratory | Submitting Laboratory |
|---|---|---|
| EPI_ISL_426444 | WA State Department of Health | Pathogen Discovery, Respiratory Viruses Branch, Division of Viral Diseases, Centers for Disease Control and Prevention |
| EPI_ISL_426445 | WA State Department of Health | Pathogen Discovery, Respiratory Viruses Branch, Division of Viral Diseases, Centers for Disease Control and Prevention |
| EPI_ISL_426446 | WA State Department of Health | Pathogen Discovery, Respiratory Viruses Branch, Division of Viral Diseases, Centers for Disease Control and Prevention |
| EPI_ISL_426447 | WA State Department of Health | Pathogen Discovery, Respiratory Viruses Branch, Division of Viral Diseases, Centers for Disease Control and Prevention |
| EPI_ISL_426448 | WA State Department of Health | Pathogen Discovery, Respiratory Viruses Branch, Division of Viral Diseases, Centers for Disease Control and Prevention |
| EPI_ISL_426449 | WA State Department of Health | Pathogen Discovery, Respiratory Viruses Branch, Division of Viral Diseases, Centers for Disease Control and Prevention |
| EPI_ISL_426450 | WA State Department of Health | Pathogen Discovery, Respiratory Viruses Branch, Division of Viral Diseases, Centers for Disease Control and Prevention |
| EPI_ISL_426451 | WA State Department of Health | Pathogen Discovery, Respiratory Viruses Branch, Division of Viral Diseases, Centers for Disease Control and Prevention |
| EPI_ISL_426452 | WA State Department of Health | Pathogen Discovery, Respiratory Viruses Branch, Division of Viral Diseases, Centers for Disease Control and Prevention |
| EPI_ISL_426453 | WA State Department of Health | Pathogen Discovery, Respiratory Viruses Branch, Division of Viral Diseases, Centers for Disease Control and Prevention |
| EPI_ISL_426454 | Virginia DCLS | Virginia DCLS |
| EPI_ISL_426455 | Virginia DCLS | Virginia DCLS |
| EPI_ISL_426456 | Virginia DCLS | Virginia DCLS |

| EPI_Accession_ID | Originating Laboratory | Submitting Laboratory |
|---|---|---|
| EPI_ISL_426457 | Virginia DCLS | Virginia DCLS |
| EPI_ISL_426458 | Virginia DCLS | Virginia DCLS |
| EPI_ISL_426459 | Virginia DCLS | Virginia DCLS |
| EPI_ISL_426460 | Virginia DCLS | Virginia DCLS |
| EPI_ISL_426461 | Virginia DCLS | Virginia DCLS |
| EPI_ISL_426462 | Virginia DCLS | Virginia DCLS |
| EPI_ISL_426463 | Virginia DCLS | Virginia DCLS |
| EPI_ISL_426464 | Virginia DCLS | Virginia DCLS |
| EPI_ISL_426465 | Virginia DCLS | Virginia DCLS |
| EPI_ISL_426466 | Virginia DCLS | Virginia DCLS |
| EPI_ISL_426467 | Virginia DCLS | Virginia DCLS |
| EPI_ISL_426468 | Virginia DCLS | Virginia DCLS |
| EPI_ISL_426469 | Virginia DCLS | Virginia DCLS |
| EPI_ISL_426470 | Virginia DCLS | Virginia DCLS |
| EPI_ISL_426471 | Virginia DCLS | Virginia DCLS |
| EPI_ISL_426472 | Virginia DCLS | Virginia DCLS |
| EPI_ISL_426473 | Virginia DCLS | Virginia DCLS |
| EPI_ISL_426474 | Virginia DCLS | Virginia DCLS |
| EPI_ISL_426475 | Virginia DCLS | Virginia DCLS |
| EPI_ISL_426476 | Microbial Genomics Laboratory, Institut Pasteur Montevideo | Microbial Genomics Laboratory, Institut Pasteur Montevideo, Uruguay |
| EPI_ISL_426477 | Microbial Genomics Laboratory, Institut Pasteur Montevideo | Microbial Genomics Laboratory, Institut Pasteur Montevideo, Uruguay |
| EPI_ISL_426479 | Microbial Genomics Laboratory, Institut Pasteur Montevideo | Microbial Genomics Laboratory, Institut Pasteur Montevideo |
| EPI_ISL_426480 | Microbial Genomics Laboratory, Institut Pasteur Montevideo | Microbial Genomics Laboratory, Institut Pasteur Montevideo |
| EPI_ISL_426481 | Microbial Genomics Laboratory, Institut Pasteur Montevideo, Uruguay | Microbial Genomics Laboratory, Institut Pasteur Montevideo |
| EPI_ISL_426482 | Microbial Genomics Laboratory, Institut Pasteur Montevideo, Uruguay | Microbial Genomics Laboratory, Institut Pasteur Montevideo |
| EPI_ISL_426483 | AZ SPHL, Arizona Department of Health Services | TGen North |

| EPI_Accession_ID | Originating Laboratory | Submitting Laboratory |
|---|---|---|
| EPI_ISL_426484 | n/a | Worobey Lab on behalf of the Arizona COVID-19 Genomics Union |
| EPI_ISL_426485 | AZ SPHL, Arizona Department of Health Services | TGen North |
| EPI_ISL_426499 | TGen North | TGen North |
| EPI_ISL_426500 | TGen North | TGen North |
| EPI_ISL_426501 | TGen North | TGen North |
| EPI_ISL_426502 | TGen North | TGen North |
| EPI_ISL_426503 | TGen North | TGen North |
| EPI_ISL_426504 | TGen North | TGen North |
| EPI_ISL_426505 | TGen North | TGen North |
| EPI_ISL_426506 | TGen North | TGen North |
| EPI_ISL_426507 | TGen North | TGen North |
| EPI_ISL_426509 | TGen North | TGen North |
| EPI_ISL_426510 | TGen North | TGen North |
| EPI_ISL_426511 | TGen North | TGen North |
| EPI_ISL_426512 | AZ SPHL, Arizona Department of Health Services | TGen North |
| EPI_ISL_426513 | AZ SPHL, Arizona Department of Health Services | TGen North |
| EPI_ISL_426514 | AZ SPHL, Arizona Department of Health Services | TGen North |
| EPI_ISL_426515 | AZ SPHL, Arizona Department of Health Services | TGen North |
| EPI_ISL_426516 | AZ SPHL, Arizona Department of Health Services | TGen North |
| EPI_ISL_426517 | AZ SPHL, Arizona Department of Health Services | TGen North |
| EPI_ISL_426518 | AZ SPHL, Arizona Department of Health Services | TGen North |
| EPI_ISL_426519 | AZ SPHL, Arizona Department of Health Services | TGen North |
| EPI_ISL_426520 | TGen North | TGen North |
| EPI_ISL_426521 | TGen North | TGen North |
| EPI_ISL_426522 | TGen North | TGen North |
| EPI_ISL_426523 | TGen North | TGen North |
| EPI_ISL_426524 | TGen North | TGen North |
| EPI_ISL_426525 | TGen North | TGen North |
| EPI_ISL_426526 | TGen North | TGen North |
| EPI_ISL_426527 | AZ SPHL, Arizona Department of Health Services | TGen North |
| EPI_ISL_426528 | AZ SPHL, Arizona Department of Health Services | TGen North |
| EPI_ISL_426529 | AZ SPHL, Arizona Department of Health Services | TGen North |

| EPI_Accession_ID | Originating Laboratory | Submitting Laboratory |
|---|---|---|
| EPI_ISL_426530 | AZ SPHL, Arizona Department of Health Services | TGen North |
| EPI_ISL_426531 | AZ SPHL, Arizona Department of Health Services | TGen North |
| EPI_ISL_426532 | TGen North | TGen North |
| EPI_ISL_426533 | TGen North | TGen North |
| EPI_ISL_426534 | TGen North | TGen North |
| EPI_ISL_426535 | TGen North | TGen North |
| EPI_ISL_426536 | TGen North | TGen North |
| EPI_ISL_426537 | AZ SPHL, Arizona Department of Health Services | TGen North |
| EPI_ISL_426538 | AZ SPHL, Arizona Department of Health Services | TGen North |
| EPI_ISL_426539 | AZ SPHL, Arizona Department of Health Services | TGen North |
| EPI_ISL_426540 | AZ SPHL, Arizona Department of Health Services | TGen North |
| EPI_ISL_426541 | AZ SPHL, Arizona Department of Health Services | TGen North |
| EPI_ISL_426542 | AZ SPHL, Arizona Department of Health Services | TGen North |
| EPI_ISL_426543 | AZ SPHL, Arizona Department of Health Services | TGen North |
| EPI_ISL_426544 | AZ SPHL, Arizona Department of Health Services | TGen North |
| EPI_ISL_426545 | AZ SPHL, Arizona Department of Health Services | TGen North |
| EPI_ISL_426546 | AZ SPHL, Arizona Department of Health Services | TGen North |
| EPI_ISL_426547 | AZ SPHL, Arizona Department of Health Services | TGen North |
| EPI_ISL_426548 | AZ SPHL, Arizona Department of Health Services | TGen North |
| EPI_ISL_426549 | AZ SPHL, Arizona Department of Health Services | TGen North |
| EPI_ISL_426550 | AZ SPHL, Arizona Department of Health Services | TGen North |
| EPI_ISL_426551 | AZ SPHL, Arizona Department of Health Services | TGen North |
| EPI_ISL_426552 | AZ SPHL, Arizona Department of Health Services | TGen North |
| EPI_ISL_426553 | AZ SPHL, Arizona Department of Health Services | TGen North |
| EPI_ISL_426554 | AZ SPHL, Arizona Department of Health Services | TGen North |
| EPI_ISL_426555 | AZ SPHL, Arizona Department of Health Services | TGen North |
| EPI_ISL_426556 | TGen North | TGen North |
| EPI_ISL_426557 | TGen North | TGen North |
| EPI_ISL_426558 | AZ SPHL, Arizona Department of Health Services | TGen North |
| EPI_ISL_426559 | AZ SPHL, Arizona Department of Health Services | TGen North |
| EPI_ISL_426560 | AZ SPHL, Arizona Department of Health Services | TGen North |
| EPI_ISL_426561 | AZ SPHL, Arizona Department of Health Services | TGen North |
| EPI_ISL_426562 | AZ SPHL, Arizona Department of Health Services | TGen North |

| EPI_Accession_ID | Originating Laboratory | Submitting Laboratory |
|---|---|---|
| EPI_ISL_426563 | AZ SPHL, Arizona Department of Health Services | TGen North |
| EPI_ISL_426564 | AZ SPHL, Arizona Department of Health Services | TGen North |
| EPI_ISL_426565 | AZ SPHL, Arizona Department of Health Services | TGen North |
| EPI_ISL_426566 | AZ SPHL, Arizona Department of Health Services | TGen North |
| EPI_ISL_426567 | AZ SPHL, Arizona Department of Health Services | TGen North |
| EPI_ISL_426568 | AZ SPHL, Arizona Department of Health Services | TGen North |
| EPI_ISL_426569 | AZ SPHL, Arizona Department of Health Services | TGen North |
| EPI_ISL_426580 | Instituto Sabin | Laboratory of Virology |
| EPI_ISL_426583 | Microbial Genomics Laboratory, Institut Pasteur Monteivdeo | Microbial Genomics Laboratory, Institut Pasteur Montevideo, Uruguay |
| EPI_ISL_426584 | Microbial Genomics Laboratory, Institut Pasteur Montevideo, Uruguay | Microbial Genomics Laboratory, Institut Pasteur Montevideo, Uruguay |
| EPI_ISL_426617 | NYU Langone Health | Departments of Pathology and Medicine, New York University School of Medicine |
| EPI_ISL_426618 | NYU Langone Health | Departments of Pathology and Medicine, New York University School of Medicine |
| EPI_ISL_426619 | NYU Langone Health | Departments of Pathology and Medicine, New York University School of Medicine |
| EPI_ISL_426620 | NYU Langone Health | Departments of Pathology and Medicine, New York University School of Medicine |
| EPI_ISL_426621 | NYU Langone Health | Departments of Pathology and Medicine, New York University School of Medicine |
| EPI_ISL_426622 | NYU Langone Health | Departments of Pathology and Medicine, New York University School of Medicine |
| EPI_ISL_426623 | NYU Langone Health | Departments of Pathology and Medicine, New York University School of Medicine |
| EPI_ISL_426624 | NYU Langone Health | Departments of Pathology and Medicine, New York University School of Medicine |
| EPI_ISL_426625 | NYU Langone Health | Departments of Pathology and Medicine, New York University School of Medicine |
| EPI_ISL_426626 | NYU Langone Health | Departments of Pathology and Medicine, New York University School of Medicine |
| EPI_ISL_426627 | Ochsner Health | BioInfoExperts, LLC |

| EPI_Accession_ID | Originating Laboratory | Submitting Laboratory |
|---|---|---|
| EPI_ISL_426628 | Ochsner Health | BioInfoExperts, LLC |
| EPI_ISL_426629 | TSGH-CP molecular lab | TSGH-CP molecular lab |
| EPI_ISL_426630 | TSGH-CP molecular lab | TSGH-CP molecular lab |
| EPI_ISL_426631 | TSGH-CP molecular lab | TSGH-CP molecular lab |
| EPI_ISL_426632 | TSGH-CP molecular lab | TSGH-CP molecular lab |
| EPI_ISL_426633 | Royal Darwin Hospital Pathology | Microbiological Diagnostic Unit Public Health Laboratory and Victorian Infectious Diseases Reference Laboratory, Doherty Institute |
| EPI_ISL_426634 | Royal Darwin Hospital Pathology | Microbiological Diagnostic Unit Public Health Laboratory and Victorian Infectious Diseases Reference Laboratory, Doherty Institute |
| EPI_ISL_426635 | Royal Darwin Hospital Pathology | Microbiological Diagnostic Unit Public Health Laboratory and Victorian Infectious Diseases Reference Laboratory, Doherty Institute |
| EPI_ISL_426636 | Royal Darwin Hospital Pathology | Microbiological Diagnostic Unit Public Health Laboratory and Victorian Infectious Diseases Reference Laboratory, Doherty Institute |
| EPI_ISL_426637 | Victorian Infectious Diseases Reference Laboratory (VIDRL) | Microbiological Diagnostic Unit Public Health Laboratory and Victorian Infectious Diseases Reference Laboratory, Doherty Institute |
| EPI_ISL_426638 | Victorian Infectious Diseases Reference Laboratory (VIDRL) | Microbiological Diagnostic Unit Public Health Laboratory and Victorian Infectious Diseases Reference Laboratory, Doherty Institute |
| EPI_ISL_426639 | Victorian Infectious Diseases Reference Laboratory (VIDRL) | Microbiological Diagnostic Unit Public Health Laboratory and Victorian Infectious Diseases Reference Laboratory, Doherty Institute |
| EPI_ISL_426640 | Victorian Infectious Diseases Reference Laboratory (VIDRL) | Microbiological Diagnostic Unit Public Health Laboratory and Victorian Infectious Diseases Reference Laboratory, Doherty Institute |
| EPI_ISL_426641 | Victorian Infectious Diseases Reference Laboratory (VIDRL) | Microbiological Diagnostic Unit Public Health Laboratory and Victorian Infectious Diseases Reference Laboratory, Doherty Institute |

| EPI_Accession_ID | Originating Laboratory | Submitting Laboratory |
|---|---|---|
| EPI_ISL_426642 | Victorian Infectious Diseases Reference Laboratory (VIDRL) | Microbiological Diagnostic Unit Public Health Laboratory and Victorian Infectious Diseases Reference Laboratory, Doherty Institute |
| EPI_ISL_426643 | Victorian Infectious Diseases Reference Laboratory (VIDRL) | Microbiological Diagnostic Unit Public Health Laboratory and Victorian Infectious Diseases Reference Laboratory, Doherty Institute |
| EPI_ISL_426644 | Victorian Infectious Diseases Reference Laboratory (VIDRL) | Microbiological Diagnostic Unit Public Health Laboratory and Victorian Infectious Diseases Reference Laboratory, Doherty Institute |
| EPI_ISL_426645 | Victorian Infectious Diseases Reference Laboratory (VIDRL) | Microbiological Diagnostic Unit Public Health Laboratory and Victorian Infectious Diseases Reference Laboratory, Doherty Institute |
| EPI_ISL_426646 | Victorian Infectious Diseases Reference Laboratory (VIDRL) | Microbiological Diagnostic Unit Public Health Laboratory and Victorian Infectious Diseases Reference Laboratory, Doherty Institute |
| EPI_ISL_426647 | Victorian Infectious Diseases Reference Laboratory (VIDRL) | Microbiological Diagnostic Unit Public Health Laboratory and Victorian Infectious Diseases Reference Laboratory, Doherty Institute |
| EPI_ISL_426648 | Victorian Infectious Diseases Reference Laboratory (VIDRL) | Microbiological Diagnostic Unit Public Health Laboratory and Victorian Infectious Diseases Reference Laboratory, Doherty Institute |
| EPI_ISL_426649 | Victorian Infectious Diseases Reference Laboratory (VIDRL) | Microbiological Diagnostic Unit Public Health Laboratory and Victorian Infectious Diseases Reference Laboratory, Doherty Institute |
| EPI_ISL_426650 | Victorian Infectious Diseases Reference Laboratory (VIDRL) | Microbiological Diagnostic Unit Public Health Laboratory and Victorian Infectious Diseases Reference Laboratory, Doherty Institute |
| EPI_ISL_426651 | Victorian Infectious Diseases Reference Laboratory (VIDRL) | Microbiological Diagnostic Unit Public Health Laboratory and Victorian Infectious Diseases Reference Laboratory, Doherty Institute |
| EPI_ISL_426652 | Victorian Infectious Diseases Reference Laboratory (VIDRL) | Microbiological Diagnostic Unit Public Health Laboratory and Victorian Infectious Diseases Reference Laboratory, Doherty Institute |

| EPI_Accession_ID | Originating Laboratory | Submitting Laboratory |
|---|---|---|
| EPI_ISL_426653 | Victorian Infectious Diseases Reference Laboratory (VIDRL) | Microbiological Diagnostic Unit Public Health Laboratory and Victorian Infectious Diseases Reference Laboratory, Doherty Institute |
| EPI_ISL_426654 | Victorian Infectious Diseases Reference Laboratory (VIDRL) | Microbiological Diagnostic Unit Public Health Laboratory and Victorian Infectious Diseases Reference Laboratory, Doherty Institute |
| EPI_ISL_426655 | Victorian Infectious Diseases Reference Laboratory (VIDRL) | Microbiological Diagnostic Unit Public Health Laboratory and Victorian Infectious Diseases Reference Laboratory, Doherty Institute |
| EPI_ISL_426656 | Victorian Infectious Diseases Reference Laboratory (VIDRL) | Microbiological Diagnostic Unit Public Health Laboratory and Victorian Infectious Diseases Reference Laboratory, Doherty Institute |
| EPI_ISL_426657 | Victorian Infectious Diseases Reference Laboratory (VIDRL) | Microbiological Diagnostic Unit Public Health Laboratory and Victorian Infectious Diseases Reference Laboratory, Doherty Institute |
| EPI_ISL_426658 | Victorian Infectious Diseases Reference Laboratory (VIDRL) | Microbiological Diagnostic Unit Public Health Laboratory and Victorian Infectious Diseases Reference Laboratory, Doherty Institute |
| EPI_ISL_426659 | Victorian Infectious Diseases Reference Laboratory (VIDRL) | Microbiological Diagnostic Unit Public Health Laboratory and Victorian Infectious Diseases Reference Laboratory, Doherty Institute |
| EPI_ISL_426660 | Victorian Infectious Diseases Reference Laboratory (VIDRL) | Microbiological Diagnostic Unit Public Health Laboratory and Victorian Infectious Diseases Reference Laboratory, Doherty Institute |
| EPI_ISL_426661 | Victorian Infectious Diseases Reference Laboratory (VIDRL) | Microbiological Diagnostic Unit Public Health Laboratory and Victorian Infectious Diseases Reference Laboratory, Doherty Institute |
| EPI_ISL_426662 | Victorian Infectious Diseases Reference Laboratory (VIDRL) | Microbiological Diagnostic Unit Public Health Laboratory and Victorian Infectious Diseases Reference Laboratory, Doherty Institute |
| EPI_ISL_426663 | Victorian Infectious Diseases Reference Laboratory (VIDRL) | Microbiological Diagnostic Unit Public Health Laboratory and Victorian Infectious Diseases Reference Laboratory, Doherty Institute |

| EPI_Accession_ID | Originating Laboratory | Submitting Laboratory |
|---|---|---|
| EPI_ISL_426664 | Victorian Infectious Diseases Reference Laboratory (VIDRL) | Microbiological Diagnostic Unit Public Health Laboratory and Victorian Infectious Diseases Reference Laboratory, Doherty Institute |
| EPI_ISL_426665 | Victorian Infectious Diseases Reference Laboratory (VIDRL) | Microbiological Diagnostic Unit Public Health Laboratory and Victorian Infectious Diseases Reference Laboratory, Doherty Institute |
| EPI_ISL_426666 | Victorian Infectious Diseases Reference Laboratory (VIDRL) | Microbiological Diagnostic Unit Public Health Laboratory and Victorian Infectious Diseases Reference Laboratory, Doherty Institute |
| EPI_ISL_426667 | Victorian Infectious Diseases Reference Laboratory (VIDRL) | Microbiological Diagnostic Unit Public Health Laboratory and Victorian Infectious Diseases Reference Laboratory, Doherty Institute |
| EPI_ISL_426668 | Victorian Infectious Diseases Reference Laboratory (VIDRL) | Microbiological Diagnostic Unit Public Health Laboratory and Victorian Infectious Diseases Reference Laboratory, Doherty Institute |
| EPI_ISL_426669 | Victorian Infectious Diseases Reference Laboratory (VIDRL) | Microbiological Diagnostic Unit Public Health Laboratory and Victorian Infectious Diseases Reference Laboratory, Doherty Institute |
| EPI_ISL_426670 | Victorian Infectious Diseases Reference Laboratory (VIDRL) | Microbiological Diagnostic Unit Public Health Laboratory and Victorian Infectious Diseases Reference Laboratory, Doherty Institute |
| EPI_ISL_426671 | Victorian Infectious Diseases Reference Laboratory (VIDRL) | Microbiological Diagnostic Unit Public Health Laboratory and Victorian Infectious Diseases Reference Laboratory, Doherty Institute |
| EPI_ISL_426672 | Victorian Infectious Diseases Reference Laboratory (VIDRL) | Microbiological Diagnostic Unit Public Health Laboratory and Victorian Infectious Diseases Reference Laboratory, Doherty Institute |
| EPI_ISL_426673 | Victorian Infectious Diseases Reference Laboratory (VIDRL) | Microbiological Diagnostic Unit Public Health Laboratory and Victorian Infectious Diseases Reference Laboratory, Doherty Institute |
| EPI_ISL_426674 | Victorian Infectious Diseases Reference Laboratory (VIDRL) | Microbiological Diagnostic Unit Public Health Laboratory and Victorian Infectious Diseases Reference Laboratory, Doherty Institute |

| EPI_Accession_ID | Originating Laboratory | Submitting Laboratory |
|---|---|---|
| EPI_ISL_426675 | Victorian Infectious Diseases Reference Laboratory (VIDRL) | Microbiological Diagnostic Unit Public Health Laboratory and Victorian Infectious Diseases Reference Laboratory, Doherty Institute |
| EPI_ISL_426676 | Victorian Infectious Diseases Reference Laboratory (VIDRL) | Microbiological Diagnostic Unit Public Health Laboratory and Victorian Infectious Diseases Reference Laboratory, Doherty Institute |
| EPI_ISL_426677 | Victorian Infectious Diseases Reference Laboratory (VIDRL) | Microbiological Diagnostic Unit Public Health Laboratory and Victorian Infectious Diseases Reference Laboratory, Doherty Institute |
| EPI_ISL_426678 | Victorian Infectious Diseases Reference Laboratory (VIDRL) | Microbiological Diagnostic Unit Public Health Laboratory and Victorian Infectious Diseases Reference Laboratory, Doherty Institute |
| EPI_ISL_426679 | Victorian Infectious Diseases Reference Laboratory (VIDRL) | Microbiological Diagnostic Unit Public Health Laboratory and Victorian Infectious Diseases Reference Laboratory, Doherty Institute |
| EPI_ISL_426680 | Victorian Infectious Diseases Reference Laboratory (VIDRL) | Microbiological Diagnostic Unit Public Health Laboratory and Victorian Infectious Diseases Reference Laboratory, Doherty Institute |
| EPI_ISL_426681 | Victorian Infectious Diseases Reference Laboratory (VIDRL) | Microbiological Diagnostic Unit Public Health Laboratory and Victorian Infectious Diseases Reference Laboratory, Doherty Institute |
| EPI_ISL_426682 | Victorian Infectious Diseases Reference Laboratory (VIDRL) | Microbiological Diagnostic Unit Public Health Laboratory and Victorian Infectious Diseases Reference Laboratory, Doherty Institute |
| EPI_ISL_426683 | Victorian Infectious Diseases Reference Laboratory (VIDRL) | Microbiological Diagnostic Unit Public Health Laboratory and Victorian Infectious Diseases Reference Laboratory, Doherty Institute |
| EPI_ISL_426684 | Victorian Infectious Diseases Reference Laboratory (VIDRL) | Microbiological Diagnostic Unit Public Health Laboratory and Victorian Infectious Diseases Reference Laboratory, Doherty Institute |
| EPI_ISL_426685 | Victorian Infectious Diseases Reference Laboratory (VIDRL) | Microbiological Diagnostic Unit Public Health Laboratory and Victorian Infectious Diseases Reference Laboratory, Doherty Institute |

| EPI_Accession_ID | Originating Laboratory | Submitting Laboratory |
|---|---|---|
| EPI_ISL_426686 | Victorian Infectious Diseases Reference Laboratory (VIDRL) | Microbiological Diagnostic Unit Public Health Laboratory and Victorian Infectious Diseases Reference Laboratory, Doherty Institute |
| EPI_ISL_426687 | Victorian Infectious Diseases Reference Laboratory (VIDRL) | Microbiological Diagnostic Unit Public Health Laboratory and Victorian Infectious Diseases Reference Laboratory, Doherty Institute |
| EPI_ISL_426688 | Victorian Infectious Diseases Reference Laboratory (VIDRL) | Microbiological Diagnostic Unit Public Health Laboratory and Victorian Infectious Diseases Reference Laboratory, Doherty Institute |
| EPI_ISL_426689 | Victorian Infectious Diseases Reference Laboratory (VIDRL) | Microbiological Diagnostic Unit Public Health Laboratory and Victorian Infectious Diseases Reference Laboratory, Doherty Institute |
| EPI_ISL_426690 | Victorian Infectious Diseases Reference Laboratory (VIDRL) | Microbiological Diagnostic Unit Public Health Laboratory and Victorian Infectious Diseases Reference Laboratory, Doherty Institute |
| EPI_ISL_426691 | Victorian Infectious Diseases Reference Laboratory (VIDRL) | Microbiological Diagnostic Unit Public Health Laboratory and Victorian Infectious Diseases Reference Laboratory, Doherty Institute |
| EPI_ISL_426692 | Victorian Infectious Diseases Reference Laboratory (VIDRL) | Microbiological Diagnostic Unit Public Health Laboratory and Victorian Infectious Diseases Reference Laboratory, Doherty Institute |
| EPI_ISL_426693 | Victorian Infectious Diseases Reference Laboratory (VIDRL) | Microbiological Diagnostic Unit Public Health Laboratory and Victorian Infectious Diseases Reference Laboratory, Doherty Institute |
| EPI_ISL_426694 | Victorian Infectious Diseases Reference Laboratory (VIDRL) | Microbiological Diagnostic Unit Public Health Laboratory and Victorian Infectious Diseases Reference Laboratory, Doherty Institute |
| EPI_ISL_426695 | Victorian Infectious Diseases Reference Laboratory (VIDRL) | Microbiological Diagnostic Unit Public Health Laboratory and Victorian Infectious Diseases Reference Laboratory, Doherty Institute |
| EPI_ISL_426696 | Victorian Infectious Diseases Reference Laboratory (VIDRL) | Microbiological Diagnostic Unit Public Health Laboratory and Victorian Infectious Diseases Reference Laboratory, Doherty Institute |

| EPI_Accession_ID | Originating Laboratory | Submitting Laboratory |
|---|---|---|
| EPI_ISL_426697 | Victorian Infectious Diseases Reference Laboratory (VIDRL) | Microbiological Diagnostic Unit Public Health Laboratory and Victorian Infectious Diseases Reference Laboratory, Doherty Institute |
| EPI_ISL_426698 | Victorian Infectious Diseases Reference Laboratory (VIDRL) | Microbiological Diagnostic Unit Public Health Laboratory and Victorian Infectious Diseases Reference Laboratory, Doherty Institute |
| EPI_ISL_426699 | Victorian Infectious Diseases Reference Laboratory (VIDRL) | Microbiological Diagnostic Unit Public Health Laboratory and Victorian Infectious Diseases Reference Laboratory, Doherty Institute |
| EPI_ISL_426700 | Victorian Infectious Diseases Reference Laboratory (VIDRL) | Microbiological Diagnostic Unit Public Health Laboratory and Victorian Infectious Diseases Reference Laboratory, Doherty Institute |
| EPI_ISL_426701 | Victorian Infectious Diseases Reference Laboratory (VIDRL) | Microbiological Diagnostic Unit Public Health Laboratory and Victorian Infectious Diseases Reference Laboratory, Doherty Institute |
| EPI_ISL_426702 | Victorian Infectious Diseases Reference Laboratory (VIDRL) | Microbiological Diagnostic Unit Public Health Laboratory and Victorian Infectious Diseases Reference Laboratory, Doherty Institute |
| EPI_ISL_426703 | Victorian Infectious Diseases Reference Laboratory (VIDRL) | Microbiological Diagnostic Unit Public Health Laboratory and Victorian Infectious Diseases Reference Laboratory, Doherty Institute |
| EPI_ISL_426704 | Victorian Infectious Diseases Reference Laboratory (VIDRL) | Microbiological Diagnostic Unit Public Health Laboratory and Victorian Infectious Diseases Reference Laboratory, Doherty Institute |
| EPI_ISL_426705 | Victorian Infectious Diseases Reference Laboratory (VIDRL) | Microbiological Diagnostic Unit Public Health Laboratory and Victorian Infectious Diseases Reference Laboratory, Doherty Institute |
| EPI_ISL_426706 | Victorian Infectious Diseases Reference Laboratory (VIDRL) | Microbiological Diagnostic Unit Public Health Laboratory and Victorian Infectious Diseases Reference Laboratory, Doherty Institute |
| EPI_ISL_426707 | Victorian Infectious Diseases Reference Laboratory (VIDRL) | Microbiological Diagnostic Unit Public Health Laboratory and Victorian Infectious Diseases Reference Laboratory, Doherty Institute |

| EPI_Accession_ID | Originating Laboratory | Submitting Laboratory |
|---|---|---|
| EPI_ISL_426708 | Victorian Infectious Diseases Reference Laboratory (VIDRL) | Microbiological Diagnostic Unit Public Health Laboratory and Victorian Infectious Diseases Reference Laboratory, Doherty Institute |
| EPI_ISL_426709 | Victorian Infectious Diseases Reference Laboratory (VIDRL) | Microbiological Diagnostic Unit Public Health Laboratory and Victorian Infectious Diseases Reference Laboratory, Doherty Institute |
| EPI_ISL_426710 | Victorian Infectious Diseases Reference Laboratory (VIDRL) | Microbiological Diagnostic Unit Public Health Laboratory and Victorian Infectious Diseases Reference Laboratory, Doherty Institute |
| EPI_ISL_426711 | Victorian Infectious Diseases Reference Laboratory (VIDRL) | Microbiological Diagnostic Unit Public Health Laboratory and Victorian Infectious Diseases Reference Laboratory, Doherty Institute |
| EPI_ISL_426712 | Victorian Infectious Diseases Reference Laboratory (VIDRL) | Microbiological Diagnostic Unit Public Health Laboratory and Victorian Infectious Diseases Reference Laboratory, Doherty Institute |
| EPI_ISL_426713 | Victorian Infectious Diseases Reference Laboratory (VIDRL) | Microbiological Diagnostic Unit Public Health Laboratory and Victorian Infectious Diseases Reference Laboratory, Doherty Institute |
| EPI_ISL_426714 | Victorian Infectious Diseases Reference Laboratory (VIDRL) | Microbiological Diagnostic Unit Public Health Laboratory and Victorian Infectious Diseases Reference Laboratory, Doherty Institute |
| EPI_ISL_426715 | Victorian Infectious Diseases Reference Laboratory (VIDRL) | Microbiological Diagnostic Unit Public Health Laboratory and Victorian Infectious Diseases Reference Laboratory, Doherty Institute |
| EPI_ISL_426716 | Victorian Infectious Diseases Reference Laboratory (VIDRL) | Microbiological Diagnostic Unit Public Health Laboratory and Victorian Infectious Diseases Reference Laboratory, Doherty Institute |
| EPI_ISL_426717 | Victorian Infectious Diseases Reference Laboratory (VIDRL) | Microbiological Diagnostic Unit Public Health Laboratory and Victorian Infectious Diseases Reference Laboratory, Doherty Institute |
| EPI_ISL_426718 | Victorian Infectious Diseases Reference Laboratory (VIDRL) | Microbiological Diagnostic Unit Public Health Laboratory and Victorian Infectious Diseases Reference Laboratory, Doherty Institute |

| EPI_Accession_ID | Originating Laboratory | Submitting Laboratory |
|---|---|---|
| EPI_ISL_426719 | Victorian Infectious Diseases Reference Laboratory (VIDRL) | Microbiological Diagnostic Unit Public Health Laboratory and Victorian Infectious Diseases Reference Laboratory, Doherty Institute |
| EPI_ISL_426720 | Victorian Infectious Diseases Reference Laboratory (VIDRL) | Microbiological Diagnostic Unit Public Health Laboratory and Victorian Infectious Diseases Reference Laboratory, Doherty Institute |
| EPI_ISL_426721 | Victorian Infectious Diseases Reference Laboratory (VIDRL) | Microbiological Diagnostic Unit Public Health Laboratory and Victorian Infectious Diseases Reference Laboratory, Doherty Institute |
| EPI_ISL_426722 | Victorian Infectious Diseases Reference Laboratory (VIDRL) | Microbiological Diagnostic Unit Public Health Laboratory and Victorian Infectious Diseases Reference Laboratory, Doherty Institute |
| EPI_ISL_426723 | Victorian Infectious Diseases Reference Laboratory (VIDRL) | Microbiological Diagnostic Unit Public Health Laboratory and Victorian Infectious Diseases Reference Laboratory, Doherty Institute |
| EPI_ISL_426724 | Victorian Infectious Diseases Reference Laboratory (VIDRL) | Microbiological Diagnostic Unit Public Health Laboratory and Victorian Infectious Diseases Reference Laboratory, Doherty Institute |
| EPI_ISL_426725 | Victorian Infectious Diseases Reference Laboratory (VIDRL) | Microbiological Diagnostic Unit Public Health Laboratory and Victorian Infectious Diseases Reference Laboratory, Doherty Institute |
| EPI_ISL_426726 | Victorian Infectious Diseases Reference Laboratory (VIDRL) | Microbiological Diagnostic Unit Public Health Laboratory and Victorian Infectious Diseases Reference Laboratory, Doherty Institute |
| EPI_ISL_426727 | Victorian Infectious Diseases Reference Laboratory (VIDRL) | Microbiological Diagnostic Unit Public Health Laboratory and Victorian Infectious Diseases Reference Laboratory, Doherty Institute |
| EPI_ISL_426728 | Victorian Infectious Diseases Reference Laboratory (VIDRL) | Microbiological Diagnostic Unit Public Health Laboratory and Victorian Infectious Diseases Reference Laboratory, Doherty Institute |
| EPI_ISL_426729 | Victorian Infectious Diseases Reference Laboratory (VIDRL) | Microbiological Diagnostic Unit Public Health Laboratory and Victorian Infectious Diseases Reference Laboratory, Doherty Institute |

| EPI_Accession_ID | Originating Laboratory | Submitting Laboratory |
|---|---|---|
| EPI_ISL_426730 | Victorian Infectious Diseases Reference Laboratory (VIDRL) | Microbiological Diagnostic Unit Public Health Laboratory and Victorian Infectious Diseases Reference Laboratory, Doherty Institute |
| EPI_ISL_426731 | Victorian Infectious Diseases Reference Laboratory (VIDRL) | Microbiological Diagnostic Unit Public Health Laboratory and Victorian Infectious Diseases Reference Laboratory, Doherty Institute |
| EPI_ISL_426732 | Victorian Infectious Diseases Reference Laboratory (VIDRL) | Microbiological Diagnostic Unit Public Health Laboratory and Victorian Infectious Diseases Reference Laboratory, Doherty Institute |
| EPI_ISL_426733 | Victorian Infectious Diseases Reference Laboratory (VIDRL) | Microbiological Diagnostic Unit Public Health Laboratory and Victorian Infectious Diseases Reference Laboratory, Doherty Institute |
| EPI_ISL_426734 | Victorian Infectious Diseases Reference Laboratory (VIDRL) | Microbiological Diagnostic Unit Public Health Laboratory and Victorian Infectious Diseases Reference Laboratory, Doherty Institute |
| EPI_ISL_426735 | Victorian Infectious Diseases Reference Laboratory (VIDRL) | Microbiological Diagnostic Unit Public Health Laboratory and Victorian Infectious Diseases Reference Laboratory, Doherty Institute |
| EPI_ISL_426736 | Victorian Infectious Diseases Reference Laboratory (VIDRL) | Microbiological Diagnostic Unit Public Health Laboratory and Victorian Infectious Diseases Reference Laboratory, Doherty Institute |
| EPI_ISL_426737 | Victorian Infectious Diseases Reference Laboratory (VIDRL) | Microbiological Diagnostic Unit Public Health Laboratory and Victorian Infectious Diseases Reference Laboratory, Doherty Institute |
| EPI_ISL_426738 | Victorian Infectious Diseases Reference Laboratory (VIDRL) | Microbiological Diagnostic Unit Public Health Laboratory and Victorian Infectious Diseases Reference Laboratory, Doherty Institute |
| EPI_ISL_426739 | Victorian Infectious Diseases Reference Laboratory (VIDRL) | Microbiological Diagnostic Unit Public Health Laboratory and Victorian Infectious Diseases Reference Laboratory, Doherty Institute |
| EPI_ISL_426740 | Victorian Infectious Diseases Reference Laboratory (VIDRL) | Microbiological Diagnostic Unit Public Health Laboratory and Victorian Infectious Diseases Reference Laboratory, Doherty Institute |

| EPI_Accession_ID | Originating Laboratory | Submitting Laboratory |
|---|---|---|
| EPI_ISL_426741 | Victorian Infectious Diseases Reference Laboratory (VIDRL) | Microbiological Diagnostic Unit Public Health Laboratory and Victorian Infectious Diseases Reference Laboratory, Doherty Institute |
| EPI_ISL_426743 | Victorian Infectious Diseases Reference Laboratory (VIDRL) | Microbiological Diagnostic Unit Public Health Laboratory and Victorian Infectious Diseases Reference Laboratory, Doherty Institute |
| EPI_ISL_426744 | Victorian Infectious Diseases Reference Laboratory (VIDRL) | Microbiological Diagnostic Unit Public Health Laboratory and Victorian Infectious Diseases Reference Laboratory, Doherty Institute |
| EPI_ISL_426745 | Victorian Infectious Diseases Reference Laboratory (VIDRL) | Microbiological Diagnostic Unit Public Health Laboratory and Victorian Infectious Diseases Reference Laboratory, Doherty Institute |
| EPI_ISL_426746 | Victorian Infectious Diseases Reference Laboratory (VIDRL) | Microbiological Diagnostic Unit Public Health Laboratory and Victorian Infectious Diseases Reference Laboratory, Doherty Institute |
| EPI_ISL_426747 | Victorian Infectious Diseases Reference Laboratory (VIDRL) | Microbiological Diagnostic Unit Public Health Laboratory and Victorian Infectious Diseases Reference Laboratory, Doherty Institute |
| EPI_ISL_426748 | Victorian Infectious Diseases Reference Laboratory (VIDRL) | Microbiological Diagnostic Unit Public Health Laboratory and Victorian Infectious Diseases Reference Laboratory, Doherty Institute |
| EPI_ISL_426749 | Victorian Infectious Diseases Reference Laboratory (VIDRL) | Microbiological Diagnostic Unit Public Health Laboratory and Victorian Infectious Diseases Reference Laboratory, Doherty Institute |
| EPI_ISL_426750 | Victorian Infectious Diseases Reference Laboratory (VIDRL) | Microbiological Diagnostic Unit Public Health Laboratory and Victorian Infectious Diseases Reference Laboratory, Doherty Institute |
| EPI_ISL_426751 | Victorian Infectious Diseases Reference Laboratory (VIDRL) | Microbiological Diagnostic Unit Public Health Laboratory and Victorian Infectious Diseases Reference Laboratory, Doherty Institute |
| EPI_ISL_426752 | Victorian Infectious Diseases Reference Laboratory (VIDRL) | Microbiological Diagnostic Unit Public Health Laboratory and Victorian Infectious Diseases Reference Laboratory, Doherty Institute |

| EPI_Accession_ID | Originating Laboratory | Submitting Laboratory |
|---|---|---|
| EPI_ISL_426753 | Victorian Infectious Diseases Reference Laboratory (VIDRL) | Microbiological Diagnostic Unit Public Health Laboratory and Victorian Infectious Diseases Reference Laboratory, Doherty Institute |
| EPI_ISL_426754 | Victorian Infectious Diseases Reference Laboratory (VIDRL) | Microbiological Diagnostic Unit Public Health Laboratory and Victorian Infectious Diseases Reference Laboratory, Doherty Institute |
| EPI_ISL_426755 | Victorian Infectious Diseases Reference Laboratory (VIDRL) | Microbiological Diagnostic Unit Public Health Laboratory and Victorian Infectious Diseases Reference Laboratory, Doherty Institute |
| EPI_ISL_426756 | Victorian Infectious Diseases Reference Laboratory (VIDRL) | Microbiological Diagnostic Unit Public Health Laboratory and Victorian Infectious Diseases Reference Laboratory, Doherty Institute |
| EPI_ISL_426757 | Victorian Infectious Diseases Reference Laboratory (VIDRL) | Microbiological Diagnostic Unit Public Health Laboratory and Victorian Infectious Diseases Reference Laboratory, Doherty Institute |
| EPI_ISL_426758 | Victorian Infectious Diseases Reference Laboratory (VIDRL) | Microbiological Diagnostic Unit Public Health Laboratory and Victorian Infectious Diseases Reference Laboratory, Doherty Institute |
| EPI_ISL_426759 | Victorian Infectious Diseases Reference Laboratory (VIDRL) | Microbiological Diagnostic Unit Public Health Laboratory and Victorian Infectious Diseases Reference Laboratory, Doherty Institute |
| EPI_ISL_426760 | Victorian Infectious Diseases Reference Laboratory (VIDRL) | Microbiological Diagnostic Unit Public Health Laboratory and Victorian Infectious Diseases Reference Laboratory, Doherty Institute |
| EPI_ISL_426761 | Victorian Infectious Diseases Reference Laboratory (VIDRL) | Microbiological Diagnostic Unit Public Health Laboratory and Victorian Infectious Diseases Reference Laboratory, Doherty Institute |
| EPI_ISL_426762 | Victorian Infectious Diseases Reference Laboratory (VIDRL) | Microbiological Diagnostic Unit Public Health Laboratory and Victorian Infectious Diseases Reference Laboratory, Doherty Institute |
| EPI_ISL_426763 | Victorian Infectious Diseases Reference Laboratory (VIDRL) | Microbiological Diagnostic Unit Public Health Laboratory and Victorian Infectious Diseases Reference Laboratory, Doherty Institute |

| EPI_Accession_ID | Originating Laboratory | Submitting Laboratory |
|---|---|---|
| EPI_ISL_426764 | Victorian Infectious Diseases Reference Laboratory (VIDRL) | Microbiological Diagnostic Unit Public Health Laboratory and Victorian Infectious Diseases Reference Laboratory, Doherty Institute |
| EPI_ISL_426765 | Victorian Infectious Diseases Reference Laboratory (VIDRL) | Microbiological Diagnostic Unit Public Health Laboratory and Victorian Infectious Diseases Reference Laboratory, Doherty Institute |
| EPI_ISL_426766 | Victorian Infectious Diseases Reference Laboratory (VIDRL) | Microbiological Diagnostic Unit Public Health Laboratory and Victorian Infectious Diseases Reference Laboratory, Doherty Institute |
| EPI_ISL_426767 | Victorian Infectious Diseases Reference Laboratory (VIDRL) | Microbiological Diagnostic Unit Public Health Laboratory and Victorian Infectious Diseases Reference Laboratory, Doherty Institute |
| EPI_ISL_426768 | Victorian Infectious Diseases Reference Laboratory (VIDRL) | Microbiological Diagnostic Unit Public Health Laboratory and Victorian Infectious Diseases Reference Laboratory, Doherty Institute |
| EPI_ISL_426769 | Victorian Infectious Diseases Reference Laboratory (VIDRL) | Microbiological Diagnostic Unit Public Health Laboratory and Victorian Infectious Diseases Reference Laboratory, Doherty Institute |
| EPI_ISL_426770 | Victorian Infectious Diseases Reference Laboratory (VIDRL) | Microbiological Diagnostic Unit Public Health Laboratory and Victorian Infectious Diseases Reference Laboratory, Doherty Institute |
| EPI_ISL_426771 | Victorian Infectious Diseases Reference Laboratory (VIDRL) | Microbiological Diagnostic Unit Public Health Laboratory and Victorian Infectious Diseases Reference Laboratory, Doherty Institute |
| EPI_ISL_426772 | Victorian Infectious Diseases Reference Laboratory (VIDRL) | Microbiological Diagnostic Unit Public Health Laboratory and Victorian Infectious Diseases Reference Laboratory, Doherty Institute |
| EPI_ISL_426773 | Victorian Infectious Diseases Reference Laboratory (VIDRL) | Microbiological Diagnostic Unit Public Health Laboratory and Victorian Infectious Diseases Reference Laboratory, Doherty Institute |
| EPI_ISL_426774 | Victorian Infectious Diseases Reference Laboratory (VIDRL) | Microbiological Diagnostic Unit Public Health Laboratory and Victorian Infectious Diseases Reference Laboratory, Doherty Institute |

| EPI_Accession_ID | Originating Laboratory | Submitting Laboratory |
|---|---|---|
| EPI_ISL_426775 | Victorian Infectious Diseases Reference Laboratory (VIDRL) | Microbiological Diagnostic Unit Public Health Laboratory and Victorian Infectious Diseases Reference Laboratory, Doherty Institute |
| EPI_ISL_426776 | Victorian Infectious Diseases Reference Laboratory (VIDRL) | Microbiological Diagnostic Unit Public Health Laboratory and Victorian Infectious Diseases Reference Laboratory, Doherty Institute |
| EPI_ISL_426777 | Victorian Infectious Diseases Reference Laboratory (VIDRL) | Microbiological Diagnostic Unit Public Health Laboratory and Victorian Infectious Diseases Reference Laboratory, Doherty Institute |
| EPI_ISL_426778 | Victorian Infectious Diseases Reference Laboratory (VIDRL) | Microbiological Diagnostic Unit Public Health Laboratory and Victorian Infectious Diseases Reference Laboratory, Doherty Institute |
| EPI_ISL_426779 | Victorian Infectious Diseases Reference Laboratory (VIDRL) | Microbiological Diagnostic Unit Public Health Laboratory and Victorian Infectious Diseases Reference Laboratory, Doherty Institute |
| EPI_ISL_426780 | Victorian Infectious Diseases Reference Laboratory (VIDRL) | Microbiological Diagnostic Unit Public Health Laboratory and Victorian Infectious Diseases Reference Laboratory, Doherty Institute |
| EPI_ISL_426781 | Victorian Infectious Diseases Reference Laboratory (VIDRL) | Microbiological Diagnostic Unit Public Health Laboratory and Victorian Infectious Diseases Reference Laboratory, Doherty Institute |
| EPI_ISL_426782 | Victorian Infectious Diseases Reference Laboratory (VIDRL) | Microbiological Diagnostic Unit Public Health Laboratory and Victorian Infectious Diseases Reference Laboratory, Doherty Institute |
| EPI_ISL_426783 | Victorian Infectious Diseases Reference Laboratory (VIDRL) | Microbiological Diagnostic Unit Public Health Laboratory and Victorian Infectious Diseases Reference Laboratory, Doherty Institute |
| EPI_ISL_426784 | Victorian Infectious Diseases Reference Laboratory (VIDRL) | Microbiological Diagnostic Unit Public Health Laboratory and Victorian Infectious Diseases Reference Laboratory, Doherty Institute |
| EPI_ISL_426785 | Victorian Infectious Diseases Reference Laboratory (VIDRL) | Microbiological Diagnostic Unit Public Health Laboratory and Victorian Infectious Diseases Reference Laboratory, Doherty Institute |

| EPI_Accession_ID | Originating Laboratory | Submitting Laboratory |
|---|---|---|
| EPI_ISL_426786 | Victorian Infectious Diseases Reference Laboratory (VIDRL) | Microbiological Diagnostic Unit Public Health Laboratory and Victorian Infectious Diseases Reference Laboratory, Doherty Institute |
| EPI_ISL_426787 | Victorian Infectious Diseases Reference Laboratory (VIDRL) | Microbiological Diagnostic Unit Public Health Laboratory and Victorian Infectious Diseases Reference Laboratory, Doherty Institute |
| EPI_ISL_426788 | Victorian Infectious Diseases Reference Laboratory (VIDRL) | Microbiological Diagnostic Unit Public Health Laboratory and Victorian Infectious Diseases Reference Laboratory, Doherty Institute |
| EPI_ISL_426789 | Victorian Infectious Diseases Reference Laboratory (VIDRL) | Microbiological Diagnostic Unit Public Health Laboratory and Victorian Infectious Diseases Reference Laboratory, Doherty Institute |
| EPI_ISL_426790 | Victorian Infectious Diseases Reference Laboratory (VIDRL) | Microbiological Diagnostic Unit Public Health Laboratory and Victorian Infectious Diseases Reference Laboratory, Doherty Institute |
| EPI_ISL_426791 | Victorian Infectious Diseases Reference Laboratory (VIDRL) | Microbiological Diagnostic Unit Public Health Laboratory and Victorian Infectious Diseases Reference Laboratory, Doherty Institute |
| EPI_ISL_426792 | Victorian Infectious Diseases Reference Laboratory (VIDRL) | Microbiological Diagnostic Unit Public Health Laboratory and Victorian Infectious Diseases Reference Laboratory, Doherty Institute |
| EPI_ISL_426793 | Victorian Infectious Diseases Reference Laboratory (VIDRL) | Microbiological Diagnostic Unit Public Health Laboratory and Victorian Infectious Diseases Reference Laboratory, Doherty Institute |
| EPI_ISL_426794 | Victorian Infectious Diseases Reference Laboratory (VIDRL) | Microbiological Diagnostic Unit Public Health Laboratory and Victorian Infectious Diseases Reference Laboratory, Doherty Institute |
| EPI_ISL_426795 | Victorian Infectious Diseases Reference Laboratory (VIDRL) | Microbiological Diagnostic Unit Public Health Laboratory and Victorian Infectious Diseases Reference Laboratory, Doherty Institute |
| EPI_ISL_426796 | Victorian Infectious Diseases Reference Laboratory (VIDRL) | Microbiological Diagnostic Unit Public Health Laboratory and Victorian Infectious Diseases Reference Laboratory, Doherty Institute |

| EPI_Accession_ID | Originating Laboratory | Submitting Laboratory |
|---|---|---|
| EPI_ISL_426797 | Victorian Infectious Diseases Reference Laboratory (VIDRL) | Microbiological Diagnostic Unit Public Health Laboratory and Victorian Infectious Diseases Reference Laboratory, Doherty Institute |
| EPI_ISL_426798 | Victorian Infectious Diseases Reference Laboratory (VIDRL) | Microbiological Diagnostic Unit Public Health Laboratory and Victorian Infectious Diseases Reference Laboratory, Doherty Institute |
| EPI_ISL_426799 | Victorian Infectious Diseases Reference Laboratory (VIDRL) | Microbiological Diagnostic Unit Public Health Laboratory and Victorian Infectious Diseases Reference Laboratory, Doherty Institute |
| EPI_ISL_426800 | Victorian Infectious Diseases Reference Laboratory (VIDRL) | Microbiological Diagnostic Unit Public Health Laboratory and Victorian Infectious Diseases Reference Laboratory, Doherty Institute |
| EPI_ISL_426801 | Victorian Infectious Diseases Reference Laboratory (VIDRL) | Microbiological Diagnostic Unit Public Health Laboratory and Victorian Infectious Diseases Reference Laboratory, Doherty Institute |
| EPI_ISL_426802 | Victorian Infectious Diseases Reference Laboratory (VIDRL) | Microbiological Diagnostic Unit Public Health Laboratory and Victorian Infectious Diseases Reference Laboratory, Doherty Institute |
| EPI_ISL_426803 | Victorian Infectious Diseases Reference Laboratory (VIDRL) | Microbiological Diagnostic Unit Public Health Laboratory and Victorian Infectious Diseases Reference Laboratory, Doherty Institute |
| EPI_ISL_426804 | Victorian Infectious Diseases Reference Laboratory (VIDRL) | Microbiological Diagnostic Unit Public Health Laboratory and Victorian Infectious Diseases Reference Laboratory, Doherty Institute |
| EPI_ISL_426805 | Victorian Infectious Diseases Reference Laboratory (VIDRL) | Microbiological Diagnostic Unit Public Health Laboratory and Victorian Infectious Diseases Reference Laboratory, Doherty Institute |
| EPI_ISL_426806 | Victorian Infectious Diseases Reference Laboratory (VIDRL) | Microbiological Diagnostic Unit Public Health Laboratory and Victorian Infectious Diseases Reference Laboratory, Doherty Institute |
| EPI_ISL_426807 | Victorian Infectious Diseases Reference Laboratory (VIDRL) | Microbiological Diagnostic Unit Public Health Laboratory and Victorian Infectious Diseases Reference Laboratory, Doherty Institute |

| EPI_Accession_ID | Originating Laboratory | Submitting Laboratory |
|---|---|---|
| EPI_ISL_426808 | Victorian Infectious Diseases Reference Laboratory (VIDRL) | Microbiological Diagnostic Unit Public Health Laboratory and Victorian Infectious Diseases Reference Laboratory, Doherty Institute |
| EPI_ISL_426809 | Victorian Infectious Diseases Reference Laboratory (VIDRL) | Microbiological Diagnostic Unit Public Health Laboratory and Victorian Infectious Diseases Reference Laboratory, Doherty Institute |
| EPI_ISL_426810 | Victorian Infectious Diseases Reference Laboratory (VIDRL) | Microbiological Diagnostic Unit Public Health Laboratory and Victorian Infectious Diseases Reference Laboratory, Doherty Institute |
| EPI_ISL_426811 | Victorian Infectious Diseases Reference Laboratory (VIDRL) | Microbiological Diagnostic Unit Public Health Laboratory and Victorian Infectious Diseases Reference Laboratory, Doherty Institute |
| EPI_ISL_426812 | Victorian Infectious Diseases Reference Laboratory (VIDRL) | Microbiological Diagnostic Unit Public Health Laboratory and Victorian Infectious Diseases Reference Laboratory, Doherty Institute |
| EPI_ISL_426813 | Victorian Infectious Diseases Reference Laboratory (VIDRL) | Microbiological Diagnostic Unit Public Health Laboratory and Victorian Infectious Diseases Reference Laboratory, Doherty Institute |
| EPI_ISL_426814 | Victorian Infectious Diseases Reference Laboratory (VIDRL) | Microbiological Diagnostic Unit Public Health Laboratory and Victorian Infectious Diseases Reference Laboratory, Doherty Institute |
| EPI_ISL_426815 | Victorian Infectious Diseases Reference Laboratory (VIDRL) | Microbiological Diagnostic Unit Public Health Laboratory and Victorian Infectious Diseases Reference Laboratory, Doherty Institute |
| EPI_ISL_426816 | Victorian Infectious Diseases Reference Laboratory (VIDRL) | Microbiological Diagnostic Unit Public Health Laboratory and Victorian Infectious Diseases Reference Laboratory, Doherty Institute |
| EPI_ISL_426817 | Victorian Infectious Diseases Reference Laboratory (VIDRL) | Microbiological Diagnostic Unit Public Health Laboratory and Victorian Infectious Diseases Reference Laboratory, Doherty Institute |
| EPI_ISL_426818 | Victorian Infectious Diseases Reference Laboratory (VIDRL) | Microbiological Diagnostic Unit Public Health Laboratory and Victorian Infectious Diseases Reference Laboratory, Doherty Institute |

| EPI_Accession_ID | Originating Laboratory | Submitting Laboratory |
|---|---|---|
| EPI_ISL_426819 | Victorian Infectious Diseases Reference Laboratory (VIDRL) | Microbiological Diagnostic Unit Public Health Laboratory and Victorian Infectious Diseases Reference Laboratory, Doherty Institute |
| EPI_ISL_426820 | Victorian Infectious Diseases Reference Laboratory (VIDRL) | Microbiological Diagnostic Unit Public Health Laboratory and Victorian Infectious Diseases Reference Laboratory, Doherty Institute |
| EPI_ISL_426821 | Victorian Infectious Diseases Reference Laboratory (VIDRL) | Microbiological Diagnostic Unit Public Health Laboratory and Victorian Infectious Diseases Reference Laboratory, Doherty Institute |
| EPI_ISL_426822 | Victorian Infectious Diseases Reference Laboratory (VIDRL) | Microbiological Diagnostic Unit Public Health Laboratory and Victorian Infectious Diseases Reference Laboratory, Doherty Institute |
| EPI_ISL_426823 | Victorian Infectious Diseases Reference Laboratory (VIDRL) | Microbiological Diagnostic Unit Public Health Laboratory and Victorian Infectious Diseases Reference Laboratory, Doherty Institute |
| EPI_ISL_426824 | Victorian Infectious Diseases Reference Laboratory (VIDRL) | Microbiological Diagnostic Unit Public Health Laboratory and Victorian Infectious Diseases Reference Laboratory, Doherty Institute |
| EPI_ISL_426825 | Victorian Infectious Diseases Reference Laboratory (VIDRL) | Microbiological Diagnostic Unit Public Health Laboratory and Victorian Infectious Diseases Reference Laboratory, Doherty Institute |
| EPI_ISL_426826 | Victorian Infectious Diseases Reference Laboratory (VIDRL) | Microbiological Diagnostic Unit Public Health Laboratory and Victorian Infectious Diseases Reference Laboratory, Doherty Institute |
| EPI_ISL_426827 | Victorian Infectious Diseases Reference Laboratory (VIDRL) | Microbiological Diagnostic Unit Public Health Laboratory and Victorian Infectious Diseases Reference Laboratory, Doherty Institute |
| EPI_ISL_426828 | Victorian Infectious Diseases Reference Laboratory (VIDRL) | Microbiological Diagnostic Unit Public Health Laboratory and Victorian Infectious Diseases Reference Laboratory, Doherty Institute |
| EPI_ISL_426829 | Victorian Infectious Diseases Reference Laboratory (VIDRL) | Microbiological Diagnostic Unit Public Health Laboratory and Victorian Infectious Diseases Reference Laboratory, Doherty Institute |

| EPI_Accession_ID | Originating Laboratory | Submitting Laboratory |
|---|---|---|
| EPI_ISL_426830 | Victorian Infectious Diseases Reference Laboratory (VIDRL) | Microbiological Diagnostic Unit Public Health Laboratory and Victorian Infectious Diseases Reference Laboratory, Doherty Institute |
| EPI_ISL_426831 | Victorian Infectious Diseases Reference Laboratory (VIDRL) | Microbiological Diagnostic Unit Public Health Laboratory and Victorian Infectious Diseases Reference Laboratory, Doherty Institute |
| EPI_ISL_426832 | Victorian Infectious Diseases Reference Laboratory (VIDRL) | Microbiological Diagnostic Unit Public Health Laboratory and Victorian Infectious Diseases Reference Laboratory, Doherty Institute |
| EPI_ISL_426833 | Victorian Infectious Diseases Reference Laboratory (VIDRL) | Microbiological Diagnostic Unit Public Health Laboratory and Victorian Infectious Diseases Reference Laboratory, Doherty Institute |
| EPI_ISL_426834 | Victorian Infectious Diseases Reference Laboratory (VIDRL) | Microbiological Diagnostic Unit Public Health Laboratory and Victorian Infectious Diseases Reference Laboratory, Doherty Institute |
| EPI_ISL_426835 | Victorian Infectious Diseases Reference Laboratory (VIDRL) | Microbiological Diagnostic Unit Public Health Laboratory and Victorian Infectious Diseases Reference Laboratory, Doherty Institute |
| EPI_ISL_426836 | Victorian Infectious Diseases Reference Laboratory (VIDRL) | Microbiological Diagnostic Unit Public Health Laboratory and Victorian Infectious Diseases Reference Laboratory, Doherty Institute |
| EPI_ISL_426837 | Victorian Infectious Diseases Reference Laboratory (VIDRL) | Microbiological Diagnostic Unit Public Health Laboratory and Victorian Infectious Diseases Reference Laboratory, Doherty Institute |
| EPI_ISL_426838 | Victorian Infectious Diseases Reference Laboratory (VIDRL) | Microbiological Diagnostic Unit Public Health Laboratory and Victorian Infectious Diseases Reference Laboratory, Doherty Institute |
| EPI_ISL_426839 | Victorian Infectious Diseases Reference Laboratory (VIDRL) | Microbiological Diagnostic Unit Public Health Laboratory and Victorian Infectious Diseases Reference Laboratory, Doherty Institute |
| EPI_ISL_426840 | Victorian Infectious Diseases Reference Laboratory (VIDRL) | Microbiological Diagnostic Unit Public Health Laboratory and Victorian Infectious Diseases Reference Laboratory, Doherty Institute |

| EPI_Accession_ID | Originating Laboratory | Submitting Laboratory |
|---|---|---|
| EPI_ISL_426841 | Victorian Infectious Diseases Reference Laboratory (VIDRL) | Microbiological Diagnostic Unit Public Health Laboratory and Victorian Infectious Diseases Reference Laboratory, Doherty Institute |
| EPI_ISL_426842 | Victorian Infectious Diseases Reference Laboratory (VIDRL) | Microbiological Diagnostic Unit Public Health Laboratory and Victorian Infectious Diseases Reference Laboratory, Doherty Institute |
| EPI_ISL_426843 | Victorian Infectious Diseases Reference Laboratory (VIDRL) | Microbiological Diagnostic Unit Public Health Laboratory and Victorian Infectious Diseases Reference Laboratory, Doherty Institute |
| EPI_ISL_426844 | Victorian Infectious Diseases Reference Laboratory (VIDRL) | Microbiological Diagnostic Unit Public Health Laboratory and Victorian Infectious Diseases Reference Laboratory, Doherty Institute |
| EPI_ISL_426845 | Victorian Infectious Diseases Reference Laboratory (VIDRL) | Microbiological Diagnostic Unit Public Health Laboratory and Victorian Infectious Diseases Reference Laboratory, Doherty Institute |
| EPI_ISL_426846 | Victorian Infectious Diseases Reference Laboratory (VIDRL) | Microbiological Diagnostic Unit Public Health Laboratory and Victorian Infectious Diseases Reference Laboratory, Doherty Institute |
| EPI_ISL_426847 | Victorian Infectious Diseases Reference Laboratory (VIDRL) | Microbiological Diagnostic Unit Public Health Laboratory and Victorian Infectious Diseases Reference Laboratory, Doherty Institute |
| EPI_ISL_426848 | Victorian Infectious Diseases Reference Laboratory (VIDRL) | Microbiological Diagnostic Unit Public Health Laboratory and Victorian Infectious Diseases Reference Laboratory, Doherty Institute |
| EPI_ISL_426849 | Victorian Infectious Diseases Reference Laboratory (VIDRL) | Microbiological Diagnostic Unit Public Health Laboratory and Victorian Infectious Diseases Reference Laboratory, Doherty Institute |
| EPI_ISL_426850 | Victorian Infectious Diseases Reference Laboratory (VIDRL) | Microbiological Diagnostic Unit Public Health Laboratory and Victorian Infectious Diseases Reference Laboratory, Doherty Institute |
| EPI_ISL_426851 | Victorian Infectious Diseases Reference Laboratory (VIDRL) | Microbiological Diagnostic Unit Public Health Laboratory and Victorian Infectious Diseases Reference Laboratory, Doherty Institute |

| EPI_Accession_ID | Originating Laboratory | Submitting Laboratory |
|---|---|---|
| EPI_ISL_426852 | Victorian Infectious Diseases Reference Laboratory (VIDRL) | Microbiological Diagnostic Unit Public Health Laboratory and Victorian Infectious Diseases Reference Laboratory, Doherty Institute |
| EPI_ISL_426853 | Victorian Infectious Diseases Reference Laboratory (VIDRL) | Microbiological Diagnostic Unit Public Health Laboratory and Victorian Infectious Diseases Reference Laboratory, Doherty Institute |
| EPI_ISL_426854 | Victorian Infectious Diseases Reference Laboratory (VIDRL) | Microbiological Diagnostic Unit Public Health Laboratory and Victorian Infectious Diseases Reference Laboratory, Doherty Institute |
| EPI_ISL_426855 | Victorian Infectious Diseases Reference Laboratory (VIDRL) | Microbiological Diagnostic Unit Public Health Laboratory and Victorian Infectious Diseases Reference Laboratory, Doherty Institute |
| EPI_ISL_426856 | Victorian Infectious Diseases Reference Laboratory (VIDRL) | Microbiological Diagnostic Unit Public Health Laboratory and Victorian Infectious Diseases Reference Laboratory, Doherty Institute |
| EPI_ISL_426857 | Victorian Infectious Diseases Reference Laboratory (VIDRL) | Microbiological Diagnostic Unit Public Health Laboratory and Victorian Infectious Diseases Reference Laboratory, Doherty Institute |
| EPI_ISL_426858 | Victorian Infectious Diseases Reference Laboratory (VIDRL) | Microbiological Diagnostic Unit Public Health Laboratory and Victorian Infectious Diseases Reference Laboratory, Doherty Institute |
| EPI_ISL_426859 | Victorian Infectious Diseases Reference Laboratory (VIDRL) | Microbiological Diagnostic Unit Public Health Laboratory and Victorian Infectious Diseases Reference Laboratory, Doherty Institute |
| EPI_ISL_426860 | Victorian Infectious Diseases Reference Laboratory (VIDRL) | Microbiological Diagnostic Unit Public Health Laboratory and Victorian Infectious Diseases Reference Laboratory, Doherty Institute |
| EPI_ISL_426861 | Victorian Infectious Diseases Reference Laboratory (VIDRL) | Microbiological Diagnostic Unit Public Health Laboratory and Victorian Infectious Diseases Reference Laboratory, Doherty Institute |
| EPI_ISL_426862 | Victorian Infectious Diseases Reference Laboratory (VIDRL) | Microbiological Diagnostic Unit Public Health Laboratory and Victorian Infectious Diseases Reference Laboratory, Doherty Institute |

| EPI_Accession_ID | Originating Laboratory | Submitting Laboratory |
|---|---|---|
| EPI_ISL_426863 | Victorian Infectious Diseases Reference Laboratory (VIDRL) | Microbiological Diagnostic Unit Public Health Laboratory and Victorian Infectious Diseases Reference Laboratory, Doherty Institute |
| EPI_ISL_426864 | Victorian Infectious Diseases Reference Laboratory (VIDRL) | Microbiological Diagnostic Unit Public Health Laboratory and Victorian Infectious Diseases Reference Laboratory, Doherty Institute |
| EPI_ISL_426865 | Victorian Infectious Diseases Reference Laboratory (VIDRL) | Microbiological Diagnostic Unit Public Health Laboratory and Victorian Infectious Diseases Reference Laboratory, Doherty Institute |
| EPI_ISL_426866 | Victorian Infectious Diseases Reference Laboratory (VIDRL) | Microbiological Diagnostic Unit Public Health Laboratory and Victorian Infectious Diseases Reference Laboratory, Doherty Institute |
| EPI_ISL_426867 | Victorian Infectious Diseases Reference Laboratory (VIDRL) | Microbiological Diagnostic Unit Public Health Laboratory and Victorian Infectious Diseases Reference Laboratory, Doherty Institute |
| EPI_ISL_426869 | Victorian Infectious Diseases Reference Laboratory (VIDRL) | Microbiological Diagnostic Unit Public Health Laboratory and Victorian Infectious Diseases Reference Laboratory, Doherty Institute |
| EPI_ISL_426870 | Victorian Infectious Diseases Reference Laboratory (VIDRL) | Microbiological Diagnostic Unit Public Health Laboratory and Victorian Infectious Diseases Reference Laboratory, Doherty Institute |
| EPI_ISL_426871 | Victorian Infectious Diseases Reference Laboratory (VIDRL) | Microbiological Diagnostic Unit Public Health Laboratory and Victorian Infectious Diseases Reference Laboratory, Doherty Institute |
| EPI_ISL_426872 | Victorian Infectious Diseases Reference Laboratory (VIDRL) | Microbiological Diagnostic Unit Public Health Laboratory and Victorian Infectious Diseases Reference Laboratory, Doherty Institute |
| EPI_ISL_426873 | Victorian Infectious Diseases Reference Laboratory (VIDRL) | Microbiological Diagnostic Unit Public Health Laboratory and Victorian Infectious Diseases Reference Laboratory, Doherty Institute |
| EPI_ISL_426874 | Victorian Infectious Diseases Reference Laboratory (VIDRL) | Microbiological Diagnostic Unit Public Health Laboratory and Victorian Infectious Diseases Reference Laboratory, Doherty Institute |

| EPI_Accession_ID | Originating Laboratory | Submitting Laboratory |
|---|---|---|
| EPI_ISL_426875 | Victorian Infectious Diseases Reference Laboratory (VIDRL) | Microbiological Diagnostic Unit Public Health Laboratory and Victorian Infectious Diseases Reference Laboratory, Doherty Institute |
| EPI_ISL_426876 | Victorian Infectious Diseases Reference Laboratory (VIDRL) | Microbiological Diagnostic Unit Public Health Laboratory and Victorian Infectious Diseases Reference Laboratory, Doherty Institute |
| EPI_ISL_426877 | Victorian Infectious Diseases Reference Laboratory (VIDRL) | Microbiological Diagnostic Unit Public Health Laboratory and Victorian Infectious Diseases Reference Laboratory, Doherty Institute |
| EPI_ISL_426878 | Victorian Infectious Diseases Reference Laboratory (VIDRL) | Microbiological Diagnostic Unit Public Health Laboratory and Victorian Infectious Diseases Reference Laboratory, Doherty Institute |
| EPI_ISL_426879 | Victorian Infectious Diseases Reference Laboratory (VIDRL) | Microbiological Diagnostic Unit Public Health Laboratory and Victorian Infectious Diseases Reference Laboratory, Doherty Institute |
| EPI_ISL_426881 | Victorian Infectious Diseases Reference Laboratory (VIDRL) | Microbiological Diagnostic Unit Public Health Laboratory and Victorian Infectious Diseases Reference Laboratory, Doherty Institute |
| EPI_ISL_426882 | Victorian Infectious Diseases Reference Laboratory (VIDRL) | Microbiological Diagnostic Unit Public Health Laboratory and Victorian Infectious Diseases Reference Laboratory, Doherty Institute |
| EPI_ISL_426883 | Motol University Hospital | Institute of Applied Biotechnologies a.s. |
| EPI_ISL_426884 | Motol University Hospital | Institute of Applied Biotechnologies a.s. |
| EPI_ISL_426885 | Motol University Hospital | Institute of Applied Biotechnologies a.s. |
| EPI_ISL_426886 | Motol University Hospital | Institute of Applied Biotechnologies a.s. |
| EPI_ISL_426887 | Motol University Hospital | Institute of Applied Biotechnologies a.s. |
| EPI_ISL_426888 | Motol University Hospital | Institute of Applied Biotechnologies a.s. |
| EPI_ISL_426889 | Motol University Hospital | Institute of Applied Biotechnologies a.s. |
| EPI_ISL_426890 | Motol University Hospital | Institute of Applied Biotechnologies a.s. |
| EPI_ISL_426891 | Motol University Hospital | Institute of Applied Biotechnologies a.s. |
| EPI_ISL_426892 | Motol University Hospital | Institute of Applied Biotechnologies a.s. |
| EPI_ISL_426893 | Motol University Hospital | Institute of Applied Biotechnologies a.s. |
| EPI_ISL_426894 | Motol University Hospital | Institute of Applied Biotechnologies a.s. |

| EPI_Accession_ID | Originating Laboratory | Submitting Laboratory |
|---|---|---|
| EPI_ISL_426895 | Motol University Hospital | Institute of Applied Biotechnologies a.s. |
| EPI_ISL_426896 | Motol University Hospital | Institute of Applied Biotechnologies a.s. |
| EPI_ISL_426897 | Motol University Hospital | Institute of Applied Biotechnologies a.s. |
| EPI_ISL_426898 | Royal Darwin Hospital Pathology | Microbiological Diagnostic Unit Public Health Laboratory and Victorian Infectious Diseases Reference Laboratory, Doherty Institute |
| EPI_ISL_426899 | Royal Darwin Hospital Pathology | Microbiological Diagnostic Unit Public Health Laboratory and Victorian Infectious Diseases Reference Laboratory, Doherty Institute |
| EPI_ISL_426900 | Royal Darwin Hospital Pathology | Microbiological Diagnostic Unit Public Health Laboratory and Victorian Infectious Diseases Reference Laboratory, Doherty Institute |
| EPI_ISL_426901 | Royal Darwin Hospital Pathology | Microbiological Diagnostic Unit Public Health Laboratory and Victorian Infectious Diseases Reference Laboratory, Doherty Institute |
| EPI_ISL_426902 | Royal Darwin Hospital Pathology | Microbiological Diagnostic Unit Public Health Laboratory and Victorian Infectious Diseases Reference Laboratory, Doherty Institute |
| EPI_ISL_426903 | Royal Darwin Hospital Pathology | Microbiological Diagnostic Unit Public Health Laboratory and Victorian Infectious Diseases Reference Laboratory, Doherty Institute |
| EPI_ISL_426904 | Royal Darwin Hospital Pathology | Microbiological Diagnostic Unit Public Health Laboratory and Victorian Infectious Diseases Reference Laboratory, Doherty Institute |
| EPI_ISL_426905 | Microbiological Diagnostic Unit Public Health Laboratory | Microbiological Diagnostic Unit Public Health Laboratory |
| EPI_ISL_426906 | Microbiological Diagnostic Unit Public Health Laboratory | Microbiological Diagnostic Unit Public Health Laboratory |
| EPI_ISL_426907 | Microbiological Diagnostic Unit Public Health Laboratory | Microbiological Diagnostic Unit Public Health Laboratory |
| EPI_ISL_426908 | Microbiological Diagnostic Unit Public Health Laboratory | Microbiological Diagnostic Unit Public Health Laboratory |

| EPI_Accession_ID | Originating Laboratory | Submitting Laboratory |
|---|---|---|
| EPI_ISL_426909 | Microbiological Diagnostic Unit Public Health Laboratory | Microbiological Diagnostic Unit Public Health Laboratory |
| EPI_ISL_426910 | Microbiological Diagnostic Unit Public Health Laboratory | Microbiological Diagnostic Unit Public Health Laboratory |
| EPI_ISL_426911 | Microbiological Diagnostic Unit Public Health Laboratory | Microbiological Diagnostic Unit Public Health Laboratory |
| EPI_ISL_426912 | Microbiological Diagnostic Unit Public Health Laboratory | Microbiological Diagnostic Unit Public Health Laboratory |
| EPI_ISL_426913 | Microbiological Diagnostic Unit Public Health Laboratory | Microbiological Diagnostic Unit Public Health Laboratory |
| EPI_ISL_426914 | Microbiological Diagnostic Unit Public Health Laboratory | Microbiological Diagnostic Unit Public Health Laboratory |
| EPI_ISL_426915 | Microbiological Diagnostic Unit Public Health Laboratory | Microbiological Diagnostic Unit Public Health Laboratory |
| EPI_ISL_426916 | Microbiological Diagnostic Unit Public Health Laboratory | Microbiological Diagnostic Unit Public Health Laboratory |
| EPI_ISL_426917 | Microbiological Diagnostic Unit Public Health Laboratory | Microbiological Diagnostic Unit Public Health Laboratory |
| EPI_ISL_426918 | Microbiological Diagnostic Unit Public Health Laboratory | Microbiological Diagnostic Unit Public Health Laboratory |
| EPI_ISL_426919 | Microbiological Diagnostic Unit Public Health Laboratory | Microbiological Diagnostic Unit Public Health Laboratory |
| EPI_ISL_426920 | Microbiological Diagnostic Unit Public Health Laboratory | Microbiological Diagnostic Unit Public Health Laboratory |
| EPI_ISL_426921 | Microbiological Diagnostic Unit Public Health Laboratory | Microbiological Diagnostic Unit Public Health Laboratory |
| EPI_ISL_426922 | Microbiological Diagnostic Unit Public Health Laboratory | Microbiological Diagnostic Unit Public Health Laboratory |
| EPI_ISL_426923 | Victorian Infectious Diseases Reference Laboratory (VIDRL) | Microbiological Diagnostic Unit Public Health Laboratory and Victorian Infectious Diseases Reference Laboratory, Doherty Institute |

| EPI_Accession_ID | Originating Laboratory | Submitting Laboratory |
|---|---|---|
| EPI_ISL_426924 | Victorian Infectious Diseases Reference Laboratory (VIDRL) | Microbiological Diagnostic Unit Public Health Laboratory and Victorian Infectious Diseases Reference Laboratory, Doherty Institute |
| EPI_ISL_426925 | Victorian Infectious Diseases Reference Laboratory (VIDRL) | Microbiological Diagnostic Unit Public Health Laboratory and Victorian Infectious Diseases Reference Laboratory, Doherty Institute |
| EPI_ISL_426926 | Victorian Infectious Diseases Reference Laboratory (VIDRL) | Microbiological Diagnostic Unit Public Health Laboratory and Victorian Infectious Diseases Reference Laboratory, Doherty Institute |
| EPI_ISL_426927 | Victorian Infectious Diseases Reference Laboratory (VIDRL) | Microbiological Diagnostic Unit Public Health Laboratory and Victorian Infectious Diseases Reference Laboratory, Doherty Institute |
| EPI_ISL_426928 | Victorian Infectious Diseases Reference Laboratory (VIDRL) | Microbiological Diagnostic Unit Public Health Laboratory and Victorian Infectious Diseases Reference Laboratory, Doherty Institute |
| EPI_ISL_426929 | Victorian Infectious Diseases Reference Laboratory (VIDRL) | Microbiological Diagnostic Unit Public Health Laboratory and Victorian Infectious Diseases Reference Laboratory, Doherty Institute |
| EPI_ISL_426930 | Victorian Infectious Diseases Reference Laboratory (VIDRL) | Microbiological Diagnostic Unit Public Health Laboratory and Victorian Infectious Diseases Reference Laboratory, Doherty Institute |
| EPI_ISL_426931 | Victorian Infectious Diseases Reference Laboratory (VIDRL) | Microbiological Diagnostic Unit Public Health Laboratory and Victorian Infectious Diseases Reference Laboratory, Doherty Institute |
| EPI_ISL_426932 | Victorian Infectious Diseases Reference Laboratory (VIDRL) | Microbiological Diagnostic Unit Public Health Laboratory and Victorian Infectious Diseases Reference Laboratory, Doherty Institute |
| EPI_ISL_426933 | Victorian Infectious Diseases Reference Laboratory (VIDRL) | Microbiological Diagnostic Unit Public Health Laboratory and Victorian Infectious Diseases Reference Laboratory, Doherty Institute |
| EPI_ISL_426934 | Victorian Infectious Diseases Reference Laboratory (VIDRL) | Microbiological Diagnostic Unit Public Health Laboratory and Victorian Infectious Diseases Reference Laboratory, Doherty Institute |

| EPI_Accession_ID | Originating Laboratory | Submitting Laboratory |
|---|---|---|
| EPI_ISL_426935 | Victorian Infectious Diseases Reference Laboratory (VIDRL) | Microbiological Diagnostic Unit Public Health Laboratory and Victorian Infectious Diseases Reference Laboratory, Doherty Institute |
| EPI_ISL_426936 | Victorian Infectious Diseases Reference Laboratory (VIDRL) | Microbiological Diagnostic Unit Public Health Laboratory and Victorian Infectious Diseases Reference Laboratory, Doherty Institute |
| EPI_ISL_426937 | Victorian Infectious Diseases Reference Laboratory (VIDRL) | Microbiological Diagnostic Unit Public Health Laboratory and Victorian Infectious Diseases Reference Laboratory, Doherty Institute |
| EPI_ISL_426938 | Victorian Infectious Diseases Reference Laboratory (VIDRL) | Microbiological Diagnostic Unit Public Health Laboratory and Victorian Infectious Diseases Reference Laboratory, Doherty Institute |
| EPI_ISL_426939 | Victorian Infectious Diseases Reference Laboratory (VIDRL) | Microbiological Diagnostic Unit Public Health Laboratory and Victorian Infectious Diseases Reference Laboratory, Doherty Institute |
| EPI_ISL_426940 | Victorian Infectious Diseases Reference Laboratory (VIDRL) | Microbiological Diagnostic Unit Public Health Laboratory and Victorian Infectious Diseases Reference Laboratory, Doherty Institute |
| EPI_ISL_426941 | Victorian Infectious Diseases Reference Laboratory (VIDRL) | Microbiological Diagnostic Unit Public Health Laboratory and Victorian Infectious Diseases Reference Laboratory, Doherty Institute |
| EPI_ISL_426942 | Victorian Infectious Diseases Reference Laboratory (VIDRL) | Microbiological Diagnostic Unit Public Health Laboratory and Victorian Infectious Diseases Reference Laboratory, Doherty Institute |
| EPI_ISL_426943 | Victorian Infectious Diseases Reference Laboratory (VIDRL) | Microbiological Diagnostic Unit Public Health Laboratory and Victorian Infectious Diseases Reference Laboratory, Doherty Institute |
| EPI_ISL_426944 | Victorian Infectious Diseases Reference Laboratory (VIDRL) | Microbiological Diagnostic Unit Public Health Laboratory and Victorian Infectious Diseases Reference Laboratory, Doherty Institute |
| EPI_ISL_426945 | Victorian Infectious Diseases Reference Laboratory (VIDRL) | Microbiological Diagnostic Unit Public Health Laboratory and Victorian Infectious Diseases Reference Laboratory, Doherty Institute |

| EPI_Accession_ID | Originating Laboratory | Submitting Laboratory |
|---|---|---|
| EPI_ISL_426947 | Victorian Infectious Diseases Reference Laboratory (VIDRL) | Microbiological Diagnostic Unit Public Health Laboratory and Victorian Infectious Diseases Reference Laboratory, Doherty Institute |
| EPI_ISL_426948 | Victorian Infectious Diseases Reference Laboratory (VIDRL) | Microbiological Diagnostic Unit Public Health Laboratory and Victorian Infectious Diseases Reference Laboratory, Doherty Institute |
| EPI_ISL_426949 | Victorian Infectious Diseases Reference Laboratory (VIDRL) | Microbiological Diagnostic Unit Public Health Laboratory and Victorian Infectious Diseases Reference Laboratory, Doherty Institute |
| EPI_ISL_426950 | Victorian Infectious Diseases Reference Laboratory (VIDRL) | Microbiological Diagnostic Unit Public Health Laboratory and Victorian Infectious Diseases Reference Laboratory, Doherty Institute |
| EPI_ISL_426951 | Victorian Infectious Diseases Reference Laboratory (VIDRL) | Microbiological Diagnostic Unit Public Health Laboratory and Victorian Infectious Diseases Reference Laboratory, Doherty Institute |
| EPI_ISL_426952 | Victorian Infectious Diseases Reference Laboratory (VIDRL) | Microbiological Diagnostic Unit Public Health Laboratory and Victorian Infectious Diseases Reference Laboratory, Doherty Institute |
| EPI_ISL_426953 | Victorian Infectious Diseases Reference Laboratory (VIDRL) | Microbiological Diagnostic Unit Public Health Laboratory and Victorian Infectious Diseases Reference Laboratory, Doherty Institute |
| EPI_ISL_426954 | Victorian Infectious Diseases Reference Laboratory (VIDRL) | Microbiological Diagnostic Unit Public Health Laboratory and Victorian Infectious Diseases Reference Laboratory, Doherty Institute |
| EPI_ISL_426955 | Victorian Infectious Diseases Reference Laboratory (VIDRL) | Microbiological Diagnostic Unit Public Health Laboratory and Victorian Infectious Diseases Reference Laboratory, Doherty Institute |
| EPI_ISL_426956 | Victorian Infectious Diseases Reference Laboratory (VIDRL) | Microbiological Diagnostic Unit Public Health Laboratory and Victorian Infectious Diseases Reference Laboratory, Doherty Institute |
| EPI_ISL_426957 | Victorian Infectious Diseases Reference Laboratory (VIDRL) | Microbiological Diagnostic Unit Public Health Laboratory and Victorian Infectious Diseases Reference Laboratory, Doherty Institute |

| EPI_Accession_ID | Originating Laboratory | Submitting Laboratory |
|---|---|---|
| EPI_ISL_426958 | Victorian Infectious Diseases Reference Laboratory (VIDRL) | Microbiological Diagnostic Unit Public Health Laboratory and Victorian Infectious Diseases Reference Laboratory, Doherty Institute |
| EPI_ISL_426959 | Victorian Infectious Diseases Reference Laboratory (VIDRL) | Microbiological Diagnostic Unit Public Health Laboratory and Victorian Infectious Diseases Reference Laboratory, Doherty Institute |
| EPI_ISL_426960 | Victorian Infectious Diseases Reference Laboratory (VIDRL) | Microbiological Diagnostic Unit Public Health Laboratory and Victorian Infectious Diseases Reference Laboratory, Doherty Institute |
| EPI_ISL_426961 | Victorian Infectious Diseases Reference Laboratory (VIDRL) | Microbiological Diagnostic Unit Public Health Laboratory and Victorian Infectious Diseases Reference Laboratory, Doherty Institute |
| EPI_ISL_426962 | Victorian Infectious Diseases Reference Laboratory (VIDRL) | Microbiological Diagnostic Unit Public Health Laboratory and Victorian Infectious Diseases Reference Laboratory, Doherty Institute |
| EPI_ISL_426963 | Victorian Infectious Diseases Reference Laboratory (VIDRL) | Microbiological Diagnostic Unit Public Health Laboratory and Victorian Infectious Diseases Reference Laboratory, Doherty Institute |
| EPI_ISL_426964 | Victorian Infectious Diseases Reference Laboratory (VIDRL) | Microbiological Diagnostic Unit Public Health Laboratory and Victorian Infectious Diseases Reference Laboratory, Doherty Institute |
| EPI_ISL_426965 | Victorian Infectious Diseases Reference Laboratory (VIDRL) | Microbiological Diagnostic Unit Public Health Laboratory and Victorian Infectious Diseases Reference Laboratory, Doherty Institute |
| EPI_ISL_426966 | Victorian Infectious Diseases Reference Laboratory (VIDRL) | Microbiological Diagnostic Unit Public Health Laboratory and Victorian Infectious Diseases Reference Laboratory, Doherty Institute |
| EPI_ISL_426967 | Victorian Infectious Diseases Reference Laboratory (VIDRL) | Microbiological Diagnostic Unit Public Health Laboratory and Victorian Infectious Diseases Reference Laboratory, Doherty Institute |
| EPI_ISL_426968 | Victorian Infectious Diseases Reference Laboratory (VIDRL) | Microbiological Diagnostic Unit Public Health Laboratory and Victorian Infectious Diseases Reference Laboratory, Doherty Institute |

| EPI_Accession_ID | Originating Laboratory | Submitting Laboratory |
|---|---|---|
| EPI_ISL_426969 | Victorian Infectious Diseases Reference Laboratory (VIDRL) | Microbiological Diagnostic Unit Public Health Laboratory and Victorian Infectious Diseases Reference Laboratory, Doherty Institute |
| EPI_ISL_426970 | Victorian Infectious Diseases Reference Laboratory (VIDRL) | Microbiological Diagnostic Unit Public Health Laboratory and Victorian Infectious Diseases Reference Laboratory, Doherty Institute |
| EPI_ISL_426971 | Victorian Infectious Diseases Reference Laboratory (VIDRL) | Microbiological Diagnostic Unit Public Health Laboratory and Victorian Infectious Diseases Reference Laboratory, Doherty Institute |
| EPI_ISL_426972 | Victorian Infectious Diseases Reference Laboratory (VIDRL) | Microbiological Diagnostic Unit Public Health Laboratory and Victorian Infectious Diseases Reference Laboratory, Doherty Institute |
| EPI_ISL_426973 | Victorian Infectious Diseases Reference Laboratory (VIDRL) | Microbiological Diagnostic Unit Public Health Laboratory and Victorian Infectious Diseases Reference Laboratory, Doherty Institute |
| EPI_ISL_426974 | Victorian Infectious Diseases Reference Laboratory (VIDRL) | Microbiological Diagnostic Unit Public Health Laboratory and Victorian Infectious Diseases Reference Laboratory, Doherty Institute |
| EPI_ISL_426975 | Victorian Infectious Diseases Reference Laboratory (VIDRL) | Microbiological Diagnostic Unit Public Health Laboratory and Victorian Infectious Diseases Reference Laboratory, Doherty Institute |
| EPI_ISL_426976 | Victorian Infectious Diseases Reference Laboratory (VIDRL) | Microbiological Diagnostic Unit Public Health Laboratory and Victorian Infectious Diseases Reference Laboratory, Doherty Institute |
| EPI_ISL_426977 | Victorian Infectious Diseases Reference Laboratory (VIDRL) | Microbiological Diagnostic Unit Public Health Laboratory and Victorian Infectious Diseases Reference Laboratory, Doherty Institute |
| EPI_ISL_426978 | Victorian Infectious Diseases Reference Laboratory (VIDRL) | Microbiological Diagnostic Unit Public Health Laboratory and Victorian Infectious Diseases Reference Laboratory, Doherty Institute |
| EPI_ISL_426979 | Victorian Infectious Diseases Reference Laboratory (VIDRL) | Microbiological Diagnostic Unit Public Health Laboratory and Victorian Infectious Diseases Reference Laboratory, Doherty Institute |

| EPI_Accession_ID | Originating Laboratory | Submitting Laboratory |
|---|---|---|
| EPI_ISL_426980 | Victorian Infectious Diseases Reference Laboratory (VIDRL) | Microbiological Diagnostic Unit Public Health Laboratory and Victorian Infectious Diseases Reference Laboratory, Doherty Institute |
| EPI_ISL_426981 | Victorian Infectious Diseases Reference Laboratory (VIDRL) | Microbiological Diagnostic Unit Public Health Laboratory and Victorian Infectious Diseases Reference Laboratory, Doherty Institute |
| EPI_ISL_426982 | Victorian Infectious Diseases Reference Laboratory (VIDRL) | Microbiological Diagnostic Unit Public Health Laboratory and Victorian Infectious Diseases Reference Laboratory, Doherty Institute |
| EPI_ISL_426983 | Victorian Infectious Diseases Reference Laboratory (VIDRL) | Microbiological Diagnostic Unit Public Health Laboratory and Victorian Infectious Diseases Reference Laboratory, Doherty Institute |
| EPI_ISL_426984 | Victorian Infectious Diseases Reference Laboratory (VIDRL) | Microbiological Diagnostic Unit Public Health Laboratory and Victorian Infectious Diseases Reference Laboratory, Doherty Institute |
| EPI_ISL_426985 | Victorian Infectious Diseases Reference Laboratory (VIDRL) | Microbiological Diagnostic Unit Public Health Laboratory and Victorian Infectious Diseases Reference Laboratory, Doherty Institute |
| EPI_ISL_426986 | Victorian Infectious Diseases Reference Laboratory (VIDRL) | Microbiological Diagnostic Unit Public Health Laboratory and Victorian Infectious Diseases Reference Laboratory, Doherty Institute |
| EPI_ISL_426987 | Victorian Infectious Diseases Reference Laboratory (VIDRL) | Microbiological Diagnostic Unit Public Health Laboratory and Victorian Infectious Diseases Reference Laboratory, Doherty Institute |
| EPI_ISL_426988 | Victorian Infectious Diseases Reference Laboratory (VIDRL) | Microbiological Diagnostic Unit Public Health Laboratory and Victorian Infectious Diseases Reference Laboratory, Doherty Institute |
| EPI_ISL_426989 | Victorian Infectious Diseases Reference Laboratory (VIDRL) | Microbiological Diagnostic Unit Public Health Laboratory and Victorian Infectious Diseases Reference Laboratory, Doherty Institute |
| EPI_ISL_426990 | Victorian Infectious Diseases Reference Laboratory (VIDRL) | Microbiological Diagnostic Unit Public Health Laboratory and Victorian Infectious Diseases Reference Laboratory, Doherty Institute |

| EPI_Accession_ID | Originating Laboratory | Submitting Laboratory |
|---|---|---|
| EPI_ISL_426991 | Victorian Infectious Diseases Reference Laboratory (VIDRL) | Microbiological Diagnostic Unit Public Health Laboratory and Victorian Infectious Diseases Reference Laboratory, Doherty Institute |
| EPI_ISL_426992 | Victorian Infectious Diseases Reference Laboratory (VIDRL) | Microbiological Diagnostic Unit Public Health Laboratory and Victorian Infectious Diseases Reference Laboratory, Doherty Institute |
| EPI_ISL_426993 | Victorian Infectious Diseases Reference Laboratory (VIDRL) | Microbiological Diagnostic Unit Public Health Laboratory and Victorian Infectious Diseases Reference Laboratory, Doherty Institute |
| EPI_ISL_426994 | Victorian Infectious Diseases Reference Laboratory (VIDRL) | Microbiological Diagnostic Unit Public Health Laboratory and Victorian Infectious Diseases Reference Laboratory, Doherty Institute |
| EPI_ISL_426995 | Victorian Infectious Diseases Reference Laboratory (VIDRL) | Microbiological Diagnostic Unit Public Health Laboratory and Victorian Infectious Diseases Reference Laboratory, Doherty Institute |
| EPI_ISL_426996 | Victorian Infectious Diseases Reference Laboratory (VIDRL) | Microbiological Diagnostic Unit Public Health Laboratory and Victorian Infectious Diseases Reference Laboratory, Doherty Institute |
| EPI_ISL_426997 | Victorian Infectious Diseases Reference Laboratory (VIDRL) | Microbiological Diagnostic Unit Public Health Laboratory and Victorian Infectious Diseases Reference Laboratory, Doherty Institute |
| EPI_ISL_426998 | Victorian Infectious Diseases Reference Laboratory (VIDRL) | Microbiological Diagnostic Unit Public Health Laboratory and Victorian Infectious Diseases Reference Laboratory, Doherty Institute |
| EPI_ISL_426999 | Victorian Infectious Diseases Reference Laboratory (VIDRL) | Microbiological Diagnostic Unit Public Health Laboratory and Victorian Infectious Diseases Reference Laboratory, Doherty Institute |
| EPI_ISL_427000 | Victorian Infectious Diseases Reference Laboratory (VIDRL) | Microbiological Diagnostic Unit Public Health Laboratory and Victorian Infectious Diseases Reference Laboratory, Doherty Institute |
| EPI_ISL_427001 | Victorian Infectious Diseases Reference Laboratory (VIDRL) | Microbiological Diagnostic Unit Public Health Laboratory and Victorian Infectious Diseases Reference Laboratory, Doherty Institute |

| EPI_Accession_ID | Originating Laboratory | Submitting Laboratory |
|---|---|---|
| EPI_ISL_427002 | Victorian Infectious Diseases Reference Laboratory (VIDRL) | Microbiological Diagnostic Unit Public Health Laboratory and Victorian Infectious Diseases Reference Laboratory, Doherty Institute |
| EPI_ISL_427003 | Victorian Infectious Diseases Reference Laboratory (VIDRL) | Microbiological Diagnostic Unit Public Health Laboratory and Victorian Infectious Diseases Reference Laboratory, Doherty Institute |
| EPI_ISL_427004 | Victorian Infectious Diseases Reference Laboratory (VIDRL) | Microbiological Diagnostic Unit Public Health Laboratory and Victorian Infectious Diseases Reference Laboratory, Doherty Institute |
| EPI_ISL_427005 | Victorian Infectious Diseases Reference Laboratory (VIDRL) | Microbiological Diagnostic Unit Public Health Laboratory and Victorian Infectious Diseases Reference Laboratory, Doherty Institute |
| EPI_ISL_427006 | Victorian Infectious Diseases Reference Laboratory (VIDRL) | Microbiological Diagnostic Unit Public Health Laboratory and Victorian Infectious Diseases Reference Laboratory, Doherty Institute |
| EPI_ISL_427007 | Victorian Infectious Diseases Reference Laboratory (VIDRL) | Microbiological Diagnostic Unit Public Health Laboratory and Victorian Infectious Diseases Reference Laboratory, Doherty Institute |
| EPI_ISL_427008 | Victorian Infectious Diseases Reference Laboratory (VIDRL) | Microbiological Diagnostic Unit Public Health Laboratory and Victorian Infectious Diseases Reference Laboratory, Doherty Institute |
| EPI_ISL_427009 | Victorian Infectious Diseases Reference Laboratory (VIDRL) | Microbiological Diagnostic Unit Public Health Laboratory and Victorian Infectious Diseases Reference Laboratory, Doherty Institute |
| EPI_ISL_427010 | Victorian Infectious Diseases Reference Laboratory (VIDRL) | Microbiological Diagnostic Unit Public Health Laboratory and Victorian Infectious Diseases Reference Laboratory, Doherty Institute |
| EPI_ISL_427011 | Victorian Infectious Diseases Reference Laboratory (VIDRL) | Microbiological Diagnostic Unit Public Health Laboratory and Victorian Infectious Diseases Reference Laboratory, Doherty Institute |
| EPI_ISL_427012 | Victorian Infectious Diseases Reference Laboratory (VIDRL) | Microbiological Diagnostic Unit Public Health Laboratory and Victorian Infectious Diseases Reference Laboratory, Doherty Institute |

| EPI_Accession_ID | Originating Laboratory | Submitting Laboratory |
|---|---|---|
| EPI_ISL_427013 | Victorian Infectious Diseases Reference Laboratory (VIDRL) | Microbiological Diagnostic Unit Public Health Laboratory and Victorian Infectious Diseases Reference Laboratory, Doherty Institute |
| EPI_ISL_427014 | Victorian Infectious Diseases Reference Laboratory (VIDRL) | Microbiological Diagnostic Unit Public Health Laboratory and Victorian Infectious Diseases Reference Laboratory, Doherty Institute |
| EPI_ISL_427015 | Victorian Infectious Diseases Reference Laboratory (VIDRL) | Microbiological Diagnostic Unit Public Health Laboratory and Victorian Infectious Diseases Reference Laboratory, Doherty Institute |
| EPI_ISL_427016 | Victorian Infectious Diseases Reference Laboratory (VIDRL) | Microbiological Diagnostic Unit Public Health Laboratory and Victorian Infectious Diseases Reference Laboratory, Doherty Institute |
| EPI_ISL_427017 | Victorian Infectious Diseases Reference Laboratory (VIDRL) | Microbiological Diagnostic Unit Public Health Laboratory and Victorian Infectious Diseases Reference Laboratory, Doherty Institute |
| EPI_ISL_427018 | Victorian Infectious Diseases Reference Laboratory (VIDRL) | Microbiological Diagnostic Unit Public Health Laboratory and Victorian Infectious Diseases Reference Laboratory, Doherty Institute |
| EPI_ISL_427019 | Victorian Infectious Diseases Reference Laboratory (VIDRL) | Microbiological Diagnostic Unit Public Health Laboratory and Victorian Infectious Diseases Reference Laboratory, Doherty Institute |
| EPI_ISL_427020 | Victorian Infectious Diseases Reference Laboratory (VIDRL) | Microbiological Diagnostic Unit Public Health Laboratory and Victorian Infectious Diseases Reference Laboratory, Doherty Institute |
| EPI_ISL_427021 | Victorian Infectious Diseases Reference Laboratory (VIDRL) | Microbiological Diagnostic Unit Public Health Laboratory and Victorian Infectious Diseases Reference Laboratory, Doherty Institute |
| EPI_ISL_427022 | Victorian Infectious Diseases Reference Laboratory (VIDRL) | Microbiological Diagnostic Unit Public Health Laboratory and Victorian Infectious Diseases Reference Laboratory, Doherty Institute |
| EPI_ISL_427023 | Victorian Infectious Diseases Reference Laboratory (VIDRL) | Microbiological Diagnostic Unit Public Health Laboratory and Victorian Infectious Diseases Reference Laboratory, Doherty Institute |

| EPI_Accession_ID | Originating Laboratory | Submitting Laboratory |
|---|---|---|
| EPI_ISL_427024 | Victorian Infectious Diseases Reference Laboratory (VIDRL) | Microbiological Diagnostic Unit Public Health Laboratory and Victorian Infectious Diseases Reference Laboratory, Doherty Institute |
| EPI_ISL_427025 | Victorian Infectious Diseases Reference Laboratory (VIDRL) | Microbiological Diagnostic Unit Public Health Laboratory and Victorian Infectious Diseases Reference Laboratory, Doherty Institute |
| EPI_ISL_427026 | Victorian Infectious Diseases Reference Laboratory (VIDRL) | Microbiological Diagnostic Unit Public Health Laboratory and Victorian Infectious Diseases Reference Laboratory, Doherty Institute |
| EPI_ISL_427027 | Victorian Infectious Diseases Reference Laboratory (VIDRL) | Microbiological Diagnostic Unit Public Health Laboratory and Victorian Infectious Diseases Reference Laboratory, Doherty Institute |
| EPI_ISL_427028 | Victorian Infectious Diseases Reference Laboratory (VIDRL) | Microbiological Diagnostic Unit Public Health Laboratory and Victorian Infectious Diseases Reference Laboratory, Doherty Institute |
| EPI_ISL_427029 | Victorian Infectious Diseases Reference Laboratory (VIDRL) | Microbiological Diagnostic Unit Public Health Laboratory and Victorian Infectious Diseases Reference Laboratory, Doherty Institute |
| EPI_ISL_427030 | Victorian Infectious Diseases Reference Laboratory (VIDRL) | Microbiological Diagnostic Unit Public Health Laboratory and Victorian Infectious Diseases Reference Laboratory, Doherty Institute |
| EPI_ISL_427031 | Victorian Infectious Diseases Reference Laboratory (VIDRL) | Microbiological Diagnostic Unit Public Health Laboratory and Victorian Infectious Diseases Reference Laboratory, Doherty Institute |
| EPI_ISL_427032 | Victorian Infectious Diseases Reference Laboratory (VIDRL) | Microbiological Diagnostic Unit Public Health Laboratory and Victorian Infectious Diseases Reference Laboratory, Doherty Institute |
| EPI_ISL_427033 | Victorian Infectious Diseases Reference Laboratory (VIDRL) | Microbiological Diagnostic Unit Public Health Laboratory and Victorian Infectious Diseases Reference Laboratory, Doherty Institute |
| EPI_ISL_427034 | Victorian Infectious Diseases Reference Laboratory (VIDRL) | Microbiological Diagnostic Unit Public Health Laboratory and Victorian Infectious Diseases Reference Laboratory, Doherty Institute |

| EPI_Accession_ID | Originating Laboratory | Submitting Laboratory |
|---|---|---|
| EPI_ISL_427035 | Victorian Infectious Diseases Reference Laboratory (VIDRL) | Microbiological Diagnostic Unit Public Health Laboratory and Victorian Infectious Diseases Reference Laboratory, Doherty Institute |
| EPI_ISL_427036 | Victorian Infectious Diseases Reference Laboratory (VIDRL) | Microbiological Diagnostic Unit Public Health Laboratory and Victorian Infectious Diseases Reference Laboratory, Doherty Institute |
| EPI_ISL_427037 | Victorian Infectious Diseases Reference Laboratory (VIDRL) | Microbiological Diagnostic Unit Public Health Laboratory and Victorian Infectious Diseases Reference Laboratory, Doherty Institute |
| EPI_ISL_427038 | Victorian Infectious Diseases Reference Laboratory (VIDRL) | Microbiological Diagnostic Unit Public Health Laboratory and Victorian Infectious Diseases Reference Laboratory, Doherty Institute |
| EPI_ISL_427039 | Victorian Infectious Diseases Reference Laboratory (VIDRL) | Microbiological Diagnostic Unit Public Health Laboratory and Victorian Infectious Diseases Reference Laboratory, Doherty Institute |
| EPI_ISL_427040 | Victorian Infectious Diseases Reference Laboratory (VIDRL) | Microbiological Diagnostic Unit Public Health Laboratory and Victorian Infectious Diseases Reference Laboratory, Doherty Institute |
| EPI_ISL_427041 | Victorian Infectious Diseases Reference Laboratory (VIDRL) | Microbiological Diagnostic Unit Public Health Laboratory and Victorian Infectious Diseases Reference Laboratory, Doherty Institute |
| EPI_ISL_427042 | Victorian Infectious Diseases Reference Laboratory (VIDRL) | Microbiological Diagnostic Unit Public Health Laboratory and Victorian Infectious Diseases Reference Laboratory, Doherty Institute |
| EPI_ISL_427043 | Laboratory of Microbiology, Medical School, National and Kapodistrian University of Athens | Laboratory of Biology, Department of Medicine, Democritus University of Thrace |
| EPI_ISL_427044 | Victorian Infectious Diseases Reference Laboratory (VIDRL) | Microbiological Diagnostic Unit Public Health Laboratory and Victorian Infectious Diseases Reference Laboratory, Doherty Institute |
| EPI_ISL_427045 | Victorian Infectious Diseases Reference Laboratory (VIDRL) | Microbiological Diagnostic Unit Public Health Laboratory and Victorian Infectious Diseases Reference Laboratory, Doherty Institute |

| EPI_Accession_ID | Originating Laboratory | Submitting Laboratory |
|---|---|---|
| EPI_ISL_427046 | Victorian Infectious Diseases Reference Laboratory (VIDRL) | Microbiological Diagnostic Unit Public Health Laboratory and Victorian Infectious Diseases Reference Laboratory, Doherty Institute |
| EPI_ISL_427047 | Victorian Infectious Diseases Reference Laboratory (VIDRL) | Microbiological Diagnostic Unit Public Health Laboratory and Victorian Infectious Diseases Reference Laboratory, Doherty Institute |
| EPI_ISL_427048 | Victorian Infectious Diseases Reference Laboratory (VIDRL) | Microbiological Diagnostic Unit Public Health Laboratory and Victorian Infectious Diseases Reference Laboratory, Doherty Institute |
| EPI_ISL_427049 | Victorian Infectious Diseases Reference Laboratory (VIDRL) | Microbiological Diagnostic Unit Public Health Laboratory and Victorian Infectious Diseases Reference Laboratory, Doherty Institute |
| EPI_ISL_427050 | Victorian Infectious Diseases Reference Laboratory (VIDRL) | Microbiological Diagnostic Unit Public Health Laboratory and Victorian Infectious Diseases Reference Laboratory, Doherty Institute |
| EPI_ISL_427051 | Victorian Infectious Diseases Reference Laboratory (VIDRL) | Microbiological Diagnostic Unit Public Health Laboratory and Victorian Infectious Diseases Reference Laboratory, Doherty Institute |
| EPI_ISL_427052 | Victorian Infectious Diseases Reference Laboratory (VIDRL) | Microbiological Diagnostic Unit Public Health Laboratory and Victorian Infectious Diseases Reference Laboratory, Doherty Institute |
| EPI_ISL_427053 | Victorian Infectious Diseases Reference Laboratory (VIDRL) | Microbiological Diagnostic Unit Public Health Laboratory and Victorian Infectious Diseases Reference Laboratory, Doherty Institute |
| EPI_ISL_427054 | Microbiological Diagnostic Unit Public Health Laboratory | Microbiological Diagnostic Unit Public Health Laboratory |
| EPI_ISL_427055 | Microbiological Diagnostic Unit Public Health Laboratory | Microbiological Diagnostic Unit Public Health Laboratory |
| EPI_ISL_427056 | Microbiological Diagnostic Unit Public Health Laboratory | Microbiological Diagnostic Unit Public Health Laboratory |
| EPI_ISL_427057 | Microbiological Diagnostic Unit Public Health Laboratory | Microbiological Diagnostic Unit Public Health Laboratory |

| EPI_Accession_ID | Originating Laboratory | Submitting Laboratory |
|---|---|---|
| EPI_ISL_427058 | Microbiological Diagnostic Unit Public Health Laboratory | Microbiological Diagnostic Unit Public Health Laboratory |
| EPI_ISL_427059 | Microbiological Diagnostic Unit Public Health Laboratory | Microbiological Diagnostic Unit Public Health Laboratory |
| EPI_ISL_427060 | Microbiological Diagnostic Unit Public Health Laboratory | Microbiological Diagnostic Unit Public Health Laboratory |
| EPI_ISL_427061 | Microbiological Diagnostic Unit Public Health Laboratory | Microbiological Diagnostic Unit Public Health Laboratory |
| EPI_ISL_427062 | Microbiological Diagnostic Unit Public Health Laboratory | Microbiological Diagnostic Unit Public Health Laboratory |
| EPI_ISL_427063 | Microbiological Diagnostic Unit Public Health Laboratory | Microbiological Diagnostic Unit Public Health Laboratory |
| EPI_ISL_427064 | Microbiological Diagnostic Unit Public Health Laboratory | Microbiological Diagnostic Unit Public Health Laboratory |
| EPI_ISL_427065 | Microbiological Diagnostic Unit Public Health Laboratory | Microbiological Diagnostic Unit Public Health Laboratory |
| EPI_ISL_427066 | Microbiological Diagnostic Unit Public Health Laboratory | Microbiological Diagnostic Unit Public Health Laboratory |
| EPI_ISL_427067 | Microbiological Diagnostic Unit Public Health Laboratory | Microbiological Diagnostic Unit Public Health Laboratory |
| EPI_ISL_427068 | Microbiological Diagnostic Unit Public Health Laboratory | Microbiological Diagnostic Unit Public Health Laboratory |
| EPI_ISL_427069 | Microbiological Diagnostic Unit Public Health Laboratory | Microbiological Diagnostic Unit Public Health Laboratory |
| EPI_ISL_427070 | Microbiological Diagnostic Unit Public Health Laboratory | Microbiological Diagnostic Unit Public Health Laboratory |
| EPI_ISL_427071 | Microbiological Diagnostic Unit Public Health Laboratory | Microbiological Diagnostic Unit Public Health Laboratory |
| EPI_ISL_427072 | Microbiological Diagnostic Unit Public Health Laboratory | Microbiological Diagnostic Unit Public Health Laboratory |
| EPI_ISL_427073 | Microbiological Diagnostic Unit Public Health Laboratory | Microbiological Diagnostic Unit Public Health Laboratory |

| EPI_Accession_ID | Originating Laboratory | Submitting Laboratory |
|---|---|---|
| EPI_ISL_427074 | Microbiological Diagnostic Unit Public Health Laboratory | Microbiological Diagnostic Unit Public Health Laboratory |
| EPI_ISL_427075 | Microbiological Diagnostic Unit Public Health Laboratory | Microbiological Diagnostic Unit Public Health Laboratory |
| EPI_ISL_427076 | Microbiological Diagnostic Unit Public Health Laboratory | Microbiological Diagnostic Unit Public Health Laboratory |
| EPI_ISL_427077 | Microbiological Diagnostic Unit Public Health Laboratory | Microbiological Diagnostic Unit Public Health Laboratory |
| EPI_ISL_427078 | Microbiological Diagnostic Unit Public Health Laboratory | Microbiological Diagnostic Unit Public Health Laboratory |
| EPI_ISL_427079 | Victorian Infectious Diseases Reference Laboratory (VIDRL) | Microbiological Diagnostic Unit Public Health Laboratory and Victorian Infectious Diseases Reference Laboratory, Doherty Institute |
| EPI_ISL_427080 | Victorian Infectious Diseases Reference Laboratory (VIDRL) | Microbiological Diagnostic Unit Public Health Laboratory and Victorian Infectious Diseases Reference Laboratory, Doherty Institute |
| EPI_ISL_427081 | Victorian Infectious Diseases Reference Laboratory (VIDRL) | Microbiological Diagnostic Unit Public Health Laboratory and Victorian Infectious Diseases Reference Laboratory, Doherty Institute |
| EPI_ISL_427082 | Victorian Infectious Diseases Reference Laboratory (VIDRL) | Microbiological Diagnostic Unit Public Health Laboratory and Victorian Infectious Diseases Reference Laboratory, Doherty Institute |
| EPI_ISL_427083 | Victorian Infectious Diseases Reference Laboratory (VIDRL) | Microbiological Diagnostic Unit Public Health Laboratory and Victorian Infectious Diseases Reference Laboratory, Doherty Institute |
| EPI_ISL_427084 | Victorian Infectious Diseases Reference Laboratory (VIDRL) | Microbiological Diagnostic Unit Public Health Laboratory and Victorian Infectious Diseases Reference Laboratory, Doherty Institute |
| EPI_ISL_427085 | Victorian Infectious Diseases Reference Laboratory (VIDRL) | Microbiological Diagnostic Unit Public Health Laboratory and Victorian Infectious Diseases Reference Laboratory, Doherty Institute |

| EPI_Accession_ID | Originating Laboratory | Submitting Laboratory |
|---|---|---|
| EPI_ISL_427086 | Victorian Infectious Diseases Reference Laboratory (VIDRL) | Microbiological Diagnostic Unit Public Health Laboratory and Victorian Infectious Diseases Reference Laboratory, Doherty Institute |
| EPI_ISL_427087 | Victorian Infectious Diseases Reference Laboratory (VIDRL) | Microbiological Diagnostic Unit Public Health Laboratory and Victorian Infectious Diseases Reference Laboratory, Doherty Institute |
| EPI_ISL_427088 | Victorian Infectious Diseases Reference Laboratory (VIDRL) | Microbiological Diagnostic Unit Public Health Laboratory and Victorian Infectious Diseases Reference Laboratory, Doherty Institute |
| EPI_ISL_427089 | Victorian Infectious Diseases Reference Laboratory (VIDRL) | Microbiological Diagnostic Unit Public Health Laboratory and Victorian Infectious Diseases Reference Laboratory, Doherty Institute |
| EPI_ISL_427090 | Victorian Infectious Diseases Reference Laboratory (VIDRL) | Microbiological Diagnostic Unit Public Health Laboratory and Victorian Infectious Diseases Reference Laboratory, Doherty Institute |
| EPI_ISL_427091 | Victorian Infectious Diseases Reference Laboratory (VIDRL) | Microbiological Diagnostic Unit Public Health Laboratory and Victorian Infectious Diseases Reference Laboratory, Doherty Institute |
| EPI_ISL_427092 | Victorian Infectious Diseases Reference Laboratory (VIDRL) | Microbiological Diagnostic Unit Public Health Laboratory and Victorian Infectious Diseases Reference Laboratory, Doherty Institute |
| EPI_ISL_427093 | Victorian Infectious Diseases Reference Laboratory (VIDRL) | Microbiological Diagnostic Unit Public Health Laboratory and Victorian Infectious Diseases Reference Laboratory, Doherty Institute |
| EPI_ISL_427094 | Victorian Infectious Diseases Reference Laboratory (VIDRL) | Microbiological Diagnostic Unit Public Health Laboratory and Victorian Infectious Diseases Reference Laboratory, Doherty Institute |
| EPI_ISL_427095 | Victorian Infectious Diseases Reference Laboratory (VIDRL) | Microbiological Diagnostic Unit Public Health Laboratory and Victorian Infectious Diseases Reference Laboratory, Doherty Institute |
| EPI_ISL_427096 | Victorian Infectious Diseases Reference Laboratory (VIDRL) | Microbiological Diagnostic Unit Public Health Laboratory and Victorian Infectious Diseases Reference Laboratory, Doherty Institute |

| EPI_Accession_ID | Originating Laboratory | Submitting Laboratory |
|---|---|---|
| EPI_ISL_427097 | Victorian Infectious Diseases Reference Laboratory (VIDRL) | Microbiological Diagnostic Unit Public Health Laboratory and Victorian Infectious Diseases Reference Laboratory, Doherty Institute |
| EPI_ISL_427099 | Victorian Infectious Diseases Reference Laboratory (VIDRL) | Microbiological Diagnostic Unit Public Health Laboratory and Victorian Infectious Diseases Reference Laboratory, Doherty Institute |
| EPI_ISL_427100 | Victorian Infectious Diseases Reference Laboratory (VIDRL) | Microbiological Diagnostic Unit Public Health Laboratory and Victorian Infectious Diseases Reference Laboratory, Doherty Institute |
| EPI_ISL_427101 | Victorian Infectious Diseases Reference Laboratory (VIDRL) | Microbiological Diagnostic Unit Public Health Laboratory and Victorian Infectious Diseases Reference Laboratory, Doherty Institute |
| EPI_ISL_427102 | Victorian Infectious Diseases Reference Laboratory (VIDRL) | Microbiological Diagnostic Unit Public Health Laboratory and Victorian Infectious Diseases Reference Laboratory, Doherty Institute |
| EPI_ISL_427103 | Victorian Infectious Diseases Reference Laboratory (VIDRL) | Microbiological Diagnostic Unit Public Health Laboratory and Victorian Infectious Diseases Reference Laboratory, Doherty Institute |
| EPI_ISL_427104 | Victorian Infectious Diseases Reference Laboratory (VIDRL) | Microbiological Diagnostic Unit Public Health Laboratory and Victorian Infectious Diseases Reference Laboratory, Doherty Institute |
| EPI_ISL_427105 | Victorian Infectious Diseases Reference Laboratory (VIDRL) | Microbiological Diagnostic Unit Public Health Laboratory and Victorian Infectious Diseases Reference Laboratory, Doherty Institute |
| EPI_ISL_427106 | Victorian Infectious Diseases Reference Laboratory (VIDRL) | Microbiological Diagnostic Unit Public Health Laboratory and Victorian Infectious Diseases Reference Laboratory, Doherty Institute |
| EPI_ISL_427107 | Victorian Infectious Diseases Reference Laboratory (VIDRL) | Microbiological Diagnostic Unit Public Health Laboratory and Victorian Infectious Diseases Reference Laboratory, Doherty Institute |
| EPI_ISL_427108 | Victorian Infectious Diseases Reference Laboratory (VIDRL) | Microbiological Diagnostic Unit Public Health Laboratory and Victorian Infectious Diseases Reference Laboratory, Doherty Institute |

| EPI_Accession_ID | Originating Laboratory | Submitting Laboratory |
|---|---|---|
| EPI_ISL_427109 | Victorian Infectious Diseases Reference Laboratory (VIDRL) | Microbiological Diagnostic Unit Public Health Laboratory and Victorian Infectious Diseases Reference Laboratory, Doherty Institute |
| EPI_ISL_427110 | Victorian Infectious Diseases Reference Laboratory (VIDRL) | Microbiological Diagnostic Unit Public Health Laboratory and Victorian Infectious Diseases Reference Laboratory, Doherty Institute |
| EPI_ISL_427111 | Victorian Infectious Diseases Reference Laboratory (VIDRL) | Microbiological Diagnostic Unit Public Health Laboratory and Victorian Infectious Diseases Reference Laboratory, Doherty Institute |
| EPI_ISL_427112 | Victorian Infectious Diseases Reference Laboratory (VIDRL) | Microbiological Diagnostic Unit Public Health Laboratory and Victorian Infectious Diseases Reference Laboratory, Doherty Institute |
| EPI_ISL_427113 | Victorian Infectious Diseases Reference Laboratory (VIDRL) | Microbiological Diagnostic Unit Public Health Laboratory and Victorian Infectious Diseases Reference Laboratory, Doherty Institute |
| EPI_ISL_427114 | Victorian Infectious Diseases Reference Laboratory (VIDRL) | Microbiological Diagnostic Unit Public Health Laboratory and Victorian Infectious Diseases Reference Laboratory, Doherty Institute |
| EPI_ISL_427115 | Victorian Infectious Diseases Reference Laboratory (VIDRL) | Microbiological Diagnostic Unit Public Health Laboratory and Victorian Infectious Diseases Reference Laboratory, Doherty Institute |
| EPI_ISL_427116 | Victorian Infectious Diseases Reference Laboratory (VIDRL) | Microbiological Diagnostic Unit Public Health Laboratory and Victorian Infectious Diseases Reference Laboratory, Doherty Institute |
| EPI_ISL_427117 | Victorian Infectious Diseases Reference Laboratory (VIDRL) | Microbiological Diagnostic Unit Public Health Laboratory and Victorian Infectious Diseases Reference Laboratory, Doherty Institute |
| EPI_ISL_427118 | Victorian Infectious Diseases Reference Laboratory (VIDRL) | Microbiological Diagnostic Unit Public Health Laboratory and Victorian Infectious Diseases Reference Laboratory, Doherty Institute |
| EPI_ISL_427119 | Victorian Infectious Diseases Reference Laboratory (VIDRL) | Microbiological Diagnostic Unit Public Health Laboratory and Victorian Infectious Diseases Reference Laboratory, Doherty Institute |

| EPI_Accession_ID | Originating Laboratory | Submitting Laboratory |
|---|---|---|
| EPI_ISL_427120 | Victorian Infectious Diseases Reference Laboratory (VIDRL) | Microbiological Diagnostic Unit Public Health Laboratory and Victorian Infectious Diseases Reference Laboratory, Doherty Institute |
| EPI_ISL_427121 | Victorian Infectious Diseases Reference Laboratory (VIDRL) | Microbiological Diagnostic Unit Public Health Laboratory and Victorian Infectious Diseases Reference Laboratory, Doherty Institute |
| EPI_ISL_427122 | Victorian Infectious Diseases Reference Laboratory (VIDRL) | Microbiological Diagnostic Unit Public Health Laboratory and Victorian Infectious Diseases Reference Laboratory, Doherty Institute |
| EPI_ISL_427123 | Victorian Infectious Diseases Reference Laboratory (VIDRL) | Microbiological Diagnostic Unit Public Health Laboratory and Victorian Infectious Diseases Reference Laboratory, Doherty Institute |
| EPI_ISL_427124 | Victorian Infectious Diseases Reference Laboratory (VIDRL) | Microbiological Diagnostic Unit Public Health Laboratory and Victorian Infectious Diseases Reference Laboratory, Doherty Institute |
| EPI_ISL_427125 | Victorian Infectious Diseases Reference Laboratory (VIDRL) | Microbiological Diagnostic Unit Public Health Laboratory and Victorian Infectious Diseases Reference Laboratory, Doherty Institute |
| EPI_ISL_427126 | Victorian Infectious Diseases Reference Laboratory (VIDRL) | Microbiological Diagnostic Unit Public Health Laboratory and Victorian Infectious Diseases Reference Laboratory, Doherty Institute |
| EPI_ISL_427127 | Victorian Infectious Diseases Reference Laboratory (VIDRL) | Microbiological Diagnostic Unit Public Health Laboratory and Victorian Infectious Diseases Reference Laboratory, Doherty Institute |
| EPI_ISL_427128 | Victorian Infectious Diseases Reference Laboratory (VIDRL) | Microbiological Diagnostic Unit Public Health Laboratory and Victorian Infectious Diseases Reference Laboratory, Doherty Institute |
| EPI_ISL_427129 | Victorian Infectious Diseases Reference Laboratory (VIDRL) | Microbiological Diagnostic Unit Public Health Laboratory and Victorian Infectious Diseases Reference Laboratory, Doherty Institute |
| EPI_ISL_427130 | Victorian Infectious Diseases Reference Laboratory (VIDRL) | Microbiological Diagnostic Unit Public Health Laboratory and Victorian Infectious Diseases Reference Laboratory, Doherty Institute |

| EPI_Accession_ID | Originating Laboratory | Submitting Laboratory |
|---|---|---|
| EPI_ISL_427131 | Victorian Infectious Diseases Reference Laboratory (VIDRL) | Microbiological Diagnostic Unit Public Health Laboratory and Victorian Infectious Diseases Reference Laboratory, Doherty Institute |
| EPI_ISL_427132 | Victorian Infectious Diseases Reference Laboratory (VIDRL) | Microbiological Diagnostic Unit Public Health Laboratory and Victorian Infectious Diseases Reference Laboratory, Doherty Institute |
| EPI_ISL_427133 | Microbiological Diagnostic Unit Public Health Laboratory | Microbiological Diagnostic Unit Public Health Laboratory |
| EPI_ISL_427134 | Victorian Infectious Diseases Reference Laboratory (VIDRL) | Microbiological Diagnostic Unit Public Health Laboratory and Victorian Infectious Diseases Reference Laboratory, Doherty Institute |
| EPI_ISL_427135 | Victorian Infectious Diseases Reference Laboratory (VIDRL) | Microbiological Diagnostic Unit Public Health Laboratory and Victorian Infectious Diseases Reference Laboratory, Doherty Institute |
| EPI_ISL_427136 | Victorian Infectious Diseases Reference Laboratory (VIDRL) | Microbiological Diagnostic Unit Public Health Laboratory and Victorian Infectious Diseases Reference Laboratory, Doherty Institute |
| EPI_ISL_427137 | Victorian Infectious Diseases Reference Laboratory (VIDRL) | Microbiological Diagnostic Unit Public Health Laboratory and Victorian Infectious Diseases Reference Laboratory, Doherty Institute |
| EPI_ISL_427138 | Victorian Infectious Diseases Reference Laboratory (VIDRL) | Microbiological Diagnostic Unit Public Health Laboratory and Victorian Infectious Diseases Reference Laboratory, Doherty Institute |
| EPI_ISL_427139 | Victorian Infectious Diseases Reference Laboratory (VIDRL) | Microbiological Diagnostic Unit Public Health Laboratory and Victorian Infectious Diseases Reference Laboratory, Doherty Institute |
| EPI_ISL_427140 | Victorian Infectious Diseases Reference Laboratory (VIDRL) | Microbiological Diagnostic Unit Public Health Laboratory and Victorian Infectious Diseases Reference Laboratory, Doherty Institute |
| EPI_ISL_427141 | Victorian Infectious Diseases Reference Laboratory (VIDRL) | Microbiological Diagnostic Unit Public Health Laboratory and Victorian Infectious Diseases Reference Laboratory, Doherty Institute |

| EPI_Accession_ID | Originating Laboratory | Submitting Laboratory |
|---|---|---|
| EPI_ISL_427142 | Victorian Infectious Diseases Reference Laboratory (VIDRL) | Microbiological Diagnostic Unit Public Health Laboratory and Victorian Infectious Diseases Reference Laboratory, Doherty Institute |
| EPI_ISL_427143 | Victorian Infectious Diseases Reference Laboratory (VIDRL) | Microbiological Diagnostic Unit Public Health Laboratory and Victorian Infectious Diseases Reference Laboratory, Doherty Institute |
| EPI_ISL_427144 | Victorian Infectious Diseases Reference Laboratory (VIDRL) | Microbiological Diagnostic Unit Public Health Laboratory and Victorian Infectious Diseases Reference Laboratory, Doherty Institute |
| EPI_ISL_427145 | Victorian Infectious Diseases Reference Laboratory (VIDRL) | Microbiological Diagnostic Unit Public Health Laboratory and Victorian Infectious Diseases Reference Laboratory, Doherty Institute |
| EPI_ISL_427146 | Victorian Infectious Diseases Reference Laboratory (VIDRL) | Microbiological Diagnostic Unit Public Health Laboratory and Victorian Infectious Diseases Reference Laboratory, Doherty Institute |
| EPI_ISL_427147 | Victorian Infectious Diseases Reference Laboratory (VIDRL) | Microbiological Diagnostic Unit Public Health Laboratory and Victorian Infectious Diseases Reference Laboratory, Doherty Institute |
| EPI_ISL_427148 | Microbiological Diagnostic Unit Public Health Laboratory | Microbiological Diagnostic Unit Public Health Laboratory |
| EPI_ISL_427149 | Microbiological Diagnostic Unit Public Health Laboratory | Microbiological Diagnostic Unit Public Health Laboratory |
| EPI_ISL_427150 | Victorian Infectious Diseases Reference Laboratory (VIDRL) | Microbiological Diagnostic Unit Public Health Laboratory and Victorian Infectious Diseases Reference Laboratory, Doherty Institute |
| EPI_ISL_427151 | Victorian Infectious Diseases Reference Laboratory (VIDRL) | Microbiological Diagnostic Unit Public Health Laboratory and Victorian Infectious Diseases Reference Laboratory, Doherty Institute |
| EPI_ISL_427152 | Victorian Infectious Diseases Reference Laboratory (VIDRL) | Microbiological Diagnostic Unit Public Health Laboratory and Victorian Infectious Diseases Reference Laboratory, Doherty Institute |

| EPI_Accession_ID | Originating Laboratory | Submitting Laboratory |
|---|---|---|
| EPI_ISL_427153 | Victorian Infectious Diseases Reference Laboratory (VIDRL) | Microbiological Diagnostic Unit Public Health Laboratory and Victorian Infectious Diseases Reference Laboratory, Doherty Institute |
| EPI_ISL_427154 | Victorian Infectious Diseases Reference Laboratory (VIDRL) | Microbiological Diagnostic Unit Public Health Laboratory and Victorian Infectious Diseases Reference Laboratory, Doherty Institute |
| EPI_ISL_427155 | Victorian Infectious Diseases Reference Laboratory (VIDRL) | Microbiological Diagnostic Unit Public Health Laboratory and Victorian Infectious Diseases Reference Laboratory, Doherty Institute |
| EPI_ISL_427156 | Victorian Infectious Diseases Reference Laboratory (VIDRL) | Microbiological Diagnostic Unit Public Health Laboratory and Victorian Infectious Diseases Reference Laboratory, Doherty Institute |
| EPI_ISL_427157 | Victorian Infectious Diseases Reference Laboratory (VIDRL) | Microbiological Diagnostic Unit Public Health Laboratory and Victorian Infectious Diseases Reference Laboratory, Doherty Institute |
| EPI_ISL_427158 | Victorian Infectious Diseases Reference Laboratory (VIDRL) | Microbiological Diagnostic Unit Public Health Laboratory and Victorian Infectious Diseases Reference Laboratory, Doherty Institute |
| EPI_ISL_427159 | Victorian Infectious Diseases Reference Laboratory (VIDRL) | Microbiological Diagnostic Unit Public Health Laboratory and Victorian Infectious Diseases Reference Laboratory, Doherty Institute |
| EPI_ISL_427160 | Victorian Infectious Diseases Reference Laboratory (VIDRL) | Microbiological Diagnostic Unit Public Health Laboratory and Victorian Infectious Diseases Reference Laboratory, Doherty Institute |
| EPI_ISL_427161 | UW Virology Lab | UW Virology Lab |
| EPI_ISL_427162 | UW Virology Lab | UW Virology Lab |
| EPI_ISL_427163 | UW Virology Lab | UW Virology Lab |
| EPI_ISL_427164 | UW Virology Lab | UW Virology Lab |
| EPI_ISL_427165 | UW Virology Lab | UW Virology Lab |
| EPI_ISL_427166 | UW Virology Lab | UW Virology Lab |
| EPI_ISL_427167 | UW Virology Lab | UW Virology Lab |
| EPI_ISL_427169 | UW Virology Lab | UW Virology Lab |
| EPI_ISL_427170 | UW Virology Lab | UW Virology Lab |

| EPI_Accession_ID | Originating Laboratory | Submitting Laboratory |
|---|---|---|
| EPI_ISL_427172 | UW Virology Lab | UW Virology Lab |
| EPI_ISL_427173 | UW Virology Lab | UW Virology Lab |
| EPI_ISL_427174 | UW Virology Lab | UW Virology Lab |
| EPI_ISL_427175 | UW Virology Lab | UW Virology Lab |
| EPI_ISL_427176 | UW Virology Lab | UW Virology Lab |
| EPI_ISL_427177 | UW Virology Lab | UW Virology Lab |
| EPI_ISL_427178 | UW Virology Lab | UW Virology Lab |
| EPI_ISL_427179 | UW Virology Lab | UW Virology Lab |
| EPI_ISL_427180 | UW Virology Lab | UW Virology Lab |
| EPI_ISL_427181 | UW Virology Lab | UW Virology Lab |
| EPI_ISL_427182 | UW Virology Lab | UW Virology Lab |
| EPI_ISL_427183 | UW Virology Lab | UW Virology Lab |
| EPI_ISL_427184 | UW Virology Lab | UW Virology Lab |
| EPI_ISL_427185 | UW Virology Lab | UW Virology Lab |
| EPI_ISL_427186 | UW Virology Lab | UW Virology Lab |
| EPI_ISL_427187 | UW Virology Lab | UW Virology Lab |
| EPI_ISL_427188 | UW Virology Lab | UW Virology Lab |
| EPI_ISL_427189 | UW Virology Lab | UW Virology Lab |
| EPI_ISL_427190 | UW Virology Lab | UW Virology Lab |
| EPI_ISL_427191 | UW Virology Lab | UW Virology Lab |
| EPI_ISL_427192 | UW Virology Lab | UW Virology Lab |
| EPI_ISL_427193 | UW Virology Lab | UW Virology Lab |
| EPI_ISL_427194 | UW Virology Lab | UW Virology Lab |
| EPI_ISL_427195 | UW Virology Lab | UW Virology Lab |
| EPI_ISL_427196 | UW Virology Lab | UW Virology Lab |
| EPI_ISL_427197 | UW Virology Lab | UW Virology Lab |
| EPI_ISL_427198 | UW Virology Lab | UW Virology Lab |
| EPI_ISL_427199 | UW Virology Lab | UW Virology Lab |
| EPI_ISL_427200 | UW Virology Lab | UW Virology Lab |
| EPI_ISL_427201 | UW Virology Lab | UW Virology Lab |
| EPI_ISL_427202 | UW Virology Lab | UW Virology Lab |
| EPI_ISL_427203 | UW Virology Lab | UW Virology Lab |
| EPI_ISL_427204 | UW Virology Lab | UW Virology Lab |

| EPI_Accession_ID | Originating Laboratory | Submitting Laboratory |
|---|---|---|
| EPI_ISL_427205 | UW Virology Lab | UW Virology Lab |
| EPI_ISL_427206 | UW Virology Lab | UW Virology Lab |
| EPI_ISL_427207 | UW Virology Lab | UW Virology Lab |
| EPI_ISL_427208 | UW Virology Lab | UW Virology Lab |
| EPI_ISL_427209 | UW Virology Lab | UW Virology Lab |
| EPI_ISL_427210 | UW Virology Lab | UW Virology Lab |
| EPI_ISL_427211 | UW Virology Lab | UW Virology Lab |
| EPI_ISL_427212 | UW Virology Lab | UW Virology Lab |
| EPI_ISL_427213 | UW Virology Lab | UW Virology Lab |
| EPI_ISL_427214 | UW Virology Lab | UW Virology Lab |
| EPI_ISL_427215 | UW Virology Lab | UW Virology Lab |
| EPI_ISL_427216 | UW Virology Lab | UW Virology Lab |
| EPI_ISL_427217 | UW Virology Lab | UW Virology Lab |
| EPI_ISL_427218 | UW Virology Lab | UW Virology Lab |
| EPI_ISL_427219 | UW Virology Lab | UW Virology Lab |
| EPI_ISL_427220 | UW Virology Lab | UW Virology Lab |
| EPI_ISL_427221 | UW Virology Lab | UW Virology Lab |
| EPI_ISL_427222 | UW Virology Lab | UW Virology Lab |
| EPI_ISL_427223 | UW Virology Lab | UW Virology Lab |
| EPI_ISL_427224 | UW Virology Lab | UW Virology Lab |
| EPI_ISL_427225 | UW Virology Lab | UW Virology Lab |
| EPI_ISL_427226 | UW Virology Lab | UW Virology Lab |
| EPI_ISL_427227 | UW Virology Lab | UW Virology Lab |
| EPI_ISL_427228 | UW Virology Lab | UW Virology Lab |
| EPI_ISL_427229 | UW Virology Lab | UW Virology Lab |
| EPI_ISL_427230 | UW Virology Lab | UW Virology Lab |
| EPI_ISL_427231 | UW Virology Lab | UW Virology Lab |
| EPI_ISL_427232 | UW Virology Lab | UW Virology Lab |
| EPI_ISL_427233 | UW Virology Lab | UW Virology Lab |
| EPI_ISL_427234 | UW Virology Lab | UW Virology Lab |
| EPI_ISL_427235 | UW Virology Lab | UW Virology Lab |
| EPI_ISL_427236 | UW Virology Lab | UW Virology Lab |
| EPI_ISL_427237 | UW Virology Lab | UW Virology Lab |

| EPI_Accession_ID | Originating Laboratory | Submitting Laboratory |
|---|---|---|
| EPI_ISL_427238 | UW Virology Lab | UW Virology Lab |
| EPI_ISL_427239 | UW Virology Lab | UW Virology Lab |
| EPI_ISL_427240 | UW Virology Lab | UW Virology Lab |
| EPI_ISL_427241 | UW Virology Lab | UW Virology Lab |
| EPI_ISL_427242 | UW Virology Lab | UW Virology Lab |
| EPI_ISL_427245 | UW Virology Lab | UW Virology Lab |
| EPI_ISL_427246 | UW Virology Lab | UW Virology Lab |
| EPI_ISL_427247 | UW Virology Lab | UW Virology Lab |
| EPI_ISL_427248 | UW Virology Lab | UW Virology Lab |
| EPI_ISL_427249 | UW Virology Lab | UW Virology Lab |
| EPI_ISL_427250 | UW Virology Lab | UW Virology Lab |
| EPI_ISL_427251 | UW Virology Lab | UW Virology Lab |
| EPI_ISL_427252 | UW Virology Lab | UW Virology Lab |
| EPI_ISL_427253 | UW Virology Lab | UW Virology Lab |
| EPI_ISL_427254 | UW Virology Lab | UW Virology Lab |
| EPI_ISL_427255 | UW Virology Lab | UW Virology Lab |
| EPI_ISL_427256 | UW Virology Lab | UW Virology Lab |
| EPI_ISL_427257 | UW Virology Lab | UW Virology Lab |
| EPI_ISL_427258 | UW Virology Lab | UW Virology Lab |
| EPI_ISL_427259 | UW Virology Lab | UW Virology Lab |
| EPI_ISL_427260 | UW Virology Lab | UW Virology Lab |
| EPI_ISL_427261 | UW Virology Lab | UW Virology Lab |
| EPI_ISL_427262 | UW Virology Lab | UW Virology Lab |
| EPI_ISL_427263 | UW Virology Lab | UW Virology Lab |
| EPI_ISL_427264 | UW Virology Lab | UW Virology Lab |
| EPI_ISL_427265 | UW Virology Lab | UW Virology Lab |
| EPI_ISL_427266 | UW Virology Lab | UW Virology Lab |
| EPI_ISL_427267 | UW Virology Lab | UW Virology Lab |
| EPI_ISL_427268 | UW Virology Lab | UW Virology Lab |
| EPI_ISL_427269 | UW Virology Lab | UW Virology Lab |
| EPI_ISL_427270 | UW Virology Lab | UW Virology Lab |
| EPI_ISL_427271 | AZ SPHL, Arizona Department of Health Services | TGen North |
| EPI_ISL_427272 | AZ SPHL, Arizona Department of Health Services | TGen North |

| EPI_Accession_ID | Originating Laboratory | Submitting Laboratory |
|---|---|---|
| EPI_ISL_427273 | Minnesota Department of Health, Public Health Laboratory | Minnesota Department of Health, Public Health Laboratory |
| EPI_ISL_427274 | Minnesota Department of Health, Public Health Laboratory | Minnesota Department of Health, Public Health Laboratory |
| EPI_ISL_427275 | Minnesota Department of Health, Public Health Laboratory | Minnesota Department of Health, Public Health Laboratory |
| EPI_ISL_427276 | Minnesota Department of Health, Public Health Laboratory | Minnesota Department of Health, Public Health Laboratory |
| EPI_ISL_427277 | Minnesota Department of Health, Public Health Laboratory | Minnesota Department of Health, Public Health Laboratory |
| EPI_ISL_427278 | Minnesota Department of Health, Public Health Laboratory | Minnesota Department of Health, Public Health Laboratory |
| EPI_ISL_427279 | Minnesota Department of Health, Public Health Laboratory | Minnesota Department of Health, Public Health Laboratory |
| EPI_ISL_427280 | Minnesota Department of Health, Public Health Laboratory | Minnesota Department of Health, Public Health Laboratory |
| EPI_ISL_427281 | Minnesota Department of Health, Public Health Laboratory | Minnesota Department of Health, Public Health Laboratory |
| EPI_ISL_427282 | Minnesota Department of Health, Public Health Laboratory | Minnesota Department of Health, Public Health Laboratory |
| EPI_ISL_427283 | Minnesota Department of Health, Public Health Laboratory | Minnesota Department of Health, Public Health Laboratory |
| EPI_ISL_427284 | Minnesota Department of Health, Public Health Laboratory | Minnesota Department of Health, Public Health Laboratory |
| EPI_ISL_427285 | Minnesota Department of Health, Public Health Laboratory | Minnesota Department of Health, Public Health Laboratory |
| EPI_ISL_427286 | Minnesota Department of Health, Public Health Laboratory | Minnesota Department of Health, Public Health Laboratory |
| EPI_ISL_427287 | Minnesota Department of Health, Public Health Laboratory | Minnesota Department of Health, Public Health Laboratory |
| EPI_ISL_427288 | The Ohio State University | The Ohio State University-James Molecular Lab at Polaris |

| EPI_Accession_ID | Originating Laboratory | Submitting Laboratory |
|---|---|---|
| EPI_ISL_427290 | The Ohio State University | The Ohio State University-James Molecular Lab at Polaris |
| EPI_ISL_427292 | LACEN-AL - Laboratorio Central de Alagoas | Instituto Oswaldo Cruz FIOCRUZ - Laboratory of Respiratory Viruses and Measles (LVRS) |
| EPI_ISL_427293 | LACEN-BA - LaboratÃ³rio Central de SaÃºde PÃºblica Professor GonÃ§alo Moniz | Instituto Oswaldo Cruz FIOCRUZ - Laboratory of Respiratory Viruses and Measles (LVRS) |
| EPI_ISL_427294 | Instituto Oswaldo Cruz FIOCRUZ - Laboratory of Respiratory Viruses and Measles (LVRS) | Instituto Oswaldo Cruz FIOCRUZ - Laboratory of Respiratory Viruses and Measles (LVRS) |
| EPI_ISL_427295 | Instituto Oswaldo Cruz FIOCRUZ - Laboratory of Respiratory Viruses and Measles (LVRS) | Instituto Oswaldo Cruz FIOCRUZ - Laboratory of Respiratory Viruses and Measles (LVRS) |
| EPI_ISL_427296 | Instituto Oswaldo Cruz FIOCRUZ - Laboratory of Respiratory Viruses and Measles (LVRS) | Instituto Oswaldo Cruz FIOCRUZ - Laboratory of Respiratory Viruses and Measles (LVRS) |
| EPI_ISL_427297 | Instituto Oswaldo Cruz FIOCRUZ - Laboratory of Respiratory Viruses and Measles (LVRS) | Instituto Oswaldo Cruz FIOCRUZ - Laboratory of Respiratory Viruses and Measles (LVRS) |
| EPI_ISL_427298 | Instituto Oswaldo Cruz FIOCRUZ - Laboratory of Respiratory Viruses and Measles (LVRS) | Instituto Oswaldo Cruz FIOCRUZ - Laboratory of Respiratory Viruses and Measles (LVRS) |
| EPI_ISL_427299 | Instituto Oswaldo Cruz FIOCRUZ - Laboratory of Respiratory Viruses and Measles (LVRS) | Instituto Oswaldo Cruz FIOCRUZ - Laboratory of Respiratory Viruses and Measles (LVRS) |
| EPI_ISL_427300 | Instituto Oswaldo Cruz FIOCRUZ - Laboratory of Respiratory Viruses and Measles (LVRS) | Instituto Oswaldo Cruz FIOCRUZ - Laboratory of Respiratory Viruses and Measles (LVRS) |
| EPI_ISL_427301 | Instituto Oswaldo Cruz FIOCRUZ - Laboratory of Respiratory Viruses and Measles (LVRS) | Instituto Oswaldo Cruz FIOCRUZ - Laboratory of Respiratory Viruses and Measles (LVRS) |
| EPI_ISL_427302 | Instituto Oswaldo Cruz FIOCRUZ - Laboratory of Respiratory Viruses and Measles (LVRS) | Instituto Oswaldo Cruz FIOCRUZ - Laboratory of Respiratory Viruses and Measles (LVRS) |
| EPI_ISL_427303 | Instituto Oswaldo Cruz FIOCRUZ - Laboratory of Respiratory Viruses and Measles (LVRS) | Instituto Oswaldo Cruz FIOCRUZ - Laboratory of Respiratory Viruses and Measles (LVRS) |
| EPI_ISL_427304 | Instituto Oswaldo Cruz FIOCRUZ - Laboratory of Respiratory Viruses and Measles (LVRS) | Instituto Oswaldo Cruz FIOCRUZ - Laboratory of Respiratory Viruses and Measles (LVRS) |
| EPI_ISL_427305 | LACEN-SC - Laboratorio Central de Santa Catarina | Instituto Oswaldo Cruz FIOCRUZ - Laboratory of Respiratory Viruses and Measles (LVRS) |
| EPI_ISL_427306 | LACEN-SC - Laboratorio Central de Santa Catarina | Instituto Oswaldo Cruz FIOCRUZ - Laboratory of Respiratory Viruses and Measles (LVRS) |
| EPI_ISL_427307 | WHO National Influenza Centre Russian Federation | WHO National Influenza Centre Russian Federation |

| EPI_Accession_ID | Originating Laboratory | Submitting Laboratory |
|---|---|---|
| EPI_ISL_427308 | WHO National Influenza Centre Russian Federation | WHO National Influenza Centre Russian Federation |
| EPI_ISL_427309 | WHO National Influenza Centre Russian Federation | WHO National Influenza Centre Russian Federation |
| EPI_ISL_427310 | WHO National Influenza Centre Russian Federation | WHO National Influenza Centre Russian Federation |
| EPI_ISL_427311 | WHO National Influenza Centre Russian Federation | WHO National Influenza Centre Russian Federation |
| EPI_ISL_427312 | WHO National Influenza Centre Russian Federation | WHO National Influenza Centre Russian Federation |
| EPI_ISL_427313 | WHO National Influenza Centre Russian Federation | WHO National Influenza Centre Russian Federation |
| EPI_ISL_427314 | WHO National Influenza Centre Russian Federation | WHO National Influenza Centre Russian Federation |
| EPI_ISL_427315 | WHO National Influenza Centre Russian Federation | WHO National Influenza Centre Russian Federation |
| EPI_ISL_427316 | WHO National Influenza Centre Russian Federation | WHO National Influenza Centre Russian Federation |
| EPI_ISL_427317 | WHO National Influenza Centre Russian Federation | WHO National Influenza Centre Russian Federation |
| EPI_ISL_427318 | WHO National Influenza Centre Russian Federation | WHO National Influenza Centre Russian Federation |
| EPI_ISL_427319 | WHO National Influenza Centre Russian Federation | WHO National Influenza Centre Russian Federation |
| EPI_ISL_427320 | WHO National Influenza Centre Russian Federation | WHO National Influenza Centre Russian Federation |
| EPI_ISL_427321 | WHO National Influenza Centre Russian Federation | WHO National Influenza Centre Russian Federation |
| EPI_ISL_427322 | WHO National Influenza Centre Russian Federation | WHO National Influenza Centre Russian Federation |
| EPI_ISL_427323 | WHO National Influenza Centre Russian Federation | WHO National Influenza Centre Russian Federation |
| EPI_ISL_427324 | WHO National Influenza Centre Russian Federation | WHO National Influenza Centre Russian Federation |
| EPI_ISL_427325 | WHO National Influenza Centre Russian Federation | WHO National Influenza Centre Russian Federation |
| EPI_ISL_427326 | WHO National Influenza Centre Russian Federation | WHO National Influenza Centre Russian Federation |
| EPI_ISL_427327 | WHO National Influenza Centre Russian Federation | WHO National Influenza Centre Russian Federation |
| EPI_ISL_427328 | WHO National Influenza Centre Russian Federation | WHO National Influenza Centre Russian Federation |
| EPI_ISL_427329 | WHO National Influenza Centre Russian Federation | WHO National Influenza Centre Russian Federation |
| EPI_ISL_427330 | WHO National Influenza Centre Russian Federation | WHO National Influenza Centre Russian Federation |
| EPI_ISL_427331 | WHO National Influenza Centre Russian Federation | WHO National Influenza Centre Russian Federation |
| EPI_ISL_427332 | WHO National Influenza Centre Russian Federation | WHO National Influenza Centre Russian Federation |
| EPI_ISL_427333 | WHO National Influenza Centre Russian Federation | WHO National Influenza Centre Russian Federation |
| EPI_ISL_427334 | WHO National Influenza Centre Russian Federation | WHO National Influenza Centre Russian Federation |
| EPI_ISL_427335 | WHO National Influenza Centre Russian Federation | WHO National Influenza Centre Russian Federation |
| EPI_ISL_427336 | WHO National Influenza Centre Russian Federation | WHO National Influenza Centre Russian Federation |
| EPI_ISL_427337 | WHO National Influenza Centre Russian Federation | WHO National Influenza Centre Russian Federation |
| EPI_ISL_427338 | WHO National Influenza Centre Russian Federation | WHO National Influenza Centre Russian Federation |
| EPI_ISL_427339 | WHO National Influenza Centre Russian Federation | WHO National Influenza Centre Russian Federation |
| EPI_ISL_427340 | Department of Clinical Microbiology | GIGA Medical Genomics |

| EPI_Accession_ID | Originating Laboratory | Submitting Laboratory |
|---|---|---|
| EPI_ISL_427341 | Department of Clinical Microbiology | GIGA Medical Genomics |
| EPI_ISL_427342 | Department of Clinical Microbiology | GIGA Medical Genomics |
| EPI_ISL_427343 | Department of Clinical Microbiology | GIGA Medical Genomics |
| EPI_ISL_427344 | Department of Clinical Microbiology | GIGA Medical Genomics |
| EPI_ISL_427345 | Department of Clinical Microbiology | GIGA Medical Genomics |
| EPI_ISL_427346 | Department of Clinical Microbiology | GIGA Medical Genomics |
| EPI_ISL_427347 | Department of Clinical Microbiology | GIGA Medical Genomics |
| EPI_ISL_427348 | Department of Clinical Microbiology | GIGA Medical Genomics |
| EPI_ISL_427349 | Department of Clinical Microbiology | GIGA Medical Genomics |
| EPI_ISL_427350 | Department of Clinical Microbiology | GIGA Medical Genomics |
| EPI_ISL_427351 | Department of Clinical Microbiology | GIGA Medical Genomics |
| EPI_ISL_427352 | Department of Clinical Microbiology | GIGA Medical Genomics |
| EPI_ISL_427353 | Department of Clinical Microbiology | GIGA Medical Genomics |
| EPI_ISL_427354 | Department of Clinical Microbiology | GIGA Medical Genomics |
| EPI_ISL_427355 | Department of Clinical Microbiology | GIGA Medical Genomics |
| EPI_ISL_427356 | Department of Clinical Microbiology | GIGA Medical Genomics |
| EPI_ISL_427357 | Department of Clinical Microbiology | GIGA Medical Genomics |
| EPI_ISL_427358 | Department of Clinical Microbiology | GIGA Medical Genomics |
| EPI_ISL_427359 | Department of Clinical Microbiology | GIGA Medical Genomics |
| EPI_ISL_427360 | Department of Clinical Microbiology | GIGA Medical Genomics |
| EPI_ISL_427361 | Department of Clinical Microbiology | GIGA Medical Genomics |
| EPI_ISL_427362 | Department of Clinical Microbiology | GIGA Medical Genomics |
| EPI_ISL_427363 | Department of Clinical Microbiology | GIGA Medical Genomics |
| EPI_ISL_427364 | Department of Clinical Microbiology | GIGA Medical Genomics |
| EPI_ISL_427365 | Department of Clinical Microbiology | GIGA Medical Genomics |
| EPI_ISL_427366 | Department of Clinical Microbiology | GIGA Medical Genomics |
| EPI_ISL_427367 | Department of Clinical Microbiology | GIGA Medical Genomics |
| EPI_ISL_427368 | Department of Clinical Microbiology | GIGA Medical Genomics |
| EPI_ISL_427369 | Department of Clinical Microbiology | GIGA Medical Genomics |
| EPI_ISL_427370 | Department of Clinical Microbiology | GIGA Medical Genomics |
| EPI_ISL_427371 | Department of Clinical Microbiology | GIGA Medical Genomics |
| EPI_ISL_427372 | Department of Clinical Microbiology | GIGA Medical Genomics |
| EPI_ISL_427373 | Department of Clinical Microbiology | GIGA Medical Genomics |

| EPI_Accession_ID | Originating Laboratory | Submitting Laboratory |
|---|---|---|
| EPI_ISL_427374 | Department of Clinical Microbiology | GIGA Medical Genomics |
| EPI_ISL_427375 | Department of Clinical Microbiology | GIGA Medical Genomics |
| EPI_ISL_427376 | Department of Clinical Microbiology | GIGA Medical Genomics |
| EPI_ISL_427377 | Department of Clinical Microbiology | GIGA Medical Genomics |
| EPI_ISL_427378 | Department of Clinical Microbiology | GIGA Medical Genomics |
| EPI_ISL_427379 | Department of Clinical Microbiology | GIGA Medical Genomics |
| EPI_ISL_427380 | Department of Clinical Microbiology | GIGA Medical Genomics |
| EPI_ISL_427381 | Department of Clinical Microbiology | GIGA Medical Genomics |
| EPI_ISL_427382 | Department of Clinical Microbiology | GIGA Medical Genomics |
| EPI_ISL_427383 | Department of Clinical Microbiology | GIGA Medical Genomics |
| EPI_ISL_427384 | Department of Clinical Microbiology | GIGA Medical Genomics |
| EPI_ISL_427385 | Department of Clinical Microbiology | GIGA Medical Genomics |
| EPI_ISL_427386 | Department of Clinical Microbiology | GIGA Medical Genomics |
| EPI_ISL_427387 | Department of Clinical Microbiology | GIGA Medical Genomics |
| EPI_ISL_427388 | Department of Clinical Microbiology | GIGA Medical Genomics |
| EPI_ISL_427389 | Department of Clinical Microbiology | GIGA Medical Genomics |
| EPI_ISL_427390 | Department of Clinical Microbiology | GIGA Medical Genomics |
| EPI_ISL_427391 | Genomic Laboratory (GLAB) (Conjoint lab of Health Directorate of Istanbul and Istanbul Technical University) | Genomic Laboratory (GLAB), Istanbul Technical University |
| EPI_ISL_427392 | TSGH-CP molecular lab | TSGH-CP molecular lab |
| EPI_ISL_427393 | TSGH-CP molecular lab | TSGH-CP molecular lab |
| EPI_ISL_427394 | TSGH-CP molecular lab | TSGH-CP molecular lab |
| EPI_ISL_427395 | TSGH-CP molecular lab | TSGH-CP molecular lab |
| EPI_ISL_427396 | TSGH-CP molecular lab | TSGH-CP molecular lab |
| EPI_ISL_427397 | TSGH-CP molecular lab | TSGH-CP molecular lab |
| EPI_ISL_427398 | TSGH-CP molecular lab | TSGH-CP molecular lab |
| EPI_ISL_427404 | Ministry of Public Health (MoPH) | Biomedical Research Center (BRC) |
| EPI_ISL_427405 | Ministry of Public Health (MoPH) | Biomedical Research Center (BRC) |
| EPI_ISL_427406 | Ministry of Public Health (MoPH) | Biomedical Research Center (BRC) |
| EPI_ISL_427407 | Ministry of Public Health (MoPH) | Biomedical Research Center (BRC) |
| EPI_ISL_427408 | Ministry of Public Health (MoPH) | Biomedical Research Center (BRC) |
| EPI_ISL_427410 | Ministry of Public Health (MoPH) | Biomedical Research Center (BRC) |

| EPI_Accession_ID | Originating Laboratory | Submitting Laboratory |
|---|---|---|
| EPI_ISL_427411 | Ministry of Public Health (MoPH) | Biomedical Research Center (BRC) |
| EPI_ISL_427412 | Ministry of Public Health (MoPH) | Biomedical Research Center (BRC) |
| EPI_ISL_427414 | Ministry of Public Health (MoPH) | Biomedical Research Center (BRC) |
| EPI_ISL_427415 | Ministry of Public Health (MoPH) | Biomedical Research Center (BRC) |
| EPI_ISL_427416 | Ministry of Public Health (MoPH) | Biomedical Research Center (BRC) |
| EPI_ISL_427417 | Ministry of Public Health (MoPH) | Biomedical Research Center (BRC) |
| EPI_ISL_427418 | Ministry of Public Health (MoPH) | Biomedical Research Center (BRC) |
| EPI_ISL_427419 | Ministry of Public Health (MoPH) | Biomedical Research Center (BRC) |
| EPI_ISL_427420 | Ministry of Public Health (MoPH) | Biomedical Research Center (BRC) |
| EPI_ISL_427427 | University of Wisconsin-Madison AIDS Vaccine Research Laboratories | University of Wisconsin-Madison AIDS Vaccine Research Laboratories |
| EPI_ISL_427428 | University of Wisconsin-Madison AIDS Vaccine Research Laboratories | University of Wisconsin-Madison AIDS Vaccine Research Laboratories |
| EPI_ISL_427429 | University of Wisconsin-Madison AIDS Vaccine Research Laboratories | University of Wisconsin-Madison AIDS Vaccine Research Laboratories |
| EPI_ISL_427430 | University of Wisconsin-Madison AIDS Vaccine Research Laboratories | University of Wisconsin-Madison AIDS Vaccine Research Laboratories |
| EPI_ISL_427431 | University of Wisconsin-Madison AIDS Vaccine Research Laboratories | University of Wisconsin-Madison AIDS Vaccine Research Laboratories |
| EPI_ISL_427432 | University of Wisconsin-Madison AIDS Vaccine Research Laboratories | University of Wisconsin-Madison AIDS Vaccine Research Laboratories |
| EPI_ISL_427433 | University of Wisconsin-Madison AIDS Vaccine Research Laboratories | University of Wisconsin-Madison AIDS Vaccine Research Laboratories |
| EPI_ISL_427434 | University of Wisconsin-Madison AIDS Vaccine Research Laboratories | University of Wisconsin-Madison AIDS Vaccine Research Laboratories |
| EPI_ISL_427435 | University of Wisconsin-Madison AIDS Vaccine Research Laboratories | University of Wisconsin-Madison AIDS Vaccine Research Laboratories |
| EPI_ISL_427436 | University of Wisconsin-Madison AIDS Vaccine Research Laboratories | University of Wisconsin-Madison AIDS Vaccine Research Laboratories |
| EPI_ISL_427437 | University of Wisconsin-Madison AIDS Vaccine Research Laboratories | University of Wisconsin-Madison AIDS Vaccine Research Laboratories |
| EPI_ISL_427438 | University of Wisconsin-Madison AIDS Vaccine Research Laboratories | University of Wisconsin-Madison AIDS Vaccine Research Laboratories |

| EPI_Accession_ID | Originating Laboratory | Submitting Laboratory |
|---|---|---|
| EPI_ISL_427439 | University of Wisconsin-Madison AIDS Vaccine Research Laboratories | University of Wisconsin-Madison AIDS Vaccine Research Laboratories |
| EPI_ISL_427440 | University of Wisconsin-Madison AIDS Vaccine Research Laboratories | University of Wisconsin-Madison AIDS Vaccine Research Laboratories |
| EPI_ISL_427441 | University of Wisconsin-Madison AIDS Vaccine Research Laboratories | University of Wisconsin-Madison AIDS Vaccine Research Laboratories |
| EPI_ISL_427442 | University of Wisconsin-Madison AIDS Vaccine Research Laboratories | University of Wisconsin-Madison AIDS Vaccine Research Laboratories |
| EPI_ISL_427443 | University of Wisconsin-Madison AIDS Vaccine Research Laboratories | University of Wisconsin-Madison AIDS Vaccine Research Laboratories |
| EPI_ISL_427444 | University of Wisconsin-Madison AIDS Vaccine Research Laboratories | University of Wisconsin-Madison AIDS Vaccine Research Laboratories |
| EPI_ISL_427445 | University of Wisconsin-Madison AIDS Vaccine Research Laboratories | University of Wisconsin-Madison AIDS Vaccine Research Laboratories |
| EPI_ISL_427446 | University of Wisconsin-Madison AIDS Vaccine Research Laboratories | University of Wisconsin-Madison AIDS Vaccine Research Laboratories |
| EPI_ISL_427447 | University of Wisconsin-Madison AIDS Vaccine Research Laboratories | University of Wisconsin-Madison AIDS Vaccine Research Laboratories |
| EPI_ISL_427448 | University of Wisconsin-Madison AIDS Vaccine Research Laboratories | University of Wisconsin-Madison AIDS Vaccine Research Laboratories |
| EPI_ISL_427449 | University of Wisconsin-Madison AIDS Vaccine Research Laboratories | University of Wisconsin-Madison AIDS Vaccine Research Laboratories |
| EPI_ISL_427450 | University of Wisconsin-Madison AIDS Vaccine Research Laboratories | University of Wisconsin-Madison AIDS Vaccine Research Laboratories |
| EPI_ISL_427451 | University of Wisconsin-Madison AIDS Vaccine Research Laboratories | University of Wisconsin-Madison AIDS Vaccine Research Laboratories |
| EPI_ISL_427452 | University of Wisconsin-Madison AIDS Vaccine Research Laboratories | University of Wisconsin-Madison AIDS Vaccine Research Laboratories |
| EPI_ISL_427453 | University of Wisconsin-Madison AIDS Vaccine Research Laboratories | University of Wisconsin-Madison AIDS Vaccine Research Laboratories |
| EPI_ISL_427454 | University of Wisconsin-Madison AIDS Vaccine Research Laboratories | University of Wisconsin-Madison AIDS Vaccine Research Laboratories |

| EPI_Accession_ID | Originating Laboratory | Submitting Laboratory |
|---|---|---|
| EPI_ISL_427455 | University of Wisconsin-Madison AIDS Vaccine Research Laboratories | University of Wisconsin-Madison AIDS Vaccine Research Laboratories |
| EPI_ISL_427456 | University of Wisconsin-Madison AIDS Vaccine Research Laboratories | University of Wisconsin-Madison AIDS Vaccine Research Laboratories |
| EPI_ISL_427457 | University of Wisconsin-Madison AIDS Vaccine Research Laboratories | University of Wisconsin-Madison AIDS Vaccine Research Laboratories |
| EPI_ISL_427458 | University of Wisconsin-Madison AIDS Vaccine Research Laboratories | University of Wisconsin-Madison AIDS Vaccine Research Laboratories |
| EPI_ISL_427459 | University of Wisconsin-Madison AIDS Vaccine Research Laboratories | University of Wisconsin-Madison AIDS Vaccine Research Laboratories |
| EPI_ISL_427460 | University of Wisconsin-Madison AIDS Vaccine Research Laboratories | University of Wisconsin-Madison AIDS Vaccine Research Laboratories |
| EPI_ISL_427461 | University of Wisconsin-Madison AIDS Vaccine Research Laboratories | University of Wisconsin-Madison AIDS Vaccine Research Laboratories |
| EPI_ISL_427462 | University of Wisconsin-Madison AIDS Vaccine Research Laboratories | University of Wisconsin-Madison AIDS Vaccine Research Laboratories |
| EPI_ISL_427469 | NYU Langone Health | Departments of Pathology and Medicine, New York University School of Medicine |
| EPI_ISL_427470 | NYU Langone Health | Departments of Pathology and Medicine, New York University School of Medicine |
| EPI_ISL_427471 | NYU Langone Health | Departments of Pathology and Medicine, New York University School of Medicine |
| EPI_ISL_427472 | NYU Langone Health | Departments of Pathology and Medicine, New York University School of Medicine |
| EPI_ISL_427473 | NYU Langone Health | Departments of Pathology and Medicine, New York University School of Medicine |
| EPI_ISL_427474 | NYU Langone Health | Departments of Pathology and Medicine, New York University School of Medicine |
| EPI_ISL_427475 | NYU Langone Health | Departments of Pathology and Medicine, New York University School of Medicine |
| EPI_ISL_427476 | NYU Langone Health | Departments of Pathology and Medicine, New York University School of Medicine |

| EPI_Accession_ID | Originating Laboratory | Submitting Laboratory |
|---|---|---|
| EPI_ISL_427477 | NYU Langone Health | Departments of Pathology and Medicine, New York University School of Medicine |
| EPI_ISL_427478 | NYU Langone Health | Departments of Pathology and Medicine, New York University School of Medicine |
| EPI_ISL_427479 | NYU Langone Health | Departments of Pathology and Medicine, New York University School of Medicine |
| EPI_ISL_427480 | NYU Langone Health | Departments of Pathology and Medicine, New York University School of Medicine |
| EPI_ISL_427481 | NYU Langone Health | Departments of Pathology and Medicine, New York University School of Medicine |
| EPI_ISL_427482 | NYU Langone Health | Departments of Pathology and Medicine, New York University School of Medicine |
| EPI_ISL_427483 | NYU Langone Health | Departments of Pathology and Medicine, New York University School of Medicine |
| EPI_ISL_427484 | NYU Langone Health | Departments of Pathology and Medicine, New York University School of Medicine |
| EPI_ISL_427485 | NYU Langone Health | Departments of Pathology and Medicine, New York University School of Medicine |
| EPI_ISL_427486 | NYU Langone Health | Departments of Pathology and Medicine, New York University School of Medicine |
| EPI_ISL_427487 | NYU Langone Health | Departments of Pathology and Medicine, New York University School of Medicine |
| EPI_ISL_427488 | NYU Langone Health | Departments of Pathology and Medicine, New York University School of Medicine |
| EPI_ISL_427489 | NYU Langone Health | Departments of Pathology and Medicine, New York University School of Medicine |
| EPI_ISL_427490 | NYU Langone Health | Departments of Pathology and Medicine, New York University School of Medicine |
| EPI_ISL_427491 | NYU Langone Health | Departments of Pathology and Medicine, New York University School of Medicine |
| EPI_ISL_427492 | NYU Langone Health | Departments of Pathology and Medicine, New York University School of Medicine |

| EPI_Accession_ID | Originating Laboratory | Submitting Laboratory |
|---|---|---|
| EPI_ISL_427493 | NYU Langone Health | Departments of Pathology and Medicine, New York University School of Medicine |
| EPI_ISL_427494 | NYU Langone Health | Departments of Pathology and Medicine, New York University School of Medicine |
| EPI_ISL_427495 | NYU Langone Health | Departments of Pathology and Medicine, New York University School of Medicine |
| EPI_ISL_427496 | NYU Langone Health | Departments of Pathology and Medicine, New York University School of Medicine |
| EPI_ISL_427497 | NYU Langone Health | Departments of Pathology and Medicine, New York University School of Medicine |
| EPI_ISL_427498 | NYU Langone Health | Departments of Pathology and Medicine, New York University School of Medicine |
| EPI_ISL_427499 | NYU Langone Health | Departments of Pathology and Medicine, New York University School of Medicine |
| EPI_ISL_427500 | NYU Langone Health | Departments of Pathology and Medicine, New York University School of Medicine |
| EPI_ISL_427501 | NYU Langone Health | Departments of Pathology and Medicine, New York University School of Medicine |
| EPI_ISL_427502 | NYU Langone Health | Departments of Pathology and Medicine, New York University School of Medicine |
| EPI_ISL_427503 | NYU Langone Health | Departments of Pathology and Medicine, New York University School of Medicine |
| EPI_ISL_427504 | NYU Langone Health | Departments of Pathology and Medicine, New York University School of Medicine |
| EPI_ISL_427505 | NYU Langone Health | Departments of Pathology and Medicine, New York University School of Medicine |
| EPI_ISL_427506 | NYU Langone Health | Departments of Pathology and Medicine, New York University School of Medicine |
| EPI_ISL_427507 | NYU Langone Health | Departments of Pathology and Medicine, New York University School of Medicine |
| EPI_ISL_427508 | NYU Langone Health | Departments of Pathology and Medicine, New York University School of Medicine |

| EPI_Accession_ID | Originating Laboratory | Submitting Laboratory |
|---|---|---|
| EPI_ISL_427509 | NYU Langone Health | Departments of Pathology and Medicine, New York University School of Medicine |
| EPI_ISL_427510 | NYU Langone Health | Departments of Pathology and Medicine, New York University School of Medicine |
| EPI_ISL_427511 | NYU Langone Health | Departments of Pathology and Medicine, New York University School of Medicine |
| EPI_ISL_427512 | NYU Langone Health | Departments of Pathology and Medicine, New York University School of Medicine |
| EPI_ISL_427513 | NYU Langone Health | Departments of Pathology and Medicine, New York University School of Medicine |
| EPI_ISL_427514 | NYU Langone Health | Departments of Pathology and Medicine, New York University School of Medicine |
| EPI_ISL_427515 | NYU Langone Health | Departments of Pathology and Medicine, New York University School of Medicine |
| EPI_ISL_427516 | NYU Langone Health | Departments of Pathology and Medicine, New York University School of Medicine |
| EPI_ISL_427517 | NYU Langone Health | Departments of Pathology and Medicine, New York University School of Medicine |
| EPI_ISL_427518 | NYU Langone Health | Departments of Pathology and Medicine, New York University School of Medicine |
| EPI_ISL_427519 | NYU Langone Health | Departments of Pathology and Medicine, New York University School of Medicine |
| EPI_ISL_427520 | NYU Langone Health | Departments of Pathology and Medicine, New York University School of Medicine |
| EPI_ISL_427521 | NYU Langone Health | Departments of Pathology and Medicine, New York University School of Medicine |
| EPI_ISL_427524 | NYU Langone Health | Departments of Pathology and Medicine, New York University School of Medicine |
| EPI_ISL_427525 | NYU Langone Health | Departments of Pathology and Medicine, New York University School of Medicine |
| EPI_ISL_427526 | NewYork-Presbyterian & Mason Lab | Mason Lab |
| EPI_ISL_427527 | NewYork-Presbyterian & Mason Lab | Mason Lab |
| EPI_ISL_427528 | NewYork-Presbyterian & Mason Lab | Mason Lab |

| EPI_Accession_ID | Originating Laboratory | Submitting Laboratory |
|---|---|---|
| EPI_ISL_427529 | NewYork-Presbyterian & Mason Lab | Mason Lab |
| EPI_ISL_427530 | NewYork-Presbyterian & Mason Lab | Mason Lab |
| EPI_ISL_427531 | NewYork-Presbyterian & Mason Lab | Mason Lab |
| EPI_ISL_427532 | NewYork-Presbyterian & Mason Lab | Mason Lab |
| EPI_ISL_427533 | NewYork-Presbyterian & Mason Lab | Mason Lab |
| EPI_ISL_427534 | NewYork-Presbyterian & Mason Lab | Mason Lab |
| EPI_ISL_427535 | NewYork-Presbyterian & Mason Lab | Mason Lab |
| EPI_ISL_427536 | NewYork-Presbyterian & Mason Lab | Mason Lab |
| EPI_ISL_427537 | NewYork-Presbyterian & Mason Lab | Mason Lab |
| EPI_ISL_427538 | NewYork-Presbyterian & Mason Lab | Mason Lab |
| EPI_ISL_427539 | NewYork-Presbyterian & Mason Lab | Mason Lab |
| EPI_ISL_427540 | NewYork-Presbyterian & Mason Lab | Mason Lab |
| EPI_ISL_427541 | NewYork-Presbyterian & Mason Lab | Mason Lab |
| EPI_ISL_427542 | NewYork-Presbyterian & Mason Lab | Mason Lab |
| EPI_ISL_427543 | NewYork-Presbyterian & Mason Lab | Mason Lab |
| EPI_ISL_427544 | NewYork-Presbyterian & Mason Lab | Mason Lab |
| EPI_ISL_427545 | NewYork-Presbyterian & Mason Lab | Mason Lab |
| EPI_ISL_427546 | NewYork-Presbyterian & Mason Lab | Mason Lab |
| EPI_ISL_427547 | NewYork-Presbyterian & Mason Lab | Mason Lab |
| EPI_ISL_427548 | NewYork-Presbyterian & Mason Lab | Mason Lab |
| EPI_ISL_427549 | NewYork-Presbyterian & Mason Lab | Mason Lab |
| EPI_ISL_427550 | NewYork-Presbyterian & Mason Lab | Mason Lab |
| EPI_ISL_427551 | NewYork-Presbyterian & Mason Lab | Mason Lab |
| EPI_ISL_427552 | NewYork-Presbyterian & Mason Lab | Mason Lab |
| EPI_ISL_427553 | NewYork-Presbyterian & Mason Lab | Mason Lab |
| EPI_ISL_427554 | NewYork-Presbyterian & Mason Lab | Mason Lab |
| EPI_ISL_427555 | NewYork-Presbyterian & Mason Lab | Mason Lab |
| EPI_ISL_427556 | NewYork-Presbyterian & Mason Lab | Mason Lab |
| EPI_ISL_427557 | NewYork-Presbyterian & Mason Lab | Mason Lab |
| EPI_ISL_427558 | NewYork-Presbyterian & Mason Lab | Mason Lab |
| EPI_ISL_427559 | NewYork-Presbyterian & Mason Lab | Mason Lab |
| EPI_ISL_427560 | NewYork-Presbyterian & Mason Lab | Mason Lab |
| EPI_ISL_427561 | NewYork-Presbyterian & Mason Lab | Mason Lab |

| EPI_Accession_ID | Originating Laboratory | Submitting Laboratory |
|---|---|---|
| EPI_ISL_427562 | NewYork-Presbyterian & Mason Lab | Mason Lab |
| EPI_ISL_427563 | NewYork-Presbyterian & Mason Lab | Mason Lab |
| EPI_ISL_427564 | NewYork-Presbyterian & Mason Lab | Mason Lab |
| EPI_ISL_427565 | NewYork-Presbyterian & Mason Lab | Mason Lab |
| EPI_ISL_427566 | NewYork-Presbyterian & Mason Lab | Mason Lab |
| EPI_ISL_427567 | NewYork-Presbyterian & Mason Lab | Mason Lab |
| EPI_ISL_427568 | NewYork-Presbyterian & Mason Lab | Mason Lab |
| EPI_ISL_427569 | NewYork-Presbyterian & Mason Lab | Mason Lab |
| EPI_ISL_427570 | NewYork-Presbyterian & Mason Lab | Mason Lab |
| EPI_ISL_427571 | NewYork-Presbyterian & Mason Lab | Mason Lab |
| EPI_ISL_427572 | NewYork-Presbyterian & Mason Lab | Mason Lab |
| EPI_ISL_427573 | NewYork-Presbyterian & Mason Lab | Mason Lab |
| EPI_ISL_427574 | NewYork-Presbyterian & Mason Lab | Mason Lab |
| EPI_ISL_427575 | NewYork-Presbyterian & Mason Lab | Mason Lab |
| EPI_ISL_427576 | NewYork-Presbyterian & Mason Lab | Mason Lab |
| EPI_ISL_427577 | NewYork-Presbyterian & Mason Lab | Mason Lab |
| EPI_ISL_427578 | NewYork-Presbyterian & Mason Lab | Mason Lab |
| EPI_ISL_427579 | NewYork-Presbyterian & Mason Lab | Mason Lab |
| EPI_ISL_427580 | NewYork-Presbyterian & Mason Lab | Mason Lab |
| EPI_ISL_427581 | NewYork-Presbyterian & Mason Lab | Mason Lab |
| EPI_ISL_427582 | NewYork-Presbyterian & Mason Lab | Mason Lab |
| EPI_ISL_427583 | NewYork-Presbyterian & Mason Lab | Mason Lab |
| EPI_ISL_427584 | NewYork-Presbyterian & Mason Lab | Mason Lab |
| EPI_ISL_427585 | NewYork-Presbyterian & Mason Lab | Mason Lab |
| EPI_ISL_427586 | NewYork-Presbyterian & Mason Lab | Mason Lab |
| EPI_ISL_427587 | NewYork-Presbyterian & Mason Lab | Mason Lab |
| EPI_ISL_427588 | NewYork-Presbyterian & Mason Lab | Mason Lab |
| EPI_ISL_427589 | NewYork-Presbyterian & Mason Lab | Mason Lab |
| EPI_ISL_427590 | NewYork-Presbyterian & Mason Lab | Mason Lab |
| EPI_ISL_427591 | NewYork-Presbyterian & Mason Lab | Mason Lab |
| EPI_ISL_427592 | NewYork-Presbyterian & Mason Lab | Mason Lab |
| EPI_ISL_427593 | NewYork-Presbyterian & Mason Lab | Mason Lab |
| EPI_ISL_427594 | NewYork-Presbyterian & Mason Lab | Mason Lab |

| EPI_Accession_ID | Originating Laboratory | Submitting Laboratory |
|---|---|---|
| EPI_ISL_427595 | NewYork-Presbyterian & Mason Lab | Mason Lab |
| EPI_ISL_427596 | NewYork-Presbyterian & Mason Lab | Mason Lab |
| EPI_ISL_427597 | NewYork-Presbyterian & Mason Lab | Mason Lab |
| EPI_ISL_427598 | NewYork-Presbyterian & Mason Lab | Mason Lab |
| EPI_ISL_427599 | NewYork-Presbyterian & Mason Lab | Mason Lab |
| EPI_ISL_427600 | NewYork-Presbyterian & Mason Lab | Mason Lab |
| EPI_ISL_427601 | NewYork-Presbyterian & Mason Lab | Mason Lab |
| EPI_ISL_427602 | NewYork-Presbyterian & Mason Lab | Mason Lab |
| EPI_ISL_427603 | NewYork-Presbyterian & Mason Lab | Mason Lab |
| EPI_ISL_427604 | NewYork-Presbyterian & Mason Lab | Mason Lab |
| EPI_ISL_427605 | NewYork-Presbyterian & Mason Lab | Mason Lab |
| EPI_ISL_427606 | NewYork-Presbyterian & Mason Lab | Mason Lab |
| EPI_ISL_427607 | NewYork-Presbyterian & Mason Lab | Mason Lab |
| EPI_ISL_427608 | NewYork-Presbyterian & Mason Lab | Mason Lab |
| EPI_ISL_427609 | NewYork-Presbyterian & Mason Lab | Mason Lab |
| EPI_ISL_427610 | NewYork-Presbyterian & Mason Lab | Mason Lab |
| EPI_ISL_427611 | NewYork-Presbyterian & Mason Lab | Mason Lab |
| EPI_ISL_427612 | NewYork-Presbyterian & Mason Lab | Mason Lab |
| EPI_ISL_427613 | NewYork-Presbyterian & Mason Lab | Mason Lab |
| EPI_ISL_427614 | NewYork-Presbyterian & Mason Lab | Mason Lab |
| EPI_ISL_427615 | NewYork-Presbyterian & Mason Lab | Mason Lab |
| EPI_ISL_427616 | NewYork-Presbyterian & Mason Lab | Mason Lab |
| EPI_ISL_427617 | NewYork-Presbyterian & Mason Lab | Mason Lab |
| EPI_ISL_427618 | NewYork-Presbyterian & Mason Lab | Mason Lab |
| EPI_ISL_427619 | Alaska State Virology Laboratory | Alaska State Virology Laboratory |
| EPI_ISL_427620 | Alaska State Virology Laboratory | Alaska State Virology Laboratory |
| EPI_ISL_427621 | Alaska State Virology Laboratory | Alaska State Virology Laboratory |
| EPI_ISL_427622 | Alaska State Virology Laboratory | Alaska State Virology Laboratory |
| EPI_ISL_427627 | NYU Langone Health | Departments of Pathology and Medicine, New York University School of Medicine |
| EPI_ISL_427628 | NYU Langone Health | Departments of Pathology and Medicine, New York University School of Medicine |

| EPI_Accession_ID | Originating Laboratory | Submitting Laboratory |
|---|---|---|
| EPI_ISL_427629 | NYU Langone Health | Departments of Pathology and Medicine, New York University School of Medicine |
| EPI_ISL_427630 | NYU Langone Health | Departments of Pathology and Medicine, New York University School of Medicine |
| EPI_ISL_427631 | NYU Langone Health | Departments of Pathology and Medicine, New York University School of Medicine |
| EPI_ISL_427632 | NYU Langone Health | Departments of Pathology and Medicine, New York University School of Medicine |
| EPI_ISL_427633 | NYU Langone Health | Departments of Pathology and Medicine, New York University School of Medicine |
| EPI_ISL_427634 | NYU Langone Health | Departments of Pathology and Medicine, New York University School of Medicine |
| EPI_ISL_427635 | NYU Langone Health | Departments of Pathology and Medicine, New York University School of Medicine |
| EPI_ISL_427636 | NYU Langone Health | Departments of Pathology and Medicine, New York University School of Medicine |
| EPI_ISL_427637 | NYU Langone Health | Departments of Pathology and Medicine, New York University School of Medicine |
| EPI_ISL_427638 | NYU Langone Health | Departments of Pathology and Medicine, New York University School of Medicine |
| EPI_ISL_427639 | NYU Langone Health | Departments of Pathology and Medicine, New York University School of Medicine |
| EPI_ISL_427640 | NYU Langone Health | Departments of Pathology and Medicine, New York University School of Medicine |
| EPI_ISL_427641 | NYU Langone Health | Departments of Pathology and Medicine, New York University School of Medicine |
| EPI_ISL_427642 | NYU Langone Health | Departments of Pathology and Medicine, New York University School of Medicine |
| EPI_ISL_427643 | Centre for Infectious Diseases and Microbiology Public Health | NSW Health Pathology - Institute of Clinical Pathology and Medical Research; Westmead Hospital; University of Sydney |

| EPI_Accession_ID | Originating Laboratory | Submitting Laboratory |
|---|---|---|
| EPI_ISL_427644 | Centre for Infectious Diseases and Microbiology Public Health | NSW Health Pathology - Institute of Clinical Pathology and Medical Research; Westmead Hospital; University of Sydney |
| EPI_ISL_427645 | Centre for Infectious Diseases and Microbiology Public Health | NSW Health Pathology - Institute of Clinical Pathology and Medical Research; Westmead Hospital; University of Sydney |
| EPI_ISL_427646 | Centre for Infectious Diseases and Microbiology Public Health | NSW Health Pathology - Institute of Clinical Pathology and Medical Research; Westmead Hospital; University of Sydney |
| EPI_ISL_427647 | Centre for Infectious Diseases and Microbiology Public Health | NSW Health Pathology - Institute of Clinical Pathology and Medical Research; Westmead Hospital; University of Sydney |
| EPI_ISL_427648 | Centre for Infectious Diseases and Microbiology Public Health | NSW Health Pathology - Institute of Clinical Pathology and Medical Research; Westmead Hospital; University of Sydney |
| EPI_ISL_427649 | Centre for Infectious Diseases and Microbiology Public Health | NSW Health Pathology - Institute of Clinical Pathology and Medical Research; Westmead Hospital; University of Sydney |
| EPI_ISL_427650 | Centre for Infectious Diseases and Microbiology Public Health | NSW Health Pathology - Institute of Clinical Pathology and Medical Research; Westmead Hospital; University of Sydney |
| EPI_ISL_427651 | Centre for Infectious Diseases and Microbiology Public Health | NSW Health Pathology - Institute of Clinical Pathology and Medical Research; Westmead Hospital; University of Sydney |
| EPI_ISL_427652 | Centre for Infectious Diseases and Microbiology Public Health | NSW Health Pathology - Institute of Clinical Pathology and Medical Research; Westmead Hospital; University of Sydney |
| EPI_ISL_427653 | Centre for Infectious Diseases and Microbiology Public Health | NSW Health Pathology - Institute of Clinical Pathology and Medical Research; Westmead Hospital; University of Sydney |
| EPI_ISL_427654 | Centre for Infectious Diseases and Microbiology Public Health | NSW Health Pathology - Institute of Clinical Pathology and Medical Research; Westmead Hospital; University of Sydney |

| EPI_Accession_ID | Originating Laboratory | Submitting Laboratory |
|---|---|---|
| EPI_ISL_427655 | Centre for Infectious Diseases and Microbiology Public Health | NSW Health Pathology - Institute of Clinical Pathology and Medical Research; Westmead Hospital; University of Sydney |
| EPI_ISL_427656 | Centre for Infectious Diseases and Microbiology Public Health | NSW Health Pathology - Institute of Clinical Pathology and Medical Research; Westmead Hospital; University of Sydney |
| EPI_ISL_427657 | Centre for Infectious Diseases and Microbiology Public Health | NSW Health Pathology - Institute of Clinical Pathology and Medical Research; Westmead Hospital; University of Sydney |
| EPI_ISL_427658 | Centre for Infectious Diseases and Microbiology Public Health | NSW Health Pathology - Institute of Clinical Pathology and Medical Research; Westmead Hospital; University of Sydney |
| EPI_ISL_427659 | Centre for Infectious Diseases and Microbiology Public Health | NSW Health Pathology - Institute of Clinical Pathology and Medical Research; Westmead Hospital; University of Sydney |
| EPI_ISL_427660 | Centre for Infectious Diseases and Microbiology Public Health | NSW Health Pathology - Institute of Clinical Pathology and Medical Research; Westmead Hospital; University of Sydney |
| EPI_ISL_427661 | Centre for Infectious Diseases and Microbiology Public Health | NSW Health Pathology - Institute of Clinical Pathology and Medical Research; Westmead Hospital; University of Sydney |
| EPI_ISL_427662 | Centre for Infectious Diseases and Microbiology Public Health | NSW Health Pathology - Institute of Clinical Pathology and Medical Research; Westmead Hospital; University of Sydney |
| EPI_ISL_427663 | Centre for Infectious Diseases and Microbiology Public Health | NSW Health Pathology - Institute of Clinical Pathology and Medical Research; Westmead Hospital; University of Sydney |
| EPI_ISL_427664 | Centre for Infectious Diseases and Microbiology Public Health | NSW Health Pathology - Institute of Clinical Pathology and Medical Research; Westmead Hospital; University of Sydney |
| EPI_ISL_427665 | Centre for Infectious Diseases and Microbiology Public Health | NSW Health Pathology - Institute of Clinical Pathology and Medical Research; Westmead Hospital; University of Sydney |

| EPI_Accession_ID | Originating Laboratory | Submitting Laboratory |
|---|---|---|
| EPI_ISL_427666 | Centre for Infectious Diseases and Microbiology Public Health | NSW Health Pathology - Institute of Clinical Pathology and Medical Research; Westmead Hospital; University of Sydney |
| EPI_ISL_427667 | Centre for Infectious Diseases and Microbiology Public Health | NSW Health Pathology - Institute of Clinical Pathology and Medical Research; Westmead Hospital; University of Sydney |
| EPI_ISL_427668 | Centre for Infectious Diseases and Microbiology Public Health | NSW Health Pathology - Institute of Clinical Pathology and Medical Research; Westmead Hospital; University of Sydney |
| EPI_ISL_427669 | Centre for Infectious Diseases and Microbiology Public Health | NSW Health Pathology - Institute of Clinical Pathology and Medical Research; Westmead Hospital; University of Sydney |
| EPI_ISL_427670 | Centre for Infectious Diseases and Microbiology Public Health | NSW Health Pathology - Institute of Clinical Pathology and Medical Research; Westmead Hospital; University of Sydney |
| EPI_ISL_427671 | Centre for Infectious Diseases and Microbiology Public Health | NSW Health Pathology - Institute of Clinical Pathology and Medical Research; Westmead Hospital; University of Sydney |
| EPI_ISL_427672 | Centre for Infectious Diseases and Microbiology Public Health | NSW Health Pathology - Institute of Clinical Pathology and Medical Research; Westmead Hospital; University of Sydney |
| EPI_ISL_427673 | Centre for Infectious Diseases and Microbiology Public Health | NSW Health Pathology - Institute of Clinical Pathology and Medical Research; Westmead Hospital; University of Sydney |
| EPI_ISL_427674 | Centre for Infectious Diseases and Microbiology Public Health | NSW Health Pathology - Institute of Clinical Pathology and Medical Research; Westmead Hospital; University of Sydney |
| EPI_ISL_427675 | Centre for Infectious Diseases and Microbiology Public Health | NSW Health Pathology - Institute of Clinical Pathology and Medical Research; Westmead Hospital; University of Sydney |
| EPI_ISL_427676 | Centre for Infectious Diseases and Microbiology Public Health | NSW Health Pathology - Institute of Clinical Pathology and Medical Research; Westmead Hospital; University of Sydney |

| EPI_Accession_ID | Originating Laboratory | Submitting Laboratory |
| --- | --- | --- |
| EPI_ISL_427677 | Centre for Infectious Diseases and Microbiology Public Health | NSW Health Pathology - Institute of Clinical Pathology and Medical Research; Westmead Hospital; University of Sydney |
| EPI_ISL_427678 | Centre for Infectious Diseases and Microbiology Public Health | NSW Health Pathology - Institute of Clinical Pathology and Medical Research; Westmead Hospital; University of Sydney |
| EPI_ISL_427679 | Centre for Infectious Diseases and Microbiology Public Health | NSW Health Pathology - Institute of Clinical Pathology and Medical Research; Westmead Hospital; University of Sydney |
| EPI_ISL_427680 | Centre for Infectious Diseases and Microbiology Public Health | NSW Health Pathology - Institute of Clinical Pathology and Medical Research; Westmead Hospital; University of Sydney |
| EPI_ISL_427681 | Centre for Infectious Diseases and Microbiology Public Health | NSW Health Pathology - Institute of Clinical Pathology and Medical Research; Westmead Hospital; University of Sydney |
| EPI_ISL_427682 | Centre for Infectious Diseases and Microbiology Public Health | NSW Health Pathology - Institute of Clinical Pathology and Medical Research; Westmead Hospital; University of Sydney |
| EPI_ISL_427683 | Centre for Infectious Diseases and Microbiology Public Health | NSW Health Pathology - Institute of Clinical Pathology and Medical Research; Westmead Hospital; University of Sydney |
| EPI_ISL_427684 | Centre for Infectious Diseases and Microbiology Public Health | NSW Health Pathology - Institute of Clinical Pathology and Medical Research; Westmead Hospital; University of Sydney |
| EPI_ISL_427685 | Centre for Infectious Diseases and Microbiology Public Health | NSW Health Pathology - Institute of Clinical Pathology and Medical Research; Westmead Hospital; University of Sydney |
| EPI_ISL_427686 | Centre for Infectious Diseases and Microbiology Public Health | NSW Health Pathology - Institute of Clinical Pathology and Medical Research; Westmead Hospital; University of Sydney |
| EPI_ISL_427687 | Centre for Infectious Diseases and Microbiology Public Health | NSW Health Pathology - Institute of Clinical Pathology and Medical Research; Westmead Hospital; University of Sydney |

| EPI_Accession_ID | Originating Laboratory | Submitting Laboratory |
|---|---|---|
| EPI_ISL_427688 | Centre for Infectious Diseases and Microbiology Public Health | NSW Health Pathology - Institute of Clinical Pathology and Medical Research; Westmead Hospital; University of Sydney |
| EPI_ISL_427689 | Centre for Infectious Diseases and Microbiology Public Health | NSW Health Pathology - Institute of Clinical Pathology and Medical Research; Westmead Hospital; University of Sydney |
| EPI_ISL_427690 | Centre for Infectious Diseases and Microbiology Public Health | NSW Health Pathology - Institute of Clinical Pathology and Medical Research; Westmead Hospital; University of Sydney |
| EPI_ISL_427691 | Centre for Infectious Diseases and Microbiology Public Health | NSW Health Pathology - Institute of Clinical Pathology and Medical Research; Westmead Hospital; University of Sydney |
| EPI_ISL_427692 | Centre for Infectious Diseases and Microbiology Public Health | NSW Health Pathology - Institute of Clinical Pathology and Medical Research; Westmead Hospital; University of Sydney |
| EPI_ISL_427693 | Centre for Infectious Diseases and Microbiology Public Health | NSW Health Pathology - Institute of Clinical Pathology and Medical Research; Westmead Hospital; University of Sydney |
| EPI_ISL_427694 | Centre for Infectious Diseases and Microbiology Public Health | NSW Health Pathology - Institute of Clinical Pathology and Medical Research; Westmead Hospital; University of Sydney |
| EPI_ISL_427695 | Centre for Infectious Diseases and Microbiology Public Health | NSW Health Pathology - Institute of Clinical Pathology and Medical Research; Westmead Hospital; University of Sydney |
| EPI_ISL_427696 | Centre for Infectious Diseases and Microbiology Public Health | NSW Health Pathology - Institute of Clinical Pathology and Medical Research; Westmead Hospital; University of Sydney |
| EPI_ISL_427697 | Centre for Infectious Diseases and Microbiology Public Health | NSW Health Pathology - Institute of Clinical Pathology and Medical Research; Westmead Hospital; University of Sydney |
| EPI_ISL_427698 | Centre for Infectious Diseases and Microbiology Public Health | NSW Health Pathology - Institute of Clinical Pathology and Medical Research; Westmead Hospital; University of Sydney |

| EPI_Accession_ID | Originating Laboratory | Submitting Laboratory |
| --- | --- | --- |
| EPI_ISL_427699 | Centre for Infectious Diseases and Microbiology Public Health | NSW Health Pathology - Institute of Clinical Pathology and Medical Research; Westmead Hospital; University of Sydney |
| EPI_ISL_427700 | Centre for Infectious Diseases and Microbiology Public Health | NSW Health Pathology - Institute of Clinical Pathology and Medical Research; Westmead Hospital; University of Sydney |
| EPI_ISL_427701 | Centre for Infectious Diseases and Microbiology Public Health | NSW Health Pathology - Institute of Clinical Pathology and Medical Research; Westmead Hospital; University of Sydney |
| EPI_ISL_427702 | Centre for Infectious Diseases and Microbiology Public Health | NSW Health Pathology - Institute of Clinical Pathology and Medical Research; Westmead Hospital; University of Sydney |
| EPI_ISL_427703 | Centre for Infectious Diseases and Microbiology Public Health | NSW Health Pathology - Institute of Clinical Pathology and Medical Research; Westmead Hospital; University of Sydney |
| EPI_ISL_427704 | Centre for Infectious Diseases and Microbiology Public Health | NSW Health Pathology - Institute of Clinical Pathology and Medical Research; Westmead Hospital; University of Sydney |
| EPI_ISL_427705 | Centre for Infectious Diseases and Microbiology Public Health | NSW Health Pathology - Institute of Clinical Pathology and Medical Research; Westmead Hospital; University of Sydney |
| EPI_ISL_427706 | Centre for Infectious Diseases and Microbiology Public Health | NSW Health Pathology - Institute of Clinical Pathology and Medical Research; Westmead Hospital; University of Sydney |
| EPI_ISL_427707 | Centre for Infectious Diseases and Microbiology Public Health | NSW Health Pathology - Institute of Clinical Pathology and Medical Research; Westmead Hospital; University of Sydney |
| EPI_ISL_427708 | Centre for Infectious Diseases and Microbiology Public Health | NSW Health Pathology - Institute of Clinical Pathology and Medical Research; Westmead Hospital; University of Sydney |
| EPI_ISL_427709 | Centre for Infectious Diseases and Microbiology Public Health | NSW Health Pathology - Institute of Clinical Pathology and Medical Research; Westmead Hospital; University of Sydney |

| EPI_Accession_ID | Originating Laboratory | Submitting Laboratory |
|---|---|---|
| EPI_ISL_427710 | Centre for Infectious Diseases and Microbiology Public Health | NSW Health Pathology - Institute of Clinical Pathology and Medical Research; Westmead Hospital; University of Sydney |
| EPI_ISL_427711 | ACT Pathology, The Canberra Hospital | NSW Health Pathology - Institute of Clinical Pathology and Medical Research; Westmead Hospital; University of Sydney |
| EPI_ISL_427712 | Centre for Infectious Diseases and Microbiology Public Health | NSW Health Pathology - Institute of Clinical Pathology and Medical Research; Westmead Hospital; University of Sydney |
| EPI_ISL_427713 | Centre for Infectious Diseases and Microbiology Public Health | NSW Health Pathology - Institute of Clinical Pathology and Medical Research; Westmead Hospital; University of Sydney |
| EPI_ISL_427714 | Centre for Infectious Diseases and Microbiology Public Health | NSW Health Pathology - Institute of Clinical Pathology and Medical Research; Westmead Hospital; University of Sydney |
| EPI_ISL_427715 | Centre for Infectious Diseases and Microbiology Public Health | NSW Health Pathology - Institute of Clinical Pathology and Medical Research; Westmead Hospital; University of Sydney |
| EPI_ISL_427716 | Centre for Infectious Diseases and Microbiology Public Health | NSW Health Pathology - Institute of Clinical Pathology and Medical Research; Westmead Hospital; University of Sydney |
| EPI_ISL_427717 | ACT Pathology, The Canberra Hospital | NSW Health Pathology - Institute of Clinical Pathology and Medical Research; Westmead Hospital; University of Sydney |
| EPI_ISL_427718 | ACT Pathology, The Canberra Hospital | NSW Health Pathology - Institute of Clinical Pathology and Medical Research; Westmead Hospital; University of Sydney |
| EPI_ISL_427719 | ACT Pathology, The Canberra Hospital | NSW Health Pathology - Institute of Clinical Pathology and Medical Research; Westmead Hospital; University of Sydney |
| EPI_ISL_427720 | Centre for Infectious Diseases and Microbiology Public Health | NSW Health Pathology - Institute of Clinical Pathology and Medical Research; Westmead Hospital; University of Sydney |

| EPI_Accession_ID | Originating Laboratory | Submitting Laboratory |
|---|---|---|
| EPI_ISL_427721 | ACT Pathology, The Canberra Hospital | NSW Health Pathology - Institute of Clinical Pathology and Medical Research; Westmead Hospital; University of Sydney |
| EPI_ISL_427722 | Centre for Infectious Diseases and Microbiology Public Health | NSW Health Pathology - Institute of Clinical Pathology and Medical Research; Westmead Hospital; University of Sydney |
| EPI_ISL_427723 | Centre for Infectious Diseases and Microbiology Public Health | NSW Health Pathology - Institute of Clinical Pathology and Medical Research; Westmead Hospital; University of Sydney |
| EPI_ISL_427724 | Centre for Infectious Diseases and Microbiology Public Health | NSW Health Pathology - Institute of Clinical Pathology and Medical Research; Westmead Hospital; University of Sydney |
| EPI_ISL_427725 | Centre for Infectious Diseases and Microbiology Public Health | NSW Health Pathology - Institute of Clinical Pathology and Medical Research; Westmead Hospital; University of Sydney |
| EPI_ISL_427726 | Centre for Infectious Diseases and Microbiology Public Health | NSW Health Pathology - Institute of Clinical Pathology and Medical Research; Westmead Hospital; University of Sydney |
| EPI_ISL_427727 | Centre for Infectious Diseases and Microbiology Public Health | NSW Health Pathology - Institute of Clinical Pathology and Medical Research; Westmead Hospital; University of Sydney |
| EPI_ISL_427728 | Centre for Infectious Diseases and Microbiology Public Health | NSW Health Pathology - Institute of Clinical Pathology and Medical Research; Westmead Hospital; University of Sydney |
| EPI_ISL_427729 | Centre for Infectious Diseases and Microbiology Public Health | NSW Health Pathology - Institute of Clinical Pathology and Medical Research; Westmead Hospital; University of Sydney |
| EPI_ISL_427730 | Centre for Infectious Diseases and Microbiology Public Health | NSW Health Pathology - Institute of Clinical Pathology and Medical Research; Westmead Hospital; University of Sydney |
| EPI_ISL_427731 | Centre for Infectious Diseases and Microbiology Public Health | NSW Health Pathology - Institute of Clinical Pathology and Medical Research; Westmead Hospital; University of Sydney |

| EPI_Accession_ID | Originating Laboratory | Submitting Laboratory |
|---|---|---|
| EPI_ISL_427732 | Centre for Infectious Diseases and Microbiology Public Health | NSW Health Pathology - Institute of Clinical Pathology and Medical Research; Westmead Hospital; University of Sydney |
| EPI_ISL_427733 | Centre for Infectious Diseases and Microbiology Public Health | NSW Health Pathology - Institute of Clinical Pathology and Medical Research; Westmead Hospital; University of Sydney |
| EPI_ISL_427734 | Centre for Infectious Diseases and Microbiology Public Health | NSW Health Pathology - Institute of Clinical Pathology and Medical Research; Westmead Hospital; University of Sydney |
| EPI_ISL_427735 | Centre for Infectious Diseases and Microbiology Public Health | NSW Health Pathology - Institute of Clinical Pathology and Medical Research; Westmead Hospital; University of Sydney |
| EPI_ISL_427736 | Centre for Infectious Diseases and Microbiology Public Health | NSW Health Pathology - Institute of Clinical Pathology and Medical Research; Westmead Hospital; University of Sydney |
| EPI_ISL_427737 | Centre for Infectious Diseases and Microbiology Public Health | NSW Health Pathology - Institute of Clinical Pathology and Medical Research; Westmead Hospital; University of Sydney |
| EPI_ISL_427738 | Centre for Infectious Diseases and Microbiology Public Health | NSW Health Pathology - Institute of Clinical Pathology and Medical Research; Westmead Hospital; University of Sydney |
| EPI_ISL_427739 | Centre for Infectious Diseases and Microbiology Public Health | NSW Health Pathology - Institute of Clinical Pathology and Medical Research; Westmead Hospital; University of Sydney |
| EPI_ISL_427740 | Centre for Infectious Diseases and Microbiology Public Health | NSW Health Pathology - Institute of Clinical Pathology and Medical Research; Westmead Hospital; University of Sydney |
| EPI_ISL_427741 | Centre for Infectious Diseases and Microbiology Public Health | NSW Health Pathology - Institute of Clinical Pathology and Medical Research; Westmead Hospital; University of Sydney |
| EPI_ISL_427742 | Centre for Infectious Diseases and Microbiology Public Health | NSW Health Pathology - Institute of Clinical Pathology and Medical Research; Westmead Hospital; University of Sydney |

| EPI_Accession_ID | Originating Laboratory | Submitting Laboratory |
|---|---|---|
| EPI_ISL_427743 | Centre for Infectious Diseases and Microbiology Public Health | NSW Health Pathology - Institute of Clinical Pathology and Medical Research; Westmead Hospital; University of Sydney |
| EPI_ISL_427744 | Centre for Infectious Diseases and Microbiology Public Health | NSW Health Pathology - Institute of Clinical Pathology and Medical Research; Westmead Hospital; University of Sydney |
| EPI_ISL_427745 | Centre for Infectious Diseases and Microbiology Public Health | NSW Health Pathology - Institute of Clinical Pathology and Medical Research; Westmead Hospital; University of Sydney |
| EPI_ISL_427746 | Centre for Infectious Diseases and Microbiology Public Health | NSW Health Pathology - Institute of Clinical Pathology and Medical Research; Westmead Hospital; University of Sydney |
| EPI_ISL_427747 | Centre for Infectious Diseases and Microbiology Public Health | NSW Health Pathology - Institute of Clinical Pathology and Medical Research; Westmead Hospital; University of Sydney |
| EPI_ISL_427748 | Centre for Infectious Diseases and Microbiology Public Health | NSW Health Pathology - Institute of Clinical Pathology and Medical Research; Westmead Hospital; University of Sydney |
| EPI_ISL_427749 | Centre for Infectious Diseases and Microbiology Public Health | NSW Health Pathology - Institute of Clinical Pathology and Medical Research; Westmead Hospital; University of Sydney |
| EPI_ISL_427750 | Centre for Infectious Diseases and Microbiology Public Health | NSW Health Pathology - Institute of Clinical Pathology and Medical Research; Westmead Hospital; University of Sydney |
| EPI_ISL_427751 | Centre for Infectious Diseases and Microbiology Public Health | NSW Health Pathology - Institute of Clinical Pathology and Medical Research; Westmead Hospital; University of Sydney |
| EPI_ISL_427752 | Centre for Infectious Diseases and Microbiology Public Health | NSW Health Pathology - Institute of Clinical Pathology and Medical Research; Westmead Hospital; University of Sydney |
| EPI_ISL_427753 | Centre for Infectious Diseases and Microbiology Public Health | NSW Health Pathology - Institute of Clinical Pathology and Medical Research; Westmead Hospital; University of Sydney |

| EPI_Accession_ID | Originating Laboratory | Submitting Laboratory |
|---|---|---|
| EPI_ISL_427754 | Centre for Infectious Diseases and Microbiology Public Health | NSW Health Pathology - Institute of Clinical Pathology and Medical Research; Westmead Hospital; University of Sydney |
| EPI_ISL_427755 | Centre for Infectious Diseases and Microbiology Public Health | NSW Health Pathology - Institute of Clinical Pathology and Medical Research; Westmead Hospital; University of Sydney |
| EPI_ISL_427756 | Centre for Infectious Diseases and Microbiology Public Health | NSW Health Pathology - Institute of Clinical Pathology and Medical Research; Westmead Hospital; University of Sydney |
| EPI_ISL_427757 | Centre for Infectious Diseases and Microbiology Public Health | NSW Health Pathology - Institute of Clinical Pathology and Medical Research; Westmead Hospital; University of Sydney |
| EPI_ISL_427758 | Centre for Infectious Diseases and Microbiology Public Health | NSW Health Pathology - Institute of Clinical Pathology and Medical Research; Westmead Hospital; University of Sydney |
| EPI_ISL_427759 | Centre for Infectious Diseases and Microbiology Public Health | NSW Health Pathology - Institute of Clinical Pathology and Medical Research; Westmead Hospital; University of Sydney |
| EPI_ISL_427760 | Centre for Infectious Diseases and Microbiology Public Health | NSW Health Pathology - Institute of Clinical Pathology and Medical Research; Westmead Hospital; University of Sydney |
| EPI_ISL_427761 | Centre for Infectious Diseases and Microbiology Public Health | NSW Health Pathology - Institute of Clinical Pathology and Medical Research; Westmead Hospital; University of Sydney |
| EPI_ISL_427762 | Centre for Infectious Diseases and Microbiology Public Health | NSW Health Pathology - Institute of Clinical Pathology and Medical Research; Westmead Hospital; University of Sydney |
| EPI_ISL_427763 | Centre for Infectious Diseases and Microbiology Public Health | NSW Health Pathology - Institute of Clinical Pathology and Medical Research; Westmead Hospital; University of Sydney |
| EPI_ISL_427764 | Centre for Infectious Diseases and Microbiology Public Health | NSW Health Pathology - Institute of Clinical Pathology and Medical Research; Westmead Hospital; University of Sydney |

| EPI_Accession_ID | Originating Laboratory | Submitting Laboratory |
|---|---|---|
| EPI_ISL_427765 | Centre for Infectious Diseases and Microbiology Public Health | NSW Health Pathology - Institute of Clinical Pathology and Medical Research; Westmead Hospital; University of Sydney |
| EPI_ISL_427766 | Centre for Infectious Diseases and Microbiology Public Health | NSW Health Pathology - Institute of Clinical Pathology and Medical Research; Westmead Hospital; University of Sydney |
| EPI_ISL_427767 | Centre for Infectious Diseases and Microbiology Public Health | NSW Health Pathology - Institute of Clinical Pathology and Medical Research; Westmead Hospital; University of Sydney |
| EPI_ISL_427768 | Centre for Infectious Diseases and Microbiology Public Health | NSW Health Pathology - Institute of Clinical Pathology and Medical Research; Westmead Hospital; University of Sydney |
| EPI_ISL_427769 | Centre for Infectious Diseases and Microbiology Public Health | NSW Health Pathology - Institute of Clinical Pathology and Medical Research; Westmead Hospital; University of Sydney |
| EPI_ISL_427770 | Centre for Infectious Diseases and Microbiology Public Health | NSW Health Pathology - Institute of Clinical Pathology and Medical Research; Westmead Hospital; University of Sydney |
| EPI_ISL_427771 | Centre for Infectious Diseases and Microbiology Public Health | NSW Health Pathology - Institute of Clinical Pathology and Medical Research; Westmead Hospital; University of Sydney |
| EPI_ISL_427772 | Centre for Infectious Diseases and Microbiology Public Health | NSW Health Pathology - Institute of Clinical Pathology and Medical Research; Westmead Hospital; University of Sydney |
| EPI_ISL_427773 | Centre for Infectious Diseases and Microbiology Public Health | NSW Health Pathology - Institute of Clinical Pathology and Medical Research; Westmead Hospital; University of Sydney |
| EPI_ISL_427774 | Centre for Infectious Diseases and Microbiology Public Health | NSW Health Pathology - Institute of Clinical Pathology and Medical Research; Westmead Hospital; University of Sydney |
| EPI_ISL_427775 | Centre for Infectious Diseases and Microbiology Public Health | NSW Health Pathology - Institute of Clinical Pathology and Medical Research; Westmead Hospital; University of Sydney |

| EPI_Accession_ID | Originating Laboratory | Submitting Laboratory |
|---|---|---|
| EPI_ISL_427776 | Centre for Infectious Diseases and Microbiology Public Health | NSW Health Pathology - Institute of Clinical Pathology and Medical Research; Westmead Hospital; University of Sydney |
| EPI_ISL_427777 | Centre for Infectious Diseases and Microbiology Public Health | NSW Health Pathology - Institute of Clinical Pathology and Medical Research; Westmead Hospital; University of Sydney |
| EPI_ISL_427778 | Centre for Infectious Diseases and Microbiology Public Health | NSW Health Pathology - Institute of Clinical Pathology and Medical Research; Westmead Hospital; University of Sydney |
| EPI_ISL_427779 | Centre for Infectious Diseases and Microbiology Public Health | NSW Health Pathology - Institute of Clinical Pathology and Medical Research; Westmead Hospital; University of Sydney |
| EPI_ISL_427780 | Centre for Infectious Diseases and Microbiology Public Health | NSW Health Pathology - Institute of Clinical Pathology and Medical Research; Westmead Hospital; University of Sydney |
| EPI_ISL_427781 | Centre for Infectious Diseases and Microbiology Public Health | NSW Health Pathology - Institute of Clinical Pathology and Medical Research; Westmead Hospital; University of Sydney |
| EPI_ISL_427782 | Centre for Infectious Diseases and Microbiology Public Health | NSW Health Pathology - Institute of Clinical Pathology and Medical Research; Westmead Hospital; University of Sydney |
| EPI_ISL_427783 | Centre for Infectious Diseases and Microbiology Public Health | NSW Health Pathology - Institute of Clinical Pathology and Medical Research; Westmead Hospital; University of Sydney |
| EPI_ISL_427784 | Centre for Infectious Diseases and Microbiology Public Health | NSW Health Pathology - Institute of Clinical Pathology and Medical Research; Westmead Hospital; University of Sydney |
| EPI_ISL_427785 | Centre for Infectious Diseases and Microbiology Public Health | NSW Health Pathology - Institute of Clinical Pathology and Medical Research; Westmead Hospital; University of Sydney |
| EPI_ISL_427786 | Centre for Infectious Diseases and Microbiology Public Health | NSW Health Pathology - Institute of Clinical Pathology and Medical Research; Westmead Hospital; University of Sydney |

| EPI_Accession_ID | Originating Laboratory | Submitting Laboratory |
|---|---|---|
| EPI_ISL_427787 | Centre for Infectious Diseases and Microbiology Public Health | NSW Health Pathology - Institute of Clinical Pathology and Medical Research; Westmead Hospital; University of Sydney |
| EPI_ISL_427788 | Centre for Infectious Diseases and Microbiology Public Health | NSW Health Pathology - Institute of Clinical Pathology and Medical Research; Westmead Hospital; University of Sydney |
| EPI_ISL_427789 | Centre for Infectious Diseases and Microbiology Public Health | NSW Health Pathology - Institute of Clinical Pathology and Medical Research; Westmead Hospital; University of Sydney |
| EPI_ISL_427790 | Centre for Infectious Diseases and Microbiology Public Health | NSW Health Pathology - Institute of Clinical Pathology and Medical Research; Westmead Hospital; University of Sydney |
| EPI_ISL_427791 | Centre for Infectious Diseases and Microbiology Public Health | NSW Health Pathology - Institute of Clinical Pathology and Medical Research; Westmead Hospital; University of Sydney |
| EPI_ISL_427792 | Centre for Infectious Diseases and Microbiology Public Health | NSW Health Pathology - Institute of Clinical Pathology and Medical Research; Westmead Hospital; University of Sydney |
| EPI_ISL_427793 | Centre for Infectious Diseases and Microbiology Public Health | NSW Health Pathology - Institute of Clinical Pathology and Medical Research; Westmead Hospital; University of Sydney |
| EPI_ISL_427794 | Centre for Infectious Diseases and Microbiology Public Health | NSW Health Pathology - Institute of Clinical Pathology and Medical Research; Westmead Hospital; University of Sydney |
| EPI_ISL_427795 | Centre for Infectious Diseases and Microbiology Public Health | NSW Health Pathology - Institute of Clinical Pathology and Medical Research; Westmead Hospital; University of Sydney |
| EPI_ISL_427796 | Centre for Infectious Diseases and Microbiology Public Health | NSW Health Pathology - Institute of Clinical Pathology and Medical Research; Westmead Hospital; University of Sydney |
| EPI_ISL_427797 | Centre for Infectious Diseases and Microbiology Public Health | NSW Health Pathology - Institute of Clinical Pathology and Medical Research; Westmead Hospital; University of Sydney |

| EPI_Accession_ID | Originating Laboratory | Submitting Laboratory |
|---|---|---|
| EPI_ISL_427798 | Centre for Infectious Diseases and Microbiology Public Health | NSW Health Pathology - Institute of Clinical Pathology and Medical Research; Westmead Hospital; University of Sydney |
| EPI_ISL_427799 | Centre for Infectious Diseases and Microbiology Public Health | NSW Health Pathology - Institute of Clinical Pathology and Medical Research; Westmead Hospital; University of Sydney |
| EPI_ISL_427800 | Centre for Infectious Diseases and Microbiology Public Health | NSW Health Pathology - Institute of Clinical Pathology and Medical Research; Westmead Hospital; University of Sydney |
| EPI_ISL_427801 | Centre for Infectious Diseases and Microbiology Public Health | NSW Health Pathology - Institute of Clinical Pathology and Medical Research; Westmead Hospital; University of Sydney |
| EPI_ISL_427802 | Centre for Infectious Diseases and Microbiology Public Health | NSW Health Pathology - Institute of Clinical Pathology and Medical Research; Westmead Hospital; University of Sydney |
| EPI_ISL_427803 | Centre for Infectious Diseases and Microbiology Public Health | NSW Health Pathology - Institute of Clinical Pathology and Medical Research; Westmead Hospital; University of Sydney |
| EPI_ISL_427804 | Centre for Infectious Diseases and Microbiology Public Health | NSW Health Pathology - Institute of Clinical Pathology and Medical Research; Westmead Hospital; University of Sydney |
| EPI_ISL_427805 | Centre for Infectious Diseases and Microbiology Public Health | NSW Health Pathology - Institute of Clinical Pathology and Medical Research; Westmead Hospital; University of Sydney |
| EPI_ISL_427806 | Centre for Infectious Diseases and Microbiology Public Health | NSW Health Pathology - Institute of Clinical Pathology and Medical Research; Westmead Hospital; University of Sydney |
| EPI_ISL_427807 | Centre for Infectious Diseases and Microbiology Public Health | NSW Health Pathology - Institute of Clinical Pathology and Medical Research; Westmead Hospital; University of Sydney |
| EPI_ISL_427808 | Centre for Infectious Diseases and Microbiology Public Health | NSW Health Pathology - Institute of Clinical Pathology and Medical Research; Westmead Hospital; University of Sydney |

| EPI_Accession_ID | Originating Laboratory | Submitting Laboratory |
|---|---|---|
| EPI_ISL_427809 | Division of Viral Diseases, Center for Laboratory Control of Infectious Diseases, Korea Centers for Diseases Control and Prevention | Division of Viral Diseases, Center for Laboratory Control of Infectious Diseases, Korea Centers for Diseases Control and Prevention |
| EPI_ISL_427810 | Division of Viral Diseases, Center for Laboratory Control of Infectious Diseases, Korea Centers for Diseases Control and Prevention | Division of Viral Diseases, Center for Laboratory Control of Infectious Diseases, Korea Centers for Diseases Control and Prevention |
| EPI_ISL_427811 | Division of Viral Diseases, Center for Laboratory Control of Infectious Diseases, Korea Centers for Diseases Control and Prevention | Division of Viral Diseases, Center for Laboratory Control of Infectious Diseases, Korea Centers for Diseases Control and Prevention |
| EPI_ISL_427812 | Division of Viral Diseases, Center for Laboratory Control of Infectious Diseases, Korea Centers for Diseases Control and Prevention | Division of Viral Diseases, Center for Laboratory Control of Infectious Diseases, Korea Centers for Diseases Control and Prevention |
| EPI_ISL_427813 | Division of Viral Diseases, Center for Laboratory Control of Infectious Diseases, Korea Centers for Diseases Control and Prevention | Division of Viral Diseases, Center for Laboratory Control of Infectious Diseases, Korea Centers for Diseases Control and Prevention |
| EPI_ISL_427815 | WHO National Influenza Centre Russian Federation | WHO National Influenza Centre Russian Federation |
| EPI_ISL_428148 | Klinisk mikrobiologi, Region VÃ¤sterbotten | Unit for Biological Agents, Department for CBRN Defence and Security, Swedish Defence Research Agency |
| EPI_ISL_428201 | Klinisk mikrobiologi, Region VÃ¤sterbotten | Unit for Biological Agents, Department for CBRN Defence and Security, Swedish Defence Research Agency |
| EPI_ISL_428202 | Nebraska Public Health Laboratory | UNMC COVID-19 Response Team |
| EPI_ISL_428203 | Nebraska Public Health Laboratory | UNMC COVID-19 Response Team |
| EPI_ISL_428204 | Nebraska Public Health Laboratory | UNMC COVID-19 Response Team |
| EPI_ISL_428205 | Nebraska Public Health Laboratory | UNMC COVID-19 Response Team |
| EPI_ISL_428206 | Nebraska Public Health Laboratory | UNMC COVID-19 Response Team |
| EPI_ISL_428209 | Laboratory of Molecular Biology, Diagnostyka sp. z o.o. | Laboratory of Recombinant Vaccines |
| EPI_ISL_428229 | TSGH-CP molecular lab | TSGH-CP molecular lab |
| EPI_ISL_428230 | TSGH-CP molecular lab | TSGH-CP molecular lab |
| EPI_ISL_428231 | TSGH-CP molecular lab | TSGH-CP molecular lab |

| EPI_Accession_ID | Originating Laboratory | Submitting Laboratory |
|---|---|---|
| EPI_ISL_428232 | Hematology Laboratory, Section of Molecular Diagnostics, University Clinical Centre, Medical University of Gdansk | Department of Virology, Faculty of Medicine, University of Helsinki, Helsinki, Finland |
| EPI_ISL_428233 | Hematology Laboratory, Section of Molecular Diagnostics, University Clinical Centre, Medical University of Gdansk | Department of Virology, Faculty of Medicine, University of Helsinki, Helsinki, Finland |
| EPI_ISL_428234 | Hematology Laboratory, Section of Molecular Diagnostics, University Clinical Centre, Medical University of Gdansk | Department of Virology, Faculty of Medicine, University of Helsinki, Helsinki, Finland |
| EPI_ISL_428235 | Hematology Laboratory, Section of Molecular Diagnostics, University Clinical Centre, Medical University of Gdansk | Department of Virology, Faculty of Medicine, University of Helsinki, Helsinki, Finland |
| EPI_ISL_428236 | Hematology Laboratory, Section of Molecular Diagnostics, University Clinical Centre, Medical University of Gdansk | Department of Virology, Faculty of Medicine, University of Helsinki, Helsinki, Finland |
| EPI_ISL_428250 | Yale Clinical Virology Laboratory | Grubaugh Lab - Yale School of Public Health |
| EPI_ISL_428252 | University of Wisconsin-Madison AIDS Vaccine Research Laboratories | University of Wisconsin-Madison AIDS Vaccine Research Laboratories |
| EPI_ISL_428253 | University of Wisconsin-Madison AIDS Vaccine Research Laboratories | University of Wisconsin-Madison AIDS Vaccine Research Laboratories |
| EPI_ISL_428254 | University of Wisconsin-Madison AIDS Vaccine Research Laboratories | University of Wisconsin-Madison AIDS Vaccine Research Laboratories |
| EPI_ISL_428255 | University of Wisconsin-Madison AIDS Vaccine Research Laboratories | University of Wisconsin-Madison AIDS Vaccine Research Laboratories |
| EPI_ISL_428256 | University of Wisconsin-Madison AIDS Vaccine Research Laboratories | University of Wisconsin-Madison AIDS Vaccine Research Laboratories |
| EPI_ISL_428257 | University of Wisconsin-Madison AIDS Vaccine Research Laboratories | University of Wisconsin-Madison AIDS Vaccine Research Laboratories |
| EPI_ISL_428258 | University of Wisconsin-Madison AIDS Vaccine Research Laboratories | University of Wisconsin-Madison AIDS Vaccine Research Laboratories |
| EPI_ISL_428259 | University of Wisconsin-Madison AIDS Vaccine Research Laboratories | University of Wisconsin-Madison AIDS Vaccine Research Laboratories |

| EPI_Accession_ID | Originating Laboratory | Submitting Laboratory |
|---|---|---|
| EPI_ISL_428260 | University of Wisconsin-Madison AIDS Vaccine Research Laboratories | University of Wisconsin-Madison AIDS Vaccine Research Laboratories |
| EPI_ISL_428261 | University of Wisconsin-Madison AIDS Vaccine Research Laboratories | University of Wisconsin-Madison AIDS Vaccine Research Laboratories |
| EPI_ISL_428262 | University of Wisconsin-Madison AIDS Vaccine Research Laboratories | University of Wisconsin-Madison AIDS Vaccine Research Laboratories |
| EPI_ISL_428263 | University of Wisconsin-Madison AIDS Vaccine Research Laboratories | University of Wisconsin-Madison AIDS Vaccine Research Laboratories |
| EPI_ISL_428264 | University of Wisconsin-Madison AIDS Vaccine Research Laboratories | University of Wisconsin-Madison AIDS Vaccine Research Laboratories |
| EPI_ISL_428265 | University of Wisconsin-Madison AIDS Vaccine Research Laboratories | University of Wisconsin-Madison AIDS Vaccine Research Laboratories |
| EPI_ISL_428266 | University of Wisconsin-Madison AIDS Vaccine Research Laboratories | University of Wisconsin-Madison AIDS Vaccine Research Laboratories |
| EPI_ISL_428267 | University of Wisconsin-Madison AIDS Vaccine Research Laboratories | University of Wisconsin-Madison AIDS Vaccine Research Laboratories |
| EPI_ISL_428268 | University of Wisconsin-Madison AIDS Vaccine Research Laboratories | University of Wisconsin-Madison AIDS Vaccine Research Laboratories |
| EPI_ISL_428269 | University of Wisconsin-Madison AIDS Vaccine Research Laboratories | University of Wisconsin-Madison AIDS Vaccine Research Laboratories |
| EPI_ISL_428270 | University of Wisconsin-Madison AIDS Vaccine Research Laboratories | University of Wisconsin-Madison AIDS Vaccine Research Laboratories |
| EPI_ISL_428271 | University of Wisconsin-Madison AIDS Vaccine Research Laboratories | University of Wisconsin-Madison AIDS Vaccine Research Laboratories |
| EPI_ISL_428272 | University of Wisconsin-Madison AIDS Vaccine Research Laboratories | University of Wisconsin-Madison AIDS Vaccine Research Laboratories |
| EPI_ISL_428273 | University of Wisconsin-Madison AIDS Vaccine Research Laboratories | University of Wisconsin-Madison AIDS Vaccine Research Laboratories |
| EPI_ISL_428274 | University of Wisconsin-Madison AIDS Vaccine Research Laboratories | University of Wisconsin-Madison AIDS Vaccine Research Laboratories |
| EPI_ISL_428275 | University of Wisconsin-Madison AIDS Vaccine Research Laboratories | University of Wisconsin-Madison AIDS Vaccine Research Laboratories |

| EPI_Accession_ID | Originating Laboratory | Submitting Laboratory |
|---|---|---|
| EPI_ISL_428276 | University of Wisconsin-Madison AIDS Vaccine Research Laboratories | University of Wisconsin-Madison AIDS Vaccine Research Laboratories |
| EPI_ISL_428277 | University of Wisconsin-Madison AIDS Vaccine Research Laboratories | University of Wisconsin-Madison AIDS Vaccine Research Laboratories |
| EPI_ISL_428278 | University of Wisconsin-Madison AIDS Vaccine Research Laboratories | University of Wisconsin-Madison AIDS Vaccine Research Laboratories |
| EPI_ISL_428279 | University of Wisconsin-Madison AIDS Vaccine Research Laboratories | University of Wisconsin-Madison AIDS Vaccine Research Laboratories |
| EPI_ISL_428280 | University of Wisconsin-Madison AIDS Vaccine Research Laboratories | University of Wisconsin-Madison AIDS Vaccine Research Laboratories |
| EPI_ISL_428281 | University of Wisconsin-Madison AIDS Vaccine Research Laboratories | University of Wisconsin-Madison AIDS Vaccine Research Laboratories |
| EPI_ISL_428282 | University of Wisconsin-Madison AIDS Vaccine Research Laboratories | University of Wisconsin-Madison AIDS Vaccine Research Laboratories |
| EPI_ISL_428283 | University of Wisconsin-Madison AIDS Vaccine Research Laboratories | University of Wisconsin-Madison AIDS Vaccine Research Laboratories |
| EPI_ISL_428284 | University of Wisconsin-Madison AIDS Vaccine Research Laboratories | University of Wisconsin-Madison AIDS Vaccine Research Laboratories |
| EPI_ISL_428285 | University of Wisconsin-Madison AIDS Vaccine Research Laboratories | University of Wisconsin-Madison AIDS Vaccine Research Laboratories |
| EPI_ISL_428286 | University of Wisconsin-Madison AIDS Vaccine Research Laboratories | University of Wisconsin-Madison AIDS Vaccine Research Laboratories |
| EPI_ISL_428287 | University of Wisconsin-Madison AIDS Vaccine Research Laboratories | University of Wisconsin-Madison AIDS Vaccine Research Laboratories |
| EPI_ISL_428288 | University of Wisconsin-Madison AIDS Vaccine Research Laboratories | University of Wisconsin-Madison AIDS Vaccine Research Laboratories |
| EPI_ISL_428289 | University of Wisconsin-Madison AIDS Vaccine Research Laboratories | University of Wisconsin-Madison AIDS Vaccine Research Laboratories |
| EPI_ISL_428290 | University of Wisconsin-Madison AIDS Vaccine Research Laboratories | University of Wisconsin-Madison AIDS Vaccine Research Laboratories |
| EPI_ISL_428291 | University of Wisconsin-Madison AIDS Vaccine Research Laboratories | University of Wisconsin-Madison AIDS Vaccine Research Laboratories |

| EPI_Accession_ID | Originating Laboratory | Submitting Laboratory |
|---|---|---|
| EPI_ISL_428292 | University of Wisconsin-Madison AIDS Vaccine Research Laboratories | University of Wisconsin-Madison AIDS Vaccine Research Laboratories |
| EPI_ISL_428293 | University of Wisconsin-Madison AIDS Vaccine Research Laboratories | University of Wisconsin-Madison AIDS Vaccine Research Laboratories |
| EPI_ISL_428294 | University of Wisconsin-Madison AIDS Vaccine Research Laboratories | University of Wisconsin-Madison AIDS Vaccine Research Laboratories |
| EPI_ISL_428295 | University of Wisconsin-Madison AIDS Vaccine Research Laboratories | University of Wisconsin-Madison AIDS Vaccine Research Laboratories |
| EPI_ISL_428296 | University of Wisconsin-Madison AIDS Vaccine Research Laboratories | University of Wisconsin-Madison AIDS Vaccine Research Laboratories |
| EPI_ISL_428297 | University of Wisconsin-Madison AIDS Vaccine Research Laboratories | University of Wisconsin-Madison AIDS Vaccine Research Laboratories |
| EPI_ISL_428298 | University of Wisconsin-Madison AIDS Vaccine Research Laboratories | University of Wisconsin-Madison AIDS Vaccine Research Laboratories |
| EPI_ISL_428299 | University of Wisconsin-Madison AIDS Vaccine Research Laboratories | University of Wisconsin-Madison AIDS Vaccine Research Laboratories |
| EPI_ISL_428300 | University of Wisconsin-Madison AIDS Vaccine Research Laboratories | University of Wisconsin-Madison AIDS Vaccine Research Laboratories |
| EPI_ISL_428301 | University of Wisconsin-Madison AIDS Vaccine Research Laboratories | University of Wisconsin-Madison AIDS Vaccine Research Laboratories |
| EPI_ISL_428302 | University of Wisconsin-Madison AIDS Vaccine Research Laboratories | University of Wisconsin-Madison AIDS Vaccine Research Laboratories |
| EPI_ISL_428303 | University of Wisconsin-Madison AIDS Vaccine Research Laboratories | University of Wisconsin-Madison AIDS Vaccine Research Laboratories |
| EPI_ISL_428304 | University of Wisconsin-Madison AIDS Vaccine Research Laboratories | University of Wisconsin-Madison AIDS Vaccine Research Laboratories |
| EPI_ISL_428305 | University of Wisconsin-Madison AIDS Vaccine Research Laboratories | University of Wisconsin-Madison AIDS Vaccine Research Laboratories |
| EPI_ISL_428306 | University of Wisconsin-Madison AIDS Vaccine Research Laboratories | University of Wisconsin-Madison AIDS Vaccine Research Laboratories |
| EPI_ISL_428307 | University of Wisconsin-Madison AIDS Vaccine Research Laboratories | University of Wisconsin-Madison AIDS Vaccine Research Laboratories |

| EPI_Accession_ID | Originating Laboratory | Submitting Laboratory |
|---|---|---|
| EPI_ISL_428308 | University of Wisconsin-Madison AIDS Vaccine Research Laboratories | University of Wisconsin-Madison AIDS Vaccine Research Laboratories |
| EPI_ISL_428309 | University of Wisconsin-Madison AIDS Vaccine Research Laboratories | University of Wisconsin-Madison AIDS Vaccine Research Laboratories |
| EPI_ISL_428310 | University of Wisconsin-Madison AIDS Vaccine Research Laboratories | University of Wisconsin-Madison AIDS Vaccine Research Laboratories |
| EPI_ISL_428311 | University of Wisconsin-Madison AIDS Vaccine Research Laboratories | University of Wisconsin-Madison AIDS Vaccine Research Laboratories |
| EPI_ISL_428312 | University of Wisconsin-Madison AIDS Vaccine Research Laboratories | University of Wisconsin-Madison AIDS Vaccine Research Laboratories |
| EPI_ISL_428313 | University of Wisconsin-Madison AIDS Vaccine Research Laboratories | University of Wisconsin-Madison AIDS Vaccine Research Laboratories |
| EPI_ISL_428314 | University of Wisconsin-Madison AIDS Vaccine Research Laboratories | University of Wisconsin-Madison AIDS Vaccine Research Laboratories |
| EPI_ISL_428315 | University of Wisconsin-Madison AIDS Vaccine Research Laboratories | University of Wisconsin-Madison AIDS Vaccine Research Laboratories |
| EPI_ISL_428316 | University of Wisconsin-Madison AIDS Vaccine Research Laboratories | University of Wisconsin-Madison AIDS Vaccine Research Laboratories |
| EPI_ISL_428317 | University of Wisconsin-Madison AIDS Vaccine Research Laboratories | University of Wisconsin-Madison AIDS Vaccine Research Laboratories |
| EPI_ISL_428318 | University of Wisconsin-Madison AIDS Vaccine Research Laboratories | University of Wisconsin-Madison AIDS Vaccine Research Laboratories |
| EPI_ISL_428319 | University of Wisconsin-Madison AIDS Vaccine Research Laboratories | University of Wisconsin-Madison AIDS Vaccine Research Laboratories |
| EPI_ISL_428320 | University of Wisconsin-Madison AIDS Vaccine Research Laboratories | University of Wisconsin-Madison AIDS Vaccine Research Laboratories |
| EPI_ISL_428321 | University of Wisconsin-Madison AIDS Vaccine Research Laboratories | University of Wisconsin-Madison AIDS Vaccine Research Laboratories |
| EPI_ISL_428322 | University of Wisconsin-Madison AIDS Vaccine Research Laboratories | University of Wisconsin-Madison AIDS Vaccine Research Laboratories |
| EPI_ISL_428323 | University of Wisconsin-Madison AIDS Vaccine Research Laboratories | University of Wisconsin-Madison AIDS Vaccine Research Laboratories |

| EPI_Accession_ID | Originating Laboratory | Submitting Laboratory |
|---|---|---|
| EPI_ISL_428324 | University of Wisconsin-Madison AIDS Vaccine Research Laboratories | University of Wisconsin-Madison AIDS Vaccine Research Laboratories |
| EPI_ISL_428325 | University of Wisconsin-Madison AIDS Vaccine Research Laboratories | University of Wisconsin-Madison AIDS Vaccine Research Laboratories |
| EPI_ISL_428326 | University of Wisconsin-Madison AIDS Vaccine Research Laboratories | University of Wisconsin-Madison AIDS Vaccine Research Laboratories |
| EPI_ISL_428327 | University of Wisconsin-Madison AIDS Vaccine Research Laboratories | University of Wisconsin-Madison AIDS Vaccine Research Laboratories |
| EPI_ISL_428328 | University of Wisconsin-Madison AIDS Vaccine Research Laboratories | University of Wisconsin-Madison AIDS Vaccine Research Laboratories |
| EPI_ISL_428329 | University of Wisconsin-Madison AIDS Vaccine Research Laboratories | University of Wisconsin-Madison AIDS Vaccine Research Laboratories |
| EPI_ISL_428330 | University of Wisconsin-Madison AIDS Vaccine Research Laboratories | University of Wisconsin-Madison AIDS Vaccine Research Laboratories |
| EPI_ISL_428331 | University of Wisconsin-Madison AIDS Vaccine Research Laboratories | University of Wisconsin-Madison AIDS Vaccine Research Laboratories |
| EPI_ISL_428332 | University of Wisconsin-Madison AIDS Vaccine Research Laboratories | University of Wisconsin-Madison AIDS Vaccine Research Laboratories |
| EPI_ISL_428333 | University of Wisconsin-Madison AIDS Vaccine Research Laboratories | University of Wisconsin-Madison AIDS Vaccine Research Laboratories |
| EPI_ISL_428334 | University of Wisconsin-Madison AIDS Vaccine Research Laboratories | University of Wisconsin-Madison AIDS Vaccine Research Laboratories |
| EPI_ISL_428335 | University of Wisconsin-Madison AIDS Vaccine Research Laboratories | University of Wisconsin-Madison AIDS Vaccine Research Laboratories |
| EPI_ISL_428336 | University of Wisconsin-Madison AIDS Vaccine Research Laboratories | University of Wisconsin-Madison AIDS Vaccine Research Laboratories |
| EPI_ISL_428337 | University of Wisconsin-Madison AIDS Vaccine Research Laboratories | University of Wisconsin-Madison AIDS Vaccine Research Laboratories |
| EPI_ISL_428338 | University of Wisconsin-Madison AIDS Vaccine Research Laboratories | University of Wisconsin-Madison AIDS Vaccine Research Laboratories |
| EPI_ISL_428339 | University of Wisconsin-Madison AIDS Vaccine Research Laboratories | University of Wisconsin-Madison AIDS Vaccine Research Laboratories |

| EPI_Accession_ID | Originating Laboratory | Submitting Laboratory |
|---|---|---|
| EPI_ISL_428340 | University of Wisconsin-Madison AIDS Vaccine Research Laboratories | University of Wisconsin-Madison AIDS Vaccine Research Laboratories |
| EPI_ISL_428341 | University of Wisconsin-Madison AIDS Vaccine Research Laboratories | University of Wisconsin-Madison AIDS Vaccine Research Laboratories |
| EPI_ISL_428342 | University of Wisconsin-Madison AIDS Vaccine Research Laboratories | University of Wisconsin-Madison AIDS Vaccine Research Laboratories |
| EPI_ISL_428343 | University of Wisconsin-Madison AIDS Vaccine Research Laboratories | University of Wisconsin-Madison AIDS Vaccine Research Laboratories |
| EPI_ISL_428344 | University of Wisconsin-Madison AIDS Vaccine Research Laboratories | University of Wisconsin-Madison AIDS Vaccine Research Laboratories |
| EPI_ISL_428345 | University of Wisconsin-Madison AIDS Vaccine Research Laboratories | University of Wisconsin-Madison AIDS Vaccine Research Laboratories |
| EPI_ISL_428346 | Genomic Laboratory (GLAB) (Conjoint lab of Health Directorate of Istanbul and Istanbul Technical University) | Genomic Laboratory (GLAB), Istanbul Technical University |
| EPI_ISL_428347 | Service de Biologie MÃ©dicale - BP 125 | National Reference Center for Viruses of Respiratory Infections, Institut Pasteur, Paris |
| EPI_ISL_428348 | Maison de SantÃ© du Val d'Ormois | National Reference Center for Viruses of Respiratory Infections, Institut Pasteur, Paris |
| EPI_ISL_428349 | Service de Biologie MÃ©dicale - BP 125 | National Reference Center for Viruses of Respiratory Infections, Institut Pasteur, Paris |
| EPI_ISL_428350 | CH Jean de Navarre Laboratoire de Biologie | National Reference Center for Viruses of Respiratory Infections, Institut Pasteur, Paris |
| EPI_ISL_428351 | GH Nord Essonne Service de Biologie clinique | National Reference Center for Viruses of Respiratory Infections, Institut Pasteur, Paris |
| EPI_ISL_428352 | GH Nord Essonne Service de Biologie clinique | National Reference Center for Viruses of Respiratory Infections, Institut Pasteur, Paris |
| EPI_ISL_428353 | CH CompiÃ¨gne Laboratoire de Biologie | National Reference Center for Viruses of Respiratory Infections, Institut Pasteur, Paris |
| EPI_ISL_428354 | LABM GH nord Essonne de Longjumeau - BP 125 | National Reference Center for Viruses of Respiratory Infections, Institut Pasteur, Paris |
| EPI_ISL_428355 | Institut MÃ©dico lÃ©gal- Hop R. PoincarÃ© | National Reference Center for Viruses of Respiratory Infections, Institut Pasteur, Paris |

| EPI_Accession_ID | Originating Laboratory | Submitting Laboratory |
|---|---|---|
| EPI_ISL_428356 | Institut MÃ©dico lÃ©gal- Hop R. PoincarÃ© | National Reference Center for Viruses of Respiratory Infections, Institut Pasteur, Paris |
| EPI_ISL_428357 | Institut MÃ©dico lÃ©gal- Hop R. PoincarÃ© | National Reference Center for Viruses of Respiratory Infections, Institut Pasteur, Paris |
| EPI_ISL_428358 | CH Jeanne de Navarre Laboratoire de Biologie | National Reference Center for Viruses of Respiratory Infections, Institut Pasteur, Paris |
| EPI_ISL_428359 | CH CompiÃ¨gne Laboratoire de Biologie | National Reference Center for Viruses of Respiratory Infections, Institut Pasteur, Paris |
| EPI_ISL_428360 | CH CompiÃ¨gne Laboratoire de Biologie | National Reference Center for Viruses of Respiratory Infections, Institut Pasteur, Paris |
| EPI_ISL_428361 | LABM GH nord Essonne de Longjumeau - BP 125 | National Reference Center for Viruses of Respiratory Infections, Institut Pasteur, Paris |
| EPI_ISL_428362 | LABM GH nord Essonne de Longjumeau - BP 125 | National Reference Center for Viruses of Respiratory Infections, Institut Pasteur, Paris |
| EPI_ISL_428363 | GH Nord Essonne Service de Biologie clinique | National Reference Center for Viruses of Respiratory Infections, Institut Pasteur, Paris |
| EPI_ISL_428364 | Cabinet MÃ©dical | National Reference Center for Viruses of Respiratory Infections, Institut Pasteur, Paris |
| EPI_ISL_428365 | LABM GH nord Essonne de Longjumeau - BP 125 | National Reference Center for Viruses of Respiratory Infections, Institut Pasteur, Paris |
| EPI_ISL_428366 | CH Jeanne de Navarre Laboratoire de Biologie | National Reference Center for Viruses of Respiratory Infections, Institut Pasteur, Paris |
| EPI_ISL_428367 | Cabinet MÃ©dical | National Reference Center for Viruses of Respiratory Infections, Institut Pasteur, Paris |
| EPI_ISL_428368 | Genomic Laboratory (GLAB) (Conjoint lab of Health Directorate of Istanbul and Istanbul Technical University) | Genomic Laboratory (GLAB), Istanbul Technical University |
| EPI_ISL_428370 | Yale Clinical Virology Laboratory | Grubaugh Lab - Yale School of Public Health |
| EPI_ISL_428371 | Yale Clinical Virology Laboratory | Grubaugh Lab - Yale School of Public Health |
| EPI_ISL_428372 | Yale Clinical Virology Laboratory | Grubaugh Lab - Yale School of Public Health |
| EPI_ISL_428373 | Yale COVID-19 Biorepository | Grubaugh Lab - Yale School of Public Health |
| EPI_ISL_428374 | Yale COVID-19 Biorepository | Grubaugh Lab - Yale School of Public Health |
| EPI_ISL_428375 | Yale COVID-19 Biorepository | Grubaugh Lab - Yale School of Public Health |

| EPI_Accession_ID | Originating Laboratory | Submitting Laboratory |
| --- | --- | --- |
| EPI_ISL_428376 | Yale COVID-19 Biorepository | Grubaugh Lab - Yale School of Public Health |
| EPI_ISL_428377 | Yale COVID-19 Biorepository | Grubaugh Lab - Yale School of Public Health |
| EPI_ISL_428378 | Yale COVID-19 Biorepository | Grubaugh Lab - Yale School of Public Health |
| EPI_ISL_428379 | Yale COVID-19 Biorepository | Grubaugh Lab - Yale School of Public Health |
| EPI_ISL_428380 | Yale COVID-19 Biorepository | Grubaugh Lab - Yale School of Public Health |
| EPI_ISL_428381 | Yale Clinical Virology Laboratory | Grubaugh Lab - Yale School of Public Health |
| EPI_ISL_428382 | Yale Clinical Virology Laboratory | Grubaugh Lab - Yale School of Public Health |
| EPI_ISL_428384 | Yale COVID-19 Biorepository | Grubaugh Lab - Yale School of Public Health |
| EPI_ISL_428385 | Yale COVID-19 Biorepository | Grubaugh Lab - Yale School of Public Health |
| EPI_ISL_428386 | Yale COVID-19 Biorepository | Grubaugh Lab - Yale School of Public Health |
| EPI_ISL_428387 | Yale COVID-19 Biorepository | Grubaugh Lab - Yale School of Public Health |
| EPI_ISL_428388 | Yale COVID-19 Biorepository | Grubaugh Lab - Yale School of Public Health |
| EPI_ISL_428389 | Yale COVID-19 Biorepository | Grubaugh Lab - Yale School of Public Health |
| EPI_ISL_428390 | Yale COVID-19 Biorepository | Grubaugh Lab - Yale School of Public Health |
| EPI_ISL_428391 | Yale COVID-19 Biorepository | Grubaugh Lab - Yale School of Public Health |
| EPI_ISL_428393 | Yale COVID-19 Biorepository | Grubaugh Lab - Yale School of Public Health |
| EPI_ISL_428394 | Yale COVID-19 Biorepository | Grubaugh Lab - Yale School of Public Health |
| EPI_ISL_428395 | Yale COVID-19 Biorepository | Grubaugh Lab - Yale School of Public Health |
| EPI_ISL_428396 | Yale COVID-19 Biorepository | Grubaugh Lab - Yale School of Public Health |
| EPI_ISL_428397 | Yale COVID-19 Biorepository | Grubaugh Lab - Yale School of Public Health |
| EPI_ISL_428398 | Yale COVID-19 Biorepository | Grubaugh Lab - Yale School of Public Health |
| EPI_ISL_428399 | Yale COVID-19 Biorepository | Grubaugh Lab - Yale School of Public Health |
| EPI_ISL_428400 | Yale COVID-19 Biorepository | Grubaugh Lab - Yale School of Public Health |
| EPI_ISL_428401 | Yale COVID-19 Biorepository | Grubaugh Lab - Yale School of Public Health |
| EPI_ISL_428402 | Yale COVID-19 Biorepository | Grubaugh Lab - Yale School of Public Health |
| EPI_ISL_428403 | Yale COVID-19 Biorepository | Grubaugh Lab - Yale School of Public Health |
| EPI_ISL_428404 | Yale COVID-19 Biorepository | Grubaugh Lab - Yale School of Public Health |
| EPI_ISL_428405 | Yale COVID-19 Biorepository | Grubaugh Lab - Yale School of Public Health |
| EPI_ISL_428440 | Guangdong Provincial Center for Diseases Control and Prevention;Guangdong Provincial Institute of Public Health | School of Public Health, The University of Hong Kong |

| EPI_Accession_ID | Originating Laboratory | Submitting Laboratory |
|---|---|---|
| EPI_ISL_428441 | Guangdong Provincial Center for Diseases Control and Prevention;Guangdong Provincial Institute of Public Health | School of Public Health, The University of Hong Kong |
| EPI_ISL_428442 | Guangdong Provincial Center for Diseases Control and Prevention;Guangdong Provincial Institute of Public Health | School of Public Health, The University of Hong Kong |
| EPI_ISL_428443 | Guangdong Provincial Center for Diseases Control and Prevention;Guangdong Provincial Institute of Public Health | School of Public Health, The University of Hong Kong |
| EPI_ISL_428444 | Guangdong Provincial Center for Diseases Control and Prevention;Guangdong Provincial Institute of Public Health | School of Public Health, The University of Hong Kong |
| EPI_ISL_428445 | Guangdong Provincial Center for Diseases Control and Prevention;Guangdong Provincial Institute of Public Health | School of Public Health, The University of Hong Kong |
| EPI_ISL_428446 | Guangdong Provincial Center for Diseases Control and Prevention;Guangdong Provincial Institute of Public Health | School of Public Health, The University of Hong Kong |
| EPI_ISL_428447 | Guangdong Provincial Center for Diseases Control and Prevention;Guangdong Provincial Institute of Public Health | School of Public Health, The University of Hong Kong |
| EPI_ISL_428448 | Guangdong Provincial Center for Diseases Control and Prevention;Guangdong Provincial Institute of Public Health | School of Public Health, The University of Hong Kong |
| EPI_ISL_428449 | Guangdong Provincial Center for Diseases Control and Prevention;Guangdong Provincial Institute of Public Health | School of Public Health, The University of Hong Kong |
| EPI_ISL_428450 | Guangdong Provincial Center for Diseases Control and Prevention;Guangdong Provincial Institute of Public Health | School of Public Health, The University of Hong Kong |
| EPI_ISL_428451 | Guangdong Provincial Center for Diseases Control and Prevention;Guangdong Provincial Institute of Public Health | School of Public Health, The University of Hong Kong |

| EPI_Accession_ID | Originating Laboratory | Submitting Laboratory |
|---|---|---|
| EPI_ISL_428452 | Guangdong Provincial Center for Diseases Control and Prevention;Guangdong Provincial Institute of Public Health | School of Public Health, The University of Hong Kong |
| EPI_ISL_428453 | Guangdong Provincial Center for Diseases Control and Prevention;Guangdong Provincial Institute of Public Health | School of Public Health, The University of Hong Kong |
| EPI_ISL_428454 | Guangdong Provincial Center for Diseases Control and Prevention;Guangdong Provincial Institute of Public Health | School of Public Health, The University of Hong Kong |
| EPI_ISL_428455 | Guangdong Provincial Center for Diseases Control and Prevention;Guangdong Provincial Institute of Public Health | School of Public Health, The University of Hong Kong |
| EPI_ISL_428456 | Guangdong Provincial Center for Diseases Control and Prevention;Guangdong Provincial Institute of Public Health | School of Public Health, The University of Hong Kong |
| EPI_ISL_428457 | Guangdong Provincial Center for Diseases Control and Prevention;Guangdong Provincial Institute of Public Health | School of Public Health, The University of Hong Kong |
| EPI_ISL_428458 | Guangdong Provincial Center for Diseases Control and Prevention;Guangdong Provincial Institute of Public Health | School of Public Health, The University of Hong Kong |
| EPI_ISL_428459 | Guangdong Provincial Center for Diseases Control and Prevention;Guangdong Provincial Institute of Public Health | School of Public Health, The University of Hong Kong |
| EPI_ISL_428460 | Guangdong Provincial Center for Diseases Control and Prevention;Guangdong Provincial Institute of Public Health | School of Public Health, The University of Hong Kong |
| EPI_ISL_428461 | Guangdong Provincial Center for Diseases Control and Prevention;Guangdong Provincial Institute of Public Health | School of Public Health, The University of Hong Kong |
| EPI_ISL_428462 | Guangdong Provincial Center for Diseases Control and Prevention;Guangdong Provincial Institute of Public Health | School of Public Health, The University of Hong Kong |

| EPI_Accession_ID | Originating Laboratory | Submitting Laboratory |
|---|---|---|
| EPI_ISL_428463 | Guangdong Provincial Center for Diseases Control and Prevention;Guangdong Provincial Institute of Public Health | School of Public Health, The University of Hong Kong |
| EPI_ISL_428464 | Guangdong Provincial Center for Diseases Control and Prevention;Guangdong Provincial Institute of Public Health | School of Public Health, The University of Hong Kong |
| EPI_ISL_428465 | Guangdong Provincial Center for Diseases Control and Prevention;Guangdong Provincial Institute of Public Health | School of Public Health, The University of Hong Kong |
| EPI_ISL_428466 | Guangdong Provincial Center for Diseases Control and Prevention;Guangdong Provincial Institute of Public Health | School of Public Health, The University of Hong Kong |
| EPI_ISL_428467 | Guangdong Provincial Center for Diseases Control and Prevention;Guangdong Provincial Institute of Public Health | School of Public Health, The University of Hong Kong |
| EPI_ISL_428468 | Guangdong Provincial Center for Diseases Control and Prevention;Guangdong Provincial Institute of Public Health | School of Public Health, The University of Hong Kong |
| EPI_ISL_428469 | Guangdong Provincial Center for Diseases Control and Prevention;Guangdong Provincial Institute of Public Health | School of Public Health, The University of Hong Kong |
| EPI_ISL_428470 | Guangdong Provincial Center for Diseases Control and Prevention;Guangdong Provincial Institute of Public Health | School of Public Health, The University of Hong Kong |
| EPI_ISL_428471 | Guangdong Provincial Center for Diseases Control and Prevention;Guangdong Provincial Institute of Public Health | School of Public Health, The University of Hong Kong |
| EPI_ISL_428472 | Guangdong Provincial Center for Diseases Control and Prevention;Guangdong Provincial Institute of Public Health | School of Public Health, The University of Hong Kong |
| EPI_ISL_428473 | Guangdong Provincial Center for Diseases Control and Prevention;Guangdong Provincial Institute of Public Health | School of Public Health, The University of Hong Kong |

| EPI_Accession_ID | Originating Laboratory | Submitting Laboratory |
|---|---|---|
| EPI_ISL_428474 | Guangdong Provincial Center for Diseases Control and Prevention;Guangdong Provincial Institute of Public Health | School of Public Health, The University of Hong Kong |
| EPI_ISL_428475 | Guangdong Provincial Center for Diseases Control and Prevention;Guangdong Provincial Institute of Public Health | School of Public Health, The University of Hong Kong |
| EPI_ISL_428476 | Guangdong Provincial Center for Diseases Control and Prevention;Guangdong Provincial Institute of Public Health | School of Public Health, The University of Hong Kong |
| EPI_ISL_428477 | Guangdong Provincial Center for Diseases Control and Prevention;Guangdong Provincial Institute of Public Health | School of Public Health, The University of Hong Kong |
| EPI_ISL_428478 | Guangdong Provincial Center for Diseases Control and Prevention;Guangdong Provincial Institute of Public Health | School of Public Health, The University of Hong Kong |
| EPI_ISL_428479 | District Surveillence Unit | Department of Neurovirology, National Institute of Mental Health and Neuroscience (NIMHANS) |
| EPI_ISL_428480 | District Surveillence Unit | Department of Neurovirology, National Institute of Mental Health and Neuroscience (NIMHANS) |
| EPI_ISL_428481 | District Surveillence Unit | Department of Neurovirology, National Institute of Mental Health and Neuroscience (NIMHANS) |
| EPI_ISL_428482 | District Surveillence Unit | Department of Neurovirology, National Institute of Mental Health and Neuroscience (NIMHANS) |
| EPI_ISL_428483 | District Surveillence Unit | Department of Neurovirology, National Institute of Mental Health and Neuroscience (NIMHANS) |
| EPI_ISL_428484 | District Surveillence Unit | Department of Neurovirology, National Institute of Mental Health and Neuroscience (NIMHANS) |
| EPI_ISL_428485 | District Surveillence Unit | Department of Neurovirology, National Institute of Mental Health and Neuroscience (NIMHANS) |
| EPI_ISL_428486 | District Surveillence Unit | Department of Neurovirology, National Institute of Mental Health and Neuroscience (NIMHANS) |
| EPI_ISL_428487 | District Surveillence Unit | Department of Neurovirology, National Institute of Mental Health and Neuroscience (NIMHANS) |

| EPI_Accession_ID | Originating Laboratory | Submitting Laboratory |
|---|---|---|
| EPI_ISL_428488 | Centers for Disease Control, R.O.C. (Taiwan) | Centers for Disease Control, R.O.C. (Taiwan) |
| EPI_ISL_428489 | Centers for Disease Control, R.O.C. (Taiwan) | Centers for Disease Control, R.O.C. (Taiwan) |
| EPI_ISL_428490 | Centers for Disease Control, R.O.C. (Taiwan) | Centers for Disease Control, R.O.C. (Taiwan) |
| EPI_ISL_428491 | Centers for Disease Control, R.O.C. (Taiwan) | Centers for Disease Control, R.O.C. (Taiwan) |
| EPI_ISL_428670 | Centre for Dengue Research | Centre for Dengue Research |
| EPI_ISL_428671 | Centre for Dengue Research | Centre for Dengue Research |
| EPI_ISL_428672 | Centre for Dengue Research | Centre for Dengue Research |
| EPI_ISL_428673 | Centre for Dengue Research | Centre for Dengue Research |
| EPI_ISL_428674 | Hospital Universitario La Paz | Hospital Universitario 12 de Octubre |
| EPI_ISL_428675 | Hospital Universitario La Paz | Hospital Universitario 12 de Octubre |
| EPI_ISL_428676 | Hospital Universitario La Paz | Hospital Universitario 12 de Octubre |
| EPI_ISL_428677 | Hospital Universitario La Paz | Hospital Universitario 12 de Octubre |
| EPI_ISL_428678 | Hospital Universitario La Paz | Hospital Universitario 12 de Octubre |
| EPI_ISL_428679 | Hospital Universitario La Paz | Hospital Universitario 12 de Octubre |
| EPI_ISL_428680 | Hospital Universitario La Paz | Hospital Universitario 12 de Octubre |
| EPI_ISL_428681 | Hospital Universitario La Paz | Hospital Universitario 12 de Octubre |
| EPI_ISL_428682 | Hospital Universitario La Paz | Hospital Universitario 12 de Octubre |
| EPI_ISL_428683 | Hospital Universitario 12 de Octubre | Hospital Universitario 12 de Octubre |
| EPI_ISL_428684 | Hospital Universitario 12 de Octubre | Hospital Universitario 12 de Octubre |
| EPI_ISL_428685 | Hospital Universitario 12 de Octubre | Hospital Universitario 12 de Octubre |
| EPI_ISL_428686 | Hospital Universitario 12 de Octubre | Hospital Universitario 12 de Octubre |
| EPI_ISL_428687 | Hospital Universitario 12 de Octubre | Hospital Universitario 12 de Octubre |
| EPI_ISL_428688 | Hospital Universitario 12 de Octubre | Hospital Universitario 12 de Octubre |
| EPI_ISL_428689 | Hospital Universitario 12 de Octubre | Hospital Universitario 12 de Octubre |
| EPI_ISL_428690 | Hospital Universitario 12 de Octubre | Hospital Universitario 12 de Octubre |
| EPI_ISL_428691 | Hospital Universitario 12 de Octubre | Hospital Universitario 12 de Octubre |
| EPI_ISL_428692 | Hospital Universitario 12 de Octubre | Hospital Universitario 12 de Octubre |
| EPI_ISL_428693 | Hospital Universitario 12 de Octubre | Hospital Universitario 12 de Octubre |
| EPI_ISL_428694 | Hospital Universitario 12 de Octubre | Hospital Universitario 12 de Octubre |
| EPI_ISL_428695 | Hospital Universitario 12 de Octubre | Hospital Universitario 12 de Octubre |
| EPI_ISL_428696 | Hospital Universitario 12 de Octubre | Hospital Universitario 12 de Octubre |
| EPI_ISL_428697 | Hospital Universitario 12 de Octubre | Hospital Universitario 12 de Octubre |
| EPI_ISL_428698 | Hospital Universitario 12 de Octubre | Hospital Universitario 12 de Octubre |

| EPI_Accession_ID | Originating Laboratory | Submitting Laboratory |
|---|---|---|
| EPI_ISL_428699 | Hospital Universitario 12 de Octubre | Hospital Universitario 12 de Octubre |
| EPI_ISL_428700 | Hospital Universitario 12 de Octubre | Hospital Universitario 12 de Octubre |
| EPI_ISL_428701 | Hospital Universitario 12 de Octubre | Hospital Universitario 12 de Octubre |
| EPI_ISL_428702 | Hospital Universitario 12 de Octubre | Hospital Universitario 12 de Octubre |
| EPI_ISL_428703 | Hospital Universitario 12 de Octubre | Hospital Universitario 12 de Octubre |
| EPI_ISL_428704 | Hospital Universitario 12 de Octubre | Hospital Universitario 12 de Octubre |
| EPI_ISL_428705 | Hospital Universitario 12 de Octubre | Hospital Universitario 12 de Octubre |
| EPI_ISL_428706 | Hospital Universitario 12 de Octubre | Hospital Universitario 12 de Octubre |
| EPI_ISL_428707 | Hospital Universitario 12 de Octubre | Hospital Universitario 12 de Octubre |
| EPI_ISL_428708 | Hospital Universitario 12 de Octubre | Hospital Universitario 12 de Octubre |
| EPI_ISL_428709 | Hospital Universitario 12 de Octubre | Hospital Universitario 12 de Octubre |
| EPI_ISL_428710 | Hospital Universitario 12 de Octubre | Hospital Universitario 12 de Octubre |
| EPI_ISL_428711 | Hospital Universitario 12 de Octubre | Hospital Universitario 12 de Octubre |
| EPI_ISL_428712 | Ministry of Health Turkey | Ministry of Health Turkey |
| EPI_ISL_428713 | Ministry of Health Turkey | Ministry of Health Turkey |
| EPI_ISL_428714 | Ministry of Health Turkey | Ministry of Health Turkey |
| EPI_ISL_428715 | Ministry of Health Turkey | Ministry of Health Turkey |
| EPI_ISL_428716 | Ministry of Health Turkey | Ministry of Health Turkey |
| EPI_ISL_428717 | Ministry of Health Turkey | Ministry of Health Turkey |
| EPI_ISL_428718 | Ministry of Health Turkey | Ministry of Health Turkey |
| EPI_ISL_428719 | Ministry of Health Turkey | Ministry of Health Turkey |
| EPI_ISL_428720 | Ministry of Health Turkey | Ministry of Health Turkey |
| EPI_ISL_428721 | Ministry of Health Turkey | Ministry of Health Turkey |
| EPI_ISL_428722 | Ministry of Health Turkey | Ministry of Health Turkey |
| EPI_ISL_428723 | Ministry of Health Turkey | Ministry of Health Turkey |
| EPI_ISL_428724 | University of Wisconsin-Madison AIDS Vaccine Research Laboratories | University of Wisconsin-Madison AIDS Vaccine Research Laboratories |
| EPI_ISL_428725 | University of Wisconsin-Madison AIDS Vaccine Research Laboratories | University of Wisconsin-Madison AIDS Vaccine Research Laboratories |
| EPI_ISL_428726 | University of Wisconsin-Madison AIDS Vaccine Research Laboratories | University of Wisconsin-Madison AIDS Vaccine Research Laboratories |
| EPI_ISL_428727 | University of Wisconsin-Madison AIDS Vaccine Research Laboratories | University of Wisconsin-Madison AIDS Vaccine Research Laboratories |

| EPI_Accession_ID | Originating Laboratory | Submitting Laboratory |
|---|---|---|
| EPI_ISL_428728 | University of Wisconsin-Madison AIDS Vaccine Research Laboratories | University of Wisconsin-Madison AIDS Vaccine Research Laboratories |
| EPI_ISL_428729 | University of Wisconsin-Madison AIDS Vaccine Research Laboratories | University of Wisconsin-Madison AIDS Vaccine Research Laboratories |
| EPI_ISL_428730 | University of Wisconsin-Madison AIDS Vaccine Research Laboratories | University of Wisconsin-Madison AIDS Vaccine Research Laboratories |
| EPI_ISL_428731 | University of Wisconsin-Madison AIDS Vaccine Research Laboratories | University of Wisconsin-Madison AIDS Vaccine Research Laboratories |
| EPI_ISL_428732 | University of Wisconsin-Madison AIDS Vaccine Research Laboratories | University of Wisconsin-Madison AIDS Vaccine Research Laboratories |
| EPI_ISL_428740 | Yale Clinical Virology Laboratory | Grubaugh Lab - Yale School of Public Health |
| EPI_ISL_428745 | Yale COVID-19 Biorepository | Grubaugh Lab - Yale School of Public Health |
| EPI_ISL_428746 | Yale COVID-19 Biorepository | Grubaugh Lab - Yale School of Public Health |
| EPI_ISL_428747 | Yale COVID-19 Biorepository | Grubaugh Lab - Yale School of Public Health |
| EPI_ISL_428748 | Yale COVID-19 Biorepository | Grubaugh Lab - Yale School of Public Health |
| EPI_ISL_428749 | Yale COVID-19 Biorepository | Grubaugh Lab - Yale School of Public Health |
| EPI_ISL_428750 | Yale COVID-19 Biorepository | Grubaugh Lab - Yale School of Public Health |
| EPI_ISL_428751 | Yale COVID-19 Biorepository | Grubaugh Lab - Yale School of Public Health |
| EPI_ISL_428752 | Yale COVID-19 Biorepository | Grubaugh Lab - Yale School of Public Health |
| EPI_ISL_428757 | NYU Langone Health | Departments of Pathology and Medicine, New York University School of Medicine |
| EPI_ISL_428758 | NYU Langone Health | Departments of Pathology and Medicine, New York University School of Medicine |
| EPI_ISL_428759 | NYU Langone Health | Departments of Pathology and Medicine, New York University School of Medicine |
| EPI_ISL_428760 | NYU Langone Health | Departments of Pathology and Medicine, New York University School of Medicine |
| EPI_ISL_428761 | NYU Langone Health | Departments of Pathology and Medicine, New York University School of Medicine |
| EPI_ISL_428762 | NYU Langone Health | Departments of Pathology and Medicine, New York University School of Medicine |
| EPI_ISL_428763 | NYU Langone Health | Departments of Pathology and Medicine, New York University School of Medicine |

| EPI_Accession_ID | Originating Laboratory | Submitting Laboratory |
|---|---|---|
| EPI_ISL_428764 | NYU Langone Health | Departments of Pathology and Medicine, New York University School of Medicine |
| EPI_ISL_428765 | NYU Langone Health | Departments of Pathology and Medicine, New York University School of Medicine |
| EPI_ISL_428766 | NYU Langone Health | Departments of Pathology and Medicine, New York University School of Medicine |
| EPI_ISL_428767 | NYU Langone Health | Departments of Pathology and Medicine, New York University School of Medicine |
| EPI_ISL_428768 | NYU Langone Health | Departments of Pathology and Medicine, New York University School of Medicine |
| EPI_ISL_428769 | NYU Langone Health | Departments of Pathology and Medicine, New York University School of Medicine |
| EPI_ISL_428770 | NYU Langone Health | Departments of Pathology and Medicine, New York University School of Medicine |
| EPI_ISL_428771 | NYU Langone Health | Departments of Pathology and Medicine, New York University School of Medicine |
| EPI_ISL_428772 | NYU Langone Health | Departments of Pathology and Medicine, New York University School of Medicine |
| EPI_ISL_428773 | NYU Langone Health | Departments of Pathology and Medicine, New York University School of Medicine |
| EPI_ISL_428774 | NYU Langone Health | Departments of Pathology and Medicine, New York University School of Medicine |
| EPI_ISL_428775 | NYU Langone Health | Departments of Pathology and Medicine, New York University School of Medicine |
| EPI_ISL_428776 | NYU Langone Health | Departments of Pathology and Medicine, New York University School of Medicine |
| EPI_ISL_428777 | NYU Langone Health | Departments of Pathology and Medicine, New York University School of Medicine |
| EPI_ISL_428778 | NYU Langone Health | Departments of Pathology and Medicine, New York University School of Medicine |
| EPI_ISL_428779 | NYU Langone Health | Departments of Pathology and Medicine, New York University School of Medicine |

| EPI_Accession_ID | Originating Laboratory | Submitting Laboratory |
|---|---|---|
| EPI_ISL_428780 | NYU Langone Health | Departments of Pathology and Medicine, New York University School of Medicine |
| EPI_ISL_428781 | NYU Langone Health | Departments of Pathology and Medicine, New York University School of Medicine |
| EPI_ISL_428782 | NYU Langone Health | Departments of Pathology and Medicine, New York University School of Medicine |
| EPI_ISL_428783 | NYU Langone Health | Departments of Pathology and Medicine, New York University School of Medicine |
| EPI_ISL_428784 | NYU Langone Health | Departments of Pathology and Medicine, New York University School of Medicine |
| EPI_ISL_428785 | NYU Langone Health | Departments of Pathology and Medicine, New York University School of Medicine |
| EPI_ISL_428786 | NYU Langone Health | Departments of Pathology and Medicine, New York University School of Medicine |
| EPI_ISL_428787 | NYU Langone Health | Departments of Pathology and Medicine, New York University School of Medicine |
| EPI_ISL_428788 | NYU Langone Health | Departments of Pathology and Medicine, New York University School of Medicine |
| EPI_ISL_428789 | NYU Langone Health | Departments of Pathology and Medicine, New York University School of Medicine |
| EPI_ISL_428790 | NYU Langone Health | Departments of Pathology and Medicine, New York University School of Medicine |
| EPI_ISL_428791 | NYU Langone Health | Departments of Pathology and Medicine, New York University School of Medicine |
| EPI_ISL_428792 | NYU Langone Health | Departments of Pathology and Medicine, New York University School of Medicine |
| EPI_ISL_428793 | NYU Langone Health | Departments of Pathology and Medicine, New York University School of Medicine |
| EPI_ISL_428794 | NYU Langone Health | Departments of Pathology and Medicine, New York University School of Medicine |
| EPI_ISL_428795 | NYU Langone Health | Departments of Pathology and Medicine, New York University School of Medicine |

| EPI_Accession_ID | Originating Laboratory | Submitting Laboratory |
| --- | --- | --- |
| EPI_ISL_428796 | NYU Langone Health | Departments of Pathology and Medicine, New York University School of Medicine |
| EPI_ISL_428797 | NYU Langone Health | Departments of Pathology and Medicine, New York University School of Medicine |
| EPI_ISL_428798 | NYU Langone Health | Departments of Pathology and Medicine, New York University School of Medicine |
| EPI_ISL_428799 | NYU Langone Health | Departments of Pathology and Medicine, New York University School of Medicine |
| EPI_ISL_428800 | NYU Langone Health | Departments of Pathology and Medicine, New York University School of Medicine |
| EPI_ISL_428801 | NYU Langone Health | Departments of Pathology and Medicine, New York University School of Medicine |
| EPI_ISL_428802 | NYU Langone Health | Departments of Pathology and Medicine, New York University School of Medicine |
| EPI_ISL_428803 | NYU Langone Health | Departments of Pathology and Medicine, New York University School of Medicine |
| EPI_ISL_428804 | NYU Langone Health | Departments of Pathology and Medicine, New York University School of Medicine |
| EPI_ISL_428805 | NYU Langone Health | Departments of Pathology and Medicine, New York University School of Medicine |
| EPI_ISL_428822 | National Public Health Laboratory, National Centre for Infectious Diseases | National Public Health Laboratory, National Centre for Infectious Diseases |
| EPI_ISL_428823 | National Public Health Laboratory, National Centre for Infectious Diseases | National Public Health Laboratory, National Centre for Infectious Diseases |
| EPI_ISL_428824 | National Public Health Laboratory, National Centre for Infectious Diseases | National Public Health Laboratory, National Centre for Infectious Diseases |
| EPI_ISL_428825 | National Public Health Laboratory, National Centre for Infectious Diseases | National Public Health Laboratory, National Centre for Infectious Diseases |
| EPI_ISL_428826 | National Public Health Laboratory, National Centre for Infectious Diseases | National Public Health Laboratory, National Centre for Infectious Diseases |
| EPI_ISL_428827 | National Public Health Laboratory, National Centre for Infectious Diseases | National Public Health Laboratory, National Centre for Infectious Diseases |

| EPI_Accession_ID | Originating Laboratory | Submitting Laboratory |
|---|---|---|
| EPI_ISL_428828 | National Public Health Laboratory, National Centre for Infectious Diseases | National Public Health Laboratory, National Centre for Infectious Diseases |
| EPI_ISL_428829 | National Public Health Laboratory, National Centre for Infectious Diseases | National Public Health Laboratory, National Centre for Infectious Diseases |
| EPI_ISL_428830 | National Public Health Laboratory, National Centre for Infectious Diseases | National Public Health Laboratory, National Centre for Infectious Diseases |
| EPI_ISL_428831 | National Public Health Laboratory, National Centre for Infectious Diseases | National Public Health Laboratory, National Centre for Infectious Diseases |
| EPI_ISL_428832 | National Public Health Laboratory, National Centre for Infectious Diseases | National Public Health Laboratory, National Centre for Infectious Diseases |
| EPI_ISL_428833 | National Public Health Laboratory, National Centre for Infectious Diseases | National Public Health Laboratory, National Centre for Infectious Diseases |
| EPI_ISL_428834 | National Public Health Laboratory, National Centre for Infectious Diseases | National Public Health Laboratory, National Centre for Infectious Diseases |
| EPI_ISL_428835 | National Public Health Laboratory, National Centre for Infectious Diseases | National Public Health Laboratory, National Centre for Infectious Diseases |
| EPI_ISL_428836 | National Public Health Laboratory, National Centre for Infectious Diseases | National Public Health Laboratory, National Centre for Infectious Diseases |
| EPI_ISL_428837 | National Public Health Laboratory, National Centre for Infectious Diseases | National Public Health Laboratory, National Centre for Infectious Diseases |
| EPI_ISL_428838 | National Public Health Laboratory, National Centre for Infectious Diseases | National Public Health Laboratory, National Centre for Infectious Diseases |
| EPI_ISL_428839 | National Public Health Laboratory, National Centre for Infectious Diseases | National Public Health Laboratory, National Centre for Infectious Diseases |
| EPI_ISL_428840 | National Public Health Laboratory, National Centre for Infectious Diseases | National Public Health Laboratory, National Centre for Infectious Diseases |
| EPI_ISL_428841 | National Public Health Laboratory, National Centre for Infectious Diseases | National Public Health Laboratory, National Centre for Infectious Diseases |
| EPI_ISL_428842 | National Public Health Laboratory, National Centre for Infectious Diseases | National Public Health Laboratory, National Centre for Infectious Diseases |
| EPI_ISL_428843 | National Public Health Laboratory, National Centre for Infectious Diseases | National Public Health Laboratory, National Centre for Infectious Diseases |

| EPI_Accession_ID | Originating Laboratory | Submitting Laboratory |
|---|---|---|
| EPI_ISL_428844 | National Public Health Laboratory, National Centre for Infectious Diseases | National Public Health Laboratory, National Centre for Infectious Diseases |
| EPI_ISL_428845 | National Public Health Laboratory, National Centre for Infectious Diseases | National Public Health Laboratory, National Centre for Infectious Diseases |
| EPI_ISL_428846 | National Public Health Laboratory, National Centre for Infectious Diseases | National Public Health Laboratory, National Centre for Infectious Diseases |
| EPI_ISL_428847 | National Public Health Laboratory, National Centre for Infectious Diseases | National Public Health Laboratory, National Centre for Infectious Diseases |
| EPI_ISL_428848 | National Public Health Laboratory, National Centre for Infectious Diseases | National Public Health Laboratory, National Centre for Infectious Diseases |
| EPI_ISL_428849 | National Public Health Laboratory, National Centre for Infectious Diseases | National Public Health Laboratory, National Centre for Infectious Diseases |
| EPI_ISL_428850 | National Public Health Laboratory, National Centre for Infectious Diseases | National Public Health Laboratory, National Centre for Infectious Diseases |
| EPI_ISL_428851 | FSBSI "Chumakov Federal Scientific Center for Research and Development of Immune-and-Biological Products of Russian Academy of Sciences" | FSBSI "Chumakov Federal Scientific Center for Research and Development of Immune-and-Biological Products of Russian Academy of Sciences" & NRC "Kurchatov institute" |
| EPI_ISL_428852 | FSBSI "Chumakov Federal Scientific Center for Research and Development of Immune-and-Biological Products of Russian Academy of Sciences" | FSBSI "Chumakov Federal Scientific Center for Research and Development of Immune-and-Biological Products of Russian Academy of Sciences" & NRC "Kurchatov institute" |
| EPI_ISL_428853 | Laboratory of Molecular Virology International Center for Genetic Engineering and Biotechnology (ICGEB) | ARGO Open Lab Platform for Genome Sequencing |
| EPI_ISL_428854 | Laboratory of Molecular Virology International Center for Genetic Engineering and Biotechnology (ICGEB) | ARGO Open Lab Platform for Genome sequencing |
| EPI_ISL_428855 | MRCG at LSHTM Geomics lab | MRCG at LSHTM Genomics lab |
| EPI_ISL_428856 | MRCG at LSHTM Genomics Lab | MRCG at LSHTM Genomics lab |
| EPI_ISL_428857 | MRCG at LSHTM Genomics lab | MRCG at LSHTM Genomics lab |
| EPI_ISL_428860 | State Research Center of Virology and Biotechnology VECTOR, Department of Collection of Microorganisms | State Research Center of Virology and Biotechnology VECTOR, Department of Collection of Microorganisms |
| EPI_ISL_428861 | State Research Center of Virology and Biotechnology VECTOR, Department of Collection of Microorganisms | State Research Center of Virology and Biotechnology VECTOR, Department of Collection of Microorganisms |

| EPI_Accession_ID | Originating Laboratory | Submitting Laboratory |
|---|---|---|
| EPI_ISL_428862 | State Research Center of Virology and Biotechnology VECTOR, Department of Collection of Microorganisms | State Research Center of Virology and Biotechnology VECTOR, Department of Collection of Microorganisms |
| EPI_ISL_428863 | State Research Center of Virology and Biotechnology VECTOR, Department of Collection of Microorganisms | State Research Center of Virology and Biotechnology VECTOR, Department of Collection of Microorganisms |
| EPI_ISL_428864 | State Research Center of Virology and Biotechnology VECTOR, Department of Collection of Microorganisms | State Research Center of Virology and Biotechnology VECTOR, Department of Collection of Microorganisms |
| EPI_ISL_428865 | State Research Center of Virology and Biotechnology VECTOR, Department of Collection of Microorganisms | State Research Center of Virology and Biotechnology VECTOR, Department of Collection of Microorganisms |
| EPI_ISL_428866 | State Research Center of Virology and Biotechnology VECTOR, Department of Collection of Microorganisms | State Research Center of Virology and Biotechnology VECTOR, Department of Collection of Microorganisms |
| EPI_ISL_428867 | State Research Center of Virology and Biotechnology VECTOR, Department of Collection of Microorganisms | State Research Center of Virology and Biotechnology VECTOR, Department of Collection of Microorganisms |
| EPI_ISL_428868 | State Research Center of Virology and Biotechnology VECTOR, Department of Collection of Microorganisms | State Research Center of Virology and Biotechnology VECTOR, Department of Collection of Microorganisms |
| EPI_ISL_428869 | State Research Center of Virology and Biotechnology VECTOR, Department of Collection of Microorganisms | State Research Center of Virology and Biotechnology VECTOR, Department of Collection of Microorganisms |
| EPI_ISL_428870 | State Research Center of Virology and Biotechnology VECTOR, Department of Collection of Microorganisms | State Research Center of Virology and Biotechnology VECTOR, Department of Collection of Microorganisms |
| EPI_ISL_428871 | State Research Center of Virology and Biotechnology VECTOR, Department of Collection of Microorganisms | State Research Center of Virology and Biotechnology VECTOR, Department of Collection of Microorganisms |
| EPI_ISL_428872 | State Research Center of Virology and Biotechnology VECTOR, Department of Collection of Microorganisms | State Research Center of Virology and Biotechnology VECTOR, Department of Collection of Microorganisms |
| EPI_ISL_428873 | State Research Center of Virology and Biotechnology VECTOR, Department of Collection of Microorganisms | State Research Center of Virology and Biotechnology VECTOR, Department of Collection of Microorganisms |
| EPI_ISL_428874 | State Research Center of Virology and Biotechnology VECTOR, Department of Collection of Microorganisms | State Research Center of Virology and Biotechnology VECTOR, Department of Collection of Microorganisms |
| EPI_ISL_428875 | State Research Center of Virology and Biotechnology VECTOR, Department of Collection of Microorganisms | State Research Center of Virology and Biotechnology VECTOR, Department of Collection of Microorganisms |
| EPI_ISL_428876 | State Research Center of Virology and Biotechnology VECTOR, Department of Collection of Microorganisms | State Research Center of Virology and Biotechnology VECTOR, Department of Collection of Microorganisms |
| EPI_ISL_428877 | State Research Center of Virology and Biotechnology VECTOR, Department of Collection of Microorganisms | State Research Center of Virology and Biotechnology VECTOR, Department of Collection of Microorganisms |

| EPI_Accession_ID | Originating Laboratory | Submitting Laboratory |
|---|---|---|
| EPI_ISL_428878 | State Research Center of Virology and Biotechnology VECTOR, Department of Collection of Microorganisms | State Research Center of Virology and Biotechnology VECTOR, Department of Collection of Microorganisms |
| EPI_ISL_428879 | State Research Center of Virology and Biotechnology VECTOR, Department of Collection of Microorganisms | State Research Center of Virology and Biotechnology VECTOR, Department of Collection of Microorganisms |
| EPI_ISL_428880 | State Research Center of Virology and Biotechnology VECTOR, Department of Collection of Microorganisms | State Research Center of Virology and Biotechnology VECTOR, Department of Collection of Microorganisms |
| EPI_ISL_428881 | State Research Center of Virology and Biotechnology VECTOR, Department of Collection of Microorganisms | State Research Center of Virology and Biotechnology VECTOR, Department of Collection of Microorganisms |
| EPI_ISL_428882 | State Research Center of Virology and Biotechnology VECTOR, Department of Collection of Microorganisms | State Research Center of Virology and Biotechnology VECTOR, Department of Collection of Microorganisms |
| EPI_ISL_428883 | State Research Center of Virology and Biotechnology VECTOR, Department of Collection of Microorganisms | State Research Center of Virology and Biotechnology VECTOR, Department of Collection of Microorganisms |
| EPI_ISL_428884 | State Research Center of Virology and Biotechnology VECTOR, Department of Collection of Microorganisms | State Research Center of Virology and Biotechnology VECTOR, Department of Collection of Microorganisms |
| EPI_ISL_428885 | State Research Center of Virology and Biotechnology VECTOR, Department of Collection of Microorganisms | State Research Center of Virology and Biotechnology VECTOR, Department of Collection of Microorganisms |
| EPI_ISL_428886 | State Research Center of Virology and Biotechnology VECTOR, Department of Collection of Microorganisms | State Research Center of Virology and Biotechnology VECTOR, Department of Collection of Microorganisms |
| EPI_ISL_428887 | State Research Center of Virology and Biotechnology VECTOR, Department of Collection of Microorganisms | State Research Center of Virology and Biotechnology VECTOR, Department of Collection of Microorganisms |
| EPI_ISL_428888 | State Research Center of Virology and Biotechnology VECTOR, Department of Collection of Microorganisms | State Research Center of Virology and Biotechnology VECTOR, Department of Collection of Microorganisms |
| EPI_ISL_428889 | State Research Center of Virology and Biotechnology VECTOR, Department of Collection of Microorganisms | State Research Center of Virology and Biotechnology VECTOR, Department of Collection of Microorganisms |
| EPI_ISL_428890 | State Research Center of Virology and Biotechnology VECTOR, Department of Collection of Microorganisms | State Research Center of Virology and Biotechnology VECTOR, Department of Collection of Microorganisms |
| EPI_ISL_428891 | State Research Center of Virology and Biotechnology VECTOR, Department of Collection of Microorganisms | State Research Center of Virology and Biotechnology VECTOR, Department of Collection of Microorganisms |
| EPI_ISL_428892 | State Research Center of Virology and Biotechnology VECTOR, Department of Collection of Microorganisms | State Research Center of Virology and Biotechnology VECTOR, Department of Collection of Microorganisms |
| EPI_ISL_428893 | State Research Center of Virology and Biotechnology VECTOR, Department of Collection of Microorganisms | State Research Center of Virology and Biotechnology VECTOR, Department of Collection of Microorganisms |

| EPI_Accession_ID | Originating Laboratory | Submitting Laboratory |
|---|---|---|
| EPI_ISL_428894 | State Research Center of Virology and Biotechnology VECTOR, Department of Collection of Microorganisms | State Research Center of Virology and Biotechnology VECTOR, Department of Collection of Microorganisms |
| EPI_ISL_428895 | State Research Center of Virology and Biotechnology VECTOR, Department of Collection of Microorganisms | State Research Center of Virology and Biotechnology VECTOR, Department of Collection of Microorganisms |
| EPI_ISL_428896 | State Research Center of Virology and Biotechnology VECTOR, Department of Collection of Microorganisms | State Research Center of Virology and Biotechnology VECTOR, Department of Collection of Microorganisms |
| EPI_ISL_428897 | State Research Center of Virology and Biotechnology VECTOR, Department of Collection of Microorganisms | State Research Center of Virology and Biotechnology VECTOR, Department of Collection of Microorganisms |
| EPI_ISL_428898 | State Research Center of Virology and Biotechnology VECTOR, Department of Collection of Microorganisms | State Research Center of Virology and Biotechnology VECTOR, Department of Collection of Microorganisms |
| EPI_ISL_428899 | State Research Center of Virology and Biotechnology VECTOR, Department of Collection of Microorganisms | State Research Center of Virology and Biotechnology VECTOR, Department of Collection of Microorganisms |
| EPI_ISL_428900 | State Research Center of Virology and Biotechnology VECTOR, Department of Collection of Microorganisms | State Research Center of Virology and Biotechnology VECTOR, Department of Collection of Microorganisms |
| EPI_ISL_428901 | State Research Center of Virology and Biotechnology VECTOR, Department of Collection of Microorganisms | State Research Center of Virology and Biotechnology VECTOR, Department of Collection of Microorganisms |
| EPI_ISL_428902 | State Research Center of Virology and Biotechnology VECTOR, Department of Collection of Microorganisms | State Research Center of Virology and Biotechnology VECTOR, Department of Collection of Microorganisms |
| EPI_ISL_428903 | State Research Center of Virology and Biotechnology VECTOR, Department of Collection of Microorganisms | State Research Center of Virology and Biotechnology VECTOR, Department of Collection of Microorganisms |
| EPI_ISL_428904 | State Research Center of Virology and Biotechnology VECTOR, Department of Collection of Microorganisms | State Research Center of Virology and Biotechnology VECTOR, Department of Collection of Microorganisms |
| EPI_ISL_428905 | State Research Center of Virology and Biotechnology VECTOR, Department of Collection of Microorganisms | State Research Center of Virology and Biotechnology VECTOR, Department of Collection of Microorganisms |
| EPI_ISL_428906 | State Research Center of Virology and Biotechnology VECTOR, Department of Collection of Microorganisms | State Research Center of Virology and Biotechnology VECTOR, Department of Collection of Microorganisms |
| EPI_ISL_428907 | State Research Center of Virology and Biotechnology VECTOR, Department of Collection of Microorganisms | State Research Center of Virology and Biotechnology VECTOR, Department of Collection of Microorganisms |
| EPI_ISL_428908 | State Research Center of Virology and Biotechnology VECTOR, Department of Collection of Microorganisms | State Research Center of Virology and Biotechnology VECTOR, Department of Collection of Microorganisms |
| EPI_ISL_428909 | State Research Center of Virology and Biotechnology VECTOR, Department of Collection of Microorganisms | State Research Center of Virology and Biotechnology VECTOR, Department of Collection of Microorganisms |

| EPI_Accession_ID | Originating Laboratory | Submitting Laboratory |
| --- | --- | --- |
| EPI_ISL_428910 | State Research Center of Virology and Biotechnology VECTOR, Department of Collection of Microorganisms | State Research Center of Virology and Biotechnology VECTOR, Department of Collection of Microorganisms |
| EPI_ISL_428911 | State Research Center of Virology and Biotechnology VECTOR, Department of Collection of Microorganisms | State Research Center of Virology and Biotechnology VECTOR, Department of Collection of Microorganisms |
| EPI_ISL_428912 | State Research Center of Virology and Biotechnology VECTOR, Department of Collection of Microorganisms | State Research Center of Virology and Biotechnology VECTOR, Department of Collection of Microorganisms |
| EPI_ISL_428913 | State Research Center of Virology and Biotechnology VECTOR, Department of Collection of Microorganisms | State Research Center of Virology and Biotechnology VECTOR, Department of Collection of Microorganisms |
| EPI_ISL_428914 | State Research Center of Virology and Biotechnology VECTOR, Department of Collection of Microorganisms | State Research Center of Virology and Biotechnology VECTOR, Department of Collection of Microorganisms |
| EPI_ISL_428915 | State Research Center of Virology and Biotechnology VECTOR, Department of Collection of Microorganisms | State Research Center of Virology and Biotechnology VECTOR, Department of Collection of Microorganisms |
| EPI_ISL_428916 | State Research Center of Virology and Biotechnology VECTOR, Department of Collection of Microorganisms | State Research Center of Virology and Biotechnology VECTOR, Department of Collection of Microorganisms |
| EPI_ISL_428917 | State Research Center of Virology and Biotechnology VECTOR, Department of Collection of Microorganisms | State Research Center of Virology and Biotechnology VECTOR, Department of Collection of Microorganisms |
| EPI_ISL_428918 | State Research Center of Virology and Biotechnology VECTOR, Department of Collection of Microorganisms | State Research Center of Virology and Biotechnology VECTOR, Department of Collection of Microorganisms |
| EPI_ISL_428919 | State Research Center of Virology and Biotechnology VECTOR, Department of Collection of Microorganisms | State Research Center of Virology and Biotechnology VECTOR, Department of Collection of Microorganisms |
| EPI_ISL_428920 | State Research Center of Virology and Biotechnology VECTOR, Department of Collection of Microorganisms | State Research Center of Virology and Biotechnology VECTOR, Department of Collection of Microorganisms |
| EPI_ISL_428921 | State Research Center of Virology and Biotechnology VECTOR, Department of Collection of Microorganisms | State Research Center of Virology and Biotechnology VECTOR, Department of Collection of Microorganisms |
| EPI_ISL_428922 | State Research Center of Virology and Biotechnology VECTOR, Department of Collection of Microorganisms | State Research Center of Virology and Biotechnology VECTOR, Department of Collection of Microorganisms |
| EPI_ISL_428923 | State Research Center of Virology and Biotechnology VECTOR, Department of Collection of Microorganisms | State Research Center of Virology and Biotechnology VECTOR, Department of Collection of Microorganisms |
| EPI_ISL_428924 | State Research Center of Virology and Biotechnology VECTOR, Department of Collection of Microorganisms | State Research Center of Virology and Biotechnology VECTOR, Department of Collection of Microorganisms |

| EPI_Accession_ID | Originating Laboratory | Submitting Laboratory |
|---|---|---|
| EPI_ISL_428925 | ViroGenetics - BSL3 Laboratory of Virology; Human Genome Variation Research Group & Genomics Centre MCB; Bioinformatics Research Group; WojewÃ³dzka Stacja Sanitarno-Epidemiologiczna w Krakowie | ViroGenetics - BSL3 Laboratory of Virology; Human Genome Variation Research Group & Genomics Centre MCB; Bioinformatics Research Group |
| EPI_ISL_428926 | ViroGenetics - BSL3 Laboratory of Virology; Human Genome Variation Research Group & Genomics Centre MCB; Bioinformatics Research Group; WojewÃ³dzka Stacja Sanitarno-Epidemiologiczna w Krakowie | ViroGenetics - BSL3 Laboratory of Virology; Human Genome Variation Research Group & Genomics Centre MCB; Bioinformatics Research Group |
| EPI_ISL_428927 | ViroGenetics - BSL3 Laboratory of Virology; Human Genome Variation Research Group & Genomics Centre MCB; Bioinformatics Research Group; WojewÃ³dzka Stacja Sanitarno-Epidemiologiczna w Krakowie | ViroGenetics - BSL3 Laboratory of Virology; Human Genome Variation Research Group & Genomics Centre MCB; Bioinformatics Research Group |
| EPI_ISL_428928 | ViroGenetics - BSL3 Laboratory of Virology; Human Genome Variation Research Group & Genomics Centre MCB; Bioinformatics Research Group; WojewÃ³dzka Stacja Sanitarno-Epidemiologiczna w Krakowie | ViroGenetics - BSL3 Laboratory of Virology; Human Genome Variation Research Group & Genomics Centre MCB; Bioinformatics Research Group |
| EPI_ISL_428929 | ViroGenetics - BSL3 Laboratory of Virology; Human Genome Variation Research Group & Genomics Centre MCB; Bioinformatics Research Group; WojewÃ³dzka Stacja Sanitarno-Epidemiologiczna w Krakowie | ViroGenetics - BSL3 Laboratory of Virology; Human Genome Variation Research Group & Genomics Centre MCB; Bioinformatics Research Group |
| EPI_ISL_428930 | ViroGenetics - BSL3 Laboratory of Virology; Human Genome Variation Research Group & Genomics Centre MCB; Bioinformatics Research Group; WojewÃ³dzka Stacja Sanitarno-Epidemiologiczna w Krakowie | ViroGenetics - BSL3 Laboratory of Virology; Human Genome Variation Research Group & Genomics Centre MCB; Bioinformatics Research Group |
| EPI_ISL_428931 | ViroGenetics - BSL3 Laboratory of Virology; Human Genome Variation Research Group & Genomics Centre MCB; Bioinformatics Research Group; WojewÃ³dzka Stacja Sanitarno-Epidemiologiczna w Krakowie | ViroGenetics - BSL3 Laboratory of Virology; Human Genome Variation Research Group & Genomics Centre MCB; Bioinformatics Research Group |
| EPI_ISL_428932 | ViroGenetics - BSL3 Laboratory of Virology; Human Genome Variation Research Group & Genomics Centre MCB; Bioinformatics Research Group; WojewÃ³dzka Stacja Sanitarno-Epidemiologiczna w Krakowie | ViroGenetics - BSL3 Laboratory of Virology; Human Genome Variation Research Group & Genomics Centre MCB; Bioinformatics Research Group |

| EPI_Accession_ID | Originating Laboratory | Submitting Laboratory |
|---|---|---|
| EPI_ISL_428935 | University of Wisconsin-Madison AIDS Vaccine Research Laboratories | University of Wisconsin-Madison AIDS Vaccine Research Laboratories |
| EPI_ISL_428936 | University of Wisconsin-Madison AIDS Vaccine Research Laboratories | University of Wisconsin-Madison AIDS Vaccine Research Laboratories |
| EPI_ISL_428939 | Laboratoire National de Sante, Microbiology, Virology | Laboratoire National de Sante, Microbiology, Epidemiology and Microbial Genomics |
| EPI_ISL_428940 | Laboratoire National de Sante, Microbiology, Virology | Laboratoire National de Sante, Microbiology, Epidemiology and Microbial Genomics |
| EPI_ISL_428941 | Laboratoire National de Sante, Microbiology, Virology | Laboratoire National de Sante, Microbiology, Epidemiology and Microbial Genomics |
| EPI_ISL_428942 | Laboratoire National de Sante, Microbiology, Virology | Laboratoire National de Sante, Microbiology, Epidemiology and Microbial Genomics |
| EPI_ISL_428943 | Laboratoire National de Sante, Microbiology, Virology | Laboratoire National de Sante, Microbiology, Epidemiology and Microbial Genomics |
| EPI_ISL_428944 | Laboratoire National de Sante, Microbiology, Virology | Laboratoire National de Sante, Microbiology, Epidemiology and Microbial Genomics |
| EPI_ISL_428945 | Laboratoire National de Sante, Microbiology, Virology | Laboratoire National de Sante, Microbiology, Epidemiology and Microbial Genomics |
| EPI_ISL_428946 | Laboratoire National de Sante, Microbiology, Virology | Laboratoire National de Sante, Microbiology, Epidemiology and Microbial Genomics |
| EPI_ISL_428947 | Laboratoire National de Sante, Microbiology, Virology | Laboratoire National de Sante, Microbiology, Epidemiology and Microbial Genomics |
| EPI_ISL_428948 | Laboratoire National de Sante, Microbiology, Virology | Laboratoire National de Sante, Microbiology, Epidemiology and Microbial Genomics |
| EPI_ISL_428949 | Laboratoire National de Sante, Microbiology, Virology | Laboratoire National de Sante, Microbiology, Epidemiology and Microbial Genomics |
| EPI_ISL_428950 | Laboratoire National de Sante, Microbiology, Virology | Laboratoire National de Sante, Microbiology, Epidemiology and Microbial Genomics |
| EPI_ISL_428951 | Laboratoire National de Sante, Microbiology, Virology | Laboratoire National de Sante, Microbiology, Epidemiology and Microbial Genomics |
| EPI_ISL_428952 | Laboratoire National de Sante, Microbiology, Virology | Laboratoire National de Sante, Microbiology, Epidemiology and Microbial Genomics |

| EPI_Accession_ID | Originating Laboratory | Submitting Laboratory |
|---|---|---|
| EPI_ISL_428953 | Laboratoire National de Sante, Microbiology, Virology | Laboratoire National de Sante, Microbiology, Epidemiology and Microbial Genomics |
| EPI_ISL_428954 | Laboratoire National de Sante, Microbiology, Virology | Laboratoire National de Sante, Microbiology, Epidemiology and Microbial Genomics |
| EPI_ISL_428955 | Laboratoire National de Sante, Microbiology, Virology | Laboratoire National de Sante, Microbiology, Epidemiology and Microbial Genomics |
| EPI_ISL_428956 | Laboratoire National de Sante, Microbiology, Virology | Laboratoire National de Sante, Microbiology, Epidemiology and Microbial Genomics |
| EPI_ISL_428957 | Laboratoire National de Sante, Microbiology, Virology | Laboratoire National de Sante, Microbiology, Epidemiology and Microbial Genomics |
| EPI_ISL_428958 | Laboratoire National de Sante, Microbiology, Virology | Laboratoire National de Sante, Microbiology, Epidemiology and Microbial Genomics |
| EPI_ISL_428959 | Laboratoire National de Sante, Microbiology, Virology | Laboratoire National de Sante, Microbiology, Epidemiology and Microbial Genomics |
| EPI_ISL_428960 | Laboratoire National de Sante, Microbiology, Virology | Laboratoire National de Sante, Microbiology, Epidemiology and Microbial Genomics |
| EPI_ISL_428961 | Laboratoire National de Sante, Microbiology, Virology | Laboratoire National de Sante, Microbiology, Epidemiology and Microbial Genomics |
| EPI_ISL_428962 | Laboratoire National de Sante, Microbiology, Virology | Laboratoire National de Sante, Microbiology, Epidemiology and Microbial Genomics |
| EPI_ISL_428990 | UCSF Clinical Microbiology Laboratory | Chan-Zuckerberg Biohub |
| EPI_ISL_428991 | UCSF Clinical Microbiology Laboratory | Chan-Zuckerberg Biohub |
| EPI_ISL_428992 | UCSF Clinical Microbiology Laboratory | Chan-Zuckerberg Biohub |
| EPI_ISL_428993 | UCSF Clinical Microbiology Laboratory | Chan-Zuckerberg Biohub |
| EPI_ISL_428994 | UCSF Clinical Microbiology Laboratory | Chan-Zuckerberg Biohub |
| EPI_ISL_428995 | UCSF Clinical Microbiology Laboratory | Chan-Zuckerberg Biohub |
| EPI_ISL_428996 | UCSF Clinical Microbiology Laboratory | Chan-Zuckerberg Biohub |
| EPI_ISL_428997 | UCSF Clinical Microbiology Laboratory | Chan-Zuckerberg Biohub |
| EPI_ISL_428998 | UCSF Clinical Microbiology Laboratory | Chan-Zuckerberg Biohub |
| EPI_ISL_428999 | UCSF Clinical Microbiology Laboratory | Chan-Zuckerberg Biohub |
| EPI_ISL_429000 | UCSF Clinical Microbiology Laboratory | Chan-Zuckerberg Biohub |
| EPI_ISL_429001 | UCSF Clinical Microbiology Laboratory | Chan-Zuckerberg Biohub |
| EPI_ISL_429002 | UCSF Clinical Microbiology Laboratory | Chan-Zuckerberg Biohub |

| EPI_Accession_ID | Originating Laboratory | Submitting Laboratory |
|---|---|---|
| EPI_ISL_429003 | UCSF Clinical Microbiology Laboratory | Chan-Zuckerberg Biohub |
| EPI_ISL_429004 | UCSF Clinical Microbiology Laboratory | Chan-Zuckerberg Biohub |
| EPI_ISL_429005 | UCSF Clinical Microbiology Laboratory | Chan-Zuckerberg Biohub |
| EPI_ISL_429006 | UCSF Clinical Microbiology Laboratory | Chan-Zuckerberg Biohub |
| EPI_ISL_429007 | UCSF Clinical Microbiology Laboratory | Chan-Zuckerberg Biohub |
| EPI_ISL_429008 | UCSF Clinical Microbiology Laboratory | Chan-Zuckerberg Biohub |
| EPI_ISL_429009 | UCSF Clinical Microbiology Laboratory | Chan-Zuckerberg Biohub |
| EPI_ISL_429010 | UCSF Clinical Microbiology Laboratory | Chan-Zuckerberg Biohub |
| EPI_ISL_429011 | UCSF Clinical Microbiology Laboratory | Chan-Zuckerberg Biohub |
| EPI_ISL_429012 | UCSF Clinical Microbiology Laboratory | Chan-Zuckerberg Biohub |
| EPI_ISL_429013 | UCSF Clinical Microbiology Laboratory | Chan-Zuckerberg Biohub |
| EPI_ISL_429014 | UCSF Clinical Microbiology Laboratory | Chan-Zuckerberg Biohub |
| EPI_ISL_429015 | UCSF Clinical Microbiology Laboratory | Chan-Zuckerberg Biohub |
| EPI_ISL_429016 | UCSF Clinical Microbiology Laboratory | Chan-Zuckerberg Biohub |
| EPI_ISL_429017 | UCSF Clinical Microbiology Laboratory | Chan-Zuckerberg Biohub |
| EPI_ISL_429018 | UCSF Clinical Microbiology Laboratory | Chan-Zuckerberg Biohub |
| EPI_ISL_429019 | UCSF Clinical Microbiology Laboratory | Chan-Zuckerberg Biohub |
| EPI_ISL_429020 | UCSF Clinical Microbiology Laboratory | Chan-Zuckerberg Biohub |
| EPI_ISL_429021 | UCSF Clinical Microbiology Laboratory | Chan-Zuckerberg Biohub |
| EPI_ISL_429022 | UCSF Clinical Microbiology Laboratory | Chan-Zuckerberg Biohub |
| EPI_ISL_429023 | UCSF Clinical Microbiology Laboratory | Chan-Zuckerberg Biohub |
| EPI_ISL_429024 | UCSF Clinical Microbiology Laboratory | Chan-Zuckerberg Biohub |
| EPI_ISL_429025 | UCSF Clinical Microbiology Laboratory | Chan-Zuckerberg Biohub |
| EPI_ISL_429026 | UCSF Clinical Microbiology Laboratory | Chan-Zuckerberg Biohub |
| EPI_ISL_429027 | UCSF Clinical Microbiology Laboratory | Chan-Zuckerberg Biohub |
| EPI_ISL_429028 | UCSF Clinical Microbiology Laboratory | Chan-Zuckerberg Biohub |
| EPI_ISL_429029 | UCSF Clinical Microbiology Laboratory | Chan-Zuckerberg Biohub |
| EPI_ISL_429030 | UCSF Clinical Microbiology Laboratory | Chan-Zuckerberg Biohub |
| EPI_ISL_429031 | UCSF Clinical Microbiology Laboratory | Chan-Zuckerberg Biohub |
| EPI_ISL_429032 | UCSF Clinical Microbiology Laboratory | Chan-Zuckerberg Biohub |
| EPI_ISL_429033 | UCSF Clinical Microbiology Laboratory | Chan-Zuckerberg Biohub |
| EPI_ISL_429034 | UCSF Clinical Microbiology Laboratory | Chan-Zuckerberg Biohub |
| EPI_ISL_429035 | UCSF Clinical Microbiology Laboratory | Chan-Zuckerberg Biohub |

| EPI_Accession_ID | Originating Laboratory | Submitting Laboratory |
|---|---|---|
| EPI_ISL_429036 | UCSF Clinical Microbiology Laboratory | Chan-Zuckerberg Biohub |
| EPI_ISL_429037 | UCSF Clinical Microbiology Laboratory | Chan-Zuckerberg Biohub |
| EPI_ISL_429038 | UCSF Clinical Microbiology Laboratory | Chan-Zuckerberg Biohub |
| EPI_ISL_429039 | UCSF Clinical Microbiology Laboratory | Chan-Zuckerberg Biohub |
| EPI_ISL_429040 | UCSF Clinical Microbiology Laboratory | Chan-Zuckerberg Biohub |
| EPI_ISL_429041 | UCSF Clinical Microbiology Laboratory | Chan-Zuckerberg Biohub |
| EPI_ISL_429042 | UCSF Clinical Microbiology Laboratory | Chan-Zuckerberg Biohub |
| EPI_ISL_429043 | UCSF Clinical Microbiology Laboratory | Chan-Zuckerberg Biohub |
| EPI_ISL_429044 | UCSF Clinical Microbiology Laboratory | Chan-Zuckerberg Biohub |
| EPI_ISL_429045 | UCSF Clinical Microbiology Laboratory | Chan-Zuckerberg Biohub |
| EPI_ISL_429046 | UCSF Clinical Microbiology Laboratory | Chan-Zuckerberg Biohub |
| EPI_ISL_429047 | UCSF Clinical Microbiology Laboratory | Chan-Zuckerberg Biohub |
| EPI_ISL_429048 | UCSF Clinical Microbiology Laboratory | Chan-Zuckerberg Biohub |
| EPI_ISL_429049 | UCSF Clinical Microbiology Laboratory | Chan-Zuckerberg Biohub |
| EPI_ISL_429050 | UCSF Clinical Microbiology Laboratory | Chan-Zuckerberg Biohub |
| EPI_ISL_429051 | UCSF Clinical Microbiology Laboratory | Chan-Zuckerberg Biohub |
| EPI_ISL_429052 | UCSF Clinical Microbiology Laboratory | Chan-Zuckerberg Biohub |
| EPI_ISL_429053 | UCSF Clinical Microbiology Laboratory | Chan-Zuckerberg Biohub |
| EPI_ISL_429054 | UCSF Clinical Microbiology Laboratory | Chan-Zuckerberg Biohub |
| EPI_ISL_429055 | UCSF Clinical Microbiology Laboratory | Chan-Zuckerberg Biohub |
| EPI_ISL_429056 | UCSF Clinical Microbiology Laboratory | Chan-Zuckerberg Biohub |
| EPI_ISL_429057 | UCSF Clinical Microbiology Laboratory | Chan-Zuckerberg Biohub |
| EPI_ISL_429058 | UCSF Clinical Microbiology Laboratory | Chan-Zuckerberg Biohub |
| EPI_ISL_429059 | UCSF Clinical Microbiology Laboratory | Chan-Zuckerberg Biohub |
| EPI_ISL_429060 | UCSF Clinical Microbiology Laboratory | Chan-Zuckerberg Biohub |
| EPI_ISL_429061 | UCSF Clinical Microbiology Laboratory | Chan-Zuckerberg Biohub |
| EPI_ISL_429062 | UCSF Clinical Microbiology Laboratory | Chan-Zuckerberg Biohub |
| EPI_ISL_429063 | UCSF Clinical Microbiology Laboratory | Chan-Zuckerberg Biohub |
| EPI_ISL_429064 | UCSF Clinical Microbiology Laboratory | Chan-Zuckerberg Biohub |
| EPI_ISL_429065 | UCSF Clinical Microbiology Laboratory | Chan-Zuckerberg Biohub |
| EPI_ISL_429066 | UCSF Clinical Microbiology Laboratory | Chan-Zuckerberg Biohub |
| EPI_ISL_429067 | UCSF Clinical Microbiology Laboratory | Chan-Zuckerberg Biohub |
| EPI_ISL_429068 | UCSF Clinical Microbiology Laboratory | Chan-Zuckerberg Biohub |

| EPI_Accession_ID | Originating Laboratory | Submitting Laboratory |
|---|---|---|
| EPI_ISL_429069 | UCSF Clinical Microbiology Laboratory | Chan-Zuckerberg Biohub |
| EPI_ISL_429070 | UCSF Clinical Microbiology Laboratory | Chan-Zuckerberg Biohub |
| EPI_ISL_429071 | UCSF Clinical Microbiology Laboratory | Chan-Zuckerberg Biohub |
| EPI_ISL_429072 | UCSF Clinical Microbiology Laboratory | Chan-Zuckerberg Biohub |
| EPI_ISL_429073 | UCSF Clinical Microbiology Laboratory | Chan-Zuckerberg Biohub |
| EPI_ISL_429074 | The First Affiliated Hospital of Guangzhou Medical University | BGI-shenzhen & The First Affiliated Hospital of Guangzhou Medical University |
| EPI_ISL_429075 | The First Affiliated Hospital of Guangzhou Medical University | BGI-shenzhen & The First Affiliated Hospital of Guangzhou Medical University |
| EPI_ISL_429076 | The First Affiliated Hospital of Guangzhou Medical University | BGI-shenzhen & The First Affiliated Hospital of Guangzhou Medical University |
| EPI_ISL_429077 | The First Affiliated Hospital of Guangzhou Medical University | BGI-shenzhen & The First Affiliated Hospital of Guangzhou Medical University |
| EPI_ISL_429078 | The First Affiliated Hospital of Guangzhou Medical University | BGI-shenzhen & The First Affiliated Hospital of Guangzhou Medical University |
| EPI_ISL_429079 | The First Affiliated Hospital of Guangzhou Medical University | BGI-shenzhen & The First Affiliated Hospital of Guangzhou Medical University |
| EPI_ISL_429080 | The First Affiliated Hospital of Guangzhou Medical University | BGI-shenzhen & The First Affiliated Hospital of Guangzhou Medical University |
| EPI_ISL_429081 | The First Affiliated Hospital of Guangzhou Medical University | BGI-shenzhen & The First Affiliated Hospital of Guangzhou Medical University |
| EPI_ISL_429082 | The First Affiliated Hospital of Guangzhou Medical University | BGI-shenzhen & The First Affiliated Hospital of Guangzhou Medical University |
| EPI_ISL_429083 | The First Affiliated Hospital of Guangzhou Medical University | BGI-shenzhen & The First Affiliated Hospital of Guangzhou Medical University |
| EPI_ISL_429084 | The First Affiliated Hospital of Guangzhou Medical University | BGI-shenzhen & The First Affiliated Hospital of Guangzhou Medical University |
| EPI_ISL_429085 | The First Affiliated Hospital of Guangzhou Medical University | BGI-shenzhen & The First Affiliated Hospital of Guangzhou Medical University |
| EPI_ISL_429086 | The First Affiliated Hospital of Guangzhou Medical University | BGI-shenzhen & The First Affiliated Hospital of Guangzhou Medical University |
| EPI_ISL_429088 | The First Affiliated Hospital of Guangzhou Medical University | BGI-shenzhen & The First Affiliated Hospital of Guangzhou Medical University |

| EPI_Accession_ID | Originating Laboratory | Submitting Laboratory |
|---|---|---|
| EPI_ISL_429089 | The First Affiliated Hospital of Guangzhou Medical University | BGI-shenzhen & The First Affiliated Hospital of Guangzhou Medical University |
| EPI_ISL_429090 | The First Affiliated Hospital of Guangzhou Medical University | BGI-shenzhen & The First Affiliated Hospital of Guangzhou Medical University |
| EPI_ISL_429091 | The First Affiliated Hospital of Guangzhou Medical University | BGI-shenzhen & The First Affiliated Hospital of Guangzhou Medical University |
| EPI_ISL_429092 | The First Affiliated Hospital of Guangzhou Medical University | BGI-shenzhen & The First Affiliated Hospital of Guangzhou Medical University |
| EPI_ISL_429093 | The First Affiliated Hospital of Guangzhou Medical University | BGI-shenzhen & The First Affiliated Hospital of Guangzhou Medical University |
| EPI_ISL_429094 | The First Affiliated Hospital of Guangzhou Medical University | BGI-shenzhen & The First Affiliated Hospital of Guangzhou Medical University |
| EPI_ISL_429095 | The First Affiliated Hospital of Guangzhou Medical University | BGI-shenzhen & The First Affiliated Hospital of Guangzhou Medical University |
| EPI_ISL_429096 | The First Affiliated Hospital of Guangzhou Medical University | BGI-shenzhen & The First Affiliated Hospital of Guangzhou Medical University |
| EPI_ISL_429097 | The First Affiliated Hospital of Guangzhou Medical University | BGI-shenzhen & The First Affiliated Hospital of Guangzhou Medical University |
| EPI_ISL_429098 | The First Affiliated Hospital of Guangzhou Medical University | BGI-shenzhen & The First Affiliated Hospital of Guangzhou Medical University |
| EPI_ISL_429099 | The First Affiliated Hospital of Guangzhou Medical University | BGI-shenzhen & The First Affiliated Hospital of Guangzhou Medical University |
| EPI_ISL_429100 | The First Affiliated Hospital of Guangzhou Medical University | BGI-shenzhen & The First Affiliated Hospital of Guangzhou Medical University |
| EPI_ISL_429101 | The First Affiliated Hospital of Guangzhou Medical University | BGI-shenzhen & The First Affiliated Hospital of Guangzhou Medical University |
| EPI_ISL_429102 | The First Affiliated Hospital of Guangzhou Medical University | BGI-shenzhen & The First Affiliated Hospital of Guangzhou Medical University |
| EPI_ISL_429103 | The First Affiliated Hospital of Guangzhou Medical University | BGI-shenzhen & The First Affiliated Hospital of Guangzhou Medical University |
| EPI_ISL_429104 | The First Affiliated Hospital of Guangzhou Medical University | BGI-shenzhen & The First Affiliated Hospital of Guangzhou Medical University |

| EPI_Accession_ID | Originating Laboratory | Submitting Laboratory |
|---|---|---|
| EPI_ISL_429105 | The First Affiliated Hospital of Guangzhou Medical University | BGI-shenzhen & The First Affiliated Hospital of Guangzhou Medical University |
| EPI_ISL_429115 | Klinisk mikrobiologi och vardhygien Halmstad | The Public Health Agency of Sweden |
| EPI_ISL_429116 | Klinisk mikrobiologi och vardhygien Halmstad | The Public Health Agency of Sweden |
| EPI_ISL_429117 | Klinisk mikrobiologi och vardhygien Halmstad | The Public Health Agency of Sweden |
| EPI_ISL_429118 | Klinisk mikrobiologi och vardhygien Halmstad | The Public Health Agency of Sweden |
| EPI_ISL_429119 | Klinisk mikrobiologi och vardhygien Halmstad | The Public Health Agency of Sweden |
| EPI_ISL_429120 | Klinisk mikrobiologi och vardhygien Halmstad | The Public Health Agency of Sweden |
| EPI_ISL_429121 | Klinisk mikrobiologi och vardhygien Halmstad | The Public Health Agency of Sweden |
| EPI_ISL_429122 | Klinisk mikrobiologi och vardhygien Halmstad | The Public Health Agency of Sweden |
| EPI_ISL_429123 | Klinisk mikrobiologi och vardhygien Halmstad | The Public Health Agency of Sweden |
| EPI_ISL_429124 | Klinisk mikrobiologi och vardhygien Halmstad | The Public Health Agency of Sweden |
| EPI_ISL_429125 | Klinisk mikrobiologi och vardhygien Halmstad | The Public Health Agency of Sweden |
| EPI_ISL_429126 | Unilabs Skovde | The Public Health Agency of Sweden |
| EPI_ISL_429127 | Unilabs Skovde | The Public Health Agency of Sweden |
| EPI_ISL_429129 | Laboratoriemedicin | The Public Health Agency of Sweden |
| EPI_ISL_429130 | Laboratoriemedicin | The Public Health Agency of Sweden |
| EPI_ISL_429131 | Laboratoriemedicin | The Public Health Agency of Sweden |
| EPI_ISL_429132 | Laboratoriemedicin | The Public Health Agency of Sweden |
| EPI_ISL_429133 | Laboratoriemedicin | The Public Health Agency of Sweden |
| EPI_ISL_429134 | Laboratoriemedicin | The Public Health Agency of Sweden |
| EPI_ISL_429135 | Laboratoriemedicin | The Public Health Agency of Sweden |
| EPI_ISL_429136 | Klinisk mikrobiologi Orebro | The Public Health Agency of Sweden |
| EPI_ISL_429137 | Klinisk mikrobiologi Orebro | The Public Health Agency of Sweden |
| EPI_ISL_429138 | Klinisk mikrobiologi Orebro | The Public Health Agency of Sweden |
| EPI_ISL_429139 | Klinisk mikrobiologi Orebro | The Public Health Agency of Sweden |
| EPI_ISL_429140 | Klinisk mikrobiologi Orebro | The Public Health Agency of Sweden |
| EPI_ISL_429141 | Klinisk mikrobiologi Orebro | The Public Health Agency of Sweden |
| EPI_ISL_429142 | Klinisk mikrobiologi Orebro | The Public Health Agency of Sweden |
| EPI_ISL_429143 | Klinisk mikrobiologi Orebro | The Public Health Agency of Sweden |
| EPI_ISL_429144 | Klinisk mikrobiologi Orebro | The Public Health Agency of Sweden |
| EPI_ISL_429145 | Klinisk mikrobiologi Orebro | The Public Health Agency of Sweden |
| EPI_ISL_429146 | Klinisk mikrobiologi Orebro | The Public Health Agency of Sweden |

| EPI_Accession_ID | Originating Laboratory | Submitting Laboratory |
|---|---|---|
| EPI_ISL_429147 | Klinisk mikrobiologi Orebro | The Public Health Agency of Sweden |
| EPI_ISL_429148 | Klinisk mikrobiologi Orebro | The Public Health Agency of Sweden |
| EPI_ISL_429150 | Klinisk mikrobiologi Orebro | The Public Health Agency of Sweden |
| EPI_ISL_429151 | Klinisk mikrobiologi Orebro | The Public Health Agency of Sweden |
| EPI_ISL_429152 | Klinisk mikrobiologi Orebro | The Public Health Agency of Sweden |
| EPI_ISL_429153 | Klinisk mikrobiologi Orebro | The Public Health Agency of Sweden |
| EPI_ISL_429154 | Klinisk mikrobiologi Orebro | The Public Health Agency of Sweden |
| EPI_ISL_429155 | Klinisk mikrobiologi Orebro | The Public Health Agency of Sweden |
| EPI_ISL_429156 | Klinisk mikrobiologi Orebro | The Public Health Agency of Sweden |
| EPI_ISL_429157 | Klinisk mikrobiologi Orebro | The Public Health Agency of Sweden |
| EPI_ISL_429158 | Klinisk mikrobiologi Orebro | The Public Health Agency of Sweden |
| EPI_ISL_429160 | Klinisk mikrobiologi Orebro | The Public Health Agency of Sweden |
| EPI_ISL_429161 | The Public Health Agency of Sweden | The Public Health Agency of Sweden |
| EPI_ISL_429162 | The Public Health Agency of Sweden | The Public Health Agency of Sweden |
| EPI_ISL_429163 | The Public Health Agency of Sweden | The Public Health Agency of Sweden |
| EPI_ISL_429164 | Ramathibodi Hospital | COVID-19 Network Investigations (CONI) Alliance |
| EPI_ISL_429165 | Ramathibodi Hospital | COVID-19 Network Investigations (CONI) Alliance |
| EPI_ISL_429166 | Ramathibodi Hospital | COVID-19 Network Investigations (CONI) Alliance |
| EPI_ISL_429167 | Ramathibodi Hospital | COVID-19 Network Investigations (CONI) Alliance |
| EPI_ISL_429168 | Ramathibodi Hospital | COVID-19 Network Investigations (CONI) Alliance |
| EPI_ISL_429169 | Ramathibodi Hospital | COVID-19 Network Investigations (CONI) Alliance |
| EPI_ISL_429170 | Ramathibodi Hospital | COVID-19 Network Investigations (CONI) Alliance |
| EPI_ISL_429171 | Ramathibodi Hospital | COVID-19 Network Investigations (CONI) Alliance |
| EPI_ISL_429172 | Ramathibodi Hospital | COVID-19 Network Investigations (CONI) Alliance |
| EPI_ISL_429173 | Ramathibodi Hospital | COVID-19 Network Investigations (CONI) Alliance |
| EPI_ISL_429174 | Ramathibodi Hospital | COVID-19 Network Investigations (CONI) Alliance |
| EPI_ISL_429175 | Ramathibodi Hospital | COVID-19 Network Investigations (CONI) Alliance |
| EPI_ISL_429176 | Ramathibodi Hospital | COVID-19 Network Investigations (CONI) Alliance |
| EPI_ISL_429177 | Ramathibodi Hospital | COVID-19 Network Investigations (CONI) Alliance |
| EPI_ISL_429178 | Ramathibodi Hospital | COVID-19 Network Investigations (CONI) Alliance |
| EPI_ISL_429179 | Ramathibodi Hospital | COVID-19 Network Investigations (CONI) Alliance |
| EPI_ISL_429180 | Ramathibodi Hospital | COVID-19 Network Investigations (CONI) Alliance |
| EPI_ISL_429181 | Ramathibodi Hospital | COVID-19 Network Investigations (CONI) Alliance |

| EPI_Accession_ID | Originating Laboratory | Submitting Laboratory |
| --- | --- | --- |
| EPI_ISL_429182 | Ramathibodi Hospital | COVID-19 Network Investigations (CONI) Alliance |
| EPI_ISL_429183 | Ramathibodi Hospital | COVID-19 Network Investigations (CONI) Alliance |
| EPI_ISL_429184 | Ramathibodi Hospital | COVID-19 Network Investigations (CONI) Alliance |
| EPI_ISL_429196 | University Hospitals of Geneva Laboratory of Virology | University Hospitals of Geneva Laboratory of Virology |
| EPI_ISL_429197 | University Hospitals of Geneva Laboratory of Virology | University Hospitals of Geneva Laboratory of Virology |
| EPI_ISL_429199 | University Hospitals of Geneva Laboratory of Virology | University Hospitals of Geneva Laboratory of Virology |
| EPI_ISL_429200 | University Hospitals of Geneva Laboratory of Virology | University Hospitals of Geneva Laboratory of Virology |
| EPI_ISL_429201 | University Hospitals of Geneva Laboratory of Virology | University Hospitals of Geneva Laboratory of Virology |
| EPI_ISL_429203 | University Hospitals of Geneva Laboratory of Virology | University Hospitals of Geneva Laboratory of Virology |
| EPI_ISL_429204 | University Hospitals of Geneva Laboratory of Virology | University Hospitals of Geneva Laboratory of Virology |
| EPI_ISL_429206 | University Hospitals of Geneva Laboratory of Virology | University Hospitals of Geneva Laboratory of Virology |
| EPI_ISL_429207 | University Hospitals of Geneva Laboratory of Virology | University Hospitals of Geneva Laboratory of Virology |
| EPI_ISL_429208 | University Hospitals of Geneva Laboratory of Virology | University Hospitals of Geneva Laboratory of Virology |
| EPI_ISL_429209 | University Hospitals of Geneva Laboratory of Virology | University Hospitals of Geneva Laboratory of Virology |
| EPI_ISL_429210 | University Hospitals of Geneva Laboratory of Virology | University Hospitals of Geneva Laboratory of Virology |
| EPI_ISL_429211 | University Hospitals of Geneva Laboratory of Virology | University Hospitals of Geneva Laboratory of Virology |
| EPI_ISL_429212 | University Hospitals of Geneva Laboratory of Virology | University Hospitals of Geneva Laboratory of Virology |
| EPI_ISL_429213 | University Hospitals of Geneva Laboratory of Virology | University Hospitals of Geneva Laboratory of Virology |
| EPI_ISL_429214 | University Hospitals of Geneva Laboratory of Virology | University Hospitals of Geneva Laboratory of Virology |
| EPI_ISL_429215 | University Hospitals of Geneva Laboratory of Virology | University Hospitals of Geneva Laboratory of Virology |
| EPI_ISL_429217 | University Hospitals of Geneva Laboratory of Virology | University Hospitals of Geneva Laboratory of Virology |
| EPI_ISL_429218 | University Hospitals of Geneva Laboratory of Virology | University Hospitals of Geneva Laboratory of Virology |
| EPI_ISL_429219 | University Hospitals of Geneva Laboratory of Virology | University Hospitals of Geneva Laboratory of Virology |
| EPI_ISL_429220 | University Hospitals of Geneva Laboratory of Virology | University Hospitals of Geneva Laboratory of Virology |
| EPI_ISL_429221 | University Hospitals of Geneva Laboratory of Virology | University Hospitals of Geneva Laboratory of Virology |
| EPI_ISL_429222 | University Hospitals of Geneva Laboratory of Virology | University Hospitals of Geneva Laboratory of Virology |
| EPI_ISL_429223 | University Hospitals of Geneva Laboratory of Virology | University Hospitals of Geneva Laboratory of Virology |
| EPI_ISL_429226 | Presidio Ospedaliero Santo Spirito | Istituto Zooprofilattico Sperimentale dell'Abruzzo e Molise "G. Caporale" |
| EPI_ISL_429227 | Presidio Ospedaliero Santo Spirito | Istituto Zooprofilattico Sperimentale dell'Abruzzo e Molise "G. Caporale" |
| EPI_ISL_429228 | Ospedale Civile Giuseppe Mazzini | Istituto Zooprofilattico Sperimentale dell'Abruzzo e Molise "G. Caporale" |

| EPI_Accession_ID | Originating Laboratory | Submitting Laboratory |
|---|---|---|
| EPI_ISL_429229 | Ospedale Regionale San Salvatore | Istituto Zooprofilattico Sperimentale dell'Abruzzo e Molise "G. Caporale" |
| EPI_ISL_429230 | Ospedale Civile Giuseppe Mazzini | Istituto Zooprofilattico Sperimentale dell'Abruzzo e Molise "G. Caporale" |
| EPI_ISL_429231 | Ospedale Civile Giuseppe Mazzini | Istituto Zooprofilattico Sperimentale dell'Abruzzo e Molise "G. Caporale" |
| EPI_ISL_429232 | Ospedale Civile Giuseppe Mazzini | Istituto Zooprofilattico Sperimentale dell'Abruzzo e Molise "G. Caporale" |
| EPI_ISL_429233 | Ospedale Civile Giuseppe Mazzini | Istituto Zooprofilattico Sperimentale dell'Abruzzo e Molise "G. Caporale" |
| EPI_ISL_429234 | Ospedale Civile Giuseppe Mazzini | Istituto Zooprofilattico Sperimentale dell'Abruzzo e Molise "G. Caporale" |
| EPI_ISL_429235 | Ospedale Civile Giuseppe Mazzini | Istituto Zooprofilattico Sperimentale dell'Abruzzo e Molise "G. Caporale" |
| EPI_ISL_429236 | Ospedale Civile S. Liberatore di Atri | Istituto Zooprofilattico Sperimentale dell'Abruzzo e Molise "G. Caporale" |
| EPI_ISL_429239 | Department of Clinical Laboratory, the First People's Hospital of Yunnan Province | Department of Clinical Laboratory, the First People's Hospital of Yunnan Province |
| EPI_ISL_429254 | Viral Respiratory Lab, National Institute for Biomedical Research (INRB) | Pathogen Sequencing Lab, National Institute for Biomedical Research (INRB) |
| EPI_ISL_429255 | Viral Respiratory Lab, National Institute for Biomedical Research (INRB) | Pathogen Sequencing Lab, National Institute for Biomedical Research (INRB) |
| EPI_ISL_429256 | Health Sciences Technology Park, Avicena, 8, 18016 Granada. Spain | Sequencing and Bioinformatics Service FISABIO-Public Health |
| EPI_ISL_429257 | Microbial Genomics Laboratory, Institut Pasteur Montevideo | Microbial Genomics Laboratory, Institut Pasteur Montevideo |
| EPI_ISL_429258 | Viral Respiratory Lab, National Institute for Biomedical Research (INRB) | Pathogen Sequencing Lab, National Institute for Biomedical Research (INRB) |
| EPI_ISL_429259 | Viral Respiratory Lab, National Institute for Biomedical Research (INRB) | Pathogen Sequencing Lab, National Institute for Biomedical Research (INRB) |
| EPI_ISL_429262 | Department of Clinical Microbiology, Copenhagen University Hospital, Hvidovre, Kettegaard Alle 30, 2650 Hvidovre. | Albertsen lab, Department of Chemistry and Bioscience, Aalborg University, Denmark |

| EPI_Accession_ID | Originating Laboratory | Submitting Laboratory |
|---|---|---|
| EPI_ISL_429263 | Department of Clinical Microbiology, Copenhagen University Hospital, Hvidovre, Kettegaard Alle 30, 2650 Hvidovre. | Albertsen lab, Department of Chemistry and Bioscience, Aalborg University, Denmark |
| EPI_ISL_429264 | Department of Clinical Microbiology, Copenhagen University Hospital, Hvidovre, Kettegaard Alle 30, 2650 Hvidovre. | Albertsen lab, Department of Chemistry and Bioscience, Aalborg University, Denmark |
| EPI_ISL_429265 | Department of Clinical Microbiology, Copenhagen University Hospital, Hvidovre, Kettegaard Alle 30, 2650 Hvidovre. | Albertsen lab, Department of Chemistry and Bioscience, Aalborg University, Denmark |
| EPI_ISL_429266 | Department of Clinical Microbiology, Copenhagen University Hospital, Hvidovre, Kettegaard Alle 30, 2650 Hvidovre. | Albertsen lab, Department of Chemistry and Bioscience, Aalborg University, Denmark |
| EPI_ISL_429267 | Department of Clinical Microbiology, Copenhagen University Hospital, Hvidovre, Kettegaard Alle 30, 2650 Hvidovre. | Albertsen lab, Department of Chemistry and Bioscience, Aalborg University, Denmark |
| EPI_ISL_429268 | Department of Clinical Microbiology, Copenhagen University Hospital, Hvidovre, Kettegaard Alle 30, 2650 Hvidovre. | Albertsen lab, Department of Chemistry and Bioscience, Aalborg University, Denmark |
| EPI_ISL_429269 | Department of Clinical Microbiology, Copenhagen University Hospital, Hvidovre, Kettegaard Alle 30, 2650 Hvidovre. | Albertsen lab, Department of Chemistry and Bioscience, Aalborg University, Denmark |
| EPI_ISL_429270 | Department of Clinical Microbiology, Copenhagen University Hospital, Hvidovre, Kettegaard Alle 30, 2650 Hvidovre. | Albertsen lab, Department of Chemistry and Bioscience, Aalborg University, Denmark |
| EPI_ISL_429271 | Department of Clinical Microbiology, Copenhagen University Hospital, Hvidovre, Kettegaard Alle 30, 2650 Hvidovre. | Albertsen lab, Department of Chemistry and Bioscience, Aalborg University, Denmark |
| EPI_ISL_429272 | Department of Clinical Microbiology, Copenhagen University Hospital, Hvidovre, Kettegaard Alle 30, 2650 Hvidovre. | Albertsen lab, Department of Chemistry and Bioscience, Aalborg University, Denmark |
| EPI_ISL_429273 | Department of Clinical Microbiology, Copenhagen University Hospital, Hvidovre, Kettegaard Alle 30, 2650 Hvidovre. | Albertsen lab, Department of Chemistry and Bioscience, Aalborg University, Denmark |

| EPI_Accession_ID | Originating Laboratory | Submitting Laboratory |
|---|---|---|
| EPI_ISL_429274 | Department of Clinical Microbiology, Copenhagen University Hospital, Hvidovre, Kettegaard Alle 30, 2650 Hvidovre. | Albertsen lab, Department of Chemistry and Bioscience, Aalborg University, Denmark |
| EPI_ISL_429275 | Department of Clinical Microbiology, Copenhagen University Hospital, Hvidovre, Kettegaard Alle 30, 2650 Hvidovre. | Albertsen lab, Department of Chemistry and Bioscience, Aalborg University, Denmark |
| EPI_ISL_429276 | Department of Clinical Microbiology, Copenhagen University Hospital, Hvidovre, Kettegaard Alle 30, 2650 Hvidovre. | Albertsen lab, Department of Chemistry and Bioscience, Aalborg University, Denmark |
| EPI_ISL_429277 | Department of Clinical Microbiology, Copenhagen University Hospital, Hvidovre, Kettegaard Alle 30, 2650 Hvidovre. | Albertsen lab, Department of Chemistry and Bioscience, Aalborg University, Denmark |
| EPI_ISL_429278 | Department of Clinical Microbiology, Copenhagen University Hospital, Hvidovre, Kettegaard Alle 30, 2650 Hvidovre. | Albertsen lab, Department of Chemistry and Bioscience, Aalborg University, Denmark |
| EPI_ISL_429279 | Department of Clinical Microbiology, Copenhagen University Hospital, Hvidovre, Kettegaard Alle 30, 2650 Hvidovre. | Albertsen lab, Department of Chemistry and Bioscience, Aalborg University, Denmark |
| EPI_ISL_429280 | Department of Clinical Microbiology, Copenhagen University Hospital, Hvidovre, Kettegaard Alle 30, 2650 Hvidovre. | Albertsen lab, Department of Chemistry and Bioscience, Aalborg University, Denmark |
| EPI_ISL_429281 | Department of Clinical Microbiology, Copenhagen University Hospital, Hvidovre, Kettegaard Alle 30, 2650 Hvidovre. | Albertsen lab, Department of Chemistry and Bioscience, Aalborg University, Denmark |
| EPI_ISL_429282 | Department of Clinical Microbiology, Copenhagen University Hospital, Hvidovre, Kettegaard Alle 30, 2650 Hvidovre. | Albertsen lab, Department of Chemistry and Bioscience, Aalborg University, Denmark |
| EPI_ISL_429283 | Department of Clinical Microbiology, Copenhagen University Hospital, Hvidovre, Kettegaard Alle 30, 2650 Hvidovre. | Albertsen lab, Department of Chemistry and Bioscience, Aalborg University, Denmark |
| EPI_ISL_429284 | Department of Clinical Microbiology, Copenhagen University Hospital, Hvidovre, Kettegaard Alle 30, 2650 Hvidovre. | Albertsen lab, Department of Chemistry and Bioscience, Aalborg University, Denmark |

| EPI_Accession_ID | Originating Laboratory | Submitting Laboratory |
|---|---|---|
| EPI_ISL_429285 | Department of Clinical Microbiology, Copenhagen University Hospital, Hvidovre, Kettegaard Alle 30, 2650 Hvidovre. | Albertsen lab, Department of Chemistry and Bioscience, Aalborg University, Denmark |
| EPI_ISL_429286 | Department of Clinical Microbiology, Copenhagen University Hospital, Hvidovre, Kettegaard Alle 30, 2650 Hvidovre. | Albertsen lab, Department of Chemistry and Bioscience, Aalborg University, Denmark |
| EPI_ISL_429287 | Department of Clinical Microbiology, Copenhagen University Hospital, Hvidovre, Kettegaard Alle 30, 2650 Hvidovre. | Albertsen lab, Department of Chemistry and Bioscience, Aalborg University, Denmark |
| EPI_ISL_429288 | Department of Clinical Microbiology, Copenhagen University Hospital, Hvidovre, Kettegaard Alle 30, 2650 Hvidovre. | Albertsen lab, Department of Chemistry and Bioscience, Aalborg University, Denmark |
| EPI_ISL_429289 | Department of Clinical Microbiology, Copenhagen University Hospital, Hvidovre, Kettegaard Alle 30, 2650 Hvidovre. | Albertsen lab, Department of Chemistry and Bioscience, Aalborg University, Denmark |
| EPI_ISL_429290 | Department of Clinical Microbiology, Copenhagen University Hospital, Hvidovre, Kettegaard Alle 30, 2650 Hvidovre. | Albertsen lab, Department of Chemistry and Bioscience, Aalborg University, Denmark |
| EPI_ISL_429291 | Department of Clinical Microbiology, Copenhagen University Hospital, Hvidovre, Kettegaard Alle 30, 2650 Hvidovre. | Albertsen lab, Department of Chemistry and Bioscience, Aalborg University, Denmark |
| EPI_ISL_429292 | Department of Clinical Microbiology, Copenhagen University Hospital, Hvidovre, Kettegaard Alle 30, 2650 Hvidovre. | Albertsen lab, Department of Chemistry and Bioscience, Aalborg University, Denmark |
| EPI_ISL_429293 | Department of Clinical Microbiology, Copenhagen University Hospital, Hvidovre, Kettegaard Alle 30, 2650 Hvidovre. | Albertsen lab, Department of Chemistry and Bioscience, Aalborg University, Denmark |
| EPI_ISL_429294 | Department of Clinical Microbiology, Copenhagen University Hospital, Hvidovre, Kettegaard Alle 30, 2650 Hvidovre. | Albertsen lab, Department of Chemistry and Bioscience, Aalborg University, Denmark |
| EPI_ISL_429295 | Department of Clinical Microbiology, Copenhagen University Hospital, Hvidovre, Kettegaard Alle 30, 2650 Hvidovre. | Albertsen lab, Department of Chemistry and Bioscience, Aalborg University, Denmark |

| EPI_Accession_ID | Originating Laboratory | Submitting Laboratory |
|---|---|---|
| EPI_ISL_429296 | Department of Clinical Microbiology, Copenhagen University Hospital, Hvidovre, Kettegaard Alle 30, 2650 Hvidovre. | Albertsen lab, Department of Chemistry and Bioscience, Aalborg University, Denmark |
| EPI_ISL_429297 | Department of Clinical Microbiology, Copenhagen University Hospital, Hvidovre, Kettegaard Alle 30, 2650 Hvidovre. | Albertsen lab, Department of Chemistry and Bioscience, Aalborg University, Denmark |
| EPI_ISL_429298 | Department of Clinical Microbiology, Copenhagen University Hospital, Hvidovre, Kettegaard Alle 30, 2650 Hvidovre. | Albertsen lab, Department of Chemistry and Bioscience, Aalborg University, Denmark |
| EPI_ISL_429299 | Department of Clinical Microbiology, Copenhagen University Hospital, Hvidovre, Kettegaard Alle 30, 2650 Hvidovre. | Albertsen lab, Department of Chemistry and Bioscience, Aalborg University, Denmark |
| EPI_ISL_429300 | Department of Clinical Microbiology, Copenhagen University Hospital, Hvidovre, Kettegaard Alle 30, 2650 Hvidovre. | Albertsen lab, Department of Chemistry and Bioscience, Aalborg University, Denmark |
| EPI_ISL_429301 | Department of Clinical Microbiology, Copenhagen University Hospital, Hvidovre, Kettegaard Alle 30, 2650 Hvidovre. | Albertsen lab, Department of Chemistry and Bioscience, Aalborg University, Denmark |
| EPI_ISL_429302 | Department of Clinical Microbiology, Copenhagen University Hospital, Hvidovre, Kettegaard Alle 30, 2650 Hvidovre. | Albertsen lab, Department of Chemistry and Bioscience, Aalborg University, Denmark |
| EPI_ISL_429303 | Department of Clinical Microbiology, Copenhagen University Hospital, Hvidovre, Kettegaard Alle 30, 2650 Hvidovre. | Albertsen lab, Department of Chemistry and Bioscience, Aalborg University, Denmark |
| EPI_ISL_429304 | Department of Clinical Microbiology, Copenhagen University Hospital, Hvidovre, Kettegaard Alle 30, 2650 Hvidovre. | Albertsen lab, Department of Chemistry and Bioscience, Aalborg University, Denmark |
| EPI_ISL_429305 | Department of Clinical Microbiology, Copenhagen University Hospital, Hvidovre, Kettegaard Alle 30, 2650 Hvidovre. | Albertsen lab, Department of Chemistry and Bioscience, Aalborg University, Denmark |
| EPI_ISL_429306 | Department of Clinical Microbiology, Copenhagen University Hospital, Hvidovre, Kettegaard Alle 30, 2650 Hvidovre. | Albertsen lab, Department of Chemistry and Bioscience, Aalborg University, Denmark |

| EPI_Accession_ID | Originating Laboratory | Submitting Laboratory |
|---|---|---|
| EPI_ISL_429307 | Department of Clinical Microbiology, Copenhagen University Hospital, Hvidovre, Kettegaard Alle 30, 2650 Hvidovre. | Albertsen lab, Department of Chemistry and Bioscience, Aalborg University, Denmark |
| EPI_ISL_429308 | Department of Clinical Microbiology, Copenhagen University Hospital, Hvidovre, Kettegaard Alle 30, 2650 Hvidovre. | Albertsen lab, Department of Chemistry and Bioscience, Aalborg University, Denmark |
| EPI_ISL_429309 | Department of Clinical Microbiology, Copenhagen University Hospital, Hvidovre, Kettegaard Alle 30, 2650 Hvidovre. | Albertsen lab, Department of Chemistry and Bioscience, Aalborg University, Denmark |
| EPI_ISL_429310 | Department of Clinical Microbiology, Copenhagen University Hospital, Hvidovre, Kettegaard Alle 30, 2650 Hvidovre. | Albertsen lab, Department of Chemistry and Bioscience, Aalborg University, Denmark |
| EPI_ISL_429311 | Department of Clinical Microbiology, Copenhagen University Hospital, Hvidovre, Kettegaard Alle 30, 2650 Hvidovre. | Albertsen lab, Department of Chemistry and Bioscience, Aalborg University, Denmark |
| EPI_ISL_429312 | Department of Clinical Microbiology, Copenhagen University Hospital, Hvidovre, Kettegaard Alle 30, 2650 Hvidovre. | Albertsen lab, Department of Chemistry and Bioscience, Aalborg University, Denmark |
| EPI_ISL_429313 | Department of Clinical Microbiology, Copenhagen University Hospital, Hvidovre, Kettegaard Alle 30, 2650 Hvidovre. | Albertsen lab, Department of Chemistry and Bioscience, Aalborg University, Denmark |
| EPI_ISL_429314 | Department of Clinical Microbiology, Copenhagen University Hospital, Hvidovre, Kettegaard Alle 30, 2650 Hvidovre. | Albertsen lab, Department of Chemistry and Bioscience, Aalborg University, Denmark |
| EPI_ISL_429315 | Department of Clinical Microbiology, Copenhagen University Hospital, Hvidovre, Kettegaard Alle 30, 2650 Hvidovre. | Albertsen lab, Department of Chemistry and Bioscience, Aalborg University, Denmark |
| EPI_ISL_429316 | Department of Clinical Microbiology, Copenhagen University Hospital, Hvidovre, Kettegaard Alle 30, 2650 Hvidovre. | Albertsen lab, Department of Chemistry and Bioscience, Aalborg University, Denmark |
| EPI_ISL_429317 | Department of Clinical Microbiology, Copenhagen University Hospital, Hvidovre, Kettegaard Alle 30, 2650 Hvidovre. | Albertsen lab, Department of Chemistry and Bioscience, Aalborg University, Denmark |

| EPI_Accession_ID | Originating Laboratory | Submitting Laboratory |
| --- | --- | --- |
| EPI_ISL_429318 | Department of Clinical Microbiology, Copenhagen University Hospital, Hvidovre, Kettegaard Alle 30, 2650 Hvidovre. | Albertsen lab, Department of Chemistry and Bioscience, Aalborg University, Denmark |
| EPI_ISL_429320 | Department of Clinical Microbiology, Copenhagen University Hospital, Hvidovre, Kettegaard Alle 30, 2650 Hvidovre. | Albertsen lab, Department of Chemistry and Bioscience, Aalborg University, Denmark |
| EPI_ISL_429321 | Department of Clinical Microbiology, Copenhagen University Hospital, Hvidovre, Kettegaard Alle 30, 2650 Hvidovre. | Albertsen lab, Department of Chemistry and Bioscience, Aalborg University, Denmark |
| EPI_ISL_429322 | Department of Clinical Microbiology, Copenhagen University Hospital, Hvidovre, Kettegaard Alle 30, 2650 Hvidovre. | Albertsen lab, Department of Chemistry and Bioscience, Aalborg University, Denmark |
| EPI_ISL_429323 | Department of Clinical Microbiology, Copenhagen University Hospital, Hvidovre, Kettegaard Alle 30, 2650 Hvidovre. | Albertsen lab, Department of Chemistry and Bioscience, Aalborg University, Denmark |
| EPI_ISL_429324 | Department of Clinical Microbiology, Copenhagen University Hospital, Hvidovre, Kettegaard Alle 30, 2650 Hvidovre. | Albertsen lab, Department of Chemistry and Bioscience, Aalborg University, Denmark |
| EPI_ISL_429325 | Department of Clinical Microbiology, Copenhagen University Hospital, Hvidovre, Kettegaard Alle 30, 2650 Hvidovre. | Albertsen lab, Department of Chemistry and Bioscience, Aalborg University, Denmark |
| EPI_ISL_429326 | Department of Clinical Microbiology, Copenhagen University Hospital, Hvidovre, Kettegaard Alle 30, 2650 Hvidovre. | Albertsen lab, Department of Chemistry and Bioscience, Aalborg University, Denmark |
| EPI_ISL_429327 | Department of Clinical Microbiology, Copenhagen University Hospital, Hvidovre, Kettegaard Alle 30, 2650 Hvidovre. | Albertsen lab, Department of Chemistry and Bioscience, Aalborg University, Denmark |
| EPI_ISL_429328 | Department of Clinical Microbiology, Copenhagen University Hospital, Hvidovre, Kettegaard Alle 30, 2650 Hvidovre. | Albertsen lab, Department of Chemistry and Bioscience, Aalborg University, Denmark |
| EPI_ISL_429329 | Department of Clinical Microbiology, Copenhagen University Hospital, Hvidovre, Kettegaard Alle 30, 2650 Hvidovre. | Albertsen lab, Department of Chemistry and Bioscience, Aalborg University, Denmark |

| EPI_Accession_ID | Originating Laboratory | Submitting Laboratory |
|---|---|---|
| EPI_ISL_429330 | Department of Clinical Microbiology, Copenhagen University Hospital, Hvidovre, Kettegaard Alle 30, 2650 Hvidovre. | Albertsen lab, Department of Chemistry and Bioscience, Aalborg University, Denmark |
| EPI_ISL_429331 | Department of Clinical Microbiology, Copenhagen University Hospital, Hvidovre, Kettegaard Alle 30, 2650 Hvidovre. | Albertsen lab, Department of Chemistry and Bioscience, Aalborg University, Denmark |
| EPI_ISL_429332 | Department of Clinical Microbiology, Copenhagen University Hospital, Hvidovre, Kettegaard Alle 30, 2650 Hvidovre. | Albertsen lab, Department of Chemistry and Bioscience, Aalborg University, Denmark |
| EPI_ISL_429333 | Department of Virus and Microbiological Special Diagnostics, Statens Serum Institut, Copenhagen, Denmark, Artillerivej 5, 2300 Copenahgen S | Albertsen lab, Department of Chemistry and Bioscience, Aalborg University, Denmark |
| EPI_ISL_429334 | Department of Virus and Microbiological Special Diagnostics, Statens Serum Institut, Copenhagen, Denmark, Artillerivej 5, 2300 Copenahgen S | Albertsen lab, Department of Chemistry and Bioscience, Aalborg University, Denmark |
| EPI_ISL_429335 | Department of Virus and Microbiological Special Diagnostics, Statens Serum Institut, Copenhagen, Denmark, Artillerivej 5, 2300 Copenahgen S | Albertsen lab, Department of Chemistry and Bioscience, Aalborg University, Denmark |
| EPI_ISL_429336 | Department of Virus and Microbiological Special Diagnostics, Statens Serum Institut, Copenhagen, Denmark, Artillerivej 5, 2300 Copenahgen S | Albertsen lab, Department of Chemistry and Bioscience, Aalborg University, Denmark |
| EPI_ISL_429337 | Department of Virus and Microbiological Special Diagnostics, Statens Serum Institut, Copenhagen, Denmark, Artillerivej 5, 2300 Copenahgen S | Albertsen lab, Department of Chemistry and Bioscience, Aalborg University, Denmark |
| EPI_ISL_429338 | Department of Virus and Microbiological Special Diagnostics, Statens Serum Institut, Copenhagen, Denmark, Artillerivej 5, 2300 Copenahgen S | Albertsen lab, Department of Chemistry and Bioscience, Aalborg University, Denmark |
| EPI_ISL_429339 | Department of Virus and Microbiological Special Diagnostics, Statens Serum Institut, Copenhagen, Denmark, Artillerivej 5, 2300 Copenahgen S | Albertsen lab, Department of Chemistry and Bioscience, Aalborg University, Denmark |
| EPI_ISL_429340 | Department of Virus and Microbiological Special Diagnostics, Statens Serum Institut, Copenhagen, Denmark, Artillerivej 5, 2300 Copenahgen S | Albertsen lab, Department of Chemistry and Bioscience, Aalborg University, Denmark |

| EPI_Accession_ID | Originating Laboratory | Submitting Laboratory |
|---|---|---|
| EPI_ISL_429341 | Department of Virus and Microbiological Special Diagnostics, Statens Serum Institut, Copenhagen, Denmark, Artillerivej 5, 2300 Copenahgen S | Albertsen lab, Department of Chemistry and Bioscience, Aalborg University, Denmark |
| EPI_ISL_429342 | Department of Virus and Microbiological Special Diagnostics, Statens Serum Institut, Copenhagen, Denmark, Artillerivej 5, 2300 Copenahgen S | Albertsen lab, Department of Chemistry and Bioscience, Aalborg University, Denmark |
| EPI_ISL_429343 | Department of Virus and Microbiological Special Diagnostics, Statens Serum Institut, Copenhagen, Denmark, Artillerivej 5, 2300 Copenahgen S | Albertsen lab, Department of Chemistry and Bioscience, Aalborg University, Denmark |
| EPI_ISL_429344 | Department of Virus and Microbiological Special Diagnostics, Statens Serum Institut, Copenhagen, Denmark, Artillerivej 5, 2300 Copenahgen S | Albertsen lab, Department of Chemistry and Bioscience, Aalborg University, Denmark |
| EPI_ISL_429345 | Department of Virus and Microbiological Special Diagnostics, Statens Serum Institut, Copenhagen, Denmark, Artillerivej 5, 2300 Copenahgen S | Albertsen lab, Department of Chemistry and Bioscience, Aalborg University, Denmark |
| EPI_ISL_429346 | Department of Virus and Microbiological Special Diagnostics, Statens Serum Institut, Copenhagen, Denmark, Artillerivej 5, 2300 Copenahgen S | Albertsen lab, Department of Chemistry and Bioscience, Aalborg University, Denmark |
| EPI_ISL_429347 | Department of Virus and Microbiological Special Diagnostics, Statens Serum Institut, Copenhagen, Denmark, Artillerivej 5, 2300 Copenahgen S | Albertsen lab, Department of Chemistry and Bioscience, Aalborg University, Denmark |
| EPI_ISL_429348 | Department of Virus and Microbiological Special Diagnostics, Statens Serum Institut, Copenhagen, Denmark, Artillerivej 5, 2300 Copenahgen S | Albertsen lab, Department of Chemistry and Bioscience, Aalborg University, Denmark |
| EPI_ISL_429349 | Department of Virus and Microbiological Special Diagnostics, Statens Serum Institut, Copenhagen, Denmark, Artillerivej 5, 2300 Copenahgen S | Albertsen lab, Department of Chemistry and Bioscience, Aalborg University, Denmark |
| EPI_ISL_429350 | Department of Virus and Microbiological Special Diagnostics, Statens Serum Institut, Copenhagen, Denmark, Artillerivej 5, 2300 Copenahgen S | Albertsen lab, Department of Chemistry and Bioscience, Aalborg University, Denmark |
| EPI_ISL_429351 | Department of Virus and Microbiological Special Diagnostics, Statens Serum Institut, Copenhagen, Denmark, Artillerivej 5, 2300 Copenahgen S | Albertsen lab, Department of Chemistry and Bioscience, Aalborg University, Denmark |

| EPI_Accession_ID | Originating Laboratory | Submitting Laboratory |
|---|---|---|
| EPI_ISL_429352 | Department of Virus and Microbiological Special Diagnostics, Statens Serum Institut, Copenhagen, Denmark, Artillerivej 5, 2300 Copenahgen S | Albertsen lab, Department of Chemistry and Bioscience, Aalborg University, Denmark |
| EPI_ISL_429353 | Department of Virus and Microbiological Special Diagnostics, Statens Serum Institut, Copenhagen, Denmark, Artillerivej 5, 2300 Copenahgen S | Albertsen lab, Department of Chemistry and Bioscience, Aalborg University, Denmark |
| EPI_ISL_429354 | Department of Virus and Microbiological Special Diagnostics, Statens Serum Institut, Copenhagen, Denmark, Artillerivej 5, 2300 Copenahgen S | Albertsen lab, Department of Chemistry and Bioscience, Aalborg University, Denmark |
| EPI_ISL_429355 | Department of Virus and Microbiological Special Diagnostics, Statens Serum Institut, Copenhagen, Denmark, Artillerivej 5, 2300 Copenahgen S | Albertsen lab, Department of Chemistry and Bioscience, Aalborg University, Denmark |
| EPI_ISL_429356 | Department of Virus and Microbiological Special Diagnostics, Statens Serum Institut, Copenhagen, Denmark, Artillerivej 5, 2300 Copenahgen S | Albertsen lab, Department of Chemistry and Bioscience, Aalborg University, Denmark |
| EPI_ISL_429357 | Department of Virus and Microbiological Special Diagnostics, Statens Serum Institut, Copenhagen, Denmark, Artillerivej 5, 2300 Copenahgen S | Albertsen lab, Department of Chemistry and Bioscience, Aalborg University, Denmark |
| EPI_ISL_429358 | Department of Virus and Microbiological Special Diagnostics, Statens Serum Institut, Copenhagen, Denmark, Artillerivej 5, 2300 Copenahgen S | Albertsen lab, Department of Chemistry and Bioscience, Aalborg University, Denmark |
| EPI_ISL_429359 | Department of Virus and Microbiological Special Diagnostics, Statens Serum Institut, Copenhagen, Denmark, Artillerivej 5, 2300 Copenahgen S | Albertsen lab, Department of Chemistry and Bioscience, Aalborg University, Denmark |
| EPI_ISL_429360 | Department of Virus and Microbiological Special Diagnostics, Statens Serum Institut, Copenhagen, Denmark, Artillerivej 5, 2300 Copenahgen S | Albertsen lab, Department of Chemistry and Bioscience, Aalborg University, Denmark |
| EPI_ISL_429361 | Department of Virus and Microbiological Special Diagnostics, Statens Serum Institut, Copenhagen, Denmark, Artillerivej 5, 2300 Copenahgen S | Albertsen lab, Department of Chemistry and Bioscience, Aalborg University, Denmark |
| EPI_ISL_429362 | Department of Virus and Microbiological Special Diagnostics, Statens Serum Institut, Copenhagen, Denmark, Artillerivej 5, 2300 Copenahgen S | Albertsen lab, Department of Chemistry and Bioscience, Aalborg University, Denmark |

| EPI_Accession_ID | Originating Laboratory | Submitting Laboratory |
|---|---|---|
| EPI_ISL_429363 | Department of Virus and Microbiological Special Diagnostics, Statens Serum Institut, Copenhagen, Denmark, Artillerivej 5, 2300 Copenahgen S | Albertsen lab, Department of Chemistry and Bioscience, Aalborg University, Denmark |
| EPI_ISL_429364 | Department of Virus and Microbiological Special Diagnostics, Statens Serum Institut, Copenhagen, Denmark, Artillerivej 5, 2300 Copenahgen S | Albertsen lab, Department of Chemistry and Bioscience, Aalborg University, Denmark |
| EPI_ISL_429365 | Department of Virus and Microbiological Special Diagnostics, Statens Serum Institut, Copenhagen, Denmark, Artillerivej 5, 2300 Copenahgen S | Albertsen lab, Department of Chemistry and Bioscience, Aalborg University, Denmark |
| EPI_ISL_429366 | Department of Virus and Microbiological Special Diagnostics, Statens Serum Institut, Copenhagen, Denmark, Artillerivej 5, 2300 Copenahgen S | Albertsen lab, Department of Chemistry and Bioscience, Aalborg University, Denmark |
| EPI_ISL_429367 | Department of Virus and Microbiological Special Diagnostics, Statens Serum Institut, Copenhagen, Denmark, Artillerivej 5, 2300 Copenahgen S | Albertsen lab, Department of Chemistry and Bioscience, Aalborg University, Denmark |
| EPI_ISL_429368 | Department of Virus and Microbiological Special Diagnostics, Statens Serum Institut, Copenhagen, Denmark, Artillerivej 5, 2300 Copenahgen S | Albertsen lab, Department of Chemistry and Bioscience, Aalborg University, Denmark |
| EPI_ISL_429369 | Department of Virus and Microbiological Special Diagnostics, Statens Serum Institut, Copenhagen, Denmark, Artillerivej 5, 2300 Copenahgen S | Albertsen lab, Department of Chemistry and Bioscience, Aalborg University, Denmark |
| EPI_ISL_429370 | Department of Virus and Microbiological Special Diagnostics, Statens Serum Institut, Copenhagen, Denmark, Artillerivej 5, 2300 Copenahgen S | Albertsen lab, Department of Chemistry and Bioscience, Aalborg University, Denmark |
| EPI_ISL_429371 | Department of Virus and Microbiological Special Diagnostics, Statens Serum Institut, Copenhagen, Denmark, Artillerivej 5, 2300 Copenahgen S | Albertsen lab, Department of Chemistry and Bioscience, Aalborg University, Denmark |
| EPI_ISL_429372 | Department of Virus and Microbiological Special Diagnostics, Statens Serum Institut, Copenhagen, Denmark, Artillerivej 5, 2300 Copenahgen S | Albertsen lab, Department of Chemistry and Bioscience, Aalborg University, Denmark |
| EPI_ISL_429373 | Department of Virus and Microbiological Special Diagnostics, Statens Serum Institut, Copenhagen, Denmark, Artillerivej 5, 2300 Copenahgen S | Albertsen lab, Department of Chemistry and Bioscience, Aalborg University, Denmark |

| EPI_Accession_ID | Originating Laboratory | Submitting Laboratory |
|---|---|---|
| EPI_ISL_429374 | Department of Virus and Microbiological Special Diagnostics, Statens Serum Institut, Copenhagen, Denmark, Artillerivej 5, 2300 Copenahgen S | Albertsen lab, Department of Chemistry and Bioscience, Aalborg University, Denmark |
| EPI_ISL_429375 | Department of Virus and Microbiological Special Diagnostics, Statens Serum Institut, Copenhagen, Denmark, Artillerivej 5, 2300 Copenahgen S | Albertsen lab, Department of Chemistry and Bioscience, Aalborg University, Denmark |
| EPI_ISL_429376 | Department of Virus and Microbiological Special Diagnostics, Statens Serum Institut, Copenhagen, Denmark, Artillerivej 5, 2300 Copenahgen S | Albertsen lab, Department of Chemistry and Bioscience, Aalborg University, Denmark |
| EPI_ISL_429377 | Department of Virus and Microbiological Special Diagnostics, Statens Serum Institut, Copenhagen, Denmark, Artillerivej 5, 2300 Copenahgen S | Albertsen lab, Department of Chemistry and Bioscience, Aalborg University, Denmark |
| EPI_ISL_429378 | Department of Virus and Microbiological Special Diagnostics, Statens Serum Institut, Copenhagen, Denmark, Artillerivej 5, 2300 Copenahgen S | Albertsen lab, Department of Chemistry and Bioscience, Aalborg University, Denmark |
| EPI_ISL_429379 | Department of Virus and Microbiological Special Diagnostics, Statens Serum Institut, Copenhagen, Denmark, Artillerivej 5, 2300 Copenahgen S | Albertsen lab, Department of Chemistry and Bioscience, Aalborg University, Denmark |
| EPI_ISL_429380 | Department of Virus and Microbiological Special Diagnostics, Statens Serum Institut, Copenhagen, Denmark, Artillerivej 5, 2300 Copenahgen S | Albertsen lab, Department of Chemistry and Bioscience, Aalborg University, Denmark |
| EPI_ISL_429381 | Department of Virus and Microbiological Special Diagnostics, Statens Serum Institut, Copenhagen, Denmark, Artillerivej 5, 2300 Copenahgen S | Albertsen lab, Department of Chemistry and Bioscience, Aalborg University, Denmark |
| EPI_ISL_429382 | Department of Virus and Microbiological Special Diagnostics, Statens Serum Institut, Copenhagen, Denmark, Artillerivej 5, 2300 Copenahgen S | Albertsen lab, Department of Chemistry and Bioscience, Aalborg University, Denmark |
| EPI_ISL_429383 | Department of Virus and Microbiological Special Diagnostics, Statens Serum Institut, Copenhagen, Denmark, Artillerivej 5, 2300 Copenahgen S | Albertsen lab, Department of Chemistry and Bioscience, Aalborg University, Denmark |
| EPI_ISL_429384 | Department of Virus and Microbiological Special Diagnostics, Statens Serum Institut, Copenhagen, Denmark, Artillerivej 5, 2300 Copenahgen S | Albertsen lab, Department of Chemistry and Bioscience, Aalborg University, Denmark |

| EPI_Accession_ID | Originating Laboratory | Submitting Laboratory |
|---|---|---|
| EPI_ISL_429385 | Department of Virus and Microbiological Special Diagnostics, Statens Serum Institut, Copenhagen, Denmark, Artillerivej 5, 2300 Copenahgen S | Albertsen lab, Department of Chemistry and Bioscience, Aalborg University, Denmark |
| EPI_ISL_429386 | Department of Virus and Microbiological Special Diagnostics, Statens Serum Institut, Copenhagen, Denmark, Artillerivej 5, 2300 Copenahgen S | Albertsen lab, Department of Chemistry and Bioscience, Aalborg University, Denmark |
| EPI_ISL_429387 | Department of Virus and Microbiological Special Diagnostics, Statens Serum Institut, Copenhagen, Denmark, Artillerivej 5, 2300 Copenahgen S | Albertsen lab, Department of Chemistry and Bioscience, Aalborg University, Denmark |
| EPI_ISL_429388 | Department of Virus and Microbiological Special Diagnostics, Statens Serum Institut, Copenhagen, Denmark, Artillerivej 5, 2300 Copenahgen S | Albertsen lab, Department of Chemistry and Bioscience, Aalborg University, Denmark |
| EPI_ISL_429389 | Department of Virus and Microbiological Special Diagnostics, Statens Serum Institut, Copenhagen, Denmark, Artillerivej 5, 2300 Copenahgen S | Albertsen lab, Department of Chemistry and Bioscience, Aalborg University, Denmark |
| EPI_ISL_429390 | Department of Virus and Microbiological Special Diagnostics, Statens Serum Institut, Copenhagen, Denmark, Artillerivej 5, 2300 Copenahgen S | Albertsen lab, Department of Chemistry and Bioscience, Aalborg University, Denmark |
| EPI_ISL_429391 | Department of Virus and Microbiological Special Diagnostics, Statens Serum Institut, Copenhagen, Denmark, Artillerivej 5, 2300 Copenahgen S | Albertsen lab, Department of Chemistry and Bioscience, Aalborg University, Denmark |
| EPI_ISL_429392 | Department of Virus and Microbiological Special Diagnostics, Statens Serum Institut, Copenhagen, Denmark, Artillerivej 5, 2300 Copenahgen S | Albertsen lab, Department of Chemistry and Bioscience, Aalborg University, Denmark |
| EPI_ISL_429393 | Department of Virus and Microbiological Special Diagnostics, Statens Serum Institut, Copenhagen, Denmark, Artillerivej 5, 2300 Copenahgen S | Albertsen lab, Department of Chemistry and Bioscience, Aalborg University, Denmark |
| EPI_ISL_429394 | Department of Virus and Microbiological Special Diagnostics, Statens Serum Institut, Copenhagen, Denmark, Artillerivej 5, 2300 Copenahgen S | Albertsen lab, Department of Chemistry and Bioscience, Aalborg University, Denmark |
| EPI_ISL_429395 | Department of Virus and Microbiological Special Diagnostics, Statens Serum Institut, Copenhagen, Denmark, Artillerivej 5, 2300 Copenahgen S | Albertsen lab, Department of Chemistry and Bioscience, Aalborg University, Denmark |

| EPI_Accession_ID | Originating Laboratory | Submitting Laboratory |
|---|---|---|
| EPI_ISL_429396 | Department of Virus and Microbiological Special Diagnostics, Statens Serum Institut, Copenhagen, Denmark, Artillerivej 5, 2300 Copenahgen S | Albertsen lab, Department of Chemistry and Bioscience, Aalborg University, Denmark |
| EPI_ISL_429397 | Department of Virus and Microbiological Special Diagnostics, Statens Serum Institut, Copenhagen, Denmark, Artillerivej 5, 2300 Copenahgen S | Albertsen lab, Department of Chemistry and Bioscience, Aalborg University, Denmark |
| EPI_ISL_429398 | Department of Virus and Microbiological Special Diagnostics, Statens Serum Institut, Copenhagen, Denmark, Artillerivej 5, 2300 Copenahgen S | Albertsen lab, Department of Chemistry and Bioscience, Aalborg University, Denmark |
| EPI_ISL_429399 | Department of Virus and Microbiological Special Diagnostics, Statens Serum Institut, Copenhagen, Denmark, Artillerivej 5, 2300 Copenahgen S | Albertsen lab, Department of Chemistry and Bioscience, Aalborg University, Denmark |
| EPI_ISL_429400 | Department of Virus and Microbiological Special Diagnostics, Statens Serum Institut, Copenhagen, Denmark, Artillerivej 5, 2300 Copenahgen S | Albertsen lab, Department of Chemistry and Bioscience, Aalborg University, Denmark |
| EPI_ISL_429401 | Department of Virus and Microbiological Special Diagnostics, Statens Serum Institut, Copenhagen, Denmark, Artillerivej 5, 2300 Copenahgen S | Albertsen lab, Department of Chemistry and Bioscience, Aalborg University, Denmark |
| EPI_ISL_429402 | Department of Virus and Microbiological Special Diagnostics, Statens Serum Institut, Copenhagen, Denmark, Artillerivej 5, 2300 Copenahgen S | Albertsen lab, Department of Chemistry and Bioscience, Aalborg University, Denmark |
| EPI_ISL_429403 | Department of Virus and Microbiological Special Diagnostics, Statens Serum Institut, Copenhagen, Denmark, Artillerivej 5, 2300 Copenahgen S | Albertsen lab, Department of Chemistry and Bioscience, Aalborg University, Denmark |
| EPI_ISL_429404 | Department of Virus and Microbiological Special Diagnostics, Statens Serum Institut, Copenhagen, Denmark, Artillerivej 5, 2300 Copenahgen S | Albertsen lab, Department of Chemistry and Bioscience, Aalborg University, Denmark |
| EPI_ISL_429405 | Department of Virus and Microbiological Special Diagnostics, Statens Serum Institut, Copenhagen, Denmark, Artillerivej 5, 2300 Copenahgen S | Albertsen lab, Department of Chemistry and Bioscience, Aalborg University, Denmark |
| EPI_ISL_429406 | Department of Virus and Microbiological Special Diagnostics, Statens Serum Institut, Copenhagen, Denmark, Artillerivej 5, 2300 Copenahgen S | Albertsen lab, Department of Chemistry and Bioscience, Aalborg University, Denmark |

| EPI_Accession_ID | Originating Laboratory | Submitting Laboratory |
|---|---|---|
| EPI_ISL_429407 | Department of Virus and Microbiological Special Diagnostics, Statens Serum Institut, Copenhagen, Denmark, Artillerivej 5, 2300 Copenahgen S | Albertsen lab, Department of Chemistry and Bioscience, Aalborg University, Denmark |
| EPI_ISL_429408 | Department of Virus and Microbiological Special Diagnostics, Statens Serum Institut, Copenhagen, Denmark, Artillerivej 5, 2300 Copenahgen S | Albertsen lab, Department of Chemistry and Bioscience, Aalborg University, Denmark |
| EPI_ISL_429409 | Department of Virus and Microbiological Special Diagnostics, Statens Serum Institut, Copenhagen, Denmark, Artillerivej 5, 2300 Copenahgen S | Albertsen lab, Department of Chemistry and Bioscience, Aalborg University, Denmark |
| EPI_ISL_429410 | Department of Virus and Microbiological Special Diagnostics, Statens Serum Institut, Copenhagen, Denmark, Artillerivej 5, 2300 Copenahgen S | Albertsen lab, Department of Chemistry and Bioscience, Aalborg University, Denmark |
| EPI_ISL_429411 | Department of Virus and Microbiological Special Diagnostics, Statens Serum Institut, Copenhagen, Denmark, Artillerivej 5, 2300 Copenahgen S | Albertsen lab, Department of Chemistry and Bioscience, Aalborg University, Denmark |
| EPI_ISL_429412 | Department of Virus and Microbiological Special Diagnostics, Statens Serum Institut, Copenhagen, Denmark, Artillerivej 5, 2300 Copenahgen S | Albertsen lab, Department of Chemistry and Bioscience, Aalborg University, Denmark |
| EPI_ISL_429413 | Department of Virus and Microbiological Special Diagnostics, Statens Serum Institut, Copenhagen, Denmark, Artillerivej 5, 2300 Copenahgen S | Albertsen lab, Department of Chemistry and Bioscience, Aalborg University, Denmark |
| EPI_ISL_429414 | Department of Virus and Microbiological Special Diagnostics, Statens Serum Institut, Copenhagen, Denmark, Artillerivej 5, 2300 Copenahgen S | Albertsen lab, Department of Chemistry and Bioscience, Aalborg University, Denmark |
| EPI_ISL_429415 | Department of Virus and Microbiological Special Diagnostics, Statens Serum Institut, Copenhagen, Denmark, Artillerivej 5, 2300 Copenahgen S | Albertsen lab, Department of Chemistry and Bioscience, Aalborg University, Denmark |
| EPI_ISL_429416 | Department of Virus and Microbiological Special Diagnostics, Statens Serum Institut, Copenhagen, Denmark, Artillerivej 5, 2300 Copenahgen S | Albertsen lab, Department of Chemistry and Bioscience, Aalborg University, Denmark |
| EPI_ISL_429417 | Department of Virus and Microbiological Special Diagnostics, Statens Serum Institut, Copenhagen, Denmark, Artillerivej 5, 2300 Copenahgen S | Albertsen lab, Department of Chemistry and Bioscience, Aalborg University, Denmark |

| EPI_Accession_ID | Originating Laboratory | Submitting Laboratory |
|---|---|---|
| EPI_ISL_429418 | Department of Virus and Microbiological Special Diagnostics, Statens Serum Institut, Copenhagen, Denmark, Artillerivej 5, 2300 Copenahgen S | Albertsen lab, Department of Chemistry and Bioscience, Aalborg University, Denmark |
| EPI_ISL_429419 | Department of Virus and Microbiological Special Diagnostics, Statens Serum Institut, Copenhagen, Denmark, Artillerivej 5, 2300 Copenahgen S | Albertsen lab, Department of Chemistry and Bioscience, Aalborg University, Denmark |
| EPI_ISL_429420 | Department of Virus and Microbiological Special Diagnostics, Statens Serum Institut, Copenhagen, Denmark, Artillerivej 5, 2300 Copenahgen S | Albertsen lab, Department of Chemistry and Bioscience, Aalborg University, Denmark |
| EPI_ISL_429421 | Department of Virus and Microbiological Special Diagnostics, Statens Serum Institut, Copenhagen, Denmark, Artillerivej 5, 2300 Copenahgen S | Albertsen lab, Department of Chemistry and Bioscience, Aalborg University, Denmark |
| EPI_ISL_429422 | Department of Virus and Microbiological Special Diagnostics, Statens Serum Institut, Copenhagen, Denmark, Artillerivej 5, 2300 Copenahgen S | Albertsen lab, Department of Chemistry and Bioscience, Aalborg University, Denmark |
| EPI_ISL_429423 | Department of Virus and Microbiological Special Diagnostics, Statens Serum Institut, Copenhagen, Denmark, Artillerivej 5, 2300 Copenahgen S | Albertsen lab, Department of Chemistry and Bioscience, Aalborg University, Denmark |
| EPI_ISL_429424 | Department of Virus and Microbiological Special Diagnostics, Statens Serum Institut, Copenhagen, Denmark, Artillerivej 5, 2300 Copenahgen S | Albertsen lab, Department of Chemistry and Bioscience, Aalborg University, Denmark |
| EPI_ISL_429425 | Department of Virus and Microbiological Special Diagnostics, Statens Serum Institut, Copenhagen, Denmark, Artillerivej 5, 2300 Copenahgen S | Albertsen lab, Department of Chemistry and Bioscience, Aalborg University, Denmark |
| EPI_ISL_429426 | Department of Virus and Microbiological Special Diagnostics, Statens Serum Institut, Copenhagen, Denmark, Artillerivej 5, 2300 Copenahgen S | Albertsen lab, Department of Chemistry and Bioscience, Aalborg University, Denmark |
| EPI_ISL_429427 | Department of Virus and Microbiological Special Diagnostics, Statens Serum Institut, Copenhagen, Denmark, Artillerivej 5, 2300 Copenahgen S | Albertsen lab, Department of Chemistry and Bioscience, Aalborg University, Denmark |
| EPI_ISL_429428 | Department of Virus and Microbiological Special Diagnostics, Statens Serum Institut, Copenhagen, Denmark, Artillerivej 5, 2300 Copenahgen S | Albertsen lab, Department of Chemistry and Bioscience, Aalborg University, Denmark |

| EPI_Accession_ID | Originating Laboratory | Submitting Laboratory |
|---|---|---|
| EPI_ISL_429429 | Department of Virus and Microbiological Special Diagnostics, Statens Serum Institut, Copenhagen, Denmark, Artillerivej 5, 2300 Copenahgen S | Albertsen lab, Department of Chemistry and Bioscience, Aalborg University, Denmark |
| EPI_ISL_429430 | Department of Virus and Microbiological Special Diagnostics, Statens Serum Institut, Copenhagen, Denmark, Artillerivej 5, 2300 Copenahgen S | Albertsen lab, Department of Chemistry and Bioscience, Aalborg University, Denmark |
| EPI_ISL_429431 | Department of Virus and Microbiological Special Diagnostics, Statens Serum Institut, Copenhagen, Denmark, Artillerivej 5, 2300 Copenahgen S | Albertsen lab, Department of Chemistry and Bioscience, Aalborg University, Denmark |
| EPI_ISL_429432 | Department of Virus and Microbiological Special Diagnostics, Statens Serum Institut, Copenhagen, Denmark, Artillerivej 5, 2300 Copenahgen S | Albertsen lab, Department of Chemistry and Bioscience, Aalborg University, Denmark |
| EPI_ISL_429433 | Department of Virus and Microbiological Special Diagnostics, Statens Serum Institut, Copenhagen, Denmark, Artillerivej 5, 2300 Copenahgen S | Albertsen lab, Department of Chemistry and Bioscience, Aalborg University, Denmark |
| EPI_ISL_429434 | Department of Virus and Microbiological Special Diagnostics, Statens Serum Institut, Copenhagen, Denmark, Artillerivej 5, 2300 Copenahgen S | Albertsen lab, Department of Chemistry and Bioscience, Aalborg University, Denmark |
| EPI_ISL_429435 | Department of Virus and Microbiological Special Diagnostics, Statens Serum Institut, Copenhagen, Denmark, Artillerivej 5, 2300 Copenahgen S | Albertsen lab, Department of Chemistry and Bioscience, Aalborg University, Denmark |
| EPI_ISL_429436 | Department of Virus and Microbiological Special Diagnostics, Statens Serum Institut, Copenhagen, Denmark, Artillerivej 5, 2300 Copenahgen S | Albertsen lab, Department of Chemistry and Bioscience, Aalborg University, Denmark |
| EPI_ISL_429437 | Department of Virus and Microbiological Special Diagnostics, Statens Serum Institut, Copenhagen, Denmark, Artillerivej 5, 2300 Copenahgen S | Albertsen lab, Department of Chemistry and Bioscience, Aalborg University, Denmark |
| EPI_ISL_429438 | Department of Virus and Microbiological Special Diagnostics, Statens Serum Institut, Copenhagen, Denmark, Artillerivej 5, 2300 Copenahgen S | Albertsen lab, Department of Chemistry and Bioscience, Aalborg University, Denmark |
| EPI_ISL_429439 | Department of Virus and Microbiological Special Diagnostics, Statens Serum Institut, Copenhagen, Denmark, Artillerivej 5, 2300 Copenahgen S | Albertsen lab, Department of Chemistry and Bioscience, Aalborg University, Denmark |

| EPI_Accession_ID | Originating Laboratory | Submitting Laboratory |
|---|---|---|
| EPI_ISL_429440 | Department of Virus and Microbiological Special Diagnostics, Statens Serum Institut, Copenhagen, Denmark, Artillerivej 5, 2300 Copenahgen S | Albertsen lab, Department of Chemistry and Bioscience, Aalborg University, Denmark |
| EPI_ISL_429441 | Department of Virus and Microbiological Special Diagnostics, Statens Serum Institut, Copenhagen, Denmark, Artillerivej 5, 2300 Copenahgen S | Albertsen lab, Department of Chemistry and Bioscience, Aalborg University, Denmark |
| EPI_ISL_429442 | Department of Virus and Microbiological Special Diagnostics, Statens Serum Institut, Copenhagen, Denmark, Artillerivej 5, 2300 Copenahgen S | Albertsen lab, Department of Chemistry and Bioscience, Aalborg University, Denmark |
| EPI_ISL_429443 | Department of Virus and Microbiological Special Diagnostics, Statens Serum Institut, Copenhagen, Denmark, Artillerivej 5, 2300 Copenahgen S | Albertsen lab, Department of Chemistry and Bioscience, Aalborg University, Denmark |
| EPI_ISL_429444 | Department of Virus and Microbiological Special Diagnostics, Statens Serum Institut, Copenhagen, Denmark, Artillerivej 5, 2300 Copenahgen S | Albertsen lab, Department of Chemistry and Bioscience, Aalborg University, Denmark |
| EPI_ISL_429445 | Department of Virus and Microbiological Special Diagnostics, Statens Serum Institut, Copenhagen, Denmark, Artillerivej 5, 2300 Copenahgen S | Albertsen lab, Department of Chemistry and Bioscience, Aalborg University, Denmark |
| EPI_ISL_429446 | Department of Virus and Microbiological Special Diagnostics, Statens Serum Institut, Copenhagen, Denmark, Artillerivej 5, 2300 Copenahgen S | Albertsen lab, Department of Chemistry and Bioscience, Aalborg University, Denmark |
| EPI_ISL_429447 | Department of Virus and Microbiological Special Diagnostics, Statens Serum Institut, Copenhagen, Denmark, Artillerivej 5, 2300 Copenahgen S | Albertsen lab, Department of Chemistry and Bioscience, Aalborg University, Denmark |
| EPI_ISL_429448 | Department of Virus and Microbiological Special Diagnostics, Statens Serum Institut, Copenhagen, Denmark, Artillerivej 5, 2300 Copenahgen S | Albertsen lab, Department of Chemistry and Bioscience, Aalborg University, Denmark |
| EPI_ISL_429449 | Department of Virus and Microbiological Special Diagnostics, Statens Serum Institut, Copenhagen, Denmark, Artillerivej 5, 2300 Copenahgen S | Albertsen lab, Department of Chemistry and Bioscience, Aalborg University, Denmark |
| EPI_ISL_429450 | Department of Virus and Microbiological Special Diagnostics, Statens Serum Institut, Copenhagen, Denmark, Artillerivej 5, 2300 Copenahgen S | Albertsen lab, Department of Chemistry and Bioscience, Aalborg University, Denmark |

| EPI_Accession_ID | Originating Laboratory | Submitting Laboratory |
|---|---|---|
| EPI_ISL_429451 | Department of Virus and Microbiological Special Diagnostics, Statens Serum Institut, Copenhagen, Denmark, Artillerivej 5, 2300 Copenahgen S | Albertsen lab, Department of Chemistry and Bioscience, Aalborg University, Denmark |
| EPI_ISL_429452 | Department of Virus and Microbiological Special Diagnostics, Statens Serum Institut, Copenhagen, Denmark, Artillerivej 5, 2300 Copenahgen S | Albertsen lab, Department of Chemistry and Bioscience, Aalborg University, Denmark |
| EPI_ISL_429453 | Department of Virus and Microbiological Special Diagnostics, Statens Serum Institut, Copenhagen, Denmark, Artillerivej 5, 2300 Copenahgen S | Albertsen lab, Department of Chemistry and Bioscience, Aalborg University, Denmark |
| EPI_ISL_429454 | Department of Virus and Microbiological Special Diagnostics, Statens Serum Institut, Copenhagen, Denmark, Artillerivej 5, 2300 Copenahgen S | Albertsen lab, Department of Chemistry and Bioscience, Aalborg University, Denmark |
| EPI_ISL_429455 | Department of Virus and Microbiological Special Diagnostics, Statens Serum Institut, Copenhagen, Denmark, Artillerivej 5, 2300 Copenahgen S | Albertsen lab, Department of Chemistry and Bioscience, Aalborg University, Denmark |
| EPI_ISL_429456 | Department of Virus and Microbiological Special Diagnostics, Statens Serum Institut, Copenhagen, Denmark, Artillerivej 5, 2300 Copenahgen S | Albertsen lab, Department of Chemistry and Bioscience, Aalborg University, Denmark |
| EPI_ISL_429457 | Department of Virus and Microbiological Special Diagnostics, Statens Serum Institut, Copenhagen, Denmark, Artillerivej 5, 2300 Copenahgen S | Albertsen lab, Department of Chemistry and Bioscience, Aalborg University, Denmark |
| EPI_ISL_429458 | Department of Virus and Microbiological Special Diagnostics, Statens Serum Institut, Copenhagen, Denmark, Artillerivej 5, 2300 Copenahgen S | Albertsen lab, Department of Chemistry and Bioscience, Aalborg University, Denmark |
| EPI_ISL_429459 | Department of Virus and Microbiological Special Diagnostics, Statens Serum Institut, Copenhagen, Denmark, Artillerivej 5, 2300 Copenahgen S | Albertsen lab, Department of Chemistry and Bioscience, Aalborg University, Denmark |
| EPI_ISL_429460 | Department of Virus and Microbiological Special Diagnostics, Statens Serum Institut, Copenhagen, Denmark, Artillerivej 5, 2300 Copenahgen S | Albertsen lab, Department of Chemistry and Bioscience, Aalborg University, Denmark |
| EPI_ISL_429461 | Department of Virus and Microbiological Special Diagnostics, Statens Serum Institut, Copenhagen, Denmark, Artillerivej 5, 2300 Copenahgen S | Albertsen lab, Department of Chemistry and Bioscience, Aalborg University, Denmark |

| EPI_Accession_ID | Originating Laboratory | Submitting Laboratory |
|---|---|---|
| EPI_ISL_429462 | Department of Virus and Microbiological Special Diagnostics, Statens Serum Institut, Copenhagen, Denmark, Artillerivej 5, 2300 Copenahgen S | Albertsen lab, Department of Chemistry and Bioscience, Aalborg University, Denmark |
| EPI_ISL_429463 | Department of Virus and Microbiological Special Diagnostics, Statens Serum Institut, Copenhagen, Denmark, Artillerivej 5, 2300 Copenahgen S | Albertsen lab, Department of Chemistry and Bioscience, Aalborg University, Denmark |
| EPI_ISL_429464 | Department of Virus and Microbiological Special Diagnostics, Statens Serum Institut, Copenhagen, Denmark, Artillerivej 5, 2300 Copenahgen S | Albertsen lab, Department of Chemistry and Bioscience, Aalborg University, Denmark |
| EPI_ISL_429465 | Department of Virus and Microbiological Special Diagnostics, Statens Serum Institut, Copenhagen, Denmark, Artillerivej 5, 2300 Copenahgen S | Albertsen lab, Department of Chemistry and Bioscience, Aalborg University, Denmark |
| EPI_ISL_429466 | Department of Virus and Microbiological Special Diagnostics, Statens Serum Institut, Copenhagen, Denmark, Artillerivej 5, 2300 Copenahgen S | Albertsen lab, Department of Chemistry and Bioscience, Aalborg University, Denmark |
| EPI_ISL_429467 | Department of Virus and Microbiological Special Diagnostics, Statens Serum Institut, Copenhagen, Denmark, Artillerivej 5, 2300 Copenahgen S | Albertsen lab, Department of Chemistry and Bioscience, Aalborg University, Denmark |
| EPI_ISL_429468 | Department of Virus and Microbiological Special Diagnostics, Statens Serum Institut, Copenhagen, Denmark, Artillerivej 5, 2300 Copenahgen S | Albertsen lab, Department of Chemistry and Bioscience, Aalborg University, Denmark |
| EPI_ISL_429469 | Department of Virus and Microbiological Special Diagnostics, Statens Serum Institut, Copenhagen, Denmark, Artillerivej 5, 2300 Copenahgen S | Albertsen lab, Department of Chemistry and Bioscience, Aalborg University, Denmark |
| EPI_ISL_429470 | Department of Virus and Microbiological Special Diagnostics, Statens Serum Institut, Copenhagen, Denmark, Artillerivej 5, 2300 Copenahgen S | Albertsen lab, Department of Chemistry and Bioscience, Aalborg University, Denmark |
| EPI_ISL_429471 | Department of Virus and Microbiological Special Diagnostics, Statens Serum Institut, Copenhagen, Denmark, Artillerivej 5, 2300 Copenahgen S | Albertsen lab, Department of Chemistry and Bioscience, Aalborg University, Denmark |
| EPI_ISL_429472 | Department of Virus and Microbiological Special Diagnostics, Statens Serum Institut, Copenhagen, Denmark, Artillerivej 5, 2300 Copenahgen S | Albertsen lab, Department of Chemistry and Bioscience, Aalborg University, Denmark |

| EPI_Accession_ID | Originating Laboratory | Submitting Laboratory |
| --- | --- | --- |
| EPI_ISL_429473 | Department of Virus and Microbiological Special Diagnostics, Statens Serum Institut, Copenhagen, Denmark, Artillerivej 5, 2300 Copenahgen S | Albertsen lab, Department of Chemistry and Bioscience, Aalborg University, Denmark |
| EPI_ISL_429474 | Department of Virus and Microbiological Special Diagnostics, Statens Serum Institut, Copenhagen, Denmark, Artillerivej 5, 2300 Copenahgen S | Albertsen lab, Department of Chemistry and Bioscience, Aalborg University, Denmark |
| EPI_ISL_429475 | Department of Virus and Microbiological Special Diagnostics, Statens Serum Institut, Copenhagen, Denmark, Artillerivej 5, 2300 Copenahgen S | Albertsen lab, Department of Chemistry and Bioscience, Aalborg University, Denmark |
| EPI_ISL_429476 | Department of Virus and Microbiological Special Diagnostics, Statens Serum Institut, Copenhagen, Denmark, Artillerivej 5, 2300 Copenahgen S | Albertsen lab, Department of Chemistry and Bioscience, Aalborg University, Denmark |
| EPI_ISL_429477 | Department of Virus and Microbiological Special Diagnostics, Statens Serum Institut, Copenhagen, Denmark, Artillerivej 5, 2300 Copenahgen S | Albertsen lab, Department of Chemistry and Bioscience, Aalborg University, Denmark |
| EPI_ISL_429478 | Department of Virus and Microbiological Special Diagnostics, Statens Serum Institut, Copenhagen, Denmark, Artillerivej 5, 2300 Copenahgen S | Albertsen lab, Department of Chemistry and Bioscience, Aalborg University, Denmark |
| EPI_ISL_429479 | Department of Virus and Microbiological Special Diagnostics, Statens Serum Institut, Copenhagen, Denmark, Artillerivej 5, 2300 Copenahgen S | Albertsen lab, Department of Chemistry and Bioscience, Aalborg University, Denmark |
| EPI_ISL_429480 | Department of Virus and Microbiological Special Diagnostics, Statens Serum Institut, Copenhagen, Denmark, Artillerivej 5, 2300 Copenahgen S | Albertsen lab, Department of Chemistry and Bioscience, Aalborg University, Denmark |
| EPI_ISL_429481 | Department of Virus and Microbiological Special Diagnostics, Statens Serum Institut, Copenhagen, Denmark, Artillerivej 5, 2300 Copenahgen S | Albertsen lab, Department of Chemistry and Bioscience, Aalborg University, Denmark |
| EPI_ISL_429482 | Department of Virus and Microbiological Special Diagnostics, Statens Serum Institut, Copenhagen, Denmark, Artillerivej 5, 2300 Copenahgen S | Albertsen lab, Department of Chemistry and Bioscience, Aalborg University, Denmark |
| EPI_ISL_429483 | Department of Virus and Microbiological Special Diagnostics, Statens Serum Institut, Copenhagen, Denmark, Artillerivej 5, 2300 Copenahgen S | Albertsen lab, Department of Chemistry and Bioscience, Aalborg University, Denmark |

| EPI_Accession_ID | Originating Laboratory | Submitting Laboratory |
| --- | --- | --- |
| EPI_ISL_429484 | Department of Virus and Microbiological Special Diagnostics, Statens Serum Institut, Copenhagen, Denmark, Artillerivej 5, 2300 Copenahgen S | Albertsen lab, Department of Chemistry and Bioscience, Aalborg University, Denmark |
| EPI_ISL_429485 | Department of Virus and Microbiological Special Diagnostics, Statens Serum Institut, Copenhagen, Denmark, Artillerivej 5, 2300 Copenahgen S | Albertsen lab, Department of Chemistry and Bioscience, Aalborg University, Denmark |
| EPI_ISL_429486 | Department of Virus and Microbiological Special Diagnostics, Statens Serum Institut, Copenhagen, Denmark, Artillerivej 5, 2300 Copenahgen S | Albertsen lab, Department of Chemistry and Bioscience, Aalborg University, Denmark |
| EPI_ISL_429487 | Department of Virus and Microbiological Special Diagnostics, Statens Serum Institut, Copenhagen, Denmark, Artillerivej 5, 2300 Copenahgen S | Albertsen lab, Department of Chemistry and Bioscience, Aalborg University, Denmark |
| EPI_ISL_429488 | Department of Virus and Microbiological Special Diagnostics, Statens Serum Institut, Copenhagen, Denmark, Artillerivej 5, 2300 Copenahgen S | Albertsen lab, Department of Chemistry and Bioscience, Aalborg University, Denmark |
| EPI_ISL_429489 | Department of Virus and Microbiological Special Diagnostics, Statens Serum Institut, Copenhagen, Denmark, Artillerivej 5, 2300 Copenahgen S | Albertsen lab, Department of Chemistry and Bioscience, Aalborg University, Denmark |
| EPI_ISL_429490 | Department of Virus and Microbiological Special Diagnostics, Statens Serum Institut, Copenhagen, Denmark, Artillerivej 5, 2300 Copenahgen S | Albertsen lab, Department of Chemistry and Bioscience, Aalborg University, Denmark |
| EPI_ISL_429491 | Department of Virus and Microbiological Special Diagnostics, Statens Serum Institut, Copenhagen, Denmark, Artillerivej 5, 2300 Copenahgen S | Albertsen lab, Department of Chemistry and Bioscience, Aalborg University, Denmark |
| EPI_ISL_429492 | Department of Virus and Microbiological Special Diagnostics, Statens Serum Institut, Copenhagen, Denmark, Artillerivej 5, 2300 Copenahgen S | Albertsen lab, Department of Chemistry and Bioscience, Aalborg University, Denmark |
| EPI_ISL_429493 | Department of Virus and Microbiological Special Diagnostics, Statens Serum Institut, Copenhagen, Denmark, Artillerivej 5, 2300 Copenahgen S | Albertsen lab, Department of Chemistry and Bioscience, Aalborg University, Denmark |
| EPI_ISL_429494 | Department of Virus and Microbiological Special Diagnostics, Statens Serum Institut, Copenhagen, Denmark, Artillerivej 5, 2300 Copenahgen S | Albertsen lab, Department of Chemistry and Bioscience, Aalborg University, Denmark |

| EPI_Accession_ID | Originating Laboratory | Submitting Laboratory |
|---|---|---|
| EPI_ISL_429495 | Department of Virus and Microbiological Special Diagnostics, Statens Serum Institut, Copenhagen, Denmark, Artillerivej 5, 2300 Copenahgen S | Albertsen lab, Department of Chemistry and Bioscience, Aalborg University, Denmark |
| EPI_ISL_429496 | Department of Virus and Microbiological Special Diagnostics, Statens Serum Institut, Copenhagen, Denmark, Artillerivej 5, 2300 Copenahgen S | Albertsen lab, Department of Chemistry and Bioscience, Aalborg University, Denmark |
| EPI_ISL_429497 | Department of Virus and Microbiological Special Diagnostics, Statens Serum Institut, Copenhagen, Denmark, Artillerivej 5, 2300 Copenahgen S | Albertsen lab, Department of Chemistry and Bioscience, Aalborg University, Denmark |
| EPI_ISL_429498 | Department of Virus and Microbiological Special Diagnostics, Statens Serum Institut, Copenhagen, Denmark, Artillerivej 5, 2300 Copenahgen S | Albertsen lab, Department of Chemistry and Bioscience, Aalborg University, Denmark |
| EPI_ISL_429499 | Department of Virus and Microbiological Special Diagnostics, Statens Serum Institut, Copenhagen, Denmark, Artillerivej 5, 2300 Copenahgen S | Albertsen lab, Department of Chemistry and Bioscience, Aalborg University, Denmark |
| EPI_ISL_429500 | Department of Virus and Microbiological Special Diagnostics, Statens Serum Institut, Copenhagen, Denmark, Artillerivej 5, 2300 Copenahgen S | Albertsen lab, Department of Chemistry and Bioscience, Aalborg University, Denmark |
| EPI_ISL_429501 | Department of Virus and Microbiological Special Diagnostics, Statens Serum Institut, Copenhagen, Denmark, Artillerivej 5, 2300 Copenahgen S | Albertsen lab, Department of Chemistry and Bioscience, Aalborg University, Denmark |
| EPI_ISL_429502 | Department of Virus and Microbiological Special Diagnostics, Statens Serum Institut, Copenhagen, Denmark, Artillerivej 5, 2300 Copenahgen S | Albertsen lab, Department of Chemistry and Bioscience, Aalborg University, Denmark |
| EPI_ISL_429503 | Department of Virus and Microbiological Special Diagnostics, Statens Serum Institut, Copenhagen, Denmark, Artillerivej 5, 2300 Copenahgen S | Albertsen lab, Department of Chemistry and Bioscience, Aalborg University, Denmark |
| EPI_ISL_429504 | Department of Virus and Microbiological Special Diagnostics, Statens Serum Institut, Copenhagen, Denmark, Artillerivej 5, 2300 Copenahgen S | Albertsen lab, Department of Chemistry and Bioscience, Aalborg University, Denmark |
| EPI_ISL_429505 | Department of Virus and Microbiological Special Diagnostics, Statens Serum Institut, Copenhagen, Denmark, Artillerivej 5, 2300 Copenahgen S | Albertsen lab, Department of Chemistry and Bioscience, Aalborg University, Denmark |

| EPI_Accession_ID | Originating Laboratory | Submitting Laboratory |
|---|---|---|
| EPI_ISL_429506 | Department of Virus and Microbiological Special Diagnostics, Statens Serum Institut, Copenhagen, Denmark, Artillerivej 5, 2300 Copenahgen S | Albertsen lab, Department of Chemistry and Bioscience, Aalborg University, Denmark |
| EPI_ISL_429507 | Department of Virus and Microbiological Special Diagnostics, Statens Serum Institut, Copenhagen, Denmark, Artillerivej 5, 2300 Copenahgen S | Albertsen lab, Department of Chemistry and Bioscience, Aalborg University, Denmark |
| EPI_ISL_429508 | Department of Virus and Microbiological Special Diagnostics, Statens Serum Institut, Copenhagen, Denmark, Artillerivej 5, 2300 Copenahgen S | Albertsen lab, Department of Chemistry and Bioscience, Aalborg University, Denmark |
| EPI_ISL_429509 | Department of Virus and Microbiological Special Diagnostics, Statens Serum Institut, Copenhagen, Denmark, Artillerivej 5, 2300 Copenahgen S | Albertsen lab, Department of Chemistry and Bioscience, Aalborg University, Denmark |
| EPI_ISL_429510 | Department of Virus and Microbiological Special Diagnostics, Statens Serum Institut, Copenhagen, Denmark, Artillerivej 5, 2300 Copenahgen S | Albertsen lab, Department of Chemistry and Bioscience, Aalborg University, Denmark |
| EPI_ISL_429511 | Department of Virus and Microbiological Special Diagnostics, Statens Serum Institut, Copenhagen, Denmark, Artillerivej 5, 2300 Copenahgen S | Albertsen lab, Department of Chemistry and Bioscience, Aalborg University, Denmark |
| EPI_ISL_429512 | Department of Virus and Microbiological Special Diagnostics, Statens Serum Institut, Copenhagen, Denmark, Artillerivej 5, 2300 Copenahgen S | Albertsen lab, Department of Chemistry and Bioscience, Aalborg University, Denmark |
| EPI_ISL_429513 | Department of Virus and Microbiological Special Diagnostics, Statens Serum Institut, Copenhagen, Denmark, Artillerivej 5, 2300 Copenahgen S | Albertsen lab, Department of Chemistry and Bioscience, Aalborg University, Denmark |
| EPI_ISL_429514 | Department of Virus and Microbiological Special Diagnostics, Statens Serum Institut, Copenhagen, Denmark, Artillerivej 5, 2300 Copenahgen S | Albertsen lab, Department of Chemistry and Bioscience, Aalborg University, Denmark |
| EPI_ISL_429515 | Department of Virus and Microbiological Special Diagnostics, Statens Serum Institut, Copenhagen, Denmark, Artillerivej 5, 2300 Copenahgen S | Albertsen lab, Department of Chemistry and Bioscience, Aalborg University, Denmark |
| EPI_ISL_429516 | Department of Virus and Microbiological Special Diagnostics, Statens Serum Institut, Copenhagen, Denmark, Artillerivej 5, 2300 Copenahgen S | Albertsen lab, Department of Chemistry and Bioscience, Aalborg University, Denmark |

| EPI_Accession_ID | Originating Laboratory | Submitting Laboratory |
|---|---|---|
| EPI_ISL_429517 | Department of Virus and Microbiological Special Diagnostics, Statens Serum Institut, Copenhagen, Denmark, Artillerivej 5, 2300 Copenahgen S | Albertsen lab, Department of Chemistry and Bioscience, Aalborg University, Denmark |
| EPI_ISL_429518 | Department of Virus and Microbiological Special Diagnostics, Statens Serum Institut, Copenhagen, Denmark, Artillerivej 5, 2300 Copenahgen S | Albertsen lab, Department of Chemistry and Bioscience, Aalborg University, Denmark |
| EPI_ISL_429519 | Department of Virus and Microbiological Special Diagnostics, Statens Serum Institut, Copenhagen, Denmark, Artillerivej 5, 2300 Copenahgen S | Albertsen lab, Department of Chemistry and Bioscience, Aalborg University, Denmark |
| EPI_ISL_429520 | Department of Virus and Microbiological Special Diagnostics, Statens Serum Institut, Copenhagen, Denmark, Artillerivej 5, 2300 Copenahgen S | Albertsen lab, Department of Chemistry and Bioscience, Aalborg University, Denmark |
| EPI_ISL_429521 | Department of Virus and Microbiological Special Diagnostics, Statens Serum Institut, Copenhagen, Denmark, Artillerivej 5, 2300 Copenahgen S | Albertsen lab, Department of Chemistry and Bioscience, Aalborg University, Denmark |
| EPI_ISL_429522 | Department of Virus and Microbiological Special Diagnostics, Statens Serum Institut, Copenhagen, Denmark, Artillerivej 5, 2300 Copenahgen S | Albertsen lab, Department of Chemistry and Bioscience, Aalborg University, Denmark |
| EPI_ISL_429524 | Department of Virus and Microbiological Special Diagnostics, Statens Serum Institut, Copenhagen, Denmark, Artillerivej 5, 2300 Copenahgen S | Albertsen lab, Department of Chemistry and Bioscience, Aalborg University, Denmark |
| EPI_ISL_429525 | Department of Virus and Microbiological Special Diagnostics, Statens Serum Institut, Copenhagen, Denmark, Artillerivej 5, 2300 Copenahgen S | Albertsen lab, Department of Chemistry and Bioscience, Aalborg University, Denmark |
| EPI_ISL_429526 | Department of Virus and Microbiological Special Diagnostics, Statens Serum Institut, Copenhagen, Denmark, Artillerivej 5, 2300 Copenahgen S | Albertsen lab, Department of Chemistry and Bioscience, Aalborg University, Denmark |
| EPI_ISL_429527 | Department of Virus and Microbiological Special Diagnostics, Statens Serum Institut, Copenhagen, Denmark, Artillerivej 5, 2300 Copenahgen S | Albertsen lab, Department of Chemistry and Bioscience, Aalborg University, Denmark |
| EPI_ISL_429528 | Department of Virus and Microbiological Special Diagnostics, Statens Serum Institut, Copenhagen, Denmark, Artillerivej 5, 2300 Copenahgen S | Albertsen lab, Department of Chemistry and Bioscience, Aalborg University, Denmark |

| EPI_Accession_ID | Originating Laboratory | Submitting Laboratory |
|---|---|---|
| EPI_ISL_429529 | Department of Virus and Microbiological Special Diagnostics, Statens Serum Institut, Copenhagen, Denmark, Artillerivej 5, 2300 Copenahgen S | Albertsen lab, Department of Chemistry and Bioscience, Aalborg University, Denmark |
| EPI_ISL_429530 | Department of Virus and Microbiological Special Diagnostics, Statens Serum Institut, Copenhagen, Denmark, Artillerivej 5, 2300 Copenahgen S | Albertsen lab, Department of Chemistry and Bioscience, Aalborg University, Denmark |
| EPI_ISL_429531 | Department of Virus and Microbiological Special Diagnostics, Statens Serum Institut, Copenhagen, Denmark, Artillerivej 5, 2300 Copenahgen S | Albertsen lab, Department of Chemistry and Bioscience, Aalborg University, Denmark |
| EPI_ISL_429532 | Department of Virus and Microbiological Special Diagnostics, Statens Serum Institut, Copenhagen, Denmark, Artillerivej 5, 2300 Copenahgen S | Albertsen lab, Department of Chemistry and Bioscience, Aalborg University, Denmark |
| EPI_ISL_429533 | Department of Virus and Microbiological Special Diagnostics, Statens Serum Institut, Copenhagen, Denmark, Artillerivej 5, 2300 Copenahgen S | Albertsen lab, Department of Chemistry and Bioscience, Aalborg University, Denmark |
| EPI_ISL_429534 | Department of Virus and Microbiological Special Diagnostics, Statens Serum Institut, Copenhagen, Denmark, Artillerivej 5, 2300 Copenahgen S | Albertsen lab, Department of Chemistry and Bioscience, Aalborg University, Denmark |
| EPI_ISL_429535 | Department of Virus and Microbiological Special Diagnostics, Statens Serum Institut, Copenhagen, Denmark, Artillerivej 5, 2300 Copenahgen S | Albertsen lab, Department of Chemistry and Bioscience, Aalborg University, Denmark |
| EPI_ISL_429536 | Department of Virus and Microbiological Special Diagnostics, Statens Serum Institut, Copenhagen, Denmark, Artillerivej 5, 2300 Copenahgen S | Albertsen lab, Department of Chemistry and Bioscience, Aalborg University, Denmark |
| EPI_ISL_429537 | Department of Virus and Microbiological Special Diagnostics, Statens Serum Institut, Copenhagen, Denmark, Artillerivej 5, 2300 Copenahgen S | Albertsen lab, Department of Chemistry and Bioscience, Aalborg University, Denmark |
| EPI_ISL_429538 | Department of Virus and Microbiological Special Diagnostics, Statens Serum Institut, Copenhagen, Denmark, Artillerivej 5, 2300 Copenahgen S | Albertsen lab, Department of Chemistry and Bioscience, Aalborg University, Denmark |
| EPI_ISL_429539 | Department of Virus and Microbiological Special Diagnostics, Statens Serum Institut, Copenhagen, Denmark, Artillerivej 5, 2300 Copenahgen S | Albertsen lab, Department of Chemistry and Bioscience, Aalborg University, Denmark |

| EPI_Accession_ID | Originating Laboratory | Submitting Laboratory |
|---|---|---|
| EPI_ISL_429540 | Department of Virus and Microbiological Special Diagnostics, Statens Serum Institut, Copenhagen, Denmark, Artillerivej 5, 2300 Copenahgen S | Albertsen lab, Department of Chemistry and Bioscience, Aalborg University, Denmark |
| EPI_ISL_429541 | Department of Virus and Microbiological Special Diagnostics, Statens Serum Institut, Copenhagen, Denmark, Artillerivej 5, 2300 Copenahgen S | Albertsen lab, Department of Chemistry and Bioscience, Aalborg University, Denmark |
| EPI_ISL_429542 | Department of Virus and Microbiological Special Diagnostics, Statens Serum Institut, Copenhagen, Denmark, Artillerivej 5, 2300 Copenahgen S | Albertsen lab, Department of Chemistry and Bioscience, Aalborg University, Denmark |
| EPI_ISL_429543 | Department of Virus and Microbiological Special Diagnostics, Statens Serum Institut, Copenhagen, Denmark, Artillerivej 5, 2300 Copenahgen S | Albertsen lab, Department of Chemistry and Bioscience, Aalborg University, Denmark |
| EPI_ISL_429544 | Department of Virus and Microbiological Special Diagnostics, Statens Serum Institut, Copenhagen, Denmark, Artillerivej 5, 2300 Copenahgen S | Albertsen lab, Department of Chemistry and Bioscience, Aalborg University, Denmark |
| EPI_ISL_429545 | Department of Virus and Microbiological Special Diagnostics, Statens Serum Institut, Copenhagen, Denmark, Artillerivej 5, 2300 Copenahgen S | Albertsen lab, Department of Chemistry and Bioscience, Aalborg University, Denmark |
| EPI_ISL_429546 | Department of Virus and Microbiological Special Diagnostics, Statens Serum Institut, Copenhagen, Denmark, Artillerivej 5, 2300 Copenahgen S | Albertsen lab, Department of Chemistry and Bioscience, Aalborg University, Denmark |
| EPI_ISL_429547 | Department of Virus and Microbiological Special Diagnostics, Statens Serum Institut, Copenhagen, Denmark, Artillerivej 5, 2300 Copenahgen S | Albertsen lab, Department of Chemistry and Bioscience, Aalborg University, Denmark |
| EPI_ISL_429548 | Department of Virus and Microbiological Special Diagnostics, Statens Serum Institut, Copenhagen, Denmark, Artillerivej 5, 2300 Copenahgen S | Albertsen lab, Department of Chemistry and Bioscience, Aalborg University, Denmark |
| EPI_ISL_429549 | Department of Virus and Microbiological Special Diagnostics, Statens Serum Institut, Copenhagen, Denmark, Artillerivej 5, 2300 Copenahgen S | Albertsen lab, Department of Chemistry and Bioscience, Aalborg University, Denmark |
| EPI_ISL_429550 | Department of Virus and Microbiological Special Diagnostics, Statens Serum Institut, Copenhagen, Denmark, Artillerivej 5, 2300 Copenahgen S | Albertsen lab, Department of Chemistry and Bioscience, Aalborg University, Denmark |

| EPI_Accession_ID | Originating Laboratory | Submitting Laboratory |
|---|---|---|
| EPI_ISL_429551 | Department of Virus and Microbiological Special Diagnostics, Statens Serum Institut, Copenhagen, Denmark, Artillerivej 5, 2300 Copenahgen S | Albertsen lab, Department of Chemistry and Bioscience, Aalborg University, Denmark |
| EPI_ISL_429552 | Department of Virus and Microbiological Special Diagnostics, Statens Serum Institut, Copenhagen, Denmark, Artillerivej 5, 2300 Copenahgen S | Albertsen lab, Department of Chemistry and Bioscience, Aalborg University, Denmark |
| EPI_ISL_429553 | Department of Virus and Microbiological Special Diagnostics, Statens Serum Institut, Copenhagen, Denmark, Artillerivej 5, 2300 Copenahgen S | Albertsen lab, Department of Chemistry and Bioscience, Aalborg University, Denmark |
| EPI_ISL_429554 | Department of Virus and Microbiological Special Diagnostics, Statens Serum Institut, Copenhagen, Denmark, Artillerivej 5, 2300 Copenahgen S | Albertsen lab, Department of Chemistry and Bioscience, Aalborg University, Denmark |
| EPI_ISL_429555 | Department of Virus and Microbiological Special Diagnostics, Statens Serum Institut, Copenhagen, Denmark, Artillerivej 5, 2300 Copenahgen S | Albertsen lab, Department of Chemistry and Bioscience, Aalborg University, Denmark |
| EPI_ISL_429556 | Department of Virus and Microbiological Special Diagnostics, Statens Serum Institut, Copenhagen, Denmark, Artillerivej 5, 2300 Copenahgen S | Albertsen lab, Department of Chemistry and Bioscience, Aalborg University, Denmark |
| EPI_ISL_429557 | Department of Virus and Microbiological Special Diagnostics, Statens Serum Institut, Copenhagen, Denmark, Artillerivej 5, 2300 Copenahgen S | Albertsen lab, Department of Chemistry and Bioscience, Aalborg University, Denmark |
| EPI_ISL_429558 | Department of Virus and Microbiological Special Diagnostics, Statens Serum Institut, Copenhagen, Denmark, Artillerivej 5, 2300 Copenahgen S | Albertsen lab, Department of Chemistry and Bioscience, Aalborg University, Denmark |
| EPI_ISL_429559 | Department of Virus and Microbiological Special Diagnostics, Statens Serum Institut, Copenhagen, Denmark, Artillerivej 5, 2300 Copenahgen S | Albertsen lab, Department of Chemistry and Bioscience, Aalborg University, Denmark |
| EPI_ISL_429560 | Department of Virus and Microbiological Special Diagnostics, Statens Serum Institut, Copenhagen, Denmark, Artillerivej 5, 2300 Copenahgen S | Albertsen lab, Department of Chemistry and Bioscience, Aalborg University, Denmark |
| EPI_ISL_429561 | Department of Virus and Microbiological Special Diagnostics, Statens Serum Institut, Copenhagen, Denmark, Artillerivej 5, 2300 Copenahgen S | Albertsen lab, Department of Chemistry and Bioscience, Aalborg University, Denmark |

| EPI_Accession_ID | Originating Laboratory | Submitting Laboratory |
|---|---|---|
| EPI_ISL_429562 | Department of Virus and Microbiological Special Diagnostics, Statens Serum Institut, Copenhagen, Denmark, Artillerivej 5, 2300 Copenahgen S | Albertsen lab, Department of Chemistry and Bioscience, Aalborg University, Denmark |
| EPI_ISL_429563 | Department of Virus and Microbiological Special Diagnostics, Statens Serum Institut, Copenhagen, Denmark, Artillerivej 5, 2300 Copenahgen S | Albertsen lab, Department of Chemistry and Bioscience, Aalborg University, Denmark |
| EPI_ISL_429564 | Department of Virus and Microbiological Special Diagnostics, Statens Serum Institut, Copenhagen, Denmark, Artillerivej 5, 2300 Copenahgen S | Albertsen lab, Department of Chemistry and Bioscience, Aalborg University, Denmark |
| EPI_ISL_429565 | Department of Virus and Microbiological Special Diagnostics, Statens Serum Institut, Copenhagen, Denmark, Artillerivej 5, 2300 Copenahgen S | Albertsen lab, Department of Chemistry and Bioscience, Aalborg University, Denmark |
| EPI_ISL_429566 | Department of Virus and Microbiological Special Diagnostics, Statens Serum Institut, Copenhagen, Denmark, Artillerivej 5, 2300 Copenahgen S | Albertsen lab, Department of Chemistry and Bioscience, Aalborg University, Denmark |
| EPI_ISL_429567 | Department of Virus and Microbiological Special Diagnostics, Statens Serum Institut, Copenhagen, Denmark, Artillerivej 5, 2300 Copenahgen S | Albertsen lab, Department of Chemistry and Bioscience, Aalborg University, Denmark |
| EPI_ISL_429568 | Department of Virus and Microbiological Special Diagnostics, Statens Serum Institut, Copenhagen, Denmark, Artillerivej 5, 2300 Copenahgen S | Albertsen lab, Department of Chemistry and Bioscience, Aalborg University, Denmark |
| EPI_ISL_429569 | Department of Virus and Microbiological Special Diagnostics, Statens Serum Institut, Copenhagen, Denmark, Artillerivej 5, 2300 Copenahgen S | Albertsen lab, Department of Chemistry and Bioscience, Aalborg University, Denmark |
| EPI_ISL_429570 | Department of Virus and Microbiological Special Diagnostics, Statens Serum Institut, Copenhagen, Denmark, Artillerivej 5, 2300 Copenahgen S | Albertsen lab, Department of Chemistry and Bioscience, Aalborg University, Denmark |
| EPI_ISL_429571 | Department of Virus and Microbiological Special Diagnostics, Statens Serum Institut, Copenhagen, Denmark, Artillerivej 5, 2300 Copenahgen S | Albertsen lab, Department of Chemistry and Bioscience, Aalborg University, Denmark |
| EPI_ISL_429572 | Department of Virus and Microbiological Special Diagnostics, Statens Serum Institut, Copenhagen, Denmark, Artillerivej 5, 2300 Copenahgen S | Albertsen lab, Department of Chemistry and Bioscience, Aalborg University, Denmark |

| EPI_Accession_ID | Originating Laboratory | Submitting Laboratory |
|---|---|---|
| EPI_ISL_429573 | Department of Virus and Microbiological Special Diagnostics, Statens Serum Institut, Copenhagen, Denmark, Artillerivej 5, 2300 Copenahgen S | Albertsen lab, Department of Chemistry and Bioscience, Aalborg University, Denmark |
| EPI_ISL_429574 | Department of Virus and Microbiological Special Diagnostics, Statens Serum Institut, Copenhagen, Denmark, Artillerivej 5, 2300 Copenahgen S | Albertsen lab, Department of Chemistry and Bioscience, Aalborg University, Denmark |
| EPI_ISL_429575 | Department of Virus and Microbiological Special Diagnostics, Statens Serum Institut, Copenhagen, Denmark, Artillerivej 5, 2300 Copenahgen S | Albertsen lab, Department of Chemistry and Bioscience, Aalborg University, Denmark |
| EPI_ISL_429576 | Department of Virus and Microbiological Special Diagnostics, Statens Serum Institut, Copenhagen, Denmark, Artillerivej 5, 2300 Copenahgen S | Albertsen lab, Department of Chemistry and Bioscience, Aalborg University, Denmark |
| EPI_ISL_429577 | Department of Virus and Microbiological Special Diagnostics, Statens Serum Institut, Copenhagen, Denmark, Artillerivej 5, 2300 Copenahgen S | Albertsen lab, Department of Chemistry and Bioscience, Aalborg University, Denmark |
| EPI_ISL_429578 | Department of Virus and Microbiological Special Diagnostics, Statens Serum Institut, Copenhagen, Denmark, Artillerivej 5, 2300 Copenahgen S | Albertsen lab, Department of Chemistry and Bioscience, Aalborg University, Denmark |
| EPI_ISL_429579 | Department of Virus and Microbiological Special Diagnostics, Statens Serum Institut, Copenhagen, Denmark, Artillerivej 5, 2300 Copenahgen S | Albertsen lab, Department of Chemistry and Bioscience, Aalborg University, Denmark |
| EPI_ISL_429580 | Department of Virus and Microbiological Special Diagnostics, Statens Serum Institut, Copenhagen, Denmark, Artillerivej 5, 2300 Copenahgen S | Albertsen lab, Department of Chemistry and Bioscience, Aalborg University, Denmark |
| EPI_ISL_429581 | Department of Virus and Microbiological Special Diagnostics, Statens Serum Institut, Copenhagen, Denmark, Artillerivej 5, 2300 Copenahgen S | Albertsen lab, Department of Chemistry and Bioscience, Aalborg University, Denmark |
| EPI_ISL_429582 | Department of Virus and Microbiological Special Diagnostics, Statens Serum Institut, Copenhagen, Denmark, Artillerivej 5, 2300 Copenahgen S | Albertsen lab, Department of Chemistry and Bioscience, Aalborg University, Denmark |
| EPI_ISL_429583 | Department of Virus and Microbiological Special Diagnostics, Statens Serum Institut, Copenhagen, Denmark, Artillerivej 5, 2300 Copenahgen S | Albertsen lab, Department of Chemistry and Bioscience, Aalborg University, Denmark |

| EPI_Accession_ID | Originating Laboratory | Submitting Laboratory |
|---|---|---|
| EPI_ISL_429584 | Department of Virus and Microbiological Special Diagnostics, Statens Serum Institut, Copenhagen, Denmark, Artillerivej 5, 2300 Copenahgen S | Albertsen lab, Department of Chemistry and Bioscience, Aalborg University, Denmark |
| EPI_ISL_429585 | Department of Virus and Microbiological Special Diagnostics, Statens Serum Institut, Copenhagen, Denmark, Artillerivej 5, 2300 Copenahgen S | Albertsen lab, Department of Chemistry and Bioscience, Aalborg University, Denmark |
| EPI_ISL_429586 | Department of Virus and Microbiological Special Diagnostics, Statens Serum Institut, Copenhagen, Denmark, Artillerivej 5, 2300 Copenahgen S | Albertsen lab, Department of Chemistry and Bioscience, Aalborg University, Denmark |
| EPI_ISL_429587 | Department of Virus and Microbiological Special Diagnostics, Statens Serum Institut, Copenhagen, Denmark, Artillerivej 5, 2300 Copenahgen S | Albertsen lab, Department of Chemistry and Bioscience, Aalborg University, Denmark |
| EPI_ISL_429588 | Department of Virus and Microbiological Special Diagnostics, Statens Serum Institut, Copenhagen, Denmark, Artillerivej 5, 2300 Copenahgen S | Albertsen lab, Department of Chemistry and Bioscience, Aalborg University, Denmark |
| EPI_ISL_429589 | Department of Virus and Microbiological Special Diagnostics, Statens Serum Institut, Copenhagen, Denmark, Artillerivej 5, 2300 Copenahgen S | Albertsen lab, Department of Chemistry and Bioscience, Aalborg University, Denmark |
| EPI_ISL_429590 | Department of Virus and Microbiological Special Diagnostics, Statens Serum Institut, Copenhagen, Denmark, Artillerivej 5, 2300 Copenahgen S | Albertsen lab, Department of Chemistry and Bioscience, Aalborg University, Denmark |
| EPI_ISL_429597 | UW Virology Lab | UW Virology Lab |
| EPI_ISL_429598 | UW Virology Lab | UW Virology Lab |
| EPI_ISL_429599 | UW Virology Lab | UW Virology Lab |
| EPI_ISL_429600 | UW Virology Lab | UW Virology Lab |
| EPI_ISL_429601 | UW Virology Lab | UW Virology Lab |
| EPI_ISL_429602 | UW Virology Lab | UW Virology Lab |
| EPI_ISL_429603 | UW Virology Lab | UW Virology Lab |
| EPI_ISL_429605 | UW Virology Lab | UW Virology Lab |
| EPI_ISL_429606 | UW Virology Lab | UW Virology Lab |
| EPI_ISL_429607 | UW Virology Lab | UW Virology Lab |
| EPI_ISL_429608 | UW Virology Lab | UW Virology Lab |
| EPI_ISL_429609 | UW Virology Lab | UW Virology Lab |

| EPI_Accession_ID | Originating Laboratory | Submitting Laboratory |
|---|---|---|
| EPI_ISL_429610 | UW Virology Lab | UW Virology Lab |
| EPI_ISL_429611 | UW Virology Lab | UW Virology Lab |
| EPI_ISL_429612 | UW Virology Lab | UW Virology Lab |
| EPI_ISL_429613 | UW Virology Lab | UW Virology Lab |
| EPI_ISL_429614 | UW Virology Lab | UW Virology Lab |
| EPI_ISL_429615 | UW Virology Lab | UW Virology Lab |
| EPI_ISL_429616 | UW Virology Lab | UW Virology Lab |
| EPI_ISL_429617 | UW Virology Lab | UW Virology Lab |
| EPI_ISL_429618 | UW Virology Lab | UW Virology Lab |
| EPI_ISL_429619 | UW Virology Lab | UW Virology Lab |
| EPI_ISL_429621 | UW Virology Lab | UW Virology Lab |
| EPI_ISL_429622 | UW Virology Lab | UW Virology Lab |
| EPI_ISL_429623 | UW Virology Lab | UW Virology Lab |
| EPI_ISL_429624 | UW Virology Lab | UW Virology Lab |
| EPI_ISL_429625 | UW Virology Lab | UW Virology Lab |
| EPI_ISL_429626 | UW Virology Lab | UW Virology Lab |
| EPI_ISL_429627 | UW Virology Lab | UW Virology Lab |
| EPI_ISL_429628 | UW Virology Lab | UW Virology Lab |
| EPI_ISL_429629 | UW Virology Lab | UW Virology Lab |
| EPI_ISL_429630 | UW Virology Lab | UW Virology Lab |
| EPI_ISL_429631 | UW Virology Lab | UW Virology Lab |
| EPI_ISL_429632 | UW Virology Lab | UW Virology Lab |
| EPI_ISL_429633 | UW Virology Lab | UW Virology Lab |
| EPI_ISL_429634 | UW Virology Lab | UW Virology Lab |
| EPI_ISL_429635 | UW Virology Lab | UW Virology Lab |
| EPI_ISL_429636 | UW Virology Lab | UW Virology Lab |
| EPI_ISL_429637 | UW Virology Lab | UW Virology Lab |
| EPI_ISL_429638 | UW Virology Lab | UW Virology Lab |
| EPI_ISL_429639 | UW Virology Lab | UW Virology Lab |
| EPI_ISL_429640 | UW Virology Lab | UW Virology Lab |
| EPI_ISL_429641 | UW Virology Lab | UW Virology Lab |
| EPI_ISL_429642 | UW Virology Lab | UW Virology Lab |
| EPI_ISL_429643 | UW Virology Lab | UW Virology Lab |

| EPI_Accession_ID | Originating Laboratory | Submitting Laboratory |
|---|---|---|
| EPI_ISL_429644 | UW Virology Lab | UW Virology Lab |
| EPI_ISL_429645 | UW Virology Lab | UW Virology Lab |
| EPI_ISL_429646 | UW Virology Lab | UW Virology Lab |
| EPI_ISL_429647 | UW Virology Lab | UW Virology Lab |
| EPI_ISL_429648 | UW Virology Lab | UW Virology Lab |
| EPI_ISL_429649 | UW Virology Lab | UW Virology Lab |
| EPI_ISL_429650 | UW Virology Lab | UW Virology Lab |
| EPI_ISL_429651 | UW Virology Lab | UW Virology Lab |
| EPI_ISL_429652 | UW Virology Lab | UW Virology Lab |
| EPI_ISL_429653 | UW Virology Lab | UW Virology Lab |
| EPI_ISL_429654 | UW Virology Lab | UW Virology Lab |
| EPI_ISL_429655 | UW Virology Lab | UW Virology Lab |
| EPI_ISL_429656 | UW Virology Lab | UW Virology Lab |
| EPI_ISL_429665 | Central Public Health Laboratory/OctÃ¡vio MagalhÃ£es Institute (IOM) from the Ezequiel Dias Foundation (FUNED) | Instituto OctÃ¡vio MagalhÃ£es / FundaÃ§Ã£o Ezequiel Dias (IOM/Funed) |
| EPI_ISL_429666 | Central Public Health Laboratory/OctÃ¡vio MagalhÃ£es Institute (IOM) from the Ezequiel Dias Foundation (FUNED) | Instituto OctÃ¡vio MagalhÃ£es / FundaÃ§Ã£o Ezequiel Dias (IOM/Funed) |
| EPI_ISL_429667 | Central Public Health Laboratory/OctÃ¡vio MagalhÃ£es Institute (IOM) from the Ezequiel Dias Foundation (FUNED) | Instituto OctÃ¡vio MagalhÃ£es / FundaÃ§Ã£o Ezequiel Dias (IOM/Funed) |
| EPI_ISL_429669 | Central Public Health Laboratory/OctÃ¡vio MagalhÃ£es Institute (IOM) from the Ezequiel Dias Foundation (FUNED) | Instituto OctÃ¡vio MagalhÃ£es / FundaÃ§Ã£o Ezequiel Dias (IOM/Funed) |
| EPI_ISL_429671 | Central Public Health Laboratory/OctÃ¡vio MagalhÃ£es Institute (IOM) from the Ezequiel Dias Foundation (FUNED) | Instituto OctÃ¡vio MagalhÃ£es / FundaÃ§Ã£o Ezequiel Dias (IOM/Funed) |
| EPI_ISL_429672 | Central Public Health Laboratory/OctÃ¡vio MagalhÃ£es Institute (IOM) from the Ezequiel Dias Foundation (FUNED) | Instituto OctÃ¡vio MagalhÃ£es / FundaÃ§Ã£o Ezequiel Dias (IOM/Funed) |

| EPI_Accession_ID | Originating Laboratory | Submitting Laboratory |
|---|---|---|
| EPI_ISL_429674 | Central Public Health Laboratory/OctÃ¡vio MagalhÃ£es Institute (IOM) from the Ezequiel Dias Foundation (FUNED) | Instituto OctÃ¡vio MagalhÃ£es / FundaÃ§Ã£o Ezequiel Dias (IOM/Funed) |
| EPI_ISL_429675 | Central Public Health Laboratory/OctÃ¡vio MagalhÃ£es Institute (IOM) from the Ezequiel Dias Foundation (FUNED) | Instituto OctÃ¡vio MagalhÃ£es / FundaÃ§Ã£o Ezequiel Dias (IOM/Funed) |
| EPI_ISL_429676 | Central Public Health Laboratory/OctÃ¡vio MagalhÃ£es Institute (IOM) from the Ezequiel Dias Foundation (FUNED) | Instituto OctÃ¡vio MagalhÃ£es / FundaÃ§Ã£o Ezequiel Dias (IOM/Funed) |
| EPI_ISL_429677 | Central Public Health Laboratory/OctÃ¡vio MagalhÃ£es Institute (IOM) from the Ezequiel Dias Foundation (FUNED) | Instituto OctÃ¡vio MagalhÃ£es / FundaÃ§Ã£o Ezequiel Dias (IOM/Funed) |
| EPI_ISL_429679 | Central Public Health Laboratory/OctÃ¡vio MagalhÃ£es Institute (IOM) from the Ezequiel Dias Foundation (FUNED) | Instituto OctÃ¡vio MagalhÃ£es / FundaÃ§Ã£o Ezequiel Dias (IOM/Funed) |
| EPI_ISL_429680 | Central Public Health Laboratory/OctÃ¡vio MagalhÃ£es Institute (IOM) from the Ezequiel Dias Foundation (FUNED) | Instituto OctÃ¡vio MagalhÃ£es / FundaÃ§Ã£o Ezequiel Dias (IOM/Funed) |
| EPI_ISL_429681 | Central Public Health Laboratory/OctÃ¡vio MagalhÃ£es Institute (IOM) from the Ezequiel Dias Foundation (FUNED) | Instituto OctÃ¡vio MagalhÃ£es / FundaÃ§Ã£o Ezequiel Dias (IOM/Funed) |
| EPI_ISL_429684 | Central Public Health Laboratory/OctÃ¡vio MagalhÃ£es Institute (IOM) from the Ezequiel Dias Foundation (FUNED) | Instituto OctÃ¡vio MagalhÃ£es / FundaÃ§Ã£o Ezequiel Dias (IOM/Funed) |
| EPI_ISL_429686 | Central Public Health Laboratory/OctÃ¡vio MagalhÃ£es Institute (IOM) from the Ezequiel Dias Foundation (FUNED) | Instituto OctÃ¡vio MagalhÃ£es / FundaÃ§Ã£o Ezequiel Dias (IOM/Funed) |
| EPI_ISL_429687 | Central Public Health Laboratory/OctÃ¡vio MagalhÃ£es Institute (IOM) from the Ezequiel Dias Foundation (FUNED) | Instituto OctÃ¡vio MagalhÃ£es / FundaÃ§Ã£o Ezequiel Dias (IOM/Funed) |
| EPI_ISL_429688 | Central Public Health Laboratory/OctÃ¡vio MagalhÃ£es Institute (IOM) from the Ezequiel Dias Foundation (FUNED) | Instituto OctÃ¡vio MagalhÃ£es / FundaÃ§Ã£o Ezequiel Dias (IOM/Funed) |

| EPI_Accession_ID | Originating Laboratory | Submitting Laboratory |
|---|---|---|
| EPI_ISL_429689 | Central Public Health Laboratory/OctÃ¡vio MagalhÃ£es Institute (IOM) from the Ezequiel Dias Foundation (FUNED) | Instituto OctÃ¡vio MagalhÃ£es / FundaÃ§Ã£o Ezequiel Dias (IOM/Funed) |
| EPI_ISL_429691 | Central Public Health Laboratory/OctÃ¡vio MagalhÃ£es Institute (IOM) from the Ezequiel Dias Foundation (FUNED) | Instituto OctÃ¡vio MagalhÃ£es / FundaÃ§Ã£o Ezequiel Dias (IOM/Funed) |
| EPI_ISL_429695 | Central Public Health Laboratory/OctÃ¡vio MagalhÃ£es Institute (IOM) from the Ezequiel Dias Foundation (FUNED) | Instituto OctÃ¡vio MagalhÃ£es / FundaÃ§Ã£o Ezequiel Dias (IOM/Funed) |
| EPI_ISL_429696 | Central Public Health Laboratory/OctÃ¡vio MagalhÃ£es Institute (IOM) from the Ezequiel Dias Foundation (FUNED) | Instituto OctÃ¡vio MagalhÃ£es / FundaÃ§Ã£o Ezequiel Dias (IOM/Funed) |
| EPI_ISL_429701 | Central Public Health Laboratory/OctÃ¡vio MagalhÃ£es Institute (IOM) from the Ezequiel Dias Foundation (FUNED) | Instituto OctÃ¡vio MagalhÃ£es / FundaÃ§Ã£o Ezequiel Dias (IOM/Funed) |
| EPI_ISL_429702 | Central Public Health Laboratory/OctÃ¡vio MagalhÃ£es Institute (IOM) from the Ezequiel Dias Foundation (FUNED) | Instituto OctÃ¡vio MagalhÃ£es / FundaÃ§Ã£o Ezequiel Dias (IOM/Funed) |
| EPI_ISL_429706 | Laboratoire National de Sante, Microbiology, Virology | Laboratoire National de Sante, Microbiology, Epidemiology and Microbial Genomics |
| EPI_ISL_429707 | Laboratoire National de Sante, Microbiology, Virology | Laboratoire National de Sante, Microbiology, Epidemiology and Microbial Genomics |
| EPI_ISL_429708 | Laboratoire National de Sante, Microbiology, Virology | Laboratoire National de Sante, Microbiology, Epidemiology and Microbial Genomics |
| EPI_ISL_429709 | Laboratoire National de Sante, Microbiology, Virology | Laboratoire National de Sante, Microbiology, Epidemiology and Microbial Genomics |
| EPI_ISL_429710 | Laboratoire National de Sante, Microbiology, Virology | Laboratoire National de Sante, Microbiology, Epidemiology and Microbial Genomics |
| EPI_ISL_429711 | Laboratoire National de Sante, Microbiology, Virology | Laboratoire National de Sante, Microbiology, Epidemiology and Microbial Genomics |
| EPI_ISL_429712 | Laboratoire National de Sante, Microbiology, Virology | Laboratoire National de Sante, Microbiology, Epidemiology and Microbial Genomics |

| EPI_Accession_ID | Originating Laboratory | Submitting Laboratory |
| --- | --- | --- |
| EPI_ISL_429713 | Laboratoire National de Sante, Microbiology, Virology | Laboratoire National de Sante, Microbiology, Epidemiology and Microbial Genomics |
| EPI_ISL_429714 | Laboratoire National de Sante, Microbiology, Virology | Laboratoire National de Sante, Microbiology, Epidemiology and Microbial Genomics |
| EPI_ISL_429715 | Laboratoire National de Sante, Microbiology, Virology | Laboratoire National de Sante, Microbiology, Epidemiology and Microbial Genomics |
| EPI_ISL_429716 | Laboratoire National de Sante, Microbiology, Virology | Laboratoire National de Sante, Microbiology, Epidemiology and Microbial Genomics |
| EPI_ISL_429717 | Laboratoire National de Sante, Microbiology, Virology | Laboratoire National de Sante, Microbiology, Epidemiology and Microbial Genomics |
| EPI_ISL_429718 | Laboratoire National de Sante, Microbiology, Virology | Laboratoire National de Sante, Microbiology, Epidemiology and Microbial Genomics |
| EPI_ISL_429719 | Laboratoire National de Sante, Microbiology, Virology | Laboratoire National de Sante, Microbiology, Epidemiology and Microbial Genomics |
| EPI_ISL_429720 | Laboratoire National de Sante, Microbiology, Virology | Laboratoire National de Sante, Microbiology, Epidemiology and Microbial Genomics |
| EPI_ISL_429721 | Laboratoire National de Sante, Microbiology, Virology | Laboratoire National de Sante, Microbiology, Epidemiology and Microbial Genomics |
| EPI_ISL_429722 | Laboratoire National de Sante, Microbiology, Virology | Laboratoire National de Sante, Microbiology, Epidemiology and Microbial Genomics |
| EPI_ISL_429723 | Laboratoire National de Sante, Microbiology, Virology | Laboratoire National de Sante, Microbiology, Epidemiology and Microbial Genomics |
| EPI_ISL_429724 | Laboratoire National de Sante, Microbiology, Virology | Laboratoire National de Sante, Microbiology, Epidemiology and Microbial Genomics |
| EPI_ISL_429725 | Laboratoire National de Sante, Microbiology, Virology | Laboratoire National de Sante, Microbiology, Epidemiology and Microbial Genomics |
| EPI_ISL_429726 | Laboratoire National de Sante, Microbiology, Virology | Laboratoire National de Sante, Microbiology, Epidemiology and Microbial Genomics |
| EPI_ISL_429727 | Laboratoire National de Sante, Microbiology, Virology | Laboratoire National de Sante, Microbiology, Epidemiology and Microbial Genomics |
| EPI_ISL_429728 | Laboratoire National de Sante, Microbiology, Virology | Laboratoire National de Sante, Microbiology, Epidemiology and Microbial Genomics |

| EPI_Accession_ID | Originating Laboratory | Submitting Laboratory |
|---|---|---|
| EPI_ISL_429729 | Laboratoire National de Sante, Microbiology, Virology | Laboratoire National de Sante, Microbiology, Epidemiology and Microbial Genomics |
| EPI_ISL_429730 | Laboratoire National de Sante, Microbiology, Virology | Laboratoire National de Sante, Microbiology, Epidemiology and Microbial Genomics |
| EPI_ISL_429731 | Laboratoire National de Sante, Microbiology, Virology | Laboratoire National de Sante, Microbiology, Epidemiology and Microbial Genomics |
| EPI_ISL_429732 | Laboratoire National de Sante, Microbiology, Virology | Laboratoire National de Sante, Microbiology, Epidemiology and Microbial Genomics |
| EPI_ISL_429733 | Laboratoire National de Sante, Microbiology, Virology | Laboratoire National de Sante, Microbiology, Epidemiology and Microbial Genomics |
| EPI_ISL_429734 | Laboratoire National de Sante, Microbiology, Virology | Laboratoire National de Sante, Microbiology, Epidemiology and Microbial Genomics |
| EPI_ISL_429735 | Laboratoire National de Sante, Microbiology, Virology | Laboratoire National de Sante, Microbiology, Epidemiology and Microbial Genomics |
| EPI_ISL_429736 | Laboratoire National de Sante, Microbiology, Virology | Laboratoire National de Sante, Microbiology, Epidemiology and Microbial Genomics |
| EPI_ISL_429737 | Laboratoire National de Sante, Microbiology, Virology | Laboratoire National de Sante, Microbiology, Epidemiology and Microbial Genomics |
| EPI_ISL_429738 | Laboratoire National de Sante, Microbiology, Virology | Laboratoire National de Sante, Microbiology, Epidemiology and Microbial Genomics |
| EPI_ISL_429739 | Laboratoire National de Sante, Microbiology, Virology | Laboratoire National de Sante, Microbiology, Epidemiology and Microbial Genomics |
| EPI_ISL_429740 | Laboratoire National de Sante, Microbiology, Virology | Laboratoire National de Sante, Microbiology, Epidemiology and Microbial Genomics |
| EPI_ISL_429741 | Laboratoire National de Sante, Microbiology, Virology | Laboratoire National de Sante, Microbiology, Epidemiology and Microbial Genomics |
| EPI_ISL_429742 | Laboratoire National de Sante, Microbiology, Virology | Laboratoire National de Sante, Microbiology, Epidemiology and Microbial Genomics |
| EPI_ISL_429743 | Laboratoire National de Sante, Microbiology, Virology | Laboratoire National de Sante, Microbiology, Epidemiology and Microbial Genomics |
| EPI_ISL_429744 | Laboratoire National de Sante, Microbiology, Virology | Laboratoire National de Sante, Microbiology, Epidemiology and Microbial Genomics |

| EPI_Accession_ID | Originating Laboratory | Submitting Laboratory |
| --- | --- | --- |
| EPI_ISL_429745 | Laboratoire National de Sante, Microbiology, Virology | Laboratoire National de Sante, Microbiology, Epidemiology and Microbial Genomics |
| EPI_ISL_429746 | Laboratoire National de Sante, Microbiology, Virology | Laboratoire National de Sante, Microbiology, Epidemiology and Microbial Genomics |
| EPI_ISL_429747 | Laboratoire National de Sante, Microbiology, Virology | Laboratoire National de Sante, Microbiology, Epidemiology and Microbial Genomics |
| EPI_ISL_429748 | Laboratoire National de Sante, Microbiology, Virology | Laboratoire National de Sante, Microbiology, Epidemiology and Microbial Genomics |
| EPI_ISL_429749 | Laboratoire National de Sante, Microbiology, Virology | Laboratoire National de Sante, Microbiology, Epidemiology and Microbial Genomics |
| EPI_ISL_429750 | Laboratoire National de Sante, Microbiology, Virology | Laboratoire National de Sante, Microbiology, Epidemiology and Microbial Genomics |
| EPI_ISL_429751 | Laboratoire National de Sante, Microbiology, Virology | Laboratoire National de Sante, Microbiology, Epidemiology and Microbial Genomics |
| EPI_ISL_429752 | Laboratoire National de Sante, Microbiology, Virology | Laboratoire National de Sante, Microbiology, Epidemiology and Microbial Genomics |
| EPI_ISL_429753 | Laboratoire National de Sante, Microbiology, Virology | Laboratoire National de Sante, Microbiology, Epidemiology and Microbial Genomics |
| EPI_ISL_429754 | Laboratoire National de Sante, Microbiology, Virology | Laboratoire National de Sante, Microbiology, Epidemiology and Microbial Genomics |
| EPI_ISL_429755 | Laboratoire National de Sante, Microbiology, Virology | Laboratoire National de Sante, Microbiology, Epidemiology and Microbial Genomics |
| EPI_ISL_429756 | Laboratoire National de Sante, Microbiology, Virology | Laboratoire National de Sante, Microbiology, Epidemiology and Microbial Genomics |
| EPI_ISL_429757 | Laboratoire National de Sante, Microbiology, Virology | Laboratoire National de Sante, Microbiology, Epidemiology and Microbial Genomics |
| EPI_ISL_429758 | Laboratoire National de Sante, Microbiology, Virology | Laboratoire National de Sante, Microbiology, Epidemiology and Microbial Genomics |
| EPI_ISL_429759 | Laboratoire National de Sante, Microbiology, Virology | Laboratoire National de Sante, Microbiology, Epidemiology and Microbial Genomics |
| EPI_ISL_429760 | Laboratoire National de Sante, Microbiology, Virology | Laboratoire National de Sante, Microbiology, Epidemiology and Microbial Genomics |

| EPI_Accession_ID | Originating Laboratory | Submitting Laboratory |
|---|---|---|
| EPI_ISL_429761 | Laboratoire National de Sante, Microbiology, Virology | Laboratoire National de Sante, Microbiology, Epidemiology and Microbial Genomics |
| EPI_ISL_429762 | Laboratoire National de Sante, Microbiology, Virology | Laboratoire National de Sante, Microbiology, Epidemiology and Microbial Genomics |
| EPI_ISL_429763 | Laboratoire National de Sante, Microbiology, Virology | Laboratoire National de Sante, Microbiology, Epidemiology and Microbial Genomics |
| EPI_ISL_429764 | Laboratoire National de Sante, Microbiology, Virology | Laboratoire National de Sante, Microbiology, Epidemiology and Microbial Genomics |
| EPI_ISL_429765 | Laboratoire National de Sante, Microbiology, Virology | Laboratoire National de Sante, Microbiology, Epidemiology and Microbial Genomics |
| EPI_ISL_429766 | Laboratoire National de Sante, Microbiology, Virology | Laboratoire National de Sante, Microbiology, Epidemiology and Microbial Genomics |
| EPI_ISL_429767 | Laboratoire National de Sante, Microbiology, Virology | Laboratoire National de Sante, Microbiology, Epidemiology and Microbial Genomics |
| EPI_ISL_429768 | Laboratoire National de Sante, Microbiology, Virology | Laboratoire National de Sante, Microbiology, Epidemiology and Microbial Genomics |
| EPI_ISL_429769 | Laboratoire National de Sante, Microbiology, Virology | Laboratoire National de Sante, Microbiology, Epidemiology and Microbial Genomics |
| EPI_ISL_429770 | Laboratoire National de Sante, Microbiology, Virology | Laboratoire National de Sante, Microbiology, Epidemiology and Microbial Genomics |
| EPI_ISL_429771 | Laboratoire National de Sante, Microbiology, Virology | Laboratoire National de Sante, Microbiology, Epidemiology and Microbial Genomics |
| EPI_ISL_429772 | Laboratoire National de Sante, Microbiology, Virology | Laboratoire National de Sante, Microbiology, Epidemiology and Microbial Genomics |
| EPI_ISL_429773 | Laboratoire National de Sante, Microbiology, Virology | Laboratoire National de Sante, Microbiology, Epidemiology and Microbial Genomics |
| EPI_ISL_429774 | Laboratoire National de Sante, Microbiology, Virology | Laboratoire National de Sante, Microbiology, Epidemiology and Microbial Genomics |
| EPI_ISL_429775 | Laboratoire National de Sante, Microbiology, Virology | Laboratoire National de Sante, Microbiology, Epidemiology and Microbial Genomics |
| EPI_ISL_429776 | Laboratoire National de Sante, Microbiology, Virology | Laboratoire National de Sante, Microbiology, Epidemiology and Microbial Genomics |

| EPI_Accession_ID | Originating Laboratory | Submitting Laboratory |
|---|---|---|
| EPI_ISL_429777 | Laboratoire National de Sante, Microbiology, Virology | Laboratoire National de Sante, Microbiology, Epidemiology and Microbial Genomics |
| EPI_ISL_429778 | Laboratoire National de Sante, Microbiology, Virology | Laboratoire National de Sante, Microbiology, Epidemiology and Microbial Genomics |
| EPI_ISL_429779 | Laboratoire National de Sante, Microbiology, Virology | Laboratoire National de Sante, Microbiology, Epidemiology and Microbial Genomics |
| EPI_ISL_429780 | Laboratoire National de Sante, Microbiology, Virology | Laboratoire National de Sante, Microbiology, Epidemiology and Microbial Genomics |
| EPI_ISL_429781 | Laboratoire National de Sante, Microbiology, Virology | Laboratoire National de Sante, Microbiology, Epidemiology and Microbial Genomics |
| EPI_ISL_429782 | Laboratoire National de Sante, Microbiology, Virology | Laboratoire National de Sante, Microbiology, Epidemiology and Microbial Genomics |
| EPI_ISL_429783 | Laboratoire National de Sante, Microbiology, Virology | Laboratoire National de Sante, Microbiology, Epidemiology and Microbial Genomics |
| EPI_ISL_429784 | Laboratoire National de Sante, Microbiology, Virology | Laboratoire National de Sante, Microbiology, Epidemiology and Microbial Genomics |
| EPI_ISL_429785 | Laboratoire National de Sante, Microbiology, Virology | Laboratoire National de Sante, Microbiology, Epidemiology and Microbial Genomics |
| EPI_ISL_429786 | Laboratoire National de Sante, Microbiology, Virology | Laboratoire National de Sante, Microbiology, Epidemiology and Microbial Genomics |
| EPI_ISL_429787 | Laboratoire National de Sante, Microbiology, Virology | Laboratoire National de Sante, Microbiology, Epidemiology and Microbial Genomics |
| EPI_ISL_429788 | Laboratoire National de Sante, Microbiology, Virology | Laboratoire National de Sante, Microbiology, Epidemiology and Microbial Genomics |
| EPI_ISL_429789 | Laboratoire National de Sante, Microbiology, Virology | Laboratoire National de Sante, Microbiology, Epidemiology and Microbial Genomics |
| EPI_ISL_429790 | Laboratoire National de Sante, Microbiology, Virology | Laboratoire National de Sante, Microbiology, Epidemiology and Microbial Genomics |
| EPI_ISL_429791 | Laboratoire National de Sante, Microbiology, Virology | Laboratoire National de Sante, Microbiology, Epidemiology and Microbial Genomics |
| EPI_ISL_429793 | Laboratoire National de Sante, Microbiology, Virology | Laboratoire National de Sante, Microbiology, Epidemiology and Microbial Genomics |

| EPI_Accession_ID | Originating Laboratory | Submitting Laboratory |
|---|---|---|
| EPI_ISL_429794 | Laboratoire National de Sante, Microbiology, Virology | Laboratoire National de Sante, Microbiology, Epidemiology and Microbial Genomics |
| EPI_ISL_429795 | Laboratoire National de Sante, Microbiology, Virology | Laboratoire National de Sante, Microbiology, Epidemiology and Microbial Genomics |
| EPI_ISL_429796 | Laboratoire National de Sante, Microbiology, Virology | Laboratoire National de Sante, Microbiology, Epidemiology and Microbial Genomics |
| EPI_ISL_429797 | Laboratoire National de Sante, Microbiology, Virology | Laboratoire National de Sante, Microbiology, Epidemiology and Microbial Genomics |
| EPI_ISL_429798 | Laboratoire National de Sante, Microbiology, Virology | Laboratoire National de Sante, Microbiology, Epidemiology and Microbial Genomics |
| EPI_ISL_429799 | Laboratoire National de Sante, Microbiology, Virology | Laboratoire National de Sante, Microbiology, Epidemiology and Microbial Genomics |
| EPI_ISL_429800 | Laboratoire National de Sante, Microbiology, Virology | Laboratoire National de Sante, Microbiology, Epidemiology and Microbial Genomics |
| EPI_ISL_429806 | Dr. Georges-L.-Dumont University Hospital Centre | National Microbiology Laboratory |
| EPI_ISL_429807 | Cadham Provincial Laboratory | National Microbiology Laboratory |
| EPI_ISL_429811 | Dr. Georges-L.-Dumont University Hospital Centre | National Microbiology Laboratory |
| EPI_ISL_429812 | Queen Elizabeth II Health Science Centre | National Microbiology Laboratory |
| EPI_ISL_429813 | Queen Elizabeth II Health Science Centre | National Microbiology Laboratory |
| EPI_ISL_429814 | Queen Elizabeth II Health Science Centre | National Microbiology Laboratory |
| EPI_ISL_429815 | Public Health Laboratory | National Microbiology Laboratory |
| EPI_ISL_429816 | Dr. Georges-L.-Dumont University Hospital Centre | National Microbiology Laboratory |
| EPI_ISL_429817 | Cadham Provincial Laboratory | National Microbiology Laboratory |
| EPI_ISL_429818 | Cadham Provincial Laboratory | National Microbiology Laboratory |
| EPI_ISL_429819 | Cadham Provincial Laboratory | National Microbiology Laboratory |
| EPI_ISL_429820 | Cadham Provincial Laboratory | National Microbiology Laboratory |
| EPI_ISL_429843 | Gundersen Molecular Diagnostics Laboratory | Kabara Cancer Research Institute |
| EPI_ISL_429844 | Gundersen Molecular Diagnostics Laboratory | Kabara Cancer Research Institute |
| EPI_ISL_429845 | Gundersen Molecular Diagnostics Laboratory | Kabara Cancer Research Institute |
| EPI_ISL_429846 | Gundersen Molecular Diagnostics Laboratory | Kabara Cancer Research Institute |
| EPI_ISL_429847 | Gundersen Molecular Diagnostics Laboratory | Kabara Cancer Research Institute |
| EPI_ISL_429848 | Gundersen Molecular Diagnostics Laboratory | Kabara Cancer Research Institute |

| EPI_Accession_ID | Originating Laboratory | Submitting Laboratory |
|---|---|---|
| EPI_ISL_429852 | Centers for Disease Control and Prevention of Lishui | Department of Inspectionï¼ŒCenters for Disease Control and Prevention of Lishui |
| EPI_ISL_429853 | Centers for Disease Control and Prevention of Lishui | Department of Inspectionï¼ŒCenters for Disease Control and Prevention of Lishui |
| EPI_ISL_429854 | Centers for Disease Control and Prevention of Lishui | Department of Inspectionï¼ŒCenters for Disease Control and Prevention of Lishui |
| EPI_ISL_429855 | Centers for Disease Control and Prevention of Lishui | Department of Inspectionï¼ŒCenters for Disease Control and Prevention of Lishui |
| EPI_ISL_429861 | Ministry of Health Turkey | Ministry of Health Turkey |
| EPI_ISL_429862 | Ministry of Health Turkey | Ministry of Health Turkey |
| EPI_ISL_429863 | Ministry of Health Turkey | Ministry of Health Turkey |
| EPI_ISL_429864 | Ministry of Health Turkey | Ministry of Health Turkey |
| EPI_ISL_429865 | Ministry of Health Turkey | Ministry of Health Turkey |
| EPI_ISL_429866 | Ministry of Health Turkey | Ministry of Health Turkey |
| EPI_ISL_429867 | Ministry of Health Turkey | Ministry of Health Turkey |
| EPI_ISL_429868 | Ministry of Health Turkey | Ministry of Health Turkey |
| EPI_ISL_429869 | Ministry of Health Turkey | Ministry of Health Turkey |
| EPI_ISL_429870 | Ministry of Health Turkey | Ministry of Health Turkey |
| EPI_ISL_429871 | Ministry of Health Turkey | Ministry of Health Turkey |
| EPI_ISL_429872 | Ministry of Health Turkey | Ministry of Health Turkey |
| EPI_ISL_429873 | Ministry of Health Turkey | Ministry of Health Turkey |
| EPI_ISL_429874 | Microbiology, Virology and Biemergency Laboratory-ASST FBF Sacco | Microbiology, Virology and Biemergency Laboratory-ASST FBF Sacco |
| EPI_ISL_429875 | California Department of Public Health | Chiu Laboratory, University of California, San Francisco |
| EPI_ISL_429877 | Chiu Laboratory, University of California, San Francisco | Chiu Laboratory, University of California, San Francisco |
| EPI_ISL_429878 | Chiu Laboratory, University of California, San Francisco | Chiu Laboratory, University of California, San Francisco |
| EPI_ISL_429879 | Santa Clara County Public Health Department | Chiu Laboratory, University of California, San Francisco |
| EPI_ISL_429880 | Santa Clara County Public Health Department | Chiu Laboratory, University of California, San Francisco |
| EPI_ISL_429881 | California Department of Public Health | Chiu Laboratory, University of California, San Francisco |
| EPI_ISL_429882 | Centers for Disease Control, R.O.C. (Taiwan) | Centers for Disease Control, R.O.C. (Taiwan) |
| EPI_ISL_429883 | Centers for Disease Control, R.O.C. (Taiwan) | Centers for Disease Control, R.O.C. (Taiwan) |
| EPI_ISL_429884 | Centers for Disease Control, R.O.C. (Taiwan) | Centers for Disease Control, R.O.C. (Taiwan) |

| EPI_Accession_ID | Originating Laboratory | Submitting Laboratory |
|---|---|---|
| EPI_ISL_429968 | Centre Hospitalier CompiÃ¨gne Laboratoire de Biologie | National Reference Center for Viruses of Respiratory Infections, Institut Pasteur, Paris |
| EPI_ISL_429969 | Virginia DCLS | Virginia DCLS |
| EPI_ISL_429970 | Virginia DCLS | Virginia DCLS |
| EPI_ISL_429971 | Virginia DCLS | Virginia DCLS |
| EPI_ISL_429972 | Virginia DCLS | Virginia DCLS |
| EPI_ISL_429973 | Virginia DCLS | Virginia DCLS |
| EPI_ISL_429974 | Virginia DCLS | Virginia DCLS |
| EPI_ISL_429975 | Virginia DCLS | Virginia DCLS |
| EPI_ISL_429976 | Virginia DCLS | Virginia DCLS |
| EPI_ISL_429977 | Virginia DCLS | Virginia DCLS |
| EPI_ISL_429978 | Virginia DCLS | Virginia DCLS |
| EPI_ISL_429979 | Virginia DCLS | Virginia DCLS |
| EPI_ISL_429980 | Virginia DCLS | Virginia DCLS |
| EPI_ISL_429981 | Virginia DCLS | Virginia DCLS |
| EPI_ISL_429982 | Virginia DCLS | Virginia DCLS |
| EPI_ISL_429983 | Virginia DCLS | Virginia DCLS |
| EPI_ISL_429984 | Virginia DCLS | Virginia DCLS |
| EPI_ISL_429985 | Virginia DCLS | Virginia DCLS |
| EPI_ISL_429986 | Virginia DCLS | Virginia DCLS |
| EPI_ISL_429987 | Virginia DCLS | Virginia DCLS |
| EPI_ISL_429988 | Virginia DCLS | Virginia DCLS |
| EPI_ISL_429989 | Virginia DCLS | Virginia DCLS |
| EPI_ISL_429990 | Rady's Childrens Hospital | Andersen lab at Scripps Research |
| EPI_ISL_429991 | Andersen lab at Scripps Research | Andersen lab at Scripps Research |
| EPI_ISL_429992 | Biolab Diagnostic Laboratories | Andersen lab at Scripps Research |
| EPI_ISL_429993 | Biolab Diagnostic Laboratories | Andersen lab at Scripps Research |
| EPI_ISL_429994 | Biolab Diagnostic Laboratories | Andersen lab at Scripps Research |
| EPI_ISL_429995 | Biolab Diagnostic Laboratories | Andersen lab at Scripps Research |
| EPI_ISL_429996 | Biolab Diagnostic Laboratories | Andersen lab at Scripps Research |
| EPI_ISL_429997 | Biolab Diagnostic Laboratories | Andersen lab at Scripps Research |
| EPI_ISL_429998 | Biolab Diagnostic Laboratories | Andersen lab at Scripps Research |
| EPI_ISL_429999 | Biolab Diagnostic Laboratories | Andersen lab at Scripps Research |

| EPI_Accession_ID | Originating Laboratory | Submitting Laboratory |
|---|---|---|
| EPI_ISL_430000 | Biolab Diagnostic Laboratories | Andersen lab at Scripps Research |
| EPI_ISL_430001 | Biolab Diagnostic Laboratories | Andersen lab at Scripps Research |
| EPI_ISL_430002 | Biolab Diagnostic Laboratories | Andersen lab at Scripps Research |
| EPI_ISL_430003 | Biolab Diagnostic Laboratories | Andersen lab at Scripps Research |
| EPI_ISL_430004 | Biolab Diagnostic Laboratories | Andersen lab at Scripps Research |
| EPI_ISL_430005 | Biolab Diagnostic Laboratories | Andersen lab at Scripps Research |
| EPI_ISL_430006 | Biolab Diagnostic Laboratories | Andersen lab at Scripps Research |
| EPI_ISL_430007 | Biolab Diagnostic Laboratories | Andersen lab at Scripps Research |
| EPI_ISL_430008 | Biolab Diagnostic Laboratories | Andersen lab at Scripps Research |
| EPI_ISL_430009 | Biolab Diagnostic Laboratories | Andersen lab at Scripps Research |
| EPI_ISL_430010 | OSU Wexner Medical Center | James Molecular Lab - OSUWMC |
| EPI_ISL_430011 | Biolab Diagnostic Laboratories | Andersen lab at Scripps Research |
| EPI_ISL_430012 | Biolab Diagnostic Laboratories | Andersen lab at Scripps Research |
| EPI_ISL_430013 | Biolab Diagnostic Laboratories | Andersen lab at Scripps Research |
| EPI_ISL_430014 | Biolab Diagnostic Laboratories | Andersen lab at Scripps Research |
| EPI_ISL_430015 | Biolab Diagnostic Laboratories | Andersen lab at Scripps Research |
| EPI_ISL_430016 | Andersen lab at Scripps Research | Andersen lab at Scripps Research |
| EPI_ISL_430018 | Molecular Pathology Division, Department of Pathology, Hong Kong Sanatorium & Hospital | Molecular Pathology Division, Department of Pathology, Hong Kong Sanatorium & Hospital |
| EPI_ISL_430019 | Utah Public Health Laboratory | Utah Public Health Laboratory |
| EPI_ISL_430020 | Utah Public Health Laboratory | Utah Public Health Laboratory |
| EPI_ISL_430021 | Utah Public Health Laboratory | Utah Public Health Laboratory |
| EPI_ISL_430022 | Utah Public Health Laboratory | Utah Public Health Laboratory |
| EPI_ISL_430023 | Utah Public Health Laboratory | Utah Public Health Laboratory |
| EPI_ISL_430024 | Utah Public Health Laboratory | Utah Public Health Laboratory |
| EPI_ISL_430025 | Utah Public Health Laboratory | Utah Public Health Laboratory |
| EPI_ISL_430026 | Utah Public Health Laboratory | Utah Public Health Laboratory |
| EPI_ISL_430027 | Utah Public Health Laboratory | Utah Public Health Laboratory |
| EPI_ISL_430028 | Utah Public Health Laboratory | Utah Public Health Laboratory |
| EPI_ISL_430029 | Utah Public Health Laboratory | Utah Public Health Laboratory |
| EPI_ISL_430030 | Utah Public Health Laboratory | Utah Public Health Laboratory |
| EPI_ISL_430031 | Utah Public Health Laboratory | Utah Public Health Laboratory |
| EPI_ISL_430032 | Utah Public Health Laboratory | Utah Public Health Laboratory |

| EPI_Accession_ID | Originating Laboratory | Submitting Laboratory |
|---|---|---|
| EPI_ISL_430033 | Utah Public Health Laboratory | Utah Public Health Laboratory |
| EPI_ISL_430034 | Utah Public Health Laboratory | Utah Public Health Laboratory |
| EPI_ISL_430035 | Utah Public Health Laboratory | Utah Public Health Laboratory |
| EPI_ISL_430036 | Utah Public Health Laboratory | Utah Public Health Laboratory |
| EPI_ISL_430037 | Utah Public Health Laboratory | Utah Public Health Laboratory |
| EPI_ISL_430038 | Utah Public Health Laboratory | Utah Public Health Laboratory |
| EPI_ISL_430039 | Utah Public Health Laboratory | Utah Public Health Laboratory |
| EPI_ISL_430040 | Utah Public Health Laboratory | Utah Public Health Laboratory |
| EPI_ISL_430041 | Utah Public Health Laboratory | Utah Public Health Laboratory |
| EPI_ISL_430042 | Utah Public Health Laboratory | Utah Public Health Laboratory |
| EPI_ISL_430043 | Utah Public Health Laboratory | Utah Public Health Laboratory |
| EPI_ISL_430044 | Utah Public Health Laboratory | Utah Public Health Laboratory |
| EPI_ISL_430045 | Utah Public Health Laboratory | Utah Public Health Laboratory |
| EPI_ISL_430046 | Utah Public Health Laboratory | Utah Public Health Laboratory |
| EPI_ISL_430047 | Utah Public Health Laboratory | Utah Public Health Laboratory |
| EPI_ISL_430048 | Utah Public Health Laboratory | Utah Public Health Laboratory |
| EPI_ISL_430049 | Utah Public Health Laboratory | Utah Public Health Laboratory |
| EPI_ISL_430050 | Utah Public Health Laboratory | Utah Public Health Laboratory |
| EPI_ISL_430051 | Utah Public Health Laboratory | Utah Public Health Laboratory |
| EPI_ISL_430052 | Utah Public Health Laboratory | Utah Public Health Laboratory |
| EPI_ISL_430053 | Utah Public Health Laboratory | Utah Public Health Laboratory |
| EPI_ISL_430054 | Utah Public Health Laboratory | Utah Public Health Laboratory |
| EPI_ISL_430055 | Utah Public Health Laboratory | Utah Public Health Laboratory |
| EPI_ISL_430056 | Utah Public Health Laboratory | Utah Public Health Laboratory |
| EPI_ISL_430057 | Utah Public Health Laboratory | Utah Public Health Laboratory |
| EPI_ISL_430058 | Utah Public Health Laboratory | Utah Public Health Laboratory |
| EPI_ISL_430059 | Utah Public Health Laboratory | Utah Public Health Laboratory |
| EPI_ISL_430060 | Utah Public Health Laboratory | Utah Public Health Laboratory |
| EPI_ISL_430061 | Utah Public Health Laboratory | Utah Public Health Laboratory |
| EPI_ISL_430062 | Utah Public Health Laboratory | Utah Public Health Laboratory |
| EPI_ISL_430063 | Molecular Pathology Division, Department of Pathology, Hong Kong Sanatorium & Hospital | Molecular Pathology Division, Department of Pathology, Hong Kong Sanatorium & Hospital |
| EPI_ISL_430064 | Geelong Centre for Emerging Infectious Diseases | Geelong Centre for Emerging Infectious Diseases |

| EPI_Accession_ID | Originating Laboratory | Submitting Laboratory |
|---|---|---|
| EPI_ISL_430065 | Geelong Centre for Emerging Infectious Diseases | Geelong Centre for Emerging Infectious Diseases |
| EPI_ISL_430066 | Geelong Centre for Emerging Infectious Diseases | Geelong Centre for Emerging Infectious Diseases |
| EPI_ISL_430067 | WHO National Influenza Centre Russian Federation | WHO National Influenza Centre Russian Federation |
| EPI_ISL_430068 | WHO National Influenza Centre Russian Federation | WHO National Influenza Centre Russian Federation |
| EPI_ISL_430069 | WHO National Influenza Centre Russian Federation | WHO National Influenza Centre Russian Federation |
| EPI_ISL_430070 | WHO National Influenza Centre Russian Federation | WHO National Influenza Centre Russian Federation |
| EPI_ISL_430071 | WHO National Influenza Centre Russian Federation | WHO National Influenza Centre Russian Federation |
| EPI_ISL_430072 | WHO National Influenza Centre Russian Federation | WHO National Influenza Centre Russian Federation |
| EPI_ISL_430073 | WHO National Influenza Centre Russian Federation | WHO National Influenza Centre Russian Federation |
| EPI_ISL_430074 | WHO National Influenza Centre Russian Federation | WHO National Influenza Centre Russian Federation |
| EPI_ISL_430075 | WHO National Influenza Centre Russian Federation | WHO National Influenza Centre Russian Federation |
| EPI_ISL_430076 | WHO National Influenza Centre Russian Federation | WHO National Influenza Centre Russian Federation |
| EPI_ISL_430077 | WHO National Influenza Centre Russian Federation | WHO National Influenza Centre Russian Federation |
| EPI_ISL_430078 | WHO National Influenza Centre Russian Federation | WHO National Influenza Centre Russian Federation |
| EPI_ISL_430079 | WHO National Influenza Centre Russian Federation | WHO National Influenza Centre Russian Federation |
| EPI_ISL_430080 | WHO National Influenza Centre Russian Federation | WHO National Influenza Centre Russian Federation |
| EPI_ISL_430081 | WHO National Influenza Centre Russian Federation | WHO National Influenza Centre Russian Federation |
| EPI_ISL_430082 | WHO National Influenza Centre Russian Federation | WHO National Influenza Centre Russian Federation |
| EPI_ISL_430083 | WHO National Influenza Centre Russian Federation | WHO National Influenza Centre Russian Federation |
| EPI_ISL_430084 | WHO National Influenza Centre Russian Federation | WHO National Influenza Centre Russian Federation |
| EPI_ISL_430085 | WHO National Influenza Centre Russian Federation | WHO National Influenza Centre Russian Federation |
| EPI_ISL_430086 | WHO National Influenza Centre Russian Federation | WHO National Influenza Centre Russian Federation |
| EPI_ISL_430087 | WHO National Influenza Centre Russian Federation | WHO National Influenza Centre Russian Federation |
| EPI_ISL_430088 | WHO National Influenza Centre Russian Federation | WHO National Influenza Centre Russian Federation |
| EPI_ISL_430089 | WHO National Influenza Centre Russian Federation | WHO National Influenza Centre Russian Federation |
| EPI_ISL_430090 | WHO National Influenza Centre Russian Federation | WHO National Influenza Centre Russian Federation |
| EPI_ISL_430091 | WHO National Influenza Centre Russian Federation | WHO National Influenza Centre Russian Federation |
| EPI_ISL_430092 | WHO National Influenza Centre Russian Federation | WHO National Influenza Centre Russian Federation |
| EPI_ISL_430093 | WHO National Influenza Centre Russian Federation | WHO National Influenza Centre Russian Federation |
| EPI_ISL_430094 | WHO National Influenza Centre Russian Federation | WHO National Influenza Centre Russian Federation |
| EPI_ISL_430095 | WHO National Influenza Centre Russian Federation | WHO National Influenza Centre Russian Federation |
| EPI_ISL_430096 | WHO National Influenza Centre Russian Federation | WHO National Influenza Centre Russian Federation |
| EPI_ISL_430097 | WHO National Influenza Centre Russian Federation | WHO National Influenza Centre Russian Federation |

| EPI_Accession_ID | Originating Laboratory | Submitting Laboratory |
|---|---|---|
| EPI_ISL_430098 | WHO National Influenza Centre Russian Federation | WHO National Influenza Centre Russian Federation |
| EPI_ISL_430099 | WHO National Influenza Centre Russian Federation | WHO National Influenza Centre Russian Federation |
| EPI_ISL_430100 | WHO National Influenza Centre Russian Federation | WHO National Influenza Centre Russian Federation |
| EPI_ISL_430101 | WHO National Influenza Centre Russian Federation | WHO National Influenza Centre Russian Federation |
| EPI_ISL_430102 | WHO National Influenza Centre Russian Federation | WHO National Influenza Centre Russian Federation |
| EPI_ISL_430103 | WHO National Influenza Centre Russian Federation | WHO National Influenza Centre Russian Federation |
| EPI_ISL_430104 | WHO National Influenza Centre Russian Federation | WHO National Influenza Centre Russian Federation |
| EPI_ISL_430105 | WHO National Influenza Centre Russian Federation | WHO National Influenza Centre Russian Federation |
| EPI_ISL_430106 | WHO National Influenza Centre Russian Federation | WHO National Influenza Centre Russian Federation |
| EPI_ISL_430107 | WHO National Influenza Centre Russian Federation | WHO National Influenza Centre Russian Federation |
| EPI_ISL_430108 | WHO National Influenza Centre Russian Federation | WHO National Influenza Centre Russian Federation |
| EPI_ISL_430109 | WHO National Influenza Centre Russian Federation | WHO National Influenza Centre Russian Federation |
| EPI_ISL_430110 | WHO National Influenza Centre Russian Federation | WHO National Influenza Centre Russian Federation |
| EPI_ISL_430111 | WHO National Influenza Centre Russian Federation | WHO National Influenza Centre Russian Federation |
| EPI_ISL_430112 | WHO National Influenza Centre Russian Federation | WHO National Influenza Centre Russian Federation |
| EPI_ISL_430113 | Seattle Flu Study | Seattle Flu Study |
| EPI_ISL_430114 | Seattle Flu Study | Seattle Flu Study |
| EPI_ISL_430115 | Seattle Flu Study | Seattle Flu Study |
| EPI_ISL_430116 | Seattle Flu Study | Seattle Flu Study |
| EPI_ISL_430117 | Seattle Flu Study | Seattle Flu Study |
| EPI_ISL_430118 | Seattle Flu Study | Seattle Flu Study |
| EPI_ISL_430119 | Seattle Flu Study | Seattle Flu Study |
| EPI_ISL_430120 | Seattle Flu Study | Seattle Flu Study |
| EPI_ISL_430121 | Seattle Flu Study | Seattle Flu Study |
| EPI_ISL_430122 | Seattle Flu Study | Seattle Flu Study |
| EPI_ISL_430123 | Seattle Flu Study | Seattle Flu Study |
| EPI_ISL_430124 | Seattle Flu Study | Seattle Flu Study |
| EPI_ISL_430125 | Seattle Flu Study | Seattle Flu Study |
| EPI_ISL_430126 | Seattle Flu Study | Seattle Flu Study |
| EPI_ISL_430127 | Seattle Flu Study | Seattle Flu Study |
| EPI_ISL_430128 | Seattle Flu Study | Seattle Flu Study |
| EPI_ISL_430129 | Seattle Flu Study | Seattle Flu Study |
| EPI_ISL_430130 | Seattle Flu Study | Seattle Flu Study |

| EPI_Accession_ID | Originating Laboratory | Submitting Laboratory |
|---|---|---|
| EPI_ISL_430131 | Seattle Flu Study | Seattle Flu Study |
| EPI_ISL_430132 | Seattle Flu Study | Seattle Flu Study |
| EPI_ISL_430133 | Seattle Flu Study | Seattle Flu Study |
| EPI_ISL_430134 | Seattle Flu Study | Seattle Flu Study |
| EPI_ISL_430135 | Seattle Flu Study | Seattle Flu Study |
| EPI_ISL_430136 | Seattle Flu Study | Seattle Flu Study |
| EPI_ISL_430137 | Seattle Flu Study | Seattle Flu Study |
| EPI_ISL_430138 | Seattle Flu Study | Seattle Flu Study |
| EPI_ISL_430139 | Seattle Flu Study | Seattle Flu Study |
| EPI_ISL_430140 | Seattle Flu Study | Seattle Flu Study |
| EPI_ISL_430141 | Seattle Flu Study | Seattle Flu Study |
| EPI_ISL_430142 | Seattle Flu Study | Seattle Flu Study |
| EPI_ISL_430143 | Seattle Flu Study | Seattle Flu Study |
| EPI_ISL_430144 | Seattle Flu Study | Seattle Flu Study |
| EPI_ISL_430145 | Seattle Flu Study | Seattle Flu Study |
| EPI_ISL_430146 | Seattle Flu Study | Seattle Flu Study |
| EPI_ISL_430147 | Seattle Flu Study | Seattle Flu Study |
| EPI_ISL_430148 | Seattle Flu Study | Seattle Flu Study |
| EPI_ISL_430149 | Seattle Flu Study | Seattle Flu Study |
| EPI_ISL_430150 | Seattle Flu Study | Seattle Flu Study |
| EPI_ISL_430151 | Seattle Flu Study | Seattle Flu Study |
| EPI_ISL_430152 | Seattle Flu Study | Seattle Flu Study |
| EPI_ISL_430153 | Seattle Flu Study | Seattle Flu Study |
| EPI_ISL_430154 | Seattle Flu Study | Seattle Flu Study |
| EPI_ISL_430155 | Seattle Flu Study | Seattle Flu Study |
| EPI_ISL_430156 | Seattle Flu Study | Seattle Flu Study |
| EPI_ISL_430160 | Washington State Department of Health | Seattle Flu Study |
| EPI_ISL_430161 | Washington State Department of Health | Seattle Flu Study |
| EPI_ISL_430162 | Washington State Department of Health | Seattle Flu Study |
| EPI_ISL_430163 | Washington State Department of Health | Seattle Flu Study |
| EPI_ISL_430164 | Washington State Department of Health | Seattle Flu Study |
| EPI_ISL_430165 | Washington State Department of Health | Seattle Flu Study |
| EPI_ISL_430166 | Washington State Department of Health | Seattle Flu Study |

| EPI_Accession_ID | Originating Laboratory | Submitting Laboratory |
|---|---|---|
| EPI_ISL_430167 | Washington State Department of Health | Seattle Flu Study |
| EPI_ISL_430168 | Washington State Department of Health | Seattle Flu Study |
| EPI_ISL_430169 | Washington State Department of Health | Seattle Flu Study |
| EPI_ISL_430170 | Washington State Department of Health | Seattle Flu Study |
| EPI_ISL_430171 | Washington State Department of Health | Seattle Flu Study |
| EPI_ISL_430172 | Washington State Department of Health | Seattle Flu Study |
| EPI_ISL_430173 | Washington State Department of Health | Seattle Flu Study |
| EPI_ISL_430174 | Washington State Department of Health | Seattle Flu Study |
| EPI_ISL_430175 | Washington State Department of Health | Seattle Flu Study |
| EPI_ISL_430176 | Washington State Department of Health | Seattle Flu Study |
| EPI_ISL_430177 | Washington State Department of Health | Seattle Flu Study |
| EPI_ISL_430178 | Washington State Department of Health | Seattle Flu Study |
| EPI_ISL_430179 | Washington State Department of Health | Seattle Flu Study |
| EPI_ISL_430180 | Washington State Department of Health | Seattle Flu Study |
| EPI_ISL_430181 | Washington State Department of Health | Seattle Flu Study |
| EPI_ISL_430182 | Washington State Department of Health | Seattle Flu Study |
| EPI_ISL_430183 | Washington State Department of Health | Seattle Flu Study |
| EPI_ISL_430184 | Washington State Department of Health | Seattle Flu Study |
| EPI_ISL_430185 | Washington State Department of Health | Seattle Flu Study |
| EPI_ISL_430186 | Washington State Department of Health | Seattle Flu Study |
| EPI_ISL_430187 | Washington State Department of Health | Seattle Flu Study |
| EPI_ISL_430188 | Washington State Department of Health | Seattle Flu Study |
| EPI_ISL_430189 | Washington State Department of Health | Seattle Flu Study |
| EPI_ISL_430190 | Washington State Department of Health | Seattle Flu Study |
| EPI_ISL_430191 | Washington State Department of Health | Seattle Flu Study |
| EPI_ISL_430192 | Washington State Department of Health | Seattle Flu Study |
| EPI_ISL_430193 | Washington State Department of Health | Seattle Flu Study |
| EPI_ISL_430194 | Washington State Department of Health | Seattle Flu Study |
| EPI_ISL_430196 | Washington State Department of Health | Seattle Flu Study |
| EPI_ISL_430197 | Washington State Department of Health | Seattle Flu Study |
| EPI_ISL_430198 | Washington State Department of Health | Seattle Flu Study |
| EPI_ISL_430199 | Washington State Department of Health | Seattle Flu Study |
| EPI_ISL_430200 | Washington State Department of Health | Seattle Flu Study |

| EPI_Accession_ID | Originating Laboratory | Submitting Laboratory |
| --- | --- | --- |
| EPI_ISL_430201 | Washington State Department of Health | Seattle Flu Study |
| EPI_ISL_430202 | Washington State Department of Health | Seattle Flu Study |
| EPI_ISL_430203 | Washington State Department of Health | Seattle Flu Study |
| EPI_ISL_430204 | Washington State Department of Health | Seattle Flu Study |
| EPI_ISL_430205 | Washington State Department of Health | Seattle Flu Study |
| EPI_ISL_430206 | Washington State Department of Health | Seattle Flu Study |
| EPI_ISL_430207 | Washington State Department of Health | Seattle Flu Study |
| EPI_ISL_430208 | Washington State Department of Health | Seattle Flu Study |
| EPI_ISL_430209 | Washington State Department of Health | Seattle Flu Study |
| EPI_ISL_430210 | Washington State Department of Health | Seattle Flu Study |
| EPI_ISL_430211 | Washington State Department of Health | Seattle Flu Study |
| EPI_ISL_430212 | Washington State Department of Health | Seattle Flu Study |
| EPI_ISL_430213 | Washington State Department of Health | Seattle Flu Study |
| EPI_ISL_430214 | Washington State Department of Health | Seattle Flu Study |
| EPI_ISL_430215 | Washington State Department of Health | Seattle Flu Study |
| EPI_ISL_430216 | Washington State Department of Health | Seattle Flu Study |
| EPI_ISL_430217 | Washington State Department of Health | Seattle Flu Study |
| EPI_ISL_430218 | Washington State Department of Health | Seattle Flu Study |
| EPI_ISL_430219 | Washington State Department of Health | Seattle Flu Study |
| EPI_ISL_430220 | Washington State Department of Health | Seattle Flu Study |
| EPI_ISL_430221 | Washington State Department of Health | Seattle Flu Study |
| EPI_ISL_430222 | Washington State Department of Health | Seattle Flu Study |
| EPI_ISL_430223 | Washington State Department of Health | Seattle Flu Study |
| EPI_ISL_430224 | Washington State Department of Health | Seattle Flu Study |
| EPI_ISL_430225 | Washington State Department of Health | Seattle Flu Study |
| EPI_ISL_430226 | Washington State Department of Health | Seattle Flu Study |
| EPI_ISL_430227 | Washington State Department of Health | Seattle Flu Study |
| EPI_ISL_430228 | Washington State Department of Health | Seattle Flu Study |
| EPI_ISL_430229 | Washington State Department of Health | Seattle Flu Study |
| EPI_ISL_430230 | Washington State Department of Health | Seattle Flu Study |
| EPI_ISL_430231 | Washington State Department of Health | Seattle Flu Study |
| EPI_ISL_430232 | Washington State Department of Health | Seattle Flu Study |
| EPI_ISL_430233 | Washington State Department of Health | Seattle Flu Study |

| EPI_Accession_ID | Originating Laboratory | Submitting Laboratory |
|---|---|---|
| EPI_ISL_430234 | Washington State Department of Health | Seattle Flu Study |
| EPI_ISL_430235 | Washington State Department of Health | Seattle Flu Study |
| EPI_ISL_430236 | Washington State Department of Health | Seattle Flu Study |
| EPI_ISL_430237 | Washington State Department of Health | Seattle Flu Study |
| EPI_ISL_430238 | Washington State Department of Health | Seattle Flu Study |
| EPI_ISL_430239 | Washington State Department of Health | Seattle Flu Study |
| EPI_ISL_430240 | Washington State Department of Health | Seattle Flu Study |
| EPI_ISL_430241 | Washington State Department of Health | Seattle Flu Study |
| EPI_ISL_430242 | Washington State Department of Health | Seattle Flu Study |
| EPI_ISL_430243 | Washington State Department of Health | Seattle Flu Study |
| EPI_ISL_430244 | Washington State Department of Health | Seattle Flu Study |
| EPI_ISL_430245 | Washington State Department of Health | Seattle Flu Study |
| EPI_ISL_430246 | Washington State Department of Health | Seattle Flu Study |
| EPI_ISL_430247 | Washington State Department of Health | Seattle Flu Study |
| EPI_ISL_430248 | Washington State Department of Health | Seattle Flu Study |
| EPI_ISL_430249 | Washington State Department of Health | Seattle Flu Study |
| EPI_ISL_430250 | Washington State Department of Health | Seattle Flu Study |
| EPI_ISL_430251 | Washington State Department of Health | Seattle Flu Study |
| EPI_ISL_430252 | Washington State Department of Health | Seattle Flu Study |
| EPI_ISL_430253 | Washington State Department of Health | Seattle Flu Study |
| EPI_ISL_430254 | Washington State Department of Health | Seattle Flu Study |
| EPI_ISL_430255 | Washington State Department of Health | Seattle Flu Study |
| EPI_ISL_430256 | Washington State Department of Health | Seattle Flu Study |
| EPI_ISL_430257 | Washington State Department of Health | Seattle Flu Study |
| EPI_ISL_430258 | Washington State Department of Health | Seattle Flu Study |
| EPI_ISL_430259 | Washington State Department of Health | Seattle Flu Study |
| EPI_ISL_430260 | Washington State Department of Health | Seattle Flu Study |
| EPI_ISL_430261 | Washington State Department of Health | Seattle Flu Study |
| EPI_ISL_430262 | Washington State Department of Health | Seattle Flu Study |
| EPI_ISL_430263 | Washington State Department of Health | Seattle Flu Study |
| EPI_ISL_430264 | Washington State Department of Health | Seattle Flu Study |
| EPI_ISL_430265 | Washington State Department of Health | Seattle Flu Study |
| EPI_ISL_430266 | Washington State Department of Health | Seattle Flu Study |

| EPI_Accession_ID | Originating Laboratory | Submitting Laboratory |
|---|---|---|
| EPI_ISL_430267 | Washington State Department of Health | Seattle Flu Study |
| EPI_ISL_430268 | Washington State Department of Health | Seattle Flu Study |
| EPI_ISL_430269 | Washington State Department of Health | Seattle Flu Study |
| EPI_ISL_430270 | Washington State Department of Health | Seattle Flu Study |
| EPI_ISL_430271 | Washington State Department of Health | Seattle Flu Study |
| EPI_ISL_430272 | Washington State Department of Health | Seattle Flu Study |
| EPI_ISL_430273 | Washington State Department of Health | Seattle Flu Study |
| EPI_ISL_430274 | Washington State Department of Health | Seattle Flu Study |
| EPI_ISL_430275 | Washington State Department of Health | Seattle Flu Study |
| EPI_ISL_430276 | Washington State Department of Health | Seattle Flu Study |
| EPI_ISL_430277 | Washington State Department of Health | Seattle Flu Study |
| EPI_ISL_430278 | Washington State Department of Health | Seattle Flu Study |
| EPI_ISL_430279 | Washington State Department of Health | Seattle Flu Study |
| EPI_ISL_430280 | Washington State Department of Health | Seattle Flu Study |
| EPI_ISL_430281 | Washington State Department of Health | Seattle Flu Study |
| EPI_ISL_430282 | Washington State Department of Health | Seattle Flu Study |
| EPI_ISL_430283 | Washington State Department of Health | Seattle Flu Study |
| EPI_ISL_430284 | Washington State Department of Health | Seattle Flu Study |
| EPI_ISL_430285 | Washington State Department of Health | Seattle Flu Study |
| EPI_ISL_430286 | Washington State Department of Health | Seattle Flu Study |
| EPI_ISL_430287 | Washington State Department of Health | Seattle Flu Study |
| EPI_ISL_430288 | Washington State Department of Health | Seattle Flu Study |
| EPI_ISL_430289 | Washington State Department of Health | Seattle Flu Study |
| EPI_ISL_430290 | Washington State Department of Health | Seattle Flu Study |
| EPI_ISL_430291 | Washington State Department of Health | Seattle Flu Study |
| EPI_ISL_430292 | Washington State Department of Health | Seattle Flu Study |
| EPI_ISL_430293 | Washington State Department of Health | Seattle Flu Study |
| EPI_ISL_430294 | Washington State Department of Health | Seattle Flu Study |
| EPI_ISL_430295 | Washington State Department of Health | Seattle Flu Study |
| EPI_ISL_430296 | Washington State Department of Health | Seattle Flu Study |
| EPI_ISL_430297 | National Institute for Communicable Diseases of the National Health Laboratory Service | National Institute for Communicable Diseases of the National Health Laboratory Service |

| EPI_Accession_ID | Originating Laboratory | Submitting Laboratory |
|---|---|---|
| EPI_ISL_430319 | NYU Langone Health | Departments of Pathology and Medicine, New York University School of Medicine |
| EPI_ISL_430322 | NYU Langone Health | Departments of Pathology and Medicine, New York University School of Medicine |
| EPI_ISL_430323 | NYU Langone Health | Departments of Pathology and Medicine, New York University School of Medicine |
| EPI_ISL_430324 | NYU Langone Health | Departments of Pathology and Medicine, New York University School of Medicine |
| EPI_ISL_430325 | NYU Langone Health | Departments of Pathology and Medicine, New York University School of Medicine |
| EPI_ISL_430326 | NYU Langone Health | Departments of Pathology and Medicine, New York University School of Medicine |
| EPI_ISL_430327 | NYU Langone Health | Departments of Pathology and Medicine, New York University School of Medicine |
| EPI_ISL_430328 | NYU Langone Health | Departments of Pathology and Medicine, New York University School of Medicine |
| EPI_ISL_430329 | NYU Langone Health | Departments of Pathology and Medicine, New York University School of Medicine |
| EPI_ISL_430330 | NYU Langone Health | Departments of Pathology and Medicine, New York University School of Medicine |
| EPI_ISL_430331 | NYU Langone Health | Departments of Pathology and Medicine, New York University School of Medicine |
| EPI_ISL_430332 | NYU Langone Health | Departments of Pathology and Medicine, New York University School of Medicine |
| EPI_ISL_430333 | NYU Langone Health | Departments of Pathology and Medicine, New York University School of Medicine |
| EPI_ISL_430334 | NYU Langone Health | Departments of Pathology and Medicine, New York University School of Medicine |
| EPI_ISL_430335 | NYU Langone Health | Departments of Pathology and Medicine, New York University School of Medicine |
| EPI_ISL_430336 | NYU Langone Health | Departments of Pathology and Medicine, New York University School of Medicine |

| EPI_Accession_ID | Originating Laboratory | Submitting Laboratory |
|---|---|---|
| EPI_ISL_430337 | NYU Langone Health | Departments of Pathology and Medicine, New York University School of Medicine |
| EPI_ISL_430338 | NYU Langone Health | Departments of Pathology and Medicine, New York University School of Medicine |
| EPI_ISL_430339 | NYU Langone Health | Departments of Pathology and Medicine, New York University School of Medicine |
| EPI_ISL_430340 | NYU Langone Health | Departments of Pathology and Medicine, New York University School of Medicine |
| EPI_ISL_430341 | NYU Langone Health | Departments of Pathology and Medicine, New York University School of Medicine |
| EPI_ISL_430342 | NYU Langone Health | Departments of Pathology and Medicine, New York University School of Medicine |
| EPI_ISL_430343 | NYU Langone Health | Departments of Pathology and Medicine, New York University School of Medicine |
| EPI_ISL_430344 | NYU Langone Health | Departments of Pathology and Medicine, New York University School of Medicine |
| EPI_ISL_430345 | NYU Langone Health | Departments of Pathology and Medicine, New York University School of Medicine |
| EPI_ISL_430346 | NYU Langone Health | Departments of Pathology and Medicine, New York University School of Medicine |
| EPI_ISL_430347 | NYU Langone Health | Departments of Pathology and Medicine, New York University School of Medicine |
| EPI_ISL_430348 | NYU Langone Health | Departments of Pathology and Medicine, New York University School of Medicine |
| EPI_ISL_430349 | NYU Langone Health | Departments of Pathology and Medicine, New York University School of Medicine |
| EPI_ISL_430350 | NYU Langone Health | Departments of Pathology and Medicine, New York University School of Medicine |
| EPI_ISL_430351 | NYU Langone Health | Departments of Pathology and Medicine, New York University School of Medicine |
| EPI_ISL_430352 | NYU Langone Health | Departments of Pathology and Medicine, New York University School of Medicine |

| EPI_Accession_ID | Originating Laboratory | Submitting Laboratory |
|---|---|---|
| EPI_ISL_430353 | NYU Langone Health | Departments of Pathology and Medicine, New York University School of Medicine |
| EPI_ISL_430354 | NYU Langone Health | Departments of Pathology and Medicine, New York University School of Medicine |
| EPI_ISL_430355 | NYU Langone Health | Departments of Pathology and Medicine, New York University School of Medicine |
| EPI_ISL_430356 | NYU Langone Health | Departments of Pathology and Medicine, New York University School of Medicine |
| EPI_ISL_430357 | NYU Langone Health | Departments of Pathology and Medicine, New York University School of Medicine |
| EPI_ISL_430358 | NYU Langone Health | Departments of Pathology and Medicine, New York University School of Medicine |
| EPI_ISL_430359 | NYU Langone Health | Departments of Pathology and Medicine, New York University School of Medicine |
| EPI_ISL_430360 | NYU Langone Health | Departments of Pathology and Medicine, New York University School of Medicine |
| EPI_ISL_430361 | NYU Langone Health | Departments of Pathology and Medicine, New York University School of Medicine |
| EPI_ISL_430362 | NYU Langone Health | Departments of Pathology and Medicine, New York University School of Medicine |
| EPI_ISL_430363 | NYU Langone Health | Departments of Pathology and Medicine, New York University School of Medicine |
| EPI_ISL_430364 | NYU Langone Health | Departments of Pathology and Medicine, New York University School of Medicine |
| EPI_ISL_430365 | NYU Langone Health | Departments of Pathology and Medicine, New York University School of Medicine |
| EPI_ISL_430366 | NYU Langone Health | Departments of Pathology and Medicine, New York University School of Medicine |
| EPI_ISL_430367 | NYU Langone Health | Departments of Pathology and Medicine, New York University School of Medicine |
| EPI_ISL_430368 | NYU Langone Health | Departments of Pathology and Medicine, New York University School of Medicine |

| EPI_Accession_ID | Originating Laboratory | Submitting Laboratory |
|---|---|---|
| EPI_ISL_430369 | NYU Langone Health | Departments of Pathology and Medicine, New York University School of Medicine |
| EPI_ISL_430370 | NYU Langone Health | Departments of Pathology and Medicine, New York University School of Medicine |
| EPI_ISL_430371 | NYU Langone Health | Departments of Pathology and Medicine, New York University School of Medicine |
| EPI_ISL_430372 | NYU Langone Health | Departments of Pathology and Medicine, New York University School of Medicine |
| EPI_ISL_430373 | NYU Langone Health | Departments of Pathology and Medicine, New York University School of Medicine |
| EPI_ISL_430374 | NYU Langone Health | Departments of Pathology and Medicine, New York University School of Medicine |
| EPI_ISL_430375 | NYU Langone Health | Departments of Pathology and Medicine, New York University School of Medicine |
| EPI_ISL_430376 | NYU Langone Health | Departments of Pathology and Medicine, New York University School of Medicine |
| EPI_ISL_430377 | NYU Langone Health | Departments of Pathology and Medicine, New York University School of Medicine |
| EPI_ISL_430378 | NYU Langone Health | Departments of Pathology and Medicine, New York University School of Medicine |
| EPI_ISL_430379 | NYU Langone Health | Departments of Pathology and Medicine, New York University School of Medicine |
| EPI_ISL_430380 | NYU Langone Health | Departments of Pathology and Medicine, New York University School of Medicine |
| EPI_ISL_430381 | NYU Langone Health | Departments of Pathology and Medicine, New York University School of Medicine |
| EPI_ISL_430382 | NYU Langone Health | Departments of Pathology and Medicine, New York University School of Medicine |
| EPI_ISL_430383 | NYU Langone Health | Departments of Pathology and Medicine, New York University School of Medicine |
| EPI_ISL_430384 | NYU Langone Health | Departments of Pathology and Medicine, New York University School of Medicine |

| EPI_Accession_ID | Originating Laboratory | Submitting Laboratory |
|---|---|---|
| EPI_ISL_430385 | NYU Langone Health | Departments of Pathology and Medicine, New York University School of Medicine |
| EPI_ISL_430386 | NYU Langone Health | Departments of Pathology and Medicine, New York University School of Medicine |
| EPI_ISL_430387 | NYU Langone Health | Departments of Pathology and Medicine, New York University School of Medicine |
| EPI_ISL_430388 | NYU Langone Health | Departments of Pathology and Medicine, New York University School of Medicine |
| EPI_ISL_430389 | NYU Langone Health | Departments of Pathology and Medicine, New York University School of Medicine |
| EPI_ISL_430390 | NYU Langone Health | Departments of Pathology and Medicine, New York University School of Medicine |
| EPI_ISL_430391 | NYU Langone Health | Departments of Pathology and Medicine, New York University School of Medicine |
| EPI_ISL_430392 | NYU Langone Health | Departments of Pathology and Medicine, New York University School of Medicine |
| EPI_ISL_430393 | NYU Langone Health | Departments of Pathology and Medicine, New York University School of Medicine |
| EPI_ISL_430394 | NYU Langone Health | Departments of Pathology and Medicine, New York University School of Medicine |
| EPI_ISL_430395 | NYU Langone Health | Departments of Pathology and Medicine, New York University School of Medicine |
| EPI_ISL_430396 | NYU Langone Health | Departments of Pathology and Medicine, New York University School of Medicine |
| EPI_ISL_430397 | NYU Langone Health | Departments of Pathology and Medicine, New York University School of Medicine |
| EPI_ISL_430398 | NYU Langone Health | Departments of Pathology and Medicine, New York University School of Medicine |
| EPI_ISL_430399 | NYU Langone Health | Departments of Pathology and Medicine, New York University School of Medicine |
| EPI_ISL_430400 | NYU Langone Health | Departments of Pathology and Medicine, New York University School of Medicine |

| EPI_Accession_ID | Originating Laboratory | Submitting Laboratory |
|---|---|---|
| EPI_ISL_430401 | NYU Langone Health | Departments of Pathology and Medicine, New York University School of Medicine |
| EPI_ISL_430402 | NYU Langone Health | Departments of Pathology and Medicine, New York University School of Medicine |
| EPI_ISL_430403 | NYU Langone Health | Departments of Pathology and Medicine, New York University School of Medicine |
| EPI_ISL_430404 | NYU Langone Health | Departments of Pathology and Medicine, New York University School of Medicine |
| EPI_ISL_430405 | NYU Langone Health | Departments of Pathology and Medicine, New York University School of Medicine |
| EPI_ISL_430406 | NYU Langone Health | Departments of Pathology and Medicine, New York University School of Medicine |
| EPI_ISL_430407 | NYU Langone Health | Departments of Pathology and Medicine, New York University School of Medicine |
| EPI_ISL_430408 | NYU Langone Health | Departments of Pathology and Medicine, New York University School of Medicine |
| EPI_ISL_430409 | NYU Langone Health | Departments of Pathology and Medicine, New York University School of Medicine |
| EPI_ISL_430410 | NYU Langone Health | Departments of Pathology and Medicine, New York University School of Medicine |
| EPI_ISL_430411 | NYU Langone Health | Departments of Pathology and Medicine, New York University School of Medicine |
| EPI_ISL_430412 | NYU Langone Health | Departments of Pathology and Medicine, New York University School of Medicine |
| EPI_ISL_430413 | NYU Langone Health | Departments of Pathology and Medicine, New York University School of Medicine |
| EPI_ISL_430414 | NYU Langone Health | Departments of Pathology and Medicine, New York University School of Medicine |
| EPI_ISL_430415 | NYU Langone Health | Departments of Pathology and Medicine, New York University School of Medicine |
| EPI_ISL_430416 | NYU Langone Health | Departments of Pathology and Medicine, New York University School of Medicine |

| EPI_Accession_ID | Originating Laboratory | Submitting Laboratory |
|---|---|---|
| EPI_ISL_430417 | NYU Langone Health | Departments of Pathology and Medicine, New York University School of Medicine |
| EPI_ISL_430418 | NYU Langone Health | Departments of Pathology and Medicine, New York University School of Medicine |
| EPI_ISL_430419 | NYU Langone Health | Departments of Pathology and Medicine, New York University School of Medicine |
| EPI_ISL_430420 | NYU Langone Health | Departments of Pathology and Medicine, New York University School of Medicine |
| EPI_ISL_430421 | NYU Langone Health | Departments of Pathology and Medicine, New York University School of Medicine |
| EPI_ISL_430422 | NYU Langone Health | Departments of Pathology and Medicine, New York University School of Medicine |
| EPI_ISL_430423 | NYU Langone Health | Departments of Pathology and Medicine, New York University School of Medicine |
| EPI_ISL_430424 | NYU Langone Health | Departments of Pathology and Medicine, New York University School of Medicine |
| EPI_ISL_430425 | NYU Langone Health | Departments of Pathology and Medicine, New York University School of Medicine |
| EPI_ISL_430426 | NYU Langone Health | Departments of Pathology and Medicine, New York University School of Medicine |
| EPI_ISL_430427 | NYU Langone Health | Departments of Pathology and Medicine, New York University School of Medicine |
| EPI_ISL_430428 | NYU Langone Health | Departments of Pathology and Medicine, New York University School of Medicine |
| EPI_ISL_430429 | NYU Langone Health | Departments of Pathology and Medicine, New York University School of Medicine |
| EPI_ISL_430430 | NYU Langone Health | Departments of Pathology and Medicine, New York University School of Medicine |
| EPI_ISL_430431 | NYU Langone Health | Departments of Pathology and Medicine, New York University School of Medicine |
| EPI_ISL_430432 | NYU Langone Health | Departments of Pathology and Medicine, New York University School of Medicine |

| EPI_Accession_ID | Originating Laboratory | Submitting Laboratory |
|---|---|---|
| EPI_ISL_430433 | NYU Langone Health | Departments of Pathology and Medicine, New York University School of Medicine |
| EPI_ISL_430434 | NYU Langone Health | Departments of Pathology and Medicine, New York University School of Medicine |
| EPI_ISL_430439 | Institute for Medical Research, Infectious Disease Research Centre, National Institutes of Health, Ministry of Health Malaysia | Institute for Medical Research Infectious Disease Research Centre, National Institutes of Health, Ministry of Health Malaysia |
| EPI_ISL_430440 | Institute for Medical Research, Infectious Disease Research Centre, National Institutes of Health, Ministry of Health Malaysia | Institute for Medical Research, Infectious Disease Research Centre, National Institutes of Health, Ministry of Health Malaysia |
| EPI_ISL_430441 | Institute for Medical Research, Infectious Disease Research Centre, National Institutes of Health, Ministry of Health Malaysia | Institute for Medical Research, Infectious Disease Research Centre, National Institutes of Health, Ministry of Health Malaysia |
| EPI_ISL_430442 | Institute for Medical Research, Infectious Disease Research Centre, National Institutes of Health, Ministry of Health Malaysia | Institute for Medical Research, Infectious Disease Research Centre, National Institutes of Health, Ministry of Health Malaysia |
| EPI_ISL_430443 | Institute for Medical Research, Infectious Disease Research Centre, National Institutes of Health, Ministry of Health Malaysia | Institute for Medical Research, Infectious Disease Research Centre, National Institutes of Health, Ministry of Health Malaysia |
| EPI_ISL_430444 | Institute for Medical Research, Infectious Disease Research Centre, National Institutes of Health, Ministry of Health Malaysia | Institute for Medical Research, Infectious Disease Research Centre, National Institutes of Health, Ministry of Health Malaysia |
| EPI_ISL_430456 | Rizal Medical Center | Research Institute for Tropical Medicine |
| EPI_ISL_430464 | ICMR-National Institute of Cholera and Enteric Diseases | National Institute of Biomedical Genomics |
| EPI_ISL_430465 | ICMR-National Institute of Cholera and Enteric Diseases | National Institute of Biomedical Genomics |
| EPI_ISL_430466 | ICMR-National Institute of Cholera and Enteric Diseases | National Institute of Biomedical Genomics |
| EPI_ISL_430467 | ICMR-National Institute of Cholera and Enteric Diseases | National Institute of Biomedical Genomics |
| EPI_ISL_430468 | ICMR-National Institute of Cholera and Enteric Diseases | National Institute of Biomedical Genomics |

| EPI_Accession_ID | Originating Laboratory | Submitting Laboratory |
|---|---|---|
| EPI_ISL_430469 | Hellenic Pasteur Institute, Public Health Laboratories | Hellenic Pasteur Institute, Public Health Laboratories, Unit of Bioinformatics and Applied Genomics |
| EPI_ISL_430470 | Microbiological Diagnostic Unit Public Health Laboratory | Microbiological Diagnostic Unit Public Health Laboratory |
| EPI_ISL_430471 | Microbiological Diagnostic Unit Public Health Laboratory | Microbiological Diagnostic Unit Public Health Laboratory |
| EPI_ISL_430472 | Microbiological Diagnostic Unit Public Health Laboratory | Microbiological Diagnostic Unit Public Health Laboratory |
| EPI_ISL_430473 | Victorian Infectious Diseases Reference Laboratory (VIDRL) | Microbiological Diagnostic Unit Public Health Laboratory and Victorian Infectious Diseases Reference Laboratory, The Peter Doherty Institute for Infection and Immunity |
| EPI_ISL_430474 | Victorian Infectious Diseases Reference Laboratory (VIDRL) | Microbiological Diagnostic Unit Public Health Laboratory and Victorian Infectious Diseases Reference Laboratory, The Peter Doherty Institute for Infection and Immunity |
| EPI_ISL_430475 | Victorian Infectious Diseases Reference Laboratory (VIDRL) | Microbiological Diagnostic Unit Public Health Laboratory and Victorian Infectious Diseases Reference Laboratory, The Peter Doherty Institute for Infection and Immunity |
| EPI_ISL_430476 | Victorian Infectious Diseases Reference Laboratory (VIDRL) | Microbiological Diagnostic Unit Public Health Laboratory and Victorian Infectious Diseases Reference Laboratory, The Peter Doherty Institute for Infection and Immunity |
| EPI_ISL_430477 | Victorian Infectious Diseases Reference Laboratory (VIDRL) | Microbiological Diagnostic Unit Public Health Laboratory and Victorian Infectious Diseases Reference Laboratory, The Peter Doherty Institute for Infection and Immunity |
| EPI_ISL_430478 | Victorian Infectious Diseases Reference Laboratory (VIDRL) | Microbiological Diagnostic Unit Public Health Laboratory and Victorian Infectious Diseases Reference Laboratory, The Peter Doherty Institute for Infection and Immunity |

| EPI_Accession_ID | Originating Laboratory | Submitting Laboratory |
|---|---|---|
| EPI_ISL_430479 | Victorian Infectious Diseases Reference Laboratory (VIDRL) | Microbiological Diagnostic Unit Public Health Laboratory and Victorian Infectious Diseases Reference Laboratory, The Peter Doherty Institute for Infection and Immunity |
| EPI_ISL_430480 | Victorian Infectious Diseases Reference Laboratory (VIDRL) | Microbiological Diagnostic Unit Public Health Laboratory and Victorian Infectious Diseases Reference Laboratory, The Peter Doherty Institute for Infection and Immunity |
| EPI_ISL_430481 | Victorian Infectious Diseases Reference Laboratory (VIDRL) | Microbiological Diagnostic Unit Public Health Laboratory and Victorian Infectious Diseases Reference Laboratory, The Peter Doherty Institute for Infection and Immunity |
| EPI_ISL_430482 | Victorian Infectious Diseases Reference Laboratory (VIDRL) | Microbiological Diagnostic Unit Public Health Laboratory and Victorian Infectious Diseases Reference Laboratory, The Peter Doherty Institute for Infection and Immunity |
| EPI_ISL_430483 | Victorian Infectious Diseases Reference Laboratory (VIDRL) | Microbiological Diagnostic Unit Public Health Laboratory and Victorian Infectious Diseases Reference Laboratory, The Peter Doherty Institute for Infection and Immunity |
| EPI_ISL_430484 | Victorian Infectious Diseases Reference Laboratory (VIDRL) | Microbiological Diagnostic Unit Public Health Laboratory and Victorian Infectious Diseases Reference Laboratory, The Peter Doherty Institute for Infection and Immunity |
| EPI_ISL_430485 | Victorian Infectious Diseases Reference Laboratory (VIDRL) | Microbiological Diagnostic Unit Public Health Laboratory and Victorian Infectious Diseases Reference Laboratory, The Peter Doherty Institute for Infection and Immunity |
| EPI_ISL_430486 | Victorian Infectious Diseases Reference Laboratory (VIDRL) | Microbiological Diagnostic Unit Public Health Laboratory and Victorian Infectious Diseases Reference Laboratory, The Peter Doherty Institute for Infection and Immunity |

| EPI_Accession_ID | Originating Laboratory | Submitting Laboratory |
|---|---|---|
| EPI_ISL_430487 | Victorian Infectious Diseases Reference Laboratory (VIDRL) | Microbiological Diagnostic Unit Public Health Laboratory and Victorian Infectious Diseases Reference Laboratory, The Peter Doherty Institute for Infection and Immunity |
| EPI_ISL_430488 | Victorian Infectious Diseases Reference Laboratory (VIDRL) | Microbiological Diagnostic Unit Public Health Laboratory and Victorian Infectious Diseases Reference Laboratory, The Peter Doherty Institute for Infection and Immunity |
| EPI_ISL_430489 | Victorian Infectious Diseases Reference Laboratory (VIDRL) | Microbiological Diagnostic Unit Public Health Laboratory and Victorian Infectious Diseases Reference Laboratory, The Peter Doherty Institute for Infection and Immunity |
| EPI_ISL_430490 | Victorian Infectious Diseases Reference Laboratory (VIDRL) | Microbiological Diagnostic Unit Public Health Laboratory and Victorian Infectious Diseases Reference Laboratory, The Peter Doherty Institute for Infection and Immunity |
| EPI_ISL_430491 | Victorian Infectious Diseases Reference Laboratory (VIDRL) | Microbiological Diagnostic Unit Public Health Laboratory and Victorian Infectious Diseases Reference Laboratory, The Peter Doherty Institute for Infection and Immunity |
| EPI_ISL_430492 | Victorian Infectious Diseases Reference Laboratory (VIDRL) | Microbiological Diagnostic Unit Public Health Laboratory and Victorian Infectious Diseases Reference Laboratory, The Peter Doherty Institute for Infection and Immunity |
| EPI_ISL_430493 | Victorian Infectious Diseases Reference Laboratory (VIDRL) | Microbiological Diagnostic Unit Public Health Laboratory and Victorian Infectious Diseases Reference Laboratory, The Peter Doherty Institute for Infection and Immunity |
| EPI_ISL_430498 | Victorian Infectious Diseases Reference Laboratory (VIDRL) | Microbiological Diagnostic Unit Public Health Laboratory and Victorian Infectious Diseases Reference Laboratory, The Peter Doherty Institute for Infection and Immunity |

| EPI_Accession_ID | Originating Laboratory | Submitting Laboratory |
|---|---|---|
| EPI_ISL_430499 | Victorian Infectious Diseases Reference Laboratory (VIDRL) | Microbiological Diagnostic Unit Public Health Laboratory and Victorian Infectious Diseases Reference Laboratory, The Peter Doherty Institute for Infection and Immunity |
| EPI_ISL_430500 | Victorian Infectious Diseases Reference Laboratory (VIDRL) | Microbiological Diagnostic Unit Public Health Laboratory and Victorian Infectious Diseases Reference Laboratory, The Peter Doherty Institute for Infection and Immunity |
| EPI_ISL_430501 | Victorian Infectious Diseases Reference Laboratory (VIDRL) | Microbiological Diagnostic Unit Public Health Laboratory and Victorian Infectious Diseases Reference Laboratory, The Peter Doherty Institute for Infection and Immunity |
| EPI_ISL_430502 | Victorian Infectious Diseases Reference Laboratory (VIDRL) | Microbiological Diagnostic Unit Public Health Laboratory and Victorian Infectious Diseases Reference Laboratory, The Peter Doherty Institute for Infection and Immunity |
| EPI_ISL_430503 | Victorian Infectious Diseases Reference Laboratory (VIDRL) | Microbiological Diagnostic Unit Public Health Laboratory and Victorian Infectious Diseases Reference Laboratory, The Peter Doherty Institute for Infection and Immunity |
| EPI_ISL_430504 | Victorian Infectious Diseases Reference Laboratory (VIDRL) | Microbiological Diagnostic Unit Public Health Laboratory and Victorian Infectious Diseases Reference Laboratory, The Peter Doherty Institute for Infection and Immunity |
| EPI_ISL_430505 | Victorian Infectious Diseases Reference Laboratory (VIDRL) | Microbiological Diagnostic Unit Public Health Laboratory and Victorian Infectious Diseases Reference Laboratory, The Peter Doherty Institute for Infection and Immunity |
| EPI_ISL_430506 | Victorian Infectious Diseases Reference Laboratory (VIDRL) | Microbiological Diagnostic Unit Public Health Laboratory and Victorian Infectious Diseases Reference Laboratory, The Peter Doherty Institute for Infection and Immunity |

| EPI_Accession_ID | Originating Laboratory | Submitting Laboratory |
|---|---|---|
| EPI_ISL_430507 | Victorian Infectious Diseases Reference Laboratory (VIDRL) | Microbiological Diagnostic Unit Public Health Laboratory and Victorian Infectious Diseases Reference Laboratory, The Peter Doherty Institute for Infection and Immunity |
| EPI_ISL_430508 | Victorian Infectious Diseases Reference Laboratory (VIDRL) | Microbiological Diagnostic Unit Public Health Laboratory and Victorian Infectious Diseases Reference Laboratory, The Peter Doherty Institute for Infection and Immunity |
| EPI_ISL_430509 | Victorian Infectious Diseases Reference Laboratory (VIDRL) | Microbiological Diagnostic Unit Public Health Laboratory and Victorian Infectious Diseases Reference Laboratory, The Peter Doherty Institute for Infection and Immunity |
| EPI_ISL_430510 | Victorian Infectious Diseases Reference Laboratory (VIDRL) | Microbiological Diagnostic Unit Public Health Laboratory and Victorian Infectious Diseases Reference Laboratory, The Peter Doherty Institute for Infection and Immunity |
| EPI_ISL_430511 | Victorian Infectious Diseases Reference Laboratory (VIDRL) | Microbiological Diagnostic Unit Public Health Laboratory and Victorian Infectious Diseases Reference Laboratory, The Peter Doherty Institute for Infection and Immunity |
| EPI_ISL_430512 | Victorian Infectious Diseases Reference Laboratory (VIDRL) | Microbiological Diagnostic Unit Public Health Laboratory and Victorian Infectious Diseases Reference Laboratory, The Peter Doherty Institute for Infection and Immunity |
| EPI_ISL_430513 | Victorian Infectious Diseases Reference Laboratory (VIDRL) | Microbiological Diagnostic Unit Public Health Laboratory and Victorian Infectious Diseases Reference Laboratory, The Peter Doherty Institute for Infection and Immunity |
| EPI_ISL_430514 | Victorian Infectious Diseases Reference Laboratory (VIDRL) | Microbiological Diagnostic Unit Public Health Laboratory and Victorian Infectious Diseases Reference Laboratory, The Peter Doherty Institute for Infection and Immunity |

| EPI_Accession_ID | Originating Laboratory | Submitting Laboratory |
|---|---|---|
| EPI_ISL_430515 | Victorian Infectious Diseases Reference Laboratory (VIDRL) | Microbiological Diagnostic Unit Public Health Laboratory and Victorian Infectious Diseases Reference Laboratory, The Peter Doherty Institute for Infection and Immunity |
| EPI_ISL_430516 | Victorian Infectious Diseases Reference Laboratory (VIDRL) | Microbiological Diagnostic Unit Public Health Laboratory and Victorian Infectious Diseases Reference Laboratory, The Peter Doherty Institute for Infection and Immunity |
| EPI_ISL_430517 | Victorian Infectious Diseases Reference Laboratory (VIDRL) | Microbiological Diagnostic Unit Public Health Laboratory and Victorian Infectious Diseases Reference Laboratory, The Peter Doherty Institute for Infection and Immunity |
| EPI_ISL_430518 | Victorian Infectious Diseases Reference Laboratory (VIDRL) | Microbiological Diagnostic Unit Public Health Laboratory and Victorian Infectious Diseases Reference Laboratory, The Peter Doherty Institute for Infection and Immunity |
| EPI_ISL_430519 | Victorian Infectious Diseases Reference Laboratory (VIDRL) | Microbiological Diagnostic Unit Public Health Laboratory and Victorian Infectious Diseases Reference Laboratory, The Peter Doherty Institute for Infection and Immunity |
| EPI_ISL_430520 | Victorian Infectious Diseases Reference Laboratory (VIDRL) | Microbiological Diagnostic Unit Public Health Laboratory and Victorian Infectious Diseases Reference Laboratory, The Peter Doherty Institute for Infection and Immunity |
| EPI_ISL_430521 | Victorian Infectious Diseases Reference Laboratory (VIDRL) | Microbiological Diagnostic Unit Public Health Laboratory and Victorian Infectious Diseases Reference Laboratory, The Peter Doherty Institute for Infection and Immunity |
| EPI_ISL_430522 | Victorian Infectious Diseases Reference Laboratory (VIDRL) | Microbiological Diagnostic Unit Public Health Laboratory and Victorian Infectious Diseases Reference Laboratory, The Peter Doherty Institute for Infection and Immunity |

| EPI_Accession_ID | Originating Laboratory | Submitting Laboratory |
| --- | --- | --- |
| EPI_ISL_430523 | Victorian Infectious Diseases Reference Laboratory (VIDRL) | Microbiological Diagnostic Unit Public Health Laboratory and Victorian Infectious Diseases Reference Laboratory, The Peter Doherty Institute for Infection and Immunity |
| EPI_ISL_430524 | Victorian Infectious Diseases Reference Laboratory (VIDRL) | Microbiological Diagnostic Unit Public Health Laboratory and Victorian Infectious Diseases Reference Laboratory, The Peter Doherty Institute for Infection and Immunity |
| EPI_ISL_430525 | Victorian Infectious Diseases Reference Laboratory (VIDRL) | Microbiological Diagnostic Unit Public Health Laboratory and Victorian Infectious Diseases Reference Laboratory, The Peter Doherty Institute for Infection and Immunity |
| EPI_ISL_430526 | Victorian Infectious Diseases Reference Laboratory (VIDRL) | Microbiological Diagnostic Unit Public Health Laboratory and Victorian Infectious Diseases Reference Laboratory, The Peter Doherty Institute for Infection and Immunity |
| EPI_ISL_430527 | Victorian Infectious Diseases Reference Laboratory (VIDRL) | Microbiological Diagnostic Unit Public Health Laboratory and Victorian Infectious Diseases Reference Laboratory, The Peter Doherty Institute for Infection and Immunity |
| EPI_ISL_430528 | Victorian Infectious Diseases Reference Laboratory (VIDRL) | Microbiological Diagnostic Unit Public Health Laboratory and Victorian Infectious Diseases Reference Laboratory, The Peter Doherty Institute for Infection and Immunity |
| EPI_ISL_430529 | Victorian Infectious Diseases Reference Laboratory (VIDRL) | Microbiological Diagnostic Unit Public Health Laboratory and Victorian Infectious Diseases Reference Laboratory, The Peter Doherty Institute for Infection and Immunity |
| EPI_ISL_430530 | Victorian Infectious Diseases Reference Laboratory (VIDRL) | Microbiological Diagnostic Unit Public Health Laboratory and Victorian Infectious Diseases Reference Laboratory, The Peter Doherty Institute for Infection and Immunity |

| EPI_Accession_ID | Originating Laboratory | Submitting Laboratory |
|---|---|---|
| EPI_ISL_430531 | Victorian Infectious Diseases Reference Laboratory (VIDRL) | Microbiological Diagnostic Unit Public Health Laboratory and Victorian Infectious Diseases Reference Laboratory, The Peter Doherty Institute for Infection and Immunity |
| EPI_ISL_430532 | Victorian Infectious Diseases Reference Laboratory (VIDRL) | Microbiological Diagnostic Unit Public Health Laboratory and Victorian Infectious Diseases Reference Laboratory, The Peter Doherty Institute for Infection and Immunity |
| EPI_ISL_430533 | Victorian Infectious Diseases Reference Laboratory (VIDRL) | Microbiological Diagnostic Unit Public Health Laboratory and Victorian Infectious Diseases Reference Laboratory, The Peter Doherty Institute for Infection and Immunity |
| EPI_ISL_430534 | Victorian Infectious Diseases Reference Laboratory (VIDRL) | Microbiological Diagnostic Unit Public Health Laboratory and Victorian Infectious Diseases Reference Laboratory, The Peter Doherty Institute for Infection and Immunity |
| EPI_ISL_430535 | Victorian Infectious Diseases Reference Laboratory (VIDRL) | Microbiological Diagnostic Unit Public Health Laboratory and Victorian Infectious Diseases Reference Laboratory, The Peter Doherty Institute for Infection and Immunity |
| EPI_ISL_430536 | Victorian Infectious Diseases Reference Laboratory (VIDRL) | Microbiological Diagnostic Unit Public Health Laboratory and Victorian Infectious Diseases Reference Laboratory, The Peter Doherty Institute for Infection and Immunity |
| EPI_ISL_430537 | Victorian Infectious Diseases Reference Laboratory (VIDRL) | Microbiological Diagnostic Unit Public Health Laboratory and Victorian Infectious Diseases Reference Laboratory, The Peter Doherty Institute for Infection and Immunity |
| EPI_ISL_430538 | Victorian Infectious Diseases Reference Laboratory (VIDRL) | Microbiological Diagnostic Unit Public Health Laboratory and Victorian Infectious Diseases Reference Laboratory, The Peter Doherty Institute for Infection and Immunity |

| EPI_Accession_ID | Originating Laboratory | Submitting Laboratory |
|---|---|---|
| EPI_ISL_430539 | Victorian Infectious Diseases Reference Laboratory (VIDRL) | Microbiological Diagnostic Unit Public Health Laboratory and Victorian Infectious Diseases Reference Laboratory, The Peter Doherty Institute for Infection and Immunity |
| EPI_ISL_430540 | Victorian Infectious Diseases Reference Laboratory (VIDRL) | Microbiological Diagnostic Unit Public Health Laboratory and Victorian Infectious Diseases Reference Laboratory, The Peter Doherty Institute for Infection and Immunity |
| EPI_ISL_430541 | Victorian Infectious Diseases Reference Laboratory (VIDRL) | Microbiological Diagnostic Unit Public Health Laboratory and Victorian Infectious Diseases Reference Laboratory, The Peter Doherty Institute for Infection and Immunity |
| EPI_ISL_430542 | Victorian Infectious Diseases Reference Laboratory (VIDRL) | Microbiological Diagnostic Unit Public Health Laboratory and Victorian Infectious Diseases Reference Laboratory, The Peter Doherty Institute for Infection and Immunity |
| EPI_ISL_430543 | Victorian Infectious Diseases Reference Laboratory (VIDRL) | Microbiological Diagnostic Unit Public Health Laboratory and Victorian Infectious Diseases Reference Laboratory, The Peter Doherty Institute for Infection and Immunity |
| EPI_ISL_430544 | Victorian Infectious Diseases Reference Laboratory (VIDRL) | Microbiological Diagnostic Unit Public Health Laboratory and Victorian Infectious Diseases Reference Laboratory, The Peter Doherty Institute for Infection and Immunity |
| EPI_ISL_430545 | Victorian Infectious Diseases Reference Laboratory (VIDRL) | Microbiological Diagnostic Unit Public Health Laboratory and Victorian Infectious Diseases Reference Laboratory, The Peter Doherty Institute for Infection and Immunity |
| EPI_ISL_430546 | Victorian Infectious Diseases Reference Laboratory (VIDRL) | Microbiological Diagnostic Unit Public Health Laboratory and Victorian Infectious Diseases Reference Laboratory, The Peter Doherty Institute for Infection and Immunity |

| EPI_Accession_ID | Originating Laboratory | Submitting Laboratory |
| --- | --- | --- |
| EPI_ISL_430547 | Victorian Infectious Diseases Reference Laboratory (VIDRL) | Microbiological Diagnostic Unit Public Health Laboratory and Victorian Infectious Diseases Reference Laboratory, The Peter Doherty Institute for Infection and Immunity |
| EPI_ISL_430548 | Victorian Infectious Diseases Reference Laboratory (VIDRL) | Microbiological Diagnostic Unit Public Health Laboratory and Victorian Infectious Diseases Reference Laboratory, The Peter Doherty Institute for Infection and Immunity |
| EPI_ISL_430549 | Victorian Infectious Diseases Reference Laboratory (VIDRL) | Microbiological Diagnostic Unit Public Health Laboratory and Victorian Infectious Diseases Reference Laboratory, The Peter Doherty Institute for Infection and Immunity |
| EPI_ISL_430550 | Victorian Infectious Diseases Reference Laboratory (VIDRL) | Microbiological Diagnostic Unit Public Health Laboratory and Victorian Infectious Diseases Reference Laboratory, The Peter Doherty Institute for Infection and Immunity |
| EPI_ISL_430551 | Victorian Infectious Diseases Reference Laboratory (VIDRL) | Microbiological Diagnostic Unit Public Health Laboratory and Victorian Infectious Diseases Reference Laboratory, The Peter Doherty Institute for Infection and Immunity |
| EPI_ISL_430552 | Victorian Infectious Diseases Reference Laboratory (VIDRL) | Microbiological Diagnostic Unit Public Health Laboratory and Victorian Infectious Diseases Reference Laboratory, The Peter Doherty Institute for Infection and Immunity |
| EPI_ISL_430553 | Victorian Infectious Diseases Reference Laboratory (VIDRL) | Microbiological Diagnostic Unit Public Health Laboratory and Victorian Infectious Diseases Reference Laboratory, The Peter Doherty Institute for Infection and Immunity |
| EPI_ISL_430554 | Victorian Infectious Diseases Reference Laboratory (VIDRL) | Microbiological Diagnostic Unit Public Health Laboratory and Victorian Infectious Diseases Reference Laboratory, The Peter Doherty Institute for Infection and Immunity |

| EPI_Accession_ID | Originating Laboratory | Submitting Laboratory |
|---|---|---|
| EPI_ISL_430555 | Victorian Infectious Diseases Reference Laboratory (VIDRL) | Microbiological Diagnostic Unit Public Health Laboratory and Victorian Infectious Diseases Reference Laboratory, The Peter Doherty Institute for Infection and Immunity |
| EPI_ISL_430556 | Victorian Infectious Diseases Reference Laboratory (VIDRL) | Microbiological Diagnostic Unit Public Health Laboratory and Victorian Infectious Diseases Reference Laboratory, The Peter Doherty Institute for Infection and Immunity |
| EPI_ISL_430557 | Victorian Infectious Diseases Reference Laboratory (VIDRL) | Microbiological Diagnostic Unit Public Health Laboratory and Victorian Infectious Diseases Reference Laboratory, The Peter Doherty Institute for Infection and Immunity |
| EPI_ISL_430558 | Victorian Infectious Diseases Reference Laboratory (VIDRL) | Microbiological Diagnostic Unit Public Health Laboratory and Victorian Infectious Diseases Reference Laboratory, The Peter Doherty Institute for Infection and Immunity |
| EPI_ISL_430559 | Victorian Infectious Diseases Reference Laboratory (VIDRL) | Microbiological Diagnostic Unit Public Health Laboratory and Victorian Infectious Diseases Reference Laboratory, The Peter Doherty Institute for Infection and Immunity |
| EPI_ISL_430560 | Victorian Infectious Diseases Reference Laboratory (VIDRL) | Microbiological Diagnostic Unit Public Health Laboratory and Victorian Infectious Diseases Reference Laboratory, The Peter Doherty Institute for Infection and Immunity |
| EPI_ISL_430561 | Victorian Infectious Diseases Reference Laboratory (VIDRL) | Microbiological Diagnostic Unit Public Health Laboratory and Victorian Infectious Diseases Reference Laboratory, The Peter Doherty Institute for Infection and Immunity |
| EPI_ISL_430562 | Victorian Infectious Diseases Reference Laboratory (VIDRL) | Microbiological Diagnostic Unit Public Health Laboratory and Victorian Infectious Diseases Reference Laboratory, The Peter Doherty Institute for Infection and Immunity |

| EPI_Accession_ID | Originating Laboratory | Submitting Laboratory |
|---|---|---|
| EPI_ISL_430563 | Victorian Infectious Diseases Reference Laboratory (VIDRL) | Microbiological Diagnostic Unit Public Health Laboratory and Victorian Infectious Diseases Reference Laboratory, The Peter Doherty Institute for Infection and Immunity |
| EPI_ISL_430564 | Victorian Infectious Diseases Reference Laboratory (VIDRL) | Microbiological Diagnostic Unit Public Health Laboratory and Victorian Infectious Diseases Reference Laboratory, The Peter Doherty Institute for Infection and Immunity |
| EPI_ISL_430565 | Victorian Infectious Diseases Reference Laboratory (VIDRL) | Microbiological Diagnostic Unit Public Health Laboratory and Victorian Infectious Diseases Reference Laboratory, The Peter Doherty Institute for Infection and Immunity |
| EPI_ISL_430566 | Victorian Infectious Diseases Reference Laboratory (VIDRL) | Microbiological Diagnostic Unit Public Health Laboratory and Victorian Infectious Diseases Reference Laboratory, The Peter Doherty Institute for Infection and Immunity |
| EPI_ISL_430567 | Victorian Infectious Diseases Reference Laboratory (VIDRL) | Microbiological Diagnostic Unit Public Health Laboratory and Victorian Infectious Diseases Reference Laboratory, The Peter Doherty Institute for Infection and Immunity |
| EPI_ISL_430568 | Victorian Infectious Diseases Reference Laboratory (VIDRL) | Microbiological Diagnostic Unit Public Health Laboratory and Victorian Infectious Diseases Reference Laboratory, The Peter Doherty Institute for Infection and Immunity |
| EPI_ISL_430569 | Victorian Infectious Diseases Reference Laboratory (VIDRL) | Microbiological Diagnostic Unit Public Health Laboratory and Victorian Infectious Diseases Reference Laboratory, The Peter Doherty Institute for Infection and Immunity |
| EPI_ISL_430570 | Victorian Infectious Diseases Reference Laboratory (VIDRL) | Microbiological Diagnostic Unit Public Health Laboratory and Victorian Infectious Diseases Reference Laboratory, The Peter Doherty Institute for Infection and Immunity |

| EPI_Accession_ID | Originating Laboratory | Submitting Laboratory |
|---|---|---|
| EPI_ISL_430571 | Victorian Infectious Diseases Reference Laboratory (VIDRL) | Microbiological Diagnostic Unit Public Health Laboratory and Victorian Infectious Diseases Reference Laboratory, The Peter Doherty Institute for Infection and Immunity |
| EPI_ISL_430572 | Victorian Infectious Diseases Reference Laboratory (VIDRL) | Microbiological Diagnostic Unit Public Health Laboratory and Victorian Infectious Diseases Reference Laboratory, The Peter Doherty Institute for Infection and Immunity |
| EPI_ISL_430573 | Victorian Infectious Diseases Reference Laboratory (VIDRL) | Microbiological Diagnostic Unit Public Health Laboratory and Victorian Infectious Diseases Reference Laboratory, The Peter Doherty Institute for Infection and Immunity |
| EPI_ISL_430574 | Victorian Infectious Diseases Reference Laboratory (VIDRL) | Microbiological Diagnostic Unit Public Health Laboratory and Victorian Infectious Diseases Reference Laboratory, The Peter Doherty Institute for Infection and Immunity |
| EPI_ISL_430575 | Victorian Infectious Diseases Reference Laboratory (VIDRL) | Microbiological Diagnostic Unit Public Health Laboratory and Victorian Infectious Diseases Reference Laboratory, The Peter Doherty Institute for Infection and Immunity |
| EPI_ISL_430576 | Victorian Infectious Diseases Reference Laboratory (VIDRL) | Microbiological Diagnostic Unit Public Health Laboratory and Victorian Infectious Diseases Reference Laboratory, The Peter Doherty Institute for Infection and Immunity |
| EPI_ISL_430577 | Victorian Infectious Diseases Reference Laboratory (VIDRL) | Microbiological Diagnostic Unit Public Health Laboratory and Victorian Infectious Diseases Reference Laboratory, The Peter Doherty Institute for Infection and Immunity |
| EPI_ISL_430578 | Victorian Infectious Diseases Reference Laboratory (VIDRL) | Microbiological Diagnostic Unit Public Health Laboratory and Victorian Infectious Diseases Reference Laboratory, The Peter Doherty Institute for Infection and Immunity |

| EPI_Accession_ID | Originating Laboratory | Submitting Laboratory |
|---|---|---|
| EPI_ISL_430579 | Victorian Infectious Diseases Reference Laboratory (VIDRL) | Microbiological Diagnostic Unit Public Health Laboratory and Victorian Infectious Diseases Reference Laboratory, The Peter Doherty Institute for Infection and Immunity |
| EPI_ISL_430580 | Victorian Infectious Diseases Reference Laboratory (VIDRL) | Microbiological Diagnostic Unit Public Health Laboratory and Victorian Infectious Diseases Reference Laboratory, The Peter Doherty Institute for Infection and Immunity |
| EPI_ISL_430581 | Victorian Infectious Diseases Reference Laboratory (VIDRL) | Microbiological Diagnostic Unit Public Health Laboratory and Victorian Infectious Diseases Reference Laboratory, The Peter Doherty Institute for Infection and Immunity |
| EPI_ISL_430582 | Victorian Infectious Diseases Reference Laboratory (VIDRL) | Microbiological Diagnostic Unit Public Health Laboratory and Victorian Infectious Diseases Reference Laboratory, The Peter Doherty Institute for Infection and Immunity |
| EPI_ISL_430583 | Victorian Infectious Diseases Reference Laboratory (VIDRL) | Microbiological Diagnostic Unit Public Health Laboratory and Victorian Infectious Diseases Reference Laboratory, The Peter Doherty Institute for Infection and Immunity |
| EPI_ISL_430584 | Victorian Infectious Diseases Reference Laboratory (VIDRL) | Microbiological Diagnostic Unit Public Health Laboratory and Victorian Infectious Diseases Reference Laboratory, The Peter Doherty Institute for Infection and Immunity |
| EPI_ISL_430585 | Victorian Infectious Diseases Reference Laboratory (VIDRL) | Microbiological Diagnostic Unit Public Health Laboratory and Victorian Infectious Diseases Reference Laboratory, The Peter Doherty Institute for Infection and Immunity |
| EPI_ISL_430586 | Victorian Infectious Diseases Reference Laboratory (VIDRL) | Microbiological Diagnostic Unit Public Health Laboratory and Victorian Infectious Diseases Reference Laboratory, The Peter Doherty Institute for Infection and Immunity |

| EPI_Accession_ID | Originating Laboratory | Submitting Laboratory |
|---|---|---|
| EPI_ISL_430587 | Victorian Infectious Diseases Reference Laboratory (VIDRL) | Microbiological Diagnostic Unit Public Health Laboratory and Victorian Infectious Diseases Reference Laboratory, The Peter Doherty Institute for Infection and Immunity |
| EPI_ISL_430588 | Victorian Infectious Diseases Reference Laboratory (VIDRL) | Microbiological Diagnostic Unit Public Health Laboratory and Victorian Infectious Diseases Reference Laboratory, The Peter Doherty Institute for Infection and Immunity |
| EPI_ISL_430589 | Victorian Infectious Diseases Reference Laboratory (VIDRL) | Microbiological Diagnostic Unit Public Health Laboratory and Victorian Infectious Diseases Reference Laboratory, The Peter Doherty Institute for Infection and Immunity |
| EPI_ISL_430590 | Victorian Infectious Diseases Reference Laboratory (VIDRL) | Microbiological Diagnostic Unit Public Health Laboratory and Victorian Infectious Diseases Reference Laboratory, The Peter Doherty Institute for Infection and Immunity |
| EPI_ISL_430591 | Victorian Infectious Diseases Reference Laboratory (VIDRL) | Microbiological Diagnostic Unit Public Health Laboratory and Victorian Infectious Diseases Reference Laboratory, The Peter Doherty Institute for Infection and Immunity |
| EPI_ISL_430592 | Victorian Infectious Diseases Reference Laboratory (VIDRL) | Microbiological Diagnostic Unit Public Health Laboratory and Victorian Infectious Diseases Reference Laboratory, The Peter Doherty Institute for Infection and Immunity |
| EPI_ISL_430593 | Victorian Infectious Diseases Reference Laboratory (VIDRL) | Microbiological Diagnostic Unit Public Health Laboratory and Victorian Infectious Diseases Reference Laboratory, The Peter Doherty Institute for Infection and Immunity |
| EPI_ISL_430594 | Victorian Infectious Diseases Reference Laboratory (VIDRL) | Microbiological Diagnostic Unit Public Health Laboratory and Victorian Infectious Diseases Reference Laboratory, The Peter Doherty Institute for Infection and Immunity |

| EPI_Accession_ID | Originating Laboratory | Submitting Laboratory |
|---|---|---|
| EPI_ISL_430595 | Victorian Infectious Diseases Reference Laboratory (VIDRL) | Microbiological Diagnostic Unit Public Health Laboratory and Victorian Infectious Diseases Reference Laboratory, The Peter Doherty Institute for Infection and Immunity |
| EPI_ISL_430596 | Victorian Infectious Diseases Reference Laboratory (VIDRL) | Microbiological Diagnostic Unit Public Health Laboratory and Victorian Infectious Diseases Reference Laboratory, The Peter Doherty Institute for Infection and Immunity |
| EPI_ISL_430597 | Victorian Infectious Diseases Reference Laboratory (VIDRL) | Microbiological Diagnostic Unit Public Health Laboratory and Victorian Infectious Diseases Reference Laboratory, The Peter Doherty Institute for Infection and Immunity |
| EPI_ISL_430598 | Victorian Infectious Diseases Reference Laboratory (VIDRL) | Microbiological Diagnostic Unit Public Health Laboratory and Victorian Infectious Diseases Reference Laboratory, The Peter Doherty Institute for Infection and Immunity |
| EPI_ISL_430599 | Victorian Infectious Diseases Reference Laboratory (VIDRL) | Microbiological Diagnostic Unit Public Health Laboratory and Victorian Infectious Diseases Reference Laboratory, The Peter Doherty Institute for Infection and Immunity |
| EPI_ISL_430600 | Victorian Infectious Diseases Reference Laboratory (VIDRL) | Microbiological Diagnostic Unit Public Health Laboratory and Victorian Infectious Diseases Reference Laboratory, The Peter Doherty Institute for Infection and Immunity |
| EPI_ISL_430601 | Victorian Infectious Diseases Reference Laboratory (VIDRL) | Microbiological Diagnostic Unit Public Health Laboratory and Victorian Infectious Diseases Reference Laboratory, The Peter Doherty Institute for Infection and Immunity |
| EPI_ISL_430602 | Victorian Infectious Diseases Reference Laboratory (VIDRL) | Microbiological Diagnostic Unit Public Health Laboratory and Victorian Infectious Diseases Reference Laboratory, The Peter Doherty Institute for Infection and Immunity |

| EPI_Accession_ID | Originating Laboratory | Submitting Laboratory |
|---|---|---|
| EPI_ISL_430603 | Victorian Infectious Diseases Reference Laboratory (VIDRL) | Microbiological Diagnostic Unit Public Health Laboratory and Victorian Infectious Diseases Reference Laboratory, The Peter Doherty Institute for Infection and Immunity |
| EPI_ISL_430604 | Victorian Infectious Diseases Reference Laboratory (VIDRL) | Microbiological Diagnostic Unit Public Health Laboratory and Victorian Infectious Diseases Reference Laboratory, The Peter Doherty Institute for Infection and Immunity |
| EPI_ISL_430605 | Victorian Infectious Diseases Reference Laboratory (VIDRL) | Microbiological Diagnostic Unit Public Health Laboratory and Victorian Infectious Diseases Reference Laboratory, The Peter Doherty Institute for Infection and Immunity |
| EPI_ISL_430606 | Victorian Infectious Diseases Reference Laboratory (VIDRL) | Microbiological Diagnostic Unit Public Health Laboratory and Victorian Infectious Diseases Reference Laboratory, The Peter Doherty Institute for Infection and Immunity |
| EPI_ISL_430607 | Victorian Infectious Diseases Reference Laboratory (VIDRL) | Microbiological Diagnostic Unit Public Health Laboratory and Victorian Infectious Diseases Reference Laboratory, The Peter Doherty Institute for Infection and Immunity |
| EPI_ISL_430608 | Victorian Infectious Diseases Reference Laboratory (VIDRL) | Microbiological Diagnostic Unit Public Health Laboratory and Victorian Infectious Diseases Reference Laboratory, The Peter Doherty Institute for Infection and Immunity |
| EPI_ISL_430609 | Victorian Infectious Diseases Reference Laboratory (VIDRL) | Microbiological Diagnostic Unit Public Health Laboratory and Victorian Infectious Diseases Reference Laboratory, The Peter Doherty Institute for Infection and Immunity |
| EPI_ISL_430610 | Victorian Infectious Diseases Reference Laboratory (VIDRL) | Microbiological Diagnostic Unit Public Health Laboratory and Victorian Infectious Diseases Reference Laboratory, The Peter Doherty Institute for Infection and Immunity |

| EPI_Accession_ID | Originating Laboratory | Submitting Laboratory |
|---|---|---|
| EPI_ISL_430611 | Victorian Infectious Diseases Reference Laboratory (VIDRL) | Microbiological Diagnostic Unit Public Health Laboratory and Victorian Infectious Diseases Reference Laboratory, The Peter Doherty Institute for Infection and Immunity |
| EPI_ISL_430612 | Victorian Infectious Diseases Reference Laboratory (VIDRL) | Microbiological Diagnostic Unit Public Health Laboratory and Victorian Infectious Diseases Reference Laboratory, The Peter Doherty Institute for Infection and Immunity |
| EPI_ISL_430613 | Victorian Infectious Diseases Reference Laboratory (VIDRL) | Microbiological Diagnostic Unit Public Health Laboratory and Victorian Infectious Diseases Reference Laboratory, The Peter Doherty Institute for Infection and Immunity |
| EPI_ISL_430614 | Victorian Infectious Diseases Reference Laboratory (VIDRL) | Microbiological Diagnostic Unit Public Health Laboratory and Victorian Infectious Diseases Reference Laboratory, The Peter Doherty Institute for Infection and Immunity |
| EPI_ISL_430615 | Victorian Infectious Diseases Reference Laboratory (VIDRL) | Microbiological Diagnostic Unit Public Health Laboratory and Victorian Infectious Diseases Reference Laboratory, The Peter Doherty Institute for Infection and Immunity |
| EPI_ISL_430616 | Victorian Infectious Diseases Reference Laboratory (VIDRL) | Microbiological Diagnostic Unit Public Health Laboratory and Victorian Infectious Diseases Reference Laboratory, The Peter Doherty Institute for Infection and Immunity |
| EPI_ISL_430617 | Victorian Infectious Diseases Reference Laboratory (VIDRL) | Microbiological Diagnostic Unit Public Health Laboratory and Victorian Infectious Diseases Reference Laboratory, The Peter Doherty Institute for Infection and Immunity |
| EPI_ISL_430618 | Victorian Infectious Diseases Reference Laboratory (VIDRL) | Microbiological Diagnostic Unit Public Health Laboratory and Victorian Infectious Diseases Reference Laboratory, The Peter Doherty Institute for Infection and Immunity |

| EPI_Accession_ID | Originating Laboratory | Submitting Laboratory |
|---|---|---|
| EPI_ISL_430619 | Victorian Infectious Diseases Reference Laboratory (VIDRL) | Microbiological Diagnostic Unit Public Health Laboratory and Victorian Infectious Diseases Reference Laboratory, The Peter Doherty Institute for Infection and Immunity |
| EPI_ISL_430620 | Victorian Infectious Diseases Reference Laboratory (VIDRL) | Microbiological Diagnostic Unit Public Health Laboratory and Victorian Infectious Diseases Reference Laboratory, The Peter Doherty Institute for Infection and Immunity |
| EPI_ISL_430621 | Victorian Infectious Diseases Reference Laboratory (VIDRL) | Microbiological Diagnostic Unit Public Health Laboratory and Victorian Infectious Diseases Reference Laboratory, The Peter Doherty Institute for Infection and Immunity |
| EPI_ISL_430622 | Victorian Infectious Diseases Reference Laboratory (VIDRL) | Microbiological Diagnostic Unit Public Health Laboratory and Victorian Infectious Diseases Reference Laboratory, The Peter Doherty Institute for Infection and Immunity |
| EPI_ISL_430623 | Victorian Infectious Diseases Reference Laboratory (VIDRL) | Microbiological Diagnostic Unit Public Health Laboratory and Victorian Infectious Diseases Reference Laboratory, The Peter Doherty Institute for Infection and Immunity |
| EPI_ISL_430624 | Victorian Infectious Diseases Reference Laboratory (VIDRL) | Microbiological Diagnostic Unit Public Health Laboratory and Victorian Infectious Diseases Reference Laboratory, The Peter Doherty Institute for Infection and Immunity |
| EPI_ISL_430625 | Victorian Infectious Diseases Reference Laboratory (VIDRL) | Microbiological Diagnostic Unit Public Health Laboratory and Victorian Infectious Diseases Reference Laboratory, The Peter Doherty Institute for Infection and Immunity |
| EPI_ISL_430626 | Victorian Infectious Diseases Reference Laboratory (VIDRL) | Microbiological Diagnostic Unit Public Health Laboratory and Victorian Infectious Diseases Reference Laboratory, The Peter Doherty Institute for Infection and Immunity |

| EPI_Accession_ID | Originating Laboratory | Submitting Laboratory |
|---|---|---|
| EPI_ISL_430627 | Victorian Infectious Diseases Reference Laboratory (VIDRL) | Microbiological Diagnostic Unit Public Health Laboratory and Victorian Infectious Diseases Reference Laboratory, The Peter Doherty Institute for Infection and Immunity |
| EPI_ISL_430628 | Victorian Infectious Diseases Reference Laboratory (VIDRL) | Microbiological Diagnostic Unit Public Health Laboratory and Victorian Infectious Diseases Reference Laboratory, The Peter Doherty Institute for Infection and Immunity |
| EPI_ISL_430629 | Victorian Infectious Diseases Reference Laboratory (VIDRL) | Microbiological Diagnostic Unit Public Health Laboratory and Victorian Infectious Diseases Reference Laboratory, The Peter Doherty Institute for Infection and Immunity |
| EPI_ISL_430630 | Victorian Infectious Diseases Reference Laboratory (VIDRL) | Microbiological Diagnostic Unit Public Health Laboratory and Victorian Infectious Diseases Reference Laboratory, The Peter Doherty Institute for Infection and Immunity |
| EPI_ISL_430637 | Victorian Infectious Diseases Reference Laboratory (VIDRL) | Microbiological Diagnostic Unit Public Health Laboratory and Victorian Infectious Diseases Reference Laboratory, The Peter Doherty Institute for Infection and Immunity |
| EPI_ISL_430638 | Victorian Infectious Diseases Reference Laboratory (VIDRL) | Microbiological Diagnostic Unit Public Health Laboratory and Victorian Infectious Diseases Reference Laboratory, The Peter Doherty Institute for Infection and Immunity |
| EPI_ISL_430639 | Microbiological Diagnostic Unit Public Health Laboratory | Microbiological Diagnostic Unit Public Health Laboratory |
| EPI_ISL_430640 | Microbiological Diagnostic Unit Public Health Laboratory | Microbiological Diagnostic Unit Public Health Laboratory |
| EPI_ISL_430641 | Microbiological Diagnostic Unit Public Health Laboratory | Microbiological Diagnostic Unit Public Health Laboratory |
| EPI_ISL_430642 | Microbiological Diagnostic Unit Public Health Laboratory | Microbiological Diagnostic Unit Public Health Laboratory |

| EPI_Accession_ID | Originating Laboratory | Submitting Laboratory |
|---|---|---|
| EPI_ISL_430643 | Microbiological Diagnostic Unit Public Health Laboratory | Microbiological Diagnostic Unit Public Health Laboratory |
| EPI_ISL_430644 | Microbiological Diagnostic Unit Public Health Laboratory | Microbiological Diagnostic Unit Public Health Laboratory |
| EPI_ISL_430645 | Microbiological Diagnostic Unit Public Health Laboratory | Microbiological Diagnostic Unit Public Health Laboratory |
| EPI_ISL_430646 | Microbiological Diagnostic Unit Public Health Laboratory | Microbiological Diagnostic Unit Public Health Laboratory |
| EPI_ISL_430647 | Microbiological Diagnostic Unit Public Health Laboratory | Microbiological Diagnostic Unit Public Health Laboratory |
| EPI_ISL_430648 | Microbiological Diagnostic Unit Public Health Laboratory | Microbiological Diagnostic Unit Public Health Laboratory |
| EPI_ISL_430649 | Microbiological Diagnostic Unit Public Health Laboratory | Microbiological Diagnostic Unit Public Health Laboratory |
| EPI_ISL_430650 | Microbiological Diagnostic Unit Public Health Laboratory | Microbiological Diagnostic Unit Public Health Laboratory |
| EPI_ISL_430651 | Microbiological Diagnostic Unit Public Health Laboratory | Microbiological Diagnostic Unit Public Health Laboratory |
| EPI_ISL_430652 | Microbiological Diagnostic Unit Public Health Laboratory | Microbiological Diagnostic Unit Public Health Laboratory |
| EPI_ISL_430653 | Microbiological Diagnostic Unit Public Health Laboratory | Microbiological Diagnostic Unit Public Health Laboratory |
| EPI_ISL_430654 | Microbiological Diagnostic Unit Public Health Laboratory | Microbiological Diagnostic Unit Public Health Laboratory |
| EPI_ISL_430655 | Microbiological Diagnostic Unit Public Health Laboratory | Microbiological Diagnostic Unit Public Health Laboratory |
| EPI_ISL_430656 | Microbiological Diagnostic Unit Public Health Laboratory | Microbiological Diagnostic Unit Public Health Laboratory |
| EPI_ISL_430657 | Microbiological Diagnostic Unit Public Health Laboratory | Microbiological Diagnostic Unit Public Health Laboratory |
| EPI_ISL_430658 | Microbiological Diagnostic Unit Public Health Laboratory | Microbiological Diagnostic Unit Public Health Laboratory |

| EPI_Accession_ID | Originating Laboratory | Submitting Laboratory |
|---|---|---|
| EPI_ISL_430659 | Microbiological Diagnostic Unit Public Health Laboratory | Microbiological Diagnostic Unit Public Health Laboratory |
| EPI_ISL_430660 | Microbiological Diagnostic Unit Public Health Laboratory | Microbiological Diagnostic Unit Public Health Laboratory |
| EPI_ISL_430661 | Microbiological Diagnostic Unit Public Health Laboratory | Microbiological Diagnostic Unit Public Health Laboratory |
| EPI_ISL_430662 | Microbiological Diagnostic Unit Public Health Laboratory | Microbiological Diagnostic Unit Public Health Laboratory |
| EPI_ISL_430663 | Microbiological Diagnostic Unit Public Health Laboratory | Microbiological Diagnostic Unit Public Health Laboratory |
| EPI_ISL_430664 | Microbiological Diagnostic Unit Public Health Laboratory | Microbiological Diagnostic Unit Public Health Laboratory |
| EPI_ISL_430665 | Microbiological Diagnostic Unit Public Health Laboratory | Microbiological Diagnostic Unit Public Health Laboratory |
| EPI_ISL_430666 | Microbiological Diagnostic Unit Public Health Laboratory | Microbiological Diagnostic Unit Public Health Laboratory |
| EPI_ISL_430667 | Microbiological Diagnostic Unit Public Health Laboratory | Microbiological Diagnostic Unit Public Health Laboratory |
| EPI_ISL_430668 | Microbiological Diagnostic Unit Public Health Laboratory | Microbiological Diagnostic Unit Public Health Laboratory |
| EPI_ISL_430669 | Microbiological Diagnostic Unit Public Health Laboratory | Microbiological Diagnostic Unit Public Health Laboratory |
| EPI_ISL_430670 | Microbiological Diagnostic Unit Public Health Laboratory | Microbiological Diagnostic Unit Public Health Laboratory |
| EPI_ISL_430671 | Microbiological Diagnostic Unit Public Health Laboratory | Microbiological Diagnostic Unit Public Health Laboratory |
| EPI_ISL_430672 | Microbiological Diagnostic Unit Public Health Laboratory | Microbiological Diagnostic Unit Public Health Laboratory |
| EPI_ISL_430673 | Microbiological Diagnostic Unit Public Health Laboratory | Microbiological Diagnostic Unit Public Health Laboratory |
| EPI_ISL_430674 | Microbiological Diagnostic Unit Public Health Laboratory | Microbiological Diagnostic Unit Public Health Laboratory |

| EPI_Accession_ID | Originating Laboratory | Submitting Laboratory |
|---|---|---|
| EPI_ISL_430675 | Microbiological Diagnostic Unit Public Health Laboratory | Microbiological Diagnostic Unit Public Health Laboratory |
| EPI_ISL_430676 | Microbiological Diagnostic Unit Public Health Laboratory | Microbiological Diagnostic Unit Public Health Laboratory |
| EPI_ISL_430677 | Microbiological Diagnostic Unit Public Health Laboratory | Microbiological Diagnostic Unit Public Health Laboratory |
| EPI_ISL_430678 | Microbiological Diagnostic Unit Public Health Laboratory | Microbiological Diagnostic Unit Public Health Laboratory |
| EPI_ISL_430679 | Microbiological Diagnostic Unit Public Health Laboratory | Microbiological Diagnostic Unit Public Health Laboratory |
| EPI_ISL_430680 | Microbiological Diagnostic Unit Public Health Laboratory | Microbiological Diagnostic Unit Public Health Laboratory |
| EPI_ISL_430681 | Microbiological Diagnostic Unit Public Health Laboratory | Microbiological Diagnostic Unit Public Health Laboratory |
| EPI_ISL_430682 | Microbiological Diagnostic Unit Public Health Laboratory | Microbiological Diagnostic Unit Public Health Laboratory |
| EPI_ISL_430683 | Microbiological Diagnostic Unit Public Health Laboratory | Microbiological Diagnostic Unit Public Health Laboratory |
| EPI_ISL_430684 | Microbiological Diagnostic Unit Public Health Laboratory | Microbiological Diagnostic Unit Public Health Laboratory |
| EPI_ISL_430685 | Microbiological Diagnostic Unit Public Health Laboratory | Microbiological Diagnostic Unit Public Health Laboratory |
| EPI_ISL_430686 | Microbiological Diagnostic Unit Public Health Laboratory | Microbiological Diagnostic Unit Public Health Laboratory |
| EPI_ISL_430687 | Microbiological Diagnostic Unit Public Health Laboratory | Microbiological Diagnostic Unit Public Health Laboratory |
| EPI_ISL_430688 | Victorian Infectious Diseases Reference Laboratory (VIDRL) | Microbiological Diagnostic Unit Public Health Laboratory and Victorian Infectious Diseases Reference Laboratory, The Peter Doherty Institute for Infection and Immunity |

| EPI_Accession_ID | Originating Laboratory | Submitting Laboratory |
|---|---|---|
| EPI_ISL_430689 | Victorian Infectious Diseases Reference Laboratory (VIDRL) | Microbiological Diagnostic Unit Public Health Laboratory and Victorian Infectious Diseases Reference Laboratory, The Peter Doherty Institute for Infection and Immunity |
| EPI_ISL_430690 | Victorian Infectious Diseases Reference Laboratory (VIDRL) | Microbiological Diagnostic Unit Public Health Laboratory and Victorian Infectious Diseases Reference Laboratory, The Peter Doherty Institute for Infection and Immunity |
| EPI_ISL_430691 | Victorian Infectious Diseases Reference Laboratory (VIDRL) | Microbiological Diagnostic Unit Public Health Laboratory and Victorian Infectious Diseases Reference Laboratory, The Peter Doherty Institute for Infection and Immunity |
| EPI_ISL_430692 | Victorian Infectious Diseases Reference Laboratory (VIDRL) | Microbiological Diagnostic Unit Public Health Laboratory and Victorian Infectious Diseases Reference Laboratory, The Peter Doherty Institute for Infection and Immunity |
| EPI_ISL_430693 | Victorian Infectious Diseases Reference Laboratory (VIDRL) | Microbiological Diagnostic Unit Public Health Laboratory and Victorian Infectious Diseases Reference Laboratory, The Peter Doherty Institute for Infection and Immunity |
| EPI_ISL_430694 | Victorian Infectious Diseases Reference Laboratory (VIDRL) | Microbiological Diagnostic Unit Public Health Laboratory and Victorian Infectious Diseases Reference Laboratory, The Peter Doherty Institute for Infection and Immunity |
| EPI_ISL_430695 | Victorian Infectious Diseases Reference Laboratory (VIDRL) | Microbiological Diagnostic Unit Public Health Laboratory and Victorian Infectious Diseases Reference Laboratory, The Peter Doherty Institute for Infection and Immunity |
| EPI_ISL_430696 | Victorian Infectious Diseases Reference Laboratory (VIDRL) | Microbiological Diagnostic Unit Public Health Laboratory and Victorian Infectious Diseases Reference Laboratory, The Peter Doherty Institute for Infection and Immunity |

| EPI_Accession_ID | Originating Laboratory | Submitting Laboratory |
|---|---|---|
| EPI_ISL_430697 | Victorian Infectious Diseases Reference Laboratory (VIDRL) | Microbiological Diagnostic Unit Public Health Laboratory and Victorian Infectious Diseases Reference Laboratory, The Peter Doherty Institute for Infection and Immunity |
| EPI_ISL_430698 | Victorian Infectious Diseases Reference Laboratory (VIDRL) | Microbiological Diagnostic Unit Public Health Laboratory and Victorian Infectious Diseases Reference Laboratory, The Peter Doherty Institute for Infection and Immunity |
| EPI_ISL_430699 | Victorian Infectious Diseases Reference Laboratory (VIDRL) | Microbiological Diagnostic Unit Public Health Laboratory and Victorian Infectious Diseases Reference Laboratory, The Peter Doherty Institute for Infection and Immunity |
| EPI_ISL_430700 | Victorian Infectious Diseases Reference Laboratory (VIDRL) | Microbiological Diagnostic Unit Public Health Laboratory and Victorian Infectious Diseases Reference Laboratory, The Peter Doherty Institute for Infection and Immunity |
| EPI_ISL_430701 | Victorian Infectious Diseases Reference Laboratory (VIDRL) | Microbiological Diagnostic Unit Public Health Laboratory and Victorian Infectious Diseases Reference Laboratory, The Peter Doherty Institute for Infection and Immunity |
| EPI_ISL_430702 | Victorian Infectious Diseases Reference Laboratory (VIDRL) | Microbiological Diagnostic Unit Public Health Laboratory and Victorian Infectious Diseases Reference Laboratory, The Peter Doherty Institute for Infection and Immunity |
| EPI_ISL_430703 | Victorian Infectious Diseases Reference Laboratory (VIDRL) | Microbiological Diagnostic Unit Public Health Laboratory and Victorian Infectious Diseases Reference Laboratory, The Peter Doherty Institute for Infection and Immunity |
| EPI_ISL_430704 | Victorian Infectious Diseases Reference Laboratory (VIDRL) | Microbiological Diagnostic Unit Public Health Laboratory and Victorian Infectious Diseases Reference Laboratory, The Peter Doherty Institute for Infection and Immunity |

| EPI_Accession_ID | Originating Laboratory | Submitting Laboratory |
|---|---|---|
| EPI_ISL_430705 | Victorian Infectious Diseases Reference Laboratory (VIDRL) | Microbiological Diagnostic Unit Public Health Laboratory and Victorian Infectious Diseases Reference Laboratory, The Peter Doherty Institute for Infection and Immunity |
| EPI_ISL_430706 | Victorian Infectious Diseases Reference Laboratory (VIDRL) | Microbiological Diagnostic Unit Public Health Laboratory and Victorian Infectious Diseases Reference Laboratory, The Peter Doherty Institute for Infection and Immunity |
| EPI_ISL_430707 | Victorian Infectious Diseases Reference Laboratory (VIDRL) | Microbiological Diagnostic Unit Public Health Laboratory and Victorian Infectious Diseases Reference Laboratory, The Peter Doherty Institute for Infection and Immunity |
| EPI_ISL_430708 | Victorian Infectious Diseases Reference Laboratory (VIDRL) | Microbiological Diagnostic Unit Public Health Laboratory and Victorian Infectious Diseases Reference Laboratory, The Peter Doherty Institute for Infection and Immunity |
| EPI_ISL_430709 | Victorian Infectious Diseases Reference Laboratory (VIDRL) | Microbiological Diagnostic Unit Public Health Laboratory and Victorian Infectious Diseases Reference Laboratory, The Peter Doherty Institute for Infection and Immunity |
| EPI_ISL_430710 | Victorian Infectious Diseases Reference Laboratory (VIDRL) | Microbiological Diagnostic Unit Public Health Laboratory and Victorian Infectious Diseases Reference Laboratory, The Peter Doherty Institute for Infection and Immunity |
| EPI_ISL_430711 | Victorian Infectious Diseases Reference Laboratory (VIDRL) | Microbiological Diagnostic Unit Public Health Laboratory and Victorian Infectious Diseases Reference Laboratory, The Peter Doherty Institute for Infection and Immunity |
| EPI_ISL_430712 | Victorian Infectious Diseases Reference Laboratory (VIDRL) | Microbiological Diagnostic Unit Public Health Laboratory and Victorian Infectious Diseases Reference Laboratory, The Peter Doherty Institute for Infection and Immunity |

| EPI_Accession_ID | Originating Laboratory | Submitting Laboratory |
|---|---|---|
| EPI_ISL_430713 | Victorian Infectious Diseases Reference Laboratory (VIDRL) | Microbiological Diagnostic Unit Public Health Laboratory and Victorian Infectious Diseases Reference Laboratory, The Peter Doherty Institute for Infection and Immunity |
| EPI_ISL_430714 | Victorian Infectious Diseases Reference Laboratory (VIDRL) | Microbiological Diagnostic Unit Public Health Laboratory and Victorian Infectious Diseases Reference Laboratory, The Peter Doherty Institute for Infection and Immunity |
| EPI_ISL_430715 | Microbiological Diagnostic Unit Public Health Laboratory | Microbiological Diagnostic Unit Public Health Laboratory |
| EPI_ISL_430716 | Microbiological Diagnostic Unit Public Health Laboratory | Microbiological Diagnostic Unit Public Health Laboratory |
| EPI_ISL_430717 | Microbiological Diagnostic Unit Public Health Laboratory | Microbiological Diagnostic Unit Public Health Laboratory |
| EPI_ISL_430718 | Hospital Universitario 12 de Octubre | Hospital Universitario 12 de Octubre |
| EPI_ISL_430719 | Hospital Universitario La Paz | Hospital Universitario 12 de Octubre |
| EPI_ISL_430720 | Hospital Universitario La Paz | Hospital Universitario 12 de Octubre |
| EPI_ISL_430721 | Hospital Universitario La Paz | Hospital Universitario 12 de Octubre |
| EPI_ISL_430722 | Chinese PLA Institute for Disease Control and Prevention | Chinese PLA Institute for Disease Control and Prevention |
| EPI_ISL_430723 | Chinese PLA Institute for Disease Control and Prevention | Chinese PLA Institute for Disease Control and Prevention |
| EPI_ISL_430724 | Chinese PLA Institute for Disease Control and Prevention | Chinese PLA Institute for Disease Control and Prevention |
| EPI_ISL_430725 | Chinese PLA Institute for Disease Control and Prevention | Chinese PLA Institute for Disease Control and Prevention |
| EPI_ISL_430727 | Chinese PLA Institute for Disease Control and Prevention | Chinese PLA Institute for Disease Control and Prevention |
| EPI_ISL_430728 | Chinese PLA Institute for Disease Control and Prevention | Chinese PLA Institute for Disease Control and Prevention |
| EPI_ISL_430729 | Chinese PLA Institute for Disease Control and Prevention | Chinese PLA Institute for Disease Control and Prevention |

| EPI_Accession_ID | Originating Laboratory | Submitting Laboratory |
|---|---|---|
| EPI_ISL_430730 | Chinese PLA Institute for Disease Control and Prevention | Chinese PLA Institute for Disease Control and Prevention |
| EPI_ISL_430731 | Chinese PLA Institute for Disease Control and Prevention | Chinese PLA Institute for Disease Control and Prevention |
| EPI_ISL_430732 | Chinese PLA Institute for Disease Control and Prevention | Chinese PLA Institute for Disease Control and Prevention |
| EPI_ISL_430733 | Chinese PLA Institute for Disease Control and Prevention | Chinese PLA Institute for Disease Control and Prevention |
| EPI_ISL_430734 | Chinese PLA Institute for Disease Control and Prevention | Chinese PLA Institute for Disease Control and Prevention |
| EPI_ISL_430735 | Chinese PLA Institute for Disease Control and Prevention | Chinese PLA Institute for Disease Control and Prevention |
| EPI_ISL_430736 | Chinese PLA Institute for Disease Control and Prevention | Chinese PLA Institute for Disease Control and Prevention |
| EPI_ISL_430737 | Chinese PLA Institute for Disease Control and Prevention | Chinese PLA Institute for Disease Control and Prevention |
| EPI_ISL_430738 | Chinese PLA Institute for Disease Control and Prevention | Chinese PLA Institute for Disease Control and Prevention |
| EPI_ISL_430739 | Chinese PLA Institute for Disease Control and Prevention | Chinese PLA Institute for Disease Control and Prevention |
| EPI_ISL_430740 | Chinese PLA Institute for Disease Control and Prevention | Chinese PLA Institute for Disease Control and Prevention |
| EPI_ISL_430741 | Chinese PLA Institute for Disease Control and Prevention | Chinese PLA Institute for Disease Control and Prevention |
| EPI_ISL_430742 | Chinese PLA Institute for Disease Control and Prevention | Chinese PLA Institute for Disease Control and Prevention |
| EPI_ISL_430743 | Chinese PLA Institute for Disease Control and Prevention | Chinese PLA Institute for Disease Control and Prevention |
| EPI_ISL_430744 | Chinese PLA Institute for Disease Control and Prevention | Chinese PLA Institute for Disease Control and Prevention |
| EPI_ISL_430745 | Chinese PLA Institute for Disease Control and Prevention | Chinese PLA Institute for Disease Control and Prevention |

| EPI_Accession_ID | Originating Laboratory | Submitting Laboratory |
| --- | --- | --- |
| EPI_ISL_430746 | Chinese PLA Institute for Disease Control and Prevention | Chinese PLA Institute for Disease Control and Prevention |
| EPI_ISL_430791 | UCSF Clinical Microbiology Laboratory | Chan-Zuckerberg Biohub |
| EPI_ISL_430792 | UCSF Clinical Microbiology Laboratory | Chan-Zuckerberg Biohub |
| EPI_ISL_430793 | Laboratorio AnÃ¡lisis ClÃ-nicos, Unidad de Servicios DiagnÃ³sticos, Swiss Medical Group | Ãrea de SecuenciaciÃ³n del Laboratorio de VirologÃ-a del Hospital de NiÃ±os Dr. Ricardo Gutierrez |
| EPI_ISL_430794 | Laboratorio AnÃ¡lisis ClÃ-nicos, Unidad de Servicios DiagnÃ³sticos, Swiss Medical Group | Ãrea de SecuenciaciÃ³n del Laboratorio de VirologÃ-a del Hospital de NiÃ±os Dr. Ricardo Gutierrez |
| EPI_ISL_430795 | Laboratorio de VirologÃ-a del Hospital de NiÃ±os Dr. Ricardo Gutierrez | Ãrea de SecuenciaciÃ³n del Laboratorio de VirologÃ-a del Hospital de NiÃ±os Dr. Ricardo Gutierrez |
| EPI_ISL_430796 | Departamento de BiologÃ-a y genÃ©tica molecular, IACA Laboratorios. | Ãrea de SecuenciaciÃ³n del Laboratorio de VirologÃ-a del Hospital de NiÃ±os Dr. Ricardo Gutierrez |
| EPI_ISL_430797 | Departamento de BiologÃ-a y genÃ©tica molecular, IACA Laboratorios. | Ãrea de SecuenciaciÃ³n del Laboratorio de VirologÃ-a del Hospital de NiÃ±os Dr. Ricardo Gutierrez |
| EPI_ISL_430798 | Departamento de BiologÃ-a y genÃ©tica molecular, IACA Laboratorios. | Ãrea de SecuenciaciÃ³n del Laboratorio de VirologÃ-a del Hospital de NiÃ±os Dr. Ricardo Gutierrez |
| EPI_ISL_430799 | Laboratorio de VirologÃ-a del Hospital de NiÃ±os Dr. Ricardo Gutierrez | Ãrea de SecuenciaciÃ³n del Laboratorio de VirologÃ-a del Hospital de NiÃ±os Dr. Ricardo Gutierrez |
| EPI_ISL_430800 | Laboratorio de VirologÃ-a del Hospital de NiÃ±os Dr. Ricardo Gutierrez | Ãrea de SecuenciaciÃ³n del Laboratorio de VirologÃ-a del Hospital de NiÃ±os Dr. Ricardo Gutierrez |
| EPI_ISL_430801 | Laboratorio de VirologÃ-a del Hospital de NiÃ±os Dr. Ricardo Gutierrez | Ãrea de SecuenciaciÃ³n del Laboratorio de VirologÃ-a del Hospital de NiÃ±os Dr. Ricardo Gutierrez |
| EPI_ISL_430802 | Departamento de BiologÃ-a y genÃ©tica molecular, IACA Laboratorios. | Ãrea de SecuenciaciÃ³n del Laboratorio de VirologÃ-a del Hospital de NiÃ±os Dr. Ricardo Gutierrez |
| EPI_ISL_430803 | Laboratorio de VirologÃ-a del Hospital de NiÃ±os Dr. Ricardo Gutierrez | Ãrea de SecuenciaciÃ³n del Laboratorio de VirologÃ-a del Hospital de NiÃ±os Dr. Ricardo Gutierrez |
| EPI_ISL_430804 | Laboratorio de VirologÃ-a del Hospital de NiÃ±os Dr. Ricardo Gutierrez | Ãrea de SecuenciaciÃ³n del Laboratorio de VirologÃ-a del Hospital de NiÃ±os Dr. Ricardo Gutierrez |
| EPI_ISL_430805 | Departamento de BiologÃ-a y genÃ©tica molecular, IACA Laboratorios. | Ãrea de SecuenciaciÃ³n del Laboratorio de VirologÃ-a del Hospital de NiÃ±os Dr. Ricardo Gutierrez |
| EPI_ISL_430806 | Departamento de BiologÃ-a y genÃ©tica molecular, IACA Laboratorios. | Ãrea de SecuenciaciÃ³n del Laboratorio de VirologÃ-a del Hospital de NiÃ±os Dr. Ricardo Gutierrez |

| EPI_Accession_ID | Originating Laboratory | Submitting Laboratory |
|---|---|---|
| EPI_ISL_430807 | Laboratorio de VirologÃ-a del Hospital de NiÃ±os Dr. Ricardo Gutierrez | Ã�ea de SecuenciaciÃ³n del Laboratorio de VirologÃ-a del Hospital de NiÃ±os Dr. Ricardo Gutierrez |
| EPI_ISL_430808 | Departamento de BiologÃ-a y genÃ©tica molecular, IACA Laboratorios. | Ã�ea de SecuenciaciÃ³n del Laboratorio de VirologÃ-a del Hospital de NiÃ±os Dr. Ricardo Gutierrez |
| EPI_ISL_430809 | Laboratorio de VirologÃ-a del Hospital de NiÃ±os Dr. Ricardo Gutierrez | Ã�ea de SecuenciaciÃ³n del Laboratorio de VirologÃ-a del Hospital de NiÃ±os Dr. Ricardo Gutierrez |
| EPI_ISL_430810 | Laboratorio de VirologÃ-a del Hospital de NiÃ±os Dr. Ricardo Gutierrez | Ã�ea de SecuenciaciÃ³n del Laboratorio de VirologÃ-a del Hospital de NiÃ±os Dr. Ricardo Gutierrez |
| EPI_ISL_430811 | Laboratorio de VirologÃ-a del Hospital de NiÃ±os Dr. Ricardo Gutierrez | Ã�ea de SecuenciaciÃ³n del Laboratorio de VirologÃ-a del Hospital de NiÃ±os Dr. Ricardo Gutierrez |
| EPI_ISL_430812 | Laboratorio de VirologÃ-a del Hospital de NiÃ±os Dr. Ricardo Gutierrez | Ã�ea de SecuenciaciÃ³n del Laboratorio de VirologÃ-a del Hospital de NiÃ±os Dr. Ricardo Gutierrez |
| EPI_ISL_430813 | Laboratorio de VirologÃ-a del Hospital de NiÃ±os Dr. Ricardo Gutierrez | Ã�ea de SecuenciaciÃ³n del Laboratorio de VirologÃ-a del Hospital de NiÃ±os Dr. Ricardo Gutierrez |
| EPI_ISL_430814 | Laboratorio de VirologÃ-a del Hospital de NiÃ±os Dr. Ricardo Gutierrez | Ã�ea de SecuenciaciÃ³n del Laboratorio de VirologÃ-a del Hospital de NiÃ±os Dr. Ricardo Gutierrez |
| EPI_ISL_430815 | Laboratorio de VirologÃ-a del Hospital de NiÃ±os Dr. Ricardo Gutierrez | Ã�ea de SecuenciaciÃ³n del Laboratorio de VirologÃ-a del Hospital de NiÃ±os Dr. Ricardo Gutierrez |
| EPI_ISL_430816 | Laboratorio de VirologÃ-a del Hospital de NiÃ±os Dr. Ricardo Gutierrez | Ã�ea de SecuenciaciÃ³n del Laboratorio de VirologÃ-a del Hospital de NiÃ±os Dr. Ricardo Gutierrez |
| EPI_ISL_430817 | Laboratorio de VirologÃ-a del Hospital de NiÃ±os Dr. Ricardo Gutierrez | Ã�ea de SecuenciaciÃ³n del Laboratorio de VirologÃ-a del Hospital de NiÃ±os Dr. Ricardo Gutierrez |
| EPI_ISL_430818 | Laboratorio de VirologÃ-a del Hospital de NiÃ±os Dr. Ricardo Gutierrez | Ã�ea de SecuenciaciÃ³n del Laboratorio de VirologÃ-a del Hospital de NiÃ±os Dr. Ricardo Gutierrez |
| EPI_ISL_430819 | Center of Scientific Excellence for Influenza Viruses,National Research Centre (NRC), Egypt. | Center of Scientific Excellence for Influenza Viruses,National Research Centre (NRC), Egypt. |
| EPI_ISL_430820 | Center of Scientific Excellence for Influenza Viruses, National Research Centre (NRC), Egypt. | Center of Scientific Excellence for Influenza Viruses, National Research Centre (NRC), Egypt. |
| EPI_ISL_430837 | n/a | Thai National Influenza Center, Department of medical Science, Ministry of Public Health, Thailand |
| EPI_ISL_430839 | Research Institute for Tropical Medicine | Research Institute for Tropical Medicine |
| EPI_ISL_430840 | Veterans Memorial Medical Center | Research Institute for Tropical Medicine |

| EPI_Accession_ID | Originating Laboratory | Submitting Laboratory |
|---|---|---|
| EPI_ISL_430841 | Praram 9 Hospital | National Institute of Health. Department of medical Sciences, Ministry of Public Health, Thailand |
| EPI_ISL_430842 | Central chest Institute of Thailand | National Institute of Health. Department of medical Sciences, Ministry of Public Health, Thailand |
| EPI_ISL_430843 | Bethany Hospital | Research Institute for Tropical Medicine |
| EPI_ISL_430844 | Lung Center of the Philippines | Research Institute for Tropical Medicine |
| EPI_ISL_430845 | Pasig City General Hospital | Research Institute for Tropical Medicine |
| EPI_ISL_430846 | General Intensive Care Unit, Raymond PoincarÃ© Hospital (AP-HP), Lab Inflammation & Infection, U1173 University Paris Saclay-UVSQ/INSERM, Garches, France. | Institut Pasteur, Laboratory for Urgent Response to biological Threats |
| EPI_ISL_430847 | HS mikrobiologi virus | The Public Health Agency of Sweden |
| EPI_ISL_430848 | Klinisk mikrobiologi och vardhygien Halmstad | The Public Health Agency of Sweden |
| EPI_ISL_430849 | Klinisk mikrobiologi och vardhygien Halmstad | The Public Health Agency of Sweden |
| EPI_ISL_430850 | Klinisk mikrobiologi och vardhygien Halmstad | The Public Health Agency of Sweden |
| EPI_ISL_430851 | Klinisk mikrobiologi och vardhygien Halmstad | The Public Health Agency of Sweden |
| EPI_ISL_430852 | Klinisk mikrobiologi och vardhygien Halmstad | The Public Health Agency of Sweden |
| EPI_ISL_430853 | Klinisk mikrobiologi och vardhygien Halmstad | The Public Health Agency of Sweden |
| EPI_ISL_430854 | Klinisk mikrobiologi och vardhygien Halmstad | The Public Health Agency of Sweden |
| EPI_ISL_430855 | Klinisk mikrobiologi och vardhygien Halmstad | The Public Health Agency of Sweden |
| EPI_ISL_430856 | Laboratoriemedicin | The Public Health Agency of Sweden |
| EPI_ISL_430857 | Laboratoriemedicin | The Public Health Agency of Sweden |
| EPI_ISL_430858 | Laboratoriemedicin | The Public Health Agency of Sweden |
| EPI_ISL_430859 | Laboratoriemedicin | The Public Health Agency of Sweden |
| EPI_ISL_430860 | Klinisk mikrobiologi Orebro | The Public Health Agency of Sweden |
| EPI_ISL_430861 | Klinisk mikrobiologi Orebro | The Public Health Agency of Sweden |
| EPI_ISL_430862 | The Public Health Agency of Sweden | The Public Health Agency of Sweden |
| EPI_ISL_430863 | Klinisk mikrobiologi Orebro | The Public Health Agency of Sweden |
| EPI_ISL_430864 | The Public Health Agency of Sweden | The Public Health Agency of Sweden |
| EPI_ISL_430867 | UW Virology Lab | UW Virology Lab |
| EPI_ISL_430868 | UW Virology Lab | UW Virology Lab |
| EPI_ISL_430869 | UW Virology Lab | UW Virology Lab |
| EPI_ISL_430870 | UW Virology Lab | UW Virology Lab |

| EPI_Accession_ID | Originating Laboratory | Submitting Laboratory |
|---|---|---|
| EPI_ISL_430871 | UW Virology Lab | UW Virology Lab |
| EPI_ISL_430872 | UW Virology Lab | UW Virology Lab |
| EPI_ISL_430873 | UW Virology Lab | UW Virology Lab |
| EPI_ISL_430874 | UW Virology Lab | UW Virology Lab |
| EPI_ISL_430875 | UW Virology Lab | UW Virology Lab |
| EPI_ISL_430876 | UW Virology Lab | UW Virology Lab |
| EPI_ISL_430877 | UW Virology Lab | UW Virology Lab |
| EPI_ISL_430878 | UW Virology Lab | UW Virology Lab |
| EPI_ISL_430879 | UW Virology Lab | UW Virology Lab |
| EPI_ISL_430880 | UW Virology Lab | UW Virology Lab |
| EPI_ISL_430881 | UW Virology Lab | UW Virology Lab |
| EPI_ISL_430882 | UW Virology Lab | UW Virology Lab |
| EPI_ISL_430883 | UW Virology Lab | UW Virology Lab |
| EPI_ISL_430884 | UW Virology Lab | UW Virology Lab |
| EPI_ISL_430885 | UW Virology Lab | UW Virology Lab |
| EPI_ISL_430886 | UW Virology Lab | UW Virology Lab |
| EPI_ISL_430887 | UW Virology Lab | UW Virology Lab |
| EPI_ISL_430888 | UW Virology Lab | UW Virology Lab |
| EPI_ISL_430889 | UW Virology Lab | UW Virology Lab |
| EPI_ISL_430890 | UW Virology Lab | UW Virology Lab |
| EPI_ISL_430891 | UW Virology Lab | UW Virology Lab |
| EPI_ISL_430892 | UW Virology Lab | UW Virology Lab |
| EPI_ISL_430893 | UW Virology Lab | UW Virology Lab |
| EPI_ISL_430894 | UW Virology Lab | UW Virology Lab |
| EPI_ISL_430895 | UW Virology Lab | UW Virology Lab |
| EPI_ISL_430896 | UW Virology Lab | UW Virology Lab |
| EPI_ISL_430897 | UW Virology Lab | UW Virology Lab |
| EPI_ISL_430898 | UW Virology Lab | UW Virology Lab |
| EPI_ISL_430899 | UW Virology Lab | UW Virology Lab |
| EPI_ISL_430900 | UW Virology Lab | UW Virology Lab |
| EPI_ISL_430901 | UW Virology Lab | UW Virology Lab |
| EPI_ISL_430902 | UW Virology Lab | UW Virology Lab |
| EPI_ISL_430903 | UW Virology Lab | UW Virology Lab |

| EPI_Accession_ID | Originating Laboratory | Submitting Laboratory |
|---|---|---|
| EPI_ISL_430904 | UW Virology Lab | UW Virology Lab |
| EPI_ISL_430905 | UW Virology Lab | UW Virology Lab |
| EPI_ISL_430906 | UW Virology Lab | UW Virology Lab |
| EPI_ISL_430907 | UW Virology Lab | UW Virology Lab |
| EPI_ISL_430908 | UW Virology Lab | UW Virology Lab |
| EPI_ISL_430909 | UW Virology Lab | UW Virology Lab |
| EPI_ISL_430910 | UW Virology Lab | UW Virology Lab |
| EPI_ISL_430911 | UW Virology Lab | UW Virology Lab |
| EPI_ISL_430912 | UW Virology Lab | UW Virology Lab |
| EPI_ISL_430913 | UW Virology Lab | UW Virology Lab |
| EPI_ISL_430914 | UW Virology Lab | UW Virology Lab |
| EPI_ISL_430915 | UW Virology Lab | UW Virology Lab |
| EPI_ISL_430916 | UW Virology Lab | UW Virology Lab |
| EPI_ISL_430917 | UW Virology Lab | UW Virology Lab |
| EPI_ISL_430918 | UW Virology Lab | UW Virology Lab |
| EPI_ISL_430919 | UW Virology Lab | UW Virology Lab |
| EPI_ISL_430920 | UW Virology Lab | UW Virology Lab |
| EPI_ISL_430921 | UW Virology Lab | UW Virology Lab |
| EPI_ISL_430922 | UW Virology Lab | UW Virology Lab |
| EPI_ISL_430923 | UW Virology Lab | UW Virology Lab |
| EPI_ISL_430924 | UW Virology Lab | UW Virology Lab |
| EPI_ISL_430925 | UW Virology Lab | UW Virology Lab |
| EPI_ISL_430926 | UW Virology Lab | UW Virology Lab |
| EPI_ISL_430927 | UW Virology Lab | UW Virology Lab |
| EPI_ISL_430928 | UW Virology Lab | UW Virology Lab |
| EPI_ISL_430929 | UW Virology Lab | UW Virology Lab |
| EPI_ISL_430930 | UW Virology Lab | UW Virology Lab |
| EPI_ISL_430931 | UW Virology Lab | UW Virology Lab |
| EPI_ISL_430932 | UW Virology Lab | UW Virology Lab |
| EPI_ISL_430933 | UW Virology Lab | UW Virology Lab |
| EPI_ISL_430934 | UW Virology Lab | UW Virology Lab |
| EPI_ISL_430935 | UW Virology Lab | UW Virology Lab |
| EPI_ISL_430936 | UW Virology Lab | UW Virology Lab |

| EPI_Accession_ID | Originating Laboratory | Submitting Laboratory |
|---|---|---|
| EPI_ISL_430937 | UW Virology Lab | UW Virology Lab |
| EPI_ISL_430938 | UW Virology Lab | UW Virology Lab |
| EPI_ISL_430939 | UW Virology Lab | UW Virology Lab |
| EPI_ISL_430940 | UW Virology Lab | UW Virology Lab |
| EPI_ISL_430941 | UW Virology Lab | UW Virology Lab |
| EPI_ISL_430942 | UW Virology Lab | UW Virology Lab |
| EPI_ISL_430943 | UW Virology Lab | UW Virology Lab |
| EPI_ISL_430944 | UW Virology Lab | UW Virology Lab |
| EPI_ISL_430945 | UW Virology Lab | UW Virology Lab |
| EPI_ISL_430946 | UW Virology Lab | UW Virology Lab |
| EPI_ISL_430947 | UW Virology Lab | UW Virology Lab |
| EPI_ISL_430948 | UW Virology Lab | UW Virology Lab |
| EPI_ISL_430949 | UW Virology Lab | UW Virology Lab |
| EPI_ISL_430950 | UW Virology Lab | UW Virology Lab |
| EPI_ISL_430951 | UW Virology Lab | UW Virology Lab |
| EPI_ISL_430952 | UW Virology Lab | UW Virology Lab |
| EPI_ISL_430953 | UW Virology Lab | UW Virology Lab |
| EPI_ISL_430954 | UW Virology Lab | UW Virology Lab |
| EPI_ISL_430955 | UW Virology Lab | UW Virology Lab |
| EPI_ISL_430956 | UW Virology Lab | UW Virology Lab |
| EPI_ISL_430957 | UW Virology Lab | UW Virology Lab |
| EPI_ISL_430958 | UW Virology Lab | UW Virology Lab |
| EPI_ISL_430959 | UW Virology Lab | UW Virology Lab |
| EPI_ISL_430960 | UW Virology Lab | UW Virology Lab |
| EPI_ISL_430961 | UW Virology Lab | UW Virology Lab |
| EPI_ISL_430962 | UW Virology Lab | UW Virology Lab |
| EPI_ISL_430963 | UW Virology Lab | UW Virology Lab |
| EPI_ISL_430964 | UW Virology Lab | UW Virology Lab |
| EPI_ISL_430965 | UW Virology Lab | UW Virology Lab |
| EPI_ISL_430966 | UW Virology Lab | UW Virology Lab |
| EPI_ISL_430967 | UW Virology Lab | UW Virology Lab |
| EPI_ISL_430968 | UW Virology Lab | UW Virology Lab |
| EPI_ISL_430969 | UW Virology Lab | UW Virology Lab |

| EPI_Accession_ID | Originating Laboratory | Submitting Laboratory |
|---|---|---|
| EPI_ISL_430970 | UW Virology Lab | UW Virology Lab |
| EPI_ISL_430971 | UW Virology Lab | UW Virology Lab |
| EPI_ISL_430972 | UW Virology Lab | UW Virology Lab |
| EPI_ISL_430973 | UW Virology Lab | UW Virology Lab |
| EPI_ISL_430974 | UW Virology Lab | UW Virology Lab |
| EPI_ISL_430975 | UW Virology Lab | UW Virology Lab |
| EPI_ISL_430976 | UW Virology Lab | UW Virology Lab |
| EPI_ISL_430977 | UW Virology Lab | UW Virology Lab |
| EPI_ISL_430978 | UW Virology Lab | UW Virology Lab |
| EPI_ISL_430979 | UW Virology Lab | UW Virology Lab |
| EPI_ISL_430980 | UW Virology Lab | UW Virology Lab |
| EPI_ISL_431011 | Viral Respiratory Lab, National Institute for Biomedical Research (INRB) | Pathogen Sequencing Lab, National Institute for Biomedical Research (INRB) |
| EPI_ISL_431012 | Viral Respiratory Lab, National Institute for Biomedical Research (INRB) | Pathogen Sequencing Lab, National Institute for Biomedical Research (INRB) |
| EPI_ISL_431013 | Alaska State Virology Laboratory | Alaska State Virology Laboratory |
| EPI_ISL_431014 | Alaska State Virology Laboratory | Alaska State Virology Laboratory |
| EPI_ISL_431015 | Alaska State Virology Laboratory | Alaska State Virology Laboratory |
| EPI_ISL_431016 | Alaska State Virology Laboratory | Alaska State Virology Laboratory |
| EPI_ISL_431017 | Alaska State Virology Laboratory | Alaska State Virology Laboratory |
| EPI_ISL_431018 | Alaska State Virology Laboratory | Alaska State Virology Laboratory |
| EPI_ISL_431019 | Alaska State Virology Laboratory | Alaska State Virology Laboratory |
| EPI_ISL_431080 | Yale COVID-19 Biorepository | Grubaugh Lab - Yale School of Public Health |
| EPI_ISL_431081 | Yale COVID-19 Biorepository | Grubaugh Lab - Yale School of Public Health |
| EPI_ISL_431082 | Yale COVID-19 Biorepository | Grubaugh Lab - Yale School of Public Health |
| EPI_ISL_431083 | Yale COVID-19 Biorepository | Grubaugh Lab - Yale School of Public Health |
| EPI_ISL_431084 | Yale COVID-19 Biorepository | Grubaugh Lab - Yale School of Public Health |
| EPI_ISL_431085 | Yale COVID-19 Biorepository | Grubaugh Lab - Yale School of Public Health |
| EPI_ISL_431086 | Yale COVID-19 Biorepository | Grubaugh Lab - Yale School of Public Health |
| EPI_ISL_431087 | Yale COVID-19 Biorepository | Grubaugh Lab - Yale School of Public Health |
| EPI_ISL_431088 | Yale COVID-19 Biorepository | Grubaugh Lab - Yale School of Public Health |
| EPI_ISL_431089 | Yale COVID-19 Biorepository | Grubaugh Lab - Yale School of Public Health |
| EPI_ISL_431090 | Yale COVID-19 Biorepository | Grubaugh Lab - Yale School of Public Health |

| EPI_Accession_ID | Originating Laboratory | Submitting Laboratory |
|---|---|---|
| EPI_ISL_431091 | Yale COVID-19 Biorepository | Grubaugh Lab - Yale School of Public Health |
| EPI_ISL_431092 | Yale COVID-19 Biorepository | Grubaugh Lab - Yale School of Public Health |
| EPI_ISL_431093 | Yale COVID-19 Biorepository | Grubaugh Lab - Yale School of Public Health |
| EPI_ISL_431094 | Yale COVID-19 Biorepository | Grubaugh Lab - Yale School of Public Health |
| EPI_ISL_431095 | Yale COVID-19 Biorepository | Grubaugh Lab - Yale School of Public Health |
| EPI_ISL_431096 | Yale COVID-19 Biorepository | Grubaugh Lab - Yale School of Public Health |
| EPI_ISL_431101 | Department of Microbiology,Gandhi Medical College and Hospital | Virus Research Laboratory, Department of Zoology, Osmania University,Hyderabad,India |
| EPI_ISL_431102 | Department of MicroBiology,Gandhi Medical College and Hospital,Secendrabad,Hyderabad,India | Department of Microbiology, Gandhi Medical College and Hospital, Secendrabad, Hyderabad |
| EPI_ISL_431103 | Department of Microbiology, Gandhi Medical College and Hospital, Secendrabad, Hyderabad, India | Department of Microbiology, Gandhi Medical College and Hospital, Secendrabad, Hyderabad, India |
| EPI_ISL_431117 | Department of Microbiology, Gandhi Medical College and Hospital, Secendrabad, Hyderabad, India | Department of Microbiology, Gandhi Medical College and Hospital, Secendrabad, Hyderabad, India |
| EPI_ISL_431118 | Fujian Center for Disease Control and Prevention | Fujian Center for Disease Control and Prevention |
| EPI_ISL_431180 | Fujian Center for Disease Control and Prevention | Fujian Center for Disease Control and Prevention |
| EPI_ISL_431240 | Fujian Center for Disease Control and Prevention | Fujian Center for Disease Control and Prevention |
| EPI_ISL_431778 | unknown | Virology, Wageningen Bioveterinary Research |
| EPI_ISL_431779 | Fujian Center for Disease Control and Prevention | Fujian Center for Disease Control and Prevention |
| EPI_ISL_431780 | Fujian Center for Disease Control and Prevention | Fujian Center for Disease Control and Prevention |
| EPI_ISL_431781 | Fujian Center for Disease Control and Prevention | Fujian Center for Disease Control and Prevention |
| EPI_ISL_431782 | Fujian Center for Disease Control and Prevention | Fujian Center for Disease Control and Prevention |
| EPI_ISL_431783 | Fujian Center for Disease Control and Prevention | Fujian Center for Disease Control and Prevention |
| EPI_ISL_431785 | Fujian Center for Disease Control and Prevention | Fujian Center for Disease Control and Prevention |
| EPI_ISL_431833 | National Institutes of Health, University of the Philippines Manila | Philippine Genome Center, University of the Philippines System |
| EPI_ISL_431901 | Wales Specialist Virology Centre | Public Health Wales Microbiology Cardiff |
| EPI_ISL_431902 | Wales Specialist Virology Centre | Public Health Wales Microbiology Cardiff |
| EPI_ISL_431903 | Wales Specialist Virology Centre | Public Health Wales Microbiology Cardiff |
| EPI_ISL_431904 | Wales Specialist Virology Centre | Public Health Wales Microbiology Cardiff |
| EPI_ISL_431905 | Wales Specialist Virology Centre | Public Health Wales Microbiology Cardiff |
| EPI_ISL_431906 | Wales Specialist Virology Centre | Public Health Wales Microbiology Cardiff |
| EPI_ISL_431907 | Wales Specialist Virology Centre | Public Health Wales Microbiology Cardiff |

| EPI_Accession_ID | Originating Laboratory | Submitting Laboratory |
|---|---|---|
| EPI_ISL_431908 | Wales Specialist Virology Centre | Public Health Wales Microbiology Cardiff |
| EPI_ISL_431909 | Wales Specialist Virology Centre | Public Health Wales Microbiology Cardiff |
| EPI_ISL_431910 | Wales Specialist Virology Centre | Public Health Wales Microbiology Cardiff |
| EPI_ISL_431911 | Wales Specialist Virology Centre | Public Health Wales Microbiology Cardiff |
| EPI_ISL_431912 | Wales Specialist Virology Centre | Public Health Wales Microbiology Cardiff |
| EPI_ISL_431913 | Wales Specialist Virology Centre | Public Health Wales Microbiology Cardiff |
| EPI_ISL_431914 | Wales Specialist Virology Centre | Public Health Wales Microbiology Cardiff |
| EPI_ISL_431916 | Wales Specialist Virology Centre | Public Health Wales Microbiology Cardiff |
| EPI_ISL_431917 | Wales Specialist Virology Centre | Public Health Wales Microbiology Cardiff |
| EPI_ISL_431918 | Wales Specialist Virology Centre | Public Health Wales Microbiology Cardiff |
| EPI_ISL_431919 | Wales Specialist Virology Centre | Public Health Wales Microbiology Cardiff |
| EPI_ISL_431920 | Wales Specialist Virology Centre | Public Health Wales Microbiology Cardiff |
| EPI_ISL_431921 | Wales Specialist Virology Centre | Public Health Wales Microbiology Cardiff |
| EPI_ISL_431922 | Wales Specialist Virology Centre | Public Health Wales Microbiology Cardiff |
| EPI_ISL_431923 | Wales Specialist Virology Centre | Public Health Wales Microbiology Cardiff |
| EPI_ISL_431924 | Wales Specialist Virology Centre | Public Health Wales Microbiology Cardiff |
| EPI_ISL_431925 | Wales Specialist Virology Centre | Public Health Wales Microbiology Cardiff |
| EPI_ISL_431926 | Wales Specialist Virology Centre | Public Health Wales Microbiology Cardiff |
| EPI_ISL_431927 | Wales Specialist Virology Centre | Public Health Wales Microbiology Cardiff |
| EPI_ISL_431928 | Wales Specialist Virology Centre | Public Health Wales Microbiology Cardiff |
| EPI_ISL_431929 | Wales Specialist Virology Centre | Public Health Wales Microbiology Cardiff |
| EPI_ISL_431930 | Wales Specialist Virology Centre | Public Health Wales Microbiology Cardiff |
| EPI_ISL_431931 | Wales Specialist Virology Centre | Public Health Wales Microbiology Cardiff |
| EPI_ISL_431932 | Wales Specialist Virology Centre | Public Health Wales Microbiology Cardiff |
| EPI_ISL_431933 | Wales Specialist Virology Centre | Public Health Wales Microbiology Cardiff |
| EPI_ISL_431934 | Wales Specialist Virology Centre | Public Health Wales Microbiology Cardiff |
| EPI_ISL_431935 | Wales Specialist Virology Centre | Public Health Wales Microbiology Cardiff |
| EPI_ISL_431936 | Wales Specialist Virology Centre | Public Health Wales Microbiology Cardiff |
| EPI_ISL_431937 | Wales Specialist Virology Centre | Public Health Wales Microbiology Cardiff |
| EPI_ISL_431938 | Wales Specialist Virology Centre | Public Health Wales Microbiology Cardiff |
| EPI_ISL_431939 | Wales Specialist Virology Centre | Public Health Wales Microbiology Cardiff |
| EPI_ISL_431940 | Wales Specialist Virology Centre | Public Health Wales Microbiology Cardiff |
| EPI_ISL_431941 | Wales Specialist Virology Centre | Public Health Wales Microbiology Cardiff |

| EPI_Accession_ID | Originating Laboratory | Submitting Laboratory |
|---|---|---|
| EPI_ISL_431942 | Wales Specialist Virology Centre | Public Health Wales Microbiology Cardiff |
| EPI_ISL_431943 | Wales Specialist Virology Centre | Public Health Wales Microbiology Cardiff |
| EPI_ISL_431944 | Wales Specialist Virology Centre | Public Health Wales Microbiology Cardiff |
| EPI_ISL_431945 | Wales Specialist Virology Centre | Public Health Wales Microbiology Cardiff |
| EPI_ISL_431946 | Wales Specialist Virology Centre | Public Health Wales Microbiology Cardiff |
| EPI_ISL_431947 | Wales Specialist Virology Centre | Public Health Wales Microbiology Cardiff |
| EPI_ISL_431948 | Wales Specialist Virology Centre | Public Health Wales Microbiology Cardiff |
| EPI_ISL_431949 | Wales Specialist Virology Centre | Public Health Wales Microbiology Cardiff |
| EPI_ISL_431950 | Wales Specialist Virology Centre | Public Health Wales Microbiology Cardiff |
| EPI_ISL_431951 | Wales Specialist Virology Centre | Public Health Wales Microbiology Cardiff |
| EPI_ISL_431952 | Wales Specialist Virology Centre | Public Health Wales Microbiology Cardiff |
| EPI_ISL_431953 | Wales Specialist Virology Centre | Public Health Wales Microbiology Cardiff |
| EPI_ISL_431954 | Wales Specialist Virology Centre | Public Health Wales Microbiology Cardiff |
| EPI_ISL_431955 | Wales Specialist Virology Centre | Public Health Wales Microbiology Cardiff |
| EPI_ISL_431956 | Wales Specialist Virology Centre | Public Health Wales Microbiology Cardiff |
| EPI_ISL_431957 | Wales Specialist Virology Centre | Public Health Wales Microbiology Cardiff |
| EPI_ISL_431958 | Wales Specialist Virology Centre | Public Health Wales Microbiology Cardiff |
| EPI_ISL_431959 | Wales Specialist Virology Centre | Public Health Wales Microbiology Cardiff |
| EPI_ISL_431960 | Wales Specialist Virology Centre | Public Health Wales Microbiology Cardiff |
| EPI_ISL_431961 | Wales Specialist Virology Centre | Public Health Wales Microbiology Cardiff |
| EPI_ISL_431962 | Wales Specialist Virology Centre | Public Health Wales Microbiology Cardiff |
| EPI_ISL_431963 | Wales Specialist Virology Centre | Public Health Wales Microbiology Cardiff |
| EPI_ISL_431964 | Wales Specialist Virology Centre | Public Health Wales Microbiology Cardiff |
| EPI_ISL_431965 | Wales Specialist Virology Centre | Public Health Wales Microbiology Cardiff |
| EPI_ISL_431966 | Wales Specialist Virology Centre | Public Health Wales Microbiology Cardiff |
| EPI_ISL_431967 | Wales Specialist Virology Centre | Public Health Wales Microbiology Cardiff |
| EPI_ISL_431968 | Wales Specialist Virology Centre | Public Health Wales Microbiology Cardiff |
| EPI_ISL_431971 | Wales Specialist Virology Centre | Public Health Wales Microbiology Cardiff |
| EPI_ISL_431972 | Wales Specialist Virology Centre | Public Health Wales Microbiology Cardiff |
| EPI_ISL_431973 | Wales Specialist Virology Centre | Public Health Wales Microbiology Cardiff |
| EPI_ISL_431974 | Wales Specialist Virology Centre | Public Health Wales Microbiology Cardiff |
| EPI_ISL_431975 | Wales Specialist Virology Centre | Public Health Wales Microbiology Cardiff |
| EPI_ISL_431976 | Wales Specialist Virology Centre | Public Health Wales Microbiology Cardiff |

| EPI_Accession_ID | Originating Laboratory | Submitting Laboratory |
|---|---|---|
| EPI_ISL_431977 | Wales Specialist Virology Centre | Public Health Wales Microbiology Cardiff |
| EPI_ISL_431978 | Wales Specialist Virology Centre | Public Health Wales Microbiology Cardiff |
| EPI_ISL_431980 | Wales Specialist Virology Centre | Public Health Wales Microbiology Cardiff |
| EPI_ISL_431981 | Wales Specialist Virology Centre | Public Health Wales Microbiology Cardiff |
| EPI_ISL_431982 | Wales Specialist Virology Centre | Public Health Wales Microbiology Cardiff |
| EPI_ISL_431983 | Wales Specialist Virology Centre | Public Health Wales Microbiology Cardiff |
| EPI_ISL_431984 | Wales Specialist Virology Centre | Public Health Wales Microbiology Cardiff |
| EPI_ISL_431985 | Wales Specialist Virology Centre | Public Health Wales Microbiology Cardiff |
| EPI_ISL_431986 | Wales Specialist Virology Centre | Public Health Wales Microbiology Cardiff |
| EPI_ISL_431987 | Wales Specialist Virology Centre | Public Health Wales Microbiology Cardiff |
| EPI_ISL_431988 | Wales Specialist Virology Centre | Public Health Wales Microbiology Cardiff |
| EPI_ISL_431989 | Wales Specialist Virology Centre | Public Health Wales Microbiology Cardiff |
| EPI_ISL_431990 | Wales Specialist Virology Centre | Public Health Wales Microbiology Cardiff |
| EPI_ISL_431991 | Wales Specialist Virology Centre | Public Health Wales Microbiology Cardiff |
| EPI_ISL_431992 | Wales Specialist Virology Centre | Public Health Wales Microbiology Cardiff |
| EPI_ISL_431994 | Wales Specialist Virology Centre | Public Health Wales Microbiology Cardiff |
| EPI_ISL_431995 | Wales Specialist Virology Centre | Public Health Wales Microbiology Cardiff |
| EPI_ISL_431996 | Wales Specialist Virology Centre | Public Health Wales Microbiology Cardiff |
| EPI_ISL_431997 | Wales Specialist Virology Centre | Public Health Wales Microbiology Cardiff |
| EPI_ISL_431998 | Wales Specialist Virology Centre | Public Health Wales Microbiology Cardiff |
| EPI_ISL_431999 | Wales Specialist Virology Centre | Public Health Wales Microbiology Cardiff |
| EPI_ISL_432000 | Wales Specialist Virology Centre | Public Health Wales Microbiology Cardiff |
| EPI_ISL_432001 | Wales Specialist Virology Centre | Public Health Wales Microbiology Cardiff |
| EPI_ISL_432002 | Wales Specialist Virology Centre | Public Health Wales Microbiology Cardiff |
| EPI_ISL_432003 | Wales Specialist Virology Centre | Public Health Wales Microbiology Cardiff |
| EPI_ISL_432004 | Wales Specialist Virology Centre | Public Health Wales Microbiology Cardiff |
| EPI_ISL_432005 | Wales Specialist Virology Centre | Public Health Wales Microbiology Cardiff |
| EPI_ISL_432006 | Wales Specialist Virology Centre | Public Health Wales Microbiology Cardiff |
| EPI_ISL_432007 | Wales Specialist Virology Centre | Public Health Wales Microbiology Cardiff |
| EPI_ISL_432008 | Wales Specialist Virology Centre | Public Health Wales Microbiology Cardiff |
| EPI_ISL_432009 | Wales Specialist Virology Centre | Public Health Wales Microbiology Cardiff |
| EPI_ISL_432010 | Wales Specialist Virology Centre | Public Health Wales Microbiology Cardiff |
| EPI_ISL_432011 | Wales Specialist Virology Centre | Public Health Wales Microbiology Cardiff |

| EPI_Accession_ID | Originating Laboratory | Submitting Laboratory |
|---|---|---|
| EPI_ISL_432012 | Wales Specialist Virology Centre | Public Health Wales Microbiology Cardiff |
| EPI_ISL_432013 | Wales Specialist Virology Centre | Public Health Wales Microbiology Cardiff |
| EPI_ISL_432014 | Wales Specialist Virology Centre | Public Health Wales Microbiology Cardiff |
| EPI_ISL_432015 | Wales Specialist Virology Centre | Public Health Wales Microbiology Cardiff |
| EPI_ISL_432016 | Wales Specialist Virology Centre | Public Health Wales Microbiology Cardiff |
| EPI_ISL_432017 | Wales Specialist Virology Centre | Public Health Wales Microbiology Cardiff |
| EPI_ISL_432018 | Wales Specialist Virology Centre | Public Health Wales Microbiology Cardiff |
| EPI_ISL_432019 | Wales Specialist Virology Centre | Public Health Wales Microbiology Cardiff |
| EPI_ISL_432021 | Wales Specialist Virology Centre | Public Health Wales Microbiology Cardiff |
| EPI_ISL_432022 | Wales Specialist Virology Centre | Public Health Wales Microbiology Cardiff |
| EPI_ISL_432023 | Wales Specialist Virology Centre | Public Health Wales Microbiology Cardiff |
| EPI_ISL_432024 | Wales Specialist Virology Centre | Public Health Wales Microbiology Cardiff |
| EPI_ISL_432025 | Wales Specialist Virology Centre | Public Health Wales Microbiology Cardiff |
| EPI_ISL_432026 | Wales Specialist Virology Centre | Public Health Wales Microbiology Cardiff |
| EPI_ISL_432027 | Wales Specialist Virology Centre | Public Health Wales Microbiology Cardiff |
| EPI_ISL_432028 | Wales Specialist Virology Centre | Public Health Wales Microbiology Cardiff |
| EPI_ISL_432029 | Wales Specialist Virology Centre | Public Health Wales Microbiology Cardiff |
| EPI_ISL_432030 | Wales Specialist Virology Centre | Public Health Wales Microbiology Cardiff |
| EPI_ISL_432031 | Wales Specialist Virology Centre | Public Health Wales Microbiology Cardiff |
| EPI_ISL_432032 | Wales Specialist Virology Centre | Public Health Wales Microbiology Cardiff |
| EPI_ISL_432033 | Wales Specialist Virology Centre | Public Health Wales Microbiology Cardiff |
| EPI_ISL_432034 | Wales Specialist Virology Centre | Public Health Wales Microbiology Cardiff |
| EPI_ISL_432035 | Wales Specialist Virology Centre | Public Health Wales Microbiology Cardiff |
| EPI_ISL_432036 | Wales Specialist Virology Centre | Public Health Wales Microbiology Cardiff |
| EPI_ISL_432037 | Wales Specialist Virology Centre | Public Health Wales Microbiology Cardiff |
| EPI_ISL_432038 | Wales Specialist Virology Centre | Public Health Wales Microbiology Cardiff |
| EPI_ISL_432039 | Wales Specialist Virology Centre | Public Health Wales Microbiology Cardiff |
| EPI_ISL_432040 | Wales Specialist Virology Centre | Public Health Wales Microbiology Cardiff |
| EPI_ISL_432041 | Wales Specialist Virology Centre | Public Health Wales Microbiology Cardiff |
| EPI_ISL_432042 | Wales Specialist Virology Centre | Public Health Wales Microbiology Cardiff |
| EPI_ISL_432043 | Wales Specialist Virology Centre | Public Health Wales Microbiology Cardiff |
| EPI_ISL_432044 | Wales Specialist Virology Centre | Public Health Wales Microbiology Cardiff |
| EPI_ISL_432045 | Wales Specialist Virology Centre | Public Health Wales Microbiology Cardiff |

| EPI_Accession_ID | Originating Laboratory | Submitting Laboratory |
|---|---|---|
| EPI_ISL_432046 | Wales Specialist Virology Centre | Public Health Wales Microbiology Cardiff |
| EPI_ISL_432047 | Wales Specialist Virology Centre | Public Health Wales Microbiology Cardiff |
| EPI_ISL_432048 | Wales Specialist Virology Centre | Public Health Wales Microbiology Cardiff |
| EPI_ISL_432049 | Wales Specialist Virology Centre | Public Health Wales Microbiology Cardiff |
| EPI_ISL_432050 | Wales Specialist Virology Centre | Public Health Wales Microbiology Cardiff |
| EPI_ISL_432051 | Wales Specialist Virology Centre | Public Health Wales Microbiology Cardiff |
| EPI_ISL_432052 | Wales Specialist Virology Centre | Public Health Wales Microbiology Cardiff |
| EPI_ISL_432053 | Wales Specialist Virology Centre | Public Health Wales Microbiology Cardiff |
| EPI_ISL_432054 | Wales Specialist Virology Centre | Public Health Wales Microbiology Cardiff |
| EPI_ISL_432055 | Wales Specialist Virology Centre | Public Health Wales Microbiology Cardiff |
| EPI_ISL_432056 | Wales Specialist Virology Centre | Public Health Wales Microbiology Cardiff |
| EPI_ISL_432057 | Wales Specialist Virology Centre | Public Health Wales Microbiology Cardiff |
| EPI_ISL_432058 | Wales Specialist Virology Centre | Public Health Wales Microbiology Cardiff |
| EPI_ISL_432059 | Wales Specialist Virology Centre | Public Health Wales Microbiology Cardiff |
| EPI_ISL_432062 | Wales Specialist Virology Centre | Public Health Wales Microbiology Cardiff |
| EPI_ISL_432065 | Wales Specialist Virology Centre | Public Health Wales Microbiology Cardiff |
| EPI_ISL_432066 | Wales Specialist Virology Centre | Public Health Wales Microbiology Cardiff |
| EPI_ISL_432067 | Wales Specialist Virology Centre | Public Health Wales Microbiology Cardiff |
| EPI_ISL_432068 | Wales Specialist Virology Centre | Public Health Wales Microbiology Cardiff |
| EPI_ISL_432069 | Wales Specialist Virology Centre | Public Health Wales Microbiology Cardiff |
| EPI_ISL_432070 | Wales Specialist Virology Centre | Public Health Wales Microbiology Cardiff |
| EPI_ISL_432071 | Wales Specialist Virology Centre | Public Health Wales Microbiology Cardiff |
| EPI_ISL_432072 | Wales Specialist Virology Centre | Public Health Wales Microbiology Cardiff |
| EPI_ISL_432073 | Wales Specialist Virology Centre | Public Health Wales Microbiology Cardiff |
| EPI_ISL_432074 | Wales Specialist Virology Centre | Public Health Wales Microbiology Cardiff |
| EPI_ISL_432075 | Wales Specialist Virology Centre | Public Health Wales Microbiology Cardiff |
| EPI_ISL_432077 | Wales Specialist Virology Centre | Public Health Wales Microbiology Cardiff |
| EPI_ISL_432078 | Wales Specialist Virology Centre | Public Health Wales Microbiology Cardiff |
| EPI_ISL_432079 | Wales Specialist Virology Centre | Public Health Wales Microbiology Cardiff |
| EPI_ISL_432080 | Wales Specialist Virology Centre | Public Health Wales Microbiology Cardiff |
| EPI_ISL_432081 | Wales Specialist Virology Centre | Public Health Wales Microbiology Cardiff |
| EPI_ISL_432082 | Wales Specialist Virology Centre | Public Health Wales Microbiology Cardiff |
| EPI_ISL_432083 | Wales Specialist Virology Centre | Public Health Wales Microbiology Cardiff |

| EPI_Accession_ID | Originating Laboratory | Submitting Laboratory |
|---|---|---|
| EPI_ISL_432084 | Wales Specialist Virology Centre | Public Health Wales Microbiology Cardiff |
| EPI_ISL_432085 | Wales Specialist Virology Centre | Public Health Wales Microbiology Cardiff |
| EPI_ISL_432086 | Wales Specialist Virology Centre | Public Health Wales Microbiology Cardiff |
| EPI_ISL_432088 | Wales Specialist Virology Centre | Public Health Wales Microbiology Cardiff |
| EPI_ISL_432089 | Wales Specialist Virology Centre | Public Health Wales Microbiology Cardiff |
| EPI_ISL_432090 | Wales Specialist Virology Centre | Public Health Wales Microbiology Cardiff |
| EPI_ISL_432091 | Wales Specialist Virology Centre | Public Health Wales Microbiology Cardiff |
| EPI_ISL_432092 | Wales Specialist Virology Centre | Public Health Wales Microbiology Cardiff |
| EPI_ISL_432093 | Wales Specialist Virology Centre | Public Health Wales Microbiology Cardiff |
| EPI_ISL_432094 | Wales Specialist Virology Centre | Public Health Wales Microbiology Cardiff |
| EPI_ISL_432095 | Wales Specialist Virology Centre | Public Health Wales Microbiology Cardiff |
| EPI_ISL_432096 | Wales Specialist Virology Centre | Public Health Wales Microbiology Cardiff |
| EPI_ISL_432097 | Wales Specialist Virology Centre | Public Health Wales Microbiology Cardiff |
| EPI_ISL_432098 | Wales Specialist Virology Centre | Public Health Wales Microbiology Cardiff |
| EPI_ISL_432099 | Wales Specialist Virology Centre | Public Health Wales Microbiology Cardiff |
| EPI_ISL_432100 | Wales Specialist Virology Centre | Public Health Wales Microbiology Cardiff |
| EPI_ISL_432103 | Wales Specialist Virology Centre | Public Health Wales Microbiology Cardiff |
| EPI_ISL_432105 | Wales Specialist Virology Centre | Public Health Wales Microbiology Cardiff |
| EPI_ISL_432106 | Wales Specialist Virology Centre | Public Health Wales Microbiology Cardiff |
| EPI_ISL_432107 | Wales Specialist Virology Centre | Public Health Wales Microbiology Cardiff |
| EPI_ISL_432108 | Wales Specialist Virology Centre | Public Health Wales Microbiology Cardiff |
| EPI_ISL_432109 | Wales Specialist Virology Centre | Public Health Wales Microbiology Cardiff |
| EPI_ISL_432110 | Wales Specialist Virology Centre | Public Health Wales Microbiology Cardiff |
| EPI_ISL_432111 | Wales Specialist Virology Centre | Public Health Wales Microbiology Cardiff |
| EPI_ISL_432112 | Wales Specialist Virology Centre | Public Health Wales Microbiology Cardiff |
| EPI_ISL_432113 | Wales Specialist Virology Centre | Public Health Wales Microbiology Cardiff |
| EPI_ISL_432115 | Wales Specialist Virology Centre | Public Health Wales Microbiology Cardiff |
| EPI_ISL_432116 | Wales Specialist Virology Centre | Public Health Wales Microbiology Cardiff |
| EPI_ISL_432117 | Wales Specialist Virology Centre | Public Health Wales Microbiology Cardiff |
| EPI_ISL_432118 | Wales Specialist Virology Centre | Public Health Wales Microbiology Cardiff |
| EPI_ISL_432119 | Wales Specialist Virology Centre | Public Health Wales Microbiology Cardiff |
| EPI_ISL_432120 | Wales Specialist Virology Centre | Public Health Wales Microbiology Cardiff |
| EPI_ISL_432121 | Wales Specialist Virology Centre | Public Health Wales Microbiology Cardiff |

| EPI_Accession_ID | Originating Laboratory | Submitting Laboratory |
|---|---|---|
| EPI_ISL_432122 | Wales Specialist Virology Centre | Public Health Wales Microbiology Cardiff |
| EPI_ISL_432123 | Wales Specialist Virology Centre | Public Health Wales Microbiology Cardiff |
| EPI_ISL_432124 | Wales Specialist Virology Centre | Public Health Wales Microbiology Cardiff |
| EPI_ISL_432125 | Wales Specialist Virology Centre | Public Health Wales Microbiology Cardiff |
| EPI_ISL_432126 | Wales Specialist Virology Centre | Public Health Wales Microbiology Cardiff |
| EPI_ISL_432127 | Wales Specialist Virology Centre | Public Health Wales Microbiology Cardiff |
| EPI_ISL_432128 | Wales Specialist Virology Centre | Public Health Wales Microbiology Cardiff |
| EPI_ISL_432129 | Wales Specialist Virology Centre | Public Health Wales Microbiology Cardiff |
| EPI_ISL_432130 | Wales Specialist Virology Centre | Public Health Wales Microbiology Cardiff |
| EPI_ISL_432131 | Wales Specialist Virology Centre | Public Health Wales Microbiology Cardiff |
| EPI_ISL_432132 | Wales Specialist Virology Centre | Public Health Wales Microbiology Cardiff |
| EPI_ISL_432133 | Wales Specialist Virology Centre | Public Health Wales Microbiology Cardiff |
| EPI_ISL_432134 | Wales Specialist Virology Centre | Public Health Wales Microbiology Cardiff |
| EPI_ISL_432135 | Wales Specialist Virology Centre | Public Health Wales Microbiology Cardiff |
| EPI_ISL_432136 | Wales Specialist Virology Centre | Public Health Wales Microbiology Cardiff |
| EPI_ISL_432137 | Wales Specialist Virology Centre | Public Health Wales Microbiology Cardiff |
| EPI_ISL_432138 | Wales Specialist Virology Centre | Public Health Wales Microbiology Cardiff |
| EPI_ISL_432139 | Wales Specialist Virology Centre | Public Health Wales Microbiology Cardiff |
| EPI_ISL_432140 | Wales Specialist Virology Centre | Public Health Wales Microbiology Cardiff |
| EPI_ISL_432141 | Wales Specialist Virology Centre | Public Health Wales Microbiology Cardiff |
| EPI_ISL_432142 | Wales Specialist Virology Centre | Public Health Wales Microbiology Cardiff |
| EPI_ISL_432143 | Wales Specialist Virology Centre | Public Health Wales Microbiology Cardiff |
| EPI_ISL_432144 | Wales Specialist Virology Centre | Public Health Wales Microbiology Cardiff |
| EPI_ISL_432145 | Wales Specialist Virology Centre | Public Health Wales Microbiology Cardiff |
| EPI_ISL_432146 | Wales Specialist Virology Centre | Public Health Wales Microbiology Cardiff |
| EPI_ISL_432147 | Wales Specialist Virology Centre | Public Health Wales Microbiology Cardiff |
| EPI_ISL_432148 | Wales Specialist Virology Centre | Public Health Wales Microbiology Cardiff |
| EPI_ISL_432150 | Wales Specialist Virology Centre | Public Health Wales Microbiology Cardiff |
| EPI_ISL_432151 | Wales Specialist Virology Centre | Public Health Wales Microbiology Cardiff |
| EPI_ISL_432152 | Wales Specialist Virology Centre | Public Health Wales Microbiology Cardiff |
| EPI_ISL_432153 | Wales Specialist Virology Centre | Public Health Wales Microbiology Cardiff |
| EPI_ISL_432154 | Wales Specialist Virology Centre | Public Health Wales Microbiology Cardiff |
| EPI_ISL_432155 | Wales Specialist Virology Centre | Public Health Wales Microbiology Cardiff |

| EPI_Accession_ID | Originating Laboratory | Submitting Laboratory |
|---|---|---|
| EPI_ISL_432156 | Wales Specialist Virology Centre | Public Health Wales Microbiology Cardiff |
| EPI_ISL_432157 | Wales Specialist Virology Centre | Public Health Wales Microbiology Cardiff |
| EPI_ISL_432158 | Wales Specialist Virology Centre | Public Health Wales Microbiology Cardiff |
| EPI_ISL_432159 | Wales Specialist Virology Centre | Public Health Wales Microbiology Cardiff |
| EPI_ISL_432160 | Wales Specialist Virology Centre | Public Health Wales Microbiology Cardiff |
| EPI_ISL_432161 | Wales Specialist Virology Centre | Public Health Wales Microbiology Cardiff |
| EPI_ISL_432162 | Wales Specialist Virology Centre | Public Health Wales Microbiology Cardiff |
| EPI_ISL_432163 | Wales Specialist Virology Centre | Public Health Wales Microbiology Cardiff |
| EPI_ISL_432165 | Wales Specialist Virology Centre | Public Health Wales Microbiology Cardiff |
| EPI_ISL_432166 | Wales Specialist Virology Centre | Public Health Wales Microbiology Cardiff |
| EPI_ISL_432167 | Wales Specialist Virology Centre | Public Health Wales Microbiology Cardiff |
| EPI_ISL_432168 | Wales Specialist Virology Centre | Public Health Wales Microbiology Cardiff |
| EPI_ISL_432169 | Wales Specialist Virology Centre | Public Health Wales Microbiology Cardiff |
| EPI_ISL_432170 | Wales Specialist Virology Centre | Public Health Wales Microbiology Cardiff |
| EPI_ISL_432171 | Wales Specialist Virology Centre | Public Health Wales Microbiology Cardiff |
| EPI_ISL_432172 | Wales Specialist Virology Centre | Public Health Wales Microbiology Cardiff |
| EPI_ISL_432173 | Wales Specialist Virology Centre | Public Health Wales Microbiology Cardiff |
| EPI_ISL_432174 | Wales Specialist Virology Centre | Public Health Wales Microbiology Cardiff |
| EPI_ISL_432175 | Wales Specialist Virology Centre | Public Health Wales Microbiology Cardiff |
| EPI_ISL_432176 | Wales Specialist Virology Centre | Public Health Wales Microbiology Cardiff |
| EPI_ISL_432177 | Wales Specialist Virology Centre | Public Health Wales Microbiology Cardiff |
| EPI_ISL_432178 | Wales Specialist Virology Centre | Public Health Wales Microbiology Cardiff |
| EPI_ISL_432179 | Wales Specialist Virology Centre | Public Health Wales Microbiology Cardiff |
| EPI_ISL_432180 | Wales Specialist Virology Centre | Public Health Wales Microbiology Cardiff |
| EPI_ISL_432181 | Wales Specialist Virology Centre | Public Health Wales Microbiology Cardiff |
| EPI_ISL_432182 | Wales Specialist Virology Centre | Public Health Wales Microbiology Cardiff |
| EPI_ISL_432183 | Wales Specialist Virology Centre | Public Health Wales Microbiology Cardiff |
| EPI_ISL_432184 | Wales Specialist Virology Centre | Public Health Wales Microbiology Cardiff |
| EPI_ISL_432185 | Wales Specialist Virology Centre | Public Health Wales Microbiology Cardiff |
| EPI_ISL_432186 | Wales Specialist Virology Centre | Public Health Wales Microbiology Cardiff |
| EPI_ISL_432187 | Wales Specialist Virology Centre | Public Health Wales Microbiology Cardiff |
| EPI_ISL_432188 | Wales Specialist Virology Centre | Public Health Wales Microbiology Cardiff |
| EPI_ISL_432189 | Wales Specialist Virology Centre | Public Health Wales Microbiology Cardiff |

| EPI_Accession_ID | Originating Laboratory | Submitting Laboratory |
|---|---|---|
| EPI_ISL_432190 | Wales Specialist Virology Centre | Public Health Wales Microbiology Cardiff |
| EPI_ISL_432191 | Wales Specialist Virology Centre | Public Health Wales Microbiology Cardiff |
| EPI_ISL_432192 | Wales Specialist Virology Centre | Public Health Wales Microbiology Cardiff |
| EPI_ISL_432193 | Wales Specialist Virology Centre | Public Health Wales Microbiology Cardiff |
| EPI_ISL_432194 | Wales Specialist Virology Centre | Public Health Wales Microbiology Cardiff |
| EPI_ISL_432195 | Wales Specialist Virology Centre | Public Health Wales Microbiology Cardiff |
| EPI_ISL_432196 | Wales Specialist Virology Centre | Public Health Wales Microbiology Cardiff |
| EPI_ISL_432197 | Wales Specialist Virology Centre | Public Health Wales Microbiology Cardiff |
| EPI_ISL_432198 | Wales Specialist Virology Centre | Public Health Wales Microbiology Cardiff |
| EPI_ISL_432199 | Wales Specialist Virology Centre | Public Health Wales Microbiology Cardiff |
| EPI_ISL_432200 | Wales Specialist Virology Centre | Public Health Wales Microbiology Cardiff |
| EPI_ISL_432201 | Wales Specialist Virology Centre | Public Health Wales Microbiology Cardiff |
| EPI_ISL_432202 | Wales Specialist Virology Centre | Public Health Wales Microbiology Cardiff |
| EPI_ISL_432203 | Wales Specialist Virology Centre | Public Health Wales Microbiology Cardiff |
| EPI_ISL_432204 | Wales Specialist Virology Centre | Public Health Wales Microbiology Cardiff |
| EPI_ISL_432205 | Wales Specialist Virology Centre | Public Health Wales Microbiology Cardiff |
| EPI_ISL_432206 | Wales Specialist Virology Centre | Public Health Wales Microbiology Cardiff |
| EPI_ISL_432207 | Wales Specialist Virology Centre | Public Health Wales Microbiology Cardiff |
| EPI_ISL_432208 | Wales Specialist Virology Centre | Public Health Wales Microbiology Cardiff |
| EPI_ISL_432209 | Wales Specialist Virology Centre | Public Health Wales Microbiology Cardiff |
| EPI_ISL_432210 | Wales Specialist Virology Centre | Public Health Wales Microbiology Cardiff |
| EPI_ISL_432211 | Wales Specialist Virology Centre | Public Health Wales Microbiology Cardiff |
| EPI_ISL_432212 | Wales Specialist Virology Centre | Public Health Wales Microbiology Cardiff |
| EPI_ISL_432213 | Wales Specialist Virology Centre | Public Health Wales Microbiology Cardiff |
| EPI_ISL_432214 | Wales Specialist Virology Centre | Public Health Wales Microbiology Cardiff |
| EPI_ISL_432215 | Wales Specialist Virology Centre | Public Health Wales Microbiology Cardiff |
| EPI_ISL_432216 | Wales Specialist Virology Centre | Public Health Wales Microbiology Cardiff |
| EPI_ISL_432217 | Wales Specialist Virology Centre | Public Health Wales Microbiology Cardiff |
| EPI_ISL_432218 | Wales Specialist Virology Centre | Public Health Wales Microbiology Cardiff |
| EPI_ISL_432219 | Wales Specialist Virology Centre | Public Health Wales Microbiology Cardiff |
| EPI_ISL_432220 | Wales Specialist Virology Centre | Public Health Wales Microbiology Cardiff |
| EPI_ISL_432221 | Wales Specialist Virology Centre | Public Health Wales Microbiology Cardiff |
| EPI_ISL_432222 | Wales Specialist Virology Centre | Public Health Wales Microbiology Cardiff |

| EPI_Accession_ID | Originating Laboratory | Submitting Laboratory |
|---|---|---|
| EPI_ISL_432223 | Wales Specialist Virology Centre | Public Health Wales Microbiology Cardiff |
| EPI_ISL_432224 | Wales Specialist Virology Centre | Public Health Wales Microbiology Cardiff |
| EPI_ISL_432225 | Wales Specialist Virology Centre | Public Health Wales Microbiology Cardiff |
| EPI_ISL_432226 | Wales Specialist Virology Centre | Public Health Wales Microbiology Cardiff |
| EPI_ISL_432227 | Wales Specialist Virology Centre | Public Health Wales Microbiology Cardiff |
| EPI_ISL_432228 | Wales Specialist Virology Centre | Public Health Wales Microbiology Cardiff |
| EPI_ISL_432229 | Wales Specialist Virology Centre | Public Health Wales Microbiology Cardiff |
| EPI_ISL_432230 | Wales Specialist Virology Centre | Public Health Wales Microbiology Cardiff |
| EPI_ISL_432231 | Wales Specialist Virology Centre | Public Health Wales Microbiology Cardiff |
| EPI_ISL_432232 | Wales Specialist Virology Centre | Public Health Wales Microbiology Cardiff |
| EPI_ISL_432233 | Wales Specialist Virology Centre | Public Health Wales Microbiology Cardiff |
| EPI_ISL_432234 | Wales Specialist Virology Centre | Public Health Wales Microbiology Cardiff |
| EPI_ISL_432235 | Wales Specialist Virology Centre | Public Health Wales Microbiology Cardiff |
| EPI_ISL_432236 | Wales Specialist Virology Centre | Public Health Wales Microbiology Cardiff |
| EPI_ISL_432237 | Wales Specialist Virology Centre | Public Health Wales Microbiology Cardiff |
| EPI_ISL_432238 | Wales Specialist Virology Centre | Public Health Wales Microbiology Cardiff |
| EPI_ISL_432239 | Wales Specialist Virology Centre | Public Health Wales Microbiology Cardiff |
| EPI_ISL_432240 | Wales Specialist Virology Centre | Public Health Wales Microbiology Cardiff |
| EPI_ISL_432241 | Wales Specialist Virology Centre | Public Health Wales Microbiology Cardiff |
| EPI_ISL_432242 | Wales Specialist Virology Centre | Public Health Wales Microbiology Cardiff |
| EPI_ISL_432243 | Wales Specialist Virology Centre | Public Health Wales Microbiology Cardiff |
| EPI_ISL_432244 | Wales Specialist Virology Centre | Public Health Wales Microbiology Cardiff |
| EPI_ISL_432245 | Wales Specialist Virology Centre | Public Health Wales Microbiology Cardiff |
| EPI_ISL_432246 | Wales Specialist Virology Centre | Public Health Wales Microbiology Cardiff |
| EPI_ISL_432247 | Wales Specialist Virology Centre | Public Health Wales Microbiology Cardiff |
| EPI_ISL_432248 | Wales Specialist Virology Centre | Public Health Wales Microbiology Cardiff |
| EPI_ISL_432249 | Wales Specialist Virology Centre | Public Health Wales Microbiology Cardiff |
| EPI_ISL_432250 | Wales Specialist Virology Centre | Public Health Wales Microbiology Cardiff |
| EPI_ISL_432251 | Wales Specialist Virology Centre | Public Health Wales Microbiology Cardiff |
| EPI_ISL_432252 | Wales Specialist Virology Centre | Public Health Wales Microbiology Cardiff |
| EPI_ISL_432253 | Wales Specialist Virology Centre | Public Health Wales Microbiology Cardiff |
| EPI_ISL_432254 | Wales Specialist Virology Centre | Public Health Wales Microbiology Cardiff |
| EPI_ISL_432255 | Wales Specialist Virology Centre | Public Health Wales Microbiology Cardiff |

| EPI_Accession_ID | Originating Laboratory | Submitting Laboratory |
|---|---|---|
| EPI_ISL_432256 | Wales Specialist Virology Centre | Public Health Wales Microbiology Cardiff |
| EPI_ISL_432257 | Wales Specialist Virology Centre | Public Health Wales Microbiology Cardiff |
| EPI_ISL_432258 | Wales Specialist Virology Centre | Public Health Wales Microbiology Cardiff |
| EPI_ISL_432259 | Wales Specialist Virology Centre | Public Health Wales Microbiology Cardiff |
| EPI_ISL_432260 | Wales Specialist Virology Centre | Public Health Wales Microbiology Cardiff |
| EPI_ISL_432261 | Wales Specialist Virology Centre | Public Health Wales Microbiology Cardiff |
| EPI_ISL_432262 | Wales Specialist Virology Centre | Public Health Wales Microbiology Cardiff |
| EPI_ISL_432263 | Wales Specialist Virology Centre | Public Health Wales Microbiology Cardiff |
| EPI_ISL_432264 | Wales Specialist Virology Centre | Public Health Wales Microbiology Cardiff |
| EPI_ISL_432265 | Wales Specialist Virology Centre | Public Health Wales Microbiology Cardiff |
| EPI_ISL_432266 | Wales Specialist Virology Centre | Public Health Wales Microbiology Cardiff |
| EPI_ISL_432267 | Wales Specialist Virology Centre | Public Health Wales Microbiology Cardiff |
| EPI_ISL_432268 | Wales Specialist Virology Centre | Public Health Wales Microbiology Cardiff |
| EPI_ISL_432269 | Wales Specialist Virology Centre | Public Health Wales Microbiology Cardiff |
| EPI_ISL_432270 | Wales Specialist Virology Centre | Public Health Wales Microbiology Cardiff |
| EPI_ISL_432271 | Wales Specialist Virology Centre | Public Health Wales Microbiology Cardiff |
| EPI_ISL_432272 | Wales Specialist Virology Centre | Public Health Wales Microbiology Cardiff |
| EPI_ISL_432273 | Wales Specialist Virology Centre | Public Health Wales Microbiology Cardiff |
| EPI_ISL_432274 | Wales Specialist Virology Centre | Public Health Wales Microbiology Cardiff |
| EPI_ISL_432275 | Wales Specialist Virology Centre | Public Health Wales Microbiology Cardiff |
| EPI_ISL_432276 | Wales Specialist Virology Centre | Public Health Wales Microbiology Cardiff |
| EPI_ISL_432277 | Wales Specialist Virology Centre | Public Health Wales Microbiology Cardiff |
| EPI_ISL_432278 | Wales Specialist Virology Centre | Public Health Wales Microbiology Cardiff |
| EPI_ISL_432279 | Wales Specialist Virology Centre | Public Health Wales Microbiology Cardiff |
| EPI_ISL_432280 | Wales Specialist Virology Centre | Public Health Wales Microbiology Cardiff |
| EPI_ISL_432281 | Wales Specialist Virology Centre | Public Health Wales Microbiology Cardiff |
| EPI_ISL_432282 | Wales Specialist Virology Centre | Public Health Wales Microbiology Cardiff |
| EPI_ISL_432283 | Wales Specialist Virology Centre | Public Health Wales Microbiology Cardiff |
| EPI_ISL_432284 | Wales Specialist Virology Centre | Public Health Wales Microbiology Cardiff |
| EPI_ISL_432285 | Wales Specialist Virology Centre | Public Health Wales Microbiology Cardiff |
| EPI_ISL_432286 | Wales Specialist Virology Centre | Public Health Wales Microbiology Cardiff |
| EPI_ISL_432287 | Wales Specialist Virology Centre | Public Health Wales Microbiology Cardiff |
| EPI_ISL_432288 | Wales Specialist Virology Centre | Public Health Wales Microbiology Cardiff |

| EPI_Accession_ID | Originating Laboratory | Submitting Laboratory |
|---|---|---|
| EPI_ISL_432289 | Wales Specialist Virology Centre | Public Health Wales Microbiology Cardiff |
| EPI_ISL_432290 | Wales Specialist Virology Centre | Public Health Wales Microbiology Cardiff |
| EPI_ISL_432291 | Wales Specialist Virology Centre | Public Health Wales Microbiology Cardiff |
| EPI_ISL_432292 | Wales Specialist Virology Centre | Public Health Wales Microbiology Cardiff |
| EPI_ISL_432293 | Wales Specialist Virology Centre | Public Health Wales Microbiology Cardiff |
| EPI_ISL_432294 | Wales Specialist Virology Centre | Public Health Wales Microbiology Cardiff |
| EPI_ISL_432295 | Wales Specialist Virology Centre | Public Health Wales Microbiology Cardiff |
| EPI_ISL_432296 | Wales Specialist Virology Centre | Public Health Wales Microbiology Cardiff |
| EPI_ISL_432297 | Wales Specialist Virology Centre | Public Health Wales Microbiology Cardiff |
| EPI_ISL_432298 | Wales Specialist Virology Centre | Public Health Wales Microbiology Cardiff |
| EPI_ISL_432299 | Wales Specialist Virology Centre | Public Health Wales Microbiology Cardiff |
| EPI_ISL_432300 | Wales Specialist Virology Centre | Public Health Wales Microbiology Cardiff |
| EPI_ISL_432301 | Wales Specialist Virology Centre | Public Health Wales Microbiology Cardiff |
| EPI_ISL_432302 | Wales Specialist Virology Centre | Public Health Wales Microbiology Cardiff |
| EPI_ISL_432303 | Wales Specialist Virology Centre | Public Health Wales Microbiology Cardiff |
| EPI_ISL_432304 | Wales Specialist Virology Centre | Public Health Wales Microbiology Cardiff |
| EPI_ISL_432305 | Wales Specialist Virology Centre | Public Health Wales Microbiology Cardiff |
| EPI_ISL_432306 | Wales Specialist Virology Centre | Public Health Wales Microbiology Cardiff |
| EPI_ISL_432307 | Wales Specialist Virology Centre | Public Health Wales Microbiology Cardiff |
| EPI_ISL_432309 | Wales Specialist Virology Centre | Public Health Wales Microbiology Cardiff |
| EPI_ISL_432310 | Wales Specialist Virology Centre | Public Health Wales Microbiology Cardiff |
| EPI_ISL_432312 | Wales Specialist Virology Centre | Public Health Wales Microbiology Cardiff |
| EPI_ISL_432313 | Wales Specialist Virology Centre | Public Health Wales Microbiology Cardiff |
| EPI_ISL_432314 | Wales Specialist Virology Centre | Public Health Wales Microbiology Cardiff |
| EPI_ISL_432315 | Wales Specialist Virology Centre | Public Health Wales Microbiology Cardiff |
| EPI_ISL_432316 | Wales Specialist Virology Centre | Public Health Wales Microbiology Cardiff |
| EPI_ISL_432317 | Wales Specialist Virology Centre | Public Health Wales Microbiology Cardiff |
| EPI_ISL_432318 | Wales Specialist Virology Centre | Public Health Wales Microbiology Cardiff |
| EPI_ISL_432319 | Wales Specialist Virology Centre | Public Health Wales Microbiology Cardiff |
| EPI_ISL_432320 | Wales Specialist Virology Centre | Public Health Wales Microbiology Cardiff |
| EPI_ISL_432321 | Wales Specialist Virology Centre | Public Health Wales Microbiology Cardiff |
| EPI_ISL_432322 | Wales Specialist Virology Centre | Public Health Wales Microbiology Cardiff |
| EPI_ISL_432323 | Wales Specialist Virology Centre | Public Health Wales Microbiology Cardiff |

| EPI_Accession_ID | Originating Laboratory | Submitting Laboratory |
|---|---|---|
| EPI_ISL_432324 | Wales Specialist Virology Centre | Public Health Wales Microbiology Cardiff |
| EPI_ISL_432325 | Wales Specialist Virology Centre | Public Health Wales Microbiology Cardiff |
| EPI_ISL_432327 | Wales Specialist Virology Centre | Public Health Wales Microbiology Cardiff |
| EPI_ISL_432328 | Wales Specialist Virology Centre | Public Health Wales Microbiology Cardiff |
| EPI_ISL_432329 | Wales Specialist Virology Centre | Public Health Wales Microbiology Cardiff |
| EPI_ISL_432330 | Wales Specialist Virology Centre | Public Health Wales Microbiology Cardiff |
| EPI_ISL_432331 | Wales Specialist Virology Centre | Public Health Wales Microbiology Cardiff |
| EPI_ISL_432332 | Wales Specialist Virology Centre | Public Health Wales Microbiology Cardiff |
| EPI_ISL_432333 | Wales Specialist Virology Centre | Public Health Wales Microbiology Cardiff |
| EPI_ISL_432334 | Wales Specialist Virology Centre | Public Health Wales Microbiology Cardiff |
| EPI_ISL_432335 | Wales Specialist Virology Centre | Public Health Wales Microbiology Cardiff |
| EPI_ISL_432337 | Wales Specialist Virology Centre | Public Health Wales Microbiology Cardiff |
| EPI_ISL_432338 | Wales Specialist Virology Centre | Public Health Wales Microbiology Cardiff |
| EPI_ISL_432339 | Wales Specialist Virology Centre | Public Health Wales Microbiology Cardiff |
| EPI_ISL_432340 | Wales Specialist Virology Centre | Public Health Wales Microbiology Cardiff |
| EPI_ISL_432341 | Wales Specialist Virology Centre | Public Health Wales Microbiology Cardiff |
| EPI_ISL_432342 | Wales Specialist Virology Centre | Public Health Wales Microbiology Cardiff |
| EPI_ISL_432343 | Wales Specialist Virology Centre | Public Health Wales Microbiology Cardiff |
| EPI_ISL_432344 | Wales Specialist Virology Centre | Public Health Wales Microbiology Cardiff |
| EPI_ISL_432345 | Wales Specialist Virology Centre | Public Health Wales Microbiology Cardiff |
| EPI_ISL_432346 | Wales Specialist Virology Centre | Public Health Wales Microbiology Cardiff |
| EPI_ISL_432347 | Wales Specialist Virology Centre | Public Health Wales Microbiology Cardiff |
| EPI_ISL_432348 | Wales Specialist Virology Centre | Public Health Wales Microbiology Cardiff |
| EPI_ISL_432349 | Wales Specialist Virology Centre | Public Health Wales Microbiology Cardiff |
| EPI_ISL_432350 | Wales Specialist Virology Centre | Public Health Wales Microbiology Cardiff |
| EPI_ISL_432351 | Wales Specialist Virology Centre | Public Health Wales Microbiology Cardiff |
| EPI_ISL_432352 | Wales Specialist Virology Centre | Public Health Wales Microbiology Cardiff |
| EPI_ISL_432353 | Wales Specialist Virology Centre | Public Health Wales Microbiology Cardiff |
| EPI_ISL_432354 | Wales Specialist Virology Centre | Public Health Wales Microbiology Cardiff |
| EPI_ISL_432355 | Wales Specialist Virology Centre | Public Health Wales Microbiology Cardiff |
| EPI_ISL_432356 | Wales Specialist Virology Centre | Public Health Wales Microbiology Cardiff |
| EPI_ISL_432357 | Wales Specialist Virology Centre | Public Health Wales Microbiology Cardiff |
| EPI_ISL_432358 | Wales Specialist Virology Centre | Public Health Wales Microbiology Cardiff |

| EPI_Accession_ID | Originating Laboratory | Submitting Laboratory |
|---|---|---|
| EPI_ISL_432359 | Wales Specialist Virology Centre | Public Health Wales Microbiology Cardiff |
| EPI_ISL_432360 | Wales Specialist Virology Centre | Public Health Wales Microbiology Cardiff |
| EPI_ISL_432361 | Wales Specialist Virology Centre | Public Health Wales Microbiology Cardiff |
| EPI_ISL_432362 | Wales Specialist Virology Centre | Public Health Wales Microbiology Cardiff |
| EPI_ISL_432363 | Wales Specialist Virology Centre | Public Health Wales Microbiology Cardiff |
| EPI_ISL_432364 | Wales Specialist Virology Centre | Public Health Wales Microbiology Cardiff |
| EPI_ISL_432365 | Wales Specialist Virology Centre | Public Health Wales Microbiology Cardiff |
| EPI_ISL_432366 | Wales Specialist Virology Centre | Public Health Wales Microbiology Cardiff |
| EPI_ISL_432367 | Wales Specialist Virology Centre | Public Health Wales Microbiology Cardiff |
| EPI_ISL_432368 | Wales Specialist Virology Centre | Public Health Wales Microbiology Cardiff |
| EPI_ISL_432369 | Wales Specialist Virology Centre | Public Health Wales Microbiology Cardiff |
| EPI_ISL_432370 | Wales Specialist Virology Centre | Public Health Wales Microbiology Cardiff |
| EPI_ISL_432371 | Wales Specialist Virology Centre | Public Health Wales Microbiology Cardiff |
| EPI_ISL_432372 | Wales Specialist Virology Centre | Public Health Wales Microbiology Cardiff |
| EPI_ISL_432373 | Wales Specialist Virology Centre | Public Health Wales Microbiology Cardiff |
| EPI_ISL_432374 | Wales Specialist Virology Centre | Public Health Wales Microbiology Cardiff |
| EPI_ISL_432377 | Wales Specialist Virology Centre | Public Health Wales Microbiology Cardiff |
| EPI_ISL_432378 | Wales Specialist Virology Centre | Public Health Wales Microbiology Cardiff |
| EPI_ISL_432379 | Wales Specialist Virology Centre | Public Health Wales Microbiology Cardiff |
| EPI_ISL_432380 | Wales Specialist Virology Centre | Public Health Wales Microbiology Cardiff |
| EPI_ISL_432381 | Wales Specialist Virology Centre | Public Health Wales Microbiology Cardiff |
| EPI_ISL_432382 | Wales Specialist Virology Centre | Public Health Wales Microbiology Cardiff |
| EPI_ISL_432383 | Wales Specialist Virology Centre | Public Health Wales Microbiology Cardiff |
| EPI_ISL_432384 | Wales Specialist Virology Centre | Public Health Wales Microbiology Cardiff |
| EPI_ISL_432385 | Wales Specialist Virology Centre | Public Health Wales Microbiology Cardiff |
| EPI_ISL_432386 | Wales Specialist Virology Centre | Public Health Wales Microbiology Cardiff |
| EPI_ISL_432387 | Wales Specialist Virology Centre | Public Health Wales Microbiology Cardiff |
| EPI_ISL_432388 | Wales Specialist Virology Centre | Public Health Wales Microbiology Cardiff |
| EPI_ISL_432389 | Wales Specialist Virology Centre | Public Health Wales Microbiology Cardiff |
| EPI_ISL_432390 | Wales Specialist Virology Centre | Public Health Wales Microbiology Cardiff |
| EPI_ISL_432391 | Wales Specialist Virology Centre | Public Health Wales Microbiology Cardiff |
| EPI_ISL_432392 | Wales Specialist Virology Centre | Public Health Wales Microbiology Cardiff |
| EPI_ISL_432393 | Wales Specialist Virology Centre | Public Health Wales Microbiology Cardiff |

| EPI_Accession_ID | Originating Laboratory | Submitting Laboratory |
|---|---|---|
| EPI_ISL_432394 | Wales Specialist Virology Centre | Public Health Wales Microbiology Cardiff |
| EPI_ISL_432395 | Wales Specialist Virology Centre | Public Health Wales Microbiology Cardiff |
| EPI_ISL_432396 | Wales Specialist Virology Centre | Public Health Wales Microbiology Cardiff |
| EPI_ISL_432397 | Wales Specialist Virology Centre | Public Health Wales Microbiology Cardiff |
| EPI_ISL_432398 | Wales Specialist Virology Centre | Public Health Wales Microbiology Cardiff |
| EPI_ISL_432399 | Wales Specialist Virology Centre | Public Health Wales Microbiology Cardiff |
| EPI_ISL_432400 | Wales Specialist Virology Centre | Public Health Wales Microbiology Cardiff |
| EPI_ISL_432401 | Wales Specialist Virology Centre | Public Health Wales Microbiology Cardiff |
| EPI_ISL_432402 | Wales Specialist Virology Centre | Public Health Wales Microbiology Cardiff |
| EPI_ISL_432403 | Wales Specialist Virology Centre | Public Health Wales Microbiology Cardiff |
| EPI_ISL_432404 | Wales Specialist Virology Centre | Public Health Wales Microbiology Cardiff |
| EPI_ISL_432407 | Wales Specialist Virology Centre | Public Health Wales Microbiology Cardiff |
| EPI_ISL_432408 | Wales Specialist Virology Centre | Public Health Wales Microbiology Cardiff |
| EPI_ISL_432409 | Wales Specialist Virology Centre | Public Health Wales Microbiology Cardiff |
| EPI_ISL_432410 | Wales Specialist Virology Centre | Public Health Wales Microbiology Cardiff |
| EPI_ISL_432411 | Wales Specialist Virology Centre | Public Health Wales Microbiology Cardiff |
| EPI_ISL_432412 | Wales Specialist Virology Centre | Public Health Wales Microbiology Cardiff |
| EPI_ISL_432413 | Wales Specialist Virology Centre | Public Health Wales Microbiology Cardiff |
| EPI_ISL_432414 | Wales Specialist Virology Centre | Public Health Wales Microbiology Cardiff |
| EPI_ISL_432416 | Wales Specialist Virology Centre | Public Health Wales Microbiology Cardiff |
| EPI_ISL_432417 | Wales Specialist Virology Centre | Public Health Wales Microbiology Cardiff |
| EPI_ISL_432418 | Wales Specialist Virology Centre | Public Health Wales Microbiology Cardiff |
| EPI_ISL_432419 | Wales Specialist Virology Centre | Public Health Wales Microbiology Cardiff |
| EPI_ISL_432420 | Wales Specialist Virology Centre | Public Health Wales Microbiology Cardiff |
| EPI_ISL_432421 | Wales Specialist Virology Centre | Public Health Wales Microbiology Cardiff |
| EPI_ISL_432422 | Wales Specialist Virology Centre | Public Health Wales Microbiology Cardiff |
| EPI_ISL_432423 | Wales Specialist Virology Centre | Public Health Wales Microbiology Cardiff |
| EPI_ISL_432424 | Wales Specialist Virology Centre | Public Health Wales Microbiology Cardiff |
| EPI_ISL_432425 | Wales Specialist Virology Centre | Public Health Wales Microbiology Cardiff |
| EPI_ISL_432426 | Wales Specialist Virology Centre | Public Health Wales Microbiology Cardiff |
| EPI_ISL_432427 | Wales Specialist Virology Centre | Public Health Wales Microbiology Cardiff |
| EPI_ISL_432428 | Wales Specialist Virology Centre | Public Health Wales Microbiology Cardiff |
| EPI_ISL_432429 | Wales Specialist Virology Centre | Public Health Wales Microbiology Cardiff |

| EPI_Accession_ID | Originating Laboratory | Submitting Laboratory |
|---|---|---|
| EPI_ISL_432430 | Wales Specialist Virology Centre | Public Health Wales Microbiology Cardiff |
| EPI_ISL_432431 | Wales Specialist Virology Centre | Public Health Wales Microbiology Cardiff |
| EPI_ISL_432432 | Wales Specialist Virology Centre | Public Health Wales Microbiology Cardiff |
| EPI_ISL_432433 | Wales Specialist Virology Centre | Public Health Wales Microbiology Cardiff |
| EPI_ISL_432434 | Wales Specialist Virology Centre | Public Health Wales Microbiology Cardiff |
| EPI_ISL_432435 | Wales Specialist Virology Centre | Public Health Wales Microbiology Cardiff |
| EPI_ISL_432436 | Wales Specialist Virology Centre | Public Health Wales Microbiology Cardiff |
| EPI_ISL_432437 | Wales Specialist Virology Centre | Public Health Wales Microbiology Cardiff |
| EPI_ISL_432438 | Wales Specialist Virology Centre | Public Health Wales Microbiology Cardiff |
| EPI_ISL_432439 | Wales Specialist Virology Centre | Public Health Wales Microbiology Cardiff |
| EPI_ISL_432440 | Wales Specialist Virology Centre | Public Health Wales Microbiology Cardiff |
| EPI_ISL_432441 | Wales Specialist Virology Centre | Public Health Wales Microbiology Cardiff |
| EPI_ISL_432442 | Wales Specialist Virology Centre | Public Health Wales Microbiology Cardiff |
| EPI_ISL_432443 | Wales Specialist Virology Centre | Public Health Wales Microbiology Cardiff |
| EPI_ISL_432444 | Wales Specialist Virology Centre | Public Health Wales Microbiology Cardiff |
| EPI_ISL_432445 | Wales Specialist Virology Centre | Public Health Wales Microbiology Cardiff |
| EPI_ISL_432446 | Wales Specialist Virology Centre | Public Health Wales Microbiology Cardiff |
| EPI_ISL_432447 | Wales Specialist Virology Centre | Public Health Wales Microbiology Cardiff |
| EPI_ISL_432448 | Wales Specialist Virology Centre | Public Health Wales Microbiology Cardiff |
| EPI_ISL_432449 | Wales Specialist Virology Centre | Public Health Wales Microbiology Cardiff |
| EPI_ISL_432450 | Wales Specialist Virology Centre | Public Health Wales Microbiology Cardiff |
| EPI_ISL_432451 | Virology Department, Sheffield Teaching Hospitals NHS Foundation Trust / Virology Department, Sheffield Teaching Hospitals NHS Foundation Trust | COVID-19 Genomics UK (COG-UK) Consortium |
| EPI_ISL_432452 | Virology Department, Sheffield Teaching Hospitals NHS Foundation Trust / Virology Department, Sheffield Teaching Hospitals NHS Foundation Trust | COVID-19 Genomics UK (COG-UK) Consortium |
| EPI_ISL_432453 | Virology Department, Sheffield Teaching Hospitals NHS Foundation Trust / Virology Department, Sheffield Teaching Hospitals NHS Foundation Trust | COVID-19 Genomics UK (COG-UK) Consortium |
| EPI_ISL_432454 | Virology Department, Sheffield Teaching Hospitals NHS Foundation Trust / Virology Department, Sheffield Teaching Hospitals NHS Foundation Trust | COVID-19 Genomics UK (COG-UK) Consortium |

| EPI_Accession_ID | Originating Laboratory | Submitting Laboratory |
|---|---|---|
| EPI_ISL_432455 | Virology Department, Sheffield Teaching Hospitals NHS Foundation Trust / Virology Department, Sheffield Teaching Hospitals NHS Foundation Trust | COVID-19 Genomics UK (COG-UK) Consortium |
| EPI_ISL_432456 | Virology Department, Sheffield Teaching Hospitals NHS Foundation Trust / Virology Department, Sheffield Teaching Hospitals NHS Foundation Trust | COVID-19 Genomics UK (COG-UK) Consortium |
| EPI_ISL_432457 | Virology Department, Sheffield Teaching Hospitals NHS Foundation Trust / Virology Department, Sheffield Teaching Hospitals NHS Foundation Trust | COVID-19 Genomics UK (COG-UK) Consortium |
| EPI_ISL_432458 | Virology Department, Sheffield Teaching Hospitals NHS Foundation Trust / Virology Department, Sheffield Teaching Hospitals NHS Foundation Trust | COVID-19 Genomics UK (COG-UK) Consortium |
| EPI_ISL_432459 | Virology Department, Sheffield Teaching Hospitals NHS Foundation Trust / Virology Department, Sheffield Teaching Hospitals NHS Foundation Trust | COVID-19 Genomics UK (COG-UK) Consortium |
| EPI_ISL_432460 | Virology Department, Sheffield Teaching Hospitals NHS Foundation Trust / Virology Department, Sheffield Teaching Hospitals NHS Foundation Trust | COVID-19 Genomics UK (COG-UK) Consortium |
| EPI_ISL_432461 | Virology Department, Sheffield Teaching Hospitals NHS Foundation Trust / Virology Department, Sheffield Teaching Hospitals NHS Foundation Trust | COVID-19 Genomics UK (COG-UK) Consortium |
| EPI_ISL_432462 | Virology Department, Sheffield Teaching Hospitals NHS Foundation Trust / Virology Department, Sheffield Teaching Hospitals NHS Foundation Trust | COVID-19 Genomics UK (COG-UK) Consortium |
| EPI_ISL_432463 | Virology Department, Sheffield Teaching Hospitals NHS Foundation Trust / Virology Department, Sheffield Teaching Hospitals NHS Foundation Trust | COVID-19 Genomics UK (COG-UK) Consortium |
| EPI_ISL_432464 | Virology Department, Sheffield Teaching Hospitals NHS Foundation Trust / Virology Department, Sheffield Teaching Hospitals NHS Foundation Trust | COVID-19 Genomics UK (COG-UK) Consortium |
| EPI_ISL_432465 | Virology Department, Sheffield Teaching Hospitals NHS Foundation Trust / Virology Department, Sheffield Teaching Hospitals NHS Foundation Trust | COVID-19 Genomics UK (COG-UK) Consortium |

| EPI_Accession_ID | Originating Laboratory | Submitting Laboratory |
|---|---|---|
| EPI_ISL_432466 | Virology Department, Sheffield Teaching Hospitals NHS Foundation Trust / Virology Department, Sheffield Teaching Hospitals NHS Foundation Trust | COVID-19 Genomics UK (COG-UK) Consortium |
| EPI_ISL_432467 | Virology Department, Sheffield Teaching Hospitals NHS Foundation Trust / Virology Department, Sheffield Teaching Hospitals NHS Foundation Trust | COVID-19 Genomics UK (COG-UK) Consortium |
| EPI_ISL_432468 | Virology Department, Sheffield Teaching Hospitals NHS Foundation Trust / Virology Department, Sheffield Teaching Hospitals NHS Foundation Trust | COVID-19 Genomics UK (COG-UK) Consortium |
| EPI_ISL_432469 | Virology Department, Sheffield Teaching Hospitals NHS Foundation Trust / Virology Department, Sheffield Teaching Hospitals NHS Foundation Trust | COVID-19 Genomics UK (COG-UK) Consortium |
| EPI_ISL_432470 | Virology Department, Sheffield Teaching Hospitals NHS Foundation Trust / Virology Department, Sheffield Teaching Hospitals NHS Foundation Trust | COVID-19 Genomics UK (COG-UK) Consortium |
| EPI_ISL_432471 | Virology Department, Sheffield Teaching Hospitals NHS Foundation Trust / Virology Department, Sheffield Teaching Hospitals NHS Foundation Trust | COVID-19 Genomics UK (COG-UK) Consortium |
| EPI_ISL_432472 | Virology Department, Sheffield Teaching Hospitals NHS Foundation Trust / Virology Department, Sheffield Teaching Hospitals NHS Foundation Trust | COVID-19 Genomics UK (COG-UK) Consortium |
| EPI_ISL_432473 | Virology Department, Sheffield Teaching Hospitals NHS Foundation Trust / Virology Department, Sheffield Teaching Hospitals NHS Foundation Trust | COVID-19 Genomics UK (COG-UK) Consortium |
| EPI_ISL_432474 | Virology Department, Sheffield Teaching Hospitals NHS Foundation Trust / Virology Department, Sheffield Teaching Hospitals NHS Foundation Trust | COVID-19 Genomics UK (COG-UK) Consortium |
| EPI_ISL_432475 | Virology Department, Sheffield Teaching Hospitals NHS Foundation Trust / Virology Department, Sheffield Teaching Hospitals NHS Foundation Trust | COVID-19 Genomics UK (COG-UK) Consortium |
| EPI_ISL_432476 | Virology Department, Sheffield Teaching Hospitals NHS Foundation Trust / Virology Department, Sheffield Teaching Hospitals NHS Foundation Trust | COVID-19 Genomics UK (COG-UK) Consortium |

| EPI_Accession_ID | Originating Laboratory | Submitting Laboratory |
|---|---|---|
| EPI_ISL_432477 | Virology Department, Sheffield Teaching Hospitals NHS Foundation Trust / Virology Department, Sheffield Teaching Hospitals NHS Foundation Trust | COVID-19 Genomics UK (COG-UK) Consortium |
| EPI_ISL_432478 | Virology Department, Sheffield Teaching Hospitals NHS Foundation Trust / Virology Department, Sheffield Teaching Hospitals NHS Foundation Trust | COVID-19 Genomics UK (COG-UK) Consortium |
| EPI_ISL_432479 | Virology Department, Sheffield Teaching Hospitals NHS Foundation Trust / Virology Department, Sheffield Teaching Hospitals NHS Foundation Trust | COVID-19 Genomics UK (COG-UK) Consortium |
| EPI_ISL_432480 | Virology Department, Sheffield Teaching Hospitals NHS Foundation Trust / Virology Department, Sheffield Teaching Hospitals NHS Foundation Trust | COVID-19 Genomics UK (COG-UK) Consortium |
| EPI_ISL_432481 | Virology Department, Sheffield Teaching Hospitals NHS Foundation Trust / Virology Department, Sheffield Teaching Hospitals NHS Foundation Trust | COVID-19 Genomics UK (COG-UK) Consortium |
| EPI_ISL_432482 | Virology Department, Sheffield Teaching Hospitals NHS Foundation Trust / Virology Department, Sheffield Teaching Hospitals NHS Foundation Trust | COVID-19 Genomics UK (COG-UK) Consortium |
| EPI_ISL_432483 | Virology Department, Sheffield Teaching Hospitals NHS Foundation Trust / Virology Department, Sheffield Teaching Hospitals NHS Foundation Trust | COVID-19 Genomics UK (COG-UK) Consortium |
| EPI_ISL_432484 | Virology Department, Sheffield Teaching Hospitals NHS Foundation Trust / Virology Department, Sheffield Teaching Hospitals NHS Foundation Trust | COVID-19 Genomics UK (COG-UK) Consortium |
| EPI_ISL_432485 | Virology Department, Sheffield Teaching Hospitals NHS Foundation Trust / Virology Department, Sheffield Teaching Hospitals NHS Foundation Trust | COVID-19 Genomics UK (COG-UK) Consortium |
| EPI_ISL_432486 | Virology Department, Sheffield Teaching Hospitals NHS Foundation Trust / Virology Department, Sheffield Teaching Hospitals NHS Foundation Trust | COVID-19 Genomics UK (COG-UK) Consortium |
| EPI_ISL_432487 | Virology Department, Sheffield Teaching Hospitals NHS Foundation Trust / Virology Department, Sheffield Teaching Hospitals NHS Foundation Trust | COVID-19 Genomics UK (COG-UK) Consortium |

| EPI_Accession_ID | Originating Laboratory | Submitting Laboratory |
|---|---|---|
| EPI_ISL_432488 | Virology Department, Sheffield Teaching Hospitals NHS Foundation Trust / Virology Department, Sheffield Teaching Hospitals NHS Foundation Trust | COVID-19 Genomics UK (COG-UK) Consortium |
| EPI_ISL_432489 | Virology Department, Sheffield Teaching Hospitals NHS Foundation Trust / Virology Department, Sheffield Teaching Hospitals NHS Foundation Trust | COVID-19 Genomics UK (COG-UK) Consortium |
| EPI_ISL_432490 | Virology Department, Sheffield Teaching Hospitals NHS Foundation Trust / Virology Department, Sheffield Teaching Hospitals NHS Foundation Trust | COVID-19 Genomics UK (COG-UK) Consortium |
| EPI_ISL_432491 | Virology Department, Sheffield Teaching Hospitals NHS Foundation Trust / Virology Department, Sheffield Teaching Hospitals NHS Foundation Trust | COVID-19 Genomics UK (COG-UK) Consortium |
| EPI_ISL_432492 | Virology Department, Sheffield Teaching Hospitals NHS Foundation Trust / Virology Department, Sheffield Teaching Hospitals NHS Foundation Trust | COVID-19 Genomics UK (COG-UK) Consortium |
| EPI_ISL_432493 | Virology Department, Sheffield Teaching Hospitals NHS Foundation Trust / Virology Department, Sheffield Teaching Hospitals NHS Foundation Trust | COVID-19 Genomics UK (COG-UK) Consortium |
| EPI_ISL_432494 | Virology Department, Sheffield Teaching Hospitals NHS Foundation Trust / Virology Department, Sheffield Teaching Hospitals NHS Foundation Trust | COVID-19 Genomics UK (COG-UK) Consortium |
| EPI_ISL_432495 | Virology Department, Sheffield Teaching Hospitals NHS Foundation Trust / Virology Department, Sheffield Teaching Hospitals NHS Foundation Trust | COVID-19 Genomics UK (COG-UK) Consortium |
| EPI_ISL_432496 | Virology Department, Sheffield Teaching Hospitals NHS Foundation Trust / Virology Department, Sheffield Teaching Hospitals NHS Foundation Trust | COVID-19 Genomics UK (COG-UK) Consortium |
| EPI_ISL_432497 | Virology Department, Sheffield Teaching Hospitals NHS Foundation Trust / Virology Department, Sheffield Teaching Hospitals NHS Foundation Trust | COVID-19 Genomics UK (COG-UK) Consortium |
| EPI_ISL_432498 | Virology Department, Sheffield Teaching Hospitals NHS Foundation Trust / Virology Department, Sheffield Teaching Hospitals NHS Foundation Trust | COVID-19 Genomics UK (COG-UK) Consortium |

| EPI_Accession_ID | Originating Laboratory | Submitting Laboratory |
|---|---|---|
| EPI_ISL_432499 | Virology Department, Sheffield Teaching Hospitals NHS Foundation Trust / Virology Department, Sheffield Teaching Hospitals NHS Foundation Trust | COVID-19 Genomics UK (COG-UK) Consortium |
| EPI_ISL_432500 | Virology Department, Sheffield Teaching Hospitals NHS Foundation Trust / Virology Department, Sheffield Teaching Hospitals NHS Foundation Trust | COVID-19 Genomics UK (COG-UK) Consortium |
| EPI_ISL_432501 | Virology Department, Sheffield Teaching Hospitals NHS Foundation Trust / Virology Department, Sheffield Teaching Hospitals NHS Foundation Trust | COVID-19 Genomics UK (COG-UK) Consortium |
| EPI_ISL_432502 | Virology Department, Sheffield Teaching Hospitals NHS Foundation Trust / Virology Department, Sheffield Teaching Hospitals NHS Foundation Trust | COVID-19 Genomics UK (COG-UK) Consortium |
| EPI_ISL_432503 | Virology Department, Sheffield Teaching Hospitals NHS Foundation Trust / Virology Department, Sheffield Teaching Hospitals NHS Foundation Trust | COVID-19 Genomics UK (COG-UK) Consortium |
| EPI_ISL_432504 | Virology Department, Sheffield Teaching Hospitals NHS Foundation Trust / Virology Department, Sheffield Teaching Hospitals NHS Foundation Trust | COVID-19 Genomics UK (COG-UK) Consortium |
| EPI_ISL_432505 | Virology Department, Sheffield Teaching Hospitals NHS Foundation Trust / Virology Department, Sheffield Teaching Hospitals NHS Foundation Trust | COVID-19 Genomics UK (COG-UK) Consortium |
| EPI_ISL_432506 | Virology Department, Sheffield Teaching Hospitals NHS Foundation Trust / Virology Department, Sheffield Teaching Hospitals NHS Foundation Trust | COVID-19 Genomics UK (COG-UK) Consortium |
| EPI_ISL_432507 | Virology Department, Sheffield Teaching Hospitals NHS Foundation Trust / Virology Department, Sheffield Teaching Hospitals NHS Foundation Trust | COVID-19 Genomics UK (COG-UK) Consortium |
| EPI_ISL_432508 | Virology Department, Sheffield Teaching Hospitals NHS Foundation Trust / Virology Department, Sheffield Teaching Hospitals NHS Foundation Trust | COVID-19 Genomics UK (COG-UK) Consortium |
| EPI_ISL_432509 | Virology Department, Sheffield Teaching Hospitals NHS Foundation Trust / Virology Department, Sheffield Teaching Hospitals NHS Foundation Trust | COVID-19 Genomics UK (COG-UK) Consortium |

| EPI_Accession_ID | Originating Laboratory | Submitting Laboratory |
|---|---|---|
| EPI_ISL_432510 | Virology Department, Sheffield Teaching Hospitals NHS Foundation Trust / Virology Department, Sheffield Teaching Hospitals NHS Foundation Trust | COVID-19 Genomics UK (COG-UK) Consortium |
| EPI_ISL_432511 | Virology Department, Sheffield Teaching Hospitals NHS Foundation Trust / Virology Department, Sheffield Teaching Hospitals NHS Foundation Trust | COVID-19 Genomics UK (COG-UK) Consortium |
| EPI_ISL_432512 | Virology Department, Sheffield Teaching Hospitals NHS Foundation Trust / Virology Department, Sheffield Teaching Hospitals NHS Foundation Trust | COVID-19 Genomics UK (COG-UK) Consortium |
| EPI_ISL_432513 | Virology Department, Sheffield Teaching Hospitals NHS Foundation Trust / Virology Department, Sheffield Teaching Hospitals NHS Foundation Trust | COVID-19 Genomics UK (COG-UK) Consortium |
| EPI_ISL_432514 | Virology Department, Sheffield Teaching Hospitals NHS Foundation Trust / Virology Department, Sheffield Teaching Hospitals NHS Foundation Trust | COVID-19 Genomics UK (COG-UK) Consortium |
| EPI_ISL_432515 | Virology Department, Sheffield Teaching Hospitals NHS Foundation Trust / Virology Department, Sheffield Teaching Hospitals NHS Foundation Trust | COVID-19 Genomics UK (COG-UK) Consortium |
| EPI_ISL_432516 | Virology Department, Sheffield Teaching Hospitals NHS Foundation Trust / Virology Department, Sheffield Teaching Hospitals NHS Foundation Trust | COVID-19 Genomics UK (COG-UK) Consortium |
| EPI_ISL_432517 | Virology Department, Sheffield Teaching Hospitals NHS Foundation Trust / Virology Department, Sheffield Teaching Hospitals NHS Foundation Trust | COVID-19 Genomics UK (COG-UK) Consortium |
| EPI_ISL_432518 | Virology Department, Sheffield Teaching Hospitals NHS Foundation Trust / Virology Department, Sheffield Teaching Hospitals NHS Foundation Trust | COVID-19 Genomics UK (COG-UK) Consortium |
| EPI_ISL_432519 | Virology Department, Sheffield Teaching Hospitals NHS Foundation Trust / Virology Department, Sheffield Teaching Hospitals NHS Foundation Trust | COVID-19 Genomics UK (COG-UK) Consortium |
| EPI_ISL_432520 | Virology Department, Sheffield Teaching Hospitals NHS Foundation Trust / Virology Department, Sheffield Teaching Hospitals NHS Foundation Trust | COVID-19 Genomics UK (COG-UK) Consortium |

| EPI_Accession_ID | Originating Laboratory | Submitting Laboratory |
|---|---|---|
| EPI_ISL_432521 | Virology Department, Sheffield Teaching Hospitals NHS Foundation Trust / Virology Department, Sheffield Teaching Hospitals NHS Foundation Trust | COVID-19 Genomics UK (COG-UK) Consortium |
| EPI_ISL_432522 | Virology Department, Sheffield Teaching Hospitals NHS Foundation Trust / Virology Department, Sheffield Teaching Hospitals NHS Foundation Trust | COVID-19 Genomics UK (COG-UK) Consortium |
| EPI_ISL_432523 | Virology Department, Sheffield Teaching Hospitals NHS Foundation Trust / Virology Department, Sheffield Teaching Hospitals NHS Foundation Trust | COVID-19 Genomics UK (COG-UK) Consortium |
| EPI_ISL_432524 | Virology Department, Sheffield Teaching Hospitals NHS Foundation Trust / Virology Department, Sheffield Teaching Hospitals NHS Foundation Trust | COVID-19 Genomics UK (COG-UK) Consortium |
| EPI_ISL_432525 | Virology Department, Sheffield Teaching Hospitals NHS Foundation Trust / Virology Department, Sheffield Teaching Hospitals NHS Foundation Trust | COVID-19 Genomics UK (COG-UK) Consortium |
| EPI_ISL_432526 | Virology Department, Sheffield Teaching Hospitals NHS Foundation Trust / Virology Department, Sheffield Teaching Hospitals NHS Foundation Trust | COVID-19 Genomics UK (COG-UK) Consortium |
| EPI_ISL_432527 | Virology Department, Sheffield Teaching Hospitals NHS Foundation Trust / Virology Department, Sheffield Teaching Hospitals NHS Foundation Trust | COVID-19 Genomics UK (COG-UK) Consortium |
| EPI_ISL_432528 | Virology Department, Sheffield Teaching Hospitals NHS Foundation Trust / Virology Department, Sheffield Teaching Hospitals NHS Foundation Trust | COVID-19 Genomics UK (COG-UK) Consortium |
| EPI_ISL_432529 | Virology Department, Sheffield Teaching Hospitals NHS Foundation Trust / Virology Department, Sheffield Teaching Hospitals NHS Foundation Trust | COVID-19 Genomics UK (COG-UK) Consortium |
| EPI_ISL_432530 | Virology Department, Sheffield Teaching Hospitals NHS Foundation Trust / Virology Department, Sheffield Teaching Hospitals NHS Foundation Trust | COVID-19 Genomics UK (COG-UK) Consortium |
| EPI_ISL_432531 | Virology Department, Sheffield Teaching Hospitals NHS Foundation Trust / Virology Department, Sheffield Teaching Hospitals NHS Foundation Trust | COVID-19 Genomics UK (COG-UK) Consortium |

| EPI_Accession_ID | Originating Laboratory | Submitting Laboratory |
|---|---|---|
| EPI_ISL_432532 | Virology Department, Sheffield Teaching Hospitals NHS Foundation Trust / Virology Department, Sheffield Teaching Hospitals NHS Foundation Trust | COVID-19 Genomics UK (COG-UK) Consortium |
| EPI_ISL_432533 | Virology Department, Sheffield Teaching Hospitals NHS Foundation Trust / Virology Department, Sheffield Teaching Hospitals NHS Foundation Trust | COVID-19 Genomics UK (COG-UK) Consortium |
| EPI_ISL_432534 | Virology Department, Sheffield Teaching Hospitals NHS Foundation Trust / Virology Department, Sheffield Teaching Hospitals NHS Foundation Trust | COVID-19 Genomics UK (COG-UK) Consortium |
| EPI_ISL_432535 | Virology Department, Sheffield Teaching Hospitals NHS Foundation Trust / Virology Department, Sheffield Teaching Hospitals NHS Foundation Trust | COVID-19 Genomics UK (COG-UK) Consortium |
| EPI_ISL_432536 | Virology Department, Sheffield Teaching Hospitals NHS Foundation Trust / Virology Department, Sheffield Teaching Hospitals NHS Foundation Trust | COVID-19 Genomics UK (COG-UK) Consortium |
| EPI_ISL_432537 | Virology Department, Sheffield Teaching Hospitals NHS Foundation Trust / Virology Department, Sheffield Teaching Hospitals NHS Foundation Trust | COVID-19 Genomics UK (COG-UK) Consortium |
| EPI_ISL_432538 | Virology Department, Sheffield Teaching Hospitals NHS Foundation Trust / Virology Department, Sheffield Teaching Hospitals NHS Foundation Trust | COVID-19 Genomics UK (COG-UK) Consortium |
| EPI_ISL_432539 | Virology Department, Sheffield Teaching Hospitals NHS Foundation Trust / Virology Department, Sheffield Teaching Hospitals NHS Foundation Trust | COVID-19 Genomics UK (COG-UK) Consortium |
| EPI_ISL_432540 | Virology Department, Sheffield Teaching Hospitals NHS Foundation Trust / Virology Department, Sheffield Teaching Hospitals NHS Foundation Trust | COVID-19 Genomics UK (COG-UK) Consortium |
| EPI_ISL_432541 | Virology Department, Sheffield Teaching Hospitals NHS Foundation Trust / Virology Department, Sheffield Teaching Hospitals NHS Foundation Trust | COVID-19 Genomics UK (COG-UK) Consortium |
| EPI_ISL_432542 | Virology Department, Sheffield Teaching Hospitals NHS Foundation Trust / Virology Department, Sheffield Teaching Hospitals NHS Foundation Trust | COVID-19 Genomics UK (COG-UK) Consortium |

| EPI_Accession_ID | Originating Laboratory | Submitting Laboratory |
|---|---|---|
| EPI_ISL_432543 | Virology Department, Sheffield Teaching Hospitals NHS Foundation Trust / Virology Department, Sheffield Teaching Hospitals NHS Foundation Trust | COVID-19 Genomics UK (COG-UK) Consortium |
| EPI_ISL_432544 | Virology Department, Sheffield Teaching Hospitals NHS Foundation Trust / Virology Department, Sheffield Teaching Hospitals NHS Foundation Trust | COVID-19 Genomics UK (COG-UK) Consortium |
| EPI_ISL_432545 | Virology Department, Sheffield Teaching Hospitals NHS Foundation Trust / Virology Department, Sheffield Teaching Hospitals NHS Foundation Trust | COVID-19 Genomics UK (COG-UK) Consortium |
| EPI_ISL_432546 | Virology Department, Sheffield Teaching Hospitals NHS Foundation Trust / Virology Department, Sheffield Teaching Hospitals NHS Foundation Trust | COVID-19 Genomics UK (COG-UK) Consortium |
| EPI_ISL_432547 | Virology Department, Sheffield Teaching Hospitals NHS Foundation Trust / Virology Department, Sheffield Teaching Hospitals NHS Foundation Trust | COVID-19 Genomics UK (COG-UK) Consortium |
| EPI_ISL_432548 | Virology Department, Sheffield Teaching Hospitals NHS Foundation Trust / Virology Department, Sheffield Teaching Hospitals NHS Foundation Trust | COVID-19 Genomics UK (COG-UK) Consortium |
| EPI_ISL_432549 | Virology Department, Sheffield Teaching Hospitals NHS Foundation Trust / Virology Department, Sheffield Teaching Hospitals NHS Foundation Trust | COVID-19 Genomics UK (COG-UK) Consortium |
| EPI_ISL_432550 | Virology Department, Sheffield Teaching Hospitals NHS Foundation Trust / Virology Department, Sheffield Teaching Hospitals NHS Foundation Trust | COVID-19 Genomics UK (COG-UK) Consortium |
| EPI_ISL_432551 | Virology Department, Sheffield Teaching Hospitals NHS Foundation Trust / Virology Department, Sheffield Teaching Hospitals NHS Foundation Trust | COVID-19 Genomics UK (COG-UK) Consortium |
| EPI_ISL_432552 | Virology Department, Sheffield Teaching Hospitals NHS Foundation Trust / Virology Department, Sheffield Teaching Hospitals NHS Foundation Trust | COVID-19 Genomics UK (COG-UK) Consortium |
| EPI_ISL_432553 | Virology Department, Sheffield Teaching Hospitals NHS Foundation Trust / Virology Department, Sheffield Teaching Hospitals NHS Foundation Trust | COVID-19 Genomics UK (COG-UK) Consortium |

| EPI_Accession_ID | Originating Laboratory | Submitting Laboratory |
|---|---|---|
| EPI_ISL_432554 | Virology Department, Sheffield Teaching Hospitals NHS Foundation Trust / Virology Department, Sheffield Teaching Hospitals NHS Foundation Trust | COVID-19 Genomics UK (COG-UK) Consortium |
| EPI_ISL_432555 | Virology Department, Sheffield Teaching Hospitals NHS Foundation Trust / Virology Department, Sheffield Teaching Hospitals NHS Foundation Trust | COVID-19 Genomics UK (COG-UK) Consortium |
| EPI_ISL_432556 | Virology Department, Sheffield Teaching Hospitals NHS Foundation Trust / Virology Department, Sheffield Teaching Hospitals NHS Foundation Trust | COVID-19 Genomics UK (COG-UK) Consortium |
| EPI_ISL_432557 | Virology Department, Sheffield Teaching Hospitals NHS Foundation Trust / Virology Department, Sheffield Teaching Hospitals NHS Foundation Trust | COVID-19 Genomics UK (COG-UK) Consortium |
| EPI_ISL_432558 | Virology Department, Sheffield Teaching Hospitals NHS Foundation Trust / Virology Department, Sheffield Teaching Hospitals NHS Foundation Trust | COVID-19 Genomics UK (COG-UK) Consortium |
| EPI_ISL_432559 | Virology Department, Sheffield Teaching Hospitals NHS Foundation Trust / Virology Department, Sheffield Teaching Hospitals NHS Foundation Trust | COVID-19 Genomics UK (COG-UK) Consortium |
| EPI_ISL_432560 | Virology Department, Sheffield Teaching Hospitals NHS Foundation Trust / Virology Department, Sheffield Teaching Hospitals NHS Foundation Trust | COVID-19 Genomics UK (COG-UK) Consortium |
| EPI_ISL_432561 | Virology Department, Sheffield Teaching Hospitals NHS Foundation Trust / Virology Department, Sheffield Teaching Hospitals NHS Foundation Trust | COVID-19 Genomics UK (COG-UK) Consortium |
| EPI_ISL_432562 | Virology Department, Sheffield Teaching Hospitals NHS Foundation Trust / Virology Department, Sheffield Teaching Hospitals NHS Foundation Trust | COVID-19 Genomics UK (COG-UK) Consortium |
| EPI_ISL_432563 | Virology Department, Sheffield Teaching Hospitals NHS Foundation Trust / Virology Department, Sheffield Teaching Hospitals NHS Foundation Trust | COVID-19 Genomics UK (COG-UK) Consortium |
| EPI_ISL_432564 | Virology Department, Sheffield Teaching Hospitals NHS Foundation Trust / Virology Department, Sheffield Teaching Hospitals NHS Foundation Trust | COVID-19 Genomics UK (COG-UK) Consortium |

| EPI_Accession_ID | Originating Laboratory | Submitting Laboratory |
|---|---|---|
| EPI_ISL_432565 | Virology Department, Sheffield Teaching Hospitals NHS Foundation Trust / Virology Department, Sheffield Teaching Hospitals NHS Foundation Trust | COVID-19 Genomics UK (COG-UK) Consortium |
| EPI_ISL_432566 | Virology Department, Sheffield Teaching Hospitals NHS Foundation Trust / Virology Department, Sheffield Teaching Hospitals NHS Foundation Trust | COVID-19 Genomics UK (COG-UK) Consortium |
| EPI_ISL_432567 | Virology Department, Sheffield Teaching Hospitals NHS Foundation Trust / Virology Department, Sheffield Teaching Hospitals NHS Foundation Trust | COVID-19 Genomics UK (COG-UK) Consortium |
| EPI_ISL_432568 | Virology Department, Sheffield Teaching Hospitals NHS Foundation Trust / Virology Department, Sheffield Teaching Hospitals NHS Foundation Trust | COVID-19 Genomics UK (COG-UK) Consortium |
| EPI_ISL_432569 | Virology Department, Sheffield Teaching Hospitals NHS Foundation Trust / Virology Department, Sheffield Teaching Hospitals NHS Foundation Trust | COVID-19 Genomics UK (COG-UK) Consortium |
| EPI_ISL_432570 | Virology Department, Sheffield Teaching Hospitals NHS Foundation Trust / Virology Department, Sheffield Teaching Hospitals NHS Foundation Trust | COVID-19 Genomics UK (COG-UK) Consortium |
| EPI_ISL_432571 | Virology Department, Sheffield Teaching Hospitals NHS Foundation Trust / Virology Department, Sheffield Teaching Hospitals NHS Foundation Trust | COVID-19 Genomics UK (COG-UK) Consortium |
| EPI_ISL_432572 | Virology Department, Sheffield Teaching Hospitals NHS Foundation Trust / Virology Department, Sheffield Teaching Hospitals NHS Foundation Trust | COVID-19 Genomics UK (COG-UK) Consortium |
| EPI_ISL_432573 | Virology Department, Sheffield Teaching Hospitals NHS Foundation Trust / Virology Department, Sheffield Teaching Hospitals NHS Foundation Trust | COVID-19 Genomics UK (COG-UK) Consortium |
| EPI_ISL_432574 | Virology Department, Sheffield Teaching Hospitals NHS Foundation Trust / Virology Department, Sheffield Teaching Hospitals NHS Foundation Trust | COVID-19 Genomics UK (COG-UK) Consortium |
| EPI_ISL_432575 | Virology Department, Sheffield Teaching Hospitals NHS Foundation Trust / Virology Department, Sheffield Teaching Hospitals NHS Foundation Trust | COVID-19 Genomics UK (COG-UK) Consortium |

| EPI_Accession_ID | Originating Laboratory | Submitting Laboratory |
|---|---|---|
| EPI_ISL_432576 | Virology Department, Sheffield Teaching Hospitals NHS Foundation Trust / Virology Department, Sheffield Teaching Hospitals NHS Foundation Trust | COVID-19 Genomics UK (COG-UK) Consortium |
| EPI_ISL_432577 | Virology Department, Sheffield Teaching Hospitals NHS Foundation Trust / Virology Department, Sheffield Teaching Hospitals NHS Foundation Trust | COVID-19 Genomics UK (COG-UK) Consortium |
| EPI_ISL_432578 | Virology Department, Sheffield Teaching Hospitals NHS Foundation Trust / Virology Department, Sheffield Teaching Hospitals NHS Foundation Trust | COVID-19 Genomics UK (COG-UK) Consortium |
| EPI_ISL_432579 | Virology Department, Sheffield Teaching Hospitals NHS Foundation Trust / Virology Department, Sheffield Teaching Hospitals NHS Foundation Trust | COVID-19 Genomics UK (COG-UK) Consortium |
| EPI_ISL_432580 | Virology Department, Sheffield Teaching Hospitals NHS Foundation Trust / Virology Department, Sheffield Teaching Hospitals NHS Foundation Trust | COVID-19 Genomics UK (COG-UK) Consortium |
| EPI_ISL_432581 | Virology Department, Sheffield Teaching Hospitals NHS Foundation Trust / Virology Department, Sheffield Teaching Hospitals NHS Foundation Trust | COVID-19 Genomics UK (COG-UK) Consortium |
| EPI_ISL_432582 | Virology Department, Sheffield Teaching Hospitals NHS Foundation Trust / Virology Department, Sheffield Teaching Hospitals NHS Foundation Trust | COVID-19 Genomics UK (COG-UK) Consortium |
| EPI_ISL_432583 | Virology Department, Sheffield Teaching Hospitals NHS Foundation Trust / Virology Department, Sheffield Teaching Hospitals NHS Foundation Trust | COVID-19 Genomics UK (COG-UK) Consortium |
| EPI_ISL_432584 | Virology Department, Sheffield Teaching Hospitals NHS Foundation Trust / Virology Department, Sheffield Teaching Hospitals NHS Foundation Trust | COVID-19 Genomics UK (COG-UK) Consortium |
| EPI_ISL_432585 | Virology Department, Sheffield Teaching Hospitals NHS Foundation Trust / Virology Department, Sheffield Teaching Hospitals NHS Foundation Trust | COVID-19 Genomics UK (COG-UK) Consortium |
| EPI_ISL_432586 | Virology Department, Sheffield Teaching Hospitals NHS Foundation Trust / Virology Department, Sheffield Teaching Hospitals NHS Foundation Trust | COVID-19 Genomics UK (COG-UK) Consortium |

| EPI_Accession_ID | Originating Laboratory | Submitting Laboratory |
|---|---|---|
| EPI_ISL_432587 | Virology Department, Sheffield Teaching Hospitals NHS Foundation Trust / Virology Department, Sheffield Teaching Hospitals NHS Foundation Trust | COVID-19 Genomics UK (COG-UK) Consortium |
| EPI_ISL_432588 | Virology Department, Sheffield Teaching Hospitals NHS Foundation Trust / Virology Department, Sheffield Teaching Hospitals NHS Foundation Trust | COVID-19 Genomics UK (COG-UK) Consortium |
| EPI_ISL_432589 | Virology Department, Sheffield Teaching Hospitals NHS Foundation Trust / Virology Department, Sheffield Teaching Hospitals NHS Foundation Trust | COVID-19 Genomics UK (COG-UK) Consortium |
| EPI_ISL_432590 | Virology Department, Sheffield Teaching Hospitals NHS Foundation Trust / Virology Department, Sheffield Teaching Hospitals NHS Foundation Trust | COVID-19 Genomics UK (COG-UK) Consortium |
| EPI_ISL_432591 | Virology Department, Sheffield Teaching Hospitals NHS Foundation Trust / Virology Department, Sheffield Teaching Hospitals NHS Foundation Trust | COVID-19 Genomics UK (COG-UK) Consortium |
| EPI_ISL_432592 | Virology Department, Sheffield Teaching Hospitals NHS Foundation Trust / Virology Department, Sheffield Teaching Hospitals NHS Foundation Trust | COVID-19 Genomics UK (COG-UK) Consortium |
| EPI_ISL_432593 | Virology Department, Sheffield Teaching Hospitals NHS Foundation Trust / Virology Department, Sheffield Teaching Hospitals NHS Foundation Trust | COVID-19 Genomics UK (COG-UK) Consortium |
| EPI_ISL_432594 | Virology Department, Sheffield Teaching Hospitals NHS Foundation Trust / Virology Department, Sheffield Teaching Hospitals NHS Foundation Trust | COVID-19 Genomics UK (COG-UK) Consortium |
| EPI_ISL_432595 | Virology Department, Sheffield Teaching Hospitals NHS Foundation Trust / Virology Department, Sheffield Teaching Hospitals NHS Foundation Trust | COVID-19 Genomics UK (COG-UK) Consortium |
| EPI_ISL_432596 | Virology Department, Sheffield Teaching Hospitals NHS Foundation Trust / Virology Department, Sheffield Teaching Hospitals NHS Foundation Trust | COVID-19 Genomics UK (COG-UK) Consortium |
| EPI_ISL_432597 | Virology Department, Sheffield Teaching Hospitals NHS Foundation Trust / Virology Department, Sheffield Teaching Hospitals NHS Foundation Trust | COVID-19 Genomics UK (COG-UK) Consortium |

| EPI_Accession_ID | Originating Laboratory | Submitting Laboratory |
|---|---|---|
| EPI_ISL_432598 | Virology Department, Sheffield Teaching Hospitals NHS Foundation Trust / Virology Department, Sheffield Teaching Hospitals NHS Foundation Trust | COVID-19 Genomics UK (COG-UK) Consortium |
| EPI_ISL_432599 | Virology Department, Sheffield Teaching Hospitals NHS Foundation Trust / Virology Department, Sheffield Teaching Hospitals NHS Foundation Trust | COVID-19 Genomics UK (COG-UK) Consortium |
| EPI_ISL_432600 | Virology Department, Sheffield Teaching Hospitals NHS Foundation Trust / Virology Department, Sheffield Teaching Hospitals NHS Foundation Trust | COVID-19 Genomics UK (COG-UK) Consortium |
| EPI_ISL_432601 | Virology Department, Sheffield Teaching Hospitals NHS Foundation Trust / Virology Department, Sheffield Teaching Hospitals NHS Foundation Trust | COVID-19 Genomics UK (COG-UK) Consortium |
| EPI_ISL_432602 | Virology Department, Sheffield Teaching Hospitals NHS Foundation Trust / Virology Department, Sheffield Teaching Hospitals NHS Foundation Trust | COVID-19 Genomics UK (COG-UK) Consortium |
| EPI_ISL_432603 | Virology Department, Sheffield Teaching Hospitals NHS Foundation Trust / Virology Department, Sheffield Teaching Hospitals NHS Foundation Trust | COVID-19 Genomics UK (COG-UK) Consortium |
| EPI_ISL_432604 | Virology Department, Sheffield Teaching Hospitals NHS Foundation Trust / Virology Department, Sheffield Teaching Hospitals NHS Foundation Trust | COVID-19 Genomics UK (COG-UK) Consortium |
| EPI_ISL_432605 | Virology Department, Sheffield Teaching Hospitals NHS Foundation Trust / Virology Department, Sheffield Teaching Hospitals NHS Foundation Trust | COVID-19 Genomics UK (COG-UK) Consortium |
| EPI_ISL_432606 | Virology Department, Sheffield Teaching Hospitals NHS Foundation Trust / Virology Department, Sheffield Teaching Hospitals NHS Foundation Trust | COVID-19 Genomics UK (COG-UK) Consortium |
| EPI_ISL_432607 | Virology Department, Sheffield Teaching Hospitals NHS Foundation Trust / Virology Department, Sheffield Teaching Hospitals NHS Foundation Trust | COVID-19 Genomics UK (COG-UK) Consortium |
| EPI_ISL_432608 | Virology Department, Sheffield Teaching Hospitals NHS Foundation Trust / Virology Department, Sheffield Teaching Hospitals NHS Foundation Trust | COVID-19 Genomics UK (COG-UK) Consortium |

| EPI_Accession_ID | Originating Laboratory | Submitting Laboratory |
|---|---|---|
| EPI_ISL_432609 | Virology Department, Sheffield Teaching Hospitals NHS Foundation Trust / Virology Department, Sheffield Teaching Hospitals NHS Foundation Trust | COVID-19 Genomics UK (COG-UK) Consortium |
| EPI_ISL_432610 | Virology Department, Sheffield Teaching Hospitals NHS Foundation Trust / Virology Department, Sheffield Teaching Hospitals NHS Foundation Trust | COVID-19 Genomics UK (COG-UK) Consortium |
| EPI_ISL_432611 | Virology Department, Sheffield Teaching Hospitals NHS Foundation Trust / Virology Department, Sheffield Teaching Hospitals NHS Foundation Trust | COVID-19 Genomics UK (COG-UK) Consortium |
| EPI_ISL_432612 | Virology Department, Sheffield Teaching Hospitals NHS Foundation Trust / Virology Department, Sheffield Teaching Hospitals NHS Foundation Trust | COVID-19 Genomics UK (COG-UK) Consortium |
| EPI_ISL_432613 | Virology Department, Sheffield Teaching Hospitals NHS Foundation Trust / Virology Department, Sheffield Teaching Hospitals NHS Foundation Trust | COVID-19 Genomics UK (COG-UK) Consortium |
| EPI_ISL_432614 | Virology Department, Sheffield Teaching Hospitals NHS Foundation Trust / Virology Department, Sheffield Teaching Hospitals NHS Foundation Trust | COVID-19 Genomics UK (COG-UK) Consortium |
| EPI_ISL_432615 | Virology Department, Sheffield Teaching Hospitals NHS Foundation Trust / Virology Department, Sheffield Teaching Hospitals NHS Foundation Trust | COVID-19 Genomics UK (COG-UK) Consortium |
| EPI_ISL_432616 | Virology Department, Sheffield Teaching Hospitals NHS Foundation Trust / Virology Department, Sheffield Teaching Hospitals NHS Foundation Trust | COVID-19 Genomics UK (COG-UK) Consortium |
| EPI_ISL_432617 | Virology Department, Sheffield Teaching Hospitals NHS Foundation Trust / Virology Department, Sheffield Teaching Hospitals NHS Foundation Trust | COVID-19 Genomics UK (COG-UK) Consortium |
| EPI_ISL_432618 | Virology Department, Sheffield Teaching Hospitals NHS Foundation Trust / Virology Department, Sheffield Teaching Hospitals NHS Foundation Trust | COVID-19 Genomics UK (COG-UK) Consortium |
| EPI_ISL_432619 | Virology Department, Sheffield Teaching Hospitals NHS Foundation Trust / Virology Department, Sheffield Teaching Hospitals NHS Foundation Trust | COVID-19 Genomics UK (COG-UK) Consortium |

| EPI_Accession_ID | Originating Laboratory | Submitting Laboratory |
|---|---|---|
| EPI_ISL_432620 | Virology Department, Sheffield Teaching Hospitals NHS Foundation Trust / Virology Department, Sheffield Teaching Hospitals NHS Foundation Trust | COVID-19 Genomics UK (COG-UK) Consortium |
| EPI_ISL_432621 | Virology Department, Sheffield Teaching Hospitals NHS Foundation Trust / Virology Department, Sheffield Teaching Hospitals NHS Foundation Trust | COVID-19 Genomics UK (COG-UK) Consortium |
| EPI_ISL_432622 | Virology Department, Sheffield Teaching Hospitals NHS Foundation Trust / Virology Department, Sheffield Teaching Hospitals NHS Foundation Trust | COVID-19 Genomics UK (COG-UK) Consortium |
| EPI_ISL_432623 | Virology Department, Sheffield Teaching Hospitals NHS Foundation Trust / Virology Department, Sheffield Teaching Hospitals NHS Foundation Trust | COVID-19 Genomics UK (COG-UK) Consortium |
| EPI_ISL_432624 | Virology Department, Sheffield Teaching Hospitals NHS Foundation Trust / Virology Department, Sheffield Teaching Hospitals NHS Foundation Trust | COVID-19 Genomics UK (COG-UK) Consortium |
| EPI_ISL_432625 | Virology Department, Sheffield Teaching Hospitals NHS Foundation Trust / Virology Department, Sheffield Teaching Hospitals NHS Foundation Trust | COVID-19 Genomics UK (COG-UK) Consortium |
| EPI_ISL_432626 | Virology Department, Sheffield Teaching Hospitals NHS Foundation Trust / Virology Department, Sheffield Teaching Hospitals NHS Foundation Trust | COVID-19 Genomics UK (COG-UK) Consortium |
| EPI_ISL_432627 | Virology Department, Sheffield Teaching Hospitals NHS Foundation Trust / Virology Department, Sheffield Teaching Hospitals NHS Foundation Trust | COVID-19 Genomics UK (COG-UK) Consortium |
| EPI_ISL_432628 | Virology Department, Sheffield Teaching Hospitals NHS Foundation Trust / Virology Department, Sheffield Teaching Hospitals NHS Foundation Trust | COVID-19 Genomics UK (COG-UK) Consortium |
| EPI_ISL_432629 | Virology Department, Sheffield Teaching Hospitals NHS Foundation Trust / Virology Department, Sheffield Teaching Hospitals NHS Foundation Trust | COVID-19 Genomics UK (COG-UK) Consortium |
| EPI_ISL_432630 | Virology Department, Sheffield Teaching Hospitals NHS Foundation Trust / Virology Department, Sheffield Teaching Hospitals NHS Foundation Trust | COVID-19 Genomics UK (COG-UK) Consortium |

| EPI_Accession_ID | Originating Laboratory | Submitting Laboratory |
|---|---|---|
| EPI_ISL_432631 | Virology Department, Sheffield Teaching Hospitals NHS Foundation Trust / Virology Department, Sheffield Teaching Hospitals NHS Foundation Trust | COVID-19 Genomics UK (COG-UK) Consortium |
| EPI_ISL_432632 | Virology Department, Sheffield Teaching Hospitals NHS Foundation Trust / Virology Department, Sheffield Teaching Hospitals NHS Foundation Trust | COVID-19 Genomics UK (COG-UK) Consortium |
| EPI_ISL_432633 | Virology Department, Sheffield Teaching Hospitals NHS Foundation Trust / Virology Department, Sheffield Teaching Hospitals NHS Foundation Trust | COVID-19 Genomics UK (COG-UK) Consortium |
| EPI_ISL_432634 | Virology Department, Sheffield Teaching Hospitals NHS Foundation Trust / Virology Department, Sheffield Teaching Hospitals NHS Foundation Trust | COVID-19 Genomics UK (COG-UK) Consortium |
| EPI_ISL_432635 | Virology Department, Sheffield Teaching Hospitals NHS Foundation Trust / Virology Department, Sheffield Teaching Hospitals NHS Foundation Trust | COVID-19 Genomics UK (COG-UK) Consortium |
| EPI_ISL_432636 | Virology Department, Sheffield Teaching Hospitals NHS Foundation Trust / Virology Department, Sheffield Teaching Hospitals NHS Foundation Trust | COVID-19 Genomics UK (COG-UK) Consortium |
| EPI_ISL_432637 | Virology Department, Sheffield Teaching Hospitals NHS Foundation Trust / Virology Department, Sheffield Teaching Hospitals NHS Foundation Trust | COVID-19 Genomics UK (COG-UK) Consortium |
| EPI_ISL_432638 | Virology Department, Sheffield Teaching Hospitals NHS Foundation Trust / Virology Department, Sheffield Teaching Hospitals NHS Foundation Trust | COVID-19 Genomics UK (COG-UK) Consortium |
| EPI_ISL_432639 | Virology Department, Sheffield Teaching Hospitals NHS Foundation Trust / Virology Department, Sheffield Teaching Hospitals NHS Foundation Trust | COVID-19 Genomics UK (COG-UK) Consortium |
| EPI_ISL_432640 | Virology Department, Sheffield Teaching Hospitals NHS Foundation Trust / Virology Department, Sheffield Teaching Hospitals NHS Foundation Trust | COVID-19 Genomics UK (COG-UK) Consortium |
| EPI_ISL_432641 | Virology Department, Sheffield Teaching Hospitals NHS Foundation Trust / Virology Department, Sheffield Teaching Hospitals NHS Foundation Trust | COVID-19 Genomics UK (COG-UK) Consortium |

| EPI_Accession_ID | Originating Laboratory | Submitting Laboratory |
|---|---|---|
| EPI_ISL_432642 | Virology Department, Sheffield Teaching Hospitals NHS Foundation Trust / Virology Department, Sheffield Teaching Hospitals NHS Foundation Trust | COVID-19 Genomics UK (COG-UK) Consortium |
| EPI_ISL_432643 | Virology Department, Sheffield Teaching Hospitals NHS Foundation Trust / Virology Department, Sheffield Teaching Hospitals NHS Foundation Trust | COVID-19 Genomics UK (COG-UK) Consortium |
| EPI_ISL_432644 | Virology Department, Sheffield Teaching Hospitals NHS Foundation Trust / Virology Department, Sheffield Teaching Hospitals NHS Foundation Trust | COVID-19 Genomics UK (COG-UK) Consortium |
| EPI_ISL_432645 | Virology Department, Sheffield Teaching Hospitals NHS Foundation Trust / Virology Department, Sheffield Teaching Hospitals NHS Foundation Trust | COVID-19 Genomics UK (COG-UK) Consortium |
| EPI_ISL_432646 | Virology Department, Sheffield Teaching Hospitals NHS Foundation Trust / Virology Department, Sheffield Teaching Hospitals NHS Foundation Trust | COVID-19 Genomics UK (COG-UK) Consortium |
| EPI_ISL_432647 | Virology Department, Sheffield Teaching Hospitals NHS Foundation Trust / Virology Department, Sheffield Teaching Hospitals NHS Foundation Trust | COVID-19 Genomics UK (COG-UK) Consortium |
| EPI_ISL_432648 | Virology Department, Sheffield Teaching Hospitals NHS Foundation Trust / Virology Department, Sheffield Teaching Hospitals NHS Foundation Trust | COVID-19 Genomics UK (COG-UK) Consortium |
| EPI_ISL_432649 | Virology Department, Sheffield Teaching Hospitals NHS Foundation Trust / Virology Department, Sheffield Teaching Hospitals NHS Foundation Trust | COVID-19 Genomics UK (COG-UK) Consortium |
| EPI_ISL_432650 | Virology Department, Sheffield Teaching Hospitals NHS Foundation Trust / Virology Department, Sheffield Teaching Hospitals NHS Foundation Trust | COVID-19 Genomics UK (COG-UK) Consortium |
| EPI_ISL_432651 | Virology Department, Sheffield Teaching Hospitals NHS Foundation Trust / Virology Department, Sheffield Teaching Hospitals NHS Foundation Trust | COVID-19 Genomics UK (COG-UK) Consortium |
| EPI_ISL_432652 | Virology Department, Sheffield Teaching Hospitals NHS Foundation Trust / Virology Department, Sheffield Teaching Hospitals NHS Foundation Trust | COVID-19 Genomics UK (COG-UK) Consortium |

| EPI_Accession_ID | Originating Laboratory | Submitting Laboratory |
|---|---|---|
| EPI_ISL_432653 | Virology Department, Sheffield Teaching Hospitals NHS Foundation Trust / Virology Department, Sheffield Teaching Hospitals NHS Foundation Trust | COVID-19 Genomics UK (COG-UK) Consortium |
| EPI_ISL_432654 | Virology Department, Sheffield Teaching Hospitals NHS Foundation Trust / Virology Department, Sheffield Teaching Hospitals NHS Foundation Trust | COVID-19 Genomics UK (COG-UK) Consortium |
| EPI_ISL_432655 | Virology Department, Sheffield Teaching Hospitals NHS Foundation Trust / Virology Department, Sheffield Teaching Hospitals NHS Foundation Trust | COVID-19 Genomics UK (COG-UK) Consortium |
| EPI_ISL_432656 | Virology Department, Sheffield Teaching Hospitals NHS Foundation Trust / Virology Department, Sheffield Teaching Hospitals NHS Foundation Trust | COVID-19 Genomics UK (COG-UK) Consortium |
| EPI_ISL_432657 | Virology Department, Sheffield Teaching Hospitals NHS Foundation Trust / Virology Department, Sheffield Teaching Hospitals NHS Foundation Trust | COVID-19 Genomics UK (COG-UK) Consortium |
| EPI_ISL_432658 | Virology Department, Sheffield Teaching Hospitals NHS Foundation Trust / Virology Department, Sheffield Teaching Hospitals NHS Foundation Trust | COVID-19 Genomics UK (COG-UK) Consortium |
| EPI_ISL_432659 | Virology Department, Sheffield Teaching Hospitals NHS Foundation Trust / Virology Department, Sheffield Teaching Hospitals NHS Foundation Trust | COVID-19 Genomics UK (COG-UK) Consortium |
| EPI_ISL_432660 | Virology Department, Sheffield Teaching Hospitals NHS Foundation Trust / Virology Department, Sheffield Teaching Hospitals NHS Foundation Trust | COVID-19 Genomics UK (COG-UK) Consortium |
| EPI_ISL_432661 | Virology Department, Sheffield Teaching Hospitals NHS Foundation Trust / Virology Department, Sheffield Teaching Hospitals NHS Foundation Trust | COVID-19 Genomics UK (COG-UK) Consortium |
| EPI_ISL_432662 | Virology Department, Sheffield Teaching Hospitals NHS Foundation Trust / Virology Department, Sheffield Teaching Hospitals NHS Foundation Trust | COVID-19 Genomics UK (COG-UK) Consortium |
| EPI_ISL_432663 | Virology Department, Sheffield Teaching Hospitals NHS Foundation Trust / Virology Department, Sheffield Teaching Hospitals NHS Foundation Trust | COVID-19 Genomics UK (COG-UK) Consortium |

| EPI_Accession_ID | Originating Laboratory | Submitting Laboratory |
|---|---|---|
| EPI_ISL_432664 | Virology Department, Sheffield Teaching Hospitals NHS Foundation Trust / Virology Department, Sheffield Teaching Hospitals NHS Foundation Trust | COVID-19 Genomics UK (COG-UK) Consortium |
| EPI_ISL_432665 | Virology Department, Sheffield Teaching Hospitals NHS Foundation Trust / Virology Department, Sheffield Teaching Hospitals NHS Foundation Trust | COVID-19 Genomics UK (COG-UK) Consortium |
| EPI_ISL_432666 | Virology Department, Sheffield Teaching Hospitals NHS Foundation Trust / Virology Department, Sheffield Teaching Hospitals NHS Foundation Trust | COVID-19 Genomics UK (COG-UK) Consortium |
| EPI_ISL_432667 | Virology Department, Sheffield Teaching Hospitals NHS Foundation Trust / Virology Department, Sheffield Teaching Hospitals NHS Foundation Trust | COVID-19 Genomics UK (COG-UK) Consortium |
| EPI_ISL_432668 | Queens Medical Centre, Clinical Microbiology Department / DeepSeq Nottingham | COVID-19 Genomics UK (COG-UK) Consortium |
| EPI_ISL_432669 | Queens Medical Centre, Clinical Microbiology Department / DeepSeq Nottingham | COVID-19 Genomics UK (COG-UK) Consortium |
| EPI_ISL_432670 | Queens Medical Centre, Clinical Microbiology Department / DeepSeq Nottingham | COVID-19 Genomics UK (COG-UK) Consortium |
| EPI_ISL_432671 | Queens Medical Centre, Clinical Microbiology Department / DeepSeq Nottingham | COVID-19 Genomics UK (COG-UK) Consortium |
| EPI_ISL_432672 | Queens Medical Centre, Clinical Microbiology Department / DeepSeq Nottingham | COVID-19 Genomics UK (COG-UK) Consortium |
| EPI_ISL_432673 | Queens Medical Centre, Clinical Microbiology Department / DeepSeq Nottingham | COVID-19 Genomics UK (COG-UK) Consortium |
| EPI_ISL_432674 | Queens Medical Centre, Clinical Microbiology Department / DeepSeq Nottingham | COVID-19 Genomics UK (COG-UK) Consortium |
| EPI_ISL_432675 | Queens Medical Centre, Clinical Microbiology Department / DeepSeq Nottingham | COVID-19 Genomics UK (COG-UK) Consortium |
| EPI_ISL_432676 | Queens Medical Centre, Clinical Microbiology Department / DeepSeq Nottingham | COVID-19 Genomics UK (COG-UK) Consortium |
| EPI_ISL_432677 | Queens Medical Centre, Clinical Microbiology Department / DeepSeq Nottingham | COVID-19 Genomics UK (COG-UK) Consortium |

| EPI_Accession_ID | Originating Laboratory | Submitting Laboratory |
|---|---|---|
| EPI_ISL_432678 | Queens Medical Centre, Clinical Microbiology Department / DeepSeq Nottingham | COVID-19 Genomics UK (COG-UK) Consortium |
| EPI_ISL_432679 | Queens Medical Centre, Clinical Microbiology Department / DeepSeq Nottingham | COVID-19 Genomics UK (COG-UK) Consortium |
| EPI_ISL_432680 | Queens Medical Centre, Clinical Microbiology Department / DeepSeq Nottingham | COVID-19 Genomics UK (COG-UK) Consortium |
| EPI_ISL_432681 | Queens Medical Centre, Clinical Microbiology Department / DeepSeq Nottingham | COVID-19 Genomics UK (COG-UK) Consortium |
| EPI_ISL_432682 | Queens Medical Centre, Clinical Microbiology Department / DeepSeq Nottingham | COVID-19 Genomics UK (COG-UK) Consortium |
| EPI_ISL_432683 | Queens Medical Centre, Clinical Microbiology Department / DeepSeq Nottingham | COVID-19 Genomics UK (COG-UK) Consortium |
| EPI_ISL_432684 | Queens Medical Centre, Clinical Microbiology Department / DeepSeq Nottingham | COVID-19 Genomics UK (COG-UK) Consortium |
| EPI_ISL_432685 | Queens Medical Centre, Clinical Microbiology Department / DeepSeq Nottingham | COVID-19 Genomics UK (COG-UK) Consortium |
| EPI_ISL_432686 | Queens Medical Centre, Clinical Microbiology Department / DeepSeq Nottingham | COVID-19 Genomics UK (COG-UK) Consortium |
| EPI_ISL_432687 | Queens Medical Centre, Clinical Microbiology Department / DeepSeq Nottingham | COVID-19 Genomics UK (COG-UK) Consortium |
| EPI_ISL_432688 | Queens Medical Centre, Clinical Microbiology Department / DeepSeq Nottingham | COVID-19 Genomics UK (COG-UK) Consortium |
| EPI_ISL_432689 | Queens Medical Centre, Clinical Microbiology Department / DeepSeq Nottingham | COVID-19 Genomics UK (COG-UK) Consortium |
| EPI_ISL_432690 | Queens Medical Centre, Clinical Microbiology Department / DeepSeq Nottingham | COVID-19 Genomics UK (COG-UK) Consortium |
| EPI_ISL_432691 | Queens Medical Centre, Clinical Microbiology Department / DeepSeq Nottingham | COVID-19 Genomics UK (COG-UK) Consortium |
| EPI_ISL_432692 | Queens Medical Centre, Clinical Microbiology Department / DeepSeq Nottingham | COVID-19 Genomics UK (COG-UK) Consortium |
| EPI_ISL_432693 | Queens Medical Centre, Clinical Microbiology Department / DeepSeq Nottingham | COVID-19 Genomics UK (COG-UK) Consortium |

| EPI_Accession_ID | Originating Laboratory | Submitting Laboratory |
|---|---|---|
| EPI_ISL_432694 | Queens Medical Centre, Clinical Microbiology Department / DeepSeq Nottingham | COVID-19 Genomics UK (COG-UK) Consortium |
| EPI_ISL_432695 | Queens Medical Centre, Clinical Microbiology Department / DeepSeq Nottingham | COVID-19 Genomics UK (COG-UK) Consortium |
| EPI_ISL_432696 | Queens Medical Centre, Clinical Microbiology Department / DeepSeq Nottingham | COVID-19 Genomics UK (COG-UK) Consortium |
| EPI_ISL_432697 | Queens Medical Centre, Clinical Microbiology Department / DeepSeq Nottingham | COVID-19 Genomics UK (COG-UK) Consortium |
| EPI_ISL_432698 | Queens Medical Centre, Clinical Microbiology Department / DeepSeq Nottingham | COVID-19 Genomics UK (COG-UK) Consortium |
| EPI_ISL_432699 | Queens Medical Centre, Clinical Microbiology Department / DeepSeq Nottingham | COVID-19 Genomics UK (COG-UK) Consortium |
| EPI_ISL_432700 | Queens Medical Centre, Clinical Microbiology Department / DeepSeq Nottingham | COVID-19 Genomics UK (COG-UK) Consortium |
| EPI_ISL_432701 | Queens Medical Centre, Clinical Microbiology Department / DeepSeq Nottingham | COVID-19 Genomics UK (COG-UK) Consortium |
| EPI_ISL_432702 | Queens Medical Centre, Clinical Microbiology Department / DeepSeq Nottingham | COVID-19 Genomics UK (COG-UK) Consortium |
| EPI_ISL_432703 | Queens Medical Centre, Clinical Microbiology Department / DeepSeq Nottingham | COVID-19 Genomics UK (COG-UK) Consortium |
| EPI_ISL_432704 | Queens Medical Centre, Clinical Microbiology Department / DeepSeq Nottingham | COVID-19 Genomics UK (COG-UK) Consortium |
| EPI_ISL_432705 | Queens Medical Centre, Clinical Microbiology Department / DeepSeq Nottingham | COVID-19 Genomics UK (COG-UK) Consortium |
| EPI_ISL_432706 | Queens Medical Centre, Clinical Microbiology Department / DeepSeq Nottingham | COVID-19 Genomics UK (COG-UK) Consortium |
| EPI_ISL_432707 | Queens Medical Centre, Clinical Microbiology Department / DeepSeq Nottingham | COVID-19 Genomics UK (COG-UK) Consortium |
| EPI_ISL_432708 | Queens Medical Centre, Clinical Microbiology Department / DeepSeq Nottingham | COVID-19 Genomics UK (COG-UK) Consortium |
| EPI_ISL_432709 | Queens Medical Centre, Clinical Microbiology Department / DeepSeq Nottingham | COVID-19 Genomics UK (COG-UK) Consortium |

| EPI_Accession_ID | Originating Laboratory | Submitting Laboratory |
|---|---|---|
| EPI_ISL_432710 | Virology Department, Sheffield Teaching Hospitals NHS Foundation Trust / Virology Department, Sheffield Teaching Hospitals NHS Foundation Trust | COVID-19 Genomics UK (COG-UK) Consortium |
| EPI_ISL_432711 | Virology Department, Sheffield Teaching Hospitals NHS Foundation Trust / Virology Department, Sheffield Teaching Hospitals NHS Foundation Trust | COVID-19 Genomics UK (COG-UK) Consortium |
| EPI_ISL_432712 | Virology Department, Sheffield Teaching Hospitals NHS Foundation Trust / Virology Department, Sheffield Teaching Hospitals NHS Foundation Trust | COVID-19 Genomics UK (COG-UK) Consortium |
| EPI_ISL_432713 | Virology Department, Sheffield Teaching Hospitals NHS Foundation Trust / Virology Department, Sheffield Teaching Hospitals NHS Foundation Trust | COVID-19 Genomics UK (COG-UK) Consortium |
| EPI_ISL_432714 | Virology Department, Sheffield Teaching Hospitals NHS Foundation Trust / Virology Department, Sheffield Teaching Hospitals NHS Foundation Trust | COVID-19 Genomics UK (COG-UK) Consortium |
| EPI_ISL_432715 | Virology Department, Sheffield Teaching Hospitals NHS Foundation Trust / Virology Department, Sheffield Teaching Hospitals NHS Foundation Trust | COVID-19 Genomics UK (COG-UK) Consortium |
| EPI_ISL_432716 | Virology Department, Sheffield Teaching Hospitals NHS Foundation Trust / Virology Department, Sheffield Teaching Hospitals NHS Foundation Trust | COVID-19 Genomics UK (COG-UK) Consortium |
| EPI_ISL_432717 | Virology Department, Sheffield Teaching Hospitals NHS Foundation Trust / Virology Department, Sheffield Teaching Hospitals NHS Foundation Trust | COVID-19 Genomics UK (COG-UK) Consortium |
| EPI_ISL_432718 | Virology Department, Sheffield Teaching Hospitals NHS Foundation Trust / Virology Department, Sheffield Teaching Hospitals NHS Foundation Trust | COVID-19 Genomics UK (COG-UK) Consortium |
| EPI_ISL_432719 | Virology Department, Sheffield Teaching Hospitals NHS Foundation Trust / Virology Department, Sheffield Teaching Hospitals NHS Foundation Trust | COVID-19 Genomics UK (COG-UK) Consortium |
| EPI_ISL_432720 | Virology Department, Sheffield Teaching Hospitals NHS Foundation Trust / Virology Department, Sheffield Teaching Hospitals NHS Foundation Trust | COVID-19 Genomics UK (COG-UK) Consortium |

| EPI_Accession_ID | Originating Laboratory | Submitting Laboratory |
|---|---|---|
| EPI_ISL_432721 | Virology Department, Sheffield Teaching Hospitals NHS Foundation Trust / Virology Department, Sheffield Teaching Hospitals NHS Foundation Trust | COVID-19 Genomics UK (COG-UK) Consortium |
| EPI_ISL_432722 | Virology Department, Sheffield Teaching Hospitals NHS Foundation Trust / Virology Department, Sheffield Teaching Hospitals NHS Foundation Trust | COVID-19 Genomics UK (COG-UK) Consortium |
| EPI_ISL_432723 | Virology Department, Sheffield Teaching Hospitals NHS Foundation Trust / Virology Department, Sheffield Teaching Hospitals NHS Foundation Trust | COVID-19 Genomics UK (COG-UK) Consortium |
| EPI_ISL_432724 | Virology Department, Sheffield Teaching Hospitals NHS Foundation Trust / Virology Department, Sheffield Teaching Hospitals NHS Foundation Trust | COVID-19 Genomics UK (COG-UK) Consortium |
| EPI_ISL_432725 | Virology Department, Sheffield Teaching Hospitals NHS Foundation Trust / Virology Department, Sheffield Teaching Hospitals NHS Foundation Trust | COVID-19 Genomics UK (COG-UK) Consortium |
| EPI_ISL_432726 | Virology Department, Sheffield Teaching Hospitals NHS Foundation Trust / Virology Department, Sheffield Teaching Hospitals NHS Foundation Trust | COVID-19 Genomics UK (COG-UK) Consortium |
| EPI_ISL_432727 | Virology Department, Sheffield Teaching Hospitals NHS Foundation Trust / Virology Department, Sheffield Teaching Hospitals NHS Foundation Trust | COVID-19 Genomics UK (COG-UK) Consortium |
| EPI_ISL_432728 | Virology Department, Sheffield Teaching Hospitals NHS Foundation Trust / Virology Department, Sheffield Teaching Hospitals NHS Foundation Trust | COVID-19 Genomics UK (COG-UK) Consortium |
| EPI_ISL_432729 | Virology Department, Sheffield Teaching Hospitals NHS Foundation Trust / Virology Department, Sheffield Teaching Hospitals NHS Foundation Trust | COVID-19 Genomics UK (COG-UK) Consortium |
| EPI_ISL_432730 | Virology Department, Sheffield Teaching Hospitals NHS Foundation Trust / Virology Department, Sheffield Teaching Hospitals NHS Foundation Trust | COVID-19 Genomics UK (COG-UK) Consortium |
| EPI_ISL_432731 | Virology Department, Sheffield Teaching Hospitals NHS Foundation Trust / Virology Department, Sheffield Teaching Hospitals NHS Foundation Trust | COVID-19 Genomics UK (COG-UK) Consortium |

| EPI_Accession_ID | Originating Laboratory | Submitting Laboratory |
|---|---|---|
| EPI_ISL_432732 | Virology Department, Sheffield Teaching Hospitals NHS Foundation Trust / Virology Department, Sheffield Teaching Hospitals NHS Foundation Trust | COVID-19 Genomics UK (COG-UK) Consortium |
| EPI_ISL_432733 | Virology Department, Sheffield Teaching Hospitals NHS Foundation Trust / Virology Department, Sheffield Teaching Hospitals NHS Foundation Trust | COVID-19 Genomics UK (COG-UK) Consortium |
| EPI_ISL_432734 | Virology Department, Sheffield Teaching Hospitals NHS Foundation Trust / Virology Department, Sheffield Teaching Hospitals NHS Foundation Trust | COVID-19 Genomics UK (COG-UK) Consortium |
| EPI_ISL_432735 | Virology Department, Sheffield Teaching Hospitals NHS Foundation Trust / Virology Department, Sheffield Teaching Hospitals NHS Foundation Trust | COVID-19 Genomics UK (COG-UK) Consortium |
| EPI_ISL_432736 | Virology Department, Sheffield Teaching Hospitals NHS Foundation Trust / Virology Department, Sheffield Teaching Hospitals NHS Foundation Trust | COVID-19 Genomics UK (COG-UK) Consortium |
| EPI_ISL_432737 | Virology Department, Sheffield Teaching Hospitals NHS Foundation Trust / Virology Department, Sheffield Teaching Hospitals NHS Foundation Trust | COVID-19 Genomics UK (COG-UK) Consortium |
| EPI_ISL_432738 | Virology Department, Sheffield Teaching Hospitals NHS Foundation Trust / Virology Department, Sheffield Teaching Hospitals NHS Foundation Trust | COVID-19 Genomics UK (COG-UK) Consortium |
| EPI_ISL_432739 | Virology Department, Sheffield Teaching Hospitals NHS Foundation Trust / Virology Department, Sheffield Teaching Hospitals NHS Foundation Trust | COVID-19 Genomics UK (COG-UK) Consortium |
| EPI_ISL_432740 | Virology Department, Sheffield Teaching Hospitals NHS Foundation Trust / Virology Department, Sheffield Teaching Hospitals NHS Foundation Trust | COVID-19 Genomics UK (COG-UK) Consortium |
| EPI_ISL_432741 | Virology Department, Sheffield Teaching Hospitals NHS Foundation Trust / Virology Department, Sheffield Teaching Hospitals NHS Foundation Trust | COVID-19 Genomics UK (COG-UK) Consortium |
| EPI_ISL_432742 | Virology Department, Sheffield Teaching Hospitals NHS Foundation Trust / Virology Department, Sheffield Teaching Hospitals NHS Foundation Trust | COVID-19 Genomics UK (COG-UK) Consortium |

| EPI_Accession_ID | Originating Laboratory | Submitting Laboratory |
|---|---|---|
| EPI_ISL_432743 | Virology Department, Sheffield Teaching Hospitals NHS Foundation Trust / Virology Department, Sheffield Teaching Hospitals NHS Foundation Trust | COVID-19 Genomics UK (COG-UK) Consortium |
| EPI_ISL_432744 | Virology Department, Sheffield Teaching Hospitals NHS Foundation Trust / Virology Department, Sheffield Teaching Hospitals NHS Foundation Trust | COVID-19 Genomics UK (COG-UK) Consortium |
| EPI_ISL_432745 | Virology Department, Sheffield Teaching Hospitals NHS Foundation Trust / Virology Department, Sheffield Teaching Hospitals NHS Foundation Trust | COVID-19 Genomics UK (COG-UK) Consortium |
| EPI_ISL_432746 | Virology Department, Sheffield Teaching Hospitals NHS Foundation Trust / Virology Department, Sheffield Teaching Hospitals NHS Foundation Trust | COVID-19 Genomics UK (COG-UK) Consortium |
| EPI_ISL_432747 | Virology Department, Sheffield Teaching Hospitals NHS Foundation Trust / Virology Department, Sheffield Teaching Hospitals NHS Foundation Trust | COVID-19 Genomics UK (COG-UK) Consortium |
| EPI_ISL_432748 | Virology Department, Sheffield Teaching Hospitals NHS Foundation Trust / Virology Department, Sheffield Teaching Hospitals NHS Foundation Trust | COVID-19 Genomics UK (COG-UK) Consortium |
| EPI_ISL_432749 | Virology Department, Sheffield Teaching Hospitals NHS Foundation Trust / Virology Department, Sheffield Teaching Hospitals NHS Foundation Trust | COVID-19 Genomics UK (COG-UK) Consortium |
| EPI_ISL_432750 | Virology Department, Sheffield Teaching Hospitals NHS Foundation Trust / Virology Department, Sheffield Teaching Hospitals NHS Foundation Trust | COVID-19 Genomics UK (COG-UK) Consortium |
| EPI_ISL_432751 | Virology Department, Sheffield Teaching Hospitals NHS Foundation Trust / Virology Department, Sheffield Teaching Hospitals NHS Foundation Trust | COVID-19 Genomics UK (COG-UK) Consortium |
| EPI_ISL_432752 | Virology Department, Sheffield Teaching Hospitals NHS Foundation Trust / Virology Department, Sheffield Teaching Hospitals NHS Foundation Trust | COVID-19 Genomics UK (COG-UK) Consortium |
| EPI_ISL_432753 | Virology Department, Sheffield Teaching Hospitals NHS Foundation Trust / Virology Department, Sheffield Teaching Hospitals NHS Foundation Trust | COVID-19 Genomics UK (COG-UK) Consortium |

| EPI_Accession_ID | Originating Laboratory | Submitting Laboratory |
|---|---|---|
| EPI_ISL_432754 | Virology Department, Sheffield Teaching Hospitals NHS Foundation Trust / Virology Department, Sheffield Teaching Hospitals NHS Foundation Trust | COVID-19 Genomics UK (COG-UK) Consortium |
| EPI_ISL_432755 | Virology Department, Sheffield Teaching Hospitals NHS Foundation Trust / Virology Department, Sheffield Teaching Hospitals NHS Foundation Trust | COVID-19 Genomics UK (COG-UK) Consortium |
| EPI_ISL_432756 | Virology Department, Sheffield Teaching Hospitals NHS Foundation Trust / Virology Department, Sheffield Teaching Hospitals NHS Foundation Trust | COVID-19 Genomics UK (COG-UK) Consortium |
| EPI_ISL_432757 | Virology Department, Sheffield Teaching Hospitals NHS Foundation Trust / Virology Department, Sheffield Teaching Hospitals NHS Foundation Trust | COVID-19 Genomics UK (COG-UK) Consortium |
| EPI_ISL_432758 | Virology Department, Sheffield Teaching Hospitals NHS Foundation Trust / Virology Department, Sheffield Teaching Hospitals NHS Foundation Trust | COVID-19 Genomics UK (COG-UK) Consortium |
| EPI_ISL_432759 | Virology Department, Sheffield Teaching Hospitals NHS Foundation Trust / Virology Department, Sheffield Teaching Hospitals NHS Foundation Trust | COVID-19 Genomics UK (COG-UK) Consortium |
| EPI_ISL_432760 | Virology Department, Sheffield Teaching Hospitals NHS Foundation Trust / Virology Department, Sheffield Teaching Hospitals NHS Foundation Trust | COVID-19 Genomics UK (COG-UK) Consortium |
| EPI_ISL_432761 | Virology Department, Sheffield Teaching Hospitals NHS Foundation Trust / Virology Department, Sheffield Teaching Hospitals NHS Foundation Trust | COVID-19 Genomics UK (COG-UK) Consortium |
| EPI_ISL_432762 | Virology Department, Sheffield Teaching Hospitals NHS Foundation Trust / Virology Department, Sheffield Teaching Hospitals NHS Foundation Trust | COVID-19 Genomics UK (COG-UK) Consortium |
| EPI_ISL_432763 | Virology Department, Sheffield Teaching Hospitals NHS Foundation Trust / Virology Department, Sheffield Teaching Hospitals NHS Foundation Trust | COVID-19 Genomics UK (COG-UK) Consortium |
| EPI_ISL_432764 | Virology Department, Sheffield Teaching Hospitals NHS Foundation Trust / Virology Department, Sheffield Teaching Hospitals NHS Foundation Trust | COVID-19 Genomics UK (COG-UK) Consortium |

| EPI_Accession_ID | Originating Laboratory | Submitting Laboratory |
|---|---|---|
| EPI_ISL_432765 | Virology Department, Sheffield Teaching Hospitals NHS Foundation Trust / Virology Department, Sheffield Teaching Hospitals NHS Foundation Trust | COVID-19 Genomics UK (COG-UK) Consortium |
| EPI_ISL_432766 | Virology Department, Sheffield Teaching Hospitals NHS Foundation Trust / Virology Department, Sheffield Teaching Hospitals NHS Foundation Trust | COVID-19 Genomics UK (COG-UK) Consortium |
| EPI_ISL_432767 | Virology Department, Sheffield Teaching Hospitals NHS Foundation Trust / Virology Department, Sheffield Teaching Hospitals NHS Foundation Trust | COVID-19 Genomics UK (COG-UK) Consortium |
| EPI_ISL_432768 | Virology Department, Sheffield Teaching Hospitals NHS Foundation Trust / Virology Department, Sheffield Teaching Hospitals NHS Foundation Trust | COVID-19 Genomics UK (COG-UK) Consortium |
| EPI_ISL_432769 | Virology Department, Sheffield Teaching Hospitals NHS Foundation Trust / Virology Department, Sheffield Teaching Hospitals NHS Foundation Trust | COVID-19 Genomics UK (COG-UK) Consortium |
| EPI_ISL_432770 | Virology Department, Sheffield Teaching Hospitals NHS Foundation Trust / Virology Department, Sheffield Teaching Hospitals NHS Foundation Trust | COVID-19 Genomics UK (COG-UK) Consortium |
| EPI_ISL_432771 | Virology Department, Sheffield Teaching Hospitals NHS Foundation Trust / Virology Department, Sheffield Teaching Hospitals NHS Foundation Trust | COVID-19 Genomics UK (COG-UK) Consortium |
| EPI_ISL_432772 | Virology Department, Sheffield Teaching Hospitals NHS Foundation Trust / Virology Department, Sheffield Teaching Hospitals NHS Foundation Trust | COVID-19 Genomics UK (COG-UK) Consortium |
| EPI_ISL_432773 | Virology Department, Sheffield Teaching Hospitals NHS Foundation Trust / Virology Department, Sheffield Teaching Hospitals NHS Foundation Trust | COVID-19 Genomics UK (COG-UK) Consortium |
| EPI_ISL_432774 | Virology Department, Sheffield Teaching Hospitals NHS Foundation Trust / Virology Department, Sheffield Teaching Hospitals NHS Foundation Trust | COVID-19 Genomics UK (COG-UK) Consortium |
| EPI_ISL_432775 | Virology Department, Sheffield Teaching Hospitals NHS Foundation Trust / Virology Department, Sheffield Teaching Hospitals NHS Foundation Trust | COVID-19 Genomics UK (COG-UK) Consortium |

| EPI_Accession_ID | Originating Laboratory | Submitting Laboratory |
|---|---|---|
| EPI_ISL_432776 | Virology Department, Sheffield Teaching Hospitals NHS Foundation Trust / Virology Department, Sheffield Teaching Hospitals NHS Foundation Trust | COVID-19 Genomics UK (COG-UK) Consortium |
| EPI_ISL_432777 | Virology Department, Sheffield Teaching Hospitals NHS Foundation Trust / Virology Department, Sheffield Teaching Hospitals NHS Foundation Trust | COVID-19 Genomics UK (COG-UK) Consortium |
| EPI_ISL_432778 | Virology Department, Sheffield Teaching Hospitals NHS Foundation Trust / Virology Department, Sheffield Teaching Hospitals NHS Foundation Trust | COVID-19 Genomics UK (COG-UK) Consortium |
| EPI_ISL_432779 | Virology Department, Sheffield Teaching Hospitals NHS Foundation Trust / Virology Department, Sheffield Teaching Hospitals NHS Foundation Trust | COVID-19 Genomics UK (COG-UK) Consortium |
| EPI_ISL_432780 | Virology Department, Sheffield Teaching Hospitals NHS Foundation Trust / Virology Department, Sheffield Teaching Hospitals NHS Foundation Trust | COVID-19 Genomics UK (COG-UK) Consortium |
| EPI_ISL_432781 | Virology Department, Sheffield Teaching Hospitals NHS Foundation Trust / Virology Department, Sheffield Teaching Hospitals NHS Foundation Trust | COVID-19 Genomics UK (COG-UK) Consortium |
| EPI_ISL_432782 | Virology Department, Sheffield Teaching Hospitals NHS Foundation Trust / Virology Department, Sheffield Teaching Hospitals NHS Foundation Trust | COVID-19 Genomics UK (COG-UK) Consortium |
| EPI_ISL_432783 | Virology Department, Sheffield Teaching Hospitals NHS Foundation Trust / Virology Department, Sheffield Teaching Hospitals NHS Foundation Trust | COVID-19 Genomics UK (COG-UK) Consortium |
| EPI_ISL_432784 | Virology Department, Sheffield Teaching Hospitals NHS Foundation Trust / Virology Department, Sheffield Teaching Hospitals NHS Foundation Trust | COVID-19 Genomics UK (COG-UK) Consortium |
| EPI_ISL_432785 | Virology Department, Sheffield Teaching Hospitals NHS Foundation Trust / Virology Department, Sheffield Teaching Hospitals NHS Foundation Trust | COVID-19 Genomics UK (COG-UK) Consortium |
| EPI_ISL_432786 | Virology Department, Sheffield Teaching Hospitals NHS Foundation Trust / Virology Department, Sheffield Teaching Hospitals NHS Foundation Trust | COVID-19 Genomics UK (COG-UK) Consortium |

| EPI_Accession_ID | Originating Laboratory | Submitting Laboratory |
|---|---|---|
| EPI_ISL_432787 | Virology Department, Sheffield Teaching Hospitals NHS Foundation Trust / Virology Department, Sheffield Teaching Hospitals NHS Foundation Trust | COVID-19 Genomics UK (COG-UK) Consortium |
| EPI_ISL_432788 | Virology Department, Sheffield Teaching Hospitals NHS Foundation Trust / Virology Department, Sheffield Teaching Hospitals NHS Foundation Trust | COVID-19 Genomics UK (COG-UK) Consortium |
| EPI_ISL_432789 | Virology Department, Sheffield Teaching Hospitals NHS Foundation Trust / Virology Department, Sheffield Teaching Hospitals NHS Foundation Trust | COVID-19 Genomics UK (COG-UK) Consortium |
| EPI_ISL_432790 | Virology Department, Sheffield Teaching Hospitals NHS Foundation Trust / Virology Department, Sheffield Teaching Hospitals NHS Foundation Trust | COVID-19 Genomics UK (COG-UK) Consortium |
| EPI_ISL_432791 | Virology Department, Sheffield Teaching Hospitals NHS Foundation Trust / Virology Department, Sheffield Teaching Hospitals NHS Foundation Trust | COVID-19 Genomics UK (COG-UK) Consortium |
| EPI_ISL_432792 | Virology Department, Sheffield Teaching Hospitals NHS Foundation Trust / Virology Department, Sheffield Teaching Hospitals NHS Foundation Trust | COVID-19 Genomics UK (COG-UK) Consortium |
| EPI_ISL_432793 | Virology Department, Sheffield Teaching Hospitals NHS Foundation Trust / Virology Department, Sheffield Teaching Hospitals NHS Foundation Trust | COVID-19 Genomics UK (COG-UK) Consortium |
| EPI_ISL_432794 | Virology Department, Sheffield Teaching Hospitals NHS Foundation Trust / Virology Department, Sheffield Teaching Hospitals NHS Foundation Trust | COVID-19 Genomics UK (COG-UK) Consortium |
| EPI_ISL_432795 | Virology Department, Sheffield Teaching Hospitals NHS Foundation Trust / Virology Department, Sheffield Teaching Hospitals NHS Foundation Trust | COVID-19 Genomics UK (COG-UK) Consortium |
| EPI_ISL_432796 | Virology Department, Sheffield Teaching Hospitals NHS Foundation Trust / Virology Department, Sheffield Teaching Hospitals NHS Foundation Trust | COVID-19 Genomics UK (COG-UK) Consortium |
| EPI_ISL_432797 | Virology Department, Sheffield Teaching Hospitals NHS Foundation Trust / Virology Department, Sheffield Teaching Hospitals NHS Foundation Trust | COVID-19 Genomics UK (COG-UK) Consortium |

| EPI_Accession_ID | Originating Laboratory | Submitting Laboratory |
|---|---|---|
| EPI_ISL_432798 | Virology Department, Sheffield Teaching Hospitals NHS Foundation Trust / Virology Department, Sheffield Teaching Hospitals NHS Foundation Trust | COVID-19 Genomics UK (COG-UK) Consortium |
| EPI_ISL_432799 | Virology Department, Sheffield Teaching Hospitals NHS Foundation Trust / Virology Department, Sheffield Teaching Hospitals NHS Foundation Trust | COVID-19 Genomics UK (COG-UK) Consortium |
| EPI_ISL_432800 | Virology Department, Sheffield Teaching Hospitals NHS Foundation Trust / Virology Department, Sheffield Teaching Hospitals NHS Foundation Trust | COVID-19 Genomics UK (COG-UK) Consortium |
| EPI_ISL_432801 | Virology Department, Sheffield Teaching Hospitals NHS Foundation Trust / Virology Department, Sheffield Teaching Hospitals NHS Foundation Trust | COVID-19 Genomics UK (COG-UK) Consortium |
| EPI_ISL_432802 | Virology Department, Sheffield Teaching Hospitals NHS Foundation Trust / Virology Department, Sheffield Teaching Hospitals NHS Foundation Trust | COVID-19 Genomics UK (COG-UK) Consortium |
| EPI_ISL_432803 | Virology Department, Sheffield Teaching Hospitals NHS Foundation Trust / Virology Department, Sheffield Teaching Hospitals NHS Foundation Trust | COVID-19 Genomics UK (COG-UK) Consortium |
| EPI_ISL_432804 | Virology Department, Sheffield Teaching Hospitals NHS Foundation Trust / Virology Department, Sheffield Teaching Hospitals NHS Foundation Trust | COVID-19 Genomics UK (COG-UK) Consortium |
| EPI_ISL_432805 | Virology Department, Sheffield Teaching Hospitals NHS Foundation Trust / Virology Department, Sheffield Teaching Hospitals NHS Foundation Trust | COVID-19 Genomics UK (COG-UK) Consortium |
| EPI_ISL_432806 | Virology Department, Sheffield Teaching Hospitals NHS Foundation Trust / Virology Department, Sheffield Teaching Hospitals NHS Foundation Trust | COVID-19 Genomics UK (COG-UK) Consortium |
| EPI_ISL_432807 | Virology Department, Sheffield Teaching Hospitals NHS Foundation Trust / Virology Department, Sheffield Teaching Hospitals NHS Foundation Trust | COVID-19 Genomics UK (COG-UK) Consortium |
| EPI_ISL_432808 | Virology Department, Sheffield Teaching Hospitals NHS Foundation Trust / Virology Department, Sheffield Teaching Hospitals NHS Foundation Trust | COVID-19 Genomics UK (COG-UK) Consortium |

| EPI_Accession_ID | Originating Laboratory | Submitting Laboratory |
|---|---|---|
| EPI_ISL_432809 | Virology Department, Sheffield Teaching Hospitals NHS Foundation Trust / Virology Department, Sheffield Teaching Hospitals NHS Foundation Trust | COVID-19 Genomics UK (COG-UK) Consortium |
| EPI_ISL_432810 | Virology Department, Sheffield Teaching Hospitals NHS Foundation Trust / Virology Department, Sheffield Teaching Hospitals NHS Foundation Trust | COVID-19 Genomics UK (COG-UK) Consortium |
| EPI_ISL_432811 | Virology Department, Sheffield Teaching Hospitals NHS Foundation Trust / Virology Department, Sheffield Teaching Hospitals NHS Foundation Trust | COVID-19 Genomics UK (COG-UK) Consortium |
| EPI_ISL_432812 | Virology Department, Sheffield Teaching Hospitals NHS Foundation Trust / Virology Department, Sheffield Teaching Hospitals NHS Foundation Trust | COVID-19 Genomics UK (COG-UK) Consortium |
| EPI_ISL_432813 | Virology Department, Sheffield Teaching Hospitals NHS Foundation Trust / Virology Department, Sheffield Teaching Hospitals NHS Foundation Trust | COVID-19 Genomics UK (COG-UK) Consortium |
| EPI_ISL_432814 | Virology Department, Sheffield Teaching Hospitals NHS Foundation Trust / Virology Department, Sheffield Teaching Hospitals NHS Foundation Trust | COVID-19 Genomics UK (COG-UK) Consortium |
| EPI_ISL_432815 | Virology Department, Sheffield Teaching Hospitals NHS Foundation Trust / Virology Department, Sheffield Teaching Hospitals NHS Foundation Trust | COVID-19 Genomics UK (COG-UK) Consortium |
| EPI_ISL_432816 | Virology Department, Sheffield Teaching Hospitals NHS Foundation Trust / Virology Department, Sheffield Teaching Hospitals NHS Foundation Trust | COVID-19 Genomics UK (COG-UK) Consortium |
| EPI_ISL_432817 | Virology Department, Sheffield Teaching Hospitals NHS Foundation Trust / Virology Department, Sheffield Teaching Hospitals NHS Foundation Trust | COVID-19 Genomics UK (COG-UK) Consortium |
| EPI_ISL_432818 | Virology Department, Sheffield Teaching Hospitals NHS Foundation Trust / Virology Department, Sheffield Teaching Hospitals NHS Foundation Trust | COVID-19 Genomics UK (COG-UK) Consortium |
| EPI_ISL_432819 | Virology Department, Sheffield Teaching Hospitals NHS Foundation Trust / Virology Department, Sheffield Teaching Hospitals NHS Foundation Trust | COVID-19 Genomics UK (COG-UK) Consortium |

| EPI_Accession_ID | Originating Laboratory | Submitting Laboratory |
|---|---|---|
| EPI_ISL_432820 | Virology Department, Sheffield Teaching Hospitals NHS Foundation Trust / Virology Department, Sheffield Teaching Hospitals NHS Foundation Trust | COVID-19 Genomics UK (COG-UK) Consortium |
| EPI_ISL_432821 | Virology Department, Sheffield Teaching Hospitals NHS Foundation Trust / Virology Department, Sheffield Teaching Hospitals NHS Foundation Trust | COVID-19 Genomics UK (COG-UK) Consortium |
| EPI_ISL_432822 | Virology Department, Sheffield Teaching Hospitals NHS Foundation Trust / Virology Department, Sheffield Teaching Hospitals NHS Foundation Trust | COVID-19 Genomics UK (COG-UK) Consortium |
| EPI_ISL_432823 | Virology Department, Sheffield Teaching Hospitals NHS Foundation Trust / Virology Department, Sheffield Teaching Hospitals NHS Foundation Trust | COVID-19 Genomics UK (COG-UK) Consortium |
| EPI_ISL_432824 | Virology Department, Sheffield Teaching Hospitals NHS Foundation Trust / Virology Department, Sheffield Teaching Hospitals NHS Foundation Trust | COVID-19 Genomics UK (COG-UK) Consortium |
| EPI_ISL_432825 | Virology Department, Sheffield Teaching Hospitals NHS Foundation Trust / Virology Department, Sheffield Teaching Hospitals NHS Foundation Trust | COVID-19 Genomics UK (COG-UK) Consortium |
| EPI_ISL_432826 | Virology Department, Sheffield Teaching Hospitals NHS Foundation Trust / Virology Department, Sheffield Teaching Hospitals NHS Foundation Trust | COVID-19 Genomics UK (COG-UK) Consortium |
| EPI_ISL_432827 | Virology Department, Sheffield Teaching Hospitals NHS Foundation Trust / Virology Department, Sheffield Teaching Hospitals NHS Foundation Trust | COVID-19 Genomics UK (COG-UK) Consortium |
| EPI_ISL_432828 | Virology Department, Sheffield Teaching Hospitals NHS Foundation Trust / Virology Department, Sheffield Teaching Hospitals NHS Foundation Trust | COVID-19 Genomics UK (COG-UK) Consortium |
| EPI_ISL_432829 | Virology Department, Sheffield Teaching Hospitals NHS Foundation Trust / Virology Department, Sheffield Teaching Hospitals NHS Foundation Trust | COVID-19 Genomics UK (COG-UK) Consortium |
| EPI_ISL_432830 | Virology Department, Sheffield Teaching Hospitals NHS Foundation Trust / Virology Department, Sheffield Teaching Hospitals NHS Foundation Trust | COVID-19 Genomics UK (COG-UK) Consortium |

| EPI_Accession_ID | Originating Laboratory | Submitting Laboratory |
|---|---|---|
| EPI_ISL_432831 | Virology Department, Sheffield Teaching Hospitals NHS Foundation Trust / Virology Department, Sheffield Teaching Hospitals NHS Foundation Trust | COVID-19 Genomics UK (COG-UK) Consortium |
| EPI_ISL_432832 | Virology Department, Sheffield Teaching Hospitals NHS Foundation Trust / Virology Department, Sheffield Teaching Hospitals NHS Foundation Trust | COVID-19 Genomics UK (COG-UK) Consortium |
| EPI_ISL_432833 | Virology Department, Sheffield Teaching Hospitals NHS Foundation Trust / Virology Department, Sheffield Teaching Hospitals NHS Foundation Trust | COVID-19 Genomics UK (COG-UK) Consortium |
| EPI_ISL_432834 | Virology Department, Sheffield Teaching Hospitals NHS Foundation Trust / Virology Department, Sheffield Teaching Hospitals NHS Foundation Trust | COVID-19 Genomics UK (COG-UK) Consortium |
| EPI_ISL_432835 | Virology Department, Sheffield Teaching Hospitals NHS Foundation Trust / Virology Department, Sheffield Teaching Hospitals NHS Foundation Trust | COVID-19 Genomics UK (COG-UK) Consortium |
| EPI_ISL_432836 | Virology Department, Sheffield Teaching Hospitals NHS Foundation Trust / Virology Department, Sheffield Teaching Hospitals NHS Foundation Trust | COVID-19 Genomics UK (COG-UK) Consortium |
| EPI_ISL_432837 | Virology Department, Sheffield Teaching Hospitals NHS Foundation Trust / Virology Department, Sheffield Teaching Hospitals NHS Foundation Trust | COVID-19 Genomics UK (COG-UK) Consortium |
| EPI_ISL_432838 | Virology Department, Sheffield Teaching Hospitals NHS Foundation Trust / Virology Department, Sheffield Teaching Hospitals NHS Foundation Trust | COVID-19 Genomics UK (COG-UK) Consortium |
| EPI_ISL_432839 | Virology Department, Sheffield Teaching Hospitals NHS Foundation Trust / Virology Department, Sheffield Teaching Hospitals NHS Foundation Trust | COVID-19 Genomics UK (COG-UK) Consortium |
| EPI_ISL_432840 | Virology Department, Sheffield Teaching Hospitals NHS Foundation Trust / Virology Department, Sheffield Teaching Hospitals NHS Foundation Trust | COVID-19 Genomics UK (COG-UK) Consortium |
| EPI_ISL_432841 | Virology Department, Sheffield Teaching Hospitals NHS Foundation Trust / Virology Department, Sheffield Teaching Hospitals NHS Foundation Trust | COVID-19 Genomics UK (COG-UK) Consortium |

| EPI_Accession_ID | Originating Laboratory | Submitting Laboratory |
|---|---|---|
| EPI_ISL_432842 | Virology Department, Sheffield Teaching Hospitals NHS Foundation Trust / Virology Department, Sheffield Teaching Hospitals NHS Foundation Trust | COVID-19 Genomics UK (COG-UK) Consortium |
| EPI_ISL_432843 | Virology Department, Sheffield Teaching Hospitals NHS Foundation Trust / Virology Department, Sheffield Teaching Hospitals NHS Foundation Trust | COVID-19 Genomics UK (COG-UK) Consortium |
| EPI_ISL_432844 | Virology Department, Sheffield Teaching Hospitals NHS Foundation Trust / Virology Department, Sheffield Teaching Hospitals NHS Foundation Trust | COVID-19 Genomics UK (COG-UK) Consortium |
| EPI_ISL_432845 | Virology Department, Sheffield Teaching Hospitals NHS Foundation Trust / Virology Department, Sheffield Teaching Hospitals NHS Foundation Trust | COVID-19 Genomics UK (COG-UK) Consortium |
| EPI_ISL_432846 | Virology Department, Sheffield Teaching Hospitals NHS Foundation Trust / Virology Department, Sheffield Teaching Hospitals NHS Foundation Trust | COVID-19 Genomics UK (COG-UK) Consortium |
| EPI_ISL_432847 | Virology Department, Sheffield Teaching Hospitals NHS Foundation Trust / Virology Department, Sheffield Teaching Hospitals NHS Foundation Trust | COVID-19 Genomics UK (COG-UK) Consortium |
| EPI_ISL_432848 | Virology Department, Sheffield Teaching Hospitals NHS Foundation Trust / Virology Department, Sheffield Teaching Hospitals NHS Foundation Trust | COVID-19 Genomics UK (COG-UK) Consortium |
| EPI_ISL_432849 | Virology Department, Sheffield Teaching Hospitals NHS Foundation Trust / Virology Department, Sheffield Teaching Hospitals NHS Foundation Trust | COVID-19 Genomics UK (COG-UK) Consortium |
| EPI_ISL_432850 | Virology Department, Sheffield Teaching Hospitals NHS Foundation Trust / Virology Department, Sheffield Teaching Hospitals NHS Foundation Trust | COVID-19 Genomics UK (COG-UK) Consortium |
| EPI_ISL_432851 | Virology Department, Sheffield Teaching Hospitals NHS Foundation Trust / Virology Department, Sheffield Teaching Hospitals NHS Foundation Trust | COVID-19 Genomics UK (COG-UK) Consortium |
| EPI_ISL_432852 | Virology Department, Sheffield Teaching Hospitals NHS Foundation Trust / Virology Department, Sheffield Teaching Hospitals NHS Foundation Trust | COVID-19 Genomics UK (COG-UK) Consortium |

| EPI_Accession_ID | Originating Laboratory | Submitting Laboratory |
| --- | --- | --- |
| EPI_ISL_432853 | Virology Department, Sheffield Teaching Hospitals NHS Foundation Trust / Virology Department, Sheffield Teaching Hospitals NHS Foundation Trust | COVID-19 Genomics UK (COG-UK) Consortium |
| EPI_ISL_432854 | Virology Department, Sheffield Teaching Hospitals NHS Foundation Trust / Virology Department, Sheffield Teaching Hospitals NHS Foundation Trust | COVID-19 Genomics UK (COG-UK) Consortium |
| EPI_ISL_432855 | Virology Department, Sheffield Teaching Hospitals NHS Foundation Trust / Virology Department, Sheffield Teaching Hospitals NHS Foundation Trust | COVID-19 Genomics UK (COG-UK) Consortium |
| EPI_ISL_432856 | Virology Department, Sheffield Teaching Hospitals NHS Foundation Trust / Virology Department, Sheffield Teaching Hospitals NHS Foundation Trust | COVID-19 Genomics UK (COG-UK) Consortium |
| EPI_ISL_432857 | Virology Department, Sheffield Teaching Hospitals NHS Foundation Trust / Virology Department, Sheffield Teaching Hospitals NHS Foundation Trust | COVID-19 Genomics UK (COG-UK) Consortium |
| EPI_ISL_432858 | Virology Department, Sheffield Teaching Hospitals NHS Foundation Trust / Virology Department, Sheffield Teaching Hospitals NHS Foundation Trust | COVID-19 Genomics UK (COG-UK) Consortium |
| EPI_ISL_432859 | Virology Department, Sheffield Teaching Hospitals NHS Foundation Trust / Virology Department, Sheffield Teaching Hospitals NHS Foundation Trust | COVID-19 Genomics UK (COG-UK) Consortium |
| EPI_ISL_432860 | Virology Department, Sheffield Teaching Hospitals NHS Foundation Trust / Virology Department, Sheffield Teaching Hospitals NHS Foundation Trust | COVID-19 Genomics UK (COG-UK) Consortium |
| EPI_ISL_432861 | Virology Department, Sheffield Teaching Hospitals NHS Foundation Trust / Virology Department, Sheffield Teaching Hospitals NHS Foundation Trust | COVID-19 Genomics UK (COG-UK) Consortium |
| EPI_ISL_432862 | Virology Department, Sheffield Teaching Hospitals NHS Foundation Trust / Virology Department, Sheffield Teaching Hospitals NHS Foundation Trust | COVID-19 Genomics UK (COG-UK) Consortium |
| EPI_ISL_432863 | Virology Department, Sheffield Teaching Hospitals NHS Foundation Trust / Virology Department, Sheffield Teaching Hospitals NHS Foundation Trust | COVID-19 Genomics UK (COG-UK) Consortium |

| EPI_Accession_ID | Originating Laboratory | Submitting Laboratory |
|---|---|---|
| EPI_ISL_432864 | Virology Department, Sheffield Teaching Hospitals NHS Foundation Trust / Virology Department, Sheffield Teaching Hospitals NHS Foundation Trust | COVID-19 Genomics UK (COG-UK) Consortium |
| EPI_ISL_432865 | Virology Department, Sheffield Teaching Hospitals NHS Foundation Trust / Virology Department, Sheffield Teaching Hospitals NHS Foundation Trust | COVID-19 Genomics UK (COG-UK) Consortium |
| EPI_ISL_432866 | Virology Department, Sheffield Teaching Hospitals NHS Foundation Trust / Virology Department, Sheffield Teaching Hospitals NHS Foundation Trust | COVID-19 Genomics UK (COG-UK) Consortium |
| EPI_ISL_432867 | Virology Department, Sheffield Teaching Hospitals NHS Foundation Trust / Virology Department, Sheffield Teaching Hospitals NHS Foundation Trust | COVID-19 Genomics UK (COG-UK) Consortium |
| EPI_ISL_432868 | Virology Department, Royal Infirmary of Edinburgh, NHS Lothian / School of Biological Sciences, University of Edinburgh / Institute of Genetics and Molecular Medicine, University of Edinburgh | COVID-19 Genomics UK (COG-UK) Consortium |
| EPI_ISL_432869 | Virology Department, Royal Infirmary of Edinburgh, NHS Lothian / School of Biological Sciences, University of Edinburgh / Institute of Genetics and Molecular Medicine, University of Edinburgh | COVID-19 Genomics UK (COG-UK) Consortium |
| EPI_ISL_432870 | Virology Department, Royal Infirmary of Edinburgh, NHS Lothian / School of Biological Sciences, University of Edinburgh / Institute of Genetics and Molecular Medicine, University of Edinburgh | COVID-19 Genomics UK (COG-UK) Consortium |
| EPI_ISL_432871 | Virology Department, Royal Infirmary of Edinburgh, NHS Lothian / School of Biological Sciences, University of Edinburgh / Institute of Genetics and Molecular Medicine, University of Edinburgh | COVID-19 Genomics UK (COG-UK) Consortium |
| EPI_ISL_432872 | Virology Department, Royal Infirmary of Edinburgh, NHS Lothian / School of Biological Sciences, University of Edinburgh / Institute of Genetics and Molecular Medicine, University of Edinburgh | COVID-19 Genomics UK (COG-UK) Consortium |

| EPI_Accession_ID | Originating Laboratory | Submitting Laboratory |
|---|---|---|
| EPI_ISL_432873 | Virology Department, Royal Infirmary of Edinburgh, NHS Lothian / School of Biological Sciences, University of Edinburgh / Institute of Genetics and Molecular Medicine, University of Edinburgh | COVID-19 Genomics UK (COG-UK) Consortium |
| EPI_ISL_432874 | Virology Department, Royal Infirmary of Edinburgh, NHS Lothian / School of Biological Sciences, University of Edinburgh / Institute of Genetics and Molecular Medicine, University of Edinburgh | COVID-19 Genomics UK (COG-UK) Consortium |
| EPI_ISL_432875 | Virology Department, Royal Infirmary of Edinburgh, NHS Lothian / School of Biological Sciences, University of Edinburgh / Institute of Genetics and Molecular Medicine, University of Edinburgh | COVID-19 Genomics UK (COG-UK) Consortium |
| EPI_ISL_432876 | Virology Department, Royal Infirmary of Edinburgh, NHS Lothian / School of Biological Sciences, University of Edinburgh / Institute of Genetics and Molecular Medicine, University of Edinburgh | COVID-19 Genomics UK (COG-UK) Consortium |
| EPI_ISL_432877 | Virology Department, Royal Infirmary of Edinburgh, NHS Lothian / School of Biological Sciences, University of Edinburgh / Institute of Genetics and Molecular Medicine, University of Edinburgh | COVID-19 Genomics UK (COG-UK) Consortium |
| EPI_ISL_432878 | Virology Department, Royal Infirmary of Edinburgh, NHS Lothian / School of Biological Sciences, University of Edinburgh / Institute of Genetics and Molecular Medicine, University of Edinburgh | COVID-19 Genomics UK (COG-UK) Consortium |
| EPI_ISL_432879 | Virology Department, Royal Infirmary of Edinburgh, NHS Lothian / School of Biological Sciences, University of Edinburgh / Institute of Genetics and Molecular Medicine, University of Edinburgh | COVID-19 Genomics UK (COG-UK) Consortium |
| EPI_ISL_432880 | Virology Department, Royal Infirmary of Edinburgh, NHS Lothian / School of Biological Sciences, University of Edinburgh / Institute of Genetics and Molecular Medicine, University of Edinburgh | COVID-19 Genomics UK (COG-UK) Consortium |

| EPI_Accession_ID | Originating Laboratory | Submitting Laboratory |
|---|---|---|
| EPI_ISL_432881 | Virology Department, Royal Infirmary of Edinburgh, NHS Lothian / School of Biological Sciences, University of Edinburgh / Institute of Genetics and Molecular Medicine, University of Edinburgh | COVID-19 Genomics UK (COG-UK) Consortium |
| EPI_ISL_432882 | Virology Department, Royal Infirmary of Edinburgh, NHS Lothian / School of Biological Sciences, University of Edinburgh / Institute of Genetics and Molecular Medicine, University of Edinburgh | COVID-19 Genomics UK (COG-UK) Consortium |
| EPI_ISL_432883 | Virology Department, Royal Infirmary of Edinburgh, NHS Lothian / School of Biological Sciences, University of Edinburgh / Institute of Genetics and Molecular Medicine, University of Edinburgh | COVID-19 Genomics UK (COG-UK) Consortium |
| EPI_ISL_432884 | Virology Department, Royal Infirmary of Edinburgh, NHS Lothian / School of Biological Sciences, University of Edinburgh / Institute of Genetics and Molecular Medicine, University of Edinburgh | COVID-19 Genomics UK (COG-UK) Consortium |
| EPI_ISL_432885 | Virology Department, Royal Infirmary of Edinburgh, NHS Lothian / School of Biological Sciences, University of Edinburgh / Institute of Genetics and Molecular Medicine, University of Edinburgh | COVID-19 Genomics UK (COG-UK) Consortium |
| EPI_ISL_432886 | Virology Department, Royal Infirmary of Edinburgh, NHS Lothian / School of Biological Sciences, University of Edinburgh / Institute of Genetics and Molecular Medicine, University of Edinburgh | COVID-19 Genomics UK (COG-UK) Consortium |
| EPI_ISL_432887 | Virology Department, Royal Infirmary of Edinburgh, NHS Lothian / School of Biological Sciences, University of Edinburgh / Institute of Genetics and Molecular Medicine, University of Edinburgh | COVID-19 Genomics UK (COG-UK) Consortium |
| EPI_ISL_432888 | Virology Department, Royal Infirmary of Edinburgh, NHS Lothian / School of Biological Sciences, University of Edinburgh / Institute of Genetics and Molecular Medicine, University of Edinburgh | COVID-19 Genomics UK (COG-UK) Consortium |

| EPI_Accession_ID | Originating Laboratory | Submitting Laboratory |
|---|---|---|
| EPI_ISL_432889 | Virology Department, Royal Infirmary of Edinburgh, NHS Lothian / School of Biological Sciences, University of Edinburgh / Institute of Genetics and Molecular Medicine, University of Edinburgh | COVID-19 Genomics UK (COG-UK) Consortium |
| EPI_ISL_432890 | Virology Department, Royal Infirmary of Edinburgh, NHS Lothian / School of Biological Sciences, University of Edinburgh / Institute of Genetics and Molecular Medicine, University of Edinburgh | COVID-19 Genomics UK (COG-UK) Consortium |
| EPI_ISL_432891 | Virology Department, Royal Infirmary of Edinburgh, NHS Lothian / School of Biological Sciences, University of Edinburgh / Institute of Genetics and Molecular Medicine, University of Edinburgh | COVID-19 Genomics UK (COG-UK) Consortium |
| EPI_ISL_432892 | Virology Department, Royal Infirmary of Edinburgh, NHS Lothian / School of Biological Sciences, University of Edinburgh / Institute of Genetics and Molecular Medicine, University of Edinburgh | COVID-19 Genomics UK (COG-UK) Consortium |
| EPI_ISL_432893 | Virology Department, Royal Infirmary of Edinburgh, NHS Lothian / School of Biological Sciences, University of Edinburgh / Institute of Genetics and Molecular Medicine, University of Edinburgh | COVID-19 Genomics UK (COG-UK) Consortium |
| EPI_ISL_432894 | Virology Department, Royal Infirmary of Edinburgh, NHS Lothian / School of Biological Sciences, University of Edinburgh / Institute of Genetics and Molecular Medicine, University of Edinburgh | COVID-19 Genomics UK (COG-UK) Consortium |
| EPI_ISL_432895 | Virology Department, Royal Infirmary of Edinburgh, NHS Lothian / School of Biological Sciences, University of Edinburgh / Institute of Genetics and Molecular Medicine, University of Edinburgh | COVID-19 Genomics UK (COG-UK) Consortium |
| EPI_ISL_432896 | Virology Department, Royal Infirmary of Edinburgh, NHS Lothian / School of Biological Sciences, University of Edinburgh / Institute of Genetics and Molecular Medicine, University of Edinburgh | COVID-19 Genomics UK (COG-UK) Consortium |

| EPI_Accession_ID | Originating Laboratory | Submitting Laboratory |
|---|---|---|
| EPI_ISL_432897 | Virology Department, Royal Infirmary of Edinburgh, NHS Lothian / School of Biological Sciences, University of Edinburgh / Institute of Genetics and Molecular Medicine, University of Edinburgh | COVID-19 Genomics UK (COG-UK) Consortium |
| EPI_ISL_432898 | Virology Department, Royal Infirmary of Edinburgh, NHS Lothian / School of Biological Sciences, University of Edinburgh / Institute of Genetics and Molecular Medicine, University of Edinburgh | COVID-19 Genomics UK (COG-UK) Consortium |
| EPI_ISL_432899 | Virology Department, Royal Infirmary of Edinburgh, NHS Lothian / School of Biological Sciences, University of Edinburgh / Institute of Genetics and Molecular Medicine, University of Edinburgh | COVID-19 Genomics UK (COG-UK) Consortium |
| EPI_ISL_432900 | Queens Medical Centre, Clinical Microbiology Department / DeepSeq Nottingham | COVID-19 Genomics UK (COG-UK) Consortium |
| EPI_ISL_432901 | Queens Medical Centre, Clinical Microbiology Department / DeepSeq Nottingham | COVID-19 Genomics UK (COG-UK) Consortium |
| EPI_ISL_432902 | Queens Medical Centre, Clinical Microbiology Department / DeepSeq Nottingham | COVID-19 Genomics UK (COG-UK) Consortium |
| EPI_ISL_432903 | Queens Medical Centre, Clinical Microbiology Department / DeepSeq Nottingham | COVID-19 Genomics UK (COG-UK) Consortium |
| EPI_ISL_432904 | Queens Medical Centre, Clinical Microbiology Department / DeepSeq Nottingham | COVID-19 Genomics UK (COG-UK) Consortium |
| EPI_ISL_432905 | Queens Medical Centre, Clinical Microbiology Department / DeepSeq Nottingham | COVID-19 Genomics UK (COG-UK) Consortium |
| EPI_ISL_432906 | Queens Medical Centre, Clinical Microbiology Department / DeepSeq Nottingham | COVID-19 Genomics UK (COG-UK) Consortium |
| EPI_ISL_432907 | Queens Medical Centre, Clinical Microbiology Department / DeepSeq Nottingham | COVID-19 Genomics UK (COG-UK) Consortium |
| EPI_ISL_432908 | Queens Medical Centre, Clinical Microbiology Department / DeepSeq Nottingham | COVID-19 Genomics UK (COG-UK) Consortium |
| EPI_ISL_432909 | Queens Medical Centre, Clinical Microbiology Department / DeepSeq Nottingham | COVID-19 Genomics UK (COG-UK) Consortium |

| EPI_Accession_ID | Originating Laboratory | Submitting Laboratory |
| --- | --- | --- |
| EPI_ISL_432910 | Queens Medical Centre, Clinical Microbiology Department / DeepSeq Nottingham | COVID-19 Genomics UK (COG-UK) Consortium |
| EPI_ISL_432911 | Queens Medical Centre, Clinical Microbiology Department / DeepSeq Nottingham | COVID-19 Genomics UK (COG-UK) Consortium |
| EPI_ISL_432912 | Queens Medical Centre, Clinical Microbiology Department / DeepSeq Nottingham | COVID-19 Genomics UK (COG-UK) Consortium |
| EPI_ISL_432913 | Queens Medical Centre, Clinical Microbiology Department / DeepSeq Nottingham | COVID-19 Genomics UK (COG-UK) Consortium |
| EPI_ISL_432914 | Queens Medical Centre, Clinical Microbiology Department / DeepSeq Nottingham | COVID-19 Genomics UK (COG-UK) Consortium |
| EPI_ISL_432915 | Queens Medical Centre, Clinical Microbiology Department / DeepSeq Nottingham | COVID-19 Genomics UK (COG-UK) Consortium |
| EPI_ISL_432916 | Queens Medical Centre, Clinical Microbiology Department / DeepSeq Nottingham | COVID-19 Genomics UK (COG-UK) Consortium |
| EPI_ISL_432917 | Queens Medical Centre, Clinical Microbiology Department / DeepSeq Nottingham | COVID-19 Genomics UK (COG-UK) Consortium |
| EPI_ISL_432918 | Queens Medical Centre, Clinical Microbiology Department / DeepSeq Nottingham | COVID-19 Genomics UK (COG-UK) Consortium |
| EPI_ISL_432919 | Queens Medical Centre, Clinical Microbiology Department / DeepSeq Nottingham | COVID-19 Genomics UK (COG-UK) Consortium |
| EPI_ISL_432920 | Queens Medical Centre, Clinical Microbiology Department / DeepSeq Nottingham | COVID-19 Genomics UK (COG-UK) Consortium |
| EPI_ISL_432921 | Queens Medical Centre, Clinical Microbiology Department / DeepSeq Nottingham | COVID-19 Genomics UK (COG-UK) Consortium |
| EPI_ISL_432922 | Queens Medical Centre, Clinical Microbiology Department / DeepSeq Nottingham | COVID-19 Genomics UK (COG-UK) Consortium |
| EPI_ISL_432923 | Queens Medical Centre, Clinical Microbiology Department / DeepSeq Nottingham | COVID-19 Genomics UK (COG-UK) Consortium |
| EPI_ISL_432924 | Queens Medical Centre, Clinical Microbiology Department / DeepSeq Nottingham | COVID-19 Genomics UK (COG-UK) Consortium |
| EPI_ISL_432925 | Queens Medical Centre, Clinical Microbiology Department / DeepSeq Nottingham | COVID-19 Genomics UK (COG-UK) Consortium |

| EPI_Accession_ID | Originating Laboratory | Submitting Laboratory |
|---|---|---|
| EPI_ISL_432926 | Queens Medical Centre, Clinical Microbiology Department / DeepSeq Nottingham | COVID-19 Genomics UK (COG-UK) Consortium |
| EPI_ISL_432927 | Queens Medical Centre, Clinical Microbiology Department / DeepSeq Nottingham | COVID-19 Genomics UK (COG-UK) Consortium |
| EPI_ISL_432928 | Queens Medical Centre, Clinical Microbiology Department / DeepSeq Nottingham | COVID-19 Genomics UK (COG-UK) Consortium |
| EPI_ISL_432929 | Queens Medical Centre, Clinical Microbiology Department / DeepSeq Nottingham | COVID-19 Genomics UK (COG-UK) Consortium |
| EPI_ISL_432930 | Queens Medical Centre, Clinical Microbiology Department / DeepSeq Nottingham | COVID-19 Genomics UK (COG-UK) Consortium |
| EPI_ISL_432931 | Queens Medical Centre, Clinical Microbiology Department / DeepSeq Nottingham | COVID-19 Genomics UK (COG-UK) Consortium |
| EPI_ISL_432932 | Queens Medical Centre, Clinical Microbiology Department / DeepSeq Nottingham | COVID-19 Genomics UK (COG-UK) Consortium |
| EPI_ISL_432933 | Queens Medical Centre, Clinical Microbiology Department / DeepSeq Nottingham | COVID-19 Genomics UK (COG-UK) Consortium |
| EPI_ISL_432934 | Queens Medical Centre, Clinical Microbiology Department / DeepSeq Nottingham | COVID-19 Genomics UK (COG-UK) Consortium |
| EPI_ISL_432935 | Queens Medical Centre, Clinical Microbiology Department / DeepSeq Nottingham | COVID-19 Genomics UK (COG-UK) Consortium |
| EPI_ISL_432936 | Queens Medical Centre, Clinical Microbiology Department / DeepSeq Nottingham | COVID-19 Genomics UK (COG-UK) Consortium |
| EPI_ISL_432937 | Queens Medical Centre, Clinical Microbiology Department / DeepSeq Nottingham | COVID-19 Genomics UK (COG-UK) Consortium |
| EPI_ISL_432938 | Queens Medical Centre, Clinical Microbiology Department / DeepSeq Nottingham | COVID-19 Genomics UK (COG-UK) Consortium |
| EPI_ISL_432939 | Queens Medical Centre, Clinical Microbiology Department / DeepSeq Nottingham | COVID-19 Genomics UK (COG-UK) Consortium |
| EPI_ISL_432940 | Queens Medical Centre, Clinical Microbiology Department / DeepSeq Nottingham | COVID-19 Genomics UK (COG-UK) Consortium |
| EPI_ISL_432941 | Queens Medical Centre, Clinical Microbiology Department / DeepSeq Nottingham | COVID-19 Genomics UK (COG-UK) Consortium |

| EPI_Accession_ID | Originating Laboratory | Submitting Laboratory |
|---|---|---|
| EPI_ISL_432942 | Queens Medical Centre, Clinical Microbiology Department / DeepSeq Nottingham | COVID-19 Genomics UK (COG-UK) Consortium |
| EPI_ISL_432943 | Queens Medical Centre, Clinical Microbiology Department / DeepSeq Nottingham | COVID-19 Genomics UK (COG-UK) Consortium |
| EPI_ISL_432944 | Queens Medical Centre, Clinical Microbiology Department / DeepSeq Nottingham | COVID-19 Genomics UK (COG-UK) Consortium |
| EPI_ISL_432945 | Queens Medical Centre, Clinical Microbiology Department / DeepSeq Nottingham | COVID-19 Genomics UK (COG-UK) Consortium |
| EPI_ISL_432946 | Queens Medical Centre, Clinical Microbiology Department / DeepSeq Nottingham | COVID-19 Genomics UK (COG-UK) Consortium |
| EPI_ISL_432947 | Queens Medical Centre, Clinical Microbiology Department / DeepSeq Nottingham | COVID-19 Genomics UK (COG-UK) Consortium |
| EPI_ISL_432948 | Queens Medical Centre, Clinical Microbiology Department / DeepSeq Nottingham | COVID-19 Genomics UK (COG-UK) Consortium |
| EPI_ISL_432949 | Queens Medical Centre, Clinical Microbiology Department / DeepSeq Nottingham | COVID-19 Genomics UK (COG-UK) Consortium |
| EPI_ISL_432950 | Queens Medical Centre, Clinical Microbiology Department / DeepSeq Nottingham | COVID-19 Genomics UK (COG-UK) Consortium |
| EPI_ISL_432951 | Queens Medical Centre, Clinical Microbiology Department / DeepSeq Nottingham | COVID-19 Genomics UK (COG-UK) Consortium |
| EPI_ISL_432952 | Queens Medical Centre, Clinical Microbiology Department / DeepSeq Nottingham | COVID-19 Genomics UK (COG-UK) Consortium |
| EPI_ISL_432953 | Queens Medical Centre, Clinical Microbiology Department / DeepSeq Nottingham | COVID-19 Genomics UK (COG-UK) Consortium |
| EPI_ISL_432954 | Queens Medical Centre, Clinical Microbiology Department / DeepSeq Nottingham | COVID-19 Genomics UK (COG-UK) Consortium |
| EPI_ISL_432955 | Queens Medical Centre, Clinical Microbiology Department / DeepSeq Nottingham | COVID-19 Genomics UK (COG-UK) Consortium |
| EPI_ISL_432956 | Queens Medical Centre, Clinical Microbiology Department / DeepSeq Nottingham | COVID-19 Genomics UK (COG-UK) Consortium |
| EPI_ISL_432957 | Queens Medical Centre, Clinical Microbiology Department / DeepSeq Nottingham | COVID-19 Genomics UK (COG-UK) Consortium |

| EPI_Accession_ID | Originating Laboratory | Submitting Laboratory |
|---|---|---|
| EPI_ISL_432958 | Queens Medical Centre, Clinical Microbiology Department / DeepSeq Nottingham | COVID-19 Genomics UK (COG-UK) Consortium |
| EPI_ISL_432959 | Queens Medical Centre, Clinical Microbiology Department / DeepSeq Nottingham | COVID-19 Genomics UK (COG-UK) Consortium |
| EPI_ISL_432960 | Queens Medical Centre, Clinical Microbiology Department / DeepSeq Nottingham | COVID-19 Genomics UK (COG-UK) Consortium |
| EPI_ISL_432961 | Queens Medical Centre, Clinical Microbiology Department / DeepSeq Nottingham | COVID-19 Genomics UK (COG-UK) Consortium |
| EPI_ISL_432962 | Queens Medical Centre, Clinical Microbiology Department / DeepSeq Nottingham | COVID-19 Genomics UK (COG-UK) Consortium |
| EPI_ISL_432963 | Queens Medical Centre, Clinical Microbiology Department / DeepSeq Nottingham | COVID-19 Genomics UK (COG-UK) Consortium |
| EPI_ISL_432964 | Queens Medical Centre, Clinical Microbiology Department / DeepSeq Nottingham | COVID-19 Genomics UK (COG-UK) Consortium |
| EPI_ISL_432965 | Queens Medical Centre, Clinical Microbiology Department / DeepSeq Nottingham | COVID-19 Genomics UK (COG-UK) Consortium |
| EPI_ISL_432966 | Queens Medical Centre, Clinical Microbiology Department / DeepSeq Nottingham | COVID-19 Genomics UK (COG-UK) Consortium |
| EPI_ISL_432967 | Queens Medical Centre, Clinical Microbiology Department / DeepSeq Nottingham | COVID-19 Genomics UK (COG-UK) Consortium |
| EPI_ISL_432968 | Queens Medical Centre, Clinical Microbiology Department / DeepSeq Nottingham | COVID-19 Genomics UK (COG-UK) Consortium |
| EPI_ISL_432969 | Queens Medical Centre, Clinical Microbiology Department / DeepSeq Nottingham | COVID-19 Genomics UK (COG-UK) Consortium |
| EPI_ISL_432970 | Queens Medical Centre, Clinical Microbiology Department / DeepSeq Nottingham | COVID-19 Genomics UK (COG-UK) Consortium |
| EPI_ISL_432971 | Queens Medical Centre, Clinical Microbiology Department / DeepSeq Nottingham | COVID-19 Genomics UK (COG-UK) Consortium |
| EPI_ISL_432972 | Queens Medical Centre, Clinical Microbiology Department / DeepSeq Nottingham | COVID-19 Genomics UK (COG-UK) Consortium |
| EPI_ISL_432973 | Queens Medical Centre, Clinical Microbiology Department / DeepSeq Nottingham | COVID-19 Genomics UK (COG-UK) Consortium |

| EPI_Accession_ID | Originating Laboratory | Submitting Laboratory |
|---|---|---|
| EPI_ISL_432974 | Queens Medical Centre, Clinical Microbiology Department / DeepSeq Nottingham | COVID-19 Genomics UK (COG-UK) Consortium |
| EPI_ISL_432975 | Queens Medical Centre, Clinical Microbiology Department / DeepSeq Nottingham | COVID-19 Genomics UK (COG-UK) Consortium |
| EPI_ISL_432976 | Queens Medical Centre, Clinical Microbiology Department / DeepSeq Nottingham | COVID-19 Genomics UK (COG-UK) Consortium |
| EPI_ISL_432977 | Queens Medical Centre, Clinical Microbiology Department / DeepSeq Nottingham | COVID-19 Genomics UK (COG-UK) Consortium |
| EPI_ISL_432978 | Queens Medical Centre, Clinical Microbiology Department / DeepSeq Nottingham | COVID-19 Genomics UK (COG-UK) Consortium |
| EPI_ISL_432979 | Queens Medical Centre, Clinical Microbiology Department / DeepSeq Nottingham | COVID-19 Genomics UK (COG-UK) Consortium |
| EPI_ISL_432980 | Queens Medical Centre, Clinical Microbiology Department / DeepSeq Nottingham | COVID-19 Genomics UK (COG-UK) Consortium |
| EPI_ISL_432981 | Queens Medical Centre, Clinical Microbiology Department / DeepSeq Nottingham | COVID-19 Genomics UK (COG-UK) Consortium |
| EPI_ISL_432982 | Queens Medical Centre, Clinical Microbiology Department / DeepSeq Nottingham | COVID-19 Genomics UK (COG-UK) Consortium |
| EPI_ISL_432983 | Queens Medical Centre, Clinical Microbiology Department / DeepSeq Nottingham | COVID-19 Genomics UK (COG-UK) Consortium |
| EPI_ISL_432984 | Queens Medical Centre, Clinical Microbiology Department / DeepSeq Nottingham | COVID-19 Genomics UK (COG-UK) Consortium |
| EPI_ISL_432985 | Queens Medical Centre, Clinical Microbiology Department / DeepSeq Nottingham | COVID-19 Genomics UK (COG-UK) Consortium |
| EPI_ISL_432986 | Queens Medical Centre, Clinical Microbiology Department / DeepSeq Nottingham | COVID-19 Genomics UK (COG-UK) Consortium |
| EPI_ISL_432987 | Queens Medical Centre, Clinical Microbiology Department / DeepSeq Nottingham | COVID-19 Genomics UK (COG-UK) Consortium |
| EPI_ISL_432988 | Queens Medical Centre, Clinical Microbiology Department / DeepSeq Nottingham | COVID-19 Genomics UK (COG-UK) Consortium |
| EPI_ISL_432989 | Queens Medical Centre, Clinical Microbiology Department / DeepSeq Nottingham | COVID-19 Genomics UK (COG-UK) Consortium |

| EPI_Accession_ID | Originating Laboratory | Submitting Laboratory |
|---|---|---|
| EPI_ISL_432990 | Queens Medical Centre, Clinical Microbiology Department / DeepSeq Nottingham | COVID-19 Genomics UK (COG-UK) Consortium |
| EPI_ISL_432991 | Queens Medical Centre, Clinical Microbiology Department / DeepSeq Nottingham | COVID-19 Genomics UK (COG-UK) Consortium |
| EPI_ISL_432992 | Queens Medical Centre, Clinical Microbiology Department / DeepSeq Nottingham | COVID-19 Genomics UK (COG-UK) Consortium |
| EPI_ISL_432993 | Queens Medical Centre, Clinical Microbiology Department / DeepSeq Nottingham | COVID-19 Genomics UK (COG-UK) Consortium |
| EPI_ISL_432994 | Queens Medical Centre, Clinical Microbiology Department / DeepSeq Nottingham | COVID-19 Genomics UK (COG-UK) Consortium |
| EPI_ISL_432995 | Queens Medical Centre, Clinical Microbiology Department / DeepSeq Nottingham | COVID-19 Genomics UK (COG-UK) Consortium |
| EPI_ISL_432996 | Queens Medical Centre, Clinical Microbiology Department / DeepSeq Nottingham | COVID-19 Genomics UK (COG-UK) Consortium |
| EPI_ISL_432997 | Queens Medical Centre, Clinical Microbiology Department / DeepSeq Nottingham | COVID-19 Genomics UK (COG-UK) Consortium |
| EPI_ISL_432998 | Queens Medical Centre, Clinical Microbiology Department / DeepSeq Nottingham | COVID-19 Genomics UK (COG-UK) Consortium |
| EPI_ISL_432999 | Queens Medical Centre, Clinical Microbiology Department / DeepSeq Nottingham | COVID-19 Genomics UK (COG-UK) Consortium |
| EPI_ISL_433000 | Queens Medical Centre, Clinical Microbiology Department / DeepSeq Nottingham | COVID-19 Genomics UK (COG-UK) Consortium |
| EPI_ISL_433001 | Queens Medical Centre, Clinical Microbiology Department / DeepSeq Nottingham | COVID-19 Genomics UK (COG-UK) Consortium |
| EPI_ISL_433002 | Queens Medical Centre, Clinical Microbiology Department / DeepSeq Nottingham | COVID-19 Genomics UK (COG-UK) Consortium |
| EPI_ISL_433003 | Queens Medical Centre, Clinical Microbiology Department / DeepSeq Nottingham | COVID-19 Genomics UK (COG-UK) Consortium |
| EPI_ISL_433004 | Queens Medical Centre, Clinical Microbiology Department / DeepSeq Nottingham | COVID-19 Genomics UK (COG-UK) Consortium |
| EPI_ISL_433005 | Queens Medical Centre, Clinical Microbiology Department / DeepSeq Nottingham | COVID-19 Genomics UK (COG-UK) Consortium |

| EPI_Accession_ID | Originating Laboratory | Submitting Laboratory |
|---|---|---|
| EPI_ISL_433006 | Queens Medical Centre, Clinical Microbiology Department / DeepSeq Nottingham | COVID-19 Genomics UK (COG-UK) Consortium |
| EPI_ISL_433007 | Queens Medical Centre, Clinical Microbiology Department / DeepSeq Nottingham | COVID-19 Genomics UK (COG-UK) Consortium |
| EPI_ISL_433008 | Queens Medical Centre, Clinical Microbiology Department / DeepSeq Nottingham | COVID-19 Genomics UK (COG-UK) Consortium |
| EPI_ISL_433009 | Queens Medical Centre, Clinical Microbiology Department / DeepSeq Nottingham | COVID-19 Genomics UK (COG-UK) Consortium |
| EPI_ISL_433010 | Queens Medical Centre, Clinical Microbiology Department / DeepSeq Nottingham | COVID-19 Genomics UK (COG-UK) Consortium |
| EPI_ISL_433011 | Queens Medical Centre, Clinical Microbiology Department / DeepSeq Nottingham | COVID-19 Genomics UK (COG-UK) Consortium |
| EPI_ISL_433012 | Queens Medical Centre, Clinical Microbiology Department / DeepSeq Nottingham | COVID-19 Genomics UK (COG-UK) Consortium |
| EPI_ISL_433013 | Queens Medical Centre, Clinical Microbiology Department / DeepSeq Nottingham | COVID-19 Genomics UK (COG-UK) Consortium |
| EPI_ISL_433014 | Queens Medical Centre, Clinical Microbiology Department / DeepSeq Nottingham | COVID-19 Genomics UK (COG-UK) Consortium |
| EPI_ISL_433015 | Queens Medical Centre, Clinical Microbiology Department / DeepSeq Nottingham | COVID-19 Genomics UK (COG-UK) Consortium |
| EPI_ISL_433016 | Queens Medical Centre, Clinical Microbiology Department / DeepSeq Nottingham | COVID-19 Genomics UK (COG-UK) Consortium |
| EPI_ISL_433017 | Queens Medical Centre, Clinical Microbiology Department / DeepSeq Nottingham | COVID-19 Genomics UK (COG-UK) Consortium |
| EPI_ISL_433018 | Queens Medical Centre, Clinical Microbiology Department / DeepSeq Nottingham | COVID-19 Genomics UK (COG-UK) Consortium |
| EPI_ISL_433019 | Queens Medical Centre, Clinical Microbiology Department / DeepSeq Nottingham | COVID-19 Genomics UK (COG-UK) Consortium |
| EPI_ISL_433020 | Queens Medical Centre, Clinical Microbiology Department / DeepSeq Nottingham | COVID-19 Genomics UK (COG-UK) Consortium |
| EPI_ISL_433021 | Queens Medical Centre, Clinical Microbiology Department / DeepSeq Nottingham | COVID-19 Genomics UK (COG-UK) Consortium |

| EPI_Accession_ID | Originating Laboratory | Submitting Laboratory |
|---|---|---|
| EPI_ISL_433022 | Queens Medical Centre, Clinical Microbiology Department / DeepSeq Nottingham | COVID-19 Genomics UK (COG-UK) Consortium |
| EPI_ISL_433023 | Queens Medical Centre, Clinical Microbiology Department / DeepSeq Nottingham | COVID-19 Genomics UK (COG-UK) Consortium |
| EPI_ISL_433024 | Queens Medical Centre, Clinical Microbiology Department / DeepSeq Nottingham | COVID-19 Genomics UK (COG-UK) Consortium |
| EPI_ISL_433025 | Queens Medical Centre, Clinical Microbiology Department / DeepSeq Nottingham | COVID-19 Genomics UK (COG-UK) Consortium |
| EPI_ISL_433026 | Queens Medical Centre, Clinical Microbiology Department / DeepSeq Nottingham | COVID-19 Genomics UK (COG-UK) Consortium |
| EPI_ISL_433027 | Queens Medical Centre, Clinical Microbiology Department / DeepSeq Nottingham | COVID-19 Genomics UK (COG-UK) Consortium |
| EPI_ISL_433028 | Queens Medical Centre, Clinical Microbiology Department / DeepSeq Nottingham | COVID-19 Genomics UK (COG-UK) Consortium |
| EPI_ISL_433029 | Queens Medical Centre, Clinical Microbiology Department / DeepSeq Nottingham | COVID-19 Genomics UK (COG-UK) Consortium |
| EPI_ISL_433030 | Queens Medical Centre, Clinical Microbiology Department / DeepSeq Nottingham | COVID-19 Genomics UK (COG-UK) Consortium |
| EPI_ISL_433031 | Queens Medical Centre, Clinical Microbiology Department / DeepSeq Nottingham | COVID-19 Genomics UK (COG-UK) Consortium |
| EPI_ISL_433032 | Queens Medical Centre, Clinical Microbiology Department / DeepSeq Nottingham | COVID-19 Genomics UK (COG-UK) Consortium |
| EPI_ISL_433033 | Queens Medical Centre, Clinical Microbiology Department / DeepSeq Nottingham | COVID-19 Genomics UK (COG-UK) Consortium |
| EPI_ISL_433034 | Queens Medical Centre, Clinical Microbiology Department / DeepSeq Nottingham | COVID-19 Genomics UK (COG-UK) Consortium |
| EPI_ISL_433035 | Queens Medical Centre, Clinical Microbiology Department / DeepSeq Nottingham | COVID-19 Genomics UK (COG-UK) Consortium |
| EPI_ISL_433036 | Queens Medical Centre, Clinical Microbiology Department / DeepSeq Nottingham | COVID-19 Genomics UK (COG-UK) Consortium |
| EPI_ISL_433037 | Queens Medical Centre, Clinical Microbiology Department / DeepSeq Nottingham | COVID-19 Genomics UK (COG-UK) Consortium |

| EPI_Accession_ID | Originating Laboratory | Submitting Laboratory |
|---|---|---|
| EPI_ISL_433038 | Queens Medical Centre, Clinical Microbiology Department / DeepSeq Nottingham | COVID-19 Genomics UK (COG-UK) Consortium |
| EPI_ISL_433039 | Queens Medical Centre, Clinical Microbiology Department / DeepSeq Nottingham | COVID-19 Genomics UK (COG-UK) Consortium |
| EPI_ISL_433040 | Queens Medical Centre, Clinical Microbiology Department / DeepSeq Nottingham | COVID-19 Genomics UK (COG-UK) Consortium |
| EPI_ISL_433041 | Queens Medical Centre, Clinical Microbiology Department / DeepSeq Nottingham | COVID-19 Genomics UK (COG-UK) Consortium |
| EPI_ISL_433042 | Queens Medical Centre, Clinical Microbiology Department / DeepSeq Nottingham | COVID-19 Genomics UK (COG-UK) Consortium |
| EPI_ISL_433043 | Queens Medical Centre, Clinical Microbiology Department / DeepSeq Nottingham | COVID-19 Genomics UK (COG-UK) Consortium |
| EPI_ISL_433044 | Queens Medical Centre, Clinical Microbiology Department / DeepSeq Nottingham | COVID-19 Genomics UK (COG-UK) Consortium |
| EPI_ISL_433045 | Queens Medical Centre, Clinical Microbiology Department / DeepSeq Nottingham | COVID-19 Genomics UK (COG-UK) Consortium |
| EPI_ISL_433046 | Queens Medical Centre, Clinical Microbiology Department / DeepSeq Nottingham | COVID-19 Genomics UK (COG-UK) Consortium |
| EPI_ISL_433047 | Queens Medical Centre, Clinical Microbiology Department / DeepSeq Nottingham | COVID-19 Genomics UK (COG-UK) Consortium |
| EPI_ISL_433048 | Queens Medical Centre, Clinical Microbiology Department / DeepSeq Nottingham | COVID-19 Genomics UK (COG-UK) Consortium |
| EPI_ISL_433049 | Queens Medical Centre, Clinical Microbiology Department / DeepSeq Nottingham | COVID-19 Genomics UK (COG-UK) Consortium |
| EPI_ISL_433050 | Queens Medical Centre, Clinical Microbiology Department / DeepSeq Nottingham | COVID-19 Genomics UK (COG-UK) Consortium |
| EPI_ISL_433051 | Queens Medical Centre, Clinical Microbiology Department / DeepSeq Nottingham | COVID-19 Genomics UK (COG-UK) Consortium |
| EPI_ISL_433052 | Queens Medical Centre, Clinical Microbiology Department / DeepSeq Nottingham | COVID-19 Genomics UK (COG-UK) Consortium |
| EPI_ISL_433053 | Queens Medical Centre, Clinical Microbiology Department / DeepSeq Nottingham | COVID-19 Genomics UK (COG-UK) Consortium |

| EPI_Accession_ID | Originating Laboratory | Submitting Laboratory |
|---|---|---|
| EPI_ISL_433054 | Queens Medical Centre, Clinical Microbiology Department / DeepSeq Nottingham | COVID-19 Genomics UK (COG-UK) Consortium |
| EPI_ISL_433055 | Queens Medical Centre, Clinical Microbiology Department / DeepSeq Nottingham | COVID-19 Genomics UK (COG-UK) Consortium |
| EPI_ISL_433056 | Queens Medical Centre, Clinical Microbiology Department / DeepSeq Nottingham | COVID-19 Genomics UK (COG-UK) Consortium |
| EPI_ISL_433057 | Queens Medical Centre, Clinical Microbiology Department / DeepSeq Nottingham | COVID-19 Genomics UK (COG-UK) Consortium |
| EPI_ISL_433058 | Queens Medical Centre, Clinical Microbiology Department / DeepSeq Nottingham | COVID-19 Genomics UK (COG-UK) Consortium |
| EPI_ISL_433059 | Queens Medical Centre, Clinical Microbiology Department / DeepSeq Nottingham | COVID-19 Genomics UK (COG-UK) Consortium |
| EPI_ISL_433060 | Queens Medical Centre, Clinical Microbiology Department / DeepSeq Nottingham | COVID-19 Genomics UK (COG-UK) Consortium |
| EPI_ISL_433061 | Queens Medical Centre, Clinical Microbiology Department / DeepSeq Nottingham | COVID-19 Genomics UK (COG-UK) Consortium |
| EPI_ISL_433062 | Queens Medical Centre, Clinical Microbiology Department / DeepSeq Nottingham | COVID-19 Genomics UK (COG-UK) Consortium |
| EPI_ISL_433063 | Queens Medical Centre, Clinical Microbiology Department / DeepSeq Nottingham | COVID-19 Genomics UK (COG-UK) Consortium |
| EPI_ISL_433064 | Queens Medical Centre, Clinical Microbiology Department / DeepSeq Nottingham | COVID-19 Genomics UK (COG-UK) Consortium |
| EPI_ISL_433065 | Queens Medical Centre, Clinical Microbiology Department / DeepSeq Nottingham | COVID-19 Genomics UK (COG-UK) Consortium |
| EPI_ISL_433066 | Queens Medical Centre, Clinical Microbiology Department / DeepSeq Nottingham | COVID-19 Genomics UK (COG-UK) Consortium |
| EPI_ISL_433067 | Queens Medical Centre, Clinical Microbiology Department / DeepSeq Nottingham | COVID-19 Genomics UK (COG-UK) Consortium |
| EPI_ISL_433068 | Virology Department, Royal Infirmary of Edinburgh, NHS Lothian / School of Biological Sciences, University of Edinburgh / Institute of Genetics and Molecular Medicine, University of Edinburgh | COVID-19 Genomics UK (COG-UK) Consortium |

| EPI_Accession_ID | Originating Laboratory | Submitting Laboratory |
|---|---|---|
| EPI_ISL_433069 | Virology Department, Royal Infirmary of Edinburgh, NHS Lothian / School of Biological Sciences, University of Edinburgh / Institute of Genetics and Molecular Medicine, University of Edinburgh | COVID-19 Genomics UK (COG-UK) Consortium |
| EPI_ISL_433070 | Virology Department, Royal Infirmary of Edinburgh, NHS Lothian / School of Biological Sciences, University of Edinburgh / Institute of Genetics and Molecular Medicine, University of Edinburgh | COVID-19 Genomics UK (COG-UK) Consortium |
| EPI_ISL_433071 | Virology Department, Royal Infirmary of Edinburgh, NHS Lothian / School of Biological Sciences, University of Edinburgh / Institute of Genetics and Molecular Medicine, University of Edinburgh | COVID-19 Genomics UK (COG-UK) Consortium |
| EPI_ISL_433072 | Virology Department, Royal Infirmary of Edinburgh, NHS Lothian / School of Biological Sciences, University of Edinburgh / Institute of Genetics and Molecular Medicine, University of Edinburgh | COVID-19 Genomics UK (COG-UK) Consortium |
| EPI_ISL_433073 | Virology Department, Royal Infirmary of Edinburgh, NHS Lothian / School of Biological Sciences, University of Edinburgh / Institute of Genetics and Molecular Medicine, University of Edinburgh | COVID-19 Genomics UK (COG-UK) Consortium |
| EPI_ISL_433074 | Virology Department, Royal Infirmary of Edinburgh, NHS Lothian / School of Biological Sciences, University of Edinburgh / Institute of Genetics and Molecular Medicine, University of Edinburgh | COVID-19 Genomics UK (COG-UK) Consortium |
| EPI_ISL_433075 | Virology Department, Royal Infirmary of Edinburgh, NHS Lothian / School of Biological Sciences, University of Edinburgh / Institute of Genetics and Molecular Medicine, University of Edinburgh | COVID-19 Genomics UK (COG-UK) Consortium |
| EPI_ISL_433076 | Virology Department, Royal Infirmary of Edinburgh, NHS Lothian / School of Biological Sciences, University of Edinburgh / Institute of Genetics and Molecular Medicine, University of Edinburgh | COVID-19 Genomics UK (COG-UK) Consortium |

| EPI_Accession_ID | Originating Laboratory | Submitting Laboratory |
|---|---|---|
| EPI_ISL_433077 | Virology Department, Royal Infirmary of Edinburgh, NHS Lothian / School of Biological Sciences, University of Edinburgh / Institute of Genetics and Molecular Medicine, University of Edinburgh | COVID-19 Genomics UK (COG-UK) Consortium |
| EPI_ISL_433078 | Virology Department, Royal Infirmary of Edinburgh, NHS Lothian / School of Biological Sciences, University of Edinburgh / Institute of Genetics and Molecular Medicine, University of Edinburgh | COVID-19 Genomics UK (COG-UK) Consortium |
| EPI_ISL_433079 | Virology Department, Royal Infirmary of Edinburgh, NHS Lothian / School of Biological Sciences, University of Edinburgh / Institute of Genetics and Molecular Medicine, University of Edinburgh | COVID-19 Genomics UK (COG-UK) Consortium |
| EPI_ISL_433080 | Virology Department, Royal Infirmary of Edinburgh, NHS Lothian / School of Biological Sciences, University of Edinburgh / Institute of Genetics and Molecular Medicine, University of Edinburgh | COVID-19 Genomics UK (COG-UK) Consortium |
| EPI_ISL_433081 | Virology Department, Royal Infirmary of Edinburgh, NHS Lothian / School of Biological Sciences, University of Edinburgh / Institute of Genetics and Molecular Medicine, University of Edinburgh | COVID-19 Genomics UK (COG-UK) Consortium |
| EPI_ISL_433082 | Virology Department, Royal Infirmary of Edinburgh, NHS Lothian / School of Biological Sciences, University of Edinburgh / Institute of Genetics and Molecular Medicine, University of Edinburgh | COVID-19 Genomics UK (COG-UK) Consortium |
| EPI_ISL_433083 | Virology Department, Royal Infirmary of Edinburgh, NHS Lothian / School of Biological Sciences, University of Edinburgh / Institute of Genetics and Molecular Medicine, University of Edinburgh | COVID-19 Genomics UK (COG-UK) Consortium |
| EPI_ISL_433084 | Virology Department, Royal Infirmary of Edinburgh, NHS Lothian / School of Biological Sciences, University of Edinburgh / Institute of Genetics and Molecular Medicine, University of Edinburgh | COVID-19 Genomics UK (COG-UK) Consortium |

| EPI_Accession_ID | Originating Laboratory | Submitting Laboratory |
|---|---|---|
| EPI_ISL_433085 | Virology Department, Royal Infirmary of Edinburgh, NHS Lothian / School of Biological Sciences, University of Edinburgh / Institute of Genetics and Molecular Medicine, University of Edinburgh | COVID-19 Genomics UK (COG-UK) Consortium |
| EPI_ISL_433086 | Virology Department, Royal Infirmary of Edinburgh, NHS Lothian / School of Biological Sciences, University of Edinburgh / Institute of Genetics and Molecular Medicine, University of Edinburgh | COVID-19 Genomics UK (COG-UK) Consortium |
| EPI_ISL_433087 | Virology Department, Royal Infirmary of Edinburgh, NHS Lothian / School of Biological Sciences, University of Edinburgh / Institute of Genetics and Molecular Medicine, University of Edinburgh | COVID-19 Genomics UK (COG-UK) Consortium |
| EPI_ISL_433088 | Virology Department, Royal Infirmary of Edinburgh, NHS Lothian / School of Biological Sciences, University of Edinburgh / Institute of Genetics and Molecular Medicine, University of Edinburgh | COVID-19 Genomics UK (COG-UK) Consortium |
| EPI_ISL_433089 | Virology Department, Royal Infirmary of Edinburgh, NHS Lothian / School of Biological Sciences, University of Edinburgh / Institute of Genetics and Molecular Medicine, University of Edinburgh | COVID-19 Genomics UK (COG-UK) Consortium |
| EPI_ISL_433090 | Virology Department, Royal Infirmary of Edinburgh, NHS Lothian / School of Biological Sciences, University of Edinburgh / Institute of Genetics and Molecular Medicine, University of Edinburgh | COVID-19 Genomics UK (COG-UK) Consortium |
| EPI_ISL_433091 | Virology Department, Royal Infirmary of Edinburgh, NHS Lothian / School of Biological Sciences, University of Edinburgh / Institute of Genetics and Molecular Medicine, University of Edinburgh | COVID-19 Genomics UK (COG-UK) Consortium |
| EPI_ISL_433092 | Virology Department, Royal Infirmary of Edinburgh, NHS Lothian / School of Biological Sciences, University of Edinburgh / Institute of Genetics and Molecular Medicine, University of Edinburgh | COVID-19 Genomics UK (COG-UK) Consortium |

| EPI_Accession_ID | Originating Laboratory | Submitting Laboratory |
|---|---|---|
| EPI_ISL_433093 | Virology Department, Royal Infirmary of Edinburgh, NHS Lothian / School of Biological Sciences, University of Edinburgh / Institute of Genetics and Molecular Medicine, University of Edinburgh | COVID-19 Genomics UK (COG-UK) Consortium |
| EPI_ISL_433094 | Virology Department, Royal Infirmary of Edinburgh, NHS Lothian / School of Biological Sciences, University of Edinburgh / Institute of Genetics and Molecular Medicine, University of Edinburgh | COVID-19 Genomics UK (COG-UK) Consortium |
| EPI_ISL_433095 | Virology Department, Royal Infirmary of Edinburgh, NHS Lothian / School of Biological Sciences, University of Edinburgh / Institute of Genetics and Molecular Medicine, University of Edinburgh | COVID-19 Genomics UK (COG-UK) Consortium |
| EPI_ISL_433096 | Virology Department, Royal Infirmary of Edinburgh, NHS Lothian / School of Biological Sciences, University of Edinburgh / Institute of Genetics and Molecular Medicine, University of Edinburgh | COVID-19 Genomics UK (COG-UK) Consortium |
| EPI_ISL_433097 | Virology Department, Royal Infirmary of Edinburgh, NHS Lothian / School of Biological Sciences, University of Edinburgh / Institute of Genetics and Molecular Medicine, University of Edinburgh | COVID-19 Genomics UK (COG-UK) Consortium |
| EPI_ISL_433098 | Virology Department, Royal Infirmary of Edinburgh, NHS Lothian / School of Biological Sciences, University of Edinburgh / Institute of Genetics and Molecular Medicine, University of Edinburgh | COVID-19 Genomics UK (COG-UK) Consortium |
| EPI_ISL_433099 | Virology Department, Royal Infirmary of Edinburgh, NHS Lothian / School of Biological Sciences, University of Edinburgh / Institute of Genetics and Molecular Medicine, University of Edinburgh | COVID-19 Genomics UK (COG-UK) Consortium |
| EPI_ISL_433100 | Virology Department, Royal Infirmary of Edinburgh, NHS Lothian / School of Biological Sciences, University of Edinburgh / Institute of Genetics and Molecular Medicine, University of Edinburgh | COVID-19 Genomics UK (COG-UK) Consortium |

| EPI_Accession_ID | Originating Laboratory | Submitting Laboratory |
|---|---|---|
| EPI_ISL_433101 | Virology Department, Royal Infirmary of Edinburgh, NHS Lothian / School of Biological Sciences, University of Edinburgh / Institute of Genetics and Molecular Medicine, University of Edinburgh | COVID-19 Genomics UK (COG-UK) Consortium |
| EPI_ISL_433102 | Virology Department, Royal Infirmary of Edinburgh, NHS Lothian / School of Biological Sciences, University of Edinburgh / Institute of Genetics and Molecular Medicine, University of Edinburgh | COVID-19 Genomics UK (COG-UK) Consortium |
| EPI_ISL_433103 | Virology Department, Royal Infirmary of Edinburgh, NHS Lothian / School of Biological Sciences, University of Edinburgh / Institute of Genetics and Molecular Medicine, University of Edinburgh | COVID-19 Genomics UK (COG-UK) Consortium |
| EPI_ISL_433104 | Virology Department, Royal Infirmary of Edinburgh, NHS Lothian / School of Biological Sciences, University of Edinburgh / Institute of Genetics and Molecular Medicine, University of Edinburgh | COVID-19 Genomics UK (COG-UK) Consortium |
| EPI_ISL_433105 | Virology Department, Royal Infirmary of Edinburgh, NHS Lothian / School of Biological Sciences, University of Edinburgh / Institute of Genetics and Molecular Medicine, University of Edinburgh | COVID-19 Genomics UK (COG-UK) Consortium |
| EPI_ISL_433106 | Virology Department, Royal Infirmary of Edinburgh, NHS Lothian / School of Biological Sciences, University of Edinburgh / Institute of Genetics and Molecular Medicine, University of Edinburgh | COVID-19 Genomics UK (COG-UK) Consortium |
| EPI_ISL_433107 | Virology Department, Royal Infirmary of Edinburgh, NHS Lothian / School of Biological Sciences, University of Edinburgh / Institute of Genetics and Molecular Medicine, University of Edinburgh | COVID-19 Genomics UK (COG-UK) Consortium |
| EPI_ISL_433108 | Virology Department, Royal Infirmary of Edinburgh, NHS Lothian / School of Biological Sciences, University of Edinburgh / Institute of Genetics and Molecular Medicine, University of Edinburgh | COVID-19 Genomics UK (COG-UK) Consortium |

| EPI_Accession_ID | Originating Laboratory | Submitting Laboratory |
|---|---|---|
| EPI_ISL_433109 | Virology Department, Royal Infirmary of Edinburgh, NHS Lothian / School of Biological Sciences, University of Edinburgh / Institute of Genetics and Molecular Medicine, University of Edinburgh | COVID-19 Genomics UK (COG-UK) Consortium |
| EPI_ISL_433110 | Virology Department, Royal Infirmary of Edinburgh, NHS Lothian / School of Biological Sciences, University of Edinburgh / Institute of Genetics and Molecular Medicine, University of Edinburgh | COVID-19 Genomics UK (COG-UK) Consortium |
| EPI_ISL_433111 | Virology Department, Royal Infirmary of Edinburgh, NHS Lothian / School of Biological Sciences, University of Edinburgh / Institute of Genetics and Molecular Medicine, University of Edinburgh | COVID-19 Genomics UK (COG-UK) Consortium |
| EPI_ISL_433112 | Virology Department, Royal Infirmary of Edinburgh, NHS Lothian / School of Biological Sciences, University of Edinburgh / Institute of Genetics and Molecular Medicine, University of Edinburgh | COVID-19 Genomics UK (COG-UK) Consortium |
| EPI_ISL_433113 | Virology Department, Royal Infirmary of Edinburgh, NHS Lothian / School of Biological Sciences, University of Edinburgh / Institute of Genetics and Molecular Medicine, University of Edinburgh | COVID-19 Genomics UK (COG-UK) Consortium |
| EPI_ISL_433114 | Virology Department, Royal Infirmary of Edinburgh, NHS Lothian / School of Biological Sciences, University of Edinburgh / Institute of Genetics and Molecular Medicine, University of Edinburgh | COVID-19 Genomics UK (COG-UK) Consortium |
| EPI_ISL_433115 | Virology Department, Royal Infirmary of Edinburgh, NHS Lothian / School of Biological Sciences, University of Edinburgh / Institute of Genetics and Molecular Medicine, University of Edinburgh | COVID-19 Genomics UK (COG-UK) Consortium |
| EPI_ISL_433116 | Virology Department, Royal Infirmary of Edinburgh, NHS Lothian / School of Biological Sciences, University of Edinburgh / Institute of Genetics and Molecular Medicine, University of Edinburgh | COVID-19 Genomics UK (COG-UK) Consortium |

| EPI_Accession_ID | Originating Laboratory | Submitting Laboratory |
|---|---|---|
| EPI_ISL_433117 | Virology Department, Royal Infirmary of Edinburgh, NHS Lothian / School of Biological Sciences, University of Edinburgh / Institute of Genetics and Molecular Medicine, University of Edinburgh | COVID-19 Genomics UK (COG-UK) Consortium |
| EPI_ISL_433118 | Virology Department, Royal Infirmary of Edinburgh, NHS Lothian / School of Biological Sciences, University of Edinburgh / Institute of Genetics and Molecular Medicine, University of Edinburgh | COVID-19 Genomics UK (COG-UK) Consortium |
| EPI_ISL_433119 | Virology Department, Royal Infirmary of Edinburgh, NHS Lothian / School of Biological Sciences, University of Edinburgh / Institute of Genetics and Molecular Medicine, University of Edinburgh | COVID-19 Genomics UK (COG-UK) Consortium |
| EPI_ISL_433120 | Virology Department, Royal Infirmary of Edinburgh, NHS Lothian / School of Biological Sciences, University of Edinburgh / Institute of Genetics and Molecular Medicine, University of Edinburgh | COVID-19 Genomics UK (COG-UK) Consortium |
| EPI_ISL_433121 | Virology Department, Royal Infirmary of Edinburgh, NHS Lothian / School of Biological Sciences, University of Edinburgh / Institute of Genetics and Molecular Medicine, University of Edinburgh | COVID-19 Genomics UK (COG-UK) Consortium |
| EPI_ISL_433122 | Virology Department, Royal Infirmary of Edinburgh, NHS Lothian / School of Biological Sciences, University of Edinburgh / Institute of Genetics and Molecular Medicine, University of Edinburgh | COVID-19 Genomics UK (COG-UK) Consortium |
| EPI_ISL_433123 | Virology Department, Royal Infirmary of Edinburgh, NHS Lothian / School of Biological Sciences, University of Edinburgh / Institute of Genetics and Molecular Medicine, University of Edinburgh | COVID-19 Genomics UK (COG-UK) Consortium |
| EPI_ISL_433124 | Virology Department, Royal Infirmary of Edinburgh, NHS Lothian / School of Biological Sciences, University of Edinburgh / Institute of Genetics and Molecular Medicine, University of Edinburgh | COVID-19 Genomics UK (COG-UK) Consortium |

| EPI_Accession_ID | Originating Laboratory | Submitting Laboratory |
|---|---|---|
| EPI_ISL_433125 | Virology Department, Royal Infirmary of Edinburgh, NHS Lothian / School of Biological Sciences, University of Edinburgh / Institute of Genetics and Molecular Medicine, University of Edinburgh | COVID-19 Genomics UK (COG-UK) Consortium |
| EPI_ISL_433126 | Virology Department, Royal Infirmary of Edinburgh, NHS Lothian / School of Biological Sciences, University of Edinburgh / Institute of Genetics and Molecular Medicine, University of Edinburgh | COVID-19 Genomics UK (COG-UK) Consortium |
| EPI_ISL_433127 | Virology Department, Royal Infirmary of Edinburgh, NHS Lothian / School of Biological Sciences, University of Edinburgh / Institute of Genetics and Molecular Medicine, University of Edinburgh | COVID-19 Genomics UK (COG-UK) Consortium |
| EPI_ISL_433128 | Virology Department, Royal Infirmary of Edinburgh, NHS Lothian / School of Biological Sciences, University of Edinburgh / Institute of Genetics and Molecular Medicine, University of Edinburgh | COVID-19 Genomics UK (COG-UK) Consortium |
| EPI_ISL_433129 | Virology Department, Royal Infirmary of Edinburgh, NHS Lothian / School of Biological Sciences, University of Edinburgh / Institute of Genetics and Molecular Medicine, University of Edinburgh | COVID-19 Genomics UK (COG-UK) Consortium |
| EPI_ISL_433130 | Virology Department, Royal Infirmary of Edinburgh, NHS Lothian / School of Biological Sciences, University of Edinburgh / Institute of Genetics and Molecular Medicine, University of Edinburgh | COVID-19 Genomics UK (COG-UK) Consortium |
| EPI_ISL_433131 | Virology Department, Royal Infirmary of Edinburgh, NHS Lothian / School of Biological Sciences, University of Edinburgh / Institute of Genetics and Molecular Medicine, University of Edinburgh | COVID-19 Genomics UK (COG-UK) Consortium |
| EPI_ISL_433132 | Virology Department, Royal Infirmary of Edinburgh, NHS Lothian / School of Biological Sciences, University of Edinburgh / Institute of Genetics and Molecular Medicine, University of Edinburgh | COVID-19 Genomics UK (COG-UK) Consortium |

| EPI_Accession_ID | Originating Laboratory | Submitting Laboratory |
|---|---|---|
| EPI_ISL_433133 | Virology Department, Royal Infirmary of Edinburgh, NHS Lothian / School of Biological Sciences, University of Edinburgh / Institute of Genetics and Molecular Medicine, University of Edinburgh | COVID-19 Genomics UK (COG-UK) Consortium |
| EPI_ISL_433134 | Virology Department, Royal Infirmary of Edinburgh, NHS Lothian / School of Biological Sciences, University of Edinburgh / Institute of Genetics and Molecular Medicine, University of Edinburgh | COVID-19 Genomics UK (COG-UK) Consortium |
| EPI_ISL_433135 | Virology Department, Royal Infirmary of Edinburgh, NHS Lothian / School of Biological Sciences, University of Edinburgh / Institute of Genetics and Molecular Medicine, University of Edinburgh | COVID-19 Genomics UK (COG-UK) Consortium |
| EPI_ISL_433136 | Virology Department, Royal Infirmary of Edinburgh, NHS Lothian / School of Biological Sciences, University of Edinburgh / Institute of Genetics and Molecular Medicine, University of Edinburgh | COVID-19 Genomics UK (COG-UK) Consortium |
| EPI_ISL_433137 | Virology Department, Royal Infirmary of Edinburgh, NHS Lothian / School of Biological Sciences, University of Edinburgh / Institute of Genetics and Molecular Medicine, University of Edinburgh | COVID-19 Genomics UK (COG-UK) Consortium |
| EPI_ISL_433138 | Virology Department, Royal Infirmary of Edinburgh, NHS Lothian / School of Biological Sciences, University of Edinburgh / Institute of Genetics and Molecular Medicine, University of Edinburgh | COVID-19 Genomics UK (COG-UK) Consortium |
| EPI_ISL_433139 | Virology Department, Royal Infirmary of Edinburgh, NHS Lothian / School of Biological Sciences, University of Edinburgh / Institute of Genetics and Molecular Medicine, University of Edinburgh | COVID-19 Genomics UK (COG-UK) Consortium |
| EPI_ISL_433140 | Virology Department, Royal Infirmary of Edinburgh, NHS Lothian / School of Biological Sciences, University of Edinburgh / Institute of Genetics and Molecular Medicine, University of Edinburgh | COVID-19 Genomics UK (COG-UK) Consortium |

| EPI_Accession_ID | Originating Laboratory | Submitting Laboratory |
|---|---|---|
| EPI_ISL_433141 | Virology Department, Royal Infirmary of Edinburgh, NHS Lothian / School of Biological Sciences, University of Edinburgh / Institute of Genetics and Molecular Medicine, University of Edinburgh | COVID-19 Genomics UK (COG-UK) Consortium |
| EPI_ISL_433142 | Virology Department, Royal Infirmary of Edinburgh, NHS Lothian / School of Biological Sciences, University of Edinburgh / Institute of Genetics and Molecular Medicine, University of Edinburgh | COVID-19 Genomics UK (COG-UK) Consortium |
| EPI_ISL_433143 | Virology Department, Royal Infirmary of Edinburgh, NHS Lothian / School of Biological Sciences, University of Edinburgh / Institute of Genetics and Molecular Medicine, University of Edinburgh | COVID-19 Genomics UK (COG-UK) Consortium |
| EPI_ISL_433144 | Virology Department, Royal Infirmary of Edinburgh, NHS Lothian / School of Biological Sciences, University of Edinburgh / Institute of Genetics and Molecular Medicine, University of Edinburgh | COVID-19 Genomics UK (COG-UK) Consortium |
| EPI_ISL_433145 | Virology Department, Royal Infirmary of Edinburgh, NHS Lothian / School of Biological Sciences, University of Edinburgh / Institute of Genetics and Molecular Medicine, University of Edinburgh | COVID-19 Genomics UK (COG-UK) Consortium |
| EPI_ISL_433146 | Virology Department, Royal Infirmary of Edinburgh, NHS Lothian / School of Biological Sciences, University of Edinburgh / Institute of Genetics and Molecular Medicine, University of Edinburgh | COVID-19 Genomics UK (COG-UK) Consortium |
| EPI_ISL_433147 | Virology Department, Royal Infirmary of Edinburgh, NHS Lothian / School of Biological Sciences, University of Edinburgh / Institute of Genetics and Molecular Medicine, University of Edinburgh | COVID-19 Genomics UK (COG-UK) Consortium |
| EPI_ISL_433148 | Virology Department, Royal Infirmary of Edinburgh, NHS Lothian / School of Biological Sciences, University of Edinburgh / Institute of Genetics and Molecular Medicine, University of Edinburgh | COVID-19 Genomics UK (COG-UK) Consortium |

| EPI_Accession_ID | Originating Laboratory | Submitting Laboratory |
|---|---|---|
| EPI_ISL_433149 | Virology Department, Royal Infirmary of Edinburgh, NHS Lothian / School of Biological Sciences, University of Edinburgh / Institute of Genetics and Molecular Medicine, University of Edinburgh | COVID-19 Genomics UK (COG-UK) Consortium |
| EPI_ISL_433150 | Virology Department, Royal Infirmary of Edinburgh, NHS Lothian / School of Biological Sciences, University of Edinburgh / Institute of Genetics and Molecular Medicine, University of Edinburgh | COVID-19 Genomics UK (COG-UK) Consortium |
| EPI_ISL_433151 | Virology Department, Royal Infirmary of Edinburgh, NHS Lothian / School of Biological Sciences, University of Edinburgh / Institute of Genetics and Molecular Medicine, University of Edinburgh | COVID-19 Genomics UK (COG-UK) Consortium |
| EPI_ISL_433152 | Virology Department, Royal Infirmary of Edinburgh, NHS Lothian / School of Biological Sciences, University of Edinburgh / Institute of Genetics and Molecular Medicine, University of Edinburgh | COVID-19 Genomics UK (COG-UK) Consortium |
| EPI_ISL_433153 | Virology Department, Royal Infirmary of Edinburgh, NHS Lothian / School of Biological Sciences, University of Edinburgh / Institute of Genetics and Molecular Medicine, University of Edinburgh | COVID-19 Genomics UK (COG-UK) Consortium |
| EPI_ISL_433154 | Virology Department, Royal Infirmary of Edinburgh, NHS Lothian / School of Biological Sciences, University of Edinburgh / Institute of Genetics and Molecular Medicine, University of Edinburgh | COVID-19 Genomics UK (COG-UK) Consortium |
| EPI_ISL_433155 | Virology Department, Royal Infirmary of Edinburgh, NHS Lothian / School of Biological Sciences, University of Edinburgh / Institute of Genetics and Molecular Medicine, University of Edinburgh | COVID-19 Genomics UK (COG-UK) Consortium |
| EPI_ISL_433156 | Virology Department, Royal Infirmary of Edinburgh, NHS Lothian / School of Biological Sciences, University of Edinburgh / Institute of Genetics and Molecular Medicine, University of Edinburgh | COVID-19 Genomics UK (COG-UK) Consortium |

| EPI_Accession_ID | Originating Laboratory | Submitting Laboratory |
|---|---|---|
| EPI_ISL_433157 | Virology Department, Royal Infirmary of Edinburgh, NHS Lothian / School of Biological Sciences, University of Edinburgh / Institute of Genetics and Molecular Medicine, University of Edinburgh | COVID-19 Genomics UK (COG-UK) Consortium |
| EPI_ISL_433158 | Virology Department, Royal Infirmary of Edinburgh, NHS Lothian / School of Biological Sciences, University of Edinburgh / Institute of Genetics and Molecular Medicine, University of Edinburgh | COVID-19 Genomics UK (COG-UK) Consortium |
| EPI_ISL_433159 | Virology Department, Royal Infirmary of Edinburgh, NHS Lothian / School of Biological Sciences, University of Edinburgh / Institute of Genetics and Molecular Medicine, University of Edinburgh | COVID-19 Genomics UK (COG-UK) Consortium |
| EPI_ISL_433160 | Virology Department, Royal Infirmary of Edinburgh, NHS Lothian / School of Biological Sciences, University of Edinburgh / Institute of Genetics and Molecular Medicine, University of Edinburgh | COVID-19 Genomics UK (COG-UK) Consortium |
| EPI_ISL_433161 | Virology Department, Royal Infirmary of Edinburgh, NHS Lothian / School of Biological Sciences, University of Edinburgh / Institute of Genetics and Molecular Medicine, University of Edinburgh | COVID-19 Genomics UK (COG-UK) Consortium |
| EPI_ISL_433162 | Virology Department, Royal Infirmary of Edinburgh, NHS Lothian / School of Biological Sciences, University of Edinburgh / Institute of Genetics and Molecular Medicine, University of Edinburgh | COVID-19 Genomics UK (COG-UK) Consortium |
| EPI_ISL_433163 | Virology Department, Royal Infirmary of Edinburgh, NHS Lothian / School of Biological Sciences, University of Edinburgh / Institute of Genetics and Molecular Medicine, University of Edinburgh | COVID-19 Genomics UK (COG-UK) Consortium |
| EPI_ISL_433164 | Virology Department, Royal Infirmary of Edinburgh, NHS Lothian / School of Biological Sciences, University of Edinburgh / Institute of Genetics and Molecular Medicine, University of Edinburgh | COVID-19 Genomics UK (COG-UK) Consortium |

| EPI_Accession_ID | Originating Laboratory | Submitting Laboratory |
|---|---|---|
| EPI_ISL_433165 | Virology Department, Royal Infirmary of Edinburgh, NHS Lothian / School of Biological Sciences, University of Edinburgh / Institute of Genetics and Molecular Medicine, University of Edinburgh | COVID-19 Genomics UK (COG-UK) Consortium |
| EPI_ISL_433166 | Virology Department, Royal Infirmary of Edinburgh, NHS Lothian / School of Biological Sciences, University of Edinburgh / Institute of Genetics and Molecular Medicine, University of Edinburgh | COVID-19 Genomics UK (COG-UK) Consortium |
| EPI_ISL_433167 | Virology Department, Royal Infirmary of Edinburgh, NHS Lothian / School of Biological Sciences, University of Edinburgh / Institute of Genetics and Molecular Medicine, University of Edinburgh | COVID-19 Genomics UK (COG-UK) Consortium |
| EPI_ISL_433168 | Virology Department, Royal Infirmary of Edinburgh, NHS Lothian / School of Biological Sciences, University of Edinburgh / Institute of Genetics and Molecular Medicine, University of Edinburgh | COVID-19 Genomics UK (COG-UK) Consortium |
| EPI_ISL_433169 | Virology Department, Royal Infirmary of Edinburgh, NHS Lothian / School of Biological Sciences, University of Edinburgh / Institute of Genetics and Molecular Medicine, University of Edinburgh | COVID-19 Genomics UK (COG-UK) Consortium |
| EPI_ISL_433170 | Virology Department, Royal Infirmary of Edinburgh, NHS Lothian / School of Biological Sciences, University of Edinburgh / Institute of Genetics and Molecular Medicine, University of Edinburgh | COVID-19 Genomics UK (COG-UK) Consortium |
| EPI_ISL_433171 | Virology Department, Royal Infirmary of Edinburgh, NHS Lothian / School of Biological Sciences, University of Edinburgh / Institute of Genetics and Molecular Medicine, University of Edinburgh | COVID-19 Genomics UK (COG-UK) Consortium |
| EPI_ISL_433172 | Virology Department, Royal Infirmary of Edinburgh, NHS Lothian / School of Biological Sciences, University of Edinburgh / Institute of Genetics and Molecular Medicine, University of Edinburgh | COVID-19 Genomics UK (COG-UK) Consortium |

| EPI_Accession_ID | Originating Laboratory | Submitting Laboratory |
|---|---|---|
| EPI_ISL_433173 | Virology Department, Royal Infirmary of Edinburgh, NHS Lothian / School of Biological Sciences, University of Edinburgh / Institute of Genetics and Molecular Medicine, University of Edinburgh | COVID-19 Genomics UK (COG-UK) Consortium |
| EPI_ISL_433174 | Virology Department, Royal Infirmary of Edinburgh, NHS Lothian / School of Biological Sciences, University of Edinburgh / Institute of Genetics and Molecular Medicine, University of Edinburgh | COVID-19 Genomics UK (COG-UK) Consortium |
| EPI_ISL_433175 | Virology Department, Royal Infirmary of Edinburgh, NHS Lothian / School of Biological Sciences, University of Edinburgh / Institute of Genetics and Molecular Medicine, University of Edinburgh | COVID-19 Genomics UK (COG-UK) Consortium |
| EPI_ISL_433176 | Virology Department, Royal Infirmary of Edinburgh, NHS Lothian / School of Biological Sciences, University of Edinburgh / Institute of Genetics and Molecular Medicine, University of Edinburgh | COVID-19 Genomics UK (COG-UK) Consortium |
| EPI_ISL_433177 | Virology Department, Royal Infirmary of Edinburgh, NHS Lothian / School of Biological Sciences, University of Edinburgh / Institute of Genetics and Molecular Medicine, University of Edinburgh | COVID-19 Genomics UK (COG-UK) Consortium |
| EPI_ISL_433178 | Virology Department, Royal Infirmary of Edinburgh, NHS Lothian / School of Biological Sciences, University of Edinburgh / Institute of Genetics and Molecular Medicine, University of Edinburgh | COVID-19 Genomics UK (COG-UK) Consortium |
| EPI_ISL_433179 | Virology Department, Royal Infirmary of Edinburgh, NHS Lothian / School of Biological Sciences, University of Edinburgh / Institute of Genetics and Molecular Medicine, University of Edinburgh | COVID-19 Genomics UK (COG-UK) Consortium |
| EPI_ISL_433180 | Virology Department, Royal Infirmary of Edinburgh, NHS Lothian / School of Biological Sciences, University of Edinburgh / Institute of Genetics and Molecular Medicine, University of Edinburgh | COVID-19 Genomics UK (COG-UK) Consortium |

| EPI_Accession_ID | Originating Laboratory | Submitting Laboratory |
|---|---|---|
| EPI_ISL_433181 | Virology Department, Royal Infirmary of Edinburgh, NHS Lothian / School of Biological Sciences, University of Edinburgh / Institute of Genetics and Molecular Medicine, University of Edinburgh | COVID-19 Genomics UK (COG-UK) Consortium |
| EPI_ISL_433182 | Virology Department, Royal Infirmary of Edinburgh, NHS Lothian / School of Biological Sciences, University of Edinburgh / Institute of Genetics and Molecular Medicine, University of Edinburgh | COVID-19 Genomics UK (COG-UK) Consortium |
| EPI_ISL_433183 | Virology Department, Royal Infirmary of Edinburgh, NHS Lothian / School of Biological Sciences, University of Edinburgh / Institute of Genetics and Molecular Medicine, University of Edinburgh | COVID-19 Genomics UK (COG-UK) Consortium |
| EPI_ISL_433184 | Virology Department, Royal Infirmary of Edinburgh, NHS Lothian / School of Biological Sciences, University of Edinburgh / Institute of Genetics and Molecular Medicine, University of Edinburgh | COVID-19 Genomics UK (COG-UK) Consortium |
| EPI_ISL_433185 | Virology Department, Royal Infirmary of Edinburgh, NHS Lothian / School of Biological Sciences, University of Edinburgh / Institute of Genetics and Molecular Medicine, University of Edinburgh | COVID-19 Genomics UK (COG-UK) Consortium |
| EPI_ISL_433186 | Virology Department, Royal Infirmary of Edinburgh, NHS Lothian / School of Biological Sciences, University of Edinburgh / Institute of Genetics and Molecular Medicine, University of Edinburgh | COVID-19 Genomics UK (COG-UK) Consortium |
| EPI_ISL_433187 | Virology Department, Royal Infirmary of Edinburgh, NHS Lothian / School of Biological Sciences, University of Edinburgh / Institute of Genetics and Molecular Medicine, University of Edinburgh | COVID-19 Genomics UK (COG-UK) Consortium |
| EPI_ISL_433188 | Virology Department, Royal Infirmary of Edinburgh, NHS Lothian / School of Biological Sciences, University of Edinburgh / Institute of Genetics and Molecular Medicine, University of Edinburgh | COVID-19 Genomics UK (COG-UK) Consortium |

| EPI_Accession_ID | Originating Laboratory | Submitting Laboratory |
|---|---|---|
| EPI_ISL_433189 | Virology Department, Royal Infirmary of Edinburgh, NHS Lothian / School of Biological Sciences, University of Edinburgh / Institute of Genetics and Molecular Medicine, University of Edinburgh | COVID-19 Genomics UK (COG-UK) Consortium |
| EPI_ISL_433190 | Virology Department, Royal Infirmary of Edinburgh, NHS Lothian / School of Biological Sciences, University of Edinburgh / Institute of Genetics and Molecular Medicine, University of Edinburgh | COVID-19 Genomics UK (COG-UK) Consortium |
| EPI_ISL_433191 | Virology Department, Royal Infirmary of Edinburgh, NHS Lothian / School of Biological Sciences, University of Edinburgh / Institute of Genetics and Molecular Medicine, University of Edinburgh | COVID-19 Genomics UK (COG-UK) Consortium |
| EPI_ISL_433192 | Virology Department, Royal Infirmary of Edinburgh, NHS Lothian / School of Biological Sciences, University of Edinburgh / Institute of Genetics and Molecular Medicine, University of Edinburgh | COVID-19 Genomics UK (COG-UK) Consortium |
| EPI_ISL_433193 | Virology Department, Royal Infirmary of Edinburgh, NHS Lothian / School of Biological Sciences, University of Edinburgh / Institute of Genetics and Molecular Medicine, University of Edinburgh | COVID-19 Genomics UK (COG-UK) Consortium |
| EPI_ISL_433194 | Virology Department, Royal Infirmary of Edinburgh, NHS Lothian / School of Biological Sciences, University of Edinburgh / Institute of Genetics and Molecular Medicine, University of Edinburgh | COVID-19 Genomics UK (COG-UK) Consortium |
| EPI_ISL_433195 | Virology Department, Royal Infirmary of Edinburgh, NHS Lothian / School of Biological Sciences, University of Edinburgh / Institute of Genetics and Molecular Medicine, University of Edinburgh | COVID-19 Genomics UK (COG-UK) Consortium |
| EPI_ISL_433196 | Virology Department, Royal Infirmary of Edinburgh, NHS Lothian / School of Biological Sciences, University of Edinburgh / Institute of Genetics and Molecular Medicine, University of Edinburgh | COVID-19 Genomics UK (COG-UK) Consortium |

| EPI_Accession_ID | Originating Laboratory | Submitting Laboratory |
|---|---|---|
| EPI_ISL_433197 | Virology Department, Royal Infirmary of Edinburgh, NHS Lothian / School of Biological Sciences, University of Edinburgh / Institute of Genetics and Molecular Medicine, University of Edinburgh | COVID-19 Genomics UK (COG-UK) Consortium |
| EPI_ISL_433198 | Virology Department, Royal Infirmary of Edinburgh, NHS Lothian / School of Biological Sciences, University of Edinburgh / Institute of Genetics and Molecular Medicine, University of Edinburgh | COVID-19 Genomics UK (COG-UK) Consortium |
| EPI_ISL_433199 | Virology Department, Royal Infirmary of Edinburgh, NHS Lothian / School of Biological Sciences, University of Edinburgh / Institute of Genetics and Molecular Medicine, University of Edinburgh | COVID-19 Genomics UK (COG-UK) Consortium |
| EPI_ISL_433200 | Virology Department, Royal Infirmary of Edinburgh, NHS Lothian / School of Biological Sciences, University of Edinburgh / Institute of Genetics and Molecular Medicine, University of Edinburgh | COVID-19 Genomics UK (COG-UK) Consortium |
| EPI_ISL_433201 | Virology Department, Royal Infirmary of Edinburgh, NHS Lothian / School of Biological Sciences, University of Edinburgh / Institute of Genetics and Molecular Medicine, University of Edinburgh | COVID-19 Genomics UK (COG-UK) Consortium |
| EPI_ISL_433202 | Virology Department, Royal Infirmary of Edinburgh, NHS Lothian / School of Biological Sciences, University of Edinburgh / Institute of Genetics and Molecular Medicine, University of Edinburgh | COVID-19 Genomics UK (COG-UK) Consortium |
| EPI_ISL_433203 | Virology Department, Royal Infirmary of Edinburgh, NHS Lothian / School of Biological Sciences, University of Edinburgh / Institute of Genetics and Molecular Medicine, University of Edinburgh | COVID-19 Genomics UK (COG-UK) Consortium |
| EPI_ISL_433204 | Virology Department, Royal Infirmary of Edinburgh, NHS Lothian / School of Biological Sciences, University of Edinburgh / Institute of Genetics and Molecular Medicine, University of Edinburgh | COVID-19 Genomics UK (COG-UK) Consortium |

| EPI_Accession_ID | Originating Laboratory | Submitting Laboratory |
|---|---|---|
| EPI_ISL_433205 | Virology Department, Royal Infirmary of Edinburgh, NHS Lothian / School of Biological Sciences, University of Edinburgh / Institute of Genetics and Molecular Medicine, University of Edinburgh | COVID-19 Genomics UK (COG-UK) Consortium |
| EPI_ISL_433206 | Virology Department, Royal Infirmary of Edinburgh, NHS Lothian / School of Biological Sciences, University of Edinburgh / Institute of Genetics and Molecular Medicine, University of Edinburgh | COVID-19 Genomics UK (COG-UK) Consortium |
| EPI_ISL_433207 | Virology Department, Royal Infirmary of Edinburgh, NHS Lothian / School of Biological Sciences, University of Edinburgh / Institute of Genetics and Molecular Medicine, University of Edinburgh | COVID-19 Genomics UK (COG-UK) Consortium |
| EPI_ISL_433208 | Virology Department, Royal Infirmary of Edinburgh, NHS Lothian / School of Biological Sciences, University of Edinburgh / Institute of Genetics and Molecular Medicine, University of Edinburgh | COVID-19 Genomics UK (COG-UK) Consortium |
| EPI_ISL_433209 | Virology Department, Royal Infirmary of Edinburgh, NHS Lothian / School of Biological Sciences, University of Edinburgh / Institute of Genetics and Molecular Medicine, University of Edinburgh | COVID-19 Genomics UK (COG-UK) Consortium |
| EPI_ISL_433210 | Virology Department, Royal Infirmary of Edinburgh, NHS Lothian / School of Biological Sciences, University of Edinburgh / Institute of Genetics and Molecular Medicine, University of Edinburgh | COVID-19 Genomics UK (COG-UK) Consortium |
| EPI_ISL_433211 | Virology Department, Royal Infirmary of Edinburgh, NHS Lothian / School of Biological Sciences, University of Edinburgh / Institute of Genetics and Molecular Medicine, University of Edinburgh | COVID-19 Genomics UK (COG-UK) Consortium |
| EPI_ISL_433212 | Virology Department, Royal Infirmary of Edinburgh, NHS Lothian / School of Biological Sciences, University of Edinburgh / Institute of Genetics and Molecular Medicine, University of Edinburgh | COVID-19 Genomics UK (COG-UK) Consortium |

| EPI_Accession_ID | Originating Laboratory | Submitting Laboratory |
|---|---|---|
| EPI_ISL_433213 | Virology Department, Royal Infirmary of Edinburgh, NHS Lothian / School of Biological Sciences, University of Edinburgh / Institute of Genetics and Molecular Medicine, University of Edinburgh | COVID-19 Genomics UK (COG-UK) Consortium |
| EPI_ISL_433214 | Virology Department, Royal Infirmary of Edinburgh, NHS Lothian / School of Biological Sciences, University of Edinburgh / Institute of Genetics and Molecular Medicine, University of Edinburgh | COVID-19 Genomics UK (COG-UK) Consortium |
| EPI_ISL_433215 | Virology Department, Royal Infirmary of Edinburgh, NHS Lothian / School of Biological Sciences, University of Edinburgh / Institute of Genetics and Molecular Medicine, University of Edinburgh | COVID-19 Genomics UK (COG-UK) Consortium |
| EPI_ISL_433216 | Virology Department, Royal Infirmary of Edinburgh, NHS Lothian / School of Biological Sciences, University of Edinburgh / Institute of Genetics and Molecular Medicine, University of Edinburgh | COVID-19 Genomics UK (COG-UK) Consortium |
| EPI_ISL_433217 | Virology Department, Royal Infirmary of Edinburgh, NHS Lothian / School of Biological Sciences, University of Edinburgh / Institute of Genetics and Molecular Medicine, University of Edinburgh | COVID-19 Genomics UK (COG-UK) Consortium |
| EPI_ISL_433218 | Virology Department, Royal Infirmary of Edinburgh, NHS Lothian / School of Biological Sciences, University of Edinburgh / Institute of Genetics and Molecular Medicine, University of Edinburgh | COVID-19 Genomics UK (COG-UK) Consortium |
| EPI_ISL_433219 | Virology Department, Royal Infirmary of Edinburgh, NHS Lothian / School of Biological Sciences, University of Edinburgh / Institute of Genetics and Molecular Medicine, University of Edinburgh | COVID-19 Genomics UK (COG-UK) Consortium |
| EPI_ISL_433220 | Virology Department, Royal Infirmary of Edinburgh, NHS Lothian / School of Biological Sciences, University of Edinburgh / Institute of Genetics and Molecular Medicine, University of Edinburgh | COVID-19 Genomics UK (COG-UK) Consortium |

| EPI_Accession_ID | Originating Laboratory | Submitting Laboratory |
|---|---|---|
| EPI_ISL_433221 | Virology Department, Royal Infirmary of Edinburgh, NHS Lothian / School of Biological Sciences, University of Edinburgh / Institute of Genetics and Molecular Medicine, University of Edinburgh | COVID-19 Genomics UK (COG-UK) Consortium |
| EPI_ISL_433222 | Virology Department, Royal Infirmary of Edinburgh, NHS Lothian / School of Biological Sciences, University of Edinburgh / Institute of Genetics and Molecular Medicine, University of Edinburgh | COVID-19 Genomics UK (COG-UK) Consortium |
| EPI_ISL_433223 | Virology Department, Royal Infirmary of Edinburgh, NHS Lothian / School of Biological Sciences, University of Edinburgh / Institute of Genetics and Molecular Medicine, University of Edinburgh | COVID-19 Genomics UK (COG-UK) Consortium |
| EPI_ISL_433224 | Virology Department, Royal Infirmary of Edinburgh, NHS Lothian / School of Biological Sciences, University of Edinburgh / Institute of Genetics and Molecular Medicine, University of Edinburgh | COVID-19 Genomics UK (COG-UK) Consortium |
| EPI_ISL_433225 | Virology Department, Royal Infirmary of Edinburgh, NHS Lothian / School of Biological Sciences, University of Edinburgh / Institute of Genetics and Molecular Medicine, University of Edinburgh | COVID-19 Genomics UK (COG-UK) Consortium |
| EPI_ISL_433226 | Virology Department, Royal Infirmary of Edinburgh, NHS Lothian / School of Biological Sciences, University of Edinburgh / Institute of Genetics and Molecular Medicine, University of Edinburgh | COVID-19 Genomics UK (COG-UK) Consortium |
| EPI_ISL_433227 | Virology Department, Royal Infirmary of Edinburgh, NHS Lothian / School of Biological Sciences, University of Edinburgh / Institute of Genetics and Molecular Medicine, University of Edinburgh | COVID-19 Genomics UK (COG-UK) Consortium |
| EPI_ISL_433228 | Virology Department, Royal Infirmary of Edinburgh, NHS Lothian / School of Biological Sciences, University of Edinburgh / Institute of Genetics and Molecular Medicine, University of Edinburgh | COVID-19 Genomics UK (COG-UK) Consortium |

| EPI_Accession_ID | Originating Laboratory | Submitting Laboratory |
|---|---|---|
| EPI_ISL_433229 | Virology Department, Royal Infirmary of Edinburgh, NHS Lothian / School of Biological Sciences, University of Edinburgh / Institute of Genetics and Molecular Medicine, University of Edinburgh | COVID-19 Genomics UK (COG-UK) Consortium |
| EPI_ISL_433230 | Virology Department, Royal Infirmary of Edinburgh, NHS Lothian / School of Biological Sciences, University of Edinburgh / Institute of Genetics and Molecular Medicine, University of Edinburgh | COVID-19 Genomics UK (COG-UK) Consortium |
| EPI_ISL_433231 | Virology Department, Royal Infirmary of Edinburgh, NHS Lothian / School of Biological Sciences, University of Edinburgh / Institute of Genetics and Molecular Medicine, University of Edinburgh | COVID-19 Genomics UK (COG-UK) Consortium |
| EPI_ISL_433232 | Virology Department, Royal Infirmary of Edinburgh, NHS Lothian / School of Biological Sciences, University of Edinburgh / Institute of Genetics and Molecular Medicine, University of Edinburgh | COVID-19 Genomics UK (COG-UK) Consortium |
| EPI_ISL_433233 | Virology Department, Royal Infirmary of Edinburgh, NHS Lothian / School of Biological Sciences, University of Edinburgh / Institute of Genetics and Molecular Medicine, University of Edinburgh | COVID-19 Genomics UK (COG-UK) Consortium |
| EPI_ISL_433234 | Virology Department, Royal Infirmary of Edinburgh, NHS Lothian / School of Biological Sciences, University of Edinburgh / Institute of Genetics and Molecular Medicine, University of Edinburgh | COVID-19 Genomics UK (COG-UK) Consortium |
| EPI_ISL_433235 | Virology Department, Royal Infirmary of Edinburgh, NHS Lothian / School of Biological Sciences, University of Edinburgh / Institute of Genetics and Molecular Medicine, University of Edinburgh | COVID-19 Genomics UK (COG-UK) Consortium |
| EPI_ISL_433236 | Virology Department, Royal Infirmary of Edinburgh, NHS Lothian / School of Biological Sciences, University of Edinburgh / Institute of Genetics and Molecular Medicine, University of Edinburgh | COVID-19 Genomics UK (COG-UK) Consortium |

| EPI_Accession_ID | Originating Laboratory | Submitting Laboratory |
|---|---|---|
| EPI_ISL_433237 | Virology Department, Royal Infirmary of Edinburgh, NHS Lothian / School of Biological Sciences, University of Edinburgh / Institute of Genetics and Molecular Medicine, University of Edinburgh | COVID-19 Genomics UK (COG-UK) Consortium |
| EPI_ISL_433238 | Virology Department, Royal Infirmary of Edinburgh, NHS Lothian / School of Biological Sciences, University of Edinburgh / Institute of Genetics and Molecular Medicine, University of Edinburgh | COVID-19 Genomics UK (COG-UK) Consortium |
| EPI_ISL_433239 | Virology Department, Royal Infirmary of Edinburgh, NHS Lothian / School of Biological Sciences, University of Edinburgh / Institute of Genetics and Molecular Medicine, University of Edinburgh | COVID-19 Genomics UK (COG-UK) Consortium |
| EPI_ISL_433240 | Virology Department, Royal Infirmary of Edinburgh, NHS Lothian / School of Biological Sciences, University of Edinburgh / Institute of Genetics and Molecular Medicine, University of Edinburgh | COVID-19 Genomics UK (COG-UK) Consortium |
| EPI_ISL_433241 | Virology Department, Royal Infirmary of Edinburgh, NHS Lothian / School of Biological Sciences, University of Edinburgh / Institute of Genetics and Molecular Medicine, University of Edinburgh | COVID-19 Genomics UK (COG-UK) Consortium |
| EPI_ISL_433242 | Virology Department, Royal Infirmary of Edinburgh, NHS Lothian / School of Biological Sciences, University of Edinburgh / Institute of Genetics and Molecular Medicine, University of Edinburgh | COVID-19 Genomics UK (COG-UK) Consortium |
| EPI_ISL_433243 | Virology Department, Royal Infirmary of Edinburgh, NHS Lothian / School of Biological Sciences, University of Edinburgh / Institute of Genetics and Molecular Medicine, University of Edinburgh | COVID-19 Genomics UK (COG-UK) Consortium |
| EPI_ISL_433244 | Virology Department, Royal Infirmary of Edinburgh, NHS Lothian / School of Biological Sciences, University of Edinburgh / Institute of Genetics and Molecular Medicine, University of Edinburgh | COVID-19 Genomics UK (COG-UK) Consortium |

| EPI_Accession_ID | Originating Laboratory | Submitting Laboratory |
|---|---|---|
| EPI_ISL_433245 | Virology Department, Royal Infirmary of Edinburgh, NHS Lothian / School of Biological Sciences, University of Edinburgh / Institute of Genetics and Molecular Medicine, University of Edinburgh | COVID-19 Genomics UK (COG-UK) Consortium |
| EPI_ISL_433246 | Virology Department, Royal Infirmary of Edinburgh, NHS Lothian / School of Biological Sciences, University of Edinburgh / Institute of Genetics and Molecular Medicine, University of Edinburgh | COVID-19 Genomics UK (COG-UK) Consortium |
| EPI_ISL_433247 | Virology Department, Royal Infirmary of Edinburgh, NHS Lothian / School of Biological Sciences, University of Edinburgh / Institute of Genetics and Molecular Medicine, University of Edinburgh | COVID-19 Genomics UK (COG-UK) Consortium |
| EPI_ISL_433248 | Virology Department, Royal Infirmary of Edinburgh, NHS Lothian / School of Biological Sciences, University of Edinburgh / Institute of Genetics and Molecular Medicine, University of Edinburgh | COVID-19 Genomics UK (COG-UK) Consortium |
| EPI_ISL_433249 | Virology Department, Royal Infirmary of Edinburgh, NHS Lothian / School of Biological Sciences, University of Edinburgh / Institute of Genetics and Molecular Medicine, University of Edinburgh | COVID-19 Genomics UK (COG-UK) Consortium |
| EPI_ISL_433250 | Virology Department, Royal Infirmary of Edinburgh, NHS Lothian / School of Biological Sciences, University of Edinburgh / Institute of Genetics and Molecular Medicine, University of Edinburgh | COVID-19 Genomics UK (COG-UK) Consortium |
| EPI_ISL_433251 | Virology Department, Royal Infirmary of Edinburgh, NHS Lothian / School of Biological Sciences, University of Edinburgh / Institute of Genetics and Molecular Medicine, University of Edinburgh | COVID-19 Genomics UK (COG-UK) Consortium |
| EPI_ISL_433252 | Virology Department, Royal Infirmary of Edinburgh, NHS Lothian / School of Biological Sciences, University of Edinburgh / Institute of Genetics and Molecular Medicine, University of Edinburgh | COVID-19 Genomics UK (COG-UK) Consortium |

| EPI_Accession_ID | Originating Laboratory | Submitting Laboratory |
|---|---|---|
| EPI_ISL_433253 | Virology Department, Royal Infirmary of Edinburgh, NHS Lothian / School of Biological Sciences, University of Edinburgh / Institute of Genetics and Molecular Medicine, University of Edinburgh | COVID-19 Genomics UK (COG-UK) Consortium |
| EPI_ISL_433254 | Virology Department, Royal Infirmary of Edinburgh, NHS Lothian / School of Biological Sciences, University of Edinburgh / Institute of Genetics and Molecular Medicine, University of Edinburgh | COVID-19 Genomics UK (COG-UK) Consortium |
| EPI_ISL_433255 | Virology Department, Royal Infirmary of Edinburgh, NHS Lothian / School of Biological Sciences, University of Edinburgh / Institute of Genetics and Molecular Medicine, University of Edinburgh | COVID-19 Genomics UK (COG-UK) Consortium |
| EPI_ISL_433256 | Virology Department, Royal Infirmary of Edinburgh, NHS Lothian / School of Biological Sciences, University of Edinburgh / Institute of Genetics and Molecular Medicine, University of Edinburgh | COVID-19 Genomics UK (COG-UK) Consortium |
| EPI_ISL_433257 | Virology Department, Royal Infirmary of Edinburgh, NHS Lothian / School of Biological Sciences, University of Edinburgh / Institute of Genetics and Molecular Medicine, University of Edinburgh | COVID-19 Genomics UK (COG-UK) Consortium |
| EPI_ISL_433258 | Virology Department, Royal Infirmary of Edinburgh, NHS Lothian / School of Biological Sciences, University of Edinburgh / Institute of Genetics and Molecular Medicine, University of Edinburgh | COVID-19 Genomics UK (COG-UK) Consortium |
| EPI_ISL_433259 | Virology Department, Royal Infirmary of Edinburgh, NHS Lothian / School of Biological Sciences, University of Edinburgh / Institute of Genetics and Molecular Medicine, University of Edinburgh | COVID-19 Genomics UK (COG-UK) Consortium |
| EPI_ISL_433260 | Virology Department, Royal Infirmary of Edinburgh, NHS Lothian / School of Biological Sciences, University of Edinburgh / Institute of Genetics and Molecular Medicine, University of Edinburgh | COVID-19 Genomics UK (COG-UK) Consortium |

| EPI_Accession_ID | Originating Laboratory | Submitting Laboratory |
| --- | --- | --- |
| EPI_ISL_433261 | Virology Department, Royal Infirmary of Edinburgh, NHS Lothian / School of Biological Sciences, University of Edinburgh / Institute of Genetics and Molecular Medicine, University of Edinburgh | COVID-19 Genomics UK (COG-UK) Consortium |
| EPI_ISL_433262 | Virology Department, Royal Infirmary of Edinburgh, NHS Lothian / School of Biological Sciences, University of Edinburgh / Institute of Genetics and Molecular Medicine, University of Edinburgh | COVID-19 Genomics UK (COG-UK) Consortium |
| EPI_ISL_433263 | Virology Department, Royal Infirmary of Edinburgh, NHS Lothian / School of Biological Sciences, University of Edinburgh / Institute of Genetics and Molecular Medicine, University of Edinburgh | COVID-19 Genomics UK (COG-UK) Consortium |
| EPI_ISL_433264 | Virology Department, Royal Infirmary of Edinburgh, NHS Lothian / School of Biological Sciences, University of Edinburgh / Institute of Genetics and Molecular Medicine, University of Edinburgh | COVID-19 Genomics UK (COG-UK) Consortium |
| EPI_ISL_433265 | Virology Department, Royal Infirmary of Edinburgh, NHS Lothian / School of Biological Sciences, University of Edinburgh / Institute of Genetics and Molecular Medicine, University of Edinburgh | COVID-19 Genomics UK (COG-UK) Consortium |
| EPI_ISL_433266 | Virology Department, Royal Infirmary of Edinburgh, NHS Lothian / School of Biological Sciences, University of Edinburgh / Institute of Genetics and Molecular Medicine, University of Edinburgh | COVID-19 Genomics UK (COG-UK) Consortium |
| EPI_ISL_433267 | Virology Department, Royal Infirmary of Edinburgh, NHS Lothian / School of Biological Sciences, University of Edinburgh / Institute of Genetics and Molecular Medicine, University of Edinburgh | COVID-19 Genomics UK (COG-UK) Consortium |
| EPI_ISL_433268 | West of Scotland Specialist Virology Centre, NHSGGC / MRC-University of Glasgow Centre for Virus Research | COVID-19 Genomics UK (COG-UK) Consortium |
| EPI_ISL_433269 | West of Scotland Specialist Virology Centre, NHSGGC / MRC-University of Glasgow Centre for Virus Research | COVID-19 Genomics UK (COG-UK) Consortium |

| EPI_Accession_ID | Originating Laboratory | Submitting Laboratory |
|---|---|---|
| EPI_ISL_433270 | West of Scotland Specialist Virology Centre, NHSGGC / MRC-University of Glasgow Centre for Virus Research | COVID-19 Genomics UK (COG-UK) Consortium |
| EPI_ISL_433271 | West of Scotland Specialist Virology Centre, NHSGGC / MRC-University of Glasgow Centre for Virus Research | COVID-19 Genomics UK (COG-UK) Consortium |
| EPI_ISL_433272 | West of Scotland Specialist Virology Centre, NHSGGC / MRC-University of Glasgow Centre for Virus Research | COVID-19 Genomics UK (COG-UK) Consortium |
| EPI_ISL_433273 | West of Scotland Specialist Virology Centre, NHSGGC / MRC-University of Glasgow Centre for Virus Research | COVID-19 Genomics UK (COG-UK) Consortium |
| EPI_ISL_433274 | West of Scotland Specialist Virology Centre, NHSGGC / MRC-University of Glasgow Centre for Virus Research | COVID-19 Genomics UK (COG-UK) Consortium |
| EPI_ISL_433275 | West of Scotland Specialist Virology Centre, NHSGGC / MRC-University of Glasgow Centre for Virus Research | COVID-19 Genomics UK (COG-UK) Consortium |
| EPI_ISL_433276 | West of Scotland Specialist Virology Centre, NHSGGC / MRC-University of Glasgow Centre for Virus Research | COVID-19 Genomics UK (COG-UK) Consortium |
| EPI_ISL_433277 | West of Scotland Specialist Virology Centre, NHSGGC / MRC-University of Glasgow Centre for Virus Research | COVID-19 Genomics UK (COG-UK) Consortium |
| EPI_ISL_433278 | West of Scotland Specialist Virology Centre, NHSGGC / MRC-University of Glasgow Centre for Virus Research | COVID-19 Genomics UK (COG-UK) Consortium |
| EPI_ISL_433279 | West of Scotland Specialist Virology Centre, NHSGGC / MRC-University of Glasgow Centre for Virus Research | COVID-19 Genomics UK (COG-UK) Consortium |
| EPI_ISL_433280 | West of Scotland Specialist Virology Centre, NHSGGC / MRC-University of Glasgow Centre for Virus Research | COVID-19 Genomics UK (COG-UK) Consortium |
| EPI_ISL_433281 | West of Scotland Specialist Virology Centre, NHSGGC / MRC-University of Glasgow Centre for Virus Research | COVID-19 Genomics UK (COG-UK) Consortium |
| EPI_ISL_433282 | West of Scotland Specialist Virology Centre, NHSGGC / MRC-University of Glasgow Centre for Virus Research | COVID-19 Genomics UK (COG-UK) Consortium |
| EPI_ISL_433283 | West of Scotland Specialist Virology Centre, NHSGGC / MRC-University of Glasgow Centre for Virus Research | COVID-19 Genomics UK (COG-UK) Consortium |
| EPI_ISL_433284 | West of Scotland Specialist Virology Centre, NHSGGC / MRC-University of Glasgow Centre for Virus Research | COVID-19 Genomics UK (COG-UK) Consortium |
| EPI_ISL_433285 | West of Scotland Specialist Virology Centre, NHSGGC / MRC-University of Glasgow Centre for Virus Research | COVID-19 Genomics UK (COG-UK) Consortium |

| EPI_Accession_ID | Originating Laboratory | Submitting Laboratory |
|---|---|---|
| EPI_ISL_433286 | West of Scotland Specialist Virology Centre, NHSGGC / MRC-University of Glasgow Centre for Virus Research | COVID-19 Genomics UK (COG-UK) Consortium |
| EPI_ISL_433287 | West of Scotland Specialist Virology Centre, NHSGGC / MRC-University of Glasgow Centre for Virus Research | COVID-19 Genomics UK (COG-UK) Consortium |
| EPI_ISL_433288 | West of Scotland Specialist Virology Centre, NHSGGC / MRC-University of Glasgow Centre for Virus Research | COVID-19 Genomics UK (COG-UK) Consortium |
| EPI_ISL_433289 | West of Scotland Specialist Virology Centre, NHSGGC / MRC-University of Glasgow Centre for Virus Research | COVID-19 Genomics UK (COG-UK) Consortium |
| EPI_ISL_433290 | West of Scotland Specialist Virology Centre, NHSGGC / MRC-University of Glasgow Centre for Virus Research | COVID-19 Genomics UK (COG-UK) Consortium |
| EPI_ISL_433291 | West of Scotland Specialist Virology Centre, NHSGGC / MRC-University of Glasgow Centre for Virus Research | COVID-19 Genomics UK (COG-UK) Consortium |
| EPI_ISL_433292 | West of Scotland Specialist Virology Centre, NHSGGC / MRC-University of Glasgow Centre for Virus Research | COVID-19 Genomics UK (COG-UK) Consortium |
| EPI_ISL_433293 | West of Scotland Specialist Virology Centre, NHSGGC / MRC-University of Glasgow Centre for Virus Research | COVID-19 Genomics UK (COG-UK) Consortium |
| EPI_ISL_433294 | West of Scotland Specialist Virology Centre, NHSGGC / MRC-University of Glasgow Centre for Virus Research | COVID-19 Genomics UK (COG-UK) Consortium |
| EPI_ISL_433295 | West of Scotland Specialist Virology Centre, NHSGGC / MRC-University of Glasgow Centre for Virus Research | COVID-19 Genomics UK (COG-UK) Consortium |
| EPI_ISL_433296 | West of Scotland Specialist Virology Centre, NHSGGC / MRC-University of Glasgow Centre for Virus Research | COVID-19 Genomics UK (COG-UK) Consortium |
| EPI_ISL_433297 | West of Scotland Specialist Virology Centre, NHSGGC / MRC-University of Glasgow Centre for Virus Research | COVID-19 Genomics UK (COG-UK) Consortium |
| EPI_ISL_433298 | West of Scotland Specialist Virology Centre, NHSGGC / MRC-University of Glasgow Centre for Virus Research | COVID-19 Genomics UK (COG-UK) Consortium |
| EPI_ISL_433299 | West of Scotland Specialist Virology Centre, NHSGGC / MRC-University of Glasgow Centre for Virus Research | COVID-19 Genomics UK (COG-UK) Consortium |
| EPI_ISL_433300 | West of Scotland Specialist Virology Centre, NHSGGC / MRC-University of Glasgow Centre for Virus Research | COVID-19 Genomics UK (COG-UK) Consortium |
| EPI_ISL_433301 | West of Scotland Specialist Virology Centre, NHSGGC / MRC-University of Glasgow Centre for Virus Research | COVID-19 Genomics UK (COG-UK) Consortium |

| EPI_Accession_ID | Originating Laboratory | Submitting Laboratory |
|---|---|---|
| EPI_ISL_433302 | West of Scotland Specialist Virology Centre, NHSGGC / MRC-University of Glasgow Centre for Virus Research | COVID-19 Genomics UK (COG-UK) Consortium |
| EPI_ISL_433303 | West of Scotland Specialist Virology Centre, NHSGGC / MRC-University of Glasgow Centre for Virus Research | COVID-19 Genomics UK (COG-UK) Consortium |
| EPI_ISL_433304 | West of Scotland Specialist Virology Centre, NHSGGC / MRC-University of Glasgow Centre for Virus Research | COVID-19 Genomics UK (COG-UK) Consortium |
| EPI_ISL_433305 | West of Scotland Specialist Virology Centre, NHSGGC / MRC-University of Glasgow Centre for Virus Research | COVID-19 Genomics UK (COG-UK) Consortium |
| EPI_ISL_433306 | West of Scotland Specialist Virology Centre, NHSGGC / MRC-University of Glasgow Centre for Virus Research | COVID-19 Genomics UK (COG-UK) Consortium |
| EPI_ISL_433307 | West of Scotland Specialist Virology Centre, NHSGGC / MRC-University of Glasgow Centre for Virus Research | COVID-19 Genomics UK (COG-UK) Consortium |
| EPI_ISL_433308 | West of Scotland Specialist Virology Centre, NHSGGC / MRC-University of Glasgow Centre for Virus Research | COVID-19 Genomics UK (COG-UK) Consortium |
| EPI_ISL_433309 | West of Scotland Specialist Virology Centre, NHSGGC / MRC-University of Glasgow Centre for Virus Research | COVID-19 Genomics UK (COG-UK) Consortium |
| EPI_ISL_433310 | West of Scotland Specialist Virology Centre, NHSGGC / MRC-University of Glasgow Centre for Virus Research | COVID-19 Genomics UK (COG-UK) Consortium |
| EPI_ISL_433311 | West of Scotland Specialist Virology Centre, NHSGGC / MRC-University of Glasgow Centre for Virus Research | COVID-19 Genomics UK (COG-UK) Consortium |
| EPI_ISL_433312 | West of Scotland Specialist Virology Centre, NHSGGC / MRC-University of Glasgow Centre for Virus Research | COVID-19 Genomics UK (COG-UK) Consortium |
| EPI_ISL_433313 | West of Scotland Specialist Virology Centre, NHSGGC / MRC-University of Glasgow Centre for Virus Research | COVID-19 Genomics UK (COG-UK) Consortium |
| EPI_ISL_433314 | West of Scotland Specialist Virology Centre, NHSGGC / MRC-University of Glasgow Centre for Virus Research | COVID-19 Genomics UK (COG-UK) Consortium |
| EPI_ISL_433315 | West of Scotland Specialist Virology Centre, NHSGGC / MRC-University of Glasgow Centre for Virus Research | COVID-19 Genomics UK (COG-UK) Consortium |
| EPI_ISL_433316 | West of Scotland Specialist Virology Centre, NHSGGC / MRC-University of Glasgow Centre for Virus Research | COVID-19 Genomics UK (COG-UK) Consortium |
| EPI_ISL_433317 | West of Scotland Specialist Virology Centre, NHSGGC / MRC-University of Glasgow Centre for Virus Research | COVID-19 Genomics UK (COG-UK) Consortium |

| EPI_Accession_ID | Originating Laboratory | Submitting Laboratory |
|---|---|---|
| EPI_ISL_433318 | West of Scotland Specialist Virology Centre, NHSGGC / MRC-University of Glasgow Centre for Virus Research | COVID-19 Genomics UK (COG-UK) Consortium |
| EPI_ISL_433319 | West of Scotland Specialist Virology Centre, NHSGGC / MRC-University of Glasgow Centre for Virus Research | COVID-19 Genomics UK (COG-UK) Consortium |
| EPI_ISL_433320 | West of Scotland Specialist Virology Centre, NHSGGC / MRC-University of Glasgow Centre for Virus Research | COVID-19 Genomics UK (COG-UK) Consortium |
| EPI_ISL_433321 | West of Scotland Specialist Virology Centre, NHSGGC / MRC-University of Glasgow Centre for Virus Research | COVID-19 Genomics UK (COG-UK) Consortium |
| EPI_ISL_433322 | West of Scotland Specialist Virology Centre, NHSGGC / MRC-University of Glasgow Centre for Virus Research | COVID-19 Genomics UK (COG-UK) Consortium |
| EPI_ISL_433323 | West of Scotland Specialist Virology Centre, NHSGGC / MRC-University of Glasgow Centre for Virus Research | COVID-19 Genomics UK (COG-UK) Consortium |
| EPI_ISL_433324 | West of Scotland Specialist Virology Centre, NHSGGC / MRC-University of Glasgow Centre for Virus Research | COVID-19 Genomics UK (COG-UK) Consortium |
| EPI_ISL_433325 | West of Scotland Specialist Virology Centre, NHSGGC / MRC-University of Glasgow Centre for Virus Research | COVID-19 Genomics UK (COG-UK) Consortium |
| EPI_ISL_433326 | West of Scotland Specialist Virology Centre, NHSGGC / MRC-University of Glasgow Centre for Virus Research | COVID-19 Genomics UK (COG-UK) Consortium |
| EPI_ISL_433327 | West of Scotland Specialist Virology Centre, NHSGGC / MRC-University of Glasgow Centre for Virus Research | COVID-19 Genomics UK (COG-UK) Consortium |
| EPI_ISL_433328 | West of Scotland Specialist Virology Centre, NHSGGC / MRC-University of Glasgow Centre for Virus Research | COVID-19 Genomics UK (COG-UK) Consortium |
| EPI_ISL_433329 | West of Scotland Specialist Virology Centre, NHSGGC / MRC-University of Glasgow Centre for Virus Research | COVID-19 Genomics UK (COG-UK) Consortium |
| EPI_ISL_433330 | West of Scotland Specialist Virology Centre, NHSGGC / MRC-University of Glasgow Centre for Virus Research | COVID-19 Genomics UK (COG-UK) Consortium |
| EPI_ISL_433331 | West of Scotland Specialist Virology Centre, NHSGGC / MRC-University of Glasgow Centre for Virus Research | COVID-19 Genomics UK (COG-UK) Consortium |
| EPI_ISL_433332 | West of Scotland Specialist Virology Centre, NHSGGC / MRC-University of Glasgow Centre for Virus Research | COVID-19 Genomics UK (COG-UK) Consortium |
| EPI_ISL_433333 | West of Scotland Specialist Virology Centre, NHSGGC / MRC-University of Glasgow Centre for Virus Research | COVID-19 Genomics UK (COG-UK) Consortium |

| EPI_Accession_ID | Originating Laboratory | Submitting Laboratory |
|---|---|---|
| EPI_ISL_433334 | West of Scotland Specialist Virology Centre, NHSGGC / MRC-University of Glasgow Centre for Virus Research | COVID-19 Genomics UK (COG-UK) Consortium |
| EPI_ISL_433335 | West of Scotland Specialist Virology Centre, NHSGGC / MRC-University of Glasgow Centre for Virus Research | COVID-19 Genomics UK (COG-UK) Consortium |
| EPI_ISL_433336 | West of Scotland Specialist Virology Centre, NHSGGC / MRC-University of Glasgow Centre for Virus Research | COVID-19 Genomics UK (COG-UK) Consortium |
| EPI_ISL_433337 | West of Scotland Specialist Virology Centre, NHSGGC / MRC-University of Glasgow Centre for Virus Research | COVID-19 Genomics UK (COG-UK) Consortium |
| EPI_ISL_433338 | West of Scotland Specialist Virology Centre, NHSGGC / MRC-University of Glasgow Centre for Virus Research | COVID-19 Genomics UK (COG-UK) Consortium |
| EPI_ISL_433339 | West of Scotland Specialist Virology Centre, NHSGGC / MRC-University of Glasgow Centre for Virus Research | COVID-19 Genomics UK (COG-UK) Consortium |
| EPI_ISL_433340 | West of Scotland Specialist Virology Centre, NHSGGC / MRC-University of Glasgow Centre for Virus Research | COVID-19 Genomics UK (COG-UK) Consortium |
| EPI_ISL_433341 | West of Scotland Specialist Virology Centre, NHSGGC / MRC-University of Glasgow Centre for Virus Research | COVID-19 Genomics UK (COG-UK) Consortium |
| EPI_ISL_433342 | West of Scotland Specialist Virology Centre, NHSGGC / MRC-University of Glasgow Centre for Virus Research | COVID-19 Genomics UK (COG-UK) Consortium |
| EPI_ISL_433343 | West of Scotland Specialist Virology Centre, NHSGGC / MRC-University of Glasgow Centre for Virus Research | COVID-19 Genomics UK (COG-UK) Consortium |
| EPI_ISL_433344 | West of Scotland Specialist Virology Centre, NHSGGC / MRC-University of Glasgow Centre for Virus Research | COVID-19 Genomics UK (COG-UK) Consortium |
| EPI_ISL_433345 | West of Scotland Specialist Virology Centre, NHSGGC / MRC-University of Glasgow Centre for Virus Research | COVID-19 Genomics UK (COG-UK) Consortium |
| EPI_ISL_433346 | West of Scotland Specialist Virology Centre, NHSGGC / MRC-University of Glasgow Centre for Virus Research | COVID-19 Genomics UK (COG-UK) Consortium |
| EPI_ISL_433347 | West of Scotland Specialist Virology Centre, NHSGGC / MRC-University of Glasgow Centre for Virus Research | COVID-19 Genomics UK (COG-UK) Consortium |
| EPI_ISL_433348 | West of Scotland Specialist Virology Centre, NHSGGC / MRC-University of Glasgow Centre for Virus Research | COVID-19 Genomics UK (COG-UK) Consortium |
| EPI_ISL_433349 | West of Scotland Specialist Virology Centre, NHSGGC / MRC-University of Glasgow Centre for Virus Research | COVID-19 Genomics UK (COG-UK) Consortium |

| EPI_Accession_ID | Originating Laboratory | Submitting Laboratory |
|---|---|---|
| EPI_ISL_433350 | West of Scotland Specialist Virology Centre, NHSGGC / MRC-University of Glasgow Centre for Virus Research | COVID-19 Genomics UK (COG-UK) Consortium |
| EPI_ISL_433351 | West of Scotland Specialist Virology Centre, NHSGGC / MRC-University of Glasgow Centre for Virus Research | COVID-19 Genomics UK (COG-UK) Consortium |
| EPI_ISL_433352 | West of Scotland Specialist Virology Centre, NHSGGC / MRC-University of Glasgow Centre for Virus Research | COVID-19 Genomics UK (COG-UK) Consortium |
| EPI_ISL_433353 | West of Scotland Specialist Virology Centre, NHSGGC / MRC-University of Glasgow Centre for Virus Research | COVID-19 Genomics UK (COG-UK) Consortium |
| EPI_ISL_433354 | West of Scotland Specialist Virology Centre, NHSGGC / MRC-University of Glasgow Centre for Virus Research | COVID-19 Genomics UK (COG-UK) Consortium |
| EPI_ISL_433355 | West of Scotland Specialist Virology Centre, NHSGGC / MRC-University of Glasgow Centre for Virus Research | COVID-19 Genomics UK (COG-UK) Consortium |
| EPI_ISL_433356 | West of Scotland Specialist Virology Centre, NHSGGC / MRC-University of Glasgow Centre for Virus Research | COVID-19 Genomics UK (COG-UK) Consortium |
| EPI_ISL_433357 | West of Scotland Specialist Virology Centre, NHSGGC / MRC-University of Glasgow Centre for Virus Research | COVID-19 Genomics UK (COG-UK) Consortium |
| EPI_ISL_433358 | West of Scotland Specialist Virology Centre, NHSGGC / MRC-University of Glasgow Centre for Virus Research | COVID-19 Genomics UK (COG-UK) Consortium |
| EPI_ISL_433359 | West of Scotland Specialist Virology Centre, NHSGGC / MRC-University of Glasgow Centre for Virus Research | COVID-19 Genomics UK (COG-UK) Consortium |
| EPI_ISL_433360 | West of Scotland Specialist Virology Centre, NHSGGC / MRC-University of Glasgow Centre for Virus Research | COVID-19 Genomics UK (COG-UK) Consortium |
| EPI_ISL_433361 | West of Scotland Specialist Virology Centre, NHSGGC / MRC-University of Glasgow Centre for Virus Research | COVID-19 Genomics UK (COG-UK) Consortium |
| EPI_ISL_433362 | West of Scotland Specialist Virology Centre, NHSGGC / MRC-University of Glasgow Centre for Virus Research | COVID-19 Genomics UK (COG-UK) Consortium |
| EPI_ISL_433363 | West of Scotland Specialist Virology Centre, NHSGGC / MRC-University of Glasgow Centre for Virus Research | COVID-19 Genomics UK (COG-UK) Consortium |
| EPI_ISL_433364 | West of Scotland Specialist Virology Centre, NHSGGC / MRC-University of Glasgow Centre for Virus Research | COVID-19 Genomics UK (COG-UK) Consortium |
| EPI_ISL_433365 | West of Scotland Specialist Virology Centre, NHSGGC / MRC-University of Glasgow Centre for Virus Research | COVID-19 Genomics UK (COG-UK) Consortium |

| EPI_Accession_ID | Originating Laboratory | Submitting Laboratory |
|---|---|---|
| EPI_ISL_433366 | West of Scotland Specialist Virology Centre, NHSGGC / MRC-University of Glasgow Centre for Virus Research | COVID-19 Genomics UK (COG-UK) Consortium |
| EPI_ISL_433367 | West of Scotland Specialist Virology Centre, NHSGGC / MRC-University of Glasgow Centre for Virus Research | COVID-19 Genomics UK (COG-UK) Consortium |
| EPI_ISL_433368 | West of Scotland Specialist Virology Centre, NHSGGC / MRC-University of Glasgow Centre for Virus Research | COVID-19 Genomics UK (COG-UK) Consortium |
| EPI_ISL_433369 | West of Scotland Specialist Virology Centre, NHSGGC / MRC-University of Glasgow Centre for Virus Research | COVID-19 Genomics UK (COG-UK) Consortium |
| EPI_ISL_433370 | West of Scotland Specialist Virology Centre, NHSGGC / MRC-University of Glasgow Centre for Virus Research | COVID-19 Genomics UK (COG-UK) Consortium |
| EPI_ISL_433371 | West of Scotland Specialist Virology Centre, NHSGGC / MRC-University of Glasgow Centre for Virus Research | COVID-19 Genomics UK (COG-UK) Consortium |
| EPI_ISL_433372 | West of Scotland Specialist Virology Centre, NHSGGC / MRC-University of Glasgow Centre for Virus Research | COVID-19 Genomics UK (COG-UK) Consortium |
| EPI_ISL_433373 | West of Scotland Specialist Virology Centre, NHSGGC / MRC-University of Glasgow Centre for Virus Research | COVID-19 Genomics UK (COG-UK) Consortium |
| EPI_ISL_433374 | Virology Department, Royal Infirmary of Edinburgh, NHS Lothian / School of Biological Sciences, University of Edinburgh / Institute of Genetics and Molecular Medicine, University of Edinburgh | COVID-19 Genomics UK (COG-UK) Consortium |
| EPI_ISL_433375 | Virology Department, Royal Infirmary of Edinburgh, NHS Lothian / School of Biological Sciences, University of Edinburgh / Institute of Genetics and Molecular Medicine, University of Edinburgh | COVID-19 Genomics UK (COG-UK) Consortium |
| EPI_ISL_433376 | Virology Department, Royal Infirmary of Edinburgh, NHS Lothian / School of Biological Sciences, University of Edinburgh / Institute of Genetics and Molecular Medicine, University of Edinburgh | COVID-19 Genomics UK (COG-UK) Consortium |
| EPI_ISL_433377 | Virology Department, Royal Infirmary of Edinburgh, NHS Lothian / School of Biological Sciences, University of Edinburgh / Institute of Genetics and Molecular Medicine, University of Edinburgh | COVID-19 Genomics UK (COG-UK) Consortium |

| EPI_Accession_ID | Originating Laboratory | Submitting Laboratory |
|---|---|---|
| EPI_ISL_433378 | Virology Department, Royal Infirmary of Edinburgh, NHS Lothian / School of Biological Sciences, University of Edinburgh / Institute of Genetics and Molecular Medicine, University of Edinburgh | COVID-19 Genomics UK (COG-UK) Consortium |
| EPI_ISL_433379 | Virology Department, Royal Infirmary of Edinburgh, NHS Lothian / School of Biological Sciences, University of Edinburgh / Institute of Genetics and Molecular Medicine, University of Edinburgh | COVID-19 Genomics UK (COG-UK) Consortium |
| EPI_ISL_433380 | Virology Department, Royal Infirmary of Edinburgh, NHS Lothian / School of Biological Sciences, University of Edinburgh / Institute of Genetics and Molecular Medicine, University of Edinburgh | COVID-19 Genomics UK (COG-UK) Consortium |
| EPI_ISL_433381 | Virology Department, Royal Infirmary of Edinburgh, NHS Lothian / School of Biological Sciences, University of Edinburgh / Institute of Genetics and Molecular Medicine, University of Edinburgh | COVID-19 Genomics UK (COG-UK) Consortium |
| EPI_ISL_433382 | Virology Department, Royal Infirmary of Edinburgh, NHS Lothian / School of Biological Sciences, University of Edinburgh / Institute of Genetics and Molecular Medicine, University of Edinburgh | COVID-19 Genomics UK (COG-UK) Consortium |
| EPI_ISL_433383 | Virology Department, Royal Infirmary of Edinburgh, NHS Lothian / School of Biological Sciences, University of Edinburgh / Institute of Genetics and Molecular Medicine, University of Edinburgh | COVID-19 Genomics UK (COG-UK) Consortium |
| EPI_ISL_433384 | Virology Department, Royal Infirmary of Edinburgh, NHS Lothian / School of Biological Sciences, University of Edinburgh / Institute of Genetics and Molecular Medicine, University of Edinburgh | COVID-19 Genomics UK (COG-UK) Consortium |
| EPI_ISL_433385 | Virology Department, Royal Infirmary of Edinburgh, NHS Lothian / School of Biological Sciences, University of Edinburgh / Institute of Genetics and Molecular Medicine, University of Edinburgh | COVID-19 Genomics UK (COG-UK) Consortium |

| EPI_Accession_ID | Originating Laboratory | Submitting Laboratory |
|---|---|---|
| EPI_ISL_433386 | Virology Department, Royal Infirmary of Edinburgh, NHS Lothian / School of Biological Sciences, University of Edinburgh / Institute of Genetics and Molecular Medicine, University of Edinburgh | COVID-19 Genomics UK (COG-UK) Consortium |
| EPI_ISL_433387 | Virology Department, Royal Infirmary of Edinburgh, NHS Lothian / School of Biological Sciences, University of Edinburgh / Institute of Genetics and Molecular Medicine, University of Edinburgh | COVID-19 Genomics UK (COG-UK) Consortium |
| EPI_ISL_433388 | Virology Department, Royal Infirmary of Edinburgh, NHS Lothian / School of Biological Sciences, University of Edinburgh / Institute of Genetics and Molecular Medicine, University of Edinburgh | COVID-19 Genomics UK (COG-UK) Consortium |
| EPI_ISL_433389 | Virology Department, Royal Infirmary of Edinburgh, NHS Lothian / School of Biological Sciences, University of Edinburgh / Institute of Genetics and Molecular Medicine, University of Edinburgh | COVID-19 Genomics UK (COG-UK) Consortium |
| EPI_ISL_433390 | Virology Department, Royal Infirmary of Edinburgh, NHS Lothian / School of Biological Sciences, University of Edinburgh / Institute of Genetics and Molecular Medicine, University of Edinburgh | COVID-19 Genomics UK (COG-UK) Consortium |
| EPI_ISL_433391 | Virology Department, Royal Infirmary of Edinburgh, NHS Lothian / School of Biological Sciences, University of Edinburgh / Institute of Genetics and Molecular Medicine, University of Edinburgh | COVID-19 Genomics UK (COG-UK) Consortium |
| EPI_ISL_433392 | Virology Department, Royal Infirmary of Edinburgh, NHS Lothian / School of Biological Sciences, University of Edinburgh / Institute of Genetics and Molecular Medicine, University of Edinburgh | COVID-19 Genomics UK (COG-UK) Consortium |
| EPI_ISL_433393 | Virology Department, Royal Infirmary of Edinburgh, NHS Lothian / School of Biological Sciences, University of Edinburgh / Institute of Genetics and Molecular Medicine, University of Edinburgh | COVID-19 Genomics UK (COG-UK) Consortium |

| EPI_Accession_ID | Originating Laboratory | Submitting Laboratory |
|---|---|---|
| EPI_ISL_433394 | Virology Department, Royal Infirmary of Edinburgh, NHS Lothian / School of Biological Sciences, University of Edinburgh / Institute of Genetics and Molecular Medicine, University of Edinburgh | COVID-19 Genomics UK (COG-UK) Consortium |
| EPI_ISL_433395 | Virology Department, Royal Infirmary of Edinburgh, NHS Lothian / School of Biological Sciences, University of Edinburgh / Institute of Genetics and Molecular Medicine, University of Edinburgh | COVID-19 Genomics UK (COG-UK) Consortium |
| EPI_ISL_433396 | Virology Department, Royal Infirmary of Edinburgh, NHS Lothian / School of Biological Sciences, University of Edinburgh / Institute of Genetics and Molecular Medicine, University of Edinburgh | COVID-19 Genomics UK (COG-UK) Consortium |
| EPI_ISL_433397 | Virology Department, Royal Infirmary of Edinburgh, NHS Lothian / School of Biological Sciences, University of Edinburgh / Institute of Genetics and Molecular Medicine, University of Edinburgh | COVID-19 Genomics UK (COG-UK) Consortium |
| EPI_ISL_433398 | Virology Department, Royal Infirmary of Edinburgh, NHS Lothian / School of Biological Sciences, University of Edinburgh / Institute of Genetics and Molecular Medicine, University of Edinburgh | COVID-19 Genomics UK (COG-UK) Consortium |
| EPI_ISL_433399 | Virology Department, Royal Infirmary of Edinburgh, NHS Lothian / School of Biological Sciences, University of Edinburgh / Institute of Genetics and Molecular Medicine, University of Edinburgh | COVID-19 Genomics UK (COG-UK) Consortium |
| EPI_ISL_433400 | Virology Department, Royal Infirmary of Edinburgh, NHS Lothian / School of Biological Sciences, University of Edinburgh / Institute of Genetics and Molecular Medicine, University of Edinburgh | COVID-19 Genomics UK (COG-UK) Consortium |
| EPI_ISL_433401 | Virology Department, Royal Infirmary of Edinburgh, NHS Lothian / School of Biological Sciences, University of Edinburgh / Institute of Genetics and Molecular Medicine, University of Edinburgh | COVID-19 Genomics UK (COG-UK) Consortium |

| EPI_Accession_ID | Originating Laboratory | Submitting Laboratory |
|---|---|---|
| EPI_ISL_433402 | Virology Department, Royal Infirmary of Edinburgh, NHS Lothian / School of Biological Sciences, University of Edinburgh / Institute of Genetics and Molecular Medicine, University of Edinburgh | COVID-19 Genomics UK (COG-UK) Consortium |
| EPI_ISL_433403 | Virology Department, Royal Infirmary of Edinburgh, NHS Lothian / School of Biological Sciences, University of Edinburgh / Institute of Genetics and Molecular Medicine, University of Edinburgh | COVID-19 Genomics UK (COG-UK) Consortium |
| EPI_ISL_433404 | Virology Department, Royal Infirmary of Edinburgh, NHS Lothian / School of Biological Sciences, University of Edinburgh / Institute of Genetics and Molecular Medicine, University of Edinburgh | COVID-19 Genomics UK (COG-UK) Consortium |
| EPI_ISL_433405 | Virology Department, Royal Infirmary of Edinburgh, NHS Lothian / School of Biological Sciences, University of Edinburgh / Institute of Genetics and Molecular Medicine, University of Edinburgh | COVID-19 Genomics UK (COG-UK) Consortium |
| EPI_ISL_433406 | Virology Department, Royal Infirmary of Edinburgh, NHS Lothian / School of Biological Sciences, University of Edinburgh / Institute of Genetics and Molecular Medicine, University of Edinburgh | COVID-19 Genomics UK (COG-UK) Consortium |
| EPI_ISL_433407 | Virology Department, Royal Infirmary of Edinburgh, NHS Lothian / School of Biological Sciences, University of Edinburgh / Institute of Genetics and Molecular Medicine, University of Edinburgh | COVID-19 Genomics UK (COG-UK) Consortium |
| EPI_ISL_433408 | Virology Department, Royal Infirmary of Edinburgh, NHS Lothian / School of Biological Sciences, University of Edinburgh / Institute of Genetics and Molecular Medicine, University of Edinburgh | COVID-19 Genomics UK (COG-UK) Consortium |
| EPI_ISL_433409 | Virology Department, Royal Infirmary of Edinburgh, NHS Lothian / School of Biological Sciences, University of Edinburgh / Institute of Genetics and Molecular Medicine, University of Edinburgh | COVID-19 Genomics UK (COG-UK) Consortium |

| EPI_Accession_ID | Originating Laboratory | Submitting Laboratory |
|---|---|---|
| EPI_ISL_433410 | Virology Department, Royal Infirmary of Edinburgh, NHS Lothian / School of Biological Sciences, University of Edinburgh / Institute of Genetics and Molecular Medicine, University of Edinburgh | COVID-19 Genomics UK (COG-UK) Consortium |
| EPI_ISL_433411 | Virology Department, Royal Infirmary of Edinburgh, NHS Lothian / School of Biological Sciences, University of Edinburgh / Institute of Genetics and Molecular Medicine, University of Edinburgh | COVID-19 Genomics UK (COG-UK) Consortium |
| EPI_ISL_433412 | Virology Department, Royal Infirmary of Edinburgh, NHS Lothian / School of Biological Sciences, University of Edinburgh / Institute of Genetics and Molecular Medicine, University of Edinburgh | COVID-19 Genomics UK (COG-UK) Consortium |
| EPI_ISL_433413 | Virology Department, Royal Infirmary of Edinburgh, NHS Lothian / School of Biological Sciences, University of Edinburgh / Institute of Genetics and Molecular Medicine, University of Edinburgh | COVID-19 Genomics UK (COG-UK) Consortium |
| EPI_ISL_433414 | Virology Department, Royal Infirmary of Edinburgh, NHS Lothian / School of Biological Sciences, University of Edinburgh / Institute of Genetics and Molecular Medicine, University of Edinburgh | COVID-19 Genomics UK (COG-UK) Consortium |
| EPI_ISL_433415 | Virology Department, Royal Infirmary of Edinburgh, NHS Lothian / School of Biological Sciences, University of Edinburgh / Institute of Genetics and Molecular Medicine, University of Edinburgh | COVID-19 Genomics UK (COG-UK) Consortium |
| EPI_ISL_433416 | Virology Department, Royal Infirmary of Edinburgh, NHS Lothian / School of Biological Sciences, University of Edinburgh / Institute of Genetics and Molecular Medicine, University of Edinburgh | COVID-19 Genomics UK (COG-UK) Consortium |
| EPI_ISL_433417 | Virology Department, Royal Infirmary of Edinburgh, NHS Lothian / School of Biological Sciences, University of Edinburgh / Institute of Genetics and Molecular Medicine, University of Edinburgh | COVID-19 Genomics UK (COG-UK) Consortium |

| EPI_Accession_ID | Originating Laboratory | Submitting Laboratory |
|---|---|---|
| EPI_ISL_433418 | Virology Department, Royal Infirmary of Edinburgh, NHS Lothian / School of Biological Sciences, University of Edinburgh / Institute of Genetics and Molecular Medicine, University of Edinburgh | COVID-19 Genomics UK (COG-UK) Consortium |
| EPI_ISL_433419 | Virology Department, Royal Infirmary of Edinburgh, NHS Lothian / School of Biological Sciences, University of Edinburgh / Institute of Genetics and Molecular Medicine, University of Edinburgh | COVID-19 Genomics UK (COG-UK) Consortium |
| EPI_ISL_433420 | Virology Department, Royal Infirmary of Edinburgh, NHS Lothian / School of Biological Sciences, University of Edinburgh / Institute of Genetics and Molecular Medicine, University of Edinburgh | COVID-19 Genomics UK (COG-UK) Consortium |
| EPI_ISL_433421 | Virology Department, Royal Infirmary of Edinburgh, NHS Lothian / School of Biological Sciences, University of Edinburgh / Institute of Genetics and Molecular Medicine, University of Edinburgh | COVID-19 Genomics UK (COG-UK) Consortium |
| EPI_ISL_433422 | Virology Department, Royal Infirmary of Edinburgh, NHS Lothian / School of Biological Sciences, University of Edinburgh / Institute of Genetics and Molecular Medicine, University of Edinburgh | COVID-19 Genomics UK (COG-UK) Consortium |
| EPI_ISL_433423 | Virology Department, Royal Infirmary of Edinburgh, NHS Lothian / School of Biological Sciences, University of Edinburgh / Institute of Genetics and Molecular Medicine, University of Edinburgh | COVID-19 Genomics UK (COG-UK) Consortium |
| EPI_ISL_433424 | Virology Department, Royal Infirmary of Edinburgh, NHS Lothian / School of Biological Sciences, University of Edinburgh / Institute of Genetics and Molecular Medicine, University of Edinburgh | COVID-19 Genomics UK (COG-UK) Consortium |
| EPI_ISL_433425 | Virology Department, Royal Infirmary of Edinburgh, NHS Lothian / School of Biological Sciences, University of Edinburgh / Institute of Genetics and Molecular Medicine, University of Edinburgh | COVID-19 Genomics UK (COG-UK) Consortium |

| EPI_Accession_ID | Originating Laboratory | Submitting Laboratory |
|---|---|---|
| EPI_ISL_433426 | Virology Department, Royal Infirmary of Edinburgh, NHS Lothian / School of Biological Sciences, University of Edinburgh / Institute of Genetics and Molecular Medicine, University of Edinburgh | COVID-19 Genomics UK (COG-UK) Consortium |
| EPI_ISL_433427 | Virology Department, Royal Infirmary of Edinburgh, NHS Lothian / School of Biological Sciences, University of Edinburgh / Institute of Genetics and Molecular Medicine, University of Edinburgh | COVID-19 Genomics UK (COG-UK) Consortium |
| EPI_ISL_433428 | Virology Department, Royal Infirmary of Edinburgh, NHS Lothian / School of Biological Sciences, University of Edinburgh / Institute of Genetics and Molecular Medicine, University of Edinburgh | COVID-19 Genomics UK (COG-UK) Consortium |
| EPI_ISL_433429 | Virology Department, Royal Infirmary of Edinburgh, NHS Lothian / School of Biological Sciences, University of Edinburgh / Institute of Genetics and Molecular Medicine, University of Edinburgh | COVID-19 Genomics UK (COG-UK) Consortium |
| EPI_ISL_433430 | Virology Department, Royal Infirmary of Edinburgh, NHS Lothian / School of Biological Sciences, University of Edinburgh / Institute of Genetics and Molecular Medicine, University of Edinburgh | COVID-19 Genomics UK (COG-UK) Consortium |
| EPI_ISL_433431 | Virology Department, Royal Infirmary of Edinburgh, NHS Lothian / School of Biological Sciences, University of Edinburgh / Institute of Genetics and Molecular Medicine, University of Edinburgh | COVID-19 Genomics UK (COG-UK) Consortium |
| EPI_ISL_433432 | Virology Department, Royal Infirmary of Edinburgh, NHS Lothian / School of Biological Sciences, University of Edinburgh / Institute of Genetics and Molecular Medicine, University of Edinburgh | COVID-19 Genomics UK (COG-UK) Consortium |
| EPI_ISL_433433 | Virology Department, Royal Infirmary of Edinburgh, NHS Lothian / School of Biological Sciences, University of Edinburgh / Institute of Genetics and Molecular Medicine, University of Edinburgh | COVID-19 Genomics UK (COG-UK) Consortium |

| EPI_Accession_ID | Originating Laboratory | Submitting Laboratory |
|---|---|---|
| EPI_ISL_433434 | Virology Department, Royal Infirmary of Edinburgh, NHS Lothian / School of Biological Sciences, University of Edinburgh / Institute of Genetics and Molecular Medicine, University of Edinburgh | COVID-19 Genomics UK (COG-UK) Consortium |
| EPI_ISL_433435 | Virology Department, Royal Infirmary of Edinburgh, NHS Lothian / School of Biological Sciences, University of Edinburgh / Institute of Genetics and Molecular Medicine, University of Edinburgh | COVID-19 Genomics UK (COG-UK) Consortium |
| EPI_ISL_433436 | Virology Department, Royal Infirmary of Edinburgh, NHS Lothian / School of Biological Sciences, University of Edinburgh / Institute of Genetics and Molecular Medicine, University of Edinburgh | COVID-19 Genomics UK (COG-UK) Consortium |
| EPI_ISL_433437 | Virology Department, Royal Infirmary of Edinburgh, NHS Lothian / School of Biological Sciences, University of Edinburgh / Institute of Genetics and Molecular Medicine, University of Edinburgh | COVID-19 Genomics UK (COG-UK) Consortium |
| EPI_ISL_433438 | Virology Department, Royal Infirmary of Edinburgh, NHS Lothian / School of Biological Sciences, University of Edinburgh / Institute of Genetics and Molecular Medicine, University of Edinburgh | COVID-19 Genomics UK (COG-UK) Consortium |
| EPI_ISL_433439 | Virology Department, Royal Infirmary of Edinburgh, NHS Lothian / School of Biological Sciences, University of Edinburgh / Institute of Genetics and Molecular Medicine, University of Edinburgh | COVID-19 Genomics UK (COG-UK) Consortium |
| EPI_ISL_433440 | Virology Department, Royal Infirmary of Edinburgh, NHS Lothian / School of Biological Sciences, University of Edinburgh / Institute of Genetics and Molecular Medicine, University of Edinburgh | COVID-19 Genomics UK (COG-UK) Consortium |
| EPI_ISL_433441 | Virology Department, Royal Infirmary of Edinburgh, NHS Lothian / School of Biological Sciences, University of Edinburgh / Institute of Genetics and Molecular Medicine, University of Edinburgh | COVID-19 Genomics UK (COG-UK) Consortium |

| EPI_Accession_ID | Originating Laboratory | Submitting Laboratory |
|---|---|---|
| EPI_ISL_433442 | Virology Department, Royal Infirmary of Edinburgh, NHS Lothian / School of Biological Sciences, University of Edinburgh / Institute of Genetics and Molecular Medicine, University of Edinburgh | COVID-19 Genomics UK (COG-UK) Consortium |
| EPI_ISL_433443 | Virology Department, Royal Infirmary of Edinburgh, NHS Lothian / School of Biological Sciences, University of Edinburgh / Institute of Genetics and Molecular Medicine, University of Edinburgh | COVID-19 Genomics UK (COG-UK) Consortium |
| EPI_ISL_433444 | Virology Department, Royal Infirmary of Edinburgh, NHS Lothian / School of Biological Sciences, University of Edinburgh / Institute of Genetics and Molecular Medicine, University of Edinburgh | COVID-19 Genomics UK (COG-UK) Consortium |
| EPI_ISL_433445 | Virology Department, Royal Infirmary of Edinburgh, NHS Lothian / School of Biological Sciences, University of Edinburgh / Institute of Genetics and Molecular Medicine, University of Edinburgh | COVID-19 Genomics UK (COG-UK) Consortium |
| EPI_ISL_433446 | Virology Department, Royal Infirmary of Edinburgh, NHS Lothian / School of Biological Sciences, University of Edinburgh / Institute of Genetics and Molecular Medicine, University of Edinburgh | COVID-19 Genomics UK (COG-UK) Consortium |
| EPI_ISL_433447 | Virology Department, Royal Infirmary of Edinburgh, NHS Lothian / School of Biological Sciences, University of Edinburgh / Institute of Genetics and Molecular Medicine, University of Edinburgh | COVID-19 Genomics UK (COG-UK) Consortium |
| EPI_ISL_433448 | Virology Department, Royal Infirmary of Edinburgh, NHS Lothian / School of Biological Sciences, University of Edinburgh / Institute of Genetics and Molecular Medicine, University of Edinburgh | COVID-19 Genomics UK (COG-UK) Consortium |
| EPI_ISL_433449 | Virology Department, Royal Infirmary of Edinburgh, NHS Lothian / School of Biological Sciences, University of Edinburgh / Institute of Genetics and Molecular Medicine, University of Edinburgh | COVID-19 Genomics UK (COG-UK) Consortium |

| EPI_Accession_ID | Originating Laboratory | Submitting Laboratory |
|---|---|---|
| EPI_ISL_433450 | Virology Department, Royal Infirmary of Edinburgh, NHS Lothian / School of Biological Sciences, University of Edinburgh / Institute of Genetics and Molecular Medicine, University of Edinburgh | COVID-19 Genomics UK (COG-UK) Consortium |
| EPI_ISL_433451 | Virology Department, Royal Infirmary of Edinburgh, NHS Lothian / School of Biological Sciences, University of Edinburgh / Institute of Genetics and Molecular Medicine, University of Edinburgh | COVID-19 Genomics UK (COG-UK) Consortium |
| EPI_ISL_433452 | Virology Department, Royal Infirmary of Edinburgh, NHS Lothian / School of Biological Sciences, University of Edinburgh / Institute of Genetics and Molecular Medicine, University of Edinburgh | COVID-19 Genomics UK (COG-UK) Consortium |
| EPI_ISL_433453 | Virology Department, Royal Infirmary of Edinburgh, NHS Lothian / School of Biological Sciences, University of Edinburgh / Institute of Genetics and Molecular Medicine, University of Edinburgh | COVID-19 Genomics UK (COG-UK) Consortium |
| EPI_ISL_433454 | Virology Department, Royal Infirmary of Edinburgh, NHS Lothian / School of Biological Sciences, University of Edinburgh / Institute of Genetics and Molecular Medicine, University of Edinburgh | COVID-19 Genomics UK (COG-UK) Consortium |
| EPI_ISL_433455 | Virology Department, Royal Infirmary of Edinburgh, NHS Lothian / School of Biological Sciences, University of Edinburgh / Institute of Genetics and Molecular Medicine, University of Edinburgh | COVID-19 Genomics UK (COG-UK) Consortium |
| EPI_ISL_433456 | Virology Department, Royal Infirmary of Edinburgh, NHS Lothian / School of Biological Sciences, University of Edinburgh / Institute of Genetics and Molecular Medicine, University of Edinburgh | COVID-19 Genomics UK (COG-UK) Consortium |
| EPI_ISL_433457 | Virology Department, Royal Infirmary of Edinburgh, NHS Lothian / School of Biological Sciences, University of Edinburgh / Institute of Genetics and Molecular Medicine, University of Edinburgh | COVID-19 Genomics UK (COG-UK) Consortium |

| EPI_Accession_ID | Originating Laboratory | Submitting Laboratory |
|---|---|---|
| EPI_ISL_433458 | Virology Department, Royal Infirmary of Edinburgh, NHS Lothian / School of Biological Sciences, University of Edinburgh / Institute of Genetics and Molecular Medicine, University of Edinburgh | COVID-19 Genomics UK (COG-UK) Consortium |
| EPI_ISL_433459 | Virology Department, Royal Infirmary of Edinburgh, NHS Lothian / School of Biological Sciences, University of Edinburgh / Institute of Genetics and Molecular Medicine, University of Edinburgh | COVID-19 Genomics UK (COG-UK) Consortium |
| EPI_ISL_433460 | Virology Department, Royal Infirmary of Edinburgh, NHS Lothian / School of Biological Sciences, University of Edinburgh / Institute of Genetics and Molecular Medicine, University of Edinburgh | COVID-19 Genomics UK (COG-UK) Consortium |
| EPI_ISL_433461 | Virology Department, Royal Infirmary of Edinburgh, NHS Lothian / School of Biological Sciences, University of Edinburgh / Institute of Genetics and Molecular Medicine, University of Edinburgh | COVID-19 Genomics UK (COG-UK) Consortium |
| EPI_ISL_433462 | Virology Department, Royal Infirmary of Edinburgh, NHS Lothian / School of Biological Sciences, University of Edinburgh / Institute of Genetics and Molecular Medicine, University of Edinburgh | COVID-19 Genomics UK (COG-UK) Consortium |
| EPI_ISL_433463 | Virology Department, Royal Infirmary of Edinburgh, NHS Lothian / School of Biological Sciences, University of Edinburgh / Institute of Genetics and Molecular Medicine, University of Edinburgh | COVID-19 Genomics UK (COG-UK) Consortium |
| EPI_ISL_433464 | Virology Department, Royal Infirmary of Edinburgh, NHS Lothian / School of Biological Sciences, University of Edinburgh / Institute of Genetics and Molecular Medicine, University of Edinburgh | COVID-19 Genomics UK (COG-UK) Consortium |
| EPI_ISL_433465 | Virology Department, Royal Infirmary of Edinburgh, NHS Lothian / School of Biological Sciences, University of Edinburgh / Institute of Genetics and Molecular Medicine, University of Edinburgh | COVID-19 Genomics UK (COG-UK) Consortium |
| EPI_ISL_433466 | Department of Pathology, University of Cambridge | COVID-19 Genomics UK (COG-UK) Consortium |

| EPI_Accession_ID | Originating Laboratory | Submitting Laboratory |
|---|---|---|
| EPI_ISL_433467 | Department of Pathology, University of Cambridge | COVID-19 Genomics UK (COG-UK) Consortium |
| EPI_ISL_433468 | Department of Pathology, University of Cambridge | COVID-19 Genomics UK (COG-UK) Consortium |
| EPI_ISL_433469 | Department of Pathology, University of Cambridge | COVID-19 Genomics UK (COG-UK) Consortium |
| EPI_ISL_433470 | Department of Pathology, University of Cambridge | COVID-19 Genomics UK (COG-UK) Consortium |
| EPI_ISL_433471 | Department of Pathology, University of Cambridge | COVID-19 Genomics UK (COG-UK) Consortium |
| EPI_ISL_433472 | Department of Pathology, University of Cambridge | COVID-19 Genomics UK (COG-UK) Consortium |
| EPI_ISL_433473 | Department of Pathology, University of Cambridge | COVID-19 Genomics UK (COG-UK) Consortium |
| EPI_ISL_433474 | Department of Pathology, University of Cambridge | COVID-19 Genomics UK (COG-UK) Consortium |
| EPI_ISL_433475 | Department of Pathology, University of Cambridge | COVID-19 Genomics UK (COG-UK) Consortium |
| EPI_ISL_433476 | Department of Pathology, University of Cambridge | COVID-19 Genomics UK (COG-UK) Consortium |
| EPI_ISL_433477 | Department of Pathology, University of Cambridge | COVID-19 Genomics UK (COG-UK) Consortium |
| EPI_ISL_433478 | Department of Pathology, University of Cambridge | COVID-19 Genomics UK (COG-UK) Consortium |
| EPI_ISL_433479 | Department of Pathology, University of Cambridge | COVID-19 Genomics UK (COG-UK) Consortium |
| EPI_ISL_433480 | Department of Pathology, University of Cambridge | COVID-19 Genomics UK (COG-UK) Consortium |
| EPI_ISL_433481 | Department of Pathology, University of Cambridge | COVID-19 Genomics UK (COG-UK) Consortium |
| EPI_ISL_433482 | Department of Pathology, University of Cambridge | COVID-19 Genomics UK (COG-UK) Consortium |
| EPI_ISL_433483 | Department of Pathology, University of Cambridge | COVID-19 Genomics UK (COG-UK) Consortium |
| EPI_ISL_433484 | Department of Pathology, University of Cambridge | COVID-19 Genomics UK (COG-UK) Consortium |
| EPI_ISL_433485 | Department of Pathology, University of Cambridge | COVID-19 Genomics UK (COG-UK) Consortium |
| EPI_ISL_433486 | Department of Pathology, University of Cambridge | COVID-19 Genomics UK (COG-UK) Consortium |
| EPI_ISL_433487 | Department of Pathology, University of Cambridge | COVID-19 Genomics UK (COG-UK) Consortium |
| EPI_ISL_433489 | Department of Pathology, University of Cambridge | COVID-19 Genomics UK (COG-UK) Consortium |
| EPI_ISL_433490 | Department of Pathology, University of Cambridge | COVID-19 Genomics UK (COG-UK) Consortium |
| EPI_ISL_433491 | Department of Pathology, University of Cambridge | COVID-19 Genomics UK (COG-UK) Consortium |
| EPI_ISL_433492 | Department of Pathology, University of Cambridge | COVID-19 Genomics UK (COG-UK) Consortium |
| EPI_ISL_433493 | Department of Pathology, University of Cambridge | COVID-19 Genomics UK (COG-UK) Consortium |
| EPI_ISL_433494 | Department of Pathology, University of Cambridge | COVID-19 Genomics UK (COG-UK) Consortium |
| EPI_ISL_433495 | Department of Pathology, University of Cambridge | COVID-19 Genomics UK (COG-UK) Consortium |
| EPI_ISL_433496 | Department of Pathology, University of Cambridge | COVID-19 Genomics UK (COG-UK) Consortium |
| EPI_ISL_433497 | Department of Pathology, University of Cambridge | COVID-19 Genomics UK (COG-UK) Consortium |
| EPI_ISL_433498 | West of Scotland Specialist Virology Centre, NHSGGC / MRC-University of Glasgow Centre for Virus Research | COVID-19 Genomics UK (COG-UK) Consortium |

| EPI_Accession_ID | Originating Laboratory | Submitting Laboratory |
|---|---|---|
| EPI_ISL_433499 | West of Scotland Specialist Virology Centre, NHSGGC / MRC-University of Glasgow Centre for Virus Research | COVID-19 Genomics UK (COG-UK) Consortium |
| EPI_ISL_433500 | West of Scotland Specialist Virology Centre, NHSGGC / MRC-University of Glasgow Centre for Virus Research | COVID-19 Genomics UK (COG-UK) Consortium |
| EPI_ISL_433501 | West of Scotland Specialist Virology Centre, NHSGGC / MRC-University of Glasgow Centre for Virus Research | COVID-19 Genomics UK (COG-UK) Consortium |
| EPI_ISL_433502 | West of Scotland Specialist Virology Centre, NHSGGC / MRC-University of Glasgow Centre for Virus Research | COVID-19 Genomics UK (COG-UK) Consortium |
| EPI_ISL_433503 | West of Scotland Specialist Virology Centre, NHSGGC / MRC-University of Glasgow Centre for Virus Research | COVID-19 Genomics UK (COG-UK) Consortium |
| EPI_ISL_433504 | West of Scotland Specialist Virology Centre, NHSGGC / MRC-University of Glasgow Centre for Virus Research | COVID-19 Genomics UK (COG-UK) Consortium |
| EPI_ISL_433505 | West of Scotland Specialist Virology Centre, NHSGGC / MRC-University of Glasgow Centre for Virus Research | COVID-19 Genomics UK (COG-UK) Consortium |
| EPI_ISL_433506 | West of Scotland Specialist Virology Centre, NHSGGC / MRC-University of Glasgow Centre for Virus Research | COVID-19 Genomics UK (COG-UK) Consortium |
| EPI_ISL_433507 | West of Scotland Specialist Virology Centre, NHSGGC / MRC-University of Glasgow Centre for Virus Research | COVID-19 Genomics UK (COG-UK) Consortium |
| EPI_ISL_433508 | West of Scotland Specialist Virology Centre, NHSGGC / MRC-University of Glasgow Centre for Virus Research | COVID-19 Genomics UK (COG-UK) Consortium |
| EPI_ISL_433509 | West of Scotland Specialist Virology Centre, NHSGGC / MRC-University of Glasgow Centre for Virus Research | COVID-19 Genomics UK (COG-UK) Consortium |
| EPI_ISL_433510 | West of Scotland Specialist Virology Centre, NHSGGC / MRC-University of Glasgow Centre for Virus Research | COVID-19 Genomics UK (COG-UK) Consortium |
| EPI_ISL_433511 | West of Scotland Specialist Virology Centre, NHSGGC / MRC-University of Glasgow Centre for Virus Research | COVID-19 Genomics UK (COG-UK) Consortium |
| EPI_ISL_433512 | West of Scotland Specialist Virology Centre, NHSGGC / MRC-University of Glasgow Centre for Virus Research | COVID-19 Genomics UK (COG-UK) Consortium |
| EPI_ISL_433513 | West of Scotland Specialist Virology Centre, NHSGGC / MRC-University of Glasgow Centre for Virus Research | COVID-19 Genomics UK (COG-UK) Consortium |
| EPI_ISL_433514 | West of Scotland Specialist Virology Centre, NHSGGC / MRC-University of Glasgow Centre for Virus Research | COVID-19 Genomics UK (COG-UK) Consortium |

| EPI_Accession_ID | Originating Laboratory | Submitting Laboratory |
|---|---|---|
| EPI_ISL_433515 | West of Scotland Specialist Virology Centre, NHSGGC / MRC-University of Glasgow Centre for Virus Research | COVID-19 Genomics UK (COG-UK) Consortium |
| EPI_ISL_433516 | West of Scotland Specialist Virology Centre, NHSGGC / MRC-University of Glasgow Centre for Virus Research | COVID-19 Genomics UK (COG-UK) Consortium |
| EPI_ISL_433517 | West of Scotland Specialist Virology Centre, NHSGGC / MRC-University of Glasgow Centre for Virus Research | COVID-19 Genomics UK (COG-UK) Consortium |
| EPI_ISL_433518 | West of Scotland Specialist Virology Centre, NHSGGC / MRC-University of Glasgow Centre for Virus Research | COVID-19 Genomics UK (COG-UK) Consortium |
| EPI_ISL_433519 | West of Scotland Specialist Virology Centre, NHSGGC / MRC-University of Glasgow Centre for Virus Research | COVID-19 Genomics UK (COG-UK) Consortium |
| EPI_ISL_433520 | West of Scotland Specialist Virology Centre, NHSGGC / MRC-University of Glasgow Centre for Virus Research | COVID-19 Genomics UK (COG-UK) Consortium |
| EPI_ISL_433521 | West of Scotland Specialist Virology Centre, NHSGGC / MRC-University of Glasgow Centre for Virus Research | COVID-19 Genomics UK (COG-UK) Consortium |
| EPI_ISL_433522 | West of Scotland Specialist Virology Centre, NHSGGC / MRC-University of Glasgow Centre for Virus Research | COVID-19 Genomics UK (COG-UK) Consortium |
| EPI_ISL_433523 | West of Scotland Specialist Virology Centre, NHSGGC / MRC-University of Glasgow Centre for Virus Research | COVID-19 Genomics UK (COG-UK) Consortium |
| EPI_ISL_433524 | West of Scotland Specialist Virology Centre, NHSGGC / MRC-University of Glasgow Centre for Virus Research | COVID-19 Genomics UK (COG-UK) Consortium |
| EPI_ISL_433525 | West of Scotland Specialist Virology Centre, NHSGGC / MRC-University of Glasgow Centre for Virus Research | COVID-19 Genomics UK (COG-UK) Consortium |
| EPI_ISL_433526 | West of Scotland Specialist Virology Centre, NHSGGC / MRC-University of Glasgow Centre for Virus Research | COVID-19 Genomics UK (COG-UK) Consortium |
| EPI_ISL_433527 | West of Scotland Specialist Virology Centre, NHSGGC / MRC-University of Glasgow Centre for Virus Research | COVID-19 Genomics UK (COG-UK) Consortium |
| EPI_ISL_433528 | West of Scotland Specialist Virology Centre, NHSGGC / MRC-University of Glasgow Centre for Virus Research | COVID-19 Genomics UK (COG-UK) Consortium |
| EPI_ISL_433529 | West of Scotland Specialist Virology Centre, NHSGGC / MRC-University of Glasgow Centre for Virus Research | COVID-19 Genomics UK (COG-UK) Consortium |
| EPI_ISL_433530 | West of Scotland Specialist Virology Centre, NHSGGC / MRC-University of Glasgow Centre for Virus Research | COVID-19 Genomics UK (COG-UK) Consortium |

| EPI_Accession_ID | Originating Laboratory | Submitting Laboratory |
| --- | --- | --- |
| EPI_ISL_433531 | West of Scotland Specialist Virology Centre, NHSGGC / MRC-University of Glasgow Centre for Virus Research | COVID-19 Genomics UK (COG-UK) Consortium |
| EPI_ISL_433532 | West of Scotland Specialist Virology Centre, NHSGGC / MRC-University of Glasgow Centre for Virus Research | COVID-19 Genomics UK (COG-UK) Consortium |
| EPI_ISL_433533 | West of Scotland Specialist Virology Centre, NHSGGC / MRC-University of Glasgow Centre for Virus Research | COVID-19 Genomics UK (COG-UK) Consortium |
| EPI_ISL_433534 | West of Scotland Specialist Virology Centre, NHSGGC / MRC-University of Glasgow Centre for Virus Research | COVID-19 Genomics UK (COG-UK) Consortium |
| EPI_ISL_433535 | West of Scotland Specialist Virology Centre, NHSGGC / MRC-University of Glasgow Centre for Virus Research | COVID-19 Genomics UK (COG-UK) Consortium |
| EPI_ISL_433536 | West of Scotland Specialist Virology Centre, NHSGGC / MRC-University of Glasgow Centre for Virus Research | COVID-19 Genomics UK (COG-UK) Consortium |
| EPI_ISL_433537 | West of Scotland Specialist Virology Centre, NHSGGC / MRC-University of Glasgow Centre for Virus Research | COVID-19 Genomics UK (COG-UK) Consortium |
| EPI_ISL_433538 | West of Scotland Specialist Virology Centre, NHSGGC / MRC-University of Glasgow Centre for Virus Research | COVID-19 Genomics UK (COG-UK) Consortium |
| EPI_ISL_433539 | West of Scotland Specialist Virology Centre, NHSGGC / MRC-University of Glasgow Centre for Virus Research | COVID-19 Genomics UK (COG-UK) Consortium |
| EPI_ISL_433540 | West of Scotland Specialist Virology Centre, NHSGGC / MRC-University of Glasgow Centre for Virus Research | COVID-19 Genomics UK (COG-UK) Consortium |
| EPI_ISL_433541 | West of Scotland Specialist Virology Centre, NHSGGC / MRC-University of Glasgow Centre for Virus Research | COVID-19 Genomics UK (COG-UK) Consortium |
| EPI_ISL_433542 | West of Scotland Specialist Virology Centre, NHSGGC / MRC-University of Glasgow Centre for Virus Research | COVID-19 Genomics UK (COG-UK) Consortium |
| EPI_ISL_433543 | West of Scotland Specialist Virology Centre, NHSGGC / MRC-University of Glasgow Centre for Virus Research | COVID-19 Genomics UK (COG-UK) Consortium |
| EPI_ISL_433544 | West of Scotland Specialist Virology Centre, NHSGGC / MRC-University of Glasgow Centre for Virus Research | COVID-19 Genomics UK (COG-UK) Consortium |
| EPI_ISL_433545 | West of Scotland Specialist Virology Centre, NHSGGC / MRC-University of Glasgow Centre for Virus Research | COVID-19 Genomics UK (COG-UK) Consortium |
| EPI_ISL_433546 | West of Scotland Specialist Virology Centre, NHSGGC / MRC-University of Glasgow Centre for Virus Research | COVID-19 Genomics UK (COG-UK) Consortium |

| EPI_Accession_ID | Originating Laboratory | Submitting Laboratory |
|---|---|---|
| EPI_ISL_433547 | West of Scotland Specialist Virology Centre, NHSGGC / MRC-University of Glasgow Centre for Virus Research | COVID-19 Genomics UK (COG-UK) Consortium |
| EPI_ISL_433548 | West of Scotland Specialist Virology Centre, NHSGGC / MRC-University of Glasgow Centre for Virus Research | COVID-19 Genomics UK (COG-UK) Consortium |
| EPI_ISL_433549 | West of Scotland Specialist Virology Centre, NHSGGC / MRC-University of Glasgow Centre for Virus Research | COVID-19 Genomics UK (COG-UK) Consortium |
| EPI_ISL_433550 | West of Scotland Specialist Virology Centre, NHSGGC / MRC-University of Glasgow Centre for Virus Research | COVID-19 Genomics UK (COG-UK) Consortium |
| EPI_ISL_433551 | West of Scotland Specialist Virology Centre, NHSGGC / MRC-University of Glasgow Centre for Virus Research | COVID-19 Genomics UK (COG-UK) Consortium |
| EPI_ISL_433552 | West of Scotland Specialist Virology Centre, NHSGGC / MRC-University of Glasgow Centre for Virus Research | COVID-19 Genomics UK (COG-UK) Consortium |
| EPI_ISL_433553 | West of Scotland Specialist Virology Centre, NHSGGC / MRC-University of Glasgow Centre for Virus Research | COVID-19 Genomics UK (COG-UK) Consortium |
| EPI_ISL_433554 | West of Scotland Specialist Virology Centre, NHSGGC / MRC-University of Glasgow Centre for Virus Research | COVID-19 Genomics UK (COG-UK) Consortium |
| EPI_ISL_433555 | West of Scotland Specialist Virology Centre, NHSGGC / MRC-University of Glasgow Centre for Virus Research | COVID-19 Genomics UK (COG-UK) Consortium |
| EPI_ISL_433556 | West of Scotland Specialist Virology Centre, NHSGGC / MRC-University of Glasgow Centre for Virus Research | COVID-19 Genomics UK (COG-UK) Consortium |
| EPI_ISL_433557 | West of Scotland Specialist Virology Centre, NHSGGC / MRC-University of Glasgow Centre for Virus Research | COVID-19 Genomics UK (COG-UK) Consortium |
| EPI_ISL_433558 | West of Scotland Specialist Virology Centre, NHSGGC / MRC-University of Glasgow Centre for Virus Research | COVID-19 Genomics UK (COG-UK) Consortium |
| EPI_ISL_433559 | West of Scotland Specialist Virology Centre, NHSGGC / MRC-University of Glasgow Centre for Virus Research | COVID-19 Genomics UK (COG-UK) Consortium |
| EPI_ISL_433560 | West of Scotland Specialist Virology Centre, NHSGGC / MRC-University of Glasgow Centre for Virus Research | COVID-19 Genomics UK (COG-UK) Consortium |
| EPI_ISL_433561 | West of Scotland Specialist Virology Centre, NHSGGC / MRC-University of Glasgow Centre for Virus Research | COVID-19 Genomics UK (COG-UK) Consortium |
| EPI_ISL_433562 | West of Scotland Specialist Virology Centre, NHSGGC / MRC-University of Glasgow Centre for Virus Research | COVID-19 Genomics UK (COG-UK) Consortium |

| EPI_Accession_ID | Originating Laboratory | Submitting Laboratory |
|---|---|---|
| EPI_ISL_433563 | West of Scotland Specialist Virology Centre, NHSGGC / MRC-University of Glasgow Centre for Virus Research | COVID-19 Genomics UK (COG-UK) Consortium |
| EPI_ISL_433564 | West of Scotland Specialist Virology Centre, NHSGGC / MRC-University of Glasgow Centre for Virus Research | COVID-19 Genomics UK (COG-UK) Consortium |
| EPI_ISL_433565 | West of Scotland Specialist Virology Centre, NHSGGC / MRC-University of Glasgow Centre for Virus Research | COVID-19 Genomics UK (COG-UK) Consortium |
| EPI_ISL_433566 | West of Scotland Specialist Virology Centre, NHSGGC / MRC-University of Glasgow Centre for Virus Research | COVID-19 Genomics UK (COG-UK) Consortium |
| EPI_ISL_433567 | West of Scotland Specialist Virology Centre, NHSGGC / MRC-University of Glasgow Centre for Virus Research | COVID-19 Genomics UK (COG-UK) Consortium |
| EPI_ISL_433568 | West of Scotland Specialist Virology Centre, NHSGGC / MRC-University of Glasgow Centre for Virus Research | COVID-19 Genomics UK (COG-UK) Consortium |
| EPI_ISL_433569 | West of Scotland Specialist Virology Centre, NHSGGC / MRC-University of Glasgow Centre for Virus Research | COVID-19 Genomics UK (COG-UK) Consortium |
| EPI_ISL_433570 | West of Scotland Specialist Virology Centre, NHSGGC / MRC-University of Glasgow Centre for Virus Research | COVID-19 Genomics UK (COG-UK) Consortium |
| EPI_ISL_433571 | West of Scotland Specialist Virology Centre, NHSGGC / MRC-University of Glasgow Centre for Virus Research | COVID-19 Genomics UK (COG-UK) Consortium |
| EPI_ISL_433572 | West of Scotland Specialist Virology Centre, NHSGGC / MRC-University of Glasgow Centre for Virus Research | COVID-19 Genomics UK (COG-UK) Consortium |
| EPI_ISL_433573 | West of Scotland Specialist Virology Centre, NHSGGC / MRC-University of Glasgow Centre for Virus Research | COVID-19 Genomics UK (COG-UK) Consortium |
| EPI_ISL_433574 | West of Scotland Specialist Virology Centre, NHSGGC / MRC-University of Glasgow Centre for Virus Research | COVID-19 Genomics UK (COG-UK) Consortium |
| EPI_ISL_433575 | West of Scotland Specialist Virology Centre, NHSGGC / MRC-University of Glasgow Centre for Virus Research | COVID-19 Genomics UK (COG-UK) Consortium |
| EPI_ISL_433576 | West of Scotland Specialist Virology Centre, NHSGGC / MRC-University of Glasgow Centre for Virus Research | COVID-19 Genomics UK (COG-UK) Consortium |
| EPI_ISL_433577 | West of Scotland Specialist Virology Centre, NHSGGC / MRC-University of Glasgow Centre for Virus Research | COVID-19 Genomics UK (COG-UK) Consortium |
| EPI_ISL_433578 | West of Scotland Specialist Virology Centre, NHSGGC / MRC-University of Glasgow Centre for Virus Research | COVID-19 Genomics UK (COG-UK) Consortium |

| EPI_Accession_ID | Originating Laboratory | Submitting Laboratory |
|---|---|---|
| EPI_ISL_433579 | West of Scotland Specialist Virology Centre, NHSGGC / MRC-University of Glasgow Centre for Virus Research | COVID-19 Genomics UK (COG-UK) Consortium |
| EPI_ISL_433580 | West of Scotland Specialist Virology Centre, NHSGGC / MRC-University of Glasgow Centre for Virus Research | COVID-19 Genomics UK (COG-UK) Consortium |
| EPI_ISL_433581 | West of Scotland Specialist Virology Centre, NHSGGC / MRC-University of Glasgow Centre for Virus Research | COVID-19 Genomics UK (COG-UK) Consortium |
| EPI_ISL_433582 | West of Scotland Specialist Virology Centre, NHSGGC / MRC-University of Glasgow Centre for Virus Research | COVID-19 Genomics UK (COG-UK) Consortium |
| EPI_ISL_433583 | West of Scotland Specialist Virology Centre, NHSGGC / MRC-University of Glasgow Centre for Virus Research | COVID-19 Genomics UK (COG-UK) Consortium |
| EPI_ISL_433584 | West of Scotland Specialist Virology Centre, NHSGGC / MRC-University of Glasgow Centre for Virus Research | COVID-19 Genomics UK (COG-UK) Consortium |
| EPI_ISL_433585 | West of Scotland Specialist Virology Centre, NHSGGC / MRC-University of Glasgow Centre for Virus Research | COVID-19 Genomics UK (COG-UK) Consortium |
| EPI_ISL_433586 | West of Scotland Specialist Virology Centre, NHSGGC / MRC-University of Glasgow Centre for Virus Research | COVID-19 Genomics UK (COG-UK) Consortium |
| EPI_ISL_433587 | West of Scotland Specialist Virology Centre, NHSGGC / MRC-University of Glasgow Centre for Virus Research | COVID-19 Genomics UK (COG-UK) Consortium |
| EPI_ISL_433588 | West of Scotland Specialist Virology Centre, NHSGGC / MRC-University of Glasgow Centre for Virus Research | COVID-19 Genomics UK (COG-UK) Consortium |
| EPI_ISL_433589 | West of Scotland Specialist Virology Centre, NHSGGC / MRC-University of Glasgow Centre for Virus Research | COVID-19 Genomics UK (COG-UK) Consortium |
| EPI_ISL_433590 | West of Scotland Specialist Virology Centre, NHSGGC / MRC-University of Glasgow Centre for Virus Research | COVID-19 Genomics UK (COG-UK) Consortium |
| EPI_ISL_433591 | West of Scotland Specialist Virology Centre, NHSGGC / MRC-University of Glasgow Centre for Virus Research | COVID-19 Genomics UK (COG-UK) Consortium |
| EPI_ISL_433592 | West of Scotland Specialist Virology Centre, NHSGGC / MRC-University of Glasgow Centre for Virus Research | COVID-19 Genomics UK (COG-UK) Consortium |
| EPI_ISL_433593 | West of Scotland Specialist Virology Centre, NHSGGC / MRC-University of Glasgow Centre for Virus Research | COVID-19 Genomics UK (COG-UK) Consortium |
| EPI_ISL_433594 | West of Scotland Specialist Virology Centre, NHSGGC / MRC-University of Glasgow Centre for Virus Research | COVID-19 Genomics UK (COG-UK) Consortium |

| EPI_Accession_ID | Originating Laboratory | Submitting Laboratory |
| --- | --- | --- |
| EPI_ISL_433595 | West of Scotland Specialist Virology Centre, NHSGGC / MRC-University of Glasgow Centre for Virus Research | COVID-19 Genomics UK (COG-UK) Consortium |
| EPI_ISL_433596 | West of Scotland Specialist Virology Centre, NHSGGC / MRC-University of Glasgow Centre for Virus Research | COVID-19 Genomics UK (COG-UK) Consortium |
| EPI_ISL_433597 | West of Scotland Specialist Virology Centre, NHSGGC / MRC-University of Glasgow Centre for Virus Research | COVID-19 Genomics UK (COG-UK) Consortium |
| EPI_ISL_433598 | West of Scotland Specialist Virology Centre, NHSGGC / MRC-University of Glasgow Centre for Virus Research | COVID-19 Genomics UK (COG-UK) Consortium |
| EPI_ISL_433599 | West of Scotland Specialist Virology Centre, NHSGGC / MRC-University of Glasgow Centre for Virus Research | COVID-19 Genomics UK (COG-UK) Consortium |
| EPI_ISL_433600 | West of Scotland Specialist Virology Centre, NHSGGC / MRC-University of Glasgow Centre for Virus Research | COVID-19 Genomics UK (COG-UK) Consortium |
| EPI_ISL_433601 | West of Scotland Specialist Virology Centre, NHSGGC / MRC-University of Glasgow Centre for Virus Research | COVID-19 Genomics UK (COG-UK) Consortium |
| EPI_ISL_433602 | West of Scotland Specialist Virology Centre, NHSGGC / MRC-University of Glasgow Centre for Virus Research | COVID-19 Genomics UK (COG-UK) Consortium |
| EPI_ISL_433603 | West of Scotland Specialist Virology Centre, NHSGGC / MRC-University of Glasgow Centre for Virus Research | COVID-19 Genomics UK (COG-UK) Consortium |
| EPI_ISL_433604 | West of Scotland Specialist Virology Centre, NHSGGC / MRC-University of Glasgow Centre for Virus Research | COVID-19 Genomics UK (COG-UK) Consortium |
| EPI_ISL_433605 | West of Scotland Specialist Virology Centre, NHSGGC / MRC-University of Glasgow Centre for Virus Research | COVID-19 Genomics UK (COG-UK) Consortium |
| EPI_ISL_433606 | West of Scotland Specialist Virology Centre, NHSGGC / MRC-University of Glasgow Centre for Virus Research | COVID-19 Genomics UK (COG-UK) Consortium |
| EPI_ISL_433607 | West of Scotland Specialist Virology Centre, NHSGGC / MRC-University of Glasgow Centre for Virus Research | COVID-19 Genomics UK (COG-UK) Consortium |
| EPI_ISL_433608 | West of Scotland Specialist Virology Centre, NHSGGC / MRC-University of Glasgow Centre for Virus Research | COVID-19 Genomics UK (COG-UK) Consortium |
| EPI_ISL_433609 | West of Scotland Specialist Virology Centre, NHSGGC / MRC-University of Glasgow Centre for Virus Research | COVID-19 Genomics UK (COG-UK) Consortium |
| EPI_ISL_433610 | West of Scotland Specialist Virology Centre, NHSGGC / MRC-University of Glasgow Centre for Virus Research | COVID-19 Genomics UK (COG-UK) Consortium |

| EPI_Accession_ID | Originating Laboratory | Submitting Laboratory |
|---|---|---|
| EPI_ISL_433611 | West of Scotland Specialist Virology Centre, NHSGGC / MRC-University of Glasgow Centre for Virus Research | COVID-19 Genomics UK (COG-UK) Consortium |
| EPI_ISL_433612 | West of Scotland Specialist Virology Centre, NHSGGC / MRC-University of Glasgow Centre for Virus Research | COVID-19 Genomics UK (COG-UK) Consortium |
| EPI_ISL_433613 | West of Scotland Specialist Virology Centre, NHSGGC / MRC-University of Glasgow Centre for Virus Research | COVID-19 Genomics UK (COG-UK) Consortium |
| EPI_ISL_433614 | West of Scotland Specialist Virology Centre, NHSGGC / MRC-University of Glasgow Centre for Virus Research | COVID-19 Genomics UK (COG-UK) Consortium |
| EPI_ISL_433615 | West of Scotland Specialist Virology Centre, NHSGGC / MRC-University of Glasgow Centre for Virus Research | COVID-19 Genomics UK (COG-UK) Consortium |
| EPI_ISL_433616 | West of Scotland Specialist Virology Centre, NHSGGC / MRC-University of Glasgow Centre for Virus Research | COVID-19 Genomics UK (COG-UK) Consortium |
| EPI_ISL_433617 | West of Scotland Specialist Virology Centre, NHSGGC / MRC-University of Glasgow Centre for Virus Research | COVID-19 Genomics UK (COG-UK) Consortium |
| EPI_ISL_433618 | West of Scotland Specialist Virology Centre, NHSGGC / MRC-University of Glasgow Centre for Virus Research | COVID-19 Genomics UK (COG-UK) Consortium |
| EPI_ISL_433619 | West of Scotland Specialist Virology Centre, NHSGGC / MRC-University of Glasgow Centre for Virus Research | COVID-19 Genomics UK (COG-UK) Consortium |
| EPI_ISL_433620 | West of Scotland Specialist Virology Centre, NHSGGC / MRC-University of Glasgow Centre for Virus Research | COVID-19 Genomics UK (COG-UK) Consortium |
| EPI_ISL_433621 | West of Scotland Specialist Virology Centre, NHSGGC / MRC-University of Glasgow Centre for Virus Research | COVID-19 Genomics UK (COG-UK) Consortium |
| EPI_ISL_433622 | West of Scotland Specialist Virology Centre, NHSGGC / MRC-University of Glasgow Centre for Virus Research | COVID-19 Genomics UK (COG-UK) Consortium |
| EPI_ISL_433623 | West of Scotland Specialist Virology Centre, NHSGGC / MRC-University of Glasgow Centre for Virus Research | COVID-19 Genomics UK (COG-UK) Consortium |
| EPI_ISL_433624 | West of Scotland Specialist Virology Centre, NHSGGC / MRC-University of Glasgow Centre for Virus Research | COVID-19 Genomics UK (COG-UK) Consortium |
| EPI_ISL_433625 | West of Scotland Specialist Virology Centre, NHSGGC / MRC-University of Glasgow Centre for Virus Research | COVID-19 Genomics UK (COG-UK) Consortium |
| EPI_ISL_433626 | West of Scotland Specialist Virology Centre, NHSGGC / MRC-University of Glasgow Centre for Virus Research | COVID-19 Genomics UK (COG-UK) Consortium |

| EPI_Accession_ID | Originating Laboratory | Submitting Laboratory |
|---|---|---|
| EPI_ISL_433627 | West of Scotland Specialist Virology Centre, NHSGGC / MRC-University of Glasgow Centre for Virus Research | COVID-19 Genomics UK (COG-UK) Consortium |
| EPI_ISL_433628 | West of Scotland Specialist Virology Centre, NHSGGC / MRC-University of Glasgow Centre for Virus Research | COVID-19 Genomics UK (COG-UK) Consortium |
| EPI_ISL_433629 | West of Scotland Specialist Virology Centre, NHSGGC / MRC-University of Glasgow Centre for Virus Research | COVID-19 Genomics UK (COG-UK) Consortium |
| EPI_ISL_433630 | West of Scotland Specialist Virology Centre, NHSGGC / MRC-University of Glasgow Centre for Virus Research | COVID-19 Genomics UK (COG-UK) Consortium |
| EPI_ISL_433631 | West of Scotland Specialist Virology Centre, NHSGGC / MRC-University of Glasgow Centre for Virus Research | COVID-19 Genomics UK (COG-UK) Consortium |
| EPI_ISL_433632 | West of Scotland Specialist Virology Centre, NHSGGC / MRC-University of Glasgow Centre for Virus Research | COVID-19 Genomics UK (COG-UK) Consortium |
| EPI_ISL_433633 | West of Scotland Specialist Virology Centre, NHSGGC / MRC-University of Glasgow Centre for Virus Research | COVID-19 Genomics UK (COG-UK) Consortium |
| EPI_ISL_433634 | West of Scotland Specialist Virology Centre, NHSGGC / MRC-University of Glasgow Centre for Virus Research | COVID-19 Genomics UK (COG-UK) Consortium |
| EPI_ISL_433635 | West of Scotland Specialist Virology Centre, NHSGGC / MRC-University of Glasgow Centre for Virus Research | COVID-19 Genomics UK (COG-UK) Consortium |
| EPI_ISL_433636 | West of Scotland Specialist Virology Centre, NHSGGC / MRC-University of Glasgow Centre for Virus Research | COVID-19 Genomics UK (COG-UK) Consortium |
| EPI_ISL_433637 | West of Scotland Specialist Virology Centre, NHSGGC / MRC-University of Glasgow Centre for Virus Research | COVID-19 Genomics UK (COG-UK) Consortium |
| EPI_ISL_433638 | West of Scotland Specialist Virology Centre, NHSGGC / MRC-University of Glasgow Centre for Virus Research | COVID-19 Genomics UK (COG-UK) Consortium |
| EPI_ISL_433639 | West of Scotland Specialist Virology Centre, NHSGGC / MRC-University of Glasgow Centre for Virus Research | COVID-19 Genomics UK (COG-UK) Consortium |
| EPI_ISL_433640 | West of Scotland Specialist Virology Centre, NHSGGC / MRC-University of Glasgow Centre for Virus Research | COVID-19 Genomics UK (COG-UK) Consortium |
| EPI_ISL_433641 | West of Scotland Specialist Virology Centre, NHSGGC / MRC-University of Glasgow Centre for Virus Research | COVID-19 Genomics UK (COG-UK) Consortium |
| EPI_ISL_433642 | West of Scotland Specialist Virology Centre, NHSGGC / MRC-University of Glasgow Centre for Virus Research | COVID-19 Genomics UK (COG-UK) Consortium |

| EPI_Accession_ID | Originating Laboratory | Submitting Laboratory |
|---|---|---|
| EPI_ISL_433643 | West of Scotland Specialist Virology Centre, NHSGGC / MRC-University of Glasgow Centre for Virus Research | COVID-19 Genomics UK (COG-UK) Consortium |
| EPI_ISL_433644 | West of Scotland Specialist Virology Centre, NHSGGC / MRC-University of Glasgow Centre for Virus Research | COVID-19 Genomics UK (COG-UK) Consortium |
| EPI_ISL_433645 | West of Scotland Specialist Virology Centre, NHSGGC / MRC-University of Glasgow Centre for Virus Research | COVID-19 Genomics UK (COG-UK) Consortium |
| EPI_ISL_433646 | West of Scotland Specialist Virology Centre, NHSGGC / MRC-University of Glasgow Centre for Virus Research | COVID-19 Genomics UK (COG-UK) Consortium |
| EPI_ISL_433647 | West of Scotland Specialist Virology Centre, NHSGGC / MRC-University of Glasgow Centre for Virus Research | COVID-19 Genomics UK (COG-UK) Consortium |
| EPI_ISL_433648 | West of Scotland Specialist Virology Centre, NHSGGC / MRC-University of Glasgow Centre for Virus Research | COVID-19 Genomics UK (COG-UK) Consortium |
| EPI_ISL_433649 | West of Scotland Specialist Virology Centre, NHSGGC / MRC-University of Glasgow Centre for Virus Research | COVID-19 Genomics UK (COG-UK) Consortium |
| EPI_ISL_433650 | West of Scotland Specialist Virology Centre, NHSGGC / MRC-University of Glasgow Centre for Virus Research | COVID-19 Genomics UK (COG-UK) Consortium |
| EPI_ISL_433651 | West of Scotland Specialist Virology Centre, NHSGGC / MRC-University of Glasgow Centre for Virus Research | COVID-19 Genomics UK (COG-UK) Consortium |
| EPI_ISL_433652 | West of Scotland Specialist Virology Centre, NHSGGC / MRC-University of Glasgow Centre for Virus Research | COVID-19 Genomics UK (COG-UK) Consortium |
| EPI_ISL_433653 | West of Scotland Specialist Virology Centre, NHSGGC / MRC-University of Glasgow Centre for Virus Research | COVID-19 Genomics UK (COG-UK) Consortium |
| EPI_ISL_433654 | West of Scotland Specialist Virology Centre, NHSGGC / MRC-University of Glasgow Centre for Virus Research | COVID-19 Genomics UK (COG-UK) Consortium |
| EPI_ISL_433655 | West of Scotland Specialist Virology Centre, NHSGGC / MRC-University of Glasgow Centre for Virus Research | COVID-19 Genomics UK (COG-UK) Consortium |
| EPI_ISL_433656 | West of Scotland Specialist Virology Centre, NHSGGC / MRC-University of Glasgow Centre for Virus Research | COVID-19 Genomics UK (COG-UK) Consortium |
| EPI_ISL_433657 | West of Scotland Specialist Virology Centre, NHSGGC / MRC-University of Glasgow Centre for Virus Research | COVID-19 Genomics UK (COG-UK) Consortium |
| EPI_ISL_433658 | West of Scotland Specialist Virology Centre, NHSGGC / MRC-University of Glasgow Centre for Virus Research | COVID-19 Genomics UK (COG-UK) Consortium |

| EPI_Accession_ID | Originating Laboratory | Submitting Laboratory |
|---|---|---|
| EPI_ISL_433659 | West of Scotland Specialist Virology Centre, NHSGGC / MRC-University of Glasgow Centre for Virus Research | COVID-19 Genomics UK (COG-UK) Consortium |
| EPI_ISL_433660 | West of Scotland Specialist Virology Centre, NHSGGC / MRC-University of Glasgow Centre for Virus Research | COVID-19 Genomics UK (COG-UK) Consortium |
| EPI_ISL_433661 | West of Scotland Specialist Virology Centre, NHSGGC / MRC-University of Glasgow Centre for Virus Research | COVID-19 Genomics UK (COG-UK) Consortium |
| EPI_ISL_433662 | West of Scotland Specialist Virology Centre, NHSGGC / MRC-University of Glasgow Centre for Virus Research | COVID-19 Genomics UK (COG-UK) Consortium |
| EPI_ISL_433663 | West of Scotland Specialist Virology Centre, NHSGGC / MRC-University of Glasgow Centre for Virus Research | COVID-19 Genomics UK (COG-UK) Consortium |
| EPI_ISL_433664 | West of Scotland Specialist Virology Centre, NHSGGC / MRC-University of Glasgow Centre for Virus Research | COVID-19 Genomics UK (COG-UK) Consortium |
| EPI_ISL_433665 | West of Scotland Specialist Virology Centre, NHSGGC / MRC-University of Glasgow Centre for Virus Research | COVID-19 Genomics UK (COG-UK) Consortium |
| EPI_ISL_433666 | Department of Pathology, University of Cambridge | COVID-19 Genomics UK (COG-UK) Consortium |
| EPI_ISL_433667 | Department of Pathology, University of Cambridge | COVID-19 Genomics UK (COG-UK) Consortium |
| EPI_ISL_433668 | Department of Pathology, University of Cambridge | COVID-19 Genomics UK (COG-UK) Consortium |
| EPI_ISL_433669 | Department of Pathology, University of Cambridge | COVID-19 Genomics UK (COG-UK) Consortium |
| EPI_ISL_433670 | Department of Pathology, University of Cambridge | COVID-19 Genomics UK (COG-UK) Consortium |
| EPI_ISL_433671 | Department of Pathology, University of Cambridge | COVID-19 Genomics UK (COG-UK) Consortium |
| EPI_ISL_433672 | Department of Pathology, University of Cambridge | COVID-19 Genomics UK (COG-UK) Consortium |
| EPI_ISL_433673 | Department of Pathology, University of Cambridge | COVID-19 Genomics UK (COG-UK) Consortium |
| EPI_ISL_433674 | Department of Pathology, University of Cambridge | COVID-19 Genomics UK (COG-UK) Consortium |
| EPI_ISL_433675 | Department of Pathology, University of Cambridge | COVID-19 Genomics UK (COG-UK) Consortium |
| EPI_ISL_433676 | Department of Pathology, University of Cambridge | COVID-19 Genomics UK (COG-UK) Consortium |
| EPI_ISL_433677 | Department of Pathology, University of Cambridge | COVID-19 Genomics UK (COG-UK) Consortium |
| EPI_ISL_433678 | Department of Pathology, University of Cambridge | COVID-19 Genomics UK (COG-UK) Consortium |
| EPI_ISL_433679 | Department of Pathology, University of Cambridge | COVID-19 Genomics UK (COG-UK) Consortium |
| EPI_ISL_433680 | Department of Pathology, University of Cambridge | COVID-19 Genomics UK (COG-UK) Consortium |
| EPI_ISL_433681 | Department of Pathology, University of Cambridge | COVID-19 Genomics UK (COG-UK) Consortium |
| EPI_ISL_433682 | Department of Pathology, University of Cambridge | COVID-19 Genomics UK (COG-UK) Consortium |
| EPI_ISL_433683 | Department of Pathology, University of Cambridge | COVID-19 Genomics UK (COG-UK) Consortium |
| EPI_ISL_433684 | Department of Pathology, University of Cambridge | COVID-19 Genomics UK (COG-UK) Consortium |

| EPI_Accession_ID | Originating Laboratory | Submitting Laboratory |
|---|---|---|
| EPI_ISL_433685 | Department of Pathology, University of Cambridge | COVID-19 Genomics UK (COG-UK) Consortium |
| EPI_ISL_433686 | Department of Pathology, University of Cambridge | COVID-19 Genomics UK (COG-UK) Consortium |
| EPI_ISL_433688 | Department of Pathology, University of Cambridge | COVID-19 Genomics UK (COG-UK) Consortium |
| EPI_ISL_433689 | Department of Pathology, University of Cambridge | COVID-19 Genomics UK (COG-UK) Consortium |
| EPI_ISL_433690 | Department of Pathology, University of Cambridge | COVID-19 Genomics UK (COG-UK) Consortium |
| EPI_ISL_433691 | Department of Pathology, University of Cambridge | COVID-19 Genomics UK (COG-UK) Consortium |
| EPI_ISL_433692 | Department of Pathology, University of Cambridge | COVID-19 Genomics UK (COG-UK) Consortium |
| EPI_ISL_433693 | Department of Pathology, University of Cambridge | COVID-19 Genomics UK (COG-UK) Consortium |
| EPI_ISL_433694 | Department of Pathology, University of Cambridge | COVID-19 Genomics UK (COG-UK) Consortium |
| EPI_ISL_433695 | Department of Pathology, University of Cambridge | COVID-19 Genomics UK (COG-UK) Consortium |
| EPI_ISL_433696 | Department of Pathology, University of Cambridge | COVID-19 Genomics UK (COG-UK) Consortium |
| EPI_ISL_433697 | Department of Pathology, University of Cambridge | COVID-19 Genomics UK (COG-UK) Consortium |
| EPI_ISL_433698 | Department of Pathology, University of Cambridge | COVID-19 Genomics UK (COG-UK) Consortium |
| EPI_ISL_433699 | Department of Pathology, University of Cambridge | COVID-19 Genomics UK (COG-UK) Consortium |
| EPI_ISL_433700 | Department of Pathology, University of Cambridge | COVID-19 Genomics UK (COG-UK) Consortium |
| EPI_ISL_433701 | Department of Pathology, University of Cambridge | COVID-19 Genomics UK (COG-UK) Consortium |
| EPI_ISL_433702 | Department of Pathology, University of Cambridge | COVID-19 Genomics UK (COG-UK) Consortium |
| EPI_ISL_433703 | Department of Pathology, University of Cambridge | COVID-19 Genomics UK (COG-UK) Consortium |
| EPI_ISL_433704 | Department of Pathology, University of Cambridge | COVID-19 Genomics UK (COG-UK) Consortium |
| EPI_ISL_433705 | Department of Pathology, University of Cambridge | COVID-19 Genomics UK (COG-UK) Consortium |
| EPI_ISL_433706 | Department of Pathology, University of Cambridge | COVID-19 Genomics UK (COG-UK) Consortium |
| EPI_ISL_433707 | Department of Pathology, University of Cambridge | COVID-19 Genomics UK (COG-UK) Consortium |
| EPI_ISL_433709 | Department of Pathology, University of Cambridge | COVID-19 Genomics UK (COG-UK) Consortium |
| EPI_ISL_433710 | Department of Pathology, University of Cambridge | COVID-19 Genomics UK (COG-UK) Consortium |
| EPI_ISL_433711 | Department of Pathology, University of Cambridge | COVID-19 Genomics UK (COG-UK) Consortium |
| EPI_ISL_433712 | Department of Pathology, University of Cambridge | COVID-19 Genomics UK (COG-UK) Consortium |
| EPI_ISL_433713 | Department of Pathology, University of Cambridge | COVID-19 Genomics UK (COG-UK) Consortium |
| EPI_ISL_433714 | Department of Pathology, University of Cambridge | COVID-19 Genomics UK (COG-UK) Consortium |
| EPI_ISL_433715 | Department of Pathology, University of Cambridge | COVID-19 Genomics UK (COG-UK) Consortium |
| EPI_ISL_433717 | Department of Pathology, University of Cambridge | COVID-19 Genomics UK (COG-UK) Consortium |
| EPI_ISL_433718 | Department of Pathology, University of Cambridge | COVID-19 Genomics UK (COG-UK) Consortium |
| EPI_ISL_433719 | Department of Pathology, University of Cambridge | COVID-19 Genomics UK (COG-UK) Consortium |
| EPI_ISL_433720 | Department of Pathology, University of Cambridge | COVID-19 Genomics UK (COG-UK) Consortium |

| EPI_Accession_ID | Originating Laboratory | Submitting Laboratory |
|---|---|---|
| EPI_ISL_433721 | Department of Pathology, University of Cambridge | COVID-19 Genomics UK (COG-UK) Consortium |
| EPI_ISL_433722 | Department of Pathology, University of Cambridge | COVID-19 Genomics UK (COG-UK) Consortium |
| EPI_ISL_433723 | Department of Pathology, University of Cambridge | COVID-19 Genomics UK (COG-UK) Consortium |
| EPI_ISL_433724 | Department of Pathology, University of Cambridge | COVID-19 Genomics UK (COG-UK) Consortium |
| EPI_ISL_433725 | Department of Pathology, University of Cambridge | COVID-19 Genomics UK (COG-UK) Consortium |
| EPI_ISL_433726 | Department of Pathology, University of Cambridge | COVID-19 Genomics UK (COG-UK) Consortium |
| EPI_ISL_433727 | Department of Pathology, University of Cambridge | COVID-19 Genomics UK (COG-UK) Consortium |
| EPI_ISL_433728 | Department of Pathology, University of Cambridge | COVID-19 Genomics UK (COG-UK) Consortium |
| EPI_ISL_433729 | Department of Pathology, University of Cambridge | COVID-19 Genomics UK (COG-UK) Consortium |
| EPI_ISL_433730 | Department of Pathology, University of Cambridge | COVID-19 Genomics UK (COG-UK) Consortium |
| EPI_ISL_433731 | Department of Pathology, University of Cambridge | COVID-19 Genomics UK (COG-UK) Consortium |
| EPI_ISL_433732 | Department of Pathology, University of Cambridge | COVID-19 Genomics UK (COG-UK) Consortium |
| EPI_ISL_433733 | Department of Pathology, University of Cambridge | COVID-19 Genomics UK (COG-UK) Consortium |
| EPI_ISL_433734 | Department of Pathology, University of Cambridge | COVID-19 Genomics UK (COG-UK) Consortium |
| EPI_ISL_433735 | Department of Pathology, University of Cambridge | COVID-19 Genomics UK (COG-UK) Consortium |
| EPI_ISL_433736 | Department of Pathology, University of Cambridge | COVID-19 Genomics UK (COG-UK) Consortium |
| EPI_ISL_433737 | Department of Pathology, University of Cambridge | COVID-19 Genomics UK (COG-UK) Consortium |
| EPI_ISL_433738 | Department of Pathology, University of Cambridge | COVID-19 Genomics UK (COG-UK) Consortium |
| EPI_ISL_433739 | Department of Pathology, University of Cambridge | COVID-19 Genomics UK (COG-UK) Consortium |
| EPI_ISL_433741 | Department of Pathology, University of Cambridge | COVID-19 Genomics UK (COG-UK) Consortium |
| EPI_ISL_433742 | Department of Pathology, University of Cambridge | COVID-19 Genomics UK (COG-UK) Consortium |
| EPI_ISL_433743 | Department of Pathology, University of Cambridge | COVID-19 Genomics UK (COG-UK) Consortium |
| EPI_ISL_433744 | Department of Pathology, University of Cambridge | COVID-19 Genomics UK (COG-UK) Consortium |
| EPI_ISL_433745 | Department of Pathology, University of Cambridge | COVID-19 Genomics UK (COG-UK) Consortium |
| EPI_ISL_433746 | Department of Pathology, University of Cambridge | COVID-19 Genomics UK (COG-UK) Consortium |
| EPI_ISL_433748 | Department of Pathology, University of Cambridge | COVID-19 Genomics UK (COG-UK) Consortium |
| EPI_ISL_433749 | Department of Pathology, University of Cambridge | COVID-19 Genomics UK (COG-UK) Consortium |
| EPI_ISL_433750 | Department of Pathology, University of Cambridge | COVID-19 Genomics UK (COG-UK) Consortium |
| EPI_ISL_433751 | Department of Pathology, University of Cambridge | COVID-19 Genomics UK (COG-UK) Consortium |
| EPI_ISL_433752 | Department of Pathology, University of Cambridge | COVID-19 Genomics UK (COG-UK) Consortium |
| EPI_ISL_433753 | Department of Pathology, University of Cambridge | COVID-19 Genomics UK (COG-UK) Consortium |
| EPI_ISL_433754 | Department of Pathology, University of Cambridge | COVID-19 Genomics UK (COG-UK) Consortium |
| EPI_ISL_433755 | Department of Pathology, University of Cambridge | COVID-19 Genomics UK (COG-UK) Consortium |

| EPI_Accession_ID | Originating Laboratory | Submitting Laboratory |
|---|---|---|
| EPI_ISL_433756 | Department of Pathology, University of Cambridge | COVID-19 Genomics UK (COG-UK) Consortium |
| EPI_ISL_433757 | Department of Pathology, University of Cambridge | COVID-19 Genomics UK (COG-UK) Consortium |
| EPI_ISL_433758 | Department of Pathology, University of Cambridge | COVID-19 Genomics UK (COG-UK) Consortium |
| EPI_ISL_433759 | Department of Pathology, University of Cambridge | COVID-19 Genomics UK (COG-UK) Consortium |
| EPI_ISL_433760 | Department of Pathology, University of Cambridge | COVID-19 Genomics UK (COG-UK) Consortium |
| EPI_ISL_433761 | Department of Pathology, University of Cambridge | COVID-19 Genomics UK (COG-UK) Consortium |
| EPI_ISL_433762 | Department of Pathology, University of Cambridge | COVID-19 Genomics UK (COG-UK) Consortium |
| EPI_ISL_433763 | Department of Pathology, University of Cambridge | COVID-19 Genomics UK (COG-UK) Consortium |
| EPI_ISL_433764 | Department of Pathology, University of Cambridge | COVID-19 Genomics UK (COG-UK) Consortium |
| EPI_ISL_433765 | Department of Pathology, University of Cambridge | COVID-19 Genomics UK (COG-UK) Consortium |
| EPI_ISL_433767 | Department of Pathology, University of Cambridge | COVID-19 Genomics UK (COG-UK) Consortium |
| EPI_ISL_433768 | Department of Pathology, University of Cambridge | COVID-19 Genomics UK (COG-UK) Consortium |
| EPI_ISL_433769 | Department of Pathology, University of Cambridge | COVID-19 Genomics UK (COG-UK) Consortium |
| EPI_ISL_433770 | Department of Pathology, University of Cambridge | COVID-19 Genomics UK (COG-UK) Consortium |
| EPI_ISL_433771 | Department of Pathology, University of Cambridge | COVID-19 Genomics UK (COG-UK) Consortium |
| EPI_ISL_433772 | Department of Pathology, University of Cambridge | COVID-19 Genomics UK (COG-UK) Consortium |
| EPI_ISL_433773 | Department of Pathology, University of Cambridge | COVID-19 Genomics UK (COG-UK) Consortium |
| EPI_ISL_433774 | Department of Pathology, University of Cambridge | COVID-19 Genomics UK (COG-UK) Consortium |
| EPI_ISL_433775 | Department of Pathology, University of Cambridge | COVID-19 Genomics UK (COG-UK) Consortium |
| EPI_ISL_433776 | Department of Pathology, University of Cambridge | COVID-19 Genomics UK (COG-UK) Consortium |
| EPI_ISL_433777 | Department of Pathology, University of Cambridge | COVID-19 Genomics UK (COG-UK) Consortium |
| EPI_ISL_433778 | Department of Pathology, University of Cambridge | COVID-19 Genomics UK (COG-UK) Consortium |
| EPI_ISL_433779 | Department of Pathology, University of Cambridge | COVID-19 Genomics UK (COG-UK) Consortium |
| EPI_ISL_433780 | Department of Pathology, University of Cambridge | COVID-19 Genomics UK (COG-UK) Consortium |
| EPI_ISL_433781 | Department of Pathology, University of Cambridge | COVID-19 Genomics UK (COG-UK) Consortium |
| EPI_ISL_433782 | Department of Pathology, University of Cambridge | COVID-19 Genomics UK (COG-UK) Consortium |
| EPI_ISL_433783 | Department of Pathology, University of Cambridge | COVID-19 Genomics UK (COG-UK) Consortium |
| EPI_ISL_433784 | Department of Pathology, University of Cambridge | COVID-19 Genomics UK (COG-UK) Consortium |
| EPI_ISL_433785 | Department of Pathology, University of Cambridge | COVID-19 Genomics UK (COG-UK) Consortium |
| EPI_ISL_433786 | Department of Pathology, University of Cambridge | COVID-19 Genomics UK (COG-UK) Consortium |
| EPI_ISL_433787 | Department of Pathology, University of Cambridge | COVID-19 Genomics UK (COG-UK) Consortium |
| EPI_ISL_433788 | Department of Pathology, University of Cambridge | COVID-19 Genomics UK (COG-UK) Consortium |
| EPI_ISL_433789 | Department of Pathology, University of Cambridge | COVID-19 Genomics UK (COG-UK) Consortium |

| EPI_Accession_ID | Originating Laboratory | Submitting Laboratory |
|---|---|---|
| EPI_ISL_433790 | Department of Pathology, University of Cambridge | COVID-19 Genomics UK (COG-UK) Consortium |
| EPI_ISL_433791 | Department of Pathology, University of Cambridge | COVID-19 Genomics UK (COG-UK) Consortium |
| EPI_ISL_433792 | Department of Pathology, University of Cambridge | COVID-19 Genomics UK (COG-UK) Consortium |
| EPI_ISL_433793 | Department of Pathology, University of Cambridge | COVID-19 Genomics UK (COG-UK) Consortium |
| EPI_ISL_433794 | Department of Pathology, University of Cambridge | COVID-19 Genomics UK (COG-UK) Consortium |
| EPI_ISL_433795 | Department of Pathology, University of Cambridge | COVID-19 Genomics UK (COG-UK) Consortium |
| EPI_ISL_433796 | Department of Pathology, University of Cambridge | COVID-19 Genomics UK (COG-UK) Consortium |
| EPI_ISL_433797 | Department of Pathology, University of Cambridge | COVID-19 Genomics UK (COG-UK) Consortium |
| EPI_ISL_433798 | Department of Pathology, University of Cambridge | COVID-19 Genomics UK (COG-UK) Consortium |
| EPI_ISL_433799 | Department of Pathology, University of Cambridge | COVID-19 Genomics UK (COG-UK) Consortium |
| EPI_ISL_433800 | Department of Pathology, University of Cambridge | COVID-19 Genomics UK (COG-UK) Consortium |
| EPI_ISL_433801 | Department of Pathology, University of Cambridge | COVID-19 Genomics UK (COG-UK) Consortium |
| EPI_ISL_433802 | Department of Pathology, University of Cambridge | COVID-19 Genomics UK (COG-UK) Consortium |
| EPI_ISL_433803 | Department of Pathology, University of Cambridge | COVID-19 Genomics UK (COG-UK) Consortium |
| EPI_ISL_433804 | Department of Pathology, University of Cambridge | COVID-19 Genomics UK (COG-UK) Consortium |
| EPI_ISL_433805 | Department of Pathology, University of Cambridge | COVID-19 Genomics UK (COG-UK) Consortium |
| EPI_ISL_433806 | Department of Pathology, University of Cambridge | COVID-19 Genomics UK (COG-UK) Consortium |
| EPI_ISL_433808 | Department of Pathology, University of Cambridge | COVID-19 Genomics UK (COG-UK) Consortium |
| EPI_ISL_433809 | Department of Pathology, University of Cambridge | COVID-19 Genomics UK (COG-UK) Consortium |
| EPI_ISL_433810 | Department of Pathology, University of Cambridge | COVID-19 Genomics UK (COG-UK) Consortium |
| EPI_ISL_433811 | Department of Pathology, University of Cambridge | COVID-19 Genomics UK (COG-UK) Consortium |
| EPI_ISL_433812 | Department of Pathology, University of Cambridge | COVID-19 Genomics UK (COG-UK) Consortium |
| EPI_ISL_433813 | Department of Pathology, University of Cambridge | COVID-19 Genomics UK (COG-UK) Consortium |
| EPI_ISL_433814 | Department of Pathology, University of Cambridge | COVID-19 Genomics UK (COG-UK) Consortium |
| EPI_ISL_433815 | Department of Pathology, University of Cambridge | COVID-19 Genomics UK (COG-UK) Consortium |
| EPI_ISL_433816 | Department of Pathology, University of Cambridge | COVID-19 Genomics UK (COG-UK) Consortium |
| EPI_ISL_433817 | Department of Pathology, University of Cambridge | COVID-19 Genomics UK (COG-UK) Consortium |
| EPI_ISL_433818 | Department of Pathology, University of Cambridge | COVID-19 Genomics UK (COG-UK) Consortium |
| EPI_ISL_433819 | Department of Pathology, University of Cambridge | COVID-19 Genomics UK (COG-UK) Consortium |
| EPI_ISL_433820 | Department of Pathology, University of Cambridge | COVID-19 Genomics UK (COG-UK) Consortium |
| EPI_ISL_433821 | Department of Pathology, University of Cambridge | COVID-19 Genomics UK (COG-UK) Consortium |
| EPI_ISL_433822 | Department of Pathology, University of Cambridge | COVID-19 Genomics UK (COG-UK) Consortium |
| EPI_ISL_433823 | Department of Pathology, University of Cambridge | COVID-19 Genomics UK (COG-UK) Consortium |

| EPI_Accession_ID | Originating Laboratory | Submitting Laboratory |
|---|---|---|
| EPI_ISL_433824 | Department of Pathology, University of Cambridge | COVID-19 Genomics UK (COG-UK) Consortium |
| EPI_ISL_433825 | Department of Pathology, University of Cambridge | COVID-19 Genomics UK (COG-UK) Consortium |
| EPI_ISL_433826 | Department of Pathology, University of Cambridge | COVID-19 Genomics UK (COG-UK) Consortium |
| EPI_ISL_433827 | Department of Pathology, University of Cambridge | COVID-19 Genomics UK (COG-UK) Consortium |
| EPI_ISL_433828 | Department of Pathology, University of Cambridge | COVID-19 Genomics UK (COG-UK) Consortium |
| EPI_ISL_433829 | Department of Pathology, University of Cambridge | COVID-19 Genomics UK (COG-UK) Consortium |
| EPI_ISL_433830 | Department of Pathology, University of Cambridge | COVID-19 Genomics UK (COG-UK) Consortium |
| EPI_ISL_433831 | Department of Pathology, University of Cambridge | COVID-19 Genomics UK (COG-UK) Consortium |
| EPI_ISL_433832 | Department of Pathology, University of Cambridge | COVID-19 Genomics UK (COG-UK) Consortium |
| EPI_ISL_433833 | Department of Pathology, University of Cambridge | COVID-19 Genomics UK (COG-UK) Consortium |
| EPI_ISL_433834 | Department of Pathology, University of Cambridge | COVID-19 Genomics UK (COG-UK) Consortium |
| EPI_ISL_433835 | Department of Pathology, University of Cambridge | COVID-19 Genomics UK (COG-UK) Consortium |
| EPI_ISL_433836 | Department of Pathology, University of Cambridge | COVID-19 Genomics UK (COG-UK) Consortium |
| EPI_ISL_433837 | Department of Pathology, University of Cambridge | COVID-19 Genomics UK (COG-UK) Consortium |
| EPI_ISL_433839 | Department of Pathology, University of Cambridge | COVID-19 Genomics UK (COG-UK) Consortium |
| EPI_ISL_433840 | Department of Pathology, University of Cambridge | COVID-19 Genomics UK (COG-UK) Consortium |
| EPI_ISL_433841 | Department of Pathology, University of Cambridge | COVID-19 Genomics UK (COG-UK) Consortium |
| EPI_ISL_433842 | Department of Pathology, University of Cambridge | COVID-19 Genomics UK (COG-UK) Consortium |
| EPI_ISL_433843 | Department of Pathology, University of Cambridge | COVID-19 Genomics UK (COG-UK) Consortium |
| EPI_ISL_433844 | Department of Pathology, University of Cambridge | COVID-19 Genomics UK (COG-UK) Consortium |
| EPI_ISL_433845 | Department of Pathology, University of Cambridge | COVID-19 Genomics UK (COG-UK) Consortium |
| EPI_ISL_433846 | Department of Pathology, University of Cambridge | COVID-19 Genomics UK (COG-UK) Consortium |
| EPI_ISL_433847 | Department of Pathology, University of Cambridge | COVID-19 Genomics UK (COG-UK) Consortium |
| EPI_ISL_433848 | Department of Pathology, University of Cambridge | COVID-19 Genomics UK (COG-UK) Consortium |
| EPI_ISL_433849 | Department of Pathology, University of Cambridge | COVID-19 Genomics UK (COG-UK) Consortium |
| EPI_ISL_433850 | Department of Pathology, University of Cambridge | COVID-19 Genomics UK (COG-UK) Consortium |
| EPI_ISL_433851 | Department of Pathology, University of Cambridge | COVID-19 Genomics UK (COG-UK) Consortium |
| EPI_ISL_433852 | Department of Pathology, University of Cambridge | COVID-19 Genomics UK (COG-UK) Consortium |
| EPI_ISL_433853 | Department of Pathology, University of Cambridge | COVID-19 Genomics UK (COG-UK) Consortium |
| EPI_ISL_433854 | Department of Pathology, University of Cambridge | COVID-19 Genomics UK (COG-UK) Consortium |
| EPI_ISL_433855 | Department of Pathology, University of Cambridge | COVID-19 Genomics UK (COG-UK) Consortium |
| EPI_ISL_433856 | Department of Pathology, University of Cambridge | COVID-19 Genomics UK (COG-UK) Consortium |
| EPI_ISL_433857 | Department of Pathology, University of Cambridge | COVID-19 Genomics UK (COG-UK) Consortium |

| EPI_Accession_ID | Originating Laboratory | Submitting Laboratory |
|---|---|---|
| EPI_ISL_433858 | Department of Pathology, University of Cambridge | COVID-19 Genomics UK (COG-UK) Consortium |
| EPI_ISL_433859 | Department of Pathology, University of Cambridge | COVID-19 Genomics UK (COG-UK) Consortium |
| EPI_ISL_433860 | Department of Pathology, University of Cambridge | COVID-19 Genomics UK (COG-UK) Consortium |
| EPI_ISL_433861 | Department of Pathology, University of Cambridge | COVID-19 Genomics UK (COG-UK) Consortium |
| EPI_ISL_433862 | Department of Pathology, University of Cambridge | COVID-19 Genomics UK (COG-UK) Consortium |
| EPI_ISL_433863 | Department of Pathology, University of Cambridge | COVID-19 Genomics UK (COG-UK) Consortium |
| EPI_ISL_433864 | Department of Pathology, University of Cambridge | COVID-19 Genomics UK (COG-UK) Consortium |
| EPI_ISL_433865 | Department of Pathology, University of Cambridge | COVID-19 Genomics UK (COG-UK) Consortium |
| EPI_ISL_433866 | Department of Pathology, University of Cambridge | COVID-19 Genomics UK (COG-UK) Consortium |
| EPI_ISL_433867 | Department of Pathology, University of Cambridge | COVID-19 Genomics UK (COG-UK) Consortium |
| EPI_ISL_433868 | Department of Pathology, University of Cambridge | COVID-19 Genomics UK (COG-UK) Consortium |
| EPI_ISL_433869 | Department of Pathology, University of Cambridge | COVID-19 Genomics UK (COG-UK) Consortium |
| EPI_ISL_433870 | Department of Pathology, University of Cambridge | COVID-19 Genomics UK (COG-UK) Consortium |
| EPI_ISL_433871 | Department of Pathology, University of Cambridge | COVID-19 Genomics UK (COG-UK) Consortium |
| EPI_ISL_433872 | Department of Pathology, University of Cambridge | COVID-19 Genomics UK (COG-UK) Consortium |
| EPI_ISL_433873 | Department of Pathology, University of Cambridge | COVID-19 Genomics UK (COG-UK) Consortium |
| EPI_ISL_433874 | Department of Pathology, University of Cambridge | COVID-19 Genomics UK (COG-UK) Consortium |
| EPI_ISL_433875 | Department of Pathology, University of Cambridge | COVID-19 Genomics UK (COG-UK) Consortium |
| EPI_ISL_433876 | Department of Pathology, University of Cambridge | COVID-19 Genomics UK (COG-UK) Consortium |
| EPI_ISL_433877 | Department of Pathology, University of Cambridge | COVID-19 Genomics UK (COG-UK) Consortium |
| EPI_ISL_433878 | Department of Pathology, University of Cambridge | COVID-19 Genomics UK (COG-UK) Consortium |
| EPI_ISL_433879 | Department of Pathology, University of Cambridge | COVID-19 Genomics UK (COG-UK) Consortium |
| EPI_ISL_433880 | Department of Pathology, University of Cambridge | COVID-19 Genomics UK (COG-UK) Consortium |
| EPI_ISL_433881 | Department of Pathology, University of Cambridge | COVID-19 Genomics UK (COG-UK) Consortium |
| EPI_ISL_433882 | Department of Pathology, University of Cambridge | COVID-19 Genomics UK (COG-UK) Consortium |
| EPI_ISL_433883 | Department of Pathology, University of Cambridge | COVID-19 Genomics UK (COG-UK) Consortium |
| EPI_ISL_433884 | Department of Pathology, University of Cambridge | COVID-19 Genomics UK (COG-UK) Consortium |
| EPI_ISL_433885 | Department of Pathology, University of Cambridge | COVID-19 Genomics UK (COG-UK) Consortium |
| EPI_ISL_433886 | Department of Pathology, University of Cambridge | COVID-19 Genomics UK (COG-UK) Consortium |
| EPI_ISL_433887 | Department of Pathology, University of Cambridge | COVID-19 Genomics UK (COG-UK) Consortium |
| EPI_ISL_433888 | Department of Pathology, University of Cambridge | COVID-19 Genomics UK (COG-UK) Consortium |
| EPI_ISL_433889 | Department of Pathology, University of Cambridge | COVID-19 Genomics UK (COG-UK) Consortium |
| EPI_ISL_433890 | Department of Pathology, University of Cambridge | COVID-19 Genomics UK (COG-UK) Consortium |

| EPI_Accession_ID | Originating Laboratory | Submitting Laboratory |
|---|---|---|
| EPI_ISL_433891 | Department of Pathology, University of Cambridge | COVID-19 Genomics UK (COG-UK) Consortium |
| EPI_ISL_433892 | Department of Pathology, University of Cambridge | COVID-19 Genomics UK (COG-UK) Consortium |
| EPI_ISL_433893 | Department of Pathology, University of Cambridge | COVID-19 Genomics UK (COG-UK) Consortium |
| EPI_ISL_433894 | Department of Pathology, University of Cambridge | COVID-19 Genomics UK (COG-UK) Consortium |
| EPI_ISL_433895 | Department of Pathology, University of Cambridge | COVID-19 Genomics UK (COG-UK) Consortium |
| EPI_ISL_433896 | Department of Pathology, University of Cambridge | COVID-19 Genomics UK (COG-UK) Consortium |
| EPI_ISL_433897 | Department of Pathology, University of Cambridge | COVID-19 Genomics UK (COG-UK) Consortium |
| EPI_ISL_433898 | Department of Pathology, University of Cambridge | COVID-19 Genomics UK (COG-UK) Consortium |
| EPI_ISL_433899 | Department of Pathology, University of Cambridge | COVID-19 Genomics UK (COG-UK) Consortium |
| EPI_ISL_433900 | Department of Pathology, University of Cambridge | COVID-19 Genomics UK (COG-UK) Consortium |
| EPI_ISL_433901 | Department of Pathology, University of Cambridge | COVID-19 Genomics UK (COG-UK) Consortium |
| EPI_ISL_433902 | Department of Pathology, University of Cambridge | COVID-19 Genomics UK (COG-UK) Consortium |
| EPI_ISL_433903 | Department of Pathology, University of Cambridge | COVID-19 Genomics UK (COG-UK) Consortium |
| EPI_ISL_433904 | Department of Pathology, University of Cambridge | COVID-19 Genomics UK (COG-UK) Consortium |
| EPI_ISL_433905 | Department of Pathology, University of Cambridge | COVID-19 Genomics UK (COG-UK) Consortium |
| EPI_ISL_433906 | Department of Pathology, University of Cambridge | COVID-19 Genomics UK (COG-UK) Consortium |
| EPI_ISL_433907 | Department of Pathology, University of Cambridge | COVID-19 Genomics UK (COG-UK) Consortium |
| EPI_ISL_433908 | Department of Pathology, University of Cambridge | COVID-19 Genomics UK (COG-UK) Consortium |
| EPI_ISL_433909 | Department of Pathology, University of Cambridge | COVID-19 Genomics UK (COG-UK) Consortium |
| EPI_ISL_433910 | Department of Pathology, University of Cambridge | COVID-19 Genomics UK (COG-UK) Consortium |
| EPI_ISL_433911 | Department of Pathology, University of Cambridge | COVID-19 Genomics UK (COG-UK) Consortium |
| EPI_ISL_433912 | Department of Pathology, University of Cambridge | COVID-19 Genomics UK (COG-UK) Consortium |
| EPI_ISL_433913 | Department of Pathology, University of Cambridge | COVID-19 Genomics UK (COG-UK) Consortium |
| EPI_ISL_433914 | Department of Pathology, University of Cambridge | COVID-19 Genomics UK (COG-UK) Consortium |
| EPI_ISL_433915 | Department of Pathology, University of Cambridge | COVID-19 Genomics UK (COG-UK) Consortium |
| EPI_ISL_433916 | Department of Pathology, University of Cambridge | COVID-19 Genomics UK (COG-UK) Consortium |
| EPI_ISL_433917 | Department of Pathology, University of Cambridge | COVID-19 Genomics UK (COG-UK) Consortium |
| EPI_ISL_433918 | Department of Pathology, University of Cambridge | COVID-19 Genomics UK (COG-UK) Consortium |
| EPI_ISL_433919 | Department of Pathology, University of Cambridge | COVID-19 Genomics UK (COG-UK) Consortium |
| EPI_ISL_433920 | Department of Pathology, University of Cambridge | COVID-19 Genomics UK (COG-UK) Consortium |
| EPI_ISL_433921 | Department of Pathology, University of Cambridge | COVID-19 Genomics UK (COG-UK) Consortium |
| EPI_ISL_433922 | Department of Pathology, University of Cambridge | COVID-19 Genomics UK (COG-UK) Consortium |
| EPI_ISL_433923 | Department of Pathology, University of Cambridge | COVID-19 Genomics UK (COG-UK) Consortium |

| EPI_Accession_ID | Originating Laboratory | Submitting Laboratory |
|---|---|---|
| EPI_ISL_433924 | Department of Pathology, University of Cambridge | COVID-19 Genomics UK (COG-UK) Consortium |
| EPI_ISL_433925 | Department of Pathology, University of Cambridge | COVID-19 Genomics UK (COG-UK) Consortium |
| EPI_ISL_433926 | Department of Pathology, University of Cambridge | COVID-19 Genomics UK (COG-UK) Consortium |
| EPI_ISL_433927 | Department of Pathology, University of Cambridge | COVID-19 Genomics UK (COG-UK) Consortium |
| EPI_ISL_433928 | Department of Pathology, University of Cambridge | COVID-19 Genomics UK (COG-UK) Consortium |
| EPI_ISL_433929 | Department of Pathology, University of Cambridge | COVID-19 Genomics UK (COG-UK) Consortium |
| EPI_ISL_433930 | Department of Pathology, University of Cambridge | COVID-19 Genomics UK (COG-UK) Consortium |
| EPI_ISL_433931 | Department of Pathology, University of Cambridge | COVID-19 Genomics UK (COG-UK) Consortium |
| EPI_ISL_433932 | Department of Pathology, University of Cambridge | COVID-19 Genomics UK (COG-UK) Consortium |
| EPI_ISL_433933 | Department of Pathology, University of Cambridge | COVID-19 Genomics UK (COG-UK) Consortium |
| EPI_ISL_433934 | Department of Pathology, University of Cambridge | COVID-19 Genomics UK (COG-UK) Consortium |
| EPI_ISL_433935 | Department of Pathology, University of Cambridge | COVID-19 Genomics UK (COG-UK) Consortium |
| EPI_ISL_433936 | Department of Pathology, University of Cambridge | COVID-19 Genomics UK (COG-UK) Consortium |
| EPI_ISL_433937 | Department of Pathology, University of Cambridge | COVID-19 Genomics UK (COG-UK) Consortium |
| EPI_ISL_433938 | Department of Pathology, University of Cambridge | COVID-19 Genomics UK (COG-UK) Consortium |
| EPI_ISL_433939 | Department of Pathology, University of Cambridge | COVID-19 Genomics UK (COG-UK) Consortium |
| EPI_ISL_433940 | Department of Pathology, University of Cambridge | COVID-19 Genomics UK (COG-UK) Consortium |
| EPI_ISL_433941 | Department of Pathology, University of Cambridge | COVID-19 Genomics UK (COG-UK) Consortium |
| EPI_ISL_433942 | Department of Pathology, University of Cambridge | COVID-19 Genomics UK (COG-UK) Consortium |
| EPI_ISL_433943 | Department of Pathology, University of Cambridge | COVID-19 Genomics UK (COG-UK) Consortium |
| EPI_ISL_433944 | Department of Pathology, University of Cambridge | COVID-19 Genomics UK (COG-UK) Consortium |
| EPI_ISL_433945 | Department of Pathology, University of Cambridge | COVID-19 Genomics UK (COG-UK) Consortium |
| EPI_ISL_433946 | Department of Pathology, University of Cambridge | COVID-19 Genomics UK (COG-UK) Consortium |
| EPI_ISL_433947 | Department of Pathology, University of Cambridge | COVID-19 Genomics UK (COG-UK) Consortium |
| EPI_ISL_433948 | Department of Pathology, University of Cambridge | COVID-19 Genomics UK (COG-UK) Consortium |
| EPI_ISL_433949 | Department of Pathology, University of Cambridge | COVID-19 Genomics UK (COG-UK) Consortium |
| EPI_ISL_433950 | Department of Pathology, University of Cambridge | COVID-19 Genomics UK (COG-UK) Consortium |
| EPI_ISL_433951 | Department of Pathology, University of Cambridge | COVID-19 Genomics UK (COG-UK) Consortium |
| EPI_ISL_433952 | Department of Pathology, University of Cambridge | COVID-19 Genomics UK (COG-UK) Consortium |
| EPI_ISL_433953 | Department of Pathology, University of Cambridge | COVID-19 Genomics UK (COG-UK) Consortium |
| EPI_ISL_433954 | Department of Pathology, University of Cambridge | COVID-19 Genomics UK (COG-UK) Consortium |
| EPI_ISL_433955 | Department of Pathology, University of Cambridge | COVID-19 Genomics UK (COG-UK) Consortium |
| EPI_ISL_433956 | Department of Pathology, University of Cambridge | COVID-19 Genomics UK (COG-UK) Consortium |

| EPI_Accession_ID | Originating Laboratory | Submitting Laboratory |
|---|---|---|
| EPI_ISL_433957 | Department of Pathology, University of Cambridge | COVID-19 Genomics UK (COG-UK) Consortium |
| EPI_ISL_433958 | Department of Pathology, University of Cambridge | COVID-19 Genomics UK (COG-UK) Consortium |
| EPI_ISL_433959 | Department of Pathology, University of Cambridge | COVID-19 Genomics UK (COG-UK) Consortium |
| EPI_ISL_433960 | Department of Pathology, University of Cambridge | COVID-19 Genomics UK (COG-UK) Consortium |
| EPI_ISL_433961 | Department of Pathology, University of Cambridge | COVID-19 Genomics UK (COG-UK) Consortium |
| EPI_ISL_433962 | Department of Pathology, University of Cambridge | COVID-19 Genomics UK (COG-UK) Consortium |
| EPI_ISL_433963 | Department of Pathology, University of Cambridge | COVID-19 Genomics UK (COG-UK) Consortium |
| EPI_ISL_433964 | Department of Pathology, University of Cambridge | COVID-19 Genomics UK (COG-UK) Consortium |
| EPI_ISL_433965 | Department of Pathology, University of Cambridge | COVID-19 Genomics UK (COG-UK) Consortium |
| EPI_ISL_433966 | Department of Pathology, University of Cambridge | COVID-19 Genomics UK (COG-UK) Consortium |
| EPI_ISL_433967 | Department of Pathology, University of Cambridge | COVID-19 Genomics UK (COG-UK) Consortium |
| EPI_ISL_433968 | Department of Pathology, University of Cambridge | COVID-19 Genomics UK (COG-UK) Consortium |
| EPI_ISL_433969 | Department of Pathology, University of Cambridge | COVID-19 Genomics UK (COG-UK) Consortium |
| EPI_ISL_433970 | Department of Pathology, University of Cambridge | COVID-19 Genomics UK (COG-UK) Consortium |
| EPI_ISL_433971 | Department of Pathology, University of Cambridge | COVID-19 Genomics UK (COG-UK) Consortium |
| EPI_ISL_433974 | Department of Pathology, University of Cambridge | COVID-19 Genomics UK (COG-UK) Consortium |
| EPI_ISL_433975 | Department of Pathology, University of Cambridge | COVID-19 Genomics UK (COG-UK) Consortium |
| EPI_ISL_433976 | Department of Pathology, University of Cambridge | COVID-19 Genomics UK (COG-UK) Consortium |
| EPI_ISL_433977 | Department of Pathology, University of Cambridge | COVID-19 Genomics UK (COG-UK) Consortium |
| EPI_ISL_433978 | Department of Pathology, University of Cambridge | COVID-19 Genomics UK (COG-UK) Consortium |
| EPI_ISL_433979 | Department of Pathology, University of Cambridge | COVID-19 Genomics UK (COG-UK) Consortium |
| EPI_ISL_433980 | Department of Pathology, University of Cambridge | COVID-19 Genomics UK (COG-UK) Consortium |
| EPI_ISL_433981 | Department of Pathology, University of Cambridge | COVID-19 Genomics UK (COG-UK) Consortium |
| EPI_ISL_433982 | Department of Pathology, University of Cambridge | COVID-19 Genomics UK (COG-UK) Consortium |
| EPI_ISL_433983 | Department of Pathology, University of Cambridge | COVID-19 Genomics UK (COG-UK) Consortium |
| EPI_ISL_433986 | Department of Pathology, University of Cambridge | COVID-19 Genomics UK (COG-UK) Consortium |
| EPI_ISL_433987 | Department of Pathology, University of Cambridge | COVID-19 Genomics UK (COG-UK) Consortium |
| EPI_ISL_433988 | Department of Pathology, University of Cambridge | COVID-19 Genomics UK (COG-UK) Consortium |
| EPI_ISL_433989 | Department of Pathology, University of Cambridge | COVID-19 Genomics UK (COG-UK) Consortium |
| EPI_ISL_433990 | Department of Pathology, University of Cambridge | COVID-19 Genomics UK (COG-UK) Consortium |
| EPI_ISL_433991 | Department of Pathology, University of Cambridge | COVID-19 Genomics UK (COG-UK) Consortium |
| EPI_ISL_433992 | Department of Pathology, University of Cambridge | COVID-19 Genomics UK (COG-UK) Consortium |
| EPI_ISL_433993 | Department of Pathology, University of Cambridge | COVID-19 Genomics UK (COG-UK) Consortium |

| EPI_Accession_ID | Originating Laboratory | Submitting Laboratory |
|---|---|---|
| EPI_ISL_433994 | Department of Pathology, University of Cambridge | COVID-19 Genomics UK (COG-UK) Consortium |
| EPI_ISL_433995 | Department of Pathology, University of Cambridge | COVID-19 Genomics UK (COG-UK) Consortium |
| EPI_ISL_433996 | Department of Pathology, University of Cambridge | COVID-19 Genomics UK (COG-UK) Consortium |
| EPI_ISL_433997 | Department of Pathology, University of Cambridge | COVID-19 Genomics UK (COG-UK) Consortium |
| EPI_ISL_433999 | Department of Pathology, University of Cambridge | COVID-19 Genomics UK (COG-UK) Consortium |
| EPI_ISL_434000 | Department of Pathology, University of Cambridge | COVID-19 Genomics UK (COG-UK) Consortium |
| EPI_ISL_434001 | Department of Pathology, University of Cambridge | COVID-19 Genomics UK (COG-UK) Consortium |
| EPI_ISL_434002 | Department of Pathology, University of Cambridge | COVID-19 Genomics UK (COG-UK) Consortium |
| EPI_ISL_434003 | Department of Pathology, University of Cambridge | COVID-19 Genomics UK (COG-UK) Consortium |
| EPI_ISL_434004 | Department of Pathology, University of Cambridge | COVID-19 Genomics UK (COG-UK) Consortium |
| EPI_ISL_434005 | Department of Pathology, University of Cambridge | COVID-19 Genomics UK (COG-UK) Consortium |
| EPI_ISL_434006 | Department of Pathology, University of Cambridge | COVID-19 Genomics UK (COG-UK) Consortium |
| EPI_ISL_434008 | Department of Pathology, University of Cambridge | COVID-19 Genomics UK (COG-UK) Consortium |
| EPI_ISL_434009 | Department of Pathology, University of Cambridge | COVID-19 Genomics UK (COG-UK) Consortium |
| EPI_ISL_434010 | Department of Pathology, University of Cambridge | COVID-19 Genomics UK (COG-UK) Consortium |
| EPI_ISL_434011 | Department of Pathology, University of Cambridge | COVID-19 Genomics UK (COG-UK) Consortium |
| EPI_ISL_434012 | Department of Pathology, University of Cambridge | COVID-19 Genomics UK (COG-UK) Consortium |
| EPI_ISL_434013 | Department of Pathology, University of Cambridge | COVID-19 Genomics UK (COG-UK) Consortium |
| EPI_ISL_434014 | Department of Pathology, University of Cambridge | COVID-19 Genomics UK (COG-UK) Consortium |
| EPI_ISL_434015 | Department of Pathology, University of Cambridge | COVID-19 Genomics UK (COG-UK) Consortium |
| EPI_ISL_434016 | Department of Pathology, University of Cambridge | COVID-19 Genomics UK (COG-UK) Consortium |
| EPI_ISL_434017 | Department of Pathology, University of Cambridge | COVID-19 Genomics UK (COG-UK) Consortium |
| EPI_ISL_434018 | Department of Pathology, University of Cambridge | COVID-19 Genomics UK (COG-UK) Consortium |
| EPI_ISL_434019 | Department of Pathology, University of Cambridge | COVID-19 Genomics UK (COG-UK) Consortium |
| EPI_ISL_434020 | Department of Pathology, University of Cambridge | COVID-19 Genomics UK (COG-UK) Consortium |
| EPI_ISL_434021 | Department of Pathology, University of Cambridge | COVID-19 Genomics UK (COG-UK) Consortium |
| EPI_ISL_434022 | Department of Pathology, University of Cambridge | COVID-19 Genomics UK (COG-UK) Consortium |
| EPI_ISL_434023 | Department of Pathology, University of Cambridge | COVID-19 Genomics UK (COG-UK) Consortium |
| EPI_ISL_434024 | Department of Pathology, University of Cambridge | COVID-19 Genomics UK (COG-UK) Consortium |
| EPI_ISL_434025 | Department of Pathology, University of Cambridge | COVID-19 Genomics UK (COG-UK) Consortium |
| EPI_ISL_434026 | Department of Pathology, University of Cambridge | COVID-19 Genomics UK (COG-UK) Consortium |
| EPI_ISL_434027 | Department of Pathology, University of Cambridge | COVID-19 Genomics UK (COG-UK) Consortium |
| EPI_ISL_434028 | Department of Pathology, University of Cambridge | COVID-19 Genomics UK (COG-UK) Consortium |

| EPI_Accession_ID | Originating Laboratory | Submitting Laboratory |
|---|---|---|
| EPI_ISL_434029 | Department of Pathology, University of Cambridge | COVID-19 Genomics UK (COG-UK) Consortium |
| EPI_ISL_434030 | Department of Pathology, University of Cambridge | COVID-19 Genomics UK (COG-UK) Consortium |
| EPI_ISL_434031 | Department of Pathology, University of Cambridge | COVID-19 Genomics UK (COG-UK) Consortium |
| EPI_ISL_434032 | Department of Pathology, University of Cambridge | COVID-19 Genomics UK (COG-UK) Consortium |
| EPI_ISL_434033 | Department of Pathology, University of Cambridge | COVID-19 Genomics UK (COG-UK) Consortium |
| EPI_ISL_434034 | Department of Pathology, University of Cambridge | COVID-19 Genomics UK (COG-UK) Consortium |
| EPI_ISL_434035 | Department of Pathology, University of Cambridge | COVID-19 Genomics UK (COG-UK) Consortium |
| EPI_ISL_434036 | Department of Pathology, University of Cambridge | COVID-19 Genomics UK (COG-UK) Consortium |
| EPI_ISL_434037 | Department of Pathology, University of Cambridge | COVID-19 Genomics UK (COG-UK) Consortium |
| EPI_ISL_434038 | Department of Pathology, University of Cambridge | COVID-19 Genomics UK (COG-UK) Consortium |
| EPI_ISL_434039 | Department of Pathology, University of Cambridge | COVID-19 Genomics UK (COG-UK) Consortium |
| EPI_ISL_434040 | Department of Pathology, University of Cambridge | COVID-19 Genomics UK (COG-UK) Consortium |
| EPI_ISL_434041 | Department of Pathology, University of Cambridge | COVID-19 Genomics UK (COG-UK) Consortium |
| EPI_ISL_434042 | Department of Pathology, University of Cambridge | COVID-19 Genomics UK (COG-UK) Consortium |
| EPI_ISL_434043 | Department of Pathology, University of Cambridge | COVID-19 Genomics UK (COG-UK) Consortium |
| EPI_ISL_434044 | Department of Pathology, University of Cambridge | COVID-19 Genomics UK (COG-UK) Consortium |
| EPI_ISL_434045 | Department of Pathology, University of Cambridge | COVID-19 Genomics UK (COG-UK) Consortium |
| EPI_ISL_434046 | Department of Pathology, University of Cambridge | COVID-19 Genomics UK (COG-UK) Consortium |
| EPI_ISL_434047 | Department of Pathology, University of Cambridge | COVID-19 Genomics UK (COG-UK) Consortium |
| EPI_ISL_434048 | Department of Pathology, University of Cambridge | COVID-19 Genomics UK (COG-UK) Consortium |
| EPI_ISL_434049 | Department of Pathology, University of Cambridge | COVID-19 Genomics UK (COG-UK) Consortium |
| EPI_ISL_434050 | Department of Pathology, University of Cambridge | COVID-19 Genomics UK (COG-UK) Consortium |
| EPI_ISL_434051 | Department of Pathology, University of Cambridge | COVID-19 Genomics UK (COG-UK) Consortium |
| EPI_ISL_434052 | Department of Pathology, University of Cambridge | COVID-19 Genomics UK (COG-UK) Consortium |
| EPI_ISL_434053 | Department of Pathology, University of Cambridge | COVID-19 Genomics UK (COG-UK) Consortium |
| EPI_ISL_434054 | Department of Pathology, University of Cambridge | COVID-19 Genomics UK (COG-UK) Consortium |
| EPI_ISL_434055 | Department of Pathology, University of Cambridge | COVID-19 Genomics UK (COG-UK) Consortium |
| EPI_ISL_434056 | Department of Pathology, University of Cambridge | COVID-19 Genomics UK (COG-UK) Consortium |
| EPI_ISL_434057 | Department of Pathology, University of Cambridge | COVID-19 Genomics UK (COG-UK) Consortium |
| EPI_ISL_434058 | Department of Pathology, University of Cambridge | COVID-19 Genomics UK (COG-UK) Consortium |
| EPI_ISL_434059 | Department of Pathology, University of Cambridge | COVID-19 Genomics UK (COG-UK) Consortium |
| EPI_ISL_434060 | Department of Pathology, University of Cambridge | COVID-19 Genomics UK (COG-UK) Consortium |
| EPI_ISL_434061 | Department of Pathology, University of Cambridge | COVID-19 Genomics UK (COG-UK) Consortium |

| EPI_Accession_ID | Originating Laboratory | Submitting Laboratory |
|---|---|---|
| EPI_ISL_434062 | Department of Pathology, University of Cambridge | COVID-19 Genomics UK (COG-UK) Consortium |
| EPI_ISL_434063 | Washington State Department of Health | Seattle Flu Study |
| EPI_ISL_434064 | Washington State Department of Health | Seattle Flu Study |
| EPI_ISL_434065 | Washington State Department of Health | Seattle Flu Study |
| EPI_ISL_434066 | Washington State Department of Health | Seattle Flu Study |
| EPI_ISL_434067 | Washington State Department of Health | Seattle Flu Study |
| EPI_ISL_434068 | Washington State Department of Health | Seattle Flu Study |
| EPI_ISL_434069 | Washington State Department of Health | Seattle Flu Study |
| EPI_ISL_434070 | Washington State Department of Health | Seattle Flu Study |
| EPI_ISL_434071 | Washington State Department of Health | Seattle Flu Study |
| EPI_ISL_434072 | Washington State Department of Health | Seattle Flu Study |
| EPI_ISL_434073 | Washington State Department of Health | Seattle Flu Study |
| EPI_ISL_434074 | Washington State Department of Health | Seattle Flu Study |
| EPI_ISL_434075 | Washington State Department of Health | Seattle Flu Study |
| EPI_ISL_434076 | Washington State Department of Health | Seattle Flu Study |
| EPI_ISL_434077 | Washington State Department of Health | Seattle Flu Study |
| EPI_ISL_434078 | Washington State Department of Health | Seattle Flu Study |
| EPI_ISL_434079 | Washington State Department of Health | Seattle Flu Study |
| EPI_ISL_434080 | Washington State Department of Health | Seattle Flu Study |
| EPI_ISL_434081 | Washington State Department of Health | Seattle Flu Study |
| EPI_ISL_434082 | Washington State Department of Health | Seattle Flu Study |
| EPI_ISL_434083 | Washington State Department of Health | Seattle Flu Study |
| EPI_ISL_434084 | Washington State Department of Health | Seattle Flu Study |
| EPI_ISL_434085 | Washington State Department of Health | Seattle Flu Study |
| EPI_ISL_434086 | Washington State Department of Health | Seattle Flu Study |
| EPI_ISL_434087 | Washington State Department of Health | Seattle Flu Study |
| EPI_ISL_434088 | Washington State Department of Health | Seattle Flu Study |
| EPI_ISL_434089 | Washington State Department of Health | Seattle Flu Study |
| EPI_ISL_434090 | Washington State Department of Health | Seattle Flu Study |
| EPI_ISL_434091 | Washington State Department of Health | Seattle Flu Study |
| EPI_ISL_434092 | Washington State Department of Health | Seattle Flu Study |
| EPI_ISL_434093 | Washington State Department of Health | Seattle Flu Study |
| EPI_ISL_434094 | Washington State Department of Health | Seattle Flu Study |

| EPI_Accession_ID | Originating Laboratory | Submitting Laboratory |
|---|---|---|
| EPI_ISL_434095 | Washington State Department of Health | Seattle Flu Study |
| EPI_ISL_434096 | Washington State Department of Health | Seattle Flu Study |
| EPI_ISL_434097 | Washington State Department of Health | Seattle Flu Study |
| EPI_ISL_434098 | Washington State Department of Health | Seattle Flu Study |
| EPI_ISL_434099 | Washington State Department of Health | Seattle Flu Study |
| EPI_ISL_434100 | Washington State Department of Health | Seattle Flu Study |
| EPI_ISL_434101 | Washington State Department of Health | Seattle Flu Study |
| EPI_ISL_434102 | Washington State Department of Health | Seattle Flu Study |
| EPI_ISL_434103 | Washington State Department of Health | Seattle Flu Study |
| EPI_ISL_434104 | Washington State Department of Health | Seattle Flu Study |
| EPI_ISL_434105 | Washington State Department of Health | Seattle Flu Study |
| EPI_ISL_434106 | Washington State Department of Health | Seattle Flu Study |
| EPI_ISL_434107 | Washington State Department of Health | Seattle Flu Study |
| EPI_ISL_434108 | Washington State Department of Health | Seattle Flu Study |
| EPI_ISL_434109 | Washington State Department of Health | Seattle Flu Study |
| EPI_ISL_434110 | Washington State Department of Health | Seattle Flu Study |
| EPI_ISL_434111 | Washington State Department of Health | Seattle Flu Study |
| EPI_ISL_434112 | Washington State Department of Health | Seattle Flu Study |
| EPI_ISL_434113 | Washington State Department of Health | Seattle Flu Study |
| EPI_ISL_434114 | Washington State Department of Health | Seattle Flu Study |
| EPI_ISL_434115 | Washington State Department of Health | Seattle Flu Study |
| EPI_ISL_434116 | Washington State Department of Health | Seattle Flu Study |
| EPI_ISL_434117 | Washington State Department of Health | Seattle Flu Study |
| EPI_ISL_434118 | Washington State Department of Health | Seattle Flu Study |
| EPI_ISL_434119 | Washington State Department of Health | Seattle Flu Study |
| EPI_ISL_434120 | Washington State Department of Health | Seattle Flu Study |
| EPI_ISL_434121 | Washington State Department of Health | Seattle Flu Study |
| EPI_ISL_434122 | Washington State Department of Health | Seattle Flu Study |
| EPI_ISL_434123 | Washington State Department of Health | Seattle Flu Study |
| EPI_ISL_434124 | Washington State Department of Health | Seattle Flu Study |
| EPI_ISL_434125 | Washington State Department of Health | Seattle Flu Study |
| EPI_ISL_434126 | Washington State Department of Health | Seattle Flu Study |
| EPI_ISL_434127 | Washington State Department of Health | Seattle Flu Study |

| EPI_Accession_ID | Originating Laboratory | Submitting Laboratory |
|---|---|---|
| EPI_ISL_434128 | Washington State Department of Health | Seattle Flu Study |
| EPI_ISL_434129 | Washington State Department of Health | Seattle Flu Study |
| EPI_ISL_434130 | Washington State Department of Health | Seattle Flu Study |
| EPI_ISL_434131 | Washington State Department of Health | Seattle Flu Study |
| EPI_ISL_434132 | Washington State Department of Health | Seattle Flu Study |
| EPI_ISL_434133 | Washington State Department of Health | Seattle Flu Study |
| EPI_ISL_434134 | Washington State Department of Health | Seattle Flu Study |
| EPI_ISL_434135 | Washington State Department of Health | Seattle Flu Study |
| EPI_ISL_434136 | Washington State Department of Health | Seattle Flu Study |
| EPI_ISL_434137 | Washington State Department of Health | Seattle Flu Study |
| EPI_ISL_434138 | Washington State Department of Health | Seattle Flu Study |
| EPI_ISL_434139 | Washington State Department of Health | Seattle Flu Study |
| EPI_ISL_434140 | Washington State Department of Health | Seattle Flu Study |
| EPI_ISL_434141 | Washington State Department of Health | Seattle Flu Study |
| EPI_ISL_434142 | Washington State Department of Health | Seattle Flu Study |
| EPI_ISL_434143 | Washington State Department of Health | Seattle Flu Study |
| EPI_ISL_434144 | Washington State Department of Health | Seattle Flu Study |
| EPI_ISL_434145 | Washington State Department of Health | Seattle Flu Study |
| EPI_ISL_434146 | Washington State Department of Health | Seattle Flu Study |
| EPI_ISL_434147 | Washington State Department of Health | Seattle Flu Study |
| EPI_ISL_434148 | Washington State Department of Health | Seattle Flu Study |
| EPI_ISL_434149 | Washington State Department of Health | Seattle Flu Study |
| EPI_ISL_434150 | Washington State Department of Health | Seattle Flu Study |
| EPI_ISL_434151 | Washington State Department of Health | Seattle Flu Study |
| EPI_ISL_434152 | Washington State Department of Health | Seattle Flu Study |
| EPI_ISL_434153 | Washington State Department of Health | Seattle Flu Study |
| EPI_ISL_434154 | Washington State Department of Health | Seattle Flu Study |
| EPI_ISL_434155 | Washington State Department of Health | Seattle Flu Study |
| EPI_ISL_434156 | Washington State Department of Health | Seattle Flu Study |
| EPI_ISL_434157 | Washington State Department of Health | Seattle Flu Study |
| EPI_ISL_434158 | Washington State Department of Health | Seattle Flu Study |
| EPI_ISL_434159 | Washington State Department of Health | Seattle Flu Study |
| EPI_ISL_434160 | Washington State Department of Health | Seattle Flu Study |

| EPI_Accession_ID | Originating Laboratory | Submitting Laboratory |
|---|---|---|
| EPI_ISL_434161 | Washington State Department of Health | Seattle Flu Study |
| EPI_ISL_434162 | Washington State Department of Health | Seattle Flu Study |
| EPI_ISL_434163 | Washington State Department of Health | Seattle Flu Study |
| EPI_ISL_434164 | Washington State Department of Health | Seattle Flu Study |
| EPI_ISL_434165 | Washington State Department of Health | Seattle Flu Study |
| EPI_ISL_434166 | Washington State Department of Health | Seattle Flu Study |
| EPI_ISL_434167 | Washington State Department of Health | Seattle Flu Study |
| EPI_ISL_434168 | Washington State Department of Health | Seattle Flu Study |
| EPI_ISL_434169 | Washington State Department of Health | Seattle Flu Study |
| EPI_ISL_434170 | Washington State Department of Health | Seattle Flu Study |
| EPI_ISL_434171 | Washington State Department of Health | Seattle Flu Study |
| EPI_ISL_434172 | Washington State Department of Health | Seattle Flu Study |
| EPI_ISL_434173 | Washington State Department of Health | Seattle Flu Study |
| EPI_ISL_434174 | Washington State Department of Health | Seattle Flu Study |
| EPI_ISL_434175 | Washington State Department of Health | Seattle Flu Study |
| EPI_ISL_434176 | Washington State Department of Health | Seattle Flu Study |
| EPI_ISL_434177 | Washington State Department of Health | Seattle Flu Study |
| EPI_ISL_434178 | Washington State Department of Health | Seattle Flu Study |
| EPI_ISL_434179 | Washington State Department of Health | Seattle Flu Study |
| EPI_ISL_434180 | Washington State Department of Health | Seattle Flu Study |
| EPI_ISL_434181 | Washington State Department of Health | Seattle Flu Study |
| EPI_ISL_434182 | Washington State Department of Health | Seattle Flu Study |
| EPI_ISL_434183 | Washington State Department of Health | Seattle Flu Study |
| EPI_ISL_434184 | Washington State Department of Health | Seattle Flu Study |
| EPI_ISL_434185 | Washington State Department of Health | Seattle Flu Study |
| EPI_ISL_434186 | Washington State Department of Health | Seattle Flu Study |
| EPI_ISL_434187 | Washington State Department of Health | Seattle Flu Study |
| EPI_ISL_434188 | Washington State Department of Health | Seattle Flu Study |
| EPI_ISL_434189 | Washington State Department of Health | Seattle Flu Study |
| EPI_ISL_434190 | Washington State Department of Health | Seattle Flu Study |
| EPI_ISL_434191 | Washington State Department of Health | Seattle Flu Study |
| EPI_ISL_434192 | Washington State Department of Health | Seattle Flu Study |
| EPI_ISL_434193 | Washington State Department of Health | Seattle Flu Study |

| EPI_Accession_ID | Originating Laboratory | Submitting Laboratory |
|---|---|---|
| EPI_ISL_434194 | Washington State Department of Health | Seattle Flu Study |
| EPI_ISL_434195 | Washington State Department of Health | Seattle Flu Study |
| EPI_ISL_434196 | Washington State Department of Health | Seattle Flu Study |
| EPI_ISL_434197 | Washington State Department of Health | Seattle Flu Study |
| EPI_ISL_434198 | Washington State Department of Health | Seattle Flu Study |
| EPI_ISL_434199 | Washington State Department of Health | Seattle Flu Study |
| EPI_ISL_434200 | Washington State Department of Health | Seattle Flu Study |
| EPI_ISL_434201 | Washington State Department of Health | Seattle Flu Study |
| EPI_ISL_434202 | Washington State Department of Health | Seattle Flu Study |
| EPI_ISL_434203 | Washington State Department of Health | Seattle Flu Study |
| EPI_ISL_434204 | Washington State Department of Health | Seattle Flu Study |
| EPI_ISL_434205 | Washington State Department of Health | Seattle Flu Study |
| EPI_ISL_434206 | Washington State Department of Health | Seattle Flu Study |
| EPI_ISL_434207 | Washington State Department of Health | Seattle Flu Study |
| EPI_ISL_434208 | Washington State Department of Health | Seattle Flu Study |
| EPI_ISL_434209 | Washington State Department of Health | Seattle Flu Study |
| EPI_ISL_434210 | Washington State Department of Health | Seattle Flu Study |
| EPI_ISL_434211 | Washington State Department of Health | Seattle Flu Study |
| EPI_ISL_434212 | Washington State Department of Health | Seattle Flu Study |
| EPI_ISL_434213 | Washington State Department of Health | Seattle Flu Study |
| EPI_ISL_434214 | Washington State Department of Health | Seattle Flu Study |
| EPI_ISL_434215 | Washington State Department of Health | Seattle Flu Study |
| EPI_ISL_434216 | Washington State Department of Health | Seattle Flu Study |
| EPI_ISL_434217 | Washington State Department of Health | Seattle Flu Study |
| EPI_ISL_434218 | Washington State Department of Health | Seattle Flu Study |
| EPI_ISL_434219 | Washington State Department of Health | Seattle Flu Study |
| EPI_ISL_434220 | Washington State Department of Health | Seattle Flu Study |
| EPI_ISL_434221 | Washington State Department of Health | Seattle Flu Study |
| EPI_ISL_434222 | Washington State Department of Health | Seattle Flu Study |
| EPI_ISL_434223 | Washington State Department of Health | Seattle Flu Study |
| EPI_ISL_434224 | Washington State Department of Health | Seattle Flu Study |
| EPI_ISL_434225 | Washington State Department of Health | Seattle Flu Study |
| EPI_ISL_434226 | Washington State Department of Health | Seattle Flu Study |

| EPI_Accession_ID | Originating Laboratory | Submitting Laboratory |
|---|---|---|
| EPI_ISL_434227 | Washington State Department of Health | Seattle Flu Study |
| EPI_ISL_434228 | Washington State Department of Health | Seattle Flu Study |
| EPI_ISL_434229 | Washington State Department of Health | Seattle Flu Study |
| EPI_ISL_434230 | Washington State Department of Health | Seattle Flu Study |
| EPI_ISL_434231 | Washington State Department of Health | Seattle Flu Study |
| EPI_ISL_434232 | Washington State Department of Health | Seattle Flu Study |
| EPI_ISL_434233 | Washington State Department of Health | Seattle Flu Study |
| EPI_ISL_434234 | Washington State Department of Health | Seattle Flu Study |
| EPI_ISL_434235 | Washington State Department of Health | Seattle Flu Study |
| EPI_ISL_434236 | Washington State Department of Health | Seattle Flu Study |
| EPI_ISL_434237 | Washington State Department of Health | Seattle Flu Study |
| EPI_ISL_434238 | Washington State Department of Health | Seattle Flu Study |
| EPI_ISL_434239 | Washington State Department of Health | Seattle Flu Study |
| EPI_ISL_434240 | Washington State Department of Health | Seattle Flu Study |
| EPI_ISL_434241 | Washington State Department of Health | Seattle Flu Study |
| EPI_ISL_434242 | Washington State Department of Health | Seattle Flu Study |
| EPI_ISL_434243 | Washington State Department of Health | Seattle Flu Study |
| EPI_ISL_434244 | Washington State Department of Health | Seattle Flu Study |
| EPI_ISL_434245 | Washington State Department of Health | Seattle Flu Study |
| EPI_ISL_434246 | Washington State Department of Health | Seattle Flu Study |
| EPI_ISL_434247 | Washington State Department of Health | Seattle Flu Study |
| EPI_ISL_434248 | Washington State Department of Health | Seattle Flu Study |
| EPI_ISL_434249 | Washington State Department of Health | Seattle Flu Study |
| EPI_ISL_434250 | Washington State Department of Health | Seattle Flu Study |
| EPI_ISL_434251 | Washington State Department of Health | Seattle Flu Study |
| EPI_ISL_434252 | Washington State Department of Health | Seattle Flu Study |
| EPI_ISL_434253 | Washington State Department of Health | Seattle Flu Study |
| EPI_ISL_434254 | Washington State Department of Health | Seattle Flu Study |
| EPI_ISL_434255 | Washington State Department of Health | Seattle Flu Study |
| EPI_ISL_434256 | Washington State Department of Health | Seattle Flu Study |
| EPI_ISL_434257 | Washington State Department of Health | Seattle Flu Study |
| EPI_ISL_434258 | Washington State Department of Health | Seattle Flu Study |
| EPI_ISL_434259 | Washington State Department of Health | Seattle Flu Study |

| EPI_Accession_ID | Originating Laboratory | Submitting Laboratory |
|---|---|---|
| EPI_ISL_434260 | Washington State Department of Health | Seattle Flu Study |
| EPI_ISL_434261 | Washington State Department of Health | Seattle Flu Study |
| EPI_ISL_434262 | Washington State Department of Health | Seattle Flu Study |
| EPI_ISL_434263 | Washington State Department of Health | Seattle Flu Study |
| EPI_ISL_434264 | Washington State Department of Health | Seattle Flu Study |
| EPI_ISL_434265 | Washington State Department of Health | Seattle Flu Study |
| EPI_ISL_434266 | Washington State Department of Health | Seattle Flu Study |
| EPI_ISL_434267 | Washington State Department of Health | Seattle Flu Study |
| EPI_ISL_434268 | Washington State Department of Health | Seattle Flu Study |
| EPI_ISL_434269 | Washington State Department of Health | Seattle Flu Study |
| EPI_ISL_434270 | Washington State Department of Health | Seattle Flu Study |
| EPI_ISL_434271 | Washington State Department of Health | Seattle Flu Study |
| EPI_ISL_434272 | Washington State Department of Health | Seattle Flu Study |
| EPI_ISL_434273 | Washington State Department of Health | Seattle Flu Study |
| EPI_ISL_434274 | Washington State Department of Health | Seattle Flu Study |
| EPI_ISL_434275 | Washington State Department of Health | Seattle Flu Study |
| EPI_ISL_434276 | Washington State Department of Health | Seattle Flu Study |
| EPI_ISL_434277 | Washington State Department of Health | Seattle Flu Study |
| EPI_ISL_434278 | Washington State Department of Health | Seattle Flu Study |
| EPI_ISL_434279 | Washington State Department of Health | Seattle Flu Study |
| EPI_ISL_434280 | Washington State Department of Health | Seattle Flu Study |
| EPI_ISL_434281 | Washington State Department of Health | Seattle Flu Study |
| EPI_ISL_434282 | Washington State Department of Health | Seattle Flu Study |
| EPI_ISL_434283 | Washington State Department of Health | Seattle Flu Study |
| EPI_ISL_434284 | Washington State Department of Health | Seattle Flu Study |
| EPI_ISL_434285 | Washington State Department of Health | Seattle Flu Study |
| EPI_ISL_434286 | Washington State Department of Health | Seattle Flu Study |
| EPI_ISL_434287 | Washington State Department of Health | Seattle Flu Study |
| EPI_ISL_434288 | Washington State Department of Health | Seattle Flu Study |
| EPI_ISL_434289 | Washington State Department of Health | Seattle Flu Study |
| EPI_ISL_434290 | Washington State Department of Health | Seattle Flu Study |
| EPI_ISL_434291 | Washington State Department of Health | Seattle Flu Study |
| EPI_ISL_434292 | Washington State Department of Health | Seattle Flu Study |

| EPI_Accession_ID | Originating Laboratory | Submitting Laboratory |
|---|---|---|
| EPI_ISL_434293 | Washington State Department of Health | Seattle Flu Study |
| EPI_ISL_434294 | Washington State Department of Health | Seattle Flu Study |
| EPI_ISL_434295 | Washington State Department of Health | Seattle Flu Study |
| EPI_ISL_434296 | Washington State Department of Health | Seattle Flu Study |
| EPI_ISL_434297 | Washington State Department of Health | Seattle Flu Study |
| EPI_ISL_434298 | Washington State Department of Health | Seattle Flu Study |
| EPI_ISL_434299 | Washington State Department of Health | Seattle Flu Study |
| EPI_ISL_434300 | Washington State Department of Health | Seattle Flu Study |
| EPI_ISL_434301 | Washington State Department of Health | Seattle Flu Study |
| EPI_ISL_434302 | Washington State Department of Health | Seattle Flu Study |
| EPI_ISL_434303 | Washington State Department of Health | Seattle Flu Study |
| EPI_ISL_434304 | Washington State Department of Health | Seattle Flu Study |
| EPI_ISL_434305 | Washington State Department of Health | Seattle Flu Study |
| EPI_ISL_434306 | Washington State Department of Health | Seattle Flu Study |
| EPI_ISL_434307 | Washington State Department of Health | Seattle Flu Study |
| EPI_ISL_434308 | Washington State Department of Health | Seattle Flu Study |
| EPI_ISL_434309 | Washington State Department of Health | Seattle Flu Study |
| EPI_ISL_434310 | Washington State Department of Health | Seattle Flu Study |
| EPI_ISL_434311 | Washington State Department of Health | Seattle Flu Study |
| EPI_ISL_434312 | Washington State Department of Health | Seattle Flu Study |
| EPI_ISL_434313 | Washington State Department of Health | Seattle Flu Study |
| EPI_ISL_434314 | Washington State Department of Health | Seattle Flu Study |
| EPI_ISL_434315 | Washington State Department of Health | Seattle Flu Study |
| EPI_ISL_434316 | Washington State Department of Health | Seattle Flu Study |
| EPI_ISL_434317 | Washington State Department of Health | Seattle Flu Study |
| EPI_ISL_434318 | Washington State Department of Health | Seattle Flu Study |
| EPI_ISL_434319 | Washington State Department of Health | Seattle Flu Study |
| EPI_ISL_434320 | Washington State Department of Health | Seattle Flu Study |
| EPI_ISL_434321 | Washington State Department of Health | Seattle Flu Study |
| EPI_ISL_434322 | Washington State Department of Health | Seattle Flu Study |
| EPI_ISL_434323 | Washington State Department of Health | Seattle Flu Study |
| EPI_ISL_434324 | Washington State Department of Health | Seattle Flu Study |
| EPI_ISL_434325 | Washington State Department of Health | Seattle Flu Study |

| EPI_Accession_ID | Originating Laboratory | Submitting Laboratory |
|---|---|---|
| EPI_ISL_434326 | Washington State Department of Health | Seattle Flu Study |
| EPI_ISL_434327 | Washington State Department of Health | Seattle Flu Study |
| EPI_ISL_434328 | Washington State Department of Health | Seattle Flu Study |
| EPI_ISL_434329 | Washington State Department of Health | Seattle Flu Study |
| EPI_ISL_434330 | Washington State Department of Health | Seattle Flu Study |
| EPI_ISL_434331 | Washington State Department of Health | Seattle Flu Study |
| EPI_ISL_434332 | Washington State Department of Health | Seattle Flu Study |
| EPI_ISL_434333 | Washington State Department of Health | Seattle Flu Study |
| EPI_ISL_434334 | Washington State Department of Health | Seattle Flu Study |
| EPI_ISL_434335 | Washington State Department of Health | Seattle Flu Study |
| EPI_ISL_434336 | Washington State Department of Health | Seattle Flu Study |
| EPI_ISL_434337 | Washington State Department of Health | Seattle Flu Study |
| EPI_ISL_434338 | Washington State Department of Health | Seattle Flu Study |
| EPI_ISL_434339 | Washington State Department of Health | Seattle Flu Study |
| EPI_ISL_434340 | Washington State Department of Health | Seattle Flu Study |
| EPI_ISL_434341 | Washington State Department of Health | Seattle Flu Study |
| EPI_ISL_434342 | Washington State Department of Health | Seattle Flu Study |
| EPI_ISL_434343 | Washington State Department of Health | Seattle Flu Study |
| EPI_ISL_434344 | Washington State Department of Health | Seattle Flu Study |
| EPI_ISL_434345 | Washington State Department of Health | Seattle Flu Study |
| EPI_ISL_434346 | Washington State Department of Health | Seattle Flu Study |
| EPI_ISL_434347 | Lab voor klinische biologie | Onderzoeksgroep Virologie |
| EPI_ISL_434348 | Lab voor klinische biologie | Onderzoeksgroep Virologie |
| EPI_ISL_434349 | Lab voor klinische biologie | Onderzoeksgroep Virologie |
| EPI_ISL_434350 | Lab voor klinische biologie | Onderzoeksgroep Virologie |
| EPI_ISL_434351 | Lab voor klinische biologie | Onderzoeksgroep Virologie |
| EPI_ISL_434352 | Lab voor klinische biologie | Onderzoeksgroep Virologie |
| EPI_ISL_434353 | Lab voor klinische biologie | Onderzoeksgroep Virologie |
| EPI_ISL_434354 | Lab voor klinische biologie | Onderzoeksgroep Virologie |
| EPI_ISL_434355 | Lab voor klinische biologie | Onderzoeksgroep Virologie |
| EPI_ISL_434356 | Lab voor klinische biologie | Onderzoeksgroep Virologie |
| EPI_ISL_434357 | Lab voor klinische biologie | Onderzoeksgroep Virologie |
| EPI_ISL_434358 | Lab voor klinische biologie | Onderzoeksgroep Virologie |

| EPI_Accession_ID | Originating Laboratory | Submitting Laboratory |
|---|---|---|
| EPI_ISL_434359 | Lab voor klinische biologie | Onderzoeksgroep Virologie |
| EPI_ISL_434360 | Lab voor klinische biologie | Onderzoeksgroep Virologie |
| EPI_ISL_434361 | Lab voor klinische biologie | Onderzoeksgroep Virologie |
| EPI_ISL_434362 | Lab voor klinische biologie | Onderzoeksgroep Virologie |
| EPI_ISL_434363 | Lab voor klinische biologie | Onderzoeksgroep Virologie |
| EPI_ISL_434364 | Lab voor klinische biologie | Onderzoeksgroep Virologie |
| EPI_ISL_434365 | Lab voor klinische biologie | Onderzoeksgroep Virologie |
| EPI_ISL_434366 | Hospital AZ Rivierenland | Institute of Tropical Medicine |
| EPI_ISL_434367 | Hospital AZ Rivierenland | Institute of Tropical Medicine |
| EPI_ISL_434368 | Hospital AZ Rivierenland | Institute of Tropical Medicine |
| EPI_ISL_434369 | Hospital AZ Rivierenland | Institute of Tropical Medicine |
| EPI_ISL_434370 | Hospital AZ Rivierenland | Institute of Tropical Medicine |
| EPI_ISL_434371 | Hospital AZ Rivierenland | Institute of Tropical Medicine |
| EPI_ISL_434372 | Hospital AZ Rivierenland | Institute of Tropical Medicine |
| EPI_ISL_434373 | Hospital AZ Rivierenland | Institute of Tropical Medicine |
| EPI_ISL_434374 | Hospital AZ Rivierenland | Institute of Tropical Medicine |
| EPI_ISL_434375 | Hospital AZ Rivierenland | Institute of Tropical Medicine |
| EPI_ISL_434376 | Hospital AZ Rivierenland | Institute of Tropical Medicine |
| EPI_ISL_434377 | Hospital AZ Rivierenland | Institute of Tropical Medicine |
| EPI_ISL_434378 | Hospital AZ Rivierenland | Institute of Tropical Medicine |
| EPI_ISL_434379 | Hospital AZ Rivierenland | Institute of Tropical Medicine |
| EPI_ISL_434380 | Hospital AZ Rivierenland | Institute of Tropical Medicine |
| EPI_ISL_434381 | Hospital AZ Rivierenland | Institute of Tropical Medicine |
| EPI_ISL_434382 | Hospital AZ Rivierenland | Institute of Tropical Medicine |
| EPI_ISL_434383 | Hospital AZ Rivierenland | Institute of Tropical Medicine |
| EPI_ISL_434384 | Hospital AZ Rivierenland | Institute of Tropical Medicine |
| EPI_ISL_434385 | Hospital AZ Rivierenland | Institute of Tropical Medicine |
| EPI_ISL_434386 | Hospital AZ Rivierenland | Institute of Tropical Medicine |
| EPI_ISL_434455 | Laboratory of Microbiology, Medical School, National and Kapodistrian University of Athens | Laboratory of Biology, Department of Medicine, Democritus University of Thrace |
| EPI_ISL_434456 | Laboratory of Microbiology, Medical School, National and Kapodistrian University of Athens | Laboratory of Biology, Department of Medicine, Democritus University of Thrace |

| EPI_Accession_ID | Originating Laboratory | Submitting Laboratory |
|---|---|---|
| EPI_ISL_434457 | Laboratory of Microbiology, Medical School, National and Kapodistrian University of Athens | Laboratory of Biology, Department of Medicine, Democritus University of Thrace |
| EPI_ISL_434458 | Laboratory of Microbiology, Medical School, National and Kapodistrian University of Athens | Laboratory of Biology, Department of Medicine, Democritus University of Thrace |
| EPI_ISL_434459 | Laboratory of Microbiology, Medical School, National and Kapodistrian University of Athens | Laboratory of Biology, Department of Medicine, Democritus University of Thrace |
| EPI_ISL_434460 | Laboratory of Microbiology, Medical School, National and Kapodistrian University of Athens | Laboratory of Biology, Department of Medicine, Democritus University of Thrace |
| EPI_ISL_434461 | Laboratory of Microbiology, Medical School, National and Kapodistrian University of Athens | Laboratory of Biology, Department of Medicine, Democritus University of Thrace |
| EPI_ISL_434462 | Laboratory of Microbiology, Medical School, National and Kapodistrian University of Athens | Laboratory of Biology, Department of Medicine, Democritus University of Thrace |
| EPI_ISL_434463 | Laboratory of Microbiology, Medical School, National and Kapodistrian University of Athens | Laboratory of Biology, Department of Medicine, Democritus University of Thrace |
| EPI_ISL_434464 | Laboratory of Microbiology, Medical School, National and Kapodistrian University of Athens | Laboratory of Biology, Department of Medicine, Democritus University of Thrace |
| EPI_ISL_434465 | Laboratory of Microbiology, Medical School, National and Kapodistrian University of Athens | Laboratory of Biology, Department of Medicine, Democritus University of Thrace |
| EPI_ISL_434466 | Laboratory of Microbiology, Medical School, National and Kapodistrian University of Athens | Laboratory of Biology, Department of Medicine, Democritus University of Thrace |
| EPI_ISL_434467 | Laboratory of Microbiology, Medical School, National and Kapodistrian University of Athens | Laboratory of Biology, Department of Medicine, Democritus University of Thrace |
| EPI_ISL_434468 | Laboratory of Microbiology, Medical School, National and Kapodistrian University of Athens | Laboratory of Biology, Department of Medicine, Democritus University of Thrace |
| EPI_ISL_434469 | Laboratory of Microbiology, Medical School, National and Kapodistrian University of Athens | Laboratory of Biology, Department of Medicine, Democritus University of Thrace |
| EPI_ISL_434470 | Laboratory of Microbiology, Medical School, National and Kapodistrian University of Athens | Laboratory of Biology, Department of Medicine, Democritus University of Thrace |
| EPI_ISL_434471 | Laboratory of Microbiology, Medical School, National and Kapodistrian University of Athens | Laboratory of Biology, Department of Medicine, Democritus University of Thrace |
| EPI_ISL_434472 | Laboratory of Microbiology, Medical School, National and Kapodistrian University of Athens | Laboratory of Biology, Department of Medicine, Democritus University of Thrace |

| EPI_Accession_ID | Originating Laboratory | Submitting Laboratory |
|---|---|---|
| EPI_ISL_434473 | Laboratory of Microbiology, Medical School, National and Kapodistrian University of Athens | Laboratory of Biology, Department of Medicine, Democritus University of Thrace |
| EPI_ISL_434474 | Laboratory of Microbiology, Medical School, National and Kapodistrian University of Athens | Laboratory of Biology, Department of Medicine, Democritus University of Thrace |
| EPI_ISL_434475 | Laboratory of Microbiology, Medical School, National and Kapodistrian University of Athens | Laboratory of Biology, Department of Medicine, Democritus University of Thrace |
| EPI_ISL_434476 | Laboratory of Microbiology, Medical School, National and Kapodistrian University of Athens | Laboratory of Biology, Department of Medicine, Democritus University of Thrace |
| EPI_ISL_434477 | Laboratory of Microbiology, Medical School, National and Kapodistrian University of Athens | Laboratory of Biology, Department of Medicine, Democritus University of Thrace |
| EPI_ISL_434478 | Laboratory of Microbiology, Medical School, National and Kapodistrian University of Athens | Laboratory of Biology, Department of Medicine, Democritus University of Thrace |
| EPI_ISL_434479 | Laboratory of Microbiology, Medical School, National and Kapodistrian University of Athens | Laboratory of Biology, Department of Medicine, Democritus University of Thrace |
| EPI_ISL_434480 | Laboratory of Microbiology, Medical School, National and Kapodistrian University of Athens | Laboratory of Biology, Department of Medicine, Democritus University of Thrace |
| EPI_ISL_434481 | Laboratory of Microbiology, Medical School, National and Kapodistrian University of Athens | Laboratory of Biology, Department of Medicine, Democritus University of Thrace |
| EPI_ISL_434482 | Laboratory of Microbiology, Medical School, National and Kapodistrian University of Athens | Laboratory of Biology, Department of Medicine, Democritus University of Thrace |
| EPI_ISL_434483 | Laboratory of Microbiology, Medical School, National and Kapodistrian University of Athens | Laboratory of Biology, Department of Medicine, Democritus University of Thrace |
| EPI_ISL_434484 | Laboratory of Microbiology, Medical School, National and Kapodistrian University of Athens | Laboratory of Biology, Department of Medicine, Democritus University of Thrace |
| EPI_ISL_434485 | Laboratory of Microbiology, Medical School, National and Kapodistrian University of Athens | Laboratory of Biology, Department of Medicine, Democritus University of Thrace |
| EPI_ISL_434486 | Laboratory of Microbiology, Medical School, National and Kapodistrian University of Athens | Laboratory of Biology, Department of Medicine, Democritus University of Thrace |
| EPI_ISL_434487 | Laboratoire National de Sante, Microbiology, Virology | Laboratoire National de Sante, Microbiology, Epidemiology and Microbial Genomics |
| EPI_ISL_434488 | Laboratoire National de Sante, Microbiology, Virology | Laboratoire National de Sante, Microbiology, Epidemiology and Microbial Genomics |

| EPI_Accession_ID | Originating Laboratory | Submitting Laboratory |
| --- | --- | --- |
| EPI_ISL_434489 | Laboratoire National de Sante, Microbiology, Virology | Laboratoire National de Sante, Microbiology, Epidemiology and Microbial Genomics |
| EPI_ISL_434490 | Laboratoire National de Sante, Microbiology, Virology | Laboratoire National de Sante, Microbiology, Epidemiology and Microbial Genomics |
| EPI_ISL_434491 | Laboratoire National de Sante, Microbiology, Virology | Laboratoire National de Sante, Microbiology, Epidemiology and Microbial Genomics |
| EPI_ISL_434492 | Laboratoire National de Sante, Microbiology, Virology | Laboratoire National de Sante, Microbiology, Epidemiology and Microbial Genomics |
| EPI_ISL_434493 | Laboratoire National de Sante, Microbiology, Virology | Laboratoire National de Sante, Microbiology, Epidemiology and Microbial Genomics |
| EPI_ISL_434494 | Laboratoire National de Sante, Microbiology, Virology | Laboratoire National de Sante, Microbiology, Epidemiology and Microbial Genomics |
| EPI_ISL_434495 | Laboratoire National de Sante, Microbiology, Virology | Laboratoire National de Sante, Microbiology, Epidemiology and Microbial Genomics |
| EPI_ISL_434496 | Laboratoire National de Sante, Microbiology, Virology | Laboratoire National de Sante, Microbiology, Epidemiology and Microbial Genomics |
| EPI_ISL_434497 | Laboratoire National de Sante, Microbiology, Virology | Laboratoire National de Sante, Microbiology, Epidemiology and Microbial Genomics |
| EPI_ISL_434498 | Laboratoire National de Sante, Microbiology, Virology | Laboratoire National de Sante, Microbiology, Epidemiology and Microbial Genomics |
| EPI_ISL_434499 | Laboratoire National de Sante, Microbiology, Virology | Laboratoire National de Sante, Microbiology, Epidemiology and Microbial Genomics |
| EPI_ISL_434500 | Laboratoire National de Sante, Microbiology, Virology | Laboratoire National de Sante, Microbiology, Epidemiology and Microbial Genomics |
| EPI_ISL_434501 | Laboratoire National de Sante, Microbiology, Virology | Laboratoire National de Sante, Microbiology, Epidemiology and Microbial Genomics |
| EPI_ISL_434502 | Laboratoire National de Sante, Microbiology, Virology | Laboratoire National de Sante, Microbiology, Epidemiology and Microbial Genomics |
| EPI_ISL_434503 | Laboratoire National de Sante, Microbiology, Virology | Laboratoire National de Sante, Microbiology, Epidemiology and Microbial Genomics |
| EPI_ISL_434504 | Laboratoire National de Sante, Microbiology, Virology | Laboratoire National de Sante, Microbiology, Epidemiology and Microbial Genomics |

| EPI_Accession_ID | Originating Laboratory | Submitting Laboratory |
|---|---|---|
| EPI_ISL_434505 | Laboratoire National de Sante, Microbiology, Virology | Laboratoire National de Sante, Microbiology, Epidemiology and Microbial Genomics |
| EPI_ISL_434506 | Laboratoire National de Sante, Microbiology, Virology | Laboratoire National de Sante, Microbiology, Epidemiology and Microbial Genomics |
| EPI_ISL_434507 | Laboratoire National de Sante, Microbiology, Virology | Laboratoire National de Sante, Microbiology, Epidemiology and Microbial Genomics |
| EPI_ISL_434508 | Laboratoire National de Sante, Microbiology, Virology | Laboratoire National de Sante, Microbiology, Epidemiology and Microbial Genomics |
| EPI_ISL_434509 | Laboratoire National de Sante, Microbiology, Virology | Laboratoire National de Sante, Microbiology, Epidemiology and Microbial Genomics |
| EPI_ISL_434510 | Laboratoire National de Sante, Microbiology, Virology | Laboratoire National de Sante, Microbiology, Epidemiology and Microbial Genomics |
| EPI_ISL_434511 | Laboratoire National de Sante, Microbiology, Virology | Laboratoire National de Sante, Microbiology, Epidemiology and Microbial Genomics |
| EPI_ISL_434512 | Laboratoire National de Sante, Microbiology, Virology | Laboratoire National de Sante, Microbiology, Epidemiology and Microbial Genomics |
| EPI_ISL_434513 | Laboratoire National de Sante, Microbiology, Virology | Laboratoire National de Sante, Microbiology, Epidemiology and Microbial Genomics |
| EPI_ISL_434514 | Laboratoire National de Sante, Microbiology, Virology | Laboratoire National de Sante, Microbiology, Epidemiology and Microbial Genomics |
| EPI_ISL_434515 | Laboratoire National de Sante, Microbiology, Virology | Laboratoire National de Sante, Microbiology, Epidemiology and Microbial Genomics |
| EPI_ISL_434516 | Biolab Diagnostic Laboratories | Andersen lab at Scripps Research |
| EPI_ISL_434517 | Robert Garry lab | Andersen lab at Scripps Research |
| EPI_ISL_434518 | Robert Garry lab | Andersen lab at Scripps Research |
| EPI_ISL_434519 | Robert Garry lab | Andersen lab at Scripps Research |
| EPI_ISL_434520 | Robert Garry lab | Andersen lab at Scripps Research |
| EPI_ISL_434521 | Robert Garry lab | Andersen lab at Scripps Research |
| EPI_ISL_434522 | Robert Garry lab | Andersen lab at Scripps Research |
| EPI_ISL_434523 | Robert Garry lab | Andersen lab at Scripps Research |
| EPI_ISL_434524 | Robert Garry lab | Andersen lab at Scripps Research |
| EPI_ISL_434525 | Robert Garry lab | Andersen lab at Scripps Research |
| EPI_ISL_434526 | Robert Garry lab | Andersen lab at Scripps Research |

| EPI_Accession_ID | Originating Laboratory | Submitting Laboratory |
|---|---|---|
| EPI_ISL_434527 | Robert Garry lab | Andersen lab at Scripps Research |
| EPI_ISL_434528 | Robert Garry lab | Andersen lab at Scripps Research |
| EPI_ISL_434529 | Robert Garry lab | Andersen lab at Scripps Research |
| EPI_ISL_434530 | Robert Garry lab | Andersen lab at Scripps Research |
| EPI_ISL_434531 | Robert Garry lab | Andersen lab at Scripps Research |
| EPI_ISL_434532 | Robert Garry lab | Andersen lab at Scripps Research |
| EPI_ISL_434533 | Area de Salud Alajuela Sur | Inciensa, Instituto Costarricense de InvestigaciÃ³n y EnseÃ±anza en NutriciÃ³n y Salud |
| EPI_ISL_434534 | National Institute for Viral Disease Control and Prevention, China CDC | National Institute for Viral Disease Control and Prevention, China CDC, Yunnan Provincial CDC |
| EPI_ISL_434535 | Area de Salud Alajuela Sur | Inciensa, Instituto Costarricense de InvestigaciÃ³n y EnseÃ±anza en NutriciÃ³n y Salud |
| EPI_ISL_434536 | Hospital San Vicente de Paul | Inciensa, Instituto Costarricense de InvestigaciÃ³n y EnseÃ±anza en NutriciÃ³n y Salud |
| EPI_ISL_434538 | COOPESAIN | Inciensa, Instituto Costarricense de InvestigaciÃ³n y EnseÃ±anza en NutriciÃ³n y Salud |
| EPI_ISL_434539 | Area de Salud Orotina | Inciensa, Instituto Costarricense de InvestigaciÃ³n y EnseÃ±anza en NutriciÃ³n y Salud |
| EPI_ISL_434540 | EBAIS ConcepciÃ³n Norte | Inciensa, Instituto Costarricense de InvestigaciÃ³n y EnseÃ±anza en NutriciÃ³n y Salud |
| EPI_ISL_434541 | Puerto Rico Department of Health | Centers for Disease Control and Prevention, Dengue Branch |
| EPI_ISL_434542 | Puerto Rico Department of Health | Centers for Disease Control and Prevention, Dengue Branch |
| EPI_ISL_434543 | Puerto Rico Department of Health | Centers for Disease Control and Prevention, Dengue Branch |
| EPI_ISL_434544 | Puerto Rico Department of Health | Centers for Disease Control and Prevention, Dengue Branch |
| EPI_ISL_434545 | Puerto Rico Department of Health | Centers for Disease Control and Prevention, Dengue Branch |
| EPI_ISL_434546 | Puerto Rico Department of Health | Centers for Disease Control and Prevention, Dengue Branch |

| EPI_Accession_ID | Originating Laboratory | Submitting Laboratory |
|---|---|---|
| EPI_ISL_434547 | Puerto Rico Department of Health | Centers for Disease Control and Prevention, Dengue Branch |
| EPI_ISL_434548 | Puerto Rico Department of Health | Centers for Disease Control and Prevention, Dengue Branch |
| EPI_ISL_434549 | Puerto Rico Department of Health | Centers for Disease Control and Prevention, Dengue Branch |
| EPI_ISL_434550 | Puerto Rico Department of Health | Centers for Disease Control and Prevention, Dengue Branch |
| EPI_ISL_434551 | Puerto Rico Department of Health | Centers for Disease Control and Prevention, Dengue Branch |
| EPI_ISL_434552 | Puerto Rico Department of Health | Centers for Disease Control and Prevention, Dengue Branch |
| EPI_ISL_434553 | Puerto Rico Department of Health | Centers for Disease Control and Prevention, Dengue Branch |
| EPI_ISL_434554 | National Institutes of Health, University of the Philippines Manila | Philippine Genome Center |
| EPI_ISL_434555 | National Institutes of Health, University of the Philippines Manila | Philippine Genome Center |
| EPI_ISL_434556 | National Institutes of Health, University of the Philippines Manila | Philippine Genome Center |
| EPI_ISL_434557 | National Institutes of Health, University of the Philippines Manila | Philippine Genome Center |
| EPI_ISL_434558 | National Institutes of Health, University of the Philippines Manila | Philippine Genome Center |
| EPI_ISL_434563 | unknown | Microbiology |
| EPI_ISL_434564 | unknown | unknown |
| EPI_ISL_434565 | unknown | Microbiology |
| EPI_ISL_434566 | unknown | Microbiology |
| EPI_ISL_434567 | unknown | Microbiology |
| EPI_ISL_434568 | unknown | Microbiology |
| EPI_ISL_434569 | unknown | Microbiology |
| EPI_ISL_434570 | unknown | Microbiology |
| EPI_ISL_434571 | unknown | Microbiology |

| EPI_Accession_ID | Originating Laboratory | Submitting Laboratory |
|---|---|---|
| EPI_ISL_434572 | The National Institute of Public Health Center for Epidemiology and Microbiology | The National Institute of Public Health Center for Epidemiology and Microbiology |
| EPI_ISL_434586 | Johns Hopkins Hospital Department of Pathology | Johns Hopkins Hospital Department of Pathology |
| EPI_ISL_434587 | Johns Hopkins Hospital Department of Pathology | Johns Hopkins Hospital Department of Pathology |
| EPI_ISL_434588 | Johns Hopkins Hospital Department of Pathology | Johns Hopkins Hospital Department of Pathology |
| EPI_ISL_434590 | Virginia DCLS | Virginia DCLS |
| EPI_ISL_434591 | Virginia DCLS | Virginia DCLS |
| EPI_ISL_434592 | Virginia DCLS | Virginia DCLS |
| EPI_ISL_434593 | Virginia DCLS | Virginia DCLS |
| EPI_ISL_434594 | Virginia DCLS | Virginia DCLS |
| EPI_ISL_434595 | Virginia DCLS | Virginia DCLS |
| EPI_ISL_434596 | Virginia DCLS | Virginia DCLS |
| EPI_ISL_434597 | Virginia DCLS | Virginia DCLS |
| EPI_ISL_434598 | Virginia DCLS | Virginia DCLS |
| EPI_ISL_434599 | Virginia DCLS | Virginia DCLS |
| EPI_ISL_434600 | Virginia DCLS | Virginia DCLS |
| EPI_ISL_434601 | Virginia DCLS | Virginia DCLS |
| EPI_ISL_434602 | Virginia DCLS | Virginia DCLS |
| EPI_ISL_434603 | Virginia DCLS | Virginia DCLS |
| EPI_ISL_434604 | Virginia DCLS | Virginia DCLS |
| EPI_ISL_434605 | Virginia DCLS | Virginia DCLS |
| EPI_ISL_434606 | Virginia DCLS | Virginia DCLS |
| EPI_ISL_434607 | University of Wisconsin-Madison AIDS Vaccine Research Laboratories | University of Wisconsin-Madison AIDS Vaccine Research Laboratories |
| EPI_ISL_434608 | University of Wisconsin-Madison AIDS Vaccine Research Laboratories | University of Wisconsin-Madison AIDS Vaccine Research Laboratories |
| EPI_ISL_434609 | University of Wisconsin-Madison AIDS Vaccine Research Laboratories | University of Wisconsin-Madison AIDS Vaccine Research Laboratories |
| EPI_ISL_434610 | University of Wisconsin-Madison AIDS Vaccine Research Laboratories | University of Wisconsin-Madison AIDS Vaccine Research Laboratories |
| EPI_ISL_434611 | University of Wisconsin-Madison AIDS Vaccine Research Laboratories | University of Wisconsin-Madison AIDS Vaccine Research Laboratories |

| EPI_Accession_ID | Originating Laboratory | Submitting Laboratory |
|---|---|---|
| EPI_ISL_434612 | University of Wisconsin-Madison AIDS Vaccine Research Laboratories | University of Wisconsin-Madison AIDS Vaccine Research Laboratories |
| EPI_ISL_434613 | University of Wisconsin-Madison AIDS Vaccine Research Laboratories | University of Wisconsin-Madison AIDS Vaccine Research Laboratories |
| EPI_ISL_434614 | University of Wisconsin-Madison AIDS Vaccine Research Laboratories | University of Wisconsin-Madison AIDS Vaccine Research Laboratories |
| EPI_ISL_434615 | University of Wisconsin-Madison AIDS Vaccine Research Laboratories | University of Wisconsin-Madison AIDS Vaccine Research Laboratories |
| EPI_ISL_434626 | CHU Purpan - Laboratoire de Virologie - Institut FÃ©dÃ©ratif de Biologie | Laboratoire de virologie - Ã‰cole Nationale VÃ©tÃ©rinaire de Toulouse |
| EPI_ISL_434627 | CHU Purpan - Laboratoire de Virologie - Institut FÃ©dÃ©ratif de Biologie | Laboratoire de virologie - Ã‰cole Nationale VÃ©tÃ©rinaire de Toulouse |
| EPI_ISL_434628 | CHU Purpan - Laboratoire de Virologie - Institut FÃ©dÃ©ratif de Biologie | Laboratoire de virologie - Ã‰cole Nationale VÃ©tÃ©rinaire de Toulouse |
| EPI_ISL_434629 | CHU Purpan - Laboratoire de Virologie - Institut FÃ©dÃ©ratif de Biologie | Laboratoire de virologie - Ã‰cole Nationale VÃ©tÃ©rinaire de Toulouse |
| EPI_ISL_434630 | CHU Purpan - Laboratoire de Virologie - Institut FÃ©dÃ©ratif de Biologie | Laboratoire de virologie - Ã‰cole Nationale VÃ©tÃ©rinaire de Toulouse |
| EPI_ISL_434631 | CHU Purpan - Laboratoire de Virologie - Institut FÃ©dÃ©ratif de Biologie | Laboratoire de virologie - Ã‰cole Nationale VÃ©tÃ©rinaire de Toulouse |
| EPI_ISL_434632 | CHU Purpan - Laboratoire de Virologie - Institut FÃ©dÃ©ratif de Biologie | Laboratoire de virologie - Ã‰cole Nationale VÃ©tÃ©rinaire de Toulouse |
| EPI_ISL_434633 | CHU Purpan - Laboratoire de Virologie - Institut FÃ©dÃ©ratif de Biologie | Laboratoire de virologie - Ã‰cole Nationale VÃ©tÃ©rinaire de Toulouse |
| EPI_ISL_434634 | CHU Purpan - Laboratoire de Virologie - Institut FÃ©dÃ©ratif de Biologie | Laboratoire de virologie - Ã‰cole Nationale VÃ©tÃ©rinaire de Toulouse |
| EPI_ISL_434635 | CHU Purpan - Laboratoire de Virologie - Institut FÃ©dÃ©ratif de Biologie | Laboratoire de virologie - Ã‰cole Nationale VÃ©tÃ©rinaire de Toulouse |
| EPI_ISL_434636 | Lednicky Laboratory, Emerging Pathogens Institute, University of Florida | Lednicky Laboratory at Emerging Pathogens Institute, University of Florida |
| EPI_ISL_434637 | Lednicky Laboratory, Emerging Pathogens Institute, University of Florida. | Lednicky Laboratory, Emerging Pathogens Institute, University of Florida. |
| EPI_ISL_434638 | Johns Hopkins Hospital Department of Pathology | Johns Hopkins Hospital Department of Pathology |

| EPI_Accession_ID | Originating Laboratory | Submitting Laboratory |
|---|---|---|
| EPI_ISL_434639 | Johns Hopkins Hospital Department of Pathology | Johns Hopkins Hospital Department of Pathology |
| EPI_ISL_434640 | Johns Hopkins Hospital Department of Pathology | Johns Hopkins Hospital Department of Pathology |
| EPI_ISL_434641 | Laboratoriemedicin | The Public Health Agency of Sweden |
| EPI_ISL_434642 | Laboratoriemedicin | The Public Health Agency of Sweden |
| EPI_ISL_434643 | Uppsala Narakut Aleris | The Public Health Agency of Sweden |
| EPI_ISL_434644 | Kungsholmsdoktorn | The Public Health Agency of Sweden |
| EPI_ISL_434645 | Svardsjo VC | The Public Health Agency of Sweden |
| EPI_ISL_434646 | Ulltuna Vardcentral | The Public Health Agency of Sweden |
| EPI_ISL_434647 | Victoria Vard och Halsa | The Public Health Agency of Sweden |
| EPI_ISL_434648 | Victoria Vard och Halsa | The Public Health Agency of Sweden |
| EPI_ISL_434649 | Svardsjo VC | The Public Health Agency of Sweden |
| EPI_ISL_434650 | Follinge Halsocentral | The Public Health Agency of Sweden |
| EPI_ISL_434651 | Krokoms Halsocentral | The Public Health Agency of Sweden |
| EPI_ISL_434652 | Ektorps Vardcentral | The Public Health Agency of Sweden |
| EPI_ISL_434653 | Trollbackens VC | The Public Health Agency of Sweden |
| EPI_ISL_434654 | Saroledens Familjelakare | The Public Health Agency of Sweden |
| EPI_ISL_434655 | Uppsala Narakut Aleris | The Public Health Agency of Sweden |
| EPI_ISL_434656 | Trollbackens VC | The Public Health Agency of Sweden |
| EPI_ISL_434657 | Kristianstadkliniken | The Public Health Agency of Sweden |
| EPI_ISL_434658 | Lundens VC | The Public Health Agency of Sweden |
| EPI_ISL_434659 | Knivsta VC | The Public Health Agency of Sweden |
| EPI_ISL_434660 | Narhalsan Backa vardcentral | The Public Health Agency of Sweden |
| EPI_ISL_434661 | Omtanken Grimmered | The Public Health Agency of Sweden |
| EPI_ISL_434662 | Omtanken Grimmered | The Public Health Agency of Sweden |
| EPI_ISL_434663 | Surbrunns VC | The Public Health Agency of Sweden |
| EPI_ISL_434664 | Omtanken Grimmered | The Public Health Agency of Sweden |
| EPI_ISL_434665 | Uppsala Narakut Aleris | The Public Health Agency of Sweden |
| EPI_ISL_434666 | Hornefors Halsocentral | The Public Health Agency of Sweden |
| EPI_ISL_434667 | Ulltuna Vardcentral | The Public Health Agency of Sweden |
| EPI_ISL_434668 | Kungsors VC | The Public Health Agency of Sweden |
| EPI_ISL_434669 | Lakargruppen | The Public Health Agency of Sweden |
| EPI_ISL_434670 | Narhalsan Molnlycke, Barn och ungdomsmedicin | The Public Health Agency of Sweden |
| EPI_ISL_434671 | Narhalsan Sjobo vardcentral | The Public Health Agency of Sweden |

| EPI_Accession_ID | Originating Laboratory | Submitting Laboratory |
|---|---|---|
| EPI_ISL_434672 | Surbrunns VC | The Public Health Agency of Sweden |
| EPI_ISL_434673 | Omtanken Grimmered | The Public Health Agency of Sweden |
| EPI_ISL_434674 | Narhalsan Backa vardcentral | The Public Health Agency of Sweden |
| EPI_ISL_434675 | Surbrunns VC | The Public Health Agency of Sweden |
| EPI_ISL_434676 | Surbrunns VC | The Public Health Agency of Sweden |
| EPI_ISL_434677 | Lednicky Laboratory at Emerging Pathogens Institute | Lednicky Laboratory at Emerging Pathogens Institute |
| EPI_ISL_434678 | Viral Respiratory Lab, National Institute for Biomedical Research (INRB) | Pathogen Sequencing Lab, National Institute for Biomedical Research (INRB) |
| EPI_ISL_434679 | Viral Respiratory Lab, National Institute for Biomedical Research (INRB) | Pathogen Sequencing Lab, National Institute for Biomedical Research (INRB) |
| EPI_ISL_434680 | Viral Respiratory Lab, National Institute for Biomedical Research (INRB) | Pathogen Sequencing Lab, National Institute for Biomedical Research (INRB) |
| EPI_ISL_434681 | Viral Respiratory Lab, National Institute for Biomedical Research (INRB) | Pathogen Sequencing Lab, National Institute for Biomedical Research (INRB) |
| EPI_ISL_434682 | Johns Hopkins Hospital Department of Pathology | Johns Hopkins Hospital Department of Pathology |
| EPI_ISL_434683 | Johns Hopkins Hospital Department of Pathology | Johns Hopkins Hospital Department of Pathology |
| EPI_ISL_434684 | Johns Hopkins Hospital Department of Pathology | Johns Hopkins Hospital Department of Pathology |
| EPI_ISL_434685 | Johns Hopkins Hospital Department of Pathology | Johns Hopkins Hospital Department of Pathology |
| EPI_ISL_434686 | Johns Hopkins Hospital Department of Pathology | Johns Hopkins Hospital Department of Pathology |
| EPI_ISL_434687 | Johns Hopkins Hospital Department of Pathology | Johns Hopkins Hospital Department of Pathology |
| EPI_ISL_434688 | Johns Hopkins Hospital Department of Pathology | Johns Hopkins Hospital Department of Pathology |
| EPI_ISL_434689 | Johns Hopkins Hospital Department of Pathology | Johns Hopkins Hospital Department of Pathology |
| EPI_ISL_434690 | Johns Hopkins Hospital Department of Pathology | Johns Hopkins Hospital Department of Pathology |
| EPI_ISL_434691 | Johns Hopkins Hospital Department of Pathology | Johns Hopkins Hospital Department of Pathology |
| EPI_ISL_434692 | Bamrasnaradura hospital | National Institute of Health. Department of medical Sciences, Ministry of Public Health, Thailand |
| EPI_ISL_434693 | Bamrasnaradura hospital | National Institute of Health. Department of medical Sciences, Ministry of Public Health, Thailand |
| EPI_ISL_434694 | Bamrasnaradura hospital | National Institute of Health. Department of medical Sciences, Ministry of Public Health, Thailand |
| EPI_ISL_434695 | unknown | National Institute of Health. Department of medical Sciences, Ministry of Public Health, Thailand |

| EPI_Accession_ID | Originating Laboratory | Submitting Laboratory |
|---|---|---|
| EPI_ISL_434696 | Bamrasnaradura hospital | National Institute of Health. Department of medical Sciences, Ministry of Public Health, Thailand |
| EPI_ISL_434697 | unknown | National Institute of Health. Department of medical Sciences, Ministry of Public Health, Thailand |
| EPI_ISL_434698 | Praram 9 Hospital | National Institute of Health. Department of medical Sciences, Ministry of Public Health, Thailand |
| EPI_ISL_434699 | Praram 9 Hospital | National Institute of Health. Department of medical Sciences, Ministry of Public Health, Thailand |
| EPI_ISL_434700 | Ramkhamhaeng Hospital | National Institute of Health. Department of medical Sciences, Ministry of Public Health, Thailand |
| EPI_ISL_434701 | Panyanunthaphikkhu Chonprathan Medical Center (PCMC) | National Institute of Health. Department of medical Sciences, Ministry of Public Health, Thailand |
| EPI_ISL_434702 | Praram 9 Hospital | National Institute of Health. Department of medical Sciences, Ministry of Public Health, Thailand |
| EPI_ISL_434703 | Praram 9 Hospital | National Institute of Health. Department of medical Sciences, Ministry of Public Health, Thailand |
| EPI_ISL_434704 | Praram 9 Hospital | National Institute of Health. Department of medical Sciences, Ministry of Public Health, Thailand |
| EPI_ISL_434705 | Praram 9 Hospital | National Institute of Health. Department of medical Sciences, Ministry of Public Health, Thailand |
| EPI_ISL_434706 | Praram 9 Hospital | National Institute of Health. Department of medical Sciences, Ministry of Public Health, Thailand |
| EPI_ISL_434707 | Thammasat University Hospital | National Institute of Health. Department of medical Sciences, Ministry of Public Health, Thailand |
| EPI_ISL_434708 | unknown | National Institute of Health. Department of medical Sciences, Ministry of Public Health, Thailand |
| EPI_ISL_434710 | Viral Respiratory Lab, National Institute for Biomedical Research (INRB) | Pathogen Sequencing Lab, National Institute for Biomedical Research (INRB) |
| EPI_ISL_434711 | Viral Respiratory Lab, National Institute for Biomedical Research (INRB) | Pathogen Sequencing Lab, National Institute for Biomedical Research (INRB) |
| EPI_ISL_434712 | Houston Methodist Hospital | Houston Methodist Hospital |
| EPI_ISL_434713 | Houston Methodist Hospital | Houston Methodist Hospital |
| EPI_ISL_434714 | Houston Methodist Hospital | Houston Methodist Hospital |

| EPI_Accession_ID | Originating Laboratory | Submitting Laboratory |
| --- | --- | --- |
| EPI_ISL_434715 | Houston Methodist Hospital | Houston Methodist Hospital |
| EPI_ISL_434716 | Houston Methodist Hospital | Houston Methodist Hospital |
| EPI_ISL_434717 | Houston Methodist Hospital | Houston Methodist Hospital |
| EPI_ISL_434718 | Houston Methodist Hospital | Houston Methodist Hospital |
| EPI_ISL_434719 | Houston Methodist Hospital | Houston Methodist Hospital |
| EPI_ISL_434720 | Houston Methodist Hospital | Houston Methodist Hospital |
| EPI_ISL_434721 | Houston Methodist Hospital | Houston Methodist Hospital |
| EPI_ISL_434722 | Houston Methodist Hospital | Houston Methodist Hospital |
| EPI_ISL_434723 | Houston Methodist Hospital | Houston Methodist Hospital |
| EPI_ISL_434724 | Houston Methodist Hospital | Houston Methodist Hospital |
| EPI_ISL_434725 | Houston Methodist Hospital | Houston Methodist Hospital |
| EPI_ISL_434726 | Houston Methodist Hospital | Houston Methodist Hospital |
| EPI_ISL_434727 | Houston Methodist Hospital | Houston Methodist Hospital |
| EPI_ISL_434728 | Houston Methodist Hospital | Houston Methodist Hospital |
| EPI_ISL_434729 | Houston Methodist Hospital | Houston Methodist Hospital |
| EPI_ISL_434730 | Houston Methodist Hospital | Houston Methodist Hospital |
| EPI_ISL_434731 | Houston Methodist Hospital | Houston Methodist Hospital |
| EPI_ISL_434732 | Houston Methodist Hospital | Houston Methodist Hospital |
| EPI_ISL_434733 | Houston Methodist Hospital | Houston Methodist Hospital |
| EPI_ISL_434734 | Houston Methodist Hospital | Houston Methodist Hospital |
| EPI_ISL_434735 | Houston Methodist Hospital | Houston Methodist Hospital |
| EPI_ISL_434736 | Houston Methodist Hospital | Houston Methodist Hospital |
| EPI_ISL_434737 | Houston Methodist Hospital | Houston Methodist Hospital |
| EPI_ISL_434738 | Houston Methodist Hospital | Houston Methodist Hospital |
| EPI_ISL_434739 | Houston Methodist Hospital | Houston Methodist Hospital |
| EPI_ISL_434740 | Houston Methodist Hospital | Houston Methodist Hospital |
| EPI_ISL_434741 | Houston Methodist Hospital | Houston Methodist Hospital |
| EPI_ISL_434742 | Houston Methodist Hospital | Houston Methodist Hospital |
| EPI_ISL_434743 | Houston Methodist Hospital | Houston Methodist Hospital |
| EPI_ISL_434744 | Houston Methodist Hospital | Houston Methodist Hospital |
| EPI_ISL_434745 | Houston Methodist Hospital | Houston Methodist Hospital |
| EPI_ISL_434746 | Houston Methodist Hospital | Houston Methodist Hospital |
| EPI_ISL_434747 | Houston Methodist Hospital | Houston Methodist Hospital |

| EPI_Accession_ID | Originating Laboratory | Submitting Laboratory |
| --- | --- | --- |
| EPI_ISL_434748 | Houston Methodist Hospital | Houston Methodist Hospital |
| EPI_ISL_434749 | Houston Methodist Hospital | Houston Methodist Hospital |
| EPI_ISL_434750 | Houston Methodist Hospital | Houston Methodist Hospital |
| EPI_ISL_434751 | Houston Methodist Hospital | Houston Methodist Hospital |
| EPI_ISL_434752 | Houston Methodist Hospital | Houston Methodist Hospital |
| EPI_ISL_434753 | Houston Methodist Hospital | Houston Methodist Hospital |
| EPI_ISL_434754 | Houston Methodist Hospital | Houston Methodist Hospital |
| EPI_ISL_434755 | Houston Methodist Hospital | Houston Methodist Hospital |
| EPI_ISL_434756 | Houston Methodist Hospital | Houston Methodist Hospital |
| EPI_ISL_434757 | Houston Methodist Hospital | Houston Methodist Hospital |
| EPI_ISL_434758 | Houston Methodist Hospital | Houston Methodist Hospital |
| EPI_ISL_434759 | Houston Methodist Hospital | Houston Methodist Hospital |
| EPI_ISL_434760 | Houston Methodist Hospital | Houston Methodist Hospital |
| EPI_ISL_434761 | Houston Methodist Hospital | Houston Methodist Hospital |
| EPI_ISL_434762 | Houston Methodist Hospital | Houston Methodist Hospital |
| EPI_ISL_434763 | Houston Methodist Hospital | Houston Methodist Hospital |
| EPI_ISL_434764 | Houston Methodist Hospital | Houston Methodist Hospital |
| EPI_ISL_434765 | Houston Methodist Hospital | Houston Methodist Hospital |
| EPI_ISL_434766 | Houston Methodist Hospital | Houston Methodist Hospital |
| EPI_ISL_434767 | Houston Methodist Hospital | Houston Methodist Hospital |
| EPI_ISL_434768 | Houston Methodist Hospital | Houston Methodist Hospital |
| EPI_ISL_434769 | Houston Methodist Hospital | Houston Methodist Hospital |
| EPI_ISL_434770 | Houston Methodist Hospital | Houston Methodist Hospital |
| EPI_ISL_434771 | Houston Methodist Hospital | Houston Methodist Hospital |
| EPI_ISL_434772 | Houston Methodist Hospital | Houston Methodist Hospital |
| EPI_ISL_434773 | Houston Methodist Hospital | Houston Methodist Hospital |
| EPI_ISL_434774 | Houston Methodist Hospital | Houston Methodist Hospital |
| EPI_ISL_434775 | Houston Methodist Hospital | Houston Methodist Hospital |
| EPI_ISL_434776 | Houston Methodist Hospital | Houston Methodist Hospital |
| EPI_ISL_434777 | Houston Methodist Hospital | Houston Methodist Hospital |
| EPI_ISL_434779 | Houston Methodist Hospital | Houston Methodist Hospital |
| EPI_ISL_434780 | Houston Methodist Hospital | Houston Methodist Hospital |
| EPI_ISL_434781 | Houston Methodist Hospital | Houston Methodist Hospital |

| EPI_Accession_ID | Originating Laboratory | Submitting Laboratory |
|---|---|---|
| EPI_ISL_434782 | Houston Methodist Hospital | Houston Methodist Hospital |
| EPI_ISL_434783 | Houston Methodist Hospital | Houston Methodist Hospital |
| EPI_ISL_434784 | Houston Methodist Hospital | Houston Methodist Hospital |
| EPI_ISL_434785 | Houston Methodist Hospital | Houston Methodist Hospital |
| EPI_ISL_434786 | Houston Methodist Hospital | Houston Methodist Hospital |
| EPI_ISL_434787 | Houston Methodist Hospital | Houston Methodist Hospital |
| EPI_ISL_434788 | Houston Methodist Hospital | Houston Methodist Hospital |
| EPI_ISL_434789 | Houston Methodist Hospital | Houston Methodist Hospital |
| EPI_ISL_434790 | Houston Methodist Hospital | Houston Methodist Hospital |
| EPI_ISL_434791 | Houston Methodist Hospital | Houston Methodist Hospital |
| EPI_ISL_434792 | Houston Methodist Hospital | Houston Methodist Hospital |
| EPI_ISL_434793 | Houston Methodist Hospital | Houston Methodist Hospital |
| EPI_ISL_434794 | Houston Methodist Hospital | Houston Methodist Hospital |
| EPI_ISL_434795 | Houston Methodist Hospital | Houston Methodist Hospital |
| EPI_ISL_434796 | Houston Methodist Hospital | Houston Methodist Hospital |
| EPI_ISL_434797 | Houston Methodist Hospital | Houston Methodist Hospital |
| EPI_ISL_434798 | Houston Methodist Hospital | Houston Methodist Hospital |
| EPI_ISL_434799 | Houston Methodist Hospital | Houston Methodist Hospital |
| EPI_ISL_434800 | Houston Methodist Hospital | Houston Methodist Hospital |
| EPI_ISL_434801 | Houston Methodist Hospital | Houston Methodist Hospital |
| EPI_ISL_434802 | Houston Methodist Hospital | Houston Methodist Hospital |
| EPI_ISL_434803 | Houston Methodist Hospital | Houston Methodist Hospital |
| EPI_ISL_434804 | Houston Methodist Hospital | Houston Methodist Hospital |
| EPI_ISL_434805 | Houston Methodist Hospital | Houston Methodist Hospital |
| EPI_ISL_434806 | Houston Methodist Hospital | Houston Methodist Hospital |
| EPI_ISL_434807 | Houston Methodist Hospital | Houston Methodist Hospital |
| EPI_ISL_434808 | Houston Methodist Hospital | Houston Methodist Hospital |
| EPI_ISL_434809 | Houston Methodist Hospital | Houston Methodist Hospital |
| EPI_ISL_434810 | Houston Methodist Hospital | Houston Methodist Hospital |
| EPI_ISL_434811 | Houston Methodist Hospital | Houston Methodist Hospital |
| EPI_ISL_434812 | Houston Methodist Hospital | Houston Methodist Hospital |
| EPI_ISL_434813 | Houston Methodist Hospital | Houston Methodist Hospital |
| EPI_ISL_434814 | Houston Methodist Hospital | Houston Methodist Hospital |

| EPI_Accession_ID | Originating Laboratory | Submitting Laboratory |
|---|---|---|
| EPI_ISL_434815 | Houston Methodist Hospital | Houston Methodist Hospital |
| EPI_ISL_434816 | Houston Methodist Hospital | Houston Methodist Hospital |
| EPI_ISL_434817 | Houston Methodist Hospital | Houston Methodist Hospital |
| EPI_ISL_434818 | Houston Methodist Hospital | Houston Methodist Hospital |
| EPI_ISL_434819 | Houston Methodist Hospital | Houston Methodist Hospital |
| EPI_ISL_434820 | Houston Methodist Hospital | Houston Methodist Hospital |
| EPI_ISL_434821 | Houston Methodist Hospital | Houston Methodist Hospital |
| EPI_ISL_434822 | Houston Methodist Hospital | Houston Methodist Hospital |
| EPI_ISL_434823 | Houston Methodist Hospital | Houston Methodist Hospital |
| EPI_ISL_434824 | Houston Methodist Hospital | Houston Methodist Hospital |
| EPI_ISL_434825 | Houston Methodist Hospital | Houston Methodist Hospital |
| EPI_ISL_434826 | Houston Methodist Hospital | Houston Methodist Hospital |
| EPI_ISL_434827 | Houston Methodist Hospital | Houston Methodist Hospital |
| EPI_ISL_434828 | Houston Methodist Hospital | Houston Methodist Hospital |
| EPI_ISL_434829 | Houston Methodist Hospital | Houston Methodist Hospital |
| EPI_ISL_434830 | Houston Methodist Hospital | Houston Methodist Hospital |
| EPI_ISL_434831 | Houston Methodist Hospital | Houston Methodist Hospital |
| EPI_ISL_434832 | Houston Methodist Hospital | Houston Methodist Hospital |
| EPI_ISL_434833 | Houston Methodist Hospital | Houston Methodist Hospital |
| EPI_ISL_434834 | Houston Methodist Hospital | Houston Methodist Hospital |
| EPI_ISL_434835 | Houston Methodist Hospital | Houston Methodist Hospital |
| EPI_ISL_434836 | Houston Methodist Hospital | Houston Methodist Hospital |
| EPI_ISL_434837 | Houston Methodist Hospital | Houston Methodist Hospital |
| EPI_ISL_434838 | Houston Methodist Hospital | Houston Methodist Hospital |
| EPI_ISL_434839 | Houston Methodist Hospital | Houston Methodist Hospital |
| EPI_ISL_434840 | Houston Methodist Hospital | Houston Methodist Hospital |
| EPI_ISL_434841 | Houston Methodist Hospital | Houston Methodist Hospital |
| EPI_ISL_434842 | Houston Methodist Hospital | Houston Methodist Hospital |
| EPI_ISL_434843 | Houston Methodist Hospital | Houston Methodist Hospital |
| EPI_ISL_434844 | Houston Methodist Hospital | Houston Methodist Hospital |
| EPI_ISL_434845 | Houston Methodist Hospital | Houston Methodist Hospital |
| EPI_ISL_434846 | Houston Methodist Hospital | Houston Methodist Hospital |
| EPI_ISL_434847 | Houston Methodist Hospital | Houston Methodist Hospital |

| EPI_Accession_ID | Originating Laboratory | Submitting Laboratory |
|---|---|---|
| EPI_ISL_434848 | Houston Methodist Hospital | Houston Methodist Hospital |
| EPI_ISL_434849 | Houston Methodist Hospital | Houston Methodist Hospital |
| EPI_ISL_434850 | Houston Methodist Hospital | Houston Methodist Hospital |
| EPI_ISL_434851 | Houston Methodist Hospital | Houston Methodist Hospital |
| EPI_ISL_434852 | Houston Methodist Hospital | Houston Methodist Hospital |
| EPI_ISL_434853 | Houston Methodist Hospital | Houston Methodist Hospital |
| EPI_ISL_434854 | Houston Methodist Hospital | Houston Methodist Hospital |
| EPI_ISL_434855 | Houston Methodist Hospital | Houston Methodist Hospital |
| EPI_ISL_434856 | Houston Methodist Hospital | Houston Methodist Hospital |
| EPI_ISL_434857 | Houston Methodist Hospital | Houston Methodist Hospital |
| EPI_ISL_434858 | Houston Methodist Hospital | Houston Methodist Hospital |
| EPI_ISL_434859 | Houston Methodist Hospital | Houston Methodist Hospital |
| EPI_ISL_434860 | Houston Methodist Hospital | Houston Methodist Hospital |
| EPI_ISL_434861 | Houston Methodist Hospital | Houston Methodist Hospital |
| EPI_ISL_434862 | Houston Methodist Hospital | Houston Methodist Hospital |
| EPI_ISL_434863 | Houston Methodist Hospital | Houston Methodist Hospital |
| EPI_ISL_434864 | Houston Methodist Hospital | Houston Methodist Hospital |
| EPI_ISL_434865 | Houston Methodist Hospital | Houston Methodist Hospital |
| EPI_ISL_434866 | Houston Methodist Hospital | Houston Methodist Hospital |
| EPI_ISL_434867 | Houston Methodist Hospital | Houston Methodist Hospital |
| EPI_ISL_434868 | Houston Methodist Hospital | Houston Methodist Hospital |
| EPI_ISL_434869 | Houston Methodist Hospital | Houston Methodist Hospital |
| EPI_ISL_434870 | Houston Methodist Hospital | Houston Methodist Hospital |
| EPI_ISL_434871 | Houston Methodist Hospital | Houston Methodist Hospital |
| EPI_ISL_434872 | Houston Methodist Hospital | Houston Methodist Hospital |
| EPI_ISL_434873 | Houston Methodist Hospital | Houston Methodist Hospital |
| EPI_ISL_434874 | Houston Methodist Hospital | Houston Methodist Hospital |
| EPI_ISL_434875 | Houston Methodist Hospital | Houston Methodist Hospital |
| EPI_ISL_434876 | Houston Methodist Hospital | Houston Methodist Hospital |
| EPI_ISL_434877 | Houston Methodist Hospital | Houston Methodist Hospital |
| EPI_ISL_434878 | Houston Methodist Hospital | Houston Methodist Hospital |
| EPI_ISL_434879 | Houston Methodist Hospital | Houston Methodist Hospital |
| EPI_ISL_434880 | Houston Methodist Hospital | Houston Methodist Hospital |

| EPI_Accession_ID | Originating Laboratory | Submitting Laboratory |
|---|---|---|
| EPI_ISL_434881 | Houston Methodist Hospital | Houston Methodist Hospital |
| EPI_ISL_434882 | Houston Methodist Hospital | Houston Methodist Hospital |
| EPI_ISL_434883 | Houston Methodist Hospital | Houston Methodist Hospital |
| EPI_ISL_434884 | Houston Methodist Hospital | Houston Methodist Hospital |
| EPI_ISL_434885 | Houston Methodist Hospital | Houston Methodist Hospital |
| EPI_ISL_434886 | Houston Methodist Hospital | Houston Methodist Hospital |
| EPI_ISL_434887 | Houston Methodist Hospital | Houston Methodist Hospital |
| EPI_ISL_434888 | Houston Methodist Hospital | Houston Methodist Hospital |
| EPI_ISL_434889 | Houston Methodist Hospital | Houston Methodist Hospital |
| EPI_ISL_434890 | Houston Methodist Hospital | Houston Methodist Hospital |
| EPI_ISL_434891 | Houston Methodist Hospital | Houston Methodist Hospital |
| EPI_ISL_434892 | Houston Methodist Hospital | Houston Methodist Hospital |
| EPI_ISL_434893 | Houston Methodist Hospital | Houston Methodist Hospital |
| EPI_ISL_434894 | Houston Methodist Hospital | Houston Methodist Hospital |
| EPI_ISL_434895 | Houston Methodist Hospital | Houston Methodist Hospital |
| EPI_ISL_434896 | Houston Methodist Hospital | Houston Methodist Hospital |
| EPI_ISL_434897 | Houston Methodist Hospital | Houston Methodist Hospital |
| EPI_ISL_434898 | Houston Methodist Hospital | Houston Methodist Hospital |
| EPI_ISL_434899 | Houston Methodist Hospital | Houston Methodist Hospital |
| EPI_ISL_434900 | Houston Methodist Hospital | Houston Methodist Hospital |
| EPI_ISL_434901 | Houston Methodist Hospital | Houston Methodist Hospital |
| EPI_ISL_434902 | Houston Methodist Hospital | Houston Methodist Hospital |
| EPI_ISL_434903 | Houston Methodist Hospital | Houston Methodist Hospital |
| EPI_ISL_434904 | Houston Methodist Hospital | Houston Methodist Hospital |
| EPI_ISL_434905 | Houston Methodist Hospital | Houston Methodist Hospital |
| EPI_ISL_434906 | Houston Methodist Hospital | Houston Methodist Hospital |
| EPI_ISL_434907 | Houston Methodist Hospital | Houston Methodist Hospital |
| EPI_ISL_434908 | Houston Methodist Hospital | Houston Methodist Hospital |
| EPI_ISL_434909 | Houston Methodist Hospital | Houston Methodist Hospital |
| EPI_ISL_434910 | Houston Methodist Hospital | Houston Methodist Hospital |
| EPI_ISL_434911 | Houston Methodist Hospital | Houston Methodist Hospital |
| EPI_ISL_434912 | Houston Methodist Hospital | Houston Methodist Hospital |
| EPI_ISL_434913 | Houston Methodist Hospital | Houston Methodist Hospital |

| EPI_Accession_ID | Originating Laboratory | Submitting Laboratory |
|---|---|---|
| EPI_ISL_434914 | Houston Methodist Hospital | Houston Methodist Hospital |
| EPI_ISL_434915 | Houston Methodist Hospital | Houston Methodist Hospital |
| EPI_ISL_434916 | Houston Methodist Hospital | Houston Methodist Hospital |
| EPI_ISL_434917 | Houston Methodist Hospital | Houston Methodist Hospital |
| EPI_ISL_434918 | Houston Methodist Hospital | Houston Methodist Hospital |
| EPI_ISL_434919 | Houston Methodist Hospital | Houston Methodist Hospital |
| EPI_ISL_434920 | Houston Methodist Hospital | Houston Methodist Hospital |
| EPI_ISL_434921 | Houston Methodist Hospital | Houston Methodist Hospital |
| EPI_ISL_434922 | Houston Methodist Hospital | Houston Methodist Hospital |
| EPI_ISL_434923 | Houston Methodist Hospital | Houston Methodist Hospital |
| EPI_ISL_434924 | Houston Methodist Hospital | Houston Methodist Hospital |
| EPI_ISL_434925 | Houston Methodist Hospital | Houston Methodist Hospital |
| EPI_ISL_434926 | Houston Methodist Hospital | Houston Methodist Hospital |
| EPI_ISL_434927 | Houston Methodist Hospital | Houston Methodist Hospital |
| EPI_ISL_434928 | Houston Methodist Hospital | Houston Methodist Hospital |
| EPI_ISL_434929 | Houston Methodist Hospital | Houston Methodist Hospital |
| EPI_ISL_434930 | Houston Methodist Hospital | Houston Methodist Hospital |
| EPI_ISL_434931 | Houston Methodist Hospital | Houston Methodist Hospital |
| EPI_ISL_434932 | Houston Methodist Hospital | Houston Methodist Hospital |
| EPI_ISL_434933 | Houston Methodist Hospital | Houston Methodist Hospital |
| EPI_ISL_434934 | Houston Methodist Hospital | Houston Methodist Hospital |
| EPI_ISL_434935 | Houston Methodist Hospital | Houston Methodist Hospital |
| EPI_ISL_434936 | Houston Methodist Hospital | Houston Methodist Hospital |
| EPI_ISL_434937 | Houston Methodist Hospital | Houston Methodist Hospital |
| EPI_ISL_434938 | Houston Methodist Hospital | Houston Methodist Hospital |
| EPI_ISL_434939 | Houston Methodist Hospital | Houston Methodist Hospital |
| EPI_ISL_434940 | Houston Methodist Hospital | Houston Methodist Hospital |
| EPI_ISL_434941 | Houston Methodist Hospital | Houston Methodist Hospital |
| EPI_ISL_434942 | Houston Methodist Hospital | Houston Methodist Hospital |
| EPI_ISL_434943 | Houston Methodist Hospital | Houston Methodist Hospital |
| EPI_ISL_434944 | Houston Methodist Hospital | Houston Methodist Hospital |
| EPI_ISL_434945 | Houston Methodist Hospital | Houston Methodist Hospital |
| EPI_ISL_434946 | Houston Methodist Hospital | Houston Methodist Hospital |

| EPI_Accession_ID | Originating Laboratory | Submitting Laboratory |
|---|---|---|
| EPI_ISL_434947 | Houston Methodist Hospital | Houston Methodist Hospital |
| EPI_ISL_434948 | Houston Methodist Hospital | Houston Methodist Hospital |
| EPI_ISL_434949 | Houston Methodist Hospital | Houston Methodist Hospital |
| EPI_ISL_434950 | Houston Methodist Hospital | Houston Methodist Hospital |
| EPI_ISL_434951 | Houston Methodist Hospital | Houston Methodist Hospital |
| EPI_ISL_434952 | Houston Methodist Hospital | Houston Methodist Hospital |
| EPI_ISL_434953 | Houston Methodist Hospital | Houston Methodist Hospital |
| EPI_ISL_434954 | Houston Methodist Hospital | Houston Methodist Hospital |
| EPI_ISL_434955 | Houston Methodist Hospital | Houston Methodist Hospital |
| EPI_ISL_434956 | Houston Methodist Hospital | Houston Methodist Hospital |
| EPI_ISL_434957 | Houston Methodist Hospital | Houston Methodist Hospital |
| EPI_ISL_434958 | Houston Methodist Hospital | Houston Methodist Hospital |
| EPI_ISL_434959 | Houston Methodist Hospital | Houston Methodist Hospital |
| EPI_ISL_434960 | Houston Methodist Hospital | Houston Methodist Hospital |
| EPI_ISL_434961 | Houston Methodist Hospital | Houston Methodist Hospital |
| EPI_ISL_434962 | Houston Methodist Hospital | Houston Methodist Hospital |
| EPI_ISL_434963 | Houston Methodist Hospital | Houston Methodist Hospital |
| EPI_ISL_434964 | Houston Methodist Hospital | Houston Methodist Hospital |
| EPI_ISL_434965 | Houston Methodist Hospital | Houston Methodist Hospital |
| EPI_ISL_434966 | Houston Methodist Hospital | Houston Methodist Hospital |
| EPI_ISL_434967 | Houston Methodist Hospital | Houston Methodist Hospital |
| EPI_ISL_434968 | Houston Methodist Hospital | Houston Methodist Hospital |
| EPI_ISL_434969 | Houston Methodist Hospital | Houston Methodist Hospital |
| EPI_ISL_434970 | Houston Methodist Hospital | Houston Methodist Hospital |
| EPI_ISL_434971 | Houston Methodist Hospital | Houston Methodist Hospital |
| EPI_ISL_434972 | Houston Methodist Hospital | Houston Methodist Hospital |
| EPI_ISL_434973 | Houston Methodist Hospital | Houston Methodist Hospital |
| EPI_ISL_434974 | Houston Methodist Hospital | Houston Methodist Hospital |
| EPI_ISL_434975 | Houston Methodist Hospital | Houston Methodist Hospital |
| EPI_ISL_434976 | Houston Methodist Hospital | Houston Methodist Hospital |
| EPI_ISL_434977 | Houston Methodist Hospital | Houston Methodist Hospital |
| EPI_ISL_434978 | Houston Methodist Hospital | Houston Methodist Hospital |
| EPI_ISL_434979 | Houston Methodist Hospital | Houston Methodist Hospital |

| EPI_Accession_ID | Originating Laboratory | Submitting Laboratory |
|---|---|---|
| EPI_ISL_434980 | Houston Methodist Hospital | Houston Methodist Hospital |
| EPI_ISL_434981 | Houston Methodist Hospital | Houston Methodist Hospital |
| EPI_ISL_434982 | Houston Methodist Hospital | Houston Methodist Hospital |
| EPI_ISL_434983 | Houston Methodist Hospital | Houston Methodist Hospital |
| EPI_ISL_434984 | Houston Methodist Hospital | Houston Methodist Hospital |
| EPI_ISL_434985 | Houston Methodist Hospital | Houston Methodist Hospital |
| EPI_ISL_434986 | Houston Methodist Hospital | Houston Methodist Hospital |
| EPI_ISL_434987 | Houston Methodist Hospital | Houston Methodist Hospital |
| EPI_ISL_434988 | Houston Methodist Hospital | Houston Methodist Hospital |
| EPI_ISL_434989 | Houston Methodist Hospital | Houston Methodist Hospital |
| EPI_ISL_434990 | Houston Methodist Hospital | Houston Methodist Hospital |
| EPI_ISL_434991 | Houston Methodist Hospital | Houston Methodist Hospital |
| EPI_ISL_434992 | Houston Methodist Hospital | Houston Methodist Hospital |
| EPI_ISL_434993 | Houston Methodist Hospital | Houston Methodist Hospital |
| EPI_ISL_434994 | Houston Methodist Hospital | Houston Methodist Hospital |
| EPI_ISL_434995 | Houston Methodist Hospital | Houston Methodist Hospital |
| EPI_ISL_434996 | Houston Methodist Hospital | Houston Methodist Hospital |
| EPI_ISL_434997 | Houston Methodist Hospital | Houston Methodist Hospital |
| EPI_ISL_434998 | Houston Methodist Hospital | Houston Methodist Hospital |
| EPI_ISL_434999 | Houston Methodist Hospital | Houston Methodist Hospital |
| EPI_ISL_435000 | Houston Methodist Hospital | Houston Methodist Hospital |
| EPI_ISL_435001 | Houston Methodist Hospital | Houston Methodist Hospital |
| EPI_ISL_435002 | Houston Methodist Hospital | Houston Methodist Hospital |
| EPI_ISL_435003 | Houston Methodist Hospital | Houston Methodist Hospital |
| EPI_ISL_435004 | Houston Methodist Hospital | Houston Methodist Hospital |
| EPI_ISL_435005 | Houston Methodist Hospital | Houston Methodist Hospital |
| EPI_ISL_435006 | Houston Methodist Hospital | Houston Methodist Hospital |
| EPI_ISL_435007 | Houston Methodist Hospital | Houston Methodist Hospital |
| EPI_ISL_435008 | Houston Methodist Hospital | Houston Methodist Hospital |
| EPI_ISL_435009 | Houston Methodist Hospital | Houston Methodist Hospital |
| EPI_ISL_435010 | Houston Methodist Hospital | Houston Methodist Hospital |
| EPI_ISL_435011 | Houston Methodist Hospital | Houston Methodist Hospital |
| EPI_ISL_435012 | Houston Methodist Hospital | Houston Methodist Hospital |

| EPI_Accession_ID | Originating Laboratory | Submitting Laboratory |
|---|---|---|
| EPI_ISL_435013 | Houston Methodist Hospital | Houston Methodist Hospital |
| EPI_ISL_435014 | Houston Methodist Hospital | Houston Methodist Hospital |
| EPI_ISL_435015 | Houston Methodist Hospital | Houston Methodist Hospital |
| EPI_ISL_435016 | Houston Methodist Hospital | Houston Methodist Hospital |
| EPI_ISL_435017 | Houston Methodist Hospital | Houston Methodist Hospital |
| EPI_ISL_435018 | Houston Methodist Hospital | Houston Methodist Hospital |
| EPI_ISL_435019 | Houston Methodist Hospital | Houston Methodist Hospital |
| EPI_ISL_435020 | Houston Methodist Hospital | Houston Methodist Hospital |
| EPI_ISL_435021 | Houston Methodist Hospital | Houston Methodist Hospital |
| EPI_ISL_435022 | Houston Methodist Hospital | Houston Methodist Hospital |
| EPI_ISL_435023 | Houston Methodist Hospital | Houston Methodist Hospital |
| EPI_ISL_435024 | Houston Methodist Hospital | Houston Methodist Hospital |
| EPI_ISL_435025 | Houston Methodist Hospital | Houston Methodist Hospital |
| EPI_ISL_435026 | Houston Methodist Hospital | Houston Methodist Hospital |
| EPI_ISL_435027 | Houston Methodist Hospital | Houston Methodist Hospital |
| EPI_ISL_435028 | Houston Methodist Hospital | Houston Methodist Hospital |
| EPI_ISL_435029 | Houston Methodist Hospital | Houston Methodist Hospital |
| EPI_ISL_435030 | Houston Methodist Hospital | Houston Methodist Hospital |
| EPI_ISL_435031 | Houston Methodist Hospital | Houston Methodist Hospital |
| EPI_ISL_435032 | Viral Respiratory Lab, National Institute for Biomedical Research (INRB) | Pathogen Sequencing Lab, National Institute for Biomedical Research (INRB) |
| EPI_ISL_435033 | Viral Respiratory Lab, National Institute for Biomedical Research (INRB) | Pathogen Sequencing Lab, National Institute for Biomedical Research (INRB) |
| EPI_ISL_435034 | LSUHS Emerging Viral Threat Laboratory | Microbial Genome Sequencing Center |
| EPI_ISL_435035 | LSUHS Emerging Viral Threat Laboratory | Microbial Genome Sequencing Center |
| EPI_ISL_435036 | LSUHS Emerging Viral Threat Laboratory | Microbial Genome Sequencing Center |
| EPI_ISL_435037 | LSUHS Emerging Viral Threat Laboratory | Microbial Genome Sequencing Center |
| EPI_ISL_435038 | LSUHS Emerging Viral Threat Laboratory | Microbial Genome Sequencing Center |
| EPI_ISL_435039 | LSUHS Emerging Viral Threat Laboratory | Microbial Genome Sequencing Center |
| EPI_ISL_435040 | LSUHS Emerging Viral Threat Laboratory | Microbial Genome Sequencing Center |
| EPI_ISL_435041 | LSUHS Emerging Viral Threat Laboratory | Microbial Genome Sequencing Center |
| EPI_ISL_435042 | LSUHS Emerging Viral Threat Laboratory | Microbial Genome Sequencing Center |
| EPI_ISL_435043 | LSUHS Emerging Viral Threat Laboratory | Microbial Genome Sequencing Center |

| EPI_Accession_ID | Originating Laboratory | Submitting Laboratory |
|---|---|---|
| EPI_ISL_435044 | LSUHS Emerging Viral Threat Laboratory | Microbial Genome Sequencing Center |
| EPI_ISL_435045 | Laboratory of Applied Genetics | RSE "National Center for Biotechnology" |
| EPI_ISL_435046 | Laboratory of Applied Genetics | RSE "National Center for Biotechnology" |
| EPI_ISL_435047 | Laboratory of Applied Genetics | RSE "National Center for Biotechnology" |
| EPI_ISL_435048 | Laboratory of Applied Genetics | RSE "National Center for Biotechnology" |
| EPI_ISL_435049 | B.J. Medical College and Civil hospital | Gujarat Biotechnology Research Centre |
| EPI_ISL_435050 | B.J. Medical College and Civil hospital | Gujarat Biotechnology Research Centre |
| EPI_ISL_435051 | B.J. Medical College and Civil hospital | Gujarat Biotechnology Research Centre |
| EPI_ISL_435052 | B.J. Medical College and Civil hospital | Gujarat Biotechnology Research Centre |
| EPI_ISL_435053 | B.J. Medical College and Civil hospital | Gujarat Biotechnology Research Centre |
| EPI_ISL_435054 | B.J. Medical College and Civil hospital | Gujarat Biotechnology Research Centre |
| EPI_ISL_435055 | Gujarat Biotechnology Research Centre | Gujarat Biotechnology Research Centre |
| EPI_ISL_435056 | Gujarat Biotechnology Research Centre | Gujarat Biotechnology Research Centre |
| EPI_ISL_435057 | T.C. SaÄŸlÄ±k BakanlÄ±ÄŸÄ± AdÄ±yaman Ä°l SaÄŸlÄ±k MÃ¼dÃ¼rlÃ¼ÄŸÃ¼ AdÄ±yaman EÄŸitim Ve AraÅŸtÄ±rma Hastanesi | VETAL Animal Health Products Company, BSL3+ Production Laboratuary, Turkey |
| EPI_ISL_435060 | National Centre for Disease control (NCDC), CSIR-Institute of Genomics and Integrative Biology (CSIR-IGIB) | NCDC/CSIR-IGIB |
| EPI_ISL_435061 | National Centre for Disease control (NCDC), CSIR-Institute of Genomics and Integrative Biology (CSIR-IGIB) | NCDC/CSIR-IGIB |
| EPI_ISL_435062 | National Centre for Disease control (NCDC), CSIR-Institute of Genomics and Integrative Biology (CSIR-IGIB) | NCDC/CSIR-IGIB |
| EPI_ISL_435063 | National Centre for Disease control (NCDC), CSIR-Institute of Genomics and Integrative Biology (CSIR-IGIB) | NCDC/CSIR-IGIB |
| EPI_ISL_435064 | National Centre for Disease control (NCDC), CSIR-Institute of Genomics and Integrative Biology (CSIR-IGIB) | NCDC/CSIR-IGIB |

| EPI_Accession_ID | Originating Laboratory | Submitting Laboratory |
|---|---|---|
| EPI_ISL_435065 | National Centre for Disease control (NCDC), CSIR-Institute of Genomics and Integrative Biology (CSIR-IGIB) | NCDC/CSIR-IGIB |
| EPI_ISL_435066 | National Centre for Disease control (NCDC), CSIR-Institute of Genomics and Integrative Biology (CSIR-IGIB) | NCDC/CSIR-IGIB |
| EPI_ISL_435067 | National Centre for Disease control (NCDC), CSIR-Institute of Genomics and Integrative Biology (CSIR-IGIB) | NCDC/CSIR-IGIB |
| EPI_ISL_435068 | National Centre for Disease control (NCDC), CSIR-Institute of Genomics and Integrative Biology (CSIR-IGIB) | NCDC/CSIR-IGIB |
| EPI_ISL_435069 | National Centre for Disease control (NCDC), CSIR-Institute of Genomics and Integrative Biology (CSIR-IGIB) | NCDC/CSIR-IGIB |
| EPI_ISL_435070 | National Centre for Disease control (NCDC), CSIR-Institute of Genomics and Integrative Biology (CSIR-IGIB) | NCDC/CSIR-IGIB |
| EPI_ISL_435071 | National Centre for Disease control (NCDC), CSIR-Institute of Genomics and Integrative Biology (CSIR-IGIB) | NCDC/CSIR-IGIB |
| EPI_ISL_435072 | National Centre for Disease control (NCDC), CSIR-Institute of Genomics and Integrative Biology (CSIR-IGIB) | NCDC/CSIR-IGIB |
| EPI_ISL_435073 | National Centre for Disease control (NCDC), CSIR-Institute of Genomics and Integrative Biology (CSIR-IGIB) | NCDC/CSIR-IGIB |
| EPI_ISL_435074 | National Centre for Disease control (NCDC), CSIR-Institute of Genomics and Integrative Biology (CSIR-IGIB) | NCDC/CSIR-IGIB |
| EPI_ISL_435075 | National Centre for Disease control (NCDC), CSIR-Institute of Genomics and Integrative Biology (CSIR-IGIB) | NCDC/CSIR-IGIB |

| EPI_Accession_ID | Originating Laboratory | Submitting Laboratory |
|---|---|---|
| EPI_ISL_435076 | National Centre for Disease control (NCDC), CSIR-Institute of Genomics and Integrative Biology (CSIR-IGIB) | NCDC/CSIR-IGIB |
| EPI_ISL_435077 | National Centre for Disease control (NCDC), CSIR-Institute of Genomics and Integrative Biology (CSIR-IGIB) | NCDC/CSIR-IGIB |
| EPI_ISL_435078 | National Centre for Disease control (NCDC), CSIR-Institute of Genomics and Integrative Biology (CSIR-IGIB) | NCDC/CSIR-IGIB |
| EPI_ISL_435079 | National Centre for Disease control (NCDC), CSIR-Institute of Genomics and Integrative Biology (CSIR-IGIB) | NCDC/CSIR-IGIB |
| EPI_ISL_435080 | National Centre for Disease control (NCDC), CSIR-Institute of Genomics and Integrative Biology (CSIR-IGIB) | NCDC/CSIR-IGIB |
| EPI_ISL_435081 | National Centre for Disease control (NCDC), CSIR-Institute of Genomics and Integrative Biology (CSIR-IGIB) | NCDC/CSIR-IGIB |
| EPI_ISL_435082 | National Centre for Disease control (NCDC), CSIR-Institute of Genomics and Integrative Biology (CSIR-IGIB) | NCDC/CSIR-IGIB |
| EPI_ISL_435083 | National Centre for Disease control (NCDC), CSIR-Institute of Genomics and Integrative Biology (CSIR-IGIB) | NCDC/CSIR-IGIB |
| EPI_ISL_435084 | National Centre for Disease control (NCDC), CSIR-Institute of Genomics and Integrative Biology (CSIR-IGIB) | NCDC/CSIR-IGIB |
| EPI_ISL_435085 | National Centre for Disease control (NCDC), CSIR-Institute of Genomics and Integrative Biology (CSIR-IGIB) | NCDC/CSIR-IGIB |
| EPI_ISL_435086 | National Centre for Disease control (NCDC), CSIR-Institute of Genomics and Integrative Biology (CSIR-IGIB) | NCDC/CSIR-IGIB |

| EPI_Accession_ID | Originating Laboratory | Submitting Laboratory |
|---|---|---|
| EPI_ISL_435087 | National Centre for Disease control (NCDC), CSIR-Institute of Genomics and Integrative Biology (CSIR-IGIB) | NCDC/CSIR-IGIB |
| EPI_ISL_435088 | National Centre for Disease control (NCDC), CSIR-Institute of Genomics and Integrative Biology (CSIR-IGIB) | NCDC/CSIR-IGIB |
| EPI_ISL_435089 | National Centre for Disease control (NCDC), CSIR-Institute of Genomics and Integrative Biology (CSIR-IGIB) | NCDC/CSIR-IGIB |
| EPI_ISL_435090 | National Centre for Disease control (NCDC), CSIR-Institute of Genomics and Integrative Biology (CSIR-IGIB) | NCDC/CSIR-IGIB |
| EPI_ISL_435091 | National Centre for Disease control (NCDC), CSIR-Institute of Genomics and Integrative Biology (CSIR-IGIB) | NCDC/CSIR-IGIB |
| EPI_ISL_435092 | National Centre for Disease control (NCDC), CSIR-Institute of Genomics and Integrative Biology (CSIR-IGIB) | NCDC/CSIR-IGIB |
| EPI_ISL_435093 | National Centre for Disease control (NCDC), CSIR-Institute of Genomics and Integrative Biology (CSIR-IGIB) | NCDC/CSIR-IGIB |
| EPI_ISL_435094 | National Centre for Disease control (NCDC), CSIR-Institute of Genomics and Integrative Biology (CSIR-IGIB) | NCDC/CSIR-IGIB |
| EPI_ISL_435095 | National Centre for Disease control (NCDC), CSIR-Institute of Genomics and Integrative Biology (CSIR-IGIB) | NCDC/CSIR-IGIB |
| EPI_ISL_435096 | National Centre for Disease control (NCDC), CSIR-Institute of Genomics and Integrative Biology (CSIR-IGIB) | NCDC/CSIR-IGIB |
| EPI_ISL_435097 | National Centre for Disease control (NCDC), CSIR-Institute of Genomics and Integrative Biology (CSIR-IGIB) | NCDC/CSIR-IGIB |

| EPI_Accession_ID | Originating Laboratory | Submitting Laboratory |
|---|---|---|
| EPI_ISL_435098 | National Centre for Disease control (NCDC), CSIR-Institute of Genomics and Integrative Biology (CSIR-IGIB) | NCDC/CSIR-IGIB |
| EPI_ISL_435099 | National Centre for Disease control (NCDC), CSIR-Institute of Genomics and Integrative Biology (CSIR-IGIB) | NCDC/CSIR-IGIB |
| EPI_ISL_435100 | National Centre for Disease control (NCDC), CSIR-Institute of Genomics and Integrative Biology (CSIR-IGIB) | NCDC/CSIR-IGIB |
| EPI_ISL_435101 | National Centre for Disease control (NCDC), CSIR-Institute of Genomics and Integrative Biology (CSIR-IGIB) | NCDC/CSIR-IGIB |
| EPI_ISL_435102 | National Centre for Disease control (NCDC), CSIR-Institute of Genomics and Integrative Biology (CSIR-IGIB) | NCDC/CSIR-IGIB |
| EPI_ISL_435103 | National Centre for Disease control (NCDC), CSIR-Institute of Genomics and Integrative Biology (CSIR-IGIB) | NCDC/CSIR-IGIB |
| EPI_ISL_435104 | National Centre for Disease control (NCDC), CSIR-Institute of Genomics and Integrative Biology (CSIR-IGIB) | NCDC/CSIR-IGIB |
| EPI_ISL_435105 | National Centre for Disease control (NCDC), CSIR-Institute of Genomics and Integrative Biology (CSIR-IGIB) | NCDC/CSIR-IGIB |
| EPI_ISL_435106 | National Centre for Disease control (NCDC), CSIR-Institute of Genomics and Integrative Biology (CSIR-IGIB) | NCDC/CSIR-IGIB |
| EPI_ISL_435107 | National Centre for Disease control (NCDC), CSIR-Institute of Genomics and Integrative Biology (CSIR-IGIB) | NCDC/CSIR-IGIB |
| EPI_ISL_435108 | National Centre for Disease control (NCDC), CSIR-Institute of Genomics and Integrative Biology (CSIR-IGIB) | NCDC/CSIR-IGIB |

| EPI_Accession_ID | Originating Laboratory | Submitting Laboratory |
|---|---|---|
| EPI_ISL_435109 | National Centre for Disease control (NCDC), CSIR-Institute of Genomics and Integrative Biology (CSIR-IGIB) | NCDC/CSIR-IGIB |
| EPI_ISL_435110 | National Centre for Disease control (NCDC), CSIR-Institute of Genomics and Integrative Biology (CSIR-IGIB) | NCDC/CSIR-IGIB |
| EPI_ISL_435111 | National Centre for Disease control (NCDC), CSIR-Institute of Genomics and Integrative Biology (CSIR-IGIB) | NCDC/CSIR-IGIB |
| EPI_ISL_435112 | National Centre for Disease control (NCDC), CSIR-Institute of Genomics and Integrative Biology (CSIR-IGIB) | NCDC/CSIR-IGIB |
| EPI_ISL_435113 | Viral Respiratory Lab, National Institute for Biomedical Research (INRB) | Pathogen Sequencing Lab, National Institute for Biomedical Research (INRB) |
| EPI_ISL_435114 | Viral Respiratory Lab, National Institute for Biomedical Research (INRB) | Pathogen Sequencing Lab, National Institute for Biomedical Research (INRB) |
| EPI_ISL_435116 | Viral Respiratory Lab, National Institute for Biomedical Research (INRB) | Pathogen Sequencing Lab, National Institute for Biomedical Research (INRB) |
| EPI_ISL_435117 | Viral Respiratory Lab, National Institute for Biomedical Research (INRB) | Pathogen Sequencing Lab, National Institute for Biomedical Research (INRB) |
| EPI_ISL_435118 | Viral Respiratory Lab, National Institute for Biomedical Research (INRB) | Pathogen Sequencing Lab, National Institute for Biomedical Research (INRB) |
| EPI_ISL_435119 | Mohammed Bin Rashid University of Medicine and Health Sciences | Al Jalila Childrenâ€™s Hospital |
| EPI_ISL_435120 | Mohammed Bin Rashid University of Medicine and Health Sciences | Al Jalila Genomics Center |
| EPI_ISL_435121 | Mohammed Bin Rashid University of Medicine and Health Sciences | Al Jalila Genomics Center |
| EPI_ISL_435122 | Mohammed Bin Rashid University of Medicine and Health Sciences | Al Jalila Genomics Center |
| EPI_ISL_435123 | Mohammed Bin Rashid University of Medicine and Health Sciences | Al Jalila Genomics Center |

| EPI_Accession_ID | Originating Laboratory | Submitting Laboratory |
|---|---|---|
| EPI_ISL_435124 | Mohammed Bin Rashid University of Medicine and Health Sciences | Al Jalila Genomics Center |
| EPI_ISL_435125 | Mohammed Bin Rashid University of Medicine and Health Sciences | Al Jalila Genomics Center |
| EPI_ISL_435126 | Mohammed Bin Rashid University of Medicine and Health Sciences | Al Jalila Genomics Center |
| EPI_ISL_435127 | Mohammed Bin Rashid University of Medicine and Health Sciences | Al Jalila Genomics Center |
| EPI_ISL_435128 | Mohammed Bin Rashid University of Medicine and Health Sciences | Al Jalila Genomics Center |
| EPI_ISL_435129 | Mohammed Bin Rashid University of Medicine and Health Sciences | Al Jalila Genomics Center |
| EPI_ISL_435130 | Mohammed Bin Rashid University of Medicine and Health Sciences | Al Jalila Genomics Center |
| EPI_ISL_435131 | Mohammed Bin Rashid University of Medicine and Health Sciences | Al Jalila Genomics Center |
| EPI_ISL_435132 | Mohammed Bin Rashid University of Medicine and Health Sciences | Al Jalila Genomics Center |
| EPI_ISL_435133 | Mohammed Bin Rashid University of Medicine and Health Sciences | Al Jalila Genomics Center |
| EPI_ISL_435134 | Mohammed Bin Rashid University of Medicine and Health Sciences | Al Jalila Genomics Center |
| EPI_ISL_435135 | Mohammed Bin Rashid University of Medicine and Health Sciences | Al Jalila Genomics Center |
| EPI_ISL_435136 | Mohammed Bin Rashid University of Medicine and Health Sciences | Al Jalila Genomics Center |
| EPI_ISL_435137 | Mohammed Bin Rashid University of Medicine and Health Sciences | Al Jalila Genomics Center |
| EPI_ISL_435138 | Mohammed Bin Rashid University of Medicine and Health Sciences | Al Jalila Genomics Center |
| EPI_ISL_435139 | Mohammed Bin Rashid University of Medicine and Health Sciences | Al Jalila Genomics Center |

| EPI_Accession_ID | Originating Laboratory | Submitting Laboratory |
| --- | --- | --- |
| EPI_ISL_435140 | Mohammed Bin Rashid University of Medicine and Health Sciences | Al Jalila Genomics Center |
| EPI_ISL_435141 | Mohammed Bin Rashid University of Medicine and Health Sciences | Al Jalila Genomics Center |
| EPI_ISL_435142 | Mohammed Bin Rashid University of Medicine and Health Sciences | Al Jalila Genomics Center |
| EPI_ISL_435143 | Mohammed Bin Rashid University of Medicine and Health Sciences | Al Jalila Genomics Center |
| EPI_ISL_435144 | Hospital Universitario La Paz | Hospital Universitario 12 de Octubre |
| EPI_ISL_435145 | Ospedale Civile Giuseppe Mazzini | Istituto Zooprofilattico Sperimentale dellâ€™Abruzzo e Molise â€œG. Caporaleâ€� |
| EPI_ISL_435146 | Villa Serena del Dr. Leonardo Petruzzi | Istituto Zooprofilattico Sperimentale dellâ€™Abruzzo e Molise â€œG. Caporaleâ€� |
| EPI_ISL_435147 | Villa Serena del Dr. Leonardo Petruzzi | Istituto Zooprofilattico Sperimentale dellâ€™Abruzzo e Molise â€œG. Caporaleâ€� |
| EPI_ISL_435148 | Ospedale SS Annunziata | Istituto Zooprofilattico Sperimentale dellâ€™Abruzzo e Molise â€œG. Caporaleâ€� |
| EPI_ISL_435149 | SERVIZIO DI IGIENE E SANITÃ€ PUBBLICA ASL Teramo | Istituto Zooprofilattico Sperimentale dellâ€™Abruzzo e Molise â€œG. Caporaleâ€� |
| EPI_ISL_435150 | Ospedale SS Annunziata | Istituto Zooprofilattico Sperimentale dellâ€™Abruzzo e Molise â€œG. Caporaleâ€� |
| EPI_ISL_435151 | Ospedale SS Annunziata | Istituto Zooprofilattico Sperimentale dellâ€™Abruzzo e Molise â€œG. Caporaleâ€� |
| EPI_ISL_435152 | Servizio di Igiene, Epidemiologia e SanitÃ Pubblica (SIESP) Avezzano | Istituto Zooprofilattico Sperimentale dellâ€™Abruzzo e Molise â€œG. Caporaleâ€� |
| EPI_ISL_435153 | SERVIZIO DI IGIENE E SANITÃ€ PUBBLICA ASL Teramo | Istituto Zooprofilattico Sperimentale dellâ€™Abruzzo e Molise â€œG. Caporaleâ€� |
| EPI_ISL_435154 | SERVIZIO DI IGIENE E SANITÃ€ PUBBLICA ASL Teramo | Istituto Zooprofilattico Sperimentale dellâ€™Abruzzo e Molise â€œG. Caporaleâ€� |
| EPI_ISL_435155 | SERVIZIO DI IGIENE E SANITÃ€ PUBBLICA ASL Teramo | Istituto Zooprofilattico Sperimentale dellâ€™Abruzzo e Molise â€œG. Caporaleâ€� |
| EPI_ISL_435156 | Viral Respiratory Lab, National Institute for Biomedical Research (INRB) | Pathogen Sequencing Lab, National Institute for Biomedical Research (INRB) |

| EPI_Accession_ID | Originating Laboratory | Submitting Laboratory |
|---|---|---|
| EPI_ISL_435157 | Viral Respiratory Lab, National Institute for Biomedical Research (INRB) | Pathogen Sequencing Lab, National Institute for Biomedical Research (INRB) |
| EPI_ISL_435158 | Viral Respiratory Lab, National Institute for Biomedical Research (INRB) | Pathogen Sequencing Lab, National Institute for Biomedical Research (INRB) |
| EPI_ISL_435159 | Viral Respiratory Lab, National Institute for Biomedical Research (INRB) | Pathogen Sequencing Lab, National Institute for Biomedical Research (INRB) |
| EPI_ISL_435160 | Viral Respiratory Lab, National Institute for Biomedical Research (INRB) | Pathogen Sequencing Lab, National Institute for Biomedical Research (INRB) |
| EPI_ISL_435161 | Viral Respiratory Lab, National Institute for Biomedical Research (INRB) | Pathogen Sequencing Lab, National Institute for Biomedical Research (INRB) |
| EPI_ISL_435162 | Viral Respiratory Lab, National Institute for Biomedical Research (INRB) | Pathogen Sequencing Lab, National Institute for Biomedical Research (INRB) |
| EPI_ISL_435163 | Viral Respiratory Lab, National Institute for Biomedical Research (INRB) | Pathogen Sequencing Lab, National Institute for Biomedical Research (INRB) |
| EPI_ISL_435164 | Viral Respiratory Lab, National Institute for Biomedical Research (INRB) | Pathogen Sequencing Lab, National Institute for Biomedical Research (INRB) |
| EPI_ISL_435165 | Viral Respiratory Lab, National Institute for Biomedical Research (INRB) | Pathogen Sequencing Lab, National Institute for Biomedical Research (INRB) |
| EPI_ISL_435166 | Viral Respiratory Lab, National Institute for Biomedical Research (INRB) | Pathogen Sequencing Lab, National Institute for Biomedical Research (INRB) |
| EPI_ISL_435167 | Viral Respiratory Lab, National Institute for Biomedical Research (INRB) | Pathogen Sequencing Lab, National Institute for Biomedical Research (INRB) |
| EPI_ISL_435168 | Viral Respiratory Lab, National Institute for Biomedical Research (INRB) | Pathogen Sequencing Lab, National Institute for Biomedical Research (INRB) |
| EPI_ISL_435281 | Medistra Hospital Jakarta | Eijkman Institute for Molecular Biology, Ministry of Research and Technology/National Agency for Research and Innovation |
| EPI_ISL_435282 | RS Pondok Indah Hospital â€“ Pondok Indah | Eijkman Institute for Molecular Biology, Ministry of Research and Technology/National Agency for Research and Innovation |
| EPI_ISL_435283 | RS Pondok Indah Hospital â€“ Pondok Indah | Eijkman Institute for Molecular Biology, Ministry of Research and Technology/National Agency for Research and Innovation |

| EPI_Accession_ID | Originating Laboratory | Submitting Laboratory |
|---|---|---|
| EPI_ISL_435303 | National Hospital of Tropical Diseases | Oxford University Clinical Research Unit, Hanoi, Vietnam |
| EPI_ISL_435305 | National Hospital of Tropical Diseases | Oxford University Clinical Research Unit, Hanoi, Vietnam |
| EPI_ISL_435308 | National Hospital of Tropical Diseases | Oxford University Clinical Research Unit, Hanoi, Vietnam |
| EPI_ISL_435310 | National Hospital of Tropical Diseases | Oxford University Clinical Research Unit, Hanoi, Vietnam |
| EPI_ISL_435311 | National Hospital of Tropical Diseases | Oxford University Clinical Research Unit, Hanoi, Vietnam |
| EPI_ISL_435312 | National Hospital of Tropical Diseases | Oxford University Clinical Research Unit, Hanoi, Vietnam |
| EPI_ISL_435313 | National Hospital of Tropical Diseases | Oxford University Clinical Research Unit, Hanoi, Vietnam |
| EPI_ISL_435314 | National Hospital of Tropical Diseases | Oxford University Clinical Research Unit, Hanoi, Vietnam |
| EPI_ISL_435315 | National Hospital of Tropical Diseases | Oxford University Clinical Research Unit, Hanoi, Vietnam |
| EPI_ISL_435316 | National Hospital of Tropical Diseases | Oxford University Clinical Research Unit, Hanoi, Vietnam |
| EPI_ISL_435317 | National Hospital of Tropical Diseases | Oxford University Clinical Research Unit, Hanoi, Vietnam |
| EPI_ISL_435348 | Utah Public Health Laboratory | Utah Public Health Laboratory |
| EPI_ISL_435349 | Utah Public Health Laboratory | Utah Public Health Laboratory |
| EPI_ISL_435350 | Utah Public Health Laboratory | Utah Public Health Laboratory |
| EPI_ISL_435351 | Utah Public Health Laboratory | Utah Public Health Laboratory |
| EPI_ISL_435352 | Utah Public Health Laboratory | Utah Public Health Laboratory |
| EPI_ISL_435353 | Utah Public Health Laboratory | Utah Public Health Laboratory |
| EPI_ISL_435354 | Utah Public Health Laboratory | Utah Public Health Laboratory |
| EPI_ISL_435355 | Utah Public Health Laboratory | Utah Public Health Laboratory |
| EPI_ISL_435356 | Utah Public Health Laboratory | Utah Public Health Laboratory |
| EPI_ISL_435357 | Utah Public Health Laboratory | Utah Public Health Laboratory |
| EPI_ISL_435358 | Utah Public Health Laboratory | Utah Public Health Laboratory |

| EPI_Accession_ID | Originating Laboratory | Submitting Laboratory |
|---|---|---|
| EPI_ISL_435359 | Utah Public Health Laboratory | Utah Public Health Laboratory |
| EPI_ISL_435360 | Utah Public Health Laboratory | Utah Public Health Laboratory |
| EPI_ISL_435361 | Utah Public Health Laboratory | Utah Public Health Laboratory |
| EPI_ISL_435362 | Utah Public Health Laboratory | Utah Public Health Laboratory |
| EPI_ISL_435363 | Utah Public Health Laboratory | Utah Public Health Laboratory |
| EPI_ISL_435364 | Utah Public Health Laboratory | Utah Public Health Laboratory |
| EPI_ISL_435365 | Utah Public Health Laboratory | Utah Public Health Laboratory |
| EPI_ISL_435366 | Utah Public Health Laboratory | Utah Public Health Laboratory |
| EPI_ISL_435367 | Utah Public Health Laboratory | Utah Public Health Laboratory |
| EPI_ISL_435368 | Utah Public Health Laboratory | Utah Public Health Laboratory |
| EPI_ISL_435369 | Utah Public Health Laboratory | Utah Public Health Laboratory |
| EPI_ISL_435370 | Utah Public Health Laboratory | Utah Public Health Laboratory |
| EPI_ISL_435371 | Utah Public Health Laboratory | Utah Public Health Laboratory |
| EPI_ISL_435372 | Utah Public Health Laboratory | Utah Public Health Laboratory |
| EPI_ISL_435373 | Utah Public Health Laboratory | Utah Public Health Laboratory |
| EPI_ISL_435374 | Utah Public Health Laboratory | Utah Public Health Laboratory |
| EPI_ISL_435375 | Utah Public Health Laboratory | Utah Public Health Laboratory |
| EPI_ISL_435376 | Utah Public Health Laboratory | Utah Public Health Laboratory |
| EPI_ISL_435377 | Utah Public Health Laboratory | Utah Public Health Laboratory |
| EPI_ISL_435378 | Utah Public Health Laboratory | Utah Public Health Laboratory |
| EPI_ISL_435379 | Utah Public Health Laboratory | Utah Public Health Laboratory |
| EPI_ISL_435380 | Utah Public Health Laboratory | Utah Public Health Laboratory |
| EPI_ISL_435381 | Utah Public Health Laboratory | Utah Public Health Laboratory |
| EPI_ISL_435382 | Utah Public Health Laboratory | Utah Public Health Laboratory |
| EPI_ISL_435383 | Utah Public Health Laboratory | Utah Public Health Laboratory |
| EPI_ISL_435384 | Utah Public Health Laboratory | Utah Public Health Laboratory |
| EPI_ISL_435385 | Utah Public Health Laboratory | Utah Public Health Laboratory |
| EPI_ISL_435386 | Utah Public Health Laboratory | Utah Public Health Laboratory |
| EPI_ISL_435387 | Utah Public Health Laboratory | Utah Public Health Laboratory |
| EPI_ISL_435388 | Utah Public Health Laboratory | Utah Public Health Laboratory |
| EPI_ISL_435389 | Utah Public Health Laboratory | Utah Public Health Laboratory |
| EPI_ISL_435390 | Utah Public Health Laboratory | Utah Public Health Laboratory |
| EPI_ISL_435391 | Utah Public Health Laboratory | Utah Public Health Laboratory |

| EPI_Accession_ID | Originating Laboratory | Submitting Laboratory |
|---|---|---|
| EPI_ISL_435392 | Utah Public Health Laboratory | Utah Public Health Laboratory |
| EPI_ISL_435393 | Utah Public Health Laboratory | Utah Public Health Laboratory |
| EPI_ISL_435394 | Gundersen Molecular Diagnostics Laboratory | Kabara Cancer Research Institute |
| EPI_ISL_435395 | Gundersen Molecular Diagnostics Laboratory | Kabara Cancer Research Institute |
| EPI_ISL_435396 | Gundersen Molecular Diagnostics Laboratory | Kabara Cancer Research Institute |
| EPI_ISL_435397 | Gundersen Molecular Diagnostics Laboratory | Kabara Cancer Research Institute |
| EPI_ISL_435398 | Gundersen Molecular Diagnostics Laboratory | Kabara Cancer Research Institute |
| EPI_ISL_435399 | Gundersen Molecular Diagnostics Laboratory | Kabara Cancer Research Institute |
| EPI_ISL_435400 | Gundersen Molecular Diagnostics Laboratory | Kabara Cancer Research Institute |
| EPI_ISL_435401 | Gundersen Molecular Diagnostics Laboratory | Kabara Cancer Research Institute |
| EPI_ISL_435402 | Gundersen Molecular Diagnostics Laboratory | Kabara Cancer Research Institute |
| EPI_ISL_435403 | Virological Research Group, SzentÃ¡gothai Research Centre | Bioinformatics Research Group, SzentÃ¡gothai Research Centre |
| EPI_ISL_435404 | Virological Research Group, SzentÃ¡gothai Research Centre | Bioinformatics Research Group, SzentÃ¡gothai Research Centre |
| EPI_ISL_435405 | Virological Research Group, SzentÃ¡gothai Research Centre | Bioinformatics Research Group, SzentÃ¡gothai Research Centre |
| EPI_ISL_435406 | Virological Research Group, SzentÃ¡gothai Research Centre | Bioinformatics Research Group, SzentÃ¡gothai Research Centre |
| EPI_ISL_435407 | Virological Research Group, SzentÃ¡gothai Research Centre | Bioinformatics Research Group, SzentÃ¡gothai Research Centre |
| EPI_ISL_435408 | Virological Research Group, SzentÃ¡gothai Research Centre | Bioinformatics Research Group, SzentÃ¡gothai Research Centre |
| EPI_ISL_435409 | Virological Research Group, SzentÃ¡gothai Research Centre | Bioinformatics Research Group, SzentÃ¡gothai Research Centre |
| EPI_ISL_435410 | Virological Research Group, SzentÃ¡gothai Research Centre | Bioinformatics Research Group, SzentÃ¡gothai Research Centre |
| EPI_ISL_435411 | Virological Research Group, SzentÃ¡gothai Research Centre | Bioinformatics Research Group, SzentÃ¡gothai Research Centre |
| EPI_ISL_435412 | Virological Research Group, SzentÃ¡gothai Research Centre | Bioinformatics Research Group, SzentÃ¡gothai Research Centre |
| EPI_ISL_435413 | Virological Research Group, SzentÃ¡gothai Research Centre | Bioinformatics Research Group, SzentÃ¡gothai Research Centre |

| EPI_Accession_ID | Originating Laboratory | Submitting Laboratory |
|---|---|---|
| EPI_ISL_435414 | Virological Research Group, SzentÃ¡gothai Research Centre | Bioinformatics Research Group, SzentÃ¡gothai Research Centre |
| EPI_ISL_435415 | Virological Research Group, SzentÃ¡gothai Research Centre | Bioinformatics Research Group, SzentÃ¡gothai Research Centre |
| EPI_ISL_435416 | Virological Research Group, SzentÃ¡gothai Research Centre | Bioinformatics Research Group, SzentÃ¡gothai Research Centre |
| EPI_ISL_435417 | Virological Research Group, SzentÃ¡gothai Research Centre | Bioinformatics Research Group, SzentÃ¡gothai Research Centre |
| EPI_ISL_435418 | Virological Research Group, SzentÃ¡gothai Research Centre | Bioinformatics Research Group, SzentÃ¡gothai Research Centre |
| EPI_ISL_435419 | Virological Research Group, SzentÃ¡gothai Research Centre | Bioinformatics Research Group, SzentÃ¡gothai Research Centre |
| EPI_ISL_435420 | Virological Research Group, SzentÃ¡gothai Research Centre | Bioinformatics Research Group, SzentÃ¡gothai Research Centre |
| EPI_ISL_435421 | Virological Research Group, SzentÃ¡gothai Research Centre | Bioinformatics Research Group, SzentÃ¡gothai Research Centre |
| EPI_ISL_435422 | Virological Research Group, SzentÃ¡gothai Research Centre | Bioinformatics Research Group, SzentÃ¡gothai Research Centre |
| EPI_ISL_435423 | Virological Research Group, SzentÃ¡gothai Research Centre | Bioinformatics Research Group, SzentÃ¡gothai Research Centre |
| EPI_ISL_435424 | Virological Research Group, SzentÃ¡gothai Research Centre | Bioinformatics Research Group, SzentÃ¡gothai Research Centre |
| EPI_ISL_435425 | Virological Research Group, SzentÃ¡gothai Research Centre | Bioinformatics Research Group, SzentÃ¡gothai Research Centre |
| EPI_ISL_435426 | Virological Research Group, SzentÃ¡gothai Research Centre | Bioinformatics Research Group, SzentÃ¡gothai Research Centre |
| EPI_ISL_435427 | Virological Research Group, SzentÃ¡gothai Research Centre | Bioinformatics Research Group, SzentÃ¡gothai Research Centre |
| EPI_ISL_435428 | Virological Research Group, SzentÃ¡gothai Research Centre | Bioinformatics Research Group, SzentÃ¡gothai Research Centre |
| EPI_ISL_435429 | Virological Research Group, SzentÃ¡gothai Research Centre | Bioinformatics Research Group, SzentÃ¡gothai Research Centre |

| EPI_Accession_ID | Originating Laboratory | Submitting Laboratory |
|---|---|---|
| EPI_ISL_435430 | Virological Research Group, SzentÃ¡gothai Research Centre | Bioinformatics Research Group, SzentÃ¡gothai Research Centre |
| EPI_ISL_435431 | Virological Research Group, SzentÃ¡gothai Research Centre | Bioinformatics Research Group, SzentÃ¡gothai Research Centre |
| EPI_ISL_435441 | Alaska State Virology Laboratory | Alaska State Virology Laboratory |
| EPI_ISL_435442 | Alaska State Virology Laboratory | Alaska State Virology Laboratory |
| EPI_ISL_435443 | Alaska State Virology Laboratory | Alaska State Virology Laboratory |
| EPI_ISL_435444 | Alaska State Virology Laboratory | Alaska State Virology Laboratory |
| EPI_ISL_435445 | Robert Garry lab | Andersen lab at Scripps Research |
| EPI_ISL_435446 | Robert Garry lab | Andersen lab at Scripps Research |
| EPI_ISL_435447 | Robert Garry lab | Andersen lab at Scripps Research |
| EPI_ISL_435448 | Robert Garry lab | Andersen lab at Scripps Research |
| EPI_ISL_435449 | Robert Garry lab | Andersen lab at Scripps Research |
| EPI_ISL_435450 | Robert Garry lab | Andersen lab at Scripps Research |
| EPI_ISL_435451 | Robert Garry lab | Andersen lab at Scripps Research |
| EPI_ISL_435452 | Robert Garry lab | Andersen lab at Scripps Research |
| EPI_ISL_435453 | Robert Garry lab | Andersen lab at Scripps Research |
| EPI_ISL_435454 | Robert Garry lab | Andersen lab at Scripps Research |
| EPI_ISL_435455 | Robert Garry lab | Andersen lab at Scripps Research |
| EPI_ISL_435456 | Robert Garry lab | Andersen lab at Scripps Research |
| EPI_ISL_435457 | Robert Garry lab | Andersen lab at Scripps Research |
| EPI_ISL_435458 | Robert Garry lab | Andersen lab at Scripps Research |
| EPI_ISL_435459 | Robert Garry lab | Andersen lab at Scripps Research |
| EPI_ISL_435460 | Robert Garry lab | Andersen lab at Scripps Research |
| EPI_ISL_435461 | Robert Garry lab | Andersen lab at Scripps Research |
| EPI_ISL_435462 | Robert Garry lab | Andersen lab at Scripps Research |
| EPI_ISL_435463 | Robert Garry lab | Andersen lab at Scripps Research |
| EPI_ISL_435464 | Robert Garry lab | Andersen lab at Scripps Research |
| EPI_ISL_435465 | Robert Garry lab | Andersen lab at Scripps Research |
| EPI_ISL_435466 | Robert Garry lab | Andersen lab at Scripps Research |
| EPI_ISL_435467 | Robert Garry lab | Andersen lab at Scripps Research |
| EPI_ISL_435468 | Robert Garry lab | Andersen lab at Scripps Research |
| EPI_ISL_435469 | Robert Garry lab | Andersen lab at Scripps Research |

| EPI_Accession_ID | Originating Laboratory | Submitting Laboratory |
|---|---|---|
| EPI_ISL_435470 | Robert Garry lab | Andersen lab at Scripps Research |
| EPI_ISL_435471 | Robert Garry lab | Andersen lab at Scripps Research |
| EPI_ISL_435472 | Robert Garry lab | Andersen lab at Scripps Research |
| EPI_ISL_435473 | Rady's Childrens Hospital | Andersen lab at Scripps Research |
| EPI_ISL_435474 | Rady's Childrens Hospital | Andersen lab at Scripps Research |
| EPI_ISL_435475 | NYU Langone Health | Departments of Pathology and Medicine, New York University School of Medicine |
| EPI_ISL_435476 | NYU Langone Health | Departments of Pathology and Medicine, New York University School of Medicine |
| EPI_ISL_435477 | NYU Langone Health | Departments of Pathology and Medicine, New York University School of Medicine |
| EPI_ISL_435478 | NYU Langone Health | Departments of Pathology and Medicine, New York University School of Medicine |
| EPI_ISL_435479 | NYU Langone Health | Departments of Pathology and Medicine, New York University School of Medicine |
| EPI_ISL_435480 | NYU Langone Health | Departments of Pathology and Medicine, New York University School of Medicine |
| EPI_ISL_435481 | NYU Langone Health | Departments of Pathology and Medicine, New York University School of Medicine |
| EPI_ISL_435482 | NYU Langone Health | Departments of Pathology and Medicine, New York University School of Medicine |
| EPI_ISL_435483 | NYU Langone Health | Departments of Pathology and Medicine, New York University School of Medicine |
| EPI_ISL_435484 | NYU Langone Health | Departments of Pathology and Medicine, New York University School of Medicine |
| EPI_ISL_435485 | NYU Langone Health | Departments of Pathology and Medicine, New York University School of Medicine |
| EPI_ISL_435486 | NYU Langone Health | Departments of Pathology and Medicine, New York University School of Medicine |
| EPI_ISL_435487 | NYU Langone Health | Departments of Pathology and Medicine, New York University School of Medicine |
| EPI_ISL_435488 | NYU Langone Health | Departments of Pathology and Medicine, New York University School of Medicine |

| EPI_Accession_ID | Originating Laboratory | Submitting Laboratory |
|---|---|---|
| EPI_ISL_435489 | NYU Langone Health | Departments of Pathology and Medicine, New York University School of Medicine |
| EPI_ISL_435490 | NYU Langone Health | Departments of Pathology and Medicine, New York University School of Medicine |
| EPI_ISL_435491 | NYU Langone Health | Departments of Pathology and Medicine, New York University School of Medicine |
| EPI_ISL_435492 | NYU Langone Health | Departments of Pathology and Medicine, New York University School of Medicine |
| EPI_ISL_435493 | NYU Langone Health | Departments of Pathology and Medicine, New York University School of Medicine |
| EPI_ISL_435494 | NYU Langone Health | Departments of Pathology and Medicine, New York University School of Medicine |
| EPI_ISL_435495 | NYU Langone Health | Departments of Pathology and Medicine, New York University School of Medicine |
| EPI_ISL_435496 | NYU Langone Health | Departments of Pathology and Medicine, New York University School of Medicine |
| EPI_ISL_435497 | NYU Langone Health | Departments of Pathology and Medicine, New York University School of Medicine |
| EPI_ISL_435498 | NYU Langone Health | Departments of Pathology and Medicine, New York University School of Medicine |
| EPI_ISL_435499 | NYU Langone Health | Departments of Pathology and Medicine, New York University School of Medicine |
| EPI_ISL_435500 | NYU Langone Health | Departments of Pathology and Medicine, New York University School of Medicine |
| EPI_ISL_435501 | NYU Langone Health | Departments of Pathology and Medicine, New York University School of Medicine |
| EPI_ISL_435502 | NYU Langone Health | Departments of Pathology and Medicine, New York University School of Medicine |
| EPI_ISL_435503 | NYU Langone Health | Departments of Pathology and Medicine, New York University School of Medicine |
| EPI_ISL_435504 | NYU Langone Health | Departments of Pathology and Medicine, New York University School of Medicine |

| EPI_Accession_ID | Originating Laboratory | Submitting Laboratory |
|---|---|---|
| EPI_ISL_435505 | NYU Langone Health | Departments of Pathology and Medicine, New York University School of Medicine |
| EPI_ISL_435506 | NYU Langone Health | Departments of Pathology and Medicine, New York University School of Medicine |
| EPI_ISL_435507 | NYU Langone Health | Departments of Pathology and Medicine, New York University School of Medicine |
| EPI_ISL_435508 | NYU Langone Health | Departments of Pathology and Medicine, New York University School of Medicine |
| EPI_ISL_435509 | NYU Langone Health | Departments of Pathology and Medicine, New York University School of Medicine |
| EPI_ISL_435510 | NYU Langone Health | Departments of Pathology and Medicine, New York University School of Medicine |
| EPI_ISL_435511 | NYU Langone Health | Departments of Pathology and Medicine, New York University School of Medicine |
| EPI_ISL_435512 | NYU Langone Health | Departments of Pathology and Medicine, New York University School of Medicine |
| EPI_ISL_435513 | NYU Langone Health | Departments of Pathology and Medicine, New York University School of Medicine |
| EPI_ISL_435514 | NYU Langone Health | Departments of Pathology and Medicine, New York University School of Medicine |
| EPI_ISL_435515 | NYU Langone Health | Departments of Pathology and Medicine, New York University School of Medicine |
| EPI_ISL_435516 | NYU Langone Health | Departments of Pathology and Medicine, New York University School of Medicine |
| EPI_ISL_435517 | NYU Langone Health | Departments of Pathology and Medicine, New York University School of Medicine |
| EPI_ISL_435518 | NYU Langone Health | Departments of Pathology and Medicine, New York University School of Medicine |
| EPI_ISL_435519 | NYU Langone Health | Departments of Pathology and Medicine, New York University School of Medicine |
| EPI_ISL_435520 | NYU Langone Health | Departments of Pathology and Medicine, New York University School of Medicine |

| EPI_Accession_ID | Originating Laboratory | Submitting Laboratory |
|---|---|---|
| EPI_ISL_435521 | NYU Langone Health | Departments of Pathology and Medicine, New York University School of Medicine |
| EPI_ISL_435522 | NYU Langone Health | Departments of Pathology and Medicine, New York University School of Medicine |
| EPI_ISL_435523 | NYU Langone Health | Departments of Pathology and Medicine, New York University School of Medicine |
| EPI_ISL_435524 | NYU Langone Health | Departments of Pathology and Medicine, New York University School of Medicine |
| EPI_ISL_435525 | NYU Langone Health | Departments of Pathology and Medicine, New York University School of Medicine |
| EPI_ISL_435526 | NYU Langone Health | Departments of Pathology and Medicine, New York University School of Medicine |
| EPI_ISL_435527 | NYU Langone Health | Departments of Pathology and Medicine, New York University School of Medicine |
| EPI_ISL_435528 | NYU Langone Health | Departments of Pathology and Medicine, New York University School of Medicine |
| EPI_ISL_435529 | NYU Langone Health | Departments of Pathology and Medicine, New York University School of Medicine |
| EPI_ISL_435530 | NYU Langone Health | Departments of Pathology and Medicine, New York University School of Medicine |
| EPI_ISL_435531 | NYU Langone Health | Departments of Pathology and Medicine, New York University School of Medicine |
| EPI_ISL_435532 | NYU Langone Health | Departments of Pathology and Medicine, New York University School of Medicine |
| EPI_ISL_435533 | NYU Langone Health | Departments of Pathology and Medicine, New York University School of Medicine |
| EPI_ISL_435534 | NYU Langone Health | Departments of Pathology and Medicine, New York University School of Medicine |
| EPI_ISL_435535 | NYU Langone Health | Departments of Pathology and Medicine, New York University School of Medicine |
| EPI_ISL_435536 | NYU Langone Health | Departments of Pathology and Medicine, New York University School of Medicine |

| EPI_Accession_ID | Originating Laboratory | Submitting Laboratory |
|---|---|---|
| EPI_ISL_435537 | NYU Langone Health | Departments of Pathology and Medicine, New York University School of Medicine |
| EPI_ISL_435538 | NYU Langone Health | Departments of Pathology and Medicine, New York University School of Medicine |
| EPI_ISL_435539 | NYU Langone Health | Departments of Pathology and Medicine, New York University School of Medicine |
| EPI_ISL_435540 | NYU Langone Health | Departments of Pathology and Medicine, New York University School of Medicine |
| EPI_ISL_435541 | NYU Langone Health | Departments of Pathology and Medicine, New York University School of Medicine |
| EPI_ISL_435542 | NYU Langone Health | Departments of Pathology and Medicine, New York University School of Medicine |
| EPI_ISL_435543 | NYU Langone Health | Departments of Pathology and Medicine, New York University School of Medicine |
| EPI_ISL_435544 | NYU Langone Health | Departments of Pathology and Medicine, New York University School of Medicine |
| EPI_ISL_435545 | NYU Langone Health | Departments of Pathology and Medicine, New York University School of Medicine |
| EPI_ISL_435546 | NYU Langone Health | Departments of Pathology and Medicine, New York University School of Medicine |
| EPI_ISL_435547 | NYU Langone Health | Departments of Pathology and Medicine, New York University School of Medicine |
| EPI_ISL_435548 | NYU Langone Health | Departments of Pathology and Medicine, New York University School of Medicine |
| EPI_ISL_435549 | NYU Langone Health | Departments of Pathology and Medicine, New York University School of Medicine |
| EPI_ISL_435550 | LSUHS Emerging Viral Threat Laboratory | Microbial Genome Sequencing Center |
| EPI_ISL_435551 | LSUHS Emerging Viral Threat Laboratory | Microbial Genome Sequencing Center |
| EPI_ISL_435552 | LSUHS Emerging Viral Threat Laboratory | Microbial Genome Sequencing Center |
| EPI_ISL_435553 | LSUHS Emerging Viral Threat Laboratory | Microbial Genome Sequencing Center |
| EPI_ISL_435554 | LSUHS Emerging Viral Threat Laboratory | Microbial Genome Sequencing Center |
| EPI_ISL_435555 | LSUHS Emerging Viral Threat Laboratory | Microbial Genome Sequencing Center |
| EPI_ISL_435556 | LSUHS Emerging Viral Threat Laboratory | Microbial Genome Sequencing Center |

| EPI_Accession_ID | Originating Laboratory | Submitting Laboratory |
|---|---|---|
| EPI_ISL_435557 | LSUHS Emerging Viral Threat Laboratory | Microbial Genome Sequencing Center |
| EPI_ISL_435558 | LSUHS Emerging Viral Threat Laboratory | Microbial Genome Sequencing Center |
| EPI_ISL_435559 | LSUHS Emerging Viral Threat Laboratory | Microbial Genome Sequencing Center |
| EPI_ISL_435560 | LSUHS Emerging Viral Threat Laboratory | Microbial Genome Sequencing Center |
| EPI_ISL_435561 | LSUHS Emerging Viral Threat Laboratory | Microbial Genome Sequencing Center |
| EPI_ISL_435562 | LSUHS Emerging Viral Threat Laboratory | Microbial Genome Sequencing Center |
| EPI_ISL_435563 | LSUHS Emerging Viral Threat Laboratory | Microbial Genome Sequencing Center |
| EPI_ISL_435564 | LSUHS Emerging Viral Threat Laboratory | Microbial Genome Sequencing Center |
| EPI_ISL_435565 | LSUHS Emerging Viral Threat Laboratory | Microbial Genome Sequencing Center |
| EPI_ISL_435566 | LSUHS Emerging Viral Threat Laboratory | Microbial Genome Sequencing Center |
| EPI_ISL_435567 | LSUHS Emerging Viral Threat Laboratory | Microbial Genome Sequencing Center |
| EPI_ISL_435568 | LSUHS Emerging Viral Threat Laboratory | Microbial Genome Sequencing Center |
| EPI_ISL_435569 | LSUHS Emerging Viral Threat Laboratory | Microbial Genome Sequencing Center |
| EPI_ISL_435570 | LSUHS Emerging Viral Threat Laboratory | Microbial Genome Sequencing Center |
| EPI_ISL_435571 | LSUHS Emerging Viral Threat Laboratory | Microbial Genome Sequencing Center |
| EPI_ISL_435572 | LSUHS Emerging Viral Threat Laboratory | Microbial Genome Sequencing Center |
| EPI_ISL_435573 | LSUHS Emerging Viral Threat Laboratory | Microbial Genome Sequencing Center |
| EPI_ISL_435574 | LSUHS Emerging Viral Threat Laboratory | Microbial Genome Sequencing Center |
| EPI_ISL_435575 | LSUHS Emerging Viral Threat Laboratory | Microbial Genome Sequencing Center |
| EPI_ISL_435576 | LSUHS Emerging Viral Threat Laboratory | Microbial Genome Sequencing Center |
| EPI_ISL_435577 | LSUHS Emerging Viral Threat Laboratory | Microbial Genome Sequencing Center |
| EPI_ISL_435578 | LSUHS Emerging Viral Threat Laboratory | Microbial Genome Sequencing Center |
| EPI_ISL_435579 | LSUHS Emerging Viral Threat Laboratory | Microbial Genome Sequencing Center |
| EPI_ISL_435580 | Santa Clara County Public Health Department | Chiu Laboratory, University of California, San Francisco |
| EPI_ISL_435581 | Santa Clara County Public Health Department | Chiu Laboratory, University of California, San Francisco |
| EPI_ISL_435582 | Santa Clara County Public Health Department | Chiu Laboratory, University of California, San Francisco |
| EPI_ISL_435583 | Santa Clara County Public Health Department | Chiu Laboratory, University of California, San Francisco |
| EPI_ISL_435584 | Santa Clara County Public Health Department | Chiu Laboratory, University of California, San Francisco |
| EPI_ISL_435585 | Santa Clara County Public Health Department | Chiu Laboratory, University of California, San Francisco |
| EPI_ISL_435586 | Santa Clara County Public Health Department | Chiu Laboratory, University of California, San Francisco |
| EPI_ISL_435587 | Santa Clara County Public Health Department | Chiu Laboratory, University of California, San Francisco |
| EPI_ISL_435588 | Santa Clara County Public Health Department | Chiu Laboratory, University of California, San Francisco |
| EPI_ISL_435589 | Santa Clara County Public Health Department | Chiu Laboratory, University of California, San Francisco |

| EPI_Accession_ID | Originating Laboratory | Submitting Laboratory |
|---|---|---|
| EPI_ISL_435590 | Santa Clara County Public Health Department | Chiu Laboratory, University of California, San Francisco |
| EPI_ISL_435591 | Santa Clara County Public Health Department | Chiu Laboratory, University of California, San Francisco |
| EPI_ISL_435592 | Santa Clara County Public Health Department | Chiu Laboratory, University of California, San Francisco |
| EPI_ISL_435594 | Santa Clara County Public Health Department | Chiu Laboratory, University of California, San Francisco |
| EPI_ISL_435595 | Santa Clara County Public Health Department | Chiu Laboratory, University of California, San Francisco |
| EPI_ISL_435596 | Santa Clara County Public Health Department | Chiu Laboratory, University of California, San Francisco |
| EPI_ISL_435597 | Santa Clara County Public Health Department | Chiu Laboratory, University of California, San Francisco |
| EPI_ISL_435598 | Santa Clara County Public Health Department | Chiu Laboratory, University of California, San Francisco |
| EPI_ISL_435599 | Santa Clara County Public Health Department | Chiu Laboratory, University of California, San Francisco |
| EPI_ISL_435600 | Santa Clara County Public Health Department | Chiu Laboratory, University of California, San Francisco |
| EPI_ISL_435601 | Santa Clara County Public Health Department | Chiu Laboratory, University of California, San Francisco |
| EPI_ISL_435602 | Santa Clara County Public Health Department | Chiu Laboratory, University of California, San Francisco |
| EPI_ISL_435603 | Santa Clara County Public Health Department | Chiu Laboratory, University of California, San Francisco |
| EPI_ISL_435604 | Santa Clara County Public Health Department | Chiu Laboratory, University of California, San Francisco |
| EPI_ISL_435605 | Santa Clara County Public Health Department | Chiu Laboratory, University of California, San Francisco |
| EPI_ISL_435607 | Santa Clara County Public Health Department | Chiu Laboratory, University of California, San Francisco |
| EPI_ISL_435608 | Santa Clara County Public Health Department | Chiu Laboratory, University of California, San Francisco |
| EPI_ISL_435609 | Santa Clara County Public Health Department | Chiu Laboratory, University of California, San Francisco |
| EPI_ISL_435610 | Santa Clara County Public Health Department | Chiu Laboratory, University of California, San Francisco |
| EPI_ISL_435611 | Santa Clara County Public Health Department | Chiu Laboratory, University of California, San Francisco |
| EPI_ISL_435612 | Santa Clara County Public Health Department | Chiu Laboratory, University of California, San Francisco |
| EPI_ISL_435615 | Santa Clara County Public Health Department | Chiu Laboratory, University of California, San Francisco |
| EPI_ISL_435616 | Santa Clara County Public Health Department | Chiu Laboratory, University of California, San Francisco |
| EPI_ISL_435617 | Santa Clara County Public Health Department | Chiu Laboratory, University of California, San Francisco |
| EPI_ISL_435618 | Santa Clara County Public Health Department | Chiu Laboratory, University of California, San Francisco |
| EPI_ISL_435619 | Santa Clara County Public Health Department | Chiu Laboratory, University of California, San Francisco |
| EPI_ISL_435620 | Santa Clara County Public Health Department | Chiu Laboratory, University of California, San Francisco |
| EPI_ISL_435621 | Santa Clara County Public Health Department | Chiu Laboratory, University of California, San Francisco |
| EPI_ISL_435624 | Santa Clara County Public Health Department | Chiu Laboratory, University of California, San Francisco |
| EPI_ISL_435625 | Santa Clara County Public Health Department | Chiu Laboratory, University of California, San Francisco |
| EPI_ISL_435626 | Santa Clara County Public Health Department | Chiu Laboratory, University of California, San Francisco |
| EPI_ISL_435627 | Santa Clara County Public Health Department | Chiu Laboratory, University of California, San Francisco |
| EPI_ISL_435628 | Santa Clara County Public Health Department | Chiu Laboratory, University of California, San Francisco |

| EPI_Accession_ID | Originating Laboratory | Submitting Laboratory |
|---|---|---|
| EPI_ISL_435629 | Santa Clara County Public Health Department | Chiu Laboratory, University of California, San Francisco |
| EPI_ISL_435630 | Santa Clara County Public Health Department | Chiu Laboratory, University of California, San Francisco |
| EPI_ISL_435631 | Santa Clara County Public Health Department | Chiu Laboratory, University of California, San Francisco |
| EPI_ISL_435632 | Santa Clara County Public Health Department | Chiu Laboratory, University of California, San Francisco |
| EPI_ISL_435633 | Santa Clara County Public Health Department | Chiu Laboratory, University of California, San Francisco |
| EPI_ISL_435634 | Santa Clara County Public Health Department | Chiu Laboratory, University of California, San Francisco |
| EPI_ISL_435635 | Santa Clara County Public Health Department | Chiu Laboratory, University of California, San Francisco |
| EPI_ISL_435636 | Santa Clara County Public Health Department | Chiu Laboratory, University of California, San Francisco |
| EPI_ISL_435637 | Santa Clara County Public Health Department | Chiu Laboratory, University of California, San Francisco |
| EPI_ISL_435638 | Santa Clara County Public Health Department | Chiu Laboratory, University of California, San Francisco |
| EPI_ISL_435639 | Santa Clara County Public Health Department | Chiu Laboratory, University of California, San Francisco |
| EPI_ISL_435640 | Santa Clara County Public Health Department | Chiu Laboratory, University of California, San Francisco |
| EPI_ISL_435641 | Santa Clara County Public Health Department | Chiu Laboratory, University of California, San Francisco |
| EPI_ISL_435643 | Santa Clara County Public Health Department | Chiu Laboratory, University of California, San Francisco |
| EPI_ISL_435644 | Santa Clara County Public Health Department | Chiu Laboratory, University of California, San Francisco |
| EPI_ISL_435645 | Santa Clara County Public Health Department | Chiu Laboratory, University of California, San Francisco |
| EPI_ISL_435646 | Santa Clara County Public Health Department | Chiu Laboratory, University of California, San Francisco |
| EPI_ISL_435647 | Santa Clara County Public Health Department | Chiu Laboratory, University of California, San Francisco |
| EPI_ISL_435648 | Santa Clara County Public Health Department | Chiu Laboratory, University of California, San Francisco |
| EPI_ISL_435649 | Santa Clara County Public Health Department | Chiu Laboratory, University of California, San Francisco |
| EPI_ISL_435650 | Santa Clara County Public Health Department | Chiu Laboratory, University of California, San Francisco |
| EPI_ISL_435651 | Santa Clara County Public Health Department | Chiu Laboratory, University of California, San Francisco |
| EPI_ISL_435652 | Santa Clara County Public Health Department | Chiu Laboratory, University of California, San Francisco |
| EPI_ISL_435653 | Santa Clara County Public Health Department | Chiu Laboratory, University of California, San Francisco |
| EPI_ISL_435654 | Santa Clara County Public Health Department | Chiu Laboratory, University of California, San Francisco |
| EPI_ISL_435655 | Santa Clara County Public Health Department | Chiu Laboratory, University of California, San Francisco |
| EPI_ISL_435656 | Santa Clara County Public Health Department | Chiu Laboratory, University of California, San Francisco |
| EPI_ISL_435657 | Santa Clara County Public Health Department | Chiu Laboratory, University of California, San Francisco |
| EPI_ISL_435658 | Santa Clara County Public Health Department | Chiu Laboratory, University of California, San Francisco |
| EPI_ISL_435659 | Santa Clara County Public Health Department | Chiu Laboratory, University of California, San Francisco |
| EPI_ISL_435660 | Santa Clara County Public Health Department | Chiu Laboratory, University of California, San Francisco |
| EPI_ISL_435661 | Santa Clara County Public Health Department | Chiu Laboratory, University of California, San Francisco |
| EPI_ISL_435662 | Santa Clara County Public Health Department | Chiu Laboratory, University of California, San Francisco |

| EPI_Accession_ID | Originating Laboratory | Submitting Laboratory |
|---|---|---|
| EPI_ISL_435663 | Santa Clara County Public Health Department | Chiu Laboratory, University of California, San Francisco |
| EPI_ISL_435666 | Santa Clara County Public Health Department | Chiu Laboratory, University of California, San Francisco |
| EPI_ISL_435667 | Santa Clara County Public Health Department | Chiu Laboratory, University of California, San Francisco |
| EPI_ISL_435668 | Santa Clara County Public Health Department | Chiu Laboratory, University of California, San Francisco |
| EPI_ISL_435669 | Santa Clara County Public Health Department | Chiu Laboratory, University of California, San Francisco |
| EPI_ISL_435670 | Santa Clara County Public Health Department | Chiu Laboratory, University of California, San Francisco |
| EPI_ISL_435671 | Santa Clara County Public Health Department | Chiu Laboratory, University of California, San Francisco |
| EPI_ISL_435673 | Santa Clara County Public Health Department | Chiu Laboratory, University of California, San Francisco |
| EPI_ISL_435674 | National Virology Reference Laboratory | National Public Health Laboratory, National Centre for Infectious Diseases |
| EPI_ISL_435675 | National Virology Reference Laboratory | National Public Health Laboratory, National Centre for Infectious Diseases |
| EPI_ISL_435676 | National Virology Reference Laboratory | National Public Health Laboratory, National Centre for Infectious Diseases |
| EPI_ISL_435677 | National Virology Reference Laboratory | National Public Health Laboratory, National Centre for Infectious Diseases |
| EPI_ISL_435678 | National Public Health Laboratory, National Centre for Infectious Diseases | National Public Health Laboratory, National Centre for Infectious Diseases |
| EPI_ISL_435679 | National Public Health Laboratory, National Centre for Infectious Diseases | National Public Health Laboratory, National Centre for Infectious Diseases |
| EPI_ISL_435680 | National Public Health Laboratory, National Centre for Infectious Diseases | National Public Health Laboratory, National Centre for Infectious Diseases |
| EPI_ISL_435681 | National Public Health Laboratory, National Centre for Infectious Diseases | National Public Health Laboratory, National Centre for Infectious Diseases |
| EPI_ISL_435682 | National Public Health Laboratory, National Centre for Infectious Diseases | National Public Health Laboratory, National Centre for Infectious Diseases |
| EPI_ISL_435683 | National Public Health Laboratory, National Centre for Infectious Diseases | National Public Health Laboratory, National Centre for Infectious Diseases |
| EPI_ISL_435684 | National Public Health Laboratory, National Centre for Infectious Diseases | National Public Health Laboratory, National Centre for Infectious Diseases |
| EPI_ISL_435685 | National Public Health Laboratory, National Centre for Infectious Diseases | National Public Health Laboratory, National Centre for Infectious Diseases |

| EPI_Accession_ID | Originating Laboratory | Submitting Laboratory |
|---|---|---|
| EPI_ISL_435686 | National Public Health Laboratory, National Centre for Infectious Diseases | National Public Health Laboratory, National Centre for Infectious Diseases |
| EPI_ISL_435687 | National Public Health Laboratory, National Centre for Infectious Diseases | National Public Health Laboratory, National Centre for Infectious Diseases |
| EPI_ISL_435688 | National Public Health Laboratory, National Centre for Infectious Diseases | National Public Health Laboratory, National Centre for Infectious Diseases |
| EPI_ISL_435689 | National Public Health Laboratory, National Centre for Infectious Diseases | National Public Health Laboratory, National Centre for Infectious Diseases |
| EPI_ISL_435690 | National Public Health Laboratory, National Centre for Infectious Diseases | National Public Health Laboratory, National Centre for Infectious Diseases |
| EPI_ISL_435691 | National Public Health Laboratory, National Centre for Infectious Diseases | National Public Health Laboratory, National Centre for Infectious Diseases |
| EPI_ISL_435692 | National Public Health Laboratory, National Centre for Infectious Diseases | National Public Health Laboratory, National Centre for Infectious Diseases |
| EPI_ISL_435693 | National Public Health Laboratory, National Centre for Infectious Diseases | National Public Health Laboratory, National Centre for Infectious Diseases |
| EPI_ISL_435694 | National Public Health Laboratory, National Centre for Infectious Diseases | National Public Health Laboratory, National Centre for Infectious Diseases |
| EPI_ISL_435695 | National Public Health Laboratory, National Centre for Infectious Diseases | National Public Health Laboratory, National Centre for Infectious Diseases |
| EPI_ISL_435696 | National Public Health Laboratory, National Centre for Infectious Diseases | National Public Health Laboratory, National Centre for Infectious Diseases |
| EPI_ISL_435697 | National Public Health Laboratory, National Centre for Infectious Diseases | National Public Health Laboratory, National Centre for Infectious Diseases |
| EPI_ISL_435698 | National Public Health Laboratory, National Centre for Infectious Diseases | National Public Health Laboratory, National Centre for Infectious Diseases |
| EPI_ISL_435699 | National Public Health Laboratory, National Centre for Infectious Diseases | National Public Health Laboratory, National Centre for Infectious Diseases |
| EPI_ISL_435700 | National Public Health Laboratory, National Centre for Infectious Diseases | National Public Health Laboratory, National Centre for Infectious Diseases |
| EPI_ISL_435702 | Yale COVID-19 Biorepository | Grubaugh Lab - Yale School of Public Health |
| EPI_ISL_435703 | Yale COVID-19 Biorepository | Grubaugh Lab - Yale School of Public Health |
| EPI_ISL_435704 | Yale COVID-19 Biorepository | Grubaugh Lab - Yale School of Public Health |

| EPI_Accession_ID | Originating Laboratory | Submitting Laboratory |
|---|---|---|
| EPI_ISL_435705 | Yale COVID-19 Biorepository | Grubaugh Lab - Yale School of Public Health |
| EPI_ISL_435706 | Yale COVID-19 Biorepository | Grubaugh Lab - Yale School of Public Health |
| EPI_ISL_435707 | Yale COVID-19 Biorepository | Grubaugh Lab - Yale School of Public Health |
| EPI_ISL_435708 | Yale COVID-19 Biorepository | Grubaugh Lab - Yale School of Public Health |
| EPI_ISL_435709 | Yale COVID-19 Biorepository | Grubaugh Lab - Yale School of Public Health |
| EPI_ISL_435710 | Connecticut State Department of Public Health | Grubaugh Lab - Yale School of Public Health |
| EPI_ISL_435711 | Connecticut State Department of Public Health | Grubaugh Lab - Yale School of Public Health |
| EPI_ISL_435712 | Connecticut State Department of Public Health | Grubaugh Lab - Yale School of Public Health |
| EPI_ISL_435713 | Connecticut State Department of Public Health | Grubaugh Lab - Yale School of Public Health |
| EPI_ISL_435714 | Connecticut State Department of Public Health | Grubaugh Lab - Yale School of Public Health |
| EPI_ISL_435715 | Connecticut State Department of Public Health | Grubaugh Lab - Yale School of Public Health |
| EPI_ISL_435716 | Connecticut State Department of Public Health | Grubaugh Lab - Yale School of Public Health |
| EPI_ISL_435717 | Connecticut State Department of Public Health | Grubaugh Lab - Yale School of Public Health |
| EPI_ISL_435718 | Connecticut State Department of Public Health | Grubaugh Lab - Yale School of Public Health |
| EPI_ISL_435719 | Connecticut State Department of Public Health | Grubaugh Lab - Yale School of Public Health |
| EPI_ISL_435720 | Yale Clinical virology | Grubaugh Lab - Yale School of Public Health |
| EPI_ISL_435721 | NYU Langone Health | Departments of Pathology and Medicine, New York University School of Medicine |
| EPI_ISL_435722 | NYU Langone Health | Departments of Pathology and Medicine, New York University School of Medicine |
| EPI_ISL_436040 | DC Public Health Lab Dept of Forensic Science | Pathogen Discovery, Respiratory Viruses Branch, Division of Viral Diseases, Centers for Disease Control and Prevention |
| EPI_ISL_436041 | DC Public Health Lab Dept of Forensic Science | Pathogen Discovery, Respiratory Viruses Branch, Division of Viral Diseases, Centers for Disease Control and Prevention |
| EPI_ISL_436042 | DC Public Health Lab Dept of Forensic Science | Pathogen Discovery, Respiratory Viruses Branch, Division of Viral Diseases, Centers for Disease Control and Prevention |
| EPI_ISL_436043 | DC Public Health Lab Dept of Forensic Science | Pathogen Discovery, Respiratory Viruses Branch, Division of Viral Diseases, Centers for Disease Control and Prevention |

| EPI_Accession_ID | Originating Laboratory | Submitting Laboratory |
|---|---|---|
| EPI_ISL_436044 | Louisiana Office of Public Health Laboratories | Pathogen Discovery, Respiratory Viruses Branch, Division of Viral Diseases, Centers for Disease Control and Prevention |
| EPI_ISL_436045 | US VI Department of Health | Pathogen Discovery, Respiratory Viruses Branch, Division of Viral Diseases, Centers for Disease Control and Prevention |
| EPI_ISL_436046 | US VI Department of Health | Pathogen Discovery, Respiratory Viruses Branch, Division of Viral Diseases, Centers for Disease Control and Prevention |
| EPI_ISL_436047 | NYC Department of Health and Mental Hygiene | Pathogen Discovery, Respiratory Viruses Branch, Division of Viral Diseases, Centers for Disease Control and Prevention |
| EPI_ISL_436048 | NYC Department of Health and Mental Hygiene | Pathogen Discovery, Respiratory Viruses Branch, Division of Viral Diseases, Centers for Disease Control and Prevention |
| EPI_ISL_436049 | NYC Department of Health and Mental Hygiene | Pathogen Discovery, Respiratory Viruses Branch, Division of Viral Diseases, Centers for Disease Control and Prevention |
| EPI_ISL_436050 | NYC Department of Health and Mental Hygiene | Pathogen Discovery, Respiratory Viruses Branch, Division of Viral Diseases, Centers for Disease Control and Prevention |
| EPI_ISL_436051 | NYC Department of Health and Mental Hygiene | Pathogen Discovery, Respiratory Viruses Branch, Division of Viral Diseases, Centers for Disease Control and Prevention |
| EPI_ISL_436052 | NYC Department of Health and Mental Hygiene | Pathogen Discovery, Respiratory Viruses Branch, Division of Viral Diseases, Centers for Disease Control and Prevention |
| EPI_ISL_436053 | NYC Department of Health and Mental Hygiene | Pathogen Discovery, Respiratory Viruses Branch, Division of Viral Diseases, Centers for Disease Control and Prevention |
| EPI_ISL_436054 | NYC Department of Health and Mental Hygiene | Pathogen Discovery, Respiratory Viruses Branch, Division of Viral Diseases, Centers for Disease Control and Prevention |

| EPI_Accession_ID | Originating Laboratory | Submitting Laboratory |
|---|---|---|
| EPI_ISL_436055 | NYC Department of Health and Mental Hygiene | Pathogen Discovery, Respiratory Viruses Branch, Division of Viral Diseases, Centers for Disease Control and Prevention |
| EPI_ISL_436056 | NYC Department of Health and Mental Hygiene | Pathogen Discovery, Respiratory Viruses Branch, Division of Viral Diseases, Centers for Disease Control and Prevention |
| EPI_ISL_436057 | NYC Department of Health and Mental Hygiene | Pathogen Discovery, Respiratory Viruses Branch, Division of Viral Diseases, Centers for Disease Control and Prevention |
| EPI_ISL_436058 | NYC Department of Health and Mental Hygiene | Pathogen Discovery, Respiratory Viruses Branch, Division of Viral Diseases, Centers for Disease Control and Prevention |
| EPI_ISL_436059 | NYC Department of Health and Mental Hygiene | Pathogen Discovery, Respiratory Viruses Branch, Division of Viral Diseases, Centers for Disease Control and Prevention |
| EPI_ISL_436060 | NYC Department of Health and Mental Hygiene | Pathogen Discovery, Respiratory Viruses Branch, Division of Viral Diseases, Centers for Disease Control and Prevention |
| EPI_ISL_436061 | NYC Department of Health and Mental Hygiene | Pathogen Discovery, Respiratory Viruses Branch, Division of Viral Diseases, Centers for Disease Control and Prevention |
| EPI_ISL_436062 | NYC Department of Health and Mental Hygiene | Pathogen Discovery, Respiratory Viruses Branch, Division of Viral Diseases, Centers for Disease Control and Prevention |
| EPI_ISL_436063 | NYC Department of Health and Mental Hygiene | Pathogen Discovery, Respiratory Viruses Branch, Division of Viral Diseases, Centers for Disease Control and Prevention |
| EPI_ISL_436064 | NYC Department of Health and Mental Hygiene | Pathogen Discovery, Respiratory Viruses Branch, Division of Viral Diseases, Centers for Disease Control and Prevention |
| EPI_ISL_436065 | NYC Department of Health and Mental Hygiene | Pathogen Discovery, Respiratory Viruses Branch, Division of Viral Diseases, Centers for Disease Control and Prevention |

| EPI_Accession_ID | Originating Laboratory | Submitting Laboratory |
|---|---|---|
| EPI_ISL_436066 | NYC Department of Health and Mental Hygiene | Pathogen Discovery, Respiratory Viruses Branch, Division of Viral Diseases, Centers for Disease Control and Prevention |
| EPI_ISL_436067 | NYC Department of Health and Mental Hygiene | Pathogen Discovery, Respiratory Viruses Branch, Division of Viral Diseases, Centers for Disease Control and Prevention |
| EPI_ISL_436068 | NYC Department of Health and Mental Hygiene | Pathogen Discovery, Respiratory Viruses Branch, Division of Viral Diseases, Centers for Disease Control and Prevention |
| EPI_ISL_436069 | NYC Department of Health and Mental Hygiene | Pathogen Discovery, Respiratory Viruses Branch, Division of Viral Diseases, Centers for Disease Control and Prevention |
| EPI_ISL_436070 | NYC Department of Health and Mental Hygiene | Pathogen Discovery, Respiratory Viruses Branch, Division of Viral Diseases, Centers for Disease Control and Prevention |
| EPI_ISL_436071 | NYC Department of Health and Mental Hygiene | Pathogen Discovery, Respiratory Viruses Branch, Division of Viral Diseases, Centers for Disease Control and Prevention |
| EPI_ISL_436072 | NYC Department of Health and Mental Hygiene | Pathogen Discovery, Respiratory Viruses Branch, Division of Viral Diseases, Centers for Disease Control and Prevention |
| EPI_ISL_436073 | NYC Department of Health and Mental Hygiene | Pathogen Discovery, Respiratory Viruses Branch, Division of Viral Diseases, Centers for Disease Control and Prevention |
| EPI_ISL_436074 | NYC Department of Health and Mental Hygiene | Pathogen Discovery, Respiratory Viruses Branch, Division of Viral Diseases, Centers for Disease Control and Prevention |
| EPI_ISL_436075 | NYC Department of Health and Mental Hygiene | Pathogen Discovery, Respiratory Viruses Branch, Division of Viral Diseases, Centers for Disease Control and Prevention |
| EPI_ISL_436076 | NYC Department of Health and Mental Hygiene | Pathogen Discovery, Respiratory Viruses Branch, Division of Viral Diseases, Centers for Disease Control and Prevention |

| EPI_Accession_ID | Originating Laboratory | Submitting Laboratory |
|---|---|---|
| EPI_ISL_436077 | NYC Department of Health and Mental Hygiene | Pathogen Discovery, Respiratory Viruses Branch, Division of Viral Diseases, Centers for Disease Control and Prevention |
| EPI_ISL_436078 | NYC Department of Health and Mental Hygiene | Pathogen Discovery, Respiratory Viruses Branch, Division of Viral Diseases, Centers for Disease Control and Prevention |
| EPI_ISL_436079 | NYC Department of Health and Mental Hygiene | Pathogen Discovery, Respiratory Viruses Branch, Division of Viral Diseases, Centers for Disease Control and Prevention |
| EPI_ISL_436080 | NYC Department of Health and Mental Hygiene | Pathogen Discovery, Respiratory Viruses Branch, Division of Viral Diseases, Centers for Disease Control and Prevention |
| EPI_ISL_436081 | NYC Department of Health and Mental Hygiene | Pathogen Discovery, Respiratory Viruses Branch, Division of Viral Diseases, Centers for Disease Control and Prevention |
| EPI_ISL_436082 | NYC Department of Health and Mental Hygiene | Pathogen Discovery, Respiratory Viruses Branch, Division of Viral Diseases, Centers for Disease Control and Prevention |
| EPI_ISL_436097 | Prince Charles Hospital | Public Health Virology Laboratory, Forensics and Scientific Services, Queensland Health |
| EPI_ISL_436098 | Royal Brisbane and Women's Hospital | Public Health Virology Laboratory, Forensic and Scientific Services, Queensland Health |
| EPI_ISL_436099 | TSGH-CP molecular lab | TSGH-CP molecular lab |
| EPI_ISL_436100 | TSGH-CP molecular lab | TSGH-CP molecular lab |
| EPI_ISL_436101 | TSGH-CP molecular lab | TSGH-CP molecular lab |
| EPI_ISL_436102 | TSGH-CP molecular lab | TSGH-CP molecular lab |
| EPI_ISL_436103 | TSGH-CP molecular lab | TSGH-CP molecular lab |
| EPI_ISL_436104 | TSGH-CP molecular lab | TSGH-CP molecular lab |
| EPI_ISL_436105 | TSGH-CP molecular lab | TSGH-CP molecular lab |
| EPI_ISL_436106 | TSGH-CP molecular lab | TSGH-CP molecular lab |
| EPI_ISL_436107 | TSGH-CP molecular lab | TSGH-CP molecular lab |
| EPI_ISL_436108 | TSGH-CP molecular lab | TSGH-CP molecular lab |

| EPI_Accession_ID | Originating Laboratory | Submitting Laboratory |
|---|---|---|
| EPI_ISL_436111 | Victorian Infectious Diseases Reference Laboratory (VIDRL) | Microbiological Diagnostic Unit Public Health Laboratory and Victorian Infectious Diseases Reference Laboratory, Doherty Institute |
| EPI_ISL_436112 | Victorian Infectious Diseases Reference Laboratory (VIDRL) | Microbiological Diagnostic Unit Public Health Laboratory and Victorian Infectious Diseases Reference Laboratory, Doherty Institute |
| EPI_ISL_436113 | Victorian Infectious Diseases Reference Laboratory (VIDRL) | Microbiological Diagnostic Unit Public Health Laboratory and Victorian Infectious Diseases Reference Laboratory, Doherty Institute |
| EPI_ISL_436114 | Victorian Infectious Diseases Reference Laboratory (VIDRL) | Microbiological Diagnostic Unit Public Health Laboratory and Victorian Infectious Diseases Reference Laboratory, Doherty Institute |
| EPI_ISL_436115 | Victorian Infectious Diseases Reference Laboratory (VIDRL) | Microbiological Diagnostic Unit Public Health Laboratory and Victorian Infectious Diseases Reference Laboratory, Doherty Institute |
| EPI_ISL_436116 | Victorian Infectious Diseases Reference Laboratory (VIDRL) | Microbiological Diagnostic Unit Public Health Laboratory and Victorian Infectious Diseases Reference Laboratory, Doherty Institute |
| EPI_ISL_436117 | Victorian Infectious Diseases Reference Laboratory (VIDRL) | Microbiological Diagnostic Unit Public Health Laboratory and Victorian Infectious Diseases Reference Laboratory, Doherty Institute |
| EPI_ISL_436118 | Victorian Infectious Diseases Reference Laboratory (VIDRL) | Microbiological Diagnostic Unit Public Health Laboratory and Victorian Infectious Diseases Reference Laboratory, Doherty Institute |
| EPI_ISL_436119 | Victorian Infectious Diseases Reference Laboratory (VIDRL) | Microbiological Diagnostic Unit Public Health Laboratory and Victorian Infectious Diseases Reference Laboratory, Doherty Institute |
| EPI_ISL_436120 | Victorian Infectious Diseases Reference Laboratory (VIDRL) | Microbiological Diagnostic Unit Public Health Laboratory and Victorian Infectious Diseases Reference Laboratory, Doherty Institute |
| EPI_ISL_436121 | Victorian Infectious Diseases Reference Laboratory (VIDRL) | Microbiological Diagnostic Unit Public Health Laboratory and Victorian Infectious Diseases Reference Laboratory, Doherty Institute |

| EPI_Accession_ID | Originating Laboratory | Submitting Laboratory |
|---|---|---|
| EPI_ISL_436122 | Victorian Infectious Diseases Reference Laboratory (VIDRL) | Microbiological Diagnostic Unit Public Health Laboratory and Victorian Infectious Diseases Reference Laboratory, Doherty Institute |
| EPI_ISL_436123 | Victorian Infectious Diseases Reference Laboratory (VIDRL) | Microbiological Diagnostic Unit Public Health Laboratory and Victorian Infectious Diseases Reference Laboratory, Doherty Institute |
| EPI_ISL_436124 | Victorian Infectious Diseases Reference Laboratory (VIDRL) | Microbiological Diagnostic Unit Public Health Laboratory and Victorian Infectious Diseases Reference Laboratory, Doherty Institute |
| EPI_ISL_436125 | Victorian Infectious Diseases Reference Laboratory (VIDRL) | Microbiological Diagnostic Unit Public Health Laboratory and Victorian Infectious Diseases Reference Laboratory, Doherty Institute |
| EPI_ISL_436126 | Victorian Infectious Diseases Reference Laboratory (VIDRL) | Microbiological Diagnostic Unit Public Health Laboratory and Victorian Infectious Diseases Reference Laboratory, Doherty Institute |
| EPI_ISL_436127 | Victorian Infectious Diseases Reference Laboratory (VIDRL) | Microbiological Diagnostic Unit Public Health Laboratory and Victorian Infectious Diseases Reference Laboratory, Doherty Institute |
| EPI_ISL_436128 | Victorian Infectious Diseases Reference Laboratory (VIDRL) | Microbiological Diagnostic Unit Public Health Laboratory and Victorian Infectious Diseases Reference Laboratory, Doherty Institute |
| EPI_ISL_436129 | Victorian Infectious Diseases Reference Laboratory (VIDRL) | Microbiological Diagnostic Unit Public Health Laboratory and Victorian Infectious Diseases Reference Laboratory, Doherty Institute |
| EPI_ISL_436130 | Victorian Infectious Diseases Reference Laboratory (VIDRL) | Microbiological Diagnostic Unit Public Health Laboratory and Victorian Infectious Diseases Reference Laboratory, Doherty Institute |
| EPI_ISL_436131 | Victorian Infectious Diseases Reference Laboratory (VIDRL) | Microbiological Diagnostic Unit Public Health Laboratory and Victorian Infectious Diseases Reference Laboratory, Doherty Institute |
| EPI_ISL_436132 | Victorian Infectious Diseases Reference Laboratory (VIDRL) | Microbiological Diagnostic Unit Public Health Laboratory and Victorian Infectious Diseases Reference Laboratory, Doherty Institute |

| EPI_Accession_ID | Originating Laboratory | Submitting Laboratory |
|---|---|---|
| EPI_ISL_436137 | District Surveillance Unit | Department of Neurovirology, National Institute of Mental Health and Neuroscience (NIMHANS) |
| EPI_ISL_436138 | District Surveillance Unit | Department of Neurovirology, National Institute of Mental Health and Neuroscience (NIMHANS) |
| EPI_ISL_436139 | District Surveillance Unit | Department of Neurovirology, National Institute of Mental Health and Neuroscience (NIMHANS) |
| EPI_ISL_436140 | District Surveillance Unit | Department of Neurovirology, National Institute of Mental Health and Neuroscience (NIMHANS) |
| EPI_ISL_436141 | District Surveillance Unit | Department of Neurovirology, National Institute of Mental Health and Neuroscience (NIMHANS) |
| EPI_ISL_436156 | District Surveillance Unit | Department of Neurovirology, National Institute of Mental Health and Neuroscience (NIMHANS) |
| EPI_ISL_436157 | District Surveillance Unit | Department of Neurovirology, National Institute of Mental Health and Neuroscience (NIMHANS) |
| EPI_ISL_436194 | Viral Respiratory Lab, National Institute for Biomedical Research (INRB) | Pathogen Sequencing Lab, National Institute for Biomedical Research (INRB) |
| EPI_ISL_436195 | Servicio de MicrobiologÃ-a. Consorcio Hospital General Universitario de Valencia | Sequencing and Bioinformatics Service and Molecular Epidemiology Research Group. FISABIO-Public Health |
| EPI_ISL_436196 | Servicio de MicrobiologÃ-a. Consorcio Hospital General Universitario de Valencia | Sequencing and Bioinformatics Service and Molecular Epidemiology Research Group. FISABIO-Public Health |
| EPI_ISL_436197 | Servicio de MicrobiologÃ-a. Consorcio Hospital General Universitario de Valencia | Sequencing and Bioinformatics Service and Molecular Epidemiology Research Group. FISABIO-Public Health |
| EPI_ISL_436198 | Servicio de MicrobiologÃ-a. Consorcio Hospital General Universitario de Valencia | Sequencing and Bioinformatics Service and Molecular Epidemiology Research Group. FISABIO-Public Health |
| EPI_ISL_436199 | Servicio de MicrobiologÃ-a. Consorcio Hospital General Universitario de Valencia | Sequencing and Bioinformatics Service and Molecular Epidemiology Research Group. FISABIO-Public Health |
| EPI_ISL_436200 | Servicio de MicrobiologÃ-a. Consorcio Hospital General Universitario de Valencia | Sequencing and Bioinformatics Service and Molecular Epidemiology Research Group. FISABIO-Public Health |
| EPI_ISL_436201 | Servicio de MicrobiologÃ-a. Consorcio Hospital General Universitario de Valencia | Sequencing and Bioinformatics Service and Molecular Epidemiology Research Group. FISABIO-Public Health |
| EPI_ISL_436202 | Servicio de MicrobiologÃ-a. Consorcio Hospital General Universitario de Valencia | Sequencing and Bioinformatics Service and Molecular Epidemiology Research Group. FISABIO-Public Health |

| EPI_Accession_ID | Originating Laboratory | Submitting Laboratory |
|---|---|---|
| EPI_ISL_436203 | Servicio de MicrobiologÃ-a. Consorcio Hospital General Universitario de Valencia | Sequencing and Bioinformatics Service and Molecular Epidemiology Research Group. FISABIO-Public Health |
| EPI_ISL_436204 | Servicio de MicrobiologÃ-a. Consorcio Hospital General Universitario de Valencia | Sequencing and Bioinformatics Service and Molecular Epidemiology Research Group. FISABIO-Public Health |
| EPI_ISL_436205 | Servicio de MicrobiologÃ-a. Consorcio Hospital General Universitario de Valencia | Sequencing and Bioinformatics Service and Molecular Epidemiology Research Group. FISABIO-Public Health |
| EPI_ISL_436206 | Servicio de MicrobiologÃ-a. Consorcio Hospital General Universitario de Valencia | Sequencing and Bioinformatics Service and Molecular Epidemiology Research Group. FISABIO-Public Health |
| EPI_ISL_436207 | Servicio de MicrobiologÃ-a. Consorcio Hospital General Universitario de Valencia | Sequencing and Bioinformatics Service and Molecular Epidemiology Research Group. FISABIO-Public Health |
| EPI_ISL_436208 | Servicio de MicrobiologÃ-a. Consorcio Hospital General Universitario de Valencia | Sequencing and Bioinformatics Service and Molecular Epidemiology Research Group. FISABIO-Public Health |
| EPI_ISL_436209 | Servicio de MicrobiologÃ-a. Consorcio Hospital General Universitario de Valencia | Sequencing and Bioinformatics Service and Molecular Epidemiology Research Group. FISABIO-Public Health |
| EPI_ISL_436210 | Servicio de MicrobiologÃ-a. Consorcio Hospital General Universitario de Valencia | Sequencing and Bioinformatics Service and Molecular Epidemiology Research Group. FISABIO-Public Health |
| EPI_ISL_436211 | Servicio de MicrobiologÃ-a. Consorcio Hospital General Universitario de Valencia | Sequencing and Bioinformatics Service and Molecular Epidemiology Research Group. FISABIO-Public Health |
| EPI_ISL_436212 | Servicio de MicrobiologÃ-a. Consorcio Hospital General Universitario de Valencia | Sequencing and Bioinformatics Service and Molecular Epidemiology Research Group. FISABIO-Public Health |
| EPI_ISL_436213 | Servicio de MicrobiologÃ-a. Consorcio Hospital General Universitario de Valencia | Sequencing and Bioinformatics Service and Molecular Epidemiology Research Group. FISABIO-Public Health |
| EPI_ISL_436214 | Servicio de MicrobiologÃ-a. Consorcio Hospital General Universitario de Valencia | Sequencing and Bioinformatics Service and Molecular Epidemiology Research Group. FISABIO-Public Health |
| EPI_ISL_436215 | Servicio de MicrobiologÃ-a. Consorcio Hospital General Universitario de Valencia | Sequencing and Bioinformatics Service and Molecular Epidemiology Research Group. FISABIO-Public Health |
| EPI_ISL_436216 | Servicio de MicrobiologÃ-a. Consorcio Hospital General Universitario de Valencia | Sequencing and Bioinformatics Service and Molecular Epidemiology Research Group. FISABIO-Public Health |
| EPI_ISL_436217 | Servicio de MicrobiologÃ-a. Consorcio Hospital General Universitario de Valencia | Sequencing and Bioinformatics Service and Molecular Epidemiology Research Group. FISABIO-Public Health |
| EPI_ISL_436218 | Servicio de MicrobiologÃ-a. Consorcio Hospital General Universitario de Valencia | Sequencing and Bioinformatics Service and Molecular Epidemiology Research Group. FISABIO-Public Health |

| EPI_Accession_ID | Originating Laboratory | Submitting Laboratory |
|---|---|---|
| EPI_ISL_436219 | Servicio de MicrobiologÃ-a. Consorcio Hospital General Universitario de Valencia | Sequencing and Bioinformatics Service and Molecular Epidemiology Research Group. FISABIO-Public Health |
| EPI_ISL_436220 | Servicio de MicrobiologÃ-a. Consorcio Hospital General Universitario de Valencia | Sequencing and Bioinformatics Service and Molecular Epidemiology Research Group. FISABIO-Public Health |
| EPI_ISL_436221 | Servicio de MicrobiologÃ-a. Consorcio Hospital General Universitario de Valencia | Sequencing and Bioinformatics Service and Molecular Epidemiology Research Group. FISABIO-Public Health |
| EPI_ISL_436222 | Servicio de MicrobiologÃ-a. Consorcio Hospital General Universitario de Valencia | Sequencing and Bioinformatics Service and Molecular Epidemiology Research Group. FISABIO-Public Health |
| EPI_ISL_436223 | Servicio de MicrobiologÃ-a. Consorcio Hospital General Universitario de Valencia | Sequencing and Bioinformatics Service and Molecular Epidemiology Research Group. FISABIO-Public Health |
| EPI_ISL_436224 | Servicio de MicrobiologÃ-a. Consorcio Hospital General Universitario de Valencia | Sequencing and Bioinformatics Service and Molecular Epidemiology Research Group. FISABIO-Public Health |
| EPI_ISL_436225 | Servicio de MicrobiologÃ-a. Consorcio Hospital General Universitario de Valencia | Sequencing and Bioinformatics Service and Molecular Epidemiology Research Group. FISABIO-Public Health |
| EPI_ISL_436226 | Servicio de MicrobiologÃ-a. Consorcio Hospital General Universitario de Valencia | Sequencing and Bioinformatics Service and Molecular Epidemiology Research Group. FISABIO-Public Health |
| EPI_ISL_436227 | Servicio de MicrobiologÃ-a. Consorcio Hospital General Universitario de Valencia | Sequencing and Bioinformatics Service and Molecular Epidemiology Research Group. FISABIO-Public Health |
| EPI_ISL_436228 | Servicio de MicrobiologÃ-a. Consorcio Hospital General Universitario de Valencia | Sequencing and Bioinformatics Service and Molecular Epidemiology Research Group. FISABIO-Public Health |
| EPI_ISL_436229 | Servicio de MicrobiologÃ-a. Consorcio Hospital General Universitario de Valencia | Sequencing and Bioinformatics Service and Molecular Epidemiology Research Group. FISABIO-Public Health |
| EPI_ISL_436230 | Servicio de MicrobiologÃ-a. Consorcio Hospital General Universitario de Valencia | Sequencing and Bioinformatics Service and Molecular Epidemiology Research Group. FISABIO-Public Health |
| EPI_ISL_436231 | Servicio de MicrobiologÃ-a. Consorcio Hospital General Universitario de Valencia | Sequencing and Bioinformatics Service and Molecular Epidemiology Research Group. FISABIO-Public Health |
| EPI_ISL_436232 | Servicio de MicrobiologÃ-a. Consorcio Hospital General Universitario de Valencia | Sequencing and Bioinformatics Service and Molecular Epidemiology Research Group. FISABIO-Public Health |
| EPI_ISL_436233 | Servicio de MicrobiologÃ-a. Consorcio Hospital General Universitario de Valencia | Sequencing and Bioinformatics Service and Molecular Epidemiology Research Group. FISABIO-Public Health |
| EPI_ISL_436234 | Servicio de MicrobiologÃ-a. Consorcio Hospital General Universitario de Valencia | Sequencing and Bioinformatics Service and Molecular Epidemiology Research Group. FISABIO-Public Health |

| EPI_Accession_ID | Originating Laboratory | Submitting Laboratory |
| --- | --- | --- |
| EPI_ISL_436235 | Servicio de Microbiologia. Hospital Universitario Doctor Peset | Sequencing and Bioinformatics Service and Molecular Epidemiology Research Group. FISABIO-Public Health |
| EPI_ISL_436236 | Servicio de Microbiologia. Hospital Universitario Doctor Peset | Sequencing and Bioinformatics Service and Molecular Epidemiology Research Group. FISABIO-Public Health |
| EPI_ISL_436237 | Servicio de Microbiologia. Hospital Universitario Doctor Peset | Sequencing and Bioinformatics Service and Molecular Epidemiology Research Group. FISABIO-Public Health |
| EPI_ISL_436238 | Servicio de Microbiologia. Hospital Universitario Doctor Peset | Sequencing and Bioinformatics Service and Molecular Epidemiology Research Group. FISABIO-Public Health |
| EPI_ISL_436239 | Servicio de Microbiologia. Hospital Universitario Doctor Peset | Sequencing and Bioinformatics Service and Molecular Epidemiology Research Group. FISABIO-Public Health |
| EPI_ISL_436240 | Servicio de Microbiologia. Hospital Universitario Doctor Peset | Sequencing and Bioinformatics Service and Molecular Epidemiology Research Group. FISABIO-Public Health |
| EPI_ISL_436241 | Servicio de Microbiologia. Hospital Universitario Doctor Peset | Sequencing and Bioinformatics Service and Molecular Epidemiology Research Group. FISABIO-Public Health |
| EPI_ISL_436242 | Servicio de Microbiologia. Hospital Universitario Doctor Peset | Sequencing and Bioinformatics Service and Molecular Epidemiology Research Group. FISABIO-Public Health |
| EPI_ISL_436243 | Servicio de Microbiologia. Hospital Universitario Doctor Peset | Sequencing and Bioinformatics Service and Molecular Epidemiology Research Group. FISABIO-Public Health |
| EPI_ISL_436244 | Servicio de Microbiologia. Hospital Universitario Doctor Peset | Sequencing and Bioinformatics Service and Molecular Epidemiology Research Group. FISABIO-Public Health |
| EPI_ISL_436245 | Servicio de Microbiologia. Hospital Universitario Doctor Peset | Sequencing and Bioinformatics Service and Molecular Epidemiology Research Group. FISABIO-Public Health |
| EPI_ISL_436246 | Servicio de Microbiologia. Hospital Universitario Doctor Peset | Sequencing and Bioinformatics Service and Molecular Epidemiology Research Group. FISABIO-Public Health |
| EPI_ISL_436247 | Servicio de Microbiologia. Hospital Universitario Doctor Peset | Sequencing and Bioinformatics Service and Molecular Epidemiology Research Group. FISABIO-Public Health |
| EPI_ISL_436248 | Servicio de Microbiologia. Hospital Universitario Doctor Peset | Sequencing and Bioinformatics Service and Molecular Epidemiology Research Group. FISABIO-Public Health |
| EPI_ISL_436249 | Servicio de Microbiologia. Hospital Universitario Doctor Peset | Sequencing and Bioinformatics Service and Molecular Epidemiology Research Group. FISABIO-Public Health |
| EPI_ISL_436250 | Servicio de Microbiologia. Hospital Universitario Doctor Peset | Sequencing and Bioinformatics Service and Molecular Epidemiology Research Group. FISABIO-Public Health |

| EPI_Accession_ID | Originating Laboratory | Submitting Laboratory |
|---|---|---|
| EPI_ISL_436251 | Servicio de Microbiologia. Hospital Universitario Doctor Peset | Sequencing and Bioinformatics Service and Molecular Epidemiology Research Group. FISABIO-Public Health |
| EPI_ISL_436252 | Servicio de Microbiologia. Hospital Universitario Doctor Peset | Sequencing and Bioinformatics Service and Molecular Epidemiology Research Group. FISABIO-Public Health |
| EPI_ISL_436253 | Servicio de Microbiologia. Hospital Universitario Doctor Peset | Sequencing and Bioinformatics Service and Molecular Epidemiology Research Group. FISABIO-Public Health |
| EPI_ISL_436254 | Servicio de Microbiologia. Hospital Universitario Doctor Peset | Sequencing and Bioinformatics Service and Molecular Epidemiology Research Group. FISABIO-Public Health |
| EPI_ISL_436255 | Servicio de Microbiologia. Hospital Universitario Doctor Peset | Sequencing and Bioinformatics Service and Molecular Epidemiology Research Group. FISABIO-Public Health |
| EPI_ISL_436256 | Servicio de Microbiologia. Hospital Universitario Doctor Peset | Sequencing and Bioinformatics Service and Molecular Epidemiology Research Group. FISABIO-Public Health |
| EPI_ISL_436257 | Servicio de Microbiologia. Hospital Universitario Doctor Peset | Sequencing and Bioinformatics Service and Molecular Epidemiology Research Group. FISABIO-Public Health |
| EPI_ISL_436258 | Servicio de Microbiologia. Hospital Universitario Doctor Peset | Sequencing and Bioinformatics Service and Molecular Epidemiology Research Group. FISABIO-Public Health |
| EPI_ISL_436259 | Servicio de Microbiologia. Hospital Universitario Doctor Peset | Sequencing and Bioinformatics Service and Molecular Epidemiology Research Group. FISABIO-Public Health |
| EPI_ISL_436260 | Servicio de Microbiologia. Hospital Universitario Doctor Peset | Sequencing and Bioinformatics Service and Molecular Epidemiology Research Group. FISABIO-Public Health |
| EPI_ISL_436261 | Servicio de Microbiologia. Hospital Universitario Doctor Peset | Sequencing and Bioinformatics Service and Molecular Epidemiology Research Group. FISABIO-Public Health |
| EPI_ISL_436262 | Servicio de Microbiologia. Hospital Universitario Doctor Peset | Sequencing and Bioinformatics Service and Molecular Epidemiology Research Group. FISABIO-Public Health |
| EPI_ISL_436263 | Servicio de Microbiologia. Hospital Universitario Doctor Peset | Sequencing and Bioinformatics Service and Molecular Epidemiology Research Group. FISABIO-Public Health |
| EPI_ISL_436264 | Servicio de Microbiologia. Hospital Universitario Doctor Peset | Sequencing and Bioinformatics Service and Molecular Epidemiology Research Group. FISABIO-Public Health |
| EPI_ISL_436265 | Servicio de Microbiologia. Hospital Universitario Doctor Peset | Sequencing and Bioinformatics Service and Molecular Epidemiology Research Group. FISABIO-Public Health |
| EPI_ISL_436266 | Servicio de Microbiologia. Hospital Universitario Doctor Peset | Sequencing and Bioinformatics Service and Molecular Epidemiology Research Group. FISABIO-Public Health |

| EPI_Accession_ID | Originating Laboratory | Submitting Laboratory |
|---|---|---|
| EPI_ISL_436267 | Servicio de Microbiologia. Hospital Universitario Doctor Peset | Sequencing and Bioinformatics Service and Molecular Epidemiology Research Group. FISABIO-Public Health |
| EPI_ISL_436268 | Servicio de Microbiologia. Hospital Universitario Doctor Peset | Sequencing and Bioinformatics Service and Molecular Epidemiology Research Group. FISABIO-Public Health |
| EPI_ISL_436269 | Servicio de Microbiologia. Hospital Universitario Doctor Peset | Sequencing and Bioinformatics Service and Molecular Epidemiology Research Group. FISABIO-Public Health |
| EPI_ISL_436270 | Servicio de Microbiologia. Hospital Universitario Doctor Peset | Sequencing and Bioinformatics Service and Molecular Epidemiology Research Group. FISABIO-Public Health |
| EPI_ISL_436271 | Servicio de MicrobiologÃ-a. Consorcio Hospital General Universitario de Valencia | Sequencing and Bioinformatics Service and Molecular Epidemiology Research Group. FISABIO-Public Health |
| EPI_ISL_436272 | Servicio de MicrobiologÃ-a. Consorcio Hospital General Universitario de Valencia | Sequencing and Bioinformatics Service and Molecular Epidemiology Research Group. FISABIO-Public Health |
| EPI_ISL_436273 | Servicio de MicrobiologÃ-a. Consorcio Hospital General Universitario de Valencia | Sequencing and Bioinformatics Service and Molecular Epidemiology Research Group. FISABIO-Public Health |
| EPI_ISL_436274 | Servicio de MicrobiologÃ-a. Consorcio Hospital General Universitario de Valencia | Sequencing and Bioinformatics Service and Molecular Epidemiology Research Group. FISABIO-Public Health |
| EPI_ISL_436275 | Servicio de MicrobiologÃ-a. Consorcio Hospital General Universitario de Valencia | Sequencing and Bioinformatics Service and Molecular Epidemiology Research Group. FISABIO-Public Health |
| EPI_ISL_436276 | Servicio de MicrobiologÃ-a. Consorcio Hospital General Universitario de Valencia | Sequencing and Bioinformatics Service and Molecular Epidemiology Research Group. FISABIO-Public Health |
| EPI_ISL_436277 | Servicio de MicrobiologÃ-a. Consorcio Hospital General Universitario de Valencia | Sequencing and Bioinformatics Service and Molecular Epidemiology Research Group. FISABIO-Public Health |
| EPI_ISL_436278 | Servicio de Microbiologia. Hospital Clinico Universitario de Valencia | Sequencing and Bioinformatics Service and Molecular Epidemiology Research Group. FISABIO-Public Health |
| EPI_ISL_436279 | Servicio de Microbiologia. Hospital Clinico Universitario de Valencia | Sequencing and Bioinformatics Service and Molecular Epidemiology Research Group. FISABIO-Public Health |
| EPI_ISL_436280 | Servicio de Microbiologia. Hospital Clinico Universitario de Valencia | Sequencing and Bioinformatics Service and Molecular Epidemiology Research Group. FISABIO-Public Health |
| EPI_ISL_436281 | Servicio de Microbiologia. Hospital Clinico Universitario de Valencia | Sequencing and Bioinformatics Service and Molecular Epidemiology Research Group. FISABIO-Public Health |
| EPI_ISL_436282 | Servicio de Microbiologia. Hospital Clinico Universitario de Valencia | Sequencing and Bioinformatics Service and Molecular Epidemiology Research Group. FISABIO-Public Health |

| EPI_Accession_ID | Originating Laboratory | Submitting Laboratory |
|---|---|---|
| EPI_ISL_436283 | Servicio de Microbiologia. Hospital Clinico Universitario de Valencia | Sequencing and Bioinformatics Service and Molecular Epidemiology Research Group. FISABIO-Public Health |
| EPI_ISL_436284 | Servicio de Microbiologia. Hospital Clinico Universitario de Valencia | Sequencing and Bioinformatics Service and Molecular Epidemiology Research Group. FISABIO-Public Health |
| EPI_ISL_436285 | Servicio de Microbiologia. Hospital Clinico Universitario de Valencia | Sequencing and Bioinformatics Service and Molecular Epidemiology Research Group. FISABIO-Public Health |
| EPI_ISL_436286 | Servicio de Microbiologia. Hospital Clinico Universitario de Valencia | Sequencing and Bioinformatics Service and Molecular Epidemiology Research Group. FISABIO-Public Health |
| EPI_ISL_436287 | Servicio de Microbiologia. Hospital Clinico Universitario de Valencia | Sequencing and Bioinformatics Service and Molecular Epidemiology Research Group. FISABIO-Public Health |
| EPI_ISL_436288 | Servicio de Microbiologia. Hospital Clinico Universitario de Valencia | Sequencing and Bioinformatics Service and Molecular Epidemiology Research Group. FISABIO-Public Health |
| EPI_ISL_436289 | Servicio de Microbiologia. Hospital Clinico Universitario de Valencia | Sequencing and Bioinformatics Service and Molecular Epidemiology Research Group. FISABIO-Public Health |
| EPI_ISL_436290 | Servicio de Microbiologia. Hospital Clinico Universitario de Valencia | Sequencing and Bioinformatics Service and Molecular Epidemiology Research Group. FISABIO-Public Health |
| EPI_ISL_436291 | Servicio de Microbiologia. Hospital Clinico Universitario de Valencia | Sequencing and Bioinformatics Service and Molecular Epidemiology Research Group. FISABIO-Public Health |
| EPI_ISL_436292 | Servicio de Microbiologia. Hospital Clinico Universitario de Valencia | Sequencing and Bioinformatics Service and Molecular Epidemiology Research Group. FISABIO-Public Health |
| EPI_ISL_436293 | Servicio de Microbiologia. Hospital Clinico Universitario de Valencia | Sequencing and Bioinformatics Service and Molecular Epidemiology Research Group. FISABIO-Public Health |
| EPI_ISL_436294 | Servicio de Microbiologia. Hospital Clinico Universitario de Valencia | Sequencing and Bioinformatics Service and Molecular Epidemiology Research Group. FISABIO-Public Health |
| EPI_ISL_436295 | Servicio de Microbiologia. Hospital Clinico Universitario de Valencia | Sequencing and Bioinformatics Service and Molecular Epidemiology Research Group. FISABIO-Public Health |
| EPI_ISL_436296 | Servicio de Microbiologia. Hospital Clinico Universitario de Valencia | Sequencing and Bioinformatics Service and Molecular Epidemiology Research Group. FISABIO-Public Health |
| EPI_ISL_436297 | Servicio de Microbiologia. Hospital Clinico Universitario de Valencia | Sequencing and Bioinformatics Service and Molecular Epidemiology Research Group. FISABIO-Public Health |
| EPI_ISL_436298 | Servicio de Microbiologia. Hospital Clinico Universitario de Valencia | Sequencing and Bioinformatics Service and Molecular Epidemiology Research Group. FISABIO-Public Health |

| EPI_Accession_ID | Originating Laboratory | Submitting Laboratory |
|---|---|---|
| EPI_ISL_436299 | Servicio de Microbiologia. Hospital Clinico Universitario de Valencia | Sequencing and Bioinformatics Service and Molecular Epidemiology Research Group. FISABIO-Public Health |
| EPI_ISL_436300 | Servicio de Microbiologia. Hospital Clinico Universitario de Valencia | Sequencing and Bioinformatics Service and Molecular Epidemiology Research Group. FISABIO-Public Health |
| EPI_ISL_436301 | Servicio de Microbiologia. Hospital Clinico Universitario de Valencia | Sequencing and Bioinformatics Service and Molecular Epidemiology Research Group. FISABIO-Public Health |
| EPI_ISL_436302 | Servicio de Microbiologia. Hospital Clinico Universitario de Valencia | Sequencing and Bioinformatics Service and Molecular Epidemiology Research Group. FISABIO-Public Health |
| EPI_ISL_436303 | Servicio de Microbiologia. Hospital Clinico Universitario de Valencia | Sequencing and Bioinformatics Service and Molecular Epidemiology Research Group. FISABIO-Public Health |
| EPI_ISL_436304 | Servicio de Microbiologia. Hospital Clinico Universitario de Valencia | Sequencing and Bioinformatics Service and Molecular Epidemiology Research Group. FISABIO-Public Health |
| EPI_ISL_436305 | Servicio de Microbiologia. Hospital Clinico Universitario de Valencia | Sequencing and Bioinformatics Service and Molecular Epidemiology Research Group. FISABIO-Public Health |
| EPI_ISL_436306 | Servicio de Microbiologia. Hospital Clinico Universitario de Valencia | Sequencing and Bioinformatics Service and Molecular Epidemiology Research Group. FISABIO-Public Health |
| EPI_ISL_436307 | Servicio de Microbiologia. Hospital Clinico Universitario de Valencia | Sequencing and Bioinformatics Service and Molecular Epidemiology Research Group. FISABIO-Public Health |
| EPI_ISL_436308 | Servicio de Microbiologia. Hospital Clinico Universitario de Valencia | Sequencing and Bioinformatics Service and Molecular Epidemiology Research Group. FISABIO-Public Health |
| EPI_ISL_436309 | Servicio de Microbiologia. Hospital Clinico Universitario de Valencia | Sequencing and Bioinformatics Service and Molecular Epidemiology Research Group. FISABIO-Public Health |
| EPI_ISL_436310 | Servicio de Microbiologia. Hospital Clinico Universitario de Valencia | Sequencing and Bioinformatics Service and Molecular Epidemiology Research Group. FISABIO-Public Health |
| EPI_ISL_436311 | Servicio de Microbiologia. Hospital Clinico Universitario de Valencia | Sequencing and Bioinformatics Service and Molecular Epidemiology Research Group. FISABIO-Public Health |
| EPI_ISL_436312 | Servicio de Microbiologia. Hospital Clinico Universitario de Valencia | Sequencing and Bioinformatics Service and Molecular Epidemiology Research Group. FISABIO-Public Health |
| EPI_ISL_436313 | Servicio de Microbiologia. Hospital Clinico Universitario de Valencia | Sequencing and Bioinformatics Service and Molecular Epidemiology Research Group. FISABIO-Public Health |
| EPI_ISL_436314 | Servicio de Microbiologia. Hospital Clinico Universitario de Valencia | Sequencing and Bioinformatics Service and Molecular Epidemiology Research Group. FISABIO-Public Health |

| EPI_Accession_ID | Originating Laboratory | Submitting Laboratory |
| --- | --- | --- |
| EPI_ISL_436315 | Servicio de Microbiologia. Hospital Clinico Universitario de Valencia | Sequencing and Bioinformatics Service and Molecular Epidemiology Research Group. FISABIO-Public Health |
| EPI_ISL_436316 | Servicio de Microbiologia. Hospital Clinico Universitario de Valencia | Sequencing and Bioinformatics Service and Molecular Epidemiology Research Group. FISABIO-Public Health |
| EPI_ISL_436317 | Servicio de Microbiologia. Hospital Clinico Universitario de Valencia | Sequencing and Bioinformatics Service and Molecular Epidemiology Research Group. FISABIO-Public Health |
| EPI_ISL_436318 | Servicio de Microbiologia. Hospital Clinico Universitario de Valencia | Sequencing and Bioinformatics Service and Molecular Epidemiology Research Group. FISABIO-Public Health |
| EPI_ISL_436319 | Servicio de Microbiologia. Hospital Clinico Universitario de Valencia | Sequencing and Bioinformatics Service and Molecular Epidemiology Research Group. FISABIO-Public Health |
| EPI_ISL_436320 | Servicio de Microbiologia. Hospital Clinico Universitario de Valencia | Sequencing and Bioinformatics Service and Molecular Epidemiology Research Group. FISABIO-Public Health |
| EPI_ISL_436321 | Servicio de Microbiologia. Hospital Clinico Universitario de Valencia | Sequencing and Bioinformatics Service and Molecular Epidemiology Research Group. FISABIO-Public Health |
| EPI_ISL_436322 | Servicio de Microbiologia. Hospital Clinico Universitario de Valencia | Sequencing and Bioinformatics Service and Molecular Epidemiology Research Group. FISABIO-Public Health |
| EPI_ISL_436323 | Servicio de Microbiologia. Hospital Clinico Universitario de Valencia | Sequencing and Bioinformatics Service and Molecular Epidemiology Research Group. FISABIO-Public Health |
| EPI_ISL_436324 | Servicio de Microbiologia. Hospital Clinico Universitario de Valencia | Sequencing and Bioinformatics Service and Molecular Epidemiology Research Group. FISABIO-Public Health |
| EPI_ISL_436325 | Servicio de Microbiologia. Hospital Clinico Universitario de Valencia | Sequencing and Bioinformatics Service and Molecular Epidemiology Research Group. FISABIO-Public Health |
| EPI_ISL_436326 | Servicio de Microbiologia. Hospital Clinico Universitario de Valencia | Sequencing and Bioinformatics Service and Molecular Epidemiology Research Group. FISABIO-Public Health |
| EPI_ISL_436327 | Servicio de Microbiologia. Hospital Clinico Universitario de Valencia | Sequencing and Bioinformatics Service and Molecular Epidemiology Research Group. FISABIO-Public Health |
| EPI_ISL_436328 | Servicio de Microbiologia. Hospital Clinico Universitario de Valencia | Sequencing and Bioinformatics Service and Molecular Epidemiology Research Group. FISABIO-Public Health |
| EPI_ISL_436329 | Servicio de Microbiologia. Hospital Clinico Universitario de Valencia | Sequencing and Bioinformatics Service and Molecular Epidemiology Research Group. FISABIO-Public Health |
| EPI_ISL_436330 | Servicio de Microbiologia. Hospital Clinico Universitario de Valencia | Sequencing and Bioinformatics Service and Molecular Epidemiology Research Group. FISABIO-Public Health |

| EPI_Accession_ID | Originating Laboratory | Submitting Laboratory |
|---|---|---|
| EPI_ISL_436331 | Servicio de Microbiologia. Hospital Clinico Universitario de Valencia | Sequencing and Bioinformatics Service and Molecular Epidemiology Research Group. FISABIO-Public Health |
| EPI_ISL_436332 | Servicio de Microbiologia. Hospital Clinico Universitario de Valencia | Sequencing and Bioinformatics Service and Molecular Epidemiology Research Group. FISABIO-Public Health |
| EPI_ISL_436333 | Servicio de Microbiologia. Hospital Clinico Universitario de Valencia | Sequencing and Bioinformatics Service and Molecular Epidemiology Research Group. FISABIO-Public Health |
| EPI_ISL_436334 | Servicio de Microbiologia. Hospital Clinico Universitario de Valencia | Sequencing and Bioinformatics Service and Molecular Epidemiology Research Group. FISABIO-Public Health |
| EPI_ISL_436335 | Servicio de Microbiologia. Hospital Clinico Universitario de Valencia | Sequencing and Bioinformatics Service and Molecular Epidemiology Research Group. FISABIO-Public Health |
| EPI_ISL_436336 | Servicio de Microbiologia. Hospital Clinico Universitario de Valencia | Sequencing and Bioinformatics Service and Molecular Epidemiology Research Group. FISABIO-Public Health |
| EPI_ISL_436337 | Servicio de Microbiologia. Hospital Clinico Universitario de Valencia | Sequencing and Bioinformatics Service and Molecular Epidemiology Research Group. FISABIO-Public Health |
| EPI_ISL_436338 | Servicio de Microbiologia. Hospital Clinico Universitario de Valencia | Sequencing and Bioinformatics Service and Molecular Epidemiology Research Group. FISABIO-Public Health |
| EPI_ISL_436339 | Servicio de Microbiologia. Hospital Clinico Universitario de Valencia | Sequencing and Bioinformatics Service and Molecular Epidemiology Research Group. FISABIO-Public Health |
| EPI_ISL_436340 | Servicio de Microbiologia. Hospital Clinico Universitario de Valencia | Sequencing and Bioinformatics Service and Molecular Epidemiology Research Group. FISABIO-Public Health |
| EPI_ISL_436341 | Servicio de Microbiologia. Hospital Clinico Universitario de Valencia | Sequencing and Bioinformatics Service and Molecular Epidemiology Research Group. FISABIO-Public Health |
| EPI_ISL_436342 | Servicio de Microbiologia. Hospital Clinico Universitario de Valencia | Sequencing and Bioinformatics Service and Molecular Epidemiology Research Group. FISABIO-Public Health |
| EPI_ISL_436343 | Servicio de Microbiologia. Hospital Clinico Universitario de Valencia | Sequencing and Bioinformatics Service and Molecular Epidemiology Research Group. FISABIO-Public Health |
| EPI_ISL_436344 | Servicio de Microbiologia. Hospital Clinico Universitario de Valencia | Sequencing and Bioinformatics Service and Molecular Epidemiology Research Group. FISABIO-Public Health |
| EPI_ISL_436345 | Servicio de Microbiologia. Hospital Clinico Universitario de Valencia | Sequencing and Bioinformatics Service and Molecular Epidemiology Research Group. FISABIO-Public Health |
| EPI_ISL_436346 | Servicio de Microbiologia. Hospital Clinico Universitario de Valencia | Sequencing and Bioinformatics Service and Molecular Epidemiology Research Group. FISABIO-Public Health |

| EPI_Accession_ID | Originating Laboratory | Submitting Laboratory |
|---|---|---|
| EPI_ISL_436347 | Servicio de Microbiologia. Hospital Clinico Universitario de Valencia | Sequencing and Bioinformatics Service and Molecular Epidemiology Research Group. FISABIO-Public Health |
| EPI_ISL_436348 | Servicio de Microbiologia. Hospital Clinico Universitario de Valencia | Sequencing and Bioinformatics Service and Molecular Epidemiology Research Group. FISABIO-Public Health |
| EPI_ISL_436349 | Servicio de Microbiologia. Hospital Clinico Universitario de Valencia | Sequencing and Bioinformatics Service and Molecular Epidemiology Research Group. FISABIO-Public Health |
| EPI_ISL_436350 | Servicio de Microbiologia. Hospital Clinico Universitario de Valencia | Sequencing and Bioinformatics Service and Molecular Epidemiology Research Group. FISABIO-Public Health |
| EPI_ISL_436351 | Servicio de Microbiologia. Hospital Clinico Universitario de Valencia | Sequencing and Bioinformatics Service and Molecular Epidemiology Research Group. FISABIO-Public Health |
| EPI_ISL_436352 | Servicio de Microbiologia. Hospital Clinico Universitario de Valencia | Sequencing and Bioinformatics Service and Molecular Epidemiology Research Group. FISABIO-Public Health |
| EPI_ISL_436353 | Servicio de Microbiologia. Hospital Clinico Universitario de Valencia | Sequencing and Bioinformatics Service and Molecular Epidemiology Research Group. FISABIO-Public Health |
| EPI_ISL_436354 | Servicio de Microbiologia. Hospital Clinico Universitario de Valencia | Sequencing and Bioinformatics Service and Molecular Epidemiology Research Group. FISABIO-Public Health |
| EPI_ISL_436355 | Servicio de Microbiologia. Hospital Clinico Universitario de Valencia | Sequencing and Bioinformatics Service and Molecular Epidemiology Research Group. FISABIO-Public Health |
| EPI_ISL_436356 | Servicio de Microbiologia. Hospital Clinico Universitario de Valencia | Sequencing and Bioinformatics Service and Molecular Epidemiology Research Group. FISABIO-Public Health |
| EPI_ISL_436357 | Servicio de Microbiologia. Hospital Clinico Universitario de Valencia | Sequencing and Bioinformatics Service and Molecular Epidemiology Research Group. FISABIO-Public Health |
| EPI_ISL_436358 | Servicio de Microbiologia. Hospital Clinico Universitario de Valencia | Sequencing and Bioinformatics Service and Molecular Epidemiology Research Group. FISABIO-Public Health |
| EPI_ISL_436359 | Servicio de Microbiologia. Hospital Clinico Universitario de Valencia | Sequencing and Bioinformatics Service and Molecular Epidemiology Research Group. FISABIO-Public Health |
| EPI_ISL_436360 | Servicio de Microbiologia. Hospital Clinico Universitario de Valencia | Sequencing and Bioinformatics Service and Molecular Epidemiology Research Group. FISABIO-Public Health |
| EPI_ISL_436361 | Servicio de MicrobiologÃ-a. Consorcio Hospital General Universitario de Valencia | Sequencing and Bioinformatics Service and Molecular Epidemiology Research Group. FISABIO-Public Health |
| EPI_ISL_436362 | Servicio de Microbiologia. Hospital Clinico Universitario de Valencia | Sequencing and Bioinformatics Service and Molecular Epidemiology Research Group. FISABIO-Public Health |

| EPI_Accession_ID | Originating Laboratory | Submitting Laboratory |
|---|---|---|
| EPI_ISL_436363 | Servicio de Microbiologia. Hospital Clinico Universitario de Valencia | Sequencing and Bioinformatics Service and Molecular Epidemiology Research Group. FISABIO-Public Health |
| EPI_ISL_436364 | Servicio de Microbiologia. Hospital Clinico Universitario de Valencia | Sequencing and Bioinformatics Service and Molecular Epidemiology Research Group. FISABIO-Public Health |
| EPI_ISL_436365 | Servicio de Microbiologia. Hospital Clinico Universitario de Valencia | Sequencing and Bioinformatics Service and Molecular Epidemiology Research Group. FISABIO-Public Health |
| EPI_ISL_436366 | Servicio de Microbiologia. Hospital Clinico Universitario de Valencia | Sequencing and Bioinformatics Service and Molecular Epidemiology Research Group. FISABIO-Public Health |
| EPI_ISL_436367 | Servicio de Microbiologia. Hospital Clinico Universitario de Valencia | Sequencing and Bioinformatics Service and Molecular Epidemiology Research Group. FISABIO-Public Health |
| EPI_ISL_436368 | Servicio de Microbiologia. Hospital Clinico Universitario de Valencia | Sequencing and Bioinformatics Service and Molecular Epidemiology Research Group. FISABIO-Public Health |
| EPI_ISL_436369 | Servicio de Microbiologia. Hospital Clinico Universitario de Valencia | Sequencing and Bioinformatics Service and Molecular Epidemiology Research Group. FISABIO-Public Health |
| EPI_ISL_436370 | Servicio de Microbiologia. Hospital Clinico Universitario de Valencia | Sequencing and Bioinformatics Service and Molecular Epidemiology Research Group. FISABIO-Public Health |
| EPI_ISL_436371 | Servicio de Microbiologia. Hospital Clinico Universitario de Valencia | Sequencing and Bioinformatics Service and Molecular Epidemiology Research Group. FISABIO-Public Health |
| EPI_ISL_436372 | Servicio de Microbiologia. Hospital Universitario Doctor Peset | Sequencing and Bioinformatics Service and Molecular Epidemiology Research Group. FISABIO-Public Health |
| EPI_ISL_436373 | Servicio de Microbiologia. Hospital Universitario Doctor Peset | Sequencing and Bioinformatics Service and Molecular Epidemiology Research Group. FISABIO-Public Health |
| EPI_ISL_436374 | Servicio de Microbiologia. Hospital Universitario Doctor Peset | Sequencing and Bioinformatics Service and Molecular Epidemiology Research Group. FISABIO-Public Health |
| EPI_ISL_436375 | Servicio de Microbiologia. Hospital Universitario Doctor Peset | Sequencing and Bioinformatics Service and Molecular Epidemiology Research Group. FISABIO-Public Health |
| EPI_ISL_436376 | Servicio de Microbiologia. Hospital Universitario Doctor Peset | Sequencing and Bioinformatics Service and Molecular Epidemiology Research Group. FISABIO-Public Health |
| EPI_ISL_436377 | Servicio de Microbiologia. Hospital Universitario Doctor Peset | Sequencing and Bioinformatics Service and Molecular Epidemiology Research Group. FISABIO-Public Health |
| EPI_ISL_436378 | Servicio de Microbiologia. Hospital Universitario Doctor Peset | Sequencing and Bioinformatics Service and Molecular Epidemiology Research Group. FISABIO-Public Health |

| EPI_Accession_ID | Originating Laboratory | Submitting Laboratory |
|---|---|---|
| EPI_ISL_436379 | Servicio de Microbiologia. Hospital Universitario Doctor Peset | Sequencing and Bioinformatics Service and Molecular Epidemiology Research Group. FISABIO-Public Health |
| EPI_ISL_436380 | Servicio de Microbiologia. Hospital Universitario Doctor Peset | Sequencing and Bioinformatics Service and Molecular Epidemiology Research Group. FISABIO-Public Health |
| EPI_ISL_436381 | Servicio de Microbiologia. Hospital Universitario Doctor Peset | Sequencing and Bioinformatics Service and Molecular Epidemiology Research Group. FISABIO-Public Health |
| EPI_ISL_436382 | Servicio de MicrobiologÃ-a. Consorcio Hospital General Universitario de Valencia | Sequencing and Bioinformatics Service and Molecular Epidemiology Research Group. FISABIO-Public Health |
| EPI_ISL_436383 | Servicio de MicrobiologÃ-a. Consorcio Hospital General Universitario de Valencia | Sequencing and Bioinformatics Service and Molecular Epidemiology Research Group. FISABIO-Public Health |
| EPI_ISL_436384 | Servicio de MicrobiologÃ-a. Consorcio Hospital General Universitario de Valencia | Sequencing and Bioinformatics Service and Molecular Epidemiology Research Group. FISABIO-Public Health |
| EPI_ISL_436385 | Servicio de MicrobiologÃ-a. Consorcio Hospital General Universitario de Valencia | Sequencing and Bioinformatics Service and Molecular Epidemiology Research Group. FISABIO-Public Health |
| EPI_ISL_436386 | Servicio de MicrobiologÃ-a. Consorcio Hospital General Universitario de Valencia | Sequencing and Bioinformatics Service and Molecular Epidemiology Research Group. FISABIO-Public Health |
| EPI_ISL_436387 | Servicio de MicrobiologÃ-a. Consorcio Hospital General Universitario de Valencia | Sequencing and Bioinformatics Service and Molecular Epidemiology Research Group. FISABIO-Public Health |
| EPI_ISL_436388 | Servicio de MicrobiologÃ-a. Consorcio Hospital General Universitario de Valencia | Sequencing and Bioinformatics Service and Molecular Epidemiology Research Group. FISABIO-Public Health |
| EPI_ISL_436389 | Servicio de MicrobiologÃ-a. Consorcio Hospital General Universitario de Valencia | Sequencing and Bioinformatics Service and Molecular Epidemiology Research Group. FISABIO-Public Health |
| EPI_ISL_436390 | Servicio de MicrobiologÃ-a. Consorcio Hospital General Universitario de Valencia | Sequencing and Bioinformatics Service and Molecular Epidemiology Research Group. FISABIO-Public Health |
| EPI_ISL_436391 | Servicio de MicrobiologÃ-a. Consorcio Hospital General Universitario de Valencia | Sequencing and Bioinformatics Service and Molecular Epidemiology Research Group. FISABIO-Public Health |
| EPI_ISL_436392 | Servicio de MicrobiologÃ-a. Consorcio Hospital General Universitario de Valencia | Sequencing and Bioinformatics Service and Molecular Epidemiology Research Group. FISABIO-Public Health |
| EPI_ISL_436393 | Servicio de MicrobiologÃ-a. Consorcio Hospital General Universitario de Valencia | Sequencing and Bioinformatics Service and Molecular Epidemiology Research Group. FISABIO-Public Health |
| EPI_ISL_436394 | Servicio de MicrobiologÃ-a. Consorcio Hospital General Universitario de Valencia | Sequencing and Bioinformatics Service and Molecular Epidemiology Research Group. FISABIO-Public Health |

| EPI_Accession_ID | Originating Laboratory | Submitting Laboratory |
|---|---|---|
| EPI_ISL_436395 | Servicio de MicrobiologÃ-a. Consorcio Hospital General Universitario de Valencia | Sequencing and Bioinformatics Service and Molecular Epidemiology Research Group. FISABIO-Public Health |
| EPI_ISL_436396 | Servicio de Microbiologia. Hospital Clinico Universitario de Valencia | Sequencing and Bioinformatics Service and Molecular Epidemiology Research Group. FISABIO-Public Health |
| EPI_ISL_436397 | Servicio de Microbiologia. Hospital Clinico Universitario de Valencia | Sequencing and Bioinformatics Service and Molecular Epidemiology Research Group. FISABIO-Public Health |
| EPI_ISL_436398 | Servicio de Microbiologia. Hospital Clinico Universitario de Valencia | Sequencing and Bioinformatics Service and Molecular Epidemiology Research Group. FISABIO-Public Health |
| EPI_ISL_436399 | Servicio de Microbiologia. Hospital Clinico Universitario de Valencia | Sequencing and Bioinformatics Service and Molecular Epidemiology Research Group. FISABIO-Public Health |
| EPI_ISL_436400 | Servicio de Microbiologia. Hospital Clinico Universitario de Valencia | Sequencing and Bioinformatics Service and Molecular Epidemiology Research Group. FISABIO-Public Health |
| EPI_ISL_436401 | Servicio de Microbiologia. Hospital Clinico Universitario de Valencia | Sequencing and Bioinformatics Service and Molecular Epidemiology Research Group. FISABIO-Public Health |
| EPI_ISL_436402 | Servicio de Microbiologia. Hospital Clinico Universitario de Valencia | Sequencing and Bioinformatics Service and Molecular Epidemiology Research Group. FISABIO-Public Health |
| EPI_ISL_436403 | Servicio de Microbiologia. Hospital Clinico Universitario de Valencia | Sequencing and Bioinformatics Service and Molecular Epidemiology Research Group. FISABIO-Public Health |
| EPI_ISL_436404 | Servicio de Microbiologia. Hospital Clinico Universitario de Valencia | Sequencing and Bioinformatics Service and Molecular Epidemiology Research Group. FISABIO-Public Health |
| EPI_ISL_436405 | Servicio de Microbiologia. Hospital Clinico Universitario de Valencia | Sequencing and Bioinformatics Service and Molecular Epidemiology Research Group. FISABIO-Public Health |
| EPI_ISL_436406 | Servicio de Microbiologia. Hospital Clinico Universitario de Valencia | Sequencing and Bioinformatics Service and Molecular Epidemiology Research Group. FISABIO-Public Health |
| EPI_ISL_436407 | Servicio de Microbiologia. Hospital Clinico Universitario de Valencia | Sequencing and Bioinformatics Service and Molecular Epidemiology Research Group. FISABIO-Public Health |
| EPI_ISL_436408 | Servicio de Microbiologia. Hospital Clinico Universitario de Valencia | Sequencing and Bioinformatics Service and Molecular Epidemiology Research Group. FISABIO-Public Health |
| EPI_ISL_436409 | Servicio de Microbiologia. Hospital Clinico Universitario de Valencia | Sequencing and Bioinformatics Service and Molecular Epidemiology Research Group. FISABIO-Public Health |
| EPI_ISL_436410 | Servicio de Microbiologia. Hospital Clinico Universitario de Valencia | Sequencing and Bioinformatics Service and Molecular Epidemiology Research Group. FISABIO-Public Health |

| EPI_Accession_ID | Originating Laboratory | Submitting Laboratory |
|---|---|---|
| EPI_ISL_436411 | Servicio de Microbiologia. Hospital Clinico Universitario de Valencia | Sequencing and Bioinformatics Service and Molecular Epidemiology Research Group. FISABIO-Public Health |
| EPI_ISL_436412 | Viral Respiratory Lab, National Institute for Biomedical Research (INRB) | Pathogen Sequencing Lab, National Institute for Biomedical Research (INRB) |
| EPI_ISL_436413 | National Centre for Disease control (NCDC) | NCDC/CSIR-IGIB |
| EPI_ISL_436414 | National Centre for Disease control (NCDC) | NCDC/CSIR-IGIB |
| EPI_ISL_436415 | National Centre for Disease control (NCDC) | NCDC/CSIR-IGIB |
| EPI_ISL_436416 | National Centre for Disease control (NCDC) | NCDC/CSIR-IGIB |
| EPI_ISL_436417 | National Centre for Disease control (NCDC) | NCDC/CSIR-IGIB |
| EPI_ISL_436418 | National Centre for Disease control (NCDC) | NCDC/CSIR-IGIB |
| EPI_ISL_436419 | National Centre for Disease control (NCDC) | NCDC/CSIR-IGIB |
| EPI_ISL_436420 | National Centre for Disease control (NCDC) | NCDC/CSIR-IGIB |
| EPI_ISL_436421 | National Centre for Disease control (NCDC) | NCDC/CSIR-IGIB |
| EPI_ISL_436422 | National Centre for Disease control (NCDC) | NCDC/CSIR-IGIB |
| EPI_ISL_436423 | National Centre for Disease control (NCDC) | NCDC/CSIR-IGIB |
| EPI_ISL_436424 | National Centre for Disease control (NCDC) | NCDC/CSIR-IGIB |
| EPI_ISL_436425 | National Centre for Disease control (NCDC) | NCDC/CSIR-IGIB |
| EPI_ISL_436426 | National Centre for Disease control (NCDC) | NCDC/CSIR-IGIB |
| EPI_ISL_436427 | National Centre for Disease control (NCDC) | NCDC/CSIR-IGIB |
| EPI_ISL_436428 | National Centre for Disease control (NCDC) | NCDC/CSIR-IGIB |
| EPI_ISL_436429 | National Centre for Disease control (NCDC) | NCDC/CSIR-IGIB |
| EPI_ISL_436430 | National Centre for Disease control (NCDC) | NCDC/CSIR-IGIB |
| EPI_ISL_436431 | National Centre for Disease control (NCDC) | NCDC/CSIR-IGIB |
| EPI_ISL_436432 | National Centre for Disease control (NCDC) | NCDC/CSIR-IGIB |
| EPI_ISL_436433 | National Centre for Disease control (NCDC) | NCDC/CSIR-IGIB |
| EPI_ISL_436434 | National Centre for Disease control (NCDC) | NCDC/CSIR-IGIB |
| EPI_ISL_436435 | National Centre for Disease control (NCDC) | NCDC/CSIR-IGIB |
| EPI_ISL_436436 | National Centre for Disease control (NCDC) | NCDC/CSIR-IGIB |
| EPI_ISL_436437 | National Centre for Disease control (NCDC) | NCDC/CSIR-IGIB |
| EPI_ISL_436438 | National Centre for Disease control (NCDC) | NCDC/CSIR-IGIB |
| EPI_ISL_436439 | National Centre for Disease control (NCDC) | NCDC/CSIR-IGIB |
| EPI_ISL_436440 | National Centre for Disease control (NCDC) | NCDC/CSIR-IGIB |
| EPI_ISL_436441 | National Centre for Disease control (NCDC) | NCDC/CSIR-IGIB |

| EPI_Accession_ID | Originating Laboratory | Submitting Laboratory |
|---|---|---|
| EPI_ISL_436442 | National Centre for Disease control (NCDC) | NCDC/CSIR-IGIB |
| EPI_ISL_436443 | National Centre for Disease control (NCDC) | NCDC/CSIR-IGIB |
| EPI_ISL_436444 | National Centre for Disease control (NCDC) | NCDC/CSIR-IGIB |
| EPI_ISL_436445 | National Centre for Disease control (NCDC) | NCDC/CSIR-IGIB |
| EPI_ISL_436446 | National Centre for Disease control (NCDC) | NCDC/CSIR-IGIB |
| EPI_ISL_436447 | National Centre for Disease control (NCDC) | NCDC/CSIR-IGIB |
| EPI_ISL_436448 | National Centre for Disease control (NCDC) | NCDC/CSIR-IGIB |
| EPI_ISL_436449 | National Centre for Disease control (NCDC) | NCDC/CSIR-IGIB |
| EPI_ISL_436450 | National Centre for Disease control (NCDC) | NCDC/CSIR-IGIB |
| EPI_ISL_436451 | National Centre for Disease control (NCDC) | NCDC/CSIR-IGIB |
| EPI_ISL_436452 | National Centre for Disease control (NCDC) | NCDC/CSIR-IGIB |
| EPI_ISL_436453 | National Centre for Disease control (NCDC) | NCDC/CSIR-IGIB |
| EPI_ISL_436454 | National Centre for Disease control (NCDC) | NCDC/CSIR-IGIB |
| EPI_ISL_436455 | National Centre for Disease control (NCDC) | NCDC/CSIR-IGIB |
| EPI_ISL_436456 | National Centre for Disease control (NCDC) | NCDC/CSIR-IGIB |
| EPI_ISL_436457 | National Centre for Disease control (NCDC) | NCDC/CSIR-IGIB |
| EPI_ISL_436458 | National Centre for Disease control (NCDC) | NCDC/CSIR-IGIB |
| EPI_ISL_436459 | National Centre for Disease control (NCDC) | NCDC/CSIR-IGIB |
| EPI_ISL_436460 | National Centre for Disease control (NCDC) | NCDC/CSIR-IGIB |
| EPI_ISL_436461 | National Centre for Disease control (NCDC) | NCDC/CSIR-IGIB |
| EPI_ISL_436462 | National Centre for Disease control (NCDC) | NCDC/CSIR-IGIB |
| EPI_ISL_436463 | National Centre for Disease control (NCDC) | NCDC/CSIR-IGIB |
| EPI_ISL_436464 | Alaska State Virology Laboratory | Alaska State Virology Laboratory |
| EPI_ISL_436466 | UPMC Clinical Laboratory | Microbial Genome Sequencing Center, Microbial Genomic Epidemiological Laboratory |
| EPI_ISL_436467 | UPMC Clinical Laboratory | Microbial Genome Sequencing Center, Microbial Genomic Epidemiological Laboratory |
| EPI_ISL_436468 | UPMC Clinical Laboratory | Microbial Genome Sequencing Center, Microbial Genomic Epidemiological Laboratory |
| EPI_ISL_436469 | UPMC Clinical Laboratory | Microbial Genome Sequencing Center, Microbial Genomic Epidemiological Laboratory |
| EPI_ISL_436470 | UPMC Clinical Laboratory | Microbial Genome Sequencing Center, Microbial Genomic Epidemiological Laboratory |

| EPI_Accession_ID | Originating Laboratory | Submitting Laboratory |
|---|---|---|
| EPI_ISL_436471 | UPMC Clinical Laboratory | Microbial Genome Sequencing Center, Microbial Genomic Epidemiological Laboratory |
| EPI_ISL_436472 | UPMC Clinical Laboratory | Microbial Genome Sequencing Center, Microbial Genomic Epidemiological Laboratory |
| EPI_ISL_436473 | UPMC Clinical Laboratory | Microbial Genome Sequencing Center, Microbial Genomic Epidemiological Laboratory |
| EPI_ISL_436474 | UPMC Clinical Laboratory | Microbial Genome Sequencing Center, Microbial Genomic Epidemiological Laboratory |
| EPI_ISL_436475 | UPMC Clinical Laboratory | Microbial Genome Sequencing Center, Microbial Genomic Epidemiological Laboratory |
| EPI_ISL_436476 | UPMC Clinical Laboratory | Microbial Genome Sequencing Center, Microbial Genomic Epidemiological Laboratory |
| EPI_ISL_436477 | UPMC Clinical Laboratory | Microbial Genome Sequencing Center, Microbial Genomic Epidemiological Laboratory |
| EPI_ISL_436478 | UPMC Clinical Laboratory | Microbial Genome Sequencing Center, Microbial Genomic Epidemiological Laboratory |
| EPI_ISL_436479 | UPMC Clinical Laboratory | Microbial Genome Sequencing Center, Microbial Genomic Epidemiological Laboratory |
| EPI_ISL_436480 | UPMC Clinical Laboratory | Microbial Genome Sequencing Center, Microbial Genomic Epidemiological Laboratory |
| EPI_ISL_436481 | UPMC Clinical Laboratory | Microbial Genome Sequencing Center, Microbial Genomic Epidemiological Laboratory |
| EPI_ISL_436482 | UPMC Clinical Laboratory | Microbial Genome Sequencing Center, Microbial Genomic Epidemiological Laboratory |
| EPI_ISL_436483 | UPMC Clinical Laboratory | Microbial Genome Sequencing Center, Microbial Genomic Epidemiological Laboratory |
| EPI_ISL_436484 | UPMC Clinical Laboratory | Microbial Genome Sequencing Center, Microbial Genomic Epidemiological Laboratory |
| EPI_ISL_436485 | UPMC Clinical Laboratory | Microbial Genome Sequencing Center, Microbial Genomic Epidemiological Laboratory |
| EPI_ISL_436486 | UPMC Clinical Laboratory | Microbial Genome Sequencing Center, Microbial Genomic Epidemiological Laboratory |

| EPI_Accession_ID | Originating Laboratory | Submitting Laboratory |
|---|---|---|
| EPI_ISL_436487 | UPMC Clinical Laboratory | Microbial Genome Sequencing Center, Microbial Genomic Epidemiological Laboratory |
| EPI_ISL_436488 | UPMC Clinical Laboratory | Microbial Genome Sequencing Center, Microbial Genomic Epidemiological Laboratory |
| EPI_ISL_436489 | UPMC Clinical Laboratory | Microbial Genome Sequencing Center, Microbial Genomic Epidemiological Laboratory |
| EPI_ISL_436490 | UPMC Clinical Laboratory | Microbial Genome Sequencing Center, Microbial Genomic Epidemiological Laboratory |
| EPI_ISL_436491 | UPMC Clinical Laboratory | Microbial Genome Sequencing Center, Microbial Genomic Epidemiological Laboratory |
| EPI_ISL_436492 | UPMC Clinical Laboratory | Microbial Genome Sequencing Center, Microbial Genomic Epidemiological Laboratory |
| EPI_ISL_436493 | UPMC Clinical Laboratory | Microbial Genome Sequencing Center, Microbial Genomic Epidemiological Laboratory |
| EPI_ISL_436494 | UPMC Clinical Laboratory | Microbial Genome Sequencing Center, Microbial Genomic Epidemiological Laboratory |
| EPI_ISL_436495 | UPMC Clinical Laboratory | Microbial Genome Sequencing Center, Microbial Genomic Epidemiological Laboratory |
| EPI_ISL_436496 | UPMC Clinical Laboratory | Microbial Genome Sequencing Center, Microbial Genomic Epidemiological Laboratory |
| EPI_ISL_436497 | UPMC Clinical Laboratory | Microbial Genome Sequencing Center, Microbial Genomic Epidemiological Laboratory |
| EPI_ISL_436498 | UPMC Clinical Laboratory | Microbial Genome Sequencing Center, Microbial Genomic Epidemiological Laboratory |
| EPI_ISL_436499 | UPMC Clinical Laboratory | Microbial Genome Sequencing Center, Microbial Genomic Epidemiological Laboratory |
| EPI_ISL_436500 | UPMC Clinical Laboratory | Microbial Genome Sequencing Center, Microbial Genomic Epidemiological Laboratory |
| EPI_ISL_436501 | UPMC Clinical Laboratory | Microbial Genome Sequencing Center, Microbial Genomic Epidemiological Laboratory |
| EPI_ISL_436502 | UPMC Clinical Laboratory | Microbial Genome Sequencing Center, Microbial Genomic Epidemiological Laboratory |

| EPI_Accession_ID | Originating Laboratory | Submitting Laboratory |
|---|---|---|
| EPI_ISL_436503 | UPMC Clinical Laboratory | Microbial Genome Sequencing Center, Microbial Genomic Epidemiological Laboratory |
| EPI_ISL_436505 | Florida Bureau of Public Health Laboratories | Florida Bureau of Public Health Laboratories |
| EPI_ISL_436506 | Florida Bureau of Public Health Laboratories | Florida Bureau of Public Health Laboratories |
| EPI_ISL_436508 | Florida Bureau of Public Health Laboratories | Florida Bureau of Public Health Laboratories |
| EPI_ISL_436509 | Florida Bureau of Public Health Laboratories | Florida Bureau of Public Health Laboratories |
| EPI_ISL_436510 | Florida Bureau of Public Health Laboratories | Florida Bureau of Public Health Laboratories |
| EPI_ISL_436511 | Florida Bureau of Public Health Laboratories | Florida Bureau of Public Health Laboratories |
| EPI_ISL_436512 | Florida Bureau of Public Health Laboratories | Florida Bureau of Public Health Laboratories |
| EPI_ISL_436513 | Florida Bureau of Public Health Laboratories | Florida Bureau of Public Health Laboratories |
| EPI_ISL_436516 | Florida Bureau of Public Health Laboratories | Florida Bureau of Public Health Laboratories |
| EPI_ISL_436517 | Florida Bureau of Public Health Laboratories | Florida Bureau of Public Health Laboratories |
| EPI_ISL_436518 | Florida Bureau of Public Health Laboratories | Florida Bureau of Public Health Laboratories |
| EPI_ISL_436519 | Florida Bureau of Public Health Laboratories | Florida Bureau of Public Health Laboratories |
| EPI_ISL_436520 | Florida Bureau of Public Health Laboratories | Florida Bureau of Public Health Laboratories |
| EPI_ISL_436521 | Florida Bureau of Public Health Laboratories | Florida Bureau of Public Health Laboratories |
| EPI_ISL_436522 | Florida Bureau of Public Health Laboratories | Florida Bureau of Public Health Laboratories |
| EPI_ISL_436523 | Florida Bureau of Public Health Laboratories | Florida Bureau of Public Health Laboratories |
| EPI_ISL_436524 | Florida Bureau of Public Health Laboratories | Florida Bureau of Public Health Laboratories |
| EPI_ISL_436525 | Florida Bureau of Public Health Laboratories | Florida Bureau of Public Health Laboratories |
| EPI_ISL_436526 | Florida Bureau of Public Health Laboratories | Florida Bureau of Public Health Laboratories |
| EPI_ISL_436527 | Florida Bureau of Public Health Laboratories | Florida Bureau of Public Health Laboratories |
| EPI_ISL_436528 | Florida Bureau of Public Health Laboratories | Florida Bureau of Public Health Laboratories |
| EPI_ISL_436529 | Florida Bureau of Public Health Laboratories | Florida Bureau of Public Health Laboratories |
| EPI_ISL_436530 | Florida Bureau of Public Health Laboratories | Florida Bureau of Public Health Laboratories |
| EPI_ISL_436531 | Florida Bureau of Public Health Laboratories | Florida Bureau of Public Health Laboratories |
| EPI_ISL_436532 | Florida Bureau of Public Health Laboratories | Florida Bureau of Public Health Laboratories |
| EPI_ISL_436533 | Florida Bureau of Public Health Laboratories | Florida Bureau of Public Health Laboratories |
| EPI_ISL_436534 | Florida Bureau of Public Health Laboratories | Florida Bureau of Public Health Laboratories |
| EPI_ISL_436535 | Florida Bureau of Public Health Laboratories | Florida Bureau of Public Health Laboratories |
| EPI_ISL_436536 | Florida Bureau of Public Health Laboratories | Florida Bureau of Public Health Laboratories |
| EPI_ISL_436537 | Florida Bureau of Public Health Laboratories | Florida Bureau of Public Health Laboratories |
| EPI_ISL_436538 | Florida Bureau of Public Health Laboratories | Florida Bureau of Public Health Laboratories |

| EPI_Accession_ID | Originating Laboratory | Submitting Laboratory |
|---|---|---|
| EPI_ISL_436539 | Florida Bureau of Public Health Laboratories | Florida Bureau of Public Health Laboratories |
| EPI_ISL_436540 | Florida Bureau of Public Health Laboratories | Florida Bureau of Public Health Laboratories |
| EPI_ISL_436541 | Florida Bureau of Public Health Laboratories | Florida Bureau of Public Health Laboratories |
| EPI_ISL_436543 | Florida Bureau of Public Health Laboratories | Florida Bureau of Public Health Laboratories |
| EPI_ISL_436544 | Florida Bureau of Public Health Laboratories | Florida Bureau of Public Health Laboratories |
| EPI_ISL_436545 | Florida Bureau of Public Health Laboratories | Florida Bureau of Public Health Laboratories |
| EPI_ISL_436546 | Florida Bureau of Public Health Laboratories | Florida Bureau of Public Health Laboratories |
| EPI_ISL_436547 | Florida Bureau of Public Health Laboratories | Florida Bureau of Public Health Laboratories |
| EPI_ISL_436549 | Florida Bureau of Public Health Laboratories | Florida Bureau of Public Health Laboratories |
| EPI_ISL_436550 | Florida Bureau of Public Health Laboratories | Florida Bureau of Public Health Laboratories |
| EPI_ISL_436551 | Florida Bureau of Public Health Laboratories | Florida Bureau of Public Health Laboratories |
| EPI_ISL_436552 | Florida Bureau of Public Health Laboratories | Florida Bureau of Public Health Laboratories |
| EPI_ISL_436553 | Florida Bureau of Public Health Laboratories | Florida Bureau of Public Health Laboratories |
| EPI_ISL_436554 | Florida Bureau of Public Health Laboratories | Florida Bureau of Public Health Laboratories |
| EPI_ISL_436555 | Florida Bureau of Public Health Laboratories | Florida Bureau of Public Health Laboratories |
| EPI_ISL_436558 | Florida Bureau of Public Health Laboratories | Florida Bureau of Public Health Laboratories |
| EPI_ISL_436559 | Florida Bureau of Public Health Laboratories | Florida Bureau of Public Health Laboratories |
| EPI_ISL_436562 | Florida Bureau of Public Health Laboratories | Florida Bureau of Public Health Laboratories |
| EPI_ISL_436563 | Florida Bureau of Public Health Laboratories | Florida Bureau of Public Health Laboratories |
| EPI_ISL_436564 | University of Wisconsin-Madison AIDS Vaccine Research Laboratories | University of Wisconsin-Madison AIDS Vaccine Research Laboratories |
| EPI_ISL_436565 | University of Wisconsin-Madison AIDS Vaccine Research Laboratories | University of Wisconsin-Madison AIDS Vaccine Research Laboratories |
| EPI_ISL_436566 | University of Wisconsin-Madison AIDS Vaccine Research Laboratories | University of Wisconsin-Madison AIDS Vaccine Research Laboratories |
| EPI_ISL_436567 | University of Wisconsin-Madison AIDS Vaccine Research Laboratories | University of Wisconsin-Madison AIDS Vaccine Research Laboratories |
| EPI_ISL_436568 | University of Wisconsin-Madison AIDS Vaccine Research Laboratories | University of Wisconsin-Madison AIDS Vaccine Research Laboratories |
| EPI_ISL_436569 | University of Wisconsin-Madison AIDS Vaccine Research Laboratories | University of Wisconsin-Madison AIDS Vaccine Research Laboratories |
| EPI_ISL_436570 | University of Wisconsin-Madison AIDS Vaccine Research Laboratories | University of Wisconsin-Madison AIDS Vaccine Research Laboratories |

| EPI_Accession_ID | Originating Laboratory | Submitting Laboratory |
|---|---|---|
| EPI_ISL_436571 | University of Wisconsin-Madison AIDS Vaccine Research Laboratories | University of Wisconsin-Madison AIDS Vaccine Research Laboratories |
| EPI_ISL_436572 | University of Wisconsin-Madison AIDS Vaccine Research Laboratories | University of Wisconsin-Madison AIDS Vaccine Research Laboratories |
| EPI_ISL_436573 | University of Wisconsin-Madison AIDS Vaccine Research Laboratories | University of Wisconsin-Madison AIDS Vaccine Research Laboratories |
| EPI_ISL_436574 | University of Wisconsin-Madison AIDS Vaccine Research Laboratories | University of Wisconsin-Madison AIDS Vaccine Research Laboratories |
| EPI_ISL_436575 | University of Wisconsin-Madison AIDS Vaccine Research Laboratories | University of Wisconsin-Madison AIDS Vaccine Research Laboratories |
| EPI_ISL_436576 | University of Wisconsin-Madison AIDS Vaccine Research Laboratories | University of Wisconsin-Madison AIDS Vaccine Research Laboratories |
| EPI_ISL_436577 | University of Wisconsin-Madison AIDS Vaccine Research Laboratories | University of Wisconsin-Madison AIDS Vaccine Research Laboratories |
| EPI_ISL_436578 | University of Wisconsin-Madison AIDS Vaccine Research Laboratories | University of Wisconsin-Madison AIDS Vaccine Research Laboratories |
| EPI_ISL_436579 | University of Wisconsin-Madison AIDS Vaccine Research Laboratories | University of Wisconsin-Madison AIDS Vaccine Research Laboratories |
| EPI_ISL_436580 | University of Wisconsin-Madison AIDS Vaccine Research Laboratories | University of Wisconsin-Madison AIDS Vaccine Research Laboratories |
| EPI_ISL_436581 | University of Wisconsin-Madison AIDS Vaccine Research Laboratories | University of Wisconsin-Madison AIDS Vaccine Research Laboratories |
| EPI_ISL_436582 | University of Wisconsin-Madison AIDS Vaccine Research Laboratories | University of Wisconsin-Madison AIDS Vaccine Research Laboratories |
| EPI_ISL_436583 | University of Wisconsin-Madison AIDS Vaccine Research Laboratories | University of Wisconsin-Madison AIDS Vaccine Research Laboratories |
| EPI_ISL_436584 | University of Wisconsin-Madison AIDS Vaccine Research Laboratories | University of Wisconsin-Madison AIDS Vaccine Research Laboratories |
| EPI_ISL_436585 | University of Wisconsin-Madison AIDS Vaccine Research Laboratories | University of Wisconsin-Madison AIDS Vaccine Research Laboratories |
| EPI_ISL_436586 | University of Wisconsin-Madison AIDS Vaccine Research Laboratories | University of Wisconsin-Madison AIDS Vaccine Research Laboratories |

| EPI_Accession_ID | Originating Laboratory | Submitting Laboratory |
|---|---|---|
| EPI_ISL_436587 | University of Wisconsin-Madison AIDS Vaccine Research Laboratories | University of Wisconsin-Madison AIDS Vaccine Research Laboratories |
| EPI_ISL_436588 | University of Wisconsin-Madison AIDS Vaccine Research Laboratories | University of Wisconsin-Madison AIDS Vaccine Research Laboratories |
| EPI_ISL_436589 | University of Wisconsin-Madison AIDS Vaccine Research Laboratories | University of Wisconsin-Madison AIDS Vaccine Research Laboratories |
| EPI_ISL_436590 | University of Wisconsin-Madison AIDS Vaccine Research Laboratories | University of Wisconsin-Madison AIDS Vaccine Research Laboratories |
| EPI_ISL_436591 | University of Wisconsin-Madison AIDS Vaccine Research Laboratories | University of Wisconsin-Madison AIDS Vaccine Research Laboratories |
| EPI_ISL_436592 | University of Wisconsin-Madison AIDS Vaccine Research Laboratories | University of Wisconsin-Madison AIDS Vaccine Research Laboratories |
| EPI_ISL_436593 | University of Wisconsin-Madison AIDS Vaccine Research Laboratories | University of Wisconsin-Madison AIDS Vaccine Research Laboratories |
| EPI_ISL_436594 | University of Wisconsin-Madison AIDS Vaccine Research Laboratories | University of Wisconsin-Madison AIDS Vaccine Research Laboratories |
| EPI_ISL_436595 | University of Wisconsin-Madison AIDS Vaccine Research Laboratories | University of Wisconsin-Madison AIDS Vaccine Research Laboratories |
| EPI_ISL_436596 | University of Wisconsin-Madison AIDS Vaccine Research Laboratories | University of Wisconsin-Madison AIDS Vaccine Research Laboratories |
| EPI_ISL_436597 | University of Wisconsin-Madison AIDS Vaccine Research Laboratories | University of Wisconsin-Madison AIDS Vaccine Research Laboratories |
| EPI_ISL_436598 | University of Wisconsin-Madison AIDS Vaccine Research Laboratories | University of Wisconsin-Madison AIDS Vaccine Research Laboratories |
| EPI_ISL_436599 | University of Wisconsin-Madison AIDS Vaccine Research Laboratories | University of Wisconsin-Madison AIDS Vaccine Research Laboratories |
| EPI_ISL_436600 | University of Wisconsin-Madison AIDS Vaccine Research Laboratories | University of Wisconsin-Madison AIDS Vaccine Research Laboratories |
| EPI_ISL_436601 | University of Wisconsin-Madison AIDS Vaccine Research Laboratories | University of Wisconsin-Madison AIDS Vaccine Research Laboratories |
| EPI_ISL_436602 | University of Wisconsin-Madison AIDS Vaccine Research Laboratories | University of Wisconsin-Madison AIDS Vaccine Research Laboratories |

| EPI_Accession_ID | Originating Laboratory | Submitting Laboratory |
|---|---|---|
| EPI_ISL_436603 | University of Wisconsin-Madison AIDS Vaccine Research Laboratories | University of Wisconsin-Madison AIDS Vaccine Research Laboratories |
| EPI_ISL_436604 | University of Wisconsin-Madison AIDS Vaccine Research Laboratories | University of Wisconsin-Madison AIDS Vaccine Research Laboratories |
| EPI_ISL_436605 | University of Wisconsin-Madison AIDS Vaccine Research Laboratories | University of Wisconsin-Madison AIDS Vaccine Research Laboratories |
| EPI_ISL_436606 | University of Wisconsin-Madison AIDS Vaccine Research Laboratories | University of Wisconsin-Madison AIDS Vaccine Research Laboratories |
| EPI_ISL_436607 | University of Wisconsin-Madison AIDS Vaccine Research Laboratories | University of Wisconsin-Madison AIDS Vaccine Research Laboratories |
| EPI_ISL_436608 | University of Wisconsin-Madison AIDS Vaccine Research Laboratories | University of Wisconsin-Madison AIDS Vaccine Research Laboratories |
| EPI_ISL_436609 | University of Wisconsin-Madison AIDS Vaccine Research Laboratories | University of Wisconsin-Madison AIDS Vaccine Research Laboratories |
| EPI_ISL_436610 | University of Wisconsin-Madison AIDS Vaccine Research Laboratories | University of Wisconsin-Madison AIDS Vaccine Research Laboratories |
| EPI_ISL_436611 | University of Wisconsin-Madison AIDS Vaccine Research Laboratories | University of Wisconsin-Madison AIDS Vaccine Research Laboratories |
| EPI_ISL_436612 | University of Wisconsin-Madison AIDS Vaccine Research Laboratories | University of Wisconsin-Madison AIDS Vaccine Research Laboratories |
| EPI_ISL_436613 | University of Wisconsin-Madison AIDS Vaccine Research Laboratories | University of Wisconsin-Madison AIDS Vaccine Research Laboratories |
| EPI_ISL_436614 | University of Wisconsin-Madison AIDS Vaccine Research Laboratories | University of Wisconsin-Madison AIDS Vaccine Research Laboratories |
| EPI_ISL_436615 | University of Wisconsin-Madison AIDS Vaccine Research Laboratories | University of Wisconsin-Madison AIDS Vaccine Research Laboratories |
| EPI_ISL_436616 | University of Wisconsin-Madison AIDS Vaccine Research Laboratories | University of Wisconsin-Madison AIDS Vaccine Research Laboratories |
| EPI_ISL_436617 | University of Wisconsin-Madison AIDS Vaccine Research Laboratories | University of Wisconsin-Madison AIDS Vaccine Research Laboratories |
| EPI_ISL_436618 | University of Wisconsin-Madison AIDS Vaccine Research Laboratories | University of Wisconsin-Madison AIDS Vaccine Research Laboratories |

| EPI_Accession_ID | Originating Laboratory | Submitting Laboratory |
|---|---|---|
| EPI_ISL_436619 | University of Wisconsin-Madison AIDS Vaccine Research Laboratories | University of Wisconsin-Madison AIDS Vaccine Research Laboratories |
| EPI_ISL_436620 | University of Wisconsin-Madison AIDS Vaccine Research Laboratories | University of Wisconsin-Madison AIDS Vaccine Research Laboratories |
| EPI_ISL_436621 | University of Wisconsin-Madison AIDS Vaccine Research Laboratories | University of Wisconsin-Madison AIDS Vaccine Research Laboratories |
| EPI_ISL_436622 | University of Wisconsin-Madison AIDS Vaccine Research Laboratories | University of Wisconsin-Madison AIDS Vaccine Research Laboratories |
| EPI_ISL_436623 | University of Wisconsin-Madison AIDS Vaccine Research Laboratories | University of Wisconsin-Madison AIDS Vaccine Research Laboratories |
| EPI_ISL_436624 | University of Wisconsin-Madison AIDS Vaccine Research Laboratories | University of Wisconsin-Madison AIDS Vaccine Research Laboratories |
| EPI_ISL_436625 | University of Wisconsin-Madison AIDS Vaccine Research Laboratories | University of Wisconsin-Madison AIDS Vaccine Research Laboratories |
| EPI_ISL_436626 | University of Wisconsin-Madison AIDS Vaccine Research Laboratories | University of Wisconsin-Madison AIDS Vaccine Research Laboratories |
| EPI_ISL_436627 | University of Wisconsin-Madison AIDS Vaccine Research Laboratories | University of Wisconsin-Madison AIDS Vaccine Research Laboratories |
| EPI_ISL_436628 | University of Wisconsin-Madison AIDS Vaccine Research Laboratories | University of Wisconsin-Madison AIDS Vaccine Research Laboratories |
| EPI_ISL_436629 | University of Wisconsin-Madison AIDS Vaccine Research Laboratories | University of Wisconsin-Madison AIDS Vaccine Research Laboratories |
| EPI_ISL_436630 | University of Wisconsin-Madison AIDS Vaccine Research Laboratories | University of Wisconsin-Madison AIDS Vaccine Research Laboratories |
| EPI_ISL_436631 | University of Wisconsin-Madison AIDS Vaccine Research Laboratories | University of Wisconsin-Madison AIDS Vaccine Research Laboratories |
| EPI_ISL_436632 | University of Wisconsin-Madison AIDS Vaccine Research Laboratories | University of Wisconsin-Madison AIDS Vaccine Research Laboratories |
| EPI_ISL_436633 | University of Wisconsin-Madison AIDS Vaccine Research Laboratories | University of Wisconsin-Madison AIDS Vaccine Research Laboratories |
| EPI_ISL_436634 | University of Wisconsin-Madison AIDS Vaccine Research Laboratories | University of Wisconsin-Madison AIDS Vaccine Research Laboratories |

| EPI_Accession_ID | Originating Laboratory | Submitting Laboratory |
|---|---|---|
| EPI_ISL_436635 | University of Wisconsin-Madison AIDS Vaccine Research Laboratories | University of Wisconsin-Madison AIDS Vaccine Research Laboratories |
| EPI_ISL_436636 | University of Wisconsin-Madison AIDS Vaccine Research Laboratories | University of Wisconsin-Madison AIDS Vaccine Research Laboratories |
| EPI_ISL_436637 | University of Wisconsin-Madison AIDS Vaccine Research Laboratories | University of Wisconsin-Madison AIDS Vaccine Research Laboratories |
| EPI_ISL_436638 | University of Wisconsin-Madison AIDS Vaccine Research Laboratories | University of Wisconsin-Madison AIDS Vaccine Research Laboratories |
| EPI_ISL_436639 | University of Wisconsin-Madison AIDS Vaccine Research Laboratories | University of Wisconsin-Madison AIDS Vaccine Research Laboratories |
| EPI_ISL_436640 | University of Wisconsin-Madison AIDS Vaccine Research Laboratories | University of Wisconsin-Madison AIDS Vaccine Research Laboratories |
| EPI_ISL_436641 | County of Santa Clara Public Health Department | Chan-Zuckerberg Biohub |
| EPI_ISL_436642 | County of Santa Clara Public Health Department | Chan-Zuckerberg Biohub |
| EPI_ISL_436643 | County of Santa Clara Public Health Department | Chan-Zuckerberg Biohub |
| EPI_ISL_436644 | County of Santa Clara Public Health Department | Chan-Zuckerberg Biohub |
| EPI_ISL_436645 | County of Santa Clara Public Health Department | Chan-Zuckerberg Biohub |
| EPI_ISL_436646 | County of Santa Clara Public Health Department | Chan-Zuckerberg Biohub |
| EPI_ISL_436647 | County of Santa Clara Public Health Department | Chan-Zuckerberg Biohub |
| EPI_ISL_436648 | County of Santa Clara Public Health Department | Chan-Zuckerberg Biohub |
| EPI_ISL_436649 | County of Santa Clara Public Health Department | Chan-Zuckerberg Biohub |
| EPI_ISL_436650 | County of Santa Clara Public Health Department | Chan-Zuckerberg Biohub |
| EPI_ISL_436651 | County of Santa Clara Public Health Department | Chan-Zuckerberg Biohub |
| EPI_ISL_436652 | County of Santa Clara Public Health Department | Chan-Zuckerberg Biohub |
| EPI_ISL_436653 | County of Santa Clara Public Health Department | Chan-Zuckerberg Biohub |
| EPI_ISL_436654 | County of Santa Clara Public Health Department | Chan-Zuckerberg Biohub |
| EPI_ISL_436655 | County of Santa Clara Public Health Department | Chan-Zuckerberg Biohub |
| EPI_ISL_436656 | County of Santa Clara Public Health Department | Chan-Zuckerberg Biohub |
| EPI_ISL_436657 | County of Santa Clara Public Health Department | Chan-Zuckerberg Biohub |
| EPI_ISL_436658 | County of Santa Clara Public Health Department | Chan-Zuckerberg Biohub |
| EPI_ISL_436659 | County of Santa Clara Public Health Department | Chan-Zuckerberg Biohub |
| EPI_ISL_436660 | County of Santa Clara Public Health Department | Chan-Zuckerberg Biohub |
| EPI_ISL_436661 | County of Santa Clara Public Health Department | Chan-Zuckerberg Biohub |

| EPI_Accession_ID | Originating Laboratory | Submitting Laboratory |
|---|---|---|
| EPI_ISL_436662 | County of Santa Clara Public Health Department | Chan-Zuckerberg Biohub |
| EPI_ISL_436663 | County of Santa Clara Public Health Department | Chan-Zuckerberg Biohub |
| EPI_ISL_436664 | County of Santa Clara Public Health Department | Chan-Zuckerberg Biohub |
| EPI_ISL_436665 | County of Santa Clara Public Health Department | Chan-Zuckerberg Biohub |
| EPI_ISL_436666 | County of Santa Clara Public Health Department | Chan-Zuckerberg Biohub |
| EPI_ISL_436667 | County of Santa Clara Public Health Department | Chan-Zuckerberg Biohub |
| EPI_ISL_436668 | County of Santa Clara Public Health Department | Chan-Zuckerberg Biohub |
| EPI_ISL_436669 | County of Santa Clara Public Health Department | Chan-Zuckerberg Biohub |
| EPI_ISL_436670 | County of Santa Clara Public Health Department | Chan-Zuckerberg Biohub |
| EPI_ISL_436671 | County of Santa Clara Public Health Department | Chan-Zuckerberg Biohub |
| EPI_ISL_436672 | County of Santa Clara Public Health Department | Chan-Zuckerberg Biohub |
| EPI_ISL_436673 | County of Santa Clara Public Health Department | Chan-Zuckerberg Biohub |
| EPI_ISL_436674 | County of Santa Clara Public Health Department | Chan-Zuckerberg Biohub |
| EPI_ISL_436675 | County of Santa Clara Public Health Department | Chan-Zuckerberg Biohub |
| EPI_ISL_436676 | County of Santa Clara Public Health Department | Chan-Zuckerberg Biohub |
| EPI_ISL_436677 | County of Santa Clara Public Health Department | Chan-Zuckerberg Biohub |
| EPI_ISL_436678 | County of Santa Clara Public Health Department | Chan-Zuckerberg Biohub |
| EPI_ISL_436679 | County of Santa Clara Public Health Department | Chan-Zuckerberg Biohub |
| EPI_ISL_436680 | County of Santa Clara Public Health Department | Chan-Zuckerberg Biohub |
| EPI_ISL_436681 | County of Santa Clara Public Health Department | Chan-Zuckerberg Biohub |
| EPI_ISL_436682 | County of Santa Clara Public Health Department | Chan-Zuckerberg Biohub |
| EPI_ISL_436683 | County of Santa Clara Public Health Department | Chan-Zuckerberg Biohub |
| EPI_ISL_436684 | KRISP, KZN Research Innovation and Sequencing Platform | KRISP, KZN Research Innovation and Sequencing Platform |
| EPI_ISL_436685 | KRISP, KZN Research Innovation and Sequencing Platform | KRISP, KZN Research Innovation and Sequencing Platform |
| EPI_ISL_436686 | KRISP, KZN Research Innovation and Sequencing Platform | KRISP, KZN Research Innovation and Sequencing Platform |
| EPI_ISL_436687 | KRISP, KZN Research Innovation and Sequencing Platform | KRISP, KZN Research Innovation and Sequencing Platform |

| EPI_Accession_ID | Originating Laboratory | Submitting Laboratory |
|---|---|---|
| EPI_ISL_436688 | Victorian Infectious Diseases Reference Laboratory (VIDRL) | Microbiological Diagnostic Unit Public Health Laboratory and Victorian Infectious Diseases Reference Laboratory, The Peter Doherty Institute for Infection & Immunity |
| EPI_ISL_436689 | Victorian Infectious Diseases Reference Laboratory (VIDRL) | Microbiological Diagnostic Unit Public Health Laboratory and Victorian Infectious Diseases Reference Laboratory, The Peter Doherty Institute for Infection & Immunity |
| EPI_ISL_436715 | Genomics and Computational Biology Lab, Scientific Research Institute of Physical-Chemical Medicine, FMBA of Russia | Genomics and Computational Biology Lab, Scientific Research Institute of Physical-Chemical Medicine, FMBA of Russia |
| EPI_ISL_436716 | Genomics and Computational Biology Lab, Scientific Research Institute of Physical-Chemical Medicine, FMBA of Russia | Genomics and Computational Biology Lab, Scientific Research Institute of Physical-Chemical Medicine, FMBA of Russia |
| EPI_ISL_436717 | Genomics and Computational Biology Lab, Scientific Research Institute of Physical-Chemical Medicine, FMBA of Russia | Genomics and Computational Biology Lab, Scientific Research Institute of Physical-Chemical Medicine, FMBA of Russia |
| EPI_ISL_436718 | Ospedale Regionale San Salvatore | Istituto Zooprofilattico Sperimentale dellâ€™Abruzzo e Molise â€œG. Caporaleâ€� |
| EPI_ISL_436719 | Ospedale Civile S. Liberatore di Atri | Istituto Zooprofilattico Sperimentale dellâ€™Abruzzo e Molise â€œG. Caporaleâ€� |
| EPI_ISL_436720 | Ospedale Civile S. Liberatore di Atri | Istituto Zooprofilattico Sperimentale dellâ€™Abruzzo e Molise â€œG. Caporaleâ€� |
| EPI_ISL_436721 | Ospedale Civile S. Liberatore di Atri | Istituto Zooprofilattico Sperimentale dellâ€™Abruzzo e Molise â€œG. Caporaleâ€� |
| EPI_ISL_436722 | Ospedale Civile S. Liberatore di Atri | Istituto Zooprofilattico Sperimentale dellâ€™Abruzzo e Molise â€œG. Caporaleâ€� |
| EPI_ISL_436723 | Ospedale Civile Giuseppe Mazzini | Istituto Zooprofilattico Sperimentale dellâ€™Abruzzo e Molise â€œG. Caporaleâ€� |
| EPI_ISL_436724 | Ospedale Civile S. Liberatore di Atri | Istituto Zooprofilattico Sperimentale dellâ€™Abruzzo e Molise â€œG. Caporaleâ€� |
| EPI_ISL_436725 | RSA/RP Villa San Giovanni - Gruppo Edos | Istituto Zooprofilattico Sperimentale dellâ€™Abruzzo e Molise â€œG. Caporaleâ€� |

| EPI_Accession_ID | Originating Laboratory | Submitting Laboratory |
| --- | --- | --- |
| EPI_ISL_436726 | SERVIZIO DI IGIENE E SANITÃ€ PUBBLICA ASL Teramo | Istituto Zooprofilattico Sperimentale dellâ€™Abruzzo e Molise â€œG. Caporaleâ€� |
| EPI_ISL_436727 | SERVIZIO DI IGIENE E SANITÃ€ PUBBLICA ASL Teramo | Istituto Zooprofilattico Sperimentale dellâ€™Abruzzo e Molise â€œG. Caporaleâ€� |
| EPI_ISL_436728 | SERVIZIO DI IGIENE E SANITÃ€ PUBBLICA ASL Teramo | Istituto Zooprofilattico Sperimentale dellâ€™Abruzzo e Molise â€œG. Caporaleâ€� |
| EPI_ISL_436729 | SERVIZIO DI IGIENE E SANITÃ€ PUBBLICA ASL Teramo | Istituto Zooprofilattico Sperimentale dellâ€™Abruzzo e Molise â€œG. Caporaleâ€� |
| EPI_ISL_436730 | Servizio di igiene epidemiologia e sanitÃ pubblica (Siesp) Chieti | Istituto Zooprofilattico Sperimentale dellâ€™Abruzzo e Molise â€œG. Caporaleâ€� |
| EPI_ISL_436731 | Ospedale Civile S. Liberatore di Atri | Istituto Zooprofilattico Sperimentale dellâ€™Abruzzo e Molise â€œG. Caporaleâ€� |
| EPI_ISL_436732 | Ospedale Civile S. Liberatore di Atri | Istituto Zooprofilattico Sperimentale dellâ€™Abruzzo e Molise â€œG. Caporaleâ€� |
| EPI_ISL_436733 | NYU Langone Health | Departments of Pathology and Medicine, New York University School of Medicine |
| EPI_ISL_436734 | NYU Langone Health | Departments of Pathology and Medicine, New York University School of Medicine |
| EPI_ISL_436735 | NYU Langone Health | Departments of Pathology and Medicine, New York University School of Medicine |
| EPI_ISL_436736 | NYU Langone Health | Departments of Pathology and Medicine, New York University School of Medicine |
| EPI_ISL_436737 | NYU Langone Health | Departments of Pathology and Medicine, New York University School of Medicine |
| EPI_ISL_436738 | NYU Langone Health | Departments of Pathology and Medicine, New York University School of Medicine |
| EPI_ISL_436739 | NYU Langone Health | Departments of Pathology and Medicine, New York University School of Medicine |
| EPI_ISL_436740 | NYU Langone Health | Departments of Pathology and Medicine, New York University School of Medicine |
| EPI_ISL_436741 | NYU Langone Health | Departments of Pathology and Medicine, New York University School of Medicine |

| EPI_Accession_ID | Originating Laboratory | Submitting Laboratory |
| --- | --- | --- |
| EPI_ISL_436742 | NYU Langone Health | Departments of Pathology and Medicine, New York University School of Medicine |
| EPI_ISL_436743 | NYU Langone Health | Departments of Pathology and Medicine, New York University School of Medicine |
| EPI_ISL_436800 | Michigan Department of Health and Human Services, Bureau of Laboratories | Michigan Department of Health and Human Services, Bureau of Laboratories |
| EPI_ISL_436801 | Michigan Department of Health and Human Services, Bureau of Laboratories | Michigan Department of Health and Human Services, Bureau of Laboratories |
| EPI_ISL_436802 | Michigan Department of Health and Human Services, Bureau of Laboratories | Michigan Department of Health and Human Services, Bureau of Laboratories |
| EPI_ISL_436803 | Michigan Department of Health and Human Services, Bureau of Laboratories | Michigan Department of Health and Human Services, Bureau of Laboratories |
| EPI_ISL_436804 | Michigan Department of Health and Human Services, Bureau of Laboratories | Michigan Department of Health and Human Services, Bureau of Laboratories |
| EPI_ISL_436805 | Michigan Department of Health and Human Services, Bureau of Laboratories | Michigan Department of Health and Human Services, Bureau of Laboratories |
| EPI_ISL_436806 | Michigan Department of Health and Human Services, Bureau of Laboratories | Michigan Department of Health and Human Services, Bureau of Laboratories |
| EPI_ISL_436807 | Michigan Department of Health and Human Services, Bureau of Laboratories | Michigan Department of Health and Human Services, Bureau of Laboratories |
| EPI_ISL_436808 | Michigan Department of Health and Human Services, Bureau of Laboratories | Michigan Department of Health and Human Services, Bureau of Laboratories |
| EPI_ISL_436809 | Michigan Department of Health and Human Services, Bureau of Laboratories | Michigan Department of Health and Human Services, Bureau of Laboratories |
| EPI_ISL_436810 | Michigan Department of Health and Human Services, Bureau of Laboratories | Michigan Department of Health and Human Services, Bureau of Laboratories |
| EPI_ISL_436811 | Michigan Department of Health and Human Services, Bureau of Laboratories | Michigan Department of Health and Human Services, Bureau of Laboratories |
| EPI_ISL_436812 | Michigan Department of Health and Human Services, Bureau of Laboratories | Michigan Department of Health and Human Services, Bureau of Laboratories |
| EPI_ISL_436813 | Michigan Department of Health and Human Services, Bureau of Laboratories | Michigan Department of Health and Human Services, Bureau of Laboratories |

| EPI_Accession_ID | Originating Laboratory | Submitting Laboratory |
|---|---|---|
| EPI_ISL_436814 | Michigan Department of Health and Human Services, Bureau of Laboratories | Michigan Department of Health and Human Services, Bureau of Laboratories |
| EPI_ISL_436815 | Michigan Department of Health and Human Services, Bureau of Laboratories | Michigan Department of Health and Human Services, Bureau of Laboratories |
| EPI_ISL_436816 | Michigan Department of Health and Human Services, Bureau of Laboratories | Michigan Department of Health and Human Services, Bureau of Laboratories |
| EPI_ISL_436817 | Michigan Department of Health and Human Services, Bureau of Laboratories | Michigan Department of Health and Human Services, Bureau of Laboratories |
| EPI_ISL_436818 | Michigan Department of Health and Human Services, Bureau of Laboratories | Michigan Department of Health and Human Services, Bureau of Laboratories |
| EPI_ISL_436819 | Michigan Department of Health and Human Services, Bureau of Laboratories | Michigan Department of Health and Human Services, Bureau of Laboratories |
| EPI_ISL_436820 | Michigan Department of Health and Human Services, Bureau of Laboratories | Michigan Department of Health and Human Services, Bureau of Laboratories |
| EPI_ISL_436821 | Michigan Department of Health and Human Services, Bureau of Laboratories | Michigan Department of Health and Human Services, Bureau of Laboratories |
| EPI_ISL_436822 | Michigan Department of Health and Human Services, Bureau of Laboratories | Michigan Department of Health and Human Services, Bureau of Laboratories |
| EPI_ISL_436823 | Michigan Department of Health and Human Services, Bureau of Laboratories | Michigan Department of Health and Human Services, Bureau of Laboratories |
| EPI_ISL_436824 | Michigan Department of Health and Human Services, Bureau of Laboratories | Michigan Department of Health and Human Services, Bureau of Laboratories |
| EPI_ISL_436825 | Michigan Department of Health and Human Services, Bureau of Laboratories | Michigan Department of Health and Human Services, Bureau of Laboratories |
| EPI_ISL_436826 | Michigan Department of Health and Human Services, Bureau of Laboratories | Michigan Department of Health and Human Services, Bureau of Laboratories |
| EPI_ISL_436827 | Michigan Department of Health and Human Services, Bureau of Laboratories | Michigan Department of Health and Human Services, Bureau of Laboratories |
| EPI_ISL_436828 | Michigan Department of Health and Human Services, Bureau of Laboratories | Michigan Department of Health and Human Services, Bureau of Laboratories |
| EPI_ISL_436829 | Michigan Department of Health and Human Services, Bureau of Laboratories | Michigan Department of Health and Human Services, Bureau of Laboratories |

| EPI_Accession_ID | Originating Laboratory | Submitting Laboratory |
|---|---|---|
| EPI_ISL_436830 | Michigan Department of Health and Human Services, Bureau of Laboratories | Michigan Department of Health and Human Services, Bureau of Laboratories |
| EPI_ISL_436831 | Michigan Department of Health and Human Services, Bureau of Laboratories | Michigan Department of Health and Human Services, Bureau of Laboratories |
| EPI_ISL_436832 | Michigan Department of Health and Human Services, Bureau of Laboratories | Michigan Department of Health and Human Services, Bureau of Laboratories |
| EPI_ISL_436833 | Michigan Department of Health and Human Services, Bureau of Laboratories | Michigan Department of Health and Human Services, Bureau of Laboratories |
| EPI_ISL_436834 | Michigan Department of Health and Human Services, Bureau of Laboratories | Michigan Department of Health and Human Services, Bureau of Laboratories |
| EPI_ISL_436835 | Michigan Department of Health and Human Services, Bureau of Laboratories | Michigan Department of Health and Human Services, Bureau of Laboratories |
| EPI_ISL_436836 | Michigan Department of Health and Human Services, Bureau of Laboratories | Michigan Department of Health and Human Services, Bureau of Laboratories |
| EPI_ISL_436837 | Michigan Department of Health and Human Services, Bureau of Laboratories | Michigan Department of Health and Human Services, Bureau of Laboratories |
| EPI_ISL_436838 | Michigan Department of Health and Human Services, Bureau of Laboratories | Michigan Department of Health and Human Services, Bureau of Laboratories |
| EPI_ISL_436839 | Michigan Department of Health and Human Services, Bureau of Laboratories | Michigan Department of Health and Human Services, Bureau of Laboratories |
| EPI_ISL_436840 | Michigan Department of Health and Human Services, Bureau of Laboratories | Michigan Department of Health and Human Services, Bureau of Laboratories |
| EPI_ISL_436841 | Michigan Department of Health and Human Services, Bureau of Laboratories | Michigan Department of Health and Human Services, Bureau of Laboratories |
| EPI_ISL_436842 | Michigan Department of Health and Human Services, Bureau of Laboratories | Michigan Department of Health and Human Services, Bureau of Laboratories |
| EPI_ISL_436843 | Michigan Department of Health and Human Services, Bureau of Laboratories | Michigan Department of Health and Human Services, Bureau of Laboratories |
| EPI_ISL_436844 | Michigan Department of Health and Human Services, Bureau of Laboratories | Michigan Department of Health and Human Services, Bureau of Laboratories |
| EPI_ISL_436845 | Michigan Department of Health and Human Services, Bureau of Laboratories | Michigan Department of Health and Human Services, Bureau of Laboratories |

| EPI_Accession_ID | Originating Laboratory | Submitting Laboratory |
| --- | --- | --- |
| EPI_ISL_436846 | Michigan Department of Health and Human Services, Bureau of Laboratories | Michigan Department of Health and Human Services, Bureau of Laboratories |
| EPI_ISL_436847 | Michigan Department of Health and Human Services, Bureau of Laboratories | Michigan Department of Health and Human Services, Bureau of Laboratories |
| EPI_ISL_436848 | Michigan Department of Health and Human Services, Bureau of Laboratories | Michigan Department of Health and Human Services, Bureau of Laboratories |
| EPI_ISL_436849 | Michigan Department of Health and Human Services, Bureau of Laboratories | Michigan Department of Health and Human Services, Bureau of Laboratories |
| EPI_ISL_436850 | Michigan Department of Health and Human Services, Bureau of Laboratories | Michigan Department of Health and Human Services, Bureau of Laboratories |
| EPI_ISL_436851 | Michigan Department of Health and Human Services, Bureau of Laboratories | Michigan Department of Health and Human Services, Bureau of Laboratories |
| EPI_ISL_436852 | Michigan Department of Health and Human Services, Bureau of Laboratories | Michigan Department of Health and Human Services, Bureau of Laboratories |
| EPI_ISL_436853 | Michigan Department of Health and Human Services, Bureau of Laboratories | Michigan Department of Health and Human Services, Bureau of Laboratories |
| EPI_ISL_436854 | Michigan Department of Health and Human Services, Bureau of Laboratories | Michigan Department of Health and Human Services, Bureau of Laboratories |
| EPI_ISL_436855 | Michigan Department of Health and Human Services, Bureau of Laboratories | Michigan Department of Health and Human Services, Bureau of Laboratories |
| EPI_ISL_436856 | Michigan Department of Health and Human Services, Bureau of Laboratories | Michigan Department of Health and Human Services, Bureau of Laboratories |
| EPI_ISL_436857 | Michigan Department of Health and Human Services, Bureau of Laboratories | Michigan Department of Health and Human Services, Bureau of Laboratories |
| EPI_ISL_436858 | Michigan Department of Health and Human Services, Bureau of Laboratories | Michigan Department of Health and Human Services, Bureau of Laboratories |
| EPI_ISL_436859 | Michigan Department of Health and Human Services, Bureau of Laboratories | Michigan Department of Health and Human Services, Bureau of Laboratories |
| EPI_ISL_436860 | Michigan Department of Health and Human Services, Bureau of Laboratories | Michigan Department of Health and Human Services, Bureau of Laboratories |
| EPI_ISL_436861 | Michigan Department of Health and Human Services, Bureau of Laboratories | Michigan Department of Health and Human Services, Bureau of Laboratories |

| EPI_Accession_ID | Originating Laboratory | Submitting Laboratory |
|---|---|---|
| EPI_ISL_436862 | Michigan Department of Health and Human Services, Bureau of Laboratories | Michigan Department of Health and Human Services, Bureau of Laboratories |
| EPI_ISL_436863 | Michigan Department of Health and Human Services, Bureau of Laboratories | Michigan Department of Health and Human Services, Bureau of Laboratories |
| EPI_ISL_436864 | Michigan Department of Health and Human Services, Bureau of Laboratories | Michigan Department of Health and Human Services, Bureau of Laboratories |
| EPI_ISL_436865 | Michigan Department of Health and Human Services, Bureau of Laboratories | Michigan Department of Health and Human Services, Bureau of Laboratories |
| EPI_ISL_436866 | Michigan Department of Health and Human Services, Bureau of Laboratories | Michigan Department of Health and Human Services, Bureau of Laboratories |
| EPI_ISL_436867 | Michigan Department of Health and Human Services, Bureau of Laboratories | Michigan Department of Health and Human Services, Bureau of Laboratories |
| EPI_ISL_436868 | Michigan Department of Health and Human Services, Bureau of Laboratories | Michigan Department of Health and Human Services, Bureau of Laboratories |
| EPI_ISL_436869 | Michigan Department of Health and Human Services, Bureau of Laboratories | Michigan Department of Health and Human Services, Bureau of Laboratories |
| EPI_ISL_436870 | Michigan Department of Health and Human Services, Bureau of Laboratories | Michigan Department of Health and Human Services, Bureau of Laboratories |
| EPI_ISL_436871 | Michigan Department of Health and Human Services, Bureau of Laboratories | Michigan Department of Health and Human Services, Bureau of Laboratories |
| EPI_ISL_436872 | Michigan Department of Health and Human Services, Bureau of Laboratories | Michigan Department of Health and Human Services, Bureau of Laboratories |
| EPI_ISL_436873 | Michigan Department of Health and Human Services, Bureau of Laboratories | Michigan Department of Health and Human Services, Bureau of Laboratories |
| EPI_ISL_436874 | Michigan Department of Health and Human Services, Bureau of Laboratories | Michigan Department of Health and Human Services, Bureau of Laboratories |
| EPI_ISL_436875 | Michigan Department of Health and Human Services, Bureau of Laboratories | Michigan Department of Health and Human Services, Bureau of Laboratories |
| EPI_ISL_436876 | Michigan Department of Health and Human Services, Bureau of Laboratories | Michigan Department of Health and Human Services, Bureau of Laboratories |
| EPI_ISL_436877 | Michigan Department of Health and Human Services, Bureau of Laboratories | Michigan Department of Health and Human Services, Bureau of Laboratories |

| EPI_Accession_ID | Originating Laboratory | Submitting Laboratory |
|---|---|---|
| EPI_ISL_436878 | Michigan Department of Health and Human Services, Bureau of Laboratories | Michigan Department of Health and Human Services, Bureau of Laboratories |
| EPI_ISL_436879 | Michigan Department of Health and Human Services, Bureau of Laboratories | Michigan Department of Health and Human Services, Bureau of Laboratories |
| EPI_ISL_436880 | Michigan Department of Health and Human Services, Bureau of Laboratories | Michigan Department of Health and Human Services, Bureau of Laboratories |
| EPI_ISL_436881 | Michigan Department of Health and Human Services, Bureau of Laboratories | Michigan Department of Health and Human Services, Bureau of Laboratories |
| EPI_ISL_436882 | Michigan Department of Health and Human Services, Bureau of Laboratories | Michigan Department of Health and Human Services, Bureau of Laboratories |
| EPI_ISL_436883 | Michigan Department of Health and Human Services, Bureau of Laboratories | Michigan Department of Health and Human Services, Bureau of Laboratories |
| EPI_ISL_436884 | Michigan Department of Health and Human Services, Bureau of Laboratories | Michigan Department of Health and Human Services, Bureau of Laboratories |
| EPI_ISL_436885 | Michigan Department of Health and Human Services, Bureau of Laboratories | Michigan Department of Health and Human Services, Bureau of Laboratories |
| EPI_ISL_436886 | Michigan Department of Health and Human Services, Bureau of Laboratories | Michigan Department of Health and Human Services, Bureau of Laboratories |
| EPI_ISL_436887 | Michigan Department of Health and Human Services, Bureau of Laboratories | Michigan Department of Health and Human Services, Bureau of Laboratories |
| EPI_ISL_436888 | Michigan Department of Health and Human Services, Bureau of Laboratories | Michigan Department of Health and Human Services, Bureau of Laboratories |
| EPI_ISL_436889 | Michigan Department of Health and Human Services, Bureau of Laboratories | Michigan Department of Health and Human Services, Bureau of Laboratories |
| EPI_ISL_436890 | Michigan Department of Health and Human Services, Bureau of Laboratories | Michigan Department of Health and Human Services, Bureau of Laboratories |
| EPI_ISL_436891 | Gundersen Molecular Diagnostics Laboratory | Kabara Cancer Research Institute |
| EPI_ISL_436892 | Gundersen Molecular Diagnostics Laboratory | Kabara Cancer Research Institute |
| EPI_ISL_436893 | Gundersen Molecular Diagnostics Laboratory | Kabara Cancer Research Institute |
| EPI_ISL_436894 | Gundersen Molecular Diagnostics Laboratory | Kabara Cancer Research Institute |
| EPI_ISL_436895 | Gundersen Molecular Diagnostics Laboratory | Kabara Cancer Research Institute |
| EPI_ISL_436896 | Gundersen Molecular Diagnostics Laboratory | Kabara Cancer Research Institute |
| EPI_ISL_436897 | Gundersen Molecular Diagnostics Laboratory | Kabara Cancer Research Institute |

| EPI_Accession_ID | Originating Laboratory | Submitting Laboratory |
|---|---|---|
| EPI_ISL_436898 | Gundersen Molecular Diagnostics Laboratory | Kabara Cancer Research Institute |
| EPI_ISL_436899 | Gundersen Molecular Diagnostics Laboratory | Kabara Cancer Research Institute |
| EPI_ISL_436900 | Gundersen Molecular Diagnostics Laboratory | Kabara Cancer Research Institute |
| EPI_ISL_436901 | Utah Public Health Laboratory | Utah Public Health Laboratory |
| EPI_ISL_436902 | Utah Public Health Laboratory | Utah Public Health Laboratory |
| EPI_ISL_436903 | Utah Public Health Laboratory | Utah Public Health Laboratory |
| EPI_ISL_436904 | Utah Public Health Laboratory | Utah Public Health Laboratory |
| EPI_ISL_436905 | Utah Public Health Laboratory | Utah Public Health Laboratory |
| EPI_ISL_436906 | Utah Public Health Laboratory | Utah Public Health Laboratory |
| EPI_ISL_436907 | Utah Public Health Laboratory | Utah Public Health Laboratory |
| EPI_ISL_436908 | Utah Public Health Laboratory | Utah Public Health Laboratory |
| EPI_ISL_436909 | Utah Public Health Laboratory | Utah Public Health Laboratory |
| EPI_ISL_436910 | Utah Public Health Laboratory | Utah Public Health Laboratory |
| EPI_ISL_436911 | Utah Public Health Laboratory | Utah Public Health Laboratory |
| EPI_ISL_436912 | Utah Public Health Laboratory | Utah Public Health Laboratory |
| EPI_ISL_436913 | Utah Public Health Laboratory | Utah Public Health Laboratory |
| EPI_ISL_436914 | Utah Public Health Laboratory | Utah Public Health Laboratory |
| EPI_ISL_436915 | Utah Public Health Laboratory | Utah Public Health Laboratory |
| EPI_ISL_436916 | Utah Public Health Laboratory | Utah Public Health Laboratory |
| EPI_ISL_436917 | Utah Public Health Laboratory | Utah Public Health Laboratory |
| EPI_ISL_436918 | Utah Public Health Laboratory | Utah Public Health Laboratory |
| EPI_ISL_436919 | Utah Public Health Laboratory | Utah Public Health Laboratory |
| EPI_ISL_436920 | Utah Public Health Laboratory | Utah Public Health Laboratory |
| EPI_ISL_436921 | Utah Public Health Laboratory | Utah Public Health Laboratory |
| EPI_ISL_436922 | Utah Public Health Laboratory | Utah Public Health Laboratory |
| EPI_ISL_436923 | Utah Public Health Laboratory | Utah Public Health Laboratory |
| EPI_ISL_436924 | Utah Public Health Laboratory | Utah Public Health Laboratory |
| EPI_ISL_436925 | Utah Public Health Laboratory | Utah Public Health Laboratory |
| EPI_ISL_436926 | xÂ² | Utah Public Health Laboratory |
| EPI_ISL_436927 | Utah Public Health Laboratory | Utah Public Health Laboratory |
| EPI_ISL_436928 | Utah Public Health Laboratory | Utah Public Health Laboratory |
| EPI_ISL_436929 | Utah Public Health Laboratory | Utah Public Health Laboratory |
| EPI_ISL_436930 | Utah Public Health Laboratory | Utah Public Health Laboratory |

| EPI_Accession_ID | Originating Laboratory | Submitting Laboratory |
|---|---|---|
| EPI_ISL_436931 | Utah Public Health Laboratory | Utah Public Health Laboratory |
| EPI_ISL_436932 | Utah Public Health Laboratory | Utah Public Health Laboratory |
| EPI_ISL_436933 | Utah Public Health Laboratory | Utah Public Health Laboratory |
| EPI_ISL_436934 | Utah Public Health Laboratory | Utah Public Health Laboratory |
| EPI_ISL_436935 | Utah Public Health Laboratory | Utah Public Health Laboratory |
| EPI_ISL_436936 | Utah Public Health Laboratory | Utah Public Health Laboratory |
| EPI_ISL_436937 | Utah Public Health Laboratory | Utah Public Health Laboratory |
| EPI_ISL_436938 | Utah Public Health Laboratory | Utah Public Health Laboratory |
| EPI_ISL_436939 | Ochsner Health | Bioinfoexperts, LLC |
| EPI_ISL_436940 | Ochsner Health | Bioinfoexperts, LLC |
| EPI_ISL_436941 | Ochsner Health | Bioinfoexperts, LLC |
| EPI_ISL_436942 | Ochsner Health | Bioinfoexperts, LLC |
| EPI_ISL_436943 | Ochsner Health | Bioinfoexperts, LLC |
| EPI_ISL_436944 | Ochsner Health | Bioinfoexperts, LLC |
| EPI_ISL_436945 | Ochsner Health | Bioinfoexperts, LLC |
| EPI_ISL_436946 | Ochsner Health | Bioinfoexperts, LLC |
| EPI_ISL_436947 | Ochsner Health | Bioinfoexperts, LLC |
| EPI_ISL_436948 | Ochsner Health | Bioinfoexperts, LLC |
| EPI_ISL_436949 | Ochsner Health | Bioinfoexperts, LLC |
| EPI_ISL_436950 | Ochsner Health | Bioinfoexperts, LLC |
| EPI_ISL_436951 | Ochsner Health | Bioinfoexperts, LLC |
| EPI_ISL_436952 | Ochsner Health | Bioinfoexperts, LLC |
| EPI_ISL_436953 | Ochsner Health | Bioinfoexperts, LLC |
| EPI_ISL_436954 | Ochsner Health | Bioinfoexperts, LLC |
| EPI_ISL_436955 | Ochsner Health | Bioinfoexperts, LLC |
| EPI_ISL_436956 | Ochsner Health | Bioinfoexperts, LLC |
| EPI_ISL_436957 | Ochsner Health | Bioinfoexperts, LLC |
| EPI_ISL_436958 | Ochsner Health | Bioinfoexperts, LLC |
| EPI_ISL_436959 | Ochsner Health | Bioinfoexperts, LLC |
| EPI_ISL_436960 | Ochsner Health | Bioinfoexperts, LLC |
| EPI_ISL_436961 | Ochsner Health | Bioinfoexperts, LLC |

| EPI_Accession_ID | Originating Laboratory | Submitting Laboratory |
|---|---|---|
| EPI_ISL_436962 | Department of Virus and Microbiological Special Diagnostics, Statens Serum Institut, Copenhagen, Denmark, Artillerivej 5, 2300 Copenahgen S | Albertsen lab, Department of Chemistry and Bioscience, Aalborg University, Denmark |
| EPI_ISL_436963 | Department of Virus and Microbiological Special Diagnostics, Statens Serum Institut, Copenhagen, Denmark, Artillerivej 5, 2300 Copenahgen S | Albertsen lab, Department of Chemistry and Bioscience, Aalborg University, Denmark |
| EPI_ISL_436964 | Department of Virus and Microbiological Special Diagnostics, Statens Serum Institut, Copenhagen, Denmark, Artillerivej 5, 2300 Copenahgen S | Albertsen lab, Department of Chemistry and Bioscience, Aalborg University, Denmark |
| EPI_ISL_436965 | Department of Virus and Microbiological Special Diagnostics, Statens Serum Institut, Copenhagen, Denmark, Artillerivej 5, 2300 Copenahgen S | Albertsen lab, Department of Chemistry and Bioscience, Aalborg University, Denmark |
| EPI_ISL_436966 | Department of Virus and Microbiological Special Diagnostics, Statens Serum Institut, Copenhagen, Denmark, Artillerivej 5, 2300 Copenahgen S | Albertsen lab, Department of Chemistry and Bioscience, Aalborg University, Denmark |
| EPI_ISL_436967 | Department of Virus and Microbiological Special Diagnostics, Statens Serum Institut, Copenhagen, Denmark, Artillerivej 5, 2300 Copenahgen S | Albertsen lab, Department of Chemistry and Bioscience, Aalborg University, Denmark |
| EPI_ISL_436968 | Department of Virus and Microbiological Special Diagnostics, Statens Serum Institut, Copenhagen, Denmark, Artillerivej 5, 2300 Copenahgen S | Albertsen lab, Department of Chemistry and Bioscience, Aalborg University, Denmark |
| EPI_ISL_436969 | Department of Virus and Microbiological Special Diagnostics, Statens Serum Institut, Copenhagen, Denmark, Artillerivej 5, 2300 Copenahgen S | Albertsen lab, Department of Chemistry and Bioscience, Aalborg University, Denmark |
| EPI_ISL_436970 | Department of Virus and Microbiological Special Diagnostics, Statens Serum Institut, Copenhagen, Denmark, Artillerivej 5, 2300 Copenahgen S | Albertsen lab, Department of Chemistry and Bioscience, Aalborg University, Denmark |
| EPI_ISL_436971 | Department of Virus and Microbiological Special Diagnostics, Statens Serum Institut, Copenhagen, Denmark, Artillerivej 5, 2300 Copenahgen S | Albertsen lab, Department of Chemistry and Bioscience, Aalborg University, Denmark |
| EPI_ISL_436972 | Department of Virus and Microbiological Special Diagnostics, Statens Serum Institut, Copenhagen, Denmark, Artillerivej 5, 2300 Copenahgen S | Albertsen lab, Department of Chemistry and Bioscience, Aalborg University, Denmark |

| EPI_Accession_ID | Originating Laboratory | Submitting Laboratory |
|---|---|---|
| EPI_ISL_436973 | Department of Virus and Microbiological Special Diagnostics, Statens Serum Institut, Copenhagen, Denmark, Artillerivej 5, 2300 Copenahgen S | Albertsen lab, Department of Chemistry and Bioscience, Aalborg University, Denmark |
| EPI_ISL_436974 | Department of Virus and Microbiological Special Diagnostics, Statens Serum Institut, Copenhagen, Denmark, Artillerivej 5, 2300 Copenahgen S | Albertsen lab, Department of Chemistry and Bioscience, Aalborg University, Denmark |
| EPI_ISL_436975 | Department of Virus and Microbiological Special Diagnostics, Statens Serum Institut, Copenhagen, Denmark, Artillerivej 5, 2300 Copenahgen S | Albertsen lab, Department of Chemistry and Bioscience, Aalborg University, Denmark |
| EPI_ISL_436976 | Department of Virus and Microbiological Special Diagnostics, Statens Serum Institut, Copenhagen, Denmark, Artillerivej 5, 2300 Copenahgen S | Albertsen lab, Department of Chemistry and Bioscience, Aalborg University, Denmark |
| EPI_ISL_436977 | Department of Virus and Microbiological Special Diagnostics, Statens Serum Institut, Copenhagen, Denmark, Artillerivej 5, 2300 Copenahgen S | Albertsen lab, Department of Chemistry and Bioscience, Aalborg University, Denmark |
| EPI_ISL_436978 | Department of Virus and Microbiological Special Diagnostics, Statens Serum Institut, Copenhagen, Denmark, Artillerivej 5, 2300 Copenahgen S | Albertsen lab, Department of Chemistry and Bioscience, Aalborg University, Denmark |
| EPI_ISL_436979 | Department of Virus and Microbiological Special Diagnostics, Statens Serum Institut, Copenhagen, Denmark, Artillerivej 5, 2300 Copenahgen S | Albertsen lab, Department of Chemistry and Bioscience, Aalborg University, Denmark |
| EPI_ISL_436980 | Department of Virus and Microbiological Special Diagnostics, Statens Serum Institut, Copenhagen, Denmark, Artillerivej 5, 2300 Copenahgen S | Albertsen lab, Department of Chemistry and Bioscience, Aalborg University, Denmark |
| EPI_ISL_436981 | Department of Virus and Microbiological Special Diagnostics, Statens Serum Institut, Copenhagen, Denmark, Artillerivej 5, 2300 Copenahgen S | Albertsen lab, Department of Chemistry and Bioscience, Aalborg University, Denmark |
| EPI_ISL_436982 | Department of Virus and Microbiological Special Diagnostics, Statens Serum Institut, Copenhagen, Denmark, Artillerivej 5, 2300 Copenahgen S | Albertsen lab, Department of Chemistry and Bioscience, Aalborg University, Denmark |
| EPI_ISL_436983 | Department of Virus and Microbiological Special Diagnostics, Statens Serum Institut, Copenhagen, Denmark, Artillerivej 5, 2300 Copenahgen S | Albertsen lab, Department of Chemistry and Bioscience, Aalborg University, Denmark |

| EPI_Accession_ID | Originating Laboratory | Submitting Laboratory |
|---|---|---|
| EPI_ISL_436984 | Department of Virus and Microbiological Special Diagnostics, Statens Serum Institut, Copenhagen, Denmark, Artillerivej 5, 2300 Copenahgen S | Albertsen lab, Department of Chemistry and Bioscience, Aalborg University, Denmark |
| EPI_ISL_436985 | Department of Virus and Microbiological Special Diagnostics, Statens Serum Institut, Copenhagen, Denmark, Artillerivej 5, 2300 Copenahgen S | Albertsen lab, Department of Chemistry and Bioscience, Aalborg University, Denmark |
| EPI_ISL_436986 | Department of Virus and Microbiological Special Diagnostics, Statens Serum Institut, Copenhagen, Denmark, Artillerivej 5, 2300 Copenahgen S | Albertsen lab, Department of Chemistry and Bioscience, Aalborg University, Denmark |
| EPI_ISL_436987 | Department of Virus and Microbiological Special Diagnostics, Statens Serum Institut, Copenhagen, Denmark, Artillerivej 5, 2300 Copenahgen S | Albertsen lab, Department of Chemistry and Bioscience, Aalborg University, Denmark |
| EPI_ISL_436988 | Department of Virus and Microbiological Special Diagnostics, Statens Serum Institut, Copenhagen, Denmark, Artillerivej 5, 2300 Copenahgen S | Albertsen lab, Department of Chemistry and Bioscience, Aalborg University, Denmark |
| EPI_ISL_436989 | Department of Virus and Microbiological Special Diagnostics, Statens Serum Institut, Copenhagen, Denmark, Artillerivej 5, 2300 Copenahgen S | Albertsen lab, Department of Chemistry and Bioscience, Aalborg University, Denmark |
| EPI_ISL_436990 | Department of Virus and Microbiological Special Diagnostics, Statens Serum Institut, Copenhagen, Denmark, Artillerivej 5, 2300 Copenahgen S | Albertsen lab, Department of Chemistry and Bioscience, Aalborg University, Denmark |
| EPI_ISL_436991 | Department of Virus and Microbiological Special Diagnostics, Statens Serum Institut, Copenhagen, Denmark, Artillerivej 5, 2300 Copenahgen S | Albertsen lab, Department of Chemistry and Bioscience, Aalborg University, Denmark |
| EPI_ISL_436992 | Department of Virus and Microbiological Special Diagnostics, Statens Serum Institut, Copenhagen, Denmark, Artillerivej 5, 2300 Copenahgen S | Albertsen lab, Department of Chemistry and Bioscience, Aalborg University, Denmark |
| EPI_ISL_436993 | Department of Virus and Microbiological Special Diagnostics, Statens Serum Institut, Copenhagen, Denmark, Artillerivej 5, 2300 Copenahgen S | Albertsen lab, Department of Chemistry and Bioscience, Aalborg University, Denmark |
| EPI_ISL_436994 | Department of Virus and Microbiological Special Diagnostics, Statens Serum Institut, Copenhagen, Denmark, Artillerivej 5, 2300 Copenahgen S | Albertsen lab, Department of Chemistry and Bioscience, Aalborg University, Denmark |

| EPI_Accession_ID | Originating Laboratory | Submitting Laboratory |
|---|---|---|
| EPI_ISL_436995 | Department of Virus and Microbiological Special Diagnostics, Statens Serum Institut, Copenhagen, Denmark, Artillerivej 5, 2300 Copenahgen S | Albertsen lab, Department of Chemistry and Bioscience, Aalborg University, Denmark |
| EPI_ISL_436996 | Department of Virus and Microbiological Special Diagnostics, Statens Serum Institut, Copenhagen, Denmark, Artillerivej 5, 2300 Copenahgen S | Albertsen lab, Department of Chemistry and Bioscience, Aalborg University, Denmark |
| EPI_ISL_436997 | Department of Virus and Microbiological Special Diagnostics, Statens Serum Institut, Copenhagen, Denmark, Artillerivej 5, 2300 Copenahgen S | Albertsen lab, Department of Chemistry and Bioscience, Aalborg University, Denmark |
| EPI_ISL_436998 | Department of Virus and Microbiological Special Diagnostics, Statens Serum Institut, Copenhagen, Denmark, Artillerivej 5, 2300 Copenahgen S | Albertsen lab, Department of Chemistry and Bioscience, Aalborg University, Denmark |
| EPI_ISL_436999 | Department of Virus and Microbiological Special Diagnostics, Statens Serum Institut, Copenhagen, Denmark, Artillerivej 5, 2300 Copenahgen S | Albertsen lab, Department of Chemistry and Bioscience, Aalborg University, Denmark |
| EPI_ISL_437000 | Department of Virus and Microbiological Special Diagnostics, Statens Serum Institut, Copenhagen, Denmark, Artillerivej 5, 2300 Copenahgen S | Albertsen lab, Department of Chemistry and Bioscience, Aalborg University, Denmark |
| EPI_ISL_437001 | Department of Virus and Microbiological Special Diagnostics, Statens Serum Institut, Copenhagen, Denmark, Artillerivej 5, 2300 Copenahgen S | Albertsen lab, Department of Chemistry and Bioscience, Aalborg University, Denmark |
| EPI_ISL_437002 | Department of Virus and Microbiological Special Diagnostics, Statens Serum Institut, Copenhagen, Denmark, Artillerivej 5, 2300 Copenahgen S | Albertsen lab, Department of Chemistry and Bioscience, Aalborg University, Denmark |
| EPI_ISL_437003 | Department of Virus and Microbiological Special Diagnostics, Statens Serum Institut, Copenhagen, Denmark, Artillerivej 5, 2300 Copenahgen S | Albertsen lab, Department of Chemistry and Bioscience, Aalborg University, Denmark |
| EPI_ISL_437004 | Department of Virus and Microbiological Special Diagnostics, Statens Serum Institut, Copenhagen, Denmark, Artillerivej 5, 2300 Copenahgen S | Albertsen lab, Department of Chemistry and Bioscience, Aalborg University, Denmark |
| EPI_ISL_437005 | Department of Virus and Microbiological Special Diagnostics, Statens Serum Institut, Copenhagen, Denmark, Artillerivej 5, 2300 Copenahgen S | Albertsen lab, Department of Chemistry and Bioscience, Aalborg University, Denmark |

| EPI_Accession_ID | Originating Laboratory | Submitting Laboratory |
|---|---|---|
| EPI_ISL_437006 | Department of Virus and Microbiological Special Diagnostics, Statens Serum Institut, Copenhagen, Denmark, Artillerivej 5, 2300 Copenahgen S | Albertsen lab, Department of Chemistry and Bioscience, Aalborg University, Denmark |
| EPI_ISL_437007 | Department of Virus and Microbiological Special Diagnostics, Statens Serum Institut, Copenhagen, Denmark, Artillerivej 5, 2300 Copenahgen S | Albertsen lab, Department of Chemistry and Bioscience, Aalborg University, Denmark |
| EPI_ISL_437008 | Department of Virus and Microbiological Special Diagnostics, Statens Serum Institut, Copenhagen, Denmark, Artillerivej 5, 2300 Copenahgen S | Albertsen lab, Department of Chemistry and Bioscience, Aalborg University, Denmark |
| EPI_ISL_437009 | Department of Virus and Microbiological Special Diagnostics, Statens Serum Institut, Copenhagen, Denmark, Artillerivej 5, 2300 Copenahgen S | Albertsen lab, Department of Chemistry and Bioscience, Aalborg University, Denmark |
| EPI_ISL_437010 | Department of Virus and Microbiological Special Diagnostics, Statens Serum Institut, Copenhagen, Denmark, Artillerivej 5, 2300 Copenahgen S | Albertsen lab, Department of Chemistry and Bioscience, Aalborg University, Denmark |
| EPI_ISL_437011 | Department of Virus and Microbiological Special Diagnostics, Statens Serum Institut, Copenhagen, Denmark, Artillerivej 5, 2300 Copenahgen S | Albertsen lab, Department of Chemistry and Bioscience, Aalborg University, Denmark |
| EPI_ISL_437012 | Department of Virus and Microbiological Special Diagnostics, Statens Serum Institut, Copenhagen, Denmark, Artillerivej 5, 2300 Copenahgen S | Albertsen lab, Department of Chemistry and Bioscience, Aalborg University, Denmark |
| EPI_ISL_437013 | Department of Virus and Microbiological Special Diagnostics, Statens Serum Institut, Copenhagen, Denmark, Artillerivej 5, 2300 Copenahgen S | Albertsen lab, Department of Chemistry and Bioscience, Aalborg University, Denmark |
| EPI_ISL_437014 | Department of Virus and Microbiological Special Diagnostics, Statens Serum Institut, Copenhagen, Denmark, Artillerivej 5, 2300 Copenahgen S | Albertsen lab, Department of Chemistry and Bioscience, Aalborg University, Denmark |
| EPI_ISL_437015 | Department of Virus and Microbiological Special Diagnostics, Statens Serum Institut, Copenhagen, Denmark, Artillerivej 5, 2300 Copenahgen S | Albertsen lab, Department of Chemistry and Bioscience, Aalborg University, Denmark |
| EPI_ISL_437016 | Department of Virus and Microbiological Special Diagnostics, Statens Serum Institut, Copenhagen, Denmark, Artillerivej 5, 2300 Copenahgen S | Albertsen lab, Department of Chemistry and Bioscience, Aalborg University, Denmark |

| EPI_Accession_ID | Originating Laboratory | Submitting Laboratory |
|---|---|---|
| EPI_ISL_437017 | Department of Virus and Microbiological Special Diagnostics, Statens Serum Institut, Copenhagen, Denmark, Artillerivej 5, 2300 Copenahgen S | Albertsen lab, Department of Chemistry and Bioscience, Aalborg University, Denmark |
| EPI_ISL_437018 | Department of Virus and Microbiological Special Diagnostics, Statens Serum Institut, Copenhagen, Denmark, Artillerivej 5, 2300 Copenahgen S | Albertsen lab, Department of Chemistry and Bioscience, Aalborg University, Denmark |
| EPI_ISL_437019 | Department of Virus and Microbiological Special Diagnostics, Statens Serum Institut, Copenhagen, Denmark, Artillerivej 5, 2300 Copenahgen S | Albertsen lab, Department of Chemistry and Bioscience, Aalborg University, Denmark |
| EPI_ISL_437020 | Department of Virus and Microbiological Special Diagnostics, Statens Serum Institut, Copenhagen, Denmark, Artillerivej 5, 2300 Copenahgen S | Albertsen lab, Department of Chemistry and Bioscience, Aalborg University, Denmark |
| EPI_ISL_437021 | Department of Virus and Microbiological Special Diagnostics, Statens Serum Institut, Copenhagen, Denmark, Artillerivej 5, 2300 Copenahgen S | Albertsen lab, Department of Chemistry and Bioscience, Aalborg University, Denmark |
| EPI_ISL_437022 | Department of Virus and Microbiological Special Diagnostics, Statens Serum Institut, Copenhagen, Denmark, Artillerivej 5, 2300 Copenahgen S | Albertsen lab, Department of Chemistry and Bioscience, Aalborg University, Denmark |
| EPI_ISL_437023 | Department of Virus and Microbiological Special Diagnostics, Statens Serum Institut, Copenhagen, Denmark, Artillerivej 5, 2300 Copenahgen S | Albertsen lab, Department of Chemistry and Bioscience, Aalborg University, Denmark |
| EPI_ISL_437024 | Department of Virus and Microbiological Special Diagnostics, Statens Serum Institut, Copenhagen, Denmark, Artillerivej 5, 2300 Copenahgen S | Albertsen lab, Department of Chemistry and Bioscience, Aalborg University, Denmark |
| EPI_ISL_437025 | Department of Virus and Microbiological Special Diagnostics, Statens Serum Institut, Copenhagen, Denmark, Artillerivej 5, 2300 Copenahgen S | Albertsen lab, Department of Chemistry and Bioscience, Aalborg University, Denmark |
| EPI_ISL_437026 | Department of Virus and Microbiological Special Diagnostics, Statens Serum Institut, Copenhagen, Denmark, Artillerivej 5, 2300 Copenahgen S | Albertsen lab, Department of Chemistry and Bioscience, Aalborg University, Denmark |
| EPI_ISL_437027 | Department of Virus and Microbiological Special Diagnostics, Statens Serum Institut, Copenhagen, Denmark, Artillerivej 5, 2300 Copenahgen S | Albertsen lab, Department of Chemistry and Bioscience, Aalborg University, Denmark |

| EPI_Accession_ID | Originating Laboratory | Submitting Laboratory |
|---|---|---|
| EPI_ISL_437028 | Department of Virus and Microbiological Special Diagnostics, Statens Serum Institut, Copenhagen, Denmark, Artillerivej 5, 2300 Copenahgen S | Albertsen lab, Department of Chemistry and Bioscience, Aalborg University, Denmark |
| EPI_ISL_437029 | Department of Virus and Microbiological Special Diagnostics, Statens Serum Institut, Copenhagen, Denmark, Artillerivej 5, 2300 Copenahgen S | Albertsen lab, Department of Chemistry and Bioscience, Aalborg University, Denmark |
| EPI_ISL_437030 | Department of Virus and Microbiological Special Diagnostics, Statens Serum Institut, Copenhagen, Denmark, Artillerivej 5, 2300 Copenahgen S | Albertsen lab, Department of Chemistry and Bioscience, Aalborg University, Denmark |
| EPI_ISL_437031 | Department of Virus and Microbiological Special Diagnostics, Statens Serum Institut, Copenhagen, Denmark, Artillerivej 5, 2300 Copenahgen S | Albertsen lab, Department of Chemistry and Bioscience, Aalborg University, Denmark |
| EPI_ISL_437032 | Department of Virus and Microbiological Special Diagnostics, Statens Serum Institut, Copenhagen, Denmark, Artillerivej 5, 2300 Copenahgen S | Albertsen lab, Department of Chemistry and Bioscience, Aalborg University, Denmark |
| EPI_ISL_437033 | Department of Virus and Microbiological Special Diagnostics, Statens Serum Institut, Copenhagen, Denmark, Artillerivej 5, 2300 Copenahgen S | Albertsen lab, Department of Chemistry and Bioscience, Aalborg University, Denmark |
| EPI_ISL_437034 | Department of Virus and Microbiological Special Diagnostics, Statens Serum Institut, Copenhagen, Denmark, Artillerivej 5, 2300 Copenahgen S | Albertsen lab, Department of Chemistry and Bioscience, Aalborg University, Denmark |
| EPI_ISL_437035 | Department of Virus and Microbiological Special Diagnostics, Statens Serum Institut, Copenhagen, Denmark, Artillerivej 5, 2300 Copenahgen S | Albertsen lab, Department of Chemistry and Bioscience, Aalborg University, Denmark |
| EPI_ISL_437036 | Department of Virus and Microbiological Special Diagnostics, Statens Serum Institut, Copenhagen, Denmark, Artillerivej 5, 2300 Copenahgen S | Albertsen lab, Department of Chemistry and Bioscience, Aalborg University, Denmark |
| EPI_ISL_437037 | Department of Virus and Microbiological Special Diagnostics, Statens Serum Institut, Copenhagen, Denmark, Artillerivej 5, 2300 Copenahgen S | Albertsen lab, Department of Chemistry and Bioscience, Aalborg University, Denmark |
| EPI_ISL_437038 | Department of Virus and Microbiological Special Diagnostics, Statens Serum Institut, Copenhagen, Denmark, Artillerivej 5, 2300 Copenahgen S | Albertsen lab, Department of Chemistry and Bioscience, Aalborg University, Denmark |

| EPI_Accession_ID | Originating Laboratory | Submitting Laboratory |
|---|---|---|
| EPI_ISL_437039 | Department of Virus and Microbiological Special Diagnostics, Statens Serum Institut, Copenhagen, Denmark, Artillerivej 5, 2300 Copenahgen S | Albertsen lab, Department of Chemistry and Bioscience, Aalborg University, Denmark |
| EPI_ISL_437040 | Department of Virus and Microbiological Special Diagnostics, Statens Serum Institut, Copenhagen, Denmark, Artillerivej 5, 2300 Copenahgen S | Albertsen lab, Department of Chemistry and Bioscience, Aalborg University, Denmark |
| EPI_ISL_437041 | Department of Virus and Microbiological Special Diagnostics, Statens Serum Institut, Copenhagen, Denmark, Artillerivej 5, 2300 Copenahgen S | Albertsen lab, Department of Chemistry and Bioscience, Aalborg University, Denmark |
| EPI_ISL_437042 | Department of Virus and Microbiological Special Diagnostics, Statens Serum Institut, Copenhagen, Denmark, Artillerivej 5, 2300 Copenahgen S | Albertsen lab, Department of Chemistry and Bioscience, Aalborg University, Denmark |
| EPI_ISL_437043 | County of Santa Clara Public Health | Chan-Zuckerberg Biohub |
| EPI_ISL_437044 | County of Santa Clara Public Health | Chan-Zuckerberg Biohub |
| EPI_ISL_437045 | County of Santa Clara Public Health | Chan-Zuckerberg Biohub |
| EPI_ISL_437046 | County of Santa Clara Public Health | Chan-Zuckerberg Biohub |
| EPI_ISL_437047 | County of Santa Clara Public Health | Chan-Zuckerberg Biohub |
| EPI_ISL_437048 | County of Santa Clara Public Health | Chan-Zuckerberg Biohub |
| EPI_ISL_437049 | County of Santa Clara Public Health | Chan-Zuckerberg Biohub |
| EPI_ISL_437050 | County of Santa Clara Public Health | Chan-Zuckerberg Biohub |
| EPI_ISL_437051 | County of Santa Clara Public Health | Chan-Zuckerberg Biohub |
| EPI_ISL_437052 | County of Santa Clara Public Health | Chan-Zuckerberg Biohub |
| EPI_ISL_437053 | County of Santa Clara Public Health | Chan-Zuckerberg Biohub |
| EPI_ISL_437054 | County of Santa Clara Public Health | Chan-Zuckerberg Biohub |
| EPI_ISL_437055 | County of Santa Clara Public Health | Chan-Zuckerberg Biohub |
| EPI_ISL_437056 | County of Santa Clara Public Health | Chan-Zuckerberg Biohub |
| EPI_ISL_437057 | County of Santa Clara Public Health | Chan-Zuckerberg Biohub |
| EPI_ISL_437058 | County of Santa Clara Public Health | Chan-Zuckerberg Biohub |
| EPI_ISL_437059 | County of Santa Clara Public Health | Chan-Zuckerberg Biohub |
| EPI_ISL_437060 | County of Santa Clara Public Health | Chan-Zuckerberg Biohub |
| EPI_ISL_437061 | County of Santa Clara Public Health | Chan-Zuckerberg Biohub |
| EPI_ISL_437062 | County of Santa Clara Public Health | Chan-Zuckerberg Biohub |
| EPI_ISL_437063 | County of Santa Clara Public Health | Chan-Zuckerberg Biohub |

| EPI_Accession_ID | Originating Laboratory | Submitting Laboratory |
|---|---|---|
| EPI_ISL_437064 | County of Santa Clara Public Health | Chan-Zuckerberg Biohub |
| EPI_ISL_437065 | County of Santa Clara Public Health | Chan-Zuckerberg Biohub |
| EPI_ISL_437066 | County of Santa Clara Public Health | Chan-Zuckerberg Biohub |
| EPI_ISL_437067 | County of Santa Clara Public Health | Chan-Zuckerberg Biohub |
| EPI_ISL_437068 | County of Santa Clara Public Health | Chan-Zuckerberg Biohub |
| EPI_ISL_437069 | County of Santa Clara Public Health | Chan-Zuckerberg Biohub |
| EPI_ISL_437070 | County of Santa Clara Public Health | Chan-Zuckerberg Biohub |
| EPI_ISL_437071 | County of Santa Clara Public Health | Chan-Zuckerberg Biohub |
| EPI_ISL_437072 | County of Santa Clara Public Health | Chan-Zuckerberg Biohub |
| EPI_ISL_437073 | County of Santa Clara Public Health | Chan-Zuckerberg Biohub |
| EPI_ISL_437074 | County of Santa Clara Public Health | Chan-Zuckerberg Biohub |
| EPI_ISL_437075 | County of Santa Clara Public Health | Chan-Zuckerberg Biohub |
| EPI_ISL_437076 | County of Santa Clara Public Health | Chan-Zuckerberg Biohub |
| EPI_ISL_437077 | County of Santa Clara Public Health | Chan-Zuckerberg Biohub |
| EPI_ISL_437078 | County of Santa Clara Public Health | Chan-Zuckerberg Biohub |
| EPI_ISL_437079 | County of Santa Clara Public Health | Chan-Zuckerberg Biohub |
| EPI_ISL_437080 | County of Santa Clara Public Health | Chan-Zuckerberg Biohub |
| EPI_ISL_437081 | County of Santa Clara Public Health | Chan-Zuckerberg Biohub |
| EPI_ISL_437082 | County of Santa Clara Public Health | Chan-Zuckerberg Biohub |
| EPI_ISL_437083 | County of Santa Clara Public Health | Chan-Zuckerberg Biohub |
| EPI_ISL_437084 | County of Santa Clara Public Health | Chan-Zuckerberg Biohub |
| EPI_ISL_437085 | County of Santa Clara Public Health | Chan-Zuckerberg Biohub |
| EPI_ISL_437086 | County of Santa Clara Public Health | Chan-Zuckerberg Biohub |
| EPI_ISL_437087 | County of Santa Clara Public Health | Chan-Zuckerberg Biohub |
| EPI_ISL_437088 | County of Santa Clara Public Health | Chan-Zuckerberg Biohub |
| EPI_ISL_437089 | Latvijas InfektoloÄ£ijas centrs | Latvian Biomedical Research and Study Centre |
| EPI_ISL_437090 | Latvijas InfektoloÄ£ijas centrs | Latvian Biomedical Research and Study Centre |
| EPI_ISL_437091 | Latvijas InfektoloÄ£ijas centrs | Latvian Biomedical Research and Study Centre |
| EPI_ISL_437092 | Latvijas InfektoloÄ£ijas centrs | Latvian Biomedical Research and Study Centre |
| EPI_ISL_437093 | Latvijas InfektoloÄ£ijas centrs | Latvian Biomedical Research and Study Centre |
| EPI_ISL_437094 | Latvijas InfektoloÄ£ijas centrs | Latvian Biomedical Research and Study Centre |
| EPI_ISL_437095 | Latvijas InfektoloÄ£ijas centrs | Latvian Biomedical Research and Study Centre |
| EPI_ISL_437096 | Latvijas InfektoloÄ£ijas centrs | Latvian Biomedical Research and Study Centre |

| EPI_Accession_ID | Originating Laboratory | Submitting Laboratory |
|---|---|---|
| EPI_ISL_437097 | Michigan Department of Health and Human Services, Bureau of Laboratories | Michigan Department of Health and Human Services, Bureau of Laboratories |
| EPI_ISL_437098 | Michigan Department of Health and Human Services, Bureau of Laboratories | Michigan Department of Health and Human Services, Bureau of Laboratories |
| EPI_ISL_437099 | Michigan Department of Health and Human Services, Bureau of Laboratories | Michigan Department of Health and Human Services, Bureau of Laboratories |
| EPI_ISL_437100 | Michigan Department of Health and Human Services, Bureau of Laboratories | Michigan Department of Health and Human Services, Bureau of Laboratories |
| EPI_ISL_437101 | Michigan Department of Health and Human Services, Bureau of Laboratories | Michigan Department of Health and Human Services, Bureau of Laboratories |
| EPI_ISL_437102 | Michigan Department of Health and Human Services, Bureau of Laboratories | Michigan Department of Health and Human Services, Bureau of Laboratories |
| EPI_ISL_437103 | Michigan Department of Health and Human Services, Bureau of Laboratories | Michigan Department of Health and Human Services, Bureau of Laboratories |
| EPI_ISL_437104 | Michigan Department of Health and Human Services, Bureau of Laboratories | Michigan Department of Health and Human Services, Bureau of Laboratories |
| EPI_ISL_437105 | Michigan Department of Health and Human Services, Bureau of Laboratories | Michigan Department of Health and Human Services, Bureau of Laboratories |
| EPI_ISL_437106 | Michigan Department of Health and Human Services, Bureau of Laboratories | Michigan Department of Health and Human Services, Bureau of Laboratories |
| EPI_ISL_437107 | Michigan Department of Health and Human Services, Bureau of Laboratories | Michigan Department of Health and Human Services, Bureau of Laboratories |
| EPI_ISL_437108 | Michigan Department of Health and Human Services, Bureau of Laboratories | Michigan Department of Health and Human Services, Bureau of Laboratories |
| EPI_ISL_437109 | Michigan Department of Health and Human Services, Bureau of Laboratories | Michigan Department of Health and Human Services, Bureau of Laboratories |
| EPI_ISL_437110 | Michigan Department of Health and Human Services, Bureau of Laboratories | Michigan Department of Health and Human Services, Bureau of Laboratories |
| EPI_ISL_437111 | Michigan Department of Health and Human Services, Bureau of Laboratories | Michigan Department of Health and Human Services, Bureau of Laboratories |
| EPI_ISL_437112 | Michigan Department of Health and Human Services, Bureau of Laboratories | Michigan Department of Health and Human Services, Bureau of Laboratories |

| EPI_Accession_ID | Originating Laboratory | Submitting Laboratory |
|---|---|---|
| EPI_ISL_437113 | Michigan Department of Health and Human Services, Bureau of Laboratories | Michigan Department of Health and Human Services, Bureau of Laboratories |
| EPI_ISL_437114 | Michigan Department of Health and Human Services, Bureau of Laboratories | Michigan Department of Health and Human Services, Bureau of Laboratories |
| EPI_ISL_437115 | Michigan Department of Health and Human Services, Bureau of Laboratories | Michigan Department of Health and Human Services, Bureau of Laboratories |
| EPI_ISL_437116 | Michigan Department of Health and Human Services, Bureau of Laboratories | Michigan Department of Health and Human Services, Bureau of Laboratories |
| EPI_ISL_437117 | Michigan Department of Health and Human Services, Bureau of Laboratories | Michigan Department of Health and Human Services, Bureau of Laboratories |
| EPI_ISL_437118 | Michigan Department of Health and Human Services, Bureau of Laboratories | Michigan Department of Health and Human Services, Bureau of Laboratories |
| EPI_ISL_437119 | Michigan Department of Health and Human Services, Bureau of Laboratories | Michigan Department of Health and Human Services, Bureau of Laboratories |
| EPI_ISL_437120 | Michigan Department of Health and Human Services, Bureau of Laboratories | Michigan Department of Health and Human Services, Bureau of Laboratories |
| EPI_ISL_437121 | Michigan Department of Health and Human Services, Bureau of Laboratories | Michigan Department of Health and Human Services, Bureau of Laboratories |
| EPI_ISL_437122 | Michigan Department of Health and Human Services, Bureau of Laboratories | Michigan Department of Health and Human Services, Bureau of Laboratories |
| EPI_ISL_437123 | Michigan Department of Health and Human Services, Bureau of Laboratories | Michigan Department of Health and Human Services, Bureau of Laboratories |
| EPI_ISL_437124 | Michigan Department of Health and Human Services, Bureau of Laboratories | Michigan Department of Health and Human Services, Bureau of Laboratories |
| EPI_ISL_437125 | Michigan Department of Health and Human Services, Bureau of Laboratories | Michigan Department of Health and Human Services, Bureau of Laboratories |
| EPI_ISL_437126 | Michigan Department of Health and Human Services, Bureau of Laboratories | Michigan Department of Health and Human Services, Bureau of Laboratories |
| EPI_ISL_437127 | Michigan Department of Health and Human Services, Bureau of Laboratories | Michigan Department of Health and Human Services, Bureau of Laboratories |
| EPI_ISL_437128 | Michigan Department of Health and Human Services, Bureau of Laboratories | Michigan Department of Health and Human Services, Bureau of Laboratories |

| EPI_Accession_ID | Originating Laboratory | Submitting Laboratory |
|---|---|---|
| EPI_ISL_437129 | Michigan Department of Health and Human Services, Bureau of Laboratories | Michigan Department of Health and Human Services, Bureau of Laboratories |
| EPI_ISL_437130 | Michigan Department of Health and Human Services, Bureau of Laboratories | Michigan Department of Health and Human Services, Bureau of Laboratories |
| EPI_ISL_437131 | Michigan Department of Health and Human Services, Bureau of Laboratories | Michigan Department of Health and Human Services, Bureau of Laboratories |
| EPI_ISL_437132 | Michigan Department of Health and Human Services, Bureau of Laboratories | Michigan Department of Health and Human Services, Bureau of Laboratories |
| EPI_ISL_437133 | Michigan Department of Health and Human Services, Bureau of Laboratories | Michigan Department of Health and Human Services, Bureau of Laboratories |
| EPI_ISL_437134 | Michigan Department of Health and Human Services, Bureau of Laboratories | Michigan Department of Health and Human Services, Bureau of Laboratories |
| EPI_ISL_437135 | Michigan Department of Health and Human Services, Bureau of Laboratories | Michigan Department of Health and Human Services, Bureau of Laboratories |
| EPI_ISL_437136 | Michigan Department of Health and Human Services, Bureau of Laboratories | Michigan Department of Health and Human Services, Bureau of Laboratories |
| EPI_ISL_437137 | Michigan Department of Health and Human Services, Bureau of Laboratories | Michigan Department of Health and Human Services, Bureau of Laboratories |
| EPI_ISL_437138 | Michigan Department of Health and Human Services, Bureau of Laboratories | Michigan Department of Health and Human Services, Bureau of Laboratories |
| EPI_ISL_437139 | Michigan Department of Health and Human Services, Bureau of Laboratories | Michigan Department of Health and Human Services, Bureau of Laboratories |
| EPI_ISL_437140 | Michigan Department of Health and Human Services, Bureau of Laboratories | Michigan Department of Health and Human Services, Bureau of Laboratories |
| EPI_ISL_437141 | Michigan Department of Health and Human Services, Bureau of Laboratories | Michigan Department of Health and Human Services, Bureau of Laboratories |
| EPI_ISL_437142 | Michigan Department of Health and Human Services, Bureau of Laboratories | Michigan Department of Health and Human Services, Bureau of Laboratories |
| EPI_ISL_437143 | Michigan Department of Health and Human Services, Bureau of Laboratories | Michigan Department of Health and Human Services, Bureau of Laboratories |
| EPI_ISL_437144 | Michigan Department of Health and Human Services, Bureau of Laboratories | Michigan Department of Health and Human Services, Bureau of Laboratories |

| EPI_Accession_ID | Originating Laboratory | Submitting Laboratory |
|---|---|---|
| EPI_ISL_437145 | Michigan Department of Health and Human Services, Bureau of Laboratories | Michigan Department of Health and Human Services, Bureau of Laboratories |
| EPI_ISL_437146 | Michigan Department of Health and Human Services, Bureau of Laboratories | Michigan Department of Health and Human Services, Bureau of Laboratories |
| EPI_ISL_437147 | Michigan Department of Health and Human Services, Bureau of Laboratories | Michigan Department of Health and Human Services, Bureau of Laboratories |
| EPI_ISL_437148 | Michigan Department of Health and Human Services, Bureau of Laboratories | Michigan Department of Health and Human Services, Bureau of Laboratories |
| EPI_ISL_437149 | Michigan Department of Health and Human Services, Bureau of Laboratories | Michigan Department of Health and Human Services, Bureau of Laboratories |
| EPI_ISL_437150 | Michigan Department of Health and Human Services, Bureau of Laboratories | Michigan Department of Health and Human Services, Bureau of Laboratories |
| EPI_ISL_437151 | Michigan Department of Health and Human Services, Bureau of Laboratories | Michigan Department of Health and Human Services, Bureau of Laboratories |
| EPI_ISL_437152 | Michigan Department of Health and Human Services, Bureau of Laboratories | Michigan Department of Health and Human Services, Bureau of Laboratories |
| EPI_ISL_437153 | Michigan Department of Health and Human Services, Bureau of Laboratories | Michigan Department of Health and Human Services, Bureau of Laboratories |
| EPI_ISL_437154 | Michigan Department of Health and Human Services, Bureau of Laboratories | Michigan Department of Health and Human Services, Bureau of Laboratories |
| EPI_ISL_437155 | Michigan Department of Health and Human Services, Bureau of Laboratories | Michigan Department of Health and Human Services, Bureau of Laboratories |
| EPI_ISL_437156 | Michigan Department of Health and Human Services, Bureau of Laboratories | Michigan Department of Health and Human Services, Bureau of Laboratories |
| EPI_ISL_437157 | Michigan Department of Health and Human Services, Bureau of Laboratories | Michigan Department of Health and Human Services, Bureau of Laboratories |
| EPI_ISL_437158 | Michigan Department of Health and Human Services, Bureau of Laboratories | Michigan Department of Health and Human Services, Bureau of Laboratories |
| EPI_ISL_437159 | Michigan Department of Health and Human Services, Bureau of Laboratories | Michigan Department of Health and Human Services, Bureau of Laboratories |
| EPI_ISL_437160 | Michigan Department of Health and Human Services, Bureau of Laboratories | Michigan Department of Health and Human Services, Bureau of Laboratories |

| EPI_Accession_ID | Originating Laboratory | Submitting Laboratory |
|---|---|---|
| EPI_ISL_437161 | Michigan Department of Health and Human Services, Bureau of Laboratories | Michigan Department of Health and Human Services, Bureau of Laboratories |
| EPI_ISL_437162 | Michigan Department of Health and Human Services, Bureau of Laboratories | Michigan Department of Health and Human Services, Bureau of Laboratories |
| EPI_ISL_437163 | Michigan Department of Health and Human Services, Bureau of Laboratories | Michigan Department of Health and Human Services, Bureau of Laboratories |
| EPI_ISL_437164 | Michigan Department of Health and Human Services, Bureau of Laboratories | Michigan Department of Health and Human Services, Bureau of Laboratories |
| EPI_ISL_437165 | Michigan Department of Health and Human Services, Bureau of Laboratories | Michigan Department of Health and Human Services, Bureau of Laboratories |
| EPI_ISL_437166 | Michigan Department of Health and Human Services, Bureau of Laboratories | Michigan Department of Health and Human Services, Bureau of Laboratories |
| EPI_ISL_437167 | Michigan Department of Health and Human Services, Bureau of Laboratories | Michigan Department of Health and Human Services, Bureau of Laboratories |
| EPI_ISL_437168 | Michigan Department of Health and Human Services, Bureau of Laboratories | Michigan Department of Health and Human Services, Bureau of Laboratories |
| EPI_ISL_437169 | Michigan Department of Health and Human Services, Bureau of Laboratories | Michigan Department of Health and Human Services, Bureau of Laboratories |
| EPI_ISL_437170 | Michigan Department of Health and Human Services, Bureau of Laboratories | Michigan Department of Health and Human Services, Bureau of Laboratories |
| EPI_ISL_437171 | Michigan Department of Health and Human Services, Bureau of Laboratories | Michigan Department of Health and Human Services, Bureau of Laboratories |
| EPI_ISL_437172 | Michigan Department of Health and Human Services, Bureau of Laboratories | Michigan Department of Health and Human Services, Bureau of Laboratories |
| EPI_ISL_437173 | Michigan Department of Health and Human Services, Bureau of Laboratories | Michigan Department of Health and Human Services, Bureau of Laboratories |
| EPI_ISL_437174 | Michigan Department of Health and Human Services, Bureau of Laboratories | Michigan Department of Health and Human Services, Bureau of Laboratories |
| EPI_ISL_437175 | Michigan Department of Health and Human Services, Bureau of Laboratories | Michigan Department of Health and Human Services, Bureau of Laboratories |
| EPI_ISL_437176 | Michigan Department of Health and Human Services, Bureau of Laboratories | Michigan Department of Health and Human Services, Bureau of Laboratories |

| EPI_Accession_ID | Originating Laboratory | Submitting Laboratory |
| --- | --- | --- |
| EPI_ISL_437177 | Michigan Department of Health and Human Services, Bureau of Laboratories | Michigan Department of Health and Human Services, Bureau of Laboratories |
| EPI_ISL_437178 | Michigan Department of Health and Human Services, Bureau of Laboratories | Michigan Department of Health and Human Services, Bureau of Laboratories |
| EPI_ISL_437179 | Michigan Department of Health and Human Services, Bureau of Laboratories | Michigan Department of Health and Human Services, Bureau of Laboratories |
| EPI_ISL_437180 | Michigan Department of Health and Human Services, Bureau of Laboratories | Michigan Department of Health and Human Services, Bureau of Laboratories |
| EPI_ISL_437181 | Michigan Department of Health and Human Services, Bureau of Laboratories | Michigan Department of Health and Human Services, Bureau of Laboratories |
| EPI_ISL_437182 | Michigan Department of Health and Human Services, Bureau of Laboratories | Michigan Department of Health and Human Services, Bureau of Laboratories |
| EPI_ISL_437183 | Michigan Department of Health and Human Services, Bureau of Laboratories | Michigan Department of Health and Human Services, Bureau of Laboratories |
| EPI_ISL_437184 | Michigan Department of Health and Human Services, Bureau of Laboratories | Michigan Department of Health and Human Services, Bureau of Laboratories |
| EPI_ISL_437185 | Michigan Department of Health and Human Services, Bureau of Laboratories | Michigan Department of Health and Human Services, Bureau of Laboratories |
| EPI_ISL_437186 | Michigan Department of Health and Human Services, Bureau of Laboratories | Michigan Department of Health and Human Services, Bureau of Laboratories |
| EPI_ISL_437187 | Siloam Hospitals | Institute of Tropical Disease, Universitas Airlangga |
| EPI_ISL_437188 | RSUD Dr. Soetomo | Institute of Tropical Disease, Universitas Airlangga |
| EPI_ISL_437189 | Pusat Pertamina Hospital | Eijkman Institute for Molecular Biology, Ministry of Research and Technology/National Agency for Research and Innovation |
| EPI_ISL_437190 | RS Pondok Indah Hospital â€“ Pondok Indah | Eijkman Institute for Molecular Biology, Ministry of Research and Technology/National Agency for Research and Innovation |
| EPI_ISL_437191 | RS Pondok Indah Hospital â€“ Pondok Indah | Eijkman Institute for Molecular Biology, Ministry of Research and Technology/National Agency for Research and Innovation |

| EPI_Accession_ID | Originating Laboratory | Submitting Laboratory |
|---|---|---|
| EPI_ISL_437192 | Mitra Keluarga Kelapa Gading Hospital | Eijkman Institute for Molecular Biology, Ministry of Research and Technology/National Agency for Research and Innovation |
| EPI_ISL_437193 | Viral Respiratory Lab, National Institute for Biomedical Research (INRB) | Pathogen Sequencing Lab, National Institute for Biomedical Research (INRB) |
| EPI_ISL_437194 | Viral Respiratory Lab, National Institute for Biomedical Research (INRB) | Pathogen Sequencing Lab, National Institute for Biomedical Research (INRB) |
| EPI_ISL_437195 | Viral Respiratory Lab, National Institute for Biomedical Research (INRB) | Pathogen Sequencing Lab, National Institute for Biomedical Research (INRB) |
| EPI_ISL_437196 | Viral Respiratory Lab, National Institute for Biomedical Research (INRB) | Pathogen Sequencing Lab, National Institute for Biomedical Research (INRB) |
| EPI_ISL_437197 | Diagnostic- and Research Institute of Pathology, Medical University of Graz | Diagnostic- and Research Institute of Pathology, Medical University of Graz |
| EPI_ISL_437198 | Diagnostic- and Research Institute of Pathology, Medical University of Graz | Diagnostic- and Research Institute of Pathology, Medical University of Graz |
| EPI_ISL_437199 | Diagnostic- and Research Institute of Pathology, Medical University of Graz | Diagnostic- and Research Institute of Pathology, Medical University of Graz |
| EPI_ISL_437200 | Diagnostic- and Research Institute of Pathology, Medical University of Graz | Diagnostic- and Research Institute of Pathology, Medical University of Graz |
| EPI_ISL_437201 | Diagnostic- and Research Institute of Pathology, Medical University of Graz | Diagnostic- and Research Institute of Pathology, Medical University of Graz |
| EPI_ISL_437202 | Diagnostic- and Research Institute of Pathology, Medical University of Graz | Diagnostic- and Research Institute of Pathology, Medical University of Graz |
| EPI_ISL_437203 | Diagnostic- and Research Institute of Pathology, Medical University of Graz | Diagnostic- and Research Institute of Pathology, Medical University of Graz |
| EPI_ISL_437204 | Max von Pettenkofer Institute, Virology, National Reference Center for Retroviruses, LMU MÃ¼nchen | Laboratory for Functional Genome Analysis, Dept. Genomics, Gene Center of the LMU Munich |
| EPI_ISL_437205 | Max von Pettenkofer Institute, Virology, National Reference Center for Retroviruses, LMU MÃ¼nchen | Laboratory for Functional Genome Analysis, Dept. Genomics, Gene Center of the LMU Munich |
| EPI_ISL_437206 | Max von Pettenkofer Institute, Virology, National Reference Center for Retroviruses, LMU MÃ¼nchen | Laboratory for Functional Genome Analysis, Dept. Genomics, Gene Center of the LMU Munich |
| EPI_ISL_437207 | Max von Pettenkofer Institute, Virology, National Reference Center for Retroviruses, LMU MÃ¼nchen | Laboratory for Functional Genome Analysis, Dept. Genomics, Gene Center of the LMU Munich |

| EPI_Accession_ID | Originating Laboratory | Submitting Laboratory |
|---|---|---|
| EPI_ISL_437208 | Max von Pettenkofer Institute, Virology, National Reference Center for Retroviruses, LMU MÃ¼nchen | Laboratory for Functional Genome Analysis, Dept. Genomics, Gene Center of the LMU Munich |
| EPI_ISL_437209 | Max von Pettenkofer Institute, Virology, National Reference Center for Retroviruses, LMU MÃ¼nchen | Laboratory for Functional Genome Analysis, Dept. Genomics, Gene Center of the LMU Munich |
| EPI_ISL_437210 | Max von Pettenkofer Institute, Virology, National Reference Center for Retroviruses, LMU MÃ¼nchen | Laboratory for Functional Genome Analysis, Dept. Genomics, Gene Center of the LMU Munich |
| EPI_ISL_437211 | Max von Pettenkofer Institute, Virology, National Reference Center for Retroviruses, LMU MÃ¼nchen | Laboratory for Functional Genome Analysis, Dept. Genomics, Gene Center of the LMU Munich |
| EPI_ISL_437212 | Max von Pettenkofer Institute, Virology, National Reference Center for Retroviruses, LMU MÃ¼nchen | Laboratory for Functional Genome Analysis, Dept. Genomics, Gene Center of the LMU Munich |
| EPI_ISL_437213 | Max von Pettenkofer Institute, Virology, National Reference Center for Retroviruses, LMU MÃ¼nchen | Laboratory for Functional Genome Analysis, Dept. Genomics, Gene Center of the LMU Munich |
| EPI_ISL_437214 | Max von Pettenkofer Institute, Virology, National Reference Center for Retroviruses, LMU MÃ¼nchen | Laboratory for Functional Genome Analysis, Dept. Genomics, Gene Center of the LMU Munich |
| EPI_ISL_437215 | Max von Pettenkofer Institute, Virology, National Reference Center for Retroviruses, LMU MÃ¼nchen | Laboratory for Functional Genome Analysis, Dept. Genomics, Gene Center of the LMU Munich |
| EPI_ISL_437216 | Max von Pettenkofer Institute, Virology, National Reference Center for Retroviruses, LMU MÃ¼nchen | Laboratory for Functional Genome Analysis, Dept. Genomics, Gene Center of the LMU Munich |
| EPI_ISL_437217 | Max von Pettenkofer Institute, Virology, National Reference Center for Retroviruses, LMU MÃ¼nchen | Laboratory for Functional Genome Analysis, Dept. Genomics, Gene Center of the LMU Munich |
| EPI_ISL_437218 | Max von Pettenkofer Institute, Virology, National Reference Center for Retroviruses, LMU MÃ¼nchen | Laboratory for Functional Genome Analysis, Dept. Genomics, Gene Center of the LMU Munich |
| EPI_ISL_437219 | Max von Pettenkofer Institute, Virology, National Reference Center for Retroviruses, LMU MÃ¼nchen | Laboratory for Functional Genome Analysis, Dept. Genomics, Gene Center of the LMU Munich |
| EPI_ISL_437220 | Max von Pettenkofer Institute, Virology, National Reference Center for Retroviruses, LMU MÃ¼nchen | Laboratory for Functional Genome Analysis, Dept. Genomics, Gene Center of the LMU Munich |
| EPI_ISL_437221 | Max von Pettenkofer Institute, Virology, National Reference Center for Retroviruses, LMU MÃ¼nchen | Laboratory for Functional Genome Analysis, Dept. Genomics, Gene Center of the LMU Munich |
| EPI_ISL_437222 | Max von Pettenkofer Institute, Virology, National Reference Center for Retroviruses, LMU MÃ¼nchen | Laboratory for Functional Genome Analysis, Dept. Genomics, Gene Center of the LMU Munich |
| EPI_ISL_437223 | Max von Pettenkofer Institute, Virology, National Reference Center for Retroviruses, LMU MÃ¼nchen | Laboratory for Functional Genome Analysis, Dept. Genomics, Gene Center of the LMU Munich |

| EPI_Accession_ID | Originating Laboratory | Submitting Laboratory |
|---|---|---|
| EPI_ISL_437224 | Max von Pettenkofer Institute, Virology, National Reference Center for Retroviruses, LMU MÃ¼nchen | Laboratory for Functional Genome Analysis, Dept. Genomics, Gene Center of the LMU Munich |
| EPI_ISL_437225 | Max von Pettenkofer Institute, Virology, National Reference Center for Retroviruses, LMU MÃ¼nchen | Laboratory for Functional Genome Analysis, Dept. Genomics, Gene Center of the LMU Munich |
| EPI_ISL_437226 | Max von Pettenkofer Institute, Virology, National Reference Center for Retroviruses, LMU MÃ¼nchen | Laboratory for Functional Genome Analysis, Dept. Genomics, Gene Center of the LMU Munich |
| EPI_ISL_437227 | Max von Pettenkofer Institute, Virology, National Reference Center for Retroviruses, LMU MÃ¼nchen | Laboratory for Functional Genome Analysis, Dept. Genomics, Gene Center of the LMU Munich |
| EPI_ISL_437228 | Max von Pettenkofer Institute, Virology, National Reference Center for Retroviruses, LMU MÃ¼nchen | Laboratory for Functional Genome Analysis, Dept. Genomics, Gene Center of the LMU Munich |
| EPI_ISL_437229 | Max von Pettenkofer Institute, Virology, National Reference Center for Retroviruses, LMU MÃ¼nchen | Laboratory for Functional Genome Analysis, Dept. Genomics, Gene Center of the LMU Munich |
| EPI_ISL_437230 | Max von Pettenkofer Institute, Virology, National Reference Center for Retroviruses, LMU MÃ¼nchen | Laboratory for Functional Genome Analysis, Dept. Genomics, Gene Center of the LMU Munich |
| EPI_ISL_437231 | Max von Pettenkofer Institute, Virology, National Reference Center for Retroviruses, LMU MÃ¼nchen | Laboratory for Functional Genome Analysis, Dept. Genomics, Gene Center of the LMU Munich |
| EPI_ISL_437232 | Max von Pettenkofer Institute, Virology, National Reference Center for Retroviruses, LMU MÃ¼nchen | Laboratory for Functional Genome Analysis, Dept. Genomics, Gene Center of the LMU Munich |
| EPI_ISL_437233 | Max von Pettenkofer Institute, Virology, National Reference Center for Retroviruses, LMU MÃ¼nchen | Laboratory for Functional Genome Analysis, Dept. Genomics, Gene Center of the LMU Munich |
| EPI_ISL_437234 | Max von Pettenkofer Institute, Virology, National Reference Center for Retroviruses, LMU MÃ¼nchen | Laboratory for Functional Genome Analysis, Dept. Genomics, Gene Center of the LMU Munich |
| EPI_ISL_437235 | Max von Pettenkofer Institute, Virology, National Reference Center for Retroviruses, LMU MÃ¼nchen | Laboratory for Functional Genome Analysis, Dept. Genomics, Gene Center of the LMU Munich |
| EPI_ISL_437236 | Max von Pettenkofer Institute, Virology, National Reference Center for Retroviruses, LMU MÃ¼nchen | Laboratory for Functional Genome Analysis, Dept. Genomics, Gene Center of the LMU Munich |
| EPI_ISL_437237 | Max von Pettenkofer Institute, Virology, National Reference Center for Retroviruses, LMU MÃ¼nchen | Laboratory for Functional Genome Analysis, Dept. Genomics, Gene Center of the LMU Munich |
| EPI_ISL_437238 | Max von Pettenkofer Institute, Virology, National Reference Center for Retroviruses, LMU MÃ¼nchen | Laboratory for Functional Genome Analysis, Dept. Genomics, Gene Center of the LMU Munich |
| EPI_ISL_437239 | Max von Pettenkofer Institute, Virology, National Reference Center for Retroviruses, LMU MÃ¼nchen | Laboratory for Functional Genome Analysis, Dept. Genomics, Gene Center of the LMU Munich |

| EPI_Accession_ID | Originating Laboratory | Submitting Laboratory |
|---|---|---|
| EPI_ISL_437240 | Max von Pettenkofer Institute, Virology, National Reference Center for Retroviruses, LMU MÃ¼nchen | Laboratory for Functional Genome Analysis, Dept. Genomics, Gene Center of the LMU Munich |
| EPI_ISL_437241 | Max von Pettenkofer Institute, Virology, National Reference Center for Retroviruses, LMU MÃ¼nchen | Laboratory for Functional Genome Analysis, Dept. Genomics, Gene Center of the LMU Munich |
| EPI_ISL_437242 | Max von Pettenkofer Institute, Virology, National Reference Center for Retroviruses, LMU MÃ¼nchen | Laboratory for Functional Genome Analysis, Dept. Genomics, Gene Center of the LMU Munich |
| EPI_ISL_437243 | Max von Pettenkofer Institute, Virology, National Reference Center for Retroviruses, LMU MÃ¼nchen | Laboratory for Functional Genome Analysis, Dept. Genomics, Gene Center of the LMU Munich |
| EPI_ISL_437244 | Max von Pettenkofer Institute, Virology, National Reference Center for Retroviruses, LMU MÃ¼nchen | Laboratory for Functional Genome Analysis, Dept. Genomics, Gene Center of the LMU Munich |
| EPI_ISL_437245 | Max von Pettenkofer Institute, Virology, National Reference Center for Retroviruses, LMU MÃ¼nchen | Laboratory for Functional Genome Analysis, Dept. Genomics, Gene Center of the LMU Munich |
| EPI_ISL_437246 | Max von Pettenkofer Institute, Virology, National Reference Center for Retroviruses, LMU MÃ¼nchen | Laboratory for Functional Genome Analysis, Dept. Genomics, Gene Center of the LMU Munich |
| EPI_ISL_437247 | Max von Pettenkofer Institute, Virology, National Reference Center for Retroviruses, LMU MÃ¼nchen | Laboratory for Functional Genome Analysis, Dept. Genomics, Gene Center of the LMU Munich |
| EPI_ISL_437248 | Max von Pettenkofer Institute, Virology, National Reference Center for Retroviruses, LMU MÃ¼nchen | Laboratory for Functional Genome Analysis, Dept. Genomics, Gene Center of the LMU Munich |
| EPI_ISL_437249 | Max von Pettenkofer Institute, Virology, National Reference Center for Retroviruses, LMU MÃ¼nchen | Laboratory for Functional Genome Analysis, Dept. Genomics, Gene Center of the LMU Munich |
| EPI_ISL_437250 | Max von Pettenkofer Institute, Virology, National Reference Center for Retroviruses, LMU MÃ¼nchen | Laboratory for Functional Genome Analysis, Dept. Genomics, Gene Center of the LMU Munich |
| EPI_ISL_437251 | Max von Pettenkofer Institute, Virology, National Reference Center for Retroviruses, LMU MÃ¼nchen | Laboratory for Functional Genome Analysis, Dept. Genomics, Gene Center of the LMU Munich |
| EPI_ISL_437252 | Max von Pettenkofer Institute, Virology, National Reference Center for Retroviruses, LMU MÃ¼nchen | Laboratory for Functional Genome Analysis, Dept. Genomics, Gene Center of the LMU Munich |
| EPI_ISL_437253 | Max von Pettenkofer Institute, Virology, National Reference Center for Retroviruses, LMU MÃ¼nchen | Laboratory for Functional Genome Analysis, Dept. Genomics, Gene Center of the LMU Munich |
| EPI_ISL_437254 | Max von Pettenkofer Institute, Virology, National Reference Center for Retroviruses, LMU MÃ¼nchen | Laboratory for Functional Genome Analysis, Dept. Genomics, Gene Center of the LMU Munich |
| EPI_ISL_437255 | Max von Pettenkofer Institute, Virology, National Reference Center for Retroviruses, LMU MÃ¼nchen | Laboratory for Functional Genome Analysis, Dept. Genomics, Gene Center of the LMU Munich |

| EPI_Accession_ID | Originating Laboratory | Submitting Laboratory |
|---|---|---|
| EPI_ISL_437256 | Max von Pettenkofer Institute, Virology, National Reference Center for Retroviruses, LMU MÃ¼nchen | Laboratory for Functional Genome Analysis, Dept. Genomics, Gene Center of the LMU Munich |
| EPI_ISL_437257 | Max von Pettenkofer Institute, Virology, National Reference Center for Retroviruses, LMU MÃ¼nchen | Laboratory for Functional Genome Analysis, Dept. Genomics, Gene Center of the LMU Munich |
| EPI_ISL_437258 | Max von Pettenkofer Institute, Virology, National Reference Center for Retroviruses, LMU MÃ¼nchen | Laboratory for Functional Genome Analysis, Dept. Genomics, Gene Center of the LMU Munich |
| EPI_ISL_437259 | Max von Pettenkofer Institute, Virology, National Reference Center for Retroviruses, LMU MÃ¼nchen | Laboratory for Functional Genome Analysis, Dept. Genomics, Gene Center of the LMU Munich |
| EPI_ISL_437260 | Max von Pettenkofer Institute, Virology, National Reference Center for Retroviruses, LMU MÃ¼nchen | Laboratory for Functional Genome Analysis, Dept. Genomics, Gene Center of the LMU Munich |
| EPI_ISL_437261 | Max von Pettenkofer Institute, Virology, National Reference Center for Retroviruses, LMU MÃ¼nchen | Laboratory for Functional Genome Analysis, Dept. Genomics, Gene Center of the LMU Munich |
| EPI_ISL_437262 | Max von Pettenkofer Institute, Virology, National Reference Center for Retroviruses, LMU MÃ¼nchen | Laboratory for Functional Genome Analysis, Dept. Genomics, Gene Center of the LMU Munich |
| EPI_ISL_437263 | Max von Pettenkofer Institute, Virology, National Reference Center for Retroviruses, LMU MÃ¼nchen | Laboratory for Functional Genome Analysis, Dept. Genomics, Gene Center of the LMU Munich |
| EPI_ISL_437264 | Max von Pettenkofer Institute, Virology, National Reference Center for Retroviruses, LMU MÃ¼nchen | Laboratory for Functional Genome Analysis, Dept. Genomics, Gene Center of the LMU Munich |
| EPI_ISL_437265 | Max von Pettenkofer Institute, Virology, National Reference Center for Retroviruses, LMU MÃ¼nchen | Laboratory for Functional Genome Analysis, Dept. Genomics, Gene Center of the LMU Munich |
| EPI_ISL_437266 | Max von Pettenkofer Institute, Virology, National Reference Center for Retroviruses, LMU MÃ¼nchen | Laboratory for Functional Genome Analysis, Dept. Genomics, Gene Center of the LMU Munich |
| EPI_ISL_437267 | Max von Pettenkofer Institute, Virology, National Reference Center for Retroviruses, LMU MÃ¼nchen | Laboratory for Functional Genome Analysis, Dept. Genomics, Gene Center of the LMU Munich |
| EPI_ISL_437268 | Max von Pettenkofer Institute, Virology, National Reference Center for Retroviruses, LMU MÃ¼nchen | Laboratory for Functional Genome Analysis, Dept. Genomics, Gene Center of the LMU Munich |
| EPI_ISL_437269 | Max von Pettenkofer Institute, Virology, National Reference Center for Retroviruses, LMU MÃ¼nchen | Laboratory for Functional Genome Analysis, Dept. Genomics, Gene Center of the LMU Munich |
| EPI_ISL_437270 | Max von Pettenkofer Institute, Virology, National Reference Center for Retroviruses, LMU MÃ¼nchen | Laboratory for Functional Genome Analysis, Dept. Genomics, Gene Center of the LMU Munich |
| EPI_ISL_437271 | Max von Pettenkofer Institute, Virology, National Reference Center for Retroviruses, LMU MÃ¼nchen | Laboratory for Functional Genome Analysis, Dept. Genomics, Gene Center of the LMU Munich |

| EPI_Accession_ID | Originating Laboratory | Submitting Laboratory |
|---|---|---|
| EPI_ISL_437272 | Max von Pettenkofer Institute, Virology, National Reference Center for Retroviruses, LMU MÃ¼nchen | Laboratory for Functional Genome Analysis, Dept. Genomics, Gene Center of the LMU Munich |
| EPI_ISL_437273 | Max von Pettenkofer Institute, Virology, National Reference Center for Retroviruses, LMU MÃ¼nchen | Laboratory for Functional Genome Analysis, Dept. Genomics, Gene Center of the LMU Munich |
| EPI_ISL_437274 | Max von Pettenkofer Institute, Virology, National Reference Center for Retroviruses, LMU MÃ¼nchen | Laboratory for Functional Genome Analysis, Dept. Genomics, Gene Center of the LMU Munich |
| EPI_ISL_437275 | Max von Pettenkofer Institute, Virology, National Reference Center for Retroviruses, LMU MÃ¼nchen | Laboratory for Functional Genome Analysis, Dept. Genomics, Gene Center of the LMU Munich |
| EPI_ISL_437276 | Max von Pettenkofer Institute, Virology, National Reference Center for Retroviruses, LMU MÃ¼nchen | Laboratory for Functional Genome Analysis, Dept. Genomics, Gene Center of the LMU Munich |
| EPI_ISL_437277 | Max von Pettenkofer Institute, Virology, National Reference Center for Retroviruses, LMU MÃ¼nchen | Laboratory for Functional Genome Analysis, Dept. Genomics, Gene Center of the LMU Munich |
| EPI_ISL_437278 | Max von Pettenkofer Institute, Virology, National Reference Center for Retroviruses, LMU MÃ¼nchen | Laboratory for Functional Genome Analysis, Dept. Genomics, Gene Center of the LMU Munich |
| EPI_ISL_437279 | Max von Pettenkofer Institute, Virology, National Reference Center for Retroviruses, LMU MÃ¼nchen | Laboratory for Functional Genome Analysis, Dept. Genomics, Gene Center of the LMU Munich |
| EPI_ISL_437280 | Max von Pettenkofer Institute, Virology, National Reference Center for Retroviruses, LMU MÃ¼nchen | Laboratory for Functional Genome Analysis, Dept. Genomics, Gene Center of the LMU Munich |
| EPI_ISL_437281 | Max von Pettenkofer Institute, Virology, National Reference Center for Retroviruses, LMU MÃ¼nchen | Laboratory for Functional Genome Analysis, Dept. Genomics, Gene Center of the LMU Munich |
| EPI_ISL_437282 | Max von Pettenkofer Institute, Virology, National Reference Center for Retroviruses, LMU MÃ¼nchen | Laboratory for Functional Genome Analysis, Dept. Genomics, Gene Center of the LMU Munich |
| EPI_ISL_437283 | Max von Pettenkofer Institute, Virology, National Reference Center for Retroviruses, LMU MÃ¼nchen | Laboratory for Functional Genome Analysis, Dept. Genomics, Gene Center of the LMU Munich |
| EPI_ISL_437284 | Max von Pettenkofer Institute, Virology, National Reference Center for Retroviruses, LMU MÃ¼nchen | Laboratory for Functional Genome Analysis, Dept. Genomics, Gene Center of the LMU Munich |
| EPI_ISL_437285 | Max von Pettenkofer Institute, Virology, National Reference Center for Retroviruses, LMU MÃ¼nchen | Laboratory for Functional Genome Analysis, Dept. Genomics, Gene Center of the LMU Munich |
| EPI_ISL_437286 | Max von Pettenkofer Institute, Virology, National Reference Center for Retroviruses, LMU MÃ¼nchen | Laboratory for Functional Genome Analysis, Dept. Genomics, Gene Center of the LMU Munich |
| EPI_ISL_437287 | Max von Pettenkofer Institute, Virology, National Reference Center for Retroviruses, LMU MÃ¼nchen | Laboratory for Functional Genome Analysis, Dept. Genomics, Gene Center of the LMU Munich |

| EPI_Accession_ID | Originating Laboratory | Submitting Laboratory |
|---|---|---|
| EPI_ISL_437288 | Max von Pettenkofer Institute, Virology, National Reference Center for Retroviruses, LMU MÃ¼nchen | Laboratory for Functional Genome Analysis, Dept. Genomics, Gene Center of the LMU Munich |
| EPI_ISL_437289 | Max von Pettenkofer Institute, Virology, National Reference Center for Retroviruses, LMU MÃ¼nchen | Laboratory for Functional Genome Analysis, Dept. Genomics, Gene Center of the LMU Munich |
| EPI_ISL_437290 | Max von Pettenkofer Institute, Virology, National Reference Center for Retroviruses, LMU MÃ¼nchen | Laboratory for Functional Genome Analysis, Dept. Genomics, Gene Center of the LMU Munich |
| EPI_ISL_437291 | Max von Pettenkofer Institute, Virology, National Reference Center for Retroviruses, LMU MÃ¼nchen | Laboratory for Functional Genome Analysis, Dept. Genomics, Gene Center of the LMU Munich |
| EPI_ISL_437292 | Max von Pettenkofer Institute, Virology, National Reference Center for Retroviruses, LMU MÃ¼nchen | Laboratory for Functional Genome Analysis, Dept. Genomics, Gene Center of the LMU Munich |
| EPI_ISL_437293 | Max von Pettenkofer Institute, Virology, National Reference Center for Retroviruses, LMU MÃ¼nchen | Laboratory for Functional Genome Analysis, Dept. Genomics, Gene Center of the LMU Munich |
| EPI_ISL_437294 | Max von Pettenkofer Institute, Virology, National Reference Center for Retroviruses, LMU MÃ¼nchen | Laboratory for Functional Genome Analysis, Dept. Genomics, Gene Center of the LMU Munich |
| EPI_ISL_437295 | Max von Pettenkofer Institute, Virology, National Reference Center for Retroviruses, LMU MÃ¼nchen | Laboratory for Functional Genome Analysis, Dept. Genomics, Gene Center of the LMU Munich |
| EPI_ISL_437296 | Max von Pettenkofer Institute, Virology, National Reference Center for Retroviruses, LMU MÃ¼nchen | Laboratory for Functional Genome Analysis, Dept. Genomics, Gene Center of the LMU Munich |
| EPI_ISL_437297 | Max von Pettenkofer Institute, Virology, National Reference Center for Retroviruses, LMU MÃ¼nchen | Laboratory for Functional Genome Analysis, Dept. Genomics, Gene Center of the LMU Munich |
| EPI_ISL_437298 | Diagnostic- and Research Institute of Pathology, Medical University of Graz | Diagnostic- and Research Institute of Pathology, Medical University of Graz |
| EPI_ISL_437299 | Diagnostic- and Research Institute of Pathology, Medical University of Graz | Diagnostic- and Research Institute of Pathology, Medical University of Graz |
| EPI_ISL_437300 | Diagnostic- and Research Institute of Pathology, Medical University of Graz | Diagnostic- and Research Institute of Pathology, Medical University of Graz |
| EPI_ISL_437301 | Diagnostic- and Research Institute of Pathology, Medical University of Graz | Diagnostic- and Research Institute of Pathology, Medical University of Graz |
| EPI_ISL_437302 | Diagnostic- and Research Institute of Pathology, Medical University of Graz | Diagnostic- and Research Institute of Pathology, Medical University of Graz |
| EPI_ISL_437303 | Diagnostic- and Research Institute of Pathology, Medical University of Graz | Diagnostic- and Research Institute of Pathology, Medical University of Graz |
| EPI_ISL_437304 | Ministry of Health Turkey | Ministry of Health Turkey |

| EPI_Accession_ID | Originating Laboratory | Submitting Laboratory |
|---|---|---|
| EPI_ISL_437305 | Ministry of Health Turkey | Ministry of Health Turkey |
| EPI_ISL_437306 | Ministry of Health Turkey | Ministry of Health Turkey |
| EPI_ISL_437307 | Ministry of Health Turkey | Ministry of Health Turkey |
| EPI_ISL_437308 | Ministry of Health Turkey | Ministry of Health Turkey |
| EPI_ISL_437309 | Ministry of Health Turkey | Ministry of Health Turkey |
| EPI_ISL_437310 | Ministry of Health Turkey | Ministry of Health Turkey |
| EPI_ISL_437311 | Ministry of Health Turkey | Ministry of Health Turkey |
| EPI_ISL_437312 | Ministry of Health Turkey | Ministry of Health Turkey |
| EPI_ISL_437313 | Ministry of Health Turkey | Ministry of Health Turkey |
| EPI_ISL_437314 | Ministry of Health Turkey | Ministry of Health Turkey |
| EPI_ISL_437315 | Ministry of Health Turkey | Ministry of Health Turkey |
| EPI_ISL_437316 | Ministry of Health Turkey | Ministry of Health Turkey |
| EPI_ISL_437317 | Ministry of Health Turkey | Ministry of Health Turkey |
| EPI_ISL_437318 | Ministry of Health Turkey | Ministry of Health Turkey |
| EPI_ISL_437319 | Ministry of Health Turkey | Ministry of Health Turkey |
| EPI_ISL_437320 | Ministry of Health Turkey | Ministry of Health Turkey |
| EPI_ISL_437321 | Ministry of Health Turkey | Ministry of Health Turkey |
| EPI_ISL_437322 | Ministry of Health Turkey | Ministry of Health Turkey |
| EPI_ISL_437323 | Ministry of Health Turkey | Ministry of Health Turkey |
| EPI_ISL_437324 | Ministry of Health Turkey | Ministry of Health Turkey |
| EPI_ISL_437325 | Ministry of Health Turkey | Ministry of Health Turkey |
| EPI_ISL_437326 | Ministry of Health Turkey | Ministry of Health Turkey |
| EPI_ISL_437327 | Ministry of Health Turkey | Ministry of Health Turkey |
| EPI_ISL_437328 | Ministry of Health Turkey | Ministry of Health Turkey |
| EPI_ISL_437329 | Ministry of Health Turkey | Ministry of Health Turkey |
| EPI_ISL_437330 | Ministry of Health Turkey | Ministry of Health Turkey |
| EPI_ISL_437331 | Ministry of Health Turkey | Ministry of Health Turkey |
| EPI_ISL_437332 | Ministry of Health Turkey | Ministry of Health Turkey |
| EPI_ISL_437333 | Ministry of Health Turkey | Ministry of Health Turkey |
| EPI_ISL_437334 | Ministry of Health Turkey | Ministry of Health Turkey |
| EPI_ISL_437335 | Ministry of Health Turkey | Ministry of Health Turkey |
| EPI_ISL_437336 | TSGH-CP molecular lab, Division of Clinical Pathology, Department of Pathology | TSGH-CP molecular lab, Division of Clinical Pathology, Department of Pathology |

| EPI_Accession_ID | Originating Laboratory | Submitting Laboratory |
|---|---|---|
| EPI_ISL_437337 | Viral Respiratory Lab, National Institute for Biomedical Research (INRB) | Pathogen Sequencing Lab, National Institute for Biomedical Research (INRB) |
| EPI_ISL_437338 | Viral Respiratory Lab, National Institute for Biomedical Research (INRB) | Pathogen Sequencing Lab, National Institute for Biomedical Research (INRB) |
| EPI_ISL_437339 | Viral Respiratory Lab, National Institute for Biomedical Research (INRB) | Pathogen Sequencing Lab, National Institute for Biomedical Research (INRB) |
| EPI_ISL_437340 | Viral Respiratory Lab, National Institute for Biomedical Research (INRB) | Pathogen Sequencing Lab, National Institute for Biomedical Research (INRB) |
| EPI_ISL_437341 | Viral Respiratory Lab, National Institute for Biomedical Research (INRB) | Pathogen Sequencing Lab, National Institute for Biomedical Research (INRB) |
| EPI_ISL_437342 | Viral Respiratory Lab, National Institute for Biomedical Research (INRB) | Pathogen Sequencing Lab, National Institute for Biomedical Research (INRB) |
| EPI_ISL_437343 | Viral Respiratory Lab, National Institute for Biomedical Research (INRB) | Pathogen Sequencing Lab, National Institute for Biomedical Research (INRB) |
| EPI_ISL_437344 | Viral Respiratory Lab, National Institute for Biomedical Research (INRB) | Pathogen Sequencing Lab, National Institute for Biomedical Research (INRB) |
| EPI_ISL_437345 | Viral Respiratory Lab, National Institute for Biomedical Research (INRB) | Pathogen Sequencing Lab, National Institute for Biomedical Research (INRB) |
| EPI_ISL_437346 | Viral Respiratory Lab, National Institute for Biomedical Research (INRB) | Pathogen Sequencing Lab, National Institute for Biomedical Research (INRB) |
| EPI_ISL_437347 | Viral Respiratory Lab, National Institute for Biomedical Research (INRB) | Pathogen Sequencing Lab, National Institute for Biomedical Research (INRB) |
| EPI_ISL_437348 | Viral Respiratory Lab, National Institute for Biomedical Research (INRB) | Pathogen Sequencing Lab, National Institute for Biomedical Research (INRB) |
| EPI_ISL_437349 | Ecole nationale vÃ©tÃ©rinaire d'Alfort-laboratoire de santÃ© animale Anses UMR 1161 de virologie ENVA-Anses-INRAE | Institut Pasteur CIBU-ERI |
| EPI_ISL_437350 | Viral Respiratory Lab, National Institute for Biomedical Research (INRB) | Pathogen Sequencing Lab, National Institute for Biomedical Research (INRB) |
| EPI_ISL_437351 | Viral Respiratory Lab, National Institute for Biomedical Research (INRB) | Pathogen Sequencing Lab, National Institute for Biomedical Research (INRB) |
| EPI_ISL_437352 | Viral Respiratory Lab, National Institute for Biomedical Research (INRB) | Pathogen Sequencing Lab, National Institute for Biomedical Research (INRB) |

| EPI_Accession_ID | Originating Laboratory | Submitting Laboratory |
|---|---|---|
| EPI_ISL_437353 | Viral Respiratory Lab, National Institute for Biomedical Research (INRB) | Pathogen Sequencing Lab, National Institute for Biomedical Research (INRB) |
| EPI_ISL_437354 | Viral Respiratory Lab, National Institute for Biomedical Research (INRB) | Pathogen Sequencing Lab, National Institute for Biomedical Research (INRB) |
| EPI_ISL_437355 | Viral Respiratory Lab, National Institute for Biomedical Research (INRB) | Pathogen Sequencing Lab, National Institute for Biomedical Research (INRB) |
| EPI_ISL_437356 | Viral Respiratory Lab, National Institute for Biomedical Research (INRB) | Pathogen Sequencing Lab, National Institute for Biomedical Research (INRB) |
| EPI_ISL_437357 | Viral Respiratory Lab, National Institute for Biomedical Research (INRB) | Pathogen Sequencing Lab, National Institute for Biomedical Research (INRB) |
| EPI_ISL_437358 | Viral Respiratory Lab, National Institute for Biomedical Research (INRB) | Pathogen Sequencing Lab, National Institute for Biomedical Research (INRB) |
| EPI_ISL_437359 | Max von Pettenkofer Institute, Virology, National Reference Center for Retroviruses, LMU MÃ¼nchen | Laboratory for Functional Genome Analysis, Dept. Genomics, Gene Center of the LMU Munich |
| EPI_ISL_437360 | Minnesota Department of Health, Public Health Laboratory | Minnesota Department of Health, Public Health Laboratory |
| EPI_ISL_437361 | Minnesota Department of Health, Public Health Laboratory | Minnesota Department of Health, Public Health Laboratory |
| EPI_ISL_437362 | Minnesota Department of Health, Public Health Laboratory | Minnesota Department of Health, Public Health Laboratory |
| EPI_ISL_437363 | Minnesota Department of Health, Public Health Laboratory | Minnesota Department of Health, Public Health Laboratory |
| EPI_ISL_437364 | Minnesota Department of Health, Public Health Laboratory | Minnesota Department of Health, Public Health Laboratory |
| EPI_ISL_437365 | Minnesota Department of Health, Public Health Laboratory | Minnesota Department of Health, Public Health Laboratory |
| EPI_ISL_437366 | Minnesota Department of Health, Public Health Laboratory | Minnesota Department of Health, Public Health Laboratory |
| EPI_ISL_437367 | Minnesota Department of Health, Public Health Laboratory | Minnesota Department of Health, Public Health Laboratory |
| EPI_ISL_437368 | Minnesota Department of Health, Public Health Laboratory | Minnesota Department of Health, Public Health Laboratory |

| EPI_Accession_ID | Originating Laboratory | Submitting Laboratory |
| --- | --- | --- |
| EPI_ISL_437369 | Minnesota Department of Health, Public Health Laboratory | Minnesota Department of Health, Public Health Laboratory |
| EPI_ISL_437370 | Minnesota Department of Health, Public Health Laboratory | Minnesota Department of Health, Public Health Laboratory |
| EPI_ISL_437371 | Minnesota Department of Health, Public Health Laboratory | Minnesota Department of Health, Public Health Laboratory |
| EPI_ISL_437372 | Minnesota Department of Health, Public Health Laboratory | Minnesota Department of Health, Public Health Laboratory |
| EPI_ISL_437373 | Minnesota Department of Health, Public Health Laboratory | Minnesota Department of Health, Public Health Laboratory |
| EPI_ISL_437374 | Minnesota Department of Health, Public Health Laboratory | Minnesota Department of Health, Public Health Laboratory |
| EPI_ISL_437375 | Minnesota Department of Health, Public Health Laboratory | Minnesota Department of Health, Public Health Laboratory |
| EPI_ISL_437376 | Minnesota Department of Health, Public Health Laboratory | Minnesota Department of Health, Public Health Laboratory |
| EPI_ISL_437377 | Minnesota Department of Health, Public Health Laboratory | Minnesota Department of Health, Public Health Laboratory |
| EPI_ISL_437378 | Minnesota Department of Health, Public Health Laboratory | Minnesota Department of Health, Public Health Laboratory |
| EPI_ISL_437379 | Minnesota Department of Health, Public Health Laboratory | Minnesota Department of Health, Public Health Laboratory |
| EPI_ISL_437380 | Minnesota Department of Health, Public Health Laboratory | Minnesota Department of Health, Public Health Laboratory |
| EPI_ISL_437381 | Minnesota Department of Health, Public Health Laboratory | Minnesota Department of Health, Public Health Laboratory |
| EPI_ISL_437382 | Minnesota Department of Health, Public Health Laboratory | Minnesota Department of Health, Public Health Laboratory |
| EPI_ISL_437383 | Minnesota Department of Health, Public Health Laboratory | Minnesota Department of Health, Public Health Laboratory |
| EPI_ISL_437384 | Minnesota Department of Health, Public Health Laboratory | Minnesota Department of Health, Public Health Laboratory |

| EPI_Accession_ID | Originating Laboratory | Submitting Laboratory |
|---|---|---|
| EPI_ISL_437385 | Minnesota Department of Health, Public Health Laboratory | Minnesota Department of Health, Public Health Laboratory |
| EPI_ISL_437386 | Minnesota Department of Health, Public Health Laboratory | Minnesota Department of Health, Public Health Laboratory |
| EPI_ISL_437387 | Virginia DCLS | Virginia DCLS |
| EPI_ISL_437388 | Virginia DCLS | Virginia DCLS |
| EPI_ISL_437389 | Virginia DCLS | Virginia DCLS |
| EPI_ISL_437390 | Virginia DCLS | Virginia DCLS |
| EPI_ISL_437391 | Virginia DCLS | Virginia DCLS |
| EPI_ISL_437392 | Virginia DCLS | Virginia DCLS |
| EPI_ISL_437393 | Virginia DCLS | Virginia DCLS |
| EPI_ISL_437394 | Virginia DCLS | Virginia DCLS |
| EPI_ISL_437395 | Virginia DCLS | Virginia DCLS |
| EPI_ISL_437396 | Virginia DCLS | Virginia DCLS |
| EPI_ISL_437397 | Virginia DCLS | Virginia DCLS |
| EPI_ISL_437398 | Virginia DCLS | Virginia DCLS |
| EPI_ISL_437399 | Virginia DCLS | Virginia DCLS |
| EPI_ISL_437400 | Virginia DCLS | Virginia DCLS |
| EPI_ISL_437401 | Virginia DCLS | Virginia DCLS |
| EPI_ISL_437402 | Virginia DCLS | Virginia DCLS |
| EPI_ISL_437403 | Virginia DCLS | Virginia DCLS |
| EPI_ISL_437404 | Virginia DCLS | Virginia DCLS |
| EPI_ISL_437405 | Virginia DCLS | Virginia DCLS |
| EPI_ISL_437406 | Virginia DCLS | Virginia DCLS |
| EPI_ISL_437407 | Virginia DCLS | Virginia DCLS |
| EPI_ISL_437408 | Virginia DCLS | Virginia DCLS |
| EPI_ISL_437409 | Virginia DCLS | Virginia DCLS |
| EPI_ISL_437410 | Virginia DCLS | Virginia DCLS |
| EPI_ISL_437411 | Virginia DCLS | Virginia DCLS |
| EPI_ISL_437412 | Virginia DCLS | Virginia DCLS |
| EPI_ISL_437413 | Virginia DCLS | Virginia DCLS |
| EPI_ISL_437414 | Virginia DCLS | Virginia DCLS |
| EPI_ISL_437415 | Virginia DCLS | Virginia DCLS |

| EPI_Accession_ID | Originating Laboratory | Submitting Laboratory |
|---|---|---|
| EPI_ISL_437416 | Virginia DCLS | Virginia DCLS |
| EPI_ISL_437417 | Virginia DCLS | Virginia DCLS |
| EPI_ISL_437418 | Virginia DCLS | Virginia DCLS |
| EPI_ISL_437419 | Virginia DCLS | Virginia DCLS |
| EPI_ISL_437420 | Virginia DCLS | Virginia DCLS |
| EPI_ISL_437421 | Virginia DCLS | Virginia DCLS |
| EPI_ISL_437422 | Virginia DCLS | Virginia DCLS |
| EPI_ISL_437423 | Virginia DCLS | Virginia DCLS |
| EPI_ISL_437424 | Virginia DCLS | Virginia DCLS |
| EPI_ISL_437425 | Virginia DCLS | Virginia DCLS |
| EPI_ISL_437426 | Virginia DCLS | Virginia DCLS |
| EPI_ISL_437427 | Virginia DCLS | Virginia DCLS |
| EPI_ISL_437428 | Virginia DCLS | Virginia DCLS |
| EPI_ISL_437429 | Virginia DCLS | Virginia DCLS |
| EPI_ISL_437430 | Virginia DCLS | Virginia DCLS |
| EPI_ISL_437431 | Virginia DCLS | Virginia DCLS |
| EPI_ISL_437432 | Virginia DCLS | Virginia DCLS |
| EPI_ISL_437433 | Bozeman Health Deaconess Hospital | Wiedenheft lab, Montana State University |
| EPI_ISL_437434 | Bozeman Water Reclamation Facility | Wiedenheft lab, Montana State University |
| EPI_ISL_437435 | Veterinary Specialized Institue Kraljevo | Veterinary Specialized Institue Kraljevo |
| EPI_ISL_437436 | Veterinary Specialized Institue Kraljevo | Veterinary Specialized Institue Kraljevo |
| EPI_ISL_437437 | Alaska State Virology Laboratory | Alaska State Virology Laboratory |
| EPI_ISL_437438 | Department of MicroBiology, Government Medical College, Surat | Gujarat Biotechnology Research Centre |
| EPI_ISL_437439 | Department of MicroBiology, Government Medical College, Surat | Gujarat Biotechnology Research Centre |
| EPI_ISL_437440 | Department of MicroBiology, Government Medical College, Surat | Gujarat Biotechnology Research Centre |
| EPI_ISL_437441 | Department of MicroBiology, Government Medical College, Surat | Gujarat Biotechnology Research Centre |
| EPI_ISL_437442 | Department of MicroBiology, Government Medical College, Surat | Gujarat Biotechnology Research Centre |

| EPI_Accession_ID | Originating Laboratory | Submitting Laboratory |
|---|---|---|
| EPI_ISL_437443 | Department of MicroBiology, Government Medical College, Surat | Gujarat Biotechnology Research Centre |
| EPI_ISL_437444 | Department of MicroBiology, Government Medical College, Surat | Gujarat Biotechnology Research Centre |
| EPI_ISL_437445 | B.J. Medical College and Civil hospital | Gujarat Biotechnology Research Centre |
| EPI_ISL_437446 | B.J. Medical College and Civil hospital | Gujarat Biotechnology Research Centre |
| EPI_ISL_437447 | B.J. Medical College and Civil hospital | Gujarat Biotechnology Research Centre |
| EPI_ISL_437448 | B.J. Medical College and Civil hospital | Gujarat Biotechnology Research Centre |
| EPI_ISL_437449 | B.J. Medical College and Civil hospital | Gujarat Biotechnology Research Centre |
| EPI_ISL_437450 | B.J. Medical College and Civil hospital | Gujarat Biotechnology Research Centre |
| EPI_ISL_437451 | B.J. Medical College and Civil hospital | Gujarat Biotechnology Research Centre |
| EPI_ISL_437452 | B.J. Medical College and Civil hospital | Gujarat Biotechnology Research Centre |
| EPI_ISL_437453 | B.J. Medical College and Civil hospital | Gujarat Biotechnology Research Centre |
| EPI_ISL_437454 | B.J. Medical College and Civil hospital | Gujarat Biotechnology Research Centre |
| EPI_ISL_437455 | Clinical Diagnostics Laboratory, Diagnostic & Experimental Pathology, Lilly Research Laboratories | Clinical Diagnostics Laboratory, Diagnostic & Experimental Pathology, Lilly Research Laboratories |
| EPI_ISL_437456 | Clinical Diagnostics Laboratory, Diagnostic & Experimental Pathology, Lilly Research Laboratories | Clinical Diagnostics Laboratory, Diagnostic & Experimental Pathology, Lilly Research Laboratories |
| EPI_ISL_437457 | Clinical Diagnostics Laboratory, Diagnostic & Experimental Pathology, Lilly Research Laboratories | Clinical Diagnostics Laboratory, Diagnostic & Experimental Pathology, Lilly Research Laboratories |
| EPI_ISL_437458 | Clinical Diagnostics Laboratory, Diagnostic & Experimental Pathology, Lilly Research Laboratories | Clinical Diagnostics Laboratory, Diagnostic & Experimental Pathology, Lilly Research Laboratories |
| EPI_ISL_437459 | Pathogen Genomics Lab King Abdullah University of Science and Technology(KAUST) | Pathogen Genomics Lab King Abdullah University of Science and Technology(KAUST) |
| EPI_ISL_437460 | Pathogen Genomics Lab King Abdullah University of Science and Technology(KAUST) | Pathogen Genomics Lab King Abdullah University of Science and Technology(KAUST) |
| EPI_ISL_437461 | Pathogen Genomics Lab King Abdullah University of Science and Technology(KAUST) | Pathogen Genomics Lab King Abdullah University of Science and Technology(KAUST) |
| EPI_ISL_437462 | Pathogen Genomics Lab King Abdullah University of Science and Technology(KAUST) | Pathogen Genomics Lab King Abdullah University of Science and Technology(KAUST) |
| EPI_ISL_437463 | Pathogen Genomics Lab King Abdullah University of Science and Technology(KAUST) | Pathogen Genomics Lab King Abdullah University of Science and Technology(KAUST) |

| EPI_Accession_ID | Originating Laboratory | Submitting Laboratory |
|---|---|---|
| EPI_ISL_437464 | Pathogen Genomics Lab King Abdullah University of Science and Technology(KAUST) | Pathogen Genomics Lab King Abdullah University of Science and Technology(KAUST) |
| EPI_ISL_437465 | Pathogen Genomics Lab King Abdullah University of Science and Technology(KAUST) | Pathogen Genomics Lab King Abdullah University of Science and Technology(KAUST) |
| EPI_ISL_437466 | Pathogen Genomics Lab King Abdullah University of Science and Technology(KAUST) | Pathogen Genomics Lab King Abdullah University of Science and Technology(KAUST) |
| EPI_ISL_437467 | Pathogen Genomics Lab King Abdullah University of Science and Technology(KAUST) | Pathogen Genomics Lab King Abdullah University of Science and Technology(KAUST) |
| EPI_ISL_437468 | Pathogen Genomics Lab King Abdullah University of Science and Technology(KAUST) | Pathogen Genomics Lab King Abdullah University of Science and Technology(KAUST) |
| EPI_ISL_437469 | Pathogen Genomics Lab King Abdullah University of Science and Technology(KAUST) | Pathogen Genomics Lab King Abdullah University of Science and Technology(KAUST) |
| EPI_ISL_437470 | Pathogen Genomics Lab King Abdullah University of Science and Technology(KAUST) | Pathogen Genomics Lab King Abdullah University of Science and Technology(KAUST) |
| EPI_ISL_437471 | Pathogen Genomics Lab King Abdullah University of Science and Technology(KAUST) | Pathogen Genomics Lab King Abdullah University of Science and Technology(KAUST) |
| EPI_ISL_437472 | Pathogen Genomics Lab King Abdullah University of Science and Technology(KAUST) | Pathogen Genomics Lab King Abdullah University of Science and Technology(KAUST) |
| EPI_ISL_437473 | Pathogen Genomics Lab King Abdullah University of Science and Technology(KAUST) | Pathogen Genomics Lab King Abdullah University of Science and Technology(KAUST) |
| EPI_ISL_437474 | Pathogen Genomics Lab King Abdullah University of Science and Technology(KAUST) | Pathogen Genomics Lab King Abdullah University of Science and Technology(KAUST) |
| EPI_ISL_437475 | Pathogen Genomics Lab King Abdullah University of Science and Technology(KAUST) | Pathogen Genomics Lab King Abdullah University of Science and Technology(KAUST) |
| EPI_ISL_437476 | Pathogen Genomics Lab King Abdullah University of Science and Technology(KAUST) | Pathogen Genomics Lab King Abdullah University of Science and Technology(KAUST) |
| EPI_ISL_437477 | Pathogen Genomics Lab King Abdullah University of Science and Technology(KAUST) | Pathogen Genomics Lab King Abdullah University of Science and Technology(KAUST) |
| EPI_ISL_437478 | Pathogen Genomics Lab King Abdullah University of Science and Technology(KAUST) | Pathogen Genomics Lab King Abdullah University of Science and Technology(KAUST) |
| EPI_ISL_437479 | Pathogen Genomics Lab King Abdullah University of Science and Technology(KAUST) | Pathogen Genomics Lab King Abdullah University of Science and Technology(KAUST) |

| EPI_Accession_ID | Originating Laboratory | Submitting Laboratory |
|---|---|---|
| EPI_ISL_437481 | Pathogen Genomics Lab King Abdullah University of Science and Technology(KAUST) | Pathogen Genomics Lab King Abdullah University of Science and Technology(KAUST) |
| EPI_ISL_437482 | Pathogen Genomics Lab King Abdullah University of Science and Technology(KAUST) | Pathogen Genomics Lab King Abdullah University of Science and Technology(KAUST) |
| EPI_ISL_437483 | Pathogen Genomics Lab King Abdullah University of Science and Technology(KAUST) | Pathogen Genomics Lab King Abdullah University of Science and Technology(KAUST) |
| EPI_ISL_437484 | Pathogen Genomics Lab King Abdullah University of Science and Technology(KAUST) | Pathogen Genomics Lab King Abdullah University of Science and Technology(KAUST) |
| EPI_ISL_437485 | Pathogen Genomics Lab King Abdullah University of Science and Technology(KAUST) | Pathogen Genomics Lab King Abdullah University of Science and Technology(KAUST) |
| EPI_ISL_437486 | Pathogen Genomics Lab King Abdullah University of Science and Technology(KAUST) | Pathogen Genomics Lab King Abdullah University of Science and Technology(KAUST) |
| EPI_ISL_437487 | Pathogen Genomics Lab King Abdullah University of Science and Technology(KAUST) | Pathogen Genomics Lab King Abdullah University of Science and Technology(KAUST) |
| EPI_ISL_437488 | Pathogen Genomics Lab King Abdullah University of Science and Technology(KAUST) | Pathogen Genomics Lab King Abdullah University of Science and Technology(KAUST) |
| EPI_ISL_437489 | Pathogen Genomics Lab King Abdullah University of Science and Technology(KAUST) | Pathogen Genomics Lab King Abdullah University of Science and Technology(KAUST) |
| EPI_ISL_437490 | Pathogen Genomics Lab King Abdullah University of Science and Technology(KAUST) | Pathogen Genomics Lab King Abdullah University of Science and Technology(KAUST) |
| EPI_ISL_437491 | Pathogen Genomics Lab King Abdullah University of Science and Technology(KAUST) | Pathogen Genomics Lab King Abdullah University of Science and Technology(KAUST) |
| EPI_ISL_437492 | Pathogen Genomics Lab King Abdullah University of Science and Technology(KAUST) | Pathogen Genomics Lab King Abdullah University of Science and Technology(KAUST) |
| EPI_ISL_437493 | Pathogen Genomics Lab King Abdullah University of Science and Technology(KAUST) | Pathogen Genomics Lab King Abdullah University of Science and Technology(KAUST) |
| EPI_ISL_437494 | Pathogen Genomics Lab King Abdullah University of Science and Technology(KAUST) | Pathogen Genomics Lab King Abdullah University of Science and Technology(KAUST) |
| EPI_ISL_437495 | Pathogen Genomics Lab King Abdullah University of Science and Technology(KAUST) | Pathogen Genomics Lab King Abdullah University of Science and Technology(KAUST) |
| EPI_ISL_437496 | Pathogen Genomics Lab King Abdullah University of Science and Technology(KAUST) | Pathogen Genomics Lab King Abdullah University of Science and Technology(KAUST) |

| EPI_Accession_ID | Originating Laboratory | Submitting Laboratory |
|---|---|---|
| EPI_ISL_437497 | Pathogen Genomics Lab King Abdullah University of Science and Technology(KAUST) | Pathogen Genomics Lab King Abdullah University of Science and Technology(KAUST) |
| EPI_ISL_437512 | unknown | Human Genetic Research Center |
| EPI_ISL_437513 | Alaska State Virology Laboratory | Alaska State Virology Laboratory |
| EPI_ISL_437514 | Alaska State Virology Laboratory | Alaska State Virology Laboratory |
| EPI_ISL_437515 | Alaska State Virology Laboratory | Alaska State Virology Laboratory |
| EPI_ISL_437516 | Alaska State Virology Laboratory | Alaska State Virology Laboratory |
| EPI_ISL_437517 | Alaska State Virology Laboratory | Alaska State Virology Laboratory |
| EPI_ISL_437518 | Alaska State Virology Laboratory | Alaska State Virology Laboratory |
| EPI_ISL_437519 | The National Institute of Public Health Center for Epidemiology and Microbiology | The National Institute of Public Health Center for Epidemiology and Microbiology |
| EPI_ISL_437520 | OHSU Lab Services Molecular Microbiology Lab | Oregon SARS-CoV-2 Genome Sequencing Center |
| EPI_ISL_437521 | OHSU Lab Services Molecular Microbiology Lab | Oregon SARS-CoV-2 Genome Sequencing Center |
| EPI_ISL_437522 | OHSU Lab Services Molecular Microbiology Lab | Oregon SARS-CoV-2 Genome Sequencing Center |
| EPI_ISL_437523 | OHSU Lab Services Molecular Microbiology Lab | Oregon SARS-CoV-2 Genome Sequencing Center |
| EPI_ISL_437524 | OHSU Lab Services Molecular Microbiology Lab | Oregon SARS-CoV-2 Genome Sequencing Center |
| EPI_ISL_437525 | OHSU Lab Services Molecular Microbiology Lab | Oregon SARS-CoV-2 Genome Sequencing Center |
| EPI_ISL_437526 | OHSU Lab Services Molecular Microbiology Lab | Oregon SARS-CoV-2 Genome Sequencing Center |
| EPI_ISL_437527 | OHSU Lab Services Molecular Microbiology Lab | Oregon SARS-CoV-2 Genome Sequencing Center |
| EPI_ISL_437528 | OHSU Lab Services Molecular Microbiology Lab | Oregon SARS-CoV-2 Genome Sequencing Center |
| EPI_ISL_437529 | OHSU Lab Services Molecular Microbiology Lab | Oregon SARS-CoV-2 Genome Sequencing Center |
| EPI_ISL_437530 | OHSU Lab Services Molecular Microbiology Lab | Oregon SARS-CoV-2 Genome Sequencing Center |
| EPI_ISL_437531 | OHSU Lab Services Molecular Microbiology Lab | Oregon SARS-CoV-2 Genome Sequencing Center |
| EPI_ISL_437532 | OHSU Lab Services Molecular Microbiology Lab | Oregon SARS-CoV-2 Genome Sequencing Center |
| EPI_ISL_437533 | OHSU Lab Services Molecular Microbiology Lab | Oregon SARS-CoV-2 Genome Sequencing Center |
| EPI_ISL_437534 | OHSU Lab Services Molecular Microbiology Lab | Oregon SARS-CoV-2 Genome Sequencing Center |
| EPI_ISL_437535 | OHSU Lab Services Molecular Microbiology Lab | Oregon SARS-CoV-2 Genome Sequencing Center |
| EPI_ISL_437537 | ICMR-National Institute of Cholera and Enteric Diseases | National Institute of Biomedical Genomics |
| EPI_ISL_437538 | ICMR-National Institute of Cholera and Enteric Diseases | National Institute of Biomedical Genomics |
| EPI_ISL_437539 | ICMR-National Institute of Cholera and Enteric Diseases | National Institute of Biomedical Genomics |

| EPI_Accession_ID | Originating Laboratory | Submitting Laboratory |
|---|---|---|
| EPI_ISL_437540 | Robert Garry lab | Andersen lab at Scripps Research |
| EPI_ISL_437541 | Robert Garry lab | Andersen lab at Scripps Research |
| EPI_ISL_437542 | Robert Garry lab | Andersen lab at Scripps Research |
| EPI_ISL_437543 | Robert Garry lab | Andersen lab at Scripps Research |
| EPI_ISL_437544 | Robert Garry lab | Andersen lab at Scripps Research |
| EPI_ISL_437545 | Robert Garry lab | Andersen lab at Scripps Research |
| EPI_ISL_437546 | Robert Garry lab | Andersen lab at Scripps Research |
| EPI_ISL_437547 | Robert Garry lab | Andersen lab at Scripps Research |
| EPI_ISL_437548 | Robert Garry lab | Andersen lab at Scripps Research |
| EPI_ISL_437549 | Scripps Medical Laboratory | Andersen lab at Scripps Research |
| EPI_ISL_437550 | Scripps Medical Laboratory | Andersen lab at Scripps Research |
| EPI_ISL_437551 | Scripps Medical Laboratory | Andersen lab at Scripps Research |
| EPI_ISL_437552 | Scripps Medical Laboratory | Andersen lab at Scripps Research |
| EPI_ISL_437553 | Scripps Medical Laboratory | Andersen lab at Scripps Research |
| EPI_ISL_437554 | Scripps Medical Laboratory | Andersen lab at Scripps Research |
| EPI_ISL_437555 | Scripps Medical Laboratory | Andersen lab at Scripps Research |
| EPI_ISL_437556 | Scripps Medical Laboratory | Andersen lab at Scripps Research |
| EPI_ISL_437557 | Scripps Medical Laboratory | Andersen lab at Scripps Research |
| EPI_ISL_437558 | Scripps Medical Laboratory | Andersen lab at Scripps Research |
| EPI_ISL_437559 | Scripps Medical Laboratory | Andersen lab at Scripps Research |
| EPI_ISL_437560 | Scripps Medical Laboratory | Andersen lab at Scripps Research |
| EPI_ISL_437561 | Scripps Medical Laboratory | Andersen lab at Scripps Research |
| EPI_ISL_437562 | Scripps Medical Laboratory | Andersen lab at Scripps Research |
| EPI_ISL_437563 | Scripps Medical Laboratory | Andersen lab at Scripps Research |
| EPI_ISL_437564 | Scripps Medical Laboratory | Andersen lab at Scripps Research |
| EPI_ISL_437565 | Scripps Medical Laboratory | Andersen lab at Scripps Research |
| EPI_ISL_437566 | Scripps Medical Laboratory | Andersen lab at Scripps Research |
| EPI_ISL_437567 | Scripps Medical Laboratory | Andersen lab at Scripps Research |
| EPI_ISL_437568 | Scripps Medical Laboratory | Andersen lab at Scripps Research |
| EPI_ISL_437569 | Scripps Medical Laboratory | Andersen lab at Scripps Research |
| EPI_ISL_437570 | Scripps Medical Laboratory | Andersen lab at Scripps Research |
| EPI_ISL_437571 | Scripps Medical Laboratory | Andersen lab at Scripps Research |
| EPI_ISL_437572 | Scripps Medical Laboratory | Andersen lab at Scripps Research |

| EPI_Accession_ID | Originating Laboratory | Submitting Laboratory |
|---|---|---|
| EPI_ISL_437573 | Scripps Medical Laboratory | Andersen lab at Scripps Research |
| EPI_ISL_437574 | Scripps Medical Laboratory | Andersen lab at Scripps Research |
| EPI_ISL_437575 | Scripps Medical Laboratory | Andersen lab at Scripps Research |
| EPI_ISL_437576 | Scripps Medical Laboratory | Andersen lab at Scripps Research |
| EPI_ISL_437577 | Scripps Medical Laboratory | Andersen lab at Scripps Research |
| EPI_ISL_437578 | Scripps Medical Laboratory | Andersen lab at Scripps Research |
| EPI_ISL_437579 | Scripps Medical Laboratory | Andersen lab at Scripps Research |
| EPI_ISL_437580 | Scripps Medical Laboratory | Andersen lab at Scripps Research |
| EPI_ISL_437581 | Scripps Medical Laboratory | Andersen lab at Scripps Research |
| EPI_ISL_437582 | Scripps Medical Laboratory | Andersen lab at Scripps Research |
| EPI_ISL_437583 | Scripps Medical Laboratory | Andersen lab at Scripps Research |
| EPI_ISL_437584 | Scripps Medical Laboratory | Andersen lab at Scripps Research |
| EPI_ISL_437585 | Scripps Medical Laboratory | Andersen lab at Scripps Research |
| EPI_ISL_437586 | Scripps Medical Laboratory | Andersen lab at Scripps Research |
| EPI_ISL_437587 | Scripps Medical Laboratory | Andersen lab at Scripps Research |
| EPI_ISL_437588 | Scripps Medical Laboratory | Andersen lab at Scripps Research |
| EPI_ISL_437589 | Scripps Medical Laboratory | Andersen lab at Scripps Research |
| EPI_ISL_437590 | Scripps Medical Laboratory | Andersen lab at Scripps Research |
| EPI_ISL_437591 | Scripps Medical Laboratory | Andersen lab at Scripps Research |
| EPI_ISL_437592 | Scripps Medical Laboratory | Andersen lab at Scripps Research |
| EPI_ISL_437593 | Scripps Medical Laboratory | Andersen lab at Scripps Research |
| EPI_ISL_437594 | Scripps Medical Laboratory | Andersen lab at Scripps Research |
| EPI_ISL_437595 | Scripps Medical Laboratory | Andersen lab at Scripps Research |
| EPI_ISL_437596 | Scripps Medical Laboratory | Andersen lab at Scripps Research |
| EPI_ISL_437597 | Scripps Medical Laboratory | Andersen lab at Scripps Research |
| EPI_ISL_437598 | Scripps Medical Laboratory | Andersen lab at Scripps Research |
| EPI_ISL_437599 | Scripps Medical Laboratory | Andersen lab at Scripps Research |
| EPI_ISL_437600 | Scripps Medical Laboratory | Andersen lab at Scripps Research |
| EPI_ISL_437601 | Keio University School of Medicine | Keio University School of Medicine |
| EPI_ISL_437602 | unknown | Faculty of Medicine |
| EPI_ISL_437603 | unknown | Faculty of Medicine |
| EPI_ISL_437604 | unknown | Faculty of Medicine |
| EPI_ISL_437605 | unknown | Faculty of Medicine |

| EPI_Accession_ID | Originating Laboratory | Submitting Laboratory |
|---|---|---|
| EPI_ISL_437606 | unknown | Faculty of Medicine |
| EPI_ISL_437607 | unknown | Faculty of Medicine |
| EPI_ISL_437608 | unknown | Faculty of Medicine |
| EPI_ISL_437609 | unknown | Faculty of Medicine |
| EPI_ISL_437610 | unknown | Faculty of Medicine |
| EPI_ISL_437611 | unknown | Faculty of Medicine |
| EPI_ISL_437612 | unknown | Faculty of Medicine |
| EPI_ISL_437613 | unknown | Faculty of Medicine |
| EPI_ISL_437614 | unknown | Faculty of Medicine |
| EPI_ISL_437615 | unknown | Faculty of Medicine |
| EPI_ISL_437616 | unknown | Faculty of Medicine |
| EPI_ISL_437617 | unknown | Faculty of Medicine |
| EPI_ISL_437618 | unknown | Faculty of Medicine |
| EPI_ISL_437619 | unknown | Faculty of Medicine |
| EPI_ISL_437620 | unknown | Faculty of Medicine |
| EPI_ISL_437621 | unknown | Faculty of Medicine |
| EPI_ISL_437622 | unknown | Faculty of Medicine |
| EPI_ISL_437623 | unknown | Faculty of Medicine |
| EPI_ISL_437624 | unknown | Faculty of Medicine |
| EPI_ISL_437625 | Laboratory of Genomics & Bioinformatics, Institute of Immunology and Experimental Therapy, Polish Academy of Sciences Oddzial Mikrobiologii Wojewodzkiej Stacji Sanitarno-Epidemiologicznej. | Laboratory of Genomics & Bioinformatics, Institute of Immunology and Experimental Therapy, Polish Academy of Sciences |
| EPI_ISL_437626 | Department of Microbiology,Gandhi Medical College and Hospital | Department of Veterinary Biotechnalogy, College of Veterinary Science, Rajendranagar, PV Narsimha Rao Telengana Veterinary University |
| EPI_ISL_437627 | Department of Virus and Microbiological Special Diagnostics, Statens Serum Institut, Copenhagen, Denmark, Artillerivej 5, 2300 Copenahgen S | Albertsen lab, Department of Chemistry and Bioscience, Aalborg University, Denmark |
| EPI_ISL_437628 | Department of Virus and Microbiological Special Diagnostics, Statens Serum Institut, Copenhagen, Denmark, Artillerivej 5, 2300 Copenahgen S | Albertsen lab, Department of Chemistry and Bioscience, Aalborg University, Denmark |

| EPI_Accession_ID | Originating Laboratory | Submitting Laboratory |
|---|---|---|
| EPI_ISL_437629 | Department of Virus and Microbiological Special Diagnostics, Statens Serum Institut, Copenhagen, Denmark, Artillerivej 5, 2300 Copenahgen S | Albertsen lab, Department of Chemistry and Bioscience, Aalborg University, Denmark |
| EPI_ISL_437630 | Department of Virus and Microbiological Special Diagnostics, Statens Serum Institut, Copenhagen, Denmark, Artillerivej 5, 2300 Copenahgen S | Albertsen lab, Department of Chemistry and Bioscience, Aalborg University, Denmark |
| EPI_ISL_437631 | Department of Virus and Microbiological Special Diagnostics, Statens Serum Institut, Copenhagen, Denmark, Artillerivej 5, 2300 Copenahgen S | Albertsen lab, Department of Chemistry and Bioscience, Aalborg University, Denmark |
| EPI_ISL_437632 | Department of Virus and Microbiological Special Diagnostics, Statens Serum Institut, Copenhagen, Denmark, Artillerivej 5, 2300 Copenahgen S | Albertsen lab, Department of Chemistry and Bioscience, Aalborg University, Denmark |
| EPI_ISL_437633 | Department of Virus and Microbiological Special Diagnostics, Statens Serum Institut, Copenhagen, Denmark, Artillerivej 5, 2300 Copenahgen S | Albertsen lab, Department of Chemistry and Bioscience, Aalborg University, Denmark |
| EPI_ISL_437634 | Department of Virus and Microbiological Special Diagnostics, Statens Serum Institut, Copenhagen, Denmark, Artillerivej 5, 2300 Copenahgen S | Albertsen lab, Department of Chemistry and Bioscience, Aalborg University, Denmark |
| EPI_ISL_437635 | Department of Virus and Microbiological Special Diagnostics, Statens Serum Institut, Copenhagen, Denmark, Artillerivej 5, 2300 Copenahgen S | Albertsen lab, Department of Chemistry and Bioscience, Aalborg University, Denmark |
| EPI_ISL_437636 | Department of Virus and Microbiological Special Diagnostics, Statens Serum Institut, Copenhagen, Denmark, Artillerivej 5, 2300 Copenahgen S | Albertsen lab, Department of Chemistry and Bioscience, Aalborg University, Denmark |
| EPI_ISL_437637 | Department of Virus and Microbiological Special Diagnostics, Statens Serum Institut, Copenhagen, Denmark, Artillerivej 5, 2300 Copenahgen S | Albertsen lab, Department of Chemistry and Bioscience, Aalborg University, Denmark |
| EPI_ISL_437638 | Department of Virus and Microbiological Special Diagnostics, Statens Serum Institut, Copenhagen, Denmark, Artillerivej 5, 2300 Copenahgen S | Albertsen lab, Department of Chemistry and Bioscience, Aalborg University, Denmark |
| EPI_ISL_437639 | Department of Virus and Microbiological Special Diagnostics, Statens Serum Institut, Copenhagen, Denmark, Artillerivej 5, 2300 Copenahgen S | Albertsen lab, Department of Chemistry and Bioscience, Aalborg University, Denmark |

| EPI_Accession_ID | Originating Laboratory | Submitting Laboratory |
|---|---|---|
| EPI_ISL_437640 | Department of Virus and Microbiological Special Diagnostics, Statens Serum Institut, Copenhagen, Denmark, Artillerivej 5, 2300 Copenahgen S | Albertsen lab, Department of Chemistry and Bioscience, Aalborg University, Denmark |
| EPI_ISL_437641 | Department of Virus and Microbiological Special Diagnostics, Statens Serum Institut, Copenhagen, Denmark, Artillerivej 5, 2300 Copenahgen S | Albertsen lab, Department of Chemistry and Bioscience, Aalborg University, Denmark |
| EPI_ISL_437642 | Department of Virus and Microbiological Special Diagnostics, Statens Serum Institut, Copenhagen, Denmark, Artillerivej 5, 2300 Copenahgen S | Albertsen lab, Department of Chemistry and Bioscience, Aalborg University, Denmark |
| EPI_ISL_437643 | Department of Virus and Microbiological Special Diagnostics, Statens Serum Institut, Copenhagen, Denmark, Artillerivej 5, 2300 Copenahgen S | Albertsen lab, Department of Chemistry and Bioscience, Aalborg University, Denmark |
| EPI_ISL_437644 | Department of Virus and Microbiological Special Diagnostics, Statens Serum Institut, Copenhagen, Denmark, Artillerivej 5, 2300 Copenahgen S | Albertsen lab, Department of Chemistry and Bioscience, Aalborg University, Denmark |
| EPI_ISL_437645 | Department of Virus and Microbiological Special Diagnostics, Statens Serum Institut, Copenhagen, Denmark, Artillerivej 5, 2300 Copenahgen S | Albertsen lab, Department of Chemistry and Bioscience, Aalborg University, Denmark |
| EPI_ISL_437646 | Department of Virus and Microbiological Special Diagnostics, Statens Serum Institut, Copenhagen, Denmark, Artillerivej 5, 2300 Copenahgen S | Albertsen lab, Department of Chemistry and Bioscience, Aalborg University, Denmark |
| EPI_ISL_437647 | Department of Virus and Microbiological Special Diagnostics, Statens Serum Institut, Copenhagen, Denmark, Artillerivej 5, 2300 Copenahgen S | Albertsen lab, Department of Chemistry and Bioscience, Aalborg University, Denmark |
| EPI_ISL_437648 | Department of Virus and Microbiological Special Diagnostics, Statens Serum Institut, Copenhagen, Denmark, Artillerivej 5, 2300 Copenahgen S | Albertsen lab, Department of Chemistry and Bioscience, Aalborg University, Denmark |
| EPI_ISL_437649 | Department of Virus and Microbiological Special Diagnostics, Statens Serum Institut, Copenhagen, Denmark, Artillerivej 5, 2300 Copenahgen S | Albertsen lab, Department of Chemistry and Bioscience, Aalborg University, Denmark |
| EPI_ISL_437650 | Department of Virus and Microbiological Special Diagnostics, Statens Serum Institut, Copenhagen, Denmark, Artillerivej 5, 2300 Copenahgen S | Albertsen lab, Department of Chemistry and Bioscience, Aalborg University, Denmark |

| EPI_Accession_ID | Originating Laboratory | Submitting Laboratory |
|---|---|---|
| EPI_ISL_437651 | Department of Virus and Microbiological Special Diagnostics, Statens Serum Institut, Copenhagen, Denmark, Artillerivej 5, 2300 Copenahgen S | Albertsen lab, Department of Chemistry and Bioscience, Aalborg University, Denmark |
| EPI_ISL_437652 | Department of Virus and Microbiological Special Diagnostics, Statens Serum Institut, Copenhagen, Denmark, Artillerivej 5, 2300 Copenahgen S | Albertsen lab, Department of Chemistry and Bioscience, Aalborg University, Denmark |
| EPI_ISL_437653 | Department of Virus and Microbiological Special Diagnostics, Statens Serum Institut, Copenhagen, Denmark, Artillerivej 5, 2300 Copenahgen S | Albertsen lab, Department of Chemistry and Bioscience, Aalborg University, Denmark |
| EPI_ISL_437654 | Department of Virus and Microbiological Special Diagnostics, Statens Serum Institut, Copenhagen, Denmark, Artillerivej 5, 2300 Copenahgen S | Albertsen lab, Department of Chemistry and Bioscience, Aalborg University, Denmark |
| EPI_ISL_437655 | Department of Virus and Microbiological Special Diagnostics, Statens Serum Institut, Copenhagen, Denmark, Artillerivej 5, 2300 Copenahgen S | Albertsen lab, Department of Chemistry and Bioscience, Aalborg University, Denmark |
| EPI_ISL_437656 | Department of Virus and Microbiological Special Diagnostics, Statens Serum Institut, Copenhagen, Denmark, Artillerivej 5, 2300 Copenahgen S | Albertsen lab, Department of Chemistry and Bioscience, Aalborg University, Denmark |
| EPI_ISL_437657 | Department of Virus and Microbiological Special Diagnostics, Statens Serum Institut, Copenhagen, Denmark, Artillerivej 5, 2300 Copenahgen S | Albertsen lab, Department of Chemistry and Bioscience, Aalborg University, Denmark |
| EPI_ISL_437658 | Department of Virus and Microbiological Special Diagnostics, Statens Serum Institut, Copenhagen, Denmark, Artillerivej 5, 2300 Copenahgen S | Albertsen lab, Department of Chemistry and Bioscience, Aalborg University, Denmark |
| EPI_ISL_437659 | Department of Virus and Microbiological Special Diagnostics, Statens Serum Institut, Copenhagen, Denmark, Artillerivej 5, 2300 Copenahgen S | Albertsen lab, Department of Chemistry and Bioscience, Aalborg University, Denmark |
| EPI_ISL_437660 | Department of Virus and Microbiological Special Diagnostics, Statens Serum Institut, Copenhagen, Denmark, Artillerivej 5, 2300 Copenahgen S | Albertsen lab, Department of Chemistry and Bioscience, Aalborg University, Denmark |
| EPI_ISL_437661 | Department of Virus and Microbiological Special Diagnostics, Statens Serum Institut, Copenhagen, Denmark, Artillerivej 5, 2300 Copenahgen S | Albertsen lab, Department of Chemistry and Bioscience, Aalborg University, Denmark |

| EPI_Accession_ID | Originating Laboratory | Submitting Laboratory |
|---|---|---|
| EPI_ISL_437662 | Department of Virus and Microbiological Special Diagnostics, Statens Serum Institut, Copenhagen, Denmark, Artillerivej 5, 2300 Copenahgen S | Albertsen lab, Department of Chemistry and Bioscience, Aalborg University, Denmark |
| EPI_ISL_437663 | Department of Virus and Microbiological Special Diagnostics, Statens Serum Institut, Copenhagen, Denmark, Artillerivej 5, 2300 Copenahgen S | Albertsen lab, Department of Chemistry and Bioscience, Aalborg University, Denmark |
| EPI_ISL_437664 | Department of Virus and Microbiological Special Diagnostics, Statens Serum Institut, Copenhagen, Denmark, Artillerivej 5, 2300 Copenahgen S | Albertsen lab, Department of Chemistry and Bioscience, Aalborg University, Denmark |
| EPI_ISL_437665 | Department of Virus and Microbiological Special Diagnostics, Statens Serum Institut, Copenhagen, Denmark, Artillerivej 5, 2300 Copenahgen S | Albertsen lab, Department of Chemistry and Bioscience, Aalborg University, Denmark |
| EPI_ISL_437666 | Department of Virus and Microbiological Special Diagnostics, Statens Serum Institut, Copenhagen, Denmark, Artillerivej 5, 2300 Copenahgen S | Albertsen lab, Department of Chemistry and Bioscience, Aalborg University, Denmark |
| EPI_ISL_437667 | Department of Virus and Microbiological Special Diagnostics, Statens Serum Institut, Copenhagen, Denmark, Artillerivej 5, 2300 Copenahgen S | Albertsen lab, Department of Chemistry and Bioscience, Aalborg University, Denmark |
| EPI_ISL_437668 | Department of Virus and Microbiological Special Diagnostics, Statens Serum Institut, Copenhagen, Denmark, Artillerivej 5, 2300 Copenahgen S | Albertsen lab, Department of Chemistry and Bioscience, Aalborg University, Denmark |
| EPI_ISL_437669 | Department of Virus and Microbiological Special Diagnostics, Statens Serum Institut, Copenhagen, Denmark, Artillerivej 5, 2300 Copenahgen S | Albertsen lab, Department of Chemistry and Bioscience, Aalborg University, Denmark |
| EPI_ISL_437670 | Department of Virus and Microbiological Special Diagnostics, Statens Serum Institut, Copenhagen, Denmark, Artillerivej 5, 2300 Copenahgen S | Albertsen lab, Department of Chemistry and Bioscience, Aalborg University, Denmark |
| EPI_ISL_437671 | Department of Virus and Microbiological Special Diagnostics, Statens Serum Institut, Copenhagen, Denmark, Artillerivej 5, 2300 Copenahgen S | Albertsen lab, Department of Chemistry and Bioscience, Aalborg University, Denmark |
| EPI_ISL_437672 | Department of Virus and Microbiological Special Diagnostics, Statens Serum Institut, Copenhagen, Denmark, Artillerivej 5, 2300 Copenahgen S | Albertsen lab, Department of Chemistry and Bioscience, Aalborg University, Denmark |

| EPI_Accession_ID | Originating Laboratory | Submitting Laboratory |
|---|---|---|
| EPI_ISL_437673 | Department of Virus and Microbiological Special Diagnostics, Statens Serum Institut, Copenhagen, Denmark, Artillerivej 5, 2300 Copenahgen S | Albertsen lab, Department of Chemistry and Bioscience, Aalborg University, Denmark |
| EPI_ISL_437674 | Department of Virus and Microbiological Special Diagnostics, Statens Serum Institut, Copenhagen, Denmark, Artillerivej 5, 2300 Copenahgen S | Albertsen lab, Department of Chemistry and Bioscience, Aalborg University, Denmark |
| EPI_ISL_437675 | Department of Virus and Microbiological Special Diagnostics, Statens Serum Institut, Copenhagen, Denmark, Artillerivej 5, 2300 Copenahgen S | Albertsen lab, Department of Chemistry and Bioscience, Aalborg University, Denmark |
| EPI_ISL_437676 | Department of Virus and Microbiological Special Diagnostics, Statens Serum Institut, Copenhagen, Denmark, Artillerivej 5, 2300 Copenahgen S | Albertsen lab, Department of Chemistry and Bioscience, Aalborg University, Denmark |
| EPI_ISL_437677 | Department of Virus and Microbiological Special Diagnostics, Statens Serum Institut, Copenhagen, Denmark, Artillerivej 5, 2300 Copenahgen S | Albertsen lab, Department of Chemistry and Bioscience, Aalborg University, Denmark |
| EPI_ISL_437678 | Department of Virus and Microbiological Special Diagnostics, Statens Serum Institut, Copenhagen, Denmark, Artillerivej 5, 2300 Copenahgen S | Albertsen lab, Department of Chemistry and Bioscience, Aalborg University, Denmark |
| EPI_ISL_437679 | Department of Virus and Microbiological Special Diagnostics, Statens Serum Institut, Copenhagen, Denmark, Artillerivej 5, 2300 Copenahgen S | Albertsen lab, Department of Chemistry and Bioscience, Aalborg University, Denmark |
| EPI_ISL_437680 | Department of Virus and Microbiological Special Diagnostics, Statens Serum Institut, Copenhagen, Denmark, Artillerivej 5, 2300 Copenahgen S | Albertsen lab, Department of Chemistry and Bioscience, Aalborg University, Denmark |
| EPI_ISL_437681 | Department of Virus and Microbiological Special Diagnostics, Statens Serum Institut, Copenhagen, Denmark, Artillerivej 5, 2300 Copenahgen S | Albertsen lab, Department of Chemistry and Bioscience, Aalborg University, Denmark |
| EPI_ISL_437682 | Department of Virus and Microbiological Special Diagnostics, Statens Serum Institut, Copenhagen, Denmark, Artillerivej 5, 2300 Copenahgen S | Albertsen lab, Department of Chemistry and Bioscience, Aalborg University, Denmark |
| EPI_ISL_437683 | Department of Virus and Microbiological Special Diagnostics, Statens Serum Institut, Copenhagen, Denmark, Artillerivej 5, 2300 Copenahgen S | Albertsen lab, Department of Chemistry and Bioscience, Aalborg University, Denmark |

| EPI_Accession_ID | Originating Laboratory | Submitting Laboratory |
|---|---|---|
| EPI_ISL_437689 | Laboratory for Urgent Response to Biological Threats | Institut Pasteur CIBU / ERI |
| EPI_ISL_437690 | Laboratory for Urgent Response to Biological Threats | Institut Pasteur CIBU /ERI |
| EPI_ISL_437691 | Pathogen Genomics Lab King Abdullah University of Science and Technology(KAUST) | Pathogen Genomics Lab King Abdullah University of Science and Technology(KAUST) |
| EPI_ISL_437692 | Pathogen Genomics Lab King Abdullah University of Science and Technology(KAUST) | Pathogen Genomics Lab King Abdullah University of Science and Technology(KAUST) |
| EPI_ISL_437693 | Pathogen Genomics Lab King Abdullah University of Science and Technology(KAUST) | Pathogen Genomics Lab King Abdullah University of Science and Technology(KAUST) |
| EPI_ISL_437694 | Pathogen Genomics Lab King Abdullah University of Science and Technology(KAUST) | Pathogen Genomics Lab King Abdullah University of Science and Technology(KAUST) |
| EPI_ISL_437695 | Pathogen Genomics Lab King Abdullah University of Science and Technology(KAUST) | Pathogen Genomics Lab King Abdullah University of Science and Technology(KAUST) |
| EPI_ISL_437696 | Pathogen Genomics Lab King Abdullah University of Science and Technology(KAUST) | Pathogen Genomics Lab King Abdullah University of Science and Technology(KAUST) |
| EPI_ISL_437697 | Pathogen Genomics Lab King Abdullah University of Science and Technology(KAUST) | Pathogen Genomics Lab King Abdullah University of Science and Technology(KAUST) |
| EPI_ISL_437698 | Pathogen Genomics Lab King Abdullah University of Science and Technology(KAUST) | Pathogen Genomics Lab King Abdullah University of Science and Technology(KAUST) |
| EPI_ISL_437699 | Pathogen Genomics Lab King Abdullah University of Science and Technology(KAUST) | Pathogen Genomics Lab King Abdullah University of Science and Technology(KAUST) |
| EPI_ISL_437700 | Pathogen Genomics Lab King Abdullah University of Science and Technology(KAUST) | Pathogen Genomics Lab King Abdullah University of Science and Technology(KAUST) |
| EPI_ISL_437701 | Pathogen Genomics Lab King Abdullah University of Science and Technology(KAUST) | Pathogen Genomics Lab King Abdullah University of Science and Technology(KAUST) |
| EPI_ISL_437702 | Pathogen Genomics Lab King Abdullah University of Science and Technology(KAUST) | Pathogen Genomics Lab King Abdullah University of Science and Technology(KAUST) |
| EPI_ISL_437703 | Pathogen Genomics Lab King Abdullah University of Science and Technology(KAUST) | Pathogen Genomics Lab King Abdullah University of Science and Technology(KAUST) |
| EPI_ISL_437704 | Pathogen Genomics Lab King Abdullah University of Science and Technology(KAUST) | Pathogen Genomics Lab King Abdullah University of Science and Technology(KAUST) |
| EPI_ISL_437705 | Pathogen Genomics Lab King Abdullah University of Science and Technology(KAUST) | Pathogen Genomics Lab King Abdullah University of Science and Technology(KAUST) |

| EPI_Accession_ID | Originating Laboratory | Submitting Laboratory |
|---|---|---|
| EPI_ISL_437706 | Pathogen Genomics Lab King Abdullah University of Science and Technology(KAUST) | Pathogen Genomics Lab King Abdullah University of Science and Technology(KAUST) |
| EPI_ISL_437707 | Pathogen Genomics Lab King Abdullah University of Science and Technology(KAUST) | Pathogen Genomics Lab King Abdullah University of Science and Technology(KAUST) |
| EPI_ISL_437708 | Pathogen Genomics Lab King Abdullah University of Science and Technology(KAUST) | Pathogen Genomics Lab King Abdullah University of Science and Technology(KAUST) |
| EPI_ISL_437709 | Pathogen Genomics Lab King Abdullah University of Science and Technology(KAUST) | Pathogen Genomics Lab King Abdullah University of Science and Technology(KAUST) |
| EPI_ISL_437710 | Pathogen Genomics Lab King Abdullah University of Science and Technology(KAUST) | Pathogen Genomics Lab King Abdullah University of Science and Technology(KAUST) |
| EPI_ISL_437711 | Pathogen Genomics Lab King Abdullah University of Science and Technology(KAUST) | Pathogen Genomics Lab King Abdullah University of Science and Technology(KAUST) |
| EPI_ISL_437712 | Pathogen Genomics Lab King Abdullah University of Science and Technology(KAUST) | Pathogen Genomics Lab King Abdullah University of Science and Technology(KAUST) |
| EPI_ISL_437713 | Pathogen Genomics Lab King Abdullah University of Science and Technology(KAUST) | Pathogen Genomics Lab King Abdullah University of Science and Technology(KAUST) |
| EPI_ISL_437714 | Pathogen Genomics Lab King Abdullah University of Science and Technology(KAUST) | Pathogen Genomics Lab King Abdullah University of Science and Technology(KAUST) |
| EPI_ISL_437715 | Pathogen Genomics Lab King Abdullah University of Science and Technology(KAUST) | Pathogen Genomics Lab King Abdullah University of Science and Technology(KAUST) |
| EPI_ISL_437716 | Pathogen Genomics Lab King Abdullah University of Science and Technology(KAUST) | Pathogen Genomics Lab King Abdullah University of Science and Technology(KAUST) |
| EPI_ISL_437717 | Pathogen Genomics Lab King Abdullah University of Science and Technology(KAUST) | Pathogen Genomics Lab King Abdullah University of Science and Technology(KAUST) |
| EPI_ISL_437718 | Pathogen Genomics Lab King Abdullah University of Science and Technology(KAUST) | Pathogen Genomics Lab King Abdullah University of Science and Technology(KAUST) |
| EPI_ISL_437719 | Pathogen Genomics Lab King Abdullah University of Science and Technology(KAUST) | Pathogen Genomics Lab King Abdullah University of Science and Technology(KAUST) |
| EPI_ISL_437720 | Pathogen Genomics Lab King Abdullah University of Science and Technology(KAUST) | Pathogen Genomics Lab King Abdullah University of Science and Technology(KAUST) |
| EPI_ISL_437721 | Pathogen Genomics Lab King Abdullah University of Science and Technology(KAUST) | Pathogen Genomics Lab King Abdullah University of Science and Technology(KAUST) |

| EPI_Accession_ID | Originating Laboratory | Submitting Laboratory |
|---|---|---|
| EPI_ISL_437722 | Pathogen Genomics Lab King Abdullah University of Science and Technology(KAUST) | Pathogen Genomics Lab King Abdullah University of Science and Technology(KAUST) |
| EPI_ISL_437723 | Pathogen Genomics Lab King Abdullah University of Science and Technology(KAUST) | Pathogen Genomics Lab King Abdullah University of Science and Technology(KAUST) |
| EPI_ISL_437724 | Pathogen Genomics Lab King Abdullah University of Science and Technology(KAUST) | Pathogen Genomics Lab King Abdullah University of Science and Technology(KAUST) |
| EPI_ISL_437725 | Pathogen Genomics Lab King Abdullah University of Science and Technology(KAUST) | Pathogen Genomics Lab King Abdullah University of Science and Technology(KAUST) |
| EPI_ISL_437726 | Pathogen Genomics Lab King Abdullah University of Science and Technology(KAUST) | Pathogen Genomics Lab King Abdullah University of Science and Technology(KAUST) |
| EPI_ISL_437727 | Pathogen Genomics Lab King Abdullah University of Science and Technology(KAUST) | Pathogen Genomics Lab King Abdullah University of Science and Technology(KAUST) |
| EPI_ISL_437728 | Pathogen Genomics Lab King Abdullah University of Science and Technology(KAUST) | Pathogen Genomics Lab King Abdullah University of Science and Technology(KAUST) |
| EPI_ISL_437729 | Pathogen Genomics Lab King Abdullah University of Science and Technology(KAUST) | Pathogen Genomics Lab King Abdullah University of Science and Technology(KAUST) |
| EPI_ISL_437730 | Pathogen Genomics Lab King Abdullah University of Science and Technology(KAUST) | Pathogen Genomics Lab King Abdullah University of Science and Technology(KAUST) |
| EPI_ISL_437731 | Pathogen Genomics Lab King Abdullah University of Science and Technology(KAUST) | Pathogen Genomics Lab King Abdullah University of Science and Technology(KAUST) |
| EPI_ISL_437732 | Pathogen Genomics Lab King Abdullah University of Science and Technology(KAUST) | Pathogen Genomics Lab King Abdullah University of Science and Technology(KAUST) |
| EPI_ISL_437733 | Pathogen Genomics Lab King Abdullah University of Science and Technology(KAUST) | Pathogen Genomics Lab King Abdullah University of Science and Technology(KAUST) |
| EPI_ISL_437734 | Pathogen Genomics Lab King Abdullah University of Science and Technology(KAUST) | Pathogen Genomics Lab King Abdullah University of Science and Technology(KAUST) |
| EPI_ISL_437735 | Pathogen Genomics Lab King Abdullah University of Science and Technology(KAUST) | Pathogen Genomics Lab King Abdullah University of Science and Technology(KAUST) |
| EPI_ISL_437736 | Pathogen Genomics Lab King Abdullah University of Science and Technology(KAUST) | Pathogen Genomics Lab King Abdullah University of Science and Technology(KAUST) |
| EPI_ISL_437737 | Pathogen Genomics Lab King Abdullah University of Science and Technology(KAUST) | Pathogen Genomics Lab King Abdullah University of Science and Technology(KAUST) |

| EPI_Accession_ID | Originating Laboratory | Submitting Laboratory |
|---|---|---|
| EPI_ISL_437738 | Pathogen Genomics Lab King Abdullah University of Science and Technology(KAUST) | Pathogen Genomics Lab King Abdullah University of Science and Technology(KAUST) |
| EPI_ISL_437739 | Pathogen Genomics Lab King Abdullah University of Science and Technology(KAUST) | Pathogen Genomics Lab King Abdullah University of Science and Technology(KAUST) |
| EPI_ISL_437740 | Pathogen Genomics Lab King Abdullah University of Science and Technology(KAUST) | Pathogen Genomics Lab King Abdullah University of Science and Technology(KAUST) |
| EPI_ISL_437741 | Pathogen Genomics Lab King Abdullah University of Science and Technology(KAUST) | Pathogen Genomics Lab King Abdullah University of Science and Technology(KAUST) |
| EPI_ISL_437742 | Pathogen Genomics Lab King Abdullah University of Science and Technology(KAUST) | Pathogen Genomics Lab King Abdullah University of Science and Technology(KAUST) |
| EPI_ISL_437743 | Pathogen Genomics Lab King Abdullah University of Science and Technology(KAUST) | Pathogen Genomics Lab King Abdullah University of Science and Technology(KAUST) |
| EPI_ISL_437744 | Pathogen Genomics Lab King Abdullah University of Science and Technology(KAUST) | Pathogen Genomics Lab King Abdullah University of Science and Technology(KAUST) |
| EPI_ISL_437745 | Pathogen Genomics Lab King Abdullah University of Science and Technology(KAUST) | Pathogen Genomics Lab King Abdullah University of Science and Technology(KAUST) |
| EPI_ISL_437746 | Pathogen Genomics Lab King Abdullah University of Science and Technology(KAUST) | Pathogen Genomics Lab King Abdullah University of Science and Technology(KAUST) |
| EPI_ISL_437747 | Pathogen Genomics Lab King Abdullah University of Science and Technology(KAUST) | Pathogen Genomics Lab King Abdullah University of Science and Technology(KAUST) |
| EPI_ISL_437748 | Pathogen Genomics Lab King Abdullah University of Science and Technology(KAUST) | Pathogen Genomics Lab King Abdullah University of Science and Technology(KAUST) |
| EPI_ISL_437749 | Pathogen Genomics Lab King Abdullah University of Science and Technology(KAUST) | Pathogen Genomics Lab King Abdullah University of Science and Technology(KAUST) |
| EPI_ISL_437750 | Pathogen Genomics Lab King Abdullah University of Science and Technology(KAUST) | Pathogen Genomics Lab King Abdullah University of Science and Technology(KAUST) |
| EPI_ISL_437751 | Pathogen Genomics Lab King Abdullah University of Science and Technology(KAUST) | Pathogen Genomics Lab King Abdullah University of Science and Technology(KAUST) |
| EPI_ISL_437752 | Pathogen Genomics Lab King Abdullah University of Science and Technology(KAUST) | Pathogen Genomics Lab King Abdullah University of Science and Technology(KAUST) |
| EPI_ISL_437753 | Pathogen Genomics Lab King Abdullah University of Science and Technology(KAUST) | Pathogen Genomics Lab King Abdullah University of Science and Technology(KAUST) |

| EPI_Accession_ID | Originating Laboratory | Submitting Laboratory |
|---|---|---|
| EPI_ISL_437754 | Pathogen Genomics Lab King Abdullah University of Science and Technology(KAUST) | Pathogen Genomics Lab King Abdullah University of Science and Technology(KAUST) |
| EPI_ISL_437755 | Pathogen Genomics Lab King Abdullah University of Science and Technology(KAUST) | Pathogen Genomics Lab King Abdullah University of Science and Technology(KAUST) |
| EPI_ISL_437756 | Pathogen Genomics Lab King Abdullah University of Science and Technology(KAUST) | Pathogen Genomics Lab King Abdullah University of Science and Technology(KAUST) |
| EPI_ISL_437757 | Pathogen Genomics Lab King Abdullah University of Science and Technology(KAUST) | Pathogen Genomics Lab King Abdullah University of Science and Technology(KAUST) |
| EPI_ISL_437758 | Pathogen Genomics Lab King Abdullah University of Science and Technology(KAUST) | Pathogen Genomics Lab King Abdullah University of Science and Technology(KAUST) |
| EPI_ISL_437759 | Pathogen Genomics Lab King Abdullah University of Science and Technology(KAUST) | Pathogen Genomics Lab King Abdullah University of Science and Technology(KAUST) |
| EPI_ISL_437760 | Pathogen Genomics Lab King Abdullah University of Science and Technology(KAUST) | Pathogen Genomics Lab King Abdullah University of Science and Technology(KAUST) |
| EPI_ISL_437761 | Pathogen Genomics Lab King Abdullah University of Science and Technology(KAUST) | Pathogen Genomics Lab King Abdullah University of Science and Technology(KAUST) |
| EPI_ISL_437762 | Pathogen Genomics Lab King Abdullah University of Science and Technology(KAUST) | Pathogen Genomics Lab King Abdullah University of Science and Technology(KAUST) |
| EPI_ISL_437763 | Virginia DCLS | Virginia DCLS |
| EPI_ISL_437764 | Virginia DCLS | Virginia DCLS |
| EPI_ISL_437765 | Virginia DCLS | Virginia DCLS |
| EPI_ISL_437766 | Virginia DCLS | Virginia DCLS |
| EPI_ISL_437767 | Virginia DCLS | Virginia DCLS |
| EPI_ISL_437768 | Virginia DCLS | Virginia DCLS |
| EPI_ISL_437769 | Virginia DCLS | Virginia DCLS |
| EPI_ISL_437770 | Virginia DCLS | Virginia DCLS |
| EPI_ISL_437771 | Virginia DCLS | Virginia DCLS |
| EPI_ISL_437772 | Virginia DCLS | Virginia DCLS |
| EPI_ISL_437773 | Virginia DCLS | Virginia DCLS |
| EPI_ISL_437774 | Virginia DCLS | Virginia DCLS |
| EPI_ISL_437775 | Virginia DCLS | Virginia DCLS |
| EPI_ISL_437776 | Virginia DCLS | Virginia DCLS |
| EPI_ISL_437777 | Virginia DCLS | Virginia DCLS |

| EPI_Accession_ID | Originating Laboratory | Submitting Laboratory |
|---|---|---|
| EPI_ISL_437778 | Virginia DCLS | Virginia DCLS |
| EPI_ISL_437779 | Virginia DCLS | Virginia DCLS |
| EPI_ISL_437780 | Virginia DCLS | Virginia DCLS |
| EPI_ISL_437781 | Virginia DCLS | Virginia DCLS |
| EPI_ISL_437782 | Virginia DCLS | Virginia DCLS |
| EPI_ISL_437783 | Virginia DCLS | Virginia DCLS |
| EPI_ISL_437784 | Virginia DCLS | Virginia DCLS |
| EPI_ISL_437785 | Virginia DCLS | Virginia DCLS |
| EPI_ISL_437786 | Virginia DCLS | Virginia DCLS |
| EPI_ISL_437787 | Virginia DCLS | Virginia DCLS |
| EPI_ISL_437788 | Virginia DCLS | Virginia DCLS |
| EPI_ISL_437789 | Virginia DCLS | Virginia DCLS |
| EPI_ISL_437790 | Virginia DCLS | Virginia DCLS |
| EPI_ISL_437791 | Virginia DCLS | Virginia DCLS |
| EPI_ISL_437792 | Virginia DCLS | Virginia DCLS |
| EPI_ISL_437793 | Virginia DCLS | Virginia DCLS |
| EPI_ISL_437794 | Virginia DCLS | Virginia DCLS |
| EPI_ISL_437795 | Virginia DCLS | Virginia DCLS |
| EPI_ISL_437796 | Virginia DCLS | Virginia DCLS |
| EPI_ISL_437797 | Virginia DCLS | Virginia DCLS |
| EPI_ISL_437798 | Virginia DCLS | Virginia DCLS |
| EPI_ISL_437799 | Virginia DCLS | Virginia DCLS |
| EPI_ISL_437800 | Virginia DCLS | Virginia DCLS |
| EPI_ISL_437801 | Virginia DCLS | Virginia DCLS |
| EPI_ISL_437802 | Virginia DCLS | Virginia DCLS |
| EPI_ISL_437803 | UW Virology Lab | UW Virology Lab |
| EPI_ISL_437804 | UW Virology Lab | UW Virology Lab |
| EPI_ISL_437805 | UW Virology Lab | UW Virology Lab |
| EPI_ISL_437806 | UW Virology Lab | UW Virology Lab |
| EPI_ISL_437807 | UW Virology Lab | UW Virology Lab |
| EPI_ISL_437808 | UW Virology Lab | UW Virology Lab |
| EPI_ISL_437809 | UW Virology Lab | UW Virology Lab |
| EPI_ISL_437810 | UW Virology Lab | UW Virology Lab |

| EPI_Accession_ID | Originating Laboratory | Submitting Laboratory |
|---|---|---|
| EPI_ISL_437811 | UW Virology Lab | UW Virology Lab |
| EPI_ISL_437812 | UW Virology Lab | UW Virology Lab |
| EPI_ISL_437813 | UW Virology Lab | UW Virology Lab |
| EPI_ISL_437814 | UW Virology Lab | UW Virology Lab |
| EPI_ISL_437815 | UW Virology Lab | UW Virology Lab |
| EPI_ISL_437816 | UW Virology Lab | UW Virology Lab |
| EPI_ISL_437817 | UW Virology Lab | UW Virology Lab |
| EPI_ISL_437818 | UW Virology Lab | UW Virology Lab |
| EPI_ISL_437819 | UW Virology Lab | UW Virology Lab |
| EPI_ISL_437820 | UW Virology Lab | UW Virology Lab |
| EPI_ISL_437821 | UW Virology Lab | UW Virology Lab |
| EPI_ISL_437822 | UW Virology Lab | UW Virology Lab |
| EPI_ISL_437823 | UW Virology Lab | UW Virology Lab |
| EPI_ISL_437824 | UW Virology Lab | UW Virology Lab |
| EPI_ISL_437825 | UW Virology Lab | UW Virology Lab |
| EPI_ISL_437826 | UW Virology Lab | UW Virology Lab |
| EPI_ISL_437827 | UW Virology Lab | UW Virology Lab |
| EPI_ISL_437828 | UW Virology Lab | UW Virology Lab |
| EPI_ISL_437829 | UW Virology Lab | UW Virology Lab |
| EPI_ISL_437830 | UW Virology Lab | UW Virology Lab |
| EPI_ISL_437831 | UW Virology Lab | UW Virology Lab |
| EPI_ISL_437832 | UW Virology Lab | UW Virology Lab |
| EPI_ISL_437833 | UW Virology Lab | UW Virology Lab |
| EPI_ISL_437834 | UW Virology Lab | UW Virology Lab |
| EPI_ISL_437835 | UW Virology Lab | UW Virology Lab |
| EPI_ISL_437836 | UW Virology Lab | UW Virology Lab |
| EPI_ISL_437837 | UW Virology Lab | UW Virology Lab |
| EPI_ISL_437838 | UW Virology Lab | UW Virology Lab |
| EPI_ISL_437839 | UW Virology Lab | UW Virology Lab |
| EPI_ISL_437840 | UW Virology Lab | UW Virology Lab |
| EPI_ISL_437841 | UW Virology Lab | UW Virology Lab |
| EPI_ISL_437842 | UW Virology Lab | UW Virology Lab |
| EPI_ISL_437843 | UW Virology Lab | UW Virology Lab |

| EPI_Accession_ID | Originating Laboratory | Submitting Laboratory |
|---|---|---|
| EPI_ISL_437844 | UW Virology Lab | UW Virology Lab |
| EPI_ISL_437845 | UW Virology Lab | UW Virology Lab |
| EPI_ISL_437846 | UW Virology Lab | UW Virology Lab |
| EPI_ISL_437847 | UW Virology Lab | UW Virology Lab |
| EPI_ISL_437848 | UW Virology Lab | UW Virology Lab |
| EPI_ISL_437849 | UW Virology Lab | UW Virology Lab |
| EPI_ISL_437850 | UW Virology Lab | UW Virology Lab |
| EPI_ISL_437851 | UW Virology Lab | UW Virology Lab |
| EPI_ISL_437852 | UW Virology Lab | UW Virology Lab |
| EPI_ISL_437853 | UW Virology Lab | UW Virology Lab |
| EPI_ISL_437854 | UW Virology Lab | UW Virology Lab |
| EPI_ISL_437855 | UW Virology Lab | UW Virology Lab |
| EPI_ISL_437856 | UW Virology Lab | UW Virology Lab |
| EPI_ISL_437857 | UW Virology Lab | UW Virology Lab |
| EPI_ISL_437858 | UW Virology Lab | UW Virology Lab |
| EPI_ISL_437859 | UW Virology Lab | UW Virology Lab |
| EPI_ISL_437860 | UW Virology Lab | UW Virology Lab |
| EPI_ISL_437861 | UW Virology Lab | UW Virology Lab |
| EPI_ISL_437862 | UW Virology Lab | UW Virology Lab |
| EPI_ISL_437863 | UW Virology Lab | UW Virology Lab |
| EPI_ISL_437864 | UW Virology Lab | UW Virology Lab |
| EPI_ISL_437865 | UW Virology Lab | UW Virology Lab |
| EPI_ISL_437866 | UW Virology Lab | UW Virology Lab |
| EPI_ISL_437867 | UW Virology Lab | UW Virology Lab |
| EPI_ISL_437868 | UW Virology Lab | UW Virology Lab |
| EPI_ISL_437869 | UW Virology Lab | UW Virology Lab |
| EPI_ISL_437870 | UW Virology Lab | UW Virology Lab |
| EPI_ISL_437871 | UW Virology Lab | UW Virology Lab |
| EPI_ISL_437872 | UW Virology Lab | UW Virology Lab |
| EPI_ISL_437873 | Alaska State Virology Laboratory | Alaska State Virology Laboratory |
| EPI_ISL_437874 | Laboratory of Microbiology, Medical School, National and Kapodistrian University of Athens | Laboratory of Biology, Department of Medicine, Democritus University of Thrace |

| EPI_Accession_ID | Originating Laboratory | Submitting Laboratory |
|---|---|---|
| EPI_ISL_437875 | Laboratory of Microbiology, Medical School, National and Kapodistrian University of Athens | Laboratory of Biology, Department of Medicine, Democritus University of Thrace |
| EPI_ISL_437876 | Laboratory of Microbiology, Medical School, National and Kapodistrian University of Athens | Laboratory of Biology, Department of Medicine, Democritus University of Thrace |
| EPI_ISL_437877 | Laboratory of Microbiology, Medical School, National and Kapodistrian University of Athens | Laboratory of Biology, Department of Medicine, Democritus University of Thrace |
| EPI_ISL_437878 | Laboratory of Microbiology, Medical School, National and Kapodistrian University of Athens | Laboratory of Biology, Department of Medicine, Democritus University of Thrace |
| EPI_ISL_437879 | Laboratory of Microbiology, Medical School, National and Kapodistrian University of Athens | Laboratory of Biology, Department of Medicine, Democritus University of Thrace |
| EPI_ISL_437880 | Laboratory of Microbiology, Medical School, National and Kapodistrian University of Athens | Laboratory of Biology, Department of Medicine, Democritus University of Thrace |
| EPI_ISL_437881 | Laboratory of Microbiology, Medical School, National and Kapodistrian University of Athens | Laboratory of Biology, Department of Medicine, Democritus University of Thrace |
| EPI_ISL_437882 | Laboratory of Microbiology, Medical School, National and Kapodistrian University of Athens | Laboratory of Biology, Department of Medicine, Democritus University of Thrace |
| EPI_ISL_437883 | Laboratory of Microbiology, Medical School, National and Kapodistrian University of Athens | Laboratory of Biology, Department of Medicine, Democritus University of Thrace |
| EPI_ISL_437884 | Laboratory of Microbiology, Medical School, National and Kapodistrian University of Athens | Laboratory of Biology, Department of Medicine, Democritus University of Thrace |
| EPI_ISL_437885 | Laboratory of Microbiology, Medical School, National and Kapodistrian University of Athens | Laboratory of Biology, Department of Medicine, Democritus University of Thrace |
| EPI_ISL_437886 | Laboratory of Microbiology, Medical School, National and Kapodistrian University of Athens | Laboratory of Biology, Department of Medicine, Democritus University of Thrace |
| EPI_ISL_437887 | Laboratory of Microbiology, Medical School, National and Kapodistrian University of Athens | Laboratory of Biology, Department of Medicine, Democritus University of Thrace |
| EPI_ISL_437888 | Laboratory of Microbiology, Medical School, National and Kapodistrian University of Athens | Laboratory of Biology, Department of Medicine, Democritus University of Thrace |
| EPI_ISL_437889 | Laboratory of Microbiology, Medical School, National and Kapodistrian University of Athens | Laboratory of Biology, Department of Medicine, Democritus University of Thrace |
| EPI_ISL_437890 | Laboratory of Microbiology, Medical School, National and Kapodistrian University of Athens | Laboratory of Biology, Department of Medicine, Democritus University of Thrace |

| EPI_Accession_ID | Originating Laboratory | Submitting Laboratory |
|---|---|---|
| EPI_ISL_437891 | Laboratory of Microbiology, Medical School, National and Kapodistrian University of Athens | Laboratory of Biology, Department of Medicine, Democritus University of Thrace |
| EPI_ISL_437892 | Laboratory of Microbiology, Medical School, National and Kapodistrian University of Athens | Laboratory of Biology, Department of Medicine, Democritus University of Thrace |
| EPI_ISL_437893 | Laboratory of Microbiology, Medical School, National and Kapodistrian University of Athens | Laboratory of Biology, Department of Medicine, Democritus University of Thrace |
| EPI_ISL_437894 | Laboratory of Microbiology, Medical School, National and Kapodistrian University of Athens | Laboratory of Biology, Department of Medicine, Democritus University of Thrace |
| EPI_ISL_437895 | Laboratory of Microbiology, Medical School, National and Kapodistrian University of Athens | Laboratory of Biology, Department of Medicine, Democritus University of Thrace |
| EPI_ISL_437896 | Laboratory of Microbiology, Medical School, National and Kapodistrian University of Athens | Laboratory of Biology, Department of Medicine, Democritus University of Thrace |
| EPI_ISL_437897 | Laboratory of Microbiology, Medical School, National and Kapodistrian University of Athens | Laboratory of Biology, Department of Medicine, Democritus University of Thrace |
| EPI_ISL_437898 | Laboratory of Microbiology, Medical School, National and Kapodistrian University of Athens | Laboratory of Biology, Department of Medicine, Democritus University of Thrace |
| EPI_ISL_437899 | Laboratory of Microbiology, Medical School, National and Kapodistrian University of Athens | Laboratory of Biology, Department of Medicine, Democritus University of Thrace |
| EPI_ISL_437900 | Laboratory of Microbiology, Medical School, National and Kapodistrian University of Athens | Laboratory of Biology, Department of Medicine, Democritus University of Thrace |
| EPI_ISL_437901 | Laboratory of Microbiology, Medical School, National and Kapodistrian University of Athens | Laboratory of Biology, Department of Medicine, Democritus University of Thrace |
| EPI_ISL_437902 | Laboratory of Microbiology, Medical School, National and Kapodistrian University of Athens | Laboratory of Biology, Department of Medicine, Democritus University of Thrace |
| EPI_ISL_437903 | Laboratory of Microbiology, Medical School, National and Kapodistrian University of Athens | Laboratory of Biology, Department of Medicine, Democritus University of Thrace |
| EPI_ISL_437904 | Laboratory of Microbiology, Medical School, National and Kapodistrian University of Athens | Laboratory of Biology, Department of Medicine, Democritus University of Thrace |
| EPI_ISL_437905 | Laboratory of Microbiology, Medical School, National and Kapodistrian University of Athens | Laboratory of Biology, Department of Medicine, Democritus University of Thrace |
| EPI_ISL_437906 | Laboratory of Microbiology, Medical School, National and Kapodistrian University of Athens | Laboratory of Biology, Department of Medicine, Democritus University of Thrace |

| EPI_Accession_ID | Originating Laboratory | Submitting Laboratory |
|---|---|---|
| EPI_ISL_437907 | Laboratory of Microbiology, Medical School, National and Kapodistrian University of Athens | Laboratory of Biology, Department of Medicine, Democritus University of Thrace |
| EPI_ISL_437908 | Laboratory of Microbiology, Medical School, National and Kapodistrian University of Athens | Laboratory of Biology, Department of Medicine, Democritus University of Thrace |
| EPI_ISL_437909 | Laboratory of Microbiology, Medical School, National and Kapodistrian University of Athens | Laboratory of Biology, Department of Medicine, Democritus University of Thrace |
| EPI_ISL_437910 | Laboratory of Microbiology, Medical School, National and Kapodistrian University of Athens | Laboratory of Biology, Department of Medicine, Democritus University of Thrace |
| EPI_ISL_437911 | Laboratory of Microbiology, Medical School, National and Kapodistrian University of Athens | Laboratory of Biology, Department of Medicine, Democritus University of Thrace |
| EPI_ISL_437912 | Child Health Research Foundation | Child Health Research Lab |
| EPI_ISL_437913 | Institut fÃ¼r Virologie am Department fÃ¼r Hygiene, Mikrobiologie und Public Health | Bergthaler laboratory, CeMM Research Center for Molecular Medicine of the Austrian Academy of Sciences |
| EPI_ISL_437914 | Institut fÃ¼r Virologie am Department fÃ¼r Hygiene, Mikrobiologie und Public Health | Bergthaler laboratory, CeMM Research Center for Molecular Medicine of the Austrian Academy of Sciences |
| EPI_ISL_437915 | Institut fÃ¼r Virologie am Department fÃ¼r Hygiene, Mikrobiologie und Public Health | Bergthaler laboratory, CeMM Research Center for Molecular Medicine of the Austrian Academy of Sciences |
| EPI_ISL_437916 | Institut fÃ¼r Virologie am Department fÃ¼r Hygiene, Mikrobiologie und Public Health | Bergthaler laboratory, CeMM Research Center for Molecular Medicine of the Austrian Academy of Sciences |
| EPI_ISL_437917 | Institut fÃ¼r Virologie am Department fÃ¼r Hygiene, Mikrobiologie und Public Health | Bergthaler laboratory, CeMM Research Center for Molecular Medicine of the Austrian Academy of Sciences |
| EPI_ISL_437918 | Institut fÃ¼r Virologie am Department fÃ¼r Hygiene, Mikrobiologie und Public Health | Bergthaler laboratory, CeMM Research Center for Molecular Medicine of the Austrian Academy of Sciences |
| EPI_ISL_437919 | Institut fÃ¼r Virologie am Department fÃ¼r Hygiene, Mikrobiologie und Public Health | Bergthaler laboratory, CeMM Research Center for Molecular Medicine of the Austrian Academy of Sciences |

| EPI_Accession_ID | Originating Laboratory | Submitting Laboratory |
|---|---|---|
| EPI_ISL_437920 | Institut fÃ¼r Virologie am Department fÃ¼r Hygiene, Mikrobiologie und Public Health | Bergthaler laboratory, CeMM Research Center for Molecular Medicine of the Austrian Academy of Sciences |
| EPI_ISL_437921 | Institut fÃ¼r Virologie am Department fÃ¼r Hygiene, Mikrobiologie und Public Health | Bergthaler laboratory, CeMM Research Center for Molecular Medicine of the Austrian Academy of Sciences |
| EPI_ISL_437922 | Institut fÃ¼r Virologie am Department fÃ¼r Hygiene, Mikrobiologie und Public Health | Bergthaler laboratory, CeMM Research Center for Molecular Medicine of the Austrian Academy of Sciences |
| EPI_ISL_437923 | Institut fÃ¼r Virologie am Department fÃ¼r Hygiene, Mikrobiologie und Public Health | Bergthaler laboratory, CeMM Research Center for Molecular Medicine of the Austrian Academy of Sciences |
| EPI_ISL_437924 | Institut fÃ¼r Virologie am Department fÃ¼r Hygiene, Mikrobiologie und Public Health | Bergthaler laboratory, CeMM Research Center for Molecular Medicine of the Austrian Academy of Sciences |
| EPI_ISL_437925 | Institut fÃ¼r Virologie am Department fÃ¼r Hygiene, Mikrobiologie und Public Health | Bergthaler laboratory, CeMM Research Center for Molecular Medicine of the Austrian Academy of Sciences |
| EPI_ISL_437926 | Institut fÃ¼r Virologie am Department fÃ¼r Hygiene, Mikrobiologie und Public Health | Bergthaler laboratory, CeMM Research Center for Molecular Medicine of the Austrian Academy of Sciences |
| EPI_ISL_437927 | Institut fÃ¼r Virologie am Department fÃ¼r Hygiene, Mikrobiologie und Public Health | Bergthaler laboratory, CeMM Research Center for Molecular Medicine of the Austrian Academy of Sciences |
| EPI_ISL_437928 | Institut fÃ¼r Virologie am Department fÃ¼r Hygiene, Mikrobiologie und Public Health | Bergthaler laboratory, CeMM Research Center for Molecular Medicine of the Austrian Academy of Sciences |
| EPI_ISL_437929 | Institut fÃ¼r Virologie am Department fÃ¼r Hygiene, Mikrobiologie und Public Health | Bergthaler laboratory, CeMM Research Center for Molecular Medicine of the Austrian Academy of Sciences |
| EPI_ISL_437930 | Institut fÃ¼r Virologie am Department fÃ¼r Hygiene, Mikrobiologie und Public Health | Bergthaler laboratory, CeMM Research Center for Molecular Medicine of the Austrian Academy of Sciences |

| EPI_Accession_ID | Originating Laboratory | Submitting Laboratory |
|---|---|---|
| EPI_ISL_437931 | Institut fÃ¼r Virologie am Department fÃ¼r Hygiene, Mikrobiologie und Public Health | Bergthaler laboratory, CeMM Research Center for Molecular Medicine of the Austrian Academy of Sciences |
| EPI_ISL_437932 | Institut fÃ¼r Virologie am Department fÃ¼r Hygiene, Mikrobiologie und Public Health | Bergthaler laboratory, CeMM Research Center for Molecular Medicine of the Austrian Academy of Sciences |
| EPI_ISL_437933 | Universitaetsklinik fÃ¼r Innere Medizin II Innsbruck | Bergthaler laboratory, CeMM Research Center for Molecular Medicine of the Austrian Academy of Sciences |
| EPI_ISL_437934 | Universitaetsklinik fÃ¼r Innere Medizin II Innsbruck | Bergthaler laboratory, CeMM Research Center for Molecular Medicine of the Austrian Academy of Sciences |
| EPI_ISL_437935 | Universitaetsklinik fÃ¼r Innere Medizin II Innsbruck | Bergthaler laboratory, CeMM Research Center for Molecular Medicine of the Austrian Academy of Sciences |
| EPI_ISL_437936 | Universitaetsklinik fÃ¼r Innere Medizin II Innsbruck | Bergthaler laboratory, CeMM Research Center for Molecular Medicine of the Austrian Academy of Sciences |
| EPI_ISL_437937 | Universitaetsklinik fÃ¼r Innere Medizin II Innsbruck | Bergthaler laboratory, CeMM Research Center for Molecular Medicine of the Austrian Academy of Sciences |
| EPI_ISL_437938 | Universitaetsklinik fÃ¼r Innere Medizin II Innsbruck | Bergthaler laboratory, CeMM Research Center for Molecular Medicine of the Austrian Academy of Sciences |
| EPI_ISL_437939 | Universitaetsklinik fÃ¼r Innere Medizin II Innsbruck | Bergthaler laboratory, CeMM Research Center for Molecular Medicine of the Austrian Academy of Sciences |
| EPI_ISL_437940 | Universitaetsklinik fÃ¼r Innere Medizin II Innsbruck | Bergthaler laboratory, CeMM Research Center for Molecular Medicine of the Austrian Academy of Sciences |
| EPI_ISL_437941 | Universitaetsklinik fÃ¼r Innere Medizin II Innsbruck | Bergthaler laboratory, CeMM Research Center for Molecular Medicine of the Austrian Academy of Sciences |

| EPI_Accession_ID | Originating Laboratory | Submitting Laboratory |
| --- | --- | --- |
| EPI_ISL_437942 | Universitaetsklinik fÃ¼r Innere Medizin II Innsbruck | Bergthaler laboratory, CeMM Research Center for Molecular Medicine of the Austrian Academy of Sciences |
| EPI_ISL_437943 | Universitaetsklinik fÃ¼r Innere Medizin II Innsbruck | Bergthaler laboratory, CeMM Research Center for Molecular Medicine of the Austrian Academy of Sciences |
| EPI_ISL_437944 | Universitaetsklinik fÃ¼r Innere Medizin II Innsbruck | Bergthaler laboratory, CeMM Research Center for Molecular Medicine of the Austrian Academy of Sciences |
| EPI_ISL_437945 | Universitaetsklinik fÃ¼r Innere Medizin II Innsbruck | Bergthaler laboratory, CeMM Research Center for Molecular Medicine of the Austrian Academy of Sciences |
| EPI_ISL_437946 | Universitaetsklinik fÃ¼r Innere Medizin II Innsbruck | Bergthaler laboratory, CeMM Research Center for Molecular Medicine of the Austrian Academy of Sciences |
| EPI_ISL_437947 | Universitaetsklinik fÃ¼r Innere Medizin II Innsbruck | Bergthaler laboratory, CeMM Research Center for Molecular Medicine of the Austrian Academy of Sciences |
| EPI_ISL_437948 | Universitaetsklinik fÃ¼r Innere Medizin II Innsbruck | Bergthaler laboratory, CeMM Research Center for Molecular Medicine of the Austrian Academy of Sciences |
| EPI_ISL_437949 | Universitaetsklinik fÃ¼r Innere Medizin II Innsbruck | Bergthaler laboratory, CeMM Research Center for Molecular Medicine of the Austrian Academy of Sciences |
| EPI_ISL_437950 | Universitaetsklinik fÃ¼r Innere Medizin II Innsbruck | Bergthaler laboratory, CeMM Research Center for Molecular Medicine of the Austrian Academy of Sciences |
| EPI_ISL_437951 | Universitaetsklinik fÃ¼r Innere Medizin II Innsbruck | Bergthaler laboratory, CeMM Research Center for Molecular Medicine of the Austrian Academy of Sciences |
| EPI_ISL_437952 | Universitaetsklinik fÃ¼r Innere Medizin II Innsbruck | Bergthaler laboratory, CeMM Research Center for Molecular Medicine of the Austrian Academy of Sciences |

| EPI_Accession_ID | Originating Laboratory | Submitting Laboratory |
|---|---|---|
| EPI_ISL_437953 | Universitaetsklinik fÃ¼r Innere Medizin II Innsbruck | Bergthaler laboratory, CeMM Research Center for Molecular Medicine of the Austrian Academy of Sciences |
| EPI_ISL_437954 | Universitaetsklinik fÃ¼r Innere Medizin II Innsbruck | Bergthaler laboratory, CeMM Research Center for Molecular Medicine of the Austrian Academy of Sciences |
| EPI_ISL_437955 | Universitaetsklinik fÃ¼r Innere Medizin II Innsbruck | Bergthaler laboratory, CeMM Research Center for Molecular Medicine of the Austrian Academy of Sciences |
| EPI_ISL_437956 | Universitaetsklinik fÃ¼r Innere Medizin II Innsbruck | Bergthaler laboratory, CeMM Research Center for Molecular Medicine of the Austrian Academy of Sciences |
| EPI_ISL_437957 | Universitaetsklinik fÃ¼r Innere Medizin II Innsbruck | Bergthaler laboratory, CeMM Research Center for Molecular Medicine of the Austrian Academy of Sciences |
| EPI_ISL_437958 | Universitaetsklinik fÃ¼r Innere Medizin II Innsbruck | Bergthaler laboratory, CeMM Research Center for Molecular Medicine of the Austrian Academy of Sciences |
| EPI_ISL_437959 | Universitaetsklinik fÃ¼r Innere Medizin II Innsbruck | Bergthaler laboratory, CeMM Research Center for Molecular Medicine of the Austrian Academy of Sciences |
| EPI_ISL_437960 | Universitaetsklinik fÃ¼r Innere Medizin II Innsbruck | Bergthaler laboratory, CeMM Research Center for Molecular Medicine of the Austrian Academy of Sciences |
| EPI_ISL_437961 | Universitaetsklinik fÃ¼r Innere Medizin II Innsbruck | Bergthaler laboratory, CeMM Research Center for Molecular Medicine of the Austrian Academy of Sciences |
| EPI_ISL_437962 | Universitaetsklinik fÃ¼r Innere Medizin II Innsbruck | Bergthaler laboratory, CeMM Research Center for Molecular Medicine of the Austrian Academy of Sciences |
| EPI_ISL_437963 | Universitaetsklinik fÃ¼r Innere Medizin II Innsbruck | Bergthaler laboratory, CeMM Research Center for Molecular Medicine of the Austrian Academy of Sciences |

| EPI_Accession_ID | Originating Laboratory | Submitting Laboratory |
|---|---|---|
| EPI_ISL_437964 | Universitaetsklinik fÃ¼r Innere Medizin II Innsbruck | Bergthaler laboratory, CeMM Research Center for Molecular Medicine of the Austrian Academy of Sciences |
| EPI_ISL_437965 | Universitaetsklinik fÃ¼r Innere Medizin II Innsbruck | Bergthaler laboratory, CeMM Research Center for Molecular Medicine of the Austrian Academy of Sciences |
| EPI_ISL_437966 | Universitaetsklinik fÃ¼r Innere Medizin II Innsbruck | Bergthaler laboratory, CeMM Research Center for Molecular Medicine of the Austrian Academy of Sciences |
| EPI_ISL_437967 | Universitaetsklinik fÃ¼r Innere Medizin II Innsbruck | Bergthaler laboratory, CeMM Research Center for Molecular Medicine of the Austrian Academy of Sciences |
| EPI_ISL_437968 | Universitaetsklinik fÃ¼r Innere Medizin II Innsbruck | Bergthaler laboratory, CeMM Research Center for Molecular Medicine of the Austrian Academy of Sciences |
| EPI_ISL_437969 | Universitaetsklinik fÃ¼r Innere Medizin II Innsbruck | Bergthaler laboratory, CeMM Research Center for Molecular Medicine of the Austrian Academy of Sciences |
| EPI_ISL_437970 | Universitaetsklinik fÃ¼r Innere Medizin II Innsbruck | Bergthaler laboratory, CeMM Research Center for Molecular Medicine of the Austrian Academy of Sciences |
| EPI_ISL_437971 | Universitaetsklinik fÃ¼r Innere Medizin II Innsbruck | Bergthaler laboratory, CeMM Research Center for Molecular Medicine of the Austrian Academy of Sciences |
| EPI_ISL_437972 | Universitaetsklinik fÃ¼r Innere Medizin II Innsbruck | Bergthaler laboratory, CeMM Research Center for Molecular Medicine of the Austrian Academy of Sciences |
| EPI_ISL_437973 | Universitaetsklinik fÃ¼r Innere Medizin II Innsbruck | Bergthaler laboratory, CeMM Research Center for Molecular Medicine of the Austrian Academy of Sciences |
| EPI_ISL_437974 | Institut fÃ¼r Virologie am Department fÃ¼r Hygiene, Mikrobiologie und Public Health | Bergthaler laboratory, CeMM Research Center for Molecular Medicine of the Austrian Academy of Sciences |

| EPI_Accession_ID | Originating Laboratory | Submitting Laboratory |
|---|---|---|
| EPI_ISL_437975 | Institut fÃ¼r Virologie am Department fÃ¼r Hygiene, Mikrobiologie und Public Health | Bergthaler laboratory, CeMM Research Center for Molecular Medicine of the Austrian Academy of Sciences |
| EPI_ISL_437976 | Institut fÃ¼r Virologie am Department fÃ¼r Hygiene, Mikrobiologie und Public Health | Bergthaler laboratory, CeMM Research Center for Molecular Medicine of the Austrian Academy of Sciences |
| EPI_ISL_437977 | Institut fÃ¼r Virologie am Department fÃ¼r Hygiene, Mikrobiologie und Public Health | Bergthaler laboratory, CeMM Research Center for Molecular Medicine of the Austrian Academy of Sciences |
| EPI_ISL_437978 | Institut fÃ¼r Virologie am Department fÃ¼r Hygiene, Mikrobiologie und Public Health | Bergthaler laboratory, CeMM Research Center for Molecular Medicine of the Austrian Academy of Sciences |
| EPI_ISL_437979 | Institut fÃ¼r Virologie am Department fÃ¼r Hygiene, Mikrobiologie und Public Health | Bergthaler laboratory, CeMM Research Center for Molecular Medicine of the Austrian Academy of Sciences |
| EPI_ISL_437980 | Institut fÃ¼r Virologie am Department fÃ¼r Hygiene, Mikrobiologie und Public Health | Bergthaler laboratory, CeMM Research Center for Molecular Medicine of the Austrian Academy of Sciences |
| EPI_ISL_437981 | Institut fÃ¼r Virologie am Department fÃ¼r Hygiene, Mikrobiologie und Public Health | Bergthaler laboratory, CeMM Research Center for Molecular Medicine of the Austrian Academy of Sciences |
| EPI_ISL_437982 | Institut fÃ¼r Virologie am Department fÃ¼r Hygiene, Mikrobiologie und Public Health | Bergthaler laboratory, CeMM Research Center for Molecular Medicine of the Austrian Academy of Sciences |
| EPI_ISL_437983 | Institut fÃ¼r Virologie am Department fÃ¼r Hygiene, Mikrobiologie und Public Health | Bergthaler laboratory, CeMM Research Center for Molecular Medicine of the Austrian Academy of Sciences |
| EPI_ISL_437984 | Institut fÃ¼r Virologie am Department fÃ¼r Hygiene, Mikrobiologie und Public Health | Bergthaler laboratory, CeMM Research Center for Molecular Medicine of the Austrian Academy of Sciences |
| EPI_ISL_437985 | Institut fÃ¼r Virologie am Department fÃ¼r Hygiene, Mikrobiologie und Public Health | Bergthaler laboratory, CeMM Research Center for Molecular Medicine of the Austrian Academy of Sciences |

| EPI_Accession_ID | Originating Laboratory | Submitting Laboratory |
|---|---|---|
| EPI_ISL_437986 | Institut fÃ¼r Virologie am Department fÃ¼r Hygiene, Mikrobiologie und Public Health | Bergthaler laboratory, CeMM Research Center for Molecular Medicine of the Austrian Academy of Sciences |
| EPI_ISL_437987 | Institut fÃ¼r Virologie am Department fÃ¼r Hygiene, Mikrobiologie und Public Health | Bergthaler laboratory, CeMM Research Center for Molecular Medicine of the Austrian Academy of Sciences |
| EPI_ISL_437988 | Institut fÃ¼r Virologie am Department fÃ¼r Hygiene, Mikrobiologie und Public Health | Bergthaler laboratory, CeMM Research Center for Molecular Medicine of the Austrian Academy of Sciences |
| EPI_ISL_437989 | Institut fÃ¼r Virologie am Department fÃ¼r Hygiene, Mikrobiologie und Public Health | Bergthaler laboratory, CeMM Research Center for Molecular Medicine of the Austrian Academy of Sciences |
| EPI_ISL_437990 | Institut fÃ¼r Virologie am Department fÃ¼r Hygiene, Mikrobiologie und Public Health | Bergthaler laboratory, CeMM Research Center for Molecular Medicine of the Austrian Academy of Sciences |
| EPI_ISL_437991 | Institut fÃ¼r Virologie am Department fÃ¼r Hygiene, Mikrobiologie und Public Health | Bergthaler laboratory, CeMM Research Center for Molecular Medicine of the Austrian Academy of Sciences |
| EPI_ISL_437992 | Institut fÃ¼r Virologie am Department fÃ¼r Hygiene, Mikrobiologie und Public Health | Bergthaler laboratory, CeMM Research Center for Molecular Medicine of the Austrian Academy of Sciences |
| EPI_ISL_437993 | Center for Virology, Medical University of Vienna | Bergthaler laboratory, CeMM Research Center for Molecular Medicine of the Austrian Academy of Sciences |
| EPI_ISL_437994 | Center for Virology, Medical University of Vienna | Bergthaler laboratory, CeMM Research Center for Molecular Medicine of the Austrian Academy of Sciences |
| EPI_ISL_437995 | Center for Virology, Medical University of Vienna | Bergthaler laboratory, CeMM Research Center for Molecular Medicine of the Austrian Academy of Sciences |
| EPI_ISL_437996 | Center for Virology, Medical University of Vienna | Bergthaler laboratory, CeMM Research Center for Molecular Medicine of the Austrian Academy of Sciences |

| EPI_Accession_ID | Originating Laboratory | Submitting Laboratory |
|---|---|---|
| EPI_ISL_437997 | Center for Virology, Medical University of Vienna | Bergthaler laboratory, CeMM Research Center for Molecular Medicine of the Austrian Academy of Sciences |
| EPI_ISL_437998 | Center for Virology, Medical University of Vienna | Bergthaler laboratory, CeMM Research Center for Molecular Medicine of the Austrian Academy of Sciences |
| EPI_ISL_437999 | Center for Virology, Medical University of Vienna | Bergthaler laboratory, CeMM Research Center for Molecular Medicine of the Austrian Academy of Sciences |
| EPI_ISL_438000 | Center for Virology, Medical University of Vienna | Bergthaler laboratory, CeMM Research Center for Molecular Medicine of the Austrian Academy of Sciences |
| EPI_ISL_438001 | Center for Virology, Medical University of Vienna | Bergthaler laboratory, CeMM Research Center for Molecular Medicine of the Austrian Academy of Sciences |
| EPI_ISL_438002 | Center for Virology, Medical University of Vienna | Bergthaler laboratory, CeMM Research Center for Molecular Medicine of the Austrian Academy of Sciences |
| EPI_ISL_438003 | Center for Virology, Medical University of Vienna | Bergthaler laboratory, CeMM Research Center for Molecular Medicine of the Austrian Academy of Sciences |
| EPI_ISL_438004 | Center for Virology, Medical University of Vienna | Bergthaler laboratory, CeMM Research Center for Molecular Medicine of the Austrian Academy of Sciences |
| EPI_ISL_438005 | Center for Virology, Medical University of Vienna | Bergthaler laboratory, CeMM Research Center for Molecular Medicine of the Austrian Academy of Sciences |
| EPI_ISL_438006 | Center for Virology, Medical University of Vienna | Bergthaler laboratory, CeMM Research Center for Molecular Medicine of the Austrian Academy of Sciences |
| EPI_ISL_438007 | Center for Virology, Medical University of Vienna | Bergthaler laboratory, CeMM Research Center for Molecular Medicine of the Austrian Academy of Sciences |

| EPI_Accession_ID | Originating Laboratory | Submitting Laboratory |
|---|---|---|
| EPI_ISL_438008 | Center for Virology, Medical University of Vienna | Bergthaler laboratory, CeMM Research Center for Molecular Medicine of the Austrian Academy of Sciences |
| EPI_ISL_438009 | Center for Virology, Medical University of Vienna | Bergthaler laboratory, CeMM Research Center for Molecular Medicine of the Austrian Academy of Sciences |
| EPI_ISL_438010 | Center for Virology, Medical University of Vienna | Bergthaler laboratory, CeMM Research Center for Molecular Medicine of the Austrian Academy of Sciences |
| EPI_ISL_438011 | Center for Virology, Medical University of Vienna | Bergthaler laboratory, CeMM Research Center for Molecular Medicine of the Austrian Academy of Sciences |
| EPI_ISL_438012 | Center for Virology, Medical University of Vienna | Bergthaler laboratory, CeMM Research Center for Molecular Medicine of the Austrian Academy of Sciences |
| EPI_ISL_438013 | Center for Virology, Medical University of Vienna | Bergthaler laboratory, CeMM Research Center for Molecular Medicine of the Austrian Academy of Sciences |
| EPI_ISL_438014 | Center for Virology, Medical University of Vienna | Bergthaler laboratory, CeMM Research Center for Molecular Medicine of the Austrian Academy of Sciences |
| EPI_ISL_438015 | Center for Virology, Medical University of Vienna | Bergthaler laboratory, CeMM Research Center for Molecular Medicine of the Austrian Academy of Sciences |
| EPI_ISL_438016 | Center for Virology, Medical University of Vienna | Bergthaler laboratory, CeMM Research Center for Molecular Medicine of the Austrian Academy of Sciences |
| EPI_ISL_438017 | Center for Virology, Medical University of Vienna | Bergthaler laboratory, CeMM Research Center for Molecular Medicine of the Austrian Academy of Sciences |
| EPI_ISL_438018 | Center for Virology, Medical University of Vienna | Bergthaler laboratory, CeMM Research Center for Molecular Medicine of the Austrian Academy of Sciences |

| EPI_Accession_ID | Originating Laboratory | Submitting Laboratory |
|---|---|---|
| EPI_ISL_438019 | Center for Virology, Medical University of Vienna | Bergthaler laboratory, CeMM Research Center for Molecular Medicine of the Austrian Academy of Sciences |
| EPI_ISL_438020 | Center for Virology, Medical University of Vienna | Bergthaler laboratory, CeMM Research Center for Molecular Medicine of the Austrian Academy of Sciences |
| EPI_ISL_438021 | Center for Virology, Medical University of Vienna | Bergthaler laboratory, CeMM Research Center for Molecular Medicine of the Austrian Academy of Sciences |
| EPI_ISL_438022 | Center for Virology, Medical University of Vienna | Bergthaler laboratory, CeMM Research Center for Molecular Medicine of the Austrian Academy of Sciences |
| EPI_ISL_438023 | Center for Virology, Medical University of Vienna | Bergthaler laboratory, CeMM Research Center for Molecular Medicine of the Austrian Academy of Sciences |
| EPI_ISL_438024 | Center for Virology, Medical University of Vienna | Bergthaler laboratory, CeMM Research Center for Molecular Medicine of the Austrian Academy of Sciences |
| EPI_ISL_438025 | Center for Virology, Medical University of Vienna | Bergthaler laboratory, CeMM Research Center for Molecular Medicine of the Austrian Academy of Sciences |
| EPI_ISL_438026 | Center for Virology, Medical University of Vienna | Bergthaler laboratory, CeMM Research Center for Molecular Medicine of the Austrian Academy of Sciences |
| EPI_ISL_438027 | Center for Virology, Medical University of Vienna | Bergthaler laboratory, CeMM Research Center for Molecular Medicine of the Austrian Academy of Sciences |
| EPI_ISL_438028 | Center for Virology, Medical University of Vienna | Bergthaler laboratory, CeMM Research Center for Molecular Medicine of the Austrian Academy of Sciences |
| EPI_ISL_438029 | Center for Virology, Medical University of Vienna | Bergthaler laboratory, CeMM Research Center for Molecular Medicine of the Austrian Academy of Sciences |

| EPI_Accession_ID | Originating Laboratory | Submitting Laboratory |
|---|---|---|
| EPI_ISL_438030 | Center for Virology, Medical University of Vienna | Bergthaler laboratory, CeMM Research Center for Molecular Medicine of the Austrian Academy of Sciences |
| EPI_ISL_438031 | Center for Virology, Medical University of Vienna | Bergthaler laboratory, CeMM Research Center for Molecular Medicine of the Austrian Academy of Sciences |
| EPI_ISL_438032 | Center for Virology, Medical University of Vienna | Bergthaler laboratory, CeMM Research Center for Molecular Medicine of the Austrian Academy of Sciences |
| EPI_ISL_438033 | Center for Virology, Medical University of Vienna | Bergthaler laboratory, CeMM Research Center for Molecular Medicine of the Austrian Academy of Sciences |
| EPI_ISL_438034 | Center for Virology, Medical University of Vienna | Bergthaler laboratory, CeMM Research Center for Molecular Medicine of the Austrian Academy of Sciences |
| EPI_ISL_438035 | Center for Virology, Medical University of Vienna | Bergthaler laboratory, CeMM Research Center for Molecular Medicine of the Austrian Academy of Sciences |
| EPI_ISL_438036 | Center for Virology, Medical University of Vienna | Bergthaler laboratory, CeMM Research Center for Molecular Medicine of the Austrian Academy of Sciences |
| EPI_ISL_438037 | Center for Virology, Medical University of Vienna | Bergthaler laboratory, CeMM Research Center for Molecular Medicine of the Austrian Academy of Sciences |
| EPI_ISL_438038 | Center for Virology, Medical University of Vienna | Bergthaler laboratory, CeMM Research Center for Molecular Medicine of the Austrian Academy of Sciences |
| EPI_ISL_438039 | Center for Virology, Medical University of Vienna | Bergthaler laboratory, CeMM Research Center for Molecular Medicine of the Austrian Academy of Sciences |
| EPI_ISL_438040 | Center for Virology, Medical University of Vienna | Bergthaler laboratory, CeMM Research Center for Molecular Medicine of the Austrian Academy of Sciences |

| EPI_Accession_ID | Originating Laboratory | Submitting Laboratory |
| --- | --- | --- |
| EPI_ISL_438041 | Center for Virology, Medical University of Vienna | Bergthaler laboratory, CeMM Research Center for Molecular Medicine of the Austrian Academy of Sciences |
| EPI_ISL_438042 | Center for Virology, Medical University of Vienna | Bergthaler laboratory, CeMM Research Center for Molecular Medicine of the Austrian Academy of Sciences |
| EPI_ISL_438043 | Center for Virology, Medical University of Vienna | Bergthaler laboratory, CeMM Research Center for Molecular Medicine of the Austrian Academy of Sciences |
| EPI_ISL_438044 | Center for Virology, Medical University of Vienna | Bergthaler laboratory, CeMM Research Center for Molecular Medicine of the Austrian Academy of Sciences |
| EPI_ISL_438045 | Center for Virology, Medical University of Vienna | Bergthaler laboratory, CeMM Research Center for Molecular Medicine of the Austrian Academy of Sciences |
| EPI_ISL_438046 | Center for Virology, Medical University of Vienna | Bergthaler laboratory, CeMM Research Center for Molecular Medicine of the Austrian Academy of Sciences |
| EPI_ISL_438047 | Center for Virology, Medical University of Vienna | Bergthaler laboratory, CeMM Research Center for Molecular Medicine of the Austrian Academy of Sciences |
| EPI_ISL_438048 | Center for Virology, Medical University of Vienna | Bergthaler laboratory, CeMM Research Center for Molecular Medicine of the Austrian Academy of Sciences |
| EPI_ISL_438049 | Center for Virology, Medical University of Vienna | Bergthaler laboratory, CeMM Research Center for Molecular Medicine of the Austrian Academy of Sciences |
| EPI_ISL_438050 | Center for Virology, Medical University of Vienna | Bergthaler laboratory, CeMM Research Center for Molecular Medicine of the Austrian Academy of Sciences |
| EPI_ISL_438051 | Center for Virology, Medical University of Vienna | Bergthaler laboratory, CeMM Research Center for Molecular Medicine of the Austrian Academy of Sciences |

| EPI_Accession_ID | Originating Laboratory | Submitting Laboratory |
|---|---|---|
| EPI_ISL_438052 | Center for Virology, Medical University of Vienna | Bergthaler laboratory, CeMM Research Center for Molecular Medicine of the Austrian Academy of Sciences |
| EPI_ISL_438053 | Center for Virology, Medical University of Vienna | Bergthaler laboratory, CeMM Research Center for Molecular Medicine of the Austrian Academy of Sciences |
| EPI_ISL_438054 | Center for Virology, Medical University of Vienna | Bergthaler laboratory, CeMM Research Center for Molecular Medicine of the Austrian Academy of Sciences |
| EPI_ISL_438055 | Center for Virology, Medical University of Vienna | Bergthaler laboratory, CeMM Research Center for Molecular Medicine of the Austrian Academy of Sciences |
| EPI_ISL_438056 | Center for Virology, Medical University of Vienna | Bergthaler laboratory, CeMM Research Center for Molecular Medicine of the Austrian Academy of Sciences |
| EPI_ISL_438057 | Center for Virology, Medical University of Vienna | Bergthaler laboratory, CeMM Research Center for Molecular Medicine of the Austrian Academy of Sciences |
| EPI_ISL_438058 | Center for Virology, Medical University of Vienna | Bergthaler laboratory, CeMM Research Center for Molecular Medicine of the Austrian Academy of Sciences |
| EPI_ISL_438059 | Center for Virology, Medical University of Vienna | Bergthaler laboratory, CeMM Research Center for Molecular Medicine of the Austrian Academy of Sciences |
| EPI_ISL_438060 | Center for Virology, Medical University of Vienna | Bergthaler laboratory, CeMM Research Center for Molecular Medicine of the Austrian Academy of Sciences |
| EPI_ISL_438061 | Center for Virology, Medical University of Vienna | Bergthaler laboratory, CeMM Research Center for Molecular Medicine of the Austrian Academy of Sciences |
| EPI_ISL_438062 | Center for Virology, Medical University of Vienna | Bergthaler laboratory, CeMM Research Center for Molecular Medicine of the Austrian Academy of Sciences |

| EPI_Accession_ID | Originating Laboratory | Submitting Laboratory |
|---|---|---|
| EPI_ISL_438063 | Center for Virology, Medical University of Vienna | Bergthaler laboratory, CeMM Research Center for Molecular Medicine of the Austrian Academy of Sciences |
| EPI_ISL_438064 | Center for Virology, Medical University of Vienna | Bergthaler laboratory, CeMM Research Center for Molecular Medicine of the Austrian Academy of Sciences |
| EPI_ISL_438065 | Center for Virology, Medical University of Vienna | Bergthaler laboratory, CeMM Research Center for Molecular Medicine of the Austrian Academy of Sciences |
| EPI_ISL_438066 | Center for Virology, Medical University of Vienna | Bergthaler laboratory, CeMM Research Center for Molecular Medicine of the Austrian Academy of Sciences |
| EPI_ISL_438067 | Center for Virology, Medical University of Vienna | Bergthaler laboratory, CeMM Research Center for Molecular Medicine of the Austrian Academy of Sciences |
| EPI_ISL_438068 | Center for Virology, Medical University of Vienna | Bergthaler laboratory, CeMM Research Center for Molecular Medicine of the Austrian Academy of Sciences |
| EPI_ISL_438069 | Center for Virology, Medical University of Vienna | Bergthaler laboratory, CeMM Research Center for Molecular Medicine of the Austrian Academy of Sciences |
| EPI_ISL_438070 | Center for Virology, Medical University of Vienna | Bergthaler laboratory, CeMM Research Center for Molecular Medicine of the Austrian Academy of Sciences |
| EPI_ISL_438071 | Center for Virology, Medical University of Vienna | Bergthaler laboratory, CeMM Research Center for Molecular Medicine of the Austrian Academy of Sciences |
| EPI_ISL_438072 | Center for Virology, Medical University of Vienna | Bergthaler laboratory, CeMM Research Center for Molecular Medicine of the Austrian Academy of Sciences |
| EPI_ISL_438073 | Center for Virology, Medical University of Vienna | Bergthaler laboratory, CeMM Research Center for Molecular Medicine of the Austrian Academy of Sciences |

| EPI_Accession_ID | Originating Laboratory | Submitting Laboratory |
|---|---|---|
| EPI_ISL_438074 | Center for Virology, Medical University of Vienna | Bergthaler laboratory, CeMM Research Center for Molecular Medicine of the Austrian Academy of Sciences |
| EPI_ISL_438075 | Center for Virology, Medical University of Vienna | Bergthaler laboratory, CeMM Research Center for Molecular Medicine of the Austrian Academy of Sciences |
| EPI_ISL_438076 | Center for Virology, Medical University of Vienna | Bergthaler laboratory, CeMM Research Center for Molecular Medicine of the Austrian Academy of Sciences |
| EPI_ISL_438077 | Center for Virology, Medical University of Vienna | Bergthaler laboratory, CeMM Research Center for Molecular Medicine of the Austrian Academy of Sciences |
| EPI_ISL_438078 | Center for Virology, Medical University of Vienna | Bergthaler laboratory, CeMM Research Center for Molecular Medicine of the Austrian Academy of Sciences |
| EPI_ISL_438079 | Center for Virology, Medical University of Vienna | Bergthaler laboratory, CeMM Research Center for Molecular Medicine of the Austrian Academy of Sciences |
| EPI_ISL_438080 | Center for Virology, Medical University of Vienna | Bergthaler laboratory, CeMM Research Center for Molecular Medicine of the Austrian Academy of Sciences |
| EPI_ISL_438081 | Center for Virology, Medical University of Vienna | Bergthaler laboratory, CeMM Research Center for Molecular Medicine of the Austrian Academy of Sciences |
| EPI_ISL_438082 | Center for Virology, Medical University of Vienna | Bergthaler laboratory, CeMM Research Center for Molecular Medicine of the Austrian Academy of Sciences |
| EPI_ISL_438083 | Center for Virology, Medical University of Vienna | Bergthaler laboratory, CeMM Research Center for Molecular Medicine of the Austrian Academy of Sciences |
| EPI_ISL_438084 | Center for Virology, Medical University of Vienna | Bergthaler laboratory, CeMM Research Center for Molecular Medicine of the Austrian Academy of Sciences |

| EPI_Accession_ID | Originating Laboratory | Submitting Laboratory |
|---|---|---|
| EPI_ISL_438085 | Center for Virology, Medical University of Vienna | Bergthaler laboratory, CeMM Research Center for Molecular Medicine of the Austrian Academy of Sciences |
| EPI_ISL_438086 | Center for Virology, Medical University of Vienna | Bergthaler laboratory, CeMM Research Center for Molecular Medicine of the Austrian Academy of Sciences |
| EPI_ISL_438087 | Center for Virology, Medical University of Vienna | Bergthaler laboratory, CeMM Research Center for Molecular Medicine of the Austrian Academy of Sciences |
| EPI_ISL_438088 | Center for Virology, Medical University of Vienna | Bergthaler laboratory, CeMM Research Center for Molecular Medicine of the Austrian Academy of Sciences |
| EPI_ISL_438089 | Center for Virology, Medical University of Vienna | Bergthaler laboratory, CeMM Research Center for Molecular Medicine of the Austrian Academy of Sciences |
| EPI_ISL_438090 | Center for Virology, Medical University of Vienna | Bergthaler laboratory, CeMM Research Center for Molecular Medicine of the Austrian Academy of Sciences |
| EPI_ISL_438091 | Center for Virology, Medical University of Vienna | Bergthaler laboratory, CeMM Research Center for Molecular Medicine of the Austrian Academy of Sciences |
| EPI_ISL_438092 | Center for Virology, Medical University of Vienna | Bergthaler laboratory, CeMM Research Center for Molecular Medicine of the Austrian Academy of Sciences |
| EPI_ISL_438093 | Center for Virology, Medical University of Vienna | Bergthaler laboratory, CeMM Research Center for Molecular Medicine of the Austrian Academy of Sciences |
| EPI_ISL_438094 | Center for Virology, Medical University of Vienna | Bergthaler laboratory, CeMM Research Center for Molecular Medicine of the Austrian Academy of Sciences |
| EPI_ISL_438095 | Center for Virology, Medical University of Vienna | Bergthaler laboratory, CeMM Research Center for Molecular Medicine of the Austrian Academy of Sciences |

| EPI_Accession_ID | Originating Laboratory | Submitting Laboratory |
|---|---|---|
| EPI_ISL_438096 | Center for Virology, Medical University of Vienna | Bergthaler laboratory, CeMM Research Center for Molecular Medicine of the Austrian Academy of Sciences |
| EPI_ISL_438097 | Center for Virology, Medical University of Vienna | Bergthaler laboratory, CeMM Research Center for Molecular Medicine of the Austrian Academy of Sciences |
| EPI_ISL_438098 | Center for Virology, Medical University of Vienna | Bergthaler laboratory, CeMM Research Center for Molecular Medicine of the Austrian Academy of Sciences |
| EPI_ISL_438099 | Center for Virology, Medical University of Vienna | Bergthaler laboratory, CeMM Research Center for Molecular Medicine of the Austrian Academy of Sciences |
| EPI_ISL_438100 | Center for Virology, Medical University of Vienna | Bergthaler laboratory, CeMM Research Center for Molecular Medicine of the Austrian Academy of Sciences |
| EPI_ISL_438101 | Center for Virology, Medical University of Vienna | Bergthaler laboratory, CeMM Research Center for Molecular Medicine of the Austrian Academy of Sciences |
| EPI_ISL_438102 | Center for Virology, Medical University of Vienna | Bergthaler laboratory, CeMM Research Center for Molecular Medicine of the Austrian Academy of Sciences |
| EPI_ISL_438103 | Center for Virology, Medical University of Vienna | Bergthaler laboratory, CeMM Research Center for Molecular Medicine of the Austrian Academy of Sciences |
| EPI_ISL_438104 | Center for Virology, Medical University of Vienna | Bergthaler laboratory, CeMM Research Center for Molecular Medicine of the Austrian Academy of Sciences |
| EPI_ISL_438105 | Center for Virology, Medical University of Vienna | Bergthaler laboratory, CeMM Research Center for Molecular Medicine of the Austrian Academy of Sciences |
| EPI_ISL_438106 | Center for Virology, Medical University of Vienna | Bergthaler laboratory, CeMM Research Center for Molecular Medicine of the Austrian Academy of Sciences |

| EPI_Accession_ID | Originating Laboratory | Submitting Laboratory |
|---|---|---|
| EPI_ISL_438107 | Center for Virology, Medical University of Vienna | Bergthaler laboratory, CeMM Research Center for Molecular Medicine of the Austrian Academy of Sciences |
| EPI_ISL_438108 | Center for Virology, Medical University of Vienna | Bergthaler laboratory, CeMM Research Center for Molecular Medicine of the Austrian Academy of Sciences |
| EPI_ISL_438109 | Center for Virology, Medical University of Vienna | Bergthaler laboratory, CeMM Research Center for Molecular Medicine of the Austrian Academy of Sciences |
| EPI_ISL_438110 | Center for Virology, Medical University of Vienna | Bergthaler laboratory, CeMM Research Center for Molecular Medicine of the Austrian Academy of Sciences |
| EPI_ISL_438111 | Center for Virology, Medical University of Vienna | Bergthaler laboratory, CeMM Research Center for Molecular Medicine of the Austrian Academy of Sciences |
| EPI_ISL_438112 | Center for Virology, Medical University of Vienna | Bergthaler laboratory, CeMM Research Center for Molecular Medicine of the Austrian Academy of Sciences |
| EPI_ISL_438113 | Center for Virology, Medical University of Vienna | Bergthaler laboratory, CeMM Research Center for Molecular Medicine of the Austrian Academy of Sciences |
| EPI_ISL_438114 | Center for Virology, Medical University of Vienna | Bergthaler laboratory, CeMM Research Center for Molecular Medicine of the Austrian Academy of Sciences |
| EPI_ISL_438115 | Center for Virology, Medical University of Vienna | Bergthaler laboratory, CeMM Research Center for Molecular Medicine of the Austrian Academy of Sciences |
| EPI_ISL_438116 | Center for Virology, Medical University of Vienna | Bergthaler laboratory, CeMM Research Center for Molecular Medicine of the Austrian Academy of Sciences |
| EPI_ISL_438117 | Center for Virology, Medical University of Vienna | Bergthaler laboratory, CeMM Research Center for Molecular Medicine of the Austrian Academy of Sciences |

| EPI_Accession_ID | Originating Laboratory | Submitting Laboratory |
|---|---|---|
| EPI_ISL_438118 | Center for Virology, Medical University of Vienna | Bergthaler laboratory, CeMM Research Center for Molecular Medicine of the Austrian Academy of Sciences |
| EPI_ISL_438119 | Center for Virology, Medical University of Vienna | Bergthaler laboratory, CeMM Research Center for Molecular Medicine of the Austrian Academy of Sciences |
| EPI_ISL_438120 | Center for Virology, Medical University of Vienna | Bergthaler laboratory, CeMM Research Center for Molecular Medicine of the Austrian Academy of Sciences |
| EPI_ISL_438121 | Center for Virology, Medical University of Vienna | Bergthaler laboratory, CeMM Research Center for Molecular Medicine of the Austrian Academy of Sciences |
| EPI_ISL_438122 | Center for Virology, Medical University of Vienna | Bergthaler laboratory, CeMM Research Center for Molecular Medicine of the Austrian Academy of Sciences |
| EPI_ISL_438123 | Center for Virology, Medical University of Vienna | Bergthaler laboratory, CeMM Research Center for Molecular Medicine of the Austrian Academy of Sciences |
| EPI_ISL_438124 | Center for Virology, Medical University of Vienna | Bergthaler laboratory, CeMM Research Center for Molecular Medicine of the Austrian Academy of Sciences |
| EPI_ISL_438125 | Center for Virology, Medical University of Vienna | Bergthaler laboratory, CeMM Research Center for Molecular Medicine of the Austrian Academy of Sciences |
| EPI_ISL_438126 | Center for Virology, Medical University of Vienna | Bergthaler laboratory, CeMM Research Center for Molecular Medicine of the Austrian Academy of Sciences |
| EPI_ISL_438127 | Center for Virology, Medical University of Vienna | Bergthaler laboratory, CeMM Research Center for Molecular Medicine of the Austrian Academy of Sciences |
| EPI_ISL_438128 | Center for Virology, Medical University of Vienna | Bergthaler laboratory, CeMM Research Center for Molecular Medicine of the Austrian Academy of Sciences |

| EPI_Accession_ID | Originating Laboratory | Submitting Laboratory |
|---|---|---|
| EPI_ISL_438138 | Department of Microbiology,Gandhi Medical College and Hospital | Department of Microbiology, Gandhi Medical College and Hospital Secendrabad, Hyderbad, India |
| EPI_ISL_438139 | Department of Microbiology,Gandhi Medical College and Hospital,Hyderabad | Virus Research Laboratory, Department of Zoology, Osmania University, Hyderabad, India |
| EPI_ISL_438140 | Seattle Flu Study | Seattle Flu Study |
| EPI_ISL_438141 | Seattle Flu Study | Seattle Flu Study |
| EPI_ISL_438142 | Seattle Flu Study | Seattle Flu Study |
| EPI_ISL_438143 | Seattle Flu Study | Seattle Flu Study |
| EPI_ISL_438144 | Seattle Flu Study | Seattle Flu Study |
| EPI_ISL_438145 | Seattle Flu Study | Seattle Flu Study |
| EPI_ISL_438146 | Seattle Flu Study | Seattle Flu Study |
| EPI_ISL_438147 | Seattle Flu Study | Seattle Flu Study |
| EPI_ISL_438148 | Seattle Flu Study | Seattle Flu Study |
| EPI_ISL_438149 | Seattle Flu Study | Seattle Flu Study |
| EPI_ISL_438150 | Seattle Flu Study | Seattle Flu Study |
| EPI_ISL_438151 | Seattle Flu Study | Seattle Flu Study |
| EPI_ISL_438152 | Seattle Flu Study | Seattle Flu Study |
| EPI_ISL_438153 | Seattle Flu Study | Seattle Flu Study |
| EPI_ISL_438154 | Seattle Flu Study | Seattle Flu Study |
| EPI_ISL_438155 | Seattle Flu Study | Seattle Flu Study |
| EPI_ISL_438156 | Seattle Flu Study | Seattle Flu Study |
| EPI_ISL_438157 | Seattle Flu Study | Seattle Flu Study |
| EPI_ISL_438158 | Seattle Flu Study | Seattle Flu Study |
| EPI_ISL_438159 | Seattle Flu Study | Seattle Flu Study |
| EPI_ISL_438160 | Seattle Flu Study | Seattle Flu Study |
| EPI_ISL_438161 | Seattle Flu Study | Seattle Flu Study |
| EPI_ISL_438162 | Seattle Flu Study | Seattle Flu Study |
| EPI_ISL_438163 | Seattle Flu Study | Seattle Flu Study |
| EPI_ISL_438164 | Seattle Flu Study | Seattle Flu Study |
| EPI_ISL_438165 | Seattle Flu Study | Seattle Flu Study |
| EPI_ISL_438166 | Seattle Flu Study | Seattle Flu Study |
| EPI_ISL_438167 | Seattle Flu Study | Seattle Flu Study |
| EPI_ISL_438168 | Seattle Flu Study | Seattle Flu Study |

| EPI_Accession_ID | Originating Laboratory | Submitting Laboratory |
|---|---|---|
| EPI_ISL_438169 | Seattle Flu Study | Seattle Flu Study |
| EPI_ISL_438170 | Seattle Flu Study | Seattle Flu Study |
| EPI_ISL_438171 | Seattle Flu Study | Seattle Flu Study |
| EPI_ISL_438172 | Seattle Flu Study | Seattle Flu Study |
| EPI_ISL_438173 | Seattle Flu Study | Seattle Flu Study |
| EPI_ISL_438174 | Seattle Flu Study | Seattle Flu Study |
| EPI_ISL_438175 | Seattle Flu Study | Seattle Flu Study |
| EPI_ISL_438176 | Washington State Department of Health | Seattle Flu Study |
| EPI_ISL_438177 | Washington State Department of Health | Seattle Flu Study |
| EPI_ISL_438178 | Washington State Department of Health | Seattle Flu Study |
| EPI_ISL_438179 | Washington State Department of Health | Seattle Flu Study |
| EPI_ISL_438180 | Washington State Department of Health | Seattle Flu Study |
| EPI_ISL_438181 | Washington State Department of Health | Seattle Flu Study |
| EPI_ISL_438182 | Washington State Department of Health | Seattle Flu Study |
| EPI_ISL_438183 | Washington State Department of Health | Seattle Flu Study |
| EPI_ISL_438184 | Washington State Department of Health | Seattle Flu Study |
| EPI_ISL_438185 | Washington State Department of Health | Seattle Flu Study |
| EPI_ISL_438186 | Washington State Department of Health | Seattle Flu Study |
| EPI_ISL_438187 | Washington State Department of Health | Seattle Flu Study |
| EPI_ISL_438188 | Washington State Department of Health | Seattle Flu Study |
| EPI_ISL_438189 | Washington State Department of Health | Seattle Flu Study |
| EPI_ISL_438190 | Washington State Department of Health | Seattle Flu Study |
| EPI_ISL_438191 | Washington State Department of Health | Seattle Flu Study |
| EPI_ISL_438192 | Washington State Department of Health | Seattle Flu Study |
| EPI_ISL_438193 | Washington State Department of Health | Seattle Flu Study |
| EPI_ISL_438194 | Washington State Department of Health | Seattle Flu Study |
| EPI_ISL_438195 | Washington State Department of Health | Seattle Flu Study |
| EPI_ISL_438196 | Washington State Department of Health | Seattle Flu Study |
| EPI_ISL_438197 | Washington State Department of Health | Seattle Flu Study |
| EPI_ISL_438198 | Washington State Department of Health | Seattle Flu Study |
| EPI_ISL_438199 | Washington State Department of Health | Seattle Flu Study |
| EPI_ISL_438200 | Washington State Department of Health | Seattle Flu Study |
| EPI_ISL_438201 | Washington State Department of Health | Seattle Flu Study |

| EPI_Accession_ID | Originating Laboratory | Submitting Laboratory |
|---|---|---|
| EPI_ISL_438202 | Washington State Department of Health | Seattle Flu Study |
| EPI_ISL_438203 | Washington State Department of Health | Seattle Flu Study |
| EPI_ISL_438204 | Washington State Department of Health | Seattle Flu Study |
| EPI_ISL_438205 | Washington State Department of Health | Seattle Flu Study |
| EPI_ISL_438206 | Washington State Department of Health | Seattle Flu Study |
| EPI_ISL_438207 | Washington State Department of Health | Seattle Flu Study |
| EPI_ISL_438208 | Washington State Department of Health | Seattle Flu Study |
| EPI_ISL_438209 | Washington State Department of Health | Seattle Flu Study |
| EPI_ISL_438210 | Washington State Department of Health | Seattle Flu Study |
| EPI_ISL_438211 | Washington State Department of Health | Seattle Flu Study |
| EPI_ISL_438212 | Washington State Department of Health | Seattle Flu Study |
| EPI_ISL_438213 | Washington State Department of Health | Seattle Flu Study |
| EPI_ISL_438214 | Washington State Department of Health | Seattle Flu Study |
| EPI_ISL_438215 | Washington State Department of Health | Seattle Flu Study |
| EPI_ISL_438216 | Washington State Department of Health | Seattle Flu Study |
| EPI_ISL_438217 | Washington State Department of Health | Seattle Flu Study |
| EPI_ISL_438218 | Washington State Department of Health | Seattle Flu Study |
| EPI_ISL_438219 | Washington State Department of Health | Seattle Flu Study |
| EPI_ISL_438220 | Washington State Department of Health | Seattle Flu Study |
| EPI_ISL_438221 | Washington State Department of Health | Seattle Flu Study |
| EPI_ISL_438222 | Johns Hopkins Hospital Department of Pathology | Johns Hopkins Hospital Department of Pathology |
| EPI_ISL_438223 | Johns Hopkins Hospital Department of Pathology | Johns Hopkins Hospital Department of Pathology |
| EPI_ISL_438224 | Johns Hopkins Hospital Department of Pathology | Johns Hopkins Hospital Department of Pathology |
| EPI_ISL_438225 | Johns Hopkins Hospital Department of Pathology | Johns Hopkins Hospital Department of Pathology |
| EPI_ISL_438226 | Johns Hopkins Hospital Department of Pathology | Johns Hopkins Hospital Department of Pathology |
| EPI_ISL_438227 | Johns Hopkins Hospital Department of Pathology | Johns Hopkins Hospital Department of Pathology |
| EPI_ISL_438228 | Johns Hopkins Hospital Department of Pathology | Johns Hopkins Hospital Department of Pathology |
| EPI_ISL_438229 | Johns Hopkins Hospital Department of Pathology | Johns Hopkins Hospital Department of Pathology |
| EPI_ISL_438230 | Johns Hopkins Hospital Department of Pathology | Johns Hopkins Hospital Department of Pathology |
| EPI_ISL_438231 | Johns Hopkins Hospital Department of Pathology | Johns Hopkins Hospital Department of Pathology |
| EPI_ISL_438232 | Johns Hopkins Hospital Department of Pathology | Johns Hopkins Hospital Department of Pathology |
| EPI_ISL_438233 | Johns Hopkins Hospital Department of Pathology | Johns Hopkins Hospital Department of Pathology |
| EPI_ISL_438234 | Johns Hopkins Hospital Department of Pathology | Johns Hopkins Hospital Department of Pathology |

| EPI_Accession_ID | Originating Laboratory | Submitting Laboratory |
|---|---|---|
| EPI_ISL_438235 | Johns Hopkins Hospital Department of Pathology | Johns Hopkins Hospital Department of Pathology |
| EPI_ISL_438236 | Johns Hopkins Hospital Department of Pathology | Johns Hopkins Hospital Department of Pathology |
| EPI_ISL_438237 | Johns Hopkins Hospital Department of Pathology | Johns Hopkins Hospital Department of Pathology |
| EPI_ISL_438238 | Johns Hopkins Hospital Department of Pathology | Johns Hopkins Hospital Department of Pathology |
| EPI_ISL_438239 | Johns Hopkins Hospital Department of Pathology | Johns Hopkins Hospital Department of Pathology |
| EPI_ISL_438240 | Johns Hopkins Hospital Department of Pathology | Johns Hopkins Hospital Department of Pathology |
| EPI_ISL_438241 | Johns Hopkins Hospital Department of Pathology | Johns Hopkins Hospital Department of Pathology |
| EPI_ISL_438242 | Johns Hopkins Hospital Department of Pathology | Johns Hopkins Hospital Department of Pathology |
| EPI_ISL_438243 | Johns Hopkins Hospital Department of Pathology | Johns Hopkins Hospital Department of Pathology |
| EPI_ISL_438244 | Johns Hopkins Hospital Department of Pathology | Johns Hopkins Hospital Department of Pathology |
| EPI_ISL_438245 | Johns Hopkins Hospital Department of Pathology | Johns Hopkins Hospital Department of Pathology |
| EPI_ISL_438246 | Johns Hopkins Hospital Department of Pathology | Johns Hopkins Hospital Department of Pathology |
| EPI_ISL_438247 | Johns Hopkins Hospital Department of Pathology | Johns Hopkins Hospital Department of Pathology |
| EPI_ISL_438248 | Department of Pathology, University of Cambridge | Wellcome Sanger Institute for the COVID-19 Genomics UK Consortium |
| EPI_ISL_438249 | Department of Pathology, University of Cambridge | Wellcome Sanger Institute for the COVID-19 Genomics UK Consortium |
| EPI_ISL_438250 | Department of Pathology, University of Cambridge | Wellcome Sanger Institute for the COVID-19 Genomics UK Consortium |
| EPI_ISL_438251 | Department of Pathology, University of Cambridge | Wellcome Sanger Institute for the COVID-19 Genomics UK Consortium |
| EPI_ISL_438252 | Department of Pathology, University of Cambridge | Wellcome Sanger Institute for the COVID-19 Genomics UK Consortium |
| EPI_ISL_438253 | Department of Pathology, University of Cambridge | Wellcome Sanger Institute for the COVID-19 Genomics UK Consortium |
| EPI_ISL_438254 | Department of Pathology, University of Cambridge | Wellcome Sanger Institute for the COVID-19 Genomics UK Consortium |
| EPI_ISL_438255 | Department of Pathology, University of Cambridge | Wellcome Sanger Institute for the COVID-19 Genomics UK Consortium |
| EPI_ISL_438256 | Department of Pathology, University of Cambridge | Wellcome Sanger Institute for the COVID-19 Genomics UK Consortium |
| EPI_ISL_438257 | Department of Pathology, University of Cambridge | Wellcome Sanger Institute for the COVID-19 Genomics UK Consortium |

| EPI_Accession_ID | Originating Laboratory | Submitting Laboratory |
|---|---|---|
| EPI_ISL_438258 | Department of Pathology, University of Cambridge | Wellcome Sanger Institute for the COVID-19 Genomics UK Consortium |
| EPI_ISL_438259 | Department of Pathology, University of Cambridge | Wellcome Sanger Institute for the COVID-19 Genomics UK Consortium |
| EPI_ISL_438260 | Department of Pathology, University of Cambridge | Wellcome Sanger Institute for the COVID-19 Genomics UK Consortium |
| EPI_ISL_438261 | Department of Pathology, University of Cambridge | Wellcome Sanger Institute for the COVID-19 Genomics UK Consortium |
| EPI_ISL_438262 | Department of Pathology, University of Cambridge | Wellcome Sanger Institute for the COVID-19 Genomics UK Consortium |
| EPI_ISL_438263 | Department of Pathology, University of Cambridge | Wellcome Sanger Institute for the COVID-19 Genomics UK Consortium |
| EPI_ISL_438264 | Department of Pathology, University of Cambridge | Wellcome Sanger Institute for the COVID-19 Genomics UK Consortium |
| EPI_ISL_438265 | Department of Pathology, University of Cambridge | Wellcome Sanger Institute for the COVID-19 Genomics UK Consortium |
| EPI_ISL_438266 | Department of Pathology, University of Cambridge | Wellcome Sanger Institute for the COVID-19 Genomics UK Consortium |
| EPI_ISL_438267 | Department of Pathology, University of Cambridge | Wellcome Sanger Institute for the COVID-19 Genomics UK Consortium |
| EPI_ISL_438268 | Department of Pathology, University of Cambridge | Wellcome Sanger Institute for the COVID-19 Genomics UK Consortium |
| EPI_ISL_438269 | Department of Pathology, University of Cambridge | Wellcome Sanger Institute for the COVID-19 Genomics UK Consortium |
| EPI_ISL_438270 | Department of Pathology, University of Cambridge | Wellcome Sanger Institute for the COVID-19 Genomics UK Consortium |
| EPI_ISL_438271 | Department of Pathology, University of Cambridge | Wellcome Sanger Institute for the COVID-19 Genomics UK Consortium |
| EPI_ISL_438272 | Department of Pathology, University of Cambridge | Wellcome Sanger Institute for the COVID-19 Genomics UK Consortium |
| EPI_ISL_438273 | Department of Pathology, University of Cambridge | Wellcome Sanger Institute for the COVID-19 Genomics UK Consortium |

| EPI_Accession_ID | Originating Laboratory | Submitting Laboratory |
|---|---|---|
| EPI_ISL_438274 | Department of Pathology, University of Cambridge | Wellcome Sanger Institute for the COVID-19 Genomics UK Consortium |
| EPI_ISL_438275 | Department of Pathology, University of Cambridge | Wellcome Sanger Institute for the COVID-19 Genomics UK Consortium |
| EPI_ISL_438276 | Department of Pathology, University of Cambridge | Wellcome Sanger Institute for the COVID-19 Genomics UK Consortium |
| EPI_ISL_438277 | Department of Pathology, University of Cambridge | Wellcome Sanger Institute for the COVID-19 Genomics UK Consortium |
| EPI_ISL_438278 | Department of Pathology, University of Cambridge | Wellcome Sanger Institute for the COVID-19 Genomics UK Consortium |
| EPI_ISL_438279 | Department of Pathology, University of Cambridge | Wellcome Sanger Institute for the COVID-19 Genomics UK Consortium |
| EPI_ISL_438280 | Department of Pathology, University of Cambridge | Wellcome Sanger Institute for the COVID-19 Genomics UK Consortium |
| EPI_ISL_438281 | Department of Pathology, University of Cambridge | Wellcome Sanger Institute for the COVID-19 Genomics UK Consortium |
| EPI_ISL_438282 | Department of Pathology, University of Cambridge | Wellcome Sanger Institute for the COVID-19 Genomics UK Consortium |
| EPI_ISL_438283 | Department of Pathology, University of Cambridge | Wellcome Sanger Institute for the COVID-19 Genomics UK Consortium |
| EPI_ISL_438284 | Department of Pathology, University of Cambridge | Wellcome Sanger Institute for the COVID-19 Genomics UK Consortium |
| EPI_ISL_438285 | Department of Pathology, University of Cambridge | Wellcome Sanger Institute for the COVID-19 Genomics UK Consortium |
| EPI_ISL_438286 | Department of Pathology, University of Cambridge | Wellcome Sanger Institute for the COVID-19 Genomics UK Consortium |
| EPI_ISL_438287 | Department of Pathology, University of Cambridge | Wellcome Sanger Institute for the COVID-19 Genomics UK Consortium |
| EPI_ISL_438288 | Department of Pathology, University of Cambridge | Wellcome Sanger Institute for the COVID-19 Genomics UK Consortium |
| EPI_ISL_438289 | Department of Pathology, University of Cambridge | Wellcome Sanger Institute for the COVID-19 Genomics UK Consortium |

| EPI_Accession_ID | Originating Laboratory | Submitting Laboratory |
|---|---|---|
| EPI_ISL_438290 | Department of Pathology, University of Cambridge | Wellcome Sanger Institute for the COVID-19 Genomics UK Consortium |
| EPI_ISL_438291 | Department of Pathology, University of Cambridge | Wellcome Sanger Institute for the COVID-19 Genomics UK Consortium |
| EPI_ISL_438292 | Department of Pathology, University of Cambridge | Wellcome Sanger Institute for the COVID-19 Genomics UK Consortium |
| EPI_ISL_438293 | Department of Pathology, University of Cambridge | Wellcome Sanger Institute for the COVID-19 Genomics UK Consortium |
| EPI_ISL_438294 | Department of Pathology, University of Cambridge | Wellcome Sanger Institute for the COVID-19 Genomics UK Consortium |
| EPI_ISL_438295 | Department of Pathology, University of Cambridge | Wellcome Sanger Institute for the COVID-19 Genomics UK Consortium |
| EPI_ISL_438296 | Department of Pathology, University of Cambridge | Wellcome Sanger Institute for the COVID-19 Genomics UK Consortium |
| EPI_ISL_438297 | Department of Pathology, University of Cambridge | Wellcome Sanger Institute for the COVID-19 Genomics UK Consortium |
| EPI_ISL_438298 | Department of Pathology, University of Cambridge | Wellcome Sanger Institute for the COVID-19 Genomics UK Consortium |
| EPI_ISL_438299 | Department of Pathology, University of Cambridge | Wellcome Sanger Institute for the COVID-19 Genomics UK Consortium |
| EPI_ISL_438300 | Department of Pathology, University of Cambridge | Wellcome Sanger Institute for the COVID-19 Genomics UK Consortium |
| EPI_ISL_438301 | Department of Pathology, University of Cambridge | Wellcome Sanger Institute for the COVID-19 Genomics UK Consortium |
| EPI_ISL_438302 | Department of Pathology, University of Cambridge | Wellcome Sanger Institute for the COVID-19 Genomics UK Consortium |
| EPI_ISL_438303 | Department of Pathology, University of Cambridge | Wellcome Sanger Institute for the COVID-19 Genomics UK Consortium |
| EPI_ISL_438304 | Department of Pathology, University of Cambridge | Wellcome Sanger Institute for the COVID-19 Genomics UK Consortium |
| EPI_ISL_438305 | Department of Pathology, University of Cambridge | Wellcome Sanger Institute for the COVID-19 Genomics UK Consortium |

| EPI_Accession_ID | Originating Laboratory | Submitting Laboratory |
|---|---|---|
| EPI_ISL_438306 | Department of Pathology, University of Cambridge | Wellcome Sanger Institute for the COVID-19 Genomics UK Consortium |
| EPI_ISL_438307 | Department of Pathology, University of Cambridge | Wellcome Sanger Institute for the COVID-19 Genomics UK Consortium |
| EPI_ISL_438308 | Department of Pathology, University of Cambridge | Wellcome Sanger Institute for the COVID-19 Genomics UK Consortium |
| EPI_ISL_438309 | Department of Pathology, University of Cambridge | Wellcome Sanger Institute for the COVID-19 Genomics UK Consortium |
| EPI_ISL_438310 | Department of Pathology, University of Cambridge | Wellcome Sanger Institute for the COVID-19 Genomics UK Consortium |
| EPI_ISL_438311 | Department of Pathology, University of Cambridge | Wellcome Sanger Institute for the COVID-19 Genomics UK Consortium |
| EPI_ISL_438312 | Department of Pathology, University of Cambridge | Wellcome Sanger Institute for the COVID-19 Genomics UK Consortium |
| EPI_ISL_438313 | Department of Pathology, University of Cambridge | Wellcome Sanger Institute for the COVID-19 Genomics UK Consortium |
| EPI_ISL_438314 | Department of Pathology, University of Cambridge | Wellcome Sanger Institute for the COVID-19 Genomics UK Consortium |
| EPI_ISL_438315 | Department of Pathology, University of Cambridge | Wellcome Sanger Institute for the COVID-19 Genomics UK Consortium |
| EPI_ISL_438316 | Department of Pathology, University of Cambridge | Wellcome Sanger Institute for the COVID-19 Genomics UK Consortium |
| EPI_ISL_438317 | Department of Pathology, University of Cambridge | Wellcome Sanger Institute for the COVID-19 Genomics UK Consortium |
| EPI_ISL_438318 | Department of Pathology, University of Cambridge | Wellcome Sanger Institute for the COVID-19 Genomics UK Consortium |
| EPI_ISL_438319 | Department of Pathology, University of Cambridge | Wellcome Sanger Institute for the COVID-19 Genomics UK Consortium |
| EPI_ISL_438320 | Department of Pathology, University of Cambridge | Wellcome Sanger Institute for the COVID-19 Genomics UK Consortium |
| EPI_ISL_438321 | Department of Pathology, University of Cambridge | Wellcome Sanger Institute for the COVID-19 Genomics UK Consortium |

| EPI_Accession_ID | Originating Laboratory | Submitting Laboratory |
|---|---|---|
| EPI_ISL_438322 | Department of Pathology, University of Cambridge | Wellcome Sanger Institute for the COVID-19 Genomics UK Consortium |
| EPI_ISL_438323 | Department of Pathology, University of Cambridge | Wellcome Sanger Institute for the COVID-19 Genomics UK Consortium |
| EPI_ISL_438324 | Department of Pathology, University of Cambridge | Wellcome Sanger Institute for the COVID-19 Genomics UK Consortium |
| EPI_ISL_438325 | Department of Pathology, University of Cambridge | Wellcome Sanger Institute for the COVID-19 Genomics UK Consortium |
| EPI_ISL_438326 | Department of Pathology, University of Cambridge | Wellcome Sanger Institute for the COVID-19 Genomics UK Consortium |
| EPI_ISL_438327 | Department of Pathology, University of Cambridge | Wellcome Sanger Institute for the COVID-19 Genomics UK Consortium |
| EPI_ISL_438328 | Department of Pathology, University of Cambridge | Wellcome Sanger Institute for the COVID-19 Genomics UK Consortium |
| EPI_ISL_438329 | Department of Pathology, University of Cambridge | Wellcome Sanger Institute for the COVID-19 Genomics UK Consortium |
| EPI_ISL_438330 | Department of Pathology, University of Cambridge | Wellcome Sanger Institute for the COVID-19 Genomics UK Consortium |
| EPI_ISL_438331 | Department of Pathology, University of Cambridge | Wellcome Sanger Institute for the COVID-19 Genomics UK Consortium |
| EPI_ISL_438332 | Department of Pathology, University of Cambridge | Wellcome Sanger Institute for the COVID-19 Genomics UK Consortium |
| EPI_ISL_438333 | Department of Pathology, University of Cambridge | Wellcome Sanger Institute for the COVID-19 Genomics UK Consortium |
| EPI_ISL_438334 | Department of Pathology, University of Cambridge | Wellcome Sanger Institute for the COVID-19 Genomics UK Consortium |
| EPI_ISL_438335 | Department of Pathology, University of Cambridge | Wellcome Sanger Institute for the COVID-19 Genomics UK Consortium |
| EPI_ISL_438336 | Department of Pathology, University of Cambridge | Wellcome Sanger Institute for the COVID-19 Genomics UK Consortium |
| EPI_ISL_438337 | Department of Pathology, University of Cambridge | Wellcome Sanger Institute for the COVID-19 Genomics UK Consortium |

| EPI_Accession_ID | Originating Laboratory | Submitting Laboratory |
|---|---|---|
| EPI_ISL_438338 | Department of Pathology, University of Cambridge | Wellcome Sanger Institute for the COVID-19 Genomics UK Consortium |
| EPI_ISL_438339 | Department of Pathology, University of Cambridge | Wellcome Sanger Institute for the COVID-19 Genomics UK Consortium |
| EPI_ISL_438340 | Department of Pathology, University of Cambridge | Wellcome Sanger Institute for the COVID-19 Genomics UK Consortium |
| EPI_ISL_438341 | Department of Pathology, University of Cambridge | Wellcome Sanger Institute for the COVID-19 Genomics UK Consortium |
| EPI_ISL_438342 | Department of Pathology, University of Cambridge | Wellcome Sanger Institute for the COVID-19 Genomics UK Consortium |
| EPI_ISL_438343 | Department of Pathology, University of Cambridge | Wellcome Sanger Institute for the COVID-19 Genomics UK Consortium |
| EPI_ISL_438344 | Department of Pathology, University of Cambridge | Wellcome Sanger Institute for the COVID-19 Genomics UK Consortium |
| EPI_ISL_438345 | Department of Pathology, University of Cambridge | Wellcome Sanger Institute for the COVID-19 Genomics UK Consortium |
| EPI_ISL_438346 | Department of Pathology, University of Cambridge | Wellcome Sanger Institute for the COVID-19 Genomics UK Consortium |
| EPI_ISL_438347 | Department of Pathology, University of Cambridge | Wellcome Sanger Institute for the COVID-19 Genomics UK Consortium |
| EPI_ISL_438348 | Department of Pathology, University of Cambridge | Wellcome Sanger Institute for the COVID-19 Genomics UK Consortium |
| EPI_ISL_438349 | Department of Pathology, University of Cambridge | Wellcome Sanger Institute for the COVID-19 Genomics UK Consortium |
| EPI_ISL_438350 | Department of Pathology, University of Cambridge | Wellcome Sanger Institute for the COVID-19 Genomics UK Consortium |
| EPI_ISL_438351 | Department of Pathology, University of Cambridge | Wellcome Sanger Institute for the COVID-19 Genomics UK Consortium |
| EPI_ISL_438352 | Department of Pathology, University of Cambridge | Wellcome Sanger Institute for the COVID-19 Genomics UK Consortium |
| EPI_ISL_438353 | Department of Pathology, University of Cambridge | Wellcome Sanger Institute for the COVID-19 Genomics UK Consortium |

| EPI_Accession_ID | Originating Laboratory | Submitting Laboratory |
|---|---|---|
| EPI_ISL_438354 | Department of Pathology, University of Cambridge | Wellcome Sanger Institute for the COVID-19 Genomics UK Consortium |
| EPI_ISL_438355 | Department of Pathology, University of Cambridge | Wellcome Sanger Institute for the COVID-19 Genomics UK Consortium |
| EPI_ISL_438356 | Department of Pathology, University of Cambridge | Wellcome Sanger Institute for the COVID-19 Genomics UK Consortium |
| EPI_ISL_438357 | Department of Pathology, University of Cambridge | Wellcome Sanger Institute for the COVID-19 Genomics UK Consortium |
| EPI_ISL_438358 | Department of Pathology, University of Cambridge | Wellcome Sanger Institute for the COVID-19 Genomics UK Consortium |
| EPI_ISL_438359 | Department of Pathology, University of Cambridge | Wellcome Sanger Institute for the COVID-19 Genomics UK Consortium |
| EPI_ISL_438360 | Department of Pathology, University of Cambridge | Wellcome Sanger Institute for the COVID-19 Genomics UK Consortium |
| EPI_ISL_438361 | Department of Pathology, University of Cambridge | Wellcome Sanger Institute for the COVID-19 Genomics UK Consortium |
| EPI_ISL_438362 | Department of Pathology, University of Cambridge | Wellcome Sanger Institute for the COVID-19 Genomics UK Consortium |
| EPI_ISL_438363 | Department of Pathology, University of Cambridge | Wellcome Sanger Institute for the COVID-19 Genomics UK Consortium |
| EPI_ISL_438364 | Department of Pathology, University of Cambridge | Wellcome Sanger Institute for the COVID-19 Genomics UK Consortium |
| EPI_ISL_438365 | Department of Pathology, University of Cambridge | Wellcome Sanger Institute for the COVID-19 Genomics UK Consortium |
| EPI_ISL_438366 | Department of Pathology, University of Cambridge | Wellcome Sanger Institute for the COVID-19 Genomics UK Consortium |
| EPI_ISL_438367 | Department of Pathology, University of Cambridge | Wellcome Sanger Institute for the COVID-19 Genomics UK Consortium |
| EPI_ISL_438368 | Department of Pathology, University of Cambridge | Wellcome Sanger Institute for the COVID-19 Genomics UK Consortium |
| EPI_ISL_438369 | Department of Pathology, University of Cambridge | Wellcome Sanger Institute for the COVID-19 Genomics UK Consortium |

| EPI_Accession_ID | Originating Laboratory | Submitting Laboratory |
|---|---|---|
| EPI_ISL_438370 | Department of Pathology, University of Cambridge | Wellcome Sanger Institute for the COVID-19 Genomics UK Consortium |
| EPI_ISL_438371 | Department of Pathology, University of Cambridge | Wellcome Sanger Institute for the COVID-19 Genomics UK Consortium |
| EPI_ISL_438372 | Department of Pathology, University of Cambridge | Wellcome Sanger Institute for the COVID-19 Genomics UK Consortium |
| EPI_ISL_438373 | Department of Pathology, University of Cambridge | Wellcome Sanger Institute for the COVID-19 Genomics UK Consortium |
| EPI_ISL_438374 | Department of Pathology, University of Cambridge | Wellcome Sanger Institute for the COVID-19 Genomics UK Consortium |
| EPI_ISL_438375 | Department of Pathology, University of Cambridge | Wellcome Sanger Institute for the COVID-19 Genomics UK Consortium |
| EPI_ISL_438376 | Department of Pathology, University of Cambridge | Wellcome Sanger Institute for the COVID-19 Genomics UK Consortium |
| EPI_ISL_438377 | Department of Pathology, University of Cambridge | Wellcome Sanger Institute for the COVID-19 Genomics UK Consortium |
| EPI_ISL_438378 | Department of Pathology, University of Cambridge | Wellcome Sanger Institute for the COVID-19 Genomics UK Consortium |
| EPI_ISL_438379 | Department of Pathology, University of Cambridge | Wellcome Sanger Institute for the COVID-19 Genomics UK Consortium |
| EPI_ISL_438380 | Department of Pathology, University of Cambridge | Wellcome Sanger Institute for the COVID-19 Genomics UK Consortium |
| EPI_ISL_438381 | Department of Pathology, University of Cambridge | Wellcome Sanger Institute for the COVID-19 Genomics UK Consortium |
| EPI_ISL_438382 | Department of Pathology, University of Cambridge | Wellcome Sanger Institute for the COVID-19 Genomics UK Consortium |
| EPI_ISL_438383 | Department of Pathology, University of Cambridge | Wellcome Sanger Institute for the COVID-19 Genomics UK Consortium |
| EPI_ISL_438384 | Department of Pathology, University of Cambridge | Wellcome Sanger Institute for the COVID-19 Genomics UK Consortium |
| EPI_ISL_438385 | Department of Pathology, University of Cambridge | Wellcome Sanger Institute for the COVID-19 Genomics UK Consortium |

| EPI_Accession_ID | Originating Laboratory | Submitting Laboratory |
|---|---|---|
| EPI_ISL_438386 | Department of Pathology, University of Cambridge | Wellcome Sanger Institute for the COVID-19 Genomics UK Consortium |
| EPI_ISL_438387 | Department of Pathology, University of Cambridge | Wellcome Sanger Institute for the COVID-19 Genomics UK Consortium |
| EPI_ISL_438388 | Department of Pathology, University of Cambridge | Wellcome Sanger Institute for the COVID-19 Genomics UK Consortium |
| EPI_ISL_438389 | Department of Pathology, University of Cambridge | Wellcome Sanger Institute for the COVID-19 Genomics UK Consortium |
| EPI_ISL_438390 | Department of Pathology, University of Cambridge | Wellcome Sanger Institute for the COVID-19 Genomics UK Consortium |
| EPI_ISL_438391 | Department of Pathology, University of Cambridge | Wellcome Sanger Institute for the COVID-19 Genomics UK Consortium |
| EPI_ISL_438392 | Department of Pathology, University of Cambridge | Wellcome Sanger Institute for the COVID-19 Genomics UK Consortium |
| EPI_ISL_438393 | Department of Pathology, University of Cambridge | Wellcome Sanger Institute for the COVID-19 Genomics UK Consortium |
| EPI_ISL_438394 | Department of Pathology, University of Cambridge | Wellcome Sanger Institute for the COVID-19 Genomics UK Consortium |
| EPI_ISL_438395 | Department of Pathology, University of Cambridge | Wellcome Sanger Institute for the COVID-19 Genomics UK Consortium |
| EPI_ISL_438396 | Department of Pathology, University of Cambridge | Wellcome Sanger Institute for the COVID-19 Genomics UK Consortium |
| EPI_ISL_438397 | Department of Pathology, University of Cambridge | Wellcome Sanger Institute for the COVID-19 Genomics UK Consortium |
| EPI_ISL_438398 | Department of Pathology, University of Cambridge | Wellcome Sanger Institute for the COVID-19 Genomics UK Consortium |
| EPI_ISL_438399 | Department of Pathology, University of Cambridge | Wellcome Sanger Institute for the COVID-19 Genomics UK Consortium |
| EPI_ISL_438400 | Department of Pathology, University of Cambridge | Wellcome Sanger Institute for the COVID-19 Genomics UK Consortium |
| EPI_ISL_438401 | Department of Pathology, University of Cambridge | Wellcome Sanger Institute for the COVID-19 Genomics UK Consortium |

| EPI_Accession_ID | Originating Laboratory | Submitting Laboratory |
|---|---|---|
| EPI_ISL_438402 | Department of Pathology, University of Cambridge | Wellcome Sanger Institute for the COVID-19 Genomics UK Consortium |
| EPI_ISL_438403 | Department of Pathology, University of Cambridge | Wellcome Sanger Institute for the COVID-19 Genomics UK Consortium |
| EPI_ISL_438404 | Department of Pathology, University of Cambridge | Wellcome Sanger Institute for the COVID-19 Genomics UK Consortium |
| EPI_ISL_438405 | Department of Pathology, University of Cambridge | Wellcome Sanger Institute for the COVID-19 Genomics UK Consortium |
| EPI_ISL_438406 | Department of Pathology, University of Cambridge | Wellcome Sanger Institute for the COVID-19 Genomics UK Consortium |
| EPI_ISL_438407 | Department of Pathology, University of Cambridge | Wellcome Sanger Institute for the COVID-19 Genomics UK Consortium |
| EPI_ISL_438408 | Department of Pathology, University of Cambridge | Wellcome Sanger Institute for the COVID-19 Genomics UK Consortium |
| EPI_ISL_438409 | Department of Pathology, University of Cambridge | Wellcome Sanger Institute for the COVID-19 Genomics UK Consortium |
| EPI_ISL_438410 | Department of Pathology, University of Cambridge | Wellcome Sanger Institute for the COVID-19 Genomics UK Consortium |
| EPI_ISL_438411 | Department of Pathology, University of Cambridge | Wellcome Sanger Institute for the COVID-19 Genomics UK Consortium |
| EPI_ISL_438412 | Department of Pathology, University of Cambridge | Wellcome Sanger Institute for the COVID-19 Genomics UK Consortium |
| EPI_ISL_438413 | Department of Pathology, University of Cambridge | Wellcome Sanger Institute for the COVID-19 Genomics UK Consortium |
| EPI_ISL_438414 | Department of Pathology, University of Cambridge | Wellcome Sanger Institute for the COVID-19 Genomics UK Consortium |
| EPI_ISL_438415 | Department of Pathology, University of Cambridge | Wellcome Sanger Institute for the COVID-19 Genomics UK Consortium |
| EPI_ISL_438416 | Department of Pathology, University of Cambridge | Wellcome Sanger Institute for the COVID-19 Genomics UK Consortium |
| EPI_ISL_438417 | Department of Pathology, University of Cambridge | Wellcome Sanger Institute for the COVID-19 Genomics UK Consortium |

| EPI_Accession_ID | Originating Laboratory | Submitting Laboratory |
|---|---|---|
| EPI_ISL_438418 | Department of Pathology, University of Cambridge | Wellcome Sanger Institute for the COVID-19 Genomics UK Consortium |
| EPI_ISL_438419 | Department of Pathology, University of Cambridge | Wellcome Sanger Institute for the COVID-19 Genomics UK Consortium |
| EPI_ISL_438420 | Department of Pathology, University of Cambridge | Wellcome Sanger Institute for the COVID-19 Genomics UK Consortium |
| EPI_ISL_438421 | Department of Pathology, University of Cambridge | Wellcome Sanger Institute for the COVID-19 Genomics UK Consortium |
| EPI_ISL_438422 | Department of Pathology, University of Cambridge | Wellcome Sanger Institute for the COVID-19 Genomics UK Consortium |
| EPI_ISL_438423 | Department of Pathology, University of Cambridge | Wellcome Sanger Institute for the COVID-19 Genomics UK Consortium |
| EPI_ISL_438424 | Department of Pathology, University of Cambridge | Wellcome Sanger Institute for the COVID-19 Genomics UK Consortium |
| EPI_ISL_438425 | Department of Pathology, University of Cambridge | Wellcome Sanger Institute for the COVID-19 Genomics UK Consortium |
| EPI_ISL_438426 | Department of Pathology, University of Cambridge | Wellcome Sanger Institute for the COVID-19 Genomics UK Consortium |
| EPI_ISL_438427 | Department of Pathology, University of Cambridge | Wellcome Sanger Institute for the COVID-19 Genomics UK Consortium |
| EPI_ISL_438428 | Department of Pathology, University of Cambridge | Wellcome Sanger Institute for the COVID-19 Genomics UK Consortium |
| EPI_ISL_438429 | Department of Pathology, University of Cambridge | Wellcome Sanger Institute for the COVID-19 Genomics UK Consortium |
| EPI_ISL_438430 | Department of Pathology, University of Cambridge | Wellcome Sanger Institute for the COVID-19 Genomics UK Consortium |
| EPI_ISL_438431 | Department of Pathology, University of Cambridge | Wellcome Sanger Institute for the COVID-19 Genomics UK Consortium |
| EPI_ISL_438432 | Department of Pathology, University of Cambridge | Wellcome Sanger Institute for the COVID-19 Genomics UK Consortium |
| EPI_ISL_438433 | Department of Pathology, University of Cambridge | Wellcome Sanger Institute for the COVID-19 Genomics UK Consortium |

| EPI_Accession_ID | Originating Laboratory | Submitting Laboratory |
|---|---|---|
| EPI_ISL_438434 | Department of Pathology, University of Cambridge | Wellcome Sanger Institute for the COVID-19 Genomics UK Consortium |
| EPI_ISL_438435 | Department of Pathology, University of Cambridge | Wellcome Sanger Institute for the COVID-19 Genomics UK Consortium |
| EPI_ISL_438436 | Department of Pathology, University of Cambridge | Wellcome Sanger Institute for the COVID-19 Genomics UK Consortium |
| EPI_ISL_438437 | Department of Pathology, University of Cambridge | Wellcome Sanger Institute for the COVID-19 Genomics UK Consortium |
| EPI_ISL_438438 | Department of Pathology, University of Cambridge | Wellcome Sanger Institute for the COVID-19 Genomics UK Consortium |
| EPI_ISL_438439 | Department of Pathology, University of Cambridge | Wellcome Sanger Institute for the COVID-19 Genomics UK Consortium |
| EPI_ISL_438440 | Department of Pathology, University of Cambridge | Wellcome Sanger Institute for the COVID-19 Genomics UK Consortium |
| EPI_ISL_438441 | Department of Pathology, University of Cambridge | Wellcome Sanger Institute for the COVID-19 Genomics UK Consortium |
| EPI_ISL_438442 | Department of Pathology, University of Cambridge | Wellcome Sanger Institute for the COVID-19 Genomics UK Consortium |
| EPI_ISL_438443 | Department of Pathology, University of Cambridge | Wellcome Sanger Institute for the COVID-19 Genomics UK Consortium |
| EPI_ISL_438444 | Department of Pathology, University of Cambridge | Wellcome Sanger Institute for the COVID-19 Genomics UK Consortium |
| EPI_ISL_438445 | Department of Pathology, University of Cambridge | Wellcome Sanger Institute for the COVID-19 Genomics UK Consortium |
| EPI_ISL_438446 | Department of Pathology, University of Cambridge | Wellcome Sanger Institute for the COVID-19 Genomics UK Consortium |
| EPI_ISL_438447 | Department of Pathology, University of Cambridge | Wellcome Sanger Institute for the COVID-19 Genomics UK Consortium |
| EPI_ISL_438448 | Department of Pathology, University of Cambridge | Wellcome Sanger Institute for the COVID-19 Genomics UK Consortium |
| EPI_ISL_438449 | Department of Pathology, University of Cambridge | Wellcome Sanger Institute for the COVID-19 Genomics UK Consortium |

| EPI_Accession_ID | Originating Laboratory | Submitting Laboratory |
|---|---|---|
| EPI_ISL_438450 | Department of Pathology, University of Cambridge | Wellcome Sanger Institute for the COVID-19 Genomics UK Consortium |
| EPI_ISL_438451 | Department of Pathology, University of Cambridge | Wellcome Sanger Institute for the COVID-19 Genomics UK Consortium |
| EPI_ISL_438452 | Department of Pathology, University of Cambridge | Wellcome Sanger Institute for the COVID-19 Genomics UK Consortium |
| EPI_ISL_438453 | Department of Pathology, University of Cambridge | Wellcome Sanger Institute for the COVID-19 Genomics UK Consortium |
| EPI_ISL_438454 | Department of Pathology, University of Cambridge | Wellcome Sanger Institute for the COVID-19 Genomics UK Consortium |
| EPI_ISL_438455 | Department of Pathology, University of Cambridge | Wellcome Sanger Institute for the COVID-19 Genomics UK Consortium |
| EPI_ISL_438456 | Department of Pathology, University of Cambridge | Wellcome Sanger Institute for the COVID-19 Genomics UK Consortium |
| EPI_ISL_438457 | Department of Pathology, University of Cambridge | Wellcome Sanger Institute for the COVID-19 Genomics UK Consortium |
| EPI_ISL_438458 | Department of Pathology, University of Cambridge | Wellcome Sanger Institute for the COVID-19 Genomics UK Consortium |
| EPI_ISL_438459 | Department of Pathology, University of Cambridge | Wellcome Sanger Institute for the COVID-19 Genomics UK Consortium |
| EPI_ISL_438460 | Department of Pathology, University of Cambridge | Wellcome Sanger Institute for the COVID-19 Genomics UK Consortium |
| EPI_ISL_438461 | Department of Pathology, University of Cambridge | Wellcome Sanger Institute for the COVID-19 Genomics UK Consortium |
| EPI_ISL_438462 | Department of Pathology, University of Cambridge | Wellcome Sanger Institute for the COVID-19 Genomics UK Consortium |
| EPI_ISL_438463 | Department of Pathology, University of Cambridge | Wellcome Sanger Institute for the COVID-19 Genomics UK Consortium |
| EPI_ISL_438464 | Department of Pathology, University of Cambridge | Wellcome Sanger Institute for the COVID-19 Genomics UK Consortium |
| EPI_ISL_438465 | Department of Pathology, University of Cambridge | Wellcome Sanger Institute for the COVID-19 Genomics UK Consortium |

| EPI_Accession_ID | Originating Laboratory | Submitting Laboratory |
|---|---|---|
| EPI_ISL_438466 | Department of Pathology, University of Cambridge | Wellcome Sanger Institute for the COVID-19 Genomics UK Consortium |
| EPI_ISL_438467 | Department of Pathology, University of Cambridge | Wellcome Sanger Institute for the COVID-19 Genomics UK Consortium |
| EPI_ISL_438468 | Department of Pathology, University of Cambridge | Wellcome Sanger Institute for the COVID-19 Genomics UK Consortium |
| EPI_ISL_438469 | Department of Pathology, University of Cambridge | Wellcome Sanger Institute for the COVID-19 Genomics UK Consortium |
| EPI_ISL_438470 | Department of Pathology, University of Cambridge | Wellcome Sanger Institute for the COVID-19 Genomics UK Consortium |
| EPI_ISL_438471 | Department of Pathology, University of Cambridge | Wellcome Sanger Institute for the COVID-19 Genomics UK Consortium |
| EPI_ISL_438472 | Department of Pathology, University of Cambridge | Wellcome Sanger Institute for the COVID-19 Genomics UK Consortium |
| EPI_ISL_438473 | Department of Pathology, University of Cambridge | Wellcome Sanger Institute for the COVID-19 Genomics UK Consortium |
| EPI_ISL_438474 | Department of Pathology, University of Cambridge | Wellcome Sanger Institute for the COVID-19 Genomics UK Consortium |
| EPI_ISL_438475 | Department of Pathology, University of Cambridge | Wellcome Sanger Institute for the COVID-19 Genomics UK Consortium |
| EPI_ISL_438476 | Department of Pathology, University of Cambridge | Wellcome Sanger Institute for the COVID-19 Genomics UK Consortium |
| EPI_ISL_438477 | Department of Pathology, University of Cambridge | Wellcome Sanger Institute for the COVID-19 Genomics UK Consortium |
| EPI_ISL_438478 | Department of Pathology, University of Cambridge | Wellcome Sanger Institute for the COVID-19 Genomics UK Consortium |
| EPI_ISL_438479 | Department of Pathology, University of Cambridge | Wellcome Sanger Institute for the COVID-19 Genomics UK Consortium |
| EPI_ISL_438480 | Department of Pathology, University of Cambridge | Wellcome Sanger Institute for the COVID-19 Genomics UK Consortium |
| EPI_ISL_438481 | Department of Pathology, University of Cambridge | Wellcome Sanger Institute for the COVID-19 Genomics UK Consortium |

| EPI_Accession_ID | Originating Laboratory | Submitting Laboratory |
|---|---|---|
| EPI_ISL_438482 | Department of Pathology, University of Cambridge | Wellcome Sanger Institute for the COVID-19 Genomics UK Consortium |
| EPI_ISL_438483 | Department of Pathology, University of Cambridge | Wellcome Sanger Institute for the COVID-19 Genomics UK Consortium |
| EPI_ISL_438484 | Department of Pathology, University of Cambridge | Wellcome Sanger Institute for the COVID-19 Genomics UK Consortium |
| EPI_ISL_438485 | Department of Pathology, University of Cambridge | Wellcome Sanger Institute for the COVID-19 Genomics UK Consortium |
| EPI_ISL_438486 | Department of Pathology, University of Cambridge | Wellcome Sanger Institute for the COVID-19 Genomics UK Consortium |
| EPI_ISL_438487 | Department of Pathology, University of Cambridge | Wellcome Sanger Institute for the COVID-19 Genomics UK Consortium |
| EPI_ISL_438488 | Department of Pathology, University of Cambridge | Wellcome Sanger Institute for the COVID-19 Genomics UK Consortium |
| EPI_ISL_438489 | Department of Pathology, University of Cambridge | Wellcome Sanger Institute for the COVID-19 Genomics UK Consortium |
| EPI_ISL_438490 | Department of Pathology, University of Cambridge | Wellcome Sanger Institute for the COVID-19 Genomics UK Consortium |
| EPI_ISL_438491 | Department of Pathology, University of Cambridge | Wellcome Sanger Institute for the COVID-19 Genomics UK Consortium |
| EPI_ISL_438492 | Department of Pathology, University of Cambridge | Wellcome Sanger Institute for the COVID-19 Genomics UK Consortium |
| EPI_ISL_438493 | Department of Pathology, University of Cambridge | Wellcome Sanger Institute for the COVID-19 Genomics UK Consortium |
| EPI_ISL_438494 | Department of Pathology, University of Cambridge | Wellcome Sanger Institute for the COVID-19 Genomics UK Consortium |
| EPI_ISL_438495 | Department of Pathology, University of Cambridge | Wellcome Sanger Institute for the COVID-19 Genomics UK Consortium |
| EPI_ISL_438496 | Department of Pathology, University of Cambridge | Wellcome Sanger Institute for the COVID-19 Genomics UK Consortium |
| EPI_ISL_438497 | Department of Pathology, University of Cambridge | Wellcome Sanger Institute for the COVID-19 Genomics UK Consortium |

| EPI_Accession_ID | Originating Laboratory | Submitting Laboratory |
| --- | --- | --- |
| EPI_ISL_438498 | Department of Pathology, University of Cambridge | Wellcome Sanger Institute for the COVID-19 Genomics UK Consortium |
| EPI_ISL_438499 | Department of Pathology, University of Cambridge | Wellcome Sanger Institute for the COVID-19 Genomics UK Consortium |
| EPI_ISL_438500 | Department of Pathology, University of Cambridge | Wellcome Sanger Institute for the COVID-19 Genomics UK Consortium |
| EPI_ISL_438501 | Department of Pathology, University of Cambridge | Wellcome Sanger Institute for the COVID-19 Genomics UK Consortium |
| EPI_ISL_438502 | Department of Pathology, University of Cambridge | Wellcome Sanger Institute for the COVID-19 Genomics UK Consortium |
| EPI_ISL_438503 | Department of Pathology, University of Cambridge | Wellcome Sanger Institute for the COVID-19 Genomics UK Consortium |
| EPI_ISL_438504 | Department of Pathology, University of Cambridge | Wellcome Sanger Institute for the COVID-19 Genomics UK Consortium |
| EPI_ISL_438505 | Department of Pathology, University of Cambridge | Wellcome Sanger Institute for the COVID-19 Genomics UK Consortium |
| EPI_ISL_438506 | Department of Pathology, University of Cambridge | Wellcome Sanger Institute for the COVID-19 Genomics UK Consortium |
| EPI_ISL_438507 | Department of Pathology, University of Cambridge | Wellcome Sanger Institute for the COVID-19 Genomics UK Consortium |
| EPI_ISL_438508 | Department of Pathology, University of Cambridge | Wellcome Sanger Institute for the COVID-19 Genomics UK Consortium |
| EPI_ISL_438509 | Department of Pathology, University of Cambridge | Wellcome Sanger Institute for the COVID-19 Genomics UK Consortium |
| EPI_ISL_438510 | Department of Pathology, University of Cambridge | Wellcome Sanger Institute for the COVID-19 Genomics UK Consortium |
| EPI_ISL_438511 | Department of Pathology, University of Cambridge | Wellcome Sanger Institute for the COVID-19 Genomics UK Consortium |
| EPI_ISL_438512 | Department of Pathology, University of Cambridge | Wellcome Sanger Institute for the COVID-19 Genomics UK Consortium |
| EPI_ISL_438513 | Department of Pathology, University of Cambridge | Wellcome Sanger Institute for the COVID-19 Genomics UK Consortium |

| EPI_Accession_ID | Originating Laboratory | Submitting Laboratory |
|---|---|---|
| EPI_ISL_438514 | Department of Pathology, University of Cambridge | Wellcome Sanger Institute for the COVID-19 Genomics UK Consortium |
| EPI_ISL_438515 | Department of Pathology, University of Cambridge | Wellcome Sanger Institute for the COVID-19 Genomics UK Consortium |
| EPI_ISL_438516 | Department of Pathology, University of Cambridge | Wellcome Sanger Institute for the COVID-19 Genomics UK Consortium |
| EPI_ISL_438517 | Department of Pathology, University of Cambridge | Wellcome Sanger Institute for the COVID-19 Genomics UK Consortium |
| EPI_ISL_438518 | Department of Pathology, University of Cambridge | Wellcome Sanger Institute for the COVID-19 Genomics UK Consortium |
| EPI_ISL_438519 | Department of Pathology, University of Cambridge | Wellcome Sanger Institute for the COVID-19 Genomics UK Consortium |
| EPI_ISL_438520 | Department of Pathology, University of Cambridge | Wellcome Sanger Institute for the COVID-19 Genomics UK Consortium |
| EPI_ISL_438521 | Department of Pathology, University of Cambridge | Wellcome Sanger Institute for the COVID-19 Genomics UK Consortium |
| EPI_ISL_438522 | Department of Pathology, University of Cambridge | Wellcome Sanger Institute for the COVID-19 Genomics UK Consortium |
| EPI_ISL_438523 | Department of Pathology, University of Cambridge | Wellcome Sanger Institute for the COVID-19 Genomics UK Consortium |
| EPI_ISL_438524 | Department of Pathology, University of Cambridge | Wellcome Sanger Institute for the COVID-19 Genomics UK Consortium |
| EPI_ISL_438525 | Department of Pathology, University of Cambridge | Wellcome Sanger Institute for the COVID-19 Genomics UK Consortium |
| EPI_ISL_438526 | Department of Pathology, University of Cambridge | Wellcome Sanger Institute for the COVID-19 Genomics UK Consortium |
| EPI_ISL_438527 | Department of Pathology, University of Cambridge | Wellcome Sanger Institute for the COVID-19 Genomics UK Consortium |
| EPI_ISL_438528 | Department of Pathology, University of Cambridge | Wellcome Sanger Institute for the COVID-19 Genomics UK Consortium |
| EPI_ISL_438529 | Department of Pathology, University of Cambridge | Wellcome Sanger Institute for the COVID-19 Genomics UK Consortium |

| EPI_Accession_ID | Originating Laboratory | Submitting Laboratory |
|---|---|---|
| EPI_ISL_438530 | Department of Pathology, University of Cambridge | Wellcome Sanger Institute for the COVID-19 Genomics UK Consortium |
| EPI_ISL_438531 | Department of Pathology, University of Cambridge | Wellcome Sanger Institute for the COVID-19 Genomics UK Consortium |
| EPI_ISL_438532 | Department of Pathology, University of Cambridge | Wellcome Sanger Institute for the COVID-19 Genomics UK Consortium |
| EPI_ISL_438533 | Department of Pathology, University of Cambridge | Wellcome Sanger Institute for the COVID-19 Genomics UK Consortium |
| EPI_ISL_438534 | Department of Pathology, University of Cambridge | Wellcome Sanger Institute for the COVID-19 Genomics UK Consortium |
| EPI_ISL_438535 | Department of Pathology, University of Cambridge | Wellcome Sanger Institute for the COVID-19 Genomics UK Consortium |
| EPI_ISL_438536 | Department of Pathology, University of Cambridge | Wellcome Sanger Institute for the COVID-19 Genomics UK Consortium |
| EPI_ISL_438537 | Department of Pathology, University of Cambridge | Wellcome Sanger Institute for the COVID-19 Genomics UK Consortium |
| EPI_ISL_438538 | Department of Pathology, University of Cambridge | Wellcome Sanger Institute for the COVID-19 Genomics UK Consortium |
| EPI_ISL_438539 | Department of Pathology, University of Cambridge | Wellcome Sanger Institute for the COVID-19 Genomics UK Consortium |
| EPI_ISL_438540 | Department of Pathology, University of Cambridge | Wellcome Sanger Institute for the COVID-19 Genomics UK Consortium |
| EPI_ISL_438541 | Department of Pathology, University of Cambridge | Wellcome Sanger Institute for the COVID-19 Genomics UK Consortium |
| EPI_ISL_438542 | Department of Pathology, University of Cambridge | Wellcome Sanger Institute for the COVID-19 Genomics UK Consortium |
| EPI_ISL_438543 | Department of Pathology, University of Cambridge | Wellcome Sanger Institute for the COVID-19 Genomics UK Consortium |
| EPI_ISL_438544 | Department of Pathology, University of Cambridge | Wellcome Sanger Institute for the COVID-19 Genomics UK Consortium |
| EPI_ISL_438545 | Department of Pathology, University of Cambridge | Wellcome Sanger Institute for the COVID-19 Genomics UK Consortium |
| EPI_ISL_438550 | Department of Pathology, University of Cambridge | COVID-19 Genomics UK (COG-UK) Consortium |

| EPI_Accession_ID | Originating Laboratory | Submitting Laboratory |
|---|---|---|
| EPI_ISL_438551 | Department of Pathology, University of Cambridge | COVID-19 Genomics UK (COG-UK) Consortium |
| EPI_ISL_438552 | Department of Pathology, University of Cambridge | COVID-19 Genomics UK (COG-UK) Consortium |
| EPI_ISL_438553 | Department of Pathology, University of Cambridge | COVID-19 Genomics UK (COG-UK) Consortium |
| EPI_ISL_438554 | Department of Pathology, University of Cambridge | COVID-19 Genomics UK (COG-UK) Consortium |
| EPI_ISL_438555 | Department of Pathology, University of Cambridge | COVID-19 Genomics UK (COG-UK) Consortium |
| EPI_ISL_438556 | Department of Pathology, University of Cambridge | COVID-19 Genomics UK (COG-UK) Consortium |
| EPI_ISL_438557 | Department of Pathology, University of Cambridge | COVID-19 Genomics UK (COG-UK) Consortium |
| EPI_ISL_438558 | Department of Pathology, University of Cambridge | COVID-19 Genomics UK (COG-UK) Consortium |
| EPI_ISL_438559 | Department of Pathology, University of Cambridge | COVID-19 Genomics UK (COG-UK) Consortium |
| EPI_ISL_438560 | Department of Pathology, University of Cambridge | COVID-19 Genomics UK (COG-UK) Consortium |
| EPI_ISL_438561 | Department of Pathology, University of Cambridge | COVID-19 Genomics UK (COG-UK) Consortium |
| EPI_ISL_438562 | Department of Pathology, University of Cambridge | COVID-19 Genomics UK (COG-UK) Consortium |
| EPI_ISL_438563 | Department of Pathology, University of Cambridge | COVID-19 Genomics UK (COG-UK) Consortium |
| EPI_ISL_438564 | Department of Pathology, University of Cambridge | COVID-19 Genomics UK (COG-UK) Consortium |
| EPI_ISL_438565 | Department of Pathology, University of Cambridge | COVID-19 Genomics UK (COG-UK) Consortium |
| EPI_ISL_438566 | Department of Pathology, University of Cambridge | COVID-19 Genomics UK (COG-UK) Consortium |
| EPI_ISL_438567 | Department of Pathology, University of Cambridge | COVID-19 Genomics UK (COG-UK) Consortium |
| EPI_ISL_438568 | Department of Pathology, University of Cambridge | COVID-19 Genomics UK (COG-UK) Consortium |
| EPI_ISL_438569 | Department of Pathology, University of Cambridge | COVID-19 Genomics UK (COG-UK) Consortium |
| EPI_ISL_438570 | Department of Pathology, University of Cambridge | COVID-19 Genomics UK (COG-UK) Consortium |
| EPI_ISL_438571 | Department of Pathology, University of Cambridge | COVID-19 Genomics UK (COG-UK) Consortium |
| EPI_ISL_438573 | Department of Pathology, University of Cambridge | COVID-19 Genomics UK (COG-UK) Consortium |
| EPI_ISL_438574 | Department of Pathology, University of Cambridge | COVID-19 Genomics UK (COG-UK) Consortium |
| EPI_ISL_438575 | Department of Pathology, University of Cambridge | COVID-19 Genomics UK (COG-UK) Consortium |
| EPI_ISL_438576 | Department of Pathology, University of Cambridge | COVID-19 Genomics UK (COG-UK) Consortium |
| EPI_ISL_438577 | Department of Pathology, University of Cambridge | COVID-19 Genomics UK (COG-UK) Consortium |
| EPI_ISL_438578 | Department of Pathology, University of Cambridge | COVID-19 Genomics UK (COG-UK) Consortium |
| EPI_ISL_438579 | Department of Pathology, University of Cambridge | COVID-19 Genomics UK (COG-UK) Consortium |
| EPI_ISL_438580 | Department of Pathology, University of Cambridge | COVID-19 Genomics UK (COG-UK) Consortium |
| EPI_ISL_438581 | Department of Pathology, University of Cambridge | COVID-19 Genomics UK (COG-UK) Consortium |
| EPI_ISL_438582 | Department of Pathology, University of Cambridge | COVID-19 Genomics UK (COG-UK) Consortium |
| EPI_ISL_438583 | Department of Pathology, University of Cambridge | COVID-19 Genomics UK (COG-UK) Consortium |
| EPI_ISL_438584 | Department of Pathology, University of Cambridge | COVID-19 Genomics UK (COG-UK) Consortium |

| EPI_Accession_ID | Originating Laboratory | Submitting Laboratory |
|---|---|---|
| EPI_ISL_438585 | Department of Pathology, University of Cambridge | COVID-19 Genomics UK (COG-UK) Consortium |
| EPI_ISL_438586 | Department of Pathology, University of Cambridge | COVID-19 Genomics UK (COG-UK) Consortium |
| EPI_ISL_438587 | Department of Pathology, University of Cambridge | COVID-19 Genomics UK (COG-UK) Consortium |
| EPI_ISL_438588 | Department of Pathology, University of Cambridge | COVID-19 Genomics UK (COG-UK) Consortium |
| EPI_ISL_438589 | Department of Pathology, University of Cambridge | COVID-19 Genomics UK (COG-UK) Consortium |
| EPI_ISL_438590 | Department of Pathology, University of Cambridge | COVID-19 Genomics UK (COG-UK) Consortium |
| EPI_ISL_438591 | Department of Pathology, University of Cambridge | COVID-19 Genomics UK (COG-UK) Consortium |
| EPI_ISL_438592 | Department of Pathology, University of Cambridge | COVID-19 Genomics UK (COG-UK) Consortium |
| EPI_ISL_438593 | Department of Pathology, University of Cambridge | COVID-19 Genomics UK (COG-UK) Consortium |
| EPI_ISL_438594 | Department of Pathology, University of Cambridge | COVID-19 Genomics UK (COG-UK) Consortium |
| EPI_ISL_438595 | Department of Pathology, University of Cambridge | COVID-19 Genomics UK (COG-UK) Consortium |
| EPI_ISL_438596 | Department of Pathology, University of Cambridge | COVID-19 Genomics UK (COG-UK) Consortium |
| EPI_ISL_438597 | Department of Pathology, University of Cambridge | COVID-19 Genomics UK (COG-UK) Consortium |
| EPI_ISL_438598 | Department of Pathology, University of Cambridge | COVID-19 Genomics UK (COG-UK) Consortium |
| EPI_ISL_438599 | Department of Pathology, University of Cambridge | COVID-19 Genomics UK (COG-UK) Consortium |
| EPI_ISL_438600 | Department of Pathology, University of Cambridge | COVID-19 Genomics UK (COG-UK) Consortium |
| EPI_ISL_438601 | Department of Pathology, University of Cambridge | COVID-19 Genomics UK (COG-UK) Consortium |
| EPI_ISL_438602 | Department of Pathology, University of Cambridge | COVID-19 Genomics UK (COG-UK) Consortium |
| EPI_ISL_438603 | Department of Pathology, University of Cambridge | COVID-19 Genomics UK (COG-UK) Consortium |
| EPI_ISL_438604 | Department of Pathology, University of Cambridge | COVID-19 Genomics UK (COG-UK) Consortium |
| EPI_ISL_438605 | Department of Pathology, University of Cambridge | COVID-19 Genomics UK (COG-UK) Consortium |
| EPI_ISL_438606 | Department of Pathology, University of Cambridge | COVID-19 Genomics UK (COG-UK) Consortium |
| EPI_ISL_438607 | Department of Pathology, University of Cambridge | COVID-19 Genomics UK (COG-UK) Consortium |
| EPI_ISL_438608 | Department of Pathology, University of Cambridge | COVID-19 Genomics UK (COG-UK) Consortium |
| EPI_ISL_438609 | Department of Pathology, University of Cambridge | COVID-19 Genomics UK (COG-UK) Consortium |
| EPI_ISL_438610 | Department of Pathology, University of Cambridge | COVID-19 Genomics UK (COG-UK) Consortium |
| EPI_ISL_438612 | Department of Pathology, University of Cambridge | COVID-19 Genomics UK (COG-UK) Consortium |
| EPI_ISL_438613 | Department of Pathology, University of Cambridge | COVID-19 Genomics UK (COG-UK) Consortium |
| EPI_ISL_438614 | Department of Pathology, University of Cambridge | COVID-19 Genomics UK (COG-UK) Consortium |
| EPI_ISL_438615 | Department of Pathology, University of Cambridge | COVID-19 Genomics UK (COG-UK) Consortium |
| EPI_ISL_438616 | Department of Pathology, University of Cambridge | COVID-19 Genomics UK (COG-UK) Consortium |
| EPI_ISL_438617 | Department of Pathology, University of Cambridge | COVID-19 Genomics UK (COG-UK) Consortium |
| EPI_ISL_438618 | Department of Pathology, University of Cambridge | COVID-19 Genomics UK (COG-UK) Consortium |

| EPI_Accession_ID | Originating Laboratory | Submitting Laboratory |
|---|---|---|
| EPI_ISL_438619 | Department of Pathology, University of Cambridge | COVID-19 Genomics UK (COG-UK) Consortium |
| EPI_ISL_438620 | Department of Pathology, University of Cambridge | COVID-19 Genomics UK (COG-UK) Consortium |
| EPI_ISL_438621 | Department of Pathology, University of Cambridge | COVID-19 Genomics UK (COG-UK) Consortium |
| EPI_ISL_438622 | Department of Pathology, University of Cambridge | COVID-19 Genomics UK (COG-UK) Consortium |
| EPI_ISL_438623 | Department of Pathology, University of Cambridge | COVID-19 Genomics UK (COG-UK) Consortium |
| EPI_ISL_438624 | Department of Pathology, University of Cambridge | COVID-19 Genomics UK (COG-UK) Consortium |
| EPI_ISL_438625 | Department of Pathology, University of Cambridge | COVID-19 Genomics UK (COG-UK) Consortium |
| EPI_ISL_438626 | Department of Pathology, University of Cambridge | COVID-19 Genomics UK (COG-UK) Consortium |
| EPI_ISL_438627 | Department of Pathology, University of Cambridge | COVID-19 Genomics UK (COG-UK) Consortium |
| EPI_ISL_438628 | Department of Pathology, University of Cambridge | COVID-19 Genomics UK (COG-UK) Consortium |
| EPI_ISL_438629 | Department of Pathology, University of Cambridge | COVID-19 Genomics UK (COG-UK) Consortium |
| EPI_ISL_438630 | Department of Pathology, University of Cambridge | COVID-19 Genomics UK (COG-UK) Consortium |
| EPI_ISL_438631 | Department of Pathology, University of Cambridge | COVID-19 Genomics UK (COG-UK) Consortium |
| EPI_ISL_438632 | Department of Pathology, University of Cambridge | COVID-19 Genomics UK (COG-UK) Consortium |
| EPI_ISL_438633 | Department of Pathology, University of Cambridge | COVID-19 Genomics UK (COG-UK) Consortium |
| EPI_ISL_438634 | Department of Pathology, University of Cambridge | COVID-19 Genomics UK (COG-UK) Consortium |
| EPI_ISL_438635 | Department of Pathology, University of Cambridge | COVID-19 Genomics UK (COG-UK) Consortium |
| EPI_ISL_438636 | Department of Pathology, University of Cambridge | COVID-19 Genomics UK (COG-UK) Consortium |
| EPI_ISL_438637 | Department of Pathology, University of Cambridge | COVID-19 Genomics UK (COG-UK) Consortium |
| EPI_ISL_438638 | Department of Pathology, University of Cambridge | COVID-19 Genomics UK (COG-UK) Consortium |
| EPI_ISL_438639 | Department of Pathology, University of Cambridge | COVID-19 Genomics UK (COG-UK) Consortium |
| EPI_ISL_438640 | Department of Pathology, University of Cambridge | COVID-19 Genomics UK (COG-UK) Consortium |
| EPI_ISL_438641 | Department of Pathology, University of Cambridge | COVID-19 Genomics UK (COG-UK) Consortium |
| EPI_ISL_438642 | Department of Pathology, University of Cambridge | COVID-19 Genomics UK (COG-UK) Consortium |
| EPI_ISL_438643 | Department of Pathology, University of Cambridge | COVID-19 Genomics UK (COG-UK) Consortium |
| EPI_ISL_438644 | Department of Pathology, University of Cambridge | COVID-19 Genomics UK (COG-UK) Consortium |
| EPI_ISL_438645 | Department of Pathology, University of Cambridge | COVID-19 Genomics UK (COG-UK) Consortium |
| EPI_ISL_438646 | Department of Pathology, University of Cambridge | COVID-19 Genomics UK (COG-UK) Consortium |
| EPI_ISL_438647 | Department of Pathology, University of Cambridge | COVID-19 Genomics UK (COG-UK) Consortium |
| EPI_ISL_438648 | Department of Pathology, University of Cambridge | COVID-19 Genomics UK (COG-UK) Consortium |
| EPI_ISL_438649 | Department of Pathology, University of Cambridge | COVID-19 Genomics UK (COG-UK) Consortium |
| EPI_ISL_438650 | Department of Pathology, University of Cambridge | COVID-19 Genomics UK (COG-UK) Consortium |
| EPI_ISL_438651 | Department of Pathology, University of Cambridge | COVID-19 Genomics UK (COG-UK) Consortium |

| EPI_Accession_ID | Originating Laboratory | Submitting Laboratory |
|---|---|---|
| EPI_ISL_438652 | Department of Pathology, University of Cambridge | COVID-19 Genomics UK (COG-UK) Consortium |
| EPI_ISL_438653 | Department of Pathology, University of Cambridge | COVID-19 Genomics UK (COG-UK) Consortium |
| EPI_ISL_438654 | Department of Pathology, University of Cambridge | COVID-19 Genomics UK (COG-UK) Consortium |
| EPI_ISL_438656 | Department of Pathology, University of Cambridge | COVID-19 Genomics UK (COG-UK) Consortium |
| EPI_ISL_438657 | Department of Pathology, University of Cambridge | COVID-19 Genomics UK (COG-UK) Consortium |
| EPI_ISL_438658 | Department of Pathology, University of Cambridge | COVID-19 Genomics UK (COG-UK) Consortium |
| EPI_ISL_438659 | Department of Pathology, University of Cambridge | COVID-19 Genomics UK (COG-UK) Consortium |
| EPI_ISL_438660 | Department of Pathology, University of Cambridge | COVID-19 Genomics UK (COG-UK) Consortium |
| EPI_ISL_438661 | Department of Pathology, University of Cambridge | COVID-19 Genomics UK (COG-UK) Consortium |
| EPI_ISL_438662 | Department of Pathology, University of Cambridge | COVID-19 Genomics UK (COG-UK) Consortium |
| EPI_ISL_438663 | Department of Pathology, University of Cambridge | COVID-19 Genomics UK (COG-UK) Consortium |
| EPI_ISL_438664 | Department of Pathology, University of Cambridge | COVID-19 Genomics UK (COG-UK) Consortium |
| EPI_ISL_438665 | Department of Pathology, University of Cambridge | COVID-19 Genomics UK (COG-UK) Consortium |
| EPI_ISL_438666 | Department of Pathology, University of Cambridge | COVID-19 Genomics UK (COG-UK) Consortium |
| EPI_ISL_438667 | Department of Pathology, University of Cambridge | COVID-19 Genomics UK (COG-UK) Consortium |
| EPI_ISL_438668 | Department of Pathology, University of Cambridge | COVID-19 Genomics UK (COG-UK) Consortium |
| EPI_ISL_438669 | Department of Pathology, University of Cambridge | COVID-19 Genomics UK (COG-UK) Consortium |
| EPI_ISL_438670 | Department of Pathology, University of Cambridge | COVID-19 Genomics UK (COG-UK) Consortium |
| EPI_ISL_438671 | Department of Pathology, University of Cambridge | COVID-19 Genomics UK (COG-UK) Consortium |
| EPI_ISL_438672 | Department of Pathology, University of Cambridge | COVID-19 Genomics UK (COG-UK) Consortium |
| EPI_ISL_438673 | Department of Pathology, University of Cambridge | COVID-19 Genomics UK (COG-UK) Consortium |
| EPI_ISL_438674 | Department of Pathology, University of Cambridge | COVID-19 Genomics UK (COG-UK) Consortium |
| EPI_ISL_438675 | Department of Pathology, University of Cambridge | COVID-19 Genomics UK (COG-UK) Consortium |
| EPI_ISL_438676 | Department of Pathology, University of Cambridge | COVID-19 Genomics UK (COG-UK) Consortium |
| EPI_ISL_438677 | Department of Pathology, University of Cambridge | COVID-19 Genomics UK (COG-UK) Consortium |
| EPI_ISL_438678 | Department of Pathology, University of Cambridge | COVID-19 Genomics UK (COG-UK) Consortium |
| EPI_ISL_438679 | Department of Pathology, University of Cambridge | COVID-19 Genomics UK (COG-UK) Consortium |
| EPI_ISL_438680 | Department of Pathology, University of Cambridge | COVID-19 Genomics UK (COG-UK) Consortium |
| EPI_ISL_438681 | Department of Pathology, University of Cambridge | COVID-19 Genomics UK (COG-UK) Consortium |
| EPI_ISL_438682 | Department of Pathology, University of Cambridge | COVID-19 Genomics UK (COG-UK) Consortium |
| EPI_ISL_438683 | Department of Pathology, University of Cambridge | COVID-19 Genomics UK (COG-UK) Consortium |
| EPI_ISL_438684 | Department of Pathology, University of Cambridge | COVID-19 Genomics UK (COG-UK) Consortium |
| EPI_ISL_438685 | Department of Pathology, University of Cambridge | COVID-19 Genomics UK (COG-UK) Consortium |

| EPI_Accession_ID | Originating Laboratory | Submitting Laboratory |
|---|---|---|
| EPI_ISL_438686 | Department of Pathology, University of Cambridge | COVID-19 Genomics UK (COG-UK) Consortium |
| EPI_ISL_438687 | Department of Pathology, University of Cambridge | COVID-19 Genomics UK (COG-UK) Consortium |
| EPI_ISL_438689 | Department of Pathology, University of Cambridge | COVID-19 Genomics UK (COG-UK) Consortium |
| EPI_ISL_438690 | Department of Pathology, University of Cambridge | COVID-19 Genomics UK (COG-UK) Consortium |
| EPI_ISL_438691 | Department of Pathology, University of Cambridge | COVID-19 Genomics UK (COG-UK) Consortium |
| EPI_ISL_438692 | Department of Pathology, University of Cambridge | COVID-19 Genomics UK (COG-UK) Consortium |
| EPI_ISL_438693 | Department of Pathology, University of Cambridge | COVID-19 Genomics UK (COG-UK) Consortium |
| EPI_ISL_438694 | Department of Pathology, University of Cambridge | COVID-19 Genomics UK (COG-UK) Consortium |
| EPI_ISL_438695 | Department of Pathology, University of Cambridge | COVID-19 Genomics UK (COG-UK) Consortium |
| EPI_ISL_438696 | Department of Pathology, University of Cambridge | COVID-19 Genomics UK (COG-UK) Consortium |
| EPI_ISL_438697 | Department of Pathology, University of Cambridge | COVID-19 Genomics UK (COG-UK) Consortium |
| EPI_ISL_438698 | Department of Pathology, University of Cambridge | COVID-19 Genomics UK (COG-UK) Consortium |
| EPI_ISL_438699 | Department of Pathology, University of Cambridge | COVID-19 Genomics UK (COG-UK) Consortium |
| EPI_ISL_438700 | Department of Pathology, University of Cambridge | COVID-19 Genomics UK (COG-UK) Consortium |
| EPI_ISL_438701 | Department of Pathology, University of Cambridge | COVID-19 Genomics UK (COG-UK) Consortium |
| EPI_ISL_438702 | Department of Pathology, University of Cambridge | COVID-19 Genomics UK (COG-UK) Consortium |
| EPI_ISL_438703 | Department of Pathology, University of Cambridge | COVID-19 Genomics UK (COG-UK) Consortium |
| EPI_ISL_438704 | Department of Pathology, University of Cambridge | COVID-19 Genomics UK (COG-UK) Consortium |
| EPI_ISL_438705 | Department of Pathology, University of Cambridge | COVID-19 Genomics UK (COG-UK) Consortium |
| EPI_ISL_438706 | Department of Pathology, University of Cambridge | COVID-19 Genomics UK (COG-UK) Consortium |
| EPI_ISL_438707 | Department of Pathology, University of Cambridge | COVID-19 Genomics UK (COG-UK) Consortium |
| EPI_ISL_438708 | Department of Pathology, University of Cambridge | COVID-19 Genomics UK (COG-UK) Consortium |
| EPI_ISL_438709 | Department of Pathology, University of Cambridge | COVID-19 Genomics UK (COG-UK) Consortium |
| EPI_ISL_438710 | Department of Pathology, University of Cambridge | COVID-19 Genomics UK (COG-UK) Consortium |
| EPI_ISL_438711 | Department of Pathology, University of Cambridge | COVID-19 Genomics UK (COG-UK) Consortium |
| EPI_ISL_438712 | Department of Pathology, University of Cambridge | COVID-19 Genomics UK (COG-UK) Consortium |
| EPI_ISL_438713 | Department of Pathology, University of Cambridge | COVID-19 Genomics UK (COG-UK) Consortium |
| EPI_ISL_438714 | Department of Pathology, University of Cambridge | COVID-19 Genomics UK (COG-UK) Consortium |
| EPI_ISL_438715 | Department of Pathology, University of Cambridge | COVID-19 Genomics UK (COG-UK) Consortium |
| EPI_ISL_438716 | Department of Pathology, University of Cambridge | COVID-19 Genomics UK (COG-UK) Consortium |
| EPI_ISL_438717 | Department of Pathology, University of Cambridge | COVID-19 Genomics UK (COG-UK) Consortium |
| EPI_ISL_438718 | Department of Pathology, University of Cambridge | COVID-19 Genomics UK (COG-UK) Consortium |
| EPI_ISL_438719 | Department of Pathology, University of Cambridge | COVID-19 Genomics UK (COG-UK) Consortium |

| EPI_Accession_ID | Originating Laboratory | Submitting Laboratory |
| --- | --- | --- |
| EPI_ISL_438720 | Department of Pathology, University of Cambridge | COVID-19 Genomics UK (COG-UK) Consortium |
| EPI_ISL_438721 | Department of Pathology, University of Cambridge | COVID-19 Genomics UK (COG-UK) Consortium |
| EPI_ISL_438722 | Department of Pathology, University of Cambridge | COVID-19 Genomics UK (COG-UK) Consortium |
| EPI_ISL_438723 | Department of Pathology, University of Cambridge | COVID-19 Genomics UK (COG-UK) Consortium |
| EPI_ISL_438724 | Department of Pathology, University of Cambridge | COVID-19 Genomics UK (COG-UK) Consortium |
| EPI_ISL_438725 | Department of Pathology, University of Cambridge | COVID-19 Genomics UK (COG-UK) Consortium |
| EPI_ISL_438726 | Department of Pathology, University of Cambridge | COVID-19 Genomics UK (COG-UK) Consortium |
| EPI_ISL_438727 | Department of Pathology, University of Cambridge | COVID-19 Genomics UK (COG-UK) Consortium |
| EPI_ISL_438728 | Department of Pathology, University of Cambridge | COVID-19 Genomics UK (COG-UK) Consortium |
| EPI_ISL_438729 | Department of Pathology, University of Cambridge | COVID-19 Genomics UK (COG-UK) Consortium |
| EPI_ISL_438730 | Department of Pathology, University of Cambridge | COVID-19 Genomics UK (COG-UK) Consortium |
| EPI_ISL_438731 | Department of Pathology, University of Cambridge | COVID-19 Genomics UK (COG-UK) Consortium |
| EPI_ISL_438732 | Department of Pathology, University of Cambridge | COVID-19 Genomics UK (COG-UK) Consortium |
| EPI_ISL_438733 | Department of Pathology, University of Cambridge | COVID-19 Genomics UK (COG-UK) Consortium |
| EPI_ISL_438734 | Department of Pathology, University of Cambridge | COVID-19 Genomics UK (COG-UK) Consortium |
| EPI_ISL_438735 | Department of Pathology, University of Cambridge | COVID-19 Genomics UK (COG-UK) Consortium |
| EPI_ISL_438736 | Department of Pathology, University of Cambridge | COVID-19 Genomics UK (COG-UK) Consortium |
| EPI_ISL_438737 | Department of Pathology, University of Cambridge | COVID-19 Genomics UK (COG-UK) Consortium |
| EPI_ISL_438738 | Department of Pathology, University of Cambridge | COVID-19 Genomics UK (COG-UK) Consortium |
| EPI_ISL_438739 | Department of Pathology, University of Cambridge | COVID-19 Genomics UK (COG-UK) Consortium |
| EPI_ISL_438740 | Department of Pathology, University of Cambridge | COVID-19 Genomics UK (COG-UK) Consortium |
| EPI_ISL_438741 | Department of Pathology, University of Cambridge | COVID-19 Genomics UK (COG-UK) Consortium |
| EPI_ISL_438742 | Department of Pathology, University of Cambridge | COVID-19 Genomics UK (COG-UK) Consortium |
| EPI_ISL_438743 | Department of Pathology, University of Cambridge | COVID-19 Genomics UK (COG-UK) Consortium |
| EPI_ISL_438744 | Department of Pathology, University of Cambridge | COVID-19 Genomics UK (COG-UK) Consortium |
| EPI_ISL_438745 | Department of Pathology, University of Cambridge | COVID-19 Genomics UK (COG-UK) Consortium |
| EPI_ISL_438746 | Department of Pathology, University of Cambridge | COVID-19 Genomics UK (COG-UK) Consortium |
| EPI_ISL_438747 | Department of Pathology, University of Cambridge | COVID-19 Genomics UK (COG-UK) Consortium |
| EPI_ISL_438748 | Department of Pathology, University of Cambridge | COVID-19 Genomics UK (COG-UK) Consortium |
| EPI_ISL_438749 | West of Scotland Specialist Virology Centre, NHSGGC / MRC-University of Glasgow Centre for Virus Research | COVID-19 Genomics UK (COG-UK) Consortium |
| EPI_ISL_438750 | West of Scotland Specialist Virology Centre, NHSGGC / MRC-University of Glasgow Centre for Virus Research | COVID-19 Genomics UK (COG-UK) Consortium |

| EPI_Accession_ID | Originating Laboratory | Submitting Laboratory |
|---|---|---|
| EPI_ISL_438751 | West of Scotland Specialist Virology Centre, NHSGGC / MRC-University of Glasgow Centre for Virus Research | COVID-19 Genomics UK (COG-UK) Consortium |
| EPI_ISL_438752 | West of Scotland Specialist Virology Centre, NHSGGC / MRC-University of Glasgow Centre for Virus Research | COVID-19 Genomics UK (COG-UK) Consortium |
| EPI_ISL_438753 | West of Scotland Specialist Virology Centre, NHSGGC / MRC-University of Glasgow Centre for Virus Research | COVID-19 Genomics UK (COG-UK) Consortium |
| EPI_ISL_438754 | West of Scotland Specialist Virology Centre, NHSGGC / MRC-University of Glasgow Centre for Virus Research | COVID-19 Genomics UK (COG-UK) Consortium |
| EPI_ISL_438755 | West of Scotland Specialist Virology Centre, NHSGGC / MRC-University of Glasgow Centre for Virus Research | COVID-19 Genomics UK (COG-UK) Consortium |
| EPI_ISL_438756 | West of Scotland Specialist Virology Centre, NHSGGC / MRC-University of Glasgow Centre for Virus Research | COVID-19 Genomics UK (COG-UK) Consortium |
| EPI_ISL_438757 | West of Scotland Specialist Virology Centre, NHSGGC / MRC-University of Glasgow Centre for Virus Research | COVID-19 Genomics UK (COG-UK) Consortium |
| EPI_ISL_438758 | West of Scotland Specialist Virology Centre, NHSGGC / MRC-University of Glasgow Centre for Virus Research | COVID-19 Genomics UK (COG-UK) Consortium |
| EPI_ISL_438759 | West of Scotland Specialist Virology Centre, NHSGGC / MRC-University of Glasgow Centre for Virus Research | COVID-19 Genomics UK (COG-UK) Consortium |
| EPI_ISL_438760 | West of Scotland Specialist Virology Centre, NHSGGC / MRC-University of Glasgow Centre for Virus Research | COVID-19 Genomics UK (COG-UK) Consortium |
| EPI_ISL_438761 | West of Scotland Specialist Virology Centre, NHSGGC / MRC-University of Glasgow Centre for Virus Research | COVID-19 Genomics UK (COG-UK) Consortium |
| EPI_ISL_438762 | West of Scotland Specialist Virology Centre, NHSGGC / MRC-University of Glasgow Centre for Virus Research | COVID-19 Genomics UK (COG-UK) Consortium |
| EPI_ISL_438763 | West of Scotland Specialist Virology Centre, NHSGGC / MRC-University of Glasgow Centre for Virus Research | COVID-19 Genomics UK (COG-UK) Consortium |
| EPI_ISL_438764 | West of Scotland Specialist Virology Centre, NHSGGC / MRC-University of Glasgow Centre for Virus Research | COVID-19 Genomics UK (COG-UK) Consortium |
| EPI_ISL_438765 | West of Scotland Specialist Virology Centre, NHSGGC / MRC-University of Glasgow Centre for Virus Research | COVID-19 Genomics UK (COG-UK) Consortium |
| EPI_ISL_438766 | West of Scotland Specialist Virology Centre, NHSGGC / MRC-University of Glasgow Centre for Virus Research | COVID-19 Genomics UK (COG-UK) Consortium |

| EPI_Accession_ID | Originating Laboratory | Submitting Laboratory |
|---|---|---|
| EPI_ISL_438767 | West of Scotland Specialist Virology Centre, NHSGGC / MRC-University of Glasgow Centre for Virus Research | COVID-19 Genomics UK (COG-UK) Consortium |
| EPI_ISL_438768 | West of Scotland Specialist Virology Centre, NHSGGC / MRC-University of Glasgow Centre for Virus Research | COVID-19 Genomics UK (COG-UK) Consortium |
| EPI_ISL_438769 | West of Scotland Specialist Virology Centre, NHSGGC / MRC-University of Glasgow Centre for Virus Research | COVID-19 Genomics UK (COG-UK) Consortium |
| EPI_ISL_438770 | West of Scotland Specialist Virology Centre, NHSGGC / MRC-University of Glasgow Centre for Virus Research | COVID-19 Genomics UK (COG-UK) Consortium |
| EPI_ISL_438771 | West of Scotland Specialist Virology Centre, NHSGGC / MRC-University of Glasgow Centre for Virus Research | COVID-19 Genomics UK (COG-UK) Consortium |
| EPI_ISL_438772 | West of Scotland Specialist Virology Centre, NHSGGC / MRC-University of Glasgow Centre for Virus Research | COVID-19 Genomics UK (COG-UK) Consortium |
| EPI_ISL_438773 | West of Scotland Specialist Virology Centre, NHSGGC / MRC-University of Glasgow Centre for Virus Research | COVID-19 Genomics UK (COG-UK) Consortium |
| EPI_ISL_438774 | West of Scotland Specialist Virology Centre, NHSGGC / MRC-University of Glasgow Centre for Virus Research | COVID-19 Genomics UK (COG-UK) Consortium |
| EPI_ISL_438775 | West of Scotland Specialist Virology Centre, NHSGGC / MRC-University of Glasgow Centre for Virus Research | COVID-19 Genomics UK (COG-UK) Consortium |
| EPI_ISL_438776 | West of Scotland Specialist Virology Centre, NHSGGC / MRC-University of Glasgow Centre for Virus Research | COVID-19 Genomics UK (COG-UK) Consortium |
| EPI_ISL_438777 | West of Scotland Specialist Virology Centre, NHSGGC / MRC-University of Glasgow Centre for Virus Research | COVID-19 Genomics UK (COG-UK) Consortium |
| EPI_ISL_438778 | West of Scotland Specialist Virology Centre, NHSGGC / MRC-University of Glasgow Centre for Virus Research | COVID-19 Genomics UK (COG-UK) Consortium |
| EPI_ISL_438779 | West of Scotland Specialist Virology Centre, NHSGGC / MRC-University of Glasgow Centre for Virus Research | COVID-19 Genomics UK (COG-UK) Consortium |
| EPI_ISL_438780 | West of Scotland Specialist Virology Centre, NHSGGC / MRC-University of Glasgow Centre for Virus Research | COVID-19 Genomics UK (COG-UK) Consortium |
| EPI_ISL_438781 | West of Scotland Specialist Virology Centre, NHSGGC / MRC-University of Glasgow Centre for Virus Research | COVID-19 Genomics UK (COG-UK) Consortium |
| EPI_ISL_438782 | West of Scotland Specialist Virology Centre, NHSGGC / MRC-University of Glasgow Centre for Virus Research | COVID-19 Genomics UK (COG-UK) Consortium |

| EPI_Accession_ID | Originating Laboratory | Submitting Laboratory |
|---|---|---|
| EPI_ISL_438783 | West of Scotland Specialist Virology Centre, NHSGGC / MRC-University of Glasgow Centre for Virus Research | COVID-19 Genomics UK (COG-UK) Consortium |
| EPI_ISL_438784 | West of Scotland Specialist Virology Centre, NHSGGC / MRC-University of Glasgow Centre for Virus Research | COVID-19 Genomics UK (COG-UK) Consortium |
| EPI_ISL_438785 | West of Scotland Specialist Virology Centre, NHSGGC / MRC-University of Glasgow Centre for Virus Research | COVID-19 Genomics UK (COG-UK) Consortium |
| EPI_ISL_438786 | West of Scotland Specialist Virology Centre, NHSGGC / MRC-University of Glasgow Centre for Virus Research | COVID-19 Genomics UK (COG-UK) Consortium |
| EPI_ISL_438787 | West of Scotland Specialist Virology Centre, NHSGGC / MRC-University of Glasgow Centre for Virus Research | COVID-19 Genomics UK (COG-UK) Consortium |
| EPI_ISL_438788 | West of Scotland Specialist Virology Centre, NHSGGC / MRC-University of Glasgow Centre for Virus Research | COVID-19 Genomics UK (COG-UK) Consortium |
| EPI_ISL_438789 | West of Scotland Specialist Virology Centre, NHSGGC / MRC-University of Glasgow Centre for Virus Research | COVID-19 Genomics UK (COG-UK) Consortium |
| EPI_ISL_438790 | West of Scotland Specialist Virology Centre, NHSGGC / MRC-University of Glasgow Centre for Virus Research | COVID-19 Genomics UK (COG-UK) Consortium |
| EPI_ISL_438791 | West of Scotland Specialist Virology Centre, NHSGGC / MRC-University of Glasgow Centre for Virus Research | COVID-19 Genomics UK (COG-UK) Consortium |
| EPI_ISL_438792 | West of Scotland Specialist Virology Centre, NHSGGC / MRC-University of Glasgow Centre for Virus Research | COVID-19 Genomics UK (COG-UK) Consortium |
| EPI_ISL_438793 | West of Scotland Specialist Virology Centre, NHSGGC / MRC-University of Glasgow Centre for Virus Research | COVID-19 Genomics UK (COG-UK) Consortium |
| EPI_ISL_438794 | West of Scotland Specialist Virology Centre, NHSGGC / MRC-University of Glasgow Centre for Virus Research | COVID-19 Genomics UK (COG-UK) Consortium |
| EPI_ISL_438795 | West of Scotland Specialist Virology Centre, NHSGGC / MRC-University of Glasgow Centre for Virus Research | COVID-19 Genomics UK (COG-UK) Consortium |
| EPI_ISL_438796 | West of Scotland Specialist Virology Centre, NHSGGC / MRC-University of Glasgow Centre for Virus Research | COVID-19 Genomics UK (COG-UK) Consortium |
| EPI_ISL_438797 | West of Scotland Specialist Virology Centre, NHSGGC / MRC-University of Glasgow Centre for Virus Research | COVID-19 Genomics UK (COG-UK) Consortium |
| EPI_ISL_438798 | West of Scotland Specialist Virology Centre, NHSGGC / MRC-University of Glasgow Centre for Virus Research | COVID-19 Genomics UK (COG-UK) Consortium |

| EPI_Accession_ID | Originating Laboratory | Submitting Laboratory |
|---|---|---|
| EPI_ISL_438799 | West of Scotland Specialist Virology Centre, NHSGGC / MRC-University of Glasgow Centre for Virus Research | COVID-19 Genomics UK (COG-UK) Consortium |
| EPI_ISL_438800 | West of Scotland Specialist Virology Centre, NHSGGC / MRC-University of Glasgow Centre for Virus Research | COVID-19 Genomics UK (COG-UK) Consortium |
| EPI_ISL_438801 | West of Scotland Specialist Virology Centre, NHSGGC / MRC-University of Glasgow Centre for Virus Research | COVID-19 Genomics UK (COG-UK) Consortium |
| EPI_ISL_438802 | West of Scotland Specialist Virology Centre, NHSGGC / MRC-University of Glasgow Centre for Virus Research | COVID-19 Genomics UK (COG-UK) Consortium |
| EPI_ISL_438803 | West of Scotland Specialist Virology Centre, NHSGGC / MRC-University of Glasgow Centre for Virus Research | COVID-19 Genomics UK (COG-UK) Consortium |
| EPI_ISL_438804 | West of Scotland Specialist Virology Centre, NHSGGC / MRC-University of Glasgow Centre for Virus Research | COVID-19 Genomics UK (COG-UK) Consortium |
| EPI_ISL_438805 | West of Scotland Specialist Virology Centre, NHSGGC / MRC-University of Glasgow Centre for Virus Research | COVID-19 Genomics UK (COG-UK) Consortium |
| EPI_ISL_438806 | West of Scotland Specialist Virology Centre, NHSGGC / MRC-University of Glasgow Centre for Virus Research | COVID-19 Genomics UK (COG-UK) Consortium |
| EPI_ISL_438807 | West of Scotland Specialist Virology Centre, NHSGGC / MRC-University of Glasgow Centre for Virus Research | COVID-19 Genomics UK (COG-UK) Consortium |
| EPI_ISL_438808 | West of Scotland Specialist Virology Centre, NHSGGC / MRC-University of Glasgow Centre for Virus Research | COVID-19 Genomics UK (COG-UK) Consortium |
| EPI_ISL_438809 | West of Scotland Specialist Virology Centre, NHSGGC / MRC-University of Glasgow Centre for Virus Research | COVID-19 Genomics UK (COG-UK) Consortium |
| EPI_ISL_438810 | West of Scotland Specialist Virology Centre, NHSGGC / MRC-University of Glasgow Centre for Virus Research | COVID-19 Genomics UK (COG-UK) Consortium |
| EPI_ISL_438811 | West of Scotland Specialist Virology Centre, NHSGGC / MRC-University of Glasgow Centre for Virus Research | COVID-19 Genomics UK (COG-UK) Consortium |
| EPI_ISL_438812 | West of Scotland Specialist Virology Centre, NHSGGC / MRC-University of Glasgow Centre for Virus Research | COVID-19 Genomics UK (COG-UK) Consortium |
| EPI_ISL_438813 | West of Scotland Specialist Virology Centre, NHSGGC / MRC-University of Glasgow Centre for Virus Research | COVID-19 Genomics UK (COG-UK) Consortium |
| EPI_ISL_438814 | West of Scotland Specialist Virology Centre, NHSGGC / MRC-University of Glasgow Centre for Virus Research | COVID-19 Genomics UK (COG-UK) Consortium |

| EPI_Accession_ID | Originating Laboratory | Submitting Laboratory |
|---|---|---|
| EPI_ISL_438815 | West of Scotland Specialist Virology Centre, NHSGGC / MRC-University of Glasgow Centre for Virus Research | COVID-19 Genomics UK (COG-UK) Consortium |
| EPI_ISL_438816 | West of Scotland Specialist Virology Centre, NHSGGC / MRC-University of Glasgow Centre for Virus Research | COVID-19 Genomics UK (COG-UK) Consortium |
| EPI_ISL_438817 | West of Scotland Specialist Virology Centre, NHSGGC / MRC-University of Glasgow Centre for Virus Research | COVID-19 Genomics UK (COG-UK) Consortium |
| EPI_ISL_438818 | West of Scotland Specialist Virology Centre, NHSGGC / MRC-University of Glasgow Centre for Virus Research | COVID-19 Genomics UK (COG-UK) Consortium |
| EPI_ISL_438819 | West of Scotland Specialist Virology Centre, NHSGGC / MRC-University of Glasgow Centre for Virus Research | COVID-19 Genomics UK (COG-UK) Consortium |
| EPI_ISL_438820 | West of Scotland Specialist Virology Centre, NHSGGC / MRC-University of Glasgow Centre for Virus Research | COVID-19 Genomics UK (COG-UK) Consortium |
| EPI_ISL_438821 | West of Scotland Specialist Virology Centre, NHSGGC / MRC-University of Glasgow Centre for Virus Research | COVID-19 Genomics UK (COG-UK) Consortium |
| EPI_ISL_438822 | West of Scotland Specialist Virology Centre, NHSGGC / MRC-University of Glasgow Centre for Virus Research | COVID-19 Genomics UK (COG-UK) Consortium |
| EPI_ISL_438823 | West of Scotland Specialist Virology Centre, NHSGGC / MRC-University of Glasgow Centre for Virus Research | COVID-19 Genomics UK (COG-UK) Consortium |
| EPI_ISL_438824 | West of Scotland Specialist Virology Centre, NHSGGC / MRC-University of Glasgow Centre for Virus Research | COVID-19 Genomics UK (COG-UK) Consortium |
| EPI_ISL_438825 | West of Scotland Specialist Virology Centre, NHSGGC / MRC-University of Glasgow Centre for Virus Research | COVID-19 Genomics UK (COG-UK) Consortium |
| EPI_ISL_438826 | West of Scotland Specialist Virology Centre, NHSGGC / MRC-University of Glasgow Centre for Virus Research | COVID-19 Genomics UK (COG-UK) Consortium |
| EPI_ISL_438827 | West of Scotland Specialist Virology Centre, NHSGGC / MRC-University of Glasgow Centre for Virus Research | COVID-19 Genomics UK (COG-UK) Consortium |
| EPI_ISL_438828 | West of Scotland Specialist Virology Centre, NHSGGC / MRC-University of Glasgow Centre for Virus Research | COVID-19 Genomics UK (COG-UK) Consortium |
| EPI_ISL_438829 | West of Scotland Specialist Virology Centre, NHSGGC / MRC-University of Glasgow Centre for Virus Research | COVID-19 Genomics UK (COG-UK) Consortium |
| EPI_ISL_438830 | West of Scotland Specialist Virology Centre, NHSGGC / MRC-University of Glasgow Centre for Virus Research | COVID-19 Genomics UK (COG-UK) Consortium |

| EPI_Accession_ID | Originating Laboratory | Submitting Laboratory |
|---|---|---|
| EPI_ISL_438831 | West of Scotland Specialist Virology Centre, NHSGGC / MRC-University of Glasgow Centre for Virus Research | COVID-19 Genomics UK (COG-UK) Consortium |
| EPI_ISL_438832 | West of Scotland Specialist Virology Centre, NHSGGC / MRC-University of Glasgow Centre for Virus Research | COVID-19 Genomics UK (COG-UK) Consortium |
| EPI_ISL_438833 | West of Scotland Specialist Virology Centre, NHSGGC / MRC-University of Glasgow Centre for Virus Research | COVID-19 Genomics UK (COG-UK) Consortium |
| EPI_ISL_438834 | West of Scotland Specialist Virology Centre, NHSGGC / MRC-University of Glasgow Centre for Virus Research | COVID-19 Genomics UK (COG-UK) Consortium |
| EPI_ISL_438835 | West of Scotland Specialist Virology Centre, NHSGGC / MRC-University of Glasgow Centre for Virus Research | COVID-19 Genomics UK (COG-UK) Consortium |
| EPI_ISL_438836 | West of Scotland Specialist Virology Centre, NHSGGC / MRC-University of Glasgow Centre for Virus Research | COVID-19 Genomics UK (COG-UK) Consortium |
| EPI_ISL_438837 | West of Scotland Specialist Virology Centre, NHSGGC / MRC-University of Glasgow Centre for Virus Research | COVID-19 Genomics UK (COG-UK) Consortium |
| EPI_ISL_438838 | West of Scotland Specialist Virology Centre, NHSGGC / MRC-University of Glasgow Centre for Virus Research | COVID-19 Genomics UK (COG-UK) Consortium |
| EPI_ISL_438839 | West of Scotland Specialist Virology Centre, NHSGGC / MRC-University of Glasgow Centre for Virus Research | COVID-19 Genomics UK (COG-UK) Consortium |
| EPI_ISL_438840 | West of Scotland Specialist Virology Centre, NHSGGC / MRC-University of Glasgow Centre for Virus Research | COVID-19 Genomics UK (COG-UK) Consortium |
| EPI_ISL_438841 | West of Scotland Specialist Virology Centre, NHSGGC / MRC-University of Glasgow Centre for Virus Research | COVID-19 Genomics UK (COG-UK) Consortium |
| EPI_ISL_438842 | West of Scotland Specialist Virology Centre, NHSGGC / MRC-University of Glasgow Centre for Virus Research | COVID-19 Genomics UK (COG-UK) Consortium |
| EPI_ISL_438843 | West of Scotland Specialist Virology Centre, NHSGGC / MRC-University of Glasgow Centre for Virus Research | COVID-19 Genomics UK (COG-UK) Consortium |
| EPI_ISL_438844 | West of Scotland Specialist Virology Centre, NHSGGC / MRC-University of Glasgow Centre for Virus Research | COVID-19 Genomics UK (COG-UK) Consortium |
| EPI_ISL_438845 | West of Scotland Specialist Virology Centre, NHSGGC / MRC-University of Glasgow Centre for Virus Research | COVID-19 Genomics UK (COG-UK) Consortium |
| EPI_ISL_438846 | West of Scotland Specialist Virology Centre, NHSGGC / MRC-University of Glasgow Centre for Virus Research | COVID-19 Genomics UK (COG-UK) Consortium |

| EPI_Accession_ID | Originating Laboratory | Submitting Laboratory |
|---|---|---|
| EPI_ISL_438847 | West of Scotland Specialist Virology Centre, NHSGGC / MRC-University of Glasgow Centre for Virus Research | COVID-19 Genomics UK (COG-UK) Consortium |
| EPI_ISL_438848 | West of Scotland Specialist Virology Centre, NHSGGC / MRC-University of Glasgow Centre for Virus Research | COVID-19 Genomics UK (COG-UK) Consortium |
| EPI_ISL_438849 | West of Scotland Specialist Virology Centre, NHSGGC / MRC-University of Glasgow Centre for Virus Research | COVID-19 Genomics UK (COG-UK) Consortium |
| EPI_ISL_438850 | West of Scotland Specialist Virology Centre, NHSGGC / MRC-University of Glasgow Centre for Virus Research | COVID-19 Genomics UK (COG-UK) Consortium |
| EPI_ISL_438851 | West of Scotland Specialist Virology Centre, NHSGGC / MRC-University of Glasgow Centre for Virus Research | COVID-19 Genomics UK (COG-UK) Consortium |
| EPI_ISL_438852 | West of Scotland Specialist Virology Centre, NHSGGC / MRC-University of Glasgow Centre for Virus Research | COVID-19 Genomics UK (COG-UK) Consortium |
| EPI_ISL_438853 | West of Scotland Specialist Virology Centre, NHSGGC / MRC-University of Glasgow Centre for Virus Research | COVID-19 Genomics UK (COG-UK) Consortium |
| EPI_ISL_438854 | West of Scotland Specialist Virology Centre, NHSGGC / MRC-University of Glasgow Centre for Virus Research | COVID-19 Genomics UK (COG-UK) Consortium |
| EPI_ISL_438855 | West of Scotland Specialist Virology Centre, NHSGGC / MRC-University of Glasgow Centre for Virus Research | COVID-19 Genomics UK (COG-UK) Consortium |
| EPI_ISL_438856 | West of Scotland Specialist Virology Centre, NHSGGC / MRC-University of Glasgow Centre for Virus Research | COVID-19 Genomics UK (COG-UK) Consortium |
| EPI_ISL_438857 | West of Scotland Specialist Virology Centre, NHSGGC / MRC-University of Glasgow Centre for Virus Research | COVID-19 Genomics UK (COG-UK) Consortium |
| EPI_ISL_438858 | West of Scotland Specialist Virology Centre, NHSGGC / MRC-University of Glasgow Centre for Virus Research | COVID-19 Genomics UK (COG-UK) Consortium |
| EPI_ISL_438859 | West of Scotland Specialist Virology Centre, NHSGGC / MRC-University of Glasgow Centre for Virus Research | COVID-19 Genomics UK (COG-UK) Consortium |
| EPI_ISL_438860 | West of Scotland Specialist Virology Centre, NHSGGC / MRC-University of Glasgow Centre for Virus Research | COVID-19 Genomics UK (COG-UK) Consortium |
| EPI_ISL_438861 | West of Scotland Specialist Virology Centre, NHSGGC / MRC-University of Glasgow Centre for Virus Research | COVID-19 Genomics UK (COG-UK) Consortium |
| EPI_ISL_438862 | West of Scotland Specialist Virology Centre, NHSGGC / MRC-University of Glasgow Centre for Virus Research | COVID-19 Genomics UK (COG-UK) Consortium |

| EPI_Accession_ID | Originating Laboratory | Submitting Laboratory |
| --- | --- | --- |
| EPI_ISL_438863 | West of Scotland Specialist Virology Centre, NHSGGC / MRC-University of Glasgow Centre for Virus Research | COVID-19 Genomics UK (COG-UK) Consortium |
| EPI_ISL_438864 | West of Scotland Specialist Virology Centre, NHSGGC / MRC-University of Glasgow Centre for Virus Research | COVID-19 Genomics UK (COG-UK) Consortium |
| EPI_ISL_438865 | West of Scotland Specialist Virology Centre, NHSGGC / MRC-University of Glasgow Centre for Virus Research | COVID-19 Genomics UK (COG-UK) Consortium |
| EPI_ISL_438866 | West of Scotland Specialist Virology Centre, NHSGGC / MRC-University of Glasgow Centre for Virus Research | COVID-19 Genomics UK (COG-UK) Consortium |
| EPI_ISL_438867 | West of Scotland Specialist Virology Centre, NHSGGC / MRC-University of Glasgow Centre for Virus Research | COVID-19 Genomics UK (COG-UK) Consortium |
| EPI_ISL_438868 | West of Scotland Specialist Virology Centre, NHSGGC / MRC-University of Glasgow Centre for Virus Research | COVID-19 Genomics UK (COG-UK) Consortium |
| EPI_ISL_438869 | West of Scotland Specialist Virology Centre, NHSGGC / MRC-University of Glasgow Centre for Virus Research | COVID-19 Genomics UK (COG-UK) Consortium |
| EPI_ISL_438870 | West of Scotland Specialist Virology Centre, NHSGGC / MRC-University of Glasgow Centre for Virus Research | COVID-19 Genomics UK (COG-UK) Consortium |
| EPI_ISL_438871 | West of Scotland Specialist Virology Centre, NHSGGC / MRC-University of Glasgow Centre for Virus Research | COVID-19 Genomics UK (COG-UK) Consortium |
| EPI_ISL_438872 | West of Scotland Specialist Virology Centre, NHSGGC / MRC-University of Glasgow Centre for Virus Research | COVID-19 Genomics UK (COG-UK) Consortium |
| EPI_ISL_438873 | West of Scotland Specialist Virology Centre, NHSGGC / MRC-University of Glasgow Centre for Virus Research | COVID-19 Genomics UK (COG-UK) Consortium |
| EPI_ISL_438874 | West of Scotland Specialist Virology Centre, NHSGGC / MRC-University of Glasgow Centre for Virus Research | COVID-19 Genomics UK (COG-UK) Consortium |
| EPI_ISL_438875 | West of Scotland Specialist Virology Centre, NHSGGC / MRC-University of Glasgow Centre for Virus Research | COVID-19 Genomics UK (COG-UK) Consortium |
| EPI_ISL_438876 | West of Scotland Specialist Virology Centre, NHSGGC / MRC-University of Glasgow Centre for Virus Research | COVID-19 Genomics UK (COG-UK) Consortium |
| EPI_ISL_438877 | West of Scotland Specialist Virology Centre, NHSGGC / MRC-University of Glasgow Centre for Virus Research | COVID-19 Genomics UK (COG-UK) Consortium |
| EPI_ISL_438878 | West of Scotland Specialist Virology Centre, NHSGGC / MRC-University of Glasgow Centre for Virus Research | COVID-19 Genomics UK (COG-UK) Consortium |

| EPI_Accession_ID | Originating Laboratory | Submitting Laboratory |
|---|---|---|
| EPI_ISL_438879 | West of Scotland Specialist Virology Centre, NHSGGC / MRC-University of Glasgow Centre for Virus Research | COVID-19 Genomics UK (COG-UK) Consortium |
| EPI_ISL_438880 | West of Scotland Specialist Virology Centre, NHSGGC / MRC-University of Glasgow Centre for Virus Research | COVID-19 Genomics UK (COG-UK) Consortium |
| EPI_ISL_438881 | West of Scotland Specialist Virology Centre, NHSGGC / MRC-University of Glasgow Centre for Virus Research | COVID-19 Genomics UK (COG-UK) Consortium |
| EPI_ISL_438882 | West of Scotland Specialist Virology Centre, NHSGGC / MRC-University of Glasgow Centre for Virus Research | COVID-19 Genomics UK (COG-UK) Consortium |
| EPI_ISL_438883 | West of Scotland Specialist Virology Centre, NHSGGC / MRC-University of Glasgow Centre for Virus Research | COVID-19 Genomics UK (COG-UK) Consortium |
| EPI_ISL_438884 | West of Scotland Specialist Virology Centre, NHSGGC / MRC-University of Glasgow Centre for Virus Research | COVID-19 Genomics UK (COG-UK) Consortium |
| EPI_ISL_438885 | West of Scotland Specialist Virology Centre, NHSGGC / MRC-University of Glasgow Centre for Virus Research | COVID-19 Genomics UK (COG-UK) Consortium |
| EPI_ISL_438886 | West of Scotland Specialist Virology Centre, NHSGGC / MRC-University of Glasgow Centre for Virus Research | COVID-19 Genomics UK (COG-UK) Consortium |
| EPI_ISL_438887 | West of Scotland Specialist Virology Centre, NHSGGC / MRC-University of Glasgow Centre for Virus Research | COVID-19 Genomics UK (COG-UK) Consortium |
| EPI_ISL_438888 | West of Scotland Specialist Virology Centre, NHSGGC / MRC-University of Glasgow Centre for Virus Research | COVID-19 Genomics UK (COG-UK) Consortium |
| EPI_ISL_438889 | West of Scotland Specialist Virology Centre, NHSGGC / MRC-University of Glasgow Centre for Virus Research | COVID-19 Genomics UK (COG-UK) Consortium |
| EPI_ISL_438890 | West of Scotland Specialist Virology Centre, NHSGGC / MRC-University of Glasgow Centre for Virus Research | COVID-19 Genomics UK (COG-UK) Consortium |
| EPI_ISL_438891 | West of Scotland Specialist Virology Centre, NHSGGC / MRC-University of Glasgow Centre for Virus Research | COVID-19 Genomics UK (COG-UK) Consortium |
| EPI_ISL_438892 | West of Scotland Specialist Virology Centre, NHSGGC / MRC-University of Glasgow Centre for Virus Research | COVID-19 Genomics UK (COG-UK) Consortium |
| EPI_ISL_438893 | West of Scotland Specialist Virology Centre, NHSGGC / MRC-University of Glasgow Centre for Virus Research | COVID-19 Genomics UK (COG-UK) Consortium |
| EPI_ISL_438894 | West of Scotland Specialist Virology Centre, NHSGGC / MRC-University of Glasgow Centre for Virus Research | COVID-19 Genomics UK (COG-UK) Consortium |

| EPI_Accession_ID | Originating Laboratory | Submitting Laboratory |
|---|---|---|
| EPI_ISL_438895 | West of Scotland Specialist Virology Centre, NHSGGC / MRC-University of Glasgow Centre for Virus Research | COVID-19 Genomics UK (COG-UK) Consortium |
| EPI_ISL_438896 | West of Scotland Specialist Virology Centre, NHSGGC / MRC-University of Glasgow Centre for Virus Research | COVID-19 Genomics UK (COG-UK) Consortium |
| EPI_ISL_438897 | West of Scotland Specialist Virology Centre, NHSGGC / MRC-University of Glasgow Centre for Virus Research | COVID-19 Genomics UK (COG-UK) Consortium |
| EPI_ISL_438898 | West of Scotland Specialist Virology Centre, NHSGGC / MRC-University of Glasgow Centre for Virus Research | COVID-19 Genomics UK (COG-UK) Consortium |
| EPI_ISL_438899 | West of Scotland Specialist Virology Centre, NHSGGC / MRC-University of Glasgow Centre for Virus Research | COVID-19 Genomics UK (COG-UK) Consortium |
| EPI_ISL_438900 | West of Scotland Specialist Virology Centre, NHSGGC / MRC-University of Glasgow Centre for Virus Research | COVID-19 Genomics UK (COG-UK) Consortium |
| EPI_ISL_438901 | West of Scotland Specialist Virology Centre, NHSGGC / MRC-University of Glasgow Centre for Virus Research | COVID-19 Genomics UK (COG-UK) Consortium |
| EPI_ISL_438902 | West of Scotland Specialist Virology Centre, NHSGGC / MRC-University of Glasgow Centre for Virus Research | COVID-19 Genomics UK (COG-UK) Consortium |
| EPI_ISL_438903 | West of Scotland Specialist Virology Centre, NHSGGC / MRC-University of Glasgow Centre for Virus Research | COVID-19 Genomics UK (COG-UK) Consortium |
| EPI_ISL_438904 | West of Scotland Specialist Virology Centre, NHSGGC / MRC-University of Glasgow Centre for Virus Research | COVID-19 Genomics UK (COG-UK) Consortium |
| EPI_ISL_438905 | West of Scotland Specialist Virology Centre, NHSGGC / MRC-University of Glasgow Centre for Virus Research | COVID-19 Genomics UK (COG-UK) Consortium |
| EPI_ISL_438906 | West of Scotland Specialist Virology Centre, NHSGGC / MRC-University of Glasgow Centre for Virus Research | COVID-19 Genomics UK (COG-UK) Consortium |
| EPI_ISL_438907 | West of Scotland Specialist Virology Centre, NHSGGC / MRC-University of Glasgow Centre for Virus Research | COVID-19 Genomics UK (COG-UK) Consortium |
| EPI_ISL_438908 | West of Scotland Specialist Virology Centre, NHSGGC / MRC-University of Glasgow Centre for Virus Research | COVID-19 Genomics UK (COG-UK) Consortium |
| EPI_ISL_438909 | West of Scotland Specialist Virology Centre, NHSGGC / MRC-University of Glasgow Centre for Virus Research | COVID-19 Genomics UK (COG-UK) Consortium |
| EPI_ISL_438910 | West of Scotland Specialist Virology Centre, NHSGGC / MRC-University of Glasgow Centre for Virus Research | COVID-19 Genomics UK (COG-UK) Consortium |

| EPI_Accession_ID | Originating Laboratory | Submitting Laboratory |
|---|---|---|
| EPI_ISL_438911 | West of Scotland Specialist Virology Centre, NHSGGC / MRC-University of Glasgow Centre for Virus Research | COVID-19 Genomics UK (COG-UK) Consortium |
| EPI_ISL_438912 | West of Scotland Specialist Virology Centre, NHSGGC / MRC-University of Glasgow Centre for Virus Research | COVID-19 Genomics UK (COG-UK) Consortium |
| EPI_ISL_438913 | West of Scotland Specialist Virology Centre, NHSGGC / MRC-University of Glasgow Centre for Virus Research | COVID-19 Genomics UK (COG-UK) Consortium |
| EPI_ISL_438914 | West of Scotland Specialist Virology Centre, NHSGGC / MRC-University of Glasgow Centre for Virus Research | COVID-19 Genomics UK (COG-UK) Consortium |
| EPI_ISL_438915 | West of Scotland Specialist Virology Centre, NHSGGC / MRC-University of Glasgow Centre for Virus Research | COVID-19 Genomics UK (COG-UK) Consortium |
| EPI_ISL_438916 | West of Scotland Specialist Virology Centre, NHSGGC / MRC-University of Glasgow Centre for Virus Research | COVID-19 Genomics UK (COG-UK) Consortium |
| EPI_ISL_438917 | West of Scotland Specialist Virology Centre, NHSGGC / MRC-University of Glasgow Centre for Virus Research | COVID-19 Genomics UK (COG-UK) Consortium |
| EPI_ISL_438918 | West of Scotland Specialist Virology Centre, NHSGGC / MRC-University of Glasgow Centre for Virus Research | COVID-19 Genomics UK (COG-UK) Consortium |
| EPI_ISL_438919 | West of Scotland Specialist Virology Centre, NHSGGC / MRC-University of Glasgow Centre for Virus Research | COVID-19 Genomics UK (COG-UK) Consortium |
| EPI_ISL_438920 | West of Scotland Specialist Virology Centre, NHSGGC / MRC-University of Glasgow Centre for Virus Research | COVID-19 Genomics UK (COG-UK) Consortium |
| EPI_ISL_438921 | West of Scotland Specialist Virology Centre, NHSGGC / MRC-University of Glasgow Centre for Virus Research | COVID-19 Genomics UK (COG-UK) Consortium |
| EPI_ISL_438922 | West of Scotland Specialist Virology Centre, NHSGGC / MRC-University of Glasgow Centre for Virus Research | COVID-19 Genomics UK (COG-UK) Consortium |
| EPI_ISL_438923 | West of Scotland Specialist Virology Centre, NHSGGC / MRC-University of Glasgow Centre for Virus Research | COVID-19 Genomics UK (COG-UK) Consortium |
| EPI_ISL_438924 | West of Scotland Specialist Virology Centre, NHSGGC / MRC-University of Glasgow Centre for Virus Research | COVID-19 Genomics UK (COG-UK) Consortium |
| EPI_ISL_438925 | West of Scotland Specialist Virology Centre, NHSGGC / MRC-University of Glasgow Centre for Virus Research | COVID-19 Genomics UK (COG-UK) Consortium |
| EPI_ISL_438926 | West of Scotland Specialist Virology Centre, NHSGGC / MRC-University of Glasgow Centre for Virus Research | COVID-19 Genomics UK (COG-UK) Consortium |

| EPI_Accession_ID | Originating Laboratory | Submitting Laboratory |
| --- | --- | --- |
| EPI_ISL_438927 | West of Scotland Specialist Virology Centre, NHSGGC / MRC-University of Glasgow Centre for Virus Research | COVID-19 Genomics UK (COG-UK) Consortium |
| EPI_ISL_438928 | West of Scotland Specialist Virology Centre, NHSGGC / MRC-University of Glasgow Centre for Virus Research | COVID-19 Genomics UK (COG-UK) Consortium |
| EPI_ISL_438929 | West of Scotland Specialist Virology Centre, NHSGGC / MRC-University of Glasgow Centre for Virus Research | COVID-19 Genomics UK (COG-UK) Consortium |
| EPI_ISL_438930 | West of Scotland Specialist Virology Centre, NHSGGC / MRC-University of Glasgow Centre for Virus Research | COVID-19 Genomics UK (COG-UK) Consortium |
| EPI_ISL_438931 | West of Scotland Specialist Virology Centre, NHSGGC / MRC-University of Glasgow Centre for Virus Research | COVID-19 Genomics UK (COG-UK) Consortium |
| EPI_ISL_438932 | West of Scotland Specialist Virology Centre, NHSGGC / MRC-University of Glasgow Centre for Virus Research | COVID-19 Genomics UK (COG-UK) Consortium |
| EPI_ISL_438933 | West of Scotland Specialist Virology Centre, NHSGGC / MRC-University of Glasgow Centre for Virus Research | COVID-19 Genomics UK (COG-UK) Consortium |
| EPI_ISL_438934 | West of Scotland Specialist Virology Centre, NHSGGC / MRC-University of Glasgow Centre for Virus Research | COVID-19 Genomics UK (COG-UK) Consortium |
| EPI_ISL_438935 | West of Scotland Specialist Virology Centre, NHSGGC / MRC-University of Glasgow Centre for Virus Research | COVID-19 Genomics UK (COG-UK) Consortium |
| EPI_ISL_438936 | West of Scotland Specialist Virology Centre, NHSGGC / MRC-University of Glasgow Centre for Virus Research | COVID-19 Genomics UK (COG-UK) Consortium |
| EPI_ISL_438937 | West of Scotland Specialist Virology Centre, NHSGGC / MRC-University of Glasgow Centre for Virus Research | COVID-19 Genomics UK (COG-UK) Consortium |
| EPI_ISL_438938 | West of Scotland Specialist Virology Centre, NHSGGC / MRC-University of Glasgow Centre for Virus Research | COVID-19 Genomics UK (COG-UK) Consortium |
| EPI_ISL_438939 | West of Scotland Specialist Virology Centre, NHSGGC / MRC-University of Glasgow Centre for Virus Research | COVID-19 Genomics UK (COG-UK) Consortium |
| EPI_ISL_438940 | West of Scotland Specialist Virology Centre, NHSGGC / MRC-University of Glasgow Centre for Virus Research | COVID-19 Genomics UK (COG-UK) Consortium |
| EPI_ISL_438941 | West of Scotland Specialist Virology Centre, NHSGGC / MRC-University of Glasgow Centre for Virus Research | COVID-19 Genomics UK (COG-UK) Consortium |
| EPI_ISL_438942 | West of Scotland Specialist Virology Centre, NHSGGC / MRC-University of Glasgow Centre for Virus Research | COVID-19 Genomics UK (COG-UK) Consortium |

| EPI_Accession_ID | Originating Laboratory | Submitting Laboratory |
|---|---|---|
| EPI_ISL_438943 | West of Scotland Specialist Virology Centre, NHSGGC / MRC-University of Glasgow Centre for Virus Research | COVID-19 Genomics UK (COG-UK) Consortium |
| EPI_ISL_438944 | West of Scotland Specialist Virology Centre, NHSGGC / MRC-University of Glasgow Centre for Virus Research | COVID-19 Genomics UK (COG-UK) Consortium |
| EPI_ISL_438945 | West of Scotland Specialist Virology Centre, NHSGGC / MRC-University of Glasgow Centre for Virus Research | COVID-19 Genomics UK (COG-UK) Consortium |
| EPI_ISL_438946 | West of Scotland Specialist Virology Centre, NHSGGC / MRC-University of Glasgow Centre for Virus Research | COVID-19 Genomics UK (COG-UK) Consortium |
| EPI_ISL_438947 | Keio University School of Medicine | Keio University School of Medicine |
| EPI_ISL_438948 | Keio University School of Medicine | Keio University School of Medicine |
| EPI_ISL_438949 | Keio University School of Medicine | Keio University School of Medicine |
| EPI_ISL_438951 | Keio University School of Medicine | Keio University School of Medicine |
| EPI_ISL_438952 | Keio University School of Medicine | Keio University School of Medicine |
| EPI_ISL_438953 | Keio University School of Medicine | Keio University School of Medicine |
| EPI_ISL_438954 | Keio University School of Medicine | Keio University School of Medicine |
| EPI_ISL_438955 | Keio University School of Medicine | Keio University School of Medicine |
| EPI_ISL_438956 | Keio University School of Medicine | Keio University School of Medicine |
| EPI_ISL_438958 | Keio University School of Medicine | Keio University School of Medicine |
| EPI_ISL_438959 | Keio University School of Medicine | Keio University School of Medicine |
| EPI_ISL_438961 | Keio University School of Medicine | Keio University School of Medicine |
| EPI_ISL_438962 | Keio University School of Medicine | Keio University School of Medicine |
| EPI_ISL_438963 | Keio University School of Medicine | Keio University School of Medicine |
| EPI_ISL_438964 | Keio University School of Medicine | Keio University School of Medicine |
| EPI_ISL_438965 | Keio University School of Medicine | Keio University School of Medicine |
| EPI_ISL_438966 | Keio University School of Medicine | Keio University School of Medicine |
| EPI_ISL_438967 | Keio University School of Medicine | Keio University School of Medicine |
| EPI_ISL_438968 | Keio University School of Medicine | Keio University School of Medicine |
| EPI_ISL_438969 | Keio University School of Medicine | Keio University School of Medicine |
| EPI_ISL_438970 | Keio University School of Medicine | Keio University School of Medicine |
| EPI_ISL_438971 | Keio University School of Medicine | Keio University School of Medicine |
| EPI_ISL_438972 | Keio University School of Medicine | Keio University School of Medicine |
| EPI_ISL_438973 | West of Scotland Specialist Virology Centre, NHSGGC / MRC-University of Glasgow Centre for Virus Research | COVID-19 Genomics UK (COG-UK) Consortium |

| EPI_Accession_ID | Originating Laboratory | Submitting Laboratory |
|---|---|---|
| EPI_ISL_438974 | West of Scotland Specialist Virology Centre, NHSGGC / MRC-University of Glasgow Centre for Virus Research | COVID-19 Genomics UK (COG-UK) Consortium |
| EPI_ISL_438975 | West of Scotland Specialist Virology Centre, NHSGGC / MRC-University of Glasgow Centre for Virus Research | COVID-19 Genomics UK (COG-UK) Consortium |
| EPI_ISL_438976 | West of Scotland Specialist Virology Centre, NHSGGC / MRC-University of Glasgow Centre for Virus Research | COVID-19 Genomics UK (COG-UK) Consortium |
| EPI_ISL_438977 | West of Scotland Specialist Virology Centre, NHSGGC / MRC-University of Glasgow Centre for Virus Research | COVID-19 Genomics UK (COG-UK) Consortium |
| EPI_ISL_438978 | West of Scotland Specialist Virology Centre, NHSGGC / MRC-University of Glasgow Centre for Virus Research | COVID-19 Genomics UK (COG-UK) Consortium |
| EPI_ISL_438979 | West of Scotland Specialist Virology Centre, NHSGGC / MRC-University of Glasgow Centre for Virus Research | COVID-19 Genomics UK (COG-UK) Consortium |
| EPI_ISL_438980 | West of Scotland Specialist Virology Centre, NHSGGC / MRC-University of Glasgow Centre for Virus Research | COVID-19 Genomics UK (COG-UK) Consortium |
| EPI_ISL_438981 | West of Scotland Specialist Virology Centre, NHSGGC / MRC-University of Glasgow Centre for Virus Research | COVID-19 Genomics UK (COG-UK) Consortium |
| EPI_ISL_438982 | West of Scotland Specialist Virology Centre, NHSGGC / MRC-University of Glasgow Centre for Virus Research | COVID-19 Genomics UK (COG-UK) Consortium |
| EPI_ISL_438983 | West of Scotland Specialist Virology Centre, NHSGGC / MRC-University of Glasgow Centre for Virus Research | COVID-19 Genomics UK (COG-UK) Consortium |
| EPI_ISL_438984 | West of Scotland Specialist Virology Centre, NHSGGC / MRC-University of Glasgow Centre for Virus Research | COVID-19 Genomics UK (COG-UK) Consortium |
| EPI_ISL_438985 | West of Scotland Specialist Virology Centre, NHSGGC / MRC-University of Glasgow Centre for Virus Research | COVID-19 Genomics UK (COG-UK) Consortium |
| EPI_ISL_438986 | West of Scotland Specialist Virology Centre, NHSGGC / MRC-University of Glasgow Centre for Virus Research | COVID-19 Genomics UK (COG-UK) Consortium |
| EPI_ISL_438987 | West of Scotland Specialist Virology Centre, NHSGGC / MRC-University of Glasgow Centre for Virus Research | COVID-19 Genomics UK (COG-UK) Consortium |
| EPI_ISL_438988 | West of Scotland Specialist Virology Centre, NHSGGC / MRC-University of Glasgow Centre for Virus Research | COVID-19 Genomics UK (COG-UK) Consortium |
| EPI_ISL_438989 | West of Scotland Specialist Virology Centre, NHSGGC / MRC-University of Glasgow Centre for Virus Research | COVID-19 Genomics UK (COG-UK) Consortium |

| EPI_Accession_ID | Originating Laboratory | Submitting Laboratory |
|---|---|---|
| EPI_ISL_438990 | West of Scotland Specialist Virology Centre, NHSGGC / MRC-University of Glasgow Centre for Virus Research | COVID-19 Genomics UK (COG-UK) Consortium |
| EPI_ISL_438991 | West of Scotland Specialist Virology Centre, NHSGGC / MRC-University of Glasgow Centre for Virus Research | COVID-19 Genomics UK (COG-UK) Consortium |
| EPI_ISL_438992 | West of Scotland Specialist Virology Centre, NHSGGC / MRC-University of Glasgow Centre for Virus Research | COVID-19 Genomics UK (COG-UK) Consortium |
| EPI_ISL_438993 | West of Scotland Specialist Virology Centre, NHSGGC / MRC-University of Glasgow Centre for Virus Research | COVID-19 Genomics UK (COG-UK) Consortium |
| EPI_ISL_438994 | West of Scotland Specialist Virology Centre, NHSGGC / MRC-University of Glasgow Centre for Virus Research | COVID-19 Genomics UK (COG-UK) Consortium |
| EPI_ISL_438995 | West of Scotland Specialist Virology Centre, NHSGGC / MRC-University of Glasgow Centre for Virus Research | COVID-19 Genomics UK (COG-UK) Consortium |
| EPI_ISL_438996 | West of Scotland Specialist Virology Centre, NHSGGC / MRC-University of Glasgow Centre for Virus Research | COVID-19 Genomics UK (COG-UK) Consortium |
| EPI_ISL_438997 | West of Scotland Specialist Virology Centre, NHSGGC / MRC-University of Glasgow Centre for Virus Research | COVID-19 Genomics UK (COG-UK) Consortium |
| EPI_ISL_438998 | West of Scotland Specialist Virology Centre, NHSGGC / MRC-University of Glasgow Centre for Virus Research | COVID-19 Genomics UK (COG-UK) Consortium |
| EPI_ISL_438999 | West of Scotland Specialist Virology Centre, NHSGGC / MRC-University of Glasgow Centre for Virus Research | COVID-19 Genomics UK (COG-UK) Consortium |
| EPI_ISL_439000 | West of Scotland Specialist Virology Centre, NHSGGC / MRC-University of Glasgow Centre for Virus Research | COVID-19 Genomics UK (COG-UK) Consortium |
| EPI_ISL_439001 | West of Scotland Specialist Virology Centre, NHSGGC / MRC-University of Glasgow Centre for Virus Research | COVID-19 Genomics UK (COG-UK) Consortium |
| EPI_ISL_439002 | West of Scotland Specialist Virology Centre, NHSGGC / MRC-University of Glasgow Centre for Virus Research | COVID-19 Genomics UK (COG-UK) Consortium |
| EPI_ISL_439003 | West of Scotland Specialist Virology Centre, NHSGGC / MRC-University of Glasgow Centre for Virus Research | COVID-19 Genomics UK (COG-UK) Consortium |
| EPI_ISL_439004 | West of Scotland Specialist Virology Centre, NHSGGC / MRC-University of Glasgow Centre for Virus Research | COVID-19 Genomics UK (COG-UK) Consortium |
| EPI_ISL_439005 | West of Scotland Specialist Virology Centre, NHSGGC / MRC-University of Glasgow Centre for Virus Research | COVID-19 Genomics UK (COG-UK) Consortium |

| EPI_Accession_ID | Originating Laboratory | Submitting Laboratory |
|---|---|---|
| EPI_ISL_439006 | West of Scotland Specialist Virology Centre, NHSGGC / MRC-University of Glasgow Centre for Virus Research | COVID-19 Genomics UK (COG-UK) Consortium |
| EPI_ISL_439007 | West of Scotland Specialist Virology Centre, NHSGGC / MRC-University of Glasgow Centre for Virus Research | COVID-19 Genomics UK (COG-UK) Consortium |
| EPI_ISL_439008 | West of Scotland Specialist Virology Centre, NHSGGC / MRC-University of Glasgow Centre for Virus Research | COVID-19 Genomics UK (COG-UK) Consortium |
| EPI_ISL_439009 | West of Scotland Specialist Virology Centre, NHSGGC / MRC-University of Glasgow Centre for Virus Research | COVID-19 Genomics UK (COG-UK) Consortium |
| EPI_ISL_439010 | West of Scotland Specialist Virology Centre, NHSGGC / MRC-University of Glasgow Centre for Virus Research | COVID-19 Genomics UK (COG-UK) Consortium |
| EPI_ISL_439011 | West of Scotland Specialist Virology Centre, NHSGGC / MRC-University of Glasgow Centre for Virus Research | COVID-19 Genomics UK (COG-UK) Consortium |
| EPI_ISL_439012 | West of Scotland Specialist Virology Centre, NHSGGC / MRC-University of Glasgow Centre for Virus Research | COVID-19 Genomics UK (COG-UK) Consortium |
| EPI_ISL_439013 | West of Scotland Specialist Virology Centre, NHSGGC / MRC-University of Glasgow Centre for Virus Research | COVID-19 Genomics UK (COG-UK) Consortium |
| EPI_ISL_439014 | West of Scotland Specialist Virology Centre, NHSGGC / MRC-University of Glasgow Centre for Virus Research | COVID-19 Genomics UK (COG-UK) Consortium |
| EPI_ISL_439015 | West of Scotland Specialist Virology Centre, NHSGGC / MRC-University of Glasgow Centre for Virus Research | COVID-19 Genomics UK (COG-UK) Consortium |
| EPI_ISL_439016 | West of Scotland Specialist Virology Centre, NHSGGC / MRC-University of Glasgow Centre for Virus Research | COVID-19 Genomics UK (COG-UK) Consortium |
| EPI_ISL_439017 | West of Scotland Specialist Virology Centre, NHSGGC / MRC-University of Glasgow Centre for Virus Research | COVID-19 Genomics UK (COG-UK) Consortium |
| EPI_ISL_439018 | West of Scotland Specialist Virology Centre, NHSGGC / MRC-University of Glasgow Centre for Virus Research | COVID-19 Genomics UK (COG-UK) Consortium |
| EPI_ISL_439019 | West of Scotland Specialist Virology Centre, NHSGGC / MRC-University of Glasgow Centre for Virus Research | COVID-19 Genomics UK (COG-UK) Consortium |
| EPI_ISL_439020 | West of Scotland Specialist Virology Centre, NHSGGC / MRC-University of Glasgow Centre for Virus Research | COVID-19 Genomics UK (COG-UK) Consortium |
| EPI_ISL_439021 | West of Scotland Specialist Virology Centre, NHSGGC / MRC-University of Glasgow Centre for Virus Research | COVID-19 Genomics UK (COG-UK) Consortium |

| EPI_Accession_ID | Originating Laboratory | Submitting Laboratory |
|---|---|---|
| EPI_ISL_439022 | West of Scotland Specialist Virology Centre, NHSGGC / MRC-University of Glasgow Centre for Virus Research | COVID-19 Genomics UK (COG-UK) Consortium |
| EPI_ISL_439023 | West of Scotland Specialist Virology Centre, NHSGGC / MRC-University of Glasgow Centre for Virus Research | COVID-19 Genomics UK (COG-UK) Consortium |
| EPI_ISL_439024 | West of Scotland Specialist Virology Centre, NHSGGC / MRC-University of Glasgow Centre for Virus Research | COVID-19 Genomics UK (COG-UK) Consortium |
| EPI_ISL_439025 | West of Scotland Specialist Virology Centre, NHSGGC / MRC-University of Glasgow Centre for Virus Research | COVID-19 Genomics UK (COG-UK) Consortium |
| EPI_ISL_439026 | West of Scotland Specialist Virology Centre, NHSGGC / MRC-University of Glasgow Centre for Virus Research | COVID-19 Genomics UK (COG-UK) Consortium |
| EPI_ISL_439027 | West of Scotland Specialist Virology Centre, NHSGGC / MRC-University of Glasgow Centre for Virus Research | COVID-19 Genomics UK (COG-UK) Consortium |
| EPI_ISL_439028 | West of Scotland Specialist Virology Centre, NHSGGC / MRC-University of Glasgow Centre for Virus Research | COVID-19 Genomics UK (COG-UK) Consortium |
| EPI_ISL_439029 | West of Scotland Specialist Virology Centre, NHSGGC / MRC-University of Glasgow Centre for Virus Research | COVID-19 Genomics UK (COG-UK) Consortium |
| EPI_ISL_439030 | West of Scotland Specialist Virology Centre, NHSGGC / MRC-University of Glasgow Centre for Virus Research | COVID-19 Genomics UK (COG-UK) Consortium |
| EPI_ISL_439031 | West of Scotland Specialist Virology Centre, NHSGGC / MRC-University of Glasgow Centre for Virus Research | COVID-19 Genomics UK (COG-UK) Consortium |
| EPI_ISL_439032 | West of Scotland Specialist Virology Centre, NHSGGC / MRC-University of Glasgow Centre for Virus Research | COVID-19 Genomics UK (COG-UK) Consortium |
| EPI_ISL_439033 | West of Scotland Specialist Virology Centre, NHSGGC / MRC-University of Glasgow Centre for Virus Research | COVID-19 Genomics UK (COG-UK) Consortium |
| EPI_ISL_439034 | West of Scotland Specialist Virology Centre, NHSGGC / MRC-University of Glasgow Centre for Virus Research | COVID-19 Genomics UK (COG-UK) Consortium |
| EPI_ISL_439035 | West of Scotland Specialist Virology Centre, NHSGGC / MRC-University of Glasgow Centre for Virus Research | COVID-19 Genomics UK (COG-UK) Consortium |
| EPI_ISL_439036 | West of Scotland Specialist Virology Centre, NHSGGC / MRC-University of Glasgow Centre for Virus Research | COVID-19 Genomics UK (COG-UK) Consortium |
| EPI_ISL_439037 | West of Scotland Specialist Virology Centre, NHSGGC / MRC-University of Glasgow Centre for Virus Research | COVID-19 Genomics UK (COG-UK) Consortium |

| EPI_Accession_ID | Originating Laboratory | Submitting Laboratory |
|---|---|---|
| EPI_ISL_439038 | West of Scotland Specialist Virology Centre, NHSGGC / MRC-University of Glasgow Centre for Virus Research | COVID-19 Genomics UK (COG-UK) Consortium |
| EPI_ISL_439039 | West of Scotland Specialist Virology Centre, NHSGGC / MRC-University of Glasgow Centre for Virus Research | COVID-19 Genomics UK (COG-UK) Consortium |
| EPI_ISL_439040 | West of Scotland Specialist Virology Centre, NHSGGC / MRC-University of Glasgow Centre for Virus Research | COVID-19 Genomics UK (COG-UK) Consortium |
| EPI_ISL_439041 | West of Scotland Specialist Virology Centre, NHSGGC / MRC-University of Glasgow Centre for Virus Research | COVID-19 Genomics UK (COG-UK) Consortium |
| EPI_ISL_439042 | West of Scotland Specialist Virology Centre, NHSGGC / MRC-University of Glasgow Centre for Virus Research | COVID-19 Genomics UK (COG-UK) Consortium |
| EPI_ISL_439043 | West of Scotland Specialist Virology Centre, NHSGGC / MRC-University of Glasgow Centre for Virus Research | COVID-19 Genomics UK (COG-UK) Consortium |
| EPI_ISL_439044 | West of Scotland Specialist Virology Centre, NHSGGC / MRC-University of Glasgow Centre for Virus Research | COVID-19 Genomics UK (COG-UK) Consortium |
| EPI_ISL_439045 | West of Scotland Specialist Virology Centre, NHSGGC / MRC-University of Glasgow Centre for Virus Research | COVID-19 Genomics UK (COG-UK) Consortium |
| EPI_ISL_439046 | West of Scotland Specialist Virology Centre, NHSGGC / MRC-University of Glasgow Centre for Virus Research | COVID-19 Genomics UK (COG-UK) Consortium |
| EPI_ISL_439047 | West of Scotland Specialist Virology Centre, NHSGGC / MRC-University of Glasgow Centre for Virus Research | COVID-19 Genomics UK (COG-UK) Consortium |
| EPI_ISL_439048 | West of Scotland Specialist Virology Centre, NHSGGC / MRC-University of Glasgow Centre for Virus Research | COVID-19 Genomics UK (COG-UK) Consortium |
| EPI_ISL_439049 | West of Scotland Specialist Virology Centre, NHSGGC / MRC-University of Glasgow Centre for Virus Research | COVID-19 Genomics UK (COG-UK) Consortium |
| EPI_ISL_439050 | West of Scotland Specialist Virology Centre, NHSGGC / MRC-University of Glasgow Centre for Virus Research | COVID-19 Genomics UK (COG-UK) Consortium |
| EPI_ISL_439051 | West of Scotland Specialist Virology Centre, NHSGGC / MRC-University of Glasgow Centre for Virus Research | COVID-19 Genomics UK (COG-UK) Consortium |
| EPI_ISL_439052 | West of Scotland Specialist Virology Centre, NHSGGC / MRC-University of Glasgow Centre for Virus Research | COVID-19 Genomics UK (COG-UK) Consortium |
| EPI_ISL_439053 | West of Scotland Specialist Virology Centre, NHSGGC / MRC-University of Glasgow Centre for Virus Research | COVID-19 Genomics UK (COG-UK) Consortium |

| EPI_Accession_ID | Originating Laboratory | Submitting Laboratory |
|---|---|---|
| EPI_ISL_439054 | West of Scotland Specialist Virology Centre, NHSGGC / MRC-University of Glasgow Centre for Virus Research | COVID-19 Genomics UK (COG-UK) Consortium |
| EPI_ISL_439055 | West of Scotland Specialist Virology Centre, NHSGGC / MRC-University of Glasgow Centre for Virus Research | COVID-19 Genomics UK (COG-UK) Consortium |
| EPI_ISL_439056 | West of Scotland Specialist Virology Centre, NHSGGC / MRC-University of Glasgow Centre for Virus Research | COVID-19 Genomics UK (COG-UK) Consortium |
| EPI_ISL_439057 | West of Scotland Specialist Virology Centre, NHSGGC / MRC-University of Glasgow Centre for Virus Research | COVID-19 Genomics UK (COG-UK) Consortium |
| EPI_ISL_439058 | West of Scotland Specialist Virology Centre, NHSGGC / MRC-University of Glasgow Centre for Virus Research | COVID-19 Genomics UK (COG-UK) Consortium |
| EPI_ISL_439059 | West of Scotland Specialist Virology Centre, NHSGGC / MRC-University of Glasgow Centre for Virus Research | COVID-19 Genomics UK (COG-UK) Consortium |
| EPI_ISL_439060 | West of Scotland Specialist Virology Centre, NHSGGC / MRC-University of Glasgow Centre for Virus Research | COVID-19 Genomics UK (COG-UK) Consortium |
| EPI_ISL_439061 | West of Scotland Specialist Virology Centre, NHSGGC / MRC-University of Glasgow Centre for Virus Research | COVID-19 Genomics UK (COG-UK) Consortium |
| EPI_ISL_439062 | West of Scotland Specialist Virology Centre, NHSGGC / MRC-University of Glasgow Centre for Virus Research | COVID-19 Genomics UK (COG-UK) Consortium |
| EPI_ISL_439063 | West of Scotland Specialist Virology Centre, NHSGGC / MRC-University of Glasgow Centre for Virus Research | COVID-19 Genomics UK (COG-UK) Consortium |
| EPI_ISL_439064 | West of Scotland Specialist Virology Centre, NHSGGC / MRC-University of Glasgow Centre for Virus Research | COVID-19 Genomics UK (COG-UK) Consortium |
| EPI_ISL_439065 | West of Scotland Specialist Virology Centre, NHSGGC / MRC-University of Glasgow Centre for Virus Research | COVID-19 Genomics UK (COG-UK) Consortium |
| EPI_ISL_439066 | West of Scotland Specialist Virology Centre, NHSGGC / MRC-University of Glasgow Centre for Virus Research | COVID-19 Genomics UK (COG-UK) Consortium |
| EPI_ISL_439067 | West of Scotland Specialist Virology Centre, NHSGGC / MRC-University of Glasgow Centre for Virus Research | COVID-19 Genomics UK (COG-UK) Consortium |
| EPI_ISL_439068 | West of Scotland Specialist Virology Centre, NHSGGC / MRC-University of Glasgow Centre for Virus Research | COVID-19 Genomics UK (COG-UK) Consortium |
| EPI_ISL_439069 | West of Scotland Specialist Virology Centre, NHSGGC / MRC-University of Glasgow Centre for Virus Research | COVID-19 Genomics UK (COG-UK) Consortium |

| EPI_Accession_ID | Originating Laboratory | Submitting Laboratory |
|---|---|---|
| EPI_ISL_439070 | West of Scotland Specialist Virology Centre, NHSGGC / MRC-University of Glasgow Centre for Virus Research | COVID-19 Genomics UK (COG-UK) Consortium |
| EPI_ISL_439071 | West of Scotland Specialist Virology Centre, NHSGGC / MRC-University of Glasgow Centre for Virus Research | COVID-19 Genomics UK (COG-UK) Consortium |
| EPI_ISL_439072 | West of Scotland Specialist Virology Centre, NHSGGC / MRC-University of Glasgow Centre for Virus Research | COVID-19 Genomics UK (COG-UK) Consortium |
| EPI_ISL_439073 | West of Scotland Specialist Virology Centre, NHSGGC / MRC-University of Glasgow Centre for Virus Research | COVID-19 Genomics UK (COG-UK) Consortium |
| EPI_ISL_439074 | West of Scotland Specialist Virology Centre, NHSGGC / MRC-University of Glasgow Centre for Virus Research | COVID-19 Genomics UK (COG-UK) Consortium |
| EPI_ISL_439075 | West of Scotland Specialist Virology Centre, NHSGGC / MRC-University of Glasgow Centre for Virus Research | COVID-19 Genomics UK (COG-UK) Consortium |
| EPI_ISL_439076 | West of Scotland Specialist Virology Centre, NHSGGC / MRC-University of Glasgow Centre for Virus Research | COVID-19 Genomics UK (COG-UK) Consortium |
| EPI_ISL_439077 | West of Scotland Specialist Virology Centre, NHSGGC / MRC-University of Glasgow Centre for Virus Research | COVID-19 Genomics UK (COG-UK) Consortium |
| EPI_ISL_439078 | West of Scotland Specialist Virology Centre, NHSGGC / MRC-University of Glasgow Centre for Virus Research | COVID-19 Genomics UK (COG-UK) Consortium |
| EPI_ISL_439079 | West of Scotland Specialist Virology Centre, NHSGGC / MRC-University of Glasgow Centre for Virus Research | COVID-19 Genomics UK (COG-UK) Consortium |
| EPI_ISL_439080 | West of Scotland Specialist Virology Centre, NHSGGC / MRC-University of Glasgow Centre for Virus Research | COVID-19 Genomics UK (COG-UK) Consortium |
| EPI_ISL_439081 | West of Scotland Specialist Virology Centre, NHSGGC / MRC-University of Glasgow Centre for Virus Research | COVID-19 Genomics UK (COG-UK) Consortium |
| EPI_ISL_439082 | West of Scotland Specialist Virology Centre, NHSGGC / MRC-University of Glasgow Centre for Virus Research | COVID-19 Genomics UK (COG-UK) Consortium |
| EPI_ISL_439083 | West of Scotland Specialist Virology Centre, NHSGGC / MRC-University of Glasgow Centre for Virus Research | COVID-19 Genomics UK (COG-UK) Consortium |
| EPI_ISL_439084 | West of Scotland Specialist Virology Centre, NHSGGC / MRC-University of Glasgow Centre for Virus Research | COVID-19 Genomics UK (COG-UK) Consortium |
| EPI_ISL_439085 | West of Scotland Specialist Virology Centre, NHSGGC / MRC-University of Glasgow Centre for Virus Research | COVID-19 Genomics UK (COG-UK) Consortium |

| EPI_Accession_ID | Originating Laboratory | Submitting Laboratory |
|---|---|---|
| EPI_ISL_439086 | West of Scotland Specialist Virology Centre, NHSGGC / MRC-University of Glasgow Centre for Virus Research | COVID-19 Genomics UK (COG-UK) Consortium |
| EPI_ISL_439087 | West of Scotland Specialist Virology Centre, NHSGGC / MRC-University of Glasgow Centre for Virus Research | COVID-19 Genomics UK (COG-UK) Consortium |
| EPI_ISL_439088 | West of Scotland Specialist Virology Centre, NHSGGC / MRC-University of Glasgow Centre for Virus Research | COVID-19 Genomics UK (COG-UK) Consortium |
| EPI_ISL_439089 | West of Scotland Specialist Virology Centre, NHSGGC / MRC-University of Glasgow Centre for Virus Research | COVID-19 Genomics UK (COG-UK) Consortium |
| EPI_ISL_439090 | West of Scotland Specialist Virology Centre, NHSGGC / MRC-University of Glasgow Centre for Virus Research | COVID-19 Genomics UK (COG-UK) Consortium |
| EPI_ISL_439091 | West of Scotland Specialist Virology Centre, NHSGGC / MRC-University of Glasgow Centre for Virus Research | COVID-19 Genomics UK (COG-UK) Consortium |
| EPI_ISL_439092 | West of Scotland Specialist Virology Centre, NHSGGC / MRC-University of Glasgow Centre for Virus Research | COVID-19 Genomics UK (COG-UK) Consortium |
| EPI_ISL_439093 | West of Scotland Specialist Virology Centre, NHSGGC / MRC-University of Glasgow Centre for Virus Research | COVID-19 Genomics UK (COG-UK) Consortium |
| EPI_ISL_439094 | West of Scotland Specialist Virology Centre, NHSGGC / MRC-University of Glasgow Centre for Virus Research | COVID-19 Genomics UK (COG-UK) Consortium |
| EPI_ISL_439095 | West of Scotland Specialist Virology Centre, NHSGGC / MRC-University of Glasgow Centre for Virus Research | COVID-19 Genomics UK (COG-UK) Consortium |
| EPI_ISL_439096 | West of Scotland Specialist Virology Centre, NHSGGC / MRC-University of Glasgow Centre for Virus Research | COVID-19 Genomics UK (COG-UK) Consortium |
| EPI_ISL_439097 | West of Scotland Specialist Virology Centre, NHSGGC / MRC-University of Glasgow Centre for Virus Research | COVID-19 Genomics UK (COG-UK) Consortium |
| EPI_ISL_439098 | West of Scotland Specialist Virology Centre, NHSGGC / MRC-University of Glasgow Centre for Virus Research | COVID-19 Genomics UK (COG-UK) Consortium |
| EPI_ISL_439099 | West of Scotland Specialist Virology Centre, NHSGGC / MRC-University of Glasgow Centre for Virus Research | COVID-19 Genomics UK (COG-UK) Consortium |
| EPI_ISL_439100 | West of Scotland Specialist Virology Centre, NHSGGC / MRC-University of Glasgow Centre for Virus Research | COVID-19 Genomics UK (COG-UK) Consortium |
| EPI_ISL_439101 | West of Scotland Specialist Virology Centre, NHSGGC / MRC-University of Glasgow Centre for Virus Research | COVID-19 Genomics UK (COG-UK) Consortium |

| EPI_Accession_ID | Originating Laboratory | Submitting Laboratory |
|---|---|---|
| EPI_ISL_439102 | West of Scotland Specialist Virology Centre, NHSGGC / MRC-University of Glasgow Centre for Virus Research | COVID-19 Genomics UK (COG-UK) Consortium |
| EPI_ISL_439103 | West of Scotland Specialist Virology Centre, NHSGGC / MRC-University of Glasgow Centre for Virus Research | COVID-19 Genomics UK (COG-UK) Consortium |
| EPI_ISL_439104 | West of Scotland Specialist Virology Centre, NHSGGC / MRC-University of Glasgow Centre for Virus Research | COVID-19 Genomics UK (COG-UK) Consortium |
| EPI_ISL_439105 | West of Scotland Specialist Virology Centre, NHSGGC / MRC-University of Glasgow Centre for Virus Research | COVID-19 Genomics UK (COG-UK) Consortium |
| EPI_ISL_439106 | West of Scotland Specialist Virology Centre, NHSGGC / MRC-University of Glasgow Centre for Virus Research | COVID-19 Genomics UK (COG-UK) Consortium |
| EPI_ISL_439107 | West of Scotland Specialist Virology Centre, NHSGGC / MRC-University of Glasgow Centre for Virus Research | COVID-19 Genomics UK (COG-UK) Consortium |
| EPI_ISL_439108 | West of Scotland Specialist Virology Centre, NHSGGC / MRC-University of Glasgow Centre for Virus Research | COVID-19 Genomics UK (COG-UK) Consortium |
| EPI_ISL_439109 | West of Scotland Specialist Virology Centre, NHSGGC / MRC-University of Glasgow Centre for Virus Research | COVID-19 Genomics UK (COG-UK) Consortium |
| EPI_ISL_439110 | West of Scotland Specialist Virology Centre, NHSGGC / MRC-University of Glasgow Centre for Virus Research | COVID-19 Genomics UK (COG-UK) Consortium |
| EPI_ISL_439111 | West of Scotland Specialist Virology Centre, NHSGGC / MRC-University of Glasgow Centre for Virus Research | COVID-19 Genomics UK (COG-UK) Consortium |
| EPI_ISL_439112 | West of Scotland Specialist Virology Centre, NHSGGC / MRC-University of Glasgow Centre for Virus Research | COVID-19 Genomics UK (COG-UK) Consortium |
| EPI_ISL_439113 | West of Scotland Specialist Virology Centre, NHSGGC / MRC-University of Glasgow Centre for Virus Research | COVID-19 Genomics UK (COG-UK) Consortium |
| EPI_ISL_439114 | West of Scotland Specialist Virology Centre, NHSGGC / MRC-University of Glasgow Centre for Virus Research | COVID-19 Genomics UK (COG-UK) Consortium |
| EPI_ISL_439115 | West of Scotland Specialist Virology Centre, NHSGGC / MRC-University of Glasgow Centre for Virus Research | COVID-19 Genomics UK (COG-UK) Consortium |
| EPI_ISL_439116 | West of Scotland Specialist Virology Centre, NHSGGC / MRC-University of Glasgow Centre for Virus Research | COVID-19 Genomics UK (COG-UK) Consortium |
| EPI_ISL_439117 | West of Scotland Specialist Virology Centre, NHSGGC / MRC-University of Glasgow Centre for Virus Research | COVID-19 Genomics UK (COG-UK) Consortium |

| EPI_Accession_ID | Originating Laboratory | Submitting Laboratory |
|---|---|---|
| EPI_ISL_439118 | West of Scotland Specialist Virology Centre, NHSGGC / MRC-University of Glasgow Centre for Virus Research | COVID-19 Genomics UK (COG-UK) Consortium |
| EPI_ISL_439119 | West of Scotland Specialist Virology Centre, NHSGGC / MRC-University of Glasgow Centre for Virus Research | COVID-19 Genomics UK (COG-UK) Consortium |
| EPI_ISL_439120 | West of Scotland Specialist Virology Centre, NHSGGC / MRC-University of Glasgow Centre for Virus Research | COVID-19 Genomics UK (COG-UK) Consortium |
| EPI_ISL_439121 | West of Scotland Specialist Virology Centre, NHSGGC / MRC-University of Glasgow Centre for Virus Research | COVID-19 Genomics UK (COG-UK) Consortium |
| EPI_ISL_439122 | West of Scotland Specialist Virology Centre, NHSGGC / MRC-University of Glasgow Centre for Virus Research | COVID-19 Genomics UK (COG-UK) Consortium |
| EPI_ISL_439123 | West of Scotland Specialist Virology Centre, NHSGGC / MRC-University of Glasgow Centre for Virus Research | COVID-19 Genomics UK (COG-UK) Consortium |
| EPI_ISL_439124 | West of Scotland Specialist Virology Centre, NHSGGC / MRC-University of Glasgow Centre for Virus Research | COVID-19 Genomics UK (COG-UK) Consortium |
| EPI_ISL_439125 | West of Scotland Specialist Virology Centre, NHSGGC / MRC-University of Glasgow Centre for Virus Research | COVID-19 Genomics UK (COG-UK) Consortium |
| EPI_ISL_439126 | West of Scotland Specialist Virology Centre, NHSGGC / MRC-University of Glasgow Centre for Virus Research | COVID-19 Genomics UK (COG-UK) Consortium |
| EPI_ISL_439127 | West of Scotland Specialist Virology Centre, NHSGGC / MRC-University of Glasgow Centre for Virus Research | COVID-19 Genomics UK (COG-UK) Consortium |
| EPI_ISL_439128 | West of Scotland Specialist Virology Centre, NHSGGC / MRC-University of Glasgow Centre for Virus Research | COVID-19 Genomics UK (COG-UK) Consortium |
| EPI_ISL_439129 | West of Scotland Specialist Virology Centre, NHSGGC / MRC-University of Glasgow Centre for Virus Research | COVID-19 Genomics UK (COG-UK) Consortium |
| EPI_ISL_439130 | West of Scotland Specialist Virology Centre, NHSGGC / MRC-University of Glasgow Centre for Virus Research | COVID-19 Genomics UK (COG-UK) Consortium |
| EPI_ISL_439131 | West of Scotland Specialist Virology Centre, NHSGGC / MRC-University of Glasgow Centre for Virus Research | COVID-19 Genomics UK (COG-UK) Consortium |
| EPI_ISL_439132 | West of Scotland Specialist Virology Centre, NHSGGC / MRC-University of Glasgow Centre for Virus Research | COVID-19 Genomics UK (COG-UK) Consortium |
| EPI_ISL_439133 | West of Scotland Specialist Virology Centre, NHSGGC / MRC-University of Glasgow Centre for Virus Research | COVID-19 Genomics UK (COG-UK) Consortium |

| EPI_Accession_ID | Originating Laboratory | Submitting Laboratory |
| --- | --- | --- |
| EPI_ISL_439134 | West of Scotland Specialist Virology Centre, NHSGGC / MRC-University of Glasgow Centre for Virus Research | COVID-19 Genomics UK (COG-UK) Consortium |
| EPI_ISL_439135 | West of Scotland Specialist Virology Centre, NHSGGC / MRC-University of Glasgow Centre for Virus Research | COVID-19 Genomics UK (COG-UK) Consortium |
| EPI_ISL_439136 | West of Scotland Specialist Virology Centre, NHSGGC / MRC-University of Glasgow Centre for Virus Research | COVID-19 Genomics UK (COG-UK) Consortium |
| EPI_ISL_439137 | West of Scotland Specialist Virology Centre, NHSGGC / MRC-University of Glasgow Centre for Virus Research | COVID-19 Genomics UK (COG-UK) Consortium |
| EPI_ISL_439138 | West of Scotland Specialist Virology Centre, NHSGGC / MRC-University of Glasgow Centre for Virus Research | COVID-19 Genomics UK (COG-UK) Consortium |
| EPI_ISL_439139 | West of Scotland Specialist Virology Centre, NHSGGC / MRC-University of Glasgow Centre for Virus Research | COVID-19 Genomics UK (COG-UK) Consortium |
| EPI_ISL_439140 | West of Scotland Specialist Virology Centre, NHSGGC / MRC-University of Glasgow Centre for Virus Research | COVID-19 Genomics UK (COG-UK) Consortium |
| EPI_ISL_439141 | West of Scotland Specialist Virology Centre, NHSGGC / MRC-University of Glasgow Centre for Virus Research | COVID-19 Genomics UK (COG-UK) Consortium |
| EPI_ISL_439142 | West of Scotland Specialist Virology Centre, NHSGGC / MRC-University of Glasgow Centre for Virus Research | COVID-19 Genomics UK (COG-UK) Consortium |
| EPI_ISL_439143 | West of Scotland Specialist Virology Centre, NHSGGC / MRC-University of Glasgow Centre for Virus Research | COVID-19 Genomics UK (COG-UK) Consortium |
| EPI_ISL_439144 | Virology Department, Royal Infirmary of Edinburgh, NHS Lothian / School of Biological Sciences, University of Edinburgh / Institute of Genetics and Molecular Medicine, University of Edinburgh | COVID-19 Genomics UK (COG-UK) Consortium |
| EPI_ISL_439145 | Virology Department, Royal Infirmary of Edinburgh, NHS Lothian / School of Biological Sciences, University of Edinburgh / Institute of Genetics and Molecular Medicine, University of Edinburgh | COVID-19 Genomics UK (COG-UK) Consortium |
| EPI_ISL_439146 | Virology Department, Royal Infirmary of Edinburgh, NHS Lothian / School of Biological Sciences, University of Edinburgh / Institute of Genetics and Molecular Medicine, University of Edinburgh | COVID-19 Genomics UK (COG-UK) Consortium |

| EPI_Accession_ID | Originating Laboratory | Submitting Laboratory |
|---|---|---|
| EPI_ISL_439147 | Virology Department, Royal Infirmary of Edinburgh, NHS Lothian / School of Biological Sciences, University of Edinburgh / Institute of Genetics and Molecular Medicine, University of Edinburgh | COVID-19 Genomics UK (COG-UK) Consortium |
| EPI_ISL_439148 | Virology Department, Royal Infirmary of Edinburgh, NHS Lothian / School of Biological Sciences, University of Edinburgh / Institute of Genetics and Molecular Medicine, University of Edinburgh | COVID-19 Genomics UK (COG-UK) Consortium |
| EPI_ISL_439149 | Virology Department, Royal Infirmary of Edinburgh, NHS Lothian / School of Biological Sciences, University of Edinburgh / Institute of Genetics and Molecular Medicine, University of Edinburgh | COVID-19 Genomics UK (COG-UK) Consortium |
| EPI_ISL_439150 | Virology Department, Royal Infirmary of Edinburgh, NHS Lothian / School of Biological Sciences, University of Edinburgh / Institute of Genetics and Molecular Medicine, University of Edinburgh | COVID-19 Genomics UK (COG-UK) Consortium |
| EPI_ISL_439151 | Virology Department, Royal Infirmary of Edinburgh, NHS Lothian / School of Biological Sciences, University of Edinburgh / Institute of Genetics and Molecular Medicine, University of Edinburgh | COVID-19 Genomics UK (COG-UK) Consortium |
| EPI_ISL_439152 | Virology Department, Royal Infirmary of Edinburgh, NHS Lothian / School of Biological Sciences, University of Edinburgh / Institute of Genetics and Molecular Medicine, University of Edinburgh | COVID-19 Genomics UK (COG-UK) Consortium |
| EPI_ISL_439153 | Virology Department, Royal Infirmary of Edinburgh, NHS Lothian / School of Biological Sciences, University of Edinburgh / Institute of Genetics and Molecular Medicine, University of Edinburgh | COVID-19 Genomics UK (COG-UK) Consortium |
| EPI_ISL_439154 | Virology Department, Royal Infirmary of Edinburgh, NHS Lothian / School of Biological Sciences, University of Edinburgh / Institute of Genetics and Molecular Medicine, University of Edinburgh | COVID-19 Genomics UK (COG-UK) Consortium |

| EPI_Accession_ID | Originating Laboratory | Submitting Laboratory |
|---|---|---|
| EPI_ISL_439155 | Virology Department, Royal Infirmary of Edinburgh, NHS Lothian / School of Biological Sciences, University of Edinburgh / Institute of Genetics and Molecular Medicine, University of Edinburgh | COVID-19 Genomics UK (COG-UK) Consortium |
| EPI_ISL_439156 | Virology Department, Royal Infirmary of Edinburgh, NHS Lothian / School of Biological Sciences, University of Edinburgh / Institute of Genetics and Molecular Medicine, University of Edinburgh | COVID-19 Genomics UK (COG-UK) Consortium |
| EPI_ISL_439157 | Virology Department, Royal Infirmary of Edinburgh, NHS Lothian / School of Biological Sciences, University of Edinburgh / Institute of Genetics and Molecular Medicine, University of Edinburgh | COVID-19 Genomics UK (COG-UK) Consortium |
| EPI_ISL_439158 | Virology Department, Royal Infirmary of Edinburgh, NHS Lothian / School of Biological Sciences, University of Edinburgh / Institute of Genetics and Molecular Medicine, University of Edinburgh | COVID-19 Genomics UK (COG-UK) Consortium |
| EPI_ISL_439159 | Virology Department, Royal Infirmary of Edinburgh, NHS Lothian / School of Biological Sciences, University of Edinburgh / Institute of Genetics and Molecular Medicine, University of Edinburgh | COVID-19 Genomics UK (COG-UK) Consortium |
| EPI_ISL_439160 | Virology Department, Royal Infirmary of Edinburgh, NHS Lothian / School of Biological Sciences, University of Edinburgh / Institute of Genetics and Molecular Medicine, University of Edinburgh | COVID-19 Genomics UK (COG-UK) Consortium |
| EPI_ISL_439161 | Virology Department, Royal Infirmary of Edinburgh, NHS Lothian / School of Biological Sciences, University of Edinburgh / Institute of Genetics and Molecular Medicine, University of Edinburgh | COVID-19 Genomics UK (COG-UK) Consortium |
| EPI_ISL_439162 | Virology Department, Royal Infirmary of Edinburgh, NHS Lothian / School of Biological Sciences, University of Edinburgh / Institute of Genetics and Molecular Medicine, University of Edinburgh | COVID-19 Genomics UK (COG-UK) Consortium |

| EPI_Accession_ID | Originating Laboratory | Submitting Laboratory |
|---|---|---|
| EPI_ISL_439163 | Virology Department, Royal Infirmary of Edinburgh, NHS Lothian / School of Biological Sciences, University of Edinburgh / Institute of Genetics and Molecular Medicine, University of Edinburgh | COVID-19 Genomics UK (COG-UK) Consortium |
| EPI_ISL_439164 | Virology Department, Royal Infirmary of Edinburgh, NHS Lothian / School of Biological Sciences, University of Edinburgh / Institute of Genetics and Molecular Medicine, University of Edinburgh | COVID-19 Genomics UK (COG-UK) Consortium |
| EPI_ISL_439165 | Virology Department, Royal Infirmary of Edinburgh, NHS Lothian / School of Biological Sciences, University of Edinburgh / Institute of Genetics and Molecular Medicine, University of Edinburgh | COVID-19 Genomics UK (COG-UK) Consortium |
| EPI_ISL_439166 | Virology Department, Royal Infirmary of Edinburgh, NHS Lothian / School of Biological Sciences, University of Edinburgh / Institute of Genetics and Molecular Medicine, University of Edinburgh | COVID-19 Genomics UK (COG-UK) Consortium |
| EPI_ISL_439167 | Virology Department, Royal Infirmary of Edinburgh, NHS Lothian / School of Biological Sciences, University of Edinburgh / Institute of Genetics and Molecular Medicine, University of Edinburgh | COVID-19 Genomics UK (COG-UK) Consortium |
| EPI_ISL_439168 | Virology Department, Royal Infirmary of Edinburgh, NHS Lothian / School of Biological Sciences, University of Edinburgh / Institute of Genetics and Molecular Medicine, University of Edinburgh | COVID-19 Genomics UK (COG-UK) Consortium |
| EPI_ISL_439169 | Virology Department, Royal Infirmary of Edinburgh, NHS Lothian / School of Biological Sciences, University of Edinburgh / Institute of Genetics and Molecular Medicine, University of Edinburgh | COVID-19 Genomics UK (COG-UK) Consortium |
| EPI_ISL_439170 | Virology Department, Royal Infirmary of Edinburgh, NHS Lothian / School of Biological Sciences, University of Edinburgh / Institute of Genetics and Molecular Medicine, University of Edinburgh | COVID-19 Genomics UK (COG-UK) Consortium |

| EPI_Accession_ID | Originating Laboratory | Submitting Laboratory |
| --- | --- | --- |
| EPI_ISL_439171 | Virology Department, Royal Infirmary of Edinburgh, NHS Lothian / School of Biological Sciences, University of Edinburgh / Institute of Genetics and Molecular Medicine, University of Edinburgh | COVID-19 Genomics UK (COG-UK) Consortium |
| EPI_ISL_439172 | Virology Department, Royal Infirmary of Edinburgh, NHS Lothian / School of Biological Sciences, University of Edinburgh / Institute of Genetics and Molecular Medicine, University of Edinburgh | COVID-19 Genomics UK (COG-UK) Consortium |
| EPI_ISL_439173 | Virology Department, Royal Infirmary of Edinburgh, NHS Lothian / School of Biological Sciences, University of Edinburgh / Institute of Genetics and Molecular Medicine, University of Edinburgh | COVID-19 Genomics UK (COG-UK) Consortium |
| EPI_ISL_439174 | Virology Department, Royal Infirmary of Edinburgh, NHS Lothian / School of Biological Sciences, University of Edinburgh / Institute of Genetics and Molecular Medicine, University of Edinburgh | COVID-19 Genomics UK (COG-UK) Consortium |
| EPI_ISL_439175 | Virology Department, Royal Infirmary of Edinburgh, NHS Lothian / School of Biological Sciences, University of Edinburgh / Institute of Genetics and Molecular Medicine, University of Edinburgh | COVID-19 Genomics UK (COG-UK) Consortium |
| EPI_ISL_439176 | Virology Department, Royal Infirmary of Edinburgh, NHS Lothian / School of Biological Sciences, University of Edinburgh / Institute of Genetics and Molecular Medicine, University of Edinburgh | COVID-19 Genomics UK (COG-UK) Consortium |
| EPI_ISL_439177 | Virology Department, Royal Infirmary of Edinburgh, NHS Lothian / School of Biological Sciences, University of Edinburgh / Institute of Genetics and Molecular Medicine, University of Edinburgh | COVID-19 Genomics UK (COG-UK) Consortium |
| EPI_ISL_439178 | Virology Department, Royal Infirmary of Edinburgh, NHS Lothian / School of Biological Sciences, University of Edinburgh / Institute of Genetics and Molecular Medicine, University of Edinburgh | COVID-19 Genomics UK (COG-UK) Consortium |

| EPI_Accession_ID | Originating Laboratory | Submitting Laboratory |
|---|---|---|
| EPI_ISL_439179 | Virology Department, Royal Infirmary of Edinburgh, NHS Lothian / School of Biological Sciences, University of Edinburgh / Institute of Genetics and Molecular Medicine, University of Edinburgh | COVID-19 Genomics UK (COG-UK) Consortium |
| EPI_ISL_439180 | Virology Department, Royal Infirmary of Edinburgh, NHS Lothian / School of Biological Sciences, University of Edinburgh / Institute of Genetics and Molecular Medicine, University of Edinburgh | COVID-19 Genomics UK (COG-UK) Consortium |
| EPI_ISL_439181 | Virology Department, Royal Infirmary of Edinburgh, NHS Lothian / School of Biological Sciences, University of Edinburgh / Institute of Genetics and Molecular Medicine, University of Edinburgh | COVID-19 Genomics UK (COG-UK) Consortium |
| EPI_ISL_439182 | Virology Department, Royal Infirmary of Edinburgh, NHS Lothian / School of Biological Sciences, University of Edinburgh / Institute of Genetics and Molecular Medicine, University of Edinburgh | COVID-19 Genomics UK (COG-UK) Consortium |
| EPI_ISL_439183 | Virology Department, Royal Infirmary of Edinburgh, NHS Lothian / School of Biological Sciences, University of Edinburgh / Institute of Genetics and Molecular Medicine, University of Edinburgh | COVID-19 Genomics UK (COG-UK) Consortium |
| EPI_ISL_439184 | Virology Department, Royal Infirmary of Edinburgh, NHS Lothian / School of Biological Sciences, University of Edinburgh / Institute of Genetics and Molecular Medicine, University of Edinburgh | COVID-19 Genomics UK (COG-UK) Consortium |
| EPI_ISL_439185 | Virology Department, Royal Infirmary of Edinburgh, NHS Lothian / School of Biological Sciences, University of Edinburgh / Institute of Genetics and Molecular Medicine, University of Edinburgh | COVID-19 Genomics UK (COG-UK) Consortium |
| EPI_ISL_439186 | Virology Department, Royal Infirmary of Edinburgh, NHS Lothian / School of Biological Sciences, University of Edinburgh / Institute of Genetics and Molecular Medicine, University of Edinburgh | COVID-19 Genomics UK (COG-UK) Consortium |

| EPI_Accession_ID | Originating Laboratory | Submitting Laboratory |
|---|---|---|
| EPI_ISL_439187 | Virology Department, Royal Infirmary of Edinburgh, NHS Lothian / School of Biological Sciences, University of Edinburgh / Institute of Genetics and Molecular Medicine, University of Edinburgh | COVID-19 Genomics UK (COG-UK) Consortium |
| EPI_ISL_439188 | Virology Department, Royal Infirmary of Edinburgh, NHS Lothian / School of Biological Sciences, University of Edinburgh / Institute of Genetics and Molecular Medicine, University of Edinburgh | COVID-19 Genomics UK (COG-UK) Consortium |
| EPI_ISL_439189 | Virology Department, Royal Infirmary of Edinburgh, NHS Lothian / School of Biological Sciences, University of Edinburgh / Institute of Genetics and Molecular Medicine, University of Edinburgh | COVID-19 Genomics UK (COG-UK) Consortium |
| EPI_ISL_439190 | Virology Department, Royal Infirmary of Edinburgh, NHS Lothian / School of Biological Sciences, University of Edinburgh / Institute of Genetics and Molecular Medicine, University of Edinburgh | COVID-19 Genomics UK (COG-UK) Consortium |
| EPI_ISL_439191 | Virology Department, Royal Infirmary of Edinburgh, NHS Lothian / School of Biological Sciences, University of Edinburgh / Institute of Genetics and Molecular Medicine, University of Edinburgh | COVID-19 Genomics UK (COG-UK) Consortium |
| EPI_ISL_439192 | Virology Department, Royal Infirmary of Edinburgh, NHS Lothian / School of Biological Sciences, University of Edinburgh / Institute of Genetics and Molecular Medicine, University of Edinburgh | COVID-19 Genomics UK (COG-UK) Consortium |
| EPI_ISL_439193 | Virology Department, Royal Infirmary of Edinburgh, NHS Lothian / School of Biological Sciences, University of Edinburgh / Institute of Genetics and Molecular Medicine, University of Edinburgh | COVID-19 Genomics UK (COG-UK) Consortium |
| EPI_ISL_439194 | Virology Department, Royal Infirmary of Edinburgh, NHS Lothian / School of Biological Sciences, University of Edinburgh / Institute of Genetics and Molecular Medicine, University of Edinburgh | COVID-19 Genomics UK (COG-UK) Consortium |

| EPI_Accession_ID | Originating Laboratory | Submitting Laboratory |
| --- | --- | --- |
| EPI_ISL_439195 | Virology Department, Royal Infirmary of Edinburgh, NHS Lothian / School of Biological Sciences, University of Edinburgh / Institute of Genetics and Molecular Medicine, University of Edinburgh | COVID-19 Genomics UK (COG-UK) Consortium |
| EPI_ISL_439196 | Virology Department, Royal Infirmary of Edinburgh, NHS Lothian / School of Biological Sciences, University of Edinburgh / Institute of Genetics and Molecular Medicine, University of Edinburgh | COVID-19 Genomics UK (COG-UK) Consortium |
| EPI_ISL_439197 | Virology Department, Royal Infirmary of Edinburgh, NHS Lothian / School of Biological Sciences, University of Edinburgh / Institute of Genetics and Molecular Medicine, University of Edinburgh | COVID-19 Genomics UK (COG-UK) Consortium |
| EPI_ISL_439198 | Virology Department, Royal Infirmary of Edinburgh, NHS Lothian / School of Biological Sciences, University of Edinburgh / Institute of Genetics and Molecular Medicine, University of Edinburgh | COVID-19 Genomics UK (COG-UK) Consortium |
| EPI_ISL_439199 | Virology Department, Royal Infirmary of Edinburgh, NHS Lothian / School of Biological Sciences, University of Edinburgh / Institute of Genetics and Molecular Medicine, University of Edinburgh | COVID-19 Genomics UK (COG-UK) Consortium |
| EPI_ISL_439200 | Virology Department, Royal Infirmary of Edinburgh, NHS Lothian / School of Biological Sciences, University of Edinburgh / Institute of Genetics and Molecular Medicine, University of Edinburgh | COVID-19 Genomics UK (COG-UK) Consortium |
| EPI_ISL_439201 | Virology Department, Royal Infirmary of Edinburgh, NHS Lothian / School of Biological Sciences, University of Edinburgh / Institute of Genetics and Molecular Medicine, University of Edinburgh | COVID-19 Genomics UK (COG-UK) Consortium |
| EPI_ISL_439202 | Virology Department, Royal Infirmary of Edinburgh, NHS Lothian / School of Biological Sciences, University of Edinburgh / Institute of Genetics and Molecular Medicine, University of Edinburgh | COVID-19 Genomics UK (COG-UK) Consortium |

| EPI_Accession_ID | Originating Laboratory | Submitting Laboratory |
|---|---|---|
| EPI_ISL_439203 | Virology Department, Royal Infirmary of Edinburgh, NHS Lothian / School of Biological Sciences, University of Edinburgh / Institute of Genetics and Molecular Medicine, University of Edinburgh | COVID-19 Genomics UK (COG-UK) Consortium |
| EPI_ISL_439204 | Virology Department, Royal Infirmary of Edinburgh, NHS Lothian / School of Biological Sciences, University of Edinburgh / Institute of Genetics and Molecular Medicine, University of Edinburgh | COVID-19 Genomics UK (COG-UK) Consortium |
| EPI_ISL_439205 | Virology Department, Royal Infirmary of Edinburgh, NHS Lothian / School of Biological Sciences, University of Edinburgh / Institute of Genetics and Molecular Medicine, University of Edinburgh | COVID-19 Genomics UK (COG-UK) Consortium |
| EPI_ISL_439206 | Virology Department, Royal Infirmary of Edinburgh, NHS Lothian / School of Biological Sciences, University of Edinburgh / Institute of Genetics and Molecular Medicine, University of Edinburgh | COVID-19 Genomics UK (COG-UK) Consortium |
| EPI_ISL_439207 | Virology Department, Royal Infirmary of Edinburgh, NHS Lothian / School of Biological Sciences, University of Edinburgh / Institute of Genetics and Molecular Medicine, University of Edinburgh | COVID-19 Genomics UK (COG-UK) Consortium |
| EPI_ISL_439208 | Virology Department, Royal Infirmary of Edinburgh, NHS Lothian / School of Biological Sciences, University of Edinburgh / Institute of Genetics and Molecular Medicine, University of Edinburgh | COVID-19 Genomics UK (COG-UK) Consortium |
| EPI_ISL_439209 | Virology Department, Royal Infirmary of Edinburgh, NHS Lothian / School of Biological Sciences, University of Edinburgh / Institute of Genetics and Molecular Medicine, University of Edinburgh | COVID-19 Genomics UK (COG-UK) Consortium |
| EPI_ISL_439210 | Virology Department, Royal Infirmary of Edinburgh, NHS Lothian / School of Biological Sciences, University of Edinburgh / Institute of Genetics and Molecular Medicine, University of Edinburgh | COVID-19 Genomics UK (COG-UK) Consortium |

| EPI_Accession_ID | Originating Laboratory | Submitting Laboratory |
|---|---|---|
| EPI_ISL_439211 | Virology Department, Royal Infirmary of Edinburgh, NHS Lothian / School of Biological Sciences, University of Edinburgh / Institute of Genetics and Molecular Medicine, University of Edinburgh | COVID-19 Genomics UK (COG-UK) Consortium |
| EPI_ISL_439212 | Virology Department, Royal Infirmary of Edinburgh, NHS Lothian / School of Biological Sciences, University of Edinburgh / Institute of Genetics and Molecular Medicine, University of Edinburgh | COVID-19 Genomics UK (COG-UK) Consortium |
| EPI_ISL_439213 | Virology Department, Royal Infirmary of Edinburgh, NHS Lothian / School of Biological Sciences, University of Edinburgh / Institute of Genetics and Molecular Medicine, University of Edinburgh | COVID-19 Genomics UK (COG-UK) Consortium |
| EPI_ISL_439214 | Virology Department, Royal Infirmary of Edinburgh, NHS Lothian / School of Biological Sciences, University of Edinburgh / Institute of Genetics and Molecular Medicine, University of Edinburgh | COVID-19 Genomics UK (COG-UK) Consortium |
| EPI_ISL_439215 | Virology Department, Royal Infirmary of Edinburgh, NHS Lothian / School of Biological Sciences, University of Edinburgh / Institute of Genetics and Molecular Medicine, University of Edinburgh | COVID-19 Genomics UK (COG-UK) Consortium |
| EPI_ISL_439216 | Virology Department, Royal Infirmary of Edinburgh, NHS Lothian / School of Biological Sciences, University of Edinburgh / Institute of Genetics and Molecular Medicine, University of Edinburgh | COVID-19 Genomics UK (COG-UK) Consortium |
| EPI_ISL_439217 | Virology Department, Royal Infirmary of Edinburgh, NHS Lothian / School of Biological Sciences, University of Edinburgh / Institute of Genetics and Molecular Medicine, University of Edinburgh | COVID-19 Genomics UK (COG-UK) Consortium |
| EPI_ISL_439218 | Virology Department, Royal Infirmary of Edinburgh, NHS Lothian / School of Biological Sciences, University of Edinburgh / Institute of Genetics and Molecular Medicine, University of Edinburgh | COVID-19 Genomics UK (COG-UK) Consortium |

| EPI_Accession_ID | Originating Laboratory | Submitting Laboratory |
| --- | --- | --- |
| EPI_ISL_439219 | Virology Department, Royal Infirmary of Edinburgh, NHS Lothian / School of Biological Sciences, University of Edinburgh / Institute of Genetics and Molecular Medicine, University of Edinburgh | COVID-19 Genomics UK (COG-UK) Consortium |
| EPI_ISL_439220 | Virology Department, Royal Infirmary of Edinburgh, NHS Lothian / School of Biological Sciences, University of Edinburgh / Institute of Genetics and Molecular Medicine, University of Edinburgh | COVID-19 Genomics UK (COG-UK) Consortium |
| EPI_ISL_439221 | Virology Department, Royal Infirmary of Edinburgh, NHS Lothian / School of Biological Sciences, University of Edinburgh / Institute of Genetics and Molecular Medicine, University of Edinburgh | COVID-19 Genomics UK (COG-UK) Consortium |
| EPI_ISL_439222 | Virology Department, Royal Infirmary of Edinburgh, NHS Lothian / School of Biological Sciences, University of Edinburgh / Institute of Genetics and Molecular Medicine, University of Edinburgh | COVID-19 Genomics UK (COG-UK) Consortium |
| EPI_ISL_439223 | Virology Department, Royal Infirmary of Edinburgh, NHS Lothian / School of Biological Sciences, University of Edinburgh / Institute of Genetics and Molecular Medicine, University of Edinburgh | COVID-19 Genomics UK (COG-UK) Consortium |
| EPI_ISL_439224 | Virology Department, Royal Infirmary of Edinburgh, NHS Lothian / School of Biological Sciences, University of Edinburgh / Institute of Genetics and Molecular Medicine, University of Edinburgh | COVID-19 Genomics UK (COG-UK) Consortium |
| EPI_ISL_439225 | Virology Department, Royal Infirmary of Edinburgh, NHS Lothian / School of Biological Sciences, University of Edinburgh / Institute of Genetics and Molecular Medicine, University of Edinburgh | COVID-19 Genomics UK (COG-UK) Consortium |
| EPI_ISL_439226 | Virology Department, Royal Infirmary of Edinburgh, NHS Lothian / School of Biological Sciences, University of Edinburgh / Institute of Genetics and Molecular Medicine, University of Edinburgh | COVID-19 Genomics UK (COG-UK) Consortium |

| EPI_Accession_ID | Originating Laboratory | Submitting Laboratory |
|---|---|---|
| EPI_ISL_439227 | Virology Department, Royal Infirmary of Edinburgh, NHS Lothian / School of Biological Sciences, University of Edinburgh / Institute of Genetics and Molecular Medicine, University of Edinburgh | COVID-19 Genomics UK (COG-UK) Consortium |
| EPI_ISL_439228 | Virology Department, Royal Infirmary of Edinburgh, NHS Lothian / School of Biological Sciences, University of Edinburgh / Institute of Genetics and Molecular Medicine, University of Edinburgh | COVID-19 Genomics UK (COG-UK) Consortium |
| EPI_ISL_439229 | Virology Department, Royal Infirmary of Edinburgh, NHS Lothian / School of Biological Sciences, University of Edinburgh / Institute of Genetics and Molecular Medicine, University of Edinburgh | COVID-19 Genomics UK (COG-UK) Consortium |
| EPI_ISL_439230 | Virology Department, Royal Infirmary of Edinburgh, NHS Lothian / School of Biological Sciences, University of Edinburgh / Institute of Genetics and Molecular Medicine, University of Edinburgh | COVID-19 Genomics UK (COG-UK) Consortium |
| EPI_ISL_439231 | Virology Department, Royal Infirmary of Edinburgh, NHS Lothian / School of Biological Sciences, University of Edinburgh / Institute of Genetics and Molecular Medicine, University of Edinburgh | COVID-19 Genomics UK (COG-UK) Consortium |
| EPI_ISL_439232 | Virology Department, Royal Infirmary of Edinburgh, NHS Lothian / School of Biological Sciences, University of Edinburgh / Institute of Genetics and Molecular Medicine, University of Edinburgh | COVID-19 Genomics UK (COG-UK) Consortium |
| EPI_ISL_439233 | Virology Department, Royal Infirmary of Edinburgh, NHS Lothian / School of Biological Sciences, University of Edinburgh / Institute of Genetics and Molecular Medicine, University of Edinburgh | COVID-19 Genomics UK (COG-UK) Consortium |
| EPI_ISL_439234 | Virology Department, Royal Infirmary of Edinburgh, NHS Lothian / School of Biological Sciences, University of Edinburgh / Institute of Genetics and Molecular Medicine, University of Edinburgh | COVID-19 Genomics UK (COG-UK) Consortium |

| EPI_Accession_ID | Originating Laboratory | Submitting Laboratory |
|---|---|---|
| EPI_ISL_439235 | Virology Department, Royal Infirmary of Edinburgh, NHS Lothian / School of Biological Sciences, University of Edinburgh / Institute of Genetics and Molecular Medicine, University of Edinburgh | COVID-19 Genomics UK (COG-UK) Consortium |
| EPI_ISL_439236 | Virology Department, Royal Infirmary of Edinburgh, NHS Lothian / School of Biological Sciences, University of Edinburgh / Institute of Genetics and Molecular Medicine, University of Edinburgh | COVID-19 Genomics UK (COG-UK) Consortium |
| EPI_ISL_439237 | Virology Department, Royal Infirmary of Edinburgh, NHS Lothian / School of Biological Sciences, University of Edinburgh / Institute of Genetics and Molecular Medicine, University of Edinburgh | COVID-19 Genomics UK (COG-UK) Consortium |
| EPI_ISL_439238 | Virology Department, Royal Infirmary of Edinburgh, NHS Lothian / School of Biological Sciences, University of Edinburgh / Institute of Genetics and Molecular Medicine, University of Edinburgh | COVID-19 Genomics UK (COG-UK) Consortium |
| EPI_ISL_439239 | Virology Department, Royal Infirmary of Edinburgh, NHS Lothian / School of Biological Sciences, University of Edinburgh / Institute of Genetics and Molecular Medicine, University of Edinburgh | COVID-19 Genomics UK (COG-UK) Consortium |
| EPI_ISL_439240 | Virology Department, Royal Infirmary of Edinburgh, NHS Lothian / School of Biological Sciences, University of Edinburgh / Institute of Genetics and Molecular Medicine, University of Edinburgh | COVID-19 Genomics UK (COG-UK) Consortium |
| EPI_ISL_439241 | Virology Department, Royal Infirmary of Edinburgh, NHS Lothian / School of Biological Sciences, University of Edinburgh / Institute of Genetics and Molecular Medicine, University of Edinburgh | COVID-19 Genomics UK (COG-UK) Consortium |
| EPI_ISL_439242 | Virology Department, Royal Infirmary of Edinburgh, NHS Lothian / School of Biological Sciences, University of Edinburgh / Institute of Genetics and Molecular Medicine, University of Edinburgh | COVID-19 Genomics UK (COG-UK) Consortium |

| EPI_Accession_ID | Originating Laboratory | Submitting Laboratory |
|---|---|---|
| EPI_ISL_439243 | Virology Department, Royal Infirmary of Edinburgh, NHS Lothian / School of Biological Sciences, University of Edinburgh / Institute of Genetics and Molecular Medicine, University of Edinburgh | COVID-19 Genomics UK (COG-UK) Consortium |
| EPI_ISL_439244 | Virology Department, Royal Infirmary of Edinburgh, NHS Lothian / School of Biological Sciences, University of Edinburgh / Institute of Genetics and Molecular Medicine, University of Edinburgh | COVID-19 Genomics UK (COG-UK) Consortium |
| EPI_ISL_439245 | Virology Department, Royal Infirmary of Edinburgh, NHS Lothian / School of Biological Sciences, University of Edinburgh / Institute of Genetics and Molecular Medicine, University of Edinburgh | COVID-19 Genomics UK (COG-UK) Consortium |
| EPI_ISL_439246 | Virology Department, Royal Infirmary of Edinburgh, NHS Lothian / School of Biological Sciences, University of Edinburgh / Institute of Genetics and Molecular Medicine, University of Edinburgh | COVID-19 Genomics UK (COG-UK) Consortium |
| EPI_ISL_439247 | Virology Department, Royal Infirmary of Edinburgh, NHS Lothian / School of Biological Sciences, University of Edinburgh / Institute of Genetics and Molecular Medicine, University of Edinburgh | COVID-19 Genomics UK (COG-UK) Consortium |
| EPI_ISL_439248 | Virology Department, Royal Infirmary of Edinburgh, NHS Lothian / School of Biological Sciences, University of Edinburgh / Institute of Genetics and Molecular Medicine, University of Edinburgh | COVID-19 Genomics UK (COG-UK) Consortium |
| EPI_ISL_439249 | Virology Department, Royal Infirmary of Edinburgh, NHS Lothian / School of Biological Sciences, University of Edinburgh / Institute of Genetics and Molecular Medicine, University of Edinburgh | COVID-19 Genomics UK (COG-UK) Consortium |
| EPI_ISL_439250 | Virology Department, Royal Infirmary of Edinburgh, NHS Lothian / School of Biological Sciences, University of Edinburgh / Institute of Genetics and Molecular Medicine, University of Edinburgh | COVID-19 Genomics UK (COG-UK) Consortium |

| EPI_Accession_ID | Originating Laboratory | Submitting Laboratory |
|---|---|---|
| EPI_ISL_439251 | Virology Department, Royal Infirmary of Edinburgh, NHS Lothian / School of Biological Sciences, University of Edinburgh / Institute of Genetics and Molecular Medicine, University of Edinburgh | COVID-19 Genomics UK (COG-UK) Consortium |
| EPI_ISL_439252 | Virology Department, Royal Infirmary of Edinburgh, NHS Lothian / School of Biological Sciences, University of Edinburgh / Institute of Genetics and Molecular Medicine, University of Edinburgh | COVID-19 Genomics UK (COG-UK) Consortium |
| EPI_ISL_439253 | Virology Department, Royal Infirmary of Edinburgh, NHS Lothian / School of Biological Sciences, University of Edinburgh / Institute of Genetics and Molecular Medicine, University of Edinburgh | COVID-19 Genomics UK (COG-UK) Consortium |
| EPI_ISL_439254 | Virology Department, Royal Infirmary of Edinburgh, NHS Lothian / School of Biological Sciences, University of Edinburgh / Institute of Genetics and Molecular Medicine, University of Edinburgh | COVID-19 Genomics UK (COG-UK) Consortium |
| EPI_ISL_439255 | Virology Department, Royal Infirmary of Edinburgh, NHS Lothian / School of Biological Sciences, University of Edinburgh / Institute of Genetics and Molecular Medicine, University of Edinburgh | COVID-19 Genomics UK (COG-UK) Consortium |
| EPI_ISL_439256 | Virology Department, Royal Infirmary of Edinburgh, NHS Lothian / School of Biological Sciences, University of Edinburgh / Institute of Genetics and Molecular Medicine, University of Edinburgh | COVID-19 Genomics UK (COG-UK) Consortium |
| EPI_ISL_439257 | Virology Department, Royal Infirmary of Edinburgh, NHS Lothian / School of Biological Sciences, University of Edinburgh / Institute of Genetics and Molecular Medicine, University of Edinburgh | COVID-19 Genomics UK (COG-UK) Consortium |
| EPI_ISL_439258 | Virology Department, Royal Infirmary of Edinburgh, NHS Lothian / School of Biological Sciences, University of Edinburgh / Institute of Genetics and Molecular Medicine, University of Edinburgh | COVID-19 Genomics UK (COG-UK) Consortium |

| EPI_Accession_ID | Originating Laboratory | Submitting Laboratory |
| --- | --- | --- |
| EPI_ISL_439259 | Virology Department, Royal Infirmary of Edinburgh, NHS Lothian / School of Biological Sciences, University of Edinburgh / Institute of Genetics and Molecular Medicine, University of Edinburgh | COVID-19 Genomics UK (COG-UK) Consortium |
| EPI_ISL_439260 | Virology Department, Royal Infirmary of Edinburgh, NHS Lothian / School of Biological Sciences, University of Edinburgh / Institute of Genetics and Molecular Medicine, University of Edinburgh | COVID-19 Genomics UK (COG-UK) Consortium |
| EPI_ISL_439261 | Virology Department, Royal Infirmary of Edinburgh, NHS Lothian / School of Biological Sciences, University of Edinburgh / Institute of Genetics and Molecular Medicine, University of Edinburgh | COVID-19 Genomics UK (COG-UK) Consortium |
| EPI_ISL_439262 | Virology Department, Royal Infirmary of Edinburgh, NHS Lothian / School of Biological Sciences, University of Edinburgh / Institute of Genetics and Molecular Medicine, University of Edinburgh | COVID-19 Genomics UK (COG-UK) Consortium |
| EPI_ISL_439263 | Virology Department, Royal Infirmary of Edinburgh, NHS Lothian / School of Biological Sciences, University of Edinburgh / Institute of Genetics and Molecular Medicine, University of Edinburgh | COVID-19 Genomics UK (COG-UK) Consortium |
| EPI_ISL_439264 | Virology Department, Royal Infirmary of Edinburgh, NHS Lothian / School of Biological Sciences, University of Edinburgh / Institute of Genetics and Molecular Medicine, University of Edinburgh | COVID-19 Genomics UK (COG-UK) Consortium |
| EPI_ISL_439265 | Virology Department, Royal Infirmary of Edinburgh, NHS Lothian / School of Biological Sciences, University of Edinburgh / Institute of Genetics and Molecular Medicine, University of Edinburgh | COVID-19 Genomics UK (COG-UK) Consortium |
| EPI_ISL_439266 | Virology Department, Royal Infirmary of Edinburgh, NHS Lothian / School of Biological Sciences, University of Edinburgh / Institute of Genetics and Molecular Medicine, University of Edinburgh | COVID-19 Genomics UK (COG-UK) Consortium |

| EPI_Accession_ID | Originating Laboratory | Submitting Laboratory |
|---|---|---|
| EPI_ISL_439267 | Virology Department, Royal Infirmary of Edinburgh, NHS Lothian / School of Biological Sciences, University of Edinburgh / Institute of Genetics and Molecular Medicine, University of Edinburgh | COVID-19 Genomics UK (COG-UK) Consortium |
| EPI_ISL_439268 | Virology Department, Royal Infirmary of Edinburgh, NHS Lothian / School of Biological Sciences, University of Edinburgh / Institute of Genetics and Molecular Medicine, University of Edinburgh | COVID-19 Genomics UK (COG-UK) Consortium |
| EPI_ISL_439269 | Virology Department, Royal Infirmary of Edinburgh, NHS Lothian / School of Biological Sciences, University of Edinburgh / Institute of Genetics and Molecular Medicine, University of Edinburgh | COVID-19 Genomics UK (COG-UK) Consortium |
| EPI_ISL_439270 | Virology Department, Royal Infirmary of Edinburgh, NHS Lothian / School of Biological Sciences, University of Edinburgh / Institute of Genetics and Molecular Medicine, University of Edinburgh | COVID-19 Genomics UK (COG-UK) Consortium |
| EPI_ISL_439271 | Virology Department, Royal Infirmary of Edinburgh, NHS Lothian / School of Biological Sciences, University of Edinburgh / Institute of Genetics and Molecular Medicine, University of Edinburgh | COVID-19 Genomics UK (COG-UK) Consortium |
| EPI_ISL_439272 | Virology Department, Royal Infirmary of Edinburgh, NHS Lothian / School of Biological Sciences, University of Edinburgh / Institute of Genetics and Molecular Medicine, University of Edinburgh | COVID-19 Genomics UK (COG-UK) Consortium |
| EPI_ISL_439273 | Virology Department, Royal Infirmary of Edinburgh, NHS Lothian / School of Biological Sciences, University of Edinburgh / Institute of Genetics and Molecular Medicine, University of Edinburgh | COVID-19 Genomics UK (COG-UK) Consortium |
| EPI_ISL_439274 | Virology Department, Royal Infirmary of Edinburgh, NHS Lothian / School of Biological Sciences, University of Edinburgh / Institute of Genetics and Molecular Medicine, University of Edinburgh | COVID-19 Genomics UK (COG-UK) Consortium |

| EPI_Accession_ID | Originating Laboratory | Submitting Laboratory |
|---|---|---|
| EPI_ISL_439275 | Virology Department, Royal Infirmary of Edinburgh, NHS Lothian / School of Biological Sciences, University of Edinburgh / Institute of Genetics and Molecular Medicine, University of Edinburgh | COVID-19 Genomics UK (COG-UK) Consortium |
| EPI_ISL_439276 | Virology Department, Royal Infirmary of Edinburgh, NHS Lothian / School of Biological Sciences, University of Edinburgh / Institute of Genetics and Molecular Medicine, University of Edinburgh | COVID-19 Genomics UK (COG-UK) Consortium |
| EPI_ISL_439277 | Virology Department, Royal Infirmary of Edinburgh, NHS Lothian / School of Biological Sciences, University of Edinburgh / Institute of Genetics and Molecular Medicine, University of Edinburgh | COVID-19 Genomics UK (COG-UK) Consortium |
| EPI_ISL_439278 | Virology Department, Royal Infirmary of Edinburgh, NHS Lothian / School of Biological Sciences, University of Edinburgh / Institute of Genetics and Molecular Medicine, University of Edinburgh | COVID-19 Genomics UK (COG-UK) Consortium |
| EPI_ISL_439279 | Virology Department, Royal Infirmary of Edinburgh, NHS Lothian / School of Biological Sciences, University of Edinburgh / Institute of Genetics and Molecular Medicine, University of Edinburgh | COVID-19 Genomics UK (COG-UK) Consortium |
| EPI_ISL_439280 | Virology Department, Royal Infirmary of Edinburgh, NHS Lothian / School of Biological Sciences, University of Edinburgh / Institute of Genetics and Molecular Medicine, University of Edinburgh | COVID-19 Genomics UK (COG-UK) Consortium |
| EPI_ISL_439281 | Virology Department, Royal Infirmary of Edinburgh, NHS Lothian / School of Biological Sciences, University of Edinburgh / Institute of Genetics and Molecular Medicine, University of Edinburgh | COVID-19 Genomics UK (COG-UK) Consortium |
| EPI_ISL_439282 | Virology Department, Royal Infirmary of Edinburgh, NHS Lothian / School of Biological Sciences, University of Edinburgh / Institute of Genetics and Molecular Medicine, University of Edinburgh | COVID-19 Genomics UK (COG-UK) Consortium |

| EPI_Accession_ID | Originating Laboratory | Submitting Laboratory |
|---|---|---|
| EPI_ISL_439283 | Virology Department, Royal Infirmary of Edinburgh, NHS Lothian / School of Biological Sciences, University of Edinburgh / Institute of Genetics and Molecular Medicine, University of Edinburgh | COVID-19 Genomics UK (COG-UK) Consortium |
| EPI_ISL_439284 | Virology Department, Royal Infirmary of Edinburgh, NHS Lothian / School of Biological Sciences, University of Edinburgh / Institute of Genetics and Molecular Medicine, University of Edinburgh | COVID-19 Genomics UK (COG-UK) Consortium |
| EPI_ISL_439285 | Virology Department, Royal Infirmary of Edinburgh, NHS Lothian / School of Biological Sciences, University of Edinburgh / Institute of Genetics and Molecular Medicine, University of Edinburgh | COVID-19 Genomics UK (COG-UK) Consortium |
| EPI_ISL_439286 | Virology Department, Royal Infirmary of Edinburgh, NHS Lothian / School of Biological Sciences, University of Edinburgh / Institute of Genetics and Molecular Medicine, University of Edinburgh | COVID-19 Genomics UK (COG-UK) Consortium |
| EPI_ISL_439287 | Virology Department, Royal Infirmary of Edinburgh, NHS Lothian / School of Biological Sciences, University of Edinburgh / Institute of Genetics and Molecular Medicine, University of Edinburgh | COVID-19 Genomics UK (COG-UK) Consortium |
| EPI_ISL_439288 | Virology Department, Royal Infirmary of Edinburgh, NHS Lothian / School of Biological Sciences, University of Edinburgh / Institute of Genetics and Molecular Medicine, University of Edinburgh | COVID-19 Genomics UK (COG-UK) Consortium |
| EPI_ISL_439289 | Virology Department, Royal Infirmary of Edinburgh, NHS Lothian / School of Biological Sciences, University of Edinburgh / Institute of Genetics and Molecular Medicine, University of Edinburgh | COVID-19 Genomics UK (COG-UK) Consortium |
| EPI_ISL_439290 | Virology Department, Royal Infirmary of Edinburgh, NHS Lothian / School of Biological Sciences, University of Edinburgh / Institute of Genetics and Molecular Medicine, University of Edinburgh | COVID-19 Genomics UK (COG-UK) Consortium |

| EPI_Accession_ID | Originating Laboratory | Submitting Laboratory |
|---|---|---|
| EPI_ISL_439291 | Virology Department, Royal Infirmary of Edinburgh, NHS Lothian / School of Biological Sciences, University of Edinburgh / Institute of Genetics and Molecular Medicine, University of Edinburgh | COVID-19 Genomics UK (COG-UK) Consortium |
| EPI_ISL_439292 | Virology Department, Royal Infirmary of Edinburgh, NHS Lothian / School of Biological Sciences, University of Edinburgh / Institute of Genetics and Molecular Medicine, University of Edinburgh | COVID-19 Genomics UK (COG-UK) Consortium |
| EPI_ISL_439293 | Virology Department, Royal Infirmary of Edinburgh, NHS Lothian / School of Biological Sciences, University of Edinburgh / Institute of Genetics and Molecular Medicine, University of Edinburgh | COVID-19 Genomics UK (COG-UK) Consortium |
| EPI_ISL_439294 | Virology Department, Royal Infirmary of Edinburgh, NHS Lothian / School of Biological Sciences, University of Edinburgh / Institute of Genetics and Molecular Medicine, University of Edinburgh | COVID-19 Genomics UK (COG-UK) Consortium |
| EPI_ISL_439295 | Virology Department, Royal Infirmary of Edinburgh, NHS Lothian / School of Biological Sciences, University of Edinburgh / Institute of Genetics and Molecular Medicine, University of Edinburgh | COVID-19 Genomics UK (COG-UK) Consortium |
| EPI_ISL_439296 | Virology Department, Royal Infirmary of Edinburgh, NHS Lothian / School of Biological Sciences, University of Edinburgh / Institute of Genetics and Molecular Medicine, University of Edinburgh | COVID-19 Genomics UK (COG-UK) Consortium |
| EPI_ISL_439297 | Virology Department, Royal Infirmary of Edinburgh, NHS Lothian / School of Biological Sciences, University of Edinburgh / Institute of Genetics and Molecular Medicine, University of Edinburgh | COVID-19 Genomics UK (COG-UK) Consortium |
| EPI_ISL_439298 | Virology Department, Royal Infirmary of Edinburgh, NHS Lothian / School of Biological Sciences, University of Edinburgh / Institute of Genetics and Molecular Medicine, University of Edinburgh | COVID-19 Genomics UK (COG-UK) Consortium |

| EPI_Accession_ID | Originating Laboratory | Submitting Laboratory |
|---|---|---|
| EPI_ISL_439299 | Virology Department, Royal Infirmary of Edinburgh, NHS Lothian / School of Biological Sciences, University of Edinburgh / Institute of Genetics and Molecular Medicine, University of Edinburgh | COVID-19 Genomics UK (COG-UK) Consortium |
| EPI_ISL_439300 | Virology Department, Royal Infirmary of Edinburgh, NHS Lothian / School of Biological Sciences, University of Edinburgh / Institute of Genetics and Molecular Medicine, University of Edinburgh | COVID-19 Genomics UK (COG-UK) Consortium |
| EPI_ISL_439301 | Virology Department, Royal Infirmary of Edinburgh, NHS Lothian / School of Biological Sciences, University of Edinburgh / Institute of Genetics and Molecular Medicine, University of Edinburgh | COVID-19 Genomics UK (COG-UK) Consortium |
| EPI_ISL_439302 | Virology Department, Royal Infirmary of Edinburgh, NHS Lothian / School of Biological Sciences, University of Edinburgh / Institute of Genetics and Molecular Medicine, University of Edinburgh | COVID-19 Genomics UK (COG-UK) Consortium |
| EPI_ISL_439303 | Virology Department, Royal Infirmary of Edinburgh, NHS Lothian / School of Biological Sciences, University of Edinburgh / Institute of Genetics and Molecular Medicine, University of Edinburgh | COVID-19 Genomics UK (COG-UK) Consortium |
| EPI_ISL_439304 | Virology Department, Royal Infirmary of Edinburgh, NHS Lothian / School of Biological Sciences, University of Edinburgh / Institute of Genetics and Molecular Medicine, University of Edinburgh | COVID-19 Genomics UK (COG-UK) Consortium |
| EPI_ISL_439305 | Virology Department, Royal Infirmary of Edinburgh, NHS Lothian / School of Biological Sciences, University of Edinburgh / Institute of Genetics and Molecular Medicine, University of Edinburgh | COVID-19 Genomics UK (COG-UK) Consortium |
| EPI_ISL_439306 | Virology Department, Royal Infirmary of Edinburgh, NHS Lothian / School of Biological Sciences, University of Edinburgh / Institute of Genetics and Molecular Medicine, University of Edinburgh | COVID-19 Genomics UK (COG-UK) Consortium |

| EPI_Accession_ID | Originating Laboratory | Submitting Laboratory |
|---|---|---|
| EPI_ISL_439307 | Virology Department, Royal Infirmary of Edinburgh, NHS Lothian / School of Biological Sciences, University of Edinburgh / Institute of Genetics and Molecular Medicine, University of Edinburgh | COVID-19 Genomics UK (COG-UK) Consortium |
| EPI_ISL_439308 | Virology Department, Royal Infirmary of Edinburgh, NHS Lothian / School of Biological Sciences, University of Edinburgh / Institute of Genetics and Molecular Medicine, University of Edinburgh | COVID-19 Genomics UK (COG-UK) Consortium |
| EPI_ISL_439309 | Virology Department, Royal Infirmary of Edinburgh, NHS Lothian / School of Biological Sciences, University of Edinburgh / Institute of Genetics and Molecular Medicine, University of Edinburgh | COVID-19 Genomics UK (COG-UK) Consortium |
| EPI_ISL_439310 | Virology Department, Royal Infirmary of Edinburgh, NHS Lothian / School of Biological Sciences, University of Edinburgh / Institute of Genetics and Molecular Medicine, University of Edinburgh | COVID-19 Genomics UK (COG-UK) Consortium |
| EPI_ISL_439311 | Virology Department, Royal Infirmary of Edinburgh, NHS Lothian / School of Biological Sciences, University of Edinburgh / Institute of Genetics and Molecular Medicine, University of Edinburgh | COVID-19 Genomics UK (COG-UK) Consortium |
| EPI_ISL_439312 | Virology Department, Royal Infirmary of Edinburgh, NHS Lothian / School of Biological Sciences, University of Edinburgh / Institute of Genetics and Molecular Medicine, University of Edinburgh | COVID-19 Genomics UK (COG-UK) Consortium |
| EPI_ISL_439313 | Virology Department, Royal Infirmary of Edinburgh, NHS Lothian / School of Biological Sciences, University of Edinburgh / Institute of Genetics and Molecular Medicine, University of Edinburgh | COVID-19 Genomics UK (COG-UK) Consortium |
| EPI_ISL_439314 | Virology Department, Royal Infirmary of Edinburgh, NHS Lothian / School of Biological Sciences, University of Edinburgh / Institute of Genetics and Molecular Medicine, University of Edinburgh | COVID-19 Genomics UK (COG-UK) Consortium |

| EPI_Accession_ID | Originating Laboratory | Submitting Laboratory |
|---|---|---|
| EPI_ISL_439315 | Virology Department, Royal Infirmary of Edinburgh, NHS Lothian / School of Biological Sciences, University of Edinburgh / Institute of Genetics and Molecular Medicine, University of Edinburgh | COVID-19 Genomics UK (COG-UK) Consortium |
| EPI_ISL_439316 | Virology Department, Royal Infirmary of Edinburgh, NHS Lothian / School of Biological Sciences, University of Edinburgh / Institute of Genetics and Molecular Medicine, University of Edinburgh | COVID-19 Genomics UK (COG-UK) Consortium |
| EPI_ISL_439317 | Virology Department, Royal Infirmary of Edinburgh, NHS Lothian / School of Biological Sciences, University of Edinburgh / Institute of Genetics and Molecular Medicine, University of Edinburgh | COVID-19 Genomics UK (COG-UK) Consortium |
| EPI_ISL_439318 | Virology Department, Royal Infirmary of Edinburgh, NHS Lothian / School of Biological Sciences, University of Edinburgh / Institute of Genetics and Molecular Medicine, University of Edinburgh | COVID-19 Genomics UK (COG-UK) Consortium |
| EPI_ISL_439319 | Virology Department, Royal Infirmary of Edinburgh, NHS Lothian / School of Biological Sciences, University of Edinburgh / Institute of Genetics and Molecular Medicine, University of Edinburgh | COVID-19 Genomics UK (COG-UK) Consortium |
| EPI_ISL_439320 | Virology Department, Royal Infirmary of Edinburgh, NHS Lothian / School of Biological Sciences, University of Edinburgh / Institute of Genetics and Molecular Medicine, University of Edinburgh | COVID-19 Genomics UK (COG-UK) Consortium |
| EPI_ISL_439321 | Virology Department, Royal Infirmary of Edinburgh, NHS Lothian / School of Biological Sciences, University of Edinburgh / Institute of Genetics and Molecular Medicine, University of Edinburgh | COVID-19 Genomics UK (COG-UK) Consortium |
| EPI_ISL_439322 | Virology Department, Royal Infirmary of Edinburgh, NHS Lothian / School of Biological Sciences, University of Edinburgh / Institute of Genetics and Molecular Medicine, University of Edinburgh | COVID-19 Genomics UK (COG-UK) Consortium |

| EPI_Accession_ID | Originating Laboratory | Submitting Laboratory |
|---|---|---|
| EPI_ISL_439323 | Virology Department, Royal Infirmary of Edinburgh, NHS Lothian / School of Biological Sciences, University of Edinburgh / Institute of Genetics and Molecular Medicine, University of Edinburgh | COVID-19 Genomics UK (COG-UK) Consortium |
| EPI_ISL_439324 | Virology Department, Royal Infirmary of Edinburgh, NHS Lothian / School of Biological Sciences, University of Edinburgh / Institute of Genetics and Molecular Medicine, University of Edinburgh | COVID-19 Genomics UK (COG-UK) Consortium |
| EPI_ISL_439325 | Virology Department, Royal Infirmary of Edinburgh, NHS Lothian / School of Biological Sciences, University of Edinburgh / Institute of Genetics and Molecular Medicine, University of Edinburgh | COVID-19 Genomics UK (COG-UK) Consortium |
| EPI_ISL_439326 | Virology Department, Royal Infirmary of Edinburgh, NHS Lothian / School of Biological Sciences, University of Edinburgh / Institute of Genetics and Molecular Medicine, University of Edinburgh | COVID-19 Genomics UK (COG-UK) Consortium |
| EPI_ISL_439327 | Virology Department, Royal Infirmary of Edinburgh, NHS Lothian / School of Biological Sciences, University of Edinburgh / Institute of Genetics and Molecular Medicine, University of Edinburgh | COVID-19 Genomics UK (COG-UK) Consortium |
| EPI_ISL_439328 | Virology Department, Royal Infirmary of Edinburgh, NHS Lothian / School of Biological Sciences, University of Edinburgh / Institute of Genetics and Molecular Medicine, University of Edinburgh | COVID-19 Genomics UK (COG-UK) Consortium |
| EPI_ISL_439329 | Virology Department, Royal Infirmary of Edinburgh, NHS Lothian / School of Biological Sciences, University of Edinburgh / Institute of Genetics and Molecular Medicine, University of Edinburgh | COVID-19 Genomics UK (COG-UK) Consortium |
| EPI_ISL_439330 | Virology Department, Royal Infirmary of Edinburgh, NHS Lothian / School of Biological Sciences, University of Edinburgh / Institute of Genetics and Molecular Medicine, University of Edinburgh | COVID-19 Genomics UK (COG-UK) Consortium |

| EPI_Accession_ID | Originating Laboratory | Submitting Laboratory |
| --- | --- | --- |
| EPI_ISL_439331 | Virology Department, Royal Infirmary of Edinburgh, NHS Lothian / School of Biological Sciences, University of Edinburgh / Institute of Genetics and Molecular Medicine, University of Edinburgh | COVID-19 Genomics UK (COG-UK) Consortium |
| EPI_ISL_439332 | Virology Department, Royal Infirmary of Edinburgh, NHS Lothian / School of Biological Sciences, University of Edinburgh / Institute of Genetics and Molecular Medicine, University of Edinburgh | COVID-19 Genomics UK (COG-UK) Consortium |
| EPI_ISL_439333 | Virology Department, Royal Infirmary of Edinburgh, NHS Lothian / School of Biological Sciences, University of Edinburgh / Institute of Genetics and Molecular Medicine, University of Edinburgh | COVID-19 Genomics UK (COG-UK) Consortium |
| EPI_ISL_439334 | Virology Department, Royal Infirmary of Edinburgh, NHS Lothian / School of Biological Sciences, University of Edinburgh / Institute of Genetics and Molecular Medicine, University of Edinburgh | COVID-19 Genomics UK (COG-UK) Consortium |
| EPI_ISL_439335 | Virology Department, Royal Infirmary of Edinburgh, NHS Lothian / School of Biological Sciences, University of Edinburgh / Institute of Genetics and Molecular Medicine, University of Edinburgh | COVID-19 Genomics UK (COG-UK) Consortium |
| EPI_ISL_439336 | Virology Department, Royal Infirmary of Edinburgh, NHS Lothian / School of Biological Sciences, University of Edinburgh / Institute of Genetics and Molecular Medicine, University of Edinburgh | COVID-19 Genomics UK (COG-UK) Consortium |
| EPI_ISL_439337 | Virology Department, Royal Infirmary of Edinburgh, NHS Lothian / School of Biological Sciences, University of Edinburgh / Institute of Genetics and Molecular Medicine, University of Edinburgh | COVID-19 Genomics UK (COG-UK) Consortium |
| EPI_ISL_439338 | Virology Department, Royal Infirmary of Edinburgh, NHS Lothian / School of Biological Sciences, University of Edinburgh / Institute of Genetics and Molecular Medicine, University of Edinburgh | COVID-19 Genomics UK (COG-UK) Consortium |

| EPI_Accession_ID | Originating Laboratory | Submitting Laboratory |
|---|---|---|
| EPI_ISL_439339 | Virology Department, Royal Infirmary of Edinburgh, NHS Lothian / School of Biological Sciences, University of Edinburgh / Institute of Genetics and Molecular Medicine, University of Edinburgh | COVID-19 Genomics UK (COG-UK) Consortium |
| EPI_ISL_439340 | Virology Department, Royal Infirmary of Edinburgh, NHS Lothian / School of Biological Sciences, University of Edinburgh / Institute of Genetics and Molecular Medicine, University of Edinburgh | COVID-19 Genomics UK (COG-UK) Consortium |
| EPI_ISL_439341 | Virology Department, Royal Infirmary of Edinburgh, NHS Lothian / School of Biological Sciences, University of Edinburgh / Institute of Genetics and Molecular Medicine, University of Edinburgh | COVID-19 Genomics UK (COG-UK) Consortium |
| EPI_ISL_439342 | Virology Department, Royal Infirmary of Edinburgh, NHS Lothian / School of Biological Sciences, University of Edinburgh / Institute of Genetics and Molecular Medicine, University of Edinburgh | COVID-19 Genomics UK (COG-UK) Consortium |
| EPI_ISL_439343 | Virology Department, Royal Infirmary of Edinburgh, NHS Lothian / School of Biological Sciences, University of Edinburgh / Institute of Genetics and Molecular Medicine, University of Edinburgh | COVID-19 Genomics UK (COG-UK) Consortium |
| EPI_ISL_439344 | Virology Department, Royal Infirmary of Edinburgh, NHS Lothian / School of Biological Sciences, University of Edinburgh / Institute of Genetics and Molecular Medicine, University of Edinburgh | COVID-19 Genomics UK (COG-UK) Consortium |
| EPI_ISL_439345 | Virology Department, Royal Infirmary of Edinburgh, NHS Lothian / School of Biological Sciences, University of Edinburgh / Institute of Genetics and Molecular Medicine, University of Edinburgh | COVID-19 Genomics UK (COG-UK) Consortium |
| EPI_ISL_439346 | Virology Department, Royal Infirmary of Edinburgh, NHS Lothian / School of Biological Sciences, University of Edinburgh / Institute of Genetics and Molecular Medicine, University of Edinburgh | COVID-19 Genomics UK (COG-UK) Consortium |

| EPI_Accession_ID | Originating Laboratory | Submitting Laboratory |
|---|---|---|
| EPI_ISL_439347 | Virology Department, Royal Infirmary of Edinburgh, NHS Lothian / School of Biological Sciences, University of Edinburgh / Institute of Genetics and Molecular Medicine, University of Edinburgh | COVID-19 Genomics UK (COG-UK) Consortium |
| EPI_ISL_439348 | Virology Department, Royal Infirmary of Edinburgh, NHS Lothian / School of Biological Sciences, University of Edinburgh / Institute of Genetics and Molecular Medicine, University of Edinburgh | COVID-19 Genomics UK (COG-UK) Consortium |
| EPI_ISL_439349 | Virology Department, Royal Infirmary of Edinburgh, NHS Lothian / School of Biological Sciences, University of Edinburgh / Institute of Genetics and Molecular Medicine, University of Edinburgh | COVID-19 Genomics UK (COG-UK) Consortium |
| EPI_ISL_439350 | Virology Department, Royal Infirmary of Edinburgh, NHS Lothian / School of Biological Sciences, University of Edinburgh / Institute of Genetics and Molecular Medicine, University of Edinburgh | COVID-19 Genomics UK (COG-UK) Consortium |
| EPI_ISL_439351 | Virology Department, Royal Infirmary of Edinburgh, NHS Lothian / School of Biological Sciences, University of Edinburgh / Institute of Genetics and Molecular Medicine, University of Edinburgh | COVID-19 Genomics UK (COG-UK) Consortium |
| EPI_ISL_439352 | Virology Department, Royal Infirmary of Edinburgh, NHS Lothian / School of Biological Sciences, University of Edinburgh / Institute of Genetics and Molecular Medicine, University of Edinburgh | COVID-19 Genomics UK (COG-UK) Consortium |
| EPI_ISL_439353 | Virology Department, Royal Infirmary of Edinburgh, NHS Lothian / School of Biological Sciences, University of Edinburgh / Institute of Genetics and Molecular Medicine, University of Edinburgh | COVID-19 Genomics UK (COG-UK) Consortium |
| EPI_ISL_439354 | Virology Department, Royal Infirmary of Edinburgh, NHS Lothian / School of Biological Sciences, University of Edinburgh / Institute of Genetics and Molecular Medicine, University of Edinburgh | COVID-19 Genomics UK (COG-UK) Consortium |

| EPI_Accession_ID | Originating Laboratory | Submitting Laboratory |
|---|---|---|
| EPI_ISL_439355 | Virology Department, Royal Infirmary of Edinburgh, NHS Lothian / School of Biological Sciences, University of Edinburgh / Institute of Genetics and Molecular Medicine, University of Edinburgh | COVID-19 Genomics UK (COG-UK) Consortium |
| EPI_ISL_439356 | Virology Department, Royal Infirmary of Edinburgh, NHS Lothian / School of Biological Sciences, University of Edinburgh / Institute of Genetics and Molecular Medicine, University of Edinburgh | COVID-19 Genomics UK (COG-UK) Consortium |
| EPI_ISL_439357 | Virology Department, Royal Infirmary of Edinburgh, NHS Lothian / School of Biological Sciences, University of Edinburgh / Institute of Genetics and Molecular Medicine, University of Edinburgh | COVID-19 Genomics UK (COG-UK) Consortium |
| EPI_ISL_439358 | Virology Department, Royal Infirmary of Edinburgh, NHS Lothian / School of Biological Sciences, University of Edinburgh / Institute of Genetics and Molecular Medicine, University of Edinburgh | COVID-19 Genomics UK (COG-UK) Consortium |
| EPI_ISL_439359 | Virology Department, Royal Infirmary of Edinburgh, NHS Lothian / School of Biological Sciences, University of Edinburgh / Institute of Genetics and Molecular Medicine, University of Edinburgh | COVID-19 Genomics UK (COG-UK) Consortium |
| EPI_ISL_439360 | Virology Department, Royal Infirmary of Edinburgh, NHS Lothian / School of Biological Sciences, University of Edinburgh / Institute of Genetics and Molecular Medicine, University of Edinburgh | COVID-19 Genomics UK (COG-UK) Consortium |
| EPI_ISL_439361 | Virology Department, Royal Infirmary of Edinburgh, NHS Lothian / School of Biological Sciences, University of Edinburgh / Institute of Genetics and Molecular Medicine, University of Edinburgh | COVID-19 Genomics UK (COG-UK) Consortium |
| EPI_ISL_439362 | Virology Department, Royal Infirmary of Edinburgh, NHS Lothian / School of Biological Sciences, University of Edinburgh / Institute of Genetics and Molecular Medicine, University of Edinburgh | COVID-19 Genomics UK (COG-UK) Consortium |

| EPI_Accession_ID | Originating Laboratory | Submitting Laboratory |
|---|---|---|
| EPI_ISL_439363 | Virology Department, Royal Infirmary of Edinburgh, NHS Lothian / School of Biological Sciences, University of Edinburgh / Institute of Genetics and Molecular Medicine, University of Edinburgh | COVID-19 Genomics UK (COG-UK) Consortium |
| EPI_ISL_439364 | Virology Department, Royal Infirmary of Edinburgh, NHS Lothian / School of Biological Sciences, University of Edinburgh / Institute of Genetics and Molecular Medicine, University of Edinburgh | COVID-19 Genomics UK (COG-UK) Consortium |
| EPI_ISL_439365 | Virology Department, Royal Infirmary of Edinburgh, NHS Lothian / School of Biological Sciences, University of Edinburgh / Institute of Genetics and Molecular Medicine, University of Edinburgh | COVID-19 Genomics UK (COG-UK) Consortium |
| EPI_ISL_439366 | Virology Department, Royal Infirmary of Edinburgh, NHS Lothian / School of Biological Sciences, University of Edinburgh / Institute of Genetics and Molecular Medicine, University of Edinburgh | COVID-19 Genomics UK (COG-UK) Consortium |
| EPI_ISL_439367 | Virology Department, Royal Infirmary of Edinburgh, NHS Lothian / School of Biological Sciences, University of Edinburgh / Institute of Genetics and Molecular Medicine, University of Edinburgh | COVID-19 Genomics UK (COG-UK) Consortium |
| EPI_ISL_439368 | Department of Pathology, University of Cambridge | Wellcome Sanger Institute for the COVID-19 Genomics UK Consortium |
| EPI_ISL_439369 | Department of Pathology, University of Cambridge | Wellcome Sanger Institute for the COVID-19 Genomics UK Consortium |
| EPI_ISL_439370 | Department of Pathology, University of Cambridge | Wellcome Sanger Institute for the COVID-19 Genomics UK Consortium |
| EPI_ISL_439371 | Department of Pathology, University of Cambridge | Wellcome Sanger Institute for the COVID-19 Genomics UK Consortium |
| EPI_ISL_439372 | Department of Pathology, University of Cambridge | Wellcome Sanger Institute for the COVID-19 Genomics UK Consortium |
| EPI_ISL_439373 | Department of Pathology, University of Cambridge | Wellcome Sanger Institute for the COVID-19 Genomics UK Consortium |

| EPI_Accession_ID | Originating Laboratory | Submitting Laboratory |
|---|---|---|
| EPI_ISL_439374 | Department of Pathology, University of Cambridge | Wellcome Sanger Institute for the COVID-19 Genomics UK Consortium |
| EPI_ISL_439375 | Department of Pathology, University of Cambridge | Wellcome Sanger Institute for the COVID-19 Genomics UK Consortium |
| EPI_ISL_439376 | Department of Pathology, University of Cambridge | Wellcome Sanger Institute for the COVID-19 Genomics UK Consortium |
| EPI_ISL_439377 | Department of Pathology, University of Cambridge | Wellcome Sanger Institute for the COVID-19 Genomics UK Consortium |
| EPI_ISL_439378 | Department of Pathology, University of Cambridge | Wellcome Sanger Institute for the COVID-19 Genomics UK Consortium |
| EPI_ISL_439379 | Department of Pathology, University of Cambridge | Wellcome Sanger Institute for the COVID-19 Genomics UK Consortium |
| EPI_ISL_439380 | Department of Pathology, University of Cambridge | Wellcome Sanger Institute for the COVID-19 Genomics UK Consortium |
| EPI_ISL_439381 | Department of Pathology, University of Cambridge | Wellcome Sanger Institute for the COVID-19 Genomics UK Consortium |
| EPI_ISL_439382 | Department of Pathology, University of Cambridge | Wellcome Sanger Institute for the COVID-19 Genomics UK Consortium |
| EPI_ISL_439383 | Department of Pathology, University of Cambridge | Wellcome Sanger Institute for the COVID-19 Genomics UK Consortium |
| EPI_ISL_439384 | Department of Pathology, University of Cambridge | Wellcome Sanger Institute for the COVID-19 Genomics UK Consortium |
| EPI_ISL_439385 | Department of Pathology, University of Cambridge | Wellcome Sanger Institute for the COVID-19 Genomics UK Consortium |
| EPI_ISL_439386 | Department of Pathology, University of Cambridge | Wellcome Sanger Institute for the COVID-19 Genomics UK Consortium |
| EPI_ISL_439387 | Department of Pathology, University of Cambridge | Wellcome Sanger Institute for the COVID-19 Genomics UK Consortium |
| EPI_ISL_439388 | Department of Pathology, University of Cambridge | Wellcome Sanger Institute for the COVID-19 Genomics UK Consortium |
| EPI_ISL_439389 | Department of Pathology, University of Cambridge | Wellcome Sanger Institute for the COVID-19 Genomics UK Consortium |

| EPI_Accession_ID | Originating Laboratory | Submitting Laboratory |
|---|---|---|
| EPI_ISL_439390 | Department of Pathology, University of Cambridge | Wellcome Sanger Institute for the COVID-19 Genomics UK Consortium |
| EPI_ISL_439391 | Department of Pathology, University of Cambridge | Wellcome Sanger Institute for the COVID-19 Genomics UK Consortium |
| EPI_ISL_439392 | Department of Pathology, University of Cambridge | Wellcome Sanger Institute for the COVID-19 Genomics UK Consortium |
| EPI_ISL_439393 | Department of Pathology, University of Cambridge | Wellcome Sanger Institute for the COVID-19 Genomics UK Consortium |
| EPI_ISL_439394 | Department of Pathology, University of Cambridge | Wellcome Sanger Institute for the COVID-19 Genomics UK Consortium |
| EPI_ISL_439395 | Department of Pathology, University of Cambridge | Wellcome Sanger Institute for the COVID-19 Genomics UK Consortium |
| EPI_ISL_439396 | Department of Pathology, University of Cambridge | Wellcome Sanger Institute for the COVID-19 Genomics UK Consortium |
| EPI_ISL_439397 | Department of Pathology, University of Cambridge | Wellcome Sanger Institute for the COVID-19 Genomics UK Consortium |
| EPI_ISL_439398 | Department of Pathology, University of Cambridge | Wellcome Sanger Institute for the COVID-19 Genomics UK Consortium |
| EPI_ISL_439399 | Department of Pathology, University of Cambridge | Wellcome Sanger Institute for the COVID-19 Genomics UK Consortium |
| EPI_ISL_439400 | Department of Pathology, University of Cambridge | Wellcome Sanger Institute for the COVID-19 Genomics UK Consortium |
| EPI_ISL_439401 | Department of Pathology, University of Cambridge | Wellcome Sanger Institute for the COVID-19 Genomics UK Consortium |
| EPI_ISL_439402 | Department of Pathology, University of Cambridge | Wellcome Sanger Institute for the COVID-19 Genomics UK Consortium |
| EPI_ISL_439403 | Department of Pathology, University of Cambridge | Wellcome Sanger Institute for the COVID-19 Genomics UK Consortium |
| EPI_ISL_439404 | Department of Pathology, University of Cambridge | Wellcome Sanger Institute for the COVID-19 Genomics UK Consortium |
| EPI_ISL_439405 | Department of Pathology, University of Cambridge | Wellcome Sanger Institute for the COVID-19 Genomics UK Consortium |

| EPI_Accession_ID | Originating Laboratory | Submitting Laboratory |
|---|---|---|
| EPI_ISL_439406 | Department of Pathology, University of Cambridge | Wellcome Sanger Institute for the COVID-19 Genomics UK Consortium |
| EPI_ISL_439407 | Department of Pathology, University of Cambridge | Wellcome Sanger Institute for the COVID-19 Genomics UK Consortium |
| EPI_ISL_439408 | Department of Pathology, University of Cambridge | Wellcome Sanger Institute for the COVID-19 Genomics UK Consortium |
| EPI_ISL_439409 | Department of Pathology, University of Cambridge | Wellcome Sanger Institute for the COVID-19 Genomics UK Consortium |
| EPI_ISL_439410 | Department of Pathology, University of Cambridge | Wellcome Sanger Institute for the COVID-19 Genomics UK Consortium |
| EPI_ISL_439411 | Department of Pathology, University of Cambridge | Wellcome Sanger Institute for the COVID-19 Genomics UK Consortium |
| EPI_ISL_439412 | Department of Pathology, University of Cambridge | Wellcome Sanger Institute for the COVID-19 Genomics UK Consortium |
| EPI_ISL_439413 | Department of Pathology, University of Cambridge | Wellcome Sanger Institute for the COVID-19 Genomics UK Consortium |
| EPI_ISL_439414 | Department of Pathology, University of Cambridge | Wellcome Sanger Institute for the COVID-19 Genomics UK Consortium |
| EPI_ISL_439415 | Department of Pathology, University of Cambridge | Wellcome Sanger Institute for the COVID-19 Genomics UK Consortium |
| EPI_ISL_439416 | Department of Pathology, University of Cambridge | Wellcome Sanger Institute for the COVID-19 Genomics UK Consortium |
| EPI_ISL_439417 | Department of Pathology, University of Cambridge | Wellcome Sanger Institute for the COVID-19 Genomics UK Consortium |
| EPI_ISL_439418 | Department of Pathology, University of Cambridge | Wellcome Sanger Institute for the COVID-19 Genomics UK Consortium |
| EPI_ISL_439419 | Department of Pathology, University of Cambridge | Wellcome Sanger Institute for the COVID-19 Genomics UK Consortium |
| EPI_ISL_439420 | Department of Pathology, University of Cambridge | Wellcome Sanger Institute for the COVID-19 Genomics UK Consortium |
| EPI_ISL_439421 | Department of Pathology, University of Cambridge | Wellcome Sanger Institute for the COVID-19 Genomics UK Consortium |

| EPI_Accession_ID | Originating Laboratory | Submitting Laboratory |
| --- | --- | --- |
| EPI_ISL_439422 | Department of Pathology, University of Cambridge | Wellcome Sanger Institute for the COVID-19 Genomics UK Consortium |
| EPI_ISL_439423 | Department of Pathology, University of Cambridge | Wellcome Sanger Institute for the COVID-19 Genomics UK Consortium |
| EPI_ISL_439424 | Department of Pathology, University of Cambridge | Wellcome Sanger Institute for the COVID-19 Genomics UK Consortium |
| EPI_ISL_439425 | Department of Pathology, University of Cambridge | Wellcome Sanger Institute for the COVID-19 Genomics UK Consortium |
| EPI_ISL_439426 | Department of Pathology, University of Cambridge | Wellcome Sanger Institute for the COVID-19 Genomics UK Consortium |
| EPI_ISL_439427 | Department of Pathology, University of Cambridge | Wellcome Sanger Institute for the COVID-19 Genomics UK Consortium |
| EPI_ISL_439428 | Department of Pathology, University of Cambridge | Wellcome Sanger Institute for the COVID-19 Genomics UK Consortium |
| EPI_ISL_439429 | Department of Pathology, University of Cambridge | Wellcome Sanger Institute for the COVID-19 Genomics UK Consortium |
| EPI_ISL_439430 | Department of Pathology, University of Cambridge | Wellcome Sanger Institute for the COVID-19 Genomics UK Consortium |
| EPI_ISL_439431 | Department of Pathology, University of Cambridge | Wellcome Sanger Institute for the COVID-19 Genomics UK Consortium |
| EPI_ISL_439432 | Department of Pathology, University of Cambridge | Wellcome Sanger Institute for the COVID-19 Genomics UK Consortium |
| EPI_ISL_439433 | Department of Pathology, University of Cambridge | Wellcome Sanger Institute for the COVID-19 Genomics UK Consortium |
| EPI_ISL_439434 | Department of Pathology, University of Cambridge | Wellcome Sanger Institute for the COVID-19 Genomics UK Consortium |
| EPI_ISL_439435 | Department of Pathology, University of Cambridge | Wellcome Sanger Institute for the COVID-19 Genomics UK Consortium |
| EPI_ISL_439436 | Department of Pathology, University of Cambridge | Wellcome Sanger Institute for the COVID-19 Genomics UK Consortium |
| EPI_ISL_439437 | Department of Pathology, University of Cambridge | Wellcome Sanger Institute for the COVID-19 Genomics UK Consortium |

| EPI_Accession_ID | Originating Laboratory | Submitting Laboratory |
|---|---|---|
| EPI_ISL_439438 | Department of Pathology, University of Cambridge | Wellcome Sanger Institute for the COVID-19 Genomics UK Consortium |
| EPI_ISL_439439 | Department of Pathology, University of Cambridge | Wellcome Sanger Institute for the COVID-19 Genomics UK Consortium |
| EPI_ISL_439440 | Department of Pathology, University of Cambridge | Wellcome Sanger Institute for the COVID-19 Genomics UK Consortium |
| EPI_ISL_439441 | Department of Pathology, University of Cambridge | Wellcome Sanger Institute for the COVID-19 Genomics UK Consortium |
| EPI_ISL_439442 | Department of Pathology, University of Cambridge | Wellcome Sanger Institute for the COVID-19 Genomics UK Consortium |
| EPI_ISL_439443 | Department of Pathology, University of Cambridge | Wellcome Sanger Institute for the COVID-19 Genomics UK Consortium |
| EPI_ISL_439444 | Department of Pathology, University of Cambridge | Wellcome Sanger Institute for the COVID-19 Genomics UK Consortium |
| EPI_ISL_439445 | Department of Pathology, University of Cambridge | Wellcome Sanger Institute for the COVID-19 Genomics UK Consortium |
| EPI_ISL_439446 | Department of Pathology, University of Cambridge | Wellcome Sanger Institute for the COVID-19 Genomics UK Consortium |
| EPI_ISL_439447 | Department of Pathology, University of Cambridge | Wellcome Sanger Institute for the COVID-19 Genomics UK Consortium |
| EPI_ISL_439448 | Department of Pathology, University of Cambridge | Wellcome Sanger Institute for the COVID-19 Genomics UK Consortium |
| EPI_ISL_439449 | Department of Pathology, University of Cambridge | Wellcome Sanger Institute for the COVID-19 Genomics UK Consortium |
| EPI_ISL_439450 | Department of Pathology, University of Cambridge | Wellcome Sanger Institute for the COVID-19 Genomics UK Consortium |
| EPI_ISL_439451 | Department of Pathology, University of Cambridge | Wellcome Sanger Institute for the COVID-19 Genomics UK Consortium |
| EPI_ISL_439452 | Department of Pathology, University of Cambridge | Wellcome Sanger Institute for the COVID-19 Genomics UK Consortium |
| EPI_ISL_439453 | Department of Pathology, University of Cambridge | Wellcome Sanger Institute for the COVID-19 Genomics UK Consortium |

| EPI_Accession_ID | Originating Laboratory | Submitting Laboratory |
|---|---|---|
| EPI_ISL_439454 | Department of Pathology, University of Cambridge | Wellcome Sanger Institute for the COVID-19 Genomics UK Consortium |
| EPI_ISL_439455 | Department of Pathology, University of Cambridge | Wellcome Sanger Institute for the COVID-19 Genomics UK Consortium |
| EPI_ISL_439456 | Department of Pathology, University of Cambridge | Wellcome Sanger Institute for the COVID-19 Genomics UK Consortium |
| EPI_ISL_439457 | Department of Pathology, University of Cambridge | Wellcome Sanger Institute for the COVID-19 Genomics UK Consortium |
| EPI_ISL_439458 | Department of Pathology, University of Cambridge | Wellcome Sanger Institute for the COVID-19 Genomics UK Consortium |
| EPI_ISL_439459 | Department of Pathology, University of Cambridge | Wellcome Sanger Institute for the COVID-19 Genomics UK Consortium |
| EPI_ISL_439460 | Department of Pathology, University of Cambridge | Wellcome Sanger Institute for the COVID-19 Genomics UK Consortium |
| EPI_ISL_439461 | Department of Pathology, University of Cambridge | Wellcome Sanger Institute for the COVID-19 Genomics UK Consortium |
| EPI_ISL_439462 | Department of Pathology, University of Cambridge | Wellcome Sanger Institute for the COVID-19 Genomics UK Consortium |
| EPI_ISL_439463 | Department of Pathology, University of Cambridge | Wellcome Sanger Institute for the COVID-19 Genomics UK Consortium |
| EPI_ISL_439464 | Department of Pathology, University of Cambridge | Wellcome Sanger Institute for the COVID-19 Genomics UK Consortium |
| EPI_ISL_439465 | Department of Pathology, University of Cambridge | Wellcome Sanger Institute for the COVID-19 Genomics UK Consortium |
| EPI_ISL_439466 | Department of Pathology, University of Cambridge | Wellcome Sanger Institute for the COVID-19 Genomics UK Consortium |
| EPI_ISL_439467 | Department of Pathology, University of Cambridge | Wellcome Sanger Institute for the COVID-19 Genomics UK Consortium |
| EPI_ISL_439468 | Department of Pathology, University of Cambridge | Wellcome Sanger Institute for the COVID-19 Genomics UK Consortium |
| EPI_ISL_439469 | Department of Pathology, University of Cambridge | Wellcome Sanger Institute for the COVID-19 Genomics UK Consortium |

| EPI_Accession_ID | Originating Laboratory | Submitting Laboratory |
|---|---|---|
| EPI_ISL_439470 | Department of Pathology, University of Cambridge | Wellcome Sanger Institute for the COVID-19 Genomics UK Consortium |
| EPI_ISL_439471 | Department of Pathology, University of Cambridge | Wellcome Sanger Institute for the COVID-19 Genomics UK Consortium |
| EPI_ISL_439472 | Department of Pathology, University of Cambridge | Wellcome Sanger Institute for the COVID-19 Genomics UK Consortium |
| EPI_ISL_439473 | Department of Pathology, University of Cambridge | Wellcome Sanger Institute for the COVID-19 Genomics UK Consortium |
| EPI_ISL_439474 | Department of Pathology, University of Cambridge | Wellcome Sanger Institute for the COVID-19 Genomics UK Consortium |
| EPI_ISL_439475 | Department of Pathology, University of Cambridge | Wellcome Sanger Institute for the COVID-19 Genomics UK Consortium |
| EPI_ISL_439476 | Department of Pathology, University of Cambridge | Wellcome Sanger Institute for the COVID-19 Genomics UK Consortium |
| EPI_ISL_439477 | Department of Pathology, University of Cambridge | Wellcome Sanger Institute for the COVID-19 Genomics UK Consortium |
| EPI_ISL_439478 | Department of Pathology, University of Cambridge | Wellcome Sanger Institute for the COVID-19 Genomics UK Consortium |
| EPI_ISL_439479 | Department of Pathology, University of Cambridge | Wellcome Sanger Institute for the COVID-19 Genomics UK Consortium |
| EPI_ISL_439480 | Department of Pathology, University of Cambridge | Wellcome Sanger Institute for the COVID-19 Genomics UK Consortium |
| EPI_ISL_439481 | Department of Pathology, University of Cambridge | Wellcome Sanger Institute for the COVID-19 Genomics UK Consortium |
| EPI_ISL_439482 | Department of Pathology, University of Cambridge | Wellcome Sanger Institute for the COVID-19 Genomics UK Consortium |
| EPI_ISL_439483 | Department of Pathology, University of Cambridge | Wellcome Sanger Institute for the COVID-19 Genomics UK Consortium |
| EPI_ISL_439484 | Department of Pathology, University of Cambridge | Wellcome Sanger Institute for the COVID-19 Genomics UK Consortium |
| EPI_ISL_439485 | Department of Pathology, University of Cambridge | Wellcome Sanger Institute for the COVID-19 Genomics UK Consortium |

| EPI_Accession_ID | Originating Laboratory | Submitting Laboratory |
| --- | --- | --- |
| EPI_ISL_439486 | Department of Pathology, University of Cambridge | Wellcome Sanger Institute for the COVID-19 Genomics UK Consortium |
| EPI_ISL_439487 | Department of Pathology, University of Cambridge | Wellcome Sanger Institute for the COVID-19 Genomics UK Consortium |
| EPI_ISL_439488 | Department of Pathology, University of Cambridge | Wellcome Sanger Institute for the COVID-19 Genomics UK Consortium |
| EPI_ISL_439489 | Department of Pathology, University of Cambridge | Wellcome Sanger Institute for the COVID-19 Genomics UK Consortium |
| EPI_ISL_439490 | Department of Pathology, University of Cambridge | Wellcome Sanger Institute for the COVID-19 Genomics UK Consortium |
| EPI_ISL_439491 | Department of Pathology, University of Cambridge | Wellcome Sanger Institute for the COVID-19 Genomics UK Consortium |
| EPI_ISL_439492 | Department of Pathology, University of Cambridge | Wellcome Sanger Institute for the COVID-19 Genomics UK Consortium |
| EPI_ISL_439493 | Department of Pathology, University of Cambridge | Wellcome Sanger Institute for the COVID-19 Genomics UK Consortium |
| EPI_ISL_439494 | Department of Pathology, University of Cambridge | Wellcome Sanger Institute for the COVID-19 Genomics UK Consortium |
| EPI_ISL_439495 | Department of Pathology, University of Cambridge | Wellcome Sanger Institute for the COVID-19 Genomics UK Consortium |
| EPI_ISL_439496 | Department of Pathology, University of Cambridge | Wellcome Sanger Institute for the COVID-19 Genomics UK Consortium |
| EPI_ISL_439497 | Department of Pathology, University of Cambridge | Wellcome Sanger Institute for the COVID-19 Genomics UK Consortium |
| EPI_ISL_439498 | Department of Pathology, University of Cambridge | Wellcome Sanger Institute for the COVID-19 Genomics UK Consortium |
| EPI_ISL_439499 | Department of Pathology, University of Cambridge | Wellcome Sanger Institute for the COVID-19 Genomics UK Consortium |
| EPI_ISL_439500 | Department of Pathology, University of Cambridge | Wellcome Sanger Institute for the COVID-19 Genomics UK Consortium |
| EPI_ISL_439501 | Department of Pathology, University of Cambridge | Wellcome Sanger Institute for the COVID-19 Genomics UK Consortium |

| EPI_Accession_ID | Originating Laboratory | Submitting Laboratory |
|---|---|---|
| EPI_ISL_439502 | Department of Pathology, University of Cambridge | Wellcome Sanger Institute for the COVID-19 Genomics UK Consortium |
| EPI_ISL_439503 | Department of Pathology, University of Cambridge | Wellcome Sanger Institute for the COVID-19 Genomics UK Consortium |
| EPI_ISL_439504 | Department of Pathology, University of Cambridge | Wellcome Sanger Institute for the COVID-19 Genomics UK Consortium |
| EPI_ISL_439505 | Department of Pathology, University of Cambridge | Wellcome Sanger Institute for the COVID-19 Genomics UK Consortium |
| EPI_ISL_439506 | Department of Pathology, University of Cambridge | Wellcome Sanger Institute for the COVID-19 Genomics UK Consortium |
| EPI_ISL_439507 | Department of Pathology, University of Cambridge | Wellcome Sanger Institute for the COVID-19 Genomics UK Consortium |
| EPI_ISL_439508 | Department of Pathology, University of Cambridge | Wellcome Sanger Institute for the COVID-19 Genomics UK Consortium |
| EPI_ISL_439509 | Department of Pathology, University of Cambridge | Wellcome Sanger Institute for the COVID-19 Genomics UK Consortium |
| EPI_ISL_439510 | Department of Pathology, University of Cambridge | Wellcome Sanger Institute for the COVID-19 Genomics UK Consortium |
| EPI_ISL_439511 | Department of Pathology, University of Cambridge | Wellcome Sanger Institute for the COVID-19 Genomics UK Consortium |
| EPI_ISL_439512 | Department of Pathology, University of Cambridge | Wellcome Sanger Institute for the COVID-19 Genomics UK Consortium |
| EPI_ISL_439513 | Department of Pathology, University of Cambridge | Wellcome Sanger Institute for the COVID-19 Genomics UK Consortium |
| EPI_ISL_439514 | Department of Pathology, University of Cambridge | Wellcome Sanger Institute for the COVID-19 Genomics UK Consortium |
| EPI_ISL_439515 | Department of Pathology, University of Cambridge | Wellcome Sanger Institute for the COVID-19 Genomics UK Consortium |
| EPI_ISL_439516 | Department of Pathology, University of Cambridge | Wellcome Sanger Institute for the COVID-19 Genomics UK Consortium |
| EPI_ISL_439517 | Department of Pathology, University of Cambridge | Wellcome Sanger Institute for the COVID-19 Genomics UK Consortium |

| EPI_Accession_ID | Originating Laboratory | Submitting Laboratory |
|---|---|---|
| EPI_ISL_439518 | Department of Pathology, University of Cambridge | Wellcome Sanger Institute for the COVID-19 Genomics UK Consortium |
| EPI_ISL_439519 | Department of Pathology, University of Cambridge | Wellcome Sanger Institute for the COVID-19 Genomics UK Consortium |
| EPI_ISL_439520 | Department of Pathology, University of Cambridge | Wellcome Sanger Institute for the COVID-19 Genomics UK Consortium |
| EPI_ISL_439521 | Department of Pathology, University of Cambridge | Wellcome Sanger Institute for the COVID-19 Genomics UK Consortium |
| EPI_ISL_439522 | Department of Pathology, University of Cambridge | Wellcome Sanger Institute for the COVID-19 Genomics UK Consortium |
| EPI_ISL_439523 | Department of Pathology, University of Cambridge | Wellcome Sanger Institute for the COVID-19 Genomics UK Consortium |
| EPI_ISL_439524 | Department of Pathology, University of Cambridge | Wellcome Sanger Institute for the COVID-19 Genomics UK Consortium |
| EPI_ISL_439525 | Department of Pathology, University of Cambridge | Wellcome Sanger Institute for the COVID-19 Genomics UK Consortium |
| EPI_ISL_439526 | Department of Pathology, University of Cambridge | Wellcome Sanger Institute for the COVID-19 Genomics UK Consortium |
| EPI_ISL_439527 | Department of Pathology, University of Cambridge | Wellcome Sanger Institute for the COVID-19 Genomics UK Consortium |
| EPI_ISL_439528 | Department of Pathology, University of Cambridge | Wellcome Sanger Institute for the COVID-19 Genomics UK Consortium |
| EPI_ISL_439529 | Department of Pathology, University of Cambridge | Wellcome Sanger Institute for the COVID-19 Genomics UK Consortium |
| EPI_ISL_439530 | Department of Pathology, University of Cambridge | Wellcome Sanger Institute for the COVID-19 Genomics UK Consortium |
| EPI_ISL_439531 | Department of Pathology, University of Cambridge | Wellcome Sanger Institute for the COVID-19 Genomics UK Consortium |
| EPI_ISL_439532 | Department of Pathology, University of Cambridge | Wellcome Sanger Institute for the COVID-19 Genomics UK Consortium |
| EPI_ISL_439533 | Department of Pathology, University of Cambridge | Wellcome Sanger Institute for the COVID-19 Genomics UK Consortium |

| EPI_Accession_ID | Originating Laboratory | Submitting Laboratory |
|---|---|---|
| EPI_ISL_439534 | Department of Pathology, University of Cambridge | Wellcome Sanger Institute for the COVID-19 Genomics UK Consortium |
| EPI_ISL_439535 | Department of Pathology, University of Cambridge | Wellcome Sanger Institute for the COVID-19 Genomics UK Consortium |
| EPI_ISL_439536 | Department of Pathology, University of Cambridge | Wellcome Sanger Institute for the COVID-19 Genomics UK Consortium |
| EPI_ISL_439537 | Department of Pathology, University of Cambridge | Wellcome Sanger Institute for the COVID-19 Genomics UK Consortium |
| EPI_ISL_439538 | Department of Pathology, University of Cambridge | Wellcome Sanger Institute for the COVID-19 Genomics UK Consortium |
| EPI_ISL_439539 | Department of Pathology, University of Cambridge | Wellcome Sanger Institute for the COVID-19 Genomics UK Consortium |
| EPI_ISL_439540 | Department of Pathology, University of Cambridge | Wellcome Sanger Institute for the COVID-19 Genomics UK Consortium |
| EPI_ISL_439541 | Department of Pathology, University of Cambridge | Wellcome Sanger Institute for the COVID-19 Genomics UK Consortium |
| EPI_ISL_439542 | Department of Pathology, University of Cambridge | Wellcome Sanger Institute for the COVID-19 Genomics UK Consortium |
| EPI_ISL_439543 | Department of Pathology, University of Cambridge | Wellcome Sanger Institute for the COVID-19 Genomics UK Consortium |
| EPI_ISL_439544 | Department of Pathology, University of Cambridge | Wellcome Sanger Institute for the COVID-19 Genomics UK Consortium |
| EPI_ISL_439545 | Department of Pathology, University of Cambridge | Wellcome Sanger Institute for the COVID-19 Genomics UK Consortium |
| EPI_ISL_439546 | Department of Pathology, University of Cambridge | Wellcome Sanger Institute for the COVID-19 Genomics UK Consortium |
| EPI_ISL_439547 | Department of Pathology, University of Cambridge | Wellcome Sanger Institute for the COVID-19 Genomics UK Consortium |
| EPI_ISL_439548 | Department of Pathology, University of Cambridge | Wellcome Sanger Institute for the COVID-19 Genomics UK Consortium |
| EPI_ISL_439549 | Department of Pathology, University of Cambridge | Wellcome Sanger Institute for the COVID-19 Genomics UK Consortium |

| EPI_Accession_ID | Originating Laboratory | Submitting Laboratory |
|---|---|---|
| EPI_ISL_439550 | Department of Pathology, University of Cambridge | Wellcome Sanger Institute for the COVID-19 Genomics UK Consortium |
| EPI_ISL_439551 | Department of Pathology, University of Cambridge | Wellcome Sanger Institute for the COVID-19 Genomics UK Consortium |
| EPI_ISL_439552 | Department of Pathology, University of Cambridge | Wellcome Sanger Institute for the COVID-19 Genomics UK Consortium |
| EPI_ISL_439553 | Department of Pathology, University of Cambridge | Wellcome Sanger Institute for the COVID-19 Genomics UK Consortium |
| EPI_ISL_439554 | Department of Pathology, University of Cambridge | Wellcome Sanger Institute for the COVID-19 Genomics UK Consortium |
| EPI_ISL_439555 | Department of Pathology, University of Cambridge | Wellcome Sanger Institute for the COVID-19 Genomics UK Consortium |
| EPI_ISL_439556 | Department of Pathology, University of Cambridge | Wellcome Sanger Institute for the COVID-19 Genomics UK Consortium |
| EPI_ISL_439557 | Department of Pathology, University of Cambridge | Wellcome Sanger Institute for the COVID-19 Genomics UK Consortium |
| EPI_ISL_439558 | Department of Pathology, University of Cambridge | Wellcome Sanger Institute for the COVID-19 Genomics UK Consortium |
| EPI_ISL_439559 | Department of Pathology, University of Cambridge | Wellcome Sanger Institute for the COVID-19 Genomics UK Consortium |
| EPI_ISL_439560 | Department of Pathology, University of Cambridge | Wellcome Sanger Institute for the COVID-19 Genomics UK Consortium |
| EPI_ISL_439561 | Department of Pathology, University of Cambridge | Wellcome Sanger Institute for the COVID-19 Genomics UK Consortium |
| EPI_ISL_439562 | Department of Pathology, University of Cambridge | Wellcome Sanger Institute for the COVID-19 Genomics UK Consortium |
| EPI_ISL_439563 | Department of Pathology, University of Cambridge | Wellcome Sanger Institute for the COVID-19 Genomics UK Consortium |
| EPI_ISL_439564 | Department of Pathology, University of Cambridge | Wellcome Sanger Institute for the COVID-19 Genomics UK Consortium |
| EPI_ISL_439565 | Department of Pathology, University of Cambridge | Wellcome Sanger Institute for the COVID-19 Genomics UK Consortium |

| EPI_Accession_ID | Originating Laboratory | Submitting Laboratory |
|---|---|---|
| EPI_ISL_439566 | Department of Pathology, University of Cambridge | Wellcome Sanger Institute for the COVID-19 Genomics UK Consortium |
| EPI_ISL_439567 | Department of Pathology, University of Cambridge | Wellcome Sanger Institute for the COVID-19 Genomics UK Consortium |
| EPI_ISL_439568 | Department of Pathology, University of Cambridge | Wellcome Sanger Institute for the COVID-19 Genomics UK Consortium |
| EPI_ISL_439569 | Department of Pathology, University of Cambridge | Wellcome Sanger Institute for the COVID-19 Genomics UK Consortium |
| EPI_ISL_439570 | Department of Pathology, University of Cambridge | Wellcome Sanger Institute for the COVID-19 Genomics UK Consortium |
| EPI_ISL_439571 | Department of Pathology, University of Cambridge | Wellcome Sanger Institute for the COVID-19 Genomics UK Consortium |
| EPI_ISL_439572 | Department of Pathology, University of Cambridge | Wellcome Sanger Institute for the COVID-19 Genomics UK Consortium |
| EPI_ISL_439573 | Department of Pathology, University of Cambridge | Wellcome Sanger Institute for the COVID-19 Genomics UK Consortium |
| EPI_ISL_439574 | Department of Pathology, University of Cambridge | Wellcome Sanger Institute for the COVID-19 Genomics UK Consortium |
| EPI_ISL_439575 | Department of Pathology, University of Cambridge | Wellcome Sanger Institute for the COVID-19 Genomics UK Consortium |
| EPI_ISL_439576 | Department of Pathology, University of Cambridge | Wellcome Sanger Institute for the COVID-19 Genomics UK Consortium |
| EPI_ISL_439577 | Department of Pathology, University of Cambridge | Wellcome Sanger Institute for the COVID-19 Genomics UK Consortium |
| EPI_ISL_439578 | Department of Pathology, University of Cambridge | Wellcome Sanger Institute for the COVID-19 Genomics UK Consortium |
| EPI_ISL_439579 | Department of Pathology, University of Cambridge | Wellcome Sanger Institute for the COVID-19 Genomics UK Consortium |
| EPI_ISL_439580 | Department of Pathology, University of Cambridge | Wellcome Sanger Institute for the COVID-19 Genomics UK Consortium |
| EPI_ISL_439581 | Department of Pathology, University of Cambridge | Wellcome Sanger Institute for the COVID-19 Genomics UK Consortium |

| EPI_Accession_ID | Originating Laboratory | Submitting Laboratory |
|---|---|---|
| EPI_ISL_439582 | Department of Pathology, University of Cambridge | Wellcome Sanger Institute for the COVID-19 Genomics UK Consortium |
| EPI_ISL_439583 | Department of Pathology, University of Cambridge | Wellcome Sanger Institute for the COVID-19 Genomics UK Consortium |
| EPI_ISL_439584 | Department of Pathology, University of Cambridge | Wellcome Sanger Institute for the COVID-19 Genomics UK Consortium |
| EPI_ISL_439585 | Department of Pathology, University of Cambridge | Wellcome Sanger Institute for the COVID-19 Genomics UK Consortium |
| EPI_ISL_439586 | Department of Pathology, University of Cambridge | Wellcome Sanger Institute for the COVID-19 Genomics UK Consortium |
| EPI_ISL_439587 | Department of Pathology, University of Cambridge | Wellcome Sanger Institute for the COVID-19 Genomics UK Consortium |
| EPI_ISL_439588 | Department of Pathology, University of Cambridge | Wellcome Sanger Institute for the COVID-19 Genomics UK Consortium |
| EPI_ISL_439589 | Department of Pathology, University of Cambridge | Wellcome Sanger Institute for the COVID-19 Genomics UK Consortium |
| EPI_ISL_439590 | Department of Pathology, University of Cambridge | Wellcome Sanger Institute for the COVID-19 Genomics UK Consortium |
| EPI_ISL_439591 | Department of Pathology, University of Cambridge | Wellcome Sanger Institute for the COVID-19 Genomics UK Consortium |
| EPI_ISL_439592 | Department of Pathology, University of Cambridge | Wellcome Sanger Institute for the COVID-19 Genomics UK Consortium |
| EPI_ISL_439593 | Department of Pathology, University of Cambridge | Wellcome Sanger Institute for the COVID-19 Genomics UK Consortium |
| EPI_ISL_439594 | Department of Pathology, University of Cambridge | Wellcome Sanger Institute for the COVID-19 Genomics UK Consortium |
| EPI_ISL_439595 | Department of Pathology, University of Cambridge | Wellcome Sanger Institute for the COVID-19 Genomics UK Consortium |
| EPI_ISL_439596 | Department of Pathology, University of Cambridge | Wellcome Sanger Institute for the COVID-19 Genomics UK Consortium |
| EPI_ISL_439597 | Department of Pathology, University of Cambridge | Wellcome Sanger Institute for the COVID-19 Genomics UK Consortium |

| EPI_Accession_ID | Originating Laboratory | Submitting Laboratory |
|---|---|---|
| EPI_ISL_439598 | Department of Pathology, University of Cambridge | Wellcome Sanger Institute for the COVID-19 Genomics UK Consortium |
| EPI_ISL_439599 | Department of Pathology, University of Cambridge | Wellcome Sanger Institute for the COVID-19 Genomics UK Consortium |
| EPI_ISL_439600 | Department of Pathology, University of Cambridge | Wellcome Sanger Institute for the COVID-19 Genomics UK Consortium |
| EPI_ISL_439601 | Department of Pathology, University of Cambridge | Wellcome Sanger Institute for the COVID-19 Genomics UK Consortium |
| EPI_ISL_439602 | Department of Pathology, University of Cambridge | Wellcome Sanger Institute for the COVID-19 Genomics UK Consortium |
| EPI_ISL_439603 | Department of Pathology, University of Cambridge | Wellcome Sanger Institute for the COVID-19 Genomics UK Consortium |
| EPI_ISL_439604 | Department of Pathology, University of Cambridge | Wellcome Sanger Institute for the COVID-19 Genomics UK Consortium |
| EPI_ISL_439605 | Department of Pathology, University of Cambridge | Wellcome Sanger Institute for the COVID-19 Genomics UK Consortium |
| EPI_ISL_439606 | Department of Pathology, University of Cambridge | Wellcome Sanger Institute for the COVID-19 Genomics UK Consortium |
| EPI_ISL_439607 | Department of Pathology, University of Cambridge | Wellcome Sanger Institute for the COVID-19 Genomics UK Consortium |
| EPI_ISL_439608 | Department of Pathology, University of Cambridge | Wellcome Sanger Institute for the COVID-19 Genomics UK Consortium |
| EPI_ISL_439609 | Department of Pathology, University of Cambridge | Wellcome Sanger Institute for the COVID-19 Genomics UK Consortium |
| EPI_ISL_439610 | Department of Pathology, University of Cambridge | Wellcome Sanger Institute for the COVID-19 Genomics UK Consortium |
| EPI_ISL_439611 | Department of Pathology, University of Cambridge | Wellcome Sanger Institute for the COVID-19 Genomics UK Consortium |
| EPI_ISL_439612 | Department of Pathology, University of Cambridge | Wellcome Sanger Institute for the COVID-19 Genomics UK Consortium |
| EPI_ISL_439613 | Department of Pathology, University of Cambridge | Wellcome Sanger Institute for the COVID-19 Genomics UK Consortium |

| EPI_Accession_ID | Originating Laboratory | Submitting Laboratory |
|---|---|---|
| EPI_ISL_439614 | Department of Pathology, University of Cambridge | Wellcome Sanger Institute for the COVID-19 Genomics UK Consortium |
| EPI_ISL_439615 | Department of Pathology, University of Cambridge | Wellcome Sanger Institute for the COVID-19 Genomics UK Consortium |
| EPI_ISL_439616 | Department of Pathology, University of Cambridge | Wellcome Sanger Institute for the COVID-19 Genomics UK Consortium |
| EPI_ISL_439617 | Department of Pathology, University of Cambridge | Wellcome Sanger Institute for the COVID-19 Genomics UK Consortium |
| EPI_ISL_439618 | Department of Pathology, University of Cambridge | Wellcome Sanger Institute for the COVID-19 Genomics UK Consortium |
| EPI_ISL_439619 | Department of Pathology, University of Cambridge | Wellcome Sanger Institute for the COVID-19 Genomics UK Consortium |
| EPI_ISL_439620 | Department of Pathology, University of Cambridge | Wellcome Sanger Institute for the COVID-19 Genomics UK Consortium |
| EPI_ISL_439621 | Department of Pathology, University of Cambridge | Wellcome Sanger Institute for the COVID-19 Genomics UK Consortium |
| EPI_ISL_439622 | Department of Pathology, University of Cambridge | Wellcome Sanger Institute for the COVID-19 Genomics UK Consortium |
| EPI_ISL_439623 | Department of Pathology, University of Cambridge | Wellcome Sanger Institute for the COVID-19 Genomics UK Consortium |
| EPI_ISL_439624 | Department of Pathology, University of Cambridge | Wellcome Sanger Institute for the COVID-19 Genomics UK Consortium |
| EPI_ISL_439625 | Department of Pathology, University of Cambridge | Wellcome Sanger Institute for the COVID-19 Genomics UK Consortium |
| EPI_ISL_439626 | Department of Pathology, University of Cambridge | Wellcome Sanger Institute for the COVID-19 Genomics UK Consortium |
| EPI_ISL_439627 | Department of Pathology, University of Cambridge | Wellcome Sanger Institute for the COVID-19 Genomics UK Consortium |
| EPI_ISL_439628 | Department of Pathology, University of Cambridge | Wellcome Sanger Institute for the COVID-19 Genomics UK Consortium |
| EPI_ISL_439629 | Department of Pathology, University of Cambridge | Wellcome Sanger Institute for the COVID-19 Genomics UK Consortium |

| EPI_Accession_ID | Originating Laboratory | Submitting Laboratory |
|---|---|---|
| EPI_ISL_439630 | Department of Pathology, University of Cambridge | Wellcome Sanger Institute for the COVID-19 Genomics UK Consortium |
| EPI_ISL_439631 | Department of Pathology, University of Cambridge | Wellcome Sanger Institute for the COVID-19 Genomics UK Consortium |
| EPI_ISL_439632 | Department of Pathology, University of Cambridge | Wellcome Sanger Institute for the COVID-19 Genomics UK Consortium |
| EPI_ISL_439633 | Department of Pathology, University of Cambridge | Wellcome Sanger Institute for the COVID-19 Genomics UK Consortium |
| EPI_ISL_439634 | Department of Pathology, University of Cambridge | Wellcome Sanger Institute for the COVID-19 Genomics UK Consortium |
| EPI_ISL_439635 | Department of Pathology, University of Cambridge | Wellcome Sanger Institute for the COVID-19 Genomics UK Consortium |
| EPI_ISL_439636 | Department of Pathology, University of Cambridge | Wellcome Sanger Institute for the COVID-19 Genomics UK Consortium |
| EPI_ISL_439637 | Department of Pathology, University of Cambridge | Wellcome Sanger Institute for the COVID-19 Genomics UK Consortium |
| EPI_ISL_439638 | Department of Pathology, University of Cambridge | Wellcome Sanger Institute for the COVID-19 Genomics UK Consortium |
| EPI_ISL_439639 | Department of Pathology, University of Cambridge | Wellcome Sanger Institute for the COVID-19 Genomics UK Consortium |
| EPI_ISL_439640 | Department of Pathology, University of Cambridge | Wellcome Sanger Institute for the COVID-19 Genomics UK Consortium |
| EPI_ISL_439641 | Department of Pathology, University of Cambridge | Wellcome Sanger Institute for the COVID-19 Genomics UK Consortium |
| EPI_ISL_439642 | Department of Pathology, University of Cambridge | Wellcome Sanger Institute for the COVID-19 Genomics UK Consortium |
| EPI_ISL_439643 | Department of Pathology, University of Cambridge | Wellcome Sanger Institute for the COVID-19 Genomics UK Consortium |
| EPI_ISL_439644 | Department of Pathology, University of Cambridge | Wellcome Sanger Institute for the COVID-19 Genomics UK Consortium |
| EPI_ISL_439645 | Department of Pathology, University of Cambridge | Wellcome Sanger Institute for the COVID-19 Genomics UK Consortium |

| EPI_Accession_ID | Originating Laboratory | Submitting Laboratory |
|---|---|---|
| EPI_ISL_439646 | Department of Pathology, University of Cambridge | Wellcome Sanger Institute for the COVID-19 Genomics UK Consortium |
| EPI_ISL_439647 | Department of Pathology, University of Cambridge | Wellcome Sanger Institute for the COVID-19 Genomics UK Consortium |
| EPI_ISL_439648 | Department of Pathology, University of Cambridge | Wellcome Sanger Institute for the COVID-19 Genomics UK Consortium |
| EPI_ISL_439649 | Department of Pathology, University of Cambridge | Wellcome Sanger Institute for the COVID-19 Genomics UK Consortium |
| EPI_ISL_439650 | Department of Pathology, University of Cambridge | Wellcome Sanger Institute for the COVID-19 Genomics UK Consortium |
| EPI_ISL_439651 | Department of Pathology, University of Cambridge | Wellcome Sanger Institute for the COVID-19 Genomics UK Consortium |
| EPI_ISL_439652 | Department of Pathology, University of Cambridge | Wellcome Sanger Institute for the COVID-19 Genomics UK Consortium |
| EPI_ISL_439653 | Department of Pathology, University of Cambridge | Wellcome Sanger Institute for the COVID-19 Genomics UK Consortium |
| EPI_ISL_439654 | Department of Pathology, University of Cambridge | Wellcome Sanger Institute for the COVID-19 Genomics UK Consortium |
| EPI_ISL_439655 | Department of Pathology, University of Cambridge | Wellcome Sanger Institute for the COVID-19 Genomics UK Consortium |
| EPI_ISL_439656 | Department of Pathology, University of Cambridge | Wellcome Sanger Institute for the COVID-19 Genomics UK Consortium |
| EPI_ISL_439657 | Department of Pathology, University of Cambridge | Wellcome Sanger Institute for the COVID-19 Genomics UK Consortium |
| EPI_ISL_439658 | Department of Pathology, University of Cambridge | Wellcome Sanger Institute for the COVID-19 Genomics UK Consortium |
| EPI_ISL_439659 | Department of Pathology, University of Cambridge | Wellcome Sanger Institute for the COVID-19 Genomics UK Consortium |
| EPI_ISL_439660 | Department of Pathology, University of Cambridge | Wellcome Sanger Institute for the COVID-19 Genomics UK Consortium |
| EPI_ISL_439661 | Department of Pathology, University of Cambridge | Wellcome Sanger Institute for the COVID-19 Genomics UK Consortium |

| EPI_Accession_ID | Originating Laboratory | Submitting Laboratory |
|---|---|---|
| EPI_ISL_439662 | Department of Pathology, University of Cambridge | Wellcome Sanger Institute for the COVID-19 Genomics UK Consortium |
| EPI_ISL_439663 | Department of Pathology, University of Cambridge | Wellcome Sanger Institute for the COVID-19 Genomics UK Consortium |
| EPI_ISL_439664 | Department of Pathology, University of Cambridge | Wellcome Sanger Institute for the COVID-19 Genomics UK Consortium |
| EPI_ISL_439665 | Virology Department, Royal Infirmary of Edinburgh, NHS Lothian / School of Biological Sciences, University of Edinburgh / Institute of Genetics and Molecular Medicine, University of Edinburgh | COVID-19 Genomics UK (COG-UK) Consortium |
| EPI_ISL_439666 | Virology Department, Royal Infirmary of Edinburgh, NHS Lothian / School of Biological Sciences, University of Edinburgh / Institute of Genetics and Molecular Medicine, University of Edinburgh | COVID-19 Genomics UK (COG-UK) Consortium |
| EPI_ISL_439667 | Virology Department, Royal Infirmary of Edinburgh, NHS Lothian / School of Biological Sciences, University of Edinburgh / Institute of Genetics and Molecular Medicine, University of Edinburgh | COVID-19 Genomics UK (COG-UK) Consortium |
| EPI_ISL_439668 | Virology Department, Royal Infirmary of Edinburgh, NHS Lothian / School of Biological Sciences, University of Edinburgh / Institute of Genetics and Molecular Medicine, University of Edinburgh | COVID-19 Genomics UK (COG-UK) Consortium |
| EPI_ISL_439669 | Virology Department, Royal Infirmary of Edinburgh, NHS Lothian / School of Biological Sciences, University of Edinburgh / Institute of Genetics and Molecular Medicine, University of Edinburgh | COVID-19 Genomics UK (COG-UK) Consortium |
| EPI_ISL_439670 | Virology Department, Royal Infirmary of Edinburgh, NHS Lothian / School of Biological Sciences, University of Edinburgh / Institute of Genetics and Molecular Medicine, University of Edinburgh | COVID-19 Genomics UK (COG-UK) Consortium |

| EPI_Accession_ID | Originating Laboratory | Submitting Laboratory |
|---|---|---|
| EPI_ISL_439671 | Virology Department, Royal Infirmary of Edinburgh, NHS Lothian / School of Biological Sciences, University of Edinburgh / Institute of Genetics and Molecular Medicine, University of Edinburgh | COVID-19 Genomics UK (COG-UK) Consortium |
| EPI_ISL_439672 | Liverpool Clinical Laboratories | COVID-19 Genomics UK (COG-UK) Consortium |
| EPI_ISL_439673 | Liverpool Clinical Laboratories | COVID-19 Genomics UK (COG-UK) Consortium |
| EPI_ISL_439674 | Liverpool Clinical Laboratories | COVID-19 Genomics UK (COG-UK) Consortium |
| EPI_ISL_439675 | Liverpool Clinical Laboratories | COVID-19 Genomics UK (COG-UK) Consortium |
| EPI_ISL_439676 | Liverpool Clinical Laboratories | COVID-19 Genomics UK (COG-UK) Consortium |
| EPI_ISL_439677 | Liverpool Clinical Laboratories | COVID-19 Genomics UK (COG-UK) Consortium |
| EPI_ISL_439678 | Liverpool Clinical Laboratories | COVID-19 Genomics UK (COG-UK) Consortium |
| EPI_ISL_439679 | Liverpool Clinical Laboratories | COVID-19 Genomics UK (COG-UK) Consortium |
| EPI_ISL_439680 | Liverpool Clinical Laboratories | COVID-19 Genomics UK (COG-UK) Consortium |
| EPI_ISL_439681 | Liverpool Clinical Laboratories | COVID-19 Genomics UK (COG-UK) Consortium |
| EPI_ISL_439682 | Liverpool Clinical Laboratories | COVID-19 Genomics UK (COG-UK) Consortium |
| EPI_ISL_439683 | Liverpool Clinical Laboratories | COVID-19 Genomics UK (COG-UK) Consortium |
| EPI_ISL_439684 | Liverpool Clinical Laboratories | COVID-19 Genomics UK (COG-UK) Consortium |
| EPI_ISL_439685 | Liverpool Clinical Laboratories | COVID-19 Genomics UK (COG-UK) Consortium |
| EPI_ISL_439686 | Liverpool Clinical Laboratories | COVID-19 Genomics UK (COG-UK) Consortium |
| EPI_ISL_439687 | Liverpool Clinical Laboratories | COVID-19 Genomics UK (COG-UK) Consortium |
| EPI_ISL_439688 | Liverpool Clinical Laboratories | COVID-19 Genomics UK (COG-UK) Consortium |
| EPI_ISL_439689 | Liverpool Clinical Laboratories | COVID-19 Genomics UK (COG-UK) Consortium |
| EPI_ISL_439690 | Liverpool Clinical Laboratories | COVID-19 Genomics UK (COG-UK) Consortium |
| EPI_ISL_439691 | Liverpool Clinical Laboratories | COVID-19 Genomics UK (COG-UK) Consortium |
| EPI_ISL_439692 | Liverpool Clinical Laboratories | COVID-19 Genomics UK (COG-UK) Consortium |
| EPI_ISL_439693 | Liverpool Clinical Laboratories | COVID-19 Genomics UK (COG-UK) Consortium |
| EPI_ISL_439694 | Liverpool Clinical Laboratories | COVID-19 Genomics UK (COG-UK) Consortium |
| EPI_ISL_439695 | Liverpool Clinical Laboratories | COVID-19 Genomics UK (COG-UK) Consortium |
| EPI_ISL_439696 | Liverpool Clinical Laboratories | COVID-19 Genomics UK (COG-UK) Consortium |
| EPI_ISL_439697 | Liverpool Clinical Laboratories | COVID-19 Genomics UK (COG-UK) Consortium |
| EPI_ISL_439698 | Liverpool Clinical Laboratories | COVID-19 Genomics UK (COG-UK) Consortium |
| EPI_ISL_439699 | Liverpool Clinical Laboratories | COVID-19 Genomics UK (COG-UK) Consortium |
| EPI_ISL_439700 | Liverpool Clinical Laboratories | COVID-19 Genomics UK (COG-UK) Consortium |

| EPI_Accession_ID | Originating Laboratory | Submitting Laboratory |
|---|---|---|
| EPI_ISL_439701 | Liverpool Clinical Laboratories | COVID-19 Genomics UK (COG-UK) Consortium |
| EPI_ISL_439702 | Liverpool Clinical Laboratories | COVID-19 Genomics UK (COG-UK) Consortium |
| EPI_ISL_439703 | Liverpool Clinical Laboratories | COVID-19 Genomics UK (COG-UK) Consortium |
| EPI_ISL_439704 | Liverpool Clinical Laboratories | COVID-19 Genomics UK (COG-UK) Consortium |
| EPI_ISL_439705 | Liverpool Clinical Laboratories | COVID-19 Genomics UK (COG-UK) Consortium |
| EPI_ISL_439706 | Liverpool Clinical Laboratories | COVID-19 Genomics UK (COG-UK) Consortium |
| EPI_ISL_439707 | Liverpool Clinical Laboratories | COVID-19 Genomics UK (COG-UK) Consortium |
| EPI_ISL_439708 | Liverpool Clinical Laboratories | COVID-19 Genomics UK (COG-UK) Consortium |
| EPI_ISL_439709 | Liverpool Clinical Laboratories | COVID-19 Genomics UK (COG-UK) Consortium |
| EPI_ISL_439710 | Liverpool Clinical Laboratories | COVID-19 Genomics UK (COG-UK) Consortium |
| EPI_ISL_439711 | Liverpool Clinical Laboratories | COVID-19 Genomics UK (COG-UK) Consortium |
| EPI_ISL_439712 | Liverpool Clinical Laboratories | COVID-19 Genomics UK (COG-UK) Consortium |
| EPI_ISL_439713 | Liverpool Clinical Laboratories | COVID-19 Genomics UK (COG-UK) Consortium |
| EPI_ISL_439714 | Liverpool Clinical Laboratories | COVID-19 Genomics UK (COG-UK) Consortium |
| EPI_ISL_439715 | Liverpool Clinical Laboratories | COVID-19 Genomics UK (COG-UK) Consortium |
| EPI_ISL_439716 | Liverpool Clinical Laboratories | COVID-19 Genomics UK (COG-UK) Consortium |
| EPI_ISL_439717 | Liverpool Clinical Laboratories | COVID-19 Genomics UK (COG-UK) Consortium |
| EPI_ISL_439718 | Liverpool Clinical Laboratories | COVID-19 Genomics UK (COG-UK) Consortium |
| EPI_ISL_439719 | Liverpool Clinical Laboratories | COVID-19 Genomics UK (COG-UK) Consortium |
| EPI_ISL_439720 | Liverpool Clinical Laboratories | COVID-19 Genomics UK (COG-UK) Consortium |
| EPI_ISL_439721 | Liverpool Clinical Laboratories | COVID-19 Genomics UK (COG-UK) Consortium |
| EPI_ISL_439722 | Liverpool Clinical Laboratories | COVID-19 Genomics UK (COG-UK) Consortium |
| EPI_ISL_439723 | Liverpool Clinical Laboratories | COVID-19 Genomics UK (COG-UK) Consortium |
| EPI_ISL_439724 | Liverpool Clinical Laboratories | COVID-19 Genomics UK (COG-UK) Consortium |
| EPI_ISL_439725 | Liverpool Clinical Laboratories | COVID-19 Genomics UK (COG-UK) Consortium |
| EPI_ISL_439726 | Liverpool Clinical Laboratories | COVID-19 Genomics UK (COG-UK) Consortium |
| EPI_ISL_439727 | Liverpool Clinical Laboratories | COVID-19 Genomics UK (COG-UK) Consortium |
| EPI_ISL_439728 | Liverpool Clinical Laboratories | COVID-19 Genomics UK (COG-UK) Consortium |
| EPI_ISL_439729 | Liverpool Clinical Laboratories | COVID-19 Genomics UK (COG-UK) Consortium |
| EPI_ISL_439730 | Liverpool Clinical Laboratories | COVID-19 Genomics UK (COG-UK) Consortium |
| EPI_ISL_439731 | Liverpool Clinical Laboratories | COVID-19 Genomics UK (COG-UK) Consortium |
| EPI_ISL_439732 | Liverpool Clinical Laboratories | COVID-19 Genomics UK (COG-UK) Consortium |
| EPI_ISL_439733 | Liverpool Clinical Laboratories | COVID-19 Genomics UK (COG-UK) Consortium |

| EPI_Accession_ID | Originating Laboratory | Submitting Laboratory |
|---|---|---|
| EPI_ISL_439734 | Liverpool Clinical Laboratories | COVID-19 Genomics UK (COG-UK) Consortium |
| EPI_ISL_439735 | Liverpool Clinical Laboratories | COVID-19 Genomics UK (COG-UK) Consortium |
| EPI_ISL_439736 | Liverpool Clinical Laboratories | COVID-19 Genomics UK (COG-UK) Consortium |
| EPI_ISL_439737 | Liverpool Clinical Laboratories | COVID-19 Genomics UK (COG-UK) Consortium |
| EPI_ISL_439738 | Liverpool Clinical Laboratories | COVID-19 Genomics UK (COG-UK) Consortium |
| EPI_ISL_439739 | Liverpool Clinical Laboratories | COVID-19 Genomics UK (COG-UK) Consortium |
| EPI_ISL_439740 | Liverpool Clinical Laboratories | COVID-19 Genomics UK (COG-UK) Consortium |
| EPI_ISL_439741 | Liverpool Clinical Laboratories | COVID-19 Genomics UK (COG-UK) Consortium |
| EPI_ISL_439742 | Liverpool Clinical Laboratories | COVID-19 Genomics UK (COG-UK) Consortium |
| EPI_ISL_439743 | Liverpool Clinical Laboratories | COVID-19 Genomics UK (COG-UK) Consortium |
| EPI_ISL_439744 | Liverpool Clinical Laboratories | COVID-19 Genomics UK (COG-UK) Consortium |
| EPI_ISL_439745 | Liverpool Clinical Laboratories | COVID-19 Genomics UK (COG-UK) Consortium |
| EPI_ISL_439746 | Liverpool Clinical Laboratories | COVID-19 Genomics UK (COG-UK) Consortium |
| EPI_ISL_439747 | Liverpool Clinical Laboratories | COVID-19 Genomics UK (COG-UK) Consortium |
| EPI_ISL_439748 | Liverpool Clinical Laboratories | COVID-19 Genomics UK (COG-UK) Consortium |
| EPI_ISL_439749 | Liverpool Clinical Laboratories | COVID-19 Genomics UK (COG-UK) Consortium |
| EPI_ISL_439750 | Liverpool Clinical Laboratories | COVID-19 Genomics UK (COG-UK) Consortium |
| EPI_ISL_439751 | Liverpool Clinical Laboratories | COVID-19 Genomics UK (COG-UK) Consortium |
| EPI_ISL_439752 | Liverpool Clinical Laboratories | COVID-19 Genomics UK (COG-UK) Consortium |
| EPI_ISL_439753 | Liverpool Clinical Laboratories | COVID-19 Genomics UK (COG-UK) Consortium |
| EPI_ISL_439754 | Liverpool Clinical Laboratories | COVID-19 Genomics UK (COG-UK) Consortium |
| EPI_ISL_439755 | Liverpool Clinical Laboratories | COVID-19 Genomics UK (COG-UK) Consortium |
| EPI_ISL_439756 | Liverpool Clinical Laboratories | COVID-19 Genomics UK (COG-UK) Consortium |
| EPI_ISL_439757 | Liverpool Clinical Laboratories | COVID-19 Genomics UK (COG-UK) Consortium |
| EPI_ISL_439758 | Liverpool Clinical Laboratories | COVID-19 Genomics UK (COG-UK) Consortium |
| EPI_ISL_439759 | Liverpool Clinical Laboratories | COVID-19 Genomics UK (COG-UK) Consortium |
| EPI_ISL_439760 | Liverpool Clinical Laboratories | COVID-19 Genomics UK (COG-UK) Consortium |
| EPI_ISL_439761 | Liverpool Clinical Laboratories | COVID-19 Genomics UK (COG-UK) Consortium |
| EPI_ISL_439762 | Liverpool Clinical Laboratories | COVID-19 Genomics UK (COG-UK) Consortium |
| EPI_ISL_439763 | Liverpool Clinical Laboratories | COVID-19 Genomics UK (COG-UK) Consortium |
| EPI_ISL_439764 | Liverpool Clinical Laboratories | COVID-19 Genomics UK (COG-UK) Consortium |
| EPI_ISL_439765 | Liverpool Clinical Laboratories | COVID-19 Genomics UK (COG-UK) Consortium |
| EPI_ISL_439766 | Liverpool Clinical Laboratories | COVID-19 Genomics UK (COG-UK) Consortium |

| EPI_Accession_ID | Originating Laboratory | Submitting Laboratory |
|---|---|---|
| EPI_ISL_439767 | Liverpool Clinical Laboratories | COVID-19 Genomics UK (COG-UK) Consortium |
| EPI_ISL_439768 | Liverpool Clinical Laboratories | COVID-19 Genomics UK (COG-UK) Consortium |
| EPI_ISL_439769 | Liverpool Clinical Laboratories | COVID-19 Genomics UK (COG-UK) Consortium |
| EPI_ISL_439770 | Liverpool Clinical Laboratories | COVID-19 Genomics UK (COG-UK) Consortium |
| EPI_ISL_439771 | Liverpool Clinical Laboratories | COVID-19 Genomics UK (COG-UK) Consortium |
| EPI_ISL_439772 | Liverpool Clinical Laboratories | COVID-19 Genomics UK (COG-UK) Consortium |
| EPI_ISL_439773 | Liverpool Clinical Laboratories | COVID-19 Genomics UK (COG-UK) Consortium |
| EPI_ISL_439774 | Liverpool Clinical Laboratories | COVID-19 Genomics UK (COG-UK) Consortium |
| EPI_ISL_439775 | Liverpool Clinical Laboratories | COVID-19 Genomics UK (COG-UK) Consortium |
| EPI_ISL_439776 | Liverpool Clinical Laboratories | COVID-19 Genomics UK (COG-UK) Consortium |
| EPI_ISL_439777 | Liverpool Clinical Laboratories | COVID-19 Genomics UK (COG-UK) Consortium |
| EPI_ISL_439778 | Liverpool Clinical Laboratories | COVID-19 Genomics UK (COG-UK) Consortium |
| EPI_ISL_439779 | Liverpool Clinical Laboratories | COVID-19 Genomics UK (COG-UK) Consortium |
| EPI_ISL_439780 | Liverpool Clinical Laboratories | COVID-19 Genomics UK (COG-UK) Consortium |
| EPI_ISL_439781 | Liverpool Clinical Laboratories | COVID-19 Genomics UK (COG-UK) Consortium |
| EPI_ISL_439782 | Liverpool Clinical Laboratories | COVID-19 Genomics UK (COG-UK) Consortium |
| EPI_ISL_439783 | Liverpool Clinical Laboratories | COVID-19 Genomics UK (COG-UK) Consortium |
| EPI_ISL_439784 | Liverpool Clinical Laboratories | COVID-19 Genomics UK (COG-UK) Consortium |
| EPI_ISL_439785 | Liverpool Clinical Laboratories | COVID-19 Genomics UK (COG-UK) Consortium |
| EPI_ISL_439786 | Liverpool Clinical Laboratories | COVID-19 Genomics UK (COG-UK) Consortium |
| EPI_ISL_439787 | Liverpool Clinical Laboratories | COVID-19 Genomics UK (COG-UK) Consortium |
| EPI_ISL_439788 | Liverpool Clinical Laboratories | COVID-19 Genomics UK (COG-UK) Consortium |
| EPI_ISL_439789 | Liverpool Clinical Laboratories | COVID-19 Genomics UK (COG-UK) Consortium |
| EPI_ISL_439790 | Liverpool Clinical Laboratories | COVID-19 Genomics UK (COG-UK) Consortium |
| EPI_ISL_439791 | Liverpool Clinical Laboratories | COVID-19 Genomics UK (COG-UK) Consortium |
| EPI_ISL_439792 | Liverpool Clinical Laboratories | COVID-19 Genomics UK (COG-UK) Consortium |
| EPI_ISL_439793 | Liverpool Clinical Laboratories | COVID-19 Genomics UK (COG-UK) Consortium |
| EPI_ISL_439794 | Liverpool Clinical Laboratories | COVID-19 Genomics UK (COG-UK) Consortium |
| EPI_ISL_439795 | Liverpool Clinical Laboratories | COVID-19 Genomics UK (COG-UK) Consortium |
| EPI_ISL_439796 | Liverpool Clinical Laboratories | COVID-19 Genomics UK (COG-UK) Consortium |
| EPI_ISL_439797 | Liverpool Clinical Laboratories | COVID-19 Genomics UK (COG-UK) Consortium |
| EPI_ISL_439798 | Liverpool Clinical Laboratories | COVID-19 Genomics UK (COG-UK) Consortium |
| EPI_ISL_439799 | Liverpool Clinical Laboratories | COVID-19 Genomics UK (COG-UK) Consortium |

| EPI_Accession_ID | Originating Laboratory | Submitting Laboratory |
|---|---|---|
| EPI_ISL_439800 | Liverpool Clinical Laboratories | COVID-19 Genomics UK (COG-UK) Consortium |
| EPI_ISL_439801 | Liverpool Clinical Laboratories | COVID-19 Genomics UK (COG-UK) Consortium |
| EPI_ISL_439802 | Liverpool Clinical Laboratories | COVID-19 Genomics UK (COG-UK) Consortium |
| EPI_ISL_439803 | Liverpool Clinical Laboratories | COVID-19 Genomics UK (COG-UK) Consortium |
| EPI_ISL_439804 | Liverpool Clinical Laboratories | COVID-19 Genomics UK (COG-UK) Consortium |
| EPI_ISL_439805 | Liverpool Clinical Laboratories | COVID-19 Genomics UK (COG-UK) Consortium |
| EPI_ISL_439806 | Liverpool Clinical Laboratories | COVID-19 Genomics UK (COG-UK) Consortium |
| EPI_ISL_439807 | Liverpool Clinical Laboratories | COVID-19 Genomics UK (COG-UK) Consortium |
| EPI_ISL_439808 | Liverpool Clinical Laboratories | COVID-19 Genomics UK (COG-UK) Consortium |
| EPI_ISL_439809 | Liverpool Clinical Laboratories | COVID-19 Genomics UK (COG-UK) Consortium |
| EPI_ISL_439810 | Liverpool Clinical Laboratories | COVID-19 Genomics UK (COG-UK) Consortium |
| EPI_ISL_439811 | Liverpool Clinical Laboratories | COVID-19 Genomics UK (COG-UK) Consortium |
| EPI_ISL_439812 | Liverpool Clinical Laboratories | COVID-19 Genomics UK (COG-UK) Consortium |
| EPI_ISL_439813 | Liverpool Clinical Laboratories | COVID-19 Genomics UK (COG-UK) Consortium |
| EPI_ISL_439814 | Liverpool Clinical Laboratories | COVID-19 Genomics UK (COG-UK) Consortium |
| EPI_ISL_439815 | Liverpool Clinical Laboratories | COVID-19 Genomics UK (COG-UK) Consortium |
| EPI_ISL_439816 | Liverpool Clinical Laboratories | COVID-19 Genomics UK (COG-UK) Consortium |
| EPI_ISL_439817 | Liverpool Clinical Laboratories | COVID-19 Genomics UK (COG-UK) Consortium |
| EPI_ISL_439818 | Liverpool Clinical Laboratories | COVID-19 Genomics UK (COG-UK) Consortium |
| EPI_ISL_439819 | Liverpool Clinical Laboratories | COVID-19 Genomics UK (COG-UK) Consortium |
| EPI_ISL_439820 | Liverpool Clinical Laboratories | COVID-19 Genomics UK (COG-UK) Consortium |
| EPI_ISL_439821 | Liverpool Clinical Laboratories | COVID-19 Genomics UK (COG-UK) Consortium |
| EPI_ISL_439822 | Liverpool Clinical Laboratories | COVID-19 Genomics UK (COG-UK) Consortium |
| EPI_ISL_439823 | Liverpool Clinical Laboratories | COVID-19 Genomics UK (COG-UK) Consortium |
| EPI_ISL_439824 | Liverpool Clinical Laboratories | COVID-19 Genomics UK (COG-UK) Consortium |
| EPI_ISL_439825 | Liverpool Clinical Laboratories | COVID-19 Genomics UK (COG-UK) Consortium |
| EPI_ISL_439826 | Liverpool Clinical Laboratories | COVID-19 Genomics UK (COG-UK) Consortium |
| EPI_ISL_439827 | Liverpool Clinical Laboratories | COVID-19 Genomics UK (COG-UK) Consortium |
| EPI_ISL_439828 | Liverpool Clinical Laboratories | COVID-19 Genomics UK (COG-UK) Consortium |
| EPI_ISL_439829 | Liverpool Clinical Laboratories | COVID-19 Genomics UK (COG-UK) Consortium |
| EPI_ISL_439830 | Liverpool Clinical Laboratories | COVID-19 Genomics UK (COG-UK) Consortium |
| EPI_ISL_439831 | Liverpool Clinical Laboratories | COVID-19 Genomics UK (COG-UK) Consortium |
| EPI_ISL_439832 | Liverpool Clinical Laboratories | COVID-19 Genomics UK (COG-UK) Consortium |

| EPI_Accession_ID | Originating Laboratory | Submitting Laboratory |
|---|---|---|
| EPI_ISL_439833 | Liverpool Clinical Laboratories | COVID-19 Genomics UK (COG-UK) Consortium |
| EPI_ISL_439834 | Liverpool Clinical Laboratories | COVID-19 Genomics UK (COG-UK) Consortium |
| EPI_ISL_439835 | Liverpool Clinical Laboratories | COVID-19 Genomics UK (COG-UK) Consortium |
| EPI_ISL_439836 | Liverpool Clinical Laboratories | COVID-19 Genomics UK (COG-UK) Consortium |
| EPI_ISL_439837 | Liverpool Clinical Laboratories | COVID-19 Genomics UK (COG-UK) Consortium |
| EPI_ISL_439838 | Liverpool Clinical Laboratories | COVID-19 Genomics UK (COG-UK) Consortium |
| EPI_ISL_439839 | Liverpool Clinical Laboratories | COVID-19 Genomics UK (COG-UK) Consortium |
| EPI_ISL_439840 | Liverpool Clinical Laboratories | COVID-19 Genomics UK (COG-UK) Consortium |
| EPI_ISL_439841 | Liverpool Clinical Laboratories | COVID-19 Genomics UK (COG-UK) Consortium |
| EPI_ISL_439842 | Liverpool Clinical Laboratories | COVID-19 Genomics UK (COG-UK) Consortium |
| EPI_ISL_439843 | Liverpool Clinical Laboratories | COVID-19 Genomics UK (COG-UK) Consortium |
| EPI_ISL_439844 | Liverpool Clinical Laboratories | COVID-19 Genomics UK (COG-UK) Consortium |
| EPI_ISL_439845 | Liverpool Clinical Laboratories | COVID-19 Genomics UK (COG-UK) Consortium |
| EPI_ISL_439846 | Liverpool Clinical Laboratories | COVID-19 Genomics UK (COG-UK) Consortium |
| EPI_ISL_439847 | Liverpool Clinical Laboratories | COVID-19 Genomics UK (COG-UK) Consortium |
| EPI_ISL_439848 | Liverpool Clinical Laboratories | COVID-19 Genomics UK (COG-UK) Consortium |
| EPI_ISL_439849 | Liverpool Clinical Laboratories | COVID-19 Genomics UK (COG-UK) Consortium |
| EPI_ISL_439850 | Liverpool Clinical Laboratories | COVID-19 Genomics UK (COG-UK) Consortium |
| EPI_ISL_439851 | Liverpool Clinical Laboratories | COVID-19 Genomics UK (COG-UK) Consortium |
| EPI_ISL_439852 | Liverpool Clinical Laboratories | COVID-19 Genomics UK (COG-UK) Consortium |
| EPI_ISL_439853 | Liverpool Clinical Laboratories | COVID-19 Genomics UK (COG-UK) Consortium |
| EPI_ISL_439854 | Liverpool Clinical Laboratories | COVID-19 Genomics UK (COG-UK) Consortium |
| EPI_ISL_439855 | Liverpool Clinical Laboratories | COVID-19 Genomics UK (COG-UK) Consortium |
| EPI_ISL_439856 | Liverpool Clinical Laboratories | COVID-19 Genomics UK (COG-UK) Consortium |
| EPI_ISL_439857 | Liverpool Clinical Laboratories | COVID-19 Genomics UK (COG-UK) Consortium |
| EPI_ISL_439858 | Liverpool Clinical Laboratories | COVID-19 Genomics UK (COG-UK) Consortium |
| EPI_ISL_439859 | Liverpool Clinical Laboratories | COVID-19 Genomics UK (COG-UK) Consortium |
| EPI_ISL_439860 | Liverpool Clinical Laboratories | COVID-19 Genomics UK (COG-UK) Consortium |
| EPI_ISL_439861 | Liverpool Clinical Laboratories | COVID-19 Genomics UK (COG-UK) Consortium |
| EPI_ISL_439862 | Liverpool Clinical Laboratories | COVID-19 Genomics UK (COG-UK) Consortium |
| EPI_ISL_439863 | Department of Pathology, University of Cambridge | Wellcome Sanger Institute for the COVID-19 Genomics UK Consortium |

| EPI_Accession_ID | Originating Laboratory | Submitting Laboratory |
|---|---|---|
| EPI_ISL_439864 | Department of Pathology, University of Cambridge | Wellcome Sanger Institute for the COVID-19 Genomics UK Consortium |
| EPI_ISL_439865 | Department of Pathology, University of Cambridge | Wellcome Sanger Institute for the COVID-19 Genomics UK Consortium |
| EPI_ISL_439866 | Department of Pathology, University of Cambridge | Wellcome Sanger Institute for the COVID-19 Genomics UK Consortium |
| EPI_ISL_439867 | Department of Pathology, University of Cambridge | Wellcome Sanger Institute for the COVID-19 Genomics UK Consortium |
| EPI_ISL_439868 | Department of Pathology, University of Cambridge | Wellcome Sanger Institute for the COVID-19 Genomics UK Consortium |
| EPI_ISL_439869 | Department of Pathology, University of Cambridge | Wellcome Sanger Institute for the COVID-19 Genomics UK Consortium |
| EPI_ISL_439870 | Department of Pathology, University of Cambridge | Wellcome Sanger Institute for the COVID-19 Genomics UK Consortium |
| EPI_ISL_439871 | Department of Pathology, University of Cambridge | Wellcome Sanger Institute for the COVID-19 Genomics UK Consortium |
| EPI_ISL_439872 | Department of Pathology, University of Cambridge | Wellcome Sanger Institute for the COVID-19 Genomics UK Consortium |
| EPI_ISL_439873 | Department of Pathology, University of Cambridge | Wellcome Sanger Institute for the COVID-19 Genomics UK Consortium |
| EPI_ISL_439874 | Department of Pathology, University of Cambridge | Wellcome Sanger Institute for the COVID-19 Genomics UK Consortium |
| EPI_ISL_439875 | Department of Pathology, University of Cambridge | Wellcome Sanger Institute for the COVID-19 Genomics UK Consortium |
| EPI_ISL_439876 | Department of Pathology, University of Cambridge | Wellcome Sanger Institute for the COVID-19 Genomics UK Consortium |
| EPI_ISL_439877 | Department of Pathology, University of Cambridge | Wellcome Sanger Institute for the COVID-19 Genomics UK Consortium |
| EPI_ISL_439878 | Department of Pathology, University of Cambridge | Wellcome Sanger Institute for the COVID-19 Genomics UK Consortium |
| EPI_ISL_439879 | Department of Pathology, University of Cambridge | Wellcome Sanger Institute for the COVID-19 Genomics UK Consortium |

| EPI_Accession_ID | Originating Laboratory | Submitting Laboratory |
|---|---|---|
| EPI_ISL_439880 | Department of Pathology, University of Cambridge | Wellcome Sanger Institute for the COVID-19 Genomics UK Consortium |
| EPI_ISL_439881 | Department of Pathology, University of Cambridge | Wellcome Sanger Institute for the COVID-19 Genomics UK Consortium |
| EPI_ISL_439882 | Department of Pathology, University of Cambridge | Wellcome Sanger Institute for the COVID-19 Genomics UK Consortium |
| EPI_ISL_439883 | Department of Pathology, University of Cambridge | Wellcome Sanger Institute for the COVID-19 Genomics UK Consortium |
| EPI_ISL_439884 | Department of Pathology, University of Cambridge | Wellcome Sanger Institute for the COVID-19 Genomics UK Consortium |
| EPI_ISL_439885 | Department of Pathology, University of Cambridge | Wellcome Sanger Institute for the COVID-19 Genomics UK Consortium |
| EPI_ISL_439886 | Department of Pathology, University of Cambridge | Wellcome Sanger Institute for the COVID-19 Genomics UK Consortium |
| EPI_ISL_439887 | Department of Pathology, University of Cambridge | Wellcome Sanger Institute for the COVID-19 Genomics UK Consortium |
| EPI_ISL_439888 | Department of Pathology, University of Cambridge | Wellcome Sanger Institute for the COVID-19 Genomics UK Consortium |
| EPI_ISL_439889 | Department of Pathology, University of Cambridge | Wellcome Sanger Institute for the COVID-19 Genomics UK Consortium |
| EPI_ISL_439890 | Department of Pathology, University of Cambridge | Wellcome Sanger Institute for the COVID-19 Genomics UK Consortium |
| EPI_ISL_439891 | Department of Pathology, University of Cambridge | Wellcome Sanger Institute for the COVID-19 Genomics UK Consortium |
| EPI_ISL_439892 | Department of Pathology, University of Cambridge | Wellcome Sanger Institute for the COVID-19 Genomics UK Consortium |
| EPI_ISL_439893 | Department of Pathology, University of Cambridge | Wellcome Sanger Institute for the COVID-19 Genomics UK Consortium |
| EPI_ISL_439894 | Department of Pathology, University of Cambridge | Wellcome Sanger Institute for the COVID-19 Genomics UK Consortium |
| EPI_ISL_439895 | Department of Pathology, University of Cambridge | Wellcome Sanger Institute for the COVID-19 Genomics UK Consortium |

| EPI_Accession_ID | Originating Laboratory | Submitting Laboratory |
|---|---|---|
| EPI_ISL_439896 | Department of Pathology, University of Cambridge | Wellcome Sanger Institute for the COVID-19 Genomics UK Consortium |
| EPI_ISL_439897 | Department of Pathology, University of Cambridge | Wellcome Sanger Institute for the COVID-19 Genomics UK Consortium |
| EPI_ISL_439898 | Department of Pathology, University of Cambridge | Wellcome Sanger Institute for the COVID-19 Genomics UK Consortium |
| EPI_ISL_439899 | Department of Pathology, University of Cambridge | Wellcome Sanger Institute for the COVID-19 Genomics UK Consortium |
| EPI_ISL_439900 | Department of Pathology, University of Cambridge | Wellcome Sanger Institute for the COVID-19 Genomics UK Consortium |
| EPI_ISL_439901 | Department of Pathology, University of Cambridge | Wellcome Sanger Institute for the COVID-19 Genomics UK Consortium |
| EPI_ISL_439902 | Department of Pathology, University of Cambridge | Wellcome Sanger Institute for the COVID-19 Genomics UK Consortium |
| EPI_ISL_439903 | Department of Pathology, University of Cambridge | Wellcome Sanger Institute for the COVID-19 Genomics UK Consortium |
| EPI_ISL_439904 | Department of Pathology, University of Cambridge | Wellcome Sanger Institute for the COVID-19 Genomics UK Consortium |
| EPI_ISL_439905 | Department of Pathology, University of Cambridge | Wellcome Sanger Institute for the COVID-19 Genomics UK Consortium |
| EPI_ISL_439906 | Department of Pathology, University of Cambridge | Wellcome Sanger Institute for the COVID-19 Genomics UK Consortium |
| EPI_ISL_439907 | Department of Pathology, University of Cambridge | Wellcome Sanger Institute for the COVID-19 Genomics UK Consortium |
| EPI_ISL_439908 | Department of Pathology, University of Cambridge | Wellcome Sanger Institute for the COVID-19 Genomics UK Consortium |
| EPI_ISL_439909 | Department of Pathology, University of Cambridge | Wellcome Sanger Institute for the COVID-19 Genomics UK Consortium |
| EPI_ISL_439910 | Department of Pathology, University of Cambridge | Wellcome Sanger Institute for the COVID-19 Genomics UK Consortium |
| EPI_ISL_439911 | Department of Pathology, University of Cambridge | Wellcome Sanger Institute for the COVID-19 Genomics UK Consortium |

| EPI_Accession_ID | Originating Laboratory | Submitting Laboratory |
|---|---|---|
| EPI_ISL_439912 | Department of Pathology, University of Cambridge | Wellcome Sanger Institute for the COVID-19 Genomics UK Consortium |
| EPI_ISL_439913 | Department of Pathology, University of Cambridge | Wellcome Sanger Institute for the COVID-19 Genomics UK Consortium |
| EPI_ISL_439914 | Department of Pathology, University of Cambridge | Wellcome Sanger Institute for the COVID-19 Genomics UK Consortium |
| EPI_ISL_439915 | Department of Pathology, University of Cambridge | Wellcome Sanger Institute for the COVID-19 Genomics UK Consortium |
| EPI_ISL_439916 | Department of Pathology, University of Cambridge | Wellcome Sanger Institute for the COVID-19 Genomics UK Consortium |
| EPI_ISL_439917 | Department of Pathology, University of Cambridge | Wellcome Sanger Institute for the COVID-19 Genomics UK Consortium |
| EPI_ISL_439918 | Department of Pathology, University of Cambridge | Wellcome Sanger Institute for the COVID-19 Genomics UK Consortium |
| EPI_ISL_439919 | Department of Pathology, University of Cambridge | Wellcome Sanger Institute for the COVID-19 Genomics UK Consortium |
| EPI_ISL_439920 | Department of Pathology, University of Cambridge | Wellcome Sanger Institute for the COVID-19 Genomics UK Consortium |
| EPI_ISL_439921 | Department of Pathology, University of Cambridge | Wellcome Sanger Institute for the COVID-19 Genomics UK Consortium |
| EPI_ISL_439922 | Department of Pathology, University of Cambridge | Wellcome Sanger Institute for the COVID-19 Genomics UK Consortium |
| EPI_ISL_439923 | Department of Pathology, University of Cambridge | Wellcome Sanger Institute for the COVID-19 Genomics UK Consortium |
| EPI_ISL_439924 | Department of Pathology, University of Cambridge | Wellcome Sanger Institute for the COVID-19 Genomics UK Consortium |
| EPI_ISL_439925 | Department of Pathology, University of Cambridge | Wellcome Sanger Institute for the COVID-19 Genomics UK Consortium |
| EPI_ISL_439926 | Department of Pathology, University of Cambridge | Wellcome Sanger Institute for the COVID-19 Genomics UK Consortium |
| EPI_ISL_439927 | Department of Pathology, University of Cambridge | Wellcome Sanger Institute for the COVID-19 Genomics UK Consortium |

| EPI_Accession_ID | Originating Laboratory | Submitting Laboratory |
|---|---|---|
| EPI_ISL_439928 | Department of Pathology, University of Cambridge | Wellcome Sanger Institute for the COVID-19 Genomics UK Consortium |
| EPI_ISL_439929 | Department of Pathology, University of Cambridge | Wellcome Sanger Institute for the COVID-19 Genomics UK Consortium |
| EPI_ISL_439930 | Department of Pathology, University of Cambridge | Wellcome Sanger Institute for the COVID-19 Genomics UK Consortium |
| EPI_ISL_439931 | Department of Pathology, University of Cambridge | Wellcome Sanger Institute for the COVID-19 Genomics UK Consortium |
| EPI_ISL_439932 | Department of Pathology, University of Cambridge | Wellcome Sanger Institute for the COVID-19 Genomics UK Consortium |
| EPI_ISL_439933 | Department of Pathology, University of Cambridge | Wellcome Sanger Institute for the COVID-19 Genomics UK Consortium |
| EPI_ISL_439934 | Department of Pathology, University of Cambridge | Wellcome Sanger Institute for the COVID-19 Genomics UK Consortium |
| EPI_ISL_439935 | Department of Pathology, University of Cambridge | Wellcome Sanger Institute for the COVID-19 Genomics UK Consortium |
| EPI_ISL_439936 | Department of Pathology, University of Cambridge | Wellcome Sanger Institute for the COVID-19 Genomics UK Consortium |
| EPI_ISL_439937 | Department of Pathology, University of Cambridge | Wellcome Sanger Institute for the COVID-19 Genomics UK Consortium |
| EPI_ISL_439938 | Department of Pathology, University of Cambridge | Wellcome Sanger Institute for the COVID-19 Genomics UK Consortium |
| EPI_ISL_439939 | Department of Pathology, University of Cambridge | Wellcome Sanger Institute for the COVID-19 Genomics UK Consortium |
| EPI_ISL_439940 | Department of Pathology, University of Cambridge | Wellcome Sanger Institute for the COVID-19 Genomics UK Consortium |
| EPI_ISL_439941 | Department of Pathology, University of Cambridge | Wellcome Sanger Institute for the COVID-19 Genomics UK Consortium |
| EPI_ISL_439942 | Department of Pathology, University of Cambridge | Wellcome Sanger Institute for the COVID-19 Genomics UK Consortium |
| EPI_ISL_439943 | Department of Pathology, University of Cambridge | Wellcome Sanger Institute for the COVID-19 Genomics UK Consortium |

| EPI_Accession_ID | Originating Laboratory | Submitting Laboratory |
|---|---|---|
| EPI_ISL_439944 | Department of Pathology, University of Cambridge | Wellcome Sanger Institute for the COVID-19 Genomics UK Consortium |
| EPI_ISL_439945 | Department of Pathology, University of Cambridge | Wellcome Sanger Institute for the COVID-19 Genomics UK Consortium |
| EPI_ISL_439946 | Department of Pathology, University of Cambridge | Wellcome Sanger Institute for the COVID-19 Genomics UK Consortium |
| EPI_ISL_439947 | Department of Pathology, University of Cambridge | Wellcome Sanger Institute for the COVID-19 Genomics UK Consortium |
| EPI_ISL_439948 | Department of Pathology, University of Cambridge | Wellcome Sanger Institute for the COVID-19 Genomics UK Consortium |
| EPI_ISL_439949 | Department of Pathology, University of Cambridge | Wellcome Sanger Institute for the COVID-19 Genomics UK Consortium |
| EPI_ISL_439950 | Department of Pathology, University of Cambridge | Wellcome Sanger Institute for the COVID-19 Genomics UK Consortium |
| EPI_ISL_439951 | Department of Pathology, University of Cambridge | Wellcome Sanger Institute for the COVID-19 Genomics UK Consortium |
| EPI_ISL_439952 | Department of Pathology, University of Cambridge | Wellcome Sanger Institute for the COVID-19 Genomics UK Consortium |
| EPI_ISL_439953 | PHE South West Regional Laboratory, National Infection Service | Wellcome Sanger Institute for the COVID-19 Genomics UK Consortium |
| EPI_ISL_439954 | Department of Pathology, University of Cambridge | Wellcome Sanger Institute for the COVID-19 Genomics UK Consortium |
| EPI_ISL_439955 | Department of Pathology, University of Cambridge | Wellcome Sanger Institute for the COVID-19 Genomics UK Consortium |
| EPI_ISL_439956 | Department of Pathology, University of Cambridge | Wellcome Sanger Institute for the COVID-19 Genomics UK Consortium |
| EPI_ISL_439957 | PHE South West Regional Laboratory, National Infection Service | Wellcome Sanger Institute for the COVID-19 Genomics UK Consortium |
| EPI_ISL_439958 | Department of Pathology, University of Cambridge | Wellcome Sanger Institute for the COVID-19 Genomics UK Consortium |
| EPI_ISL_439959 | PHE South West Regional Laboratory, National Infection Service | Wellcome Sanger Institute for the COVID-19 Genomics UK Consortium |

| EPI_Accession_ID | Originating Laboratory | Submitting Laboratory |
|---|---|---|
| EPI_ISL_439960 | PHE South West Regional Laboratory, National Infection Service | Wellcome Sanger Institute for the COVID-19 Genomics UK Consortium |
| EPI_ISL_439961 | Department of Pathology, University of Cambridge | Wellcome Sanger Institute for the COVID-19 Genomics UK Consortium |
| EPI_ISL_439962 | PHE South West Regional Laboratory, National Infection Service | Wellcome Sanger Institute for the COVID-19 Genomics UK Consortium |
| EPI_ISL_439963 | Department of Pathology, University of Cambridge | Wellcome Sanger Institute for the COVID-19 Genomics UK Consortium |
| EPI_ISL_439964 | PHE South West Regional Laboratory, National Infection Service | Wellcome Sanger Institute for the COVID-19 Genomics UK Consortium |
| EPI_ISL_439965 | PHE South West Regional Laboratory, National Infection Service | Wellcome Sanger Institute for the COVID-19 Genomics UK Consortium |
| EPI_ISL_439966 | PHE South West Regional Laboratory, National Infection Service | Wellcome Sanger Institute for the COVID-19 Genomics UK Consortium |
| EPI_ISL_439967 | PHE South West Regional Laboratory, National Infection Service | Wellcome Sanger Institute for the COVID-19 Genomics UK Consortium |
| EPI_ISL_439968 | Department of Pathology, University of Cambridge | Wellcome Sanger Institute for the COVID-19 Genomics UK Consortium |
| EPI_ISL_439969 | PHE South West Regional Laboratory, National Infection Service | Wellcome Sanger Institute for the COVID-19 Genomics UK Consortium |
| EPI_ISL_439970 | Department of Pathology, University of Cambridge | Wellcome Sanger Institute for the COVID-19 Genomics UK Consortium |
| EPI_ISL_439971 | PHE South West Regional Laboratory, National Infection Service | Wellcome Sanger Institute for the COVID-19 Genomics UK Consortium |
| EPI_ISL_439972 | PHE South West Regional Laboratory, National Infection Service | Wellcome Sanger Institute for the COVID-19 Genomics UK Consortium |
| EPI_ISL_439973 | PHE South West Regional Laboratory, National Infection Service | Wellcome Sanger Institute for the COVID-19 Genomics UK Consortium |
| EPI_ISL_439974 | PHE South West Regional Laboratory, National Infection Service | Wellcome Sanger Institute for the COVID-19 Genomics UK Consortium |
| EPI_ISL_439975 | Department of Pathology, University of Cambridge | Wellcome Sanger Institute for the COVID-19 Genomics UK Consortium |

| EPI_Accession_ID | Originating Laboratory | Submitting Laboratory |
|---|---|---|
| EPI_ISL_439976 | PHE South West Regional Laboratory, National Infection Service | Wellcome Sanger Institute for the COVID-19 Genomics UK Consortium |
| EPI_ISL_439977 | Department of Pathology, University of Cambridge | Wellcome Sanger Institute for the COVID-19 Genomics UK Consortium |
| EPI_ISL_439978 | PHE South West Regional Laboratory, National Infection Service | Wellcome Sanger Institute for the COVID-19 Genomics UK Consortium |
| EPI_ISL_439979 | PHE South West Regional Laboratory, National Infection Service | Wellcome Sanger Institute for the COVID-19 Genomics UK Consortium |
| EPI_ISL_439980 | Department of Pathology, University of Cambridge | Wellcome Sanger Institute for the COVID-19 Genomics UK Consortium |
| EPI_ISL_439981 | PHE South West Regional Laboratory, National Infection Service | Wellcome Sanger Institute for the COVID-19 Genomics UK Consortium |
| EPI_ISL_439982 | PHE South West Regional Laboratory, National Infection Service | Wellcome Sanger Institute for the COVID-19 Genomics UK Consortium |
| EPI_ISL_439983 | PHE South West Regional Laboratory, National Infection Service | Wellcome Sanger Institute for the COVID-19 Genomics UK Consortium |
| EPI_ISL_439984 | PHE South West Regional Laboratory, National Infection Service | Wellcome Sanger Institute for the COVID-19 Genomics UK Consortium |
| EPI_ISL_439985 | PHE South West Regional Laboratory, National Infection Service | Wellcome Sanger Institute for the COVID-19 Genomics UK Consortium |
| EPI_ISL_439986 | PHE South West Regional Laboratory, National Infection Service | Wellcome Sanger Institute for the COVID-19 Genomics UK Consortium |
| EPI_ISL_439987 | Department of Pathology, University of Cambridge | Wellcome Sanger Institute for the COVID-19 Genomics UK Consortium |
| EPI_ISL_439988 | PHE South West Regional Laboratory, National Infection Service | Wellcome Sanger Institute for the COVID-19 Genomics UK Consortium |
| EPI_ISL_439989 | Department of Pathology, University of Cambridge | Wellcome Sanger Institute for the COVID-19 Genomics UK Consortium |
| EPI_ISL_439990 | Department of Pathology, University of Cambridge | Wellcome Sanger Institute for the COVID-19 Genomics UK Consortium |
| EPI_ISL_439991 | Department of Pathology, University of Cambridge | Wellcome Sanger Institute for the COVID-19 Genomics UK Consortium |

| EPI_Accession_ID | Originating Laboratory | Submitting Laboratory |
| --- | --- | --- |
| EPI_ISL_439992 | PHE South West Regional Laboratory, National Infection Service | Wellcome Sanger Institute for the COVID-19 Genomics UK Consortium |
| EPI_ISL_439993 | Department of Pathology, University of Cambridge | Wellcome Sanger Institute for the COVID-19 Genomics UK Consortium |
| EPI_ISL_439994 | Department of Pathology, University of Cambridge | Wellcome Sanger Institute for the COVID-19 Genomics UK Consortium |
| EPI_ISL_439995 | Department of Pathology, University of Cambridge | Wellcome Sanger Institute for the COVID-19 Genomics UK Consortium |
| EPI_ISL_439996 | Department of Pathology, University of Cambridge | Wellcome Sanger Institute for the COVID-19 Genomics UK Consortium |
| EPI_ISL_439997 | Department of Pathology, University of Cambridge | Wellcome Sanger Institute for the COVID-19 Genomics UK Consortium |
| EPI_ISL_439998 | PHE South West Regional Laboratory, National Infection Service | Wellcome Sanger Institute for the COVID-19 Genomics UK Consortium |
| EPI_ISL_439999 | Department of Pathology, University of Cambridge | Wellcome Sanger Institute for the COVID-19 Genomics UK Consortium |
| EPI_ISL_440000 | PHE South West Regional Laboratory, National Infection Service | Wellcome Sanger Institute for the COVID-19 Genomics UK Consortium |
| EPI_ISL_440001 | PHE South West Regional Laboratory, National Infection Service | Wellcome Sanger Institute for the COVID-19 Genomics UK Consortium |
| EPI_ISL_440002 | PHE South West Regional Laboratory, National Infection Service | Wellcome Sanger Institute for the COVID-19 Genomics UK Consortium |
| EPI_ISL_440003 | Department of Pathology, University of Cambridge | Wellcome Sanger Institute for the COVID-19 Genomics UK Consortium |
| EPI_ISL_440004 | Department of Pathology, University of Cambridge | Wellcome Sanger Institute for the COVID-19 Genomics UK Consortium |
| EPI_ISL_440005 | PHE South West Regional Laboratory, National Infection Service | Wellcome Sanger Institute for the COVID-19 Genomics UK Consortium |
| EPI_ISL_440006 | PHE South West Regional Laboratory, National Infection Service | Wellcome Sanger Institute for the COVID-19 Genomics UK Consortium |
| EPI_ISL_440007 | Department of Pathology, University of Cambridge | Wellcome Sanger Institute for the COVID-19 Genomics UK Consortium |

| EPI_Accession_ID | Originating Laboratory | Submitting Laboratory |
|---|---|---|
| EPI_ISL_440008 | PHE South West Regional Laboratory, National Infection Service | Wellcome Sanger Institute for the COVID-19 Genomics UK Consortium |
| EPI_ISL_440009 | Department of Pathology, University of Cambridge | Wellcome Sanger Institute for the COVID-19 Genomics UK Consortium |
| EPI_ISL_440010 | Department of Pathology, University of Cambridge | Wellcome Sanger Institute for the COVID-19 Genomics UK Consortium |
| EPI_ISL_440011 | Department of Pathology, University of Cambridge | Wellcome Sanger Institute for the COVID-19 Genomics UK Consortium |
| EPI_ISL_440012 | Department of Pathology, University of Cambridge | Wellcome Sanger Institute for the COVID-19 Genomics UK Consortium |
| EPI_ISL_440013 | PHE South West Regional Laboratory, National Infection Service | Wellcome Sanger Institute for the COVID-19 Genomics UK Consortium |
| EPI_ISL_440014 | Department of Pathology, University of Cambridge | Wellcome Sanger Institute for the COVID-19 Genomics UK Consortium |
| EPI_ISL_440015 | Department of Pathology, University of Cambridge | Wellcome Sanger Institute for the COVID-19 Genomics UK Consortium |
| EPI_ISL_440016 | PHE South West Regional Laboratory, National Infection Service | Wellcome Sanger Institute for the COVID-19 Genomics UK Consortium |
| EPI_ISL_440017 | Department of Pathology, University of Cambridge | Wellcome Sanger Institute for the COVID-19 Genomics UK Consortium |
| EPI_ISL_440018 | PHE South West Regional Laboratory, National Infection Service | Wellcome Sanger Institute for the COVID-19 Genomics UK Consortium |
| EPI_ISL_440019 | Department of Pathology, University of Cambridge | Wellcome Sanger Institute for the COVID-19 Genomics UK Consortium |
| EPI_ISL_440020 | PHE South West Regional Laboratory, National Infection Service | Wellcome Sanger Institute for the COVID-19 Genomics UK Consortium |
| EPI_ISL_440021 | PHE South West Regional Laboratory, National Infection Service | Wellcome Sanger Institute for the COVID-19 Genomics UK Consortium |
| EPI_ISL_440022 | PHE South West Regional Laboratory, National Infection Service | Wellcome Sanger Institute for the COVID-19 Genomics UK Consortium |
| EPI_ISL_440023 | PHE South West Regional Laboratory, National Infection Service | Wellcome Sanger Institute for the COVID-19 Genomics UK Consortium |

| EPI_Accession_ID | Originating Laboratory | Submitting Laboratory |
|---|---|---|
| EPI_ISL_440024 | Department of Pathology, University of Cambridge | Wellcome Sanger Institute for the COVID-19 Genomics UK Consortium |
| EPI_ISL_440025 | Department of Pathology, University of Cambridge | Wellcome Sanger Institute for the COVID-19 Genomics UK Consortium |
| EPI_ISL_440026 | PHE South West Regional Laboratory, National Infection Service | Wellcome Sanger Institute for the COVID-19 Genomics UK Consortium |
| EPI_ISL_440027 | PHE South West Regional Laboratory, National Infection Service | Wellcome Sanger Institute for the COVID-19 Genomics UK Consortium |
| EPI_ISL_440028 | PHE South West Regional Laboratory, National Infection Service | Wellcome Sanger Institute for the COVID-19 Genomics UK Consortium |
| EPI_ISL_440029 | PHE South West Regional Laboratory, National Infection Service | Wellcome Sanger Institute for the COVID-19 Genomics UK Consortium |
| EPI_ISL_440030 | Department of Pathology, University of Cambridge | Wellcome Sanger Institute for the COVID-19 Genomics UK Consortium |
| EPI_ISL_440031 | PHE South West Regional Laboratory, National Infection Service | Wellcome Sanger Institute for the COVID-19 Genomics UK Consortium |
| EPI_ISL_440032 | PHE South West Regional Laboratory, National Infection Service | Wellcome Sanger Institute for the COVID-19 Genomics UK Consortium |
| EPI_ISL_440033 | PHE South West Regional Laboratory, National Infection Service | Wellcome Sanger Institute for the COVID-19 Genomics UK Consortium |
| EPI_ISL_440034 | Department of Pathology, University of Cambridge | Wellcome Sanger Institute for the COVID-19 Genomics UK Consortium |
| EPI_ISL_440035 | PHE South West Regional Laboratory, National Infection Service | Wellcome Sanger Institute for the COVID-19 Genomics UK Consortium |
| EPI_ISL_440036 | PHE South West Regional Laboratory, National Infection Service | Wellcome Sanger Institute for the COVID-19 Genomics UK Consortium |
| EPI_ISL_440037 | PHE South West Regional Laboratory, National Infection Service | Wellcome Sanger Institute for the COVID-19 Genomics UK Consortium |
| EPI_ISL_440038 | PHE South West Regional Laboratory, National Infection Service | Wellcome Sanger Institute for the COVID-19 Genomics UK Consortium |
| EPI_ISL_440039 | PHE South West Regional Laboratory, National Infection Service | Wellcome Sanger Institute for the COVID-19 Genomics UK Consortium |

| EPI_Accession_ID | Originating Laboratory | Submitting Laboratory |
|---|---|---|
| EPI_ISL_440040 | Department of Pathology, University of Cambridge | Wellcome Sanger Institute for the COVID-19 Genomics UK Consortium |
| EPI_ISL_440041 | Department of Pathology, University of Cambridge | Wellcome Sanger Institute for the COVID-19 Genomics UK Consortium |
| EPI_ISL_440042 | PHE South West Regional Laboratory, National Infection Service | Wellcome Sanger Institute for the COVID-19 Genomics UK Consortium |
| EPI_ISL_440043 | PHE South West Regional Laboratory, National Infection Service | Wellcome Sanger Institute for the COVID-19 Genomics UK Consortium |
| EPI_ISL_440044 | Department of Pathology, University of Cambridge | Wellcome Sanger Institute for the COVID-19 Genomics UK Consortium |
| EPI_ISL_440045 | Department of Pathology, University of Cambridge | Wellcome Sanger Institute for the COVID-19 Genomics UK Consortium |
| EPI_ISL_440046 | Department of Pathology, University of Cambridge | Wellcome Sanger Institute for the COVID-19 Genomics UK Consortium |
| EPI_ISL_440047 | Department of Pathology, University of Cambridge | Wellcome Sanger Institute for the COVID-19 Genomics UK Consortium |
| EPI_ISL_440048 | Department of Pathology, University of Cambridge | Wellcome Sanger Institute for the COVID-19 Genomics UK Consortium |
| EPI_ISL_440049 | Department of Pathology, University of Cambridge | Wellcome Sanger Institute for the COVID-19 Genomics UK Consortium |
| EPI_ISL_440050 | PHE South West Regional Laboratory, National Infection Service | Wellcome Sanger Institute for the COVID-19 Genomics UK Consortium |
| EPI_ISL_440051 | PHE South West Regional Laboratory, National Infection Service | Wellcome Sanger Institute for the COVID-19 Genomics UK Consortium |
| EPI_ISL_440052 | PHE South West Regional Laboratory, National Infection Service | Wellcome Sanger Institute for the COVID-19 Genomics UK Consortium |
| EPI_ISL_440053 | Department of Pathology, University of Cambridge | Wellcome Sanger Institute for the COVID-19 Genomics UK Consortium |
| EPI_ISL_440054 | PHE South West Regional Laboratory, National Infection Service | Wellcome Sanger Institute for the COVID-19 Genomics UK Consortium |
| EPI_ISL_440055 | Department of Pathology, University of Cambridge | Wellcome Sanger Institute for the COVID-19 Genomics UK Consortium |

| EPI_Accession_ID | Originating Laboratory | Submitting Laboratory |
|---|---|---|
| EPI_ISL_440056 | PHE South West Regional Laboratory, National Infection Service | Wellcome Sanger Institute for the COVID-19 Genomics UK Consortium |
| EPI_ISL_440057 | Department of Pathology, University of Cambridge | Wellcome Sanger Institute for the COVID-19 Genomics UK Consortium |
| EPI_ISL_440058 | PHE South West Regional Laboratory, National Infection Service | Wellcome Sanger Institute for the COVID-19 Genomics UK Consortium |
| EPI_ISL_440059 | Department of Pathology, University of Cambridge | Wellcome Sanger Institute for the COVID-19 Genomics UK Consortium |
| EPI_ISL_440060 | Department of Pathology, University of Cambridge | Wellcome Sanger Institute for the COVID-19 Genomics UK Consortium |
| EPI_ISL_440061 | Department of Pathology, University of Cambridge | Wellcome Sanger Institute for the COVID-19 Genomics UK Consortium |
| EPI_ISL_440062 | Department of Pathology, University of Cambridge | Wellcome Sanger Institute for the COVID-19 Genomics UK Consortium |
| EPI_ISL_440063 | PHE South West Regional Laboratory, National Infection Service | Wellcome Sanger Institute for the COVID-19 Genomics UK Consortium |
| EPI_ISL_440064 | Department of Pathology, University of Cambridge | Wellcome Sanger Institute for the COVID-19 Genomics UK Consortium |
| EPI_ISL_440065 | PHE South West Regional Laboratory, National Infection Service | Wellcome Sanger Institute for the COVID-19 Genomics UK Consortium |
| EPI_ISL_440066 | Department of Pathology, University of Cambridge | Wellcome Sanger Institute for the COVID-19 Genomics UK Consortium |
| EPI_ISL_440067 | Department of Pathology, University of Cambridge | Wellcome Sanger Institute for the COVID-19 Genomics UK Consortium |
| EPI_ISL_440068 | PHE South West Regional Laboratory, National Infection Service | Wellcome Sanger Institute for the COVID-19 Genomics UK Consortium |
| EPI_ISL_440069 | Department of Pathology, University of Cambridge | Wellcome Sanger Institute for the COVID-19 Genomics UK Consortium |
| EPI_ISL_440070 | PHE South West Regional Laboratory, National Infection Service | Wellcome Sanger Institute for the COVID-19 Genomics UK Consortium |
| EPI_ISL_440071 | PHE South West Regional Laboratory, National Infection Service | Wellcome Sanger Institute for the COVID-19 Genomics UK Consortium |

| EPI_Accession_ID | Originating Laboratory | Submitting Laboratory |
|---|---|---|
| EPI_ISL_440072 | Department of Pathology, University of Cambridge | Wellcome Sanger Institute for the COVID-19 Genomics UK Consortium |
| EPI_ISL_440073 | Department of Pathology, University of Cambridge | Wellcome Sanger Institute for the COVID-19 Genomics UK Consortium |
| EPI_ISL_440074 | Department of Pathology, University of Cambridge | Wellcome Sanger Institute for the COVID-19 Genomics UK Consortium |
| EPI_ISL_440075 | Department of Pathology, University of Cambridge | Wellcome Sanger Institute for the COVID-19 Genomics UK Consortium |
| EPI_ISL_440076 | Department of Pathology, University of Cambridge | Wellcome Sanger Institute for the COVID-19 Genomics UK Consortium |
| EPI_ISL_440077 | Department of Pathology, University of Cambridge | Wellcome Sanger Institute for the COVID-19 Genomics UK Consortium |
| EPI_ISL_440078 | Department of Pathology, University of Cambridge | Wellcome Sanger Institute for the COVID-19 Genomics UK Consortium |
| EPI_ISL_440079 | Department of Pathology, University of Cambridge | Wellcome Sanger Institute for the COVID-19 Genomics UK Consortium |
| EPI_ISL_440080 | Department of Pathology, University of Cambridge | Wellcome Sanger Institute for the COVID-19 Genomics UK Consortium |
| EPI_ISL_440081 | PHE South West Regional Laboratory, National Infection Service | Wellcome Sanger Institute for the COVID-19 Genomics UK Consortium |
| EPI_ISL_440082 | Department of Pathology, University of Cambridge | Wellcome Sanger Institute for the COVID-19 Genomics UK Consortium |
| EPI_ISL_440083 | Department of Pathology, University of Cambridge | Wellcome Sanger Institute for the COVID-19 Genomics UK Consortium |
| EPI_ISL_440084 | PHE South West Regional Laboratory, National Infection Service | Wellcome Sanger Institute for the COVID-19 Genomics UK Consortium |
| EPI_ISL_440085 | Department of Pathology, University of Cambridge | Wellcome Sanger Institute for the COVID-19 Genomics UK Consortium |
| EPI_ISL_440086 | Department of Pathology, University of Cambridge | Wellcome Sanger Institute for the COVID-19 Genomics UK Consortium |
| EPI_ISL_440087 | PHE South West Regional Laboratory, National Infection Service | Wellcome Sanger Institute for the COVID-19 Genomics UK Consortium |

| EPI_Accession_ID | Originating Laboratory | Submitting Laboratory |
|---|---|---|
| EPI_ISL_440088 | Department of Pathology, University of Cambridge | Wellcome Sanger Institute for the COVID-19 Genomics UK Consortium |
| EPI_ISL_440089 | Department of Pathology, University of Cambridge | Wellcome Sanger Institute for the COVID-19 Genomics UK Consortium |
| EPI_ISL_440090 | Department of Pathology, University of Cambridge | Wellcome Sanger Institute for the COVID-19 Genomics UK Consortium |
| EPI_ISL_440091 | PHE South West Regional Laboratory, National Infection Service | Wellcome Sanger Institute for the COVID-19 Genomics UK Consortium |
| EPI_ISL_440092 | PHE South West Regional Laboratory, National Infection Service | Wellcome Sanger Institute for the COVID-19 Genomics UK Consortium |
| EPI_ISL_440093 | PHE South West Regional Laboratory, National Infection Service | Wellcome Sanger Institute for the COVID-19 Genomics UK Consortium |
| EPI_ISL_440094 | PHE South West Regional Laboratory, National Infection Service | Wellcome Sanger Institute for the COVID-19 Genomics UK Consortium |
| EPI_ISL_440095 | PHE South West Regional Laboratory, National Infection Service | Wellcome Sanger Institute for the COVID-19 Genomics UK Consortium |
| EPI_ISL_440096 | Department of Pathology, University of Cambridge | Wellcome Sanger Institute for the COVID-19 Genomics UK Consortium |
| EPI_ISL_440097 | Department of Pathology, University of Cambridge | Wellcome Sanger Institute for the COVID-19 Genomics UK Consortium |
| EPI_ISL_440098 | PHE South West Regional Laboratory, National Infection Service | Wellcome Sanger Institute for the COVID-19 Genomics UK Consortium |
| EPI_ISL_440099 | Department of Pathology, University of Cambridge | Wellcome Sanger Institute for the COVID-19 Genomics UK Consortium |
| EPI_ISL_440100 | PHE South West Regional Laboratory, National Infection Service | Wellcome Sanger Institute for the COVID-19 Genomics UK Consortium |
| EPI_ISL_440101 | PHE South West Regional Laboratory, National Infection Service | Wellcome Sanger Institute for the COVID-19 Genomics UK Consortium |
| EPI_ISL_440102 | PHE South West Regional Laboratory, National Infection Service | Wellcome Sanger Institute for the COVID-19 Genomics UK Consortium |
| EPI_ISL_440103 | PHE South West Regional Laboratory, National Infection Service | Wellcome Sanger Institute for the COVID-19 Genomics UK Consortium |

| EPI_Accession_ID | Originating Laboratory | Submitting Laboratory |
| --- | --- | --- |
| EPI_ISL_440104 | Department of Pathology, University of Cambridge | Wellcome Sanger Institute for the COVID-19 Genomics UK Consortium |
| EPI_ISL_440105 | PHE South West Regional Laboratory, National Infection Service | Wellcome Sanger Institute for the COVID-19 Genomics UK Consortium |
| EPI_ISL_440106 | PHE South West Regional Laboratory, National Infection Service | Wellcome Sanger Institute for the COVID-19 Genomics UK Consortium |
| EPI_ISL_440107 | Department of Pathology, University of Cambridge | Wellcome Sanger Institute for the COVID-19 Genomics UK Consortium |
| EPI_ISL_440108 | Department of Pathology, University of Cambridge | Wellcome Sanger Institute for the COVID-19 Genomics UK Consortium |
| EPI_ISL_440109 | Department of Pathology, University of Cambridge | Wellcome Sanger Institute for the COVID-19 Genomics UK Consortium |
| EPI_ISL_440110 | Department of Pathology, University of Cambridge | Wellcome Sanger Institute for the COVID-19 Genomics UK Consortium |
| EPI_ISL_440111 | Department of Pathology, University of Cambridge | Wellcome Sanger Institute for the COVID-19 Genomics UK Consortium |
| EPI_ISL_440112 | Department of Pathology, University of Cambridge | Wellcome Sanger Institute for the COVID-19 Genomics UK Consortium |
| EPI_ISL_440113 | PHE South West Regional Laboratory, National Infection Service | Wellcome Sanger Institute for the COVID-19 Genomics UK Consortium |
| EPI_ISL_440114 | PHE South West Regional Laboratory, National Infection Service | Wellcome Sanger Institute for the COVID-19 Genomics UK Consortium |
| EPI_ISL_440115 | PHE South West Regional Laboratory, National Infection Service | Wellcome Sanger Institute for the COVID-19 Genomics UK Consortium |
| EPI_ISL_440116 | PHE South West Regional Laboratory, National Infection Service | Wellcome Sanger Institute for the COVID-19 Genomics UK Consortium |
| EPI_ISL_440117 | Department of Pathology, University of Cambridge | Wellcome Sanger Institute for the COVID-19 Genomics UK Consortium |
| EPI_ISL_440118 | PHE South West Regional Laboratory, National Infection Service | Wellcome Sanger Institute for the COVID-19 Genomics UK Consortium |
| EPI_ISL_440119 | PHE South West Regional Laboratory, National Infection Service | Wellcome Sanger Institute for the COVID-19 Genomics UK Consortium |

| EPI_Accession_ID | Originating Laboratory | Submitting Laboratory |
|---|---|---|
| EPI_ISL_440120 | Department of Pathology, University of Cambridge | Wellcome Sanger Institute for the COVID-19 Genomics UK Consortium |
| EPI_ISL_440121 | Department of Pathology, University of Cambridge | Wellcome Sanger Institute for the COVID-19 Genomics UK Consortium |
| EPI_ISL_440122 | Department of Pathology, University of Cambridge | Wellcome Sanger Institute for the COVID-19 Genomics UK Consortium |
| EPI_ISL_440123 | PHE South West Regional Laboratory, National Infection Service | Wellcome Sanger Institute for the COVID-19 Genomics UK Consortium |
| EPI_ISL_440124 | PHE South West Regional Laboratory, National Infection Service | Wellcome Sanger Institute for the COVID-19 Genomics UK Consortium |
| EPI_ISL_440125 | Department of Pathology, University of Cambridge | Wellcome Sanger Institute for the COVID-19 Genomics UK Consortium |
| EPI_ISL_440126 | PHE South West Regional Laboratory, National Infection Service | Wellcome Sanger Institute for the COVID-19 Genomics UK Consortium |
| EPI_ISL_440127 | PHE South West Regional Laboratory, National Infection Service | Wellcome Sanger Institute for the COVID-19 Genomics UK Consortium |
| EPI_ISL_440128 | PHE South West Regional Laboratory, National Infection Service | Wellcome Sanger Institute for the COVID-19 Genomics UK Consortium |
| EPI_ISL_440129 | Department of Pathology, University of Cambridge | Wellcome Sanger Institute for the COVID-19 Genomics UK Consortium |
| EPI_ISL_440130 | PHE South West Regional Laboratory, National Infection Service | Wellcome Sanger Institute for the COVID-19 Genomics UK Consortium |
| EPI_ISL_440131 | PHE South West Regional Laboratory, National Infection Service | Wellcome Sanger Institute for the COVID-19 Genomics UK Consortium |
| EPI_ISL_440132 | Department of Pathology, University of Cambridge | Wellcome Sanger Institute for the COVID-19 Genomics UK Consortium |
| EPI_ISL_440133 | Department of Pathology, University of Cambridge | Wellcome Sanger Institute for the COVID-19 Genomics UK Consortium |
| EPI_ISL_440134 | PHE South West Regional Laboratory, National Infection Service | Wellcome Sanger Institute for the COVID-19 Genomics UK Consortium |
| EPI_ISL_440135 | PHE South West Regional Laboratory, National Infection Service | Wellcome Sanger Institute for the COVID-19 Genomics UK Consortium |

| EPI_Accession_ID | Originating Laboratory | Submitting Laboratory |
|---|---|---|
| EPI_ISL_440136 | PHE South West Regional Laboratory, National Infection Service | Wellcome Sanger Institute for the COVID-19 Genomics UK Consortium |
| EPI_ISL_440137 | PHE South West Regional Laboratory, National Infection Service | Wellcome Sanger Institute for the COVID-19 Genomics UK Consortium |
| EPI_ISL_440138 | Department of Pathology, University of Cambridge | Wellcome Sanger Institute for the COVID-19 Genomics UK Consortium |
| EPI_ISL_440139 | PHE South West Regional Laboratory, National Infection Service | Wellcome Sanger Institute for the COVID-19 Genomics UK Consortium |
| EPI_ISL_440140 | PHE South West Regional Laboratory, National Infection Service | Wellcome Sanger Institute for the COVID-19 Genomics UK Consortium |
| EPI_ISL_440141 | PHE South West Regional Laboratory, National Infection Service | Wellcome Sanger Institute for the COVID-19 Genomics UK Consortium |
| EPI_ISL_440142 | PHE South West Regional Laboratory, National Infection Service | Wellcome Sanger Institute for the COVID-19 Genomics UK Consortium |
| EPI_ISL_440143 | Department of Pathology, University of Cambridge | Wellcome Sanger Institute for the COVID-19 Genomics UK Consortium |
| EPI_ISL_440144 | Department of Pathology, University of Cambridge | Wellcome Sanger Institute for the COVID-19 Genomics UK Consortium |
| EPI_ISL_440145 | PHE South West Regional Laboratory, National Infection Service | Wellcome Sanger Institute for the COVID-19 Genomics UK Consortium |
| EPI_ISL_440146 | Department of Pathology, University of Cambridge | Wellcome Sanger Institute for the COVID-19 Genomics UK Consortium |
| EPI_ISL_440147 | Department of Pathology, University of Cambridge | Wellcome Sanger Institute for the COVID-19 Genomics UK Consortium |
| EPI_ISL_440148 | PHE South West Regional Laboratory, National Infection Service | Wellcome Sanger Institute for the COVID-19 Genomics UK Consortium |
| EPI_ISL_440149 | PHE South West Regional Laboratory, National Infection Service | Wellcome Sanger Institute for the COVID-19 Genomics UK Consortium |
| EPI_ISL_440150 | Department of Pathology, University of Cambridge | Wellcome Sanger Institute for the COVID-19 Genomics UK Consortium |
| EPI_ISL_440151 | PHE South West Regional Laboratory, National Infection Service | Wellcome Sanger Institute for the COVID-19 Genomics UK Consortium |

| EPI_Accession_ID | Originating Laboratory | Submitting Laboratory |
|---|---|---|
| EPI_ISL_440152 | Department of Pathology, University of Cambridge | Wellcome Sanger Institute for the COVID-19 Genomics UK Consortium |
| EPI_ISL_440153 | PHE South West Regional Laboratory, National Infection Service | Wellcome Sanger Institute for the COVID-19 Genomics UK Consortium |
| EPI_ISL_440154 | PHE South West Regional Laboratory, National Infection Service | Wellcome Sanger Institute for the COVID-19 Genomics UK Consortium |
| EPI_ISL_440155 | Department of Pathology, University of Cambridge | Wellcome Sanger Institute for the COVID-19 Genomics UK Consortium |
| EPI_ISL_440156 | PHE South West Regional Laboratory, National Infection Service | Wellcome Sanger Institute for the COVID-19 Genomics UK Consortium |
| EPI_ISL_440157 | PHE South West Regional Laboratory, National Infection Service | Wellcome Sanger Institute for the COVID-19 Genomics UK Consortium |
| EPI_ISL_440158 | PHE South West Regional Laboratory, National Infection Service | Wellcome Sanger Institute for the COVID-19 Genomics UK Consortium |
| EPI_ISL_440159 | Department of Pathology, University of Cambridge | Wellcome Sanger Institute for the COVID-19 Genomics UK Consortium |
| EPI_ISL_440160 | PHE South West Regional Laboratory, National Infection Service | Wellcome Sanger Institute for the COVID-19 Genomics UK Consortium |
| EPI_ISL_440161 | Department of Pathology, University of Cambridge | Wellcome Sanger Institute for the COVID-19 Genomics UK Consortium |
| EPI_ISL_440162 | PHE South West Regional Laboratory, National Infection Service | Wellcome Sanger Institute for the COVID-19 Genomics UK Consortium |
| EPI_ISL_440163 | Department of Pathology, University of Cambridge | Wellcome Sanger Institute for the COVID-19 Genomics UK Consortium |
| EPI_ISL_440164 | Department of Pathology, University of Cambridge | Wellcome Sanger Institute for the COVID-19 Genomics UK Consortium |
| EPI_ISL_440165 | PHE South West Regional Laboratory, National Infection Service | Wellcome Sanger Institute for the COVID-19 Genomics UK Consortium |
| EPI_ISL_440166 | Department of Pathology, University of Cambridge | Wellcome Sanger Institute for the COVID-19 Genomics UK Consortium |
| EPI_ISL_440167 | PHE South West Regional Laboratory, National Infection Service | Wellcome Sanger Institute for the COVID-19 Genomics UK Consortium |

| EPI_Accession_ID | Originating Laboratory | Submitting Laboratory |
|---|---|---|
| EPI_ISL_440168 | PHE South West Regional Laboratory, National Infection Service | Wellcome Sanger Institute for the COVID-19 Genomics UK Consortium |
| EPI_ISL_440169 | Department of Pathology, University of Cambridge | Wellcome Sanger Institute for the COVID-19 Genomics UK Consortium |
| EPI_ISL_440170 | Department of Pathology, University of Cambridge | Wellcome Sanger Institute for the COVID-19 Genomics UK Consortium |
| EPI_ISL_440171 | Department of Pathology, University of Cambridge | Wellcome Sanger Institute for the COVID-19 Genomics UK Consortium |
| EPI_ISL_440172 | PHE South West Regional Laboratory, National Infection Service | Wellcome Sanger Institute for the COVID-19 Genomics UK Consortium |
| EPI_ISL_440173 | Department of Pathology, University of Cambridge | Wellcome Sanger Institute for the COVID-19 Genomics UK Consortium |
| EPI_ISL_440174 | PHE South West Regional Laboratory, National Infection Service | Wellcome Sanger Institute for the COVID-19 Genomics UK Consortium |
| EPI_ISL_440175 | PHE South West Regional Laboratory, National Infection Service | Wellcome Sanger Institute for the COVID-19 Genomics UK Consortium |
| EPI_ISL_440176 | Department of Pathology, University of Cambridge | Wellcome Sanger Institute for the COVID-19 Genomics UK Consortium |
| EPI_ISL_440177 | Department of Pathology, University of Cambridge | Wellcome Sanger Institute for the COVID-19 Genomics UK Consortium |
| EPI_ISL_440178 | PHE South West Regional Laboratory, National Infection Service | Wellcome Sanger Institute for the COVID-19 Genomics UK Consortium |
| EPI_ISL_440179 | Department of Pathology, University of Cambridge | Wellcome Sanger Institute for the COVID-19 Genomics UK Consortium |
| EPI_ISL_440180 | Department of Pathology, University of Cambridge | Wellcome Sanger Institute for the COVID-19 Genomics UK Consortium |
| EPI_ISL_440181 | Department of Pathology, University of Cambridge | Wellcome Sanger Institute for the COVID-19 Genomics UK Consortium |
| EPI_ISL_440182 | Department of Pathology, University of Cambridge | Wellcome Sanger Institute for the COVID-19 Genomics UK Consortium |
| EPI_ISL_440183 | Department of Pathology, University of Cambridge | Wellcome Sanger Institute for the COVID-19 Genomics UK Consortium |

| EPI_Accession_ID | Originating Laboratory | Submitting Laboratory |
|---|---|---|
| EPI_ISL_440184 | Department of Pathology, University of Cambridge | Wellcome Sanger Institute for the COVID-19 Genomics UK Consortium |
| EPI_ISL_440185 | Department of Pathology, University of Cambridge | Wellcome Sanger Institute for the COVID-19 Genomics UK Consortium |
| EPI_ISL_440186 | Department of Pathology, University of Cambridge | Wellcome Sanger Institute for the COVID-19 Genomics UK Consortium |
| EPI_ISL_440187 | PHE South West Regional Laboratory, National Infection Service | Wellcome Sanger Institute for the COVID-19 Genomics UK Consortium |
| EPI_ISL_440188 | Department of Pathology, University of Cambridge | Wellcome Sanger Institute for the COVID-19 Genomics UK Consortium |
| EPI_ISL_440189 | PHE South West Regional Laboratory, National Infection Service | Wellcome Sanger Institute for the COVID-19 Genomics UK Consortium |
| EPI_ISL_440190 | PHE South West Regional Laboratory, National Infection Service | Wellcome Sanger Institute for the COVID-19 Genomics UK Consortium |
| EPI_ISL_440191 | PHE South West Regional Laboratory, National Infection Service | Wellcome Sanger Institute for the COVID-19 Genomics UK Consortium |
| EPI_ISL_440192 | PHE South West Regional Laboratory, National Infection Service | Wellcome Sanger Institute for the COVID-19 Genomics UK Consortium |
| EPI_ISL_440193 | Department of Pathology, University of Cambridge | Wellcome Sanger Institute for the COVID-19 Genomics UK Consortium |
| EPI_ISL_440194 | PHE South West Regional Laboratory, National Infection Service | Wellcome Sanger Institute for the COVID-19 Genomics UK Consortium |
| EPI_ISL_440195 | Department of Pathology, University of Cambridge | Wellcome Sanger Institute for the COVID-19 Genomics UK Consortium |
| EPI_ISL_440196 | PHE South West Regional Laboratory, National Infection Service | Wellcome Sanger Institute for the COVID-19 Genomics UK Consortium |
| EPI_ISL_440197 | Department of Pathology, University of Cambridge | Wellcome Sanger Institute for the COVID-19 Genomics UK Consortium |
| EPI_ISL_440198 | PHE South West Regional Laboratory, National Infection Service | Wellcome Sanger Institute for the COVID-19 Genomics UK Consortium |
| EPI_ISL_440199 | PHE South West Regional Laboratory, National Infection Service | Wellcome Sanger Institute for the COVID-19 Genomics UK Consortium |

| EPI_Accession_ID | Originating Laboratory | Submitting Laboratory |
|---|---|---|
| EPI_ISL_440200 | PHE South West Regional Laboratory, National Infection Service | Wellcome Sanger Institute for the COVID-19 Genomics UK Consortium |
| EPI_ISL_440201 | PHE South West Regional Laboratory, National Infection Service | Wellcome Sanger Institute for the COVID-19 Genomics UK Consortium |
| EPI_ISL_440202 | Department of Pathology, University of Cambridge | Wellcome Sanger Institute for the COVID-19 Genomics UK Consortium |
| EPI_ISL_440203 | Department of Pathology, University of Cambridge | Wellcome Sanger Institute for the COVID-19 Genomics UK Consortium |
| EPI_ISL_440204 | PHE South West Regional Laboratory, National Infection Service | Wellcome Sanger Institute for the COVID-19 Genomics UK Consortium |
| EPI_ISL_440205 | PHE South West Regional Laboratory, National Infection Service | Wellcome Sanger Institute for the COVID-19 Genomics UK Consortium |
| EPI_ISL_440206 | PHE South West Regional Laboratory, National Infection Service | Wellcome Sanger Institute for the COVID-19 Genomics UK Consortium |
| EPI_ISL_440207 | Department of Pathology, University of Cambridge | Wellcome Sanger Institute for the COVID-19 Genomics UK Consortium |
| EPI_ISL_440208 | Department of Pathology, University of Cambridge | Wellcome Sanger Institute for the COVID-19 Genomics UK Consortium |
| EPI_ISL_440209 | Department of Pathology, University of Cambridge | Wellcome Sanger Institute for the COVID-19 Genomics UK Consortium |
| EPI_ISL_440210 | PHE South West Regional Laboratory, National Infection Service | Wellcome Sanger Institute for the COVID-19 Genomics UK Consortium |
| EPI_ISL_440211 | Department of Pathology, University of Cambridge | Wellcome Sanger Institute for the COVID-19 Genomics UK Consortium |
| EPI_ISL_440212 | PHE South West Regional Laboratory, National Infection Service | Wellcome Sanger Institute for the COVID-19 Genomics UK Consortium |
| EPI_ISL_440213 | Department of Pathology, University of Cambridge | Wellcome Sanger Institute for the COVID-19 Genomics UK Consortium |
| EPI_ISL_440214 | PHE South West Regional Laboratory, National Infection Service | Wellcome Sanger Institute for the COVID-19 Genomics UK Consortium |
| EPI_ISL_440215 | PHE South West Regional Laboratory, National Infection Service | Wellcome Sanger Institute for the COVID-19 Genomics UK Consortium |

| EPI_Accession_ID | Originating Laboratory | Submitting Laboratory |
|---|---|---|
| EPI_ISL_440216 | PHE South West Regional Laboratory, National Infection Service | Wellcome Sanger Institute for the COVID-19 Genomics UK Consortium |
| EPI_ISL_440217 | Department of Pathology, University of Cambridge | Wellcome Sanger Institute for the COVID-19 Genomics UK Consortium |
| EPI_ISL_440218 | PHE South West Regional Laboratory, National Infection Service | Wellcome Sanger Institute for the COVID-19 Genomics UK Consortium |
| EPI_ISL_440219 | PHE South West Regional Laboratory, National Infection Service | Wellcome Sanger Institute for the COVID-19 Genomics UK Consortium |
| EPI_ISL_440220 | PHE South West Regional Laboratory, National Infection Service | Wellcome Sanger Institute for the COVID-19 Genomics UK Consortium |
| EPI_ISL_440221 | PHE South West Regional Laboratory, National Infection Service | Wellcome Sanger Institute for the COVID-19 Genomics UK Consortium |
| EPI_ISL_440222 | PHE South West Regional Laboratory, National Infection Service | Wellcome Sanger Institute for the COVID-19 Genomics UK Consortium |
| EPI_ISL_440223 | Department of Pathology, University of Cambridge | Wellcome Sanger Institute for the COVID-19 Genomics UK Consortium |
| EPI_ISL_440224 | PHE South West Regional Laboratory, National Infection Service | Wellcome Sanger Institute for the COVID-19 Genomics UK Consortium |
| EPI_ISL_440225 | PHE South West Regional Laboratory, National Infection Service | Wellcome Sanger Institute for the COVID-19 Genomics UK Consortium |
| EPI_ISL_440226 | Department of Pathology, University of Cambridge | Wellcome Sanger Institute for the COVID-19 Genomics UK Consortium |
| EPI_ISL_440227 | PHE South West Regional Laboratory, National Infection Service | Wellcome Sanger Institute for the COVID-19 Genomics UK Consortium |
| EPI_ISL_440228 | PHE South West Regional Laboratory, National Infection Service | Wellcome Sanger Institute for the COVID-19 Genomics UK Consortium |
| EPI_ISL_440229 | PHE South West Regional Laboratory, National Infection Service | Wellcome Sanger Institute for the COVID-19 Genomics UK Consortium |
| EPI_ISL_440230 | Department of Pathology, University of Cambridge | Wellcome Sanger Institute for the COVID-19 Genomics UK Consortium |
| EPI_ISL_440231 | Department of Pathology, University of Cambridge | Wellcome Sanger Institute for the COVID-19 Genomics UK Consortium |

| EPI_Accession_ID | Originating Laboratory | Submitting Laboratory |
|---|---|---|
| EPI_ISL_440232 | PHE South West Regional Laboratory, National Infection Service | Wellcome Sanger Institute for the COVID-19 Genomics UK Consortium |
| EPI_ISL_440233 | PHE South West Regional Laboratory, National Infection Service | Wellcome Sanger Institute for the COVID-19 Genomics UK Consortium |
| EPI_ISL_440234 | Department of Pathology, University of Cambridge | Wellcome Sanger Institute for the COVID-19 Genomics UK Consortium |
| EPI_ISL_440235 | PHE South West Regional Laboratory, National Infection Service | Wellcome Sanger Institute for the COVID-19 Genomics UK Consortium |
| EPI_ISL_440236 | PHE South West Regional Laboratory, National Infection Service | Wellcome Sanger Institute for the COVID-19 Genomics UK Consortium |
| EPI_ISL_440237 | Department of Pathology, University of Cambridge | Wellcome Sanger Institute for the COVID-19 Genomics UK Consortium |
| EPI_ISL_440238 | PHE South West Regional Laboratory, National Infection Service | Wellcome Sanger Institute for the COVID-19 Genomics UK Consortium |
| EPI_ISL_440239 | Department of Pathology, University of Cambridge | Wellcome Sanger Institute for the COVID-19 Genomics UK Consortium |
| EPI_ISL_440240 | Department of Pathology, University of Cambridge | Wellcome Sanger Institute for the COVID-19 Genomics UK Consortium |
| EPI_ISL_440241 | PHE South West Regional Laboratory, National Infection Service | Wellcome Sanger Institute for the COVID-19 Genomics UK Consortium |
| EPI_ISL_440242 | PHE South West Regional Laboratory, National Infection Service | Wellcome Sanger Institute for the COVID-19 Genomics UK Consortium |
| EPI_ISL_440243 | PHE South West Regional Laboratory, National Infection Service | Wellcome Sanger Institute for the COVID-19 Genomics UK Consortium |
| EPI_ISL_440244 | Department of Pathology, University of Cambridge | Wellcome Sanger Institute for the COVID-19 Genomics UK Consortium |
| EPI_ISL_440245 | PHE South West Regional Laboratory, National Infection Service | Wellcome Sanger Institute for the COVID-19 Genomics UK Consortium |
| EPI_ISL_440246 | Department of Pathology, University of Cambridge | Wellcome Sanger Institute for the COVID-19 Genomics UK Consortium |
| EPI_ISL_440247 | Department of Pathology, University of Cambridge | Wellcome Sanger Institute for the COVID-19 Genomics UK Consortium |

| EPI_Accession_ID | Originating Laboratory | Submitting Laboratory |
|---|---|---|
| EPI_ISL_440248 | PHE South West Regional Laboratory, National Infection Service | Wellcome Sanger Institute for the COVID-19 Genomics UK Consortium |
| EPI_ISL_440249 | PHE South West Regional Laboratory, National Infection Service | Wellcome Sanger Institute for the COVID-19 Genomics UK Consortium |
| EPI_ISL_440250 | PHE South West Regional Laboratory, National Infection Service | Wellcome Sanger Institute for the COVID-19 Genomics UK Consortium |
| EPI_ISL_440251 | PHE South West Regional Laboratory, National Infection Service | Wellcome Sanger Institute for the COVID-19 Genomics UK Consortium |
| EPI_ISL_440252 | PHE South West Regional Laboratory, National Infection Service | Wellcome Sanger Institute for the COVID-19 Genomics UK Consortium |
| EPI_ISL_440253 | PHE South West Regional Laboratory, National Infection Service | Wellcome Sanger Institute for the COVID-19 Genomics UK Consortium |
| EPI_ISL_440254 | Department of Pathology, University of Cambridge | Wellcome Sanger Institute for the COVID-19 Genomics UK Consortium |
| EPI_ISL_440255 | PHE South West Regional Laboratory, National Infection Service | Wellcome Sanger Institute for the COVID-19 Genomics UK Consortium |
| EPI_ISL_440256 | PHE South West Regional Laboratory, National Infection Service | Wellcome Sanger Institute for the COVID-19 Genomics UK Consortium |
| EPI_ISL_440257 | Department of Pathology, University of Cambridge | Wellcome Sanger Institute for the COVID-19 Genomics UK Consortium |
| EPI_ISL_440258 | Department of Pathology, University of Cambridge | Wellcome Sanger Institute for the COVID-19 Genomics UK Consortium |
| EPI_ISL_440259 | Department of Pathology, University of Cambridge | Wellcome Sanger Institute for the COVID-19 Genomics UK Consortium |
| EPI_ISL_440260 | Department of Pathology, University of Cambridge | Wellcome Sanger Institute for the COVID-19 Genomics UK Consortium |
| EPI_ISL_440261 | Department of Pathology, University of Cambridge | Wellcome Sanger Institute for the COVID-19 Genomics UK Consortium |
| EPI_ISL_440262 | PHE South West Regional Laboratory, National Infection Service | Wellcome Sanger Institute for the COVID-19 Genomics UK Consortium |
| EPI_ISL_440263 | Department of Pathology, University of Cambridge | Wellcome Sanger Institute for the COVID-19 Genomics UK Consortium |

| EPI_Accession_ID | Originating Laboratory | Submitting Laboratory |
|---|---|---|
| EPI_ISL_440264 | Department of Pathology, University of Cambridge | Wellcome Sanger Institute for the COVID-19 Genomics UK Consortium |
| EPI_ISL_440265 | Department of Pathology, University of Cambridge | Wellcome Sanger Institute for the COVID-19 Genomics UK Consortium |
| EPI_ISL_440266 | PHE South West Regional Laboratory, National Infection Service | Wellcome Sanger Institute for the COVID-19 Genomics UK Consortium |
| EPI_ISL_440267 | Department of Pathology, University of Cambridge | Wellcome Sanger Institute for the COVID-19 Genomics UK Consortium |
| EPI_ISL_440268 | Department of Pathology, University of Cambridge | Wellcome Sanger Institute for the COVID-19 Genomics UK Consortium |
| EPI_ISL_440269 | PHE South West Regional Laboratory, National Infection Service | Wellcome Sanger Institute for the COVID-19 Genomics UK Consortium |
| EPI_ISL_440270 | Department of Pathology, University of Cambridge | Wellcome Sanger Institute for the COVID-19 Genomics UK Consortium |
| EPI_ISL_440271 | Department of Pathology, University of Cambridge | Wellcome Sanger Institute for the COVID-19 Genomics UK Consortium |
| EPI_ISL_440272 | PHE South West Regional Laboratory, National Infection Service | Wellcome Sanger Institute for the COVID-19 Genomics UK Consortium |
| EPI_ISL_440273 | PHE South West Regional Laboratory, National Infection Service | Wellcome Sanger Institute for the COVID-19 Genomics UK Consortium |
| EPI_ISL_440274 | PHE South West Regional Laboratory, National Infection Service | Wellcome Sanger Institute for the COVID-19 Genomics UK Consortium |
| EPI_ISL_440275 | PHE South West Regional Laboratory, National Infection Service | Wellcome Sanger Institute for the COVID-19 Genomics UK Consortium |
| EPI_ISL_440276 | PHE South West Regional Laboratory, National Infection Service | Wellcome Sanger Institute for the COVID-19 Genomics UK Consortium |
| EPI_ISL_440277 | PHE South West Regional Laboratory, National Infection Service | Wellcome Sanger Institute for the COVID-19 Genomics UK Consortium |
| EPI_ISL_440278 | Department of Pathology, University of Cambridge | Wellcome Sanger Institute for the COVID-19 Genomics UK Consortium |
| EPI_ISL_440279 | Department of Pathology, University of Cambridge | Wellcome Sanger Institute for the COVID-19 Genomics UK Consortium |

| EPI_Accession_ID | Originating Laboratory | Submitting Laboratory |
|---|---|---|
| EPI_ISL_440280 | Department of Pathology, University of Cambridge | Wellcome Sanger Institute for the COVID-19 Genomics UK Consortium |
| EPI_ISL_440281 | Department of Pathology, University of Cambridge | Wellcome Sanger Institute for the COVID-19 Genomics UK Consortium |
| EPI_ISL_440282 | Department of Pathology, University of Cambridge | Wellcome Sanger Institute for the COVID-19 Genomics UK Consortium |
| EPI_ISL_440283 | Department of Pathology, University of Cambridge | Wellcome Sanger Institute for the COVID-19 Genomics UK Consortium |
| EPI_ISL_440284 | Department of Pathology, University of Cambridge | Wellcome Sanger Institute for the COVID-19 Genomics UK Consortium |
| EPI_ISL_440285 | Department of Pathology, University of Cambridge | Wellcome Sanger Institute for the COVID-19 Genomics UK Consortium |
| EPI_ISL_440286 | PHE South West Regional Laboratory, National Infection Service | Wellcome Sanger Institute for the COVID-19 Genomics UK Consortium |
| EPI_ISL_440287 | Department of Pathology, University of Cambridge | Wellcome Sanger Institute for the COVID-19 Genomics UK Consortium |
| EPI_ISL_440288 | Department of Pathology, University of Cambridge | Wellcome Sanger Institute for the COVID-19 Genomics UK Consortium |
| EPI_ISL_440289 | Department of Pathology, University of Cambridge | Wellcome Sanger Institute for the COVID-19 Genomics UK Consortium |
| EPI_ISL_440290 | Department of Pathology, University of Cambridge | Wellcome Sanger Institute for the COVID-19 Genomics UK Consortium |
| EPI_ISL_440291 | PHE South West Regional Laboratory, National Infection Service | Wellcome Sanger Institute for the COVID-19 Genomics UK Consortium |
| EPI_ISL_440292 | PHE South West Regional Laboratory, National Infection Service | Wellcome Sanger Institute for the COVID-19 Genomics UK Consortium |
| EPI_ISL_440293 | Department of Pathology, University of Cambridge | Wellcome Sanger Institute for the COVID-19 Genomics UK Consortium |
| EPI_ISL_440294 | PHE South West Regional Laboratory, National Infection Service | Wellcome Sanger Institute for the COVID-19 Genomics UK Consortium |
| EPI_ISL_440295 | Department of Pathology, University of Cambridge | Wellcome Sanger Institute for the COVID-19 Genomics UK Consortium |

| EPI_Accession_ID | Originating Laboratory | Submitting Laboratory |
|---|---|---|
| EPI_ISL_440296 | Department of Pathology, University of Cambridge | Wellcome Sanger Institute for the COVID-19 Genomics UK Consortium |
| EPI_ISL_440297 | Department of Pathology, University of Cambridge | Wellcome Sanger Institute for the COVID-19 Genomics UK Consortium |
| EPI_ISL_440298 | PHE South West Regional Laboratory, National Infection Service | Wellcome Sanger Institute for the COVID-19 Genomics UK Consortium |
| EPI_ISL_440299 | PHE South West Regional Laboratory, National Infection Service | Wellcome Sanger Institute for the COVID-19 Genomics UK Consortium |
| EPI_ISL_440300 | Department of Pathology, University of Cambridge | Wellcome Sanger Institute for the COVID-19 Genomics UK Consortium |
| EPI_ISL_440301 | Department of Pathology, University of Cambridge | Wellcome Sanger Institute for the COVID-19 Genomics UK Consortium |
| EPI_ISL_440302 | Department of Pathology, University of Cambridge | Wellcome Sanger Institute for the COVID-19 Genomics UK Consortium |
| EPI_ISL_440303 | Department of Pathology, University of Cambridge | Wellcome Sanger Institute for the COVID-19 Genomics UK Consortium |
| EPI_ISL_440304 | Department of Pathology, University of Cambridge | Wellcome Sanger Institute for the COVID-19 Genomics UK Consortium |
| EPI_ISL_440305 | Department of Pathology, University of Cambridge | Wellcome Sanger Institute for the COVID-19 Genomics UK Consortium |
| EPI_ISL_440306 | Department of Pathology, University of Cambridge | Wellcome Sanger Institute for the COVID-19 Genomics UK Consortium |
| EPI_ISL_440307 | Department of Pathology, University of Cambridge | Wellcome Sanger Institute for the COVID-19 Genomics UK Consortium |
| EPI_ISL_440308 | Department of Pathology, University of Cambridge | Wellcome Sanger Institute for the COVID-19 Genomics UK Consortium |
| EPI_ISL_440309 | Department of Pathology, University of Cambridge | Wellcome Sanger Institute for the COVID-19 Genomics UK Consortium |
| EPI_ISL_440310 | Department of Pathology, University of Cambridge | Wellcome Sanger Institute for the COVID-19 Genomics UK Consortium |
| EPI_ISL_440311 | Department of Pathology, University of Cambridge | Wellcome Sanger Institute for the COVID-19 Genomics UK Consortium |

| EPI_Accession_ID | Originating Laboratory | Submitting Laboratory |
|---|---|---|
| EPI_ISL_440312 | Department of Pathology, University of Cambridge | Wellcome Sanger Institute for the COVID-19 Genomics UK Consortium |
| EPI_ISL_440313 | Department of Pathology, University of Cambridge | Wellcome Sanger Institute for the COVID-19 Genomics UK Consortium |
| EPI_ISL_440314 | Department of Pathology, University of Cambridge | Wellcome Sanger Institute for the COVID-19 Genomics UK Consortium |
| EPI_ISL_440315 | Department of Pathology, University of Cambridge | Wellcome Sanger Institute for the COVID-19 Genomics UK Consortium |
| EPI_ISL_440316 | Department of Pathology, University of Cambridge | Wellcome Sanger Institute for the COVID-19 Genomics UK Consortium |
| EPI_ISL_440317 | Department of Pathology, University of Cambridge | Wellcome Sanger Institute for the COVID-19 Genomics UK Consortium |
| EPI_ISL_440318 | Department of Pathology, University of Cambridge | Wellcome Sanger Institute for the COVID-19 Genomics UK Consortium |
| EPI_ISL_440319 | Department of Pathology, University of Cambridge | Wellcome Sanger Institute for the COVID-19 Genomics UK Consortium |
| EPI_ISL_440320 | Department of Pathology, University of Cambridge | Wellcome Sanger Institute for the COVID-19 Genomics UK Consortium |
| EPI_ISL_440321 | Department of Pathology, University of Cambridge | Wellcome Sanger Institute for the COVID-19 Genomics UK Consortium |
| EPI_ISL_440322 | Department of Pathology, University of Cambridge | Wellcome Sanger Institute for the COVID-19 Genomics UK Consortium |
| EPI_ISL_440323 | Department of Pathology, University of Cambridge | Wellcome Sanger Institute for the COVID-19 Genomics UK Consortium |
| EPI_ISL_440324 | Department of Pathology, University of Cambridge | Wellcome Sanger Institute for the COVID-19 Genomics UK Consortium |
| EPI_ISL_440325 | Department of Pathology, University of Cambridge | Wellcome Sanger Institute for the COVID-19 Genomics UK Consortium |
| EPI_ISL_440326 | Department of Pathology, University of Cambridge | Wellcome Sanger Institute for the COVID-19 Genomics UK Consortium |
| EPI_ISL_440327 | Department of Pathology, University of Cambridge | Wellcome Sanger Institute for the COVID-19 Genomics UK Consortium |

| EPI_Accession_ID | Originating Laboratory | Submitting Laboratory |
|---|---|---|
| EPI_ISL_440328 | Department of Pathology, University of Cambridge | Wellcome Sanger Institute for the COVID-19 Genomics UK Consortium |
| EPI_ISL_440329 | Department of Pathology, University of Cambridge | Wellcome Sanger Institute for the COVID-19 Genomics UK Consortium |
| EPI_ISL_440330 | Department of Pathology, University of Cambridge | Wellcome Sanger Institute for the COVID-19 Genomics UK Consortium |
| EPI_ISL_440331 | Department of Pathology, University of Cambridge | Wellcome Sanger Institute for the COVID-19 Genomics UK Consortium |
| EPI_ISL_440332 | Department of Pathology, University of Cambridge | Wellcome Sanger Institute for the COVID-19 Genomics UK Consortium |
| EPI_ISL_440333 | Department of Pathology, University of Cambridge | Wellcome Sanger Institute for the COVID-19 Genomics UK Consortium |
| EPI_ISL_440334 | Department of Pathology, University of Cambridge | Wellcome Sanger Institute for the COVID-19 Genomics UK Consortium |
| EPI_ISL_440335 | Department of Pathology, University of Cambridge | Wellcome Sanger Institute for the COVID-19 Genomics UK Consortium |
| EPI_ISL_440336 | Department of Pathology, University of Cambridge | Wellcome Sanger Institute for the COVID-19 Genomics UK Consortium |
| EPI_ISL_440337 | Department of Pathology, University of Cambridge | Wellcome Sanger Institute for the COVID-19 Genomics UK Consortium |
| EPI_ISL_440338 | Department of Pathology, University of Cambridge | Wellcome Sanger Institute for the COVID-19 Genomics UK Consortium |
| EPI_ISL_440339 | Department of Pathology, University of Cambridge | Wellcome Sanger Institute for the COVID-19 Genomics UK Consortium |
| EPI_ISL_440340 | Department of Pathology, University of Cambridge | Wellcome Sanger Institute for the COVID-19 Genomics UK Consortium |
| EPI_ISL_440341 | Department of Pathology, University of Cambridge | Wellcome Sanger Institute for the COVID-19 Genomics UK Consortium |
| EPI_ISL_440342 | Department of Pathology, University of Cambridge | Wellcome Sanger Institute for the COVID-19 Genomics UK Consortium |
| EPI_ISL_440343 | Department of Pathology, University of Cambridge | Wellcome Sanger Institute for the COVID-19 Genomics UK Consortium |

| EPI_Accession_ID | Originating Laboratory | Submitting Laboratory |
|---|---|---|
| EPI_ISL_440344 | Department of Pathology, University of Cambridge | Wellcome Sanger Institute for the COVID-19 Genomics UK Consortium |
| EPI_ISL_440345 | Department of Pathology, University of Cambridge | Wellcome Sanger Institute for the COVID-19 Genomics UK Consortium |
| EPI_ISL_440346 | Department of Pathology, University of Cambridge | Wellcome Sanger Institute for the COVID-19 Genomics UK Consortium |
| EPI_ISL_440347 | Department of Pathology, University of Cambridge | Wellcome Sanger Institute for the COVID-19 Genomics UK Consortium |
| EPI_ISL_440348 | Department of Pathology, University of Cambridge | Wellcome Sanger Institute for the COVID-19 Genomics UK Consortium |
| EPI_ISL_440349 | Department of Pathology, University of Cambridge | Wellcome Sanger Institute for the COVID-19 Genomics UK Consortium |
| EPI_ISL_440350 | Department of Pathology, University of Cambridge | Wellcome Sanger Institute for the COVID-19 Genomics UK Consortium |
| EPI_ISL_440351 | Department of Pathology, University of Cambridge | Wellcome Sanger Institute for the COVID-19 Genomics UK Consortium |
| EPI_ISL_440352 | Department of Pathology, University of Cambridge | Wellcome Sanger Institute for the COVID-19 Genomics UK Consortium |
| EPI_ISL_440353 | Department of Pathology, University of Cambridge | Wellcome Sanger Institute for the COVID-19 Genomics UK Consortium |
| EPI_ISL_440354 | Department of Pathology, University of Cambridge | Wellcome Sanger Institute for the COVID-19 Genomics UK Consortium |
| EPI_ISL_440355 | Department of Pathology, University of Cambridge | Wellcome Sanger Institute for the COVID-19 Genomics UK Consortium |
| EPI_ISL_440356 | Department of Pathology, University of Cambridge | Wellcome Sanger Institute for the COVID-19 Genomics UK Consortium |
| EPI_ISL_440357 | Department of Pathology, University of Cambridge | Wellcome Sanger Institute for the COVID-19 Genomics UK Consortium |
| EPI_ISL_440358 | Department of Pathology, University of Cambridge | Wellcome Sanger Institute for the COVID-19 Genomics UK Consortium |
| EPI_ISL_440359 | Department of Pathology, University of Cambridge | Wellcome Sanger Institute for the COVID-19 Genomics UK Consortium |

| EPI_Accession_ID | Originating Laboratory | Submitting Laboratory |
|---|---|---|
| EPI_ISL_440360 | Department of Pathology, University of Cambridge | Wellcome Sanger Institute for the COVID-19 Genomics UK Consortium |
| EPI_ISL_440361 | Department of Pathology, University of Cambridge | Wellcome Sanger Institute for the COVID-19 Genomics UK Consortium |
| EPI_ISL_440362 | Department of Pathology, University of Cambridge | Wellcome Sanger Institute for the COVID-19 Genomics UK Consortium |
| EPI_ISL_440363 | Department of Pathology, University of Cambridge | Wellcome Sanger Institute for the COVID-19 Genomics UK Consortium |
| EPI_ISL_440364 | Department of Pathology, University of Cambridge | Wellcome Sanger Institute for the COVID-19 Genomics UK Consortium |
| EPI_ISL_440365 | Department of Pathology, University of Cambridge | Wellcome Sanger Institute for the COVID-19 Genomics UK Consortium |
| EPI_ISL_440366 | Department of Pathology, University of Cambridge | Wellcome Sanger Institute for the COVID-19 Genomics UK Consortium |
| EPI_ISL_440367 | Department of Pathology, University of Cambridge | Wellcome Sanger Institute for the COVID-19 Genomics UK Consortium |
| EPI_ISL_440368 | Department of Pathology, University of Cambridge | Wellcome Sanger Institute for the COVID-19 Genomics UK Consortium |
| EPI_ISL_440369 | Department of Pathology, University of Cambridge | Wellcome Sanger Institute for the COVID-19 Genomics UK Consortium |
| EPI_ISL_440370 | Department of Pathology, University of Cambridge | Wellcome Sanger Institute for the COVID-19 Genomics UK Consortium |
| EPI_ISL_440371 | Department of Pathology, University of Cambridge | Wellcome Sanger Institute for the COVID-19 Genomics UK Consortium |
| EPI_ISL_440372 | Department of Pathology, University of Cambridge | Wellcome Sanger Institute for the COVID-19 Genomics UK Consortium |
| EPI_ISL_440373 | Department of Pathology, University of Cambridge | Wellcome Sanger Institute for the COVID-19 Genomics UK Consortium |
| EPI_ISL_440374 | Department of Pathology, University of Cambridge | Wellcome Sanger Institute for the COVID-19 Genomics UK Consortium |
| EPI_ISL_440375 | Department of Pathology, University of Cambridge | Wellcome Sanger Institute for the COVID-19 Genomics UK Consortium |

| EPI_Accession_ID | Originating Laboratory | Submitting Laboratory |
|---|---|---|
| EPI_ISL_440376 | Department of Pathology, University of Cambridge | Wellcome Sanger Institute for the COVID-19 Genomics UK Consortium |
| EPI_ISL_440377 | Department of Pathology, University of Cambridge | Wellcome Sanger Institute for the COVID-19 Genomics UK Consortium |
| EPI_ISL_440378 | Department of Pathology, University of Cambridge | Wellcome Sanger Institute for the COVID-19 Genomics UK Consortium |
| EPI_ISL_440379 | Department of Pathology, University of Cambridge | Wellcome Sanger Institute for the COVID-19 Genomics UK Consortium |
| EPI_ISL_440380 | Department of Pathology, University of Cambridge | Wellcome Sanger Institute for the COVID-19 Genomics UK Consortium |
| EPI_ISL_440381 | Department of Pathology, University of Cambridge | Wellcome Sanger Institute for the COVID-19 Genomics UK Consortium |
| EPI_ISL_440382 | Department of Pathology, University of Cambridge | Wellcome Sanger Institute for the COVID-19 Genomics UK Consortium |
| EPI_ISL_440383 | Department of Pathology, University of Cambridge | Wellcome Sanger Institute for the COVID-19 Genomics UK Consortium |
| EPI_ISL_440384 | Department of Pathology, University of Cambridge | Wellcome Sanger Institute for the COVID-19 Genomics UK Consortium |
| EPI_ISL_440385 | Department of Pathology, University of Cambridge | Wellcome Sanger Institute for the COVID-19 Genomics UK Consortium |
| EPI_ISL_440386 | Department of Pathology, University of Cambridge | Wellcome Sanger Institute for the COVID-19 Genomics UK Consortium |
| EPI_ISL_440387 | Department of Pathology, University of Cambridge | Wellcome Sanger Institute for the COVID-19 Genomics UK Consortium |
| EPI_ISL_440388 | Department of Pathology, University of Cambridge | Wellcome Sanger Institute for the COVID-19 Genomics UK Consortium |
| EPI_ISL_440389 | Department of Pathology, University of Cambridge | Wellcome Sanger Institute for the COVID-19 Genomics UK Consortium |
| EPI_ISL_440390 | Department of Pathology, University of Cambridge | Wellcome Sanger Institute for the COVID-19 Genomics UK Consortium |
| EPI_ISL_440391 | Department of Pathology, University of Cambridge | Wellcome Sanger Institute for the COVID-19 Genomics UK Consortium |

| EPI_Accession_ID | Originating Laboratory | Submitting Laboratory |
|---|---|---|
| EPI_ISL_440392 | Department of Pathology, University of Cambridge | Wellcome Sanger Institute for the COVID-19 Genomics UK Consortium |
| EPI_ISL_440393 | Department of Pathology, University of Cambridge | Wellcome Sanger Institute for the COVID-19 Genomics UK Consortium |
| EPI_ISL_440394 | Department of Pathology, University of Cambridge | Wellcome Sanger Institute for the COVID-19 Genomics UK Consortium |
| EPI_ISL_440395 | Department of Pathology, University of Cambridge | Wellcome Sanger Institute for the COVID-19 Genomics UK Consortium |
| EPI_ISL_440396 | Department of Pathology, University of Cambridge | Wellcome Sanger Institute for the COVID-19 Genomics UK Consortium |
| EPI_ISL_440397 | Department of Pathology, University of Cambridge | Wellcome Sanger Institute for the COVID-19 Genomics UK Consortium |
| EPI_ISL_440398 | Department of Pathology, University of Cambridge | Wellcome Sanger Institute for the COVID-19 Genomics UK Consortium |
| EPI_ISL_440399 | Department of Pathology, University of Cambridge | Wellcome Sanger Institute for the COVID-19 Genomics UK Consortium |
| EPI_ISL_440400 | Department of Pathology, University of Cambridge | Wellcome Sanger Institute for the COVID-19 Genomics UK Consortium |
| EPI_ISL_440401 | Department of Pathology, University of Cambridge | Wellcome Sanger Institute for the COVID-19 Genomics UK Consortium |
| EPI_ISL_440402 | Department of Pathology, University of Cambridge | Wellcome Sanger Institute for the COVID-19 Genomics UK Consortium |
| EPI_ISL_440403 | Department of Pathology, University of Cambridge | Wellcome Sanger Institute for the COVID-19 Genomics UK Consortium |
| EPI_ISL_440404 | Department of Pathology, University of Cambridge | Wellcome Sanger Institute for the COVID-19 Genomics UK Consortium |
| EPI_ISL_440405 | Department of Pathology, University of Cambridge | Wellcome Sanger Institute for the COVID-19 Genomics UK Consortium |
| EPI_ISL_440406 | Department of Pathology, University of Cambridge | Wellcome Sanger Institute for the COVID-19 Genomics UK Consortium |
| EPI_ISL_440407 | Department of Pathology, University of Cambridge | Wellcome Sanger Institute for the COVID-19 Genomics UK Consortium |

| EPI_Accession_ID | Originating Laboratory | Submitting Laboratory |
|---|---|---|
| EPI_ISL_440408 | Department of Pathology, University of Cambridge | Wellcome Sanger Institute for the COVID-19 Genomics UK Consortium |
| EPI_ISL_440409 | Department of Pathology, University of Cambridge | Wellcome Sanger Institute for the COVID-19 Genomics UK Consortium |
| EPI_ISL_440410 | Department of Pathology, University of Cambridge | Wellcome Sanger Institute for the COVID-19 Genomics UK Consortium |
| EPI_ISL_440411 | Department of Pathology, University of Cambridge | Wellcome Sanger Institute for the COVID-19 Genomics UK Consortium |
| EPI_ISL_440412 | Department of Pathology, University of Cambridge | Wellcome Sanger Institute for the COVID-19 Genomics UK Consortium |
| EPI_ISL_440413 | Department of Pathology, University of Cambridge | Wellcome Sanger Institute for the COVID-19 Genomics UK Consortium |
| EPI_ISL_440414 | Department of Pathology, University of Cambridge | Wellcome Sanger Institute for the COVID-19 Genomics UK Consortium |
| EPI_ISL_440415 | Department of Pathology, University of Cambridge | Wellcome Sanger Institute for the COVID-19 Genomics UK Consortium |
| EPI_ISL_440416 | Department of Pathology, University of Cambridge | Wellcome Sanger Institute for the COVID-19 Genomics UK Consortium |
| EPI_ISL_440417 | Department of Pathology, University of Cambridge | Wellcome Sanger Institute for the COVID-19 Genomics UK Consortium |
| EPI_ISL_440418 | Department of Pathology, University of Cambridge | Wellcome Sanger Institute for the COVID-19 Genomics UK Consortium |
| EPI_ISL_440419 | Department of Pathology, University of Cambridge | Wellcome Sanger Institute for the COVID-19 Genomics UK Consortium |
| EPI_ISL_440420 | Department of Pathology, University of Cambridge | Wellcome Sanger Institute for the COVID-19 Genomics UK Consortium |
| EPI_ISL_440421 | Department of Pathology, University of Cambridge | Wellcome Sanger Institute for the COVID-19 Genomics UK Consortium |
| EPI_ISL_440422 | Department of Pathology, University of Cambridge | Wellcome Sanger Institute for the COVID-19 Genomics UK Consortium |
| EPI_ISL_440423 | Department of Pathology, University of Cambridge | Wellcome Sanger Institute for the COVID-19 Genomics UK Consortium |

| EPI_Accession_ID | Originating Laboratory | Submitting Laboratory |
|---|---|---|
| EPI_ISL_440424 | Department of Pathology, University of Cambridge | Wellcome Sanger Institute for the COVID-19 Genomics UK Consortium |
| EPI_ISL_440425 | Department of Pathology, University of Cambridge | Wellcome Sanger Institute for the COVID-19 Genomics UK Consortium |
| EPI_ISL_440426 | Department of Pathology, University of Cambridge | Wellcome Sanger Institute for the COVID-19 Genomics UK Consortium |
| EPI_ISL_440427 | Department of Pathology, University of Cambridge | Wellcome Sanger Institute for the COVID-19 Genomics UK Consortium |
| EPI_ISL_440428 | Department of Pathology, University of Cambridge | Wellcome Sanger Institute for the COVID-19 Genomics UK Consortium |
| EPI_ISL_440429 | Department of Pathology, University of Cambridge | Wellcome Sanger Institute for the COVID-19 Genomics UK Consortium |
| EPI_ISL_440430 | Department of Pathology, University of Cambridge | Wellcome Sanger Institute for the COVID-19 Genomics UK Consortium |
| EPI_ISL_440431 | Department of Pathology, University of Cambridge | Wellcome Sanger Institute for the COVID-19 Genomics UK Consortium |
| EPI_ISL_440432 | Department of Pathology, University of Cambridge | Wellcome Sanger Institute for the COVID-19 Genomics UK Consortium |
| EPI_ISL_440433 | Department of Pathology, University of Cambridge | Wellcome Sanger Institute for the COVID-19 Genomics UK Consortium |
| EPI_ISL_440434 | Department of Pathology, University of Cambridge | Wellcome Sanger Institute for the COVID-19 Genomics UK Consortium |
| EPI_ISL_440435 | Department of Pathology, University of Cambridge | Wellcome Sanger Institute for the COVID-19 Genomics UK Consortium |
| EPI_ISL_440436 | Department of Pathology, University of Cambridge | Wellcome Sanger Institute for the COVID-19 Genomics UK Consortium |
| EPI_ISL_440437 | Department of Pathology, University of Cambridge | Wellcome Sanger Institute for the COVID-19 Genomics UK Consortium |
| EPI_ISL_440438 | Department of Pathology, University of Cambridge | Wellcome Sanger Institute for the COVID-19 Genomics UK Consortium |
| EPI_ISL_440439 | Department of Pathology, University of Cambridge | Wellcome Sanger Institute for the COVID-19 Genomics UK Consortium |

| EPI_Accession_ID | Originating Laboratory | Submitting Laboratory |
|---|---|---|
| EPI_ISL_440440 | Department of Pathology, University of Cambridge | Wellcome Sanger Institute for the COVID-19 Genomics UK Consortium |
| EPI_ISL_440441 | Department of Pathology, University of Cambridge | Wellcome Sanger Institute for the COVID-19 Genomics UK Consortium |
| EPI_ISL_440442 | Department of Pathology, University of Cambridge | Wellcome Sanger Institute for the COVID-19 Genomics UK Consortium |
| EPI_ISL_440443 | Department of Pathology, University of Cambridge | Wellcome Sanger Institute for the COVID-19 Genomics UK Consortium |
| EPI_ISL_440444 | Department of Pathology, University of Cambridge | Wellcome Sanger Institute for the COVID-19 Genomics UK Consortium |
| EPI_ISL_440445 | Department of Pathology, University of Cambridge | Wellcome Sanger Institute for the COVID-19 Genomics UK Consortium |
| EPI_ISL_440446 | Department of Pathology, University of Cambridge | Wellcome Sanger Institute for the COVID-19 Genomics UK Consortium |
| EPI_ISL_440447 | Department of Pathology, University of Cambridge | Wellcome Sanger Institute for the COVID-19 Genomics UK Consortium |
| EPI_ISL_440448 | Department of Pathology, University of Cambridge | Wellcome Sanger Institute for the COVID-19 Genomics UK Consortium |
| EPI_ISL_440449 | Department of Pathology, University of Cambridge | Wellcome Sanger Institute for the COVID-19 Genomics UK Consortium |
| EPI_ISL_440450 | Department of Pathology, University of Cambridge | Wellcome Sanger Institute for the COVID-19 Genomics UK Consortium |
| EPI_ISL_440451 | Department of Pathology, University of Cambridge | Wellcome Sanger Institute for the COVID-19 Genomics UK Consortium |
| EPI_ISL_440452 | Department of Pathology, University of Cambridge | Wellcome Sanger Institute for the COVID-19 Genomics UK Consortium |
| EPI_ISL_440453 | Department of Pathology, University of Cambridge | Wellcome Sanger Institute for the COVID-19 Genomics UK Consortium |
| EPI_ISL_440454 | Department of Pathology, University of Cambridge | Wellcome Sanger Institute for the COVID-19 Genomics UK Consortium |
| EPI_ISL_440455 | Department of Pathology, University of Cambridge | Wellcome Sanger Institute for the COVID-19 Genomics UK Consortium |

| EPI_Accession_ID | Originating Laboratory | Submitting Laboratory |
|---|---|---|
| EPI_ISL_440456 | Department of Pathology, University of Cambridge | Wellcome Sanger Institute for the COVID-19 Genomics UK Consortium |
| EPI_ISL_440457 | Department of Pathology, University of Cambridge | Wellcome Sanger Institute for the COVID-19 Genomics UK Consortium |
| EPI_ISL_440458 | Department of Pathology, University of Cambridge | Wellcome Sanger Institute for the COVID-19 Genomics UK Consortium |
| EPI_ISL_440459 | Department of Pathology, University of Cambridge | Wellcome Sanger Institute for the COVID-19 Genomics UK Consortium |
| EPI_ISL_440460 | Department of Pathology, University of Cambridge | Wellcome Sanger Institute for the COVID-19 Genomics UK Consortium |
| EPI_ISL_440461 | Department of Pathology, University of Cambridge | Wellcome Sanger Institute for the COVID-19 Genomics UK Consortium |
| EPI_ISL_440462 | Department of Pathology, University of Cambridge | Wellcome Sanger Institute for the COVID-19 Genomics UK Consortium |
| EPI_ISL_440463 | Department of Pathology, University of Cambridge | Wellcome Sanger Institute for the COVID-19 Genomics UK Consortium |
| EPI_ISL_440464 | Department of Pathology, University of Cambridge | Wellcome Sanger Institute for the COVID-19 Genomics UK Consortium |
| EPI_ISL_440465 | Department of Pathology, University of Cambridge | Wellcome Sanger Institute for the COVID-19 Genomics UK Consortium |
| EPI_ISL_440466 | Department of Pathology, University of Cambridge | Wellcome Sanger Institute for the COVID-19 Genomics UK Consortium |
| EPI_ISL_440467 | Department of Pathology, University of Cambridge | Wellcome Sanger Institute for the COVID-19 Genomics UK Consortium |
| EPI_ISL_440468 | Department of Pathology, University of Cambridge | Wellcome Sanger Institute for the COVID-19 Genomics UK Consortium |
| EPI_ISL_440469 | Department of Pathology, University of Cambridge | Wellcome Sanger Institute for the COVID-19 Genomics UK Consortium |
| EPI_ISL_440470 | Department of Pathology, University of Cambridge | Wellcome Sanger Institute for the COVID-19 Genomics UK Consortium |
| EPI_ISL_440471 | Department of Pathology, University of Cambridge | Wellcome Sanger Institute for the COVID-19 Genomics UK Consortium |

| EPI_Accession_ID | Originating Laboratory | Submitting Laboratory |
|---|---|---|
| EPI_ISL_440472 | Department of Pathology, University of Cambridge | Wellcome Sanger Institute for the COVID-19 Genomics UK Consortium |
| EPI_ISL_440473 | Department of Pathology, University of Cambridge | Wellcome Sanger Institute for the COVID-19 Genomics UK Consortium |
| EPI_ISL_440474 | Department of Pathology, University of Cambridge | Wellcome Sanger Institute for the COVID-19 Genomics UK Consortium |
| EPI_ISL_440475 | Department of Pathology, University of Cambridge | Wellcome Sanger Institute for the COVID-19 Genomics UK Consortium |
| EPI_ISL_440476 | Department of Pathology, University of Cambridge | Wellcome Sanger Institute for the COVID-19 Genomics UK Consortium |
| EPI_ISL_440477 | Department of Pathology, University of Cambridge | Wellcome Sanger Institute for the COVID-19 Genomics UK Consortium |
| EPI_ISL_440478 | Department of Pathology, University of Cambridge | Wellcome Sanger Institute for the COVID-19 Genomics UK Consortium |
| EPI_ISL_440479 | Department of Pathology, University of Cambridge | Wellcome Sanger Institute for the COVID-19 Genomics UK Consortium |
| EPI_ISL_440480 | Department of Pathology, University of Cambridge | Wellcome Sanger Institute for the COVID-19 Genomics UK Consortium |
| EPI_ISL_440481 | Department of Pathology, University of Cambridge | Wellcome Sanger Institute for the COVID-19 Genomics UK Consortium |
| EPI_ISL_440482 | Department of Pathology, University of Cambridge | Wellcome Sanger Institute for the COVID-19 Genomics UK Consortium |
| EPI_ISL_440483 | Department of Pathology, University of Cambridge | Wellcome Sanger Institute for the COVID-19 Genomics UK Consortium |
| EPI_ISL_440484 | Department of Pathology, University of Cambridge | Wellcome Sanger Institute for the COVID-19 Genomics UK Consortium |
| EPI_ISL_440485 | Department of Pathology, University of Cambridge | Wellcome Sanger Institute for the COVID-19 Genomics UK Consortium |
| EPI_ISL_440486 | Department of Pathology, University of Cambridge | Wellcome Sanger Institute for the COVID-19 Genomics UK Consortium |
| EPI_ISL_440487 | Department of Pathology, University of Cambridge | Wellcome Sanger Institute for the COVID-19 Genomics UK Consortium |

| EPI_Accession_ID | Originating Laboratory | Submitting Laboratory |
|---|---|---|
| EPI_ISL_440488 | Department of Pathology, University of Cambridge | Wellcome Sanger Institute for the COVID-19 Genomics UK Consortium |
| EPI_ISL_440489 | Department of Pathology, University of Cambridge | Wellcome Sanger Institute for the COVID-19 Genomics UK Consortium |
| EPI_ISL_440490 | Department of Pathology, University of Cambridge | Wellcome Sanger Institute for the COVID-19 Genomics UK Consortium |
| EPI_ISL_440491 | Department of Pathology, University of Cambridge | Wellcome Sanger Institute for the COVID-19 Genomics UK Consortium |
| EPI_ISL_440492 | Department of Pathology, University of Cambridge | Wellcome Sanger Institute for the COVID-19 Genomics UK Consortium |
| EPI_ISL_440493 | Department of Pathology, University of Cambridge | Wellcome Sanger Institute for the COVID-19 Genomics UK Consortium |
| EPI_ISL_440494 | Department of Pathology, University of Cambridge | Wellcome Sanger Institute for the COVID-19 Genomics UK Consortium |
| EPI_ISL_440495 | Department of Pathology, University of Cambridge | Wellcome Sanger Institute for the COVID-19 Genomics UK Consortium |
| EPI_ISL_440496 | Department of Pathology, University of Cambridge | Wellcome Sanger Institute for the COVID-19 Genomics UK Consortium |
| EPI_ISL_440497 | Department of Pathology, University of Cambridge | Wellcome Sanger Institute for the COVID-19 Genomics UK Consortium |
| EPI_ISL_440498 | Department of Pathology, University of Cambridge | Wellcome Sanger Institute for the COVID-19 Genomics UK Consortium |
| EPI_ISL_440499 | Department of Pathology, University of Cambridge | Wellcome Sanger Institute for the COVID-19 Genomics UK Consortium |
| EPI_ISL_440500 | Department of Pathology, University of Cambridge | Wellcome Sanger Institute for the COVID-19 Genomics UK Consortium |
| EPI_ISL_440501 | Department of Pathology, University of Cambridge | Wellcome Sanger Institute for the COVID-19 Genomics UK Consortium |
| EPI_ISL_440502 | Department of Pathology, University of Cambridge | Wellcome Sanger Institute for the COVID-19 Genomics UK Consortium |
| EPI_ISL_440503 | Department of Pathology, University of Cambridge | Wellcome Sanger Institute for the COVID-19 Genomics UK Consortium |

| EPI_Accession_ID | Originating Laboratory | Submitting Laboratory |
|---|---|---|
| EPI_ISL_440504 | Department of Pathology, University of Cambridge | Wellcome Sanger Institute for the COVID-19 Genomics UK Consortium |
| EPI_ISL_440505 | Department of Pathology, University of Cambridge | Wellcome Sanger Institute for the COVID-19 Genomics UK Consortium |
| EPI_ISL_440506 | Department of Pathology, University of Cambridge | Wellcome Sanger Institute for the COVID-19 Genomics UK Consortium |
| EPI_ISL_440507 | Department of Pathology, University of Cambridge | Wellcome Sanger Institute for the COVID-19 Genomics UK Consortium |
| EPI_ISL_440508 | Department of Pathology, University of Cambridge | Wellcome Sanger Institute for the COVID-19 Genomics UK Consortium |
| EPI_ISL_440509 | Department of Pathology, University of Cambridge | Wellcome Sanger Institute for the COVID-19 Genomics UK Consortium |
| EPI_ISL_440510 | Department of Pathology, University of Cambridge | Wellcome Sanger Institute for the COVID-19 Genomics UK Consortium |
| EPI_ISL_440511 | Department of Pathology, University of Cambridge | Wellcome Sanger Institute for the COVID-19 Genomics UK Consortium |
| EPI_ISL_440512 | Department of Pathology, University of Cambridge | Wellcome Sanger Institute for the COVID-19 Genomics UK Consortium |
| EPI_ISL_440513 | Department of Pathology, University of Cambridge | Wellcome Sanger Institute for the COVID-19 Genomics UK Consortium |
| EPI_ISL_440514 | Department of Pathology, University of Cambridge | Wellcome Sanger Institute for the COVID-19 Genomics UK Consortium |
| EPI_ISL_440515 | Department of Pathology, University of Cambridge | Wellcome Sanger Institute for the COVID-19 Genomics UK Consortium |
| EPI_ISL_440516 | Department of Pathology, University of Cambridge | Wellcome Sanger Institute for the COVID-19 Genomics UK Consortium |
| EPI_ISL_440517 | Department of Pathology, University of Cambridge | Wellcome Sanger Institute for the COVID-19 Genomics UK Consortium |
| EPI_ISL_440518 | Department of Pathology, University of Cambridge | Wellcome Sanger Institute for the COVID-19 Genomics UK Consortium |
| EPI_ISL_440519 | Department of Pathology, University of Cambridge | Wellcome Sanger Institute for the COVID-19 Genomics UK Consortium |

| EPI_Accession_ID | Originating Laboratory | Submitting Laboratory |
|---|---|---|
| EPI_ISL_440520 | Department of Pathology, University of Cambridge | Wellcome Sanger Institute for the COVID-19 Genomics UK Consortium |
| EPI_ISL_440521 | Department of Pathology, University of Cambridge | Wellcome Sanger Institute for the COVID-19 Genomics UK Consortium |
| EPI_ISL_440522 | Department of Pathology, University of Cambridge | Wellcome Sanger Institute for the COVID-19 Genomics UK Consortium |
| EPI_ISL_440523 | Department of Pathology, University of Cambridge | Wellcome Sanger Institute for the COVID-19 Genomics UK Consortium |
| EPI_ISL_440524 | Department of Pathology, University of Cambridge | Wellcome Sanger Institute for the COVID-19 Genomics UK Consortium |
| EPI_ISL_440525 | Department of Pathology, University of Cambridge | Wellcome Sanger Institute for the COVID-19 Genomics UK Consortium |
| EPI_ISL_440526 | Department of Pathology, University of Cambridge | Wellcome Sanger Institute for the COVID-19 Genomics UK Consortium |
| EPI_ISL_440527 | Department of Pathology, University of Cambridge | Wellcome Sanger Institute for the COVID-19 Genomics UK Consortium |
| EPI_ISL_440528 | Department of Pathology, University of Cambridge | Wellcome Sanger Institute for the COVID-19 Genomics UK Consortium |
| EPI_ISL_440529 | Department of Pathology, University of Cambridge | Wellcome Sanger Institute for the COVID-19 Genomics UK Consortium |
| EPI_ISL_440530 | Department of Pathology, University of Cambridge | Wellcome Sanger Institute for the COVID-19 Genomics UK Consortium |
| EPI_ISL_440531 | Department of Pathology, University of Cambridge | Wellcome Sanger Institute for the COVID-19 Genomics UK Consortium |
| EPI_ISL_440532 | Department of Pathology, University of Cambridge | Wellcome Sanger Institute for the COVID-19 Genomics UK Consortium |
| EPI_ISL_440533 | Department of Pathology, University of Cambridge | Wellcome Sanger Institute for the COVID-19 Genomics UK Consortium |
| EPI_ISL_440534 | Department of Pathology, University of Cambridge | Wellcome Sanger Institute for the COVID-19 Genomics UK Consortium |
| EPI_ISL_440535 | Department of Pathology, University of Cambridge | Wellcome Sanger Institute for the COVID-19 Genomics UK Consortium |

| EPI_Accession_ID | Originating Laboratory | Submitting Laboratory |
|---|---|---|
| EPI_ISL_440536 | Department of Pathology, University of Cambridge | Wellcome Sanger Institute for the COVID-19 Genomics UK Consortium |
| EPI_ISL_440537 | Department of Pathology, University of Cambridge | Wellcome Sanger Institute for the COVID-19 Genomics UK Consortium |
| EPI_ISL_440538 | Department of Pathology, University of Cambridge | Wellcome Sanger Institute for the COVID-19 Genomics UK Consortium |
| EPI_ISL_440539 | Department of Pathology, University of Cambridge | Wellcome Sanger Institute for the COVID-19 Genomics UK Consortium |
| EPI_ISL_440540 | Department of Pathology, University of Cambridge | Wellcome Sanger Institute for the COVID-19 Genomics UK Consortium |
| EPI_ISL_440541 | Department of Pathology, University of Cambridge | Wellcome Sanger Institute for the COVID-19 Genomics UK Consortium |
| EPI_ISL_440542 | Department of Pathology, University of Cambridge | Wellcome Sanger Institute for the COVID-19 Genomics UK Consortium |
| EPI_ISL_440543 | Department of Pathology, University of Cambridge | Wellcome Sanger Institute for the COVID-19 Genomics UK Consortium |
| EPI_ISL_440544 | Department of Pathology, University of Cambridge | Wellcome Sanger Institute for the COVID-19 Genomics UK Consortium |
| EPI_ISL_440545 | Department of Pathology, University of Cambridge | Wellcome Sanger Institute for the COVID-19 Genomics UK Consortium |
| EPI_ISL_440546 | Department of Pathology, University of Cambridge | Wellcome Sanger Institute for the COVID-19 Genomics UK Consortium |
| EPI_ISL_440547 | Department of Pathology, University of Cambridge | Wellcome Sanger Institute for the COVID-19 Genomics UK Consortium |
| EPI_ISL_440548 | Department of Pathology, University of Cambridge | Wellcome Sanger Institute for the COVID-19 Genomics UK Consortium |
| EPI_ISL_440549 | Department of Pathology, University of Cambridge | Wellcome Sanger Institute for the COVID-19 Genomics UK Consortium |
| EPI_ISL_440550 | Department of Pathology, University of Cambridge | Wellcome Sanger Institute for the COVID-19 Genomics UK Consortium |
| EPI_ISL_440551 | Department of Pathology, University of Cambridge | Wellcome Sanger Institute for the COVID-19 Genomics UK Consortium |

| EPI_Accession_ID | Originating Laboratory | Submitting Laboratory |
|---|---|---|
| EPI_ISL_440552 | Department of Pathology, University of Cambridge | Wellcome Sanger Institute for the COVID-19 Genomics UK Consortium |
| EPI_ISL_440553 | Department of Pathology, University of Cambridge | Wellcome Sanger Institute for the COVID-19 Genomics UK Consortium |
| EPI_ISL_440554 | Department of Pathology, University of Cambridge | Wellcome Sanger Institute for the COVID-19 Genomics UK Consortium |
| EPI_ISL_440555 | Department of Pathology, University of Cambridge | Wellcome Sanger Institute for the COVID-19 Genomics UK Consortium |
| EPI_ISL_440556 | Department of Pathology, University of Cambridge | Wellcome Sanger Institute for the COVID-19 Genomics UK Consortium |
| EPI_ISL_440557 | Department of Pathology, University of Cambridge | Wellcome Sanger Institute for the COVID-19 Genomics UK Consortium |
| EPI_ISL_440558 | Department of Pathology, University of Cambridge | Wellcome Sanger Institute for the COVID-19 Genomics UK Consortium |
| EPI_ISL_440559 | Department of Pathology, University of Cambridge | Wellcome Sanger Institute for the COVID-19 Genomics UK Consortium |
| EPI_ISL_440560 | Department of Pathology, University of Cambridge | Wellcome Sanger Institute for the COVID-19 Genomics UK Consortium |
| EPI_ISL_440561 | Department of Pathology, University of Cambridge | Wellcome Sanger Institute for the COVID-19 Genomics UK Consortium |
| EPI_ISL_440562 | Department of Pathology, University of Cambridge | Wellcome Sanger Institute for the COVID-19 Genomics UK Consortium |
| EPI_ISL_440563 | Department of Pathology, University of Cambridge | Wellcome Sanger Institute for the COVID-19 Genomics UK Consortium |
| EPI_ISL_440564 | Department of Pathology, University of Cambridge | Wellcome Sanger Institute for the COVID-19 Genomics UK Consortium |
| EPI_ISL_440565 | Department of Pathology, University of Cambridge | Wellcome Sanger Institute for the COVID-19 Genomics UK Consortium |
| EPI_ISL_440566 | Department of Pathology, University of Cambridge | Wellcome Sanger Institute for the COVID-19 Genomics UK Consortium |
| EPI_ISL_440567 | Department of Pathology, University of Cambridge | Wellcome Sanger Institute for the COVID-19 Genomics UK Consortium |

| EPI_Accession_ID | Originating Laboratory | Submitting Laboratory |
|---|---|---|
| EPI_ISL_440568 | Department of Pathology, University of Cambridge | Wellcome Sanger Institute for the COVID-19 Genomics UK Consortium |
| EPI_ISL_440569 | Department of Pathology, University of Cambridge | Wellcome Sanger Institute for the COVID-19 Genomics UK Consortium |
| EPI_ISL_440570 | Department of Pathology, University of Cambridge | Wellcome Sanger Institute for the COVID-19 Genomics UK Consortium |
| EPI_ISL_440571 | Department of Pathology, University of Cambridge | Wellcome Sanger Institute for the COVID-19 Genomics UK Consortium |
| EPI_ISL_440572 | Department of Pathology, University of Cambridge | Wellcome Sanger Institute for the COVID-19 Genomics UK Consortium |
| EPI_ISL_440573 | Department of Pathology, University of Cambridge | Wellcome Sanger Institute for the COVID-19 Genomics UK Consortium |
| EPI_ISL_440574 | Department of Pathology, University of Cambridge | Wellcome Sanger Institute for the COVID-19 Genomics UK Consortium |
| EPI_ISL_440575 | Department of Pathology, University of Cambridge | Wellcome Sanger Institute for the COVID-19 Genomics UK Consortium |
| EPI_ISL_440576 | Department of Pathology, University of Cambridge | Wellcome Sanger Institute for the COVID-19 Genomics UK Consortium |
| EPI_ISL_440577 | Department of Pathology, University of Cambridge | Wellcome Sanger Institute for the COVID-19 Genomics UK Consortium |
| EPI_ISL_440578 | Department of Pathology, University of Cambridge | Wellcome Sanger Institute for the COVID-19 Genomics UK Consortium |
| EPI_ISL_440579 | Department of Pathology, University of Cambridge | Wellcome Sanger Institute for the COVID-19 Genomics UK Consortium |
| EPI_ISL_440580 | Department of Pathology, University of Cambridge | Wellcome Sanger Institute for the COVID-19 Genomics UK Consortium |
| EPI_ISL_440581 | Department of Pathology, University of Cambridge | Wellcome Sanger Institute for the COVID-19 Genomics UK Consortium |
| EPI_ISL_440582 | Department of Pathology, University of Cambridge | Wellcome Sanger Institute for the COVID-19 Genomics UK Consortium |
| EPI_ISL_440583 | Department of Pathology, University of Cambridge | Wellcome Sanger Institute for the COVID-19 Genomics UK Consortium |

| EPI_Accession_ID | Originating Laboratory | Submitting Laboratory |
|---|---|---|
| EPI_ISL_440584 | Department of Pathology, University of Cambridge | Wellcome Sanger Institute for the COVID-19 Genomics UK Consortium |
| EPI_ISL_440585 | Department of Pathology, University of Cambridge | Wellcome Sanger Institute for the COVID-19 Genomics UK Consortium |
| EPI_ISL_440586 | Department of Pathology, University of Cambridge | Wellcome Sanger Institute for the COVID-19 Genomics UK Consortium |
| EPI_ISL_440587 | Department of Pathology, University of Cambridge | Wellcome Sanger Institute for the COVID-19 Genomics UK Consortium |
| EPI_ISL_440588 | Department of Pathology, University of Cambridge | Wellcome Sanger Institute for the COVID-19 Genomics UK Consortium |
| EPI_ISL_440589 | Department of Pathology, University of Cambridge | Wellcome Sanger Institute for the COVID-19 Genomics UK Consortium |
| EPI_ISL_440590 | Department of Pathology, University of Cambridge | Wellcome Sanger Institute for the COVID-19 Genomics UK Consortium |
| EPI_ISL_440591 | Department of Pathology, University of Cambridge | Wellcome Sanger Institute for the COVID-19 Genomics UK Consortium |
| EPI_ISL_440592 | Department of Pathology, University of Cambridge | Wellcome Sanger Institute for the COVID-19 Genomics UK Consortium |
| EPI_ISL_440593 | Department of Pathology, University of Cambridge | Wellcome Sanger Institute for the COVID-19 Genomics UK Consortium |
| EPI_ISL_440594 | Department of Pathology, University of Cambridge | Wellcome Sanger Institute for the COVID-19 Genomics UK Consortium |
| EPI_ISL_440595 | Department of Pathology, University of Cambridge | Wellcome Sanger Institute for the COVID-19 Genomics UK Consortium |
| EPI_ISL_440596 | Department of Pathology, University of Cambridge | Wellcome Sanger Institute for the COVID-19 Genomics UK Consortium |
| EPI_ISL_440597 | Department of Pathology, University of Cambridge | Wellcome Sanger Institute for the COVID-19 Genomics UK Consortium |
| EPI_ISL_440598 | Department of Pathology, University of Cambridge | Wellcome Sanger Institute for the COVID-19 Genomics UK Consortium |
| EPI_ISL_440599 | Department of Pathology, University of Cambridge | Wellcome Sanger Institute for the COVID-19 Genomics UK Consortium |

| EPI_Accession_ID | Originating Laboratory | Submitting Laboratory |
|---|---|---|
| EPI_ISL_440600 | Department of Pathology, University of Cambridge | Wellcome Sanger Institute for the COVID-19 Genomics UK Consortium |
| EPI_ISL_440601 | Department of Pathology, University of Cambridge | Wellcome Sanger Institute for the COVID-19 Genomics UK Consortium |
| EPI_ISL_440602 | Department of Pathology, University of Cambridge | Wellcome Sanger Institute for the COVID-19 Genomics UK Consortium |
| EPI_ISL_440603 | Department of Pathology, University of Cambridge | Wellcome Sanger Institute for the COVID-19 Genomics UK Consortium |
| EPI_ISL_440604 | Department of Pathology, University of Cambridge | Wellcome Sanger Institute for the COVID-19 Genomics UK Consortium |
| EPI_ISL_440605 | Department of Pathology, University of Cambridge | Wellcome Sanger Institute for the COVID-19 Genomics UK Consortium |
| EPI_ISL_440606 | Department of Pathology, University of Cambridge | Wellcome Sanger Institute for the COVID-19 Genomics UK Consortium |
| EPI_ISL_440607 | Department of Pathology, University of Cambridge | Wellcome Sanger Institute for the COVID-19 Genomics UK Consortium |
| EPI_ISL_440608 | Department of Pathology, University of Cambridge | Wellcome Sanger Institute for the COVID-19 Genomics UK Consortium |
| EPI_ISL_440609 | Department of Pathology, University of Cambridge | Wellcome Sanger Institute for the COVID-19 Genomics UK Consortium |
| EPI_ISL_440610 | Department of Pathology, University of Cambridge | Wellcome Sanger Institute for the COVID-19 Genomics UK Consortium |
| EPI_ISL_440611 | Department of Pathology, University of Cambridge | Wellcome Sanger Institute for the COVID-19 Genomics UK Consortium |
| EPI_ISL_440612 | Department of Pathology, University of Cambridge | Wellcome Sanger Institute for the COVID-19 Genomics UK Consortium |
| EPI_ISL_440613 | Department of Pathology, University of Cambridge | Wellcome Sanger Institute for the COVID-19 Genomics UK Consortium |
| EPI_ISL_440614 | Department of Pathology, University of Cambridge | Wellcome Sanger Institute for the COVID-19 Genomics UK Consortium |
| EPI_ISL_440615 | Department of Pathology, University of Cambridge | Wellcome Sanger Institute for the COVID-19 Genomics UK Consortium |

| EPI_Accession_ID | Originating Laboratory | Submitting Laboratory |
|---|---|---|
| EPI_ISL_440616 | Department of Pathology, University of Cambridge | Wellcome Sanger Institute for the COVID-19 Genomics UK Consortium |
| EPI_ISL_440617 | Department of Pathology, University of Cambridge | Wellcome Sanger Institute for the COVID-19 Genomics UK Consortium |
| EPI_ISL_440618 | Department of Pathology, University of Cambridge | Wellcome Sanger Institute for the COVID-19 Genomics UK Consortium |
| EPI_ISL_440619 | Department of Pathology, University of Cambridge | Wellcome Sanger Institute for the COVID-19 Genomics UK Consortium |
| EPI_ISL_440620 | Department of Pathology, University of Cambridge | Wellcome Sanger Institute for the COVID-19 Genomics UK Consortium |
| EPI_ISL_440621 | Department of Pathology, University of Cambridge | Wellcome Sanger Institute for the COVID-19 Genomics UK Consortium |
| EPI_ISL_440622 | Department of Pathology, University of Cambridge | Wellcome Sanger Institute for the COVID-19 Genomics UK Consortium |
| EPI_ISL_440623 | PHE South West Regional Laboratory, National Infection Service | Wellcome Sanger Institute for the COVID-19 Genomics UK Consortium |
| EPI_ISL_440624 | PHE South West Regional Laboratory, National Infection Service | Wellcome Sanger Institute for the COVID-19 Genomics UK Consortium |
| EPI_ISL_440625 | PHE South West Regional Laboratory, National Infection Service | Wellcome Sanger Institute for the COVID-19 Genomics UK Consortium |
| EPI_ISL_440626 | PHE South West Regional Laboratory, National Infection Service | Wellcome Sanger Institute for the COVID-19 Genomics UK Consortium |
| EPI_ISL_440627 | PHE South West Regional Laboratory, National Infection Service | Wellcome Sanger Institute for the COVID-19 Genomics UK Consortium |
| EPI_ISL_440628 | PHE South West Regional Laboratory, National Infection Service | Wellcome Sanger Institute for the COVID-19 Genomics UK Consortium |
| EPI_ISL_440629 | PHE South West Regional Laboratory, National Infection Service | Wellcome Sanger Institute for the COVID-19 Genomics UK Consortium |
| EPI_ISL_440630 | PHE South West Regional Laboratory, National Infection Service | Wellcome Sanger Institute for the COVID-19 Genomics UK Consortium |
| EPI_ISL_440631 | PHE South West Regional Laboratory, National Infection Service | Wellcome Sanger Institute for the COVID-19 Genomics UK Consortium |

| EPI_Accession_ID | Originating Laboratory | Submitting Laboratory |
|---|---|---|
| EPI_ISL_440632 | PHE South West Regional Laboratory, National Infection Service | Wellcome Sanger Institute for the COVID-19 Genomics UK Consortium |
| EPI_ISL_440633 | PHE South West Regional Laboratory, National Infection Service | Wellcome Sanger Institute for the COVID-19 Genomics UK Consortium |
| EPI_ISL_440634 | PHE South West Regional Laboratory, National Infection Service | Wellcome Sanger Institute for the COVID-19 Genomics UK Consortium |
| EPI_ISL_440635 | PHE South West Regional Laboratory, National Infection Service | Wellcome Sanger Institute for the COVID-19 Genomics UK Consortium |
| EPI_ISL_440636 | PHE South West Regional Laboratory, National Infection Service | Wellcome Sanger Institute for the COVID-19 Genomics UK Consortium |
| EPI_ISL_440637 | PHE South West Regional Laboratory, National Infection Service | Wellcome Sanger Institute for the COVID-19 Genomics UK Consortium |
| EPI_ISL_440638 | PHE South West Regional Laboratory, National Infection Service | Wellcome Sanger Institute for the COVID-19 Genomics UK Consortium |
| EPI_ISL_440639 | PHE South West Regional Laboratory, National Infection Service | Wellcome Sanger Institute for the COVID-19 Genomics UK Consortium |
| EPI_ISL_440640 | PHE South West Regional Laboratory, National Infection Service | Wellcome Sanger Institute for the COVID-19 Genomics UK Consortium |
| EPI_ISL_440641 | PHE South West Regional Laboratory, National Infection Service | Wellcome Sanger Institute for the COVID-19 Genomics UK Consortium |
| EPI_ISL_440642 | PHE South West Regional Laboratory, National Infection Service | Wellcome Sanger Institute for the COVID-19 Genomics UK Consortium |
| EPI_ISL_440643 | PHE South West Regional Laboratory, National Infection Service | Wellcome Sanger Institute for the COVID-19 Genomics UK Consortium |
| EPI_ISL_440644 | PHE South West Regional Laboratory, National Infection Service | Wellcome Sanger Institute for the COVID-19 Genomics UK Consortium |
| EPI_ISL_440645 | PHE South West Regional Laboratory, National Infection Service | Wellcome Sanger Institute for the COVID-19 Genomics UK Consortium |
| EPI_ISL_440646 | PHE South West Regional Laboratory, National Infection Service | Wellcome Sanger Institute for the COVID-19 Genomics UK Consortium |
| EPI_ISL_440647 | PHE South West Regional Laboratory, National Infection Service | Wellcome Sanger Institute for the COVID-19 Genomics UK Consortium |

| EPI_Accession_ID | Originating Laboratory | Submitting Laboratory |
|---|---|---|
| EPI_ISL_440648 | PHE South West Regional Laboratory, National Infection Service | Wellcome Sanger Institute for the COVID-19 Genomics UK Consortium |
| EPI_ISL_440649 | PHE South West Regional Laboratory, National Infection Service | Wellcome Sanger Institute for the COVID-19 Genomics UK Consortium |
| EPI_ISL_440650 | PHE South West Regional Laboratory, National Infection Service | Wellcome Sanger Institute for the COVID-19 Genomics UK Consortium |
| EPI_ISL_440651 | PHE South West Regional Laboratory, National Infection Service | Wellcome Sanger Institute for the COVID-19 Genomics UK Consortium |
| EPI_ISL_440652 | PHE South West Regional Laboratory, National Infection Service | Wellcome Sanger Institute for the COVID-19 Genomics UK Consortium |
| EPI_ISL_440653 | PHE South West Regional Laboratory, National Infection Service | Wellcome Sanger Institute for the COVID-19 Genomics UK Consortium |
| EPI_ISL_440654 | PHE South West Regional Laboratory, National Infection Service | Wellcome Sanger Institute for the COVID-19 Genomics UK Consortium |
| EPI_ISL_440655 | PHE South West Regional Laboratory, National Infection Service | Wellcome Sanger Institute for the COVID-19 Genomics UK Consortium |
| EPI_ISL_440656 | PHE South West Regional Laboratory, National Infection Service | Wellcome Sanger Institute for the COVID-19 Genomics UK Consortium |
| EPI_ISL_440657 | PHE South West Regional Laboratory, National Infection Service | Wellcome Sanger Institute for the COVID-19 Genomics UK Consortium |
| EPI_ISL_440658 | PHE South West Regional Laboratory, National Infection Service | Wellcome Sanger Institute for the COVID-19 Genomics UK Consortium |
| EPI_ISL_440659 | PHE South West Regional Laboratory, National Infection Service | Wellcome Sanger Institute for the COVID-19 Genomics UK Consortium |
| EPI_ISL_440660 | PHE South West Regional Laboratory, National Infection Service | Wellcome Sanger Institute for the COVID-19 Genomics UK Consortium |
| EPI_ISL_440661 | PHE South West Regional Laboratory, National Infection Service | Wellcome Sanger Institute for the COVID-19 Genomics UK Consortium |
| EPI_ISL_440662 | PHE South West Regional Laboratory, National Infection Service | Wellcome Sanger Institute for the COVID-19 Genomics UK Consortium |
| EPI_ISL_440663 | PHE South West Regional Laboratory, National Infection Service | Wellcome Sanger Institute for the COVID-19 Genomics UK Consortium |

| EPI_Accession_ID | Originating Laboratory | Submitting Laboratory |
| --- | --- | --- |
| EPI_ISL_440664 | PHE South West Regional Laboratory, National Infection Service | Wellcome Sanger Institute for the COVID-19 Genomics UK Consortium |
| EPI_ISL_440665 | PHE South West Regional Laboratory, National Infection Service | Wellcome Sanger Institute for the COVID-19 Genomics UK Consortium |
| EPI_ISL_440666 | PHE South West Regional Laboratory, National Infection Service | Wellcome Sanger Institute for the COVID-19 Genomics UK Consortium |
| EPI_ISL_440667 | PHE South West Regional Laboratory, National Infection Service | Wellcome Sanger Institute for the COVID-19 Genomics UK Consortium |
| EPI_ISL_440668 | PHE South West Regional Laboratory, National Infection Service | Wellcome Sanger Institute for the COVID-19 Genomics UK Consortium |
| EPI_ISL_440669 | PHE South West Regional Laboratory, National Infection Service | Wellcome Sanger Institute for the COVID-19 Genomics UK Consortium |
| EPI_ISL_440670 | PHE South West Regional Laboratory, National Infection Service | Wellcome Sanger Institute for the COVID-19 Genomics UK Consortium |
| EPI_ISL_440671 | PHE South West Regional Laboratory, National Infection Service | Wellcome Sanger Institute for the COVID-19 Genomics UK Consortium |
| EPI_ISL_440672 | PHE South West Regional Laboratory, National Infection Service | Wellcome Sanger Institute for the COVID-19 Genomics UK Consortium |
| EPI_ISL_440673 | PHE South West Regional Laboratory, National Infection Service | Wellcome Sanger Institute for the COVID-19 Genomics UK Consortium |
| EPI_ISL_440674 | PHE South West Regional Laboratory, National Infection Service | Wellcome Sanger Institute for the COVID-19 Genomics UK Consortium |
| EPI_ISL_440675 | PHE South West Regional Laboratory, National Infection Service | Wellcome Sanger Institute for the COVID-19 Genomics UK Consortium |
| EPI_ISL_440676 | PHE South West Regional Laboratory, National Infection Service | Wellcome Sanger Institute for the COVID-19 Genomics UK Consortium |
| EPI_ISL_440677 | PHE South West Regional Laboratory, National Infection Service | Wellcome Sanger Institute for the COVID-19 Genomics UK Consortium |
| EPI_ISL_440678 | PHE South West Regional Laboratory, National Infection Service | Wellcome Sanger Institute for the COVID-19 Genomics UK Consortium |
| EPI_ISL_440679 | PHE South West Regional Laboratory, National Infection Service | Wellcome Sanger Institute for the COVID-19 Genomics UK Consortium |

| EPI_Accession_ID | Originating Laboratory | Submitting Laboratory |
|---|---|---|
| EPI_ISL_440680 | PHE South West Regional Laboratory, National Infection Service | Wellcome Sanger Institute for the COVID-19 Genomics UK Consortium |
| EPI_ISL_440681 | PHE South West Regional Laboratory, National Infection Service | Wellcome Sanger Institute for the COVID-19 Genomics UK Consortium |
| EPI_ISL_440682 | PHE South West Regional Laboratory, National Infection Service | Wellcome Sanger Institute for the COVID-19 Genomics UK Consortium |
| EPI_ISL_440683 | PHE South West Regional Laboratory, National Infection Service | Wellcome Sanger Institute for the COVID-19 Genomics UK Consortium |
| EPI_ISL_440684 | PHE South West Regional Laboratory, National Infection Service | Wellcome Sanger Institute for the COVID-19 Genomics UK Consortium |
| EPI_ISL_440685 | PHE South West Regional Laboratory, National Infection Service | Wellcome Sanger Institute for the COVID-19 Genomics UK Consortium |
| EPI_ISL_440686 | PHE South West Regional Laboratory, National Infection Service | Wellcome Sanger Institute for the COVID-19 Genomics UK Consortium |
| EPI_ISL_440687 | PHE South West Regional Laboratory, National Infection Service | Wellcome Sanger Institute for the COVID-19 Genomics UK Consortium |
| EPI_ISL_440688 | PHE South West Regional Laboratory, National Infection Service | Wellcome Sanger Institute for the COVID-19 Genomics UK Consortium |
| EPI_ISL_440689 | PHE South West Regional Laboratory, National Infection Service | Wellcome Sanger Institute for the COVID-19 Genomics UK Consortium |
| EPI_ISL_440690 | PHE South West Regional Laboratory, National Infection Service | Wellcome Sanger Institute for the COVID-19 Genomics UK Consortium |
| EPI_ISL_440691 | PHE South West Regional Laboratory, National Infection Service | Wellcome Sanger Institute for the COVID-19 Genomics UK Consortium |
| EPI_ISL_440692 | PHE South West Regional Laboratory, National Infection Service | Wellcome Sanger Institute for the COVID-19 Genomics UK Consortium |
| EPI_ISL_440693 | PHE South West Regional Laboratory, National Infection Service | Wellcome Sanger Institute for the COVID-19 Genomics UK Consortium |
| EPI_ISL_440694 | PHE South West Regional Laboratory, National Infection Service | Wellcome Sanger Institute for the COVID-19 Genomics UK Consortium |
| EPI_ISL_440695 | PHE South West Regional Laboratory, National Infection Service | Wellcome Sanger Institute for the COVID-19 Genomics UK Consortium |

| EPI_Accession_ID | Originating Laboratory | Submitting Laboratory |
|---|---|---|
| EPI_ISL_440696 | PHE South West Regional Laboratory, National Infection Service | Wellcome Sanger Institute for the COVID-19 Genomics UK Consortium |
| EPI_ISL_440697 | PHE South West Regional Laboratory, National Infection Service | Wellcome Sanger Institute for the COVID-19 Genomics UK Consortium |
| EPI_ISL_440698 | PHE South West Regional Laboratory, National Infection Service | Wellcome Sanger Institute for the COVID-19 Genomics UK Consortium |
| EPI_ISL_440699 | PHE South West Regional Laboratory, National Infection Service | Wellcome Sanger Institute for the COVID-19 Genomics UK Consortium |
| EPI_ISL_440700 | PHE South West Regional Laboratory, National Infection Service | Wellcome Sanger Institute for the COVID-19 Genomics UK Consortium |
| EPI_ISL_440701 | PHE South West Regional Laboratory, National Infection Service | Wellcome Sanger Institute for the COVID-19 Genomics UK Consortium |
| EPI_ISL_440702 | PHE South West Regional Laboratory, National Infection Service | Wellcome Sanger Institute for the COVID-19 Genomics UK Consortium |
| EPI_ISL_440703 | PHE South West Regional Laboratory, National Infection Service | Wellcome Sanger Institute for the COVID-19 Genomics UK Consortium |
| EPI_ISL_440704 | PHE South West Regional Laboratory, National Infection Service | Wellcome Sanger Institute for the COVID-19 Genomics UK Consortium |
| EPI_ISL_440705 | PHE South West Regional Laboratory, National Infection Service | Wellcome Sanger Institute for the COVID-19 Genomics UK Consortium |
| EPI_ISL_440706 | PHE South West Regional Laboratory, National Infection Service | Wellcome Sanger Institute for the COVID-19 Genomics UK Consortium |
| EPI_ISL_440707 | PHE South West Regional Laboratory, National Infection Service | Wellcome Sanger Institute for the COVID-19 Genomics UK Consortium |
| EPI_ISL_440708 | PHE South West Regional Laboratory, National Infection Service | Wellcome Sanger Institute for the COVID-19 Genomics UK Consortium |
| EPI_ISL_440709 | PHE South West Regional Laboratory, National Infection Service | Wellcome Sanger Institute for the COVID-19 Genomics UK Consortium |
| EPI_ISL_440710 | PHE South West Regional Laboratory, National Infection Service | Wellcome Sanger Institute for the COVID-19 Genomics UK Consortium |
| EPI_ISL_440711 | PHE South West Regional Laboratory, National Infection Service | Wellcome Sanger Institute for the COVID-19 Genomics UK Consortium |

| EPI_Accession_ID | Originating Laboratory | Submitting Laboratory |
|---|---|---|
| EPI_ISL_440712 | PHE South West Regional Laboratory, National Infection Service | Wellcome Sanger Institute for the COVID-19 Genomics UK Consortium |
| EPI_ISL_440713 | PHE South West Regional Laboratory, National Infection Service | Wellcome Sanger Institute for the COVID-19 Genomics UK Consortium |
| EPI_ISL_440714 | PHE South West Regional Laboratory, National Infection Service | Wellcome Sanger Institute for the COVID-19 Genomics UK Consortium |
| EPI_ISL_440715 | PHE South West Regional Laboratory, National Infection Service | Wellcome Sanger Institute for the COVID-19 Genomics UK Consortium |
| EPI_ISL_440716 | PHE South West Regional Laboratory, National Infection Service | Wellcome Sanger Institute for the COVID-19 Genomics UK Consortium |
| EPI_ISL_440717 | PHE South West Regional Laboratory, National Infection Service | Wellcome Sanger Institute for the COVID-19 Genomics UK Consortium |
| EPI_ISL_440718 | PHE South West Regional Laboratory, National Infection Service | Wellcome Sanger Institute for the COVID-19 Genomics UK Consortium |
| EPI_ISL_440719 | PHE South West Regional Laboratory, National Infection Service | Wellcome Sanger Institute for the COVID-19 Genomics UK Consortium |
| EPI_ISL_440720 | PHE South West Regional Laboratory, National Infection Service | Wellcome Sanger Institute for the COVID-19 Genomics UK Consortium |
| EPI_ISL_440721 | PHE South West Regional Laboratory, National Infection Service | Wellcome Sanger Institute for the COVID-19 Genomics UK Consortium |
| EPI_ISL_440722 | PHE South West Regional Laboratory, National Infection Service | Wellcome Sanger Institute for the COVID-19 Genomics UK Consortium |
| EPI_ISL_440723 | PHE South West Regional Laboratory, National Infection Service | Wellcome Sanger Institute for the COVID-19 Genomics UK Consortium |
| EPI_ISL_440724 | PHE South West Regional Laboratory, National Infection Service | Wellcome Sanger Institute for the COVID-19 Genomics UK Consortium |
| EPI_ISL_440725 | PHE South West Regional Laboratory, National Infection Service | Wellcome Sanger Institute for the COVID-19 Genomics UK Consortium |
| EPI_ISL_440726 | PHE South West Regional Laboratory, National Infection Service | Wellcome Sanger Institute for the COVID-19 Genomics UK Consortium |
| EPI_ISL_440727 | PHE South West Regional Laboratory, National Infection Service | Wellcome Sanger Institute for the COVID-19 Genomics UK Consortium |

| EPI_Accession_ID | Originating Laboratory | Submitting Laboratory |
|---|---|---|
| EPI_ISL_440728 | PHE South West Regional Laboratory, National Infection Service | Wellcome Sanger Institute for the COVID-19 Genomics UK Consortium |
| EPI_ISL_440729 | PHE South West Regional Laboratory, National Infection Service | Wellcome Sanger Institute for the COVID-19 Genomics UK Consortium |
| EPI_ISL_440730 | PHE South West Regional Laboratory, National Infection Service | Wellcome Sanger Institute for the COVID-19 Genomics UK Consortium |
| EPI_ISL_440731 | PHE South West Regional Laboratory, National Infection Service | Wellcome Sanger Institute for the COVID-19 Genomics UK Consortium |
| EPI_ISL_440732 | PHE South West Regional Laboratory, National Infection Service | Wellcome Sanger Institute for the COVID-19 Genomics UK Consortium |
| EPI_ISL_440733 | PHE South West Regional Laboratory, National Infection Service | Wellcome Sanger Institute for the COVID-19 Genomics UK Consortium |
| EPI_ISL_440734 | PHE South West Regional Laboratory, National Infection Service | Wellcome Sanger Institute for the COVID-19 Genomics UK Consortium |
| EPI_ISL_440735 | PHE South West Regional Laboratory, National Infection Service | Wellcome Sanger Institute for the COVID-19 Genomics UK Consortium |
| EPI_ISL_440736 | PHE South West Regional Laboratory, National Infection Service | Wellcome Sanger Institute for the COVID-19 Genomics UK Consortium |
| EPI_ISL_440737 | PHE South West Regional Laboratory, National Infection Service | Wellcome Sanger Institute for the COVID-19 Genomics UK Consortium |
| EPI_ISL_440738 | PHE South West Regional Laboratory, National Infection Service | Wellcome Sanger Institute for the COVID-19 Genomics UK Consortium |
| EPI_ISL_440739 | PHE South West Regional Laboratory, National Infection Service | Wellcome Sanger Institute for the COVID-19 Genomics UK Consortium |
| EPI_ISL_440740 | PHE South West Regional Laboratory, National Infection Service | Wellcome Sanger Institute for the COVID-19 Genomics UK Consortium |
| EPI_ISL_440741 | PHE South West Regional Laboratory, National Infection Service | Wellcome Sanger Institute for the COVID-19 Genomics UK Consortium |
| EPI_ISL_440742 | PHE South West Regional Laboratory, National Infection Service | Wellcome Sanger Institute for the COVID-19 Genomics UK Consortium |
| EPI_ISL_440743 | PHE South West Regional Laboratory, National Infection Service | Wellcome Sanger Institute for the COVID-19 Genomics UK Consortium |

| EPI_Accession_ID | Originating Laboratory | Submitting Laboratory |
|---|---|---|
| EPI_ISL_440744 | PHE South West Regional Laboratory, National Infection Service | Wellcome Sanger Institute for the COVID-19 Genomics UK Consortium |
| EPI_ISL_440745 | PHE South West Regional Laboratory, National Infection Service | Wellcome Sanger Institute for the COVID-19 Genomics UK Consortium |
| EPI_ISL_440746 | PHE South West Regional Laboratory, National Infection Service | Wellcome Sanger Institute for the COVID-19 Genomics UK Consortium |
| EPI_ISL_440747 | PHE South West Regional Laboratory, National Infection Service | Wellcome Sanger Institute for the COVID-19 Genomics UK Consortium |
| EPI_ISL_440748 | PHE South West Regional Laboratory, National Infection Service | Wellcome Sanger Institute for the COVID-19 Genomics UK Consortium |
| EPI_ISL_440749 | PHE South West Regional Laboratory, National Infection Service | Wellcome Sanger Institute for the COVID-19 Genomics UK Consortium |
| EPI_ISL_440750 | PHE South West Regional Laboratory, National Infection Service | Wellcome Sanger Institute for the COVID-19 Genomics UK Consortium |
| EPI_ISL_440751 | PHE South West Regional Laboratory, National Infection Service | Wellcome Sanger Institute for the COVID-19 Genomics UK Consortium |
| EPI_ISL_440752 | PHE South West Regional Laboratory, National Infection Service | Wellcome Sanger Institute for the COVID-19 Genomics UK Consortium |
| EPI_ISL_440753 | PHE South West Regional Laboratory, National Infection Service | Wellcome Sanger Institute for the COVID-19 Genomics UK Consortium |
| EPI_ISL_440754 | PHE South West Regional Laboratory, National Infection Service | Wellcome Sanger Institute for the COVID-19 Genomics UK Consortium |
| EPI_ISL_440755 | PHE South West Regional Laboratory, National Infection Service | Wellcome Sanger Institute for the COVID-19 Genomics UK Consortium |
| EPI_ISL_440756 | PHE South West Regional Laboratory, National Infection Service | Wellcome Sanger Institute for the COVID-19 Genomics UK Consortium |
| EPI_ISL_440757 | PHE South West Regional Laboratory, National Infection Service | Wellcome Sanger Institute for the COVID-19 Genomics UK Consortium |
| EPI_ISL_440758 | PHE South West Regional Laboratory, National Infection Service | Wellcome Sanger Institute for the COVID-19 Genomics UK Consortium |
| EPI_ISL_440759 | PHE South West Regional Laboratory, National Infection Service | Wellcome Sanger Institute for the COVID-19 Genomics UK Consortium |

| EPI_Accession_ID | Originating Laboratory | Submitting Laboratory |
|---|---|---|
| EPI_ISL_440760 | PHE South West Regional Laboratory, National Infection Service | Wellcome Sanger Institute for the COVID-19 Genomics UK Consortium |
| EPI_ISL_440761 | PHE South West Regional Laboratory, National Infection Service | Wellcome Sanger Institute for the COVID-19 Genomics UK Consortium |
| EPI_ISL_440762 | PHE South West Regional Laboratory, National Infection Service | Wellcome Sanger Institute for the COVID-19 Genomics UK Consortium |
| EPI_ISL_440763 | PHE South West Regional Laboratory, National Infection Service | Wellcome Sanger Institute for the COVID-19 Genomics UK Consortium |
| EPI_ISL_440764 | PHE South West Regional Laboratory, National Infection Service | Wellcome Sanger Institute for the COVID-19 Genomics UK Consortium |
| EPI_ISL_440765 | PHE South West Regional Laboratory, National Infection Service | Wellcome Sanger Institute for the COVID-19 Genomics UK Consortium |
| EPI_ISL_440766 | PHE South West Regional Laboratory, National Infection Service | Wellcome Sanger Institute for the COVID-19 Genomics UK Consortium |
| EPI_ISL_440767 | PHE South West Regional Laboratory, National Infection Service | Wellcome Sanger Institute for the COVID-19 Genomics UK Consortium |
| EPI_ISL_440768 | PHE South West Regional Laboratory, National Infection Service | Wellcome Sanger Institute for the COVID-19 Genomics UK Consortium |
| EPI_ISL_440769 | PHE South West Regional Laboratory, National Infection Service | Wellcome Sanger Institute for the COVID-19 Genomics UK Consortium |
| EPI_ISL_440770 | PHE South West Regional Laboratory, National Infection Service | Wellcome Sanger Institute for the COVID-19 Genomics UK Consortium |
| EPI_ISL_440771 | PHE South West Regional Laboratory, National Infection Service | Wellcome Sanger Institute for the COVID-19 Genomics UK Consortium |
| EPI_ISL_440772 | PHE South West Regional Laboratory, National Infection Service | Wellcome Sanger Institute for the COVID-19 Genomics UK Consortium |
| EPI_ISL_440773 | PHE South West Regional Laboratory, National Infection Service | Wellcome Sanger Institute for the COVID-19 Genomics UK Consortium |
| EPI_ISL_440774 | PHE South West Regional Laboratory, National Infection Service | Wellcome Sanger Institute for the COVID-19 Genomics UK Consortium |
| EPI_ISL_440775 | PHE South West Regional Laboratory, National Infection Service | Wellcome Sanger Institute for the COVID-19 Genomics UK Consortium |

| EPI_Accession_ID | Originating Laboratory | Submitting Laboratory |
|---|---|---|
| EPI_ISL_440776 | PHE South West Regional Laboratory, National Infection Service | Wellcome Sanger Institute for the COVID-19 Genomics UK Consortium |
| EPI_ISL_440777 | PHE South West Regional Laboratory, National Infection Service | Wellcome Sanger Institute for the COVID-19 Genomics UK Consortium |
| EPI_ISL_440778 | PHE South West Regional Laboratory, National Infection Service | Wellcome Sanger Institute for the COVID-19 Genomics UK Consortium |
| EPI_ISL_440779 | PHE South West Regional Laboratory, National Infection Service | Wellcome Sanger Institute for the COVID-19 Genomics UK Consortium |
| EPI_ISL_440780 | PHE South West Regional Laboratory, National Infection Service | Wellcome Sanger Institute for the COVID-19 Genomics UK Consortium |
| EPI_ISL_440781 | PHE South West Regional Laboratory, National Infection Service | Wellcome Sanger Institute for the COVID-19 Genomics UK Consortium |
| EPI_ISL_440782 | PHE South West Regional Laboratory, National Infection Service | Wellcome Sanger Institute for the COVID-19 Genomics UK Consortium |
| EPI_ISL_440783 | PHE South West Regional Laboratory, National Infection Service | Wellcome Sanger Institute for the COVID-19 Genomics UK Consortium |
| EPI_ISL_440784 | PHE South West Regional Laboratory, National Infection Service | Wellcome Sanger Institute for the COVID-19 Genomics UK Consortium |
| EPI_ISL_440785 | PHE South West Regional Laboratory, National Infection Service | Wellcome Sanger Institute for the COVID-19 Genomics UK Consortium |
| EPI_ISL_440786 | PHE South West Regional Laboratory, National Infection Service | Wellcome Sanger Institute for the COVID-19 Genomics UK Consortium |
| EPI_ISL_440787 | PHE South West Regional Laboratory, National Infection Service | Wellcome Sanger Institute for the COVID-19 Genomics UK Consortium |
| EPI_ISL_440788 | PHE South West Regional Laboratory, National Infection Service | Wellcome Sanger Institute for the COVID-19 Genomics UK Consortium |
| EPI_ISL_440789 | PHE South West Regional Laboratory, National Infection Service | Wellcome Sanger Institute for the COVID-19 Genomics UK Consortium |
| EPI_ISL_440790 | PHE South West Regional Laboratory, National Infection Service | Wellcome Sanger Institute for the COVID-19 Genomics UK Consortium |
| EPI_ISL_440791 | PHE South West Regional Laboratory, National Infection Service | Wellcome Sanger Institute for the COVID-19 Genomics UK Consortium |

| EPI_Accession_ID | Originating Laboratory | Submitting Laboratory |
|---|---|---|
| EPI_ISL_440792 | PHE South West Regional Laboratory, National Infection Service | Wellcome Sanger Institute for the COVID-19 Genomics UK Consortium |
| EPI_ISL_440793 | PHE South West Regional Laboratory, National Infection Service | Wellcome Sanger Institute for the COVID-19 Genomics UK Consortium |
| EPI_ISL_440794 | PHE South West Regional Laboratory, National Infection Service | Wellcome Sanger Institute for the COVID-19 Genomics UK Consortium |
| EPI_ISL_440795 | PHE South West Regional Laboratory, National Infection Service | Wellcome Sanger Institute for the COVID-19 Genomics UK Consortium |
| EPI_ISL_440796 | PHE South West Regional Laboratory, National Infection Service | Wellcome Sanger Institute for the COVID-19 Genomics UK Consortium |
| EPI_ISL_440797 | PHE South West Regional Laboratory, National Infection Service | Wellcome Sanger Institute for the COVID-19 Genomics UK Consortium |
| EPI_ISL_440798 | PHE South West Regional Laboratory, National Infection Service | Wellcome Sanger Institute for the COVID-19 Genomics UK Consortium |
| EPI_ISL_440799 | PHE South West Regional Laboratory, National Infection Service | Wellcome Sanger Institute for the COVID-19 Genomics UK Consortium |
| EPI_ISL_440800 | PHE South West Regional Laboratory, National Infection Service | Wellcome Sanger Institute for the COVID-19 Genomics UK Consortium |
| EPI_ISL_440801 | PHE South West Regional Laboratory, National Infection Service | Wellcome Sanger Institute for the COVID-19 Genomics UK Consortium |
| EPI_ISL_440802 | PHE South West Regional Laboratory, National Infection Service | Wellcome Sanger Institute for the COVID-19 Genomics UK Consortium |
| EPI_ISL_440803 | PHE South West Regional Laboratory, National Infection Service | Wellcome Sanger Institute for the COVID-19 Genomics UK Consortium |
| EPI_ISL_440804 | PHE South West Regional Laboratory, National Infection Service | Wellcome Sanger Institute for the COVID-19 Genomics UK Consortium |
| EPI_ISL_440805 | PHE South West Regional Laboratory, National Infection Service | Wellcome Sanger Institute for the COVID-19 Genomics UK Consortium |
| EPI_ISL_440806 | PHE South West Regional Laboratory, National Infection Service | Wellcome Sanger Institute for the COVID-19 Genomics UK Consortium |
| EPI_ISL_440807 | PHE South West Regional Laboratory, National Infection Service | Wellcome Sanger Institute for the COVID-19 Genomics UK Consortium |

| EPI_Accession_ID | Originating Laboratory | Submitting Laboratory |
|---|---|---|
| EPI_ISL_440808 | PHE South West Regional Laboratory, National Infection Service | Wellcome Sanger Institute for the COVID-19 Genomics UK Consortium |
| EPI_ISL_440809 | PHE South West Regional Laboratory, National Infection Service | Wellcome Sanger Institute for the COVID-19 Genomics UK Consortium |
| EPI_ISL_440810 | Department of Pathology, University of Cambridge | Wellcome Sanger Institute for the COVID-19 Genomics UK Consortium |
| EPI_ISL_440811 | Department of Pathology, University of Cambridge | Wellcome Sanger Institute for the COVID-19 Genomics UK Consortium |
| EPI_ISL_440812 | Department of Pathology, University of Cambridge | Wellcome Sanger Institute for the COVID-19 Genomics UK Consortium |
| EPI_ISL_440813 | Department of Pathology, University of Cambridge | Wellcome Sanger Institute for the COVID-19 Genomics UK Consortium |
| EPI_ISL_440814 | Department of Pathology, University of Cambridge | Wellcome Sanger Institute for the COVID-19 Genomics UK Consortium |
| EPI_ISL_440815 | Department of Pathology, University of Cambridge | Wellcome Sanger Institute for the COVID-19 Genomics UK Consortium |
| EPI_ISL_440816 | Department of Pathology, University of Cambridge | Wellcome Sanger Institute for the COVID-19 Genomics UK Consortium |
| EPI_ISL_440817 | Department of Pathology, University of Cambridge | Wellcome Sanger Institute for the COVID-19 Genomics UK Consortium |
| EPI_ISL_440818 | Department of Pathology, University of Cambridge | Wellcome Sanger Institute for the COVID-19 Genomics UK Consortium |
| EPI_ISL_440819 | Department of Pathology, University of Cambridge | Wellcome Sanger Institute for the COVID-19 Genomics UK Consortium |
| EPI_ISL_440820 | Department of Pathology, University of Cambridge | Wellcome Sanger Institute for the COVID-19 Genomics UK Consortium |
| EPI_ISL_440821 | Department of Pathology, University of Cambridge | Wellcome Sanger Institute for the COVID-19 Genomics UK Consortium |
| EPI_ISL_440822 | Department of Pathology, University of Cambridge | Wellcome Sanger Institute for the COVID-19 Genomics UK Consortium |
| EPI_ISL_440823 | Department of Pathology, University of Cambridge | Wellcome Sanger Institute for the COVID-19 Genomics UK Consortium |

| EPI_Accession_ID | Originating Laboratory | Submitting Laboratory |
|---|---|---|
| EPI_ISL_440824 | Department of Pathology, University of Cambridge | Wellcome Sanger Institute for the COVID-19 Genomics UK Consortium |
| EPI_ISL_440825 | Department of Pathology, University of Cambridge | Wellcome Sanger Institute for the COVID-19 Genomics UK Consortium |
| EPI_ISL_440826 | Department of Pathology, University of Cambridge | Wellcome Sanger Institute for the COVID-19 Genomics UK Consortium |
| EPI_ISL_440827 | Department of Pathology, University of Cambridge | Wellcome Sanger Institute for the COVID-19 Genomics UK Consortium |
| EPI_ISL_440828 | Department of Pathology, University of Cambridge | Wellcome Sanger Institute for the COVID-19 Genomics UK Consortium |
| EPI_ISL_440829 | Department of Pathology, University of Cambridge | Wellcome Sanger Institute for the COVID-19 Genomics UK Consortium |
| EPI_ISL_440830 | Department of Pathology, University of Cambridge | Wellcome Sanger Institute for the COVID-19 Genomics UK Consortium |
| EPI_ISL_440831 | Department of Pathology, University of Cambridge | Wellcome Sanger Institute for the COVID-19 Genomics UK Consortium |
| EPI_ISL_440832 | Department of Pathology, University of Cambridge | Wellcome Sanger Institute for the COVID-19 Genomics UK Consortium |
| EPI_ISL_440833 | Department of Pathology, University of Cambridge | Wellcome Sanger Institute for the COVID-19 Genomics UK Consortium |
| EPI_ISL_440834 | Department of Pathology, University of Cambridge | Wellcome Sanger Institute for the COVID-19 Genomics UK Consortium |
| EPI_ISL_440835 | Department of Pathology, University of Cambridge | Wellcome Sanger Institute for the COVID-19 Genomics UK Consortium |
| EPI_ISL_440836 | Department of Pathology, University of Cambridge | Wellcome Sanger Institute for the COVID-19 Genomics UK Consortium |
| EPI_ISL_440837 | Department of Pathology, University of Cambridge | Wellcome Sanger Institute for the COVID-19 Genomics UK Consortium |
| EPI_ISL_440838 | Department of Pathology, University of Cambridge | Wellcome Sanger Institute for the COVID-19 Genomics UK Consortium |
| EPI_ISL_440839 | Department of Pathology, University of Cambridge | Wellcome Sanger Institute for the COVID-19 Genomics UK Consortium |

| EPI_Accession_ID | Originating Laboratory | Submitting Laboratory |
|---|---|---|
| EPI_ISL_440840 | Department of Pathology, University of Cambridge | Wellcome Sanger Institute for the COVID-19 Genomics UK Consortium |
| EPI_ISL_440841 | Department of Pathology, University of Cambridge | Wellcome Sanger Institute for the COVID-19 Genomics UK Consortium |
| EPI_ISL_440842 | Department of Pathology, University of Cambridge | Wellcome Sanger Institute for the COVID-19 Genomics UK Consortium |
| EPI_ISL_440843 | Department of Pathology, University of Cambridge | Wellcome Sanger Institute for the COVID-19 Genomics UK Consortium |
| EPI_ISL_440844 | Department of Pathology, University of Cambridge | Wellcome Sanger Institute for the COVID-19 Genomics UK Consortium |
| EPI_ISL_440845 | Department of Pathology, University of Cambridge | Wellcome Sanger Institute for the COVID-19 Genomics UK Consortium |
| EPI_ISL_440846 | Department of Pathology, University of Cambridge | Wellcome Sanger Institute for the COVID-19 Genomics UK Consortium |
| EPI_ISL_440847 | Department of Pathology, University of Cambridge | Wellcome Sanger Institute for the COVID-19 Genomics UK Consortium |
| EPI_ISL_440848 | Department of Pathology, University of Cambridge | Wellcome Sanger Institute for the COVID-19 Genomics UK Consortium |
| EPI_ISL_440849 | Department of Pathology, University of Cambridge | Wellcome Sanger Institute for the COVID-19 Genomics UK Consortium |
| EPI_ISL_440850 | Department of Pathology, University of Cambridge | Wellcome Sanger Institute for the COVID-19 Genomics UK Consortium |
| EPI_ISL_440851 | Department of Pathology, University of Cambridge | Wellcome Sanger Institute for the COVID-19 Genomics UK Consortium |
| EPI_ISL_440852 | Liverpool Clinical Laboratories | COVID-19 Genomics UK (COG-UK) Consortium |
| EPI_ISL_440853 | Liverpool Clinical Laboratories | COVID-19 Genomics UK (COG-UK) Consortium |
| EPI_ISL_440854 | Liverpool Clinical Laboratories | COVID-19 Genomics UK (COG-UK) Consortium |
| EPI_ISL_440855 | Liverpool Clinical Laboratories | COVID-19 Genomics UK (COG-UK) Consortium |
| EPI_ISL_440856 | Liverpool Clinical Laboratories | COVID-19 Genomics UK (COG-UK) Consortium |
| EPI_ISL_440857 | Liverpool Clinical Laboratories | COVID-19 Genomics UK (COG-UK) Consortium |
| EPI_ISL_440858 | Liverpool Clinical Laboratories | COVID-19 Genomics UK (COG-UK) Consortium |
| EPI_ISL_440859 | Liverpool Clinical Laboratories | COVID-19 Genomics UK (COG-UK) Consortium |
| EPI_ISL_440860 | Liverpool Clinical Laboratories | COVID-19 Genomics UK (COG-UK) Consortium |

| EPI_Accession_ID | Originating Laboratory | Submitting Laboratory |
|---|---|---|
| EPI_ISL_440861 | Liverpool Clinical Laboratories | COVID-19 Genomics UK (COG-UK) Consortium |
| EPI_ISL_440862 | Liverpool Clinical Laboratories | COVID-19 Genomics UK (COG-UK) Consortium |
| EPI_ISL_440863 | Liverpool Clinical Laboratories | COVID-19 Genomics UK (COG-UK) Consortium |
| EPI_ISL_440864 | Liverpool Clinical Laboratories | COVID-19 Genomics UK (COG-UK) Consortium |
| EPI_ISL_440865 | Liverpool Clinical Laboratories | COVID-19 Genomics UK (COG-UK) Consortium |
| EPI_ISL_440866 | Liverpool Clinical Laboratories | COVID-19 Genomics UK (COG-UK) Consortium |
| EPI_ISL_440867 | Liverpool Clinical Laboratories | COVID-19 Genomics UK (COG-UK) Consortium |
| EPI_ISL_440868 | Liverpool Clinical Laboratories | COVID-19 Genomics UK (COG-UK) Consortium |
| EPI_ISL_440869 | Liverpool Clinical Laboratories | COVID-19 Genomics UK (COG-UK) Consortium |
| EPI_ISL_440870 | Liverpool Clinical Laboratories | COVID-19 Genomics UK (COG-UK) Consortium |
| EPI_ISL_440871 | Liverpool Clinical Laboratories | COVID-19 Genomics UK (COG-UK) Consortium |
| EPI_ISL_440872 | Liverpool Clinical Laboratories | COVID-19 Genomics UK (COG-UK) Consortium |
| EPI_ISL_440873 | Liverpool Clinical Laboratories | COVID-19 Genomics UK (COG-UK) Consortium |
| EPI_ISL_440874 | Liverpool Clinical Laboratories | COVID-19 Genomics UK (COG-UK) Consortium |
| EPI_ISL_440875 | Liverpool Clinical Laboratories | COVID-19 Genomics UK (COG-UK) Consortium |
| EPI_ISL_440876 | Liverpool Clinical Laboratories | COVID-19 Genomics UK (COG-UK) Consortium |
| EPI_ISL_440877 | Liverpool Clinical Laboratories | COVID-19 Genomics UK (COG-UK) Consortium |
| EPI_ISL_440878 | Liverpool Clinical Laboratories | COVID-19 Genomics UK (COG-UK) Consortium |
| EPI_ISL_440879 | Liverpool Clinical Laboratories | COVID-19 Genomics UK (COG-UK) Consortium |
| EPI_ISL_440880 | Liverpool Clinical Laboratories | COVID-19 Genomics UK (COG-UK) Consortium |
| EPI_ISL_440881 | Liverpool Clinical Laboratories | COVID-19 Genomics UK (COG-UK) Consortium |
| EPI_ISL_440882 | Liverpool Clinical Laboratories | COVID-19 Genomics UK (COG-UK) Consortium |
| EPI_ISL_440883 | Liverpool Clinical Laboratories | COVID-19 Genomics UK (COG-UK) Consortium |
| EPI_ISL_440884 | Liverpool Clinical Laboratories | COVID-19 Genomics UK (COG-UK) Consortium |
| EPI_ISL_440885 | Liverpool Clinical Laboratories | COVID-19 Genomics UK (COG-UK) Consortium |
| EPI_ISL_440886 | Liverpool Clinical Laboratories | COVID-19 Genomics UK (COG-UK) Consortium |
| EPI_ISL_440887 | Liverpool Clinical Laboratories | COVID-19 Genomics UK (COG-UK) Consortium |
| EPI_ISL_440888 | Liverpool Clinical Laboratories | COVID-19 Genomics UK (COG-UK) Consortium |
| EPI_ISL_440889 | Liverpool Clinical Laboratories | COVID-19 Genomics UK (COG-UK) Consortium |
| EPI_ISL_440890 | Liverpool Clinical Laboratories | COVID-19 Genomics UK (COG-UK) Consortium |
| EPI_ISL_440891 | Liverpool Clinical Laboratories | COVID-19 Genomics UK (COG-UK) Consortium |
| EPI_ISL_440892 | Liverpool Clinical Laboratories | COVID-19 Genomics UK (COG-UK) Consortium |
| EPI_ISL_440893 | Liverpool Clinical Laboratories | COVID-19 Genomics UK (COG-UK) Consortium |

| EPI_Accession_ID | Originating Laboratory | Submitting Laboratory |
|---|---|---|
| EPI_ISL_440894 | Liverpool Clinical Laboratories | COVID-19 Genomics UK (COG-UK) Consortium |
| EPI_ISL_440895 | Liverpool Clinical Laboratories | COVID-19 Genomics UK (COG-UK) Consortium |
| EPI_ISL_440896 | Liverpool Clinical Laboratories | COVID-19 Genomics UK (COG-UK) Consortium |
| EPI_ISL_440897 | Liverpool Clinical Laboratories | COVID-19 Genomics UK (COG-UK) Consortium |
| EPI_ISL_440898 | Liverpool Clinical Laboratories | COVID-19 Genomics UK (COG-UK) Consortium |
| EPI_ISL_440899 | Liverpool Clinical Laboratories | COVID-19 Genomics UK (COG-UK) Consortium |
| EPI_ISL_440900 | Liverpool Clinical Laboratories | COVID-19 Genomics UK (COG-UK) Consortium |
| EPI_ISL_440901 | Liverpool Clinical Laboratories | COVID-19 Genomics UK (COG-UK) Consortium |
| EPI_ISL_440902 | Liverpool Clinical Laboratories | COVID-19 Genomics UK (COG-UK) Consortium |
| EPI_ISL_440903 | Liverpool Clinical Laboratories | COVID-19 Genomics UK (COG-UK) Consortium |
| EPI_ISL_440904 | Liverpool Clinical Laboratories | COVID-19 Genomics UK (COG-UK) Consortium |
| EPI_ISL_440905 | Liverpool Clinical Laboratories | COVID-19 Genomics UK (COG-UK) Consortium |
| EPI_ISL_440906 | Liverpool Clinical Laboratories | COVID-19 Genomics UK (COG-UK) Consortium |
| EPI_ISL_440907 | Liverpool Clinical Laboratories | COVID-19 Genomics UK (COG-UK) Consortium |
| EPI_ISL_440908 | Liverpool Clinical Laboratories | COVID-19 Genomics UK (COG-UK) Consortium |
| EPI_ISL_440909 | Liverpool Clinical Laboratories | COVID-19 Genomics UK (COG-UK) Consortium |
| EPI_ISL_440910 | Liverpool Clinical Laboratories | COVID-19 Genomics UK (COG-UK) Consortium |
| EPI_ISL_440911 | Liverpool Clinical Laboratories | COVID-19 Genomics UK (COG-UK) Consortium |
| EPI_ISL_440912 | Liverpool Clinical Laboratories | COVID-19 Genomics UK (COG-UK) Consortium |
| EPI_ISL_440913 | Liverpool Clinical Laboratories | COVID-19 Genomics UK (COG-UK) Consortium |
| EPI_ISL_440914 | Liverpool Clinical Laboratories | COVID-19 Genomics UK (COG-UK) Consortium |
| EPI_ISL_440915 | Liverpool Clinical Laboratories | COVID-19 Genomics UK (COG-UK) Consortium |
| EPI_ISL_440916 | Liverpool Clinical Laboratories | COVID-19 Genomics UK (COG-UK) Consortium |
| EPI_ISL_440917 | Liverpool Clinical Laboratories | COVID-19 Genomics UK (COG-UK) Consortium |
| EPI_ISL_440918 | Liverpool Clinical Laboratories | COVID-19 Genomics UK (COG-UK) Consortium |
| EPI_ISL_440919 | Liverpool Clinical Laboratories | COVID-19 Genomics UK (COG-UK) Consortium |
| EPI_ISL_440920 | Liverpool Clinical Laboratories | COVID-19 Genomics UK (COG-UK) Consortium |
| EPI_ISL_440921 | Liverpool Clinical Laboratories | COVID-19 Genomics UK (COG-UK) Consortium |
| EPI_ISL_440922 | Liverpool Clinical Laboratories | COVID-19 Genomics UK (COG-UK) Consortium |
| EPI_ISL_440923 | Liverpool Clinical Laboratories | COVID-19 Genomics UK (COG-UK) Consortium |
| EPI_ISL_440924 | Liverpool Clinical Laboratories | COVID-19 Genomics UK (COG-UK) Consortium |
| EPI_ISL_440925 | Liverpool Clinical Laboratories | COVID-19 Genomics UK (COG-UK) Consortium |
| EPI_ISL_440926 | Liverpool Clinical Laboratories | COVID-19 Genomics UK (COG-UK) Consortium |

| EPI_Accession_ID | Originating Laboratory | Submitting Laboratory |
|---|---|---|
| EPI_ISL_440927 | Liverpool Clinical Laboratories | COVID-19 Genomics UK (COG-UK) Consortium |
| EPI_ISL_440928 | Liverpool Clinical Laboratories | COVID-19 Genomics UK (COG-UK) Consortium |
| EPI_ISL_440929 | Liverpool Clinical Laboratories | COVID-19 Genomics UK (COG-UK) Consortium |
| EPI_ISL_440930 | Liverpool Clinical Laboratories | COVID-19 Genomics UK (COG-UK) Consortium |
| EPI_ISL_440931 | Liverpool Clinical Laboratories | COVID-19 Genomics UK (COG-UK) Consortium |
| EPI_ISL_440932 | Liverpool Clinical Laboratories | COVID-19 Genomics UK (COG-UK) Consortium |
| EPI_ISL_440933 | Liverpool Clinical Laboratories | COVID-19 Genomics UK (COG-UK) Consortium |
| EPI_ISL_440934 | Liverpool Clinical Laboratories | COVID-19 Genomics UK (COG-UK) Consortium |
| EPI_ISL_440935 | Liverpool Clinical Laboratories | COVID-19 Genomics UK (COG-UK) Consortium |
| EPI_ISL_440936 | Liverpool Clinical Laboratories | COVID-19 Genomics UK (COG-UK) Consortium |
| EPI_ISL_440937 | Liverpool Clinical Laboratories | COVID-19 Genomics UK (COG-UK) Consortium |
| EPI_ISL_440938 | Liverpool Clinical Laboratories | COVID-19 Genomics UK (COG-UK) Consortium |
| EPI_ISL_440939 | Liverpool Clinical Laboratories | COVID-19 Genomics UK (COG-UK) Consortium |
| EPI_ISL_440940 | Liverpool Clinical Laboratories | COVID-19 Genomics UK (COG-UK) Consortium |
| EPI_ISL_440941 | Liverpool Clinical Laboratories | COVID-19 Genomics UK (COG-UK) Consortium |
| EPI_ISL_440942 | Liverpool Clinical Laboratories | COVID-19 Genomics UK (COG-UK) Consortium |
| EPI_ISL_440943 | Liverpool Clinical Laboratories | COVID-19 Genomics UK (COG-UK) Consortium |
| EPI_ISL_440944 | Liverpool Clinical Laboratories | COVID-19 Genomics UK (COG-UK) Consortium |
| EPI_ISL_440945 | Liverpool Clinical Laboratories | COVID-19 Genomics UK (COG-UK) Consortium |
| EPI_ISL_440946 | Liverpool Clinical Laboratories | COVID-19 Genomics UK (COG-UK) Consortium |
| EPI_ISL_440947 | Liverpool Clinical Laboratories | COVID-19 Genomics UK (COG-UK) Consortium |
| EPI_ISL_440948 | Liverpool Clinical Laboratories | COVID-19 Genomics UK (COG-UK) Consortium |
| EPI_ISL_440949 | Liverpool Clinical Laboratories | COVID-19 Genomics UK (COG-UK) Consortium |
| EPI_ISL_440950 | Liverpool Clinical Laboratories | COVID-19 Genomics UK (COG-UK) Consortium |
| EPI_ISL_440951 | University College London, Great Ormond Street Hospital for Children NHS Foundation Trust, Imperial College Healthcare NHS Trust | COVID-19 Genomics UK (COG-UK) Consortium |
| EPI_ISL_440952 | University College London, Great Ormond Street Hospital for Children NHS Foundation Trust, Imperial College Healthcare NHS Trust | COVID-19 Genomics UK (COG-UK) Consortium |
| EPI_ISL_440953 | University College London, Great Ormond Street Hospital for Children NHS Foundation Trust, Imperial College Healthcare NHS Trust | COVID-19 Genomics UK (COG-UK) Consortium |

| EPI_Accession_ID | Originating Laboratory | Submitting Laboratory |
|---|---|---|
| EPI_ISL_440954 | University College London, Great Ormond Street Hospital for Children NHS Foundation Trust, Imperial College Healthcare NHS Trust | COVID-19 Genomics UK (COG-UK) Consortium |
| EPI_ISL_440955 | University College London, Great Ormond Street Hospital for Children NHS Foundation Trust, Imperial College Healthcare NHS Trust | COVID-19 Genomics UK (COG-UK) Consortium |
| EPI_ISL_440956 | University College London, Great Ormond Street Hospital for Children NHS Foundation Trust, Imperial College Healthcare NHS Trust | COVID-19 Genomics UK (COG-UK) Consortium |
| EPI_ISL_440957 | University College London, Great Ormond Street Hospital for Children NHS Foundation Trust, Imperial College Healthcare NHS Trust | COVID-19 Genomics UK (COG-UK) Consortium |
| EPI_ISL_440958 | University College London, Great Ormond Street Hospital for Children NHS Foundation Trust, Imperial College Healthcare NHS Trust | COVID-19 Genomics UK (COG-UK) Consortium |
| EPI_ISL_440959 | University College London, Great Ormond Street Hospital for Children NHS Foundation Trust, Imperial College Healthcare NHS Trust | COVID-19 Genomics UK (COG-UK) Consortium |
| EPI_ISL_440960 | University College London, Great Ormond Street Hospital for Children NHS Foundation Trust, Imperial College Healthcare NHS Trust | COVID-19 Genomics UK (COG-UK) Consortium |
| EPI_ISL_440961 | University College London, Great Ormond Street Hospital for Children NHS Foundation Trust, Imperial College Healthcare NHS Trust | COVID-19 Genomics UK (COG-UK) Consortium |
| EPI_ISL_440962 | University College London, Great Ormond Street Hospital for Children NHS Foundation Trust, Imperial College Healthcare NHS Trust | COVID-19 Genomics UK (COG-UK) Consortium |
| EPI_ISL_440963 | University College London, Great Ormond Street Hospital for Children NHS Foundation Trust, Imperial College Healthcare NHS Trust | COVID-19 Genomics UK (COG-UK) Consortium |
| EPI_ISL_440964 | University College London, Great Ormond Street Hospital for Children NHS Foundation Trust, Imperial College Healthcare NHS Trust | COVID-19 Genomics UK (COG-UK) Consortium |

| EPI_Accession_ID | Originating Laboratory | Submitting Laboratory |
|---|---|---|
| EPI_ISL_440965 | University College London, Great Ormond Street Hospital for Children NHS Foundation Trust, Imperial College Healthcare NHS Trust | COVID-19 Genomics UK (COG-UK) Consortium |
| EPI_ISL_440966 | University College London, Great Ormond Street Hospital for Children NHS Foundation Trust, Imperial College Healthcare NHS Trust | COVID-19 Genomics UK (COG-UK) Consortium |
| EPI_ISL_440967 | University College London, Great Ormond Street Hospital for Children NHS Foundation Trust, Imperial College Healthcare NHS Trust | COVID-19 Genomics UK (COG-UK) Consortium |
| EPI_ISL_440968 | University College London, Great Ormond Street Hospital for Children NHS Foundation Trust, Imperial College Healthcare NHS Trust | COVID-19 Genomics UK (COG-UK) Consortium |
| EPI_ISL_440969 | University College London, Great Ormond Street Hospital for Children NHS Foundation Trust, Imperial College Healthcare NHS Trust | COVID-19 Genomics UK (COG-UK) Consortium |
| EPI_ISL_440970 | University College London, Great Ormond Street Hospital for Children NHS Foundation Trust, Imperial College Healthcare NHS Trust | COVID-19 Genomics UK (COG-UK) Consortium |
| EPI_ISL_440971 | University College London, Great Ormond Street Hospital for Children NHS Foundation Trust, Imperial College Healthcare NHS Trust | COVID-19 Genomics UK (COG-UK) Consortium |
| EPI_ISL_440972 | University College London, Great Ormond Street Hospital for Children NHS Foundation Trust, Imperial College Healthcare NHS Trust | COVID-19 Genomics UK (COG-UK) Consortium |
| EPI_ISL_440973 | University College London, Great Ormond Street Hospital for Children NHS Foundation Trust, Imperial College Healthcare NHS Trust | COVID-19 Genomics UK (COG-UK) Consortium |
| EPI_ISL_440974 | University College London, Great Ormond Street Hospital for Children NHS Foundation Trust, Imperial College Healthcare NHS Trust | COVID-19 Genomics UK (COG-UK) Consortium |
| EPI_ISL_440975 | University College London, Great Ormond Street Hospital for Children NHS Foundation Trust, Imperial College Healthcare NHS Trust | COVID-19 Genomics UK (COG-UK) Consortium |

| EPI_Accession_ID | Originating Laboratory | Submitting Laboratory |
|---|---|---|
| EPI_ISL_440976 | University College London, Great Ormond Street Hospital for Children NHS Foundation Trust, Imperial College Healthcare NHS Trust | COVID-19 Genomics UK (COG-UK) Consortium |
| EPI_ISL_440977 | University College London, Great Ormond Street Hospital for Children NHS Foundation Trust, Imperial College Healthcare NHS Trust | COVID-19 Genomics UK (COG-UK) Consortium |
| EPI_ISL_440978 | University College London, Great Ormond Street Hospital for Children NHS Foundation Trust, Imperial College Healthcare NHS Trust | COVID-19 Genomics UK (COG-UK) Consortium |
| EPI_ISL_440979 | University College London, Great Ormond Street Hospital for Children NHS Foundation Trust, Imperial College Healthcare NHS Trust | COVID-19 Genomics UK (COG-UK) Consortium |
| EPI_ISL_440980 | University College London, Great Ormond Street Hospital for Children NHS Foundation Trust, Imperial College Healthcare NHS Trust | COVID-19 Genomics UK (COG-UK) Consortium |
| EPI_ISL_440981 | University College London, Great Ormond Street Hospital for Children NHS Foundation Trust, Imperial College Healthcare NHS Trust | COVID-19 Genomics UK (COG-UK) Consortium |
| EPI_ISL_440982 | University College London, Great Ormond Street Hospital for Children NHS Foundation Trust, Imperial College Healthcare NHS Trust | COVID-19 Genomics UK (COG-UK) Consortium |
| EPI_ISL_440983 | University College London, Great Ormond Street Hospital for Children NHS Foundation Trust, Imperial College Healthcare NHS Trust | COVID-19 Genomics UK (COG-UK) Consortium |
| EPI_ISL_440984 | University College London, Great Ormond Street Hospital for Children NHS Foundation Trust, Imperial College Healthcare NHS Trust | COVID-19 Genomics UK (COG-UK) Consortium |
| EPI_ISL_440985 | University College London, Great Ormond Street Hospital for Children NHS Foundation Trust, Imperial College Healthcare NHS Trust | COVID-19 Genomics UK (COG-UK) Consortium |
| EPI_ISL_440986 | University College London, Great Ormond Street Hospital for Children NHS Foundation Trust, Imperial College Healthcare NHS Trust | COVID-19 Genomics UK (COG-UK) Consortium |

| EPI_Accession_ID | Originating Laboratory | Submitting Laboratory |
|---|---|---|
| EPI_ISL_440987 | University College London, Great Ormond Street Hospital for Children NHS Foundation Trust, Imperial College Healthcare NHS Trust | COVID-19 Genomics UK (COG-UK) Consortium |
| EPI_ISL_440988 | University College London, Great Ormond Street Hospital for Children NHS Foundation Trust, Imperial College Healthcare NHS Trust | COVID-19 Genomics UK (COG-UK) Consortium |
| EPI_ISL_440989 | University College London, Great Ormond Street Hospital for Children NHS Foundation Trust, Imperial College Healthcare NHS Trust | COVID-19 Genomics UK (COG-UK) Consortium |
| EPI_ISL_440990 | University College London, Great Ormond Street Hospital for Children NHS Foundation Trust, Imperial College Healthcare NHS Trust | COVID-19 Genomics UK (COG-UK) Consortium |
| EPI_ISL_440991 | University College London, Great Ormond Street Hospital for Children NHS Foundation Trust, Imperial College Healthcare NHS Trust | COVID-19 Genomics UK (COG-UK) Consortium |
| EPI_ISL_440992 | University College London, Great Ormond Street Hospital for Children NHS Foundation Trust, Imperial College Healthcare NHS Trust | COVID-19 Genomics UK (COG-UK) Consortium |
| EPI_ISL_440993 | University College London, Great Ormond Street Hospital for Children NHS Foundation Trust, Imperial College Healthcare NHS Trust | COVID-19 Genomics UK (COG-UK) Consortium |
| EPI_ISL_440994 | University College London, Great Ormond Street Hospital for Children NHS Foundation Trust, Imperial College Healthcare NHS Trust | COVID-19 Genomics UK (COG-UK) Consortium |
| EPI_ISL_440995 | University College London, Great Ormond Street Hospital for Children NHS Foundation Trust, Imperial College Healthcare NHS Trust | COVID-19 Genomics UK (COG-UK) Consortium |
| EPI_ISL_440996 | University College London, Great Ormond Street Hospital for Children NHS Foundation Trust, Imperial College Healthcare NHS Trust | COVID-19 Genomics UK (COG-UK) Consortium |
| EPI_ISL_440997 | University College London, Great Ormond Street Hospital for Children NHS Foundation Trust, Imperial College Healthcare NHS Trust | COVID-19 Genomics UK (COG-UK) Consortium |

| EPI_Accession_ID | Originating Laboratory | Submitting Laboratory |
| --- | --- | --- |
| EPI_ISL_440998 | University College London, Great Ormond Street Hospital for Children NHS Foundation Trust, Imperial College Healthcare NHS Trust | COVID-19 Genomics UK (COG-UK) Consortium |
| EPI_ISL_440999 | University College London, Great Ormond Street Hospital for Children NHS Foundation Trust, Imperial College Healthcare NHS Trust | COVID-19 Genomics UK (COG-UK) Consortium |
| EPI_ISL_441000 | University College London, Great Ormond Street Hospital for Children NHS Foundation Trust, Imperial College Healthcare NHS Trust | COVID-19 Genomics UK (COG-UK) Consortium |
| EPI_ISL_441001 | University College London, Great Ormond Street Hospital for Children NHS Foundation Trust, Imperial College Healthcare NHS Trust | COVID-19 Genomics UK (COG-UK) Consortium |
| EPI_ISL_441002 | University College London, Great Ormond Street Hospital for Children NHS Foundation Trust, Imperial College Healthcare NHS Trust | COVID-19 Genomics UK (COG-UK) Consortium |
| EPI_ISL_441003 | University College London, Great Ormond Street Hospital for Children NHS Foundation Trust, Imperial College Healthcare NHS Trust | COVID-19 Genomics UK (COG-UK) Consortium |
| EPI_ISL_441004 | University College London, Great Ormond Street Hospital for Children NHS Foundation Trust, Imperial College Healthcare NHS Trust | COVID-19 Genomics UK (COG-UK) Consortium |
| EPI_ISL_441005 | University College London, Great Ormond Street Hospital for Children NHS Foundation Trust, Imperial College Healthcare NHS Trust | COVID-19 Genomics UK (COG-UK) Consortium |
| EPI_ISL_441006 | University College London, Great Ormond Street Hospital for Children NHS Foundation Trust, Imperial College Healthcare NHS Trust | COVID-19 Genomics UK (COG-UK) Consortium |
| EPI_ISL_441007 | University College London, Great Ormond Street Hospital for Children NHS Foundation Trust, Imperial College Healthcare NHS Trust | COVID-19 Genomics UK (COG-UK) Consortium |
| EPI_ISL_441008 | University College London, Great Ormond Street Hospital for Children NHS Foundation Trust, Imperial College Healthcare NHS Trust | COVID-19 Genomics UK (COG-UK) Consortium |

| EPI_Accession_ID | Originating Laboratory | Submitting Laboratory |
|---|---|---|
| EPI_ISL_441009 | University College London, Great Ormond Street Hospital for Children NHS Foundation Trust, Imperial College Healthcare NHS Trust | COVID-19 Genomics UK (COG-UK) Consortium |
| EPI_ISL_441010 | University College London, Great Ormond Street Hospital for Children NHS Foundation Trust, Imperial College Healthcare NHS Trust | COVID-19 Genomics UK (COG-UK) Consortium |
| EPI_ISL_441011 | University College London, Great Ormond Street Hospital for Children NHS Foundation Trust, Imperial College Healthcare NHS Trust | COVID-19 Genomics UK (COG-UK) Consortium |
| EPI_ISL_441012 | University College London, Great Ormond Street Hospital for Children NHS Foundation Trust, Imperial College Healthcare NHS Trust | COVID-19 Genomics UK (COG-UK) Consortium |
| EPI_ISL_441013 | University College London, Great Ormond Street Hospital for Children NHS Foundation Trust, Imperial College Healthcare NHS Trust | COVID-19 Genomics UK (COG-UK) Consortium |
| EPI_ISL_441014 | University College London, Great Ormond Street Hospital for Children NHS Foundation Trust, Imperial College Healthcare NHS Trust | COVID-19 Genomics UK (COG-UK) Consortium |
| EPI_ISL_441015 | University College London, Great Ormond Street Hospital for Children NHS Foundation Trust, Imperial College Healthcare NHS Trust | COVID-19 Genomics UK (COG-UK) Consortium |
| EPI_ISL_441016 | University College London, Great Ormond Street Hospital for Children NHS Foundation Trust, Imperial College Healthcare NHS Trust | COVID-19 Genomics UK (COG-UK) Consortium |
| EPI_ISL_441017 | University College London, Great Ormond Street Hospital for Children NHS Foundation Trust, Imperial College Healthcare NHS Trust | COVID-19 Genomics UK (COG-UK) Consortium |
| EPI_ISL_441018 | University College London, Great Ormond Street Hospital for Children NHS Foundation Trust, Imperial College Healthcare NHS Trust | COVID-19 Genomics UK (COG-UK) Consortium |
| EPI_ISL_441019 | University College London, Great Ormond Street Hospital for Children NHS Foundation Trust, Imperial College Healthcare NHS Trust | COVID-19 Genomics UK (COG-UK) Consortium |

| EPI_Accession_ID | Originating Laboratory | Submitting Laboratory |
|---|---|---|
| EPI_ISL_441020 | University College London, Great Ormond Street Hospital for Children NHS Foundation Trust, Imperial College Healthcare NHS Trust | COVID-19 Genomics UK (COG-UK) Consortium |
| EPI_ISL_441021 | University College London, Great Ormond Street Hospital for Children NHS Foundation Trust, Imperial College Healthcare NHS Trust | COVID-19 Genomics UK (COG-UK) Consortium |
| EPI_ISL_441022 | University College London, Great Ormond Street Hospital for Children NHS Foundation Trust, Imperial College Healthcare NHS Trust | COVID-19 Genomics UK (COG-UK) Consortium |
| EPI_ISL_441023 | University College London, Great Ormond Street Hospital for Children NHS Foundation Trust, Imperial College Healthcare NHS Trust | COVID-19 Genomics UK (COG-UK) Consortium |
| EPI_ISL_441024 | University College London, Great Ormond Street Hospital for Children NHS Foundation Trust, Imperial College Healthcare NHS Trust | COVID-19 Genomics UK (COG-UK) Consortium |
| EPI_ISL_441025 | University College London, Great Ormond Street Hospital for Children NHS Foundation Trust, Imperial College Healthcare NHS Trust | COVID-19 Genomics UK (COG-UK) Consortium |
| EPI_ISL_441026 | University College London, Great Ormond Street Hospital for Children NHS Foundation Trust, Imperial College Healthcare NHS Trust | COVID-19 Genomics UK (COG-UK) Consortium |
| EPI_ISL_441027 | University College London, Great Ormond Street Hospital for Children NHS Foundation Trust, Imperial College Healthcare NHS Trust | COVID-19 Genomics UK (COG-UK) Consortium |
| EPI_ISL_441028 | University College London, Great Ormond Street Hospital for Children NHS Foundation Trust, Imperial College Healthcare NHS Trust | COVID-19 Genomics UK (COG-UK) Consortium |
| EPI_ISL_441029 | University College London, Great Ormond Street Hospital for Children NHS Foundation Trust, Imperial College Healthcare NHS Trust | COVID-19 Genomics UK (COG-UK) Consortium |
| EPI_ISL_441030 | University College London, Great Ormond Street Hospital for Children NHS Foundation Trust, Imperial College Healthcare NHS Trust | COVID-19 Genomics UK (COG-UK) Consortium |

| EPI_Accession_ID | Originating Laboratory | Submitting Laboratory |
|---|---|---|
| EPI_ISL_441031 | University College London, Great Ormond Street Hospital for Children NHS Foundation Trust, Imperial College Healthcare NHS Trust | COVID-19 Genomics UK (COG-UK) Consortium |
| EPI_ISL_441032 | University College London, Great Ormond Street Hospital for Children NHS Foundation Trust, Imperial College Healthcare NHS Trust | COVID-19 Genomics UK (COG-UK) Consortium |
| EPI_ISL_441033 | University College London, Great Ormond Street Hospital for Children NHS Foundation Trust, Imperial College Healthcare NHS Trust | COVID-19 Genomics UK (COG-UK) Consortium |
| EPI_ISL_441034 | University College London, Great Ormond Street Hospital for Children NHS Foundation Trust, Imperial College Healthcare NHS Trust | COVID-19 Genomics UK (COG-UK) Consortium |
| EPI_ISL_441035 | University College London, Great Ormond Street Hospital for Children NHS Foundation Trust, Imperial College Healthcare NHS Trust | COVID-19 Genomics UK (COG-UK) Consortium |
| EPI_ISL_441036 | University College London, Great Ormond Street Hospital for Children NHS Foundation Trust, Imperial College Healthcare NHS Trust | COVID-19 Genomics UK (COG-UK) Consortium |
| EPI_ISL_441037 | University College London, Great Ormond Street Hospital for Children NHS Foundation Trust, Imperial College Healthcare NHS Trust | COVID-19 Genomics UK (COG-UK) Consortium |
| EPI_ISL_441038 | University College London, Great Ormond Street Hospital for Children NHS Foundation Trust, Imperial College Healthcare NHS Trust | COVID-19 Genomics UK (COG-UK) Consortium |
| EPI_ISL_441039 | University College London, Great Ormond Street Hospital for Children NHS Foundation Trust, Imperial College Healthcare NHS Trust | COVID-19 Genomics UK (COG-UK) Consortium |
| EPI_ISL_441040 | University College London, Great Ormond Street Hospital for Children NHS Foundation Trust, Imperial College Healthcare NHS Trust | COVID-19 Genomics UK (COG-UK) Consortium |
| EPI_ISL_441041 | University College London, Great Ormond Street Hospital for Children NHS Foundation Trust, Imperial College Healthcare NHS Trust | COVID-19 Genomics UK (COG-UK) Consortium |

| EPI_Accession_ID | Originating Laboratory | Submitting Laboratory |
|---|---|---|
| EPI_ISL_441042 | University College London, Great Ormond Street Hospital for Children NHS Foundation Trust, Imperial College Healthcare NHS Trust | COVID-19 Genomics UK (COG-UK) Consortium |
| EPI_ISL_441043 | University College London, Great Ormond Street Hospital for Children NHS Foundation Trust, Imperial College Healthcare NHS Trust | COVID-19 Genomics UK (COG-UK) Consortium |
| EPI_ISL_441044 | University College London, Great Ormond Street Hospital for Children NHS Foundation Trust, Imperial College Healthcare NHS Trust | COVID-19 Genomics UK (COG-UK) Consortium |
| EPI_ISL_441045 | University College London, Great Ormond Street Hospital for Children NHS Foundation Trust, Imperial College Healthcare NHS Trust | COVID-19 Genomics UK (COG-UK) Consortium |
| EPI_ISL_441046 | University College London, Great Ormond Street Hospital for Children NHS Foundation Trust, Imperial College Healthcare NHS Trust | COVID-19 Genomics UK (COG-UK) Consortium |
| EPI_ISL_441047 | University College London, Great Ormond Street Hospital for Children NHS Foundation Trust, Imperial College Healthcare NHS Trust | COVID-19 Genomics UK (COG-UK) Consortium |
| EPI_ISL_441048 | University College London, Great Ormond Street Hospital for Children NHS Foundation Trust, Imperial College Healthcare NHS Trust | COVID-19 Genomics UK (COG-UK) Consortium |
| EPI_ISL_441049 | University College London, Great Ormond Street Hospital for Children NHS Foundation Trust, Imperial College Healthcare NHS Trust | COVID-19 Genomics UK (COG-UK) Consortium |
| EPI_ISL_441050 | University College London, Great Ormond Street Hospital for Children NHS Foundation Trust, Imperial College Healthcare NHS Trust | COVID-19 Genomics UK (COG-UK) Consortium |
| EPI_ISL_441051 | University College London, Great Ormond Street Hospital for Children NHS Foundation Trust, Imperial College Healthcare NHS Trust | COVID-19 Genomics UK (COG-UK) Consortium |
| EPI_ISL_441052 | Department of Pathology, University of Cambridge | Wellcome Sanger Institute for the COVID-19 Genomics UK Consortium |

| EPI_Accession_ID | Originating Laboratory | Submitting Laboratory |
|---|---|---|
| EPI_ISL_441053 | Department of Pathology, University of Cambridge | Wellcome Sanger Institute for the COVID-19 Genomics UK Consortium |
| EPI_ISL_441054 | Department of Pathology, University of Cambridge | Wellcome Sanger Institute for the COVID-19 Genomics UK Consortium |
| EPI_ISL_441055 | Department of Pathology, University of Cambridge | Wellcome Sanger Institute for the COVID-19 Genomics UK Consortium |
| EPI_ISL_441056 | Department of Pathology, University of Cambridge | Wellcome Sanger Institute for the COVID-19 Genomics UK Consortium |
| EPI_ISL_441057 | Department of Pathology, University of Cambridge | Wellcome Sanger Institute for the COVID-19 Genomics UK Consortium |
| EPI_ISL_441058 | Department of Pathology, University of Cambridge | Wellcome Sanger Institute for the COVID-19 Genomics UK Consortium |
| EPI_ISL_441059 | Department of Pathology, University of Cambridge | Wellcome Sanger Institute for the COVID-19 Genomics UK Consortium |
| EPI_ISL_441060 | Department of Pathology, University of Cambridge | Wellcome Sanger Institute for the COVID-19 Genomics UK Consortium |
| EPI_ISL_441061 | Department of Pathology, University of Cambridge | Wellcome Sanger Institute for the COVID-19 Genomics UK Consortium |
| EPI_ISL_441062 | Department of Pathology, University of Cambridge | Wellcome Sanger Institute for the COVID-19 Genomics UK Consortium |
| EPI_ISL_441063 | Department of Pathology, University of Cambridge | Wellcome Sanger Institute for the COVID-19 Genomics UK Consortium |
| EPI_ISL_441064 | Department of Pathology, University of Cambridge | Wellcome Sanger Institute for the COVID-19 Genomics UK Consortium |
| EPI_ISL_441065 | Department of Pathology, University of Cambridge | Wellcome Sanger Institute for the COVID-19 Genomics UK Consortium |
| EPI_ISL_441066 | Department of Pathology, University of Cambridge | Wellcome Sanger Institute for the COVID-19 Genomics UK Consortium |
| EPI_ISL_441067 | Department of Pathology, University of Cambridge | Wellcome Sanger Institute for the COVID-19 Genomics UK Consortium |
| EPI_ISL_441068 | Department of Pathology, University of Cambridge | Wellcome Sanger Institute for the COVID-19 Genomics UK Consortium |

| EPI_Accession_ID | Originating Laboratory | Submitting Laboratory |
|---|---|---|
| EPI_ISL_441069 | Department of Pathology, University of Cambridge | Wellcome Sanger Institute for the COVID-19 Genomics UK Consortium |
| EPI_ISL_441070 | Department of Pathology, University of Cambridge | Wellcome Sanger Institute for the COVID-19 Genomics UK Consortium |
| EPI_ISL_441071 | Department of Pathology, University of Cambridge | Wellcome Sanger Institute for the COVID-19 Genomics UK Consortium |
| EPI_ISL_441072 | Department of Pathology, University of Cambridge | Wellcome Sanger Institute for the COVID-19 Genomics UK Consortium |
| EPI_ISL_441073 | Department of Pathology, University of Cambridge | Wellcome Sanger Institute for the COVID-19 Genomics UK Consortium |
| EPI_ISL_441074 | Department of Pathology, University of Cambridge | Wellcome Sanger Institute for the COVID-19 Genomics UK Consortium |
| EPI_ISL_441075 | Department of Pathology, University of Cambridge | Wellcome Sanger Institute for the COVID-19 Genomics UK Consortium |
| EPI_ISL_441076 | Department of Pathology, University of Cambridge | Wellcome Sanger Institute for the COVID-19 Genomics UK Consortium |
| EPI_ISL_441077 | Department of Pathology, University of Cambridge | Wellcome Sanger Institute for the COVID-19 Genomics UK Consortium |
| EPI_ISL_441078 | Department of Pathology, University of Cambridge | Wellcome Sanger Institute for the COVID-19 Genomics UK Consortium |
| EPI_ISL_441079 | Department of Pathology, University of Cambridge | Wellcome Sanger Institute for the COVID-19 Genomics UK Consortium |
| EPI_ISL_441080 | Department of Pathology, University of Cambridge | Wellcome Sanger Institute for the COVID-19 Genomics UK Consortium |
| EPI_ISL_441081 | Department of Pathology, University of Cambridge | Wellcome Sanger Institute for the COVID-19 Genomics UK Consortium |
| EPI_ISL_441082 | Department of Pathology, University of Cambridge | Wellcome Sanger Institute for the COVID-19 Genomics UK Consortium |
| EPI_ISL_441083 | Department of Pathology, University of Cambridge | Wellcome Sanger Institute for the COVID-19 Genomics UK Consortium |
| EPI_ISL_441084 | Department of Pathology, University of Cambridge | Wellcome Sanger Institute for the COVID-19 Genomics UK Consortium |

| EPI_Accession_ID | Originating Laboratory | Submitting Laboratory |
|---|---|---|
| EPI_ISL_441085 | Department of Pathology, University of Cambridge | Wellcome Sanger Institute for the COVID-19 Genomics UK Consortium |
| EPI_ISL_441086 | Department of Pathology, University of Cambridge | Wellcome Sanger Institute for the COVID-19 Genomics UK Consortium |
| EPI_ISL_441087 | Department of Pathology, University of Cambridge | Wellcome Sanger Institute for the COVID-19 Genomics UK Consortium |
| EPI_ISL_441088 | Department of Pathology, University of Cambridge | Wellcome Sanger Institute for the COVID-19 Genomics UK Consortium |
| EPI_ISL_441089 | Department of Pathology, University of Cambridge | Wellcome Sanger Institute for the COVID-19 Genomics UK Consortium |
| EPI_ISL_441090 | Department of Pathology, University of Cambridge | Wellcome Sanger Institute for the COVID-19 Genomics UK Consortium |
| EPI_ISL_441091 | Department of Pathology, University of Cambridge | Wellcome Sanger Institute for the COVID-19 Genomics UK Consortium |
| EPI_ISL_441092 | Department of Pathology, University of Cambridge | Wellcome Sanger Institute for the COVID-19 Genomics UK Consortium |
| EPI_ISL_441093 | Department of Pathology, University of Cambridge | Wellcome Sanger Institute for the COVID-19 Genomics UK Consortium |
| EPI_ISL_441094 | Department of Pathology, University of Cambridge | Wellcome Sanger Institute for the COVID-19 Genomics UK Consortium |
| EPI_ISL_441095 | Department of Pathology, University of Cambridge | Wellcome Sanger Institute for the COVID-19 Genomics UK Consortium |
| EPI_ISL_441096 | Department of Pathology, University of Cambridge | Wellcome Sanger Institute for the COVID-19 Genomics UK Consortium |
| EPI_ISL_441097 | Department of Pathology, University of Cambridge | Wellcome Sanger Institute for the COVID-19 Genomics UK Consortium |
| EPI_ISL_441098 | Department of Pathology, University of Cambridge | Wellcome Sanger Institute for the COVID-19 Genomics UK Consortium |
| EPI_ISL_441099 | Department of Pathology, University of Cambridge | Wellcome Sanger Institute for the COVID-19 Genomics UK Consortium |
| EPI_ISL_441100 | Department of Pathology, University of Cambridge | Wellcome Sanger Institute for the COVID-19 Genomics UK Consortium |

| EPI_Accession_ID | Originating Laboratory | Submitting Laboratory |
| --- | --- | --- |
| EPI_ISL_441101 | Department of Pathology, University of Cambridge | Wellcome Sanger Institute for the COVID-19 Genomics UK Consortium |
| EPI_ISL_441102 | Department of Pathology, University of Cambridge | Wellcome Sanger Institute for the COVID-19 Genomics UK Consortium |
| EPI_ISL_441103 | Department of Pathology, University of Cambridge | Wellcome Sanger Institute for the COVID-19 Genomics UK Consortium |
| EPI_ISL_441104 | Department of Pathology, University of Cambridge | Wellcome Sanger Institute for the COVID-19 Genomics UK Consortium |
| EPI_ISL_441105 | Department of Pathology, University of Cambridge | Wellcome Sanger Institute for the COVID-19 Genomics UK Consortium |
| EPI_ISL_441106 | Department of Pathology, University of Cambridge | Wellcome Sanger Institute for the COVID-19 Genomics UK Consortium |
| EPI_ISL_441107 | Department of Pathology, University of Cambridge | Wellcome Sanger Institute for the COVID-19 Genomics UK Consortium |
| EPI_ISL_441108 | Department of Pathology, University of Cambridge | Wellcome Sanger Institute for the COVID-19 Genomics UK Consortium |
| EPI_ISL_441109 | Department of Pathology, University of Cambridge | Wellcome Sanger Institute for the COVID-19 Genomics UK Consortium |
| EPI_ISL_441110 | Department of Pathology, University of Cambridge | Wellcome Sanger Institute for the COVID-19 Genomics UK Consortium |
| EPI_ISL_441111 | Department of Pathology, University of Cambridge | Wellcome Sanger Institute for the COVID-19 Genomics UK Consortium |
| EPI_ISL_441112 | Department of Pathology, University of Cambridge | Wellcome Sanger Institute for the COVID-19 Genomics UK Consortium |
| EPI_ISL_441113 | Department of Pathology, University of Cambridge | Wellcome Sanger Institute for the COVID-19 Genomics UK Consortium |
| EPI_ISL_441114 | Department of Pathology, University of Cambridge | Wellcome Sanger Institute for the COVID-19 Genomics UK Consortium |
| EPI_ISL_441115 | Department of Pathology, University of Cambridge | Wellcome Sanger Institute for the COVID-19 Genomics UK Consortium |
| EPI_ISL_441116 | Department of Pathology, University of Cambridge | Wellcome Sanger Institute for the COVID-19 Genomics UK Consortium |

| EPI_Accession_ID | Originating Laboratory | Submitting Laboratory |
|---|---|---|
| EPI_ISL_441117 | Department of Pathology, University of Cambridge | Wellcome Sanger Institute for the COVID-19 Genomics UK Consortium |
| EPI_ISL_441118 | Department of Pathology, University of Cambridge | Wellcome Sanger Institute for the COVID-19 Genomics UK Consortium |
| EPI_ISL_441119 | Department of Pathology, University of Cambridge | Wellcome Sanger Institute for the COVID-19 Genomics UK Consortium |
| EPI_ISL_441120 | Department of Pathology, University of Cambridge | Wellcome Sanger Institute for the COVID-19 Genomics UK Consortium |
| EPI_ISL_441121 | Department of Pathology, University of Cambridge | Wellcome Sanger Institute for the COVID-19 Genomics UK Consortium |
| EPI_ISL_441122 | Department of Pathology, University of Cambridge | Wellcome Sanger Institute for the COVID-19 Genomics UK Consortium |
| EPI_ISL_441123 | Department of Pathology, University of Cambridge | Wellcome Sanger Institute for the COVID-19 Genomics UK Consortium |
| EPI_ISL_441124 | Department of Pathology, University of Cambridge | Wellcome Sanger Institute for the COVID-19 Genomics UK Consortium |
| EPI_ISL_441125 | Department of Pathology, University of Cambridge | Wellcome Sanger Institute for the COVID-19 Genomics UK Consortium |
| EPI_ISL_441126 | Department of Pathology, University of Cambridge | Wellcome Sanger Institute for the COVID-19 Genomics UK Consortium |
| EPI_ISL_441127 | Department of Pathology, University of Cambridge | Wellcome Sanger Institute for the COVID-19 Genomics UK Consortium |
| EPI_ISL_441128 | Department of Pathology, University of Cambridge | Wellcome Sanger Institute for the COVID-19 Genomics UK Consortium |
| EPI_ISL_441129 | Department of Pathology, University of Cambridge | Wellcome Sanger Institute for the COVID-19 Genomics UK Consortium |
| EPI_ISL_441130 | Department of Pathology, University of Cambridge | Wellcome Sanger Institute for the COVID-19 Genomics UK Consortium |
| EPI_ISL_441131 | Department of Pathology, University of Cambridge | Wellcome Sanger Institute for the COVID-19 Genomics UK Consortium |
| EPI_ISL_441132 | Department of Pathology, University of Cambridge | Wellcome Sanger Institute for the COVID-19 Genomics UK Consortium |

| EPI_Accession_ID | Originating Laboratory | Submitting Laboratory |
|---|---|---|
| EPI_ISL_441133 | Department of Pathology, University of Cambridge | Wellcome Sanger Institute for the COVID-19 Genomics UK Consortium |
| EPI_ISL_441134 | Department of Pathology, University of Cambridge | Wellcome Sanger Institute for the COVID-19 Genomics UK Consortium |
| EPI_ISL_441135 | Department of Pathology, University of Cambridge | Wellcome Sanger Institute for the COVID-19 Genomics UK Consortium |
| EPI_ISL_441136 | Department of Pathology, University of Cambridge | Wellcome Sanger Institute for the COVID-19 Genomics UK Consortium |
| EPI_ISL_441137 | Department of Pathology, University of Cambridge | Wellcome Sanger Institute for the COVID-19 Genomics UK Consortium |
| EPI_ISL_441138 | Department of Pathology, University of Cambridge | Wellcome Sanger Institute for the COVID-19 Genomics UK Consortium |
| EPI_ISL_441139 | Department of Pathology, University of Cambridge | Wellcome Sanger Institute for the COVID-19 Genomics UK Consortium |
| EPI_ISL_441140 | Department of Pathology, University of Cambridge | Wellcome Sanger Institute for the COVID-19 Genomics UK Consortium |
| EPI_ISL_441141 | Department of Pathology, University of Cambridge | Wellcome Sanger Institute for the COVID-19 Genomics UK Consortium |
| EPI_ISL_441142 | Department of Pathology, University of Cambridge | Wellcome Sanger Institute for the COVID-19 Genomics UK Consortium |
| EPI_ISL_441143 | Department of Pathology, University of Cambridge | Wellcome Sanger Institute for the COVID-19 Genomics UK Consortium |
| EPI_ISL_441144 | Department of Pathology, University of Cambridge | Wellcome Sanger Institute for the COVID-19 Genomics UK Consortium |
| EPI_ISL_441145 | Department of Pathology, University of Cambridge | Wellcome Sanger Institute for the COVID-19 Genomics UK Consortium |
| EPI_ISL_441146 | Department of Pathology, University of Cambridge | Wellcome Sanger Institute for the COVID-19 Genomics UK Consortium |
| EPI_ISL_441147 | Department of Pathology, University of Cambridge | Wellcome Sanger Institute for the COVID-19 Genomics UK Consortium |
| EPI_ISL_441148 | Department of Pathology, University of Cambridge | Wellcome Sanger Institute for the COVID-19 Genomics UK Consortium |

| EPI_Accession_ID | Originating Laboratory | Submitting Laboratory |
|---|---|---|
| EPI_ISL_441149 | Department of Pathology, University of Cambridge | Wellcome Sanger Institute for the COVID-19 Genomics UK Consortium |
| EPI_ISL_441150 | Department of Pathology, University of Cambridge | Wellcome Sanger Institute for the COVID-19 Genomics UK Consortium |
| EPI_ISL_441151 | Department of Pathology, University of Cambridge | Wellcome Sanger Institute for the COVID-19 Genomics UK Consortium |
| EPI_ISL_441152 | Department of Pathology, University of Cambridge | Wellcome Sanger Institute for the COVID-19 Genomics UK Consortium |
| EPI_ISL_441153 | Department of Pathology, University of Cambridge | Wellcome Sanger Institute for the COVID-19 Genomics UK Consortium |
| EPI_ISL_441154 | Department of Pathology, University of Cambridge | Wellcome Sanger Institute for the COVID-19 Genomics UK Consortium |
| EPI_ISL_441155 | Department of Pathology, University of Cambridge | Wellcome Sanger Institute for the COVID-19 Genomics UK Consortium |
| EPI_ISL_441156 | Department of Pathology, University of Cambridge | Wellcome Sanger Institute for the COVID-19 Genomics UK Consortium |
| EPI_ISL_441157 | Department of Pathology, University of Cambridge | Wellcome Sanger Institute for the COVID-19 Genomics UK Consortium |
| EPI_ISL_441158 | Department of Pathology, University of Cambridge | Wellcome Sanger Institute for the COVID-19 Genomics UK Consortium |
| EPI_ISL_441159 | Department of Pathology, University of Cambridge | Wellcome Sanger Institute for the COVID-19 Genomics UK Consortium |
| EPI_ISL_441160 | Department of Pathology, University of Cambridge | Wellcome Sanger Institute for the COVID-19 Genomics UK Consortium |
| EPI_ISL_441161 | Department of Pathology, University of Cambridge | Wellcome Sanger Institute for the COVID-19 Genomics UK Consortium |
| EPI_ISL_441162 | Department of Pathology, University of Cambridge | Wellcome Sanger Institute for the COVID-19 Genomics UK Consortium |
| EPI_ISL_441163 | Department of Pathology, University of Cambridge | Wellcome Sanger Institute for the COVID-19 Genomics UK Consortium |
| EPI_ISL_441164 | Department of Pathology, University of Cambridge | Wellcome Sanger Institute for the COVID-19 Genomics UK Consortium |

| EPI_Accession_ID | Originating Laboratory | Submitting Laboratory |
|---|---|---|
| EPI_ISL_441165 | Department of Pathology, University of Cambridge | Wellcome Sanger Institute for the COVID-19 Genomics UK Consortium |
| EPI_ISL_441166 | Department of Pathology, University of Cambridge | Wellcome Sanger Institute for the COVID-19 Genomics UK Consortium |
| EPI_ISL_441167 | Department of Pathology, University of Cambridge | Wellcome Sanger Institute for the COVID-19 Genomics UK Consortium |
| EPI_ISL_441168 | Department of Pathology, University of Cambridge | Wellcome Sanger Institute for the COVID-19 Genomics UK Consortium |
| EPI_ISL_441169 | Department of Pathology, University of Cambridge | Wellcome Sanger Institute for the COVID-19 Genomics UK Consortium |
| EPI_ISL_441170 | Department of Pathology, University of Cambridge | Wellcome Sanger Institute for the COVID-19 Genomics UK Consortium |
| EPI_ISL_441171 | Department of Pathology, University of Cambridge | Wellcome Sanger Institute for the COVID-19 Genomics UK Consortium |
| EPI_ISL_441172 | Department of Pathology, University of Cambridge | Wellcome Sanger Institute for the COVID-19 Genomics UK Consortium |
| EPI_ISL_441173 | Department of Pathology, University of Cambridge | Wellcome Sanger Institute for the COVID-19 Genomics UK Consortium |
| EPI_ISL_441174 | Department of Pathology, University of Cambridge | Wellcome Sanger Institute for the COVID-19 Genomics UK Consortium |
| EPI_ISL_441175 | Department of Pathology, University of Cambridge | Wellcome Sanger Institute for the COVID-19 Genomics UK Consortium |
| EPI_ISL_441176 | Department of Pathology, University of Cambridge | Wellcome Sanger Institute for the COVID-19 Genomics UK Consortium |
| EPI_ISL_441177 | Department of Pathology, University of Cambridge | Wellcome Sanger Institute for the COVID-19 Genomics UK Consortium |
| EPI_ISL_441178 | Department of Pathology, University of Cambridge | Wellcome Sanger Institute for the COVID-19 Genomics UK Consortium |
| EPI_ISL_441179 | Department of Pathology, University of Cambridge | Wellcome Sanger Institute for the COVID-19 Genomics UK Consortium |
| EPI_ISL_441180 | Department of Pathology, University of Cambridge | Wellcome Sanger Institute for the COVID-19 Genomics UK Consortium |

| EPI_Accession_ID | Originating Laboratory | Submitting Laboratory |
| --- | --- | --- |
| EPI_ISL_441181 | Department of Pathology, University of Cambridge | Wellcome Sanger Institute for the COVID-19 Genomics UK Consortium |
| EPI_ISL_441182 | Department of Pathology, University of Cambridge | Wellcome Sanger Institute for the COVID-19 Genomics UK Consortium |
| EPI_ISL_441183 | Department of Pathology, University of Cambridge | Wellcome Sanger Institute for the COVID-19 Genomics UK Consortium |
| EPI_ISL_441184 | Department of Pathology, University of Cambridge | Wellcome Sanger Institute for the COVID-19 Genomics UK Consortium |
| EPI_ISL_441185 | Department of Pathology, University of Cambridge | Wellcome Sanger Institute for the COVID-19 Genomics UK Consortium |
| EPI_ISL_441186 | Department of Pathology, University of Cambridge | Wellcome Sanger Institute for the COVID-19 Genomics UK Consortium |
| EPI_ISL_441187 | Department of Pathology, University of Cambridge | Wellcome Sanger Institute for the COVID-19 Genomics UK Consortium |
| EPI_ISL_441188 | Department of Pathology, University of Cambridge | Wellcome Sanger Institute for the COVID-19 Genomics UK Consortium |
| EPI_ISL_441189 | Department of Pathology, University of Cambridge | Wellcome Sanger Institute for the COVID-19 Genomics UK Consortium |
| EPI_ISL_441190 | Department of Pathology, University of Cambridge | Wellcome Sanger Institute for the COVID-19 Genomics UK Consortium |
| EPI_ISL_441191 | Department of Pathology, University of Cambridge | Wellcome Sanger Institute for the COVID-19 Genomics UK Consortium |
| EPI_ISL_441192 | Department of Pathology, University of Cambridge | Wellcome Sanger Institute for the COVID-19 Genomics UK Consortium |
| EPI_ISL_441193 | Department of Pathology, University of Cambridge | Wellcome Sanger Institute for the COVID-19 Genomics UK Consortium |
| EPI_ISL_441194 | Department of Pathology, University of Cambridge | Wellcome Sanger Institute for the COVID-19 Genomics UK Consortium |
| EPI_ISL_441195 | Department of Pathology, University of Cambridge | Wellcome Sanger Institute for the COVID-19 Genomics UK Consortium |
| EPI_ISL_441196 | Department of Pathology, University of Cambridge | Wellcome Sanger Institute for the COVID-19 Genomics UK Consortium |

| EPI_Accession_ID | Originating Laboratory | Submitting Laboratory |
|---|---|---|
| EPI_ISL_441197 | Department of Pathology, University of Cambridge | Wellcome Sanger Institute for the COVID-19 Genomics UK Consortium |
| EPI_ISL_441198 | Department of Pathology, University of Cambridge | Wellcome Sanger Institute for the COVID-19 Genomics UK Consortium |
| EPI_ISL_441199 | Department of Pathology, University of Cambridge | Wellcome Sanger Institute for the COVID-19 Genomics UK Consortium |
| EPI_ISL_441200 | Department of Pathology, University of Cambridge | Wellcome Sanger Institute for the COVID-19 Genomics UK Consortium |
| EPI_ISL_441201 | Department of Pathology, University of Cambridge | Wellcome Sanger Institute for the COVID-19 Genomics UK Consortium |
| EPI_ISL_441202 | Department of Pathology, University of Cambridge | Wellcome Sanger Institute for the COVID-19 Genomics UK Consortium |
| EPI_ISL_441203 | Department of Pathology, University of Cambridge | Wellcome Sanger Institute for the COVID-19 Genomics UK Consortium |
| EPI_ISL_441204 | Department of Pathology, University of Cambridge | Wellcome Sanger Institute for the COVID-19 Genomics UK Consortium |
| EPI_ISL_441205 | Department of Pathology, University of Cambridge | Wellcome Sanger Institute for the COVID-19 Genomics UK Consortium |
| EPI_ISL_441206 | Department of Pathology, University of Cambridge | Wellcome Sanger Institute for the COVID-19 Genomics UK Consortium |
| EPI_ISL_441207 | Department of Pathology, University of Cambridge | Wellcome Sanger Institute for the COVID-19 Genomics UK Consortium |
| EPI_ISL_441208 | Department of Pathology, University of Cambridge | Wellcome Sanger Institute for the COVID-19 Genomics UK Consortium |
| EPI_ISL_441209 | Department of Pathology, University of Cambridge | Wellcome Sanger Institute for the COVID-19 Genomics UK Consortium |
| EPI_ISL_441210 | Department of Pathology, University of Cambridge | Wellcome Sanger Institute for the COVID-19 Genomics UK Consortium |
| EPI_ISL_441211 | Department of Pathology, University of Cambridge | Wellcome Sanger Institute for the COVID-19 Genomics UK Consortium |
| EPI_ISL_441212 | Department of Pathology, University of Cambridge | Wellcome Sanger Institute for the COVID-19 Genomics UK Consortium |

| EPI_Accession_ID | Originating Laboratory | Submitting Laboratory |
|---|---|---|
| EPI_ISL_441213 | Department of Pathology, University of Cambridge | Wellcome Sanger Institute for the COVID-19 Genomics UK Consortium |
| EPI_ISL_441214 | Department of Pathology, University of Cambridge | Wellcome Sanger Institute for the COVID-19 Genomics UK Consortium |
| EPI_ISL_441215 | Department of Pathology, University of Cambridge | Wellcome Sanger Institute for the COVID-19 Genomics UK Consortium |
| EPI_ISL_441216 | Department of Pathology, University of Cambridge | Wellcome Sanger Institute for the COVID-19 Genomics UK Consortium |
| EPI_ISL_441217 | Department of Pathology, University of Cambridge | Wellcome Sanger Institute for the COVID-19 Genomics UK Consortium |
| EPI_ISL_441218 | Department of Pathology, University of Cambridge | Wellcome Sanger Institute for the COVID-19 Genomics UK Consortium |
| EPI_ISL_441219 | Department of Pathology, University of Cambridge | Wellcome Sanger Institute for the COVID-19 Genomics UK Consortium |
| EPI_ISL_441220 | Department of Pathology, University of Cambridge | Wellcome Sanger Institute for the COVID-19 Genomics UK Consortium |
| EPI_ISL_441221 | Department of Pathology, University of Cambridge | Wellcome Sanger Institute for the COVID-19 Genomics UK Consortium |
| EPI_ISL_441222 | Department of Pathology, University of Cambridge | Wellcome Sanger Institute for the COVID-19 Genomics UK Consortium |
| EPI_ISL_441223 | Department of Pathology, University of Cambridge | Wellcome Sanger Institute for the COVID-19 Genomics UK Consortium |
| EPI_ISL_441224 | Department of Pathology, University of Cambridge | Wellcome Sanger Institute for the COVID-19 Genomics UK Consortium |
| EPI_ISL_441225 | Department of Pathology, University of Cambridge | Wellcome Sanger Institute for the COVID-19 Genomics UK Consortium |
| EPI_ISL_441226 | Department of Pathology, University of Cambridge | Wellcome Sanger Institute for the COVID-19 Genomics UK Consortium |
| EPI_ISL_441227 | Department of Pathology, University of Cambridge | Wellcome Sanger Institute for the COVID-19 Genomics UK Consortium |
| EPI_ISL_441228 | Department of Pathology, University of Cambridge | Wellcome Sanger Institute for the COVID-19 Genomics UK Consortium |

| EPI_Accession_ID | Originating Laboratory | Submitting Laboratory |
| --- | --- | --- |
| EPI_ISL_441229 | Department of Pathology, University of Cambridge | Wellcome Sanger Institute for the COVID-19 Genomics UK Consortium |
| EPI_ISL_441230 | Department of Pathology, University of Cambridge | Wellcome Sanger Institute for the COVID-19 Genomics UK Consortium |
| EPI_ISL_441231 | Department of Pathology, University of Cambridge | Wellcome Sanger Institute for the COVID-19 Genomics UK Consortium |
| EPI_ISL_441232 | Department of Pathology, University of Cambridge | Wellcome Sanger Institute for the COVID-19 Genomics UK Consortium |
| EPI_ISL_441233 | Department of Pathology, University of Cambridge | Wellcome Sanger Institute for the COVID-19 Genomics UK Consortium |
| EPI_ISL_441234 | Department of Pathology, University of Cambridge | Wellcome Sanger Institute for the COVID-19 Genomics UK Consortium |
| EPI_ISL_441235 | Department of Pathology, University of Cambridge | Wellcome Sanger Institute for the COVID-19 Genomics UK Consortium |
| EPI_ISL_441236 | Department of Pathology, University of Cambridge | Wellcome Sanger Institute for the COVID-19 Genomics UK Consortium |
| EPI_ISL_441237 | Department of Pathology, University of Cambridge | Wellcome Sanger Institute for the COVID-19 Genomics UK Consortium |
| EPI_ISL_441238 | Department of Pathology, University of Cambridge | Wellcome Sanger Institute for the COVID-19 Genomics UK Consortium |
| EPI_ISL_441239 | Department of Pathology, University of Cambridge | Wellcome Sanger Institute for the COVID-19 Genomics UK Consortium |
| EPI_ISL_441240 | Department of Pathology, University of Cambridge | Wellcome Sanger Institute for the COVID-19 Genomics UK Consortium |
| EPI_ISL_441241 | Department of Pathology, University of Cambridge | Wellcome Sanger Institute for the COVID-19 Genomics UK Consortium |
| EPI_ISL_441242 | Department of Pathology, University of Cambridge | Wellcome Sanger Institute for the COVID-19 Genomics UK Consortium |
| EPI_ISL_441243 | Department of Pathology, University of Cambridge | Wellcome Sanger Institute for the COVID-19 Genomics UK Consortium |
| EPI_ISL_441244 | Department of Pathology, University of Cambridge | Wellcome Sanger Institute for the COVID-19 Genomics UK Consortium |

| EPI_Accession_ID | Originating Laboratory | Submitting Laboratory |
|---|---|---|
| EPI_ISL_441245 | Department of Pathology, University of Cambridge | Wellcome Sanger Institute for the COVID-19 Genomics UK Consortium |
| EPI_ISL_441246 | Department of Pathology, University of Cambridge | Wellcome Sanger Institute for the COVID-19 Genomics UK Consortium |
| EPI_ISL_441247 | Department of Pathology, University of Cambridge | Wellcome Sanger Institute for the COVID-19 Genomics UK Consortium |
| EPI_ISL_441248 | Department of Pathology, University of Cambridge | Wellcome Sanger Institute for the COVID-19 Genomics UK Consortium |
| EPI_ISL_441249 | Department of Pathology, University of Cambridge | Wellcome Sanger Institute for the COVID-19 Genomics UK Consortium |
| EPI_ISL_441250 | Department of Pathology, University of Cambridge | Wellcome Sanger Institute for the COVID-19 Genomics UK Consortium |
| EPI_ISL_441251 | Department of Pathology, University of Cambridge | Wellcome Sanger Institute for the COVID-19 Genomics UK Consortium |
| EPI_ISL_441252 | Department of Pathology, University of Cambridge | Wellcome Sanger Institute for the COVID-19 Genomics UK Consortium |
| EPI_ISL_441253 | Department of Pathology, University of Cambridge | Wellcome Sanger Institute for the COVID-19 Genomics UK Consortium |
| EPI_ISL_441254 | Department of Pathology, University of Cambridge | Wellcome Sanger Institute for the COVID-19 Genomics UK Consortium |
| EPI_ISL_441255 | Department of Pathology, University of Cambridge | Wellcome Sanger Institute for the COVID-19 Genomics UK Consortium |
| EPI_ISL_441256 | Department of Pathology, University of Cambridge | Wellcome Sanger Institute for the COVID-19 Genomics UK Consortium |
| EPI_ISL_441257 | Department of Pathology, University of Cambridge | Wellcome Sanger Institute for the COVID-19 Genomics UK Consortium |
| EPI_ISL_441258 | Department of Pathology, University of Cambridge | Wellcome Sanger Institute for the COVID-19 Genomics UK Consortium |
| EPI_ISL_441259 | Department of Pathology, University of Cambridge | Wellcome Sanger Institute for the COVID-19 Genomics UK Consortium |
| EPI_ISL_441260 | Department of Pathology, University of Cambridge | Wellcome Sanger Institute for the COVID-19 Genomics UK Consortium |

| EPI_Accession_ID | Originating Laboratory | Submitting Laboratory |
|---|---|---|
| EPI_ISL_441261 | Department of Pathology, University of Cambridge | Wellcome Sanger Institute for the COVID-19 Genomics UK Consortium |
| EPI_ISL_441262 | Department of Pathology, University of Cambridge | Wellcome Sanger Institute for the COVID-19 Genomics UK Consortium |
| EPI_ISL_441263 | Department of Pathology, University of Cambridge | Wellcome Sanger Institute for the COVID-19 Genomics UK Consortium |
| EPI_ISL_441264 | Department of Pathology, University of Cambridge | Wellcome Sanger Institute for the COVID-19 Genomics UK Consortium |
| EPI_ISL_441265 | Department of Pathology, University of Cambridge | Wellcome Sanger Institute for the COVID-19 Genomics UK Consortium |
| EPI_ISL_441266 | Department of Pathology, University of Cambridge | Wellcome Sanger Institute for the COVID-19 Genomics UK Consortium |
| EPI_ISL_441267 | Department of Pathology, University of Cambridge | Wellcome Sanger Institute for the COVID-19 Genomics UK Consortium |
| EPI_ISL_441268 | Department of Pathology, University of Cambridge | Wellcome Sanger Institute for the COVID-19 Genomics UK Consortium |
| EPI_ISL_441269 | Department of Pathology, University of Cambridge | Wellcome Sanger Institute for the COVID-19 Genomics UK Consortium |
| EPI_ISL_441270 | Department of Pathology, University of Cambridge | Wellcome Sanger Institute for the COVID-19 Genomics UK Consortium |
| EPI_ISL_441271 | Department of Pathology, University of Cambridge | Wellcome Sanger Institute for the COVID-19 Genomics UK Consortium |
| EPI_ISL_441272 | Department of Pathology, University of Cambridge | Wellcome Sanger Institute for the COVID-19 Genomics UK Consortium |
| EPI_ISL_441273 | Department of Pathology, University of Cambridge | Wellcome Sanger Institute for the COVID-19 Genomics UK Consortium |
| EPI_ISL_441274 | Department of Pathology, University of Cambridge | Wellcome Sanger Institute for the COVID-19 Genomics UK Consortium |
| EPI_ISL_441275 | Department of Pathology, University of Cambridge | Wellcome Sanger Institute for the COVID-19 Genomics UK Consortium |
| EPI_ISL_441276 | Department of Pathology, University of Cambridge | Wellcome Sanger Institute for the COVID-19 Genomics UK Consortium |

| EPI_Accession_ID | Originating Laboratory | Submitting Laboratory |
| --- | --- | --- |
| EPI_ISL_441277 | Department of Pathology, University of Cambridge | Wellcome Sanger Institute for the COVID-19 Genomics UK Consortium |
| EPI_ISL_441278 | Department of Pathology, University of Cambridge | Wellcome Sanger Institute for the COVID-19 Genomics UK Consortium |
| EPI_ISL_441279 | Department of Pathology, University of Cambridge | Wellcome Sanger Institute for the COVID-19 Genomics UK Consortium |
| EPI_ISL_441280 | Department of Pathology, University of Cambridge | Wellcome Sanger Institute for the COVID-19 Genomics UK Consortium |
| EPI_ISL_441281 | Department of Pathology, University of Cambridge | Wellcome Sanger Institute for the COVID-19 Genomics UK Consortium |
| EPI_ISL_441282 | Department of Pathology, University of Cambridge | Wellcome Sanger Institute for the COVID-19 Genomics UK Consortium |
| EPI_ISL_441283 | Department of Pathology, University of Cambridge | Wellcome Sanger Institute for the COVID-19 Genomics UK Consortium |
| EPI_ISL_441284 | Department of Pathology, University of Cambridge | Wellcome Sanger Institute for the COVID-19 Genomics UK Consortium |
| EPI_ISL_441285 | Department of Pathology, University of Cambridge | Wellcome Sanger Institute for the COVID-19 Genomics UK Consortium |
| EPI_ISL_441286 | Department of Pathology, University of Cambridge | Wellcome Sanger Institute for the COVID-19 Genomics UK Consortium |
| EPI_ISL_441287 | Department of Pathology, University of Cambridge | Wellcome Sanger Institute for the COVID-19 Genomics UK Consortium |
| EPI_ISL_441288 | Department of Pathology, University of Cambridge | Wellcome Sanger Institute for the COVID-19 Genomics UK Consortium |
| EPI_ISL_441289 | Department of Pathology, University of Cambridge | Wellcome Sanger Institute for the COVID-19 Genomics UK Consortium |
| EPI_ISL_441290 | Department of Pathology, University of Cambridge | Wellcome Sanger Institute for the COVID-19 Genomics UK Consortium |
| EPI_ISL_441291 | Department of Pathology, University of Cambridge | Wellcome Sanger Institute for the COVID-19 Genomics UK Consortium |
| EPI_ISL_441292 | Department of Pathology, University of Cambridge | Wellcome Sanger Institute for the COVID-19 Genomics UK Consortium |

| EPI_Accession_ID | Originating Laboratory | Submitting Laboratory |
|---|---|---|
| EPI_ISL_441293 | Department of Pathology, University of Cambridge | Wellcome Sanger Institute for the COVID-19 Genomics UK Consortium |
| EPI_ISL_441294 | Department of Pathology, University of Cambridge | Wellcome Sanger Institute for the COVID-19 Genomics UK Consortium |
| EPI_ISL_441295 | Department of Pathology, University of Cambridge | Wellcome Sanger Institute for the COVID-19 Genomics UK Consortium |
| EPI_ISL_441296 | Department of Pathology, University of Cambridge | Wellcome Sanger Institute for the COVID-19 Genomics UK Consortium |
| EPI_ISL_441297 | Department of Pathology, University of Cambridge | Wellcome Sanger Institute for the COVID-19 Genomics UK Consortium |
| EPI_ISL_441298 | Department of Pathology, University of Cambridge | Wellcome Sanger Institute for the COVID-19 Genomics UK Consortium |
| EPI_ISL_441299 | Department of Pathology, University of Cambridge | Wellcome Sanger Institute for the COVID-19 Genomics UK Consortium |
| EPI_ISL_441300 | Department of Pathology, University of Cambridge | Wellcome Sanger Institute for the COVID-19 Genomics UK Consortium |
| EPI_ISL_441301 | Department of Pathology, University of Cambridge | Wellcome Sanger Institute for the COVID-19 Genomics UK Consortium |
| EPI_ISL_441302 | Department of Pathology, University of Cambridge | Wellcome Sanger Institute for the COVID-19 Genomics UK Consortium |
| EPI_ISL_441303 | Department of Pathology, University of Cambridge | Wellcome Sanger Institute for the COVID-19 Genomics UK Consortium |
| EPI_ISL_441304 | Department of Pathology, University of Cambridge | Wellcome Sanger Institute for the COVID-19 Genomics UK Consortium |
| EPI_ISL_441305 | Department of Pathology, University of Cambridge | Wellcome Sanger Institute for the COVID-19 Genomics UK Consortium |
| EPI_ISL_441306 | Department of Pathology, University of Cambridge | Wellcome Sanger Institute for the COVID-19 Genomics UK Consortium |
| EPI_ISL_441307 | Department of Pathology, University of Cambridge | Wellcome Sanger Institute for the COVID-19 Genomics UK Consortium |
| EPI_ISL_441308 | Department of Pathology, University of Cambridge | Wellcome Sanger Institute for the COVID-19 Genomics UK Consortium |

| EPI_Accession_ID | Originating Laboratory | Submitting Laboratory |
|---|---|---|
| EPI_ISL_441309 | Department of Pathology, University of Cambridge | Wellcome Sanger Institute for the COVID-19 Genomics UK Consortium |
| EPI_ISL_441310 | Department of Pathology, University of Cambridge | Wellcome Sanger Institute for the COVID-19 Genomics UK Consortium |
| EPI_ISL_441311 | Department of Pathology, University of Cambridge | Wellcome Sanger Institute for the COVID-19 Genomics UK Consortium |
| EPI_ISL_441312 | Department of Pathology, University of Cambridge | Wellcome Sanger Institute for the COVID-19 Genomics UK Consortium |
| EPI_ISL_441313 | Department of Pathology, University of Cambridge | Wellcome Sanger Institute for the COVID-19 Genomics UK Consortium |
| EPI_ISL_441314 | Department of Pathology, University of Cambridge | Wellcome Sanger Institute for the COVID-19 Genomics UK Consortium |
| EPI_ISL_441315 | Department of Pathology, University of Cambridge | Wellcome Sanger Institute for the COVID-19 Genomics UK Consortium |
| EPI_ISL_441316 | Department of Pathology, University of Cambridge | Wellcome Sanger Institute for the COVID-19 Genomics UK Consortium |
| EPI_ISL_441317 | Department of Pathology, University of Cambridge | Wellcome Sanger Institute for the COVID-19 Genomics UK Consortium |
| EPI_ISL_441318 | Department of Pathology, University of Cambridge | Wellcome Sanger Institute for the COVID-19 Genomics UK Consortium |
| EPI_ISL_441319 | Department of Pathology, University of Cambridge | Wellcome Sanger Institute for the COVID-19 Genomics UK Consortium |
| EPI_ISL_441320 | Department of Pathology, University of Cambridge | Wellcome Sanger Institute for the COVID-19 Genomics UK Consortium |
| EPI_ISL_441321 | Department of Pathology, University of Cambridge | Wellcome Sanger Institute for the COVID-19 Genomics UK Consortium |
| EPI_ISL_441322 | Department of Pathology, University of Cambridge | Wellcome Sanger Institute for the COVID-19 Genomics UK Consortium |
| EPI_ISL_441323 | Department of Pathology, University of Cambridge | Wellcome Sanger Institute for the COVID-19 Genomics UK Consortium |
| EPI_ISL_441324 | Department of Pathology, University of Cambridge | Wellcome Sanger Institute for the COVID-19 Genomics UK Consortium |

| EPI_Accession_ID | Originating Laboratory | Submitting Laboratory |
|---|---|---|
| EPI_ISL_441325 | Department of Pathology, University of Cambridge | Wellcome Sanger Institute for the COVID-19 Genomics UK Consortium |
| EPI_ISL_441326 | Department of Pathology, University of Cambridge | Wellcome Sanger Institute for the COVID-19 Genomics UK Consortium |
| EPI_ISL_441327 | Department of Pathology, University of Cambridge | Wellcome Sanger Institute for the COVID-19 Genomics UK Consortium |
| EPI_ISL_441328 | Department of Pathology, University of Cambridge | Wellcome Sanger Institute for the COVID-19 Genomics UK Consortium |
| EPI_ISL_441329 | Department of Pathology, University of Cambridge | Wellcome Sanger Institute for the COVID-19 Genomics UK Consortium |
| EPI_ISL_441330 | Department of Pathology, University of Cambridge | Wellcome Sanger Institute for the COVID-19 Genomics UK Consortium |
| EPI_ISL_441331 | Department of Pathology, University of Cambridge | Wellcome Sanger Institute for the COVID-19 Genomics UK Consortium |
| EPI_ISL_441332 | Department of Pathology, University of Cambridge | Wellcome Sanger Institute for the COVID-19 Genomics UK Consortium |
| EPI_ISL_441333 | Department of Pathology, University of Cambridge | Wellcome Sanger Institute for the COVID-19 Genomics UK Consortium |
| EPI_ISL_441334 | Department of Pathology, University of Cambridge | Wellcome Sanger Institute for the COVID-19 Genomics UK Consortium |
| EPI_ISL_441335 | Department of Pathology, University of Cambridge | Wellcome Sanger Institute for the COVID-19 Genomics UK Consortium |
| EPI_ISL_441336 | Department of Pathology, University of Cambridge | Wellcome Sanger Institute for the COVID-19 Genomics UK Consortium |
| EPI_ISL_441337 | Department of Pathology, University of Cambridge | Wellcome Sanger Institute for the COVID-19 Genomics UK Consortium |
| EPI_ISL_441338 | Department of Pathology, University of Cambridge | Wellcome Sanger Institute for the COVID-19 Genomics UK Consortium |
| EPI_ISL_441339 | Department of Pathology, University of Cambridge | Wellcome Sanger Institute for the COVID-19 Genomics UK Consortium |
| EPI_ISL_441340 | Department of Pathology, University of Cambridge | Wellcome Sanger Institute for the COVID-19 Genomics UK Consortium |

| EPI_Accession_ID | Originating Laboratory | Submitting Laboratory |
|---|---|---|
| EPI_ISL_441341 | Department of Pathology, University of Cambridge | Wellcome Sanger Institute for the COVID-19 Genomics UK Consortium |
| EPI_ISL_441342 | Department of Pathology, University of Cambridge | Wellcome Sanger Institute for the COVID-19 Genomics UK Consortium |
| EPI_ISL_441343 | Department of Pathology, University of Cambridge | Wellcome Sanger Institute for the COVID-19 Genomics UK Consortium |
| EPI_ISL_441344 | Department of Pathology, University of Cambridge | Wellcome Sanger Institute for the COVID-19 Genomics UK Consortium |
| EPI_ISL_441345 | Department of Pathology, University of Cambridge | Wellcome Sanger Institute for the COVID-19 Genomics UK Consortium |
| EPI_ISL_441346 | Department of Pathology, University of Cambridge | Wellcome Sanger Institute for the COVID-19 Genomics UK Consortium |
| EPI_ISL_441347 | Department of Pathology, University of Cambridge | Wellcome Sanger Institute for the COVID-19 Genomics UK Consortium |
| EPI_ISL_441348 | Department of Pathology, University of Cambridge | Wellcome Sanger Institute for the COVID-19 Genomics UK Consortium |
| EPI_ISL_441349 | Department of Pathology, University of Cambridge | Wellcome Sanger Institute for the COVID-19 Genomics UK Consortium |
| EPI_ISL_441350 | University College London, Great Ormond Street Hospital for Children NHS Foundation Trust, Imperial College Healthcare NHS Trust | COVID-19 Genomics UK (COG-UK) Consortium |
| EPI_ISL_441351 | University College London, Great Ormond Street Hospital for Children NHS Foundation Trust, Imperial College Healthcare NHS Trust | COVID-19 Genomics UK (COG-UK) Consortium |
| EPI_ISL_441352 | University College London, Great Ormond Street Hospital for Children NHS Foundation Trust, Imperial College Healthcare NHS Trust | COVID-19 Genomics UK (COG-UK) Consortium |
| EPI_ISL_441353 | University College London, Great Ormond Street Hospital for Children NHS Foundation Trust, Imperial College Healthcare NHS Trust | COVID-19 Genomics UK (COG-UK) Consortium |
| EPI_ISL_441354 | University College London, Great Ormond Street Hospital for Children NHS Foundation Trust, Imperial College Healthcare NHS Trust | COVID-19 Genomics UK (COG-UK) Consortium |

| EPI_Accession_ID | Originating Laboratory | Submitting Laboratory |
|---|---|---|
| EPI_ISL_441355 | Regional Virus Laboratory, Belfast Health and Social Care Trust | COVID-19 Genomics UK (COG-UK) Consortium |
| EPI_ISL_441356 | Regional Virus Laboratory, Belfast Health and Social Care Trust | COVID-19 Genomics UK (COG-UK) Consortium |
| EPI_ISL_441357 | Regional Virus Laboratory, Belfast Health and Social Care Trust | COVID-19 Genomics UK (COG-UK) Consortium |
| EPI_ISL_441358 | Regional Virus Laboratory, Belfast Health and Social Care Trust | COVID-19 Genomics UK (COG-UK) Consortium |
| EPI_ISL_441359 | Regional Virus Laboratory, Belfast Health and Social Care Trust | COVID-19 Genomics UK (COG-UK) Consortium |
| EPI_ISL_441360 | Regional Virus Laboratory, Belfast Health and Social Care Trust | COVID-19 Genomics UK (COG-UK) Consortium |
| EPI_ISL_441361 | Regional Virus Laboratory, Belfast Health and Social Care Trust | COVID-19 Genomics UK (COG-UK) Consortium |
| EPI_ISL_441362 | Regional Virus Laboratory, Belfast Health and Social Care Trust | COVID-19 Genomics UK (COG-UK) Consortium |
| EPI_ISL_441363 | Regional Virus Laboratory, Belfast Health and Social Care Trust | COVID-19 Genomics UK (COG-UK) Consortium |
| EPI_ISL_441364 | Regional Virus Laboratory, Belfast Health and Social Care Trust | COVID-19 Genomics UK (COG-UK) Consortium |
| EPI_ISL_441365 | Regional Virus Laboratory, Belfast Health and Social Care Trust | COVID-19 Genomics UK (COG-UK) Consortium |
| EPI_ISL_441366 | Regional Virus Laboratory, Belfast Health and Social Care Trust | COVID-19 Genomics UK (COG-UK) Consortium |
| EPI_ISL_441367 | Regional Virus Laboratory, Belfast Health and Social Care Trust | COVID-19 Genomics UK (COG-UK) Consortium |
| EPI_ISL_441368 | Regional Virus Laboratory, Belfast Health and Social Care Trust | COVID-19 Genomics UK (COG-UK) Consortium |
| EPI_ISL_441369 | Regional Virus Laboratory, Belfast Health and Social Care Trust | COVID-19 Genomics UK (COG-UK) Consortium |
| EPI_ISL_441370 | Regional Virus Laboratory, Belfast Health and Social Care Trust | COVID-19 Genomics UK (COG-UK) Consortium |

| EPI_Accession_ID | Originating Laboratory | Submitting Laboratory |
|---|---|---|
| EPI_ISL_441371 | Regional Virus Laboratory, Belfast Health and Social Care Trust | COVID-19 Genomics UK (COG-UK) Consortium |
| EPI_ISL_441372 | Regional Virus Laboratory, Belfast Health and Social Care Trust | COVID-19 Genomics UK (COG-UK) Consortium |
| EPI_ISL_441373 | Regional Virus Laboratory, Belfast Health and Social Care Trust | COVID-19 Genomics UK (COG-UK) Consortium |
| EPI_ISL_441374 | Regional Virus Laboratory, Belfast Health and Social Care Trust | COVID-19 Genomics UK (COG-UK) Consortium |
| EPI_ISL_441375 | Regional Virus Laboratory, Belfast Health and Social Care Trust | COVID-19 Genomics UK (COG-UK) Consortium |
| EPI_ISL_441376 | Regional Virus Laboratory, Belfast Health and Social Care Trust | COVID-19 Genomics UK (COG-UK) Consortium |
| EPI_ISL_441377 | Regional Virus Laboratory, Belfast Health and Social Care Trust | COVID-19 Genomics UK (COG-UK) Consortium |
| EPI_ISL_441378 | Regional Virus Laboratory, Belfast Health and Social Care Trust | COVID-19 Genomics UK (COG-UK) Consortium |
| EPI_ISL_441379 | Regional Virus Laboratory, Belfast Health and Social Care Trust | COVID-19 Genomics UK (COG-UK) Consortium |
| EPI_ISL_441380 | Regional Virus Laboratory, Belfast Health and Social Care Trust | COVID-19 Genomics UK (COG-UK) Consortium |
| EPI_ISL_441381 | Regional Virus Laboratory, Belfast Health and Social Care Trust | COVID-19 Genomics UK (COG-UK) Consortium |
| EPI_ISL_441382 | Regional Virus Laboratory, Belfast Health and Social Care Trust | COVID-19 Genomics UK (COG-UK) Consortium |
| EPI_ISL_441383 | Regional Virus Laboratory, Belfast Health and Social Care Trust | COVID-19 Genomics UK (COG-UK) Consortium |
| EPI_ISL_441384 | Regional Virus Laboratory, Belfast Health and Social Care Trust | COVID-19 Genomics UK (COG-UK) Consortium |
| EPI_ISL_441385 | Regional Virus Laboratory, Belfast Health and Social Care Trust | COVID-19 Genomics UK (COG-UK) Consortium |
| EPI_ISL_441386 | Regional Virus Laboratory, Belfast Health and Social Care Trust | COVID-19 Genomics UK (COG-UK) Consortium |

| EPI_Accession_ID | Originating Laboratory | Submitting Laboratory |
|---|---|---|
| EPI_ISL_441387 | Regional Virus Laboratory, Belfast Health and Social Care Trust | COVID-19 Genomics UK (COG-UK) Consortium |
| EPI_ISL_441388 | Regional Virus Laboratory, Belfast Health and Social Care Trust | COVID-19 Genomics UK (COG-UK) Consortium |
| EPI_ISL_441389 | Regional Virus Laboratory, Belfast Health and Social Care Trust | COVID-19 Genomics UK (COG-UK) Consortium |
| EPI_ISL_441390 | Regional Virus Laboratory, Belfast Health and Social Care Trust | COVID-19 Genomics UK (COG-UK) Consortium |
| EPI_ISL_441391 | Regional Virus Laboratory, Belfast Health and Social Care Trust | COVID-19 Genomics UK (COG-UK) Consortium |
| EPI_ISL_441392 | Regional Virus Laboratory, Belfast Health and Social Care Trust | COVID-19 Genomics UK (COG-UK) Consortium |
| EPI_ISL_441393 | Regional Virus Laboratory, Belfast Health and Social Care Trust | COVID-19 Genomics UK (COG-UK) Consortium |
| EPI_ISL_441394 | Regional Virus Laboratory, Belfast Health and Social Care Trust | COVID-19 Genomics UK (COG-UK) Consortium |
| EPI_ISL_441395 | Regional Virus Laboratory, Belfast Health and Social Care Trust | COVID-19 Genomics UK (COG-UK) Consortium |
| EPI_ISL_441396 | Regional Virus Laboratory, Belfast Health and Social Care Trust | COVID-19 Genomics UK (COG-UK) Consortium |
| EPI_ISL_441397 | Regional Virus Laboratory, Belfast Health and Social Care Trust | COVID-19 Genomics UK (COG-UK) Consortium |
| EPI_ISL_441398 | Regional Virus Laboratory, Belfast Health and Social Care Trust | COVID-19 Genomics UK (COG-UK) Consortium |
| EPI_ISL_441399 | Regional Virus Laboratory, Belfast Health and Social Care Trust | COVID-19 Genomics UK (COG-UK) Consortium |
| EPI_ISL_441400 | Regional Virus Laboratory, Belfast Health and Social Care Trust | COVID-19 Genomics UK (COG-UK) Consortium |
| EPI_ISL_441401 | Regional Virus Laboratory, Belfast Health and Social Care Trust | COVID-19 Genomics UK (COG-UK) Consortium |
| EPI_ISL_441402 | Regional Virus Laboratory, Belfast Health and Social Care Trust | COVID-19 Genomics UK (COG-UK) Consortium |

| EPI_Accession_ID | Originating Laboratory | Submitting Laboratory |
|---|---|---|
| EPI_ISL_441403 | Regional Virus Laboratory, Belfast Health and Social Care Trust | COVID-19 Genomics UK (COG-UK) Consortium |
| EPI_ISL_441404 | Regional Virus Laboratory, Belfast Health and Social Care Trust | COVID-19 Genomics UK (COG-UK) Consortium |
| EPI_ISL_441405 | Regional Virus Laboratory, Belfast Health and Social Care Trust | COVID-19 Genomics UK (COG-UK) Consortium |
| EPI_ISL_441406 | Regional Virus Laboratory, Belfast Health and Social Care Trust | COVID-19 Genomics UK (COG-UK) Consortium |
| EPI_ISL_441407 | Regional Virus Laboratory, Belfast Health and Social Care Trust | COVID-19 Genomics UK (COG-UK) Consortium |
| EPI_ISL_441408 | Regional Virus Laboratory, Belfast Health and Social Care Trust | COVID-19 Genomics UK (COG-UK) Consortium |
| EPI_ISL_441409 | Regional Virus Laboratory, Belfast Health and Social Care Trust | COVID-19 Genomics UK (COG-UK) Consortium |
| EPI_ISL_441410 | Regional Virus Laboratory, Belfast Health and Social Care Trust | COVID-19 Genomics UK (COG-UK) Consortium |
| EPI_ISL_441411 | Regional Virus Laboratory, Belfast Health and Social Care Trust | COVID-19 Genomics UK (COG-UK) Consortium |
| EPI_ISL_441412 | Regional Virus Laboratory, Belfast Health and Social Care Trust | COVID-19 Genomics UK (COG-UK) Consortium |
| EPI_ISL_441413 | Regional Virus Laboratory, Belfast Health and Social Care Trust | COVID-19 Genomics UK (COG-UK) Consortium |
| EPI_ISL_441414 | Regional Virus Laboratory, Belfast Health and Social Care Trust | COVID-19 Genomics UK (COG-UK) Consortium |
| EPI_ISL_441415 | Regional Virus Laboratory, Belfast Health and Social Care Trust | COVID-19 Genomics UK (COG-UK) Consortium |
| EPI_ISL_441416 | Regional Virus Laboratory, Belfast Health and Social Care Trust | COVID-19 Genomics UK (COG-UK) Consortium |
| EPI_ISL_441417 | Regional Virus Laboratory, Belfast Health and Social Care Trust | COVID-19 Genomics UK (COG-UK) Consortium |
| EPI_ISL_441418 | Regional Virus Laboratory, Belfast Health and Social Care Trust | COVID-19 Genomics UK (COG-UK) Consortium |

| EPI_Accession_ID | Originating Laboratory | Submitting Laboratory |
|---|---|---|
| EPI_ISL_441419 | Regional Virus Laboratory, Belfast Health and Social Care Trust | COVID-19 Genomics UK (COG-UK) Consortium |
| EPI_ISL_441420 | Regional Virus Laboratory, Belfast Health and Social Care Trust | COVID-19 Genomics UK (COG-UK) Consortium |
| EPI_ISL_441421 | Regional Virus Laboratory, Belfast Health and Social Care Trust | COVID-19 Genomics UK (COG-UK) Consortium |
| EPI_ISL_441422 | Regional Virus Laboratory, Belfast Health and Social Care Trust | COVID-19 Genomics UK (COG-UK) Consortium |
| EPI_ISL_441423 | Regional Virus Laboratory, Belfast Health and Social Care Trust | COVID-19 Genomics UK (COG-UK) Consortium |
| EPI_ISL_441424 | Regional Virus Laboratory, Belfast Health and Social Care Trust | COVID-19 Genomics UK (COG-UK) Consortium |
| EPI_ISL_441425 | Regional Virus Laboratory, Belfast Health and Social Care Trust | COVID-19 Genomics UK (COG-UK) Consortium |
| EPI_ISL_441426 | Regional Virus Laboratory, Belfast Health and Social Care Trust | COVID-19 Genomics UK (COG-UK) Consortium |
| EPI_ISL_441427 | Regional Virus Laboratory, Belfast Health and Social Care Trust | COVID-19 Genomics UK (COG-UK) Consortium |
| EPI_ISL_441428 | Regional Virus Laboratory, Belfast Health and Social Care Trust | COVID-19 Genomics UK (COG-UK) Consortium |
| EPI_ISL_441429 | Regional Virus Laboratory, Belfast Health and Social Care Trust | COVID-19 Genomics UK (COG-UK) Consortium |
| EPI_ISL_441430 | Regional Virus Laboratory, Belfast Health and Social Care Trust | COVID-19 Genomics UK (COG-UK) Consortium |
| EPI_ISL_441431 | Regional Virus Laboratory, Belfast Health and Social Care Trust | COVID-19 Genomics UK (COG-UK) Consortium |
| EPI_ISL_441432 | Regional Virus Laboratory, Belfast Health and Social Care Trust | COVID-19 Genomics UK (COG-UK) Consortium |
| EPI_ISL_441433 | Regional Virus Laboratory, Belfast Health and Social Care Trust | COVID-19 Genomics UK (COG-UK) Consortium |
| EPI_ISL_441434 | Regional Virus Laboratory, Belfast Health and Social Care Trust | COVID-19 Genomics UK (COG-UK) Consortium |

| EPI_Accession_ID | Originating Laboratory | Submitting Laboratory |
| --- | --- | --- |
| EPI_ISL_441435 | Regional Virus Laboratory, Belfast Health and Social Care Trust | COVID-19 Genomics UK (COG-UK) Consortium |
| EPI_ISL_441436 | Regional Virus Laboratory, Belfast Health and Social Care Trust | COVID-19 Genomics UK (COG-UK) Consortium |
| EPI_ISL_441437 | Queens Medical Centre, Clinical Microbiology Department / DeepSeq Nottingham | COVID-19 Genomics UK (COG-UK) Consortium |
| EPI_ISL_441438 | Queens Medical Centre, Clinical Microbiology Department / DeepSeq Nottingham | COVID-19 Genomics UK (COG-UK) Consortium |
| EPI_ISL_441439 | Queens Medical Centre, Clinical Microbiology Department / DeepSeq Nottingham | COVID-19 Genomics UK (COG-UK) Consortium |
| EPI_ISL_441440 | Queens Medical Centre, Clinical Microbiology Department / DeepSeq Nottingham | COVID-19 Genomics UK (COG-UK) Consortium |
| EPI_ISL_441441 | Queens Medical Centre, Clinical Microbiology Department / DeepSeq Nottingham | COVID-19 Genomics UK (COG-UK) Consortium |
| EPI_ISL_441442 | Queens Medical Centre, Clinical Microbiology Department / DeepSeq Nottingham | COVID-19 Genomics UK (COG-UK) Consortium |
| EPI_ISL_441443 | Queens Medical Centre, Clinical Microbiology Department / DeepSeq Nottingham | COVID-19 Genomics UK (COG-UK) Consortium |
| EPI_ISL_441444 | Queens Medical Centre, Clinical Microbiology Department / DeepSeq Nottingham | COVID-19 Genomics UK (COG-UK) Consortium |
| EPI_ISL_441445 | Queens Medical Centre, Clinical Microbiology Department / DeepSeq Nottingham | COVID-19 Genomics UK (COG-UK) Consortium |
| EPI_ISL_441446 | Queens Medical Centre, Clinical Microbiology Department / DeepSeq Nottingham | COVID-19 Genomics UK (COG-UK) Consortium |
| EPI_ISL_441447 | Queens Medical Centre, Clinical Microbiology Department / DeepSeq Nottingham | COVID-19 Genomics UK (COG-UK) Consortium |
| EPI_ISL_441448 | Queens Medical Centre, Clinical Microbiology Department / DeepSeq Nottingham | COVID-19 Genomics UK (COG-UK) Consortium |
| EPI_ISL_441449 | Queens Medical Centre, Clinical Microbiology Department / DeepSeq Nottingham | COVID-19 Genomics UK (COG-UK) Consortium |
| EPI_ISL_441450 | Queens Medical Centre, Clinical Microbiology Department / DeepSeq Nottingham | COVID-19 Genomics UK (COG-UK) Consortium |

| EPI_Accession_ID | Originating Laboratory | Submitting Laboratory |
|---|---|---|
| EPI_ISL_441451 | Queens Medical Centre, Clinical Microbiology Department / DeepSeq Nottingham | COVID-19 Genomics UK (COG-UK) Consortium |
| EPI_ISL_441452 | Queens Medical Centre, Clinical Microbiology Department / DeepSeq Nottingham | COVID-19 Genomics UK (COG-UK) Consortium |
| EPI_ISL_441453 | Queens Medical Centre, Clinical Microbiology Department / DeepSeq Nottingham | COVID-19 Genomics UK (COG-UK) Consortium |
| EPI_ISL_441454 | Queens Medical Centre, Clinical Microbiology Department / DeepSeq Nottingham | COVID-19 Genomics UK (COG-UK) Consortium |
| EPI_ISL_441455 | Queens Medical Centre, Clinical Microbiology Department / DeepSeq Nottingham | COVID-19 Genomics UK (COG-UK) Consortium |
| EPI_ISL_441456 | Queens Medical Centre, Clinical Microbiology Department / DeepSeq Nottingham | COVID-19 Genomics UK (COG-UK) Consortium |
| EPI_ISL_441457 | Queens Medical Centre, Clinical Microbiology Department / DeepSeq Nottingham | COVID-19 Genomics UK (COG-UK) Consortium |
| EPI_ISL_441458 | Queens Medical Centre, Clinical Microbiology Department / DeepSeq Nottingham | COVID-19 Genomics UK (COG-UK) Consortium |
| EPI_ISL_441459 | Queens Medical Centre, Clinical Microbiology Department / DeepSeq Nottingham | COVID-19 Genomics UK (COG-UK) Consortium |
| EPI_ISL_441460 | Queens Medical Centre, Clinical Microbiology Department / DeepSeq Nottingham | COVID-19 Genomics UK (COG-UK) Consortium |
| EPI_ISL_441461 | Queens Medical Centre, Clinical Microbiology Department / DeepSeq Nottingham | COVID-19 Genomics UK (COG-UK) Consortium |
| EPI_ISL_441462 | Queens Medical Centre, Clinical Microbiology Department / DeepSeq Nottingham | COVID-19 Genomics UK (COG-UK) Consortium |
| EPI_ISL_441463 | Queens Medical Centre, Clinical Microbiology Department / DeepSeq Nottingham | COVID-19 Genomics UK (COG-UK) Consortium |
| EPI_ISL_441464 | Queens Medical Centre, Clinical Microbiology Department / DeepSeq Nottingham | COVID-19 Genomics UK (COG-UK) Consortium |
| EPI_ISL_441465 | Queens Medical Centre, Clinical Microbiology Department / DeepSeq Nottingham | COVID-19 Genomics UK (COG-UK) Consortium |
| EPI_ISL_441466 | Queens Medical Centre, Clinical Microbiology Department / DeepSeq Nottingham | COVID-19 Genomics UK (COG-UK) Consortium |

| EPI_Accession_ID | Originating Laboratory | Submitting Laboratory |
|---|---|---|
| EPI_ISL_441467 | Queens Medical Centre, Clinical Microbiology Department / DeepSeq Nottingham | COVID-19 Genomics UK (COG-UK) Consortium |
| EPI_ISL_441468 | Queens Medical Centre, Clinical Microbiology Department / DeepSeq Nottingham | COVID-19 Genomics UK (COG-UK) Consortium |
| EPI_ISL_441469 | Queens Medical Centre, Clinical Microbiology Department / DeepSeq Nottingham | COVID-19 Genomics UK (COG-UK) Consortium |
| EPI_ISL_441470 | Queens Medical Centre, Clinical Microbiology Department / DeepSeq Nottingham | COVID-19 Genomics UK (COG-UK) Consortium |
| EPI_ISL_441471 | Queens Medical Centre, Clinical Microbiology Department / DeepSeq Nottingham | COVID-19 Genomics UK (COG-UK) Consortium |
| EPI_ISL_441472 | Queens Medical Centre, Clinical Microbiology Department / DeepSeq Nottingham | COVID-19 Genomics UK (COG-UK) Consortium |
| EPI_ISL_441473 | Queens Medical Centre, Clinical Microbiology Department / DeepSeq Nottingham | COVID-19 Genomics UK (COG-UK) Consortium |
| EPI_ISL_441474 | Queens Medical Centre, Clinical Microbiology Department / DeepSeq Nottingham | COVID-19 Genomics UK (COG-UK) Consortium |
| EPI_ISL_441475 | Queens Medical Centre, Clinical Microbiology Department / DeepSeq Nottingham | COVID-19 Genomics UK (COG-UK) Consortium |
| EPI_ISL_441476 | Queens Medical Centre, Clinical Microbiology Department / DeepSeq Nottingham | COVID-19 Genomics UK (COG-UK) Consortium |
| EPI_ISL_441477 | Queens Medical Centre, Clinical Microbiology Department / DeepSeq Nottingham | COVID-19 Genomics UK (COG-UK) Consortium |
| EPI_ISL_441478 | Queens Medical Centre, Clinical Microbiology Department / DeepSeq Nottingham | COVID-19 Genomics UK (COG-UK) Consortium |
| EPI_ISL_441479 | Queens Medical Centre, Clinical Microbiology Department / DeepSeq Nottingham | COVID-19 Genomics UK (COG-UK) Consortium |
| EPI_ISL_441480 | Queens Medical Centre, Clinical Microbiology Department / DeepSeq Nottingham | COVID-19 Genomics UK (COG-UK) Consortium |
| EPI_ISL_441481 | Queens Medical Centre, Clinical Microbiology Department / DeepSeq Nottingham | COVID-19 Genomics UK (COG-UK) Consortium |
| EPI_ISL_441482 | Queens Medical Centre, Clinical Microbiology Department / DeepSeq Nottingham | COVID-19 Genomics UK (COG-UK) Consortium |

| EPI_Accession_ID | Originating Laboratory | Submitting Laboratory |
|---|---|---|
| EPI_ISL_441483 | Queens Medical Centre, Clinical Microbiology Department / DeepSeq Nottingham | COVID-19 Genomics UK (COG-UK) Consortium |
| EPI_ISL_441484 | Queens Medical Centre, Clinical Microbiology Department / DeepSeq Nottingham | COVID-19 Genomics UK (COG-UK) Consortium |
| EPI_ISL_441485 | Queens Medical Centre, Clinical Microbiology Department / DeepSeq Nottingham | COVID-19 Genomics UK (COG-UK) Consortium |
| EPI_ISL_441486 | Queens Medical Centre, Clinical Microbiology Department / DeepSeq Nottingham | COVID-19 Genomics UK (COG-UK) Consortium |
| EPI_ISL_441487 | Queens Medical Centre, Clinical Microbiology Department / DeepSeq Nottingham | COVID-19 Genomics UK (COG-UK) Consortium |
| EPI_ISL_441488 | Queens Medical Centre, Clinical Microbiology Department / DeepSeq Nottingham | COVID-19 Genomics UK (COG-UK) Consortium |
| EPI_ISL_441489 | Queens Medical Centre, Clinical Microbiology Department / DeepSeq Nottingham | COVID-19 Genomics UK (COG-UK) Consortium |
| EPI_ISL_441490 | Queens Medical Centre, Clinical Microbiology Department / DeepSeq Nottingham | COVID-19 Genomics UK (COG-UK) Consortium |
| EPI_ISL_441491 | Queens Medical Centre, Clinical Microbiology Department / DeepSeq Nottingham | COVID-19 Genomics UK (COG-UK) Consortium |
| EPI_ISL_441492 | Queens Medical Centre, Clinical Microbiology Department / DeepSeq Nottingham | COVID-19 Genomics UK (COG-UK) Consortium |
| EPI_ISL_441493 | Queens Medical Centre, Clinical Microbiology Department / DeepSeq Nottingham | COVID-19 Genomics UK (COG-UK) Consortium |
| EPI_ISL_441494 | Queens Medical Centre, Clinical Microbiology Department / DeepSeq Nottingham | COVID-19 Genomics UK (COG-UK) Consortium |
| EPI_ISL_441495 | Queens Medical Centre, Clinical Microbiology Department / DeepSeq Nottingham | COVID-19 Genomics UK (COG-UK) Consortium |
| EPI_ISL_441496 | Queens Medical Centre, Clinical Microbiology Department / DeepSeq Nottingham | COVID-19 Genomics UK (COG-UK) Consortium |
| EPI_ISL_441497 | Queens Medical Centre, Clinical Microbiology Department / DeepSeq Nottingham | COVID-19 Genomics UK (COG-UK) Consortium |
| EPI_ISL_441498 | Queens Medical Centre, Clinical Microbiology Department / DeepSeq Nottingham | COVID-19 Genomics UK (COG-UK) Consortium |

| EPI_Accession_ID | Originating Laboratory | Submitting Laboratory |
|---|---|---|
| EPI_ISL_441499 | Queens Medical Centre, Clinical Microbiology Department / DeepSeq Nottingham | COVID-19 Genomics UK (COG-UK) Consortium |
| EPI_ISL_441500 | Queens Medical Centre, Clinical Microbiology Department / DeepSeq Nottingham | COVID-19 Genomics UK (COG-UK) Consortium |
| EPI_ISL_441501 | Queens Medical Centre, Clinical Microbiology Department / DeepSeq Nottingham | COVID-19 Genomics UK (COG-UK) Consortium |
| EPI_ISL_441502 | Queens Medical Centre, Clinical Microbiology Department / DeepSeq Nottingham | COVID-19 Genomics UK (COG-UK) Consortium |
| EPI_ISL_441503 | Queens Medical Centre, Clinical Microbiology Department / DeepSeq Nottingham | COVID-19 Genomics UK (COG-UK) Consortium |
| EPI_ISL_441504 | Queens Medical Centre, Clinical Microbiology Department / DeepSeq Nottingham | COVID-19 Genomics UK (COG-UK) Consortium |
| EPI_ISL_441505 | Queens Medical Centre, Clinical Microbiology Department / DeepSeq Nottingham | COVID-19 Genomics UK (COG-UK) Consortium |
| EPI_ISL_441506 | Queens Medical Centre, Clinical Microbiology Department / DeepSeq Nottingham | COVID-19 Genomics UK (COG-UK) Consortium |
| EPI_ISL_441507 | Queens Medical Centre, Clinical Microbiology Department / DeepSeq Nottingham | COVID-19 Genomics UK (COG-UK) Consortium |
| EPI_ISL_441508 | Queens Medical Centre, Clinical Microbiology Department / DeepSeq Nottingham | COVID-19 Genomics UK (COG-UK) Consortium |
| EPI_ISL_441509 | Queens Medical Centre, Clinical Microbiology Department / DeepSeq Nottingham | COVID-19 Genomics UK (COG-UK) Consortium |
| EPI_ISL_441510 | Queens Medical Centre, Clinical Microbiology Department / DeepSeq Nottingham | COVID-19 Genomics UK (COG-UK) Consortium |
| EPI_ISL_441511 | Queens Medical Centre, Clinical Microbiology Department / DeepSeq Nottingham | COVID-19 Genomics UK (COG-UK) Consortium |
| EPI_ISL_441512 | Queens Medical Centre, Clinical Microbiology Department / DeepSeq Nottingham | COVID-19 Genomics UK (COG-UK) Consortium |
| EPI_ISL_441513 | Queens Medical Centre, Clinical Microbiology Department / DeepSeq Nottingham | COVID-19 Genomics UK (COG-UK) Consortium |
| EPI_ISL_441514 | Queens Medical Centre, Clinical Microbiology Department / DeepSeq Nottingham | COVID-19 Genomics UK (COG-UK) Consortium |

| EPI_Accession_ID | Originating Laboratory | Submitting Laboratory |
|---|---|---|
| EPI_ISL_441515 | Queens Medical Centre, Clinical Microbiology Department / DeepSeq Nottingham | COVID-19 Genomics UK (COG-UK) Consortium |
| EPI_ISL_441516 | Queens Medical Centre, Clinical Microbiology Department / DeepSeq Nottingham | COVID-19 Genomics UK (COG-UK) Consortium |
| EPI_ISL_441517 | Queens Medical Centre, Clinical Microbiology Department / DeepSeq Nottingham | COVID-19 Genomics UK (COG-UK) Consortium |
| EPI_ISL_441518 | Queens Medical Centre, Clinical Microbiology Department / DeepSeq Nottingham | COVID-19 Genomics UK (COG-UK) Consortium |
| EPI_ISL_441519 | Queens Medical Centre, Clinical Microbiology Department / DeepSeq Nottingham | COVID-19 Genomics UK (COG-UK) Consortium |
| EPI_ISL_441520 | Queens Medical Centre, Clinical Microbiology Department / DeepSeq Nottingham | COVID-19 Genomics UK (COG-UK) Consortium |
| EPI_ISL_441521 | Queens Medical Centre, Clinical Microbiology Department / DeepSeq Nottingham | COVID-19 Genomics UK (COG-UK) Consortium |
| EPI_ISL_441522 | Queens Medical Centre, Clinical Microbiology Department / DeepSeq Nottingham | COVID-19 Genomics UK (COG-UK) Consortium |
| EPI_ISL_441523 | Queens Medical Centre, Clinical Microbiology Department / DeepSeq Nottingham | COVID-19 Genomics UK (COG-UK) Consortium |
| EPI_ISL_441524 | Queens Medical Centre, Clinical Microbiology Department / DeepSeq Nottingham | COVID-19 Genomics UK (COG-UK) Consortium |
| EPI_ISL_441525 | Queens Medical Centre, Clinical Microbiology Department / DeepSeq Nottingham | COVID-19 Genomics UK (COG-UK) Consortium |
| EPI_ISL_441526 | Queens Medical Centre, Clinical Microbiology Department / DeepSeq Nottingham | COVID-19 Genomics UK (COG-UK) Consortium |
| EPI_ISL_441527 | Queens Medical Centre, Clinical Microbiology Department / DeepSeq Nottingham | COVID-19 Genomics UK (COG-UK) Consortium |
| EPI_ISL_441528 | Queens Medical Centre, Clinical Microbiology Department / DeepSeq Nottingham | COVID-19 Genomics UK (COG-UK) Consortium |
| EPI_ISL_441529 | Queens Medical Centre, Clinical Microbiology Department / DeepSeq Nottingham | COVID-19 Genomics UK (COG-UK) Consortium |
| EPI_ISL_441530 | Queens Medical Centre, Clinical Microbiology Department / DeepSeq Nottingham | COVID-19 Genomics UK (COG-UK) Consortium |

| EPI_Accession_ID | Originating Laboratory | Submitting Laboratory |
|---|---|---|
| EPI_ISL_441531 | Queens Medical Centre, Clinical Microbiology Department / DeepSeq Nottingham | COVID-19 Genomics UK (COG-UK) Consortium |
| EPI_ISL_441532 | Queens Medical Centre, Clinical Microbiology Department / DeepSeq Nottingham | COVID-19 Genomics UK (COG-UK) Consortium |
| EPI_ISL_441533 | Queens Medical Centre, Clinical Microbiology Department / DeepSeq Nottingham | COVID-19 Genomics UK (COG-UK) Consortium |
| EPI_ISL_441534 | Queens Medical Centre, Clinical Microbiology Department / DeepSeq Nottingham | COVID-19 Genomics UK (COG-UK) Consortium |
| EPI_ISL_441535 | Queens Medical Centre, Clinical Microbiology Department / DeepSeq Nottingham | COVID-19 Genomics UK (COG-UK) Consortium |
| EPI_ISL_441536 | Queens Medical Centre, Clinical Microbiology Department / DeepSeq Nottingham | COVID-19 Genomics UK (COG-UK) Consortium |
| EPI_ISL_441537 | Queens Medical Centre, Clinical Microbiology Department / DeepSeq Nottingham | COVID-19 Genomics UK (COG-UK) Consortium |
| EPI_ISL_441538 | Queens Medical Centre, Clinical Microbiology Department / DeepSeq Nottingham | COVID-19 Genomics UK (COG-UK) Consortium |
| EPI_ISL_441539 | Queens Medical Centre, Clinical Microbiology Department / DeepSeq Nottingham | COVID-19 Genomics UK (COG-UK) Consortium |
| EPI_ISL_441540 | Queens Medical Centre, Clinical Microbiology Department / DeepSeq Nottingham | COVID-19 Genomics UK (COG-UK) Consortium |
| EPI_ISL_441541 | Queens Medical Centre, Clinical Microbiology Department / DeepSeq Nottingham | COVID-19 Genomics UK (COG-UK) Consortium |
| EPI_ISL_441542 | Queens Medical Centre, Clinical Microbiology Department / DeepSeq Nottingham | COVID-19 Genomics UK (COG-UK) Consortium |
| EPI_ISL_441543 | Queens Medical Centre, Clinical Microbiology Department / DeepSeq Nottingham | COVID-19 Genomics UK (COG-UK) Consortium |
| EPI_ISL_441544 | Queens Medical Centre, Clinical Microbiology Department / DeepSeq Nottingham | COVID-19 Genomics UK (COG-UK) Consortium |
| EPI_ISL_441545 | Queens Medical Centre, Clinical Microbiology Department / DeepSeq Nottingham | COVID-19 Genomics UK (COG-UK) Consortium |
| EPI_ISL_441546 | Queens Medical Centre, Clinical Microbiology Department / DeepSeq Nottingham | COVID-19 Genomics UK (COG-UK) Consortium |

| EPI_Accession_ID | Originating Laboratory | Submitting Laboratory |
|---|---|---|
| EPI_ISL_441547 | Department of Pathology, University of Cambridge | Wellcome Sanger Institute for the COVID-19 Genomics UK Consortium |
| EPI_ISL_441548 | Department of Pathology, University of Cambridge | Wellcome Sanger Institute for the COVID-19 Genomics UK Consortium |
| EPI_ISL_441549 | Department of Pathology, University of Cambridge | Wellcome Sanger Institute for the COVID-19 Genomics UK Consortium |
| EPI_ISL_441550 | Department of Pathology, University of Cambridge | Wellcome Sanger Institute for the COVID-19 Genomics UK Consortium |
| EPI_ISL_441551 | Department of Pathology, University of Cambridge | Wellcome Sanger Institute for the COVID-19 Genomics UK Consortium |
| EPI_ISL_441552 | Department of Pathology, University of Cambridge | Wellcome Sanger Institute for the COVID-19 Genomics UK Consortium |
| EPI_ISL_441553 | Department of Pathology, University of Cambridge | Wellcome Sanger Institute for the COVID-19 Genomics UK Consortium |
| EPI_ISL_441554 | Department of Pathology, University of Cambridge | Wellcome Sanger Institute for the COVID-19 Genomics UK Consortium |
| EPI_ISL_441555 | Department of Pathology, University of Cambridge | Wellcome Sanger Institute for the COVID-19 Genomics UK Consortium |
| EPI_ISL_441556 | Department of Pathology, University of Cambridge | Wellcome Sanger Institute for the COVID-19 Genomics UK Consortium |
| EPI_ISL_441557 | Department of Pathology, University of Cambridge | Wellcome Sanger Institute for the COVID-19 Genomics UK Consortium |
| EPI_ISL_441558 | Department of Pathology, University of Cambridge | Wellcome Sanger Institute for the COVID-19 Genomics UK Consortium |
| EPI_ISL_441559 | Department of Pathology, University of Cambridge | Wellcome Sanger Institute for the COVID-19 Genomics UK Consortium |
| EPI_ISL_441560 | Department of Pathology, University of Cambridge | Wellcome Sanger Institute for the COVID-19 Genomics UK Consortium |
| EPI_ISL_441561 | Department of Pathology, University of Cambridge | Wellcome Sanger Institute for the COVID-19 Genomics UK Consortium |
| EPI_ISL_441562 | Department of Pathology, University of Cambridge | Wellcome Sanger Institute for the COVID-19 Genomics UK Consortium |

| EPI_Accession_ID | Originating Laboratory | Submitting Laboratory |
|---|---|---|
| EPI_ISL_441563 | Department of Pathology, University of Cambridge | Wellcome Sanger Institute for the COVID-19 Genomics UK Consortium |
| EPI_ISL_441564 | Department of Pathology, University of Cambridge | Wellcome Sanger Institute for the COVID-19 Genomics UK Consortium |
| EPI_ISL_441565 | Department of Pathology, University of Cambridge | Wellcome Sanger Institute for the COVID-19 Genomics UK Consortium |
| EPI_ISL_441566 | Department of Pathology, University of Cambridge | Wellcome Sanger Institute for the COVID-19 Genomics UK Consortium |
| EPI_ISL_441567 | Department of Pathology, University of Cambridge | Wellcome Sanger Institute for the COVID-19 Genomics UK Consortium |
| EPI_ISL_441568 | Department of Pathology, University of Cambridge | Wellcome Sanger Institute for the COVID-19 Genomics UK Consortium |
| EPI_ISL_441569 | Department of Pathology, University of Cambridge | Wellcome Sanger Institute for the COVID-19 Genomics UK Consortium |
| EPI_ISL_441570 | Department of Pathology, University of Cambridge | Wellcome Sanger Institute for the COVID-19 Genomics UK Consortium |
| EPI_ISL_441571 | Department of Pathology, University of Cambridge | Wellcome Sanger Institute for the COVID-19 Genomics UK Consortium |
| EPI_ISL_441572 | Department of Pathology, University of Cambridge | Wellcome Sanger Institute for the COVID-19 Genomics UK Consortium |
| EPI_ISL_441573 | Department of Pathology, University of Cambridge | Wellcome Sanger Institute for the COVID-19 Genomics UK Consortium |
| EPI_ISL_441574 | Department of Pathology, University of Cambridge | Wellcome Sanger Institute for the COVID-19 Genomics UK Consortium |
| EPI_ISL_441575 | Department of Pathology, University of Cambridge | Wellcome Sanger Institute for the COVID-19 Genomics UK Consortium |
| EPI_ISL_441576 | Department of Pathology, University of Cambridge | Wellcome Sanger Institute for the COVID-19 Genomics UK Consortium |
| EPI_ISL_441577 | Department of Pathology, University of Cambridge | Wellcome Sanger Institute for the COVID-19 Genomics UK Consortium |
| EPI_ISL_441578 | Department of Pathology, University of Cambridge | Wellcome Sanger Institute for the COVID-19 Genomics UK Consortium |

| EPI_Accession_ID | Originating Laboratory | Submitting Laboratory |
|---|---|---|
| EPI_ISL_441579 | Department of Pathology, University of Cambridge | Wellcome Sanger Institute for the COVID-19 Genomics UK Consortium |
| EPI_ISL_441580 | Department of Pathology, University of Cambridge | Wellcome Sanger Institute for the COVID-19 Genomics UK Consortium |
| EPI_ISL_441581 | Department of Pathology, University of Cambridge | Wellcome Sanger Institute for the COVID-19 Genomics UK Consortium |
| EPI_ISL_441582 | Department of Pathology, University of Cambridge | Wellcome Sanger Institute for the COVID-19 Genomics UK Consortium |
| EPI_ISL_441583 | Department of Pathology, University of Cambridge | Wellcome Sanger Institute for the COVID-19 Genomics UK Consortium |
| EPI_ISL_441584 | Department of Pathology, University of Cambridge | Wellcome Sanger Institute for the COVID-19 Genomics UK Consortium |
| EPI_ISL_441585 | Department of Pathology, University of Cambridge | Wellcome Sanger Institute for the COVID-19 Genomics UK Consortium |
| EPI_ISL_441586 | Department of Pathology, University of Cambridge | Wellcome Sanger Institute for the COVID-19 Genomics UK Consortium |
| EPI_ISL_441587 | Department of Pathology, University of Cambridge | Wellcome Sanger Institute for the COVID-19 Genomics UK Consortium |
| EPI_ISL_441588 | Department of Pathology, University of Cambridge | Wellcome Sanger Institute for the COVID-19 Genomics UK Consortium |
| EPI_ISL_441589 | Department of Pathology, University of Cambridge | Wellcome Sanger Institute for the COVID-19 Genomics UK Consortium |
| EPI_ISL_441590 | Department of Pathology, University of Cambridge | Wellcome Sanger Institute for the COVID-19 Genomics UK Consortium |
| EPI_ISL_441591 | Department of Pathology, University of Cambridge | Wellcome Sanger Institute for the COVID-19 Genomics UK Consortium |
| EPI_ISL_441592 | Department of Pathology, University of Cambridge | Wellcome Sanger Institute for the COVID-19 Genomics UK Consortium |
| EPI_ISL_441593 | Department of Pathology, University of Cambridge | Wellcome Sanger Institute for the COVID-19 Genomics UK Consortium |
| EPI_ISL_441594 | Department of Pathology, University of Cambridge | Wellcome Sanger Institute for the COVID-19 Genomics UK Consortium |

| EPI_Accession_ID | Originating Laboratory | Submitting Laboratory |
|---|---|---|
| EPI_ISL_441595 | Department of Pathology, University of Cambridge | Wellcome Sanger Institute for the COVID-19 Genomics UK Consortium |
| EPI_ISL_441596 | Department of Pathology, University of Cambridge | Wellcome Sanger Institute for the COVID-19 Genomics UK Consortium |
| EPI_ISL_441597 | Department of Pathology, University of Cambridge | Wellcome Sanger Institute for the COVID-19 Genomics UK Consortium |
| EPI_ISL_441598 | Department of Pathology, University of Cambridge | Wellcome Sanger Institute for the COVID-19 Genomics UK Consortium |
| EPI_ISL_441599 | Department of Pathology, University of Cambridge | Wellcome Sanger Institute for the COVID-19 Genomics UK Consortium |
| EPI_ISL_441600 | Department of Pathology, University of Cambridge | Wellcome Sanger Institute for the COVID-19 Genomics UK Consortium |
| EPI_ISL_441601 | Department of Pathology, University of Cambridge | Wellcome Sanger Institute for the COVID-19 Genomics UK Consortium |
| EPI_ISL_441602 | Department of Pathology, University of Cambridge | Wellcome Sanger Institute for the COVID-19 Genomics UK Consortium |
| EPI_ISL_441603 | Department of Pathology, University of Cambridge | Wellcome Sanger Institute for the COVID-19 Genomics UK Consortium |
| EPI_ISL_441604 | Department of Pathology, University of Cambridge | Wellcome Sanger Institute for the COVID-19 Genomics UK Consortium |
| EPI_ISL_441605 | Department of Pathology, University of Cambridge | Wellcome Sanger Institute for the COVID-19 Genomics UK Consortium |
| EPI_ISL_441606 | Department of Pathology, University of Cambridge | Wellcome Sanger Institute for the COVID-19 Genomics UK Consortium |
| EPI_ISL_441607 | Department of Pathology, University of Cambridge | Wellcome Sanger Institute for the COVID-19 Genomics UK Consortium |
| EPI_ISL_441608 | Department of Pathology, University of Cambridge | Wellcome Sanger Institute for the COVID-19 Genomics UK Consortium |
| EPI_ISL_441609 | Department of Pathology, University of Cambridge | Wellcome Sanger Institute for the COVID-19 Genomics UK Consortium |
| EPI_ISL_441610 | Department of Pathology, University of Cambridge | Wellcome Sanger Institute for the COVID-19 Genomics UK Consortium |

| EPI_Accession_ID | Originating Laboratory | Submitting Laboratory |
|---|---|---|
| EPI_ISL_441611 | Department of Pathology, University of Cambridge | Wellcome Sanger Institute for the COVID-19 Genomics UK Consortium |
| EPI_ISL_441612 | Department of Pathology, University of Cambridge | Wellcome Sanger Institute for the COVID-19 Genomics UK Consortium |
| EPI_ISL_441613 | Department of Pathology, University of Cambridge | Wellcome Sanger Institute for the COVID-19 Genomics UK Consortium |
| EPI_ISL_441614 | Department of Pathology, University of Cambridge | Wellcome Sanger Institute for the COVID-19 Genomics UK Consortium |
| EPI_ISL_441615 | Department of Pathology, University of Cambridge | Wellcome Sanger Institute for the COVID-19 Genomics UK Consortium |
| EPI_ISL_441616 | Department of Pathology, University of Cambridge | Wellcome Sanger Institute for the COVID-19 Genomics UK Consortium |
| EPI_ISL_441617 | Department of Pathology, University of Cambridge | Wellcome Sanger Institute for the COVID-19 Genomics UK Consortium |
| EPI_ISL_441618 | Department of Pathology, University of Cambridge | Wellcome Sanger Institute for the COVID-19 Genomics UK Consortium |
| EPI_ISL_441619 | Department of Pathology, University of Cambridge | Wellcome Sanger Institute for the COVID-19 Genomics UK Consortium |
| EPI_ISL_441620 | Department of Pathology, University of Cambridge | Wellcome Sanger Institute for the COVID-19 Genomics UK Consortium |
| EPI_ISL_441621 | Department of Pathology, University of Cambridge | Wellcome Sanger Institute for the COVID-19 Genomics UK Consortium |
| EPI_ISL_441622 | Department of Pathology, University of Cambridge | Wellcome Sanger Institute for the COVID-19 Genomics UK Consortium |
| EPI_ISL_441623 | Department of Pathology, University of Cambridge | Wellcome Sanger Institute for the COVID-19 Genomics UK Consortium |
| EPI_ISL_441624 | Department of Pathology, University of Cambridge | Wellcome Sanger Institute for the COVID-19 Genomics UK Consortium |
| EPI_ISL_441625 | Department of Pathology, University of Cambridge | Wellcome Sanger Institute for the COVID-19 Genomics UK Consortium |
| EPI_ISL_441626 | Department of Pathology, University of Cambridge | Wellcome Sanger Institute for the COVID-19 Genomics UK Consortium |

| EPI_Accession_ID | Originating Laboratory | Submitting Laboratory |
|---|---|---|
| EPI_ISL_441627 | Department of Pathology, University of Cambridge | Wellcome Sanger Institute for the COVID-19 Genomics UK Consortium |
| EPI_ISL_441628 | Department of Pathology, University of Cambridge | Wellcome Sanger Institute for the COVID-19 Genomics UK Consortium |
| EPI_ISL_441629 | Department of Pathology, University of Cambridge | Wellcome Sanger Institute for the COVID-19 Genomics UK Consortium |
| EPI_ISL_441630 | Department of Pathology, University of Cambridge | Wellcome Sanger Institute for the COVID-19 Genomics UK Consortium |
| EPI_ISL_441631 | Department of Pathology, University of Cambridge | Wellcome Sanger Institute for the COVID-19 Genomics UK Consortium |
| EPI_ISL_441632 | Department of Pathology, University of Cambridge | Wellcome Sanger Institute for the COVID-19 Genomics UK Consortium |
| EPI_ISL_441633 | Department of Pathology, University of Cambridge | Wellcome Sanger Institute for the COVID-19 Genomics UK Consortium |
| EPI_ISL_441634 | Department of Pathology, University of Cambridge | Wellcome Sanger Institute for the COVID-19 Genomics UK Consortium |
| EPI_ISL_441635 | Department of Pathology, University of Cambridge | Wellcome Sanger Institute for the COVID-19 Genomics UK Consortium |
| EPI_ISL_441636 | Department of Pathology, University of Cambridge | Wellcome Sanger Institute for the COVID-19 Genomics UK Consortium |
| EPI_ISL_441637 | Department of Pathology, University of Cambridge | Wellcome Sanger Institute for the COVID-19 Genomics UK Consortium |
| EPI_ISL_441638 | Department of Pathology, University of Cambridge | Wellcome Sanger Institute for the COVID-19 Genomics UK Consortium |
| EPI_ISL_441639 | Department of Pathology, University of Cambridge | Wellcome Sanger Institute for the COVID-19 Genomics UK Consortium |
| EPI_ISL_441640 | Department of Pathology, University of Cambridge | Wellcome Sanger Institute for the COVID-19 Genomics UK Consortium |
| EPI_ISL_441641 | Department of Pathology, University of Cambridge | Wellcome Sanger Institute for the COVID-19 Genomics UK Consortium |
| EPI_ISL_441642 | Department of Pathology, University of Cambridge | Wellcome Sanger Institute for the COVID-19 Genomics UK Consortium |

| EPI_Accession_ID | Originating Laboratory | Submitting Laboratory |
|---|---|---|
| EPI_ISL_441643 | Department of Pathology, University of Cambridge | Wellcome Sanger Institute for the COVID-19 Genomics UK Consortium |
| EPI_ISL_441644 | Department of Pathology, University of Cambridge | Wellcome Sanger Institute for the COVID-19 Genomics UK Consortium |
| EPI_ISL_441645 | Department of Pathology, University of Cambridge | Wellcome Sanger Institute for the COVID-19 Genomics UK Consortium |
| EPI_ISL_441646 | Department of Pathology, University of Cambridge | Wellcome Sanger Institute for the COVID-19 Genomics UK Consortium |
| EPI_ISL_441647 | Department of Pathology, University of Cambridge | Wellcome Sanger Institute for the COVID-19 Genomics UK Consortium |
| EPI_ISL_441648 | Department of Pathology, University of Cambridge | Wellcome Sanger Institute for the COVID-19 Genomics UK Consortium |
| EPI_ISL_441649 | Department of Pathology, University of Cambridge | Wellcome Sanger Institute for the COVID-19 Genomics UK Consortium |
| EPI_ISL_441650 | Department of Pathology, University of Cambridge | Wellcome Sanger Institute for the COVID-19 Genomics UK Consortium |
| EPI_ISL_441651 | Department of Pathology, University of Cambridge | Wellcome Sanger Institute for the COVID-19 Genomics UK Consortium |
| EPI_ISL_441652 | Department of Pathology, University of Cambridge | Wellcome Sanger Institute for the COVID-19 Genomics UK Consortium |
| EPI_ISL_441653 | Department of Pathology, University of Cambridge | Wellcome Sanger Institute for the COVID-19 Genomics UK Consortium |
| EPI_ISL_441654 | Department of Pathology, University of Cambridge | Wellcome Sanger Institute for the COVID-19 Genomics UK Consortium |
| EPI_ISL_441655 | Department of Pathology, University of Cambridge | Wellcome Sanger Institute for the COVID-19 Genomics UK Consortium |
| EPI_ISL_441656 | Department of Pathology, University of Cambridge | Wellcome Sanger Institute for the COVID-19 Genomics UK Consortium |
| EPI_ISL_441657 | Department of Pathology, University of Cambridge | Wellcome Sanger Institute for the COVID-19 Genomics UK Consortium |
| EPI_ISL_441658 | Department of Pathology, University of Cambridge | Wellcome Sanger Institute for the COVID-19 Genomics UK Consortium |

| EPI_Accession_ID | Originating Laboratory | Submitting Laboratory |
|---|---|---|
| EPI_ISL_441659 | Regional Virus Laboratory, Belfast Health and Social Care Trust | Wellcome Sanger Institute for the COVID-19 Genomics UK Consortium |
| EPI_ISL_441660 | Department of Pathology, University of Cambridge | Wellcome Sanger Institute for the COVID-19 Genomics UK Consortium |
| EPI_ISL_441661 | Department of Pathology, University of Cambridge | Wellcome Sanger Institute for the COVID-19 Genomics UK Consortium |
| EPI_ISL_441662 | Regional Virus Laboratory, Belfast Health and Social Care Trust | Wellcome Sanger Institute for the COVID-19 Genomics UK Consortium |
| EPI_ISL_441663 | Regional Virus Laboratory, Belfast Health and Social Care Trust | Wellcome Sanger Institute for the COVID-19 Genomics UK Consortium |
| EPI_ISL_441664 | Department of Pathology, University of Cambridge | Wellcome Sanger Institute for the COVID-19 Genomics UK Consortium |
| EPI_ISL_441665 | Department of Pathology, University of Cambridge | Wellcome Sanger Institute for the COVID-19 Genomics UK Consortium |
| EPI_ISL_441666 | Department of Pathology, University of Cambridge | Wellcome Sanger Institute for the COVID-19 Genomics UK Consortium |
| EPI_ISL_441667 | Department of Pathology, University of Cambridge | Wellcome Sanger Institute for the COVID-19 Genomics UK Consortium |
| EPI_ISL_441668 | Department of Pathology, University of Cambridge | Wellcome Sanger Institute for the COVID-19 Genomics UK Consortium |
| EPI_ISL_441669 | Department of Pathology, University of Cambridge | Wellcome Sanger Institute for the COVID-19 Genomics UK Consortium |
| EPI_ISL_441670 | Regional Virus Laboratory, Belfast Health and Social Care Trust | Wellcome Sanger Institute for the COVID-19 Genomics UK Consortium |
| EPI_ISL_441671 | Department of Pathology, University of Cambridge | Wellcome Sanger Institute for the COVID-19 Genomics UK Consortium |
| EPI_ISL_441672 | Department of Pathology, University of Cambridge | Wellcome Sanger Institute for the COVID-19 Genomics UK Consortium |
| EPI_ISL_441673 | Department of Pathology, University of Cambridge | Wellcome Sanger Institute for the COVID-19 Genomics UK Consortium |
| EPI_ISL_441674 | Department of Pathology, University of Cambridge | Wellcome Sanger Institute for the COVID-19 Genomics UK Consortium |

| EPI_Accession_ID | Originating Laboratory | Submitting Laboratory |
| --- | --- | --- |
| EPI_ISL_441675 | Regional Virus Laboratory, Belfast Health and Social Care Trust | Wellcome Sanger Institute for the COVID-19 Genomics UK Consortium |
| EPI_ISL_441676 | Regional Virus Laboratory, Belfast Health and Social Care Trust | Wellcome Sanger Institute for the COVID-19 Genomics UK Consortium |
| EPI_ISL_441677 | Department of Pathology, University of Cambridge | Wellcome Sanger Institute for the COVID-19 Genomics UK Consortium |
| EPI_ISL_441678 | Department of Pathology, University of Cambridge | Wellcome Sanger Institute for the COVID-19 Genomics UK Consortium |
| EPI_ISL_441679 | Department of Pathology, University of Cambridge | Wellcome Sanger Institute for the COVID-19 Genomics UK Consortium |
| EPI_ISL_441680 | Department of Pathology, University of Cambridge | Wellcome Sanger Institute for the COVID-19 Genomics UK Consortium |
| EPI_ISL_441681 | Department of Pathology, University of Cambridge | Wellcome Sanger Institute for the COVID-19 Genomics UK Consortium |
| EPI_ISL_441682 | Department of Pathology, University of Cambridge | Wellcome Sanger Institute for the COVID-19 Genomics UK Consortium |
| EPI_ISL_441683 | Department of Pathology, University of Cambridge | Wellcome Sanger Institute for the COVID-19 Genomics UK Consortium |
| EPI_ISL_441684 | Department of Pathology, University of Cambridge | Wellcome Sanger Institute for the COVID-19 Genomics UK Consortium |
| EPI_ISL_441685 | Regional Virus Laboratory, Belfast Health and Social Care Trust | Wellcome Sanger Institute for the COVID-19 Genomics UK Consortium |
| EPI_ISL_441686 | Department of Pathology, University of Cambridge | Wellcome Sanger Institute for the COVID-19 Genomics UK Consortium |
| EPI_ISL_441687 | Department of Pathology, University of Cambridge | Wellcome Sanger Institute for the COVID-19 Genomics UK Consortium |
| EPI_ISL_441688 | Department of Pathology, University of Cambridge | Wellcome Sanger Institute for the COVID-19 Genomics UK Consortium |
| EPI_ISL_441689 | Department of Pathology, University of Cambridge | Wellcome Sanger Institute for the COVID-19 Genomics UK Consortium |
| EPI_ISL_441690 | Regional Virus Laboratory, Belfast Health and Social Care Trust | Wellcome Sanger Institute for the COVID-19 Genomics UK Consortium |

| EPI_Accession_ID | Originating Laboratory | Submitting Laboratory |
|---|---|---|
| EPI_ISL_441691 | Regional Virus Laboratory, Belfast Health and Social Care Trust | Wellcome Sanger Institute for the COVID-19 Genomics UK Consortium |
| EPI_ISL_441692 | Department of Pathology, University of Cambridge | Wellcome Sanger Institute for the COVID-19 Genomics UK Consortium |
| EPI_ISL_441693 | Regional Virus Laboratory, Belfast Health and Social Care Trust | Wellcome Sanger Institute for the COVID-19 Genomics UK Consortium |
| EPI_ISL_441694 | Regional Virus Laboratory, Belfast Health and Social Care Trust | Wellcome Sanger Institute for the COVID-19 Genomics UK Consortium |
| EPI_ISL_441695 | Department of Pathology, University of Cambridge | Wellcome Sanger Institute for the COVID-19 Genomics UK Consortium |
| EPI_ISL_441696 | Regional Virus Laboratory, Belfast Health and Social Care Trust | Wellcome Sanger Institute for the COVID-19 Genomics UK Consortium |
| EPI_ISL_441697 | Regional Virus Laboratory, Belfast Health and Social Care Trust | Wellcome Sanger Institute for the COVID-19 Genomics UK Consortium |
| EPI_ISL_441698 | Regional Virus Laboratory, Belfast Health and Social Care Trust | Wellcome Sanger Institute for the COVID-19 Genomics UK Consortium |
| EPI_ISL_441699 | Regional Virus Laboratory, Belfast Health and Social Care Trust | Wellcome Sanger Institute for the COVID-19 Genomics UK Consortium |
| EPI_ISL_441700 | Department of Pathology, University of Cambridge | Wellcome Sanger Institute for the COVID-19 Genomics UK Consortium |
| EPI_ISL_441701 | Department of Pathology, University of Cambridge | Wellcome Sanger Institute for the COVID-19 Genomics UK Consortium |
| EPI_ISL_441702 | Department of Pathology, University of Cambridge | Wellcome Sanger Institute for the COVID-19 Genomics UK Consortium |
| EPI_ISL_441703 | Department of Pathology, University of Cambridge | Wellcome Sanger Institute for the COVID-19 Genomics UK Consortium |
| EPI_ISL_441704 | Regional Virus Laboratory, Belfast Health and Social Care Trust | Wellcome Sanger Institute for the COVID-19 Genomics UK Consortium |
| EPI_ISL_441705 | Department of Pathology, University of Cambridge | Wellcome Sanger Institute for the COVID-19 Genomics UK Consortium |
| EPI_ISL_441706 | Department of Pathology, University of Cambridge | Wellcome Sanger Institute for the COVID-19 Genomics UK Consortium |

| EPI_Accession_ID | Originating Laboratory | Submitting Laboratory |
|---|---|---|
| EPI_ISL_441707 | Department of Pathology, University of Cambridge | Wellcome Sanger Institute for the COVID-19 Genomics UK Consortium |
| EPI_ISL_441708 | Regional Virus Laboratory, Belfast Health and Social Care Trust | Wellcome Sanger Institute for the COVID-19 Genomics UK Consortium |
| EPI_ISL_441709 | Department of Pathology, University of Cambridge | Wellcome Sanger Institute for the COVID-19 Genomics UK Consortium |
| EPI_ISL_441710 | Department of Pathology, University of Cambridge | Wellcome Sanger Institute for the COVID-19 Genomics UK Consortium |
| EPI_ISL_441711 | Department of Pathology, University of Cambridge | Wellcome Sanger Institute for the COVID-19 Genomics UK Consortium |
| EPI_ISL_441712 | Regional Virus Laboratory, Belfast Health and Social Care Trust | Wellcome Sanger Institute for the COVID-19 Genomics UK Consortium |
| EPI_ISL_441713 | Department of Pathology, University of Cambridge | Wellcome Sanger Institute for the COVID-19 Genomics UK Consortium |
| EPI_ISL_441714 | Regional Virus Laboratory, Belfast Health and Social Care Trust | Wellcome Sanger Institute for the COVID-19 Genomics UK Consortium |
| EPI_ISL_441715 | Department of Pathology, University of Cambridge | Wellcome Sanger Institute for the COVID-19 Genomics UK Consortium |
| EPI_ISL_441716 | Regional Virus Laboratory, Belfast Health and Social Care Trust | Wellcome Sanger Institute for the COVID-19 Genomics UK Consortium |
| EPI_ISL_441717 | Regional Virus Laboratory, Belfast Health and Social Care Trust | Wellcome Sanger Institute for the COVID-19 Genomics UK Consortium |
| EPI_ISL_441718 | Department of Pathology, University of Cambridge | Wellcome Sanger Institute for the COVID-19 Genomics UK Consortium |
| EPI_ISL_441719 | Regional Virus Laboratory, Belfast Health and Social Care Trust | Wellcome Sanger Institute for the COVID-19 Genomics UK Consortium |
| EPI_ISL_441720 | Department of Pathology, University of Cambridge | Wellcome Sanger Institute for the COVID-19 Genomics UK Consortium |
| EPI_ISL_441721 | Department of Pathology, University of Cambridge | Wellcome Sanger Institute for the COVID-19 Genomics UK Consortium |
| EPI_ISL_441722 | Department of Pathology, University of Cambridge | Wellcome Sanger Institute for the COVID-19 Genomics UK Consortium |

| EPI_Accession_ID | Originating Laboratory | Submitting Laboratory |
|---|---|---|
| EPI_ISL_441723 | Department of Pathology, University of Cambridge | Wellcome Sanger Institute for the COVID-19 Genomics UK Consortium |
| EPI_ISL_441724 | Department of Pathology, University of Cambridge | Wellcome Sanger Institute for the COVID-19 Genomics UK Consortium |
| EPI_ISL_441725 | Department of Pathology, University of Cambridge | Wellcome Sanger Institute for the COVID-19 Genomics UK Consortium |
| EPI_ISL_441726 | Department of Pathology, University of Cambridge | Wellcome Sanger Institute for the COVID-19 Genomics UK Consortium |
| EPI_ISL_441727 | Department of Pathology, University of Cambridge | Wellcome Sanger Institute for the COVID-19 Genomics UK Consortium |
| EPI_ISL_441728 | Department of Pathology, University of Cambridge | Wellcome Sanger Institute for the COVID-19 Genomics UK Consortium |
| EPI_ISL_441729 | Department of Pathology, University of Cambridge | Wellcome Sanger Institute for the COVID-19 Genomics UK Consortium |
| EPI_ISL_441730 | Regional Virus Laboratory, Belfast Health and Social Care Trust | Wellcome Sanger Institute for the COVID-19 Genomics UK Consortium |
| EPI_ISL_441731 | Department of Pathology, University of Cambridge | Wellcome Sanger Institute for the COVID-19 Genomics UK Consortium |
| EPI_ISL_441732 | Department of Pathology, University of Cambridge | Wellcome Sanger Institute for the COVID-19 Genomics UK Consortium |
| EPI_ISL_441733 | Regional Virus Laboratory, Belfast Health and Social Care Trust | Wellcome Sanger Institute for the COVID-19 Genomics UK Consortium |
| EPI_ISL_441734 | Department of Pathology, University of Cambridge | Wellcome Sanger Institute for the COVID-19 Genomics UK Consortium |
| EPI_ISL_441735 | Department of Pathology, University of Cambridge | Wellcome Sanger Institute for the COVID-19 Genomics UK Consortium |
| EPI_ISL_441736 | Department of Pathology, University of Cambridge | Wellcome Sanger Institute for the COVID-19 Genomics UK Consortium |
| EPI_ISL_441737 | Regional Virus Laboratory, Belfast Health and Social Care Trust | Wellcome Sanger Institute for the COVID-19 Genomics UK Consortium |
| EPI_ISL_441738 | Department of Pathology, University of Cambridge | Wellcome Sanger Institute for the COVID-19 Genomics UK Consortium |

| EPI_Accession_ID | Originating Laboratory | Submitting Laboratory |
|---|---|---|
| EPI_ISL_441739 | Department of Pathology, University of Cambridge | Wellcome Sanger Institute for the COVID-19 Genomics UK Consortium |
| EPI_ISL_441740 | Department of Pathology, University of Cambridge | Wellcome Sanger Institute for the COVID-19 Genomics UK Consortium |
| EPI_ISL_441741 | Department of Pathology, University of Cambridge | Wellcome Sanger Institute for the COVID-19 Genomics UK Consortium |
| EPI_ISL_441742 | Department of Pathology, University of Cambridge | Wellcome Sanger Institute for the COVID-19 Genomics UK Consortium |
| EPI_ISL_441743 | Department of Pathology, University of Cambridge | Wellcome Sanger Institute for the COVID-19 Genomics UK Consortium |
| EPI_ISL_441744 | Department of Pathology, University of Cambridge | Wellcome Sanger Institute for the COVID-19 Genomics UK Consortium |
| EPI_ISL_441745 | Department of Pathology, University of Cambridge | Wellcome Sanger Institute for the COVID-19 Genomics UK Consortium |
| EPI_ISL_441746 | Department of Pathology, University of Cambridge | Wellcome Sanger Institute for the COVID-19 Genomics UK Consortium |
| EPI_ISL_441747 | Regional Virus Laboratory, Belfast Health and Social Care Trust | Wellcome Sanger Institute for the COVID-19 Genomics UK Consortium |
| EPI_ISL_441748 | Department of Pathology, University of Cambridge | Wellcome Sanger Institute for the COVID-19 Genomics UK Consortium |
| EPI_ISL_441749 | Department of Pathology, University of Cambridge | Wellcome Sanger Institute for the COVID-19 Genomics UK Consortium |
| EPI_ISL_441750 | Department of Pathology, University of Cambridge | Wellcome Sanger Institute for the COVID-19 Genomics UK Consortium |
| EPI_ISL_441751 | Regional Virus Laboratory, Belfast Health and Social Care Trust | Wellcome Sanger Institute for the COVID-19 Genomics UK Consortium |
| EPI_ISL_441752 | Department of Pathology, University of Cambridge | Wellcome Sanger Institute for the COVID-19 Genomics UK Consortium |
| EPI_ISL_441753 | Regional Virus Laboratory, Belfast Health and Social Care Trust | Wellcome Sanger Institute for the COVID-19 Genomics UK Consortium |
| EPI_ISL_441754 | Department of Pathology, University of Cambridge | Wellcome Sanger Institute for the COVID-19 Genomics UK Consortium |

| EPI_Accession_ID | Originating Laboratory | Submitting Laboratory |
|---|---|---|
| EPI_ISL_441755 | Department of Pathology, University of Cambridge | Wellcome Sanger Institute for the COVID-19 Genomics UK Consortium |
| EPI_ISL_441756 | Department of Pathology, University of Cambridge | Wellcome Sanger Institute for the COVID-19 Genomics UK Consortium |
| EPI_ISL_441757 | Department of Pathology, University of Cambridge | Wellcome Sanger Institute for the COVID-19 Genomics UK Consortium |
| EPI_ISL_441758 | Department of Pathology, University of Cambridge | Wellcome Sanger Institute for the COVID-19 Genomics UK Consortium |
| EPI_ISL_441759 | Department of Pathology, University of Cambridge | Wellcome Sanger Institute for the COVID-19 Genomics UK Consortium |
| EPI_ISL_441760 | Department of Pathology, University of Cambridge | Wellcome Sanger Institute for the COVID-19 Genomics UK Consortium |
| EPI_ISL_441761 | Regional Virus Laboratory, Belfast Health and Social Care Trust | Wellcome Sanger Institute for the COVID-19 Genomics UK Consortium |
| EPI_ISL_441762 | Department of Pathology, University of Cambridge | Wellcome Sanger Institute for the COVID-19 Genomics UK Consortium |
| EPI_ISL_441763 | Department of Pathology, University of Cambridge | Wellcome Sanger Institute for the COVID-19 Genomics UK Consortium |
| EPI_ISL_441764 | Department of Pathology, University of Cambridge | Wellcome Sanger Institute for the COVID-19 Genomics UK Consortium |
| EPI_ISL_441765 | Regional Virus Laboratory, Belfast Health and Social Care Trust | Wellcome Sanger Institute for the COVID-19 Genomics UK Consortium |
| EPI_ISL_441766 | Regional Virus Laboratory, Belfast Health and Social Care Trust | Wellcome Sanger Institute for the COVID-19 Genomics UK Consortium |
| EPI_ISL_441767 | Department of Pathology, University of Cambridge | Wellcome Sanger Institute for the COVID-19 Genomics UK Consortium |
| EPI_ISL_441768 | Department of Pathology, University of Cambridge | Wellcome Sanger Institute for the COVID-19 Genomics UK Consortium |
| EPI_ISL_441769 | Department of Pathology, University of Cambridge | Wellcome Sanger Institute for the COVID-19 Genomics UK Consortium |
| EPI_ISL_441770 | Regional Virus Laboratory, Belfast Health and Social Care Trust | Wellcome Sanger Institute for the COVID-19 Genomics UK Consortium |

| EPI_Accession_ID | Originating Laboratory | Submitting Laboratory |
|---|---|---|
| EPI_ISL_441771 | Regional Virus Laboratory, Belfast Health and Social Care Trust | Wellcome Sanger Institute for the COVID-19 Genomics UK Consortium |
| EPI_ISL_441772 | Department of Pathology, University of Cambridge | Wellcome Sanger Institute for the COVID-19 Genomics UK Consortium |
| EPI_ISL_441773 | Department of Pathology, University of Cambridge | Wellcome Sanger Institute for the COVID-19 Genomics UK Consortium |
| EPI_ISL_441774 | Department of Pathology, University of Cambridge | Wellcome Sanger Institute for the COVID-19 Genomics UK Consortium |
| EPI_ISL_441775 | Regional Virus Laboratory, Belfast Health and Social Care Trust | Wellcome Sanger Institute for the COVID-19 Genomics UK Consortium |
| EPI_ISL_441776 | Regional Virus Laboratory, Belfast Health and Social Care Trust | Wellcome Sanger Institute for the COVID-19 Genomics UK Consortium |
| EPI_ISL_441777 | Department of Pathology, University of Cambridge | Wellcome Sanger Institute for the COVID-19 Genomics UK Consortium |
| EPI_ISL_441778 | Department of Pathology, University of Cambridge | Wellcome Sanger Institute for the COVID-19 Genomics UK Consortium |
| EPI_ISL_441779 | Department of Pathology, University of Cambridge | Wellcome Sanger Institute for the COVID-19 Genomics UK Consortium |
| EPI_ISL_441780 | Department of Pathology, University of Cambridge | Wellcome Sanger Institute for the COVID-19 Genomics UK Consortium |
| EPI_ISL_441781 | Regional Virus Laboratory, Belfast Health and Social Care Trust | Wellcome Sanger Institute for the COVID-19 Genomics UK Consortium |
| EPI_ISL_441782 | Department of Pathology, University of Cambridge | Wellcome Sanger Institute for the COVID-19 Genomics UK Consortium |
| EPI_ISL_441783 | Department of Pathology, University of Cambridge | Wellcome Sanger Institute for the COVID-19 Genomics UK Consortium |
| EPI_ISL_441784 | Department of Pathology, University of Cambridge | Wellcome Sanger Institute for the COVID-19 Genomics UK Consortium |
| EPI_ISL_441785 | Department of Pathology, University of Cambridge | Wellcome Sanger Institute for the COVID-19 Genomics UK Consortium |
| EPI_ISL_441786 | Department of Pathology, University of Cambridge | Wellcome Sanger Institute for the COVID-19 Genomics UK Consortium |

| EPI_Accession_ID | Originating Laboratory | Submitting Laboratory |
|---|---|---|
| EPI_ISL_441787 | Department of Pathology, University of Cambridge | Wellcome Sanger Institute for the COVID-19 Genomics UK Consortium |
| EPI_ISL_441788 | Department of Pathology, University of Cambridge | Wellcome Sanger Institute for the COVID-19 Genomics UK Consortium |
| EPI_ISL_441789 | Department of Pathology, University of Cambridge | Wellcome Sanger Institute for the COVID-19 Genomics UK Consortium |
| EPI_ISL_441790 | Department of Pathology, University of Cambridge | Wellcome Sanger Institute for the COVID-19 Genomics UK Consortium |
| EPI_ISL_441791 | Department of Pathology, University of Cambridge | Wellcome Sanger Institute for the COVID-19 Genomics UK Consortium |
| EPI_ISL_441792 | Department of Pathology, University of Cambridge | Wellcome Sanger Institute for the COVID-19 Genomics UK Consortium |
| EPI_ISL_441793 | Department of Pathology, University of Cambridge | Wellcome Sanger Institute for the COVID-19 Genomics UK Consortium |
| EPI_ISL_441794 | Department of Pathology, University of Cambridge | Wellcome Sanger Institute for the COVID-19 Genomics UK Consortium |
| EPI_ISL_441795 | Department of Pathology, University of Cambridge | Wellcome Sanger Institute for the COVID-19 Genomics UK Consortium |
| EPI_ISL_441796 | Department of Pathology, University of Cambridge | Wellcome Sanger Institute for the COVID-19 Genomics UK Consortium |
| EPI_ISL_441797 | Department of Pathology, University of Cambridge | Wellcome Sanger Institute for the COVID-19 Genomics UK Consortium |
| EPI_ISL_441798 | Department of Pathology, University of Cambridge | Wellcome Sanger Institute for the COVID-19 Genomics UK Consortium |
| EPI_ISL_441799 | Department of Pathology, University of Cambridge | Wellcome Sanger Institute for the COVID-19 Genomics UK Consortium |
| EPI_ISL_441800 | Department of Pathology, University of Cambridge | Wellcome Sanger Institute for the COVID-19 Genomics UK Consortium |
| EPI_ISL_441801 | Department of Pathology, University of Cambridge | Wellcome Sanger Institute for the COVID-19 Genomics UK Consortium |
| EPI_ISL_441802 | Department of Pathology, University of Cambridge | Wellcome Sanger Institute for the COVID-19 Genomics UK Consortium |

| EPI_Accession_ID | Originating Laboratory | Submitting Laboratory |
|---|---|---|
| EPI_ISL_441803 | Department of Pathology, University of Cambridge | Wellcome Sanger Institute for the COVID-19 Genomics UK Consortium |
| EPI_ISL_441804 | Department of Pathology, University of Cambridge | Wellcome Sanger Institute for the COVID-19 Genomics UK Consortium |
| EPI_ISL_441805 | Department of Pathology, University of Cambridge | Wellcome Sanger Institute for the COVID-19 Genomics UK Consortium |
| EPI_ISL_441806 | Department of Pathology, University of Cambridge | Wellcome Sanger Institute for the COVID-19 Genomics UK Consortium |
| EPI_ISL_441807 | Department of Pathology, University of Cambridge | Wellcome Sanger Institute for the COVID-19 Genomics UK Consortium |
| EPI_ISL_441808 | Department of Pathology, University of Cambridge | Wellcome Sanger Institute for the COVID-19 Genomics UK Consortium |
| EPI_ISL_441809 | Department of Pathology, University of Cambridge | Wellcome Sanger Institute for the COVID-19 Genomics UK Consortium |
| EPI_ISL_441810 | Department of Pathology, University of Cambridge | Wellcome Sanger Institute for the COVID-19 Genomics UK Consortium |
| EPI_ISL_441811 | Department of Pathology, University of Cambridge | Wellcome Sanger Institute for the COVID-19 Genomics UK Consortium |
| EPI_ISL_441812 | Department of Pathology, University of Cambridge | Wellcome Sanger Institute for the COVID-19 Genomics UK Consortium |
| EPI_ISL_441813 | Department of Pathology, University of Cambridge | Wellcome Sanger Institute for the COVID-19 Genomics UK Consortium |
| EPI_ISL_441814 | Department of Pathology, University of Cambridge | Wellcome Sanger Institute for the COVID-19 Genomics UK Consortium |
| EPI_ISL_441815 | Department of Pathology, University of Cambridge | Wellcome Sanger Institute for the COVID-19 Genomics UK Consortium |
| EPI_ISL_441816 | Department of Pathology, University of Cambridge | Wellcome Sanger Institute for the COVID-19 Genomics UK Consortium |
| EPI_ISL_441817 | Department of Pathology, University of Cambridge | Wellcome Sanger Institute for the COVID-19 Genomics UK Consortium |
| EPI_ISL_441818 | Department of Pathology, University of Cambridge | Wellcome Sanger Institute for the COVID-19 Genomics UK Consortium |

| EPI_Accession_ID | Originating Laboratory | Submitting Laboratory |
|---|---|---|
| EPI_ISL_441819 | Department of Pathology, University of Cambridge | Wellcome Sanger Institute for the COVID-19 Genomics UK Consortium |
| EPI_ISL_441820 | Department of Pathology, University of Cambridge | Wellcome Sanger Institute for the COVID-19 Genomics UK Consortium |
| EPI_ISL_441821 | Department of Pathology, University of Cambridge | Wellcome Sanger Institute for the COVID-19 Genomics UK Consortium |
| EPI_ISL_441822 | Department of Pathology, University of Cambridge | Wellcome Sanger Institute for the COVID-19 Genomics UK Consortium |
| EPI_ISL_441823 | Department of Pathology, University of Cambridge | Wellcome Sanger Institute for the COVID-19 Genomics UK Consortium |
| EPI_ISL_441824 | Department of Pathology, University of Cambridge | Wellcome Sanger Institute for the COVID-19 Genomics UK Consortium |
| EPI_ISL_441825 | Department of Pathology, University of Cambridge | Wellcome Sanger Institute for the COVID-19 Genomics UK Consortium |
| EPI_ISL_441826 | Department of Pathology, University of Cambridge | Wellcome Sanger Institute for the COVID-19 Genomics UK Consortium |
| EPI_ISL_441827 | Department of Pathology, University of Cambridge | Wellcome Sanger Institute for the COVID-19 Genomics UK Consortium |
| EPI_ISL_441828 | Department of Pathology, University of Cambridge | Wellcome Sanger Institute for the COVID-19 Genomics UK Consortium |
| EPI_ISL_441829 | Department of Pathology, University of Cambridge | Wellcome Sanger Institute for the COVID-19 Genomics UK Consortium |
| EPI_ISL_441830 | Department of Pathology, University of Cambridge | Wellcome Sanger Institute for the COVID-19 Genomics UK Consortium |
| EPI_ISL_441831 | Department of Pathology, University of Cambridge | Wellcome Sanger Institute for the COVID-19 Genomics UK Consortium |
| EPI_ISL_441832 | Department of Pathology, University of Cambridge | Wellcome Sanger Institute for the COVID-19 Genomics UK Consortium |
| EPI_ISL_441833 | Department of Pathology, University of Cambridge | Wellcome Sanger Institute for the COVID-19 Genomics UK Consortium |
| EPI_ISL_441834 | Department of Pathology, University of Cambridge | Wellcome Sanger Institute for the COVID-19 Genomics UK Consortium |

| EPI_Accession_ID | Originating Laboratory | Submitting Laboratory |
|---|---|---|
| EPI_ISL_441835 | Department of Pathology, University of Cambridge | Wellcome Sanger Institute for the COVID-19 Genomics UK Consortium |
| EPI_ISL_441836 | Department of Pathology, University of Cambridge | Wellcome Sanger Institute for the COVID-19 Genomics UK Consortium |
| EPI_ISL_441837 | Department of Pathology, University of Cambridge | Wellcome Sanger Institute for the COVID-19 Genomics UK Consortium |
| EPI_ISL_441838 | Department of Pathology, University of Cambridge | Wellcome Sanger Institute for the COVID-19 Genomics UK Consortium |
| EPI_ISL_441839 | Department of Pathology, University of Cambridge | Wellcome Sanger Institute for the COVID-19 Genomics UK Consortium |
| EPI_ISL_441840 | Department of Pathology, University of Cambridge | Wellcome Sanger Institute for the COVID-19 Genomics UK Consortium |
| EPI_ISL_441841 | Department of Pathology, University of Cambridge | Wellcome Sanger Institute for the COVID-19 Genomics UK Consortium |
| EPI_ISL_441842 | Department of Pathology, University of Cambridge | Wellcome Sanger Institute for the COVID-19 Genomics UK Consortium |
| EPI_ISL_441843 | Department of Pathology, University of Cambridge | Wellcome Sanger Institute for the COVID-19 Genomics UK Consortium |
| EPI_ISL_441844 | Department of Pathology, University of Cambridge | Wellcome Sanger Institute for the COVID-19 Genomics UK Consortium |
| EPI_ISL_441845 | Department of Pathology, University of Cambridge | Wellcome Sanger Institute for the COVID-19 Genomics UK Consortium |
| EPI_ISL_441846 | Department of Pathology, University of Cambridge | Wellcome Sanger Institute for the COVID-19 Genomics UK Consortium |
| EPI_ISL_441847 | Queens Medical Centre, Clinical Microbiology Department / DeepSeq Nottingham | COVID-19 Genomics UK (COG-UK) Consortium |
| EPI_ISL_441848 | Queens Medical Centre, Clinical Microbiology Department / DeepSeq Nottingham | COVID-19 Genomics UK (COG-UK) Consortium |
| EPI_ISL_441849 | Queens Medical Centre, Clinical Microbiology Department / DeepSeq Nottingham | COVID-19 Genomics UK (COG-UK) Consortium |
| EPI_ISL_441850 | Queens Medical Centre, Clinical Microbiology Department / DeepSeq Nottingham | COVID-19 Genomics UK (COG-UK) Consortium |

| EPI_Accession_ID | Originating Laboratory | Submitting Laboratory |
|---|---|---|
| EPI_ISL_441851 | Queens Medical Centre, Clinical Microbiology Department / DeepSeq Nottingham | COVID-19 Genomics UK (COG-UK) Consortium |
| EPI_ISL_441852 | Queens Medical Centre, Clinical Microbiology Department / DeepSeq Nottingham | COVID-19 Genomics UK (COG-UK) Consortium |
| EPI_ISL_441853 | Queens Medical Centre, Clinical Microbiology Department / DeepSeq Nottingham | COVID-19 Genomics UK (COG-UK) Consortium |
| EPI_ISL_441854 | Queens Medical Centre, Clinical Microbiology Department / DeepSeq Nottingham | COVID-19 Genomics UK (COG-UK) Consortium |
| EPI_ISL_441855 | Queens Medical Centre, Clinical Microbiology Department / DeepSeq Nottingham | COVID-19 Genomics UK (COG-UK) Consortium |
| EPI_ISL_441856 | Queens Medical Centre, Clinical Microbiology Department / DeepSeq Nottingham | COVID-19 Genomics UK (COG-UK) Consortium |
| EPI_ISL_441857 | Queens Medical Centre, Clinical Microbiology Department / DeepSeq Nottingham | COVID-19 Genomics UK (COG-UK) Consortium |
| EPI_ISL_441858 | Queens Medical Centre, Clinical Microbiology Department / DeepSeq Nottingham | COVID-19 Genomics UK (COG-UK) Consortium |
| EPI_ISL_441859 | Queens Medical Centre, Clinical Microbiology Department / DeepSeq Nottingham | COVID-19 Genomics UK (COG-UK) Consortium |
| EPI_ISL_441860 | Queens Medical Centre, Clinical Microbiology Department / DeepSeq Nottingham | COVID-19 Genomics UK (COG-UK) Consortium |
| EPI_ISL_441861 | Queens Medical Centre, Clinical Microbiology Department / DeepSeq Nottingham | COVID-19 Genomics UK (COG-UK) Consortium |
| EPI_ISL_441862 | Queens Medical Centre, Clinical Microbiology Department / DeepSeq Nottingham | COVID-19 Genomics UK (COG-UK) Consortium |
| EPI_ISL_441863 | Queens Medical Centre, Clinical Microbiology Department / DeepSeq Nottingham | COVID-19 Genomics UK (COG-UK) Consortium |
| EPI_ISL_441864 | Queens Medical Centre, Clinical Microbiology Department / DeepSeq Nottingham | COVID-19 Genomics UK (COG-UK) Consortium |
| EPI_ISL_441865 | Queens Medical Centre, Clinical Microbiology Department / DeepSeq Nottingham | COVID-19 Genomics UK (COG-UK) Consortium |
| EPI_ISL_441866 | Queens Medical Centre, Clinical Microbiology Department / DeepSeq Nottingham | COVID-19 Genomics UK (COG-UK) Consortium |

| EPI_Accession_ID | Originating Laboratory | Submitting Laboratory |
|---|---|---|
| EPI_ISL_441867 | Queens Medical Centre, Clinical Microbiology Department / DeepSeq Nottingham | COVID-19 Genomics UK (COG-UK) Consortium |
| EPI_ISL_441868 | Queens Medical Centre, Clinical Microbiology Department / DeepSeq Nottingham | COVID-19 Genomics UK (COG-UK) Consortium |
| EPI_ISL_441869 | Queens Medical Centre, Clinical Microbiology Department / DeepSeq Nottingham | COVID-19 Genomics UK (COG-UK) Consortium |
| EPI_ISL_441870 | Queens Medical Centre, Clinical Microbiology Department / DeepSeq Nottingham | COVID-19 Genomics UK (COG-UK) Consortium |
| EPI_ISL_441871 | Queens Medical Centre, Clinical Microbiology Department / DeepSeq Nottingham | COVID-19 Genomics UK (COG-UK) Consortium |
| EPI_ISL_441872 | Queens Medical Centre, Clinical Microbiology Department / DeepSeq Nottingham | COVID-19 Genomics UK (COG-UK) Consortium |
| EPI_ISL_441873 | Queens Medical Centre, Clinical Microbiology Department / DeepSeq Nottingham | COVID-19 Genomics UK (COG-UK) Consortium |
| EPI_ISL_441874 | Queens Medical Centre, Clinical Microbiology Department / DeepSeq Nottingham | COVID-19 Genomics UK (COG-UK) Consortium |
| EPI_ISL_441875 | Queens Medical Centre, Clinical Microbiology Department / DeepSeq Nottingham | COVID-19 Genomics UK (COG-UK) Consortium |
| EPI_ISL_441876 | Queens Medical Centre, Clinical Microbiology Department / DeepSeq Nottingham | COVID-19 Genomics UK (COG-UK) Consortium |
| EPI_ISL_441877 | Queens Medical Centre, Clinical Microbiology Department / DeepSeq Nottingham | COVID-19 Genomics UK (COG-UK) Consortium |
| EPI_ISL_441878 | Queens Medical Centre, Clinical Microbiology Department / DeepSeq Nottingham | COVID-19 Genomics UK (COG-UK) Consortium |
| EPI_ISL_441879 | Queens Medical Centre, Clinical Microbiology Department / DeepSeq Nottingham | COVID-19 Genomics UK (COG-UK) Consortium |
| EPI_ISL_441880 | Queens Medical Centre, Clinical Microbiology Department / DeepSeq Nottingham | COVID-19 Genomics UK (COG-UK) Consortium |
| EPI_ISL_441881 | Queens Medical Centre, Clinical Microbiology Department / DeepSeq Nottingham | COVID-19 Genomics UK (COG-UK) Consortium |
| EPI_ISL_441882 | Queens Medical Centre, Clinical Microbiology Department / DeepSeq Nottingham | COVID-19 Genomics UK (COG-UK) Consortium |

| EPI_Accession_ID | Originating Laboratory | Submitting Laboratory |
|---|---|---|
| EPI_ISL_441883 | Queens Medical Centre, Clinical Microbiology Department / DeepSeq Nottingham | COVID-19 Genomics UK (COG-UK) Consortium |
| EPI_ISL_441884 | Queens Medical Centre, Clinical Microbiology Department / DeepSeq Nottingham | COVID-19 Genomics UK (COG-UK) Consortium |
| EPI_ISL_441885 | Queens Medical Centre, Clinical Microbiology Department / DeepSeq Nottingham | COVID-19 Genomics UK (COG-UK) Consortium |
| EPI_ISL_441886 | Queens Medical Centre, Clinical Microbiology Department / DeepSeq Nottingham | COVID-19 Genomics UK (COG-UK) Consortium |
| EPI_ISL_441887 | Queens Medical Centre, Clinical Microbiology Department / DeepSeq Nottingham | COVID-19 Genomics UK (COG-UK) Consortium |
| EPI_ISL_441888 | Queens Medical Centre, Clinical Microbiology Department / DeepSeq Nottingham | COVID-19 Genomics UK (COG-UK) Consortium |
| EPI_ISL_441889 | Queens Medical Centre, Clinical Microbiology Department / DeepSeq Nottingham | COVID-19 Genomics UK (COG-UK) Consortium |
| EPI_ISL_441890 | Queens Medical Centre, Clinical Microbiology Department / DeepSeq Nottingham | COVID-19 Genomics UK (COG-UK) Consortium |
| EPI_ISL_441891 | Queens Medical Centre, Clinical Microbiology Department / DeepSeq Nottingham | COVID-19 Genomics UK (COG-UK) Consortium |
| EPI_ISL_441892 | Queens Medical Centre, Clinical Microbiology Department / DeepSeq Nottingham | COVID-19 Genomics UK (COG-UK) Consortium |
| EPI_ISL_441893 | Queens Medical Centre, Clinical Microbiology Department / DeepSeq Nottingham | COVID-19 Genomics UK (COG-UK) Consortium |
| EPI_ISL_441894 | Queens Medical Centre, Clinical Microbiology Department / DeepSeq Nottingham | COVID-19 Genomics UK (COG-UK) Consortium |
| EPI_ISL_441895 | Queens Medical Centre, Clinical Microbiology Department / DeepSeq Nottingham | COVID-19 Genomics UK (COG-UK) Consortium |
| EPI_ISL_441896 | Queens Medical Centre, Clinical Microbiology Department / DeepSeq Nottingham | COVID-19 Genomics UK (COG-UK) Consortium |
| EPI_ISL_441897 | Queens Medical Centre, Clinical Microbiology Department / DeepSeq Nottingham | COVID-19 Genomics UK (COG-UK) Consortium |

| EPI_Accession_ID | Originating Laboratory | Submitting Laboratory |
|---|---|---|
| EPI_ISL_441898 | Virology Department, Sheffield Teaching Hospitals NHS Foundation Trust/Department of Infection, Immunity and Cardiovascular Disease, The Medical School, University of Sheffield | COVID-19 Genomics UK (COG-UK) Consortium |
| EPI_ISL_441899 | Virology Department, Sheffield Teaching Hospitals NHS Foundation Trust/Department of Infection, Immunity and Cardiovascular Disease, The Medical School, University of Sheffield | COVID-19 Genomics UK (COG-UK) Consortium |
| EPI_ISL_441900 | Virology Department, Sheffield Teaching Hospitals NHS Foundation Trust/Department of Infection, Immunity and Cardiovascular Disease, The Medical School, University of Sheffield | COVID-19 Genomics UK (COG-UK) Consortium |
| EPI_ISL_441901 | Virology Department, Sheffield Teaching Hospitals NHS Foundation Trust/Department of Infection, Immunity and Cardiovascular Disease, The Medical School, University of Sheffield | COVID-19 Genomics UK (COG-UK) Consortium |
| EPI_ISL_441902 | Virology Department, Sheffield Teaching Hospitals NHS Foundation Trust/Department of Infection, Immunity and Cardiovascular Disease, The Medical School, University of Sheffield | COVID-19 Genomics UK (COG-UK) Consortium |
| EPI_ISL_441903 | Virology Department, Sheffield Teaching Hospitals NHS Foundation Trust/Department of Infection, Immunity and Cardiovascular Disease, The Medical School, University of Sheffield | COVID-19 Genomics UK (COG-UK) Consortium |
| EPI_ISL_441904 | Virology Department, Sheffield Teaching Hospitals NHS Foundation Trust/Department of Infection, Immunity and Cardiovascular Disease, The Medical School, University of Sheffield | COVID-19 Genomics UK (COG-UK) Consortium |
| EPI_ISL_441905 | Virology Department, Sheffield Teaching Hospitals NHS Foundation Trust/Department of Infection, Immunity and Cardiovascular Disease, The Medical School, University of Sheffield | COVID-19 Genomics UK (COG-UK) Consortium |

| EPI_Accession_ID | Originating Laboratory | Submitting Laboratory |
| --- | --- | --- |
| EPI_ISL_441906 | Virology Department, Sheffield Teaching Hospitals NHS Foundation Trust/Department of Infection, Immunity and Cardiovascular Disease, The Medical School, University of Sheffield | COVID-19 Genomics UK (COG-UK) Consortium |
| EPI_ISL_441907 | Virology Department, Sheffield Teaching Hospitals NHS Foundation Trust/Department of Infection, Immunity and Cardiovascular Disease, The Medical School, University of Sheffield | COVID-19 Genomics UK (COG-UK) Consortium |
| EPI_ISL_441908 | Virology Department, Sheffield Teaching Hospitals NHS Foundation Trust/Department of Infection, Immunity and Cardiovascular Disease, The Medical School, University of Sheffield | COVID-19 Genomics UK (COG-UK) Consortium |
| EPI_ISL_441909 | Virology Department, Sheffield Teaching Hospitals NHS Foundation Trust/Department of Infection, Immunity and Cardiovascular Disease, The Medical School, University of Sheffield | COVID-19 Genomics UK (COG-UK) Consortium |
| EPI_ISL_441910 | Virology Department, Sheffield Teaching Hospitals NHS Foundation Trust/Department of Infection, Immunity and Cardiovascular Disease, The Medical School, University of Sheffield | COVID-19 Genomics UK (COG-UK) Consortium |
| EPI_ISL_441911 | Virology Department, Sheffield Teaching Hospitals NHS Foundation Trust/Department of Infection, Immunity and Cardiovascular Disease, The Medical School, University of Sheffield | COVID-19 Genomics UK (COG-UK) Consortium |
| EPI_ISL_441912 | Virology Department, Sheffield Teaching Hospitals NHS Foundation Trust/Department of Infection, Immunity and Cardiovascular Disease, The Medical School, University of Sheffield | COVID-19 Genomics UK (COG-UK) Consortium |
| EPI_ISL_441913 | Virology Department, Sheffield Teaching Hospitals NHS Foundation Trust/Department of Infection, Immunity and Cardiovascular Disease, The Medical School, University of Sheffield | COVID-19 Genomics UK (COG-UK) Consortium |

| EPI_Accession_ID | Originating Laboratory | Submitting Laboratory |
|---|---|---|
| EPI_ISL_441914 | Virology Department, Sheffield Teaching Hospitals NHS Foundation Trust/Department of Infection, Immunity and Cardiovascular Disease, The Medical School, University of Sheffield | COVID-19 Genomics UK (COG-UK) Consortium |
| EPI_ISL_441915 | Virology Department, Sheffield Teaching Hospitals NHS Foundation Trust/Department of Infection, Immunity and Cardiovascular Disease, The Medical School, University of Sheffield | COVID-19 Genomics UK (COG-UK) Consortium |
| EPI_ISL_441916 | Virology Department, Sheffield Teaching Hospitals NHS Foundation Trust/Department of Infection, Immunity and Cardiovascular Disease, The Medical School, University of Sheffield | COVID-19 Genomics UK (COG-UK) Consortium |
| EPI_ISL_441917 | Virology Department, Sheffield Teaching Hospitals NHS Foundation Trust/Department of Infection, Immunity and Cardiovascular Disease, The Medical School, University of Sheffield | COVID-19 Genomics UK (COG-UK) Consortium |
| EPI_ISL_441918 | Virology Department, Sheffield Teaching Hospitals NHS Foundation Trust/Department of Infection, Immunity and Cardiovascular Disease, The Medical School, University of Sheffield | COVID-19 Genomics UK (COG-UK) Consortium |
| EPI_ISL_441919 | Virology Department, Sheffield Teaching Hospitals NHS Foundation Trust/Department of Infection, Immunity and Cardiovascular Disease, The Medical School, University of Sheffield | COVID-19 Genomics UK (COG-UK) Consortium |
| EPI_ISL_441920 | Virology Department, Sheffield Teaching Hospitals NHS Foundation Trust/Department of Infection, Immunity and Cardiovascular Disease, The Medical School, University of Sheffield | COVID-19 Genomics UK (COG-UK) Consortium |
| EPI_ISL_441921 | Virology Department, Sheffield Teaching Hospitals NHS Foundation Trust/Department of Infection, Immunity and Cardiovascular Disease, The Medical School, University of Sheffield | COVID-19 Genomics UK (COG-UK) Consortium |

| EPI_Accession_ID | Originating Laboratory | Submitting Laboratory |
|---|---|---|
| EPI_ISL_441922 | Virology Department, Sheffield Teaching Hospitals NHS Foundation Trust/Department of Infection, Immunity and Cardiovascular Disease, The Medical School, University of Sheffield | COVID-19 Genomics UK (COG-UK) Consortium |
| EPI_ISL_441923 | Virology Department, Sheffield Teaching Hospitals NHS Foundation Trust/Department of Infection, Immunity and Cardiovascular Disease, The Medical School, University of Sheffield | COVID-19 Genomics UK (COG-UK) Consortium |
| EPI_ISL_441924 | Virology Department, Sheffield Teaching Hospitals NHS Foundation Trust/Department of Infection, Immunity and Cardiovascular Disease, The Medical School, University of Sheffield | COVID-19 Genomics UK (COG-UK) Consortium |
| EPI_ISL_441925 | Virology Department, Sheffield Teaching Hospitals NHS Foundation Trust/Department of Infection, Immunity and Cardiovascular Disease, The Medical School, University of Sheffield | COVID-19 Genomics UK (COG-UK) Consortium |
| EPI_ISL_441926 | Virology Department, Sheffield Teaching Hospitals NHS Foundation Trust/Department of Infection, Immunity and Cardiovascular Disease, The Medical School, University of Sheffield | COVID-19 Genomics UK (COG-UK) Consortium |
| EPI_ISL_441927 | Virology Department, Sheffield Teaching Hospitals NHS Foundation Trust/Department of Infection, Immunity and Cardiovascular Disease, The Medical School, University of Sheffield | COVID-19 Genomics UK (COG-UK) Consortium |
| EPI_ISL_441928 | Virology Department, Sheffield Teaching Hospitals NHS Foundation Trust/Department of Infection, Immunity and Cardiovascular Disease, The Medical School, University of Sheffield | COVID-19 Genomics UK (COG-UK) Consortium |
| EPI_ISL_441929 | Virology Department, Sheffield Teaching Hospitals NHS Foundation Trust/Department of Infection, Immunity and Cardiovascular Disease, The Medical School, University of Sheffield | COVID-19 Genomics UK (COG-UK) Consortium |

| EPI_Accession_ID | Originating Laboratory | Submitting Laboratory |
| --- | --- | --- |
| EPI_ISL_441930 | Virology Department, Sheffield Teaching Hospitals NHS Foundation Trust/Department of Infection, Immunity and Cardiovascular Disease, The Medical School, University of Sheffield | COVID-19 Genomics UK (COG-UK) Consortium |
| EPI_ISL_441931 | Virology Department, Sheffield Teaching Hospitals NHS Foundation Trust/Department of Infection, Immunity and Cardiovascular Disease, The Medical School, University of Sheffield | COVID-19 Genomics UK (COG-UK) Consortium |
| EPI_ISL_441932 | Virology Department, Sheffield Teaching Hospitals NHS Foundation Trust/Department of Infection, Immunity and Cardiovascular Disease, The Medical School, University of Sheffield | COVID-19 Genomics UK (COG-UK) Consortium |
| EPI_ISL_441933 | Virology Department, Sheffield Teaching Hospitals NHS Foundation Trust/Department of Infection, Immunity and Cardiovascular Disease, The Medical School, University of Sheffield | COVID-19 Genomics UK (COG-UK) Consortium |
| EPI_ISL_441934 | Virology Department, Sheffield Teaching Hospitals NHS Foundation Trust/Department of Infection, Immunity and Cardiovascular Disease, The Medical School, University of Sheffield | COVID-19 Genomics UK (COG-UK) Consortium |
| EPI_ISL_441935 | Virology Department, Sheffield Teaching Hospitals NHS Foundation Trust/Department of Infection, Immunity and Cardiovascular Disease, The Medical School, University of Sheffield | COVID-19 Genomics UK (COG-UK) Consortium |
| EPI_ISL_441936 | Virology Department, Sheffield Teaching Hospitals NHS Foundation Trust/Department of Infection, Immunity and Cardiovascular Disease, The Medical School, University of Sheffield | COVID-19 Genomics UK (COG-UK) Consortium |
| EPI_ISL_441937 | Virology Department, Sheffield Teaching Hospitals NHS Foundation Trust/Department of Infection, Immunity and Cardiovascular Disease, The Medical School, University of Sheffield | COVID-19 Genomics UK (COG-UK) Consortium |

| EPI_Accession_ID | Originating Laboratory | Submitting Laboratory |
|---|---|---|
| EPI_ISL_441938 | Virology Department, Sheffield Teaching Hospitals NHS Foundation Trust/Department of Infection, Immunity and Cardiovascular Disease, The Medical School, University of Sheffield | COVID-19 Genomics UK (COG-UK) Consortium |
| EPI_ISL_441939 | Virology Department, Sheffield Teaching Hospitals NHS Foundation Trust/Department of Infection, Immunity and Cardiovascular Disease, The Medical School, University of Sheffield | COVID-19 Genomics UK (COG-UK) Consortium |
| EPI_ISL_441940 | Virology Department, Sheffield Teaching Hospitals NHS Foundation Trust/Department of Infection, Immunity and Cardiovascular Disease, The Medical School, University of Sheffield | COVID-19 Genomics UK (COG-UK) Consortium |
| EPI_ISL_441941 | Virology Department, Sheffield Teaching Hospitals NHS Foundation Trust/Department of Infection, Immunity and Cardiovascular Disease, The Medical School, University of Sheffield | COVID-19 Genomics UK (COG-UK) Consortium |
| EPI_ISL_441942 | Virology Department, Sheffield Teaching Hospitals NHS Foundation Trust/Department of Infection, Immunity and Cardiovascular Disease, The Medical School, University of Sheffield | COVID-19 Genomics UK (COG-UK) Consortium |
| EPI_ISL_441943 | Virology Department, Sheffield Teaching Hospitals NHS Foundation Trust/Department of Infection, Immunity and Cardiovascular Disease, The Medical School, University of Sheffield | COVID-19 Genomics UK (COG-UK) Consortium |
| EPI_ISL_441944 | Virology Department, Sheffield Teaching Hospitals NHS Foundation Trust/Department of Infection, Immunity and Cardiovascular Disease, The Medical School, University of Sheffield | COVID-19 Genomics UK (COG-UK) Consortium |
| EPI_ISL_441945 | Virology Department, Sheffield Teaching Hospitals NHS Foundation Trust/Department of Infection, Immunity and Cardiovascular Disease, The Medical School, University of Sheffield | COVID-19 Genomics UK (COG-UK) Consortium |

| EPI_Accession_ID | Originating Laboratory | Submitting Laboratory |
|---|---|---|
| EPI_ISL_441946 | Virology Department, Sheffield Teaching Hospitals NHS Foundation Trust/Department of Infection, Immunity and Cardiovascular Disease, The Medical School, University of Sheffield | COVID-19 Genomics UK (COG-UK) Consortium |
| EPI_ISL_441947 | Virology Department, Sheffield Teaching Hospitals NHS Foundation Trust/Department of Infection, Immunity and Cardiovascular Disease, The Medical School, University of Sheffield | COVID-19 Genomics UK (COG-UK) Consortium |
| EPI_ISL_441948 | Virology Department, Sheffield Teaching Hospitals NHS Foundation Trust/Department of Infection, Immunity and Cardiovascular Disease, The Medical School, University of Sheffield | COVID-19 Genomics UK (COG-UK) Consortium |
| EPI_ISL_441949 | Virology Department, Sheffield Teaching Hospitals NHS Foundation Trust/Department of Infection, Immunity and Cardiovascular Disease, The Medical School, University of Sheffield | COVID-19 Genomics UK (COG-UK) Consortium |
| EPI_ISL_441950 | Virology Department, Sheffield Teaching Hospitals NHS Foundation Trust/Department of Infection, Immunity and Cardiovascular Disease, The Medical School, University of Sheffield | COVID-19 Genomics UK (COG-UK) Consortium |
| EPI_ISL_441951 | Virology Department, Sheffield Teaching Hospitals NHS Foundation Trust/Department of Infection, Immunity and Cardiovascular Disease, The Medical School, University of Sheffield | COVID-19 Genomics UK (COG-UK) Consortium |
| EPI_ISL_441952 | Virology Department, Sheffield Teaching Hospitals NHS Foundation Trust/Department of Infection, Immunity and Cardiovascular Disease, The Medical School, University of Sheffield | COVID-19 Genomics UK (COG-UK) Consortium |
| EPI_ISL_441953 | Virology Department, Sheffield Teaching Hospitals NHS Foundation Trust/Department of Infection, Immunity and Cardiovascular Disease, The Medical School, University of Sheffield | COVID-19 Genomics UK (COG-UK) Consortium |

| EPI_Accession_ID | Originating Laboratory | Submitting Laboratory |
|---|---|---|
| EPI_ISL_441954 | Virology Department, Sheffield Teaching Hospitals NHS Foundation Trust/Department of Infection, Immunity and Cardiovascular Disease, The Medical School, University of Sheffield | COVID-19 Genomics UK (COG-UK) Consortium |
| EPI_ISL_441955 | Virology Department, Sheffield Teaching Hospitals NHS Foundation Trust/Department of Infection, Immunity and Cardiovascular Disease, The Medical School, University of Sheffield | COVID-19 Genomics UK (COG-UK) Consortium |
| EPI_ISL_441956 | Virology Department, Sheffield Teaching Hospitals NHS Foundation Trust/Department of Infection, Immunity and Cardiovascular Disease, The Medical School, University of Sheffield | COVID-19 Genomics UK (COG-UK) Consortium |
| EPI_ISL_441957 | Virology Department, Sheffield Teaching Hospitals NHS Foundation Trust/Department of Infection, Immunity and Cardiovascular Disease, The Medical School, University of Sheffield | COVID-19 Genomics UK (COG-UK) Consortium |
| EPI_ISL_441958 | Virology Department, Sheffield Teaching Hospitals NHS Foundation Trust/Department of Infection, Immunity and Cardiovascular Disease, The Medical School, University of Sheffield | COVID-19 Genomics UK (COG-UK) Consortium |
| EPI_ISL_441959 | Virology Department, Sheffield Teaching Hospitals NHS Foundation Trust/Department of Infection, Immunity and Cardiovascular Disease, The Medical School, University of Sheffield | COVID-19 Genomics UK (COG-UK) Consortium |
| EPI_ISL_441960 | Virology Department, Sheffield Teaching Hospitals NHS Foundation Trust/Department of Infection, Immunity and Cardiovascular Disease, The Medical School, University of Sheffield | COVID-19 Genomics UK (COG-UK) Consortium |
| EPI_ISL_441961 | Virology Department, Sheffield Teaching Hospitals NHS Foundation Trust/Department of Infection, Immunity and Cardiovascular Disease, The Medical School, University of Sheffield | COVID-19 Genomics UK (COG-UK) Consortium |

| EPI_Accession_ID | Originating Laboratory | Submitting Laboratory |
|---|---|---|
| EPI_ISL_441962 | Virology Department, Sheffield Teaching Hospitals NHS Foundation Trust/Department of Infection, Immunity and Cardiovascular Disease, The Medical School, University of Sheffield | COVID-19 Genomics UK (COG-UK) Consortium |
| EPI_ISL_441963 | Virology Department, Sheffield Teaching Hospitals NHS Foundation Trust/Department of Infection, Immunity and Cardiovascular Disease, The Medical School, University of Sheffield | COVID-19 Genomics UK (COG-UK) Consortium |
| EPI_ISL_441964 | Virology Department, Sheffield Teaching Hospitals NHS Foundation Trust/Department of Infection, Immunity and Cardiovascular Disease, The Medical School, University of Sheffield | COVID-19 Genomics UK (COG-UK) Consortium |
| EPI_ISL_441965 | Virology Department, Sheffield Teaching Hospitals NHS Foundation Trust/Department of Infection, Immunity and Cardiovascular Disease, The Medical School, University of Sheffield | COVID-19 Genomics UK (COG-UK) Consortium |
| EPI_ISL_441966 | Virology Department, Sheffield Teaching Hospitals NHS Foundation Trust/Department of Infection, Immunity and Cardiovascular Disease, The Medical School, University of Sheffield | COVID-19 Genomics UK (COG-UK) Consortium |
| EPI_ISL_441967 | Virology Department, Sheffield Teaching Hospitals NHS Foundation Trust/Department of Infection, Immunity and Cardiovascular Disease, The Medical School, University of Sheffield | COVID-19 Genomics UK (COG-UK) Consortium |
| EPI_ISL_441968 | Virology Department, Sheffield Teaching Hospitals NHS Foundation Trust/Department of Infection, Immunity and Cardiovascular Disease, The Medical School, University of Sheffield | COVID-19 Genomics UK (COG-UK) Consortium |
| EPI_ISL_441969 | Virology Department, Sheffield Teaching Hospitals NHS Foundation Trust/Department of Infection, Immunity and Cardiovascular Disease, The Medical School, University of Sheffield | COVID-19 Genomics UK (COG-UK) Consortium |

| EPI_Accession_ID | Originating Laboratory | Submitting Laboratory |
|---|---|---|
| EPI_ISL_441970 | Virology Department, Sheffield Teaching Hospitals NHS Foundation Trust/Department of Infection, Immunity and Cardiovascular Disease, The Medical School, University of Sheffield | COVID-19 Genomics UK (COG-UK) Consortium |
| EPI_ISL_441971 | Virology Department, Sheffield Teaching Hospitals NHS Foundation Trust/Department of Infection, Immunity and Cardiovascular Disease, The Medical School, University of Sheffield | COVID-19 Genomics UK (COG-UK) Consortium |
| EPI_ISL_441972 | Virology Department, Sheffield Teaching Hospitals NHS Foundation Trust/Department of Infection, Immunity and Cardiovascular Disease, The Medical School, University of Sheffield | COVID-19 Genomics UK (COG-UK) Consortium |
| EPI_ISL_441973 | Virology Department, Sheffield Teaching Hospitals NHS Foundation Trust/Department of Infection, Immunity and Cardiovascular Disease, The Medical School, University of Sheffield | COVID-19 Genomics UK (COG-UK) Consortium |
| EPI_ISL_441974 | Virology Department, Sheffield Teaching Hospitals NHS Foundation Trust/Department of Infection, Immunity and Cardiovascular Disease, The Medical School, University of Sheffield | COVID-19 Genomics UK (COG-UK) Consortium |
| EPI_ISL_441975 | Virology Department, Sheffield Teaching Hospitals NHS Foundation Trust/Department of Infection, Immunity and Cardiovascular Disease, The Medical School, University of Sheffield | COVID-19 Genomics UK (COG-UK) Consortium |
| EPI_ISL_441976 | Virology Department, Sheffield Teaching Hospitals NHS Foundation Trust/Department of Infection, Immunity and Cardiovascular Disease, The Medical School, University of Sheffield | COVID-19 Genomics UK (COG-UK) Consortium |
| EPI_ISL_441977 | Virology Department, Sheffield Teaching Hospitals NHS Foundation Trust/Department of Infection, Immunity and Cardiovascular Disease, The Medical School, University of Sheffield | COVID-19 Genomics UK (COG-UK) Consortium |

| EPI_Accession_ID | Originating Laboratory | Submitting Laboratory |
|---|---|---|
| EPI_ISL_441978 | Virology Department, Sheffield Teaching Hospitals NHS Foundation Trust/Department of Infection, Immunity and Cardiovascular Disease, The Medical School, University of Sheffield | COVID-19 Genomics UK (COG-UK) Consortium |
| EPI_ISL_441979 | Virology Department, Sheffield Teaching Hospitals NHS Foundation Trust/Department of Infection, Immunity and Cardiovascular Disease, The Medical School, University of Sheffield | COVID-19 Genomics UK (COG-UK) Consortium |
| EPI_ISL_441980 | Virology Department, Sheffield Teaching Hospitals NHS Foundation Trust/Department of Infection, Immunity and Cardiovascular Disease, The Medical School, University of Sheffield | COVID-19 Genomics UK (COG-UK) Consortium |
| EPI_ISL_441981 | Virology Department, Sheffield Teaching Hospitals NHS Foundation Trust/Department of Infection, Immunity and Cardiovascular Disease, The Medical School, University of Sheffield | COVID-19 Genomics UK (COG-UK) Consortium |
| EPI_ISL_441982 | Virology Department, Sheffield Teaching Hospitals NHS Foundation Trust/Department of Infection, Immunity and Cardiovascular Disease, The Medical School, University of Sheffield | COVID-19 Genomics UK (COG-UK) Consortium |
| EPI_ISL_441983 | Virology Department, Sheffield Teaching Hospitals NHS Foundation Trust/Department of Infection, Immunity and Cardiovascular Disease, The Medical School, University of Sheffield | COVID-19 Genomics UK (COG-UK) Consortium |
| EPI_ISL_441984 | Virology Department, Sheffield Teaching Hospitals NHS Foundation Trust/Department of Infection, Immunity and Cardiovascular Disease, The Medical School, University of Sheffield | COVID-19 Genomics UK (COG-UK) Consortium |
| EPI_ISL_441985 | Virology Department, Sheffield Teaching Hospitals NHS Foundation Trust/Department of Infection, Immunity and Cardiovascular Disease, The Medical School, University of Sheffield | COVID-19 Genomics UK (COG-UK) Consortium |

| EPI_Accession_ID | Originating Laboratory | Submitting Laboratory |
|---|---|---|
| EPI_ISL_441986 | Virology Department, Sheffield Teaching Hospitals NHS Foundation Trust/Department of Infection, Immunity and Cardiovascular Disease, The Medical School, University of Sheffield | COVID-19 Genomics UK (COG-UK) Consortium |
| EPI_ISL_441987 | Virology Department, Sheffield Teaching Hospitals NHS Foundation Trust/Department of Infection, Immunity and Cardiovascular Disease, The Medical School, University of Sheffield | COVID-19 Genomics UK (COG-UK) Consortium |
| EPI_ISL_441988 | Virology Department, Sheffield Teaching Hospitals NHS Foundation Trust/Department of Infection, Immunity and Cardiovascular Disease, The Medical School, University of Sheffield | COVID-19 Genomics UK (COG-UK) Consortium |
| EPI_ISL_441989 | Virology Department, Sheffield Teaching Hospitals NHS Foundation Trust/Department of Infection, Immunity and Cardiovascular Disease, The Medical School, University of Sheffield | COVID-19 Genomics UK (COG-UK) Consortium |
| EPI_ISL_441990 | Virology Department, Sheffield Teaching Hospitals NHS Foundation Trust/Department of Infection, Immunity and Cardiovascular Disease, The Medical School, University of Sheffield | COVID-19 Genomics UK (COG-UK) Consortium |
| EPI_ISL_441991 | Virology Department, Sheffield Teaching Hospitals NHS Foundation Trust/Department of Infection, Immunity and Cardiovascular Disease, The Medical School, University of Sheffield | COVID-19 Genomics UK (COG-UK) Consortium |
| EPI_ISL_441992 | Virology Department, Sheffield Teaching Hospitals NHS Foundation Trust/Department of Infection, Immunity and Cardiovascular Disease, The Medical School, University of Sheffield | COVID-19 Genomics UK (COG-UK) Consortium |
| EPI_ISL_441993 | Virology Department, Sheffield Teaching Hospitals NHS Foundation Trust/Department of Infection, Immunity and Cardiovascular Disease, The Medical School, University of Sheffield | COVID-19 Genomics UK (COG-UK) Consortium |

| EPI_Accession_ID | Originating Laboratory | Submitting Laboratory |
| --- | --- | --- |
| EPI_ISL_441994 | Virology Department, Sheffield Teaching Hospitals NHS Foundation Trust/Department of Infection, Immunity and Cardiovascular Disease, The Medical School, University of Sheffield | COVID-19 Genomics UK (COG-UK) Consortium |
| EPI_ISL_441995 | Virology Department, Sheffield Teaching Hospitals NHS Foundation Trust/Department of Infection, Immunity and Cardiovascular Disease, The Medical School, University of Sheffield | COVID-19 Genomics UK (COG-UK) Consortium |
| EPI_ISL_441996 | Virology Department, Sheffield Teaching Hospitals NHS Foundation Trust/Department of Infection, Immunity and Cardiovascular Disease, The Medical School, University of Sheffield | COVID-19 Genomics UK (COG-UK) Consortium |
| EPI_ISL_441997 | Virology Department, Sheffield Teaching Hospitals NHS Foundation Trust/Department of Infection, Immunity and Cardiovascular Disease, The Medical School, University of Sheffield | COVID-19 Genomics UK (COG-UK) Consortium |
| EPI_ISL_441998 | Virology Department, Sheffield Teaching Hospitals NHS Foundation Trust/Department of Infection, Immunity and Cardiovascular Disease, The Medical School, University of Sheffield | COVID-19 Genomics UK (COG-UK) Consortium |
| EPI_ISL_441999 | Virology Department, Sheffield Teaching Hospitals NHS Foundation Trust/Department of Infection, Immunity and Cardiovascular Disease, The Medical School, University of Sheffield | COVID-19 Genomics UK (COG-UK) Consortium |
| EPI_ISL_442000 | Virology Department, Sheffield Teaching Hospitals NHS Foundation Trust/Department of Infection, Immunity and Cardiovascular Disease, The Medical School, University of Sheffield | COVID-19 Genomics UK (COG-UK) Consortium |
| EPI_ISL_442001 | Virology Department, Sheffield Teaching Hospitals NHS Foundation Trust/Department of Infection, Immunity and Cardiovascular Disease, The Medical School, University of Sheffield | COVID-19 Genomics UK (COG-UK) Consortium |

| EPI_Accession_ID | Originating Laboratory | Submitting Laboratory |
|---|---|---|
| EPI_ISL_442002 | Virology Department, Sheffield Teaching Hospitals NHS Foundation Trust/Department of Infection, Immunity and Cardiovascular Disease, The Medical School, University of Sheffield | COVID-19 Genomics UK (COG-UK) Consortium |
| EPI_ISL_442003 | Virology Department, Sheffield Teaching Hospitals NHS Foundation Trust/Department of Infection, Immunity and Cardiovascular Disease, The Medical School, University of Sheffield | COVID-19 Genomics UK (COG-UK) Consortium |
| EPI_ISL_442004 | Virology Department, Sheffield Teaching Hospitals NHS Foundation Trust/Department of Infection, Immunity and Cardiovascular Disease, The Medical School, University of Sheffield | COVID-19 Genomics UK (COG-UK) Consortium |
| EPI_ISL_442005 | Virology Department, Sheffield Teaching Hospitals NHS Foundation Trust/Department of Infection, Immunity and Cardiovascular Disease, The Medical School, University of Sheffield | COVID-19 Genomics UK (COG-UK) Consortium |
| EPI_ISL_442006 | Virology Department, Sheffield Teaching Hospitals NHS Foundation Trust/Department of Infection, Immunity and Cardiovascular Disease, The Medical School, University of Sheffield | COVID-19 Genomics UK (COG-UK) Consortium |
| EPI_ISL_442007 | Virology Department, Sheffield Teaching Hospitals NHS Foundation Trust/Department of Infection, Immunity and Cardiovascular Disease, The Medical School, University of Sheffield | COVID-19 Genomics UK (COG-UK) Consortium |
| EPI_ISL_442008 | Virology Department, Sheffield Teaching Hospitals NHS Foundation Trust/Department of Infection, Immunity and Cardiovascular Disease, The Medical School, University of Sheffield | COVID-19 Genomics UK (COG-UK) Consortium |
| EPI_ISL_442009 | Virology Department, Sheffield Teaching Hospitals NHS Foundation Trust/Department of Infection, Immunity and Cardiovascular Disease, The Medical School, University of Sheffield | COVID-19 Genomics UK (COG-UK) Consortium |

| EPI_Accession_ID | Originating Laboratory | Submitting Laboratory |
|---|---|---|
| EPI_ISL_442010 | Virology Department, Sheffield Teaching Hospitals NHS Foundation Trust/Department of Infection, Immunity and Cardiovascular Disease, The Medical School, University of Sheffield | COVID-19 Genomics UK (COG-UK) Consortium |
| EPI_ISL_442011 | Virology Department, Sheffield Teaching Hospitals NHS Foundation Trust/Department of Infection, Immunity and Cardiovascular Disease, The Medical School, University of Sheffield | COVID-19 Genomics UK (COG-UK) Consortium |
| EPI_ISL_442012 | Virology Department, Sheffield Teaching Hospitals NHS Foundation Trust/Department of Infection, Immunity and Cardiovascular Disease, The Medical School, University of Sheffield | COVID-19 Genomics UK (COG-UK) Consortium |
| EPI_ISL_442013 | Virology Department, Sheffield Teaching Hospitals NHS Foundation Trust/Department of Infection, Immunity and Cardiovascular Disease, The Medical School, University of Sheffield | COVID-19 Genomics UK (COG-UK) Consortium |
| EPI_ISL_442014 | Virology Department, Sheffield Teaching Hospitals NHS Foundation Trust/Department of Infection, Immunity and Cardiovascular Disease, The Medical School, University of Sheffield | COVID-19 Genomics UK (COG-UK) Consortium |
| EPI_ISL_442015 | Virology Department, Sheffield Teaching Hospitals NHS Foundation Trust/Department of Infection, Immunity and Cardiovascular Disease, The Medical School, University of Sheffield | COVID-19 Genomics UK (COG-UK) Consortium |
| EPI_ISL_442016 | Virology Department, Sheffield Teaching Hospitals NHS Foundation Trust/Department of Infection, Immunity and Cardiovascular Disease, The Medical School, University of Sheffield | COVID-19 Genomics UK (COG-UK) Consortium |
| EPI_ISL_442017 | Virology Department, Sheffield Teaching Hospitals NHS Foundation Trust/Department of Infection, Immunity and Cardiovascular Disease, The Medical School, University of Sheffield | COVID-19 Genomics UK (COG-UK) Consortium |

| EPI_Accession_ID | Originating Laboratory | Submitting Laboratory |
|---|---|---|
| EPI_ISL_442018 | Virology Department, Sheffield Teaching Hospitals NHS Foundation Trust/Department of Infection, Immunity and Cardiovascular Disease, The Medical School, University of Sheffield | COVID-19 Genomics UK (COG-UK) Consortium |
| EPI_ISL_442019 | Virology Department, Sheffield Teaching Hospitals NHS Foundation Trust/Department of Infection, Immunity and Cardiovascular Disease, The Medical School, University of Sheffield | COVID-19 Genomics UK (COG-UK) Consortium |
| EPI_ISL_442020 | Virology Department, Sheffield Teaching Hospitals NHS Foundation Trust/Department of Infection, Immunity and Cardiovascular Disease, The Medical School, University of Sheffield | COVID-19 Genomics UK (COG-UK) Consortium |
| EPI_ISL_442021 | Virology Department, Sheffield Teaching Hospitals NHS Foundation Trust/Department of Infection, Immunity and Cardiovascular Disease, The Medical School, University of Sheffield | COVID-19 Genomics UK (COG-UK) Consortium |
| EPI_ISL_442022 | Virology Department, Sheffield Teaching Hospitals NHS Foundation Trust/Department of Infection, Immunity and Cardiovascular Disease, The Medical School, University of Sheffield | COVID-19 Genomics UK (COG-UK) Consortium |
| EPI_ISL_442023 | Virology Department, Sheffield Teaching Hospitals NHS Foundation Trust/Department of Infection, Immunity and Cardiovascular Disease, The Medical School, University of Sheffield | COVID-19 Genomics UK (COG-UK) Consortium |
| EPI_ISL_442024 | Virology Department, Sheffield Teaching Hospitals NHS Foundation Trust/Department of Infection, Immunity and Cardiovascular Disease, The Medical School, University of Sheffield | COVID-19 Genomics UK (COG-UK) Consortium |
| EPI_ISL_442025 | Virology Department, Sheffield Teaching Hospitals NHS Foundation Trust/Department of Infection, Immunity and Cardiovascular Disease, The Medical School, University of Sheffield | COVID-19 Genomics UK (COG-UK) Consortium |

| EPI_Accession_ID | Originating Laboratory | Submitting Laboratory |
|---|---|---|
| EPI_ISL_442026 | Virology Department, Sheffield Teaching Hospitals NHS Foundation Trust/Department of Infection, Immunity and Cardiovascular Disease, The Medical School, University of Sheffield | COVID-19 Genomics UK (COG-UK) Consortium |
| EPI_ISL_442027 | Virology Department, Sheffield Teaching Hospitals NHS Foundation Trust/Department of Infection, Immunity and Cardiovascular Disease, The Medical School, University of Sheffield | COVID-19 Genomics UK (COG-UK) Consortium |
| EPI_ISL_442028 | Virology Department, Sheffield Teaching Hospitals NHS Foundation Trust/Department of Infection, Immunity and Cardiovascular Disease, The Medical School, University of Sheffield | COVID-19 Genomics UK (COG-UK) Consortium |
| EPI_ISL_442029 | Virology Department, Sheffield Teaching Hospitals NHS Foundation Trust/Department of Infection, Immunity and Cardiovascular Disease, The Medical School, University of Sheffield | COVID-19 Genomics UK (COG-UK) Consortium |
| EPI_ISL_442030 | Virology Department, Sheffield Teaching Hospitals NHS Foundation Trust/Department of Infection, Immunity and Cardiovascular Disease, The Medical School, University of Sheffield | COVID-19 Genomics UK (COG-UK) Consortium |
| EPI_ISL_442031 | Virology Department, Sheffield Teaching Hospitals NHS Foundation Trust/Department of Infection, Immunity and Cardiovascular Disease, The Medical School, University of Sheffield | COVID-19 Genomics UK (COG-UK) Consortium |
| EPI_ISL_442032 | Virology Department, Sheffield Teaching Hospitals NHS Foundation Trust/Department of Infection, Immunity and Cardiovascular Disease, The Medical School, University of Sheffield | COVID-19 Genomics UK (COG-UK) Consortium |
| EPI_ISL_442033 | Virology Department, Sheffield Teaching Hospitals NHS Foundation Trust/Department of Infection, Immunity and Cardiovascular Disease, The Medical School, University of Sheffield | COVID-19 Genomics UK (COG-UK) Consortium |

| EPI_Accession_ID | Originating Laboratory | Submitting Laboratory |
|---|---|---|
| EPI_ISL_442034 | Virology Department, Sheffield Teaching Hospitals NHS Foundation Trust/Department of Infection, Immunity and Cardiovascular Disease, The Medical School, University of Sheffield | COVID-19 Genomics UK (COG-UK) Consortium |
| EPI_ISL_442035 | Virology Department, Sheffield Teaching Hospitals NHS Foundation Trust/Department of Infection, Immunity and Cardiovascular Disease, The Medical School, University of Sheffield | COVID-19 Genomics UK (COG-UK) Consortium |
| EPI_ISL_442036 | Virology Department, Sheffield Teaching Hospitals NHS Foundation Trust/Department of Infection, Immunity and Cardiovascular Disease, The Medical School, University of Sheffield | COVID-19 Genomics UK (COG-UK) Consortium |
| EPI_ISL_442037 | Virology Department, Sheffield Teaching Hospitals NHS Foundation Trust/Department of Infection, Immunity and Cardiovascular Disease, The Medical School, University of Sheffield | COVID-19 Genomics UK (COG-UK) Consortium |
| EPI_ISL_442038 | Virology Department, Sheffield Teaching Hospitals NHS Foundation Trust/Department of Infection, Immunity and Cardiovascular Disease, The Medical School, University of Sheffield | COVID-19 Genomics UK (COG-UK) Consortium |
| EPI_ISL_442039 | Virology Department, Sheffield Teaching Hospitals NHS Foundation Trust/Department of Infection, Immunity and Cardiovascular Disease, The Medical School, University of Sheffield | COVID-19 Genomics UK (COG-UK) Consortium |
| EPI_ISL_442040 | Virology Department, Sheffield Teaching Hospitals NHS Foundation Trust/Department of Infection, Immunity and Cardiovascular Disease, The Medical School, University of Sheffield | COVID-19 Genomics UK (COG-UK) Consortium |
| EPI_ISL_442041 | Virology Department, Sheffield Teaching Hospitals NHS Foundation Trust/Department of Infection, Immunity and Cardiovascular Disease, The Medical School, University of Sheffield | COVID-19 Genomics UK (COG-UK) Consortium |

| EPI_Accession_ID | Originating Laboratory | Submitting Laboratory |
|---|---|---|
| EPI_ISL_442042 | Virology Department, Sheffield Teaching Hospitals NHS Foundation Trust/Department of Infection, Immunity and Cardiovascular Disease, The Medical School, University of Sheffield | COVID-19 Genomics UK (COG-UK) Consortium |
| EPI_ISL_442043 | Virology Department, Sheffield Teaching Hospitals NHS Foundation Trust/Department of Infection, Immunity and Cardiovascular Disease, The Medical School, University of Sheffield | COVID-19 Genomics UK (COG-UK) Consortium |
| EPI_ISL_442044 | Kawsar Human Genetic Research Center | Kawsar Human Genetic Research Center |
| EPI_ISL_442045 | Department of Pathology, University of Cambridge | Wellcome Sanger Institute for the COVID-19 Genomics UK Consortium |
| EPI_ISL_442046 | Department of Pathology, University of Cambridge | Wellcome Sanger Institute for the COVID-19 Genomics UK Consortium |
| EPI_ISL_442047 | Department of Pathology, University of Cambridge | Wellcome Sanger Institute for the COVID-19 Genomics UK Consortium |
| EPI_ISL_442048 | Department of Pathology, University of Cambridge | Wellcome Sanger Institute for the COVID-19 Genomics UK Consortium |
| EPI_ISL_442049 | Department of Pathology, University of Cambridge | Wellcome Sanger Institute for the COVID-19 Genomics UK Consortium |
| EPI_ISL_442050 | Department of Pathology, University of Cambridge | Wellcome Sanger Institute for the COVID-19 Genomics UK Consortium |
| EPI_ISL_442051 | Department of Pathology, University of Cambridge | Wellcome Sanger Institute for the COVID-19 Genomics UK Consortium |
| EPI_ISL_442052 | Department of Pathology, University of Cambridge | Wellcome Sanger Institute for the COVID-19 Genomics UK Consortium |
| EPI_ISL_442053 | Department of Pathology, University of Cambridge | Wellcome Sanger Institute for the COVID-19 Genomics UK Consortium |
| EPI_ISL_442054 | Department of Pathology, University of Cambridge | Wellcome Sanger Institute for the COVID-19 Genomics UK Consortium |
| EPI_ISL_442055 | Department of Pathology, University of Cambridge | Wellcome Sanger Institute for the COVID-19 Genomics UK Consortium |
| EPI_ISL_442056 | Department of Pathology, University of Cambridge | Wellcome Sanger Institute for the COVID-19 Genomics UK Consortium |

| EPI_Accession_ID | Originating Laboratory | Submitting Laboratory |
|---|---|---|
| EPI_ISL_442057 | Department of Pathology, University of Cambridge | Wellcome Sanger Institute for the COVID-19 Genomics UK Consortium |
| EPI_ISL_442058 | Department of Pathology, University of Cambridge | Wellcome Sanger Institute for the COVID-19 Genomics UK Consortium |
| EPI_ISL_442059 | Department of Pathology, University of Cambridge | Wellcome Sanger Institute for the COVID-19 Genomics UK Consortium |
| EPI_ISL_442060 | Department of Pathology, University of Cambridge | Wellcome Sanger Institute for the COVID-19 Genomics UK Consortium |
| EPI_ISL_442061 | Department of Pathology, University of Cambridge | Wellcome Sanger Institute for the COVID-19 Genomics UK Consortium |
| EPI_ISL_442062 | Department of Pathology, University of Cambridge | Wellcome Sanger Institute for the COVID-19 Genomics UK Consortium |
| EPI_ISL_442063 | Department of Pathology, University of Cambridge | Wellcome Sanger Institute for the COVID-19 Genomics UK Consortium |
| EPI_ISL_442064 | Department of Pathology, University of Cambridge | Wellcome Sanger Institute for the COVID-19 Genomics UK Consortium |
| EPI_ISL_442065 | Department of Pathology, University of Cambridge | Wellcome Sanger Institute for the COVID-19 Genomics UK Consortium |
| EPI_ISL_442066 | Department of Pathology, University of Cambridge | Wellcome Sanger Institute for the COVID-19 Genomics UK Consortium |
| EPI_ISL_442067 | Department of Pathology, University of Cambridge | Wellcome Sanger Institute for the COVID-19 Genomics UK Consortium |
| EPI_ISL_442068 | Department of Pathology, University of Cambridge | Wellcome Sanger Institute for the COVID-19 Genomics UK Consortium |
| EPI_ISL_442069 | Department of Pathology, University of Cambridge | Wellcome Sanger Institute for the COVID-19 Genomics UK Consortium |
| EPI_ISL_442070 | Department of Pathology, University of Cambridge | Wellcome Sanger Institute for the COVID-19 Genomics UK Consortium |
| EPI_ISL_442071 | Department of Pathology, University of Cambridge | Wellcome Sanger Institute for the COVID-19 Genomics UK Consortium |
| EPI_ISL_442072 | Department of Pathology, University of Cambridge | Wellcome Sanger Institute for the COVID-19 Genomics UK Consortium |

| EPI_Accession_ID | Originating Laboratory | Submitting Laboratory |
|---|---|---|
| EPI_ISL_442073 | Department of Pathology, University of Cambridge | Wellcome Sanger Institute for the COVID-19 Genomics UK Consortium |
| EPI_ISL_442074 | Department of Pathology, University of Cambridge | Wellcome Sanger Institute for the COVID-19 Genomics UK Consortium |
| EPI_ISL_442075 | Department of Pathology, University of Cambridge | Wellcome Sanger Institute for the COVID-19 Genomics UK Consortium |
| EPI_ISL_442076 | Department of Pathology, University of Cambridge | Wellcome Sanger Institute for the COVID-19 Genomics UK Consortium |
| EPI_ISL_442077 | Department of Pathology, University of Cambridge | Wellcome Sanger Institute for the COVID-19 Genomics UK Consortium |
| EPI_ISL_442078 | Department of Pathology, University of Cambridge | Wellcome Sanger Institute for the COVID-19 Genomics UK Consortium |
| EPI_ISL_442079 | Department of Pathology, University of Cambridge | Wellcome Sanger Institute for the COVID-19 Genomics UK Consortium |
| EPI_ISL_442080 | Department of Pathology, University of Cambridge | Wellcome Sanger Institute for the COVID-19 Genomics UK Consortium |
| EPI_ISL_442081 | Department of Pathology, University of Cambridge | Wellcome Sanger Institute for the COVID-19 Genomics UK Consortium |
| EPI_ISL_442082 | Department of Pathology, University of Cambridge | Wellcome Sanger Institute for the COVID-19 Genomics UK Consortium |
| EPI_ISL_442083 | Department of Pathology, University of Cambridge | Wellcome Sanger Institute for the COVID-19 Genomics UK Consortium |
| EPI_ISL_442084 | Department of Pathology, University of Cambridge | Wellcome Sanger Institute for the COVID-19 Genomics UK Consortium |
| EPI_ISL_442085 | Department of Pathology, University of Cambridge | Wellcome Sanger Institute for the COVID-19 Genomics UK Consortium |
| EPI_ISL_442086 | Department of Pathology, University of Cambridge | Wellcome Sanger Institute for the COVID-19 Genomics UK Consortium |
| EPI_ISL_442087 | Department of Pathology, University of Cambridge | Wellcome Sanger Institute for the COVID-19 Genomics UK Consortium |
| EPI_ISL_442088 | Department of Pathology, University of Cambridge | Wellcome Sanger Institute for the COVID-19 Genomics UK Consortium |

| EPI_Accession_ID | Originating Laboratory | Submitting Laboratory |
| --- | --- | --- |
| EPI_ISL_442089 | Department of Pathology, University of Cambridge | Wellcome Sanger Institute for the COVID-19 Genomics UK Consortium |
| EPI_ISL_442090 | Department of Pathology, University of Cambridge | Wellcome Sanger Institute for the COVID-19 Genomics UK Consortium |
| EPI_ISL_442091 | Department of Pathology, University of Cambridge | Wellcome Sanger Institute for the COVID-19 Genomics UK Consortium |
| EPI_ISL_442092 | Department of Pathology, University of Cambridge | Wellcome Sanger Institute for the COVID-19 Genomics UK Consortium |
| EPI_ISL_442093 | Department of Pathology, University of Cambridge | Wellcome Sanger Institute for the COVID-19 Genomics UK Consortium |
| EPI_ISL_442094 | Department of Pathology, University of Cambridge | Wellcome Sanger Institute for the COVID-19 Genomics UK Consortium |
| EPI_ISL_442095 | Department of Pathology, University of Cambridge | Wellcome Sanger Institute for the COVID-19 Genomics UK Consortium |
| EPI_ISL_442096 | Department of Pathology, University of Cambridge | Wellcome Sanger Institute for the COVID-19 Genomics UK Consortium |
| EPI_ISL_442097 | Department of Pathology, University of Cambridge | Wellcome Sanger Institute for the COVID-19 Genomics UK Consortium |
| EPI_ISL_442098 | Department of Pathology, University of Cambridge | Wellcome Sanger Institute for the COVID-19 Genomics UK Consortium |
| EPI_ISL_442099 | Department of Pathology, University of Cambridge | Wellcome Sanger Institute for the COVID-19 Genomics UK Consortium |
| EPI_ISL_442100 | Department of Pathology, University of Cambridge | Wellcome Sanger Institute for the COVID-19 Genomics UK Consortium |
| EPI_ISL_442101 | Department of Pathology, University of Cambridge | Wellcome Sanger Institute for the COVID-19 Genomics UK Consortium |
| EPI_ISL_442102 | Department of Pathology, University of Cambridge | Wellcome Sanger Institute for the COVID-19 Genomics UK Consortium |
| EPI_ISL_442103 | Department of Pathology, University of Cambridge | Wellcome Sanger Institute for the COVID-19 Genomics UK Consortium |
| EPI_ISL_442104 | Department of Pathology, University of Cambridge | Wellcome Sanger Institute for the COVID-19 Genomics UK Consortium |

| EPI_Accession_ID | Originating Laboratory | Submitting Laboratory |
|---|---|---|
| EPI_ISL_442105 | Department of Pathology, University of Cambridge | Wellcome Sanger Institute for the COVID-19 Genomics UK Consortium |
| EPI_ISL_442106 | Department of Pathology, University of Cambridge | Wellcome Sanger Institute for the COVID-19 Genomics UK Consortium |
| EPI_ISL_442107 | Department of Pathology, University of Cambridge | Wellcome Sanger Institute for the COVID-19 Genomics UK Consortium |
| EPI_ISL_442108 | Department of Pathology, University of Cambridge | Wellcome Sanger Institute for the COVID-19 Genomics UK Consortium |
| EPI_ISL_442109 | Department of Pathology, University of Cambridge | Wellcome Sanger Institute for the COVID-19 Genomics UK Consortium |
| EPI_ISL_442110 | Department of Pathology, University of Cambridge | Wellcome Sanger Institute for the COVID-19 Genomics UK Consortium |
| EPI_ISL_442111 | Department of Pathology, University of Cambridge | Wellcome Sanger Institute for the COVID-19 Genomics UK Consortium |
| EPI_ISL_442112 | Department of Pathology, University of Cambridge | Wellcome Sanger Institute for the COVID-19 Genomics UK Consortium |
| EPI_ISL_442113 | Department of Pathology, University of Cambridge | Wellcome Sanger Institute for the COVID-19 Genomics UK Consortium |
| EPI_ISL_442114 | Department of Pathology, University of Cambridge | Wellcome Sanger Institute for the COVID-19 Genomics UK Consortium |
| EPI_ISL_442115 | Department of Pathology, University of Cambridge | Wellcome Sanger Institute for the COVID-19 Genomics UK Consortium |
| EPI_ISL_442116 | Department of Pathology, University of Cambridge | Wellcome Sanger Institute for the COVID-19 Genomics UK Consortium |
| EPI_ISL_442117 | Department of Pathology, University of Cambridge | Wellcome Sanger Institute for the COVID-19 Genomics UK Consortium |
| EPI_ISL_442118 | Department of Pathology, University of Cambridge | Wellcome Sanger Institute for the COVID-19 Genomics UK Consortium |
| EPI_ISL_442119 | Department of Pathology, University of Cambridge | Wellcome Sanger Institute for the COVID-19 Genomics UK Consortium |
| EPI_ISL_442120 | Department of Pathology, University of Cambridge | Wellcome Sanger Institute for the COVID-19 Genomics UK Consortium |

| EPI_Accession_ID | Originating Laboratory | Submitting Laboratory |
| --- | --- | --- |
| EPI_ISL_442121 | Department of Pathology, University of Cambridge | Wellcome Sanger Institute for the COVID-19 Genomics UK Consortium |
| EPI_ISL_442122 | Department of Pathology, University of Cambridge | Wellcome Sanger Institute for the COVID-19 Genomics UK Consortium |
| EPI_ISL_442123 | Department of Pathology, University of Cambridge | Wellcome Sanger Institute for the COVID-19 Genomics UK Consortium |
| EPI_ISL_442124 | Department of Pathology, University of Cambridge | Wellcome Sanger Institute for the COVID-19 Genomics UK Consortium |
| EPI_ISL_442125 | Department of Pathology, University of Cambridge | Wellcome Sanger Institute for the COVID-19 Genomics UK Consortium |
| EPI_ISL_442126 | Department of Pathology, University of Cambridge | Wellcome Sanger Institute for the COVID-19 Genomics UK Consortium |
| EPI_ISL_442127 | Department of Pathology, University of Cambridge | Wellcome Sanger Institute for the COVID-19 Genomics UK Consortium |
| EPI_ISL_442128 | Department of Pathology, University of Cambridge | Wellcome Sanger Institute for the COVID-19 Genomics UK Consortium |
| EPI_ISL_442129 | Department of Pathology, University of Cambridge | Wellcome Sanger Institute for the COVID-19 Genomics UK Consortium |
| EPI_ISL_442130 | Department of Pathology, University of Cambridge | Wellcome Sanger Institute for the COVID-19 Genomics UK Consortium |
| EPI_ISL_442131 | Department of Pathology, University of Cambridge | Wellcome Sanger Institute for the COVID-19 Genomics UK Consortium |
| EPI_ISL_442132 | Department of Pathology, University of Cambridge | Wellcome Sanger Institute for the COVID-19 Genomics UK Consortium |
| EPI_ISL_442133 | Department of Pathology, University of Cambridge | Wellcome Sanger Institute for the COVID-19 Genomics UK Consortium |
| EPI_ISL_442134 | Department of Pathology, University of Cambridge | Wellcome Sanger Institute for the COVID-19 Genomics UK Consortium |
| EPI_ISL_442135 | Department of Pathology, University of Cambridge | Wellcome Sanger Institute for the COVID-19 Genomics UK Consortium |
| EPI_ISL_442136 | Department of Pathology, University of Cambridge | Wellcome Sanger Institute for the COVID-19 Genomics UK Consortium |

| EPI_Accession_ID | Originating Laboratory | Submitting Laboratory |
| --- | --- | --- |
| EPI_ISL_442137 | Department of Pathology, University of Cambridge | Wellcome Sanger Institute for the COVID-19 Genomics UK Consortium |
| EPI_ISL_442138 | Department of Pathology, University of Cambridge | Wellcome Sanger Institute for the COVID-19 Genomics UK Consortium |
| EPI_ISL_442139 | Department of Pathology, University of Cambridge | Wellcome Sanger Institute for the COVID-19 Genomics UK Consortium |
| EPI_ISL_442140 | Department of Pathology, University of Cambridge | Wellcome Sanger Institute for the COVID-19 Genomics UK Consortium |
| EPI_ISL_442141 | Department of Pathology, University of Cambridge | Wellcome Sanger Institute for the COVID-19 Genomics UK Consortium |
| EPI_ISL_442142 | Department of Pathology, University of Cambridge | Wellcome Sanger Institute for the COVID-19 Genomics UK Consortium |
| EPI_ISL_442143 | Department of Pathology, University of Cambridge | Wellcome Sanger Institute for the COVID-19 Genomics UK Consortium |
| EPI_ISL_442144 | Department of Pathology, University of Cambridge | Wellcome Sanger Institute for the COVID-19 Genomics UK Consortium |
| EPI_ISL_442145 | Department of Pathology, University of Cambridge | Wellcome Sanger Institute for the COVID-19 Genomics UK Consortium |
| EPI_ISL_442146 | Department of Pathology, University of Cambridge | Wellcome Sanger Institute for the COVID-19 Genomics UK Consortium |
| EPI_ISL_442147 | Department of Pathology, University of Cambridge | Wellcome Sanger Institute for the COVID-19 Genomics UK Consortium |
| EPI_ISL_442148 | Department of Pathology, University of Cambridge | Wellcome Sanger Institute for the COVID-19 Genomics UK Consortium |
| EPI_ISL_442149 | Department of Pathology, University of Cambridge | Wellcome Sanger Institute for the COVID-19 Genomics UK Consortium |
| EPI_ISL_442150 | Department of Pathology, University of Cambridge | Wellcome Sanger Institute for the COVID-19 Genomics UK Consortium |
| EPI_ISL_442151 | Department of Pathology, University of Cambridge | Wellcome Sanger Institute for the COVID-19 Genomics UK Consortium |
| EPI_ISL_442152 | Department of Pathology, University of Cambridge | Wellcome Sanger Institute for the COVID-19 Genomics UK Consortium |

| EPI_Accession_ID | Originating Laboratory | Submitting Laboratory |
|---|---|---|
| EPI_ISL_442153 | Department of Pathology, University of Cambridge | Wellcome Sanger Institute for the COVID-19 Genomics UK Consortium |
| EPI_ISL_442154 | Department of Pathology, University of Cambridge | Wellcome Sanger Institute for the COVID-19 Genomics UK Consortium |
| EPI_ISL_442155 | Department of Pathology, University of Cambridge | Wellcome Sanger Institute for the COVID-19 Genomics UK Consortium |
| EPI_ISL_442156 | Department of Pathology, University of Cambridge | Wellcome Sanger Institute for the COVID-19 Genomics UK Consortium |
| EPI_ISL_442157 | Department of Pathology, University of Cambridge | Wellcome Sanger Institute for the COVID-19 Genomics UK Consortium |
| EPI_ISL_442158 | Department of Pathology, University of Cambridge | Wellcome Sanger Institute for the COVID-19 Genomics UK Consortium |
| EPI_ISL_442159 | Department of Pathology, University of Cambridge | Wellcome Sanger Institute for the COVID-19 Genomics UK Consortium |
| EPI_ISL_442160 | Department of Pathology, University of Cambridge | Wellcome Sanger Institute for the COVID-19 Genomics UK Consortium |
| EPI_ISL_442161 | Department of Pathology, University of Cambridge | Wellcome Sanger Institute for the COVID-19 Genomics UK Consortium |
| EPI_ISL_442162 | Department of Pathology, University of Cambridge | Wellcome Sanger Institute for the COVID-19 Genomics UK Consortium |
| EPI_ISL_442163 | Department of Pathology, University of Cambridge | Wellcome Sanger Institute for the COVID-19 Genomics UK Consortium |
| EPI_ISL_442164 | Department of Pathology, University of Cambridge | Wellcome Sanger Institute for the COVID-19 Genomics UK Consortium |
| EPI_ISL_442165 | Department of Pathology, University of Cambridge | Wellcome Sanger Institute for the COVID-19 Genomics UK Consortium |
| EPI_ISL_442166 | Department of Pathology, University of Cambridge | Wellcome Sanger Institute for the COVID-19 Genomics UK Consortium |
| EPI_ISL_442167 | Department of Pathology, University of Cambridge | Wellcome Sanger Institute for the COVID-19 Genomics UK Consortium |
| EPI_ISL_442168 | Department of Pathology, University of Cambridge | Wellcome Sanger Institute for the COVID-19 Genomics UK Consortium |

| EPI_Accession_ID | Originating Laboratory | Submitting Laboratory |
|---|---|---|
| EPI_ISL_442169 | Department of Pathology, University of Cambridge | Wellcome Sanger Institute for the COVID-19 Genomics UK Consortium |
| EPI_ISL_442170 | Department of Pathology, University of Cambridge | Wellcome Sanger Institute for the COVID-19 Genomics UK Consortium |
| EPI_ISL_442171 | Department of Pathology, University of Cambridge | Wellcome Sanger Institute for the COVID-19 Genomics UK Consortium |
| EPI_ISL_442172 | Department of Pathology, University of Cambridge | Wellcome Sanger Institute for the COVID-19 Genomics UK Consortium |
| EPI_ISL_442173 | Department of Pathology, University of Cambridge | Wellcome Sanger Institute for the COVID-19 Genomics UK Consortium |
| EPI_ISL_442174 | Department of Pathology, University of Cambridge | Wellcome Sanger Institute for the COVID-19 Genomics UK Consortium |
| EPI_ISL_442175 | Department of Pathology, University of Cambridge | Wellcome Sanger Institute for the COVID-19 Genomics UK Consortium |
| EPI_ISL_442176 | Department of Pathology, University of Cambridge | Wellcome Sanger Institute for the COVID-19 Genomics UK Consortium |
| EPI_ISL_442177 | Department of Pathology, University of Cambridge | Wellcome Sanger Institute for the COVID-19 Genomics UK Consortium |
| EPI_ISL_442178 | Department of Pathology, University of Cambridge | Wellcome Sanger Institute for the COVID-19 Genomics UK Consortium |
| EPI_ISL_442179 | Department of Pathology, University of Cambridge | Wellcome Sanger Institute for the COVID-19 Genomics UK Consortium |
| EPI_ISL_442180 | Department of Pathology, University of Cambridge | Wellcome Sanger Institute for the COVID-19 Genomics UK Consortium |
| EPI_ISL_442181 | Department of Pathology, University of Cambridge | Wellcome Sanger Institute for the COVID-19 Genomics UK Consortium |
| EPI_ISL_442182 | Department of Pathology, University of Cambridge | Wellcome Sanger Institute for the COVID-19 Genomics UK Consortium |
| EPI_ISL_442183 | Department of Pathology, University of Cambridge | Wellcome Sanger Institute for the COVID-19 Genomics UK Consortium |
| EPI_ISL_442184 | Department of Pathology, University of Cambridge | Wellcome Sanger Institute for the COVID-19 Genomics UK Consortium |

| EPI_Accession_ID | Originating Laboratory | Submitting Laboratory |
|---|---|---|
| EPI_ISL_442185 | Department of Pathology, University of Cambridge | Wellcome Sanger Institute for the COVID-19 Genomics UK Consortium |
| EPI_ISL_442186 | Department of Pathology, University of Cambridge | Wellcome Sanger Institute for the COVID-19 Genomics UK Consortium |
| EPI_ISL_442187 | Department of Pathology, University of Cambridge | Wellcome Sanger Institute for the COVID-19 Genomics UK Consortium |
| EPI_ISL_442188 | Department of Pathology, University of Cambridge | Wellcome Sanger Institute for the COVID-19 Genomics UK Consortium |
| EPI_ISL_442189 | Department of Pathology, University of Cambridge | Wellcome Sanger Institute for the COVID-19 Genomics UK Consortium |
| EPI_ISL_442190 | Department of Pathology, University of Cambridge | Wellcome Sanger Institute for the COVID-19 Genomics UK Consortium |
| EPI_ISL_442191 | Department of Pathology, University of Cambridge | Wellcome Sanger Institute for the COVID-19 Genomics UK Consortium |
| EPI_ISL_442192 | Department of Pathology, University of Cambridge | Wellcome Sanger Institute for the COVID-19 Genomics UK Consortium |
| EPI_ISL_442193 | Department of Pathology, University of Cambridge | Wellcome Sanger Institute for the COVID-19 Genomics UK Consortium |
| EPI_ISL_442194 | Department of Pathology, University of Cambridge | Wellcome Sanger Institute for the COVID-19 Genomics UK Consortium |
| EPI_ISL_442195 | Department of Pathology, University of Cambridge | Wellcome Sanger Institute for the COVID-19 Genomics UK Consortium |
| EPI_ISL_442196 | Department of Pathology, University of Cambridge | Wellcome Sanger Institute for the COVID-19 Genomics UK Consortium |
| EPI_ISL_442197 | Department of Pathology, University of Cambridge | Wellcome Sanger Institute for the COVID-19 Genomics UK Consortium |
| EPI_ISL_442198 | Department of Pathology, University of Cambridge | Wellcome Sanger Institute for the COVID-19 Genomics UK Consortium |
| EPI_ISL_442199 | Department of Pathology, University of Cambridge | Wellcome Sanger Institute for the COVID-19 Genomics UK Consortium |
| EPI_ISL_442200 | Department of Pathology, University of Cambridge | Wellcome Sanger Institute for the COVID-19 Genomics UK Consortium |

| EPI_Accession_ID | Originating Laboratory | Submitting Laboratory |
|---|---|---|
| EPI_ISL_442201 | Department of Pathology, University of Cambridge | Wellcome Sanger Institute for the COVID-19 Genomics UK Consortium |
| EPI_ISL_442202 | Department of Pathology, University of Cambridge | Wellcome Sanger Institute for the COVID-19 Genomics UK Consortium |
| EPI_ISL_442203 | Department of Pathology, University of Cambridge | Wellcome Sanger Institute for the COVID-19 Genomics UK Consortium |
| EPI_ISL_442204 | Department of Pathology, University of Cambridge | Wellcome Sanger Institute for the COVID-19 Genomics UK Consortium |
| EPI_ISL_442205 | Department of Pathology, University of Cambridge | Wellcome Sanger Institute for the COVID-19 Genomics UK Consortium |
| EPI_ISL_442206 | Department of Pathology, University of Cambridge | Wellcome Sanger Institute for the COVID-19 Genomics UK Consortium |
| EPI_ISL_442207 | Department of Pathology, University of Cambridge | Wellcome Sanger Institute for the COVID-19 Genomics UK Consortium |
| EPI_ISL_442208 | Department of Pathology, University of Cambridge | Wellcome Sanger Institute for the COVID-19 Genomics UK Consortium |
| EPI_ISL_442209 | Department of Pathology, University of Cambridge | Wellcome Sanger Institute for the COVID-19 Genomics UK Consortium |
| EPI_ISL_442210 | Department of Pathology, University of Cambridge | Wellcome Sanger Institute for the COVID-19 Genomics UK Consortium |
| EPI_ISL_442211 | Department of Pathology, University of Cambridge | Wellcome Sanger Institute for the COVID-19 Genomics UK Consortium |
| EPI_ISL_442212 | Department of Pathology, University of Cambridge | Wellcome Sanger Institute for the COVID-19 Genomics UK Consortium |
| EPI_ISL_442213 | Department of Pathology, University of Cambridge | Wellcome Sanger Institute for the COVID-19 Genomics UK Consortium |
| EPI_ISL_442214 | Department of Pathology, University of Cambridge | Wellcome Sanger Institute for the COVID-19 Genomics UK Consortium |
| EPI_ISL_442215 | Department of Pathology, University of Cambridge | Wellcome Sanger Institute for the COVID-19 Genomics UK Consortium |
| EPI_ISL_442216 | Department of Pathology, University of Cambridge | Wellcome Sanger Institute for the COVID-19 Genomics UK Consortium |

| EPI_Accession_ID | Originating Laboratory | Submitting Laboratory |
|---|---|---|
| EPI_ISL_442217 | Department of Pathology, University of Cambridge | Wellcome Sanger Institute for the COVID-19 Genomics UK Consortium |
| EPI_ISL_442218 | Department of Pathology, University of Cambridge | Wellcome Sanger Institute for the COVID-19 Genomics UK Consortium |
| EPI_ISL_442219 | Department of Pathology, University of Cambridge | Wellcome Sanger Institute for the COVID-19 Genomics UK Consortium |
| EPI_ISL_442220 | Department of Pathology, University of Cambridge | Wellcome Sanger Institute for the COVID-19 Genomics UK Consortium |
| EPI_ISL_442221 | Department of Pathology, University of Cambridge | Wellcome Sanger Institute for the COVID-19 Genomics UK Consortium |
| EPI_ISL_442222 | Department of Pathology, University of Cambridge | Wellcome Sanger Institute for the COVID-19 Genomics UK Consortium |
| EPI_ISL_442223 | Department of Pathology, University of Cambridge | Wellcome Sanger Institute for the COVID-19 Genomics UK Consortium |
| EPI_ISL_442224 | Department of Pathology, University of Cambridge | Wellcome Sanger Institute for the COVID-19 Genomics UK Consortium |
| EPI_ISL_442225 | Department of Pathology, University of Cambridge | Wellcome Sanger Institute for the COVID-19 Genomics UK Consortium |
| EPI_ISL_442226 | Department of Pathology, University of Cambridge | Wellcome Sanger Institute for the COVID-19 Genomics UK Consortium |
| EPI_ISL_442227 | Department of Pathology, University of Cambridge | Wellcome Sanger Institute for the COVID-19 Genomics UK Consortium |
| EPI_ISL_442228 | Department of Pathology, University of Cambridge | Wellcome Sanger Institute for the COVID-19 Genomics UK Consortium |
| EPI_ISL_442229 | Department of Pathology, University of Cambridge | Wellcome Sanger Institute for the COVID-19 Genomics UK Consortium |
| EPI_ISL_442230 | Department of Pathology, University of Cambridge | Wellcome Sanger Institute for the COVID-19 Genomics UK Consortium |
| EPI_ISL_442231 | Department of Pathology, University of Cambridge | Wellcome Sanger Institute for the COVID-19 Genomics UK Consortium |
| EPI_ISL_442232 | Department of Pathology, University of Cambridge | Wellcome Sanger Institute for the COVID-19 Genomics UK Consortium |

| EPI_Accession_ID | Originating Laboratory | Submitting Laboratory |
|---|---|---|
| EPI_ISL_442233 | Department of Pathology, University of Cambridge | Wellcome Sanger Institute for the COVID-19 Genomics UK Consortium |
| EPI_ISL_442234 | Department of Pathology, University of Cambridge | Wellcome Sanger Institute for the COVID-19 Genomics UK Consortium |
| EPI_ISL_442235 | Department of Pathology, University of Cambridge | Wellcome Sanger Institute for the COVID-19 Genomics UK Consortium |
| EPI_ISL_442236 | Department of Pathology, University of Cambridge | Wellcome Sanger Institute for the COVID-19 Genomics UK Consortium |
| EPI_ISL_442237 | Department of Pathology, University of Cambridge | Wellcome Sanger Institute for the COVID-19 Genomics UK Consortium |
| EPI_ISL_442238 | Department of Pathology, University of Cambridge | Wellcome Sanger Institute for the COVID-19 Genomics UK Consortium |
| EPI_ISL_442239 | Department of Pathology, University of Cambridge | Wellcome Sanger Institute for the COVID-19 Genomics UK Consortium |
| EPI_ISL_442240 | Department of Pathology, University of Cambridge | Wellcome Sanger Institute for the COVID-19 Genomics UK Consortium |
| EPI_ISL_442241 | Department of Pathology, University of Cambridge | Wellcome Sanger Institute for the COVID-19 Genomics UK Consortium |
| EPI_ISL_442242 | Department of Pathology, University of Cambridge | Wellcome Sanger Institute for the COVID-19 Genomics UK Consortium |
| EPI_ISL_442243 | Department of Pathology, University of Cambridge | Wellcome Sanger Institute for the COVID-19 Genomics UK Consortium |
| EPI_ISL_442244 | Department of Pathology, University of Cambridge | Wellcome Sanger Institute for the COVID-19 Genomics UK Consortium |
| EPI_ISL_442245 | Department of Pathology, University of Cambridge | Wellcome Sanger Institute for the COVID-19 Genomics UK Consortium |
| EPI_ISL_442246 | Department of Pathology, University of Cambridge | Wellcome Sanger Institute for the COVID-19 Genomics UK Consortium |
| EPI_ISL_442247 | Department of Pathology, University of Cambridge | Wellcome Sanger Institute for the COVID-19 Genomics UK Consortium |
| EPI_ISL_442248 | Department of Pathology, University of Cambridge | Wellcome Sanger Institute for the COVID-19 Genomics UK Consortium |

| EPI_Accession_ID | Originating Laboratory | Submitting Laboratory |
|---|---|---|
| EPI_ISL_442249 | Department of Pathology, University of Cambridge | Wellcome Sanger Institute for the COVID-19 Genomics UK Consortium |
| EPI_ISL_442250 | Department of Pathology, University of Cambridge | Wellcome Sanger Institute for the COVID-19 Genomics UK Consortium |
| EPI_ISL_442251 | Department of Pathology, University of Cambridge | Wellcome Sanger Institute for the COVID-19 Genomics UK Consortium |
| EPI_ISL_442252 | Department of Pathology, University of Cambridge | Wellcome Sanger Institute for the COVID-19 Genomics UK Consortium |
| EPI_ISL_442253 | Department of Pathology, University of Cambridge | Wellcome Sanger Institute for the COVID-19 Genomics UK Consortium |
| EPI_ISL_442254 | Department of Pathology, University of Cambridge | Wellcome Sanger Institute for the COVID-19 Genomics UK Consortium |
| EPI_ISL_442255 | Department of Pathology, University of Cambridge | Wellcome Sanger Institute for the COVID-19 Genomics UK Consortium |
| EPI_ISL_442256 | Department of Pathology, University of Cambridge | Wellcome Sanger Institute for the COVID-19 Genomics UK Consortium |
| EPI_ISL_442257 | Department of Pathology, University of Cambridge | Wellcome Sanger Institute for the COVID-19 Genomics UK Consortium |
| EPI_ISL_442258 | Department of Pathology, University of Cambridge | Wellcome Sanger Institute for the COVID-19 Genomics UK Consortium |
| EPI_ISL_442259 | Department of Pathology, University of Cambridge | Wellcome Sanger Institute for the COVID-19 Genomics UK Consortium |
| EPI_ISL_442260 | Department of Pathology, University of Cambridge | Wellcome Sanger Institute for the COVID-19 Genomics UK Consortium |
| EPI_ISL_442261 | Department of Pathology, University of Cambridge | Wellcome Sanger Institute for the COVID-19 Genomics UK Consortium |
| EPI_ISL_442262 | Department of Pathology, University of Cambridge | Wellcome Sanger Institute for the COVID-19 Genomics UK Consortium |
| EPI_ISL_442263 | Department of Pathology, University of Cambridge | Wellcome Sanger Institute for the COVID-19 Genomics UK Consortium |
| EPI_ISL_442264 | Department of Pathology, University of Cambridge | Wellcome Sanger Institute for the COVID-19 Genomics UK Consortium |

| EPI_Accession_ID | Originating Laboratory | Submitting Laboratory |
|---|---|---|
| EPI_ISL_442265 | Department of Pathology, University of Cambridge | Wellcome Sanger Institute for the COVID-19 Genomics UK Consortium |
| EPI_ISL_442266 | Department of Pathology, University of Cambridge | Wellcome Sanger Institute for the COVID-19 Genomics UK Consortium |
| EPI_ISL_442267 | Department of Pathology, University of Cambridge | Wellcome Sanger Institute for the COVID-19 Genomics UK Consortium |
| EPI_ISL_442268 | Department of Pathology, University of Cambridge | Wellcome Sanger Institute for the COVID-19 Genomics UK Consortium |
| EPI_ISL_442269 | Department of Pathology, University of Cambridge | Wellcome Sanger Institute for the COVID-19 Genomics UK Consortium |
| EPI_ISL_442270 | Department of Pathology, University of Cambridge | Wellcome Sanger Institute for the COVID-19 Genomics UK Consortium |
| EPI_ISL_442271 | Department of Pathology, University of Cambridge | Wellcome Sanger Institute for the COVID-19 Genomics UK Consortium |
| EPI_ISL_442272 | Department of Pathology, University of Cambridge | Wellcome Sanger Institute for the COVID-19 Genomics UK Consortium |
| EPI_ISL_442273 | Department of Pathology, University of Cambridge | Wellcome Sanger Institute for the COVID-19 Genomics UK Consortium |
| EPI_ISL_442274 | Department of Pathology, University of Cambridge | Wellcome Sanger Institute for the COVID-19 Genomics UK Consortium |
| EPI_ISL_442275 | Department of Pathology, University of Cambridge | Wellcome Sanger Institute for the COVID-19 Genomics UK Consortium |
| EPI_ISL_442276 | Department of Pathology, University of Cambridge | Wellcome Sanger Institute for the COVID-19 Genomics UK Consortium |
| EPI_ISL_442277 | Department of Pathology, University of Cambridge | Wellcome Sanger Institute for the COVID-19 Genomics UK Consortium |
| EPI_ISL_442278 | Department of Pathology, University of Cambridge | Wellcome Sanger Institute for the COVID-19 Genomics UK Consortium |
| EPI_ISL_442279 | Department of Pathology, University of Cambridge | Wellcome Sanger Institute for the COVID-19 Genomics UK Consortium |
| EPI_ISL_442280 | Department of Pathology, University of Cambridge | Wellcome Sanger Institute for the COVID-19 Genomics UK Consortium |

| EPI_Accession_ID | Originating Laboratory | Submitting Laboratory |
|---|---|---|
| EPI_ISL_442281 | Department of Pathology, University of Cambridge | Wellcome Sanger Institute for the COVID-19 Genomics UK Consortium |
| EPI_ISL_442282 | Department of Pathology, University of Cambridge | Wellcome Sanger Institute for the COVID-19 Genomics UK Consortium |
| EPI_ISL_442283 | Department of Pathology, University of Cambridge | Wellcome Sanger Institute for the COVID-19 Genomics UK Consortium |
| EPI_ISL_442284 | Department of Pathology, University of Cambridge | Wellcome Sanger Institute for the COVID-19 Genomics UK Consortium |
| EPI_ISL_442285 | Department of Pathology, University of Cambridge | Wellcome Sanger Institute for the COVID-19 Genomics UK Consortium |
| EPI_ISL_442286 | Department of Pathology, University of Cambridge | Wellcome Sanger Institute for the COVID-19 Genomics UK Consortium |
| EPI_ISL_442287 | Department of Pathology, University of Cambridge | Wellcome Sanger Institute for the COVID-19 Genomics UK Consortium |
| EPI_ISL_442288 | Department of Pathology, University of Cambridge | Wellcome Sanger Institute for the COVID-19 Genomics UK Consortium |
| EPI_ISL_442289 | Department of Pathology, University of Cambridge | Wellcome Sanger Institute for the COVID-19 Genomics UK Consortium |
| EPI_ISL_442290 | Department of Pathology, University of Cambridge | Wellcome Sanger Institute for the COVID-19 Genomics UK Consortium |
| EPI_ISL_442291 | Department of Pathology, University of Cambridge | Wellcome Sanger Institute for the COVID-19 Genomics UK Consortium |
| EPI_ISL_442292 | Department of Pathology, University of Cambridge | Wellcome Sanger Institute for the COVID-19 Genomics UK Consortium |
| EPI_ISL_442293 | Department of Pathology, University of Cambridge | Wellcome Sanger Institute for the COVID-19 Genomics UK Consortium |
| EPI_ISL_442294 | Department of Pathology, University of Cambridge | Wellcome Sanger Institute for the COVID-19 Genomics UK Consortium |
| EPI_ISL_442295 | Department of Pathology, University of Cambridge | Wellcome Sanger Institute for the COVID-19 Genomics UK Consortium |
| EPI_ISL_442296 | Department of Pathology, University of Cambridge | Wellcome Sanger Institute for the COVID-19 Genomics UK Consortium |

| EPI_Accession_ID | Originating Laboratory | Submitting Laboratory |
|---|---|---|
| EPI_ISL_442297 | Department of Pathology, University of Cambridge | Wellcome Sanger Institute for the COVID-19 Genomics UK Consortium |
| EPI_ISL_442298 | Department of Pathology, University of Cambridge | Wellcome Sanger Institute for the COVID-19 Genomics UK Consortium |
| EPI_ISL_442299 | Department of Pathology, University of Cambridge | Wellcome Sanger Institute for the COVID-19 Genomics UK Consortium |
| EPI_ISL_442300 | Department of Pathology, University of Cambridge | Wellcome Sanger Institute for the COVID-19 Genomics UK Consortium |
| EPI_ISL_442301 | Department of Pathology, University of Cambridge | Wellcome Sanger Institute for the COVID-19 Genomics UK Consortium |
| EPI_ISL_442302 | Department of Pathology, University of Cambridge | Wellcome Sanger Institute for the COVID-19 Genomics UK Consortium |
| EPI_ISL_442303 | Department of Pathology, University of Cambridge | Wellcome Sanger Institute for the COVID-19 Genomics UK Consortium |
| EPI_ISL_442304 | Department of Pathology, University of Cambridge | Wellcome Sanger Institute for the COVID-19 Genomics UK Consortium |
| EPI_ISL_442305 | Department of Pathology, University of Cambridge | Wellcome Sanger Institute for the COVID-19 Genomics UK Consortium |
| EPI_ISL_442306 | Department of Pathology, University of Cambridge | Wellcome Sanger Institute for the COVID-19 Genomics UK Consortium |
| EPI_ISL_442307 | Department of Pathology, University of Cambridge | Wellcome Sanger Institute for the COVID-19 Genomics UK Consortium |
| EPI_ISL_442308 | Department of Pathology, University of Cambridge | Wellcome Sanger Institute for the COVID-19 Genomics UK Consortium |
| EPI_ISL_442309 | Department of Pathology, University of Cambridge | Wellcome Sanger Institute for the COVID-19 Genomics UK Consortium |
| EPI_ISL_442310 | Department of Pathology, University of Cambridge | Wellcome Sanger Institute for the COVID-19 Genomics UK Consortium |
| EPI_ISL_442311 | Department of Pathology, University of Cambridge | Wellcome Sanger Institute for the COVID-19 Genomics UK Consortium |
| EPI_ISL_442312 | Department of Pathology, University of Cambridge | Wellcome Sanger Institute for the COVID-19 Genomics UK Consortium |

| EPI_Accession_ID | Originating Laboratory | Submitting Laboratory |
|---|---|---|
| EPI_ISL_442313 | Department of Pathology, University of Cambridge | Wellcome Sanger Institute for the COVID-19 Genomics UK Consortium |
| EPI_ISL_442314 | Department of Pathology, University of Cambridge | Wellcome Sanger Institute for the COVID-19 Genomics UK Consortium |
| EPI_ISL_442315 | Department of Pathology, University of Cambridge | Wellcome Sanger Institute for the COVID-19 Genomics UK Consortium |
| EPI_ISL_442316 | Department of Pathology, University of Cambridge | Wellcome Sanger Institute for the COVID-19 Genomics UK Consortium |
| EPI_ISL_442317 | Department of Pathology, University of Cambridge | Wellcome Sanger Institute for the COVID-19 Genomics UK Consortium |
| EPI_ISL_442318 | Department of Pathology, University of Cambridge | Wellcome Sanger Institute for the COVID-19 Genomics UK Consortium |
| EPI_ISL_442319 | Department of Pathology, University of Cambridge | Wellcome Sanger Institute for the COVID-19 Genomics UK Consortium |
| EPI_ISL_442320 | Department of Pathology, University of Cambridge | Wellcome Sanger Institute for the COVID-19 Genomics UK Consortium |
| EPI_ISL_442321 | Department of Pathology, University of Cambridge | Wellcome Sanger Institute for the COVID-19 Genomics UK Consortium |
| EPI_ISL_442322 | Department of Pathology, University of Cambridge | Wellcome Sanger Institute for the COVID-19 Genomics UK Consortium |
| EPI_ISL_442323 | Department of Pathology, University of Cambridge | Wellcome Sanger Institute for the COVID-19 Genomics UK Consortium |
| EPI_ISL_442324 | Department of Pathology, University of Cambridge | Wellcome Sanger Institute for the COVID-19 Genomics UK Consortium |
| EPI_ISL_442325 | Department of Pathology, University of Cambridge | Wellcome Sanger Institute for the COVID-19 Genomics UK Consortium |
| EPI_ISL_442326 | Department of Pathology, University of Cambridge | Wellcome Sanger Institute for the COVID-19 Genomics UK Consortium |
| EPI_ISL_442327 | Department of Pathology, University of Cambridge | Wellcome Sanger Institute for the COVID-19 Genomics UK Consortium |
| EPI_ISL_442328 | Department of Pathology, University of Cambridge | Wellcome Sanger Institute for the COVID-19 Genomics UK Consortium |

| EPI_Accession_ID | Originating Laboratory | Submitting Laboratory |
|---|---|---|
| EPI_ISL_442329 | Department of Pathology, University of Cambridge | Wellcome Sanger Institute for the COVID-19 Genomics UK Consortium |
| EPI_ISL_442330 | Department of Pathology, University of Cambridge | Wellcome Sanger Institute for the COVID-19 Genomics UK Consortium |
| EPI_ISL_442331 | Department of Pathology, University of Cambridge | Wellcome Sanger Institute for the COVID-19 Genomics UK Consortium |
| EPI_ISL_442332 | Department of Pathology, University of Cambridge | Wellcome Sanger Institute for the COVID-19 Genomics UK Consortium |
| EPI_ISL_442333 | Department of Pathology, University of Cambridge | Wellcome Sanger Institute for the COVID-19 Genomics UK Consortium |
| EPI_ISL_442334 | Department of Pathology, University of Cambridge | Wellcome Sanger Institute for the COVID-19 Genomics UK Consortium |
| EPI_ISL_442335 | Department of Pathology, University of Cambridge | Wellcome Sanger Institute for the COVID-19 Genomics UK Consortium |
| EPI_ISL_442336 | Department of Pathology, University of Cambridge | Wellcome Sanger Institute for the COVID-19 Genomics UK Consortium |
| EPI_ISL_442337 | Department of Pathology, University of Cambridge | Wellcome Sanger Institute for the COVID-19 Genomics UK Consortium |
| EPI_ISL_442338 | Department of Pathology, University of Cambridge | Wellcome Sanger Institute for the COVID-19 Genomics UK Consortium |
| EPI_ISL_442339 | Department of Pathology, University of Cambridge | Wellcome Sanger Institute for the COVID-19 Genomics UK Consortium |
| EPI_ISL_442340 | Department of Pathology, University of Cambridge | Wellcome Sanger Institute for the COVID-19 Genomics UK Consortium |
| EPI_ISL_442341 | Department of Pathology, University of Cambridge | Wellcome Sanger Institute for the COVID-19 Genomics UK Consortium |
| EPI_ISL_442342 | Department of Pathology, University of Cambridge | Wellcome Sanger Institute for the COVID-19 Genomics UK Consortium |
| EPI_ISL_442343 | Department of Pathology, University of Cambridge | Wellcome Sanger Institute for the COVID-19 Genomics UK Consortium |

| EPI_Accession_ID | Originating Laboratory | Submitting Laboratory |
|---|---|---|
| EPI_ISL_442344 | Virology Department, Sheffield Teaching Hospitals NHS Foundation Trust/Department of Infection, Immunity and Cardiovascular Disease, The Medical School, University of Sheffield | COVID-19 Genomics UK (COG-UK) Consortium |
| EPI_ISL_442345 | Virology Department, Sheffield Teaching Hospitals NHS Foundation Trust/Department of Infection, Immunity and Cardiovascular Disease, The Medical School, University of Sheffield | COVID-19 Genomics UK (COG-UK) Consortium |
| EPI_ISL_442346 | Virology Department, Sheffield Teaching Hospitals NHS Foundation Trust/Department of Infection, Immunity and Cardiovascular Disease, The Medical School, University of Sheffield | COVID-19 Genomics UK (COG-UK) Consortium |
| EPI_ISL_442347 | Virology Department, Sheffield Teaching Hospitals NHS Foundation Trust/Department of Infection, Immunity and Cardiovascular Disease, The Medical School, University of Sheffield | COVID-19 Genomics UK (COG-UK) Consortium |
| EPI_ISL_442348 | Virology Department, Sheffield Teaching Hospitals NHS Foundation Trust/Department of Infection, Immunity and Cardiovascular Disease, The Medical School, University of Sheffield | COVID-19 Genomics UK (COG-UK) Consortium |
| EPI_ISL_442349 | Virology Department, Sheffield Teaching Hospitals NHS Foundation Trust/Department of Infection, Immunity and Cardiovascular Disease, The Medical School, University of Sheffield | COVID-19 Genomics UK (COG-UK) Consortium |
| EPI_ISL_442350 | Virology Department, Sheffield Teaching Hospitals NHS Foundation Trust/Department of Infection, Immunity and Cardiovascular Disease, The Medical School, University of Sheffield | COVID-19 Genomics UK (COG-UK) Consortium |
| EPI_ISL_442351 | Virology Department, Sheffield Teaching Hospitals NHS Foundation Trust/Department of Infection, Immunity and Cardiovascular Disease, The Medical School, University of Sheffield | COVID-19 Genomics UK (COG-UK) Consortium |

| EPI_Accession_ID | Originating Laboratory | Submitting Laboratory |
|---|---|---|
| EPI_ISL_442352 | Virology Department, Sheffield Teaching Hospitals NHS Foundation Trust/Department of Infection, Immunity and Cardiovascular Disease, The Medical School, University of Sheffield | COVID-19 Genomics UK (COG-UK) Consortium |
| EPI_ISL_442353 | Virology Department, Sheffield Teaching Hospitals NHS Foundation Trust/Department of Infection, Immunity and Cardiovascular Disease, The Medical School, University of Sheffield | COVID-19 Genomics UK (COG-UK) Consortium |
| EPI_ISL_442354 | Virology Department, Sheffield Teaching Hospitals NHS Foundation Trust/Department of Infection, Immunity and Cardiovascular Disease, The Medical School, University of Sheffield | COVID-19 Genomics UK (COG-UK) Consortium |
| EPI_ISL_442355 | Virology Department, Sheffield Teaching Hospitals NHS Foundation Trust/Department of Infection, Immunity and Cardiovascular Disease, The Medical School, University of Sheffield | COVID-19 Genomics UK (COG-UK) Consortium |
| EPI_ISL_442356 | Virology Department, Sheffield Teaching Hospitals NHS Foundation Trust/Department of Infection, Immunity and Cardiovascular Disease, The Medical School, University of Sheffield | COVID-19 Genomics UK (COG-UK) Consortium |
| EPI_ISL_442357 | Virology Department, Sheffield Teaching Hospitals NHS Foundation Trust/Department of Infection, Immunity and Cardiovascular Disease, The Medical School, University of Sheffield | COVID-19 Genomics UK (COG-UK) Consortium |
| EPI_ISL_442358 | Virology Department, Sheffield Teaching Hospitals NHS Foundation Trust/Department of Infection, Immunity and Cardiovascular Disease, The Medical School, University of Sheffield | COVID-19 Genomics UK (COG-UK) Consortium |
| EPI_ISL_442359 | Virology Department, Sheffield Teaching Hospitals NHS Foundation Trust/Department of Infection, Immunity and Cardiovascular Disease, The Medical School, University of Sheffield | COVID-19 Genomics UK (COG-UK) Consortium |

| EPI_Accession_ID | Originating Laboratory | Submitting Laboratory |
|---|---|---|
| EPI_ISL_442360 | Virology Department, Sheffield Teaching Hospitals NHS Foundation Trust/Department of Infection, Immunity and Cardiovascular Disease, The Medical School, University of Sheffield | COVID-19 Genomics UK (COG-UK) Consortium |
| EPI_ISL_442361 | Virology Department, Sheffield Teaching Hospitals NHS Foundation Trust/Department of Infection, Immunity and Cardiovascular Disease, The Medical School, University of Sheffield | COVID-19 Genomics UK (COG-UK) Consortium |
| EPI_ISL_442362 | Virology Department, Sheffield Teaching Hospitals NHS Foundation Trust/Department of Infection, Immunity and Cardiovascular Disease, The Medical School, University of Sheffield | COVID-19 Genomics UK (COG-UK) Consortium |
| EPI_ISL_442363 | Virology Department, Sheffield Teaching Hospitals NHS Foundation Trust/Department of Infection, Immunity and Cardiovascular Disease, The Medical School, University of Sheffield | COVID-19 Genomics UK (COG-UK) Consortium |
| EPI_ISL_442364 | Virology Department, Sheffield Teaching Hospitals NHS Foundation Trust/Department of Infection, Immunity and Cardiovascular Disease, The Medical School, University of Sheffield | COVID-19 Genomics UK (COG-UK) Consortium |
| EPI_ISL_442365 | Virology Department, Sheffield Teaching Hospitals NHS Foundation Trust/Department of Infection, Immunity and Cardiovascular Disease, The Medical School, University of Sheffield | COVID-19 Genomics UK (COG-UK) Consortium |
| EPI_ISL_442366 | Virology Department, Sheffield Teaching Hospitals NHS Foundation Trust/Department of Infection, Immunity and Cardiovascular Disease, The Medical School, University of Sheffield | COVID-19 Genomics UK (COG-UK) Consortium |
| EPI_ISL_442367 | Virology Department, Sheffield Teaching Hospitals NHS Foundation Trust/Department of Infection, Immunity and Cardiovascular Disease, The Medical School, University of Sheffield | COVID-19 Genomics UK (COG-UK) Consortium |

| EPI_Accession_ID | Originating Laboratory | Submitting Laboratory |
| --- | --- | --- |
| EPI_ISL_442368 | Virology Department, Sheffield Teaching Hospitals NHS Foundation Trust/Department of Infection, Immunity and Cardiovascular Disease, The Medical School, University of Sheffield | COVID-19 Genomics UK (COG-UK) Consortium |
| EPI_ISL_442369 | Virology Department, Sheffield Teaching Hospitals NHS Foundation Trust/Department of Infection, Immunity and Cardiovascular Disease, The Medical School, University of Sheffield | COVID-19 Genomics UK (COG-UK) Consortium |
| EPI_ISL_442370 | Virology Department, Sheffield Teaching Hospitals NHS Foundation Trust/Department of Infection, Immunity and Cardiovascular Disease, The Medical School, University of Sheffield | COVID-19 Genomics UK (COG-UK) Consortium |
| EPI_ISL_442371 | Virology Department, Sheffield Teaching Hospitals NHS Foundation Trust/Department of Infection, Immunity and Cardiovascular Disease, The Medical School, University of Sheffield | COVID-19 Genomics UK (COG-UK) Consortium |
| EPI_ISL_442372 | Virology Department, Sheffield Teaching Hospitals NHS Foundation Trust/Department of Infection, Immunity and Cardiovascular Disease, The Medical School, University of Sheffield | COVID-19 Genomics UK (COG-UK) Consortium |
| EPI_ISL_442373 | Virology Department, Sheffield Teaching Hospitals NHS Foundation Trust/Department of Infection, Immunity and Cardiovascular Disease, The Medical School, University of Sheffield | COVID-19 Genomics UK (COG-UK) Consortium |
| EPI_ISL_442374 | Virology Department, Sheffield Teaching Hospitals NHS Foundation Trust/Department of Infection, Immunity and Cardiovascular Disease, The Medical School, University of Sheffield | COVID-19 Genomics UK (COG-UK) Consortium |
| EPI_ISL_442375 | Virology Department, Sheffield Teaching Hospitals NHS Foundation Trust/Department of Infection, Immunity and Cardiovascular Disease, The Medical School, University of Sheffield | COVID-19 Genomics UK (COG-UK) Consortium |

| EPI_Accession_ID | Originating Laboratory | Submitting Laboratory |
|---|---|---|
| EPI_ISL_442376 | Virology Department, Sheffield Teaching Hospitals NHS Foundation Trust/Department of Infection, Immunity and Cardiovascular Disease, The Medical School, University of Sheffield | COVID-19 Genomics UK (COG-UK) Consortium |
| EPI_ISL_442377 | Virology Department, Sheffield Teaching Hospitals NHS Foundation Trust/Department of Infection, Immunity and Cardiovascular Disease, The Medical School, University of Sheffield | COVID-19 Genomics UK (COG-UK) Consortium |
| EPI_ISL_442378 | Virology Department, Sheffield Teaching Hospitals NHS Foundation Trust/Department of Infection, Immunity and Cardiovascular Disease, The Medical School, University of Sheffield | COVID-19 Genomics UK (COG-UK) Consortium |
| EPI_ISL_442379 | Virology Department, Sheffield Teaching Hospitals NHS Foundation Trust/Department of Infection, Immunity and Cardiovascular Disease, The Medical School, University of Sheffield | COVID-19 Genomics UK (COG-UK) Consortium |
| EPI_ISL_442380 | Virology Department, Sheffield Teaching Hospitals NHS Foundation Trust/Department of Infection, Immunity and Cardiovascular Disease, The Medical School, University of Sheffield | COVID-19 Genomics UK (COG-UK) Consortium |
| EPI_ISL_442381 | Virology Department, Sheffield Teaching Hospitals NHS Foundation Trust/Department of Infection, Immunity and Cardiovascular Disease, The Medical School, University of Sheffield | COVID-19 Genomics UK (COG-UK) Consortium |
| EPI_ISL_442382 | Virology Department, Sheffield Teaching Hospitals NHS Foundation Trust/Department of Infection, Immunity and Cardiovascular Disease, The Medical School, University of Sheffield | COVID-19 Genomics UK (COG-UK) Consortium |
| EPI_ISL_442383 | Virology Department, Sheffield Teaching Hospitals NHS Foundation Trust/Department of Infection, Immunity and Cardiovascular Disease, The Medical School, University of Sheffield | COVID-19 Genomics UK (COG-UK) Consortium |

| EPI_Accession_ID | Originating Laboratory | Submitting Laboratory |
|---|---|---|
| EPI_ISL_442384 | Virology Department, Sheffield Teaching Hospitals NHS Foundation Trust/Department of Infection, Immunity and Cardiovascular Disease, The Medical School, University of Sheffield | COVID-19 Genomics UK (COG-UK) Consortium |
| EPI_ISL_442385 | Virology Department, Sheffield Teaching Hospitals NHS Foundation Trust/Department of Infection, Immunity and Cardiovascular Disease, The Medical School, University of Sheffield | COVID-19 Genomics UK (COG-UK) Consortium |
| EPI_ISL_442386 | Virology Department, Sheffield Teaching Hospitals NHS Foundation Trust/Department of Infection, Immunity and Cardiovascular Disease, The Medical School, University of Sheffield | COVID-19 Genomics UK (COG-UK) Consortium |
| EPI_ISL_442387 | Virology Department, Sheffield Teaching Hospitals NHS Foundation Trust/Department of Infection, Immunity and Cardiovascular Disease, The Medical School, University of Sheffield | COVID-19 Genomics UK (COG-UK) Consortium |
| EPI_ISL_442388 | Virology Department, Sheffield Teaching Hospitals NHS Foundation Trust/Department of Infection, Immunity and Cardiovascular Disease, The Medical School, University of Sheffield | COVID-19 Genomics UK (COG-UK) Consortium |
| EPI_ISL_442389 | Virology Department, Sheffield Teaching Hospitals NHS Foundation Trust/Department of Infection, Immunity and Cardiovascular Disease, The Medical School, University of Sheffield | COVID-19 Genomics UK (COG-UK) Consortium |
| EPI_ISL_442390 | Virology Department, Sheffield Teaching Hospitals NHS Foundation Trust/Department of Infection, Immunity and Cardiovascular Disease, The Medical School, University of Sheffield | COVID-19 Genomics UK (COG-UK) Consortium |
| EPI_ISL_442391 | Virology Department, Sheffield Teaching Hospitals NHS Foundation Trust/Department of Infection, Immunity and Cardiovascular Disease, The Medical School, University of Sheffield | COVID-19 Genomics UK (COG-UK) Consortium |

| EPI_Accession_ID | Originating Laboratory | Submitting Laboratory |
|---|---|---|
| EPI_ISL_442392 | Virology Department, Sheffield Teaching Hospitals NHS Foundation Trust/Department of Infection, Immunity and Cardiovascular Disease, The Medical School, University of Sheffield | COVID-19 Genomics UK (COG-UK) Consortium |
| EPI_ISL_442393 | Virology Department, Sheffield Teaching Hospitals NHS Foundation Trust/Department of Infection, Immunity and Cardiovascular Disease, The Medical School, University of Sheffield | COVID-19 Genomics UK (COG-UK) Consortium |
| EPI_ISL_442394 | Virology Department, Sheffield Teaching Hospitals NHS Foundation Trust/Department of Infection, Immunity and Cardiovascular Disease, The Medical School, University of Sheffield | COVID-19 Genomics UK (COG-UK) Consortium |
| EPI_ISL_442395 | Virology Department, Sheffield Teaching Hospitals NHS Foundation Trust/Department of Infection, Immunity and Cardiovascular Disease, The Medical School, University of Sheffield | COVID-19 Genomics UK (COG-UK) Consortium |
| EPI_ISL_442396 | Virology Department, Sheffield Teaching Hospitals NHS Foundation Trust/Department of Infection, Immunity and Cardiovascular Disease, The Medical School, University of Sheffield | COVID-19 Genomics UK (COG-UK) Consortium |
| EPI_ISL_442397 | Virology Department, Sheffield Teaching Hospitals NHS Foundation Trust/Department of Infection, Immunity and Cardiovascular Disease, The Medical School, University of Sheffield | COVID-19 Genomics UK (COG-UK) Consortium |
| EPI_ISL_442398 | Virology Department, Sheffield Teaching Hospitals NHS Foundation Trust/Department of Infection, Immunity and Cardiovascular Disease, The Medical School, University of Sheffield | COVID-19 Genomics UK (COG-UK) Consortium |
| EPI_ISL_442399 | Virology Department, Sheffield Teaching Hospitals NHS Foundation Trust/Department of Infection, Immunity and Cardiovascular Disease, The Medical School, University of Sheffield | COVID-19 Genomics UK (COG-UK) Consortium |

| EPI_Accession_ID | Originating Laboratory | Submitting Laboratory |
|---|---|---|
| EPI_ISL_442400 | Virology Department, Sheffield Teaching Hospitals NHS Foundation Trust/Department of Infection, Immunity and Cardiovascular Disease, The Medical School, University of Sheffield | COVID-19 Genomics UK (COG-UK) Consortium |
| EPI_ISL_442401 | Virology Department, Sheffield Teaching Hospitals NHS Foundation Trust/Department of Infection, Immunity and Cardiovascular Disease, The Medical School, University of Sheffield | COVID-19 Genomics UK (COG-UK) Consortium |
| EPI_ISL_442402 | Virology Department, Sheffield Teaching Hospitals NHS Foundation Trust/Department of Infection, Immunity and Cardiovascular Disease, The Medical School, University of Sheffield | COVID-19 Genomics UK (COG-UK) Consortium |
| EPI_ISL_442403 | Virology Department, Sheffield Teaching Hospitals NHS Foundation Trust/Department of Infection, Immunity and Cardiovascular Disease, The Medical School, University of Sheffield | COVID-19 Genomics UK (COG-UK) Consortium |
| EPI_ISL_442404 | Virology Department, Sheffield Teaching Hospitals NHS Foundation Trust/Department of Infection, Immunity and Cardiovascular Disease, The Medical School, University of Sheffield | COVID-19 Genomics UK (COG-UK) Consortium |
| EPI_ISL_442405 | Virology Department, Sheffield Teaching Hospitals NHS Foundation Trust/Department of Infection, Immunity and Cardiovascular Disease, The Medical School, University of Sheffield | COVID-19 Genomics UK (COG-UK) Consortium |
| EPI_ISL_442406 | Virology Department, Sheffield Teaching Hospitals NHS Foundation Trust/Department of Infection, Immunity and Cardiovascular Disease, The Medical School, University of Sheffield | COVID-19 Genomics UK (COG-UK) Consortium |
| EPI_ISL_442407 | Virology Department, Sheffield Teaching Hospitals NHS Foundation Trust/Department of Infection, Immunity and Cardiovascular Disease, The Medical School, University of Sheffield | COVID-19 Genomics UK (COG-UK) Consortium |

| EPI_Accession_ID | Originating Laboratory | Submitting Laboratory |
|---|---|---|
| EPI_ISL_442408 | Virology Department, Sheffield Teaching Hospitals NHS Foundation Trust/Department of Infection, Immunity and Cardiovascular Disease, The Medical School, University of Sheffield | COVID-19 Genomics UK (COG-UK) Consortium |
| EPI_ISL_442409 | Virology Department, Sheffield Teaching Hospitals NHS Foundation Trust/Department of Infection, Immunity and Cardiovascular Disease, The Medical School, University of Sheffield | COVID-19 Genomics UK (COG-UK) Consortium |
| EPI_ISL_442410 | Virology Department, Sheffield Teaching Hospitals NHS Foundation Trust/Department of Infection, Immunity and Cardiovascular Disease, The Medical School, University of Sheffield | COVID-19 Genomics UK (COG-UK) Consortium |
| EPI_ISL_442411 | Virology Department, Sheffield Teaching Hospitals NHS Foundation Trust/Department of Infection, Immunity and Cardiovascular Disease, The Medical School, University of Sheffield | COVID-19 Genomics UK (COG-UK) Consortium |
| EPI_ISL_442412 | Virology Department, Sheffield Teaching Hospitals NHS Foundation Trust/Department of Infection, Immunity and Cardiovascular Disease, The Medical School, University of Sheffield | COVID-19 Genomics UK (COG-UK) Consortium |
| EPI_ISL_442413 | Virology Department, Sheffield Teaching Hospitals NHS Foundation Trust/Department of Infection, Immunity and Cardiovascular Disease, The Medical School, University of Sheffield | COVID-19 Genomics UK (COG-UK) Consortium |
| EPI_ISL_442414 | Virology Department, Sheffield Teaching Hospitals NHS Foundation Trust/Department of Infection, Immunity and Cardiovascular Disease, The Medical School, University of Sheffield | COVID-19 Genomics UK (COG-UK) Consortium |
| EPI_ISL_442415 | Virology Department, Sheffield Teaching Hospitals NHS Foundation Trust/Department of Infection, Immunity and Cardiovascular Disease, The Medical School, University of Sheffield | COVID-19 Genomics UK (COG-UK) Consortium |

| EPI_Accession_ID | Originating Laboratory | Submitting Laboratory |
| --- | --- | --- |
| EPI_ISL_442416 | Virology Department, Sheffield Teaching Hospitals NHS Foundation Trust/Department of Infection, Immunity and Cardiovascular Disease, The Medical School, University of Sheffield | COVID-19 Genomics UK (COG-UK) Consortium |
| EPI_ISL_442417 | Virology Department, Sheffield Teaching Hospitals NHS Foundation Trust/Department of Infection, Immunity and Cardiovascular Disease, The Medical School, University of Sheffield | COVID-19 Genomics UK (COG-UK) Consortium |
| EPI_ISL_442418 | Virology Department, Sheffield Teaching Hospitals NHS Foundation Trust/Department of Infection, Immunity and Cardiovascular Disease, The Medical School, University of Sheffield | COVID-19 Genomics UK (COG-UK) Consortium |
| EPI_ISL_442419 | Virology Department, Sheffield Teaching Hospitals NHS Foundation Trust/Department of Infection, Immunity and Cardiovascular Disease, The Medical School, University of Sheffield | COVID-19 Genomics UK (COG-UK) Consortium |
| EPI_ISL_442420 | Virology Department, Sheffield Teaching Hospitals NHS Foundation Trust/Department of Infection, Immunity and Cardiovascular Disease, The Medical School, University of Sheffield | COVID-19 Genomics UK (COG-UK) Consortium |
| EPI_ISL_442421 | Virology Department, Sheffield Teaching Hospitals NHS Foundation Trust/Department of Infection, Immunity and Cardiovascular Disease, The Medical School, University of Sheffield | COVID-19 Genomics UK (COG-UK) Consortium |
| EPI_ISL_442422 | Virology Department, Sheffield Teaching Hospitals NHS Foundation Trust/Department of Infection, Immunity and Cardiovascular Disease, The Medical School, University of Sheffield | COVID-19 Genomics UK (COG-UK) Consortium |
| EPI_ISL_442423 | Virology Department, Sheffield Teaching Hospitals NHS Foundation Trust/Department of Infection, Immunity and Cardiovascular Disease, The Medical School, University of Sheffield | COVID-19 Genomics UK (COG-UK) Consortium |

| EPI_Accession_ID | Originating Laboratory | Submitting Laboratory |
|---|---|---|
| EPI_ISL_442424 | Virology Department, Sheffield Teaching Hospitals NHS Foundation Trust/Department of Infection, Immunity and Cardiovascular Disease, The Medical School, University of Sheffield | COVID-19 Genomics UK (COG-UK) Consortium |
| EPI_ISL_442425 | Virology Department, Sheffield Teaching Hospitals NHS Foundation Trust/Department of Infection, Immunity and Cardiovascular Disease, The Medical School, University of Sheffield | COVID-19 Genomics UK (COG-UK) Consortium |
| EPI_ISL_442426 | Virology Department, Sheffield Teaching Hospitals NHS Foundation Trust/Department of Infection, Immunity and Cardiovascular Disease, The Medical School, University of Sheffield | COVID-19 Genomics UK (COG-UK) Consortium |
| EPI_ISL_442427 | Virology Department, Sheffield Teaching Hospitals NHS Foundation Trust/Department of Infection, Immunity and Cardiovascular Disease, The Medical School, University of Sheffield | COVID-19 Genomics UK (COG-UK) Consortium |
| EPI_ISL_442428 | Virology Department, Sheffield Teaching Hospitals NHS Foundation Trust/Department of Infection, Immunity and Cardiovascular Disease, The Medical School, University of Sheffield | COVID-19 Genomics UK (COG-UK) Consortium |
| EPI_ISL_442429 | Virology Department, Sheffield Teaching Hospitals NHS Foundation Trust/Department of Infection, Immunity and Cardiovascular Disease, The Medical School, University of Sheffield | COVID-19 Genomics UK (COG-UK) Consortium |
| EPI_ISL_442430 | Virology Department, Sheffield Teaching Hospitals NHS Foundation Trust/Department of Infection, Immunity and Cardiovascular Disease, The Medical School, University of Sheffield | COVID-19 Genomics UK (COG-UK) Consortium |
| EPI_ISL_442431 | Virology Department, Sheffield Teaching Hospitals NHS Foundation Trust/Department of Infection, Immunity and Cardiovascular Disease, The Medical School, University of Sheffield | COVID-19 Genomics UK (COG-UK) Consortium |

| EPI_Accession_ID | Originating Laboratory | Submitting Laboratory |
|---|---|---|
| EPI_ISL_442432 | Virology Department, Sheffield Teaching Hospitals NHS Foundation Trust/Department of Infection, Immunity and Cardiovascular Disease, The Medical School, University of Sheffield | COVID-19 Genomics UK (COG-UK) Consortium |
| EPI_ISL_442433 | Virology Department, Sheffield Teaching Hospitals NHS Foundation Trust/Department of Infection, Immunity and Cardiovascular Disease, The Medical School, University of Sheffield | COVID-19 Genomics UK (COG-UK) Consortium |
| EPI_ISL_442434 | Virology Department, Sheffield Teaching Hospitals NHS Foundation Trust/Department of Infection, Immunity and Cardiovascular Disease, The Medical School, University of Sheffield | COVID-19 Genomics UK (COG-UK) Consortium |
| EPI_ISL_442435 | Virology Department, Sheffield Teaching Hospitals NHS Foundation Trust/Department of Infection, Immunity and Cardiovascular Disease, The Medical School, University of Sheffield | COVID-19 Genomics UK (COG-UK) Consortium |
| EPI_ISL_442436 | Virology Department, Sheffield Teaching Hospitals NHS Foundation Trust/Department of Infection, Immunity and Cardiovascular Disease, The Medical School, University of Sheffield | COVID-19 Genomics UK (COG-UK) Consortium |
| EPI_ISL_442437 | Virology Department, Sheffield Teaching Hospitals NHS Foundation Trust/Department of Infection, Immunity and Cardiovascular Disease, The Medical School, University of Sheffield | COVID-19 Genomics UK (COG-UK) Consortium |
| EPI_ISL_442438 | Virology Department, Sheffield Teaching Hospitals NHS Foundation Trust/Department of Infection, Immunity and Cardiovascular Disease, The Medical School, University of Sheffield | COVID-19 Genomics UK (COG-UK) Consortium |
| EPI_ISL_442439 | Virology Department, Sheffield Teaching Hospitals NHS Foundation Trust/Department of Infection, Immunity and Cardiovascular Disease, The Medical School, University of Sheffield | COVID-19 Genomics UK (COG-UK) Consortium |

| EPI_Accession_ID | Originating Laboratory | Submitting Laboratory |
|---|---|---|
| EPI_ISL_442440 | Virology Department, Sheffield Teaching Hospitals NHS Foundation Trust/Department of Infection, Immunity and Cardiovascular Disease, The Medical School, University of Sheffield | COVID-19 Genomics UK (COG-UK) Consortium |
| EPI_ISL_442441 | Virology Department, Sheffield Teaching Hospitals NHS Foundation Trust/Department of Infection, Immunity and Cardiovascular Disease, The Medical School, University of Sheffield | COVID-19 Genomics UK (COG-UK) Consortium |
| EPI_ISL_442442 | Virology Department, Sheffield Teaching Hospitals NHS Foundation Trust/Department of Infection, Immunity and Cardiovascular Disease, The Medical School, University of Sheffield | COVID-19 Genomics UK (COG-UK) Consortium |
| EPI_ISL_442443 | Virology Department, Sheffield Teaching Hospitals NHS Foundation Trust/Department of Infection, Immunity and Cardiovascular Disease, The Medical School, University of Sheffield | COVID-19 Genomics UK (COG-UK) Consortium |
| EPI_ISL_442444 | Virology Department, Sheffield Teaching Hospitals NHS Foundation Trust/Department of Infection, Immunity and Cardiovascular Disease, The Medical School, University of Sheffield | COVID-19 Genomics UK (COG-UK) Consortium |
| EPI_ISL_442445 | Virology Department, Sheffield Teaching Hospitals NHS Foundation Trust/Department of Infection, Immunity and Cardiovascular Disease, The Medical School, University of Sheffield | COVID-19 Genomics UK (COG-UK) Consortium |
| EPI_ISL_442446 | Virology Department, Sheffield Teaching Hospitals NHS Foundation Trust/Department of Infection, Immunity and Cardiovascular Disease, The Medical School, University of Sheffield | COVID-19 Genomics UK (COG-UK) Consortium |
| EPI_ISL_442447 | Virology Department, Sheffield Teaching Hospitals NHS Foundation Trust/Department of Infection, Immunity and Cardiovascular Disease, The Medical School, University of Sheffield | COVID-19 Genomics UK (COG-UK) Consortium |

| EPI_Accession_ID | Originating Laboratory | Submitting Laboratory |
| --- | --- | --- |
| EPI_ISL_442448 | Virology Department, Sheffield Teaching Hospitals NHS Foundation Trust/Department of Infection, Immunity and Cardiovascular Disease, The Medical School, University of Sheffield | COVID-19 Genomics UK (COG-UK) Consortium |
| EPI_ISL_442449 | Virology Department, Sheffield Teaching Hospitals NHS Foundation Trust/Department of Infection, Immunity and Cardiovascular Disease, The Medical School, University of Sheffield | COVID-19 Genomics UK (COG-UK) Consortium |
| EPI_ISL_442450 | Virology Department, Sheffield Teaching Hospitals NHS Foundation Trust/Department of Infection, Immunity and Cardiovascular Disease, The Medical School, University of Sheffield | COVID-19 Genomics UK (COG-UK) Consortium |
| EPI_ISL_442451 | Virology Department, Sheffield Teaching Hospitals NHS Foundation Trust/Department of Infection, Immunity and Cardiovascular Disease, The Medical School, University of Sheffield | COVID-19 Genomics UK (COG-UK) Consortium |
| EPI_ISL_442452 | Virology Department, Sheffield Teaching Hospitals NHS Foundation Trust/Department of Infection, Immunity and Cardiovascular Disease, The Medical School, University of Sheffield | COVID-19 Genomics UK (COG-UK) Consortium |
| EPI_ISL_442453 | Virology Department, Sheffield Teaching Hospitals NHS Foundation Trust/Department of Infection, Immunity and Cardiovascular Disease, The Medical School, University of Sheffield | COVID-19 Genomics UK (COG-UK) Consortium |
| EPI_ISL_442454 | Virology Department, Sheffield Teaching Hospitals NHS Foundation Trust/Department of Infection, Immunity and Cardiovascular Disease, The Medical School, University of Sheffield | COVID-19 Genomics UK (COG-UK) Consortium |
| EPI_ISL_442455 | Virology Department, Sheffield Teaching Hospitals NHS Foundation Trust/Department of Infection, Immunity and Cardiovascular Disease, The Medical School, University of Sheffield | COVID-19 Genomics UK (COG-UK) Consortium |

| EPI_Accession_ID | Originating Laboratory | Submitting Laboratory |
|---|---|---|
| EPI_ISL_442456 | Virology Department, Sheffield Teaching Hospitals NHS Foundation Trust/Department of Infection, Immunity and Cardiovascular Disease, The Medical School, University of Sheffield | COVID-19 Genomics UK (COG-UK) Consortium |
| EPI_ISL_442457 | Virology Department, Sheffield Teaching Hospitals NHS Foundation Trust/Department of Infection, Immunity and Cardiovascular Disease, The Medical School, University of Sheffield | COVID-19 Genomics UK (COG-UK) Consortium |
| EPI_ISL_442458 | Virology Department, Sheffield Teaching Hospitals NHS Foundation Trust/Department of Infection, Immunity and Cardiovascular Disease, The Medical School, University of Sheffield | COVID-19 Genomics UK (COG-UK) Consortium |
| EPI_ISL_442459 | Virology Department, Sheffield Teaching Hospitals NHS Foundation Trust/Department of Infection, Immunity and Cardiovascular Disease, The Medical School, University of Sheffield | COVID-19 Genomics UK (COG-UK) Consortium |
| EPI_ISL_442460 | Virology Department, Sheffield Teaching Hospitals NHS Foundation Trust/Department of Infection, Immunity and Cardiovascular Disease, The Medical School, University of Sheffield | COVID-19 Genomics UK (COG-UK) Consortium |
| EPI_ISL_442461 | Virology Department, Sheffield Teaching Hospitals NHS Foundation Trust/Department of Infection, Immunity and Cardiovascular Disease, The Medical School, University of Sheffield | COVID-19 Genomics UK (COG-UK) Consortium |
| EPI_ISL_442462 | Virology Department, Sheffield Teaching Hospitals NHS Foundation Trust/Department of Infection, Immunity and Cardiovascular Disease, The Medical School, University of Sheffield | COVID-19 Genomics UK (COG-UK) Consortium |
| EPI_ISL_442463 | Virology Department, Sheffield Teaching Hospitals NHS Foundation Trust/Department of Infection, Immunity and Cardiovascular Disease, The Medical School, University of Sheffield | COVID-19 Genomics UK (COG-UK) Consortium |

| EPI_Accession_ID | Originating Laboratory | Submitting Laboratory |
|---|---|---|
| EPI_ISL_442464 | Virology Department, Sheffield Teaching Hospitals NHS Foundation Trust/Department of Infection, Immunity and Cardiovascular Disease, The Medical School, University of Sheffield | COVID-19 Genomics UK (COG-UK) Consortium |
| EPI_ISL_442465 | Virology Department, Sheffield Teaching Hospitals NHS Foundation Trust/Department of Infection, Immunity and Cardiovascular Disease, The Medical School, University of Sheffield | COVID-19 Genomics UK (COG-UK) Consortium |
| EPI_ISL_442466 | Virology Department, Sheffield Teaching Hospitals NHS Foundation Trust/Department of Infection, Immunity and Cardiovascular Disease, The Medical School, University of Sheffield | COVID-19 Genomics UK (COG-UK) Consortium |
| EPI_ISL_442467 | Virology Department, Sheffield Teaching Hospitals NHS Foundation Trust/Department of Infection, Immunity and Cardiovascular Disease, The Medical School, University of Sheffield | COVID-19 Genomics UK (COG-UK) Consortium |
| EPI_ISL_442468 | Virology Department, Sheffield Teaching Hospitals NHS Foundation Trust/Department of Infection, Immunity and Cardiovascular Disease, The Medical School, University of Sheffield | COVID-19 Genomics UK (COG-UK) Consortium |
| EPI_ISL_442469 | Virology Department, Sheffield Teaching Hospitals NHS Foundation Trust/Department of Infection, Immunity and Cardiovascular Disease, The Medical School, University of Sheffield | COVID-19 Genomics UK (COG-UK) Consortium |
| EPI_ISL_442470 | Virology Department, Sheffield Teaching Hospitals NHS Foundation Trust/Department of Infection, Immunity and Cardiovascular Disease, The Medical School, University of Sheffield | COVID-19 Genomics UK (COG-UK) Consortium |
| EPI_ISL_442471 | Virology Department, Sheffield Teaching Hospitals NHS Foundation Trust/Department of Infection, Immunity and Cardiovascular Disease, The Medical School, University of Sheffield | COVID-19 Genomics UK (COG-UK) Consortium |

| EPI_Accession_ID | Originating Laboratory | Submitting Laboratory |
| --- | --- | --- |
| EPI_ISL_442472 | Virology Department, Sheffield Teaching Hospitals NHS Foundation Trust/Department of Infection, Immunity and Cardiovascular Disease, The Medical School, University of Sheffield | COVID-19 Genomics UK (COG-UK) Consortium |
| EPI_ISL_442473 | Virology Department, Sheffield Teaching Hospitals NHS Foundation Trust/Department of Infection, Immunity and Cardiovascular Disease, The Medical School, University of Sheffield | COVID-19 Genomics UK (COG-UK) Consortium |
| EPI_ISL_442474 | Virology Department, Sheffield Teaching Hospitals NHS Foundation Trust/Department of Infection, Immunity and Cardiovascular Disease, The Medical School, University of Sheffield | COVID-19 Genomics UK (COG-UK) Consortium |
| EPI_ISL_442475 | Virology Department, Sheffield Teaching Hospitals NHS Foundation Trust/Department of Infection, Immunity and Cardiovascular Disease, The Medical School, University of Sheffield | COVID-19 Genomics UK (COG-UK) Consortium |
| EPI_ISL_442476 | Virology Department, Sheffield Teaching Hospitals NHS Foundation Trust/Department of Infection, Immunity and Cardiovascular Disease, The Medical School, University of Sheffield | COVID-19 Genomics UK (COG-UK) Consortium |
| EPI_ISL_442477 | Virology Department, Sheffield Teaching Hospitals NHS Foundation Trust/Department of Infection, Immunity and Cardiovascular Disease, The Medical School, University of Sheffield | COVID-19 Genomics UK (COG-UK) Consortium |
| EPI_ISL_442478 | Virology Department, Sheffield Teaching Hospitals NHS Foundation Trust/Department of Infection, Immunity and Cardiovascular Disease, The Medical School, University of Sheffield | COVID-19 Genomics UK (COG-UK) Consortium |
| EPI_ISL_442479 | Virology Department, Sheffield Teaching Hospitals NHS Foundation Trust/Department of Infection, Immunity and Cardiovascular Disease, The Medical School, University of Sheffield | COVID-19 Genomics UK (COG-UK) Consortium |

| EPI_Accession_ID | Originating Laboratory | Submitting Laboratory |
|---|---|---|
| EPI_ISL_442480 | Virology Department, Sheffield Teaching Hospitals NHS Foundation Trust/Department of Infection, Immunity and Cardiovascular Disease, The Medical School, University of Sheffield | COVID-19 Genomics UK (COG-UK) Consortium |
| EPI_ISL_442481 | Virology Department, Sheffield Teaching Hospitals NHS Foundation Trust/Department of Infection, Immunity and Cardiovascular Disease, The Medical School, University of Sheffield | COVID-19 Genomics UK (COG-UK) Consortium |
| EPI_ISL_442482 | Virology Department, Sheffield Teaching Hospitals NHS Foundation Trust/Department of Infection, Immunity and Cardiovascular Disease, The Medical School, University of Sheffield | COVID-19 Genomics UK (COG-UK) Consortium |
| EPI_ISL_442483 | Virology Department, Sheffield Teaching Hospitals NHS Foundation Trust/Department of Infection, Immunity and Cardiovascular Disease, The Medical School, University of Sheffield | COVID-19 Genomics UK (COG-UK) Consortium |
| EPI_ISL_442484 | Virology Department, Sheffield Teaching Hospitals NHS Foundation Trust/Department of Infection, Immunity and Cardiovascular Disease, The Medical School, University of Sheffield | COVID-19 Genomics UK (COG-UK) Consortium |
| EPI_ISL_442485 | Virology Department, Sheffield Teaching Hospitals NHS Foundation Trust/Department of Infection, Immunity and Cardiovascular Disease, The Medical School, University of Sheffield | COVID-19 Genomics UK (COG-UK) Consortium |
| EPI_ISL_442486 | Virology Department, Sheffield Teaching Hospitals NHS Foundation Trust/Department of Infection, Immunity and Cardiovascular Disease, The Medical School, University of Sheffield | COVID-19 Genomics UK (COG-UK) Consortium |
| EPI_ISL_442487 | Virology Department, Sheffield Teaching Hospitals NHS Foundation Trust/Department of Infection, Immunity and Cardiovascular Disease, The Medical School, University of Sheffield | COVID-19 Genomics UK (COG-UK) Consortium |

| EPI_Accession_ID | Originating Laboratory | Submitting Laboratory |
|---|---|---|
| EPI_ISL_442488 | Virology Department, Sheffield Teaching Hospitals NHS Foundation Trust/Department of Infection, Immunity and Cardiovascular Disease, The Medical School, University of Sheffield | COVID-19 Genomics UK (COG-UK) Consortium |
| EPI_ISL_442489 | Virology Department, Sheffield Teaching Hospitals NHS Foundation Trust/Department of Infection, Immunity and Cardiovascular Disease, The Medical School, University of Sheffield | COVID-19 Genomics UK (COG-UK) Consortium |
| EPI_ISL_442490 | Virology Department, Sheffield Teaching Hospitals NHS Foundation Trust/Department of Infection, Immunity and Cardiovascular Disease, The Medical School, University of Sheffield | COVID-19 Genomics UK (COG-UK) Consortium |
| EPI_ISL_442491 | Virology Department, Sheffield Teaching Hospitals NHS Foundation Trust/Department of Infection, Immunity and Cardiovascular Disease, The Medical School, University of Sheffield | COVID-19 Genomics UK (COG-UK) Consortium |
| EPI_ISL_442492 | Virology Department, Sheffield Teaching Hospitals NHS Foundation Trust/Department of Infection, Immunity and Cardiovascular Disease, The Medical School, University of Sheffield | COVID-19 Genomics UK (COG-UK) Consortium |
| EPI_ISL_442493 | Virology Department, Sheffield Teaching Hospitals NHS Foundation Trust/Department of Infection, Immunity and Cardiovascular Disease, The Medical School, University of Sheffield | COVID-19 Genomics UK (COG-UK) Consortium |
| EPI_ISL_442494 | Virology Department, Sheffield Teaching Hospitals NHS Foundation Trust/Department of Infection, Immunity and Cardiovascular Disease, The Medical School, University of Sheffield | COVID-19 Genomics UK (COG-UK) Consortium |
| EPI_ISL_442495 | Virology Department, Sheffield Teaching Hospitals NHS Foundation Trust/Department of Infection, Immunity and Cardiovascular Disease, The Medical School, University of Sheffield | COVID-19 Genomics UK (COG-UK) Consortium |

| EPI_Accession_ID | Originating Laboratory | Submitting Laboratory |
|---|---|---|
| EPI_ISL_442496 | Virology Department, Sheffield Teaching Hospitals NHS Foundation Trust/Department of Infection, Immunity and Cardiovascular Disease, The Medical School, University of Sheffield | COVID-19 Genomics UK (COG-UK) Consortium |
| EPI_ISL_442497 | Virology Department, Sheffield Teaching Hospitals NHS Foundation Trust/Department of Infection, Immunity and Cardiovascular Disease, The Medical School, University of Sheffield | COVID-19 Genomics UK (COG-UK) Consortium |
| EPI_ISL_442498 | Virology Department, Sheffield Teaching Hospitals NHS Foundation Trust/Department of Infection, Immunity and Cardiovascular Disease, The Medical School, University of Sheffield | COVID-19 Genomics UK (COG-UK) Consortium |
| EPI_ISL_442499 | Virology Department, Sheffield Teaching Hospitals NHS Foundation Trust/Department of Infection, Immunity and Cardiovascular Disease, The Medical School, University of Sheffield | COVID-19 Genomics UK (COG-UK) Consortium |
| EPI_ISL_442500 | Virology Department, Sheffield Teaching Hospitals NHS Foundation Trust/Department of Infection, Immunity and Cardiovascular Disease, The Medical School, University of Sheffield | COVID-19 Genomics UK (COG-UK) Consortium |
| EPI_ISL_442501 | Virology Department, Sheffield Teaching Hospitals NHS Foundation Trust/Department of Infection, Immunity and Cardiovascular Disease, The Medical School, University of Sheffield | COVID-19 Genomics UK (COG-UK) Consortium |
| EPI_ISL_442502 | Virology Department, Sheffield Teaching Hospitals NHS Foundation Trust/Department of Infection, Immunity and Cardiovascular Disease, The Medical School, University of Sheffield | COVID-19 Genomics UK (COG-UK) Consortium |
| EPI_ISL_442503 | Virology Department, Sheffield Teaching Hospitals NHS Foundation Trust/Department of Infection, Immunity and Cardiovascular Disease, The Medical School, University of Sheffield | COVID-19 Genomics UK (COG-UK) Consortium |

| EPI_Accession_ID | Originating Laboratory | Submitting Laboratory |
|---|---|---|
| EPI_ISL_442504 | Virology Department, Sheffield Teaching Hospitals NHS Foundation Trust/Department of Infection, Immunity and Cardiovascular Disease, The Medical School, University of Sheffield | COVID-19 Genomics UK (COG-UK) Consortium |
| EPI_ISL_442505 | Virology Department, Sheffield Teaching Hospitals NHS Foundation Trust/Department of Infection, Immunity and Cardiovascular Disease, The Medical School, University of Sheffield | COVID-19 Genomics UK (COG-UK) Consortium |
| EPI_ISL_442506 | Virology Department, Sheffield Teaching Hospitals NHS Foundation Trust/Department of Infection, Immunity and Cardiovascular Disease, The Medical School, University of Sheffield | COVID-19 Genomics UK (COG-UK) Consortium |
| EPI_ISL_442507 | Virology Department, Sheffield Teaching Hospitals NHS Foundation Trust/Department of Infection, Immunity and Cardiovascular Disease, The Medical School, University of Sheffield | COVID-19 Genomics UK (COG-UK) Consortium |
| EPI_ISL_442508 | Virology Department, Sheffield Teaching Hospitals NHS Foundation Trust/Department of Infection, Immunity and Cardiovascular Disease, The Medical School, University of Sheffield | COVID-19 Genomics UK (COG-UK) Consortium |
| EPI_ISL_442509 | Virology Department, Sheffield Teaching Hospitals NHS Foundation Trust/Department of Infection, Immunity and Cardiovascular Disease, The Medical School, University of Sheffield | COVID-19 Genomics UK (COG-UK) Consortium |
| EPI_ISL_442510 | Virology Department, Sheffield Teaching Hospitals NHS Foundation Trust/Department of Infection, Immunity and Cardiovascular Disease, The Medical School, University of Sheffield | COVID-19 Genomics UK (COG-UK) Consortium |
| EPI_ISL_442511 | Virology Department, Sheffield Teaching Hospitals NHS Foundation Trust/Department of Infection, Immunity and Cardiovascular Disease, The Medical School, University of Sheffield | COVID-19 Genomics UK (COG-UK) Consortium |

| EPI_Accession_ID | Originating Laboratory | Submitting Laboratory |
|---|---|---|
| EPI_ISL_442512 | Virology Department, Sheffield Teaching Hospitals NHS Foundation Trust/Department of Infection, Immunity and Cardiovascular Disease, The Medical School, University of Sheffield | COVID-19 Genomics UK (COG-UK) Consortium |
| EPI_ISL_442513 | Virology Department, Sheffield Teaching Hospitals NHS Foundation Trust/Department of Infection, Immunity and Cardiovascular Disease, The Medical School, University of Sheffield | COVID-19 Genomics UK (COG-UK) Consortium |
| EPI_ISL_442514 | Virology Department, Sheffield Teaching Hospitals NHS Foundation Trust/Department of Infection, Immunity and Cardiovascular Disease, The Medical School, University of Sheffield | COVID-19 Genomics UK (COG-UK) Consortium |
| EPI_ISL_442515 | Virology Department, Sheffield Teaching Hospitals NHS Foundation Trust/Department of Infection, Immunity and Cardiovascular Disease, The Medical School, University of Sheffield | COVID-19 Genomics UK (COG-UK) Consortium |
| EPI_ISL_442516 | Virology Department, Sheffield Teaching Hospitals NHS Foundation Trust/Department of Infection, Immunity and Cardiovascular Disease, The Medical School, University of Sheffield | COVID-19 Genomics UK (COG-UK) Consortium |
| EPI_ISL_442517 | Virology Department, Sheffield Teaching Hospitals NHS Foundation Trust/Department of Infection, Immunity and Cardiovascular Disease, The Medical School, University of Sheffield | COVID-19 Genomics UK (COG-UK) Consortium |
| EPI_ISL_442518 | Virology Department, Sheffield Teaching Hospitals NHS Foundation Trust/Department of Infection, Immunity and Cardiovascular Disease, The Medical School, University of Sheffield | COVID-19 Genomics UK (COG-UK) Consortium |
| EPI_ISL_442519 | Virology Department, Sheffield Teaching Hospitals NHS Foundation Trust/Department of Infection, Immunity and Cardiovascular Disease, The Medical School, University of Sheffield | COVID-19 Genomics UK (COG-UK) Consortium |

| EPI_Accession_ID | Originating Laboratory | Submitting Laboratory |
|---|---|---|
| EPI_ISL_442520 | Virology Department, Sheffield Teaching Hospitals NHS Foundation Trust/Department of Infection, Immunity and Cardiovascular Disease, The Medical School, University of Sheffield | COVID-19 Genomics UK (COG-UK) Consortium |
| EPI_ISL_442521 | Virology Department, Sheffield Teaching Hospitals NHS Foundation Trust/Department of Infection, Immunity and Cardiovascular Disease, The Medical School, University of Sheffield | COVID-19 Genomics UK (COG-UK) Consortium |
| EPI_ISL_442522 | Virology Department, Sheffield Teaching Hospitals NHS Foundation Trust/Department of Infection, Immunity and Cardiovascular Disease, The Medical School, University of Sheffield | COVID-19 Genomics UK (COG-UK) Consortium |
| EPI_ISL_442523 | Pasteur Institute of Iran | Kawsar Human Genetic Research Company |
| EPI_ISL_442524 | Department of Pathology, University of Cambridge | Wellcome Sanger Institute for the COVID-19 Genomics UK Consortium |
| EPI_ISL_442525 | Department of Pathology, University of Cambridge | Wellcome Sanger Institute for the COVID-19 Genomics UK Consortium |
| EPI_ISL_442526 | Department of Pathology, University of Cambridge | Wellcome Sanger Institute for the COVID-19 Genomics UK Consortium |
| EPI_ISL_442527 | Department of Pathology, University of Cambridge | Wellcome Sanger Institute for the COVID-19 Genomics UK Consortium |
| EPI_ISL_442528 | Department of Pathology, University of Cambridge | Wellcome Sanger Institute for the COVID-19 Genomics UK Consortium |
| EPI_ISL_442529 | Department of Pathology, University of Cambridge | Wellcome Sanger Institute for the COVID-19 Genomics UK Consortium |
| EPI_ISL_442530 | Department of Pathology, University of Cambridge | Wellcome Sanger Institute for the COVID-19 Genomics UK Consortium |
| EPI_ISL_442531 | Department of Pathology, University of Cambridge | Wellcome Sanger Institute for the COVID-19 Genomics UK Consortium |
| EPI_ISL_442532 | Department of Pathology, University of Cambridge | Wellcome Sanger Institute for the COVID-19 Genomics UK Consortium |
| EPI_ISL_442533 | Department of Pathology, University of Cambridge | Wellcome Sanger Institute for the COVID-19 Genomics UK Consortium |

| EPI_Accession_ID | Originating Laboratory | Submitting Laboratory |
|---|---|---|
| EPI_ISL_442534 | Department of Pathology, University of Cambridge | Wellcome Sanger Institute for the COVID-19 Genomics UK Consortium |
| EPI_ISL_442535 | Department of Pathology, University of Cambridge | Wellcome Sanger Institute for the COVID-19 Genomics UK Consortium |
| EPI_ISL_442536 | Department of Pathology, University of Cambridge | Wellcome Sanger Institute for the COVID-19 Genomics UK Consortium |
| EPI_ISL_442537 | Department of Pathology, University of Cambridge | Wellcome Sanger Institute for the COVID-19 Genomics UK Consortium |
| EPI_ISL_442538 | Department of Pathology, University of Cambridge | Wellcome Sanger Institute for the COVID-19 Genomics UK Consortium |
| EPI_ISL_442539 | Department of Pathology, University of Cambridge | Wellcome Sanger Institute for the COVID-19 Genomics UK Consortium |
| EPI_ISL_442540 | Department of Pathology, University of Cambridge | Wellcome Sanger Institute for the COVID-19 Genomics UK Consortium |
| EPI_ISL_442541 | Department of Pathology, University of Cambridge | Wellcome Sanger Institute for the COVID-19 Genomics UK Consortium |
| EPI_ISL_442542 | Department of Pathology, University of Cambridge | Wellcome Sanger Institute for the COVID-19 Genomics UK Consortium |
| EPI_ISL_442543 | Department of Pathology, University of Cambridge | Wellcome Sanger Institute for the COVID-19 Genomics UK Consortium |
| EPI_ISL_442544 | Department of Pathology, University of Cambridge | Wellcome Sanger Institute for the COVID-19 Genomics UK Consortium |
| EPI_ISL_442545 | Department of Pathology, University of Cambridge | Wellcome Sanger Institute for the COVID-19 Genomics UK Consortium |
| EPI_ISL_442546 | Department of Pathology, University of Cambridge | Wellcome Sanger Institute for the COVID-19 Genomics UK Consortium |
| EPI_ISL_442547 | Department of Pathology, University of Cambridge | Wellcome Sanger Institute for the COVID-19 Genomics UK Consortium |
| EPI_ISL_442548 | Department of Pathology, University of Cambridge | Wellcome Sanger Institute for the COVID-19 Genomics UK Consortium |
| EPI_ISL_442549 | Department of Pathology, University of Cambridge | Wellcome Sanger Institute for the COVID-19 Genomics UK Consortium |

| EPI_Accession_ID | Originating Laboratory | Submitting Laboratory |
| --- | --- | --- |
| EPI_ISL_442550 | Department of Pathology, University of Cambridge | Wellcome Sanger Institute for the COVID-19 Genomics UK Consortium |
| EPI_ISL_442551 | Department of Pathology, University of Cambridge | Wellcome Sanger Institute for the COVID-19 Genomics UK Consortium |
| EPI_ISL_442552 | Department of Pathology, University of Cambridge | Wellcome Sanger Institute for the COVID-19 Genomics UK Consortium |
| EPI_ISL_442553 | Department of Pathology, University of Cambridge | Wellcome Sanger Institute for the COVID-19 Genomics UK Consortium |
| EPI_ISL_442554 | Department of Pathology, University of Cambridge | Wellcome Sanger Institute for the COVID-19 Genomics UK Consortium |
| EPI_ISL_442555 | Department of Pathology, University of Cambridge | Wellcome Sanger Institute for the COVID-19 Genomics UK Consortium |
| EPI_ISL_442556 | Department of Pathology, University of Cambridge | Wellcome Sanger Institute for the COVID-19 Genomics UK Consortium |
| EPI_ISL_442557 | Department of Pathology, University of Cambridge | Wellcome Sanger Institute for the COVID-19 Genomics UK Consortium |
| EPI_ISL_442558 | Department of Pathology, University of Cambridge | Wellcome Sanger Institute for the COVID-19 Genomics UK Consortium |
| EPI_ISL_442559 | Department of Pathology, University of Cambridge | Wellcome Sanger Institute for the COVID-19 Genomics UK Consortium |
| EPI_ISL_442560 | Department of Pathology, University of Cambridge | Wellcome Sanger Institute for the COVID-19 Genomics UK Consortium |
| EPI_ISL_442561 | Department of Pathology, University of Cambridge | Wellcome Sanger Institute for the COVID-19 Genomics UK Consortium |
| EPI_ISL_442562 | Department of Pathology, University of Cambridge | Wellcome Sanger Institute for the COVID-19 Genomics UK Consortium |
| EPI_ISL_442563 | Department of Pathology, University of Cambridge | Wellcome Sanger Institute for the COVID-19 Genomics UK Consortium |
| EPI_ISL_442564 | Department of Pathology, University of Cambridge | Wellcome Sanger Institute for the COVID-19 Genomics UK Consortium |
| EPI_ISL_442565 | Department of Pathology, University of Cambridge | Wellcome Sanger Institute for the COVID-19 Genomics UK Consortium |

| EPI_Accession_ID | Originating Laboratory | Submitting Laboratory |
|---|---|---|
| EPI_ISL_442566 | Department of Pathology, University of Cambridge | Wellcome Sanger Institute for the COVID-19 Genomics UK Consortium |
| EPI_ISL_442567 | Department of Pathology, University of Cambridge | Wellcome Sanger Institute for the COVID-19 Genomics UK Consortium |
| EPI_ISL_442568 | Department of Pathology, University of Cambridge | Wellcome Sanger Institute for the COVID-19 Genomics UK Consortium |
| EPI_ISL_442569 | Department of Pathology, University of Cambridge | Wellcome Sanger Institute for the COVID-19 Genomics UK Consortium |
| EPI_ISL_442570 | Department of Pathology, University of Cambridge | Wellcome Sanger Institute for the COVID-19 Genomics UK Consortium |
| EPI_ISL_442571 | Department of Pathology, University of Cambridge | Wellcome Sanger Institute for the COVID-19 Genomics UK Consortium |
| EPI_ISL_442572 | Department of Pathology, University of Cambridge | Wellcome Sanger Institute for the COVID-19 Genomics UK Consortium |
| EPI_ISL_442573 | Department of Pathology, University of Cambridge | Wellcome Sanger Institute for the COVID-19 Genomics UK Consortium |
| EPI_ISL_442574 | Department of Pathology, University of Cambridge | Wellcome Sanger Institute for the COVID-19 Genomics UK Consortium |
| EPI_ISL_442575 | Department of Pathology, University of Cambridge | Wellcome Sanger Institute for the COVID-19 Genomics UK Consortium |
| EPI_ISL_442576 | Department of Pathology, University of Cambridge | Wellcome Sanger Institute for the COVID-19 Genomics UK Consortium |
| EPI_ISL_442577 | Department of Pathology, University of Cambridge | Wellcome Sanger Institute for the COVID-19 Genomics UK Consortium |
| EPI_ISL_442578 | Department of Pathology, University of Cambridge | Wellcome Sanger Institute for the COVID-19 Genomics UK Consortium |
| EPI_ISL_442579 | Department of Pathology, University of Cambridge | Wellcome Sanger Institute for the COVID-19 Genomics UK Consortium |
| EPI_ISL_442580 | Department of Pathology, University of Cambridge | Wellcome Sanger Institute for the COVID-19 Genomics UK Consortium |
| EPI_ISL_442581 | Department of Pathology, University of Cambridge | Wellcome Sanger Institute for the COVID-19 Genomics UK Consortium |

| EPI_Accession_ID | Originating Laboratory | Submitting Laboratory |
|---|---|---|
| EPI_ISL_442582 | Department of Pathology, University of Cambridge | Wellcome Sanger Institute for the COVID-19 Genomics UK Consortium |
| EPI_ISL_442583 | Department of Pathology, University of Cambridge | Wellcome Sanger Institute for the COVID-19 Genomics UK Consortium |
| EPI_ISL_442584 | Department of Pathology, University of Cambridge | Wellcome Sanger Institute for the COVID-19 Genomics UK Consortium |
| EPI_ISL_442585 | Department of Pathology, University of Cambridge | Wellcome Sanger Institute for the COVID-19 Genomics UK Consortium |
| EPI_ISL_442586 | Department of Pathology, University of Cambridge | Wellcome Sanger Institute for the COVID-19 Genomics UK Consortium |
| EPI_ISL_442587 | Department of Pathology, University of Cambridge | Wellcome Sanger Institute for the COVID-19 Genomics UK Consortium |
| EPI_ISL_442588 | Department of Pathology, University of Cambridge | Wellcome Sanger Institute for the COVID-19 Genomics UK Consortium |
| EPI_ISL_442589 | Department of Pathology, University of Cambridge | Wellcome Sanger Institute for the COVID-19 Genomics UK Consortium |
| EPI_ISL_442590 | Department of Pathology, University of Cambridge | Wellcome Sanger Institute for the COVID-19 Genomics UK Consortium |
| EPI_ISL_442591 | Department of Pathology, University of Cambridge | Wellcome Sanger Institute for the COVID-19 Genomics UK Consortium |
| EPI_ISL_442592 | Department of Pathology, University of Cambridge | Wellcome Sanger Institute for the COVID-19 Genomics UK Consortium |
| EPI_ISL_442593 | Department of Pathology, University of Cambridge | Wellcome Sanger Institute for the COVID-19 Genomics UK Consortium |
| EPI_ISL_442594 | Department of Pathology, University of Cambridge | Wellcome Sanger Institute for the COVID-19 Genomics UK Consortium |
| EPI_ISL_442595 | Department of Pathology, University of Cambridge | Wellcome Sanger Institute for the COVID-19 Genomics UK Consortium |
| EPI_ISL_442596 | Department of Pathology, University of Cambridge | Wellcome Sanger Institute for the COVID-19 Genomics UK Consortium |
| EPI_ISL_442597 | Department of Pathology, University of Cambridge | Wellcome Sanger Institute for the COVID-19 Genomics UK Consortium |

| EPI_Accession_ID | Originating Laboratory | Submitting Laboratory |
|---|---|---|
| EPI_ISL_442598 | Department of Pathology, University of Cambridge | Wellcome Sanger Institute for the COVID-19 Genomics UK Consortium |
| EPI_ISL_442599 | Department of Pathology, University of Cambridge | Wellcome Sanger Institute for the COVID-19 Genomics UK Consortium |
| EPI_ISL_442600 | Department of Pathology, University of Cambridge | Wellcome Sanger Institute for the COVID-19 Genomics UK Consortium |
| EPI_ISL_442601 | Department of Pathology, University of Cambridge | Wellcome Sanger Institute for the COVID-19 Genomics UK Consortium |
| EPI_ISL_442602 | Department of Pathology, University of Cambridge | Wellcome Sanger Institute for the COVID-19 Genomics UK Consortium |
| EPI_ISL_442603 | Department of Pathology, University of Cambridge | Wellcome Sanger Institute for the COVID-19 Genomics UK Consortium |
| EPI_ISL_442604 | Department of Pathology, University of Cambridge | Wellcome Sanger Institute for the COVID-19 Genomics UK Consortium |
| EPI_ISL_442605 | Department of Pathology, University of Cambridge | Wellcome Sanger Institute for the COVID-19 Genomics UK Consortium |
| EPI_ISL_442606 | Department of Pathology, University of Cambridge | Wellcome Sanger Institute for the COVID-19 Genomics UK Consortium |
| EPI_ISL_442607 | Department of Pathology, University of Cambridge | Wellcome Sanger Institute for the COVID-19 Genomics UK Consortium |
| EPI_ISL_442608 | Department of Pathology, University of Cambridge | Wellcome Sanger Institute for the COVID-19 Genomics UK Consortium |
| EPI_ISL_442609 | Department of Pathology, University of Cambridge | Wellcome Sanger Institute for the COVID-19 Genomics UK Consortium |
| EPI_ISL_442610 | Department of Pathology, University of Cambridge | Wellcome Sanger Institute for the COVID-19 Genomics UK Consortium |
| EPI_ISL_442611 | Department of Pathology, University of Cambridge | Wellcome Sanger Institute for the COVID-19 Genomics UK Consortium |
| EPI_ISL_442612 | Department of Pathology, University of Cambridge | Wellcome Sanger Institute for the COVID-19 Genomics UK Consortium |
| EPI_ISL_442613 | Department of Pathology, University of Cambridge | Wellcome Sanger Institute for the COVID-19 Genomics UK Consortium |

| EPI_Accession_ID | Originating Laboratory | Submitting Laboratory |
|---|---|---|
| EPI_ISL_442614 | Department of Pathology, University of Cambridge | Wellcome Sanger Institute for the COVID-19 Genomics UK Consortium |
| EPI_ISL_442615 | Department of Pathology, University of Cambridge | Wellcome Sanger Institute for the COVID-19 Genomics UK Consortium |
| EPI_ISL_442616 | Department of Pathology, University of Cambridge | Wellcome Sanger Institute for the COVID-19 Genomics UK Consortium |
| EPI_ISL_442617 | Department of Pathology, University of Cambridge | Wellcome Sanger Institute for the COVID-19 Genomics UK Consortium |
| EPI_ISL_442618 | Department of Pathology, University of Cambridge | Wellcome Sanger Institute for the COVID-19 Genomics UK Consortium |
| EPI_ISL_442619 | Department of Pathology, University of Cambridge | Wellcome Sanger Institute for the COVID-19 Genomics UK Consortium |
| EPI_ISL_442620 | Department of Pathology, University of Cambridge | Wellcome Sanger Institute for the COVID-19 Genomics UK Consortium |
| EPI_ISL_442621 | Department of Pathology, University of Cambridge | Wellcome Sanger Institute for the COVID-19 Genomics UK Consortium |
| EPI_ISL_442622 | Department of Pathology, University of Cambridge | Wellcome Sanger Institute for the COVID-19 Genomics UK Consortium |
| EPI_ISL_442623 | Department of Pathology, University of Cambridge | Wellcome Sanger Institute for the COVID-19 Genomics UK Consortium |
| EPI_ISL_442624 | PHE South West Regional Laboratory, National Infection Service | Wellcome Sanger Institute for the COVID-19 Genomics UK Consortium |
| EPI_ISL_442625 | PHE South West Regional Laboratory, National Infection Service | Wellcome Sanger Institute for the COVID-19 Genomics UK Consortium |
| EPI_ISL_442626 | PHE South West Regional Laboratory, National Infection Service | Wellcome Sanger Institute for the COVID-19 Genomics UK Consortium |
| EPI_ISL_442627 | PHE South West Regional Laboratory, National Infection Service | Wellcome Sanger Institute for the COVID-19 Genomics UK Consortium |
| EPI_ISL_442628 | PHE South West Regional Laboratory, National Infection Service | Wellcome Sanger Institute for the COVID-19 Genomics UK Consortium |
| EPI_ISL_442629 | PHE South West Regional Laboratory, National Infection Service | Wellcome Sanger Institute for the COVID-19 Genomics UK Consortium |

| EPI_Accession_ID | Originating Laboratory | Submitting Laboratory |
|---|---|---|
| EPI_ISL_442630 | PHE South West Regional Laboratory, National Infection Service | Wellcome Sanger Institute for the COVID-19 Genomics UK Consortium |
| EPI_ISL_442631 | PHE South West Regional Laboratory, National Infection Service | Wellcome Sanger Institute for the COVID-19 Genomics UK Consortium |
| EPI_ISL_442632 | PHE South West Regional Laboratory, National Infection Service | Wellcome Sanger Institute for the COVID-19 Genomics UK Consortium |
| EPI_ISL_442633 | PHE South West Regional Laboratory, National Infection Service | Wellcome Sanger Institute for the COVID-19 Genomics UK Consortium |
| EPI_ISL_442634 | PHE South West Regional Laboratory, National Infection Service | Wellcome Sanger Institute for the COVID-19 Genomics UK Consortium |
| EPI_ISL_442635 | PHE South West Regional Laboratory, National Infection Service | Wellcome Sanger Institute for the COVID-19 Genomics UK Consortium |
| EPI_ISL_442636 | PHE South West Regional Laboratory, National Infection Service | Wellcome Sanger Institute for the COVID-19 Genomics UK Consortium |
| EPI_ISL_442637 | PHE South West Regional Laboratory, National Infection Service | Wellcome Sanger Institute for the COVID-19 Genomics UK Consortium |
| EPI_ISL_442638 | PHE South West Regional Laboratory, National Infection Service | Wellcome Sanger Institute for the COVID-19 Genomics UK Consortium |
| EPI_ISL_442639 | PHE South West Regional Laboratory, National Infection Service | Wellcome Sanger Institute for the COVID-19 Genomics UK Consortium |
| EPI_ISL_442640 | PHE South West Regional Laboratory, National Infection Service | Wellcome Sanger Institute for the COVID-19 Genomics UK Consortium |
| EPI_ISL_442641 | PHE South West Regional Laboratory, National Infection Service | Wellcome Sanger Institute for the COVID-19 Genomics UK Consortium |
| EPI_ISL_442642 | PHE South West Regional Laboratory, National Infection Service | Wellcome Sanger Institute for the COVID-19 Genomics UK Consortium |
| EPI_ISL_442643 | PHE South West Regional Laboratory, National Infection Service | Wellcome Sanger Institute for the COVID-19 Genomics UK Consortium |
| EPI_ISL_442644 | PHE South West Regional Laboratory, National Infection Service | Wellcome Sanger Institute for the COVID-19 Genomics UK Consortium |
| EPI_ISL_442645 | PHE South West Regional Laboratory, National Infection Service | Wellcome Sanger Institute for the COVID-19 Genomics UK Consortium |

| EPI_Accession_ID | Originating Laboratory | Submitting Laboratory |
|---|---|---|
| EPI_ISL_442646 | PHE South West Regional Laboratory, National Infection Service | Wellcome Sanger Institute for the COVID-19 Genomics UK Consortium |
| EPI_ISL_442647 | PHE South West Regional Laboratory, National Infection Service | Wellcome Sanger Institute for the COVID-19 Genomics UK Consortium |
| EPI_ISL_442648 | PHE South West Regional Laboratory, National Infection Service | Wellcome Sanger Institute for the COVID-19 Genomics UK Consortium |
| EPI_ISL_442649 | PHE South West Regional Laboratory, National Infection Service | Wellcome Sanger Institute for the COVID-19 Genomics UK Consortium |
| EPI_ISL_442650 | PHE South West Regional Laboratory, National Infection Service | Wellcome Sanger Institute for the COVID-19 Genomics UK Consortium |
| EPI_ISL_442651 | PHE South West Regional Laboratory, National Infection Service | Wellcome Sanger Institute for the COVID-19 Genomics UK Consortium |
| EPI_ISL_442652 | PHE South West Regional Laboratory, National Infection Service | Wellcome Sanger Institute for the COVID-19 Genomics UK Consortium |
| EPI_ISL_442653 | PHE South West Regional Laboratory, National Infection Service | Wellcome Sanger Institute for the COVID-19 Genomics UK Consortium |
| EPI_ISL_442654 | PHE South West Regional Laboratory, National Infection Service | Wellcome Sanger Institute for the COVID-19 Genomics UK Consortium |
| EPI_ISL_442655 | PHE South West Regional Laboratory, National Infection Service | Wellcome Sanger Institute for the COVID-19 Genomics UK Consortium |
| EPI_ISL_442656 | PHE South West Regional Laboratory, National Infection Service | Wellcome Sanger Institute for the COVID-19 Genomics UK Consortium |
| EPI_ISL_442657 | PHE South West Regional Laboratory, National Infection Service | Wellcome Sanger Institute for the COVID-19 Genomics UK Consortium |
| EPI_ISL_442658 | PHE South West Regional Laboratory, National Infection Service | Wellcome Sanger Institute for the COVID-19 Genomics UK Consortium |
| EPI_ISL_442659 | PHE South West Regional Laboratory, National Infection Service | Wellcome Sanger Institute for the COVID-19 Genomics UK Consortium |
| EPI_ISL_442660 | PHE South West Regional Laboratory, National Infection Service | Wellcome Sanger Institute for the COVID-19 Genomics UK Consortium |
| EPI_ISL_442661 | PHE South West Regional Laboratory, National Infection Service | Wellcome Sanger Institute for the COVID-19 Genomics UK Consortium |

| EPI_Accession_ID | Originating Laboratory | Submitting Laboratory |
|---|---|---|
| EPI_ISL_442662 | PHE South West Regional Laboratory, National Infection Service | Wellcome Sanger Institute for the COVID-19 Genomics UK Consortium |
| EPI_ISL_442663 | PHE South West Regional Laboratory, National Infection Service | Wellcome Sanger Institute for the COVID-19 Genomics UK Consortium |
| EPI_ISL_442664 | PHE South West Regional Laboratory, National Infection Service | Wellcome Sanger Institute for the COVID-19 Genomics UK Consortium |
| EPI_ISL_442665 | PHE South West Regional Laboratory, National Infection Service | Wellcome Sanger Institute for the COVID-19 Genomics UK Consortium |
| EPI_ISL_442666 | PHE South West Regional Laboratory, National Infection Service | Wellcome Sanger Institute for the COVID-19 Genomics UK Consortium |
| EPI_ISL_442667 | PHE South West Regional Laboratory, National Infection Service | Wellcome Sanger Institute for the COVID-19 Genomics UK Consortium |
| EPI_ISL_442668 | PHE South West Regional Laboratory, National Infection Service | Wellcome Sanger Institute for the COVID-19 Genomics UK Consortium |
| EPI_ISL_442669 | PHE South West Regional Laboratory, National Infection Service | Wellcome Sanger Institute for the COVID-19 Genomics UK Consortium |
| EPI_ISL_442670 | PHE South West Regional Laboratory, National Infection Service | Wellcome Sanger Institute for the COVID-19 Genomics UK Consortium |
| EPI_ISL_442671 | PHE South West Regional Laboratory, National Infection Service | Wellcome Sanger Institute for the COVID-19 Genomics UK Consortium |
| EPI_ISL_442672 | PHE South West Regional Laboratory, National Infection Service | Wellcome Sanger Institute for the COVID-19 Genomics UK Consortium |
| EPI_ISL_442673 | PHE South West Regional Laboratory, National Infection Service | Wellcome Sanger Institute for the COVID-19 Genomics UK Consortium |
| EPI_ISL_442674 | PHE South West Regional Laboratory, National Infection Service | Wellcome Sanger Institute for the COVID-19 Genomics UK Consortium |
| EPI_ISL_442675 | PHE South West Regional Laboratory, National Infection Service | Wellcome Sanger Institute for the COVID-19 Genomics UK Consortium |
| EPI_ISL_442676 | PHE South West Regional Laboratory, National Infection Service | Wellcome Sanger Institute for the COVID-19 Genomics UK Consortium |
| EPI_ISL_442677 | PHE South West Regional Laboratory, National Infection Service | Wellcome Sanger Institute for the COVID-19 Genomics UK Consortium |

| EPI_Accession_ID | Originating Laboratory | Submitting Laboratory |
| --- | --- | --- |
| EPI_ISL_442678 | PHE South West Regional Laboratory, National Infection Service | Wellcome Sanger Institute for the COVID-19 Genomics UK Consortium |
| EPI_ISL_442679 | PHE South West Regional Laboratory, National Infection Service | Wellcome Sanger Institute for the COVID-19 Genomics UK Consortium |
| EPI_ISL_442680 | PHE South West Regional Laboratory, National Infection Service | Wellcome Sanger Institute for the COVID-19 Genomics UK Consortium |
| EPI_ISL_442681 | PHE South West Regional Laboratory, National Infection Service | Wellcome Sanger Institute for the COVID-19 Genomics UK Consortium |
| EPI_ISL_442682 | PHE South West Regional Laboratory, National Infection Service | Wellcome Sanger Institute for the COVID-19 Genomics UK Consortium |
| EPI_ISL_442683 | PHE South West Regional Laboratory, National Infection Service | Wellcome Sanger Institute for the COVID-19 Genomics UK Consortium |
| EPI_ISL_442684 | PHE South West Regional Laboratory, National Infection Service | Wellcome Sanger Institute for the COVID-19 Genomics UK Consortium |
| EPI_ISL_442685 | PHE South West Regional Laboratory, National Infection Service | Wellcome Sanger Institute for the COVID-19 Genomics UK Consortium |
| EPI_ISL_442686 | PHE South West Regional Laboratory, National Infection Service | Wellcome Sanger Institute for the COVID-19 Genomics UK Consortium |
| EPI_ISL_442687 | PHE South West Regional Laboratory, National Infection Service | Wellcome Sanger Institute for the COVID-19 Genomics UK Consortium |
| EPI_ISL_442688 | PHE South West Regional Laboratory, National Infection Service | Wellcome Sanger Institute for the COVID-19 Genomics UK Consortium |
| EPI_ISL_442689 | PHE South West Regional Laboratory, National Infection Service | Wellcome Sanger Institute for the COVID-19 Genomics UK Consortium |
| EPI_ISL_442690 | PHE South West Regional Laboratory, National Infection Service | Wellcome Sanger Institute for the COVID-19 Genomics UK Consortium |
| EPI_ISL_442691 | PHE South West Regional Laboratory, National Infection Service | Wellcome Sanger Institute for the COVID-19 Genomics UK Consortium |
| EPI_ISL_442692 | PHE South West Regional Laboratory, National Infection Service | Wellcome Sanger Institute for the COVID-19 Genomics UK Consortium |
| EPI_ISL_442693 | PHE South West Regional Laboratory, National Infection Service | Wellcome Sanger Institute for the COVID-19 Genomics UK Consortium |

| EPI_Accession_ID | Originating Laboratory | Submitting Laboratory |
| --- | --- | --- |
| EPI_ISL_442694 | PHE South West Regional Laboratory, National Infection Service | Wellcome Sanger Institute for the COVID-19 Genomics UK Consortium |
| EPI_ISL_442695 | PHE South West Regional Laboratory, National Infection Service | Wellcome Sanger Institute for the COVID-19 Genomics UK Consortium |
| EPI_ISL_442696 | PHE South West Regional Laboratory, National Infection Service | Wellcome Sanger Institute for the COVID-19 Genomics UK Consortium |
| EPI_ISL_442697 | PHE South West Regional Laboratory, National Infection Service | Wellcome Sanger Institute for the COVID-19 Genomics UK Consortium |
| EPI_ISL_442698 | PHE South West Regional Laboratory, National Infection Service | Wellcome Sanger Institute for the COVID-19 Genomics UK Consortium |
| EPI_ISL_442699 | PHE South West Regional Laboratory, National Infection Service | Wellcome Sanger Institute for the COVID-19 Genomics UK Consortium |
| EPI_ISL_442700 | PHE South West Regional Laboratory, National Infection Service | Wellcome Sanger Institute for the COVID-19 Genomics UK Consortium |
| EPI_ISL_442701 | PHE South West Regional Laboratory, National Infection Service | Wellcome Sanger Institute for the COVID-19 Genomics UK Consortium |
| EPI_ISL_442702 | PHE South West Regional Laboratory, National Infection Service | Wellcome Sanger Institute for the COVID-19 Genomics UK Consortium |
| EPI_ISL_442703 | PHE South West Regional Laboratory, National Infection Service | Wellcome Sanger Institute for the COVID-19 Genomics UK Consortium |
| EPI_ISL_442704 | PHE South West Regional Laboratory, National Infection Service | Wellcome Sanger Institute for the COVID-19 Genomics UK Consortium |
| EPI_ISL_442705 | PHE South West Regional Laboratory, National Infection Service | Wellcome Sanger Institute for the COVID-19 Genomics UK Consortium |
| EPI_ISL_442706 | PHE South West Regional Laboratory, National Infection Service | Wellcome Sanger Institute for the COVID-19 Genomics UK Consortium |
| EPI_ISL_442707 | PHE South West Regional Laboratory, National Infection Service | Wellcome Sanger Institute for the COVID-19 Genomics UK Consortium |
| EPI_ISL_442708 | PHE South West Regional Laboratory, National Infection Service | Wellcome Sanger Institute for the COVID-19 Genomics UK Consortium |
| EPI_ISL_442709 | PHE South West Regional Laboratory, National Infection Service | Wellcome Sanger Institute for the COVID-19 Genomics UK Consortium |

| EPI_Accession_ID | Originating Laboratory | Submitting Laboratory |
|---|---|---|
| EPI_ISL_442710 | PHE South West Regional Laboratory, National Infection Service | Wellcome Sanger Institute for the COVID-19 Genomics UK Consortium |
| EPI_ISL_442711 | PHE South West Regional Laboratory, National Infection Service | Wellcome Sanger Institute for the COVID-19 Genomics UK Consortium |
| EPI_ISL_442712 | PHE South West Regional Laboratory, National Infection Service | Wellcome Sanger Institute for the COVID-19 Genomics UK Consortium |
| EPI_ISL_442713 | PHE South West Regional Laboratory, National Infection Service | Wellcome Sanger Institute for the COVID-19 Genomics UK Consortium |
| EPI_ISL_442714 | PHE South West Regional Laboratory, National Infection Service | Wellcome Sanger Institute for the COVID-19 Genomics UK Consortium |
| EPI_ISL_442715 | PHE South West Regional Laboratory, National Infection Service | Wellcome Sanger Institute for the COVID-19 Genomics UK Consortium |
| EPI_ISL_442716 | PHE South West Regional Laboratory, National Infection Service | Wellcome Sanger Institute for the COVID-19 Genomics UK Consortium |
| EPI_ISL_442717 | PHE South West Regional Laboratory, National Infection Service | Wellcome Sanger Institute for the COVID-19 Genomics UK Consortium |
| EPI_ISL_442718 | PHE South West Regional Laboratory, National Infection Service | Wellcome Sanger Institute for the COVID-19 Genomics UK Consortium |
| EPI_ISL_442719 | PHE South West Regional Laboratory, National Infection Service | Wellcome Sanger Institute for the COVID-19 Genomics UK Consortium |
| EPI_ISL_442720 | PHE South West Regional Laboratory, National Infection Service | Wellcome Sanger Institute for the COVID-19 Genomics UK Consortium |
| EPI_ISL_442721 | PHE South West Regional Laboratory, National Infection Service | Wellcome Sanger Institute for the COVID-19 Genomics UK Consortium |
| EPI_ISL_442722 | PHE South West Regional Laboratory, National Infection Service | Wellcome Sanger Institute for the COVID-19 Genomics UK Consortium |
| EPI_ISL_442723 | PHE South West Regional Laboratory, National Infection Service | Wellcome Sanger Institute for the COVID-19 Genomics UK Consortium |
| EPI_ISL_442724 | PHE South West Regional Laboratory, National Infection Service | Wellcome Sanger Institute for the COVID-19 Genomics UK Consortium |
| EPI_ISL_442725 | PHE South West Regional Laboratory, National Infection Service | Wellcome Sanger Institute for the COVID-19 Genomics UK Consortium |

| EPI_Accession_ID | Originating Laboratory | Submitting Laboratory |
|---|---|---|
| EPI_ISL_442726 | PHE South West Regional Laboratory, National Infection Service | Wellcome Sanger Institute for the COVID-19 Genomics UK Consortium |
| EPI_ISL_442727 | PHE South West Regional Laboratory, National Infection Service | Wellcome Sanger Institute for the COVID-19 Genomics UK Consortium |
| EPI_ISL_442728 | PHE South West Regional Laboratory, National Infection Service | Wellcome Sanger Institute for the COVID-19 Genomics UK Consortium |
| EPI_ISL_442729 | PHE South West Regional Laboratory, National Infection Service | Wellcome Sanger Institute for the COVID-19 Genomics UK Consortium |
| EPI_ISL_442730 | PHE South West Regional Laboratory, National Infection Service | Wellcome Sanger Institute for the COVID-19 Genomics UK Consortium |
| EPI_ISL_442731 | PHE South West Regional Laboratory, National Infection Service | Wellcome Sanger Institute for the COVID-19 Genomics UK Consortium |
| EPI_ISL_442732 | PHE South West Regional Laboratory, National Infection Service | Wellcome Sanger Institute for the COVID-19 Genomics UK Consortium |
| EPI_ISL_442733 | PHE South West Regional Laboratory, National Infection Service | Wellcome Sanger Institute for the COVID-19 Genomics UK Consortium |
| EPI_ISL_442734 | PHE South West Regional Laboratory, National Infection Service | Wellcome Sanger Institute for the COVID-19 Genomics UK Consortium |
| EPI_ISL_442735 | PHE South West Regional Laboratory, National Infection Service | Wellcome Sanger Institute for the COVID-19 Genomics UK Consortium |
| EPI_ISL_442736 | PHE South West Regional Laboratory, National Infection Service | Wellcome Sanger Institute for the COVID-19 Genomics UK Consortium |
| EPI_ISL_442737 | PHE South West Regional Laboratory, National Infection Service | Wellcome Sanger Institute for the COVID-19 Genomics UK Consortium |
| EPI_ISL_442738 | PHE South West Regional Laboratory, National Infection Service | Wellcome Sanger Institute for the COVID-19 Genomics UK Consortium |
| EPI_ISL_442739 | PHE South West Regional Laboratory, National Infection Service | Wellcome Sanger Institute for the COVID-19 Genomics UK Consortium |
| EPI_ISL_442740 | PHE South West Regional Laboratory, National Infection Service | Wellcome Sanger Institute for the COVID-19 Genomics UK Consortium |
| EPI_ISL_442741 | PHE South West Regional Laboratory, National Infection Service | Wellcome Sanger Institute for the COVID-19 Genomics UK Consortium |

| EPI_Accession_ID | Originating Laboratory | Submitting Laboratory |
|---|---|---|
| EPI_ISL_442742 | PHE South West Regional Laboratory, National Infection Service | Wellcome Sanger Institute for the COVID-19 Genomics UK Consortium |
| EPI_ISL_442743 | PHE South West Regional Laboratory, National Infection Service | Wellcome Sanger Institute for the COVID-19 Genomics UK Consortium |
| EPI_ISL_442744 | PHE South West Regional Laboratory, National Infection Service | Wellcome Sanger Institute for the COVID-19 Genomics UK Consortium |
| EPI_ISL_442745 | PHE South West Regional Laboratory, National Infection Service | Wellcome Sanger Institute for the COVID-19 Genomics UK Consortium |
| EPI_ISL_442746 | PHE South West Regional Laboratory, National Infection Service | Wellcome Sanger Institute for the COVID-19 Genomics UK Consortium |
| EPI_ISL_442747 | PHE South West Regional Laboratory, National Infection Service | Wellcome Sanger Institute for the COVID-19 Genomics UK Consortium |
| EPI_ISL_442748 | PHE South West Regional Laboratory, National Infection Service | Wellcome Sanger Institute for the COVID-19 Genomics UK Consortium |
| EPI_ISL_442749 | PHE South West Regional Laboratory, National Infection Service | Wellcome Sanger Institute for the COVID-19 Genomics UK Consortium |
| EPI_ISL_442750 | PHE South West Regional Laboratory, National Infection Service | Wellcome Sanger Institute for the COVID-19 Genomics UK Consortium |
| EPI_ISL_442751 | PHE South West Regional Laboratory, National Infection Service | Wellcome Sanger Institute for the COVID-19 Genomics UK Consortium |
| EPI_ISL_442752 | PHE South West Regional Laboratory, National Infection Service | Wellcome Sanger Institute for the COVID-19 Genomics UK Consortium |
| EPI_ISL_442753 | PHE South West Regional Laboratory, National Infection Service | Wellcome Sanger Institute for the COVID-19 Genomics UK Consortium |
| EPI_ISL_442754 | PHE South West Regional Laboratory, National Infection Service | Wellcome Sanger Institute for the COVID-19 Genomics UK Consortium |
| EPI_ISL_442755 | PHE South West Regional Laboratory, National Infection Service | Wellcome Sanger Institute for the COVID-19 Genomics UK Consortium |
| EPI_ISL_442756 | PHE South West Regional Laboratory, National Infection Service | Wellcome Sanger Institute for the COVID-19 Genomics UK Consortium |
| EPI_ISL_442757 | PHE South West Regional Laboratory, National Infection Service | Wellcome Sanger Institute for the COVID-19 Genomics UK Consortium |

| EPI_Accession_ID | Originating Laboratory | Submitting Laboratory |
| --- | --- | --- |
| EPI_ISL_442758 | PHE South West Regional Laboratory, National Infection Service | Wellcome Sanger Institute for the COVID-19 Genomics UK Consortium |
| EPI_ISL_442759 | PHE South West Regional Laboratory, National Infection Service | Wellcome Sanger Institute for the COVID-19 Genomics UK Consortium |
| EPI_ISL_442760 | PHE South West Regional Laboratory, National Infection Service | Wellcome Sanger Institute for the COVID-19 Genomics UK Consortium |
| EPI_ISL_442761 | PHE South West Regional Laboratory, National Infection Service | Wellcome Sanger Institute for the COVID-19 Genomics UK Consortium |
| EPI_ISL_442762 | PHE South West Regional Laboratory, National Infection Service | Wellcome Sanger Institute for the COVID-19 Genomics UK Consortium |
| EPI_ISL_442763 | PHE South West Regional Laboratory, National Infection Service | Wellcome Sanger Institute for the COVID-19 Genomics UK Consortium |
| EPI_ISL_442764 | PHE South West Regional Laboratory, National Infection Service | Wellcome Sanger Institute for the COVID-19 Genomics UK Consortium |
| EPI_ISL_442765 | PHE South West Regional Laboratory, National Infection Service | Wellcome Sanger Institute for the COVID-19 Genomics UK Consortium |
| EPI_ISL_442766 | PHE South West Regional Laboratory, National Infection Service | Wellcome Sanger Institute for the COVID-19 Genomics UK Consortium |
| EPI_ISL_442767 | PHE South West Regional Laboratory, National Infection Service | Wellcome Sanger Institute for the COVID-19 Genomics UK Consortium |
| EPI_ISL_442768 | PHE South West Regional Laboratory, National Infection Service | Wellcome Sanger Institute for the COVID-19 Genomics UK Consortium |
| EPI_ISL_442769 | PHE South West Regional Laboratory, National Infection Service | Wellcome Sanger Institute for the COVID-19 Genomics UK Consortium |
| EPI_ISL_442770 | PHE South West Regional Laboratory, National Infection Service | Wellcome Sanger Institute for the COVID-19 Genomics UK Consortium |
| EPI_ISL_442771 | PHE South West Regional Laboratory, National Infection Service | Wellcome Sanger Institute for the COVID-19 Genomics UK Consortium |
| EPI_ISL_442772 | PHE South West Regional Laboratory, National Infection Service | Wellcome Sanger Institute for the COVID-19 Genomics UK Consortium |
| EPI_ISL_442773 | PHE South West Regional Laboratory, National Infection Service | Wellcome Sanger Institute for the COVID-19 Genomics UK Consortium |

| EPI_Accession_ID | Originating Laboratory | Submitting Laboratory |
| --- | --- | --- |
| EPI_ISL_442774 | PHE South West Regional Laboratory, National Infection Service | Wellcome Sanger Institute for the COVID-19 Genomics UK Consortium |
| EPI_ISL_442775 | PHE South West Regional Laboratory, National Infection Service | Wellcome Sanger Institute for the COVID-19 Genomics UK Consortium |
| EPI_ISL_442776 | PHE South West Regional Laboratory, National Infection Service | Wellcome Sanger Institute for the COVID-19 Genomics UK Consortium |
| EPI_ISL_442777 | PHE South West Regional Laboratory, National Infection Service | Wellcome Sanger Institute for the COVID-19 Genomics UK Consortium |
| EPI_ISL_442778 | PHE South West Regional Laboratory, National Infection Service | Wellcome Sanger Institute for the COVID-19 Genomics UK Consortium |
| EPI_ISL_442779 | PHE South West Regional Laboratory, National Infection Service | Wellcome Sanger Institute for the COVID-19 Genomics UK Consortium |
| EPI_ISL_442780 | PHE South West Regional Laboratory, National Infection Service | Wellcome Sanger Institute for the COVID-19 Genomics UK Consortium |
| EPI_ISL_442781 | PHE South West Regional Laboratory, National Infection Service | Wellcome Sanger Institute for the COVID-19 Genomics UK Consortium |
| EPI_ISL_442782 | PHE South West Regional Laboratory, National Infection Service | Wellcome Sanger Institute for the COVID-19 Genomics UK Consortium |
| EPI_ISL_442783 | PHE South West Regional Laboratory, National Infection Service | Wellcome Sanger Institute for the COVID-19 Genomics UK Consortium |
| EPI_ISL_442784 | PHE South West Regional Laboratory, National Infection Service | Wellcome Sanger Institute for the COVID-19 Genomics UK Consortium |
| EPI_ISL_442785 | PHE South West Regional Laboratory, National Infection Service | Wellcome Sanger Institute for the COVID-19 Genomics UK Consortium |
| EPI_ISL_442786 | PHE South West Regional Laboratory, National Infection Service | Wellcome Sanger Institute for the COVID-19 Genomics UK Consortium |
| EPI_ISL_442787 | PHE South West Regional Laboratory, National Infection Service | Wellcome Sanger Institute for the COVID-19 Genomics UK Consortium |
| EPI_ISL_442788 | PHE South West Regional Laboratory, National Infection Service | Wellcome Sanger Institute for the COVID-19 Genomics UK Consortium |
| EPI_ISL_442789 | PHE South West Regional Laboratory, National Infection Service | Wellcome Sanger Institute for the COVID-19 Genomics UK Consortium |

| EPI_Accession_ID | Originating Laboratory | Submitting Laboratory |
|---|---|---|
| EPI_ISL_442790 | PHE South West Regional Laboratory, National Infection Service | Wellcome Sanger Institute for the COVID-19 Genomics UK Consortium |
| EPI_ISL_442791 | PHE South West Regional Laboratory, National Infection Service | Wellcome Sanger Institute for the COVID-19 Genomics UK Consortium |
| EPI_ISL_442792 | PHE South West Regional Laboratory, National Infection Service | Wellcome Sanger Institute for the COVID-19 Genomics UK Consortium |
| EPI_ISL_442793 | PHE South West Regional Laboratory, National Infection Service | Wellcome Sanger Institute for the COVID-19 Genomics UK Consortium |
| EPI_ISL_442794 | PHE South West Regional Laboratory, National Infection Service | Wellcome Sanger Institute for the COVID-19 Genomics UK Consortium |
| EPI_ISL_442795 | PHE South West Regional Laboratory, National Infection Service | Wellcome Sanger Institute for the COVID-19 Genomics UK Consortium |
| EPI_ISL_442796 | PHE South West Regional Laboratory, National Infection Service | Wellcome Sanger Institute for the COVID-19 Genomics UK Consortium |
| EPI_ISL_442797 | PHE South West Regional Laboratory, National Infection Service | Wellcome Sanger Institute for the COVID-19 Genomics UK Consortium |
| EPI_ISL_442798 | PHE South West Regional Laboratory, National Infection Service | Wellcome Sanger Institute for the COVID-19 Genomics UK Consortium |
| EPI_ISL_442799 | PHE South West Regional Laboratory, National Infection Service | Wellcome Sanger Institute for the COVID-19 Genomics UK Consortium |
| EPI_ISL_442800 | PHE South West Regional Laboratory, National Infection Service | Wellcome Sanger Institute for the COVID-19 Genomics UK Consortium |
| EPI_ISL_442801 | PHE South West Regional Laboratory, National Infection Service | Wellcome Sanger Institute for the COVID-19 Genomics UK Consortium |
| EPI_ISL_442802 | PHE South West Regional Laboratory, National Infection Service | Wellcome Sanger Institute for the COVID-19 Genomics UK Consortium |
| EPI_ISL_442803 | PHE South West Regional Laboratory, National Infection Service | Wellcome Sanger Institute for the COVID-19 Genomics UK Consortium |
| EPI_ISL_442804 | PHE South West Regional Laboratory, National Infection Service | Wellcome Sanger Institute for the COVID-19 Genomics UK Consortium |
| EPI_ISL_442805 | PHE South West Regional Laboratory, National Infection Service | Wellcome Sanger Institute for the COVID-19 Genomics UK Consortium |

| EPI_Accession_ID | Originating Laboratory | Submitting Laboratory |
| --- | --- | --- |
| EPI_ISL_442806 | PHE South West Regional Laboratory, National Infection Service | Wellcome Sanger Institute for the COVID-19 Genomics UK Consortium |
| EPI_ISL_442807 | PHE South West Regional Laboratory, National Infection Service | Wellcome Sanger Institute for the COVID-19 Genomics UK Consortium |
| EPI_ISL_442808 | PHE South West Regional Laboratory, National Infection Service | Wellcome Sanger Institute for the COVID-19 Genomics UK Consortium |
| EPI_ISL_442809 | PHE South West Regional Laboratory, National Infection Service | Wellcome Sanger Institute for the COVID-19 Genomics UK Consortium |
| EPI_ISL_442810 | PHE South West Regional Laboratory, National Infection Service | Wellcome Sanger Institute for the COVID-19 Genomics UK Consortium |
| EPI_ISL_442811 | Department of Pathology, University of Cambridge | Wellcome Sanger Institute for the COVID-19 Genomics UK Consortium |
| EPI_ISL_442812 | Department of Pathology, University of Cambridge | Wellcome Sanger Institute for the COVID-19 Genomics UK Consortium |
| EPI_ISL_442813 | Department of Pathology, University of Cambridge | Wellcome Sanger Institute for the COVID-19 Genomics UK Consortium |
| EPI_ISL_442814 | Department of Pathology, University of Cambridge | Wellcome Sanger Institute for the COVID-19 Genomics UK Consortium |
| EPI_ISL_442815 | Department of Pathology, University of Cambridge | Wellcome Sanger Institute for the COVID-19 Genomics UK Consortium |
| EPI_ISL_442816 | Department of Pathology, University of Cambridge | Wellcome Sanger Institute for the COVID-19 Genomics UK Consortium |
| EPI_ISL_442817 | Department of Pathology, University of Cambridge | Wellcome Sanger Institute for the COVID-19 Genomics UK Consortium |
| EPI_ISL_442818 | Department of Pathology, University of Cambridge | Wellcome Sanger Institute for the COVID-19 Genomics UK Consortium |
| EPI_ISL_442819 | Department of Pathology, University of Cambridge | Wellcome Sanger Institute for the COVID-19 Genomics UK Consortium |
| EPI_ISL_442820 | Department of Pathology, University of Cambridge | Wellcome Sanger Institute for the COVID-19 Genomics UK Consortium |
| EPI_ISL_442821 | Department of Pathology, University of Cambridge | Wellcome Sanger Institute for the COVID-19 Genomics UK Consortium |

| EPI_Accession_ID | Originating Laboratory | Submitting Laboratory |
|---|---|---|
| EPI_ISL_442822 | Department of Pathology, University of Cambridge | Wellcome Sanger Institute for the COVID-19 Genomics UK Consortium |
| EPI_ISL_442823 | Department of Pathology, University of Cambridge | Wellcome Sanger Institute for the COVID-19 Genomics UK Consortium |
| EPI_ISL_442824 | Department of Pathology, University of Cambridge | Wellcome Sanger Institute for the COVID-19 Genomics UK Consortium |
| EPI_ISL_442825 | Department of Pathology, University of Cambridge | Wellcome Sanger Institute for the COVID-19 Genomics UK Consortium |
| EPI_ISL_442826 | Department of Pathology, University of Cambridge | Wellcome Sanger Institute for the COVID-19 Genomics UK Consortium |
| EPI_ISL_442827 | Department of Pathology, University of Cambridge | Wellcome Sanger Institute for the COVID-19 Genomics UK Consortium |
| EPI_ISL_442828 | Department of Pathology, University of Cambridge | Wellcome Sanger Institute for the COVID-19 Genomics UK Consortium |
| EPI_ISL_442829 | Department of Pathology, University of Cambridge | Wellcome Sanger Institute for the COVID-19 Genomics UK Consortium |
| EPI_ISL_442830 | Department of Pathology, University of Cambridge | Wellcome Sanger Institute for the COVID-19 Genomics UK Consortium |
| EPI_ISL_442831 | Department of Pathology, University of Cambridge | Wellcome Sanger Institute for the COVID-19 Genomics UK Consortium |
| EPI_ISL_442832 | Department of Pathology, University of Cambridge | Wellcome Sanger Institute for the COVID-19 Genomics UK Consortium |
| EPI_ISL_442833 | Department of Pathology, University of Cambridge | Wellcome Sanger Institute for the COVID-19 Genomics UK Consortium |
| EPI_ISL_442834 | Department of Pathology, University of Cambridge | Wellcome Sanger Institute for the COVID-19 Genomics UK Consortium |
| EPI_ISL_442835 | Department of Pathology, University of Cambridge | Wellcome Sanger Institute for the COVID-19 Genomics UK Consortium |
| EPI_ISL_442836 | Department of Pathology, University of Cambridge | Wellcome Sanger Institute for the COVID-19 Genomics UK Consortium |
| EPI_ISL_442837 | Department of Pathology, University of Cambridge | Wellcome Sanger Institute for the COVID-19 Genomics UK Consortium |

| EPI_Accession_ID | Originating Laboratory | Submitting Laboratory |
|---|---|---|
| EPI_ISL_442838 | Department of Pathology, University of Cambridge | Wellcome Sanger Institute for the COVID-19 Genomics UK Consortium |
| EPI_ISL_442839 | Department of Pathology, University of Cambridge | Wellcome Sanger Institute for the COVID-19 Genomics UK Consortium |
| EPI_ISL_442840 | Department of Pathology, University of Cambridge | Wellcome Sanger Institute for the COVID-19 Genomics UK Consortium |
| EPI_ISL_442841 | Department of Pathology, University of Cambridge | Wellcome Sanger Institute for the COVID-19 Genomics UK Consortium |
| EPI_ISL_442842 | Department of Pathology, University of Cambridge | Wellcome Sanger Institute for the COVID-19 Genomics UK Consortium |
| EPI_ISL_442843 | Department of Pathology, University of Cambridge | Wellcome Sanger Institute for the COVID-19 Genomics UK Consortium |
| EPI_ISL_442844 | Department of Pathology, University of Cambridge | Wellcome Sanger Institute for the COVID-19 Genomics UK Consortium |
| EPI_ISL_442845 | Department of Pathology, University of Cambridge | Wellcome Sanger Institute for the COVID-19 Genomics UK Consortium |
| EPI_ISL_442846 | Department of Pathology, University of Cambridge | Wellcome Sanger Institute for the COVID-19 Genomics UK Consortium |
| EPI_ISL_442847 | Department of Pathology, University of Cambridge | Wellcome Sanger Institute for the COVID-19 Genomics UK Consortium |
| EPI_ISL_442848 | Department of Pathology, University of Cambridge | Wellcome Sanger Institute for the COVID-19 Genomics UK Consortium |
| EPI_ISL_442849 | Department of Pathology, University of Cambridge | Wellcome Sanger Institute for the COVID-19 Genomics UK Consortium |
| EPI_ISL_442850 | Department of Pathology, University of Cambridge | Wellcome Sanger Institute for the COVID-19 Genomics UK Consortium |
| EPI_ISL_442851 | Department of Pathology, University of Cambridge | Wellcome Sanger Institute for the COVID-19 Genomics UK Consortium |
| EPI_ISL_442852 | Department of Pathology, University of Cambridge | Wellcome Sanger Institute for the COVID-19 Genomics UK Consortium |
| EPI_ISL_442853 | Department of Pathology, University of Cambridge | Wellcome Sanger Institute for the COVID-19 Genomics UK Consortium |

| EPI_Accession_ID | Originating Laboratory | Submitting Laboratory |
|---|---|---|
| EPI_ISL_442854 | Department of Pathology, University of Cambridge | Wellcome Sanger Institute for the COVID-19 Genomics UK Consortium |
| EPI_ISL_442855 | Department of Pathology, University of Cambridge | Wellcome Sanger Institute for the COVID-19 Genomics UK Consortium |
| EPI_ISL_442856 | Department of Pathology, University of Cambridge | Wellcome Sanger Institute for the COVID-19 Genomics UK Consortium |
| EPI_ISL_442857 | Department of Pathology, University of Cambridge | Wellcome Sanger Institute for the COVID-19 Genomics UK Consortium |
| EPI_ISL_442858 | Department of Pathology, University of Cambridge | Wellcome Sanger Institute for the COVID-19 Genomics UK Consortium |
| EPI_ISL_442859 | Department of Pathology, University of Cambridge | Wellcome Sanger Institute for the COVID-19 Genomics UK Consortium |
| EPI_ISL_442860 | Department of Pathology, University of Cambridge | Wellcome Sanger Institute for the COVID-19 Genomics UK Consortium |
| EPI_ISL_442861 | Department of Pathology, University of Cambridge | Wellcome Sanger Institute for the COVID-19 Genomics UK Consortium |
| EPI_ISL_442862 | Department of Pathology, University of Cambridge | Wellcome Sanger Institute for the COVID-19 Genomics UK Consortium |
| EPI_ISL_442863 | Department of Pathology, University of Cambridge | Wellcome Sanger Institute for the COVID-19 Genomics UK Consortium |
| EPI_ISL_442864 | Department of Pathology, University of Cambridge | Wellcome Sanger Institute for the COVID-19 Genomics UK Consortium |
| EPI_ISL_442865 | Department of Pathology, University of Cambridge | Wellcome Sanger Institute for the COVID-19 Genomics UK Consortium |
| EPI_ISL_442866 | Department of Pathology, University of Cambridge | Wellcome Sanger Institute for the COVID-19 Genomics UK Consortium |
| EPI_ISL_442867 | Department of Pathology, University of Cambridge | Wellcome Sanger Institute for the COVID-19 Genomics UK Consortium |
| EPI_ISL_442868 | Department of Pathology, University of Cambridge | Wellcome Sanger Institute for the COVID-19 Genomics UK Consortium |
| EPI_ISL_442869 | Department of Pathology, University of Cambridge | Wellcome Sanger Institute for the COVID-19 Genomics UK Consortium |

| EPI_Accession_ID | Originating Laboratory | Submitting Laboratory |
|---|---|---|
| EPI_ISL_442870 | Department of Pathology, University of Cambridge | Wellcome Sanger Institute for the COVID-19 Genomics UK Consortium |
| EPI_ISL_442871 | Department of Pathology, University of Cambridge | Wellcome Sanger Institute for the COVID-19 Genomics UK Consortium |
| EPI_ISL_442872 | Department of Pathology, University of Cambridge | Wellcome Sanger Institute for the COVID-19 Genomics UK Consortium |
| EPI_ISL_442873 | Department of Pathology, University of Cambridge | Wellcome Sanger Institute for the COVID-19 Genomics UK Consortium |
| EPI_ISL_442874 | Department of Pathology, University of Cambridge | Wellcome Sanger Institute for the COVID-19 Genomics UK Consortium |
| EPI_ISL_442875 | Department of Pathology, University of Cambridge | Wellcome Sanger Institute for the COVID-19 Genomics UK Consortium |
| EPI_ISL_442876 | Department of Pathology, University of Cambridge | Wellcome Sanger Institute for the COVID-19 Genomics UK Consortium |
| EPI_ISL_442877 | Department of Pathology, University of Cambridge | Wellcome Sanger Institute for the COVID-19 Genomics UK Consortium |
| EPI_ISL_442878 | Department of Pathology, University of Cambridge | Wellcome Sanger Institute for the COVID-19 Genomics UK Consortium |
| EPI_ISL_442879 | Department of Pathology, University of Cambridge | Wellcome Sanger Institute for the COVID-19 Genomics UK Consortium |
| EPI_ISL_442880 | Department of Pathology, University of Cambridge | Wellcome Sanger Institute for the COVID-19 Genomics UK Consortium |
| EPI_ISL_442881 | Department of Pathology, University of Cambridge | Wellcome Sanger Institute for the COVID-19 Genomics UK Consortium |
| EPI_ISL_442882 | Department of Pathology, University of Cambridge | Wellcome Sanger Institute for the COVID-19 Genomics UK Consortium |
| EPI_ISL_442883 | Department of Pathology, University of Cambridge | Wellcome Sanger Institute for the COVID-19 Genomics UK Consortium |
| EPI_ISL_442884 | Department of Pathology, University of Cambridge | Wellcome Sanger Institute for the COVID-19 Genomics UK Consortium |
| EPI_ISL_442885 | Department of Pathology, University of Cambridge | Wellcome Sanger Institute for the COVID-19 Genomics UK Consortium |

| EPI_Accession_ID | Originating Laboratory | Submitting Laboratory |
|---|---|---|
| EPI_ISL_442886 | Department of Pathology, University of Cambridge | Wellcome Sanger Institute for the COVID-19 Genomics UK Consortium |
| EPI_ISL_442887 | Department of Pathology, University of Cambridge | Wellcome Sanger Institute for the COVID-19 Genomics UK Consortium |
| EPI_ISL_442888 | Department of Pathology, University of Cambridge | Wellcome Sanger Institute for the COVID-19 Genomics UK Consortium |
| EPI_ISL_442889 | Department of Pathology, University of Cambridge | Wellcome Sanger Institute for the COVID-19 Genomics UK Consortium |
| EPI_ISL_442890 | Department of Pathology, University of Cambridge | Wellcome Sanger Institute for the COVID-19 Genomics UK Consortium |
| EPI_ISL_442891 | Department of Pathology, University of Cambridge | Wellcome Sanger Institute for the COVID-19 Genomics UK Consortium |
| EPI_ISL_442892 | Department of Pathology, University of Cambridge | Wellcome Sanger Institute for the COVID-19 Genomics UK Consortium |
| EPI_ISL_442893 | Department of Pathology, University of Cambridge | Wellcome Sanger Institute for the COVID-19 Genomics UK Consortium |
| EPI_ISL_442894 | Department of Pathology, University of Cambridge | Wellcome Sanger Institute for the COVID-19 Genomics UK Consortium |
| EPI_ISL_442895 | Department of Pathology, University of Cambridge | Wellcome Sanger Institute for the COVID-19 Genomics UK Consortium |
| EPI_ISL_442896 | Department of Pathology, University of Cambridge | Wellcome Sanger Institute for the COVID-19 Genomics UK Consortium |
| EPI_ISL_442897 | Department of Pathology, University of Cambridge | Wellcome Sanger Institute for the COVID-19 Genomics UK Consortium |
| EPI_ISL_442898 | Department of Pathology, University of Cambridge | Wellcome Sanger Institute for the COVID-19 Genomics UK Consortium |
| EPI_ISL_442899 | Department of Pathology, University of Cambridge | Wellcome Sanger Institute for the COVID-19 Genomics UK Consortium |
| EPI_ISL_442900 | Department of Pathology, University of Cambridge | Wellcome Sanger Institute for the COVID-19 Genomics UK Consortium |
| EPI_ISL_442901 | Department of Pathology, University of Cambridge | Wellcome Sanger Institute for the COVID-19 Genomics UK Consortium |

| EPI_Accession_ID | Originating Laboratory | Submitting Laboratory |
|---|---|---|
| EPI_ISL_442902 | Department of Pathology, University of Cambridge | Wellcome Sanger Institute for the COVID-19 Genomics UK Consortium |
| EPI_ISL_442903 | Department of Pathology, University of Cambridge | Wellcome Sanger Institute for the COVID-19 Genomics UK Consortium |
| EPI_ISL_442904 | Department of Pathology, University of Cambridge | Wellcome Sanger Institute for the COVID-19 Genomics UK Consortium |
| EPI_ISL_442905 | Department of Pathology, University of Cambridge | Wellcome Sanger Institute for the COVID-19 Genomics UK Consortium |
| EPI_ISL_442906 | Department of Pathology, University of Cambridge | Wellcome Sanger Institute for the COVID-19 Genomics UK Consortium |
| EPI_ISL_442907 | Department of Pathology, University of Cambridge | Wellcome Sanger Institute for the COVID-19 Genomics UK Consortium |
| EPI_ISL_442908 | Department of Pathology, University of Cambridge | Wellcome Sanger Institute for the COVID-19 Genomics UK Consortium |
| EPI_ISL_442909 | Department of Pathology, University of Cambridge | Wellcome Sanger Institute for the COVID-19 Genomics UK Consortium |
| EPI_ISL_442910 | Department of Pathology, University of Cambridge | Wellcome Sanger Institute for the COVID-19 Genomics UK Consortium |
| EPI_ISL_442911 | Department of Pathology, University of Cambridge | Wellcome Sanger Institute for the COVID-19 Genomics UK Consortium |
| EPI_ISL_442912 | Department of Pathology, University of Cambridge | Wellcome Sanger Institute for the COVID-19 Genomics UK Consortium |
| EPI_ISL_442913 | Department of Pathology, University of Cambridge | Wellcome Sanger Institute for the COVID-19 Genomics UK Consortium |
| EPI_ISL_442914 | Department of Pathology, University of Cambridge | Wellcome Sanger Institute for the COVID-19 Genomics UK Consortium |
| EPI_ISL_442915 | Department of Pathology, University of Cambridge | Wellcome Sanger Institute for the COVID-19 Genomics UK Consortium |
| EPI_ISL_442916 | Department of Pathology, University of Cambridge | Wellcome Sanger Institute for the COVID-19 Genomics UK Consortium |
| EPI_ISL_442917 | Department of Pathology, University of Cambridge | Wellcome Sanger Institute for the COVID-19 Genomics UK Consortium |

| EPI_Accession_ID | Originating Laboratory | Submitting Laboratory |
| --- | --- | --- |
| EPI_ISL_442918 | Department of Pathology, University of Cambridge | Wellcome Sanger Institute for the COVID-19 Genomics UK Consortium |
| EPI_ISL_442919 | Department of Pathology, University of Cambridge | Wellcome Sanger Institute for the COVID-19 Genomics UK Consortium |
| EPI_ISL_442920 | Department of Pathology, University of Cambridge | Wellcome Sanger Institute for the COVID-19 Genomics UK Consortium |
| EPI_ISL_442921 | Department of Pathology, University of Cambridge | Wellcome Sanger Institute for the COVID-19 Genomics UK Consortium |
| EPI_ISL_442922 | Department of Pathology, University of Cambridge | Wellcome Sanger Institute for the COVID-19 Genomics UK Consortium |
| EPI_ISL_442923 | Department of Pathology, University of Cambridge | Wellcome Sanger Institute for the COVID-19 Genomics UK Consortium |
| EPI_ISL_442924 | Department of Pathology, University of Cambridge | Wellcome Sanger Institute for the COVID-19 Genomics UK Consortium |
| EPI_ISL_442925 | Department of Pathology, University of Cambridge | Wellcome Sanger Institute for the COVID-19 Genomics UK Consortium |
| EPI_ISL_442926 | Department of Pathology, University of Cambridge | Wellcome Sanger Institute for the COVID-19 Genomics UK Consortium |
| EPI_ISL_442927 | Department of Pathology, University of Cambridge | Wellcome Sanger Institute for the COVID-19 Genomics UK Consortium |
| EPI_ISL_442928 | Department of Pathology, University of Cambridge | Wellcome Sanger Institute for the COVID-19 Genomics UK Consortium |
| EPI_ISL_442929 | Department of Pathology, University of Cambridge | Wellcome Sanger Institute for the COVID-19 Genomics UK Consortium |
| EPI_ISL_442930 | Department of Pathology, University of Cambridge | Wellcome Sanger Institute for the COVID-19 Genomics UK Consortium |
| EPI_ISL_442931 | Department of Pathology, University of Cambridge | Wellcome Sanger Institute for the COVID-19 Genomics UK Consortium |
| EPI_ISL_442932 | Department of Pathology, University of Cambridge | Wellcome Sanger Institute for the COVID-19 Genomics UK Consortium |
| EPI_ISL_442933 | Department of Pathology, University of Cambridge | Wellcome Sanger Institute for the COVID-19 Genomics UK Consortium |

| EPI_Accession_ID | Originating Laboratory | Submitting Laboratory |
|---|---|---|
| EPI_ISL_442934 | Department of Pathology, University of Cambridge | Wellcome Sanger Institute for the COVID-19 Genomics UK Consortium |
| EPI_ISL_442935 | Department of Pathology, University of Cambridge | Wellcome Sanger Institute for the COVID-19 Genomics UK Consortium |
| EPI_ISL_442936 | Department of Pathology, University of Cambridge | Wellcome Sanger Institute for the COVID-19 Genomics UK Consortium |
| EPI_ISL_442937 | Department of Pathology, University of Cambridge | Wellcome Sanger Institute for the COVID-19 Genomics UK Consortium |
| EPI_ISL_442938 | Department of Pathology, University of Cambridge | Wellcome Sanger Institute for the COVID-19 Genomics UK Consortium |
| EPI_ISL_442939 | Department of Pathology, University of Cambridge | Wellcome Sanger Institute for the COVID-19 Genomics UK Consortium |
| EPI_ISL_442940 | Department of Pathology, University of Cambridge | Wellcome Sanger Institute for the COVID-19 Genomics UK Consortium |
| EPI_ISL_442941 | Department of Pathology, University of Cambridge | Wellcome Sanger Institute for the COVID-19 Genomics UK Consortium |
| EPI_ISL_442942 | Department of Pathology, University of Cambridge | Wellcome Sanger Institute for the COVID-19 Genomics UK Consortium |
| EPI_ISL_442943 | Department of Pathology, University of Cambridge | Wellcome Sanger Institute for the COVID-19 Genomics UK Consortium |
| EPI_ISL_442944 | Department of Pathology, University of Cambridge | Wellcome Sanger Institute for the COVID-19 Genomics UK Consortium |
| EPI_ISL_442945 | Department of Pathology, University of Cambridge | Wellcome Sanger Institute for the COVID-19 Genomics UK Consortium |
| EPI_ISL_442946 | Department of Pathology, University of Cambridge | Wellcome Sanger Institute for the COVID-19 Genomics UK Consortium |
| EPI_ISL_442947 | Department of Pathology, University of Cambridge | Wellcome Sanger Institute for the COVID-19 Genomics UK Consortium |
| EPI_ISL_442948 | Department of Pathology, University of Cambridge | Wellcome Sanger Institute for the COVID-19 Genomics UK Consortium |
| EPI_ISL_442949 | Department of Pathology, University of Cambridge | Wellcome Sanger Institute for the COVID-19 Genomics UK Consortium |

| EPI_Accession_ID | Originating Laboratory | Submitting Laboratory |
|---|---|---|
| EPI_ISL_442950 | Department of Pathology, University of Cambridge | Wellcome Sanger Institute for the COVID-19 Genomics UK Consortium |
| EPI_ISL_442951 | Department of Pathology, University of Cambridge | Wellcome Sanger Institute for the COVID-19 Genomics UK Consortium |
| EPI_ISL_442952 | Department of Pathology, University of Cambridge | Wellcome Sanger Institute for the COVID-19 Genomics UK Consortium |
| EPI_ISL_442953 | Department of Pathology, University of Cambridge | Wellcome Sanger Institute for the COVID-19 Genomics UK Consortium |
| EPI_ISL_442954 | Department of Pathology, University of Cambridge | Wellcome Sanger Institute for the COVID-19 Genomics UK Consortium |
| EPI_ISL_442955 | Department of Pathology, University of Cambridge | Wellcome Sanger Institute for the COVID-19 Genomics UK Consortium |
| EPI_ISL_442956 | Department of Pathology, University of Cambridge | Wellcome Sanger Institute for the COVID-19 Genomics UK Consortium |
| EPI_ISL_442957 | Department of Pathology, University of Cambridge | Wellcome Sanger Institute for the COVID-19 Genomics UK Consortium |
| EPI_ISL_442958 | Department of Pathology, University of Cambridge | Wellcome Sanger Institute for the COVID-19 Genomics UK Consortium |
| EPI_ISL_442959 | Department of Pathology, University of Cambridge | Wellcome Sanger Institute for the COVID-19 Genomics UK Consortium |
| EPI_ISL_442960 | Department of Pathology, University of Cambridge | Wellcome Sanger Institute for the COVID-19 Genomics UK Consortium |
| EPI_ISL_442961 | Department of Pathology, University of Cambridge | Wellcome Sanger Institute for the COVID-19 Genomics UK Consortium |
| EPI_ISL_442962 | Department of Pathology, University of Cambridge | Wellcome Sanger Institute for the COVID-19 Genomics UK Consortium |
| EPI_ISL_442963 | Department of Pathology, University of Cambridge | Wellcome Sanger Institute for the COVID-19 Genomics UK Consortium |
| EPI_ISL_442964 | Department of Pathology, University of Cambridge | Wellcome Sanger Institute for the COVID-19 Genomics UK Consortium |
| EPI_ISL_442965 | Department of Pathology, University of Cambridge | Wellcome Sanger Institute for the COVID-19 Genomics UK Consortium |

| EPI_Accession_ID | Originating Laboratory | Submitting Laboratory |
|---|---|---|
| EPI_ISL_442966 | Department of Pathology, University of Cambridge | Wellcome Sanger Institute for the COVID-19 Genomics UK Consortium |
| EPI_ISL_442967 | Department of Pathology, University of Cambridge | Wellcome Sanger Institute for the COVID-19 Genomics UK Consortium |
| EPI_ISL_442968 | Department of Pathology, University of Cambridge | Wellcome Sanger Institute for the COVID-19 Genomics UK Consortium |
| EPI_ISL_442969 | Department of Pathology, University of Cambridge | Wellcome Sanger Institute for the COVID-19 Genomics UK Consortium |
| EPI_ISL_442970 | Department of Pathology, University of Cambridge | Wellcome Sanger Institute for the COVID-19 Genomics UK Consortium |
| EPI_ISL_442971 | Department of Pathology, University of Cambridge | Wellcome Sanger Institute for the COVID-19 Genomics UK Consortium |
| EPI_ISL_442972 | Department of Pathology, University of Cambridge | Wellcome Sanger Institute for the COVID-19 Genomics UK Consortium |
| EPI_ISL_442973 | Department of Pathology, University of Cambridge | Wellcome Sanger Institute for the COVID-19 Genomics UK Consortium |
| EPI_ISL_442974 | Department of Pathology, University of Cambridge | Wellcome Sanger Institute for the COVID-19 Genomics UK Consortium |
| EPI_ISL_442975 | Department of Pathology, University of Cambridge | Wellcome Sanger Institute for the COVID-19 Genomics UK Consortium |
| EPI_ISL_442976 | Department of Pathology, University of Cambridge | Wellcome Sanger Institute for the COVID-19 Genomics UK Consortium |
| EPI_ISL_442977 | Department of Pathology, University of Cambridge | Wellcome Sanger Institute for the COVID-19 Genomics UK Consortium |
| EPI_ISL_442978 | Department of Pathology, University of Cambridge | Wellcome Sanger Institute for the COVID-19 Genomics UK Consortium |
| EPI_ISL_442979 | Department of Pathology, University of Cambridge | Wellcome Sanger Institute for the COVID-19 Genomics UK Consortium |
| EPI_ISL_442980 | Department of Pathology, University of Cambridge | Wellcome Sanger Institute for the COVID-19 Genomics UK Consortium |
| EPI_ISL_442981 | Department of Pathology, University of Cambridge | Wellcome Sanger Institute for the COVID-19 Genomics UK Consortium |

| EPI_Accession_ID | Originating Laboratory | Submitting Laboratory |
|---|---|---|
| EPI_ISL_442982 | Department of Pathology, University of Cambridge | Wellcome Sanger Institute for the COVID-19 Genomics UK Consortium |
| EPI_ISL_442983 | Department of Pathology, University of Cambridge | Wellcome Sanger Institute for the COVID-19 Genomics UK Consortium |
| EPI_ISL_442984 | Department of Pathology, University of Cambridge | Wellcome Sanger Institute for the COVID-19 Genomics UK Consortium |
| EPI_ISL_442985 | Department of Pathology, University of Cambridge | Wellcome Sanger Institute for the COVID-19 Genomics UK Consortium |
| EPI_ISL_442986 | Department of Pathology, University of Cambridge | Wellcome Sanger Institute for the COVID-19 Genomics UK Consortium |
| EPI_ISL_442987 | Department of Pathology, University of Cambridge | Wellcome Sanger Institute for the COVID-19 Genomics UK Consortium |
| EPI_ISL_442988 | Department of Pathology, University of Cambridge | Wellcome Sanger Institute for the COVID-19 Genomics UK Consortium |
| EPI_ISL_442989 | Department of Pathology, University of Cambridge | Wellcome Sanger Institute for the COVID-19 Genomics UK Consortium |
| EPI_ISL_442990 | Department of Pathology, University of Cambridge | Wellcome Sanger Institute for the COVID-19 Genomics UK Consortium |
| EPI_ISL_442991 | Department of Pathology, University of Cambridge | Wellcome Sanger Institute for the COVID-19 Genomics UK Consortium |
| EPI_ISL_442992 | Department of Pathology, University of Cambridge | Wellcome Sanger Institute for the COVID-19 Genomics UK Consortium |
| EPI_ISL_442993 | Department of Pathology, University of Cambridge | Wellcome Sanger Institute for the COVID-19 Genomics UK Consortium |
| EPI_ISL_442994 | Department of Pathology, University of Cambridge | Wellcome Sanger Institute for the COVID-19 Genomics UK Consortium |
| EPI_ISL_442995 | Department of Pathology, University of Cambridge | Wellcome Sanger Institute for the COVID-19 Genomics UK Consortium |
| EPI_ISL_442996 | Department of Pathology, University of Cambridge | Wellcome Sanger Institute for the COVID-19 Genomics UK Consortium |
| EPI_ISL_442997 | Department of Pathology, University of Cambridge | Wellcome Sanger Institute for the COVID-19 Genomics UK Consortium |

| EPI_Accession_ID | Originating Laboratory | Submitting Laboratory |
|---|---|---|
| EPI_ISL_442998 | Department of Pathology, University of Cambridge | Wellcome Sanger Institute for the COVID-19 Genomics UK Consortium |
| EPI_ISL_442999 | Department of Pathology, University of Cambridge | Wellcome Sanger Institute for the COVID-19 Genomics UK Consortium |
| EPI_ISL_443000 | Department of Pathology, University of Cambridge | Wellcome Sanger Institute for the COVID-19 Genomics UK Consortium |
| EPI_ISL_443001 | Department of Pathology, University of Cambridge | Wellcome Sanger Institute for the COVID-19 Genomics UK Consortium |
| EPI_ISL_443002 | Department of Pathology, University of Cambridge | Wellcome Sanger Institute for the COVID-19 Genomics UK Consortium |
| EPI_ISL_443003 | Department of Pathology, University of Cambridge | Wellcome Sanger Institute for the COVID-19 Genomics UK Consortium |
| EPI_ISL_443004 | Department of Pathology, University of Cambridge | Wellcome Sanger Institute for the COVID-19 Genomics UK Consortium |
| EPI_ISL_443005 | Department of Pathology, University of Cambridge | Wellcome Sanger Institute for the COVID-19 Genomics UK Consortium |
| EPI_ISL_443006 | Department of Pathology, University of Cambridge | Wellcome Sanger Institute for the COVID-19 Genomics UK Consortium |
| EPI_ISL_443007 | Department of Pathology, University of Cambridge | Wellcome Sanger Institute for the COVID-19 Genomics UK Consortium |
| EPI_ISL_443008 | Department of Pathology, University of Cambridge | Wellcome Sanger Institute for the COVID-19 Genomics UK Consortium |
| EPI_ISL_443009 | Department of Pathology, University of Cambridge | Wellcome Sanger Institute for the COVID-19 Genomics UK Consortium |
| EPI_ISL_443010 | Department of Pathology, University of Cambridge | Wellcome Sanger Institute for the COVID-19 Genomics UK Consortium |
| EPI_ISL_443011 | Department of Pathology, University of Cambridge | Wellcome Sanger Institute for the COVID-19 Genomics UK Consortium |
| EPI_ISL_443012 | Department of Pathology, University of Cambridge | Wellcome Sanger Institute for the COVID-19 Genomics UK Consortium |
| EPI_ISL_443013 | Department of Pathology, University of Cambridge | Wellcome Sanger Institute for the COVID-19 Genomics UK Consortium |

| EPI_Accession_ID | Originating Laboratory | Submitting Laboratory |
|---|---|---|
| EPI_ISL_443014 | Department of Pathology, University of Cambridge | Wellcome Sanger Institute for the COVID-19 Genomics UK Consortium |
| EPI_ISL_443015 | Department of Pathology, University of Cambridge | Wellcome Sanger Institute for the COVID-19 Genomics UK Consortium |
| EPI_ISL_443016 | Department of Pathology, University of Cambridge | Wellcome Sanger Institute for the COVID-19 Genomics UK Consortium |
| EPI_ISL_443017 | Department of Pathology, University of Cambridge | Wellcome Sanger Institute for the COVID-19 Genomics UK Consortium |
| EPI_ISL_443018 | Department of Pathology, University of Cambridge | Wellcome Sanger Institute for the COVID-19 Genomics UK Consortium |
| EPI_ISL_443019 | Department of Pathology, University of Cambridge | Wellcome Sanger Institute for the COVID-19 Genomics UK Consortium |
| EPI_ISL_443020 | Department of Pathology, University of Cambridge | Wellcome Sanger Institute for the COVID-19 Genomics UK Consortium |
| EPI_ISL_443021 | Department of Pathology, University of Cambridge | Wellcome Sanger Institute for the COVID-19 Genomics UK Consortium |
| EPI_ISL_443022 | Department of Pathology, University of Cambridge | Wellcome Sanger Institute for the COVID-19 Genomics UK Consortium |
| EPI_ISL_443023 | Department of Pathology, University of Cambridge | Wellcome Sanger Institute for the COVID-19 Genomics UK Consortium |
| EPI_ISL_443024 | Department of Pathology, University of Cambridge | Wellcome Sanger Institute for the COVID-19 Genomics UK Consortium |
| EPI_ISL_443025 | Department of Pathology, University of Cambridge | Wellcome Sanger Institute for the COVID-19 Genomics UK Consortium |
| EPI_ISL_443026 | Department of Pathology, University of Cambridge | Wellcome Sanger Institute for the COVID-19 Genomics UK Consortium |
| EPI_ISL_443027 | Department of Pathology, University of Cambridge | Wellcome Sanger Institute for the COVID-19 Genomics UK Consortium |
| EPI_ISL_443028 | Department of Pathology, University of Cambridge | Wellcome Sanger Institute for the COVID-19 Genomics UK Consortium |
| EPI_ISL_443029 | Department of Pathology, University of Cambridge | Wellcome Sanger Institute for the COVID-19 Genomics UK Consortium |

| EPI_Accession_ID | Originating Laboratory | Submitting Laboratory |
|---|---|---|
| EPI_ISL_443030 | Department of Pathology, University of Cambridge | Wellcome Sanger Institute for the COVID-19 Genomics UK Consortium |
| EPI_ISL_443031 | Department of Pathology, University of Cambridge | Wellcome Sanger Institute for the COVID-19 Genomics UK Consortium |
| EPI_ISL_443032 | Department of Pathology, University of Cambridge | Wellcome Sanger Institute for the COVID-19 Genomics UK Consortium |
| EPI_ISL_443033 | Department of Pathology, University of Cambridge | Wellcome Sanger Institute for the COVID-19 Genomics UK Consortium |
| EPI_ISL_443034 | Department of Pathology, University of Cambridge | Wellcome Sanger Institute for the COVID-19 Genomics UK Consortium |
| EPI_ISL_443035 | Department of Pathology, University of Cambridge | Wellcome Sanger Institute for the COVID-19 Genomics UK Consortium |
| EPI_ISL_443036 | Department of Pathology, University of Cambridge | Wellcome Sanger Institute for the COVID-19 Genomics UK Consortium |
| EPI_ISL_443037 | Department of Pathology, University of Cambridge | Wellcome Sanger Institute for the COVID-19 Genomics UK Consortium |
| EPI_ISL_443038 | Department of Pathology, University of Cambridge | Wellcome Sanger Institute for the COVID-19 Genomics UK Consortium |
| EPI_ISL_443039 | Department of Pathology, University of Cambridge | Wellcome Sanger Institute for the COVID-19 Genomics UK Consortium |
| EPI_ISL_443040 | Department of Pathology, University of Cambridge | Wellcome Sanger Institute for the COVID-19 Genomics UK Consortium |
| EPI_ISL_443041 | Department of Pathology, University of Cambridge | Wellcome Sanger Institute for the COVID-19 Genomics UK Consortium |
| EPI_ISL_443042 | Department of Pathology, University of Cambridge | Wellcome Sanger Institute for the COVID-19 Genomics UK Consortium |
| EPI_ISL_443043 | Department of Pathology, University of Cambridge | Wellcome Sanger Institute for the COVID-19 Genomics UK Consortium |
| EPI_ISL_443044 | Department of Pathology, University of Cambridge | Wellcome Sanger Institute for the COVID-19 Genomics UK Consortium |
| EPI_ISL_443045 | Department of Pathology, University of Cambridge | Wellcome Sanger Institute for the COVID-19 Genomics UK Consortium |

| EPI_Accession_ID | Originating Laboratory | Submitting Laboratory |
|---|---|---|
| EPI_ISL_443046 | Department of Pathology, University of Cambridge | Wellcome Sanger Institute for the COVID-19 Genomics UK Consortium |
| EPI_ISL_443047 | Department of Pathology, University of Cambridge | Wellcome Sanger Institute for the COVID-19 Genomics UK Consortium |
| EPI_ISL_443048 | Department of Pathology, University of Cambridge | Wellcome Sanger Institute for the COVID-19 Genomics UK Consortium |
| EPI_ISL_443049 | Department of Pathology, University of Cambridge | Wellcome Sanger Institute for the COVID-19 Genomics UK Consortium |
| EPI_ISL_443050 | Department of Pathology, University of Cambridge | Wellcome Sanger Institute for the COVID-19 Genomics UK Consortium |
| EPI_ISL_443051 | Department of Pathology, University of Cambridge | Wellcome Sanger Institute for the COVID-19 Genomics UK Consortium |
| EPI_ISL_443052 | Department of Pathology, University of Cambridge | Wellcome Sanger Institute for the COVID-19 Genomics UK Consortium |
| EPI_ISL_443053 | Department of Pathology, University of Cambridge | Wellcome Sanger Institute for the COVID-19 Genomics UK Consortium |
| EPI_ISL_443054 | Department of Pathology, University of Cambridge | Wellcome Sanger Institute for the COVID-19 Genomics UK Consortium |
| EPI_ISL_443055 | Department of Pathology, University of Cambridge | Wellcome Sanger Institute for the COVID-19 Genomics UK Consortium |
| EPI_ISL_443056 | Department of Pathology, University of Cambridge | Wellcome Sanger Institute for the COVID-19 Genomics UK Consortium |
| EPI_ISL_443057 | Department of Pathology, University of Cambridge | Wellcome Sanger Institute for the COVID-19 Genomics UK Consortium |
| EPI_ISL_443058 | Department of Pathology, University of Cambridge | Wellcome Sanger Institute for the COVID-19 Genomics UK Consortium |
| EPI_ISL_443059 | Department of Pathology, University of Cambridge | Wellcome Sanger Institute for the COVID-19 Genomics UK Consortium |
| EPI_ISL_443060 | Department of Pathology, University of Cambridge | Wellcome Sanger Institute for the COVID-19 Genomics UK Consortium |
| EPI_ISL_443061 | Department of Pathology, University of Cambridge | Wellcome Sanger Institute for the COVID-19 Genomics UK Consortium |

| EPI_Accession_ID | Originating Laboratory | Submitting Laboratory |
|---|---|---|
| EPI_ISL_443062 | Department of Pathology, University of Cambridge | Wellcome Sanger Institute for the COVID-19 Genomics UK Consortium |
| EPI_ISL_443063 | Department of Pathology, University of Cambridge | Wellcome Sanger Institute for the COVID-19 Genomics UK Consortium |
| EPI_ISL_443064 | Department of Pathology, University of Cambridge | Wellcome Sanger Institute for the COVID-19 Genomics UK Consortium |
| EPI_ISL_443065 | Department of Pathology, University of Cambridge | Wellcome Sanger Institute for the COVID-19 Genomics UK Consortium |
| EPI_ISL_443066 | Department of Pathology, University of Cambridge | Wellcome Sanger Institute for the COVID-19 Genomics UK Consortium |
| EPI_ISL_443067 | Department of Pathology, University of Cambridge | Wellcome Sanger Institute for the COVID-19 Genomics UK Consortium |
| EPI_ISL_443068 | Department of Pathology, University of Cambridge | Wellcome Sanger Institute for the COVID-19 Genomics UK Consortium |
| EPI_ISL_443069 | Department of Pathology, University of Cambridge | Wellcome Sanger Institute for the COVID-19 Genomics UK Consortium |
| EPI_ISL_443070 | Department of Pathology, University of Cambridge | Wellcome Sanger Institute for the COVID-19 Genomics UK Consortium |
| EPI_ISL_443071 | Department of Pathology, University of Cambridge | Wellcome Sanger Institute for the COVID-19 Genomics UK Consortium |
| EPI_ISL_443072 | Department of Pathology, University of Cambridge | Wellcome Sanger Institute for the COVID-19 Genomics UK Consortium |
| EPI_ISL_443073 | Department of Pathology, University of Cambridge | Wellcome Sanger Institute for the COVID-19 Genomics UK Consortium |
| EPI_ISL_443074 | Department of Pathology, University of Cambridge | Wellcome Sanger Institute for the COVID-19 Genomics UK Consortium |
| EPI_ISL_443075 | Department of Pathology, University of Cambridge | Wellcome Sanger Institute for the COVID-19 Genomics UK Consortium |
| EPI_ISL_443076 | Department of Pathology, University of Cambridge | Wellcome Sanger Institute for the COVID-19 Genomics UK Consortium |
| EPI_ISL_443077 | Department of Pathology, University of Cambridge | Wellcome Sanger Institute for the COVID-19 Genomics UK Consortium |

| EPI_Accession_ID | Originating Laboratory | Submitting Laboratory |
|---|---|---|
| EPI_ISL_443078 | Department of Pathology, University of Cambridge | Wellcome Sanger Institute for the COVID-19 Genomics UK Consortium |
| EPI_ISL_443079 | Department of Pathology, University of Cambridge | Wellcome Sanger Institute for the COVID-19 Genomics UK Consortium |
| EPI_ISL_443080 | Department of Pathology, University of Cambridge | Wellcome Sanger Institute for the COVID-19 Genomics UK Consortium |
| EPI_ISL_443081 | Department of Pathology, University of Cambridge | Wellcome Sanger Institute for the COVID-19 Genomics UK Consortium |
| EPI_ISL_443082 | Department of Pathology, University of Cambridge | Wellcome Sanger Institute for the COVID-19 Genomics UK Consortium |
| EPI_ISL_443083 | Department of Pathology, University of Cambridge | Wellcome Sanger Institute for the COVID-19 Genomics UK Consortium |
| EPI_ISL_443084 | Department of Pathology, University of Cambridge | Wellcome Sanger Institute for the COVID-19 Genomics UK Consortium |
| EPI_ISL_443085 | Department of Pathology, University of Cambridge | Wellcome Sanger Institute for the COVID-19 Genomics UK Consortium |
| EPI_ISL_443086 | Department of Pathology, University of Cambridge | Wellcome Sanger Institute for the COVID-19 Genomics UK Consortium |
| EPI_ISL_443087 | Department of Pathology, University of Cambridge | Wellcome Sanger Institute for the COVID-19 Genomics UK Consortium |
| EPI_ISL_443088 | Department of Pathology, University of Cambridge | Wellcome Sanger Institute for the COVID-19 Genomics UK Consortium |
| EPI_ISL_443089 | Department of Pathology, University of Cambridge | Wellcome Sanger Institute for the COVID-19 Genomics UK Consortium |
| EPI_ISL_443090 | Department of Pathology, University of Cambridge | Wellcome Sanger Institute for the COVID-19 Genomics UK Consortium |
| EPI_ISL_443091 | Department of Pathology, University of Cambridge | Wellcome Sanger Institute for the COVID-19 Genomics UK Consortium |
| EPI_ISL_443092 | Department of Pathology, University of Cambridge | Wellcome Sanger Institute for the COVID-19 Genomics UK Consortium |
| EPI_ISL_443093 | Department of Pathology, University of Cambridge | Wellcome Sanger Institute for the COVID-19 Genomics UK Consortium |

| EPI_Accession_ID | Originating Laboratory | Submitting Laboratory |
|---|---|---|
| EPI_ISL_443094 | Department of Pathology, University of Cambridge | Wellcome Sanger Institute for the COVID-19 Genomics UK Consortium |
| EPI_ISL_443095 | Department of Pathology, University of Cambridge | Wellcome Sanger Institute for the COVID-19 Genomics UK Consortium |
| EPI_ISL_443096 | Department of Pathology, University of Cambridge | Wellcome Sanger Institute for the COVID-19 Genomics UK Consortium |
| EPI_ISL_443097 | Department of Pathology, University of Cambridge | Wellcome Sanger Institute for the COVID-19 Genomics UK Consortium |
| EPI_ISL_443098 | Department of Pathology, University of Cambridge | Wellcome Sanger Institute for the COVID-19 Genomics UK Consortium |
| EPI_ISL_443099 | Department of Pathology, University of Cambridge | Wellcome Sanger Institute for the COVID-19 Genomics UK Consortium |
| EPI_ISL_443100 | Department of Pathology, University of Cambridge | Wellcome Sanger Institute for the COVID-19 Genomics UK Consortium |
| EPI_ISL_443101 | Department of Pathology, University of Cambridge | Wellcome Sanger Institute for the COVID-19 Genomics UK Consortium |
| EPI_ISL_443102 | Department of Pathology, University of Cambridge | Wellcome Sanger Institute for the COVID-19 Genomics UK Consortium |
| EPI_ISL_443103 | Department of Pathology, University of Cambridge | Wellcome Sanger Institute for the COVID-19 Genomics UK Consortium |
| EPI_ISL_443104 | Department of Pathology, University of Cambridge | Wellcome Sanger Institute for the COVID-19 Genomics UK Consortium |
| EPI_ISL_443105 | Department of Pathology, University of Cambridge | Wellcome Sanger Institute for the COVID-19 Genomics UK Consortium |
| EPI_ISL_443106 | Department of Pathology, University of Cambridge | Wellcome Sanger Institute for the COVID-19 Genomics UK Consortium |
| EPI_ISL_443107 | Department of Pathology, University of Cambridge | Wellcome Sanger Institute for the COVID-19 Genomics UK Consortium |
| EPI_ISL_443108 | Department of Pathology, University of Cambridge | Wellcome Sanger Institute for the COVID-19 Genomics UK Consortium |
| EPI_ISL_443109 | Department of Pathology, University of Cambridge | Wellcome Sanger Institute for the COVID-19 Genomics UK Consortium |

| EPI_Accession_ID | Originating Laboratory | Submitting Laboratory |
|---|---|---|
| EPI_ISL_443110 | Department of Pathology, University of Cambridge | Wellcome Sanger Institute for the COVID-19 Genomics UK Consortium |
| EPI_ISL_443111 | Department of Pathology, University of Cambridge | Wellcome Sanger Institute for the COVID-19 Genomics UK Consortium |
| EPI_ISL_443112 | Department of Pathology, University of Cambridge | Wellcome Sanger Institute for the COVID-19 Genomics UK Consortium |
| EPI_ISL_443113 | Department of Pathology, University of Cambridge | Wellcome Sanger Institute for the COVID-19 Genomics UK Consortium |
| EPI_ISL_443114 | Department of Pathology, University of Cambridge | Wellcome Sanger Institute for the COVID-19 Genomics UK Consortium |
| EPI_ISL_443115 | Department of Pathology, University of Cambridge | Wellcome Sanger Institute for the COVID-19 Genomics UK Consortium |
| EPI_ISL_443116 | Department of Pathology, University of Cambridge | Wellcome Sanger Institute for the COVID-19 Genomics UK Consortium |
| EPI_ISL_443117 | Department of Pathology, University of Cambridge | Wellcome Sanger Institute for the COVID-19 Genomics UK Consortium |
| EPI_ISL_443118 | Department of Pathology, University of Cambridge | Wellcome Sanger Institute for the COVID-19 Genomics UK Consortium |
| EPI_ISL_443183 | M Health Fairview | University of Minnesota Genomics Center |
| EPI_ISL_443184 | UW Virology Lab | UW Virology Lab |
| EPI_ISL_443185 | UW Virology Lab | UW Virology Lab |
| EPI_ISL_443186 | UW Virology Lab | UW Virology Lab |
| EPI_ISL_443187 | National Virology Reference Laboratory | National Public Health Laboratory, National Centre for Infectious Diseases |
| EPI_ISL_443188 | National Public Health Laboratory, National Centre for Infectious Diseases | National Public Health Laboratory, National Centre for Infectious Diseases |
| EPI_ISL_443189 | National Public Health Laboratory, National Centre for Infectious Diseases | National Public Health Laboratory, National Centre for Infectious Diseases |
| EPI_ISL_443190 | National Public Health Laboratory, National Centre for Infectious Diseases | National Public Health Laboratory, National Centre for Infectious Diseases |
| EPI_ISL_443191 | National Public Health Laboratory, National Centre for Infectious Diseases | National Public Health Laboratory, National Centre for Infectious Diseases |

| EPI_Accession_ID | Originating Laboratory | Submitting Laboratory |
|---|---|---|
| EPI_ISL_443192 | National Public Health Laboratory, National Centre for Infectious Diseases | National Public Health Laboratory, National Centre for Infectious Diseases |
| EPI_ISL_443193 | National Public Health Laboratory, National Centre for Infectious Diseases | National Public Health Laboratory, National Centre for Infectious Diseases |
| EPI_ISL_443194 | National Public Health Laboratory, National Centre for Infectious Diseases | National Public Health Laboratory, National Centre for Infectious Diseases |
| EPI_ISL_443195 | National Public Health Laboratory, National Centre for Infectious Diseases | National Public Health Laboratory, National Centre for Infectious Diseases |
| EPI_ISL_443196 | National Public Health Laboratory, National Centre for Infectious Diseases | National Public Health Laboratory, National Centre for Infectious Diseases |
| EPI_ISL_443197 | National Public Health Laboratory, National Centre for Infectious Diseases | National Public Health Laboratory, National Centre for Infectious Diseases |
| EPI_ISL_443198 | National Public Health Laboratory, National Centre for Infectious Diseases | National Public Health Laboratory, National Centre for Infectious Diseases |
| EPI_ISL_443199 | National Public Health Laboratory, National Centre for Infectious Diseases | National Public Health Laboratory, National Centre for Infectious Diseases |
| EPI_ISL_443200 | National Public Health Laboratory, National Centre for Infectious Diseases | National Public Health Laboratory, National Centre for Infectious Diseases |
| EPI_ISL_443201 | National Public Health Laboratory, National Centre for Infectious Diseases | National Public Health Laboratory, National Centre for Infectious Diseases |
| EPI_ISL_443202 | National Public Health Laboratory, National Centre for Infectious Diseases | National Public Health Laboratory, National Centre for Infectious Diseases |
| EPI_ISL_443203 | National Public Health Laboratory, National Centre for Infectious Diseases | National Public Health Laboratory, National Centre for Infectious Diseases |
| EPI_ISL_443204 | National Public Health Laboratory, National Centre for Infectious Diseases | National Public Health Laboratory, National Centre for Infectious Diseases |
| EPI_ISL_443205 | National Public Health Laboratory, National Centre for Infectious Diseases | National Public Health Laboratory, National Centre for Infectious Diseases |
| EPI_ISL_443206 | National Public Health Laboratory, National Centre for Infectious Diseases | National Public Health Laboratory, National Centre for Infectious Diseases |
| EPI_ISL_443207 | National Public Health Laboratory, National Centre for Infectious Diseases | National Public Health Laboratory, National Centre for Infectious Diseases |

| EPI_Accession_ID | Originating Laboratory | Submitting Laboratory |
|---|---|---|
| EPI_ISL_443208 | National Public Health Laboratory, National Centre for Infectious Diseases | National Public Health Laboratory, National Centre for Infectious Diseases |
| EPI_ISL_443209 | National Public Health Laboratory, National Centre for Infectious Diseases | National Public Health Laboratory, National Centre for Infectious Diseases |
| EPI_ISL_443210 | National Public Health Laboratory, National Centre for Infectious Diseases | National Public Health Laboratory, National Centre for Infectious Diseases |
| EPI_ISL_443211 | National Public Health Laboratory, National Centre for Infectious Diseases | National Public Health Laboratory, National Centre for Infectious Diseases |
| EPI_ISL_443212 | National Public Health Laboratory, National Centre for Infectious Diseases | National Public Health Laboratory, National Centre for Infectious Diseases |
| EPI_ISL_443213 | National Public Health Laboratory, National Centre for Infectious Diseases | National Public Health Laboratory, National Centre for Infectious Diseases |
| EPI_ISL_443214 | National Public Health Laboratory, National Centre for Infectious Diseases | National Public Health Laboratory, National Centre for Infectious Diseases |
| EPI_ISL_443215 | National Public Health Laboratory, National Centre for Infectious Diseases | National Public Health Laboratory, National Centre for Infectious Diseases |
| EPI_ISL_443216 | National Public Health Laboratory, National Centre for Infectious Diseases | National Public Health Laboratory, National Centre for Infectious Diseases |
| EPI_ISL_443217 | National Public Health Laboratory, National Centre for Infectious Diseases | National Public Health Laboratory, National Centre for Infectious Diseases |
| EPI_ISL_443218 | National Public Health Laboratory, National Centre for Infectious Diseases | National Public Health Laboratory, National Centre for Infectious Diseases |
| EPI_ISL_443219 | National Public Health Laboratory, National Centre for Infectious Diseases | National Public Health Laboratory, National Centre for Infectious Diseases |
| EPI_ISL_443220 | National Public Health Laboratory, National Centre for Infectious Diseases | National Public Health Laboratory, National Centre for Infectious Diseases |
| EPI_ISL_443221 | National Public Health Laboratory, National Centre for Infectious Diseases | National Public Health Laboratory, National Centre for Infectious Diseases |
| EPI_ISL_443222 | National Public Health Laboratory, National Centre for Infectious Diseases | National Public Health Laboratory, National Centre for Infectious Diseases |
| EPI_ISL_443223 | National Public Health Laboratory, National Centre for Infectious Diseases | National Public Health Laboratory, National Centre for Infectious Diseases |

| EPI_Accession_ID | Originating Laboratory | Submitting Laboratory |
|---|---|---|
| EPI_ISL_443224 | National Public Health Laboratory, National Centre for Infectious Diseases | National Public Health Laboratory, National Centre for Infectious Diseases |
| EPI_ISL_443225 | National Public Health Laboratory, National Centre for Infectious Diseases | National Public Health Laboratory, National Centre for Infectious Diseases |
| EPI_ISL_443226 | National Public Health Laboratory, National Centre for Infectious Diseases | National Public Health Laboratory, National Centre for Infectious Diseases |
| EPI_ISL_443227 | National Public Health Laboratory, National Centre for Infectious Diseases | National Public Health Laboratory, National Centre for Infectious Diseases |
| EPI_ISL_443228 | National Public Health Laboratory, National Centre for Infectious Diseases | National Public Health Laboratory, National Centre for Infectious Diseases |
| EPI_ISL_443229 | National Public Health Laboratory, National Centre for Infectious Diseases | National Public Health Laboratory, National Centre for Infectious Diseases |
| EPI_ISL_443230 | National Public Health Laboratory, National Centre for Infectious Diseases | National Public Health Laboratory, National Centre for Infectious Diseases |
| EPI_ISL_443231 | National Public Health Laboratory, National Centre for Infectious Diseases | National Public Health Laboratory, National Centre for Infectious Diseases |
| EPI_ISL_443232 | National Public Health Laboratory, National Centre for Infectious Diseases | National Public Health Laboratory, National Centre for Infectious Diseases |
| EPI_ISL_443233 | National Public Health Laboratory, National Centre for Infectious Diseases | National Public Health Laboratory, National Centre for Infectious Diseases |
| EPI_ISL_443234 | National Public Health Laboratory, National Centre for Infectious Diseases | National Public Health Laboratory, National Centre for Infectious Diseases |
| EPI_ISL_443235 | National Public Health Laboratory, National Centre for Infectious Diseases | National Public Health Laboratory, National Centre for Infectious Diseases |
| EPI_ISL_443236 | National Public Health Laboratory, National Centre for Infectious Diseases | National Public Health Laboratory, National Centre for Infectious Diseases |
| EPI_ISL_443237 | National Public Health Laboratory, National Centre for Infectious Diseases | National Public Health Laboratory, National Centre for Infectious Diseases |
| EPI_ISL_443238 | National Public Health Laboratory, National Centre for Infectious Diseases | National Public Health Laboratory, National Centre for Infectious Diseases |
| EPI_ISL_443239 | National Public Health Laboratory, National Centre for Infectious Diseases | National Public Health Laboratory, National Centre for Infectious Diseases |

| EPI_Accession_ID | Originating Laboratory | Submitting Laboratory |
|---|---|---|
| EPI_ISL_443240 | National Public Health Laboratory, National Centre for Infectious Diseases | National Public Health Laboratory, National Centre for Infectious Diseases |
| EPI_ISL_443241 | National Public Health Laboratory, National Centre for Infectious Diseases | National Public Health Laboratory, National Centre for Infectious Diseases |
| EPI_ISL_443242 | National Public Health Laboratory, National Centre for Infectious Diseases | National Public Health Laboratory, National Centre for Infectious Diseases |
| EPI_ISL_443243 | National Public Health Laboratory, National Centre for Infectious Diseases | National Public Health Laboratory, National Centre for Infectious Diseases |
| EPI_ISL_443244 | National Public Health Laboratory, National Centre for Infectious Diseases | National Public Health Laboratory, National Centre for Infectious Diseases |
| EPI_ISL_443245 | National Public Health Laboratory, National Centre for Infectious Diseases | National Public Health Laboratory, National Centre for Infectious Diseases |
| EPI_ISL_443246 | National Public Health Laboratory, National Centre for Infectious Diseases | National Public Health Laboratory, National Centre for Infectious Diseases |
| EPI_ISL_443247 | National Public Health Laboratory, National Centre for Infectious Diseases | National Public Health Laboratory, National Centre for Infectious Diseases |
| EPI_ISL_443248 | National Public Health Laboratory, National Centre for Infectious Diseases | National Public Health Laboratory, National Centre for Infectious Diseases |
| EPI_ISL_443249 | National Public Health Laboratory, National Centre for Infectious Diseases | National Public Health Laboratory, National Centre for Infectious Diseases |
| EPI_ISL_443253 | M Health Fairview | University of Minnesota Genomics Center |
| EPI_ISL_443254 | M Health Fairview | University of Minnesota Genomics Center |
| EPI_ISL_443255 | M Health Fairview | University of Minnesota Genomics Center |
| EPI_ISL_443256 | M Health Fairview | University of Minnesota Genomics Center |
| EPI_ISL_443257 | M Health Fairview | University of Minnesota Genomics Center |
| EPI_ISL_443258 | RÃ©sidence Ornano | National Reference Center for Viruses of Respiratory Infections, Institut Pasteur, Paris |
| EPI_ISL_443259 | RÃ©sidence Ornano | National Reference Center for Viruses of Respiratory Infections, Institut Pasteur, Paris |
| EPI_ISL_443260 | LABM GH nord Essonne de Longjumeau - BP 125 | National Reference Center for Viruses of Respiratory Infections, Institut Pasteur, Paris |
| EPI_ISL_443261 | CHU de Dijon - Laboratoire de Virologie | National Reference Center for Viruses of Respiratory Infections, Institut Pasteur, Paris |

| EPI_Accession_ID | Originating Laboratory | Submitting Laboratory |
| --- | --- | --- |
| EPI_ISL_443262 | CHU de Dijon - Laboratoire de Virologie | National Reference Center for Viruses of Respiratory Infections, Institut Pasteur, Paris |
| EPI_ISL_443263 | CHU de Dijon - Laboratoire de Virologie | National Reference Center for Viruses of Respiratory Infections, Institut Pasteur, Paris |
| EPI_ISL_443264 | CHU de Dijon - Laboratoire de Virologie | National Reference Center for Viruses of Respiratory Infections, Institut Pasteur, Paris |
| EPI_ISL_443265 | CHU - HÃ´pital Cavale Blanche - Labo. de Virologie | National Reference Center for Viruses of Respiratory Infections, Institut Pasteur, Paris |
| EPI_ISL_443266 | CHU - HÃ´pital Cavale Blanche - Labo. de Virologie | National Reference Center for Viruses of Respiratory Infections, Institut Pasteur, Paris |
| EPI_ISL_443267 | CHU - HÃ´pital Cavale Blanche - Labo. de Virologie | National Reference Center for Viruses of Respiratory Infections, Institut Pasteur, Paris |
| EPI_ISL_443268 | CHU - HÃ´pital Cavale Blanche - Labo. de Virologie | National Reference Center for Viruses of Respiratory Infections, Institut Pasteur, Paris |
| EPI_ISL_443269 | CHU - HÃ´pital Cavale Blanche - Labo. de Virologie | National Reference Center for Viruses of Respiratory Infections, Institut Pasteur, Paris |
| EPI_ISL_443270 | CHU - HÃ´pital Cavale Blanche - Labo. de Virologie | National Reference Center for Viruses of Respiratory Infections, Institut Pasteur, Paris |
| EPI_ISL_443271 | CHU - HÃ´pital Cavale Blanche - Labo. de Virologie | National Reference Center for Viruses of Respiratory Infections, Institut Pasteur, Paris |
| EPI_ISL_443272 | CHU - HÃ´pital Cavale Blanche - Labo. de Virologie | National Reference Center for Viruses of Respiratory Infections, Institut Pasteur, Paris |
| EPI_ISL_443273 | CHU - HÃ´pital Cavale Blanche - Labo. de Virologie | National Reference Center for Viruses of Respiratory Infections, Institut Pasteur, Paris |
| EPI_ISL_443274 | CHU - HÃ´pital Cavale Blanche - Labo. de Virologie | National Reference Center for Viruses of Respiratory Infections, Institut Pasteur, Paris |
| EPI_ISL_443275 | CHU - HÃ´pital Cavale Blanche - Labo. de Virologie | National Reference Center for Viruses of Respiratory Infections, Institut Pasteur, Paris |
| EPI_ISL_443276 | CHU - HÃ´pital Cavale Blanche - Labo. de Virologie | National Reference Center for Viruses of Respiratory Infections, Institut Pasteur, Paris |
| EPI_ISL_443277 | CHU - HÃ´pital Cavale Blanche - Labo. de Virologie | National Reference Center for Viruses of Respiratory Infections, Institut Pasteur, Paris |

| EPI_Accession_ID | Originating Laboratory | Submitting Laboratory |
|---|---|---|
| EPI_ISL_443278 | CHU - HÃ´pital Cavale Blanche - Labo. de Virologie | National Reference Center for Viruses of Respiratory Infections, Institut Pasteur, Paris |
| EPI_ISL_443279 | CHU - HÃ´pital Cavale Blanche - Labo. de Virologie | National Reference Center for Viruses of Respiratory Infections, Institut Pasteur, Paris |
| EPI_ISL_443280 | CHU - HÃ´pital Cavale Blanche - Labo. de Virologie | National Reference Center for Viruses of Respiratory Infections, Institut Pasteur, Paris |
| EPI_ISL_443281 | CHU - HÃ´pital Cavale Blanche - Labo. de Virologie | National Reference Center for Viruses of Respiratory Infections, Institut Pasteur, Paris |
| EPI_ISL_443282 | CHU - HÃ´pital Cavale Blanche - Labo. de Virologie | National Reference Center for Viruses of Respiratory Infections, Institut Pasteur, Paris |
| EPI_ISL_443283 | CHU - HÃ´pital Cavale Blanche - Labo. de Virologie | National Reference Center for Viruses of Respiratory Infections, Institut Pasteur, Paris |
| EPI_ISL_443284 | Laboratoire de Microbiologie - BÃ¢t A - CH RenÃ© Dubois | National Reference Center for Viruses of Respiratory Infections, Institut Pasteur, Paris |
| EPI_ISL_443285 | Laboratoire de Microbiologie - BÃ¢t A - CH RenÃ© Dubois | National Reference Center for Viruses of Respiratory Infections, Institut Pasteur, Paris |
| EPI_ISL_443286 | Laboratoire de Microbiologie - BÃ¢t A - CH RenÃ© Dubois | National Reference Center for Viruses of Respiratory Infections, Institut Pasteur, Paris |
| EPI_ISL_443287 | Laboratoire de Microbiologie - BÃ¢t A - CH RenÃ© Dubois | National Reference Center for Viruses of Respiratory Infections, Institut Pasteur, Paris |
| EPI_ISL_443288 | Laboratoire de Microbiologie - BÃ¢t A - CH RenÃ© Dubois | National Reference Center for Viruses of Respiratory Infections, Institut Pasteur, Paris |
| EPI_ISL_443289 | CHRU Pontchaillou - Laboratoire de Virologie | National Reference Center for Viruses of Respiratory Infections, Institut Pasteur, Paris |
| EPI_ISL_443290 | CHRU Pontchaillou - Laboratoire de Virologie | National Reference Center for Viruses of Respiratory Infections, Institut Pasteur, Paris |
| EPI_ISL_443291 | CHRU Pontchaillou - Laboratoire de Virologie | National Reference Center for Viruses of Respiratory Infections, Institut Pasteur, Paris |
| EPI_ISL_443292 | CHRU Pontchaillou - Laboratoire de Virologie | National Reference Center for Viruses of Respiratory Infections, Institut Pasteur, Paris |
| EPI_ISL_443293 | CHRU Pontchaillou - Laboratoire de Virologie | National Reference Center for Viruses of Respiratory Infections, Institut Pasteur, Paris |

| EPI_Accession_ID | Originating Laboratory | Submitting Laboratory |
|---|---|---|
| EPI_ISL_443294 | CHRU Pontchaillou - Laboratoire de Virologie | National Reference Center for Viruses of Respiratory Infections, Institut Pasteur, Paris |
| EPI_ISL_443295 | HÃ´pital Necker - Enfants - Malades Laboratoire de Virologie | National Reference Center for Viruses of Respiratory Infections, Institut Pasteur, Paris |
| EPI_ISL_443296 | HÃ´pital Necker - Enfants - Malades Laboratoire de Virologie | National Reference Center for Viruses of Respiratory Infections, Institut Pasteur, Paris |
| EPI_ISL_443297 | HÃ´pital Necker - Enfants - Malades Laboratoire de Virologie | National Reference Center for Viruses of Respiratory Infections, Institut Pasteur, Paris |
| EPI_ISL_443298 | HÃ´pital Necker - Enfants - Malades Laboratoire de Virologie | National Reference Center for Viruses of Respiratory Infections, Institut Pasteur, Paris |
| EPI_ISL_443299 | HÃ´pital Necker - Enfants - Malades Laboratoire de Virologie | National Reference Center for Viruses of Respiratory Infections, Institut Pasteur, Paris |
| EPI_ISL_443300 | Cabinet MÃ©dical | National Reference Center for Viruses of Respiratory Infections, Institut Pasteur, Paris |
| EPI_ISL_443301 | Cabinet MÃ©dical | National Reference Center for Viruses of Respiratory Infections, Institut Pasteur, Paris |
| EPI_ISL_443302 | Cabinet MÃ©dical | National Reference Center for Viruses of Respiratory Infections, Institut Pasteur, Paris |
| EPI_ISL_443303 | RÃ©sidence Les Marines | National Reference Center for Viruses of Respiratory Infections, Institut Pasteur, Paris |
| EPI_ISL_443304 | RÃ©sidence Esterel | National Reference Center for Viruses of Respiratory Infections, Institut Pasteur, Paris |
| EPI_ISL_443305 | LABM GH nord Essonne de Longjumeau - BP 125 | National Reference Center for Viruses of Respiratory Infections, Institut Pasteur, Paris |
| EPI_ISL_443306 | Cabinet MÃ©dical | National Reference Center for Viruses of Respiratory Infections, Institut Pasteur, Paris |
| EPI_ISL_443307 | La Villa Papyri | National Reference Center for Viruses of Respiratory Infections, Institut Pasteur, Paris |
| EPI_ISL_443308 | Plaisance | National Reference Center for Viruses of Respiratory Infections, Institut Pasteur, Paris |
| EPI_ISL_443309 | CH CompiÃ¨gne Laboratoire de Biologie | National Reference Center for Viruses of Respiratory Infections, Institut Pasteur, Paris |

| EPI_Accession_ID | Originating Laboratory | Submitting Laboratory |
|---|---|---|
| EPI_ISL_443310 | Centre de santÃ© Filieris | National Reference Center for Viruses of Respiratory Infections, Institut Pasteur, Paris |
| EPI_ISL_443311 | Cabinet MÃ©dical | National Reference Center for Viruses of Respiratory Infections, Institut Pasteur, Paris |
| EPI_ISL_443312 | Cabinet MÃ©dical | National Reference Center for Viruses of Respiratory Infections, Institut Pasteur, Paris |
| EPI_ISL_443313 | Cabinet MÃ©dical | National Reference Center for Viruses of Respiratory Infections, Institut Pasteur, Paris |
| EPI_ISL_443314 | LABM GH nord Essonne de Longjumeau - BP 125 | National Reference Center for Viruses of Respiratory Infections, Institut Pasteur, Paris |
| EPI_ISL_443315 | ChÃ¢teau de la Source | National Reference Center for Viruses of Respiratory Infections, Institut Pasteur, Paris |
| EPI_ISL_443316 | CH CompiÃ¨gne Laboratoire de Biologie | National Reference Center for Viruses of Respiratory Infections, Institut Pasteur, Paris |
| EPI_ISL_443317 | Cabinet MÃ©dical | National Reference Center for Viruses of Respiratory Infections, Institut Pasteur, Paris |
| EPI_ISL_443318 | Department of Pathology, University of Cambridge | Wellcome Sanger Institute for the COVID-19 Genomics UK Consortium |
| EPI_ISL_443319 | Department of Pathology, University of Cambridge | Wellcome Sanger Institute for the COVID-19 Genomics UK Consortium |
| EPI_ISL_443320 | Department of Pathology, University of Cambridge | Wellcome Sanger Institute for the COVID-19 Genomics UK Consortium |
| EPI_ISL_443321 | Department of Pathology, University of Cambridge | Wellcome Sanger Institute for the COVID-19 Genomics UK Consortium |
| EPI_ISL_443322 | Department of Pathology, University of Cambridge | Wellcome Sanger Institute for the COVID-19 Genomics UK Consortium |
| EPI_ISL_443323 | Department of Pathology, University of Cambridge | Wellcome Sanger Institute for the COVID-19 Genomics UK Consortium |
| EPI_ISL_443324 | Department of Pathology, University of Cambridge | Wellcome Sanger Institute for the COVID-19 Genomics UK Consortium |
| EPI_ISL_443325 | Department of Pathology, University of Cambridge | Wellcome Sanger Institute for the COVID-19 Genomics UK Consortium |

| EPI_Accession_ID | Originating Laboratory | Submitting Laboratory |
|---|---|---|
| EPI_ISL_443326 | Department of Pathology, University of Cambridge | Wellcome Sanger Institute for the COVID-19 Genomics UK Consortium |
| EPI_ISL_443327 | Department of Pathology, University of Cambridge | Wellcome Sanger Institute for the COVID-19 Genomics UK Consortium |
| EPI_ISL_443328 | Department of Pathology, University of Cambridge | Wellcome Sanger Institute for the COVID-19 Genomics UK Consortium |
| EPI_ISL_443329 | Department of Pathology, University of Cambridge | Wellcome Sanger Institute for the COVID-19 Genomics UK Consortium |
| EPI_ISL_443330 | Department of Pathology, University of Cambridge | Wellcome Sanger Institute for the COVID-19 Genomics UK Consortium |
| EPI_ISL_443331 | Department of Pathology, University of Cambridge | Wellcome Sanger Institute for the COVID-19 Genomics UK Consortium |
| EPI_ISL_443332 | Department of Pathology, University of Cambridge | Wellcome Sanger Institute for the COVID-19 Genomics UK Consortium |
| EPI_ISL_443333 | Department of Pathology, University of Cambridge | Wellcome Sanger Institute for the COVID-19 Genomics UK Consortium |
| EPI_ISL_443334 | Department of Pathology, University of Cambridge | Wellcome Sanger Institute for the COVID-19 Genomics UK Consortium |
| EPI_ISL_443335 | Department of Pathology, University of Cambridge | Wellcome Sanger Institute for the COVID-19 Genomics UK Consortium |
| EPI_ISL_443336 | Department of Pathology, University of Cambridge | Wellcome Sanger Institute for the COVID-19 Genomics UK Consortium |
| EPI_ISL_443337 | Department of Pathology, University of Cambridge | Wellcome Sanger Institute for the COVID-19 Genomics UK Consortium |
| EPI_ISL_443338 | Department of Pathology, University of Cambridge | Wellcome Sanger Institute for the COVID-19 Genomics UK Consortium |
| EPI_ISL_443339 | Department of Pathology, University of Cambridge | Wellcome Sanger Institute for the COVID-19 Genomics UK Consortium |
| EPI_ISL_443340 | Department of Pathology, University of Cambridge | Wellcome Sanger Institute for the COVID-19 Genomics UK Consortium |
| EPI_ISL_443341 | Department of Pathology, University of Cambridge | Wellcome Sanger Institute for the COVID-19 Genomics UK Consortium |

| EPI_Accession_ID | Originating Laboratory | Submitting Laboratory |
|---|---|---|
| EPI_ISL_443342 | Department of Pathology, University of Cambridge | Wellcome Sanger Institute for the COVID-19 Genomics UK Consortium |
| EPI_ISL_443343 | Department of Pathology, University of Cambridge | Wellcome Sanger Institute for the COVID-19 Genomics UK Consortium |
| EPI_ISL_443344 | Department of Pathology, University of Cambridge | Wellcome Sanger Institute for the COVID-19 Genomics UK Consortium |
| EPI_ISL_443345 | Department of Pathology, University of Cambridge | Wellcome Sanger Institute for the COVID-19 Genomics UK Consortium |
| EPI_ISL_443346 | Department of Pathology, University of Cambridge | Wellcome Sanger Institute for the COVID-19 Genomics UK Consortium |
| EPI_ISL_443347 | Department of Pathology, University of Cambridge | Wellcome Sanger Institute for the COVID-19 Genomics UK Consortium |
| EPI_ISL_443348 | Department of Pathology, University of Cambridge | Wellcome Sanger Institute for the COVID-19 Genomics UK Consortium |
| EPI_ISL_443349 | Department of Pathology, University of Cambridge | Wellcome Sanger Institute for the COVID-19 Genomics UK Consortium |
| EPI_ISL_443350 | Department of Pathology, University of Cambridge | Wellcome Sanger Institute for the COVID-19 Genomics UK Consortium |
| EPI_ISL_443351 | Department of Pathology, University of Cambridge | Wellcome Sanger Institute for the COVID-19 Genomics UK Consortium |
| EPI_ISL_443352 | Department of Pathology, University of Cambridge | Wellcome Sanger Institute for the COVID-19 Genomics UK Consortium |
| EPI_ISL_443353 | Department of Pathology, University of Cambridge | Wellcome Sanger Institute for the COVID-19 Genomics UK Consortium |
| EPI_ISL_443354 | Department of Pathology, University of Cambridge | Wellcome Sanger Institute for the COVID-19 Genomics UK Consortium |
| EPI_ISL_443355 | Department of Pathology, University of Cambridge | Wellcome Sanger Institute for the COVID-19 Genomics UK Consortium |
| EPI_ISL_443356 | Department of Pathology, University of Cambridge | Wellcome Sanger Institute for the COVID-19 Genomics UK Consortium |
| EPI_ISL_443357 | Department of Pathology, University of Cambridge | Wellcome Sanger Institute for the COVID-19 Genomics UK Consortium |

| EPI_Accession_ID | Originating Laboratory | Submitting Laboratory |
|---|---|---|
| EPI_ISL_443358 | Department of Pathology, University of Cambridge | Wellcome Sanger Institute for the COVID-19 Genomics UK Consortium |
| EPI_ISL_443359 | Department of Pathology, University of Cambridge | Wellcome Sanger Institute for the COVID-19 Genomics UK Consortium |
| EPI_ISL_443360 | Department of Pathology, University of Cambridge | Wellcome Sanger Institute for the COVID-19 Genomics UK Consortium |
| EPI_ISL_443361 | Department of Pathology, University of Cambridge | Wellcome Sanger Institute for the COVID-19 Genomics UK Consortium |
| EPI_ISL_443362 | Department of Pathology, University of Cambridge | Wellcome Sanger Institute for the COVID-19 Genomics UK Consortium |
| EPI_ISL_443363 | Department of Pathology, University of Cambridge | Wellcome Sanger Institute for the COVID-19 Genomics UK Consortium |
| EPI_ISL_443364 | Department of Pathology, University of Cambridge | Wellcome Sanger Institute for the COVID-19 Genomics UK Consortium |
| EPI_ISL_443365 | Department of Pathology, University of Cambridge | Wellcome Sanger Institute for the COVID-19 Genomics UK Consortium |
| EPI_ISL_443366 | Department of Pathology, University of Cambridge | Wellcome Sanger Institute for the COVID-19 Genomics UK Consortium |
| EPI_ISL_443367 | Department of Pathology, University of Cambridge | Wellcome Sanger Institute for the COVID-19 Genomics UK Consortium |
| EPI_ISL_443368 | Department of Pathology, University of Cambridge | Wellcome Sanger Institute for the COVID-19 Genomics UK Consortium |
| EPI_ISL_443369 | Department of Pathology, University of Cambridge | Wellcome Sanger Institute for the COVID-19 Genomics UK Consortium |
| EPI_ISL_443370 | Department of Pathology, University of Cambridge | Wellcome Sanger Institute for the COVID-19 Genomics UK Consortium |
| EPI_ISL_443371 | Department of Pathology, University of Cambridge | Wellcome Sanger Institute for the COVID-19 Genomics UK Consortium |
| EPI_ISL_443372 | Department of Pathology, University of Cambridge | Wellcome Sanger Institute for the COVID-19 Genomics UK Consortium |
| EPI_ISL_443373 | Department of Pathology, University of Cambridge | Wellcome Sanger Institute for the COVID-19 Genomics UK Consortium |

| EPI_Accession_ID | Originating Laboratory | Submitting Laboratory |
|---|---|---|
| EPI_ISL_443374 | Department of Pathology, University of Cambridge | Wellcome Sanger Institute for the COVID-19 Genomics UK Consortium |
| EPI_ISL_443375 | Department of Pathology, University of Cambridge | Wellcome Sanger Institute for the COVID-19 Genomics UK Consortium |
| EPI_ISL_443376 | Department of Pathology, University of Cambridge | Wellcome Sanger Institute for the COVID-19 Genomics UK Consortium |
| EPI_ISL_443377 | Department of Pathology, University of Cambridge | Wellcome Sanger Institute for the COVID-19 Genomics UK Consortium |
| EPI_ISL_443378 | Department of Pathology, University of Cambridge | Wellcome Sanger Institute for the COVID-19 Genomics UK Consortium |
| EPI_ISL_443379 | Department of Pathology, University of Cambridge | Wellcome Sanger Institute for the COVID-19 Genomics UK Consortium |
| EPI_ISL_443380 | Department of Pathology, University of Cambridge | Wellcome Sanger Institute for the COVID-19 Genomics UK Consortium |
| EPI_ISL_443381 | Department of Pathology, University of Cambridge | Wellcome Sanger Institute for the COVID-19 Genomics UK Consortium |
| EPI_ISL_443382 | Department of Pathology, University of Cambridge | Wellcome Sanger Institute for the COVID-19 Genomics UK Consortium |
| EPI_ISL_443383 | Department of Pathology, University of Cambridge | Wellcome Sanger Institute for the COVID-19 Genomics UK Consortium |
| EPI_ISL_443384 | Department of Pathology, University of Cambridge | Wellcome Sanger Institute for the COVID-19 Genomics UK Consortium |
| EPI_ISL_443385 | Department of Pathology, University of Cambridge | Wellcome Sanger Institute for the COVID-19 Genomics UK Consortium |
| EPI_ISL_443386 | Department of Pathology, University of Cambridge | Wellcome Sanger Institute for the COVID-19 Genomics UK Consortium |
| EPI_ISL_443387 | Department of Pathology, University of Cambridge | Wellcome Sanger Institute for the COVID-19 Genomics UK Consortium |
| EPI_ISL_443388 | Department of Pathology, University of Cambridge | Wellcome Sanger Institute for the COVID-19 Genomics UK Consortium |
| EPI_ISL_443389 | Department of Pathology, University of Cambridge | Wellcome Sanger Institute for the COVID-19 Genomics UK Consortium |

| EPI_Accession_ID | Originating Laboratory | Submitting Laboratory |
|---|---|---|
| EPI_ISL_443390 | Department of Pathology, University of Cambridge | Wellcome Sanger Institute for the COVID-19 Genomics UK Consortium |
| EPI_ISL_443391 | Department of Pathology, University of Cambridge | Wellcome Sanger Institute for the COVID-19 Genomics UK Consortium |
| EPI_ISL_443392 | Department of Pathology, University of Cambridge | Wellcome Sanger Institute for the COVID-19 Genomics UK Consortium |
| EPI_ISL_443393 | Department of Pathology, University of Cambridge | Wellcome Sanger Institute for the COVID-19 Genomics UK Consortium |
| EPI_ISL_443394 | Department of Pathology, University of Cambridge | Wellcome Sanger Institute for the COVID-19 Genomics UK Consortium |
| EPI_ISL_443395 | Department of Pathology, University of Cambridge | Wellcome Sanger Institute for the COVID-19 Genomics UK Consortium |
| EPI_ISL_443396 | Department of Pathology, University of Cambridge | Wellcome Sanger Institute for the COVID-19 Genomics UK Consortium |
| EPI_ISL_443397 | Department of Pathology, University of Cambridge | Wellcome Sanger Institute for the COVID-19 Genomics UK Consortium |
| EPI_ISL_443398 | Department of Pathology, University of Cambridge | Wellcome Sanger Institute for the COVID-19 Genomics UK Consortium |
| EPI_ISL_443399 | Department of Pathology, University of Cambridge | Wellcome Sanger Institute for the COVID-19 Genomics UK Consortium |
| EPI_ISL_443400 | Department of Pathology, University of Cambridge | Wellcome Sanger Institute for the COVID-19 Genomics UK Consortium |
| EPI_ISL_443401 | Department of Pathology, University of Cambridge | Wellcome Sanger Institute for the COVID-19 Genomics UK Consortium |
| EPI_ISL_443402 | Department of Pathology, University of Cambridge | Wellcome Sanger Institute for the COVID-19 Genomics UK Consortium |
| EPI_ISL_443403 | Department of Pathology, University of Cambridge | Wellcome Sanger Institute for the COVID-19 Genomics UK Consortium |
| EPI_ISL_443404 | Department of Pathology, University of Cambridge | Wellcome Sanger Institute for the COVID-19 Genomics UK Consortium |
| EPI_ISL_443405 | Department of Pathology, University of Cambridge | Wellcome Sanger Institute for the COVID-19 Genomics UK Consortium |

| EPI_Accession_ID | Originating Laboratory | Submitting Laboratory |
|---|---|---|
| EPI_ISL_443406 | Department of Pathology, University of Cambridge | Wellcome Sanger Institute for the COVID-19 Genomics UK Consortium |
| EPI_ISL_443407 | Department of Pathology, University of Cambridge | Wellcome Sanger Institute for the COVID-19 Genomics UK Consortium |
| EPI_ISL_443408 | Department of Pathology, University of Cambridge | Wellcome Sanger Institute for the COVID-19 Genomics UK Consortium |
| EPI_ISL_443409 | Department of Pathology, University of Cambridge | Wellcome Sanger Institute for the COVID-19 Genomics UK Consortium |
| EPI_ISL_443410 | Department of Pathology, University of Cambridge | Wellcome Sanger Institute for the COVID-19 Genomics UK Consortium |
| EPI_ISL_443411 | Department of Pathology, University of Cambridge | Wellcome Sanger Institute for the COVID-19 Genomics UK Consortium |
| EPI_ISL_443412 | Department of Pathology, University of Cambridge | Wellcome Sanger Institute for the COVID-19 Genomics UK Consortium |
| EPI_ISL_443413 | Department of Pathology, University of Cambridge | Wellcome Sanger Institute for the COVID-19 Genomics UK Consortium |
| EPI_ISL_443414 | Department of Pathology, University of Cambridge | Wellcome Sanger Institute for the COVID-19 Genomics UK Consortium |
| EPI_ISL_443415 | Department of Pathology, University of Cambridge | Wellcome Sanger Institute for the COVID-19 Genomics UK Consortium |
| EPI_ISL_443416 | Department of Pathology, University of Cambridge | Wellcome Sanger Institute for the COVID-19 Genomics UK Consortium |
| EPI_ISL_443417 | Department of Pathology, University of Cambridge | Wellcome Sanger Institute for the COVID-19 Genomics UK Consortium |
| EPI_ISL_443418 | Department of Pathology, University of Cambridge | Wellcome Sanger Institute for the COVID-19 Genomics UK Consortium |
| EPI_ISL_443419 | Department of Pathology, University of Cambridge | Wellcome Sanger Institute for the COVID-19 Genomics UK Consortium |
| EPI_ISL_443420 | Department of Pathology, University of Cambridge | Wellcome Sanger Institute for the COVID-19 Genomics UK Consortium |
| EPI_ISL_443421 | Department of Pathology, University of Cambridge | Wellcome Sanger Institute for the COVID-19 Genomics UK Consortium |

| EPI_Accession_ID | Originating Laboratory | Submitting Laboratory |
| --- | --- | --- |
| EPI_ISL_443422 | Department of Pathology, University of Cambridge | Wellcome Sanger Institute for the COVID-19 Genomics UK Consortium |
| EPI_ISL_443423 | Department of Pathology, University of Cambridge | Wellcome Sanger Institute for the COVID-19 Genomics UK Consortium |
| EPI_ISL_443424 | Department of Pathology, University of Cambridge | Wellcome Sanger Institute for the COVID-19 Genomics UK Consortium |
| EPI_ISL_443425 | Department of Pathology, University of Cambridge | Wellcome Sanger Institute for the COVID-19 Genomics UK Consortium |
| EPI_ISL_443426 | Department of Pathology, University of Cambridge | Wellcome Sanger Institute for the COVID-19 Genomics UK Consortium |
| EPI_ISL_443427 | Department of Pathology, University of Cambridge | Wellcome Sanger Institute for the COVID-19 Genomics UK Consortium |
| EPI_ISL_443428 | Department of Pathology, University of Cambridge | Wellcome Sanger Institute for the COVID-19 Genomics UK Consortium |
| EPI_ISL_443429 | Department of Pathology, University of Cambridge | Wellcome Sanger Institute for the COVID-19 Genomics UK Consortium |
| EPI_ISL_443430 | Department of Pathology, University of Cambridge | Wellcome Sanger Institute for the COVID-19 Genomics UK Consortium |
| EPI_ISL_443431 | Department of Pathology, University of Cambridge | Wellcome Sanger Institute for the COVID-19 Genomics UK Consortium |
| EPI_ISL_443432 | Department of Pathology, University of Cambridge | Wellcome Sanger Institute for the COVID-19 Genomics UK Consortium |
| EPI_ISL_443433 | Department of Pathology, University of Cambridge | Wellcome Sanger Institute for the COVID-19 Genomics UK Consortium |
| EPI_ISL_443434 | Department of Pathology, University of Cambridge | Wellcome Sanger Institute for the COVID-19 Genomics UK Consortium |
| EPI_ISL_443435 | Department of Pathology, University of Cambridge | Wellcome Sanger Institute for the COVID-19 Genomics UK Consortium |
| EPI_ISL_443436 | Department of Pathology, University of Cambridge | Wellcome Sanger Institute for the COVID-19 Genomics UK Consortium |
| EPI_ISL_443437 | Department of Pathology, University of Cambridge | Wellcome Sanger Institute for the COVID-19 Genomics UK Consortium |

| EPI_Accession_ID | Originating Laboratory | Submitting Laboratory |
|---|---|---|
| EPI_ISL_443438 | Department of Pathology, University of Cambridge | Wellcome Sanger Institute for the COVID-19 Genomics UK Consortium |
| EPI_ISL_443439 | Department of Pathology, University of Cambridge | Wellcome Sanger Institute for the COVID-19 Genomics UK Consortium |
| EPI_ISL_443440 | Department of Pathology, University of Cambridge | Wellcome Sanger Institute for the COVID-19 Genomics UK Consortium |
| EPI_ISL_443441 | Department of Pathology, University of Cambridge | Wellcome Sanger Institute for the COVID-19 Genomics UK Consortium |
| EPI_ISL_443442 | Department of Pathology, University of Cambridge | Wellcome Sanger Institute for the COVID-19 Genomics UK Consortium |
| EPI_ISL_443443 | Department of Pathology, University of Cambridge | Wellcome Sanger Institute for the COVID-19 Genomics UK Consortium |
| EPI_ISL_443444 | Department of Pathology, University of Cambridge | Wellcome Sanger Institute for the COVID-19 Genomics UK Consortium |
| EPI_ISL_443445 | Department of Pathology, University of Cambridge | Wellcome Sanger Institute for the COVID-19 Genomics UK Consortium |
| EPI_ISL_443446 | Department of Pathology, University of Cambridge | Wellcome Sanger Institute for the COVID-19 Genomics UK Consortium |
| EPI_ISL_443447 | Department of Pathology, University of Cambridge | Wellcome Sanger Institute for the COVID-19 Genomics UK Consortium |
| EPI_ISL_443448 | Department of Pathology, University of Cambridge | Wellcome Sanger Institute for the COVID-19 Genomics UK Consortium |
| EPI_ISL_443449 | Department of Pathology, University of Cambridge | Wellcome Sanger Institute for the COVID-19 Genomics UK Consortium |
| EPI_ISL_443450 | Department of Pathology, University of Cambridge | Wellcome Sanger Institute for the COVID-19 Genomics UK Consortium |
| EPI_ISL_443451 | Department of Pathology, University of Cambridge | Wellcome Sanger Institute for the COVID-19 Genomics UK Consortium |
| EPI_ISL_443452 | Department of Pathology, University of Cambridge | Wellcome Sanger Institute for the COVID-19 Genomics UK Consortium |
| EPI_ISL_443453 | Department of Pathology, University of Cambridge | Wellcome Sanger Institute for the COVID-19 Genomics UK Consortium |

| EPI_Accession_ID | Originating Laboratory | Submitting Laboratory |
|---|---|---|
| EPI_ISL_443454 | Department of Pathology, University of Cambridge | Wellcome Sanger Institute for the COVID-19 Genomics UK Consortium |
| EPI_ISL_443455 | Department of Pathology, University of Cambridge | Wellcome Sanger Institute for the COVID-19 Genomics UK Consortium |
| EPI_ISL_443456 | Department of Pathology, University of Cambridge | Wellcome Sanger Institute for the COVID-19 Genomics UK Consortium |
| EPI_ISL_443457 | Department of Pathology, University of Cambridge | Wellcome Sanger Institute for the COVID-19 Genomics UK Consortium |
| EPI_ISL_443458 | Department of Pathology, University of Cambridge | Wellcome Sanger Institute for the COVID-19 Genomics UK Consortium |
| EPI_ISL_443459 | Department of Pathology, University of Cambridge | Wellcome Sanger Institute for the COVID-19 Genomics UK Consortium |
| EPI_ISL_443460 | Department of Pathology, University of Cambridge | Wellcome Sanger Institute for the COVID-19 Genomics UK Consortium |
| EPI_ISL_443461 | Department of Pathology, University of Cambridge | Wellcome Sanger Institute for the COVID-19 Genomics UK Consortium |
| EPI_ISL_443462 | Department of Pathology, University of Cambridge | Wellcome Sanger Institute for the COVID-19 Genomics UK Consortium |
| EPI_ISL_443463 | Department of Pathology, University of Cambridge | Wellcome Sanger Institute for the COVID-19 Genomics UK Consortium |
| EPI_ISL_443464 | Department of Pathology, University of Cambridge | Wellcome Sanger Institute for the COVID-19 Genomics UK Consortium |
| EPI_ISL_443465 | Department of Pathology, University of Cambridge | Wellcome Sanger Institute for the COVID-19 Genomics UK Consortium |
| EPI_ISL_443466 | Department of Pathology, University of Cambridge | Wellcome Sanger Institute for the COVID-19 Genomics UK Consortium |
| EPI_ISL_443467 | Department of Pathology, University of Cambridge | Wellcome Sanger Institute for the COVID-19 Genomics UK Consortium |
| EPI_ISL_443468 | Department of Pathology, University of Cambridge | Wellcome Sanger Institute for the COVID-19 Genomics UK Consortium |
| EPI_ISL_443469 | Department of Pathology, University of Cambridge | Wellcome Sanger Institute for the COVID-19 Genomics UK Consortium |

| EPI_Accession_ID | Originating Laboratory | Submitting Laboratory |
|---|---|---|
| EPI_ISL_443470 | Department of Pathology, University of Cambridge | Wellcome Sanger Institute for the COVID-19 Genomics UK Consortium |
| EPI_ISL_443471 | Department of Pathology, University of Cambridge | Wellcome Sanger Institute for the COVID-19 Genomics UK Consortium |
| EPI_ISL_443472 | Department of Pathology, University of Cambridge | Wellcome Sanger Institute for the COVID-19 Genomics UK Consortium |
| EPI_ISL_443473 | Department of Pathology, University of Cambridge | Wellcome Sanger Institute for the COVID-19 Genomics UK Consortium |
| EPI_ISL_443474 | Department of Pathology, University of Cambridge | Wellcome Sanger Institute for the COVID-19 Genomics UK Consortium |
| EPI_ISL_443475 | Department of Pathology, University of Cambridge | Wellcome Sanger Institute for the COVID-19 Genomics UK Consortium |
| EPI_ISL_443476 | Department of Pathology, University of Cambridge | Wellcome Sanger Institute for the COVID-19 Genomics UK Consortium |
| EPI_ISL_443477 | Department of Pathology, University of Cambridge | Wellcome Sanger Institute for the COVID-19 Genomics UK Consortium |
| EPI_ISL_443478 | Department of Pathology, University of Cambridge | Wellcome Sanger Institute for the COVID-19 Genomics UK Consortium |
| EPI_ISL_443479 | Department of Pathology, University of Cambridge | Wellcome Sanger Institute for the COVID-19 Genomics UK Consortium |
| EPI_ISL_443480 | Department of Pathology, University of Cambridge | Wellcome Sanger Institute for the COVID-19 Genomics UK Consortium |
| EPI_ISL_443481 | Department of Pathology, University of Cambridge | Wellcome Sanger Institute for the COVID-19 Genomics UK Consortium |
| EPI_ISL_443482 | Department of Pathology, University of Cambridge | Wellcome Sanger Institute for the COVID-19 Genomics UK Consortium |
| EPI_ISL_443483 | Department of Pathology, University of Cambridge | Wellcome Sanger Institute for the COVID-19 Genomics UK Consortium |
| EPI_ISL_443484 | Department of Pathology, University of Cambridge | Wellcome Sanger Institute for the COVID-19 Genomics UK Consortium |
| EPI_ISL_443485 | Department of Pathology, University of Cambridge | Wellcome Sanger Institute for the COVID-19 Genomics UK Consortium |

| EPI_Accession_ID | Originating Laboratory | Submitting Laboratory |
| --- | --- | --- |
| EPI_ISL_443486 | Department of Pathology, University of Cambridge | Wellcome Sanger Institute for the COVID-19 Genomics UK Consortium |
| EPI_ISL_443487 | Department of Pathology, University of Cambridge | Wellcome Sanger Institute for the COVID-19 Genomics UK Consortium |
| EPI_ISL_443488 | Department of Pathology, University of Cambridge | Wellcome Sanger Institute for the COVID-19 Genomics UK Consortium |
| EPI_ISL_443489 | Department of Pathology, University of Cambridge | Wellcome Sanger Institute for the COVID-19 Genomics UK Consortium |
| EPI_ISL_443490 | Department of Pathology, University of Cambridge | Wellcome Sanger Institute for the COVID-19 Genomics UK Consortium |
| EPI_ISL_443491 | Department of Pathology, University of Cambridge | Wellcome Sanger Institute for the COVID-19 Genomics UK Consortium |
| EPI_ISL_443492 | Department of Pathology, University of Cambridge | Wellcome Sanger Institute for the COVID-19 Genomics UK Consortium |
| EPI_ISL_443493 | Department of Pathology, University of Cambridge | Wellcome Sanger Institute for the COVID-19 Genomics UK Consortium |
| EPI_ISL_443494 | Department of Pathology, University of Cambridge | Wellcome Sanger Institute for the COVID-19 Genomics UK Consortium |
| EPI_ISL_443495 | Department of Pathology, University of Cambridge | Wellcome Sanger Institute for the COVID-19 Genomics UK Consortium |
| EPI_ISL_443496 | Department of Pathology, University of Cambridge | Wellcome Sanger Institute for the COVID-19 Genomics UK Consortium |
| EPI_ISL_443497 | Department of Pathology, University of Cambridge | Wellcome Sanger Institute for the COVID-19 Genomics UK Consortium |
| EPI_ISL_443498 | Department of Pathology, University of Cambridge | Wellcome Sanger Institute for the COVID-19 Genomics UK Consortium |
| EPI_ISL_443499 | Department of Pathology, University of Cambridge | Wellcome Sanger Institute for the COVID-19 Genomics UK Consortium |
| EPI_ISL_443500 | Department of Pathology, University of Cambridge | Wellcome Sanger Institute for the COVID-19 Genomics UK Consortium |
| EPI_ISL_443501 | Department of Pathology, University of Cambridge | Wellcome Sanger Institute for the COVID-19 Genomics UK Consortium |

| EPI_Accession_ID | Originating Laboratory | Submitting Laboratory |
|---|---|---|
| EPI_ISL_443502 | Department of Pathology, University of Cambridge | Wellcome Sanger Institute for the COVID-19 Genomics UK Consortium |
| EPI_ISL_443503 | Department of Pathology, University of Cambridge | Wellcome Sanger Institute for the COVID-19 Genomics UK Consortium |
| EPI_ISL_443504 | Department of Pathology, University of Cambridge | Wellcome Sanger Institute for the COVID-19 Genomics UK Consortium |
| EPI_ISL_443505 | Department of Pathology, University of Cambridge | Wellcome Sanger Institute for the COVID-19 Genomics UK Consortium |
| EPI_ISL_443506 | Department of Pathology, University of Cambridge | Wellcome Sanger Institute for the COVID-19 Genomics UK Consortium |
| EPI_ISL_443507 | Department of Pathology, University of Cambridge | Wellcome Sanger Institute for the COVID-19 Genomics UK Consortium |
| EPI_ISL_443508 | Department of Pathology, University of Cambridge | Wellcome Sanger Institute for the COVID-19 Genomics UK Consortium |
| EPI_ISL_443509 | Department of Pathology, University of Cambridge | Wellcome Sanger Institute for the COVID-19 Genomics UK Consortium |
| EPI_ISL_443510 | Department of Pathology, University of Cambridge | Wellcome Sanger Institute for the COVID-19 Genomics UK Consortium |
| EPI_ISL_443511 | Department of Pathology, University of Cambridge | Wellcome Sanger Institute for the COVID-19 Genomics UK Consortium |
| EPI_ISL_443512 | Department of Pathology, University of Cambridge | Wellcome Sanger Institute for the COVID-19 Genomics UK Consortium |
| EPI_ISL_443513 | Department of Pathology, University of Cambridge | Wellcome Sanger Institute for the COVID-19 Genomics UK Consortium |
| EPI_ISL_443514 | Department of Pathology, University of Cambridge | Wellcome Sanger Institute for the COVID-19 Genomics UK Consortium |
| EPI_ISL_443515 | Department of Pathology, University of Cambridge | Wellcome Sanger Institute for the COVID-19 Genomics UK Consortium |
| EPI_ISL_443516 | Department of Pathology, University of Cambridge | Wellcome Sanger Institute for the COVID-19 Genomics UK Consortium |
| EPI_ISL_443517 | Department of Pathology, University of Cambridge | Wellcome Sanger Institute for the COVID-19 Genomics UK Consortium |

| EPI_Accession_ID | Originating Laboratory | Submitting Laboratory |
|---|---|---|
| EPI_ISL_443518 | Department of Pathology, University of Cambridge | Wellcome Sanger Institute for the COVID-19 Genomics UK Consortium |
| EPI_ISL_443519 | Department of Pathology, University of Cambridge | Wellcome Sanger Institute for the COVID-19 Genomics UK Consortium |
| EPI_ISL_443520 | Department of Pathology, University of Cambridge | Wellcome Sanger Institute for the COVID-19 Genomics UK Consortium |
| EPI_ISL_443521 | Department of Pathology, University of Cambridge | Wellcome Sanger Institute for the COVID-19 Genomics UK Consortium |
| EPI_ISL_443522 | Department of Pathology, University of Cambridge | Wellcome Sanger Institute for the COVID-19 Genomics UK Consortium |
| EPI_ISL_443523 | Department of Pathology, University of Cambridge | Wellcome Sanger Institute for the COVID-19 Genomics UK Consortium |
| EPI_ISL_443524 | Department of Pathology, University of Cambridge | Wellcome Sanger Institute for the COVID-19 Genomics UK Consortium |
| EPI_ISL_443525 | Department of Pathology, University of Cambridge | Wellcome Sanger Institute for the COVID-19 Genomics UK Consortium |
| EPI_ISL_443526 | Department of Pathology, University of Cambridge | Wellcome Sanger Institute for the COVID-19 Genomics UK Consortium |
| EPI_ISL_443527 | Department of Pathology, University of Cambridge | Wellcome Sanger Institute for the COVID-19 Genomics UK Consortium |
| EPI_ISL_443528 | Department of Pathology, University of Cambridge | Wellcome Sanger Institute for the COVID-19 Genomics UK Consortium |
| EPI_ISL_443529 | Department of Pathology, University of Cambridge | Wellcome Sanger Institute for the COVID-19 Genomics UK Consortium |
| EPI_ISL_443530 | Department of Pathology, University of Cambridge | Wellcome Sanger Institute for the COVID-19 Genomics UK Consortium |
| EPI_ISL_443531 | Department of Pathology, University of Cambridge | Wellcome Sanger Institute for the COVID-19 Genomics UK Consortium |
| EPI_ISL_443532 | Department of Pathology, University of Cambridge | Wellcome Sanger Institute for the COVID-19 Genomics UK Consortium |
| EPI_ISL_443533 | Department of Pathology, University of Cambridge | Wellcome Sanger Institute for the COVID-19 Genomics UK Consortium |

| EPI_Accession_ID | Originating Laboratory | Submitting Laboratory |
|---|---|---|
| EPI_ISL_443534 | Department of Pathology, University of Cambridge | Wellcome Sanger Institute for the COVID-19 Genomics UK Consortium |
| EPI_ISL_443535 | Department of Pathology, University of Cambridge | Wellcome Sanger Institute for the COVID-19 Genomics UK Consortium |
| EPI_ISL_443536 | Department of Pathology, University of Cambridge | Wellcome Sanger Institute for the COVID-19 Genomics UK Consortium |
| EPI_ISL_443537 | Department of Pathology, University of Cambridge | Wellcome Sanger Institute for the COVID-19 Genomics UK Consortium |
| EPI_ISL_443538 | Department of Pathology, University of Cambridge | Wellcome Sanger Institute for the COVID-19 Genomics UK Consortium |
| EPI_ISL_443539 | Department of Pathology, University of Cambridge | Wellcome Sanger Institute for the COVID-19 Genomics UK Consortium |
| EPI_ISL_443540 | Department of Pathology, University of Cambridge | Wellcome Sanger Institute for the COVID-19 Genomics UK Consortium |
| EPI_ISL_443541 | Department of Pathology, University of Cambridge | Wellcome Sanger Institute for the COVID-19 Genomics UK Consortium |
| EPI_ISL_443542 | Department of Pathology, University of Cambridge | Wellcome Sanger Institute for the COVID-19 Genomics UK Consortium |
| EPI_ISL_443543 | Department of Pathology, University of Cambridge | Wellcome Sanger Institute for the COVID-19 Genomics UK Consortium |
| EPI_ISL_443544 | Department of Pathology, University of Cambridge | Wellcome Sanger Institute for the COVID-19 Genomics UK Consortium |
| EPI_ISL_443545 | Department of Pathology, University of Cambridge | Wellcome Sanger Institute for the COVID-19 Genomics UK Consortium |
| EPI_ISL_443546 | Department of Pathology, University of Cambridge | Wellcome Sanger Institute for the COVID-19 Genomics UK Consortium |
| EPI_ISL_443547 | Department of Pathology, University of Cambridge | Wellcome Sanger Institute for the COVID-19 Genomics UK Consortium |
| EPI_ISL_443548 | Department of Pathology, University of Cambridge | Wellcome Sanger Institute for the COVID-19 Genomics UK Consortium |
| EPI_ISL_443549 | Department of Pathology, University of Cambridge | Wellcome Sanger Institute for the COVID-19 Genomics UK Consortium |

| EPI_Accession_ID | Originating Laboratory | Submitting Laboratory |
|---|---|---|
| EPI_ISL_443550 | Department of Pathology, University of Cambridge | Wellcome Sanger Institute for the COVID-19 Genomics UK Consortium |
| EPI_ISL_443551 | Department of Pathology, University of Cambridge | Wellcome Sanger Institute for the COVID-19 Genomics UK Consortium |
| EPI_ISL_443552 | Department of Pathology, University of Cambridge | Wellcome Sanger Institute for the COVID-19 Genomics UK Consortium |
| EPI_ISL_443553 | Department of Pathology, University of Cambridge | Wellcome Sanger Institute for the COVID-19 Genomics UK Consortium |
| EPI_ISL_443554 | Department of Pathology, University of Cambridge | Wellcome Sanger Institute for the COVID-19 Genomics UK Consortium |
| EPI_ISL_443555 | Department of Pathology, University of Cambridge | Wellcome Sanger Institute for the COVID-19 Genomics UK Consortium |
| EPI_ISL_443556 | Department of Pathology, University of Cambridge | Wellcome Sanger Institute for the COVID-19 Genomics UK Consortium |
| EPI_ISL_443557 | Department of Pathology, University of Cambridge | Wellcome Sanger Institute for the COVID-19 Genomics UK Consortium |
| EPI_ISL_443558 | Department of Pathology, University of Cambridge | Wellcome Sanger Institute for the COVID-19 Genomics UK Consortium |
| EPI_ISL_443559 | Department of Pathology, University of Cambridge | Wellcome Sanger Institute for the COVID-19 Genomics UK Consortium |
| EPI_ISL_443560 | Department of Pathology, University of Cambridge | Wellcome Sanger Institute for the COVID-19 Genomics UK Consortium |
| EPI_ISL_443561 | Department of Pathology, University of Cambridge | Wellcome Sanger Institute for the COVID-19 Genomics UK Consortium |
| EPI_ISL_443562 | Department of Pathology, University of Cambridge | Wellcome Sanger Institute for the COVID-19 Genomics UK Consortium |
| EPI_ISL_443563 | Department of Pathology, University of Cambridge | Wellcome Sanger Institute for the COVID-19 Genomics UK Consortium |
| EPI_ISL_443564 | Department of Pathology, University of Cambridge | Wellcome Sanger Institute for the COVID-19 Genomics UK Consortium |
| EPI_ISL_443565 | Department of Pathology, University of Cambridge | Wellcome Sanger Institute for the COVID-19 Genomics UK Consortium |

| EPI_Accession_ID | Originating Laboratory | Submitting Laboratory |
| --- | --- | --- |
| EPI_ISL_443566 | Department of Pathology, University of Cambridge | Wellcome Sanger Institute for the COVID-19 Genomics UK Consortium |
| EPI_ISL_443567 | Department of Pathology, University of Cambridge | Wellcome Sanger Institute for the COVID-19 Genomics UK Consortium |
| EPI_ISL_443568 | Department of Pathology, University of Cambridge | Wellcome Sanger Institute for the COVID-19 Genomics UK Consortium |
| EPI_ISL_443569 | Department of Pathology, University of Cambridge | Wellcome Sanger Institute for the COVID-19 Genomics UK Consortium |
| EPI_ISL_443570 | Department of Pathology, University of Cambridge | Wellcome Sanger Institute for the COVID-19 Genomics UK Consortium |
| EPI_ISL_443571 | Department of Pathology, University of Cambridge | Wellcome Sanger Institute for the COVID-19 Genomics UK Consortium |
| EPI_ISL_443572 | Department of Pathology, University of Cambridge | Wellcome Sanger Institute for the COVID-19 Genomics UK Consortium |
| EPI_ISL_443573 | Department of Pathology, University of Cambridge | Wellcome Sanger Institute for the COVID-19 Genomics UK Consortium |
| EPI_ISL_443574 | Department of Pathology, University of Cambridge | Wellcome Sanger Institute for the COVID-19 Genomics UK Consortium |
| EPI_ISL_443575 | Department of Pathology, University of Cambridge | Wellcome Sanger Institute for the COVID-19 Genomics UK Consortium |
| EPI_ISL_443576 | Department of Pathology, University of Cambridge | Wellcome Sanger Institute for the COVID-19 Genomics UK Consortium |
| EPI_ISL_443577 | Department of Pathology, University of Cambridge | Wellcome Sanger Institute for the COVID-19 Genomics UK Consortium |
| EPI_ISL_443578 | Department of Pathology, University of Cambridge | Wellcome Sanger Institute for the COVID-19 Genomics UK Consortium |
| EPI_ISL_443579 | Department of Pathology, University of Cambridge | Wellcome Sanger Institute for the COVID-19 Genomics UK Consortium |
| EPI_ISL_443580 | Department of Pathology, University of Cambridge | Wellcome Sanger Institute for the COVID-19 Genomics UK Consortium |
| EPI_ISL_443581 | Department of Pathology, University of Cambridge | Wellcome Sanger Institute for the COVID-19 Genomics UK Consortium |

| EPI_Accession_ID | Originating Laboratory | Submitting Laboratory |
|---|---|---|
| EPI_ISL_443582 | Department of Pathology, University of Cambridge | Wellcome Sanger Institute for the COVID-19 Genomics UK Consortium |
| EPI_ISL_443583 | Department of Pathology, University of Cambridge | Wellcome Sanger Institute for the COVID-19 Genomics UK Consortium |
| EPI_ISL_443584 | Department of Pathology, University of Cambridge | Wellcome Sanger Institute for the COVID-19 Genomics UK Consortium |
| EPI_ISL_443585 | Department of Pathology, University of Cambridge | Wellcome Sanger Institute for the COVID-19 Genomics UK Consortium |
| EPI_ISL_443586 | Department of Pathology, University of Cambridge | Wellcome Sanger Institute for the COVID-19 Genomics UK Consortium |
| EPI_ISL_443587 | Department of Pathology, University of Cambridge | Wellcome Sanger Institute for the COVID-19 Genomics UK Consortium |
| EPI_ISL_443588 | Department of Pathology, University of Cambridge | Wellcome Sanger Institute for the COVID-19 Genomics UK Consortium |
| EPI_ISL_443589 | Department of Pathology, University of Cambridge | Wellcome Sanger Institute for the COVID-19 Genomics UK Consortium |
| EPI_ISL_443590 | Department of Pathology, University of Cambridge | Wellcome Sanger Institute for the COVID-19 Genomics UK Consortium |
| EPI_ISL_443591 | Department of Pathology, University of Cambridge | Wellcome Sanger Institute for the COVID-19 Genomics UK Consortium |
| EPI_ISL_443592 | Department of Pathology, University of Cambridge | Wellcome Sanger Institute for the COVID-19 Genomics UK Consortium |
| EPI_ISL_443593 | Department of Pathology, University of Cambridge | Wellcome Sanger Institute for the COVID-19 Genomics UK Consortium |
| EPI_ISL_443594 | Department of Pathology, University of Cambridge | Wellcome Sanger Institute for the COVID-19 Genomics UK Consortium |
| EPI_ISL_443595 | Department of Pathology, University of Cambridge | Wellcome Sanger Institute for the COVID-19 Genomics UK Consortium |
| EPI_ISL_443596 | Department of Pathology, University of Cambridge | Wellcome Sanger Institute for the COVID-19 Genomics UK Consortium |
| EPI_ISL_443597 | Department of Pathology, University of Cambridge | Wellcome Sanger Institute for the COVID-19 Genomics UK Consortium |

| EPI_Accession_ID | Originating Laboratory | Submitting Laboratory |
|---|---|---|
| EPI_ISL_443598 | Department of Pathology, University of Cambridge | Wellcome Sanger Institute for the COVID-19 Genomics UK Consortium |
| EPI_ISL_443599 | Department of Pathology, University of Cambridge | Wellcome Sanger Institute for the COVID-19 Genomics UK Consortium |
| EPI_ISL_443600 | Department of Pathology, University of Cambridge | Wellcome Sanger Institute for the COVID-19 Genomics UK Consortium |
| EPI_ISL_443601 | Department of Pathology, University of Cambridge | Wellcome Sanger Institute for the COVID-19 Genomics UK Consortium |
| EPI_ISL_443602 | Department of Pathology, University of Cambridge | Wellcome Sanger Institute for the COVID-19 Genomics UK Consortium |
| EPI_ISL_443603 | Department of Pathology, University of Cambridge | Wellcome Sanger Institute for the COVID-19 Genomics UK Consortium |
| EPI_ISL_443604 | Department of Pathology, University of Cambridge | Wellcome Sanger Institute for the COVID-19 Genomics UK Consortium |
| EPI_ISL_443605 | Department of Pathology, University of Cambridge | Wellcome Sanger Institute for the COVID-19 Genomics UK Consortium |
| EPI_ISL_443606 | Department of Pathology, University of Cambridge | Wellcome Sanger Institute for the COVID-19 Genomics UK Consortium |
| EPI_ISL_443607 | Department of Pathology, University of Cambridge | Wellcome Sanger Institute for the COVID-19 Genomics UK Consortium |
| EPI_ISL_443608 | Department of Pathology, University of Cambridge | Wellcome Sanger Institute for the COVID-19 Genomics UK Consortium |
| EPI_ISL_443609 | Department of Pathology, University of Cambridge | Wellcome Sanger Institute for the COVID-19 Genomics UK Consortium |
| EPI_ISL_443610 | Department of Pathology, University of Cambridge | Wellcome Sanger Institute for the COVID-19 Genomics UK Consortium |
| EPI_ISL_443611 | Department of Pathology, University of Cambridge | Wellcome Sanger Institute for the COVID-19 Genomics UK Consortium |
| EPI_ISL_443612 | Department of Pathology, University of Cambridge | Wellcome Sanger Institute for the COVID-19 Genomics UK Consortium |
| EPI_ISL_443613 | Department of Pathology, University of Cambridge | Wellcome Sanger Institute for the COVID-19 Genomics UK Consortium |

| EPI_Accession_ID | Originating Laboratory | Submitting Laboratory |
|---|---|---|
| EPI_ISL_443614 | Department of Pathology, University of Cambridge | Wellcome Sanger Institute for the COVID-19 Genomics UK Consortium |
| EPI_ISL_443615 | Department of Pathology, University of Cambridge | Wellcome Sanger Institute for the COVID-19 Genomics UK Consortium |
| EPI_ISL_443616 | Department of Pathology, University of Cambridge | Wellcome Sanger Institute for the COVID-19 Genomics UK Consortium |
| EPI_ISL_443617 | Department of Pathology, University of Cambridge | Wellcome Sanger Institute for the COVID-19 Genomics UK Consortium |
| EPI_ISL_443618 | Department of Pathology, University of Cambridge | Wellcome Sanger Institute for the COVID-19 Genomics UK Consortium |
| EPI_ISL_443619 | Department of Pathology, University of Cambridge | Wellcome Sanger Institute for the COVID-19 Genomics UK Consortium |
| EPI_ISL_443620 | Department of Pathology, University of Cambridge | Wellcome Sanger Institute for the COVID-19 Genomics UK Consortium |
| EPI_ISL_443621 | Department of Pathology, University of Cambridge | Wellcome Sanger Institute for the COVID-19 Genomics UK Consortium |
| EPI_ISL_443622 | Department of Pathology, University of Cambridge | Wellcome Sanger Institute for the COVID-19 Genomics UK Consortium |
| EPI_ISL_443623 | Department of Pathology, University of Cambridge | Wellcome Sanger Institute for the COVID-19 Genomics UK Consortium |
| EPI_ISL_443624 | Department of Pathology, University of Cambridge | Wellcome Sanger Institute for the COVID-19 Genomics UK Consortium |
| EPI_ISL_443625 | Department of Pathology, University of Cambridge | Wellcome Sanger Institute for the COVID-19 Genomics UK Consortium |
| EPI_ISL_443626 | Department of Pathology, University of Cambridge | Wellcome Sanger Institute for the COVID-19 Genomics UK Consortium |
| EPI_ISL_443627 | Department of Pathology, University of Cambridge | Wellcome Sanger Institute for the COVID-19 Genomics UK Consortium |
| EPI_ISL_443628 | Department of Pathology, University of Cambridge | Wellcome Sanger Institute for the COVID-19 Genomics UK Consortium |
| EPI_ISL_443629 | Department of Pathology, University of Cambridge | Wellcome Sanger Institute for the COVID-19 Genomics UK Consortium |

| EPI_Accession_ID | Originating Laboratory | Submitting Laboratory |
|---|---|---|
| EPI_ISL_443630 | Department of Pathology, University of Cambridge | Wellcome Sanger Institute for the COVID-19 Genomics UK Consortium |
| EPI_ISL_443631 | Department of Pathology, University of Cambridge | Wellcome Sanger Institute for the COVID-19 Genomics UK Consortium |
| EPI_ISL_443632 | Department of Pathology, University of Cambridge | Wellcome Sanger Institute for the COVID-19 Genomics UK Consortium |
| EPI_ISL_443633 | Department of Pathology, University of Cambridge | Wellcome Sanger Institute for the COVID-19 Genomics UK Consortium |
| EPI_ISL_443634 | Department of Pathology, University of Cambridge | Wellcome Sanger Institute for the COVID-19 Genomics UK Consortium |
| EPI_ISL_443635 | Department of Pathology, University of Cambridge | Wellcome Sanger Institute for the COVID-19 Genomics UK Consortium |
| EPI_ISL_443636 | Department of Pathology, University of Cambridge | Wellcome Sanger Institute for the COVID-19 Genomics UK Consortium |
| EPI_ISL_443637 | Department of Pathology, University of Cambridge | Wellcome Sanger Institute for the COVID-19 Genomics UK Consortium |
| EPI_ISL_443638 | Department of Pathology, University of Cambridge | Wellcome Sanger Institute for the COVID-19 Genomics UK Consortium |
| EPI_ISL_443639 | Department of Pathology, University of Cambridge | Wellcome Sanger Institute for the COVID-19 Genomics UK Consortium |
| EPI_ISL_443640 | Department of Pathology, University of Cambridge | Wellcome Sanger Institute for the COVID-19 Genomics UK Consortium |
| EPI_ISL_443641 | Department of Pathology, University of Cambridge | Wellcome Sanger Institute for the COVID-19 Genomics UK Consortium |
| EPI_ISL_443642 | Department of Pathology, University of Cambridge | Wellcome Sanger Institute for the COVID-19 Genomics UK Consortium |
| EPI_ISL_443643 | Department of Pathology, University of Cambridge | Wellcome Sanger Institute for the COVID-19 Genomics UK Consortium |
| EPI_ISL_443644 | Department of Pathology, University of Cambridge | Wellcome Sanger Institute for the COVID-19 Genomics UK Consortium |
| EPI_ISL_443645 | Department of Pathology, University of Cambridge | Wellcome Sanger Institute for the COVID-19 Genomics UK Consortium |

| EPI_Accession_ID | Originating Laboratory | Submitting Laboratory |
|---|---|---|
| EPI_ISL_443646 | Department of Pathology, University of Cambridge | Wellcome Sanger Institute for the COVID-19 Genomics UK Consortium |
| EPI_ISL_443647 | Department of Pathology, University of Cambridge | Wellcome Sanger Institute for the COVID-19 Genomics UK Consortium |
| EPI_ISL_443648 | Department of Pathology, University of Cambridge | Wellcome Sanger Institute for the COVID-19 Genomics UK Consortium |
| EPI_ISL_443649 | Department of Pathology, University of Cambridge | Wellcome Sanger Institute for the COVID-19 Genomics UK Consortium |
| EPI_ISL_443650 | Department of Pathology, University of Cambridge | Wellcome Sanger Institute for the COVID-19 Genomics UK Consortium |
| EPI_ISL_443651 | Department of Pathology, University of Cambridge | Wellcome Sanger Institute for the COVID-19 Genomics UK Consortium |
| EPI_ISL_443652 | Department of Pathology, University of Cambridge | Wellcome Sanger Institute for the COVID-19 Genomics UK Consortium |
| EPI_ISL_443653 | Department of Pathology, University of Cambridge | Wellcome Sanger Institute for the COVID-19 Genomics UK Consortium |
| EPI_ISL_443654 | Department of Pathology, University of Cambridge | Wellcome Sanger Institute for the COVID-19 Genomics UK Consortium |
| EPI_ISL_443655 | Department of Pathology, University of Cambridge | Wellcome Sanger Institute for the COVID-19 Genomics UK Consortium |
| EPI_ISL_443656 | Department of Pathology, University of Cambridge | Wellcome Sanger Institute for the COVID-19 Genomics UK Consortium |
| EPI_ISL_443657 | Department of Pathology, University of Cambridge | Wellcome Sanger Institute for the COVID-19 Genomics UK Consortium |
| EPI_ISL_443658 | Department of Pathology, University of Cambridge | Wellcome Sanger Institute for the COVID-19 Genomics UK Consortium |
| EPI_ISL_443659 | Department of Pathology, University of Cambridge | Wellcome Sanger Institute for the COVID-19 Genomics UK Consortium |
| EPI_ISL_443660 | Department of Pathology, University of Cambridge | Wellcome Sanger Institute for the COVID-19 Genomics UK Consortium |
| EPI_ISL_443661 | Department of Pathology, University of Cambridge | Wellcome Sanger Institute for the COVID-19 Genomics UK Consortium |

| EPI_Accession_ID | Originating Laboratory | Submitting Laboratory |
|---|---|---|
| EPI_ISL_443662 | Department of Pathology, University of Cambridge | Wellcome Sanger Institute for the COVID-19 Genomics UK Consortium |
| EPI_ISL_443663 | Department of Pathology, University of Cambridge | Wellcome Sanger Institute for the COVID-19 Genomics UK Consortium |
| EPI_ISL_443664 | Department of Pathology, University of Cambridge | Wellcome Sanger Institute for the COVID-19 Genomics UK Consortium |
| EPI_ISL_443665 | Department of Pathology, University of Cambridge | Wellcome Sanger Institute for the COVID-19 Genomics UK Consortium |
| EPI_ISL_443666 | Department of Pathology, University of Cambridge | Wellcome Sanger Institute for the COVID-19 Genomics UK Consortium |
| EPI_ISL_443667 | Department of Pathology, University of Cambridge | Wellcome Sanger Institute for the COVID-19 Genomics UK Consortium |
| EPI_ISL_443668 | Department of Pathology, University of Cambridge | Wellcome Sanger Institute for the COVID-19 Genomics UK Consortium |
| EPI_ISL_443669 | Department of Pathology, University of Cambridge | Wellcome Sanger Institute for the COVID-19 Genomics UK Consortium |
| EPI_ISL_443670 | Department of Pathology, University of Cambridge | Wellcome Sanger Institute for the COVID-19 Genomics UK Consortium |
| EPI_ISL_443671 | Department of Pathology, University of Cambridge | Wellcome Sanger Institute for the COVID-19 Genomics UK Consortium |
| EPI_ISL_443672 | Department of Pathology, University of Cambridge | Wellcome Sanger Institute for the COVID-19 Genomics UK Consortium |
| EPI_ISL_443673 | Department of Pathology, University of Cambridge | Wellcome Sanger Institute for the COVID-19 Genomics UK Consortium |
| EPI_ISL_443674 | Department of Pathology, University of Cambridge | Wellcome Sanger Institute for the COVID-19 Genomics UK Consortium |
| EPI_ISL_443675 | Department of Pathology, University of Cambridge | Wellcome Sanger Institute for the COVID-19 Genomics UK Consortium |
| EPI_ISL_443676 | Department of Pathology, University of Cambridge | Wellcome Sanger Institute for the COVID-19 Genomics UK Consortium |
| EPI_ISL_443677 | Department of Pathology, University of Cambridge | Wellcome Sanger Institute for the COVID-19 Genomics UK Consortium |

| EPI_Accession_ID | Originating Laboratory | Submitting Laboratory |
|---|---|---|
| EPI_ISL_443678 | Department of Pathology, University of Cambridge | Wellcome Sanger Institute for the COVID-19 Genomics UK Consortium |
| EPI_ISL_443679 | Department of Pathology, University of Cambridge | Wellcome Sanger Institute for the COVID-19 Genomics UK Consortium |
| EPI_ISL_443680 | Department of Pathology, University of Cambridge | Wellcome Sanger Institute for the COVID-19 Genomics UK Consortium |
| EPI_ISL_443681 | Department of Pathology, University of Cambridge | Wellcome Sanger Institute for the COVID-19 Genomics UK Consortium |
| EPI_ISL_443682 | Department of Pathology, University of Cambridge | Wellcome Sanger Institute for the COVID-19 Genomics UK Consortium |
| EPI_ISL_443683 | Department of Pathology, University of Cambridge | Wellcome Sanger Institute for the COVID-19 Genomics UK Consortium |
| EPI_ISL_443684 | Department of Pathology, University of Cambridge | Wellcome Sanger Institute for the COVID-19 Genomics UK Consortium |
| EPI_ISL_443685 | Department of Pathology, University of Cambridge | Wellcome Sanger Institute for the COVID-19 Genomics UK Consortium |
| EPI_ISL_443686 | PHE South West Regional Laboratory, National Infection Service | Wellcome Sanger Institute for the COVID-19 Genomics UK Consortium |
| EPI_ISL_443687 | PHE South West Regional Laboratory, National Infection Service | Wellcome Sanger Institute for the COVID-19 Genomics UK Consortium |
| EPI_ISL_443688 | PHE South West Regional Laboratory, National Infection Service | Wellcome Sanger Institute for the COVID-19 Genomics UK Consortium |
| EPI_ISL_443689 | PHE South West Regional Laboratory, National Infection Service | Wellcome Sanger Institute for the COVID-19 Genomics UK Consortium |
| EPI_ISL_443690 | PHE South West Regional Laboratory, National Infection Service | Wellcome Sanger Institute for the COVID-19 Genomics UK Consortium |
| EPI_ISL_443691 | PHE South West Regional Laboratory, National Infection Service | Wellcome Sanger Institute for the COVID-19 Genomics UK Consortium |
| EPI_ISL_443692 | PHE South West Regional Laboratory, National Infection Service | Wellcome Sanger Institute for the COVID-19 Genomics UK Consortium |
| EPI_ISL_443693 | PHE South West Regional Laboratory, National Infection Service | Wellcome Sanger Institute for the COVID-19 Genomics UK Consortium |

| EPI_Accession_ID | Originating Laboratory | Submitting Laboratory |
|---|---|---|
| EPI_ISL_443694 | PHE South West Regional Laboratory, National Infection Service | Wellcome Sanger Institute for the COVID-19 Genomics UK Consortium |
| EPI_ISL_443695 | PHE South West Regional Laboratory, National Infection Service | Wellcome Sanger Institute for the COVID-19 Genomics UK Consortium |
| EPI_ISL_443696 | PHE South West Regional Laboratory, National Infection Service | Wellcome Sanger Institute for the COVID-19 Genomics UK Consortium |
| EPI_ISL_443697 | PHE South West Regional Laboratory, National Infection Service | Wellcome Sanger Institute for the COVID-19 Genomics UK Consortium |
| EPI_ISL_443698 | PHE South West Regional Laboratory, National Infection Service | Wellcome Sanger Institute for the COVID-19 Genomics UK Consortium |
| EPI_ISL_443699 | PHE South West Regional Laboratory, National Infection Service | Wellcome Sanger Institute for the COVID-19 Genomics UK Consortium |
| EPI_ISL_443700 | PHE South West Regional Laboratory, National Infection Service | Wellcome Sanger Institute for the COVID-19 Genomics UK Consortium |
| EPI_ISL_443701 | PHE South West Regional Laboratory, National Infection Service | Wellcome Sanger Institute for the COVID-19 Genomics UK Consortium |
| EPI_ISL_443702 | PHE South West Regional Laboratory, National Infection Service | Wellcome Sanger Institute for the COVID-19 Genomics UK Consortium |
| EPI_ISL_443703 | PHE South West Regional Laboratory, National Infection Service | Wellcome Sanger Institute for the COVID-19 Genomics UK Consortium |
| EPI_ISL_443704 | PHE South West Regional Laboratory, National Infection Service | Wellcome Sanger Institute for the COVID-19 Genomics UK Consortium |
| EPI_ISL_443705 | PHE South West Regional Laboratory, National Infection Service | Wellcome Sanger Institute for the COVID-19 Genomics UK Consortium |
| EPI_ISL_443706 | PHE South West Regional Laboratory, National Infection Service | Wellcome Sanger Institute for the COVID-19 Genomics UK Consortium |
| EPI_ISL_443707 | PHE South West Regional Laboratory, National Infection Service | Wellcome Sanger Institute for the COVID-19 Genomics UK Consortium |
| EPI_ISL_443708 | PHE South West Regional Laboratory, National Infection Service | Wellcome Sanger Institute for the COVID-19 Genomics UK Consortium |
| EPI_ISL_443709 | PHE South West Regional Laboratory, National Infection Service | Wellcome Sanger Institute for the COVID-19 Genomics UK Consortium |

| EPI_Accession_ID | Originating Laboratory | Submitting Laboratory |
|---|---|---|
| EPI_ISL_443710 | PHE South West Regional Laboratory, National Infection Service | Wellcome Sanger Institute for the COVID-19 Genomics UK Consortium |
| EPI_ISL_443711 | PHE South West Regional Laboratory, National Infection Service | Wellcome Sanger Institute for the COVID-19 Genomics UK Consortium |
| EPI_ISL_443712 | PHE South West Regional Laboratory, National Infection Service | Wellcome Sanger Institute for the COVID-19 Genomics UK Consortium |
| EPI_ISL_443713 | PHE South West Regional Laboratory, National Infection Service | Wellcome Sanger Institute for the COVID-19 Genomics UK Consortium |
| EPI_ISL_443714 | PHE South West Regional Laboratory, National Infection Service | Wellcome Sanger Institute for the COVID-19 Genomics UK Consortium |
| EPI_ISL_443715 | PHE South West Regional Laboratory, National Infection Service | Wellcome Sanger Institute for the COVID-19 Genomics UK Consortium |
| EPI_ISL_443716 | PHE South West Regional Laboratory, National Infection Service | Wellcome Sanger Institute for the COVID-19 Genomics UK Consortium |
| EPI_ISL_443717 | PHE South West Regional Laboratory, National Infection Service | Wellcome Sanger Institute for the COVID-19 Genomics UK Consortium |
| EPI_ISL_443718 | PHE South West Regional Laboratory, National Infection Service | Wellcome Sanger Institute for the COVID-19 Genomics UK Consortium |
| EPI_ISL_443719 | PHE South West Regional Laboratory, National Infection Service | Wellcome Sanger Institute for the COVID-19 Genomics UK Consortium |
| EPI_ISL_443720 | PHE South West Regional Laboratory, National Infection Service | Wellcome Sanger Institute for the COVID-19 Genomics UK Consortium |
| EPI_ISL_443721 | PHE South West Regional Laboratory, National Infection Service | Wellcome Sanger Institute for the COVID-19 Genomics UK Consortium |
| EPI_ISL_443722 | PHE South West Regional Laboratory, National Infection Service | Wellcome Sanger Institute for the COVID-19 Genomics UK Consortium |
| EPI_ISL_443723 | PHE South West Regional Laboratory, National Infection Service | Wellcome Sanger Institute for the COVID-19 Genomics UK Consortium |
| EPI_ISL_443724 | PHE South West Regional Laboratory, National Infection Service | Wellcome Sanger Institute for the COVID-19 Genomics UK Consortium |
| EPI_ISL_443725 | PHE South West Regional Laboratory, National Infection Service | Wellcome Sanger Institute for the COVID-19 Genomics UK Consortium |

| EPI_Accession_ID | Originating Laboratory | Submitting Laboratory |
|---|---|---|
| EPI_ISL_443726 | PHE South West Regional Laboratory, National Infection Service | Wellcome Sanger Institute for the COVID-19 Genomics UK Consortium |
| EPI_ISL_443727 | PHE South West Regional Laboratory, National Infection Service | Wellcome Sanger Institute for the COVID-19 Genomics UK Consortium |
| EPI_ISL_443728 | PHE South West Regional Laboratory, National Infection Service | Wellcome Sanger Institute for the COVID-19 Genomics UK Consortium |
| EPI_ISL_443729 | PHE South West Regional Laboratory, National Infection Service | Wellcome Sanger Institute for the COVID-19 Genomics UK Consortium |
| EPI_ISL_443730 | PHE South West Regional Laboratory, National Infection Service | Wellcome Sanger Institute for the COVID-19 Genomics UK Consortium |
| EPI_ISL_443731 | PHE South West Regional Laboratory, National Infection Service | Wellcome Sanger Institute for the COVID-19 Genomics UK Consortium |
| EPI_ISL_443732 | PHE South West Regional Laboratory, National Infection Service | Wellcome Sanger Institute for the COVID-19 Genomics UK Consortium |
| EPI_ISL_443733 | PHE South West Regional Laboratory, National Infection Service | Wellcome Sanger Institute for the COVID-19 Genomics UK Consortium |
| EPI_ISL_443734 | PHE South West Regional Laboratory, National Infection Service | Wellcome Sanger Institute for the COVID-19 Genomics UK Consortium |
| EPI_ISL_443735 | PHE South West Regional Laboratory, National Infection Service | Wellcome Sanger Institute for the COVID-19 Genomics UK Consortium |
| EPI_ISL_443736 | PHE South West Regional Laboratory, National Infection Service | Wellcome Sanger Institute for the COVID-19 Genomics UK Consortium |
| EPI_ISL_443737 | PHE South West Regional Laboratory, National Infection Service | Wellcome Sanger Institute for the COVID-19 Genomics UK Consortium |
| EPI_ISL_443738 | PHE South West Regional Laboratory, National Infection Service | Wellcome Sanger Institute for the COVID-19 Genomics UK Consortium |
| EPI_ISL_443739 | PHE South West Regional Laboratory, National Infection Service | Wellcome Sanger Institute for the COVID-19 Genomics UK Consortium |
| EPI_ISL_443740 | PHE South West Regional Laboratory, National Infection Service | Wellcome Sanger Institute for the COVID-19 Genomics UK Consortium |
| EPI_ISL_443741 | PHE South West Regional Laboratory, National Infection Service | Wellcome Sanger Institute for the COVID-19 Genomics UK Consortium |

| EPI_Accession_ID | Originating Laboratory | Submitting Laboratory |
|---|---|---|
| EPI_ISL_443742 | PHE South West Regional Laboratory, National Infection Service | Wellcome Sanger Institute for the COVID-19 Genomics UK Consortium |
| EPI_ISL_443743 | PHE South West Regional Laboratory, National Infection Service | Wellcome Sanger Institute for the COVID-19 Genomics UK Consortium |
| EPI_ISL_443744 | PHE South West Regional Laboratory, National Infection Service | Wellcome Sanger Institute for the COVID-19 Genomics UK Consortium |
| EPI_ISL_443745 | PHE South West Regional Laboratory, National Infection Service | Wellcome Sanger Institute for the COVID-19 Genomics UK Consortium |
| EPI_ISL_443746 | PHE South West Regional Laboratory, National Infection Service | Wellcome Sanger Institute for the COVID-19 Genomics UK Consortium |
| EPI_ISL_443747 | PHE South West Regional Laboratory, National Infection Service | Wellcome Sanger Institute for the COVID-19 Genomics UK Consortium |
| EPI_ISL_443748 | PHE South West Regional Laboratory, National Infection Service | Wellcome Sanger Institute for the COVID-19 Genomics UK Consortium |
| EPI_ISL_443749 | PHE South West Regional Laboratory, National Infection Service | Wellcome Sanger Institute for the COVID-19 Genomics UK Consortium |
| EPI_ISL_443750 | PHE South West Regional Laboratory, National Infection Service | Wellcome Sanger Institute for the COVID-19 Genomics UK Consortium |
| EPI_ISL_443751 | PHE South West Regional Laboratory, National Infection Service | Wellcome Sanger Institute for the COVID-19 Genomics UK Consortium |
| EPI_ISL_443752 | PHE South West Regional Laboratory, National Infection Service | Wellcome Sanger Institute for the COVID-19 Genomics UK Consortium |
| EPI_ISL_443753 | PHE South West Regional Laboratory, National Infection Service | Wellcome Sanger Institute for the COVID-19 Genomics UK Consortium |
| EPI_ISL_443754 | PHE South West Regional Laboratory, National Infection Service | Wellcome Sanger Institute for the COVID-19 Genomics UK Consortium |
| EPI_ISL_443755 | PHE South West Regional Laboratory, National Infection Service | Wellcome Sanger Institute for the COVID-19 Genomics UK Consortium |
| EPI_ISL_443756 | PHE South West Regional Laboratory, National Infection Service | Wellcome Sanger Institute for the COVID-19 Genomics UK Consortium |
| EPI_ISL_443757 | PHE South West Regional Laboratory, National Infection Service | Wellcome Sanger Institute for the COVID-19 Genomics UK Consortium |

| EPI_Accession_ID | Originating Laboratory | Submitting Laboratory |
|---|---|---|
| EPI_ISL_443758 | PHE South West Regional Laboratory, National Infection Service | Wellcome Sanger Institute for the COVID-19 Genomics UK Consortium |
| EPI_ISL_443759 | PHE South West Regional Laboratory, National Infection Service | Wellcome Sanger Institute for the COVID-19 Genomics UK Consortium |
| EPI_ISL_443760 | PHE South West Regional Laboratory, National Infection Service | Wellcome Sanger Institute for the COVID-19 Genomics UK Consortium |
| EPI_ISL_443761 | PHE South West Regional Laboratory, National Infection Service | Wellcome Sanger Institute for the COVID-19 Genomics UK Consortium |
| EPI_ISL_443762 | PHE South West Regional Laboratory, National Infection Service | Wellcome Sanger Institute for the COVID-19 Genomics UK Consortium |
| EPI_ISL_443763 | PHE South West Regional Laboratory, National Infection Service | Wellcome Sanger Institute for the COVID-19 Genomics UK Consortium |
| EPI_ISL_443764 | PHE South West Regional Laboratory, National Infection Service | Wellcome Sanger Institute for the COVID-19 Genomics UK Consortium |
| EPI_ISL_443765 | PHE South West Regional Laboratory, National Infection Service | Wellcome Sanger Institute for the COVID-19 Genomics UK Consortium |
| EPI_ISL_443766 | PHE South West Regional Laboratory, National Infection Service | Wellcome Sanger Institute for the COVID-19 Genomics UK Consortium |
| EPI_ISL_443767 | PHE South West Regional Laboratory, National Infection Service | Wellcome Sanger Institute for the COVID-19 Genomics UK Consortium |
| EPI_ISL_443768 | PHE South West Regional Laboratory, National Infection Service | Wellcome Sanger Institute for the COVID-19 Genomics UK Consortium |
| EPI_ISL_443769 | PHE South West Regional Laboratory, National Infection Service | Wellcome Sanger Institute for the COVID-19 Genomics UK Consortium |
| EPI_ISL_443770 | PHE South West Regional Laboratory, National Infection Service | Wellcome Sanger Institute for the COVID-19 Genomics UK Consortium |
| EPI_ISL_443771 | PHE South West Regional Laboratory, National Infection Service | Wellcome Sanger Institute for the COVID-19 Genomics UK Consortium |
| EPI_ISL_443772 | PHE South West Regional Laboratory, National Infection Service | Wellcome Sanger Institute for the COVID-19 Genomics UK Consortium |
| EPI_ISL_443773 | PHE South West Regional Laboratory, National Infection Service | Wellcome Sanger Institute for the COVID-19 Genomics UK Consortium |

| EPI_Accession_ID | Originating Laboratory | Submitting Laboratory |
|---|---|---|
| EPI_ISL_443774 | PHE South West Regional Laboratory, National Infection Service | Wellcome Sanger Institute for the COVID-19 Genomics UK Consortium |
| EPI_ISL_443775 | PHE South West Regional Laboratory, National Infection Service | Wellcome Sanger Institute for the COVID-19 Genomics UK Consortium |
| EPI_ISL_443776 | PHE South West Regional Laboratory, National Infection Service | Wellcome Sanger Institute for the COVID-19 Genomics UK Consortium |
| EPI_ISL_443777 | PHE South West Regional Laboratory, National Infection Service | Wellcome Sanger Institute for the COVID-19 Genomics UK Consortium |
| EPI_ISL_443778 | PHE South West Regional Laboratory, National Infection Service | Wellcome Sanger Institute for the COVID-19 Genomics UK Consortium |
| EPI_ISL_443779 | PHE South West Regional Laboratory, National Infection Service | Wellcome Sanger Institute for the COVID-19 Genomics UK Consortium |
| EPI_ISL_443780 | PHE South West Regional Laboratory, National Infection Service | Wellcome Sanger Institute for the COVID-19 Genomics UK Consortium |
| EPI_ISL_443781 | PHE South West Regional Laboratory, National Infection Service | Wellcome Sanger Institute for the COVID-19 Genomics UK Consortium |
| EPI_ISL_443782 | PHE South West Regional Laboratory, National Infection Service | Wellcome Sanger Institute for the COVID-19 Genomics UK Consortium |
| EPI_ISL_443783 | PHE South West Regional Laboratory, National Infection Service | Wellcome Sanger Institute for the COVID-19 Genomics UK Consortium |
| EPI_ISL_443784 | PHE South West Regional Laboratory, National Infection Service | Wellcome Sanger Institute for the COVID-19 Genomics UK Consortium |
| EPI_ISL_443785 | PHE South West Regional Laboratory, National Infection Service | Wellcome Sanger Institute for the COVID-19 Genomics UK Consortium |
| EPI_ISL_443786 | PHE South West Regional Laboratory, National Infection Service | Wellcome Sanger Institute for the COVID-19 Genomics UK Consortium |
| EPI_ISL_443787 | PHE South West Regional Laboratory, National Infection Service | Wellcome Sanger Institute for the COVID-19 Genomics UK Consortium |
| EPI_ISL_443788 | PHE South West Regional Laboratory, National Infection Service | Wellcome Sanger Institute for the COVID-19 Genomics UK Consortium |
| EPI_ISL_443789 | PHE South West Regional Laboratory, National Infection Service | Wellcome Sanger Institute for the COVID-19 Genomics UK Consortium |

| EPI_Accession_ID | Originating Laboratory | Submitting Laboratory |
|---|---|---|
| EPI_ISL_443790 | PHE South West Regional Laboratory, National Infection Service | Wellcome Sanger Institute for the COVID-19 Genomics UK Consortium |
| EPI_ISL_443791 | PHE South West Regional Laboratory, National Infection Service | Wellcome Sanger Institute for the COVID-19 Genomics UK Consortium |
| EPI_ISL_443792 | PHE South West Regional Laboratory, National Infection Service | Wellcome Sanger Institute for the COVID-19 Genomics UK Consortium |
| EPI_ISL_443793 | PHE South West Regional Laboratory, National Infection Service | Wellcome Sanger Institute for the COVID-19 Genomics UK Consortium |
| EPI_ISL_443794 | PHE South West Regional Laboratory, National Infection Service | Wellcome Sanger Institute for the COVID-19 Genomics UK Consortium |
| EPI_ISL_443795 | PHE South West Regional Laboratory, National Infection Service | Wellcome Sanger Institute for the COVID-19 Genomics UK Consortium |
| EPI_ISL_443796 | PHE South West Regional Laboratory, National Infection Service | Wellcome Sanger Institute for the COVID-19 Genomics UK Consortium |
| EPI_ISL_443797 | PHE South West Regional Laboratory, National Infection Service | Wellcome Sanger Institute for the COVID-19 Genomics UK Consortium |
| EPI_ISL_443798 | PHE South West Regional Laboratory, National Infection Service | Wellcome Sanger Institute for the COVID-19 Genomics UK Consortium |
| EPI_ISL_443799 | PHE South West Regional Laboratory, National Infection Service | Wellcome Sanger Institute for the COVID-19 Genomics UK Consortium |
| EPI_ISL_443800 | PHE South West Regional Laboratory, National Infection Service | Wellcome Sanger Institute for the COVID-19 Genomics UK Consortium |
| EPI_ISL_443801 | PHE South West Regional Laboratory, National Infection Service | Wellcome Sanger Institute for the COVID-19 Genomics UK Consortium |
| EPI_ISL_443802 | PHE South West Regional Laboratory, National Infection Service | Wellcome Sanger Institute for the COVID-19 Genomics UK Consortium |
| EPI_ISL_443803 | PHE South West Regional Laboratory, National Infection Service | Wellcome Sanger Institute for the COVID-19 Genomics UK Consortium |
| EPI_ISL_443804 | PHE South West Regional Laboratory, National Infection Service | Wellcome Sanger Institute for the COVID-19 Genomics UK Consortium |
| EPI_ISL_443805 | PHE South West Regional Laboratory, National Infection Service | Wellcome Sanger Institute for the COVID-19 Genomics UK Consortium |

| EPI_Accession_ID | Originating Laboratory | Submitting Laboratory |
| --- | --- | --- |
| EPI_ISL_443806 | PHE South West Regional Laboratory, National Infection Service | Wellcome Sanger Institute for the COVID-19 Genomics UK Consortium |
| EPI_ISL_443807 | PHE South West Regional Laboratory, National Infection Service | Wellcome Sanger Institute for the COVID-19 Genomics UK Consortium |
| EPI_ISL_443808 | PHE South West Regional Laboratory, National Infection Service | Wellcome Sanger Institute for the COVID-19 Genomics UK Consortium |
| EPI_ISL_443809 | PHE South West Regional Laboratory, National Infection Service | Wellcome Sanger Institute for the COVID-19 Genomics UK Consortium |
| EPI_ISL_443810 | PHE South West Regional Laboratory, National Infection Service | Wellcome Sanger Institute for the COVID-19 Genomics UK Consortium |
| EPI_ISL_443811 | PHE South West Regional Laboratory, National Infection Service | Wellcome Sanger Institute for the COVID-19 Genomics UK Consortium |
| EPI_ISL_443812 | PHE South West Regional Laboratory, National Infection Service | Wellcome Sanger Institute for the COVID-19 Genomics UK Consortium |
| EPI_ISL_443813 | PHE South West Regional Laboratory, National Infection Service | Wellcome Sanger Institute for the COVID-19 Genomics UK Consortium |
| EPI_ISL_443814 | PHE South West Regional Laboratory, National Infection Service | Wellcome Sanger Institute for the COVID-19 Genomics UK Consortium |
| EPI_ISL_443815 | PHE South West Regional Laboratory, National Infection Service | Wellcome Sanger Institute for the COVID-19 Genomics UK Consortium |
| EPI_ISL_443816 | PHE South West Regional Laboratory, National Infection Service | Wellcome Sanger Institute for the COVID-19 Genomics UK Consortium |
| EPI_ISL_443817 | PHE South West Regional Laboratory, National Infection Service | Wellcome Sanger Institute for the COVID-19 Genomics UK Consortium |
| EPI_ISL_443818 | PHE South West Regional Laboratory, National Infection Service | Wellcome Sanger Institute for the COVID-19 Genomics UK Consortium |
| EPI_ISL_443819 | PHE South West Regional Laboratory, National Infection Service | Wellcome Sanger Institute for the COVID-19 Genomics UK Consortium |
| EPI_ISL_443820 | PHE South West Regional Laboratory, National Infection Service | Wellcome Sanger Institute for the COVID-19 Genomics UK Consortium |
| EPI_ISL_443821 | PHE South West Regional Laboratory, National Infection Service | Wellcome Sanger Institute for the COVID-19 Genomics UK Consortium |

| EPI_Accession_ID | Originating Laboratory | Submitting Laboratory |
|---|---|---|
| EPI_ISL_443822 | PHE South West Regional Laboratory, National Infection Service | Wellcome Sanger Institute for the COVID-19 Genomics UK Consortium |
| EPI_ISL_443823 | PHE South West Regional Laboratory, National Infection Service | Wellcome Sanger Institute for the COVID-19 Genomics UK Consortium |
| EPI_ISL_443824 | PHE South West Regional Laboratory, National Infection Service | Wellcome Sanger Institute for the COVID-19 Genomics UK Consortium |
| EPI_ISL_443825 | PHE South West Regional Laboratory, National Infection Service | Wellcome Sanger Institute for the COVID-19 Genomics UK Consortium |
| EPI_ISL_443826 | PHE South West Regional Laboratory, National Infection Service | Wellcome Sanger Institute for the COVID-19 Genomics UK Consortium |
| EPI_ISL_443827 | PHE South West Regional Laboratory, National Infection Service | Wellcome Sanger Institute for the COVID-19 Genomics UK Consortium |
| EPI_ISL_443828 | PHE South West Regional Laboratory, National Infection Service | Wellcome Sanger Institute for the COVID-19 Genomics UK Consortium |
| EPI_ISL_443829 | PHE South West Regional Laboratory, National Infection Service | Wellcome Sanger Institute for the COVID-19 Genomics UK Consortium |
| EPI_ISL_443830 | PHE South West Regional Laboratory, National Infection Service | Wellcome Sanger Institute for the COVID-19 Genomics UK Consortium |
| EPI_ISL_443831 | PHE South West Regional Laboratory, National Infection Service | Wellcome Sanger Institute for the COVID-19 Genomics UK Consortium |
| EPI_ISL_443832 | PHE South West Regional Laboratory, National Infection Service | Wellcome Sanger Institute for the COVID-19 Genomics UK Consortium |
| EPI_ISL_443833 | PHE South West Regional Laboratory, National Infection Service | Wellcome Sanger Institute for the COVID-19 Genomics UK Consortium |
| EPI_ISL_443834 | PHE South West Regional Laboratory, National Infection Service | Wellcome Sanger Institute for the COVID-19 Genomics UK Consortium |
| EPI_ISL_443835 | PHE South West Regional Laboratory, National Infection Service | Wellcome Sanger Institute for the COVID-19 Genomics UK Consortium |
| EPI_ISL_443836 | PHE South West Regional Laboratory, National Infection Service | Wellcome Sanger Institute for the COVID-19 Genomics UK Consortium |
| EPI_ISL_443837 | PHE South West Regional Laboratory, National Infection Service | Wellcome Sanger Institute for the COVID-19 Genomics UK Consortium |

| EPI_Accession_ID | Originating Laboratory | Submitting Laboratory |
|---|---|---|
| EPI_ISL_443838 | PHE South West Regional Laboratory, National Infection Service | Wellcome Sanger Institute for the COVID-19 Genomics UK Consortium |
| EPI_ISL_443839 | PHE South West Regional Laboratory, National Infection Service | Wellcome Sanger Institute for the COVID-19 Genomics UK Consortium |
| EPI_ISL_443840 | PHE South West Regional Laboratory, National Infection Service | Wellcome Sanger Institute for the COVID-19 Genomics UK Consortium |
| EPI_ISL_443841 | PHE South West Regional Laboratory, National Infection Service | Wellcome Sanger Institute for the COVID-19 Genomics UK Consortium |
| EPI_ISL_443842 | PHE South West Regional Laboratory, National Infection Service | Wellcome Sanger Institute for the COVID-19 Genomics UK Consortium |
| EPI_ISL_443843 | PHE South West Regional Laboratory, National Infection Service | Wellcome Sanger Institute for the COVID-19 Genomics UK Consortium |
| EPI_ISL_443844 | PHE South West Regional Laboratory, National Infection Service | Wellcome Sanger Institute for the COVID-19 Genomics UK Consortium |
| EPI_ISL_443845 | PHE South West Regional Laboratory, National Infection Service | Wellcome Sanger Institute for the COVID-19 Genomics UK Consortium |
| EPI_ISL_443846 | PHE South West Regional Laboratory, National Infection Service | Wellcome Sanger Institute for the COVID-19 Genomics UK Consortium |
| EPI_ISL_443847 | PHE South West Regional Laboratory, National Infection Service | Wellcome Sanger Institute for the COVID-19 Genomics UK Consortium |
| EPI_ISL_443848 | PHE South West Regional Laboratory, National Infection Service | Wellcome Sanger Institute for the COVID-19 Genomics UK Consortium |
| EPI_ISL_443849 | PHE South West Regional Laboratory, National Infection Service | Wellcome Sanger Institute for the COVID-19 Genomics UK Consortium |
| EPI_ISL_443850 | PHE South West Regional Laboratory, National Infection Service | Wellcome Sanger Institute for the COVID-19 Genomics UK Consortium |
| EPI_ISL_443851 | PHE South West Regional Laboratory, National Infection Service | Wellcome Sanger Institute for the COVID-19 Genomics UK Consortium |
| EPI_ISL_443852 | PHE South West Regional Laboratory, National Infection Service | Wellcome Sanger Institute for the COVID-19 Genomics UK Consortium |
| EPI_ISL_443853 | PHE South West Regional Laboratory, National Infection Service | Wellcome Sanger Institute for the COVID-19 Genomics UK Consortium |

| EPI_Accession_ID | Originating Laboratory | Submitting Laboratory |
|---|---|---|
| EPI_ISL_443854 | PHE South West Regional Laboratory, National Infection Service | Wellcome Sanger Institute for the COVID-19 Genomics UK Consortium |
| EPI_ISL_443855 | PHE South West Regional Laboratory, National Infection Service | Wellcome Sanger Institute for the COVID-19 Genomics UK Consortium |
| EPI_ISL_443856 | PHE South West Regional Laboratory, National Infection Service | Wellcome Sanger Institute for the COVID-19 Genomics UK Consortium |
| EPI_ISL_443857 | PHE South West Regional Laboratory, National Infection Service | Wellcome Sanger Institute for the COVID-19 Genomics UK Consortium |
| EPI_ISL_443858 | PHE South West Regional Laboratory, National Infection Service | Wellcome Sanger Institute for the COVID-19 Genomics UK Consortium |
| EPI_ISL_443859 | PHE South West Regional Laboratory, National Infection Service | Wellcome Sanger Institute for the COVID-19 Genomics UK Consortium |
| EPI_ISL_443860 | PHE South West Regional Laboratory, National Infection Service | Wellcome Sanger Institute for the COVID-19 Genomics UK Consortium |
| EPI_ISL_443861 | PHE South West Regional Laboratory, National Infection Service | Wellcome Sanger Institute for the COVID-19 Genomics UK Consortium |
| EPI_ISL_443862 | PHE South West Regional Laboratory, National Infection Service | Wellcome Sanger Institute for the COVID-19 Genomics UK Consortium |
| EPI_ISL_443863 | PHE South West Regional Laboratory, National Infection Service | Wellcome Sanger Institute for the COVID-19 Genomics UK Consortium |
| EPI_ISL_443864 | PHE South West Regional Laboratory, National Infection Service | Wellcome Sanger Institute for the COVID-19 Genomics UK Consortium |
| EPI_ISL_443865 | PHE South West Regional Laboratory, National Infection Service | Wellcome Sanger Institute for the COVID-19 Genomics UK Consortium |
| EPI_ISL_443866 | PHE South West Regional Laboratory, National Infection Service | Wellcome Sanger Institute for the COVID-19 Genomics UK Consortium |
| EPI_ISL_443867 | PHE South West Regional Laboratory, National Infection Service | Wellcome Sanger Institute for the COVID-19 Genomics UK Consortium |
| EPI_ISL_443868 | PHE South West Regional Laboratory, National Infection Service | Wellcome Sanger Institute for the COVID-19 Genomics UK Consortium |
| EPI_ISL_443869 | PHE South West Regional Laboratory, National Infection Service | Wellcome Sanger Institute for the COVID-19 Genomics UK Consortium |

| EPI_Accession_ID | Originating Laboratory | Submitting Laboratory |
| --- | --- | --- |
| EPI_ISL_443870 | PHE South West Regional Laboratory, National Infection Service | Wellcome Sanger Institute for the COVID-19 Genomics UK Consortium |
| EPI_ISL_443871 | PHE South West Regional Laboratory, National Infection Service | Wellcome Sanger Institute for the COVID-19 Genomics UK Consortium |
| EPI_ISL_443872 | PHE South West Regional Laboratory, National Infection Service | Wellcome Sanger Institute for the COVID-19 Genomics UK Consortium |
| EPI_ISL_443873 | PHE South West Regional Laboratory, National Infection Service | Wellcome Sanger Institute for the COVID-19 Genomics UK Consortium |
| EPI_ISL_443874 | PHE South West Regional Laboratory, National Infection Service | Wellcome Sanger Institute for the COVID-19 Genomics UK Consortium |
| EPI_ISL_443875 | PHE South West Regional Laboratory, National Infection Service | Wellcome Sanger Institute for the COVID-19 Genomics UK Consortium |
| EPI_ISL_443876 | PHE South West Regional Laboratory, National Infection Service | Wellcome Sanger Institute for the COVID-19 Genomics UK Consortium |
| EPI_ISL_443877 | PHE South West Regional Laboratory, National Infection Service | Wellcome Sanger Institute for the COVID-19 Genomics UK Consortium |
| EPI_ISL_443878 | PHE South West Regional Laboratory, National Infection Service | Wellcome Sanger Institute for the COVID-19 Genomics UK Consortium |
| EPI_ISL_443879 | PHE South West Regional Laboratory, National Infection Service | Wellcome Sanger Institute for the COVID-19 Genomics UK Consortium |
| EPI_ISL_443880 | PHE South West Regional Laboratory, National Infection Service | Wellcome Sanger Institute for the COVID-19 Genomics UK Consortium |
| EPI_ISL_443881 | PHE South West Regional Laboratory, National Infection Service | Wellcome Sanger Institute for the COVID-19 Genomics UK Consortium |
| EPI_ISL_443882 | PHE South West Regional Laboratory, National Infection Service | Wellcome Sanger Institute for the COVID-19 Genomics UK Consortium |
| EPI_ISL_443883 | PHE South West Regional Laboratory, National Infection Service | Wellcome Sanger Institute for the COVID-19 Genomics UK Consortium |
| EPI_ISL_443884 | PHE South West Regional Laboratory, National Infection Service | Wellcome Sanger Institute for the COVID-19 Genomics UK Consortium |
| EPI_ISL_443885 | PHE South West Regional Laboratory, National Infection Service | Wellcome Sanger Institute for the COVID-19 Genomics UK Consortium |

| EPI_Accession_ID | Originating Laboratory | Submitting Laboratory |
|---|---|---|
| EPI_ISL_443886 | PHE South West Regional Laboratory, National Infection Service | Wellcome Sanger Institute for the COVID-19 Genomics UK Consortium |
| EPI_ISL_443887 | PHE South West Regional Laboratory, National Infection Service | Wellcome Sanger Institute for the COVID-19 Genomics UK Consortium |
| EPI_ISL_443888 | PHE South West Regional Laboratory, National Infection Service | Wellcome Sanger Institute for the COVID-19 Genomics UK Consortium |
| EPI_ISL_443889 | PHE South West Regional Laboratory, National Infection Service | Wellcome Sanger Institute for the COVID-19 Genomics UK Consortium |
| EPI_ISL_443890 | PHE South West Regional Laboratory, National Infection Service | Wellcome Sanger Institute for the COVID-19 Genomics UK Consortium |
| EPI_ISL_443891 | PHE South West Regional Laboratory, National Infection Service | Wellcome Sanger Institute for the COVID-19 Genomics UK Consortium |
| EPI_ISL_443892 | PHE South West Regional Laboratory, National Infection Service | Wellcome Sanger Institute for the COVID-19 Genomics UK Consortium |
| EPI_ISL_443893 | PHE South West Regional Laboratory, National Infection Service | Wellcome Sanger Institute for the COVID-19 Genomics UK Consortium |
| EPI_ISL_443894 | PHE South West Regional Laboratory, National Infection Service | Wellcome Sanger Institute for the COVID-19 Genomics UK Consortium |
| EPI_ISL_443895 | PHE South West Regional Laboratory, National Infection Service | Wellcome Sanger Institute for the COVID-19 Genomics UK Consortium |
| EPI_ISL_443896 | PHE South West Regional Laboratory, National Infection Service | Wellcome Sanger Institute for the COVID-19 Genomics UK Consortium |
| EPI_ISL_443897 | PHE South West Regional Laboratory, National Infection Service | Wellcome Sanger Institute for the COVID-19 Genomics UK Consortium |
| EPI_ISL_443898 | PHE South West Regional Laboratory, National Infection Service | Wellcome Sanger Institute for the COVID-19 Genomics UK Consortium |
| EPI_ISL_443899 | PHE South West Regional Laboratory, National Infection Service | Wellcome Sanger Institute for the COVID-19 Genomics UK Consortium |
| EPI_ISL_443900 | PHE South West Regional Laboratory, National Infection Service | Wellcome Sanger Institute for the COVID-19 Genomics UK Consortium |
| EPI_ISL_443901 | PHE South West Regional Laboratory, National Infection Service | Wellcome Sanger Institute for the COVID-19 Genomics UK Consortium |

| EPI_Accession_ID | Originating Laboratory | Submitting Laboratory |
|---|---|---|
| EPI_ISL_443902 | PHE South West Regional Laboratory, National Infection Service | Wellcome Sanger Institute for the COVID-19 Genomics UK Consortium |
| EPI_ISL_443903 | PHE South West Regional Laboratory, National Infection Service | Wellcome Sanger Institute for the COVID-19 Genomics UK Consortium |
| EPI_ISL_443904 | PHE South West Regional Laboratory, National Infection Service | Wellcome Sanger Institute for the COVID-19 Genomics UK Consortium |
| EPI_ISL_443905 | PHE South West Regional Laboratory, National Infection Service | Wellcome Sanger Institute for the COVID-19 Genomics UK Consortium |
| EPI_ISL_443906 | PHE South West Regional Laboratory, National Infection Service | Wellcome Sanger Institute for the COVID-19 Genomics UK Consortium |
| EPI_ISL_443907 | PHE South West Regional Laboratory, National Infection Service | Wellcome Sanger Institute for the COVID-19 Genomics UK Consortium |
| EPI_ISL_443908 | PHE South West Regional Laboratory, National Infection Service | Wellcome Sanger Institute for the COVID-19 Genomics UK Consortium |
| EPI_ISL_443909 | PHE South West Regional Laboratory, National Infection Service | Wellcome Sanger Institute for the COVID-19 Genomics UK Consortium |
| EPI_ISL_443910 | PHE South West Regional Laboratory, National Infection Service | Wellcome Sanger Institute for the COVID-19 Genomics UK Consortium |
| EPI_ISL_443911 | PHE South West Regional Laboratory, National Infection Service | Wellcome Sanger Institute for the COVID-19 Genomics UK Consortium |
| EPI_ISL_443912 | PHE South West Regional Laboratory, National Infection Service | Wellcome Sanger Institute for the COVID-19 Genomics UK Consortium |
| EPI_ISL_443913 | PHE South West Regional Laboratory, National Infection Service | Wellcome Sanger Institute for the COVID-19 Genomics UK Consortium |
| EPI_ISL_443914 | PHE South West Regional Laboratory, National Infection Service | Wellcome Sanger Institute for the COVID-19 Genomics UK Consortium |
| EPI_ISL_443915 | PHE South West Regional Laboratory, National Infection Service | Wellcome Sanger Institute for the COVID-19 Genomics UK Consortium |
| EPI_ISL_443916 | PHE South West Regional Laboratory, National Infection Service | Wellcome Sanger Institute for the COVID-19 Genomics UK Consortium |
| EPI_ISL_443917 | PHE South West Regional Laboratory, National Infection Service | Wellcome Sanger Institute for the COVID-19 Genomics UK Consortium |

| EPI_Accession_ID | Originating Laboratory | Submitting Laboratory |
|---|---|---|
| EPI_ISL_443918 | PHE South West Regional Laboratory, National Infection Service | Wellcome Sanger Institute for the COVID-19 Genomics UK Consortium |
| EPI_ISL_443919 | PHE South West Regional Laboratory, National Infection Service | Wellcome Sanger Institute for the COVID-19 Genomics UK Consortium |
| EPI_ISL_443920 | PHE South West Regional Laboratory, National Infection Service | Wellcome Sanger Institute for the COVID-19 Genomics UK Consortium |
| EPI_ISL_443921 | PHE South West Regional Laboratory, National Infection Service | Wellcome Sanger Institute for the COVID-19 Genomics UK Consortium |
| EPI_ISL_443922 | PHE South West Regional Laboratory, National Infection Service | Wellcome Sanger Institute for the COVID-19 Genomics UK Consortium |
| EPI_ISL_443923 | PHE South West Regional Laboratory, National Infection Service | Wellcome Sanger Institute for the COVID-19 Genomics UK Consortium |
| EPI_ISL_443924 | PHE South West Regional Laboratory, National Infection Service | Wellcome Sanger Institute for the COVID-19 Genomics UK Consortium |
| EPI_ISL_443925 | PHE South West Regional Laboratory, National Infection Service | Wellcome Sanger Institute for the COVID-19 Genomics UK Consortium |
| EPI_ISL_443926 | PHE South West Regional Laboratory, National Infection Service | Wellcome Sanger Institute for the COVID-19 Genomics UK Consortium |
| EPI_ISL_443927 | PHE South West Regional Laboratory, National Infection Service | Wellcome Sanger Institute for the COVID-19 Genomics UK Consortium |
| EPI_ISL_443928 | PHE South West Regional Laboratory, National Infection Service | Wellcome Sanger Institute for the COVID-19 Genomics UK Consortium |
| EPI_ISL_443929 | PHE South West Regional Laboratory, National Infection Service | Wellcome Sanger Institute for the COVID-19 Genomics UK Consortium |
| EPI_ISL_443930 | PHE South West Regional Laboratory, National Infection Service | Wellcome Sanger Institute for the COVID-19 Genomics UK Consortium |
| EPI_ISL_443931 | PHE South West Regional Laboratory, National Infection Service | Wellcome Sanger Institute for the COVID-19 Genomics UK Consortium |
| EPI_ISL_443932 | PHE South West Regional Laboratory, National Infection Service | Wellcome Sanger Institute for the COVID-19 Genomics UK Consortium |
| EPI_ISL_443933 | PHE South West Regional Laboratory, National Infection Service | Wellcome Sanger Institute for the COVID-19 Genomics UK Consortium |

| EPI_Accession_ID | Originating Laboratory | Submitting Laboratory |
|---|---|---|
| EPI_ISL_443934 | PHE South West Regional Laboratory, National Infection Service | Wellcome Sanger Institute for the COVID-19 Genomics UK Consortium |
| EPI_ISL_443935 | PHE South West Regional Laboratory, National Infection Service | Wellcome Sanger Institute for the COVID-19 Genomics UK Consortium |
| EPI_ISL_443936 | PHE South West Regional Laboratory, National Infection Service | Wellcome Sanger Institute for the COVID-19 Genomics UK Consortium |
| EPI_ISL_443937 | PHE South West Regional Laboratory, National Infection Service | Wellcome Sanger Institute for the COVID-19 Genomics UK Consortium |
| EPI_ISL_443938 | PHE South West Regional Laboratory, National Infection Service | Wellcome Sanger Institute for the COVID-19 Genomics UK Consortium |
| EPI_ISL_443939 | PHE South West Regional Laboratory, National Infection Service | Wellcome Sanger Institute for the COVID-19 Genomics UK Consortium |
| EPI_ISL_443940 | PHE South West Regional Laboratory, National Infection Service | Wellcome Sanger Institute for the COVID-19 Genomics UK Consortium |
| EPI_ISL_443941 | PHE South West Regional Laboratory, National Infection Service | Wellcome Sanger Institute for the COVID-19 Genomics UK Consortium |
| EPI_ISL_443942 | PHE South West Regional Laboratory, National Infection Service | Wellcome Sanger Institute for the COVID-19 Genomics UK Consortium |
| EPI_ISL_443943 | PHE South West Regional Laboratory, National Infection Service | Wellcome Sanger Institute for the COVID-19 Genomics UK Consortium |
| EPI_ISL_443944 | PHE South West Regional Laboratory, National Infection Service | Wellcome Sanger Institute for the COVID-19 Genomics UK Consortium |
| EPI_ISL_443945 | PHE South West Regional Laboratory, National Infection Service | Wellcome Sanger Institute for the COVID-19 Genomics UK Consortium |
| EPI_ISL_443946 | PHE South West Regional Laboratory, National Infection Service | Wellcome Sanger Institute for the COVID-19 Genomics UK Consortium |
| EPI_ISL_443947 | PHE South West Regional Laboratory, National Infection Service | Wellcome Sanger Institute for the COVID-19 Genomics UK Consortium |
| EPI_ISL_443948 | PHE South West Regional Laboratory, National Infection Service | Wellcome Sanger Institute for the COVID-19 Genomics UK Consortium |
| EPI_ISL_443949 | PHE South West Regional Laboratory, National Infection Service | Wellcome Sanger Institute for the COVID-19 Genomics UK Consortium |

| EPI_Accession_ID | Originating Laboratory | Submitting Laboratory |
|---|---|---|
| EPI_ISL_443950 | PHE South West Regional Laboratory, National Infection Service | Wellcome Sanger Institute for the COVID-19 Genomics UK Consortium |
| EPI_ISL_443951 | PHE South West Regional Laboratory, National Infection Service | Wellcome Sanger Institute for the COVID-19 Genomics UK Consortium |
| EPI_ISL_443952 | PHE South West Regional Laboratory, National Infection Service | Wellcome Sanger Institute for the COVID-19 Genomics UK Consortium |
| EPI_ISL_443953 | PHE South West Regional Laboratory, National Infection Service | Wellcome Sanger Institute for the COVID-19 Genomics UK Consortium |
| EPI_ISL_443954 | PHE South West Regional Laboratory, National Infection Service | Wellcome Sanger Institute for the COVID-19 Genomics UK Consortium |
| EPI_ISL_443955 | PHE South West Regional Laboratory, National Infection Service | Wellcome Sanger Institute for the COVID-19 Genomics UK Consortium |
| EPI_ISL_443956 | PHE South West Regional Laboratory, National Infection Service | Wellcome Sanger Institute for the COVID-19 Genomics UK Consortium |
| EPI_ISL_443957 | PHE South West Regional Laboratory, National Infection Service | Wellcome Sanger Institute for the COVID-19 Genomics UK Consortium |
| EPI_ISL_443958 | PHE South West Regional Laboratory, National Infection Service | Wellcome Sanger Institute for the COVID-19 Genomics UK Consortium |
| EPI_ISL_443959 | PHE South West Regional Laboratory, National Infection Service | Wellcome Sanger Institute for the COVID-19 Genomics UK Consortium |
| EPI_ISL_443960 | PHE South West Regional Laboratory, National Infection Service | Wellcome Sanger Institute for the COVID-19 Genomics UK Consortium |
| EPI_ISL_443961 | PHE South West Regional Laboratory, National Infection Service | Wellcome Sanger Institute for the COVID-19 Genomics UK Consortium |
| EPI_ISL_443962 | PHE South West Regional Laboratory, National Infection Service | Wellcome Sanger Institute for the COVID-19 Genomics UK Consortium |
| EPI_ISL_443963 | PHE South West Regional Laboratory, National Infection Service | Wellcome Sanger Institute for the COVID-19 Genomics UK Consortium |
| EPI_ISL_443964 | PHE South West Regional Laboratory, National Infection Service | Wellcome Sanger Institute for the COVID-19 Genomics UK Consortium |
| EPI_ISL_443965 | PHE South West Regional Laboratory, National Infection Service | Wellcome Sanger Institute for the COVID-19 Genomics UK Consortium |

| EPI_Accession_ID | Originating Laboratory | Submitting Laboratory |
|---|---|---|
| EPI_ISL_443966 | PHE South West Regional Laboratory, National Infection Service | Wellcome Sanger Institute for the COVID-19 Genomics UK Consortium |
| EPI_ISL_443967 | PHE South West Regional Laboratory, National Infection Service | Wellcome Sanger Institute for the COVID-19 Genomics UK Consortium |
| EPI_ISL_443968 | PHE South West Regional Laboratory, National Infection Service | Wellcome Sanger Institute for the COVID-19 Genomics UK Consortium |
| EPI_ISL_443969 | PHE South West Regional Laboratory, National Infection Service | Wellcome Sanger Institute for the COVID-19 Genomics UK Consortium |
| EPI_ISL_443970 | PHE South West Regional Laboratory, National Infection Service | Wellcome Sanger Institute for the COVID-19 Genomics UK Consortium |
| EPI_ISL_443971 | PHE South West Regional Laboratory, National Infection Service | Wellcome Sanger Institute for the COVID-19 Genomics UK Consortium |
| EPI_ISL_443972 | PHE South West Regional Laboratory, National Infection Service | Wellcome Sanger Institute for the COVID-19 Genomics UK Consortium |
| EPI_ISL_443973 | PHE South West Regional Laboratory, National Infection Service | Wellcome Sanger Institute for the COVID-19 Genomics UK Consortium |
| EPI_ISL_443974 | PHE South West Regional Laboratory, National Infection Service | Wellcome Sanger Institute for the COVID-19 Genomics UK Consortium |
| EPI_ISL_443975 | PHE South West Regional Laboratory, National Infection Service | Wellcome Sanger Institute for the COVID-19 Genomics UK Consortium |
| EPI_ISL_443976 | PHE South West Regional Laboratory, National Infection Service | Wellcome Sanger Institute for the COVID-19 Genomics UK Consortium |
| EPI_ISL_443977 | PHE South West Regional Laboratory, National Infection Service | Wellcome Sanger Institute for the COVID-19 Genomics UK Consortium |
| EPI_ISL_443978 | PHE South West Regional Laboratory, National Infection Service | Wellcome Sanger Institute for the COVID-19 Genomics UK Consortium |
| EPI_ISL_443979 | PHE South West Regional Laboratory, National Infection Service | Wellcome Sanger Institute for the COVID-19 Genomics UK Consortium |
| EPI_ISL_443980 | PHE South West Regional Laboratory, National Infection Service | Wellcome Sanger Institute for the COVID-19 Genomics UK Consortium |
| EPI_ISL_443981 | PHE South West Regional Laboratory, National Infection Service | Wellcome Sanger Institute for the COVID-19 Genomics UK Consortium |

| EPI_Accession_ID | Originating Laboratory | Submitting Laboratory |
|---|---|---|
| EPI_ISL_443982 | PHE South West Regional Laboratory, National Infection Service | Wellcome Sanger Institute for the COVID-19 Genomics UK Consortium |
| EPI_ISL_443983 | PHE South West Regional Laboratory, National Infection Service | Wellcome Sanger Institute for the COVID-19 Genomics UK Consortium |
| EPI_ISL_443984 | PHE South West Regional Laboratory, National Infection Service | Wellcome Sanger Institute for the COVID-19 Genomics UK Consortium |
| EPI_ISL_443985 | PHE South West Regional Laboratory, National Infection Service | Wellcome Sanger Institute for the COVID-19 Genomics UK Consortium |
| EPI_ISL_443986 | PHE South West Regional Laboratory, National Infection Service | Wellcome Sanger Institute for the COVID-19 Genomics UK Consortium |
| EPI_ISL_443987 | PHE South West Regional Laboratory, National Infection Service | Wellcome Sanger Institute for the COVID-19 Genomics UK Consortium |
| EPI_ISL_443988 | PHE South West Regional Laboratory, National Infection Service | Wellcome Sanger Institute for the COVID-19 Genomics UK Consortium |
| EPI_ISL_443989 | PHE South West Regional Laboratory, National Infection Service | Wellcome Sanger Institute for the COVID-19 Genomics UK Consortium |
| EPI_ISL_443990 | PHE South West Regional Laboratory, National Infection Service | Wellcome Sanger Institute for the COVID-19 Genomics UK Consortium |
| EPI_ISL_443991 | PHE South West Regional Laboratory, National Infection Service | Wellcome Sanger Institute for the COVID-19 Genomics UK Consortium |
| EPI_ISL_443992 | PHE South West Regional Laboratory, National Infection Service | Wellcome Sanger Institute for the COVID-19 Genomics UK Consortium |
| EPI_ISL_443993 | PHE South West Regional Laboratory, National Infection Service | Wellcome Sanger Institute for the COVID-19 Genomics UK Consortium |
| EPI_ISL_443994 | PHE South West Regional Laboratory, National Infection Service | Wellcome Sanger Institute for the COVID-19 Genomics UK Consortium |
| EPI_ISL_443995 | PHE South West Regional Laboratory, National Infection Service | Wellcome Sanger Institute for the COVID-19 Genomics UK Consortium |
| EPI_ISL_443996 | PHE South West Regional Laboratory, National Infection Service | Wellcome Sanger Institute for the COVID-19 Genomics UK Consortium |
| EPI_ISL_443997 | PHE South West Regional Laboratory, National Infection Service | Wellcome Sanger Institute for the COVID-19 Genomics UK Consortium |

| EPI_Accession_ID | Originating Laboratory | Submitting Laboratory |
| --- | --- | --- |
| EPI_ISL_443998 | PHE South West Regional Laboratory, National Infection Service | Wellcome Sanger Institute for the COVID-19 Genomics UK Consortium |
| EPI_ISL_443999 | PHE South West Regional Laboratory, National Infection Service | Wellcome Sanger Institute for the COVID-19 Genomics UK Consortium |
| EPI_ISL_444000 | PHE South West Regional Laboratory, National Infection Service | Wellcome Sanger Institute for the COVID-19 Genomics UK Consortium |
| EPI_ISL_444001 | PHE South West Regional Laboratory, National Infection Service | Wellcome Sanger Institute for the COVID-19 Genomics UK Consortium |
| EPI_ISL_444002 | PHE South West Regional Laboratory, National Infection Service | Wellcome Sanger Institute for the COVID-19 Genomics UK Consortium |
| EPI_ISL_444003 | PHE South West Regional Laboratory, National Infection Service | Wellcome Sanger Institute for the COVID-19 Genomics UK Consortium |
| EPI_ISL_444004 | PHE South West Regional Laboratory, National Infection Service | Wellcome Sanger Institute for the COVID-19 Genomics UK Consortium |
| EPI_ISL_444005 | PHE South West Regional Laboratory, National Infection Service | Wellcome Sanger Institute for the COVID-19 Genomics UK Consortium |
| EPI_ISL_444006 | PHE South West Regional Laboratory, National Infection Service | Wellcome Sanger Institute for the COVID-19 Genomics UK Consortium |
| EPI_ISL_444007 | PHE South West Regional Laboratory, National Infection Service | Wellcome Sanger Institute for the COVID-19 Genomics UK Consortium |
| EPI_ISL_444008 | PHE South West Regional Laboratory, National Infection Service | Wellcome Sanger Institute for the COVID-19 Genomics UK Consortium |
| EPI_ISL_444009 | PHE South West Regional Laboratory, National Infection Service | Wellcome Sanger Institute for the COVID-19 Genomics UK Consortium |
| EPI_ISL_444010 | PHE South West Regional Laboratory, National Infection Service | Wellcome Sanger Institute for the COVID-19 Genomics UK Consortium |
| EPI_ISL_444011 | PHE South West Regional Laboratory, National Infection Service | Wellcome Sanger Institute for the COVID-19 Genomics UK Consortium |
| EPI_ISL_444012 | PHE South West Regional Laboratory, National Infection Service | Wellcome Sanger Institute for the COVID-19 Genomics UK Consortium |
| EPI_ISL_444013 | PHE South West Regional Laboratory, National Infection Service | Wellcome Sanger Institute for the COVID-19 Genomics UK Consortium |

| EPI_Accession_ID | Originating Laboratory | Submitting Laboratory |
|---|---|---|
| EPI_ISL_444014 | PHE South West Regional Laboratory, National Infection Service | Wellcome Sanger Institute for the COVID-19 Genomics UK Consortium |
| EPI_ISL_444015 | PHE South West Regional Laboratory, National Infection Service | Wellcome Sanger Institute for the COVID-19 Genomics UK Consortium |
| EPI_ISL_444016 | PHE South West Regional Laboratory, National Infection Service | Wellcome Sanger Institute for the COVID-19 Genomics UK Consortium |
| EPI_ISL_444017 | PHE South West Regional Laboratory, National Infection Service | Wellcome Sanger Institute for the COVID-19 Genomics UK Consortium |
| EPI_ISL_444018 | PHE South West Regional Laboratory, National Infection Service | Wellcome Sanger Institute for the COVID-19 Genomics UK Consortium |
| EPI_ISL_444019 | PHE South West Regional Laboratory, National Infection Service | Wellcome Sanger Institute for the COVID-19 Genomics UK Consortium |
| EPI_ISL_444020 | PHE South West Regional Laboratory, National Infection Service | Wellcome Sanger Institute for the COVID-19 Genomics UK Consortium |
| EPI_ISL_444021 | PHE South West Regional Laboratory, National Infection Service | Wellcome Sanger Institute for the COVID-19 Genomics UK Consortium |
| EPI_ISL_444022 | Baylor College of Medicine | Baylor College of Medicine: HGSC |
| EPI_ISL_444023 | County of Santa Clara Public Health | Chan-Zuckerberg Biohub |
| EPI_ISL_444024 | County of Santa Clara Public Health | Chan-Zuckerberg Biohub |
| EPI_ISL_444025 | County of Santa Clara Public Health | Chan-Zuckerberg Biohub |
| EPI_ISL_444026 | County of Santa Clara Public Health | Chan-Zuckerberg Biohub |
| EPI_ISL_444051 | UCSF Clinical Microbiology Laboratory | Chan-Zuckerberg Biohub |
| EPI_ISL_444052 | UCSF Clinical Microbiology Laboratory | Chan-Zuckerberg Biohub |
| EPI_ISL_444053 | UCSF Clinical Microbiology Laboratory | Chan-Zuckerberg Biohub |
| EPI_ISL_444054 | UCSF Clinical Microbiology Laboratory | Chan-Zuckerberg Biohub |
| EPI_ISL_444055 | UCSF Clinical Microbiology Laboratory | Chan-Zuckerberg Biohub |
| EPI_ISL_444056 | UCSF Clinical Microbiology Laboratory | Chan-Zuckerberg Biohub |
| EPI_ISL_444057 | UCSF Clinical Microbiology Laboratory | Chan-Zuckerberg Biohub |
| EPI_ISL_444058 | UCSF Clinical Microbiology Laboratory | Chan-Zuckerberg Biohub |
| EPI_ISL_444059 | UCSF Clinical Microbiology Laboratory | Chan-Zuckerberg Biohub |
| EPI_ISL_444060 | UCSF Clinical Microbiology Laboratory | Chan-Zuckerberg Biohub |
| EPI_ISL_444061 | UCSF Clinical Microbiology Laboratory | Chan-Zuckerberg Biohub |
| EPI_ISL_444062 | UCSF Clinical Microbiology Laboratory | Chan-Zuckerberg Biohub |

| EPI_Accession_ID | Originating Laboratory | Submitting Laboratory |
|---|---|---|
| EPI_ISL_444063 | UCSF Clinical Microbiology Laboratory | Chan-Zuckerberg Biohub |
| EPI_ISL_444064 | UCSF Clinical Microbiology Laboratory | Chan-Zuckerberg Biohub |
| EPI_ISL_444065 | UCSF Clinical Microbiology Laboratory | Chan-Zuckerberg Biohub |
| EPI_ISL_444066 | UCSF Clinical Microbiology Laboratory | Chan-Zuckerberg Biohub |
| EPI_ISL_444067 | UCSF Clinical Microbiology Laboratory | Chan-Zuckerberg Biohub |
| EPI_ISL_444068 | UCSF Clinical Microbiology Laboratory | Chan-Zuckerberg Biohub |
| EPI_ISL_444069 | UCSF Clinical Microbiology Laboratory | Chan-Zuckerberg Biohub |
| EPI_ISL_444070 | UCSF Clinical Microbiology Laboratory | Chan-Zuckerberg Biohub |
| EPI_ISL_444071 | UCSF Clinical Microbiology Laboratory | Chan-Zuckerberg Biohub |
| EPI_ISL_444072 | UCSF Clinical Microbiology Laboratory | Chan-Zuckerberg Biohub |
| EPI_ISL_444073 | UCSF Clinical Microbiology Laboratory | Chan-Zuckerberg Biohub |
| EPI_ISL_444074 | UCSF Clinical Microbiology Laboratory | Chan-Zuckerberg Biohub |
| EPI_ISL_444075 | UCSF Clinical Microbiology Laboratory | Chan-Zuckerberg Biohub |
| EPI_ISL_444076 | UCSF Clinical Microbiology Laboratory | Chan-Zuckerberg Biohub |
| EPI_ISL_444077 | UCSF Clinical Microbiology Laboratory | Chan-Zuckerberg Biohub |
| EPI_ISL_444078 | UCSF Clinical Microbiology Laboratory | Chan-Zuckerberg Biohub |
| EPI_ISL_444079 | University College London, Great Ormond Street Hospital for Children NHS Foundation Trust, Imperial College Healthcare NHS Trust | COVID-19 Genomics UK (COG-UK) Consortium |
| EPI_ISL_444080 | University College London, Great Ormond Street Hospital for Children NHS Foundation Trust, Imperial College Healthcare NHS Trust | COVID-19 Genomics UK (COG-UK) Consortium |
| EPI_ISL_444081 | University College London, Great Ormond Street Hospital for Children NHS Foundation Trust, Imperial College Healthcare NHS Trust | COVID-19 Genomics UK (COG-UK) Consortium |
| EPI_ISL_444082 | University College London, Great Ormond Street Hospital for Children NHS Foundation Trust, Imperial College Healthcare NHS Trust | COVID-19 Genomics UK (COG-UK) Consortium |
| EPI_ISL_444083 | University College London, Great Ormond Street Hospital for Children NHS Foundation Trust, Imperial College Healthcare NHS Trust | COVID-19 Genomics UK (COG-UK) Consortium |

| EPI_Accession_ID | Originating Laboratory | Submitting Laboratory |
|---|---|---|
| EPI_ISL_444084 | University College London, Great Ormond Street Hospital for Children NHS Foundation Trust, Imperial College Healthcare NHS Trust | COVID-19 Genomics UK (COG-UK) Consortium |
| EPI_ISL_444085 | University College London, Great Ormond Street Hospital for Children NHS Foundation Trust, Imperial College Healthcare NHS Trust | COVID-19 Genomics UK (COG-UK) Consortium |
| EPI_ISL_444086 | University College London, Great Ormond Street Hospital for Children NHS Foundation Trust, Imperial College Healthcare NHS Trust | COVID-19 Genomics UK (COG-UK) Consortium |
| EPI_ISL_444087 | University College London, Great Ormond Street Hospital for Children NHS Foundation Trust, Imperial College Healthcare NHS Trust | COVID-19 Genomics UK (COG-UK) Consortium |
| EPI_ISL_444088 | University College London, Great Ormond Street Hospital for Children NHS Foundation Trust, Imperial College Healthcare NHS Trust | COVID-19 Genomics UK (COG-UK) Consortium |
| EPI_ISL_444089 | University College London, Great Ormond Street Hospital for Children NHS Foundation Trust, Imperial College Healthcare NHS Trust | COVID-19 Genomics UK (COG-UK) Consortium |
| EPI_ISL_444090 | University College London, Great Ormond Street Hospital for Children NHS Foundation Trust, Imperial College Healthcare NHS Trust | COVID-19 Genomics UK (COG-UK) Consortium |
| EPI_ISL_444091 | University College London, Great Ormond Street Hospital for Children NHS Foundation Trust, Imperial College Healthcare NHS Trust | COVID-19 Genomics UK (COG-UK) Consortium |
| EPI_ISL_444092 | University College London, Great Ormond Street Hospital for Children NHS Foundation Trust, Imperial College Healthcare NHS Trust | COVID-19 Genomics UK (COG-UK) Consortium |
| EPI_ISL_444093 | University College London, Great Ormond Street Hospital for Children NHS Foundation Trust, Imperial College Healthcare NHS Trust | COVID-19 Genomics UK (COG-UK) Consortium |
| EPI_ISL_444094 | University College London, Great Ormond Street Hospital for Children NHS Foundation Trust, Imperial College Healthcare NHS Trust | COVID-19 Genomics UK (COG-UK) Consortium |

| EPI_Accession_ID | Originating Laboratory | Submitting Laboratory |
|---|---|---|
| EPI_ISL_444095 | University College London, Great Ormond Street Hospital for Children NHS Foundation Trust, Imperial College Healthcare NHS Trust | COVID-19 Genomics UK (COG-UK) Consortium |
| EPI_ISL_444096 | University College London, Great Ormond Street Hospital for Children NHS Foundation Trust, Imperial College Healthcare NHS Trust | COVID-19 Genomics UK (COG-UK) Consortium |
| EPI_ISL_444097 | University College London, Great Ormond Street Hospital for Children NHS Foundation Trust, Imperial College Healthcare NHS Trust | COVID-19 Genomics UK (COG-UK) Consortium |
| EPI_ISL_444098 | University College London, Great Ormond Street Hospital for Children NHS Foundation Trust, Imperial College Healthcare NHS Trust | COVID-19 Genomics UK (COG-UK) Consortium |
| EPI_ISL_444099 | University College London, Great Ormond Street Hospital for Children NHS Foundation Trust, Imperial College Healthcare NHS Trust | COVID-19 Genomics UK (COG-UK) Consortium |
| EPI_ISL_444100 | University College London, Great Ormond Street Hospital for Children NHS Foundation Trust, Imperial College Healthcare NHS Trust | COVID-19 Genomics UK (COG-UK) Consortium |
| EPI_ISL_444101 | University College London, Great Ormond Street Hospital for Children NHS Foundation Trust, Imperial College Healthcare NHS Trust | COVID-19 Genomics UK (COG-UK) Consortium |
| EPI_ISL_444102 | University College London, Great Ormond Street Hospital for Children NHS Foundation Trust, Imperial College Healthcare NHS Trust | COVID-19 Genomics UK (COG-UK) Consortium |
| EPI_ISL_444103 | University College London, Great Ormond Street Hospital for Children NHS Foundation Trust, Imperial College Healthcare NHS Trust | COVID-19 Genomics UK (COG-UK) Consortium |
| EPI_ISL_444104 | University College London, Great Ormond Street Hospital for Children NHS Foundation Trust, Imperial College Healthcare NHS Trust | COVID-19 Genomics UK (COG-UK) Consortium |
| EPI_ISL_444105 | University College London, Great Ormond Street Hospital for Children NHS Foundation Trust, Imperial College Healthcare NHS Trust | COVID-19 Genomics UK (COG-UK) Consortium |

| EPI_Accession_ID | Originating Laboratory | Submitting Laboratory |
|---|---|---|
| EPI_ISL_444106 | University College London, Great Ormond Street Hospital for Children NHS Foundation Trust, Imperial College Healthcare NHS Trust | COVID-19 Genomics UK (COG-UK) Consortium |
| EPI_ISL_444107 | University College London, Great Ormond Street Hospital for Children NHS Foundation Trust, Imperial College Healthcare NHS Trust | COVID-19 Genomics UK (COG-UK) Consortium |
| EPI_ISL_444108 | University College London, Great Ormond Street Hospital for Children NHS Foundation Trust, Imperial College Healthcare NHS Trust | COVID-19 Genomics UK (COG-UK) Consortium |
| EPI_ISL_444109 | University College London, Great Ormond Street Hospital for Children NHS Foundation Trust, Imperial College Healthcare NHS Trust | COVID-19 Genomics UK (COG-UK) Consortium |
| EPI_ISL_444110 | University College London, Great Ormond Street Hospital for Children NHS Foundation Trust, Imperial College Healthcare NHS Trust | COVID-19 Genomics UK (COG-UK) Consortium |
| EPI_ISL_444111 | University College London, Great Ormond Street Hospital for Children NHS Foundation Trust, Imperial College Healthcare NHS Trust | COVID-19 Genomics UK (COG-UK) Consortium |
| EPI_ISL_444112 | University College London, Great Ormond Street Hospital for Children NHS Foundation Trust, Imperial College Healthcare NHS Trust | COVID-19 Genomics UK (COG-UK) Consortium |
| EPI_ISL_444113 | University College London, Great Ormond Street Hospital for Children NHS Foundation Trust, Imperial College Healthcare NHS Trust | COVID-19 Genomics UK (COG-UK) Consortium |
| EPI_ISL_444114 | University College London, Great Ormond Street Hospital for Children NHS Foundation Trust, Imperial College Healthcare NHS Trust | COVID-19 Genomics UK (COG-UK) Consortium |
| EPI_ISL_444115 | University College London, Great Ormond Street Hospital for Children NHS Foundation Trust, Imperial College Healthcare NHS Trust | COVID-19 Genomics UK (COG-UK) Consortium |
| EPI_ISL_444116 | University College London, Great Ormond Street Hospital for Children NHS Foundation Trust, Imperial College Healthcare NHS Trust | COVID-19 Genomics UK (COG-UK) Consortium |

| EPI_Accession_ID | Originating Laboratory | Submitting Laboratory |
| --- | --- | --- |
| EPI_ISL_444117 | University College London, Great Ormond Street Hospital for Children NHS Foundation Trust, Imperial College Healthcare NHS Trust | COVID-19 Genomics UK (COG-UK) Consortium |
| EPI_ISL_444118 | University College London, Great Ormond Street Hospital for Children NHS Foundation Trust, Imperial College Healthcare NHS Trust | COVID-19 Genomics UK (COG-UK) Consortium |
| EPI_ISL_444119 | University College London, Great Ormond Street Hospital for Children NHS Foundation Trust, Imperial College Healthcare NHS Trust | COVID-19 Genomics UK (COG-UK) Consortium |
| EPI_ISL_444120 | University College London, Great Ormond Street Hospital for Children NHS Foundation Trust, Imperial College Healthcare NHS Trust | COVID-19 Genomics UK (COG-UK) Consortium |
| EPI_ISL_444121 | University College London, Great Ormond Street Hospital for Children NHS Foundation Trust, Imperial College Healthcare NHS Trust | COVID-19 Genomics UK (COG-UK) Consortium |
| EPI_ISL_444122 | University College London, Great Ormond Street Hospital for Children NHS Foundation Trust, Imperial College Healthcare NHS Trust | COVID-19 Genomics UK (COG-UK) Consortium |
| EPI_ISL_444123 | University College London, Great Ormond Street Hospital for Children NHS Foundation Trust, Imperial College Healthcare NHS Trust | COVID-19 Genomics UK (COG-UK) Consortium |
| EPI_ISL_444124 | University College London, Great Ormond Street Hospital for Children NHS Foundation Trust, Imperial College Healthcare NHS Trust | COVID-19 Genomics UK (COG-UK) Consortium |
| EPI_ISL_444125 | University College London, Great Ormond Street Hospital for Children NHS Foundation Trust, Imperial College Healthcare NHS Trust | COVID-19 Genomics UK (COG-UK) Consortium |
| EPI_ISL_444126 | University College London, Great Ormond Street Hospital for Children NHS Foundation Trust, Imperial College Healthcare NHS Trust | COVID-19 Genomics UK (COG-UK) Consortium |
| EPI_ISL_444127 | University College London, Great Ormond Street Hospital for Children NHS Foundation Trust, Imperial College Healthcare NHS Trust | COVID-19 Genomics UK (COG-UK) Consortium |

| EPI_Accession_ID | Originating Laboratory | Submitting Laboratory |
|---|---|---|
| EPI_ISL_444128 | University College London, Great Ormond Street Hospital for Children NHS Foundation Trust, Imperial College Healthcare NHS Trust | COVID-19 Genomics UK (COG-UK) Consortium |
| EPI_ISL_444129 | University College London, Great Ormond Street Hospital for Children NHS Foundation Trust, Imperial College Healthcare NHS Trust | COVID-19 Genomics UK (COG-UK) Consortium |
| EPI_ISL_444130 | University College London, Great Ormond Street Hospital for Children NHS Foundation Trust, Imperial College Healthcare NHS Trust | COVID-19 Genomics UK (COG-UK) Consortium |
| EPI_ISL_444131 | University College London, Great Ormond Street Hospital for Children NHS Foundation Trust, Imperial College Healthcare NHS Trust | COVID-19 Genomics UK (COG-UK) Consortium |
| EPI_ISL_444132 | University College London, Great Ormond Street Hospital for Children NHS Foundation Trust, Imperial College Healthcare NHS Trust | COVID-19 Genomics UK (COG-UK) Consortium |
| EPI_ISL_444133 | University College London, Great Ormond Street Hospital for Children NHS Foundation Trust, Imperial College Healthcare NHS Trust | COVID-19 Genomics UK (COG-UK) Consortium |
| EPI_ISL_444134 | University College London, Great Ormond Street Hospital for Children NHS Foundation Trust, Imperial College Healthcare NHS Trust | COVID-19 Genomics UK (COG-UK) Consortium |
| EPI_ISL_444135 | University College London, Great Ormond Street Hospital for Children NHS Foundation Trust, Imperial College Healthcare NHS Trust | COVID-19 Genomics UK (COG-UK) Consortium |
| EPI_ISL_444136 | University College London, Great Ormond Street Hospital for Children NHS Foundation Trust, Imperial College Healthcare NHS Trust | COVID-19 Genomics UK (COG-UK) Consortium |
| EPI_ISL_444137 | University College London, Great Ormond Street Hospital for Children NHS Foundation Trust, Imperial College Healthcare NHS Trust | COVID-19 Genomics UK (COG-UK) Consortium |
| EPI_ISL_444138 | University College London, Great Ormond Street Hospital for Children NHS Foundation Trust, Imperial College Healthcare NHS Trust | COVID-19 Genomics UK (COG-UK) Consortium |

| EPI_Accession_ID | Originating Laboratory | Submitting Laboratory |
|---|---|---|
| EPI_ISL_444139 | University College London, Great Ormond Street Hospital for Children NHS Foundation Trust, Imperial College Healthcare NHS Trust | COVID-19 Genomics UK (COG-UK) Consortium |
| EPI_ISL_444140 | University College London, Great Ormond Street Hospital for Children NHS Foundation Trust, Imperial College Healthcare NHS Trust | COVID-19 Genomics UK (COG-UK) Consortium |
| EPI_ISL_444141 | University College London, Great Ormond Street Hospital for Children NHS Foundation Trust, Imperial College Healthcare NHS Trust | COVID-19 Genomics UK (COG-UK) Consortium |
| EPI_ISL_444142 | University College London, Great Ormond Street Hospital for Children NHS Foundation Trust, Imperial College Healthcare NHS Trust | COVID-19 Genomics UK (COG-UK) Consortium |
| EPI_ISL_444143 | University College London, Great Ormond Street Hospital for Children NHS Foundation Trust, Imperial College Healthcare NHS Trust | COVID-19 Genomics UK (COG-UK) Consortium |
| EPI_ISL_444144 | University College London, Great Ormond Street Hospital for Children NHS Foundation Trust, Imperial College Healthcare NHS Trust | COVID-19 Genomics UK (COG-UK) Consortium |
| EPI_ISL_444145 | University College London, Great Ormond Street Hospital for Children NHS Foundation Trust, Imperial College Healthcare NHS Trust | COVID-19 Genomics UK (COG-UK) Consortium |
| EPI_ISL_444146 | University College London, Great Ormond Street Hospital for Children NHS Foundation Trust, Imperial College Healthcare NHS Trust | COVID-19 Genomics UK (COG-UK) Consortium |
| EPI_ISL_444147 | University College London, Great Ormond Street Hospital for Children NHS Foundation Trust, Imperial College Healthcare NHS Trust | COVID-19 Genomics UK (COG-UK) Consortium |
| EPI_ISL_444148 | University College London, Great Ormond Street Hospital for Children NHS Foundation Trust, Imperial College Healthcare NHS Trust | COVID-19 Genomics UK (COG-UK) Consortium |
| EPI_ISL_444149 | University College London, Great Ormond Street Hospital for Children NHS Foundation Trust, Imperial College Healthcare NHS Trust | COVID-19 Genomics UK (COG-UK) Consortium |

| EPI_Accession_ID | Originating Laboratory | Submitting Laboratory |
|---|---|---|
| EPI_ISL_444150 | University College London, Great Ormond Street Hospital for Children NHS Foundation Trust, Imperial College Healthcare NHS Trust | COVID-19 Genomics UK (COG-UK) Consortium |
| EPI_ISL_444151 | University College London, Great Ormond Street Hospital for Children NHS Foundation Trust, Imperial College Healthcare NHS Trust | COVID-19 Genomics UK (COG-UK) Consortium |
| EPI_ISL_444152 | University College London, Great Ormond Street Hospital for Children NHS Foundation Trust, Imperial College Healthcare NHS Trust | COVID-19 Genomics UK (COG-UK) Consortium |
| EPI_ISL_444153 | University College London, Great Ormond Street Hospital for Children NHS Foundation Trust, Imperial College Healthcare NHS Trust | COVID-19 Genomics UK (COG-UK) Consortium |
| EPI_ISL_444154 | University College London, Great Ormond Street Hospital for Children NHS Foundation Trust, Imperial College Healthcare NHS Trust | COVID-19 Genomics UK (COG-UK) Consortium |
| EPI_ISL_444155 | University College London, Great Ormond Street Hospital for Children NHS Foundation Trust, Imperial College Healthcare NHS Trust | COVID-19 Genomics UK (COG-UK) Consortium |
| EPI_ISL_444156 | University College London, Great Ormond Street Hospital for Children NHS Foundation Trust, Imperial College Healthcare NHS Trust | COVID-19 Genomics UK (COG-UK) Consortium |
| EPI_ISL_444157 | University College London, Great Ormond Street Hospital for Children NHS Foundation Trust, Imperial College Healthcare NHS Trust | COVID-19 Genomics UK (COG-UK) Consortium |
| EPI_ISL_444158 | University College London, Great Ormond Street Hospital for Children NHS Foundation Trust, Imperial College Healthcare NHS Trust | COVID-19 Genomics UK (COG-UK) Consortium |
| EPI_ISL_444159 | University College London, Great Ormond Street Hospital for Children NHS Foundation Trust, Imperial College Healthcare NHS Trust | COVID-19 Genomics UK (COG-UK) Consortium |
| EPI_ISL_444160 | University College London, Great Ormond Street Hospital for Children NHS Foundation Trust, Imperial College Healthcare NHS Trust | COVID-19 Genomics UK (COG-UK) Consortium |

| EPI_Accession_ID | Originating Laboratory | Submitting Laboratory |
|---|---|---|
| EPI_ISL_444161 | University College London, Great Ormond Street Hospital for Children NHS Foundation Trust, Imperial College Healthcare NHS Trust | COVID-19 Genomics UK (COG-UK) Consortium |
| EPI_ISL_444162 | University College London, Great Ormond Street Hospital for Children NHS Foundation Trust, Imperial College Healthcare NHS Trust | COVID-19 Genomics UK (COG-UK) Consortium |
| EPI_ISL_444163 | University College London, Great Ormond Street Hospital for Children NHS Foundation Trust, Imperial College Healthcare NHS Trust | COVID-19 Genomics UK (COG-UK) Consortium |
| EPI_ISL_444164 | University College London, Great Ormond Street Hospital for Children NHS Foundation Trust, Imperial College Healthcare NHS Trust | COVID-19 Genomics UK (COG-UK) Consortium |
| EPI_ISL_444165 | University College London, Great Ormond Street Hospital for Children NHS Foundation Trust, Imperial College Healthcare NHS Trust | COVID-19 Genomics UK (COG-UK) Consortium |
| EPI_ISL_444166 | University College London, Great Ormond Street Hospital for Children NHS Foundation Trust, Imperial College Healthcare NHS Trust | COVID-19 Genomics UK (COG-UK) Consortium |
| EPI_ISL_444167 | University College London, Great Ormond Street Hospital for Children NHS Foundation Trust, Imperial College Healthcare NHS Trust | COVID-19 Genomics UK (COG-UK) Consortium |
| EPI_ISL_444168 | University College London, Great Ormond Street Hospital for Children NHS Foundation Trust, Imperial College Healthcare NHS Trust | COVID-19 Genomics UK (COG-UK) Consortium |
| EPI_ISL_444169 | University College London, Great Ormond Street Hospital for Children NHS Foundation Trust, Imperial College Healthcare NHS Trust | COVID-19 Genomics UK (COG-UK) Consortium |
| EPI_ISL_444170 | University College London, Great Ormond Street Hospital for Children NHS Foundation Trust, Imperial College Healthcare NHS Trust | COVID-19 Genomics UK (COG-UK) Consortium |
| EPI_ISL_444171 | University College London, Great Ormond Street Hospital for Children NHS Foundation Trust, Imperial College Healthcare NHS Trust | COVID-19 Genomics UK (COG-UK) Consortium |

| EPI_Accession_ID | Originating Laboratory | Submitting Laboratory |
|---|---|---|
| EPI_ISL_444172 | University College London, Great Ormond Street Hospital for Children NHS Foundation Trust, Imperial College Healthcare NHS Trust | COVID-19 Genomics UK (COG-UK) Consortium |
| EPI_ISL_444173 | University College London, Great Ormond Street Hospital for Children NHS Foundation Trust, Imperial College Healthcare NHS Trust | COVID-19 Genomics UK (COG-UK) Consortium |
| EPI_ISL_444174 | University College London, Great Ormond Street Hospital for Children NHS Foundation Trust, Imperial College Healthcare NHS Trust | COVID-19 Genomics UK (COG-UK) Consortium |
| EPI_ISL_444175 | University College London, Great Ormond Street Hospital for Children NHS Foundation Trust, Imperial College Healthcare NHS Trust | COVID-19 Genomics UK (COG-UK) Consortium |
| EPI_ISL_444176 | University College London, Great Ormond Street Hospital for Children NHS Foundation Trust, Imperial College Healthcare NHS Trust | COVID-19 Genomics UK (COG-UK) Consortium |
| EPI_ISL_444177 | University College London, Great Ormond Street Hospital for Children NHS Foundation Trust, Imperial College Healthcare NHS Trust | COVID-19 Genomics UK (COG-UK) Consortium |
| EPI_ISL_444178 | University College London, Great Ormond Street Hospital for Children NHS Foundation Trust, Imperial College Healthcare NHS Trust | COVID-19 Genomics UK (COG-UK) Consortium |
| EPI_ISL_444179 | University College London, Great Ormond Street Hospital for Children NHS Foundation Trust, Imperial College Healthcare NHS Trust | COVID-19 Genomics UK (COG-UK) Consortium |
| EPI_ISL_444180 | University College London, Great Ormond Street Hospital for Children NHS Foundation Trust, Imperial College Healthcare NHS Trust | COVID-19 Genomics UK (COG-UK) Consortium |
| EPI_ISL_444181 | University College London, Great Ormond Street Hospital for Children NHS Foundation Trust, Imperial College Healthcare NHS Trust | COVID-19 Genomics UK (COG-UK) Consortium |
| EPI_ISL_444182 | University College London, Great Ormond Street Hospital for Children NHS Foundation Trust, Imperial College Healthcare NHS Trust | COVID-19 Genomics UK (COG-UK) Consortium |

| EPI_Accession_ID | Originating Laboratory | Submitting Laboratory |
|---|---|---|
| EPI_ISL_444183 | University College London, Great Ormond Street Hospital for Children NHS Foundation Trust, Imperial College Healthcare NHS Trust | COVID-19 Genomics UK (COG-UK) Consortium |
| EPI_ISL_444184 | University College London, Great Ormond Street Hospital for Children NHS Foundation Trust, Imperial College Healthcare NHS Trust | COVID-19 Genomics UK (COG-UK) Consortium |
| EPI_ISL_444185 | University College London, Great Ormond Street Hospital for Children NHS Foundation Trust, Imperial College Healthcare NHS Trust | COVID-19 Genomics UK (COG-UK) Consortium |
| EPI_ISL_444186 | University College London, Great Ormond Street Hospital for Children NHS Foundation Trust, Imperial College Healthcare NHS Trust | COVID-19 Genomics UK (COG-UK) Consortium |
| EPI_ISL_444187 | University College London, Great Ormond Street Hospital for Children NHS Foundation Trust, Imperial College Healthcare NHS Trust | COVID-19 Genomics UK (COG-UK) Consortium |
| EPI_ISL_444188 | University College London, Great Ormond Street Hospital for Children NHS Foundation Trust, Imperial College Healthcare NHS Trust | COVID-19 Genomics UK (COG-UK) Consortium |
| EPI_ISL_444189 | University College London, Great Ormond Street Hospital for Children NHS Foundation Trust, Imperial College Healthcare NHS Trust | COVID-19 Genomics UK (COG-UK) Consortium |
| EPI_ISL_444190 | University College London, Great Ormond Street Hospital for Children NHS Foundation Trust, Imperial College Healthcare NHS Trust | COVID-19 Genomics UK (COG-UK) Consortium |
| EPI_ISL_444191 | University College London, Great Ormond Street Hospital for Children NHS Foundation Trust, Imperial College Healthcare NHS Trust | COVID-19 Genomics UK (COG-UK) Consortium |
| EPI_ISL_444192 | University College London, Great Ormond Street Hospital for Children NHS Foundation Trust, Imperial College Healthcare NHS Trust | COVID-19 Genomics UK (COG-UK) Consortium |
| EPI_ISL_444193 | University College London, Great Ormond Street Hospital for Children NHS Foundation Trust, Imperial College Healthcare NHS Trust | COVID-19 Genomics UK (COG-UK) Consortium |

| EPI_Accession_ID | Originating Laboratory | Submitting Laboratory |
|---|---|---|
| EPI_ISL_444194 | University College London, Great Ormond Street Hospital for Children NHS Foundation Trust, Imperial College Healthcare NHS Trust | COVID-19 Genomics UK (COG-UK) Consortium |
| EPI_ISL_444195 | University College London, Great Ormond Street Hospital for Children NHS Foundation Trust, Imperial College Healthcare NHS Trust | COVID-19 Genomics UK (COG-UK) Consortium |
| EPI_ISL_444196 | University College London, Great Ormond Street Hospital for Children NHS Foundation Trust, Imperial College Healthcare NHS Trust | COVID-19 Genomics UK (COG-UK) Consortium |
| EPI_ISL_444197 | University College London, Great Ormond Street Hospital for Children NHS Foundation Trust, Imperial College Healthcare NHS Trust | COVID-19 Genomics UK (COG-UK) Consortium |
| EPI_ISL_444198 | University College London, Great Ormond Street Hospital for Children NHS Foundation Trust, Imperial College Healthcare NHS Trust | COVID-19 Genomics UK (COG-UK) Consortium |
| EPI_ISL_444199 | University College London, Great Ormond Street Hospital for Children NHS Foundation Trust, Imperial College Healthcare NHS Trust | COVID-19 Genomics UK (COG-UK) Consortium |
| EPI_ISL_444200 | University College London, Great Ormond Street Hospital for Children NHS Foundation Trust, Imperial College Healthcare NHS Trust | COVID-19 Genomics UK (COG-UK) Consortium |
| EPI_ISL_444201 | University College London, Great Ormond Street Hospital for Children NHS Foundation Trust, Imperial College Healthcare NHS Trust | COVID-19 Genomics UK (COG-UK) Consortium |
| EPI_ISL_444202 | University College London, Great Ormond Street Hospital for Children NHS Foundation Trust, Imperial College Healthcare NHS Trust | COVID-19 Genomics UK (COG-UK) Consortium |
| EPI_ISL_444203 | University College London, Great Ormond Street Hospital for Children NHS Foundation Trust, Imperial College Healthcare NHS Trust | COVID-19 Genomics UK (COG-UK) Consortium |
| EPI_ISL_444204 | University College London, Great Ormond Street Hospital for Children NHS Foundation Trust, Imperial College Healthcare NHS Trust | COVID-19 Genomics UK (COG-UK) Consortium |

| EPI_Accession_ID | Originating Laboratory | Submitting Laboratory |
| --- | --- | --- |
| EPI_ISL_444205 | University College London, Great Ormond Street Hospital for Children NHS Foundation Trust, Imperial College Healthcare NHS Trust | COVID-19 Genomics UK (COG-UK) Consortium |
| EPI_ISL_444206 | University College London, Great Ormond Street Hospital for Children NHS Foundation Trust, Imperial College Healthcare NHS Trust | COVID-19 Genomics UK (COG-UK) Consortium |
| EPI_ISL_444207 | University College London, Great Ormond Street Hospital for Children NHS Foundation Trust, Imperial College Healthcare NHS Trust | COVID-19 Genomics UK (COG-UK) Consortium |
| EPI_ISL_444208 | University College London, Great Ormond Street Hospital for Children NHS Foundation Trust, Imperial College Healthcare NHS Trust | COVID-19 Genomics UK (COG-UK) Consortium |
| EPI_ISL_444209 | University College London, Great Ormond Street Hospital for Children NHS Foundation Trust, Imperial College Healthcare NHS Trust | COVID-19 Genomics UK (COG-UK) Consortium |
| EPI_ISL_444210 | University College London, Great Ormond Street Hospital for Children NHS Foundation Trust, Imperial College Healthcare NHS Trust | COVID-19 Genomics UK (COG-UK) Consortium |
| EPI_ISL_444211 | University College London, Great Ormond Street Hospital for Children NHS Foundation Trust, Imperial College Healthcare NHS Trust | COVID-19 Genomics UK (COG-UK) Consortium |
| EPI_ISL_444212 | University College London, Great Ormond Street Hospital for Children NHS Foundation Trust, Imperial College Healthcare NHS Trust | COVID-19 Genomics UK (COG-UK) Consortium |
| EPI_ISL_444213 | University College London, Great Ormond Street Hospital for Children NHS Foundation Trust, Imperial College Healthcare NHS Trust | COVID-19 Genomics UK (COG-UK) Consortium |
| EPI_ISL_444214 | University College London, Great Ormond Street Hospital for Children NHS Foundation Trust, Imperial College Healthcare NHS Trust | COVID-19 Genomics UK (COG-UK) Consortium |
| EPI_ISL_444215 | University College London, Great Ormond Street Hospital for Children NHS Foundation Trust, Imperial College Healthcare NHS Trust | COVID-19 Genomics UK (COG-UK) Consortium |

| EPI_Accession_ID | Originating Laboratory | Submitting Laboratory |
|---|---|---|
| EPI_ISL_444216 | University College London, Great Ormond Street Hospital for Children NHS Foundation Trust, Imperial College Healthcare NHS Trust | COVID-19 Genomics UK (COG-UK) Consortium |
| EPI_ISL_444217 | University College London, Great Ormond Street Hospital for Children NHS Foundation Trust, Imperial College Healthcare NHS Trust | COVID-19 Genomics UK (COG-UK) Consortium |
| EPI_ISL_444218 | University College London, Great Ormond Street Hospital for Children NHS Foundation Trust, Imperial College Healthcare NHS Trust | COVID-19 Genomics UK (COG-UK) Consortium |
| EPI_ISL_444219 | University College London, Great Ormond Street Hospital for Children NHS Foundation Trust, Imperial College Healthcare NHS Trust | COVID-19 Genomics UK (COG-UK) Consortium |
| EPI_ISL_444220 | University College London, Great Ormond Street Hospital for Children NHS Foundation Trust, Imperial College Healthcare NHS Trust | COVID-19 Genomics UK (COG-UK) Consortium |
| EPI_ISL_444221 | University College London, Great Ormond Street Hospital for Children NHS Foundation Trust, Imperial College Healthcare NHS Trust | COVID-19 Genomics UK (COG-UK) Consortium |
| EPI_ISL_444222 | University College London, Great Ormond Street Hospital for Children NHS Foundation Trust, Imperial College Healthcare NHS Trust | COVID-19 Genomics UK (COG-UK) Consortium |
| EPI_ISL_444223 | University College London, Great Ormond Street Hospital for Children NHS Foundation Trust, Imperial College Healthcare NHS Trust | COVID-19 Genomics UK (COG-UK) Consortium |
| EPI_ISL_444224 | University College London, Great Ormond Street Hospital for Children NHS Foundation Trust, Imperial College Healthcare NHS Trust | COVID-19 Genomics UK (COG-UK) Consortium |
| EPI_ISL_444225 | University College London, Great Ormond Street Hospital for Children NHS Foundation Trust, Imperial College Healthcare NHS Trust | COVID-19 Genomics UK (COG-UK) Consortium |
| EPI_ISL_444226 | University College London, Great Ormond Street Hospital for Children NHS Foundation Trust, Imperial College Healthcare NHS Trust | COVID-19 Genomics UK (COG-UK) Consortium |

| EPI_Accession_ID | Originating Laboratory | Submitting Laboratory |
|---|---|---|
| EPI_ISL_444227 | University College London, Great Ormond Street Hospital for Children NHS Foundation Trust, Imperial College Healthcare NHS Trust | COVID-19 Genomics UK (COG-UK) Consortium |
| EPI_ISL_444228 | University College London, Great Ormond Street Hospital for Children NHS Foundation Trust, Imperial College Healthcare NHS Trust | COVID-19 Genomics UK (COG-UK) Consortium |
| EPI_ISL_444229 | University College London, Great Ormond Street Hospital for Children NHS Foundation Trust, Imperial College Healthcare NHS Trust | COVID-19 Genomics UK (COG-UK) Consortium |
| EPI_ISL_444230 | University College London, Great Ormond Street Hospital for Children NHS Foundation Trust, Imperial College Healthcare NHS Trust | COVID-19 Genomics UK (COG-UK) Consortium |
| EPI_ISL_444231 | University College London, Great Ormond Street Hospital for Children NHS Foundation Trust, Imperial College Healthcare NHS Trust | COVID-19 Genomics UK (COG-UK) Consortium |
| EPI_ISL_444232 | University College London, Great Ormond Street Hospital for Children NHS Foundation Trust, Imperial College Healthcare NHS Trust | COVID-19 Genomics UK (COG-UK) Consortium |
| EPI_ISL_444233 | University College London, Great Ormond Street Hospital for Children NHS Foundation Trust, Imperial College Healthcare NHS Trust | COVID-19 Genomics UK (COG-UK) Consortium |
| EPI_ISL_444234 | University College London, Great Ormond Street Hospital for Children NHS Foundation Trust, Imperial College Healthcare NHS Trust | COVID-19 Genomics UK (COG-UK) Consortium |
| EPI_ISL_444235 | University College London, Great Ormond Street Hospital for Children NHS Foundation Trust, Imperial College Healthcare NHS Trust | COVID-19 Genomics UK (COG-UK) Consortium |
| EPI_ISL_444236 | University College London, Great Ormond Street Hospital for Children NHS Foundation Trust, Imperial College Healthcare NHS Trust | COVID-19 Genomics UK (COG-UK) Consortium |
| EPI_ISL_444237 | University College London, Great Ormond Street Hospital for Children NHS Foundation Trust, Imperial College Healthcare NHS Trust | COVID-19 Genomics UK (COG-UK) Consortium |

| EPI_Accession_ID | Originating Laboratory | Submitting Laboratory |
|---|---|---|
| EPI_ISL_444238 | University College London, Great Ormond Street Hospital for Children NHS Foundation Trust, Imperial College Healthcare NHS Trust | COVID-19 Genomics UK (COG-UK) Consortium |
| EPI_ISL_444239 | University College London, Great Ormond Street Hospital for Children NHS Foundation Trust, Imperial College Healthcare NHS Trust | COVID-19 Genomics UK (COG-UK) Consortium |
| EPI_ISL_444240 | University College London, Great Ormond Street Hospital for Children NHS Foundation Trust, Imperial College Healthcare NHS Trust | COVID-19 Genomics UK (COG-UK) Consortium |
| EPI_ISL_444241 | University College London, Great Ormond Street Hospital for Children NHS Foundation Trust, Imperial College Healthcare NHS Trust | COVID-19 Genomics UK (COG-UK) Consortium |
| EPI_ISL_444242 | University College London, Great Ormond Street Hospital for Children NHS Foundation Trust, Imperial College Healthcare NHS Trust | COVID-19 Genomics UK (COG-UK) Consortium |
| EPI_ISL_444243 | University College London, Great Ormond Street Hospital for Children NHS Foundation Trust, Imperial College Healthcare NHS Trust | COVID-19 Genomics UK (COG-UK) Consortium |
| EPI_ISL_444244 | University College London, Great Ormond Street Hospital for Children NHS Foundation Trust, Imperial College Healthcare NHS Trust | COVID-19 Genomics UK (COG-UK) Consortium |
| EPI_ISL_444245 | University College London, Great Ormond Street Hospital for Children NHS Foundation Trust, Imperial College Healthcare NHS Trust | COVID-19 Genomics UK (COG-UK) Consortium |
| EPI_ISL_444246 | University College London, Great Ormond Street Hospital for Children NHS Foundation Trust, Imperial College Healthcare NHS Trust | COVID-19 Genomics UK (COG-UK) Consortium |
| EPI_ISL_444247 | University College London, Great Ormond Street Hospital for Children NHS Foundation Trust, Imperial College Healthcare NHS Trust | COVID-19 Genomics UK (COG-UK) Consortium |
| EPI_ISL_444248 | University College London, Great Ormond Street Hospital for Children NHS Foundation Trust, Imperial College Healthcare NHS Trust | COVID-19 Genomics UK (COG-UK) Consortium |

| EPI_Accession_ID | Originating Laboratory | Submitting Laboratory |
|---|---|---|
| EPI_ISL_444249 | University College London, Great Ormond Street Hospital for Children NHS Foundation Trust, Imperial College Healthcare NHS Trust | COVID-19 Genomics UK (COG-UK) Consortium |
| EPI_ISL_444250 | University College London, Great Ormond Street Hospital for Children NHS Foundation Trust, Imperial College Healthcare NHS Trust | COVID-19 Genomics UK (COG-UK) Consortium |
| EPI_ISL_444251 | University College London, Great Ormond Street Hospital for Children NHS Foundation Trust, Imperial College Healthcare NHS Trust | COVID-19 Genomics UK (COG-UK) Consortium |
| EPI_ISL_444252 | University College London, Great Ormond Street Hospital for Children NHS Foundation Trust, Imperial College Healthcare NHS Trust | COVID-19 Genomics UK (COG-UK) Consortium |
| EPI_ISL_444253 | University College London, Great Ormond Street Hospital for Children NHS Foundation Trust, Imperial College Healthcare NHS Trust | COVID-19 Genomics UK (COG-UK) Consortium |
| EPI_ISL_444254 | University College London, Great Ormond Street Hospital for Children NHS Foundation Trust, Imperial College Healthcare NHS Trust | COVID-19 Genomics UK (COG-UK) Consortium |
| EPI_ISL_444255 | University College London, Great Ormond Street Hospital for Children NHS Foundation Trust, Imperial College Healthcare NHS Trust | COVID-19 Genomics UK (COG-UK) Consortium |
| EPI_ISL_444256 | University College London, Great Ormond Street Hospital for Children NHS Foundation Trust, Imperial College Healthcare NHS Trust | COVID-19 Genomics UK (COG-UK) Consortium |
| EPI_ISL_444257 | University College London, Great Ormond Street Hospital for Children NHS Foundation Trust, Imperial College Healthcare NHS Trust | COVID-19 Genomics UK (COG-UK) Consortium |
| EPI_ISL_444258 | University College London, Great Ormond Street Hospital for Children NHS Foundation Trust, Imperial College Healthcare NHS Trust | COVID-19 Genomics UK (COG-UK) Consortium |
| EPI_ISL_444259 | University College London, Great Ormond Street Hospital for Children NHS Foundation Trust, Imperial College Healthcare NHS Trust | COVID-19 Genomics UK (COG-UK) Consortium |

| EPI_Accession_ID | Originating Laboratory | Submitting Laboratory |
|---|---|---|
| EPI_ISL_444260 | University College London, Great Ormond Street Hospital for Children NHS Foundation Trust, Imperial College Healthcare NHS Trust | COVID-19 Genomics UK (COG-UK) Consortium |
| EPI_ISL_444261 | University College London, Great Ormond Street Hospital for Children NHS Foundation Trust, Imperial College Healthcare NHS Trust | COVID-19 Genomics UK (COG-UK) Consortium |
| EPI_ISL_444262 | University College London, Great Ormond Street Hospital for Children NHS Foundation Trust, Imperial College Healthcare NHS Trust | COVID-19 Genomics UK (COG-UK) Consortium |
| EPI_ISL_444263 | University College London, Great Ormond Street Hospital for Children NHS Foundation Trust, Imperial College Healthcare NHS Trust | COVID-19 Genomics UK (COG-UK) Consortium |
| EPI_ISL_444264 | University College London, Great Ormond Street Hospital for Children NHS Foundation Trust, Imperial College Healthcare NHS Trust | COVID-19 Genomics UK (COG-UK) Consortium |
| EPI_ISL_444265 | University College London, Great Ormond Street Hospital for Children NHS Foundation Trust, Imperial College Healthcare NHS Trust | COVID-19 Genomics UK (COG-UK) Consortium |
| EPI_ISL_444266 | University College London, Great Ormond Street Hospital for Children NHS Foundation Trust, Imperial College Healthcare NHS Trust | COVID-19 Genomics UK (COG-UK) Consortium |
| EPI_ISL_444267 | University College London, Great Ormond Street Hospital for Children NHS Foundation Trust, Imperial College Healthcare NHS Trust | COVID-19 Genomics UK (COG-UK) Consortium |
| EPI_ISL_444268 | University College London, Great Ormond Street Hospital for Children NHS Foundation Trust, Imperial College Healthcare NHS Trust | COVID-19 Genomics UK (COG-UK) Consortium |
| EPI_ISL_444269 | University College London, Great Ormond Street Hospital for Children NHS Foundation Trust, Imperial College Healthcare NHS Trust | COVID-19 Genomics UK (COG-UK) Consortium |
| EPI_ISL_444270 | University College London, Great Ormond Street Hospital for Children NHS Foundation Trust, Imperial College Healthcare NHS Trust | COVID-19 Genomics UK (COG-UK) Consortium |

| EPI_Accession_ID | Originating Laboratory | Submitting Laboratory |
|---|---|---|
| EPI_ISL_444271 | University College London, Great Ormond Street Hospital for Children NHS Foundation Trust, Imperial College Healthcare NHS Trust | COVID-19 Genomics UK (COG-UK) Consortium |
| EPI_ISL_444272 | University College London, Great Ormond Street Hospital for Children NHS Foundation Trust, Imperial College Healthcare NHS Trust | COVID-19 Genomics UK (COG-UK) Consortium |
| EPI_ISL_444273 | unknown | State Key Laboratory of Respiratory Disease |
| EPI_ISL_444274 | Laboratory Medicine | Department of Laboratory Medicine, Lin-Kou Chang Gung Memorial Hospital, Taoyuan, Taiwan |
| EPI_ISL_444275 | Laboratory Medicine | Department of Laboratory Medicine, Lin-Kou Chang Gung Memorial Hospital, Taoyuan, Taiwan |
| EPI_ISL_444276 | Laboratory Medicine | Department of Laboratory Medicine, Lin-Kou Chang Gung Memorial Hospital, Taoyuan, Taiwan |
| EPI_ISL_444277 | Laboratory Medicine | Department of Laboratory Medicine, Lin-Kou Chang Gung Memorial Hospital, Taoyuan, Taiwan |
| EPI_ISL_444278 | Laboratory Medicine | Department of Laboratory Medicine, Lin-Kou Chang Gung Memorial Hospital, Taoyuan, Taiwan |
| EPI_ISL_444279 | University of Birmingham | COVID-19 Genomics UK (COG-UK) Consortium |
| EPI_ISL_444280 | University of Birmingham | COVID-19 Genomics UK (COG-UK) Consortium |
| EPI_ISL_444281 | University of Birmingham | COVID-19 Genomics UK (COG-UK) Consortium |
| EPI_ISL_444282 | University of Birmingham | COVID-19 Genomics UK (COG-UK) Consortium |
| EPI_ISL_444283 | University of Birmingham | COVID-19 Genomics UK (COG-UK) Consortium |
| EPI_ISL_444284 | University of Birmingham | COVID-19 Genomics UK (COG-UK) Consortium |
| EPI_ISL_444285 | University of Birmingham | COVID-19 Genomics UK (COG-UK) Consortium |
| EPI_ISL_444286 | University of Birmingham | COVID-19 Genomics UK (COG-UK) Consortium |
| EPI_ISL_444287 | University of Birmingham | COVID-19 Genomics UK (COG-UK) Consortium |
| EPI_ISL_444288 | University of Birmingham | COVID-19 Genomics UK (COG-UK) Consortium |
| EPI_ISL_444289 | University of Birmingham | COVID-19 Genomics UK (COG-UK) Consortium |
| EPI_ISL_444290 | University of Birmingham | COVID-19 Genomics UK (COG-UK) Consortium |
| EPI_ISL_444291 | University of Birmingham | COVID-19 Genomics UK (COG-UK) Consortium |
| EPI_ISL_444292 | University of Birmingham | COVID-19 Genomics UK (COG-UK) Consortium |
| EPI_ISL_444293 | University of Birmingham | COVID-19 Genomics UK (COG-UK) Consortium |
| EPI_ISL_444294 | University of Birmingham | COVID-19 Genomics UK (COG-UK) Consortium |

| EPI_Accession_ID | Originating Laboratory | Submitting Laboratory |
|---|---|---|
| EPI_ISL_444295 | University of Birmingham | COVID-19 Genomics UK (COG-UK) Consortium |
| EPI_ISL_444296 | University of Birmingham | COVID-19 Genomics UK (COG-UK) Consortium |
| EPI_ISL_444297 | University of Birmingham | COVID-19 Genomics UK (COG-UK) Consortium |
| EPI_ISL_444298 | University of Birmingham | COVID-19 Genomics UK (COG-UK) Consortium |
| EPI_ISL_444299 | University of Birmingham | COVID-19 Genomics UK (COG-UK) Consortium |
| EPI_ISL_444300 | University of Birmingham | COVID-19 Genomics UK (COG-UK) Consortium |
| EPI_ISL_444301 | University of Birmingham | COVID-19 Genomics UK (COG-UK) Consortium |
| EPI_ISL_444302 | University of Birmingham | COVID-19 Genomics UK (COG-UK) Consortium |
| EPI_ISL_444303 | University of Birmingham | COVID-19 Genomics UK (COG-UK) Consortium |
| EPI_ISL_444304 | University of Birmingham | COVID-19 Genomics UK (COG-UK) Consortium |
| EPI_ISL_444305 | University of Birmingham | COVID-19 Genomics UK (COG-UK) Consortium |
| EPI_ISL_444306 | University of Birmingham | COVID-19 Genomics UK (COG-UK) Consortium |
| EPI_ISL_444307 | University of Birmingham | COVID-19 Genomics UK (COG-UK) Consortium |
| EPI_ISL_444308 | University of Birmingham | COVID-19 Genomics UK (COG-UK) Consortium |
| EPI_ISL_444309 | University of Birmingham | COVID-19 Genomics UK (COG-UK) Consortium |
| EPI_ISL_444310 | University of Birmingham | COVID-19 Genomics UK (COG-UK) Consortium |
| EPI_ISL_444311 | University of Birmingham | COVID-19 Genomics UK (COG-UK) Consortium |
| EPI_ISL_444312 | University of Birmingham | COVID-19 Genomics UK (COG-UK) Consortium |
| EPI_ISL_444313 | Department of Pathology, University of Cambridge | COVID-19 Genomics UK (COG-UK) Consortium |
| EPI_ISL_444314 | Department of Pathology, University of Cambridge | COVID-19 Genomics UK (COG-UK) Consortium |
| EPI_ISL_444315 | Department of Pathology, University of Cambridge | COVID-19 Genomics UK (COG-UK) Consortium |
| EPI_ISL_444316 | Department of Pathology, University of Cambridge | COVID-19 Genomics UK (COG-UK) Consortium |
| EPI_ISL_444317 | Department of Pathology, University of Cambridge | COVID-19 Genomics UK (COG-UK) Consortium |
| EPI_ISL_444318 | Department of Pathology, University of Cambridge | COVID-19 Genomics UK (COG-UK) Consortium |
| EPI_ISL_444319 | Department of Pathology, University of Cambridge | COVID-19 Genomics UK (COG-UK) Consortium |
| EPI_ISL_444320 | Department of Pathology, University of Cambridge | COVID-19 Genomics UK (COG-UK) Consortium |
| EPI_ISL_444321 | Department of Pathology, University of Cambridge | COVID-19 Genomics UK (COG-UK) Consortium |
| EPI_ISL_444322 | Department of Pathology, University of Cambridge | COVID-19 Genomics UK (COG-UK) Consortium |
| EPI_ISL_444323 | Department of Pathology, University of Cambridge | COVID-19 Genomics UK (COG-UK) Consortium |
| EPI_ISL_444324 | Department of Pathology, University of Cambridge | COVID-19 Genomics UK (COG-UK) Consortium |
| EPI_ISL_444325 | Department of Pathology, University of Cambridge | COVID-19 Genomics UK (COG-UK) Consortium |
| EPI_ISL_444326 | Department of Pathology, University of Cambridge | COVID-19 Genomics UK (COG-UK) Consortium |
| EPI_ISL_444327 | Department of Pathology, University of Cambridge | COVID-19 Genomics UK (COG-UK) Consortium |

| EPI_Accession_ID | Originating Laboratory | Submitting Laboratory |
|---|---|---|
| EPI_ISL_444328 | Department of Pathology, University of Cambridge | COVID-19 Genomics UK (COG-UK) Consortium |
| EPI_ISL_444329 | Department of Pathology, University of Cambridge | COVID-19 Genomics UK (COG-UK) Consortium |
| EPI_ISL_444330 | Department of Pathology, University of Cambridge | COVID-19 Genomics UK (COG-UK) Consortium |
| EPI_ISL_444331 | Department of Pathology, University of Cambridge | COVID-19 Genomics UK (COG-UK) Consortium |
| EPI_ISL_444332 | Department of Pathology, University of Cambridge | COVID-19 Genomics UK (COG-UK) Consortium |
| EPI_ISL_444333 | Department of Pathology, University of Cambridge | COVID-19 Genomics UK (COG-UK) Consortium |
| EPI_ISL_444334 | Department of Pathology, University of Cambridge | COVID-19 Genomics UK (COG-UK) Consortium |
| EPI_ISL_444335 | Department of Pathology, University of Cambridge | COVID-19 Genomics UK (COG-UK) Consortium |
| EPI_ISL_444336 | Department of Pathology, University of Cambridge | COVID-19 Genomics UK (COG-UK) Consortium |
| EPI_ISL_444337 | Department of Pathology, University of Cambridge | COVID-19 Genomics UK (COG-UK) Consortium |
| EPI_ISL_444338 | Department of Pathology, University of Cambridge | COVID-19 Genomics UK (COG-UK) Consortium |
| EPI_ISL_444339 | Department of Pathology, University of Cambridge | COVID-19 Genomics UK (COG-UK) Consortium |
| EPI_ISL_444340 | Department of Pathology, University of Cambridge | COVID-19 Genomics UK (COG-UK) Consortium |
| EPI_ISL_444341 | Department of Pathology, University of Cambridge | COVID-19 Genomics UK (COG-UK) Consortium |
| EPI_ISL_444342 | Department of Pathology, University of Cambridge | COVID-19 Genomics UK (COG-UK) Consortium |
| EPI_ISL_444344 | Department of Pathology, University of Cambridge | COVID-19 Genomics UK (COG-UK) Consortium |
| EPI_ISL_444346 | Department of Pathology, University of Cambridge | COVID-19 Genomics UK (COG-UK) Consortium |
| EPI_ISL_444347 | Department of Pathology, University of Cambridge | COVID-19 Genomics UK (COG-UK) Consortium |
| EPI_ISL_444348 | Department of Pathology, University of Cambridge | COVID-19 Genomics UK (COG-UK) Consortium |
| EPI_ISL_444350 | Department of Pathology, University of Cambridge | COVID-19 Genomics UK (COG-UK) Consortium |
| EPI_ISL_444351 | Department of Pathology, University of Cambridge | COVID-19 Genomics UK (COG-UK) Consortium |
| EPI_ISL_444352 | Department of Pathology, University of Cambridge | COVID-19 Genomics UK (COG-UK) Consortium |
| EPI_ISL_444353 | Department of Pathology, University of Cambridge | COVID-19 Genomics UK (COG-UK) Consortium |
| EPI_ISL_444354 | Department of Pathology, University of Cambridge | COVID-19 Genomics UK (COG-UK) Consortium |
| EPI_ISL_444356 | Department of Pathology, University of Cambridge | COVID-19 Genomics UK (COG-UK) Consortium |
| EPI_ISL_444357 | Department of Pathology, University of Cambridge | COVID-19 Genomics UK (COG-UK) Consortium |
| EPI_ISL_444358 | Department of Pathology, University of Cambridge | COVID-19 Genomics UK (COG-UK) Consortium |
| EPI_ISL_444359 | Department of Pathology, University of Cambridge | COVID-19 Genomics UK (COG-UK) Consortium |
| EPI_ISL_444360 | Department of Pathology, University of Cambridge | COVID-19 Genomics UK (COG-UK) Consortium |
| EPI_ISL_444361 | Department of Pathology, University of Cambridge | COVID-19 Genomics UK (COG-UK) Consortium |
| EPI_ISL_444362 | Department of Pathology, University of Cambridge | COVID-19 Genomics UK (COG-UK) Consortium |
| EPI_ISL_444363 | Department of Pathology, University of Cambridge | COVID-19 Genomics UK (COG-UK) Consortium |
| EPI_ISL_444364 | Department of Pathology, University of Cambridge | COVID-19 Genomics UK (COG-UK) Consortium |

| EPI_Accession_ID | Originating Laboratory | Submitting Laboratory |
|---|---|---|
| EPI_ISL_444365 | Department of Pathology, University of Cambridge | COVID-19 Genomics UK (COG-UK) Consortium |
| EPI_ISL_444366 | Department of Pathology, University of Cambridge | COVID-19 Genomics UK (COG-UK) Consortium |
| EPI_ISL_444367 | Department of Pathology, University of Cambridge | COVID-19 Genomics UK (COG-UK) Consortium |
| EPI_ISL_444368 | Department of Pathology, University of Cambridge | COVID-19 Genomics UK (COG-UK) Consortium |
| EPI_ISL_444369 | Department of Pathology, University of Cambridge | COVID-19 Genomics UK (COG-UK) Consortium |
| EPI_ISL_444370 | Department of Pathology, University of Cambridge | COVID-19 Genomics UK (COG-UK) Consortium |
| EPI_ISL_444371 | Department of Pathology, University of Cambridge | COVID-19 Genomics UK (COG-UK) Consortium |
| EPI_ISL_444372 | Department of Pathology, University of Cambridge | COVID-19 Genomics UK (COG-UK) Consortium |
| EPI_ISL_444373 | Department of Pathology, University of Cambridge | COVID-19 Genomics UK (COG-UK) Consortium |
| EPI_ISL_444374 | Department of Pathology, University of Cambridge | COVID-19 Genomics UK (COG-UK) Consortium |
| EPI_ISL_444376 | Department of Pathology, University of Cambridge | COVID-19 Genomics UK (COG-UK) Consortium |
| EPI_ISL_444377 | Department of Pathology, University of Cambridge | COVID-19 Genomics UK (COG-UK) Consortium |
| EPI_ISL_444378 | Department of Pathology, University of Cambridge | COVID-19 Genomics UK (COG-UK) Consortium |
| EPI_ISL_444379 | Department of Pathology, University of Cambridge | COVID-19 Genomics UK (COG-UK) Consortium |
| EPI_ISL_444380 | Department of Pathology, University of Cambridge | COVID-19 Genomics UK (COG-UK) Consortium |
| EPI_ISL_444381 | Department of Pathology, University of Cambridge | COVID-19 Genomics UK (COG-UK) Consortium |
| EPI_ISL_444382 | Department of Pathology, University of Cambridge | COVID-19 Genomics UK (COG-UK) Consortium |
| EPI_ISL_444384 | Department of Pathology, University of Cambridge | COVID-19 Genomics UK (COG-UK) Consortium |
| EPI_ISL_444385 | Department of Pathology, University of Cambridge | COVID-19 Genomics UK (COG-UK) Consortium |
| EPI_ISL_444386 | Department of Pathology, University of Cambridge | COVID-19 Genomics UK (COG-UK) Consortium |
| EPI_ISL_444387 | Department of Pathology, University of Cambridge | COVID-19 Genomics UK (COG-UK) Consortium |
| EPI_ISL_444388 | Department of Pathology, University of Cambridge | COVID-19 Genomics UK (COG-UK) Consortium |
| EPI_ISL_444389 | Department of Pathology, University of Cambridge | COVID-19 Genomics UK (COG-UK) Consortium |
| EPI_ISL_444390 | Department of Pathology, University of Cambridge | COVID-19 Genomics UK (COG-UK) Consortium |
| EPI_ISL_444391 | Department of Pathology, University of Cambridge | COVID-19 Genomics UK (COG-UK) Consortium |
| EPI_ISL_444392 | Department of Pathology, University of Cambridge | COVID-19 Genomics UK (COG-UK) Consortium |
| EPI_ISL_444393 | Department of Pathology, University of Cambridge | COVID-19 Genomics UK (COG-UK) Consortium |
| EPI_ISL_444394 | Department of Pathology, University of Cambridge | COVID-19 Genomics UK (COG-UK) Consortium |
| EPI_ISL_444395 | Department of Pathology, University of Cambridge | COVID-19 Genomics UK (COG-UK) Consortium |
| EPI_ISL_444396 | Department of Pathology, University of Cambridge | COVID-19 Genomics UK (COG-UK) Consortium |
| EPI_ISL_444397 | Department of Pathology, University of Cambridge | COVID-19 Genomics UK (COG-UK) Consortium |
| EPI_ISL_444398 | Department of Pathology, University of Cambridge | COVID-19 Genomics UK (COG-UK) Consortium |
| EPI_ISL_444399 | Department of Pathology, University of Cambridge | COVID-19 Genomics UK (COG-UK) Consortium |

| EPI_Accession_ID | Originating Laboratory | Submitting Laboratory |
|---|---|---|
| EPI_ISL_444400 | Department of Pathology, University of Cambridge | COVID-19 Genomics UK (COG-UK) Consortium |
| EPI_ISL_444401 | Department of Pathology, University of Cambridge | COVID-19 Genomics UK (COG-UK) Consortium |
| EPI_ISL_444402 | Department of Pathology, University of Cambridge | COVID-19 Genomics UK (COG-UK) Consortium |
| EPI_ISL_444403 | Department of Pathology, University of Cambridge | COVID-19 Genomics UK (COG-UK) Consortium |
| EPI_ISL_444404 | Department of Pathology, University of Cambridge | COVID-19 Genomics UK (COG-UK) Consortium |
| EPI_ISL_444405 | Department of Pathology, University of Cambridge | COVID-19 Genomics UK (COG-UK) Consortium |
| EPI_ISL_444406 | Department of Pathology, University of Cambridge | COVID-19 Genomics UK (COG-UK) Consortium |
| EPI_ISL_444407 | Department of Pathology, University of Cambridge | COVID-19 Genomics UK (COG-UK) Consortium |
| EPI_ISL_444408 | Department of Pathology, University of Cambridge | COVID-19 Genomics UK (COG-UK) Consortium |
| EPI_ISL_444410 | Department of Pathology, University of Cambridge | COVID-19 Genomics UK (COG-UK) Consortium |
| EPI_ISL_444411 | Department of Pathology, University of Cambridge | COVID-19 Genomics UK (COG-UK) Consortium |
| EPI_ISL_444412 | Department of Pathology, University of Cambridge | COVID-19 Genomics UK (COG-UK) Consortium |
| EPI_ISL_444413 | Department of Pathology, University of Cambridge | COVID-19 Genomics UK (COG-UK) Consortium |
| EPI_ISL_444414 | Department of Pathology, University of Cambridge | COVID-19 Genomics UK (COG-UK) Consortium |
| EPI_ISL_444415 | Department of Pathology, University of Cambridge | COVID-19 Genomics UK (COG-UK) Consortium |
| EPI_ISL_444416 | Department of Pathology, University of Cambridge | COVID-19 Genomics UK (COG-UK) Consortium |
| EPI_ISL_444417 | Department of Pathology, University of Cambridge | COVID-19 Genomics UK (COG-UK) Consortium |
| EPI_ISL_444418 | Department of Pathology, University of Cambridge | COVID-19 Genomics UK (COG-UK) Consortium |
| EPI_ISL_444419 | Department of Pathology, University of Cambridge | COVID-19 Genomics UK (COG-UK) Consortium |
| EPI_ISL_444420 | Department of Pathology, University of Cambridge | COVID-19 Genomics UK (COG-UK) Consortium |
| EPI_ISL_444421 | Department of Pathology, University of Cambridge | COVID-19 Genomics UK (COG-UK) Consortium |
| EPI_ISL_444422 | Department of Pathology, University of Cambridge | COVID-19 Genomics UK (COG-UK) Consortium |
| EPI_ISL_444423 | Department of Pathology, University of Cambridge | COVID-19 Genomics UK (COG-UK) Consortium |
| EPI_ISL_444424 | Department of Pathology, University of Cambridge | COVID-19 Genomics UK (COG-UK) Consortium |
| EPI_ISL_444425 | Department of Pathology, University of Cambridge | COVID-19 Genomics UK (COG-UK) Consortium |
| EPI_ISL_444426 | Department of Pathology, University of Cambridge | COVID-19 Genomics UK (COG-UK) Consortium |
| EPI_ISL_444427 | Department of Pathology, University of Cambridge | COVID-19 Genomics UK (COG-UK) Consortium |
| EPI_ISL_444428 | Department of Pathology, University of Cambridge | COVID-19 Genomics UK (COG-UK) Consortium |
| EPI_ISL_444429 | Department of Pathology, University of Cambridge | COVID-19 Genomics UK (COG-UK) Consortium |
| EPI_ISL_444430 | Department of Pathology, University of Cambridge | COVID-19 Genomics UK (COG-UK) Consortium |
| EPI_ISL_444431 | Department of Pathology, University of Cambridge | COVID-19 Genomics UK (COG-UK) Consortium |
| EPI_ISL_444432 | Department of Pathology, University of Cambridge | COVID-19 Genomics UK (COG-UK) Consortium |
| EPI_ISL_444433 | Department of Pathology, University of Cambridge | COVID-19 Genomics UK (COG-UK) Consortium |

| EPI_Accession_ID | Originating Laboratory | Submitting Laboratory |
|---|---|---|
| EPI_ISL_444435 | Department of Pathology, University of Cambridge | COVID-19 Genomics UK (COG-UK) Consortium |
| EPI_ISL_444436 | Department of Pathology, University of Cambridge | COVID-19 Genomics UK (COG-UK) Consortium |
| EPI_ISL_444437 | Department of Pathology, University of Cambridge | COVID-19 Genomics UK (COG-UK) Consortium |
| EPI_ISL_444438 | Department of Pathology, University of Cambridge | COVID-19 Genomics UK (COG-UK) Consortium |
| EPI_ISL_444439 | Department of Pathology, University of Cambridge | COVID-19 Genomics UK (COG-UK) Consortium |
| EPI_ISL_444440 | Department of Pathology, University of Cambridge | COVID-19 Genomics UK (COG-UK) Consortium |
| EPI_ISL_444441 | Department of Pathology, University of Cambridge | COVID-19 Genomics UK (COG-UK) Consortium |
| EPI_ISL_444442 | Department of Pathology, University of Cambridge | COVID-19 Genomics UK (COG-UK) Consortium |
| EPI_ISL_444443 | Department of Pathology, University of Cambridge | COVID-19 Genomics UK (COG-UK) Consortium |
| EPI_ISL_444446 | Department of Pathology, University of Cambridge | COVID-19 Genomics UK (COG-UK) Consortium |
| EPI_ISL_444447 | Department of Pathology, University of Cambridge | COVID-19 Genomics UK (COG-UK) Consortium |
| EPI_ISL_444448 | Department of Pathology, University of Cambridge | COVID-19 Genomics UK (COG-UK) Consortium |
| EPI_ISL_444449 | Department of Pathology, University of Cambridge | COVID-19 Genomics UK (COG-UK) Consortium |
| EPI_ISL_444450 | Department of Pathology, University of Cambridge | COVID-19 Genomics UK (COG-UK) Consortium |
| EPI_ISL_444451 | Department of Pathology, University of Cambridge | COVID-19 Genomics UK (COG-UK) Consortium |
| EPI_ISL_444452 | Department of Pathology, University of Cambridge | COVID-19 Genomics UK (COG-UK) Consortium |
| EPI_ISL_444453 | Department of Pathology, University of Cambridge | COVID-19 Genomics UK (COG-UK) Consortium |
| EPI_ISL_444455 | unknown | Molecular Infectious Disease |
| EPI_ISL_444456 | B.J. Medical College and Civil hospital | Gujarat Biotechnology Research Centre |
| EPI_ISL_444457 | B.J. Medical College and Civil hospital | Gujarat Biotechnology Research Centre |
| EPI_ISL_444458 | B.J. Medical College and Civil hospital | Gujarat Biotechnology Research Centre |
| EPI_ISL_444459 | B.J. Medical College and Civil hospital | Gujarat Biotechnology Research Centre |
| EPI_ISL_444460 | B.J. Medical College and Civil hospital | Gujarat Biotechnology Research Centre |
| EPI_ISL_444461 | B.J. Medical College and Civil hospital | Gujarat Biotechnology Research Centre |
| EPI_ISL_444462 | B.J. Medical College and Civil hospital | Gujarat Biotechnology Research Centre |
| EPI_ISL_444463 | B.J. Medical College and Civil hospital | Gujarat Biotechnology Research Centre |
| EPI_ISL_444464 | B.J. Medical College and Civil hospital | Gujarat Biotechnology Research Centre |
| EPI_ISL_444465 | B.J. Medical College and Civil hospital | Gujarat Biotechnology Research Centre |
| EPI_ISL_444466 | B.J. Medical College and Civil hospital | Gujarat Biotechnology Research Centre |
| EPI_ISL_444467 | B.J. Medical College and Civil hospital | Gujarat Biotechnology Research Centre |
| EPI_ISL_444468 | B.J. Medical College and Civil hospital | Gujarat Biotechnology Research Centre |
| EPI_ISL_444469 | B.J. Medical College and Civil hospital | Gujarat Biotechnology Research Centre |
| EPI_ISL_444470 | B.J. Medical College and Civil hospital | Gujarat Biotechnology Research Centre |

| EPI_Accession_ID | Originating Laboratory | Submitting Laboratory |
| --- | --- | --- |
| EPI_ISL_444471 | B.J. Medical College and Civil hospital | Gujarat Biotechnology Research Centre |
| EPI_ISL_444472 | B.J. Medical College and Civil hospital | Gujarat Biotechnology Research Centre |
| EPI_ISL_444473 | B.J. Medical College and Civil hospital | Gujarat Biotechnology Research Centre |
| EPI_ISL_444474 | B.J. Medical College and Civil hospital | Gujarat Biotechnology Research Centre |
| EPI_ISL_444475 | B.J. Medical College and Civil hospital | Gujarat Biotechnology Research Centre |
| EPI_ISL_444476 | B.J. Medical College and Civil hospital | Gujarat Biotechnology Research Centre |
| EPI_ISL_444477 | B.J. Medical College and Civil hospital | Gujarat Biotechnology Research Centre |
| EPI_ISL_444478 | B.J. Medical College and Civil hospital | Gujarat Biotechnology Research Centre |
| EPI_ISL_444479 | B.J. Medical College and Civil hospital | Gujarat Biotechnology Research Centre |
| EPI_ISL_444480 | B.J. Medical College and Civil hospital | Gujarat Biotechnology Research Centre |
| EPI_ISL_444481 | B.J. Medical College and Civil hospital | Gujarat Biotechnology Research Centre |
| EPI_ISL_444482 | Gujarat Biotechnology Research Centre | Gujarat Biotechnology Research Centre |
| EPI_ISL_444483 | Gujarat Biotechnology Research Centre | Gujarat Biotechnology Research Centre |
| EPI_ISL_444484 | Gujarat Biotechnology Research Centre | Gujarat Biotechnology Research Centre |
| EPI_ISL_444485 | Gujarat Biotechnology Research Centre | Gujarat Biotechnology Research Centre |
| EPI_ISL_444486 | Gujarat Biotechnology Research Centre | Gujarat Biotechnology Research Centre |
| EPI_ISL_444493 | Departamento de Laboratorios de Salud Publica (DLSP, Division Epidemiologia, Ministerio de Salud Publica) | Facultad de Ciencias (SecciÃ³n GenÃ©tica Evolutiva, SecciÃ³n VirologÃ-a). |
| EPI_ISL_444494 | Laboratoire de microbiologie, Hopital de Verdun | Smith Laboratory, Centre de Recherche CHU Sainte-Justine |
| EPI_ISL_444495 | Laboratoire de microbiologie, Hopital de Verdun | Smith Laboratory, Centre de Recherche CHU Sainte-Justine |
| EPI_ISL_444496 | Laboratoire de microbiologie, Hopital de Verdun | Smith Laboratory, Centre de Recherche CHU Sainte-Justine |
| EPI_ISL_444497 | Laboratoire de microbiologie, Hopital de Verdun | Smith Laboratory, Centre de Recherche CHU Sainte-Justine |
| EPI_ISL_444498 | Laboratoire de microbiologie, Hopital de Verdun | Smith Laboratory, Centre de Recherche CHU Sainte-Justine |
| EPI_ISL_444499 | Laboratoire de microbiologie, Hopital de Verdun | Smith Laboratory, Centre de Recherche CHU Sainte-Justine |
| EPI_ISL_444500 | Laboratoire de microbiologie, Hopital de Verdun | Smith Laboratory, Centre de Recherche CHU Sainte-Justine |

| EPI_Accession_ID | Originating Laboratory | Submitting Laboratory |
|---|---|---|
| EPI_ISL_444501 | Laboratoire de microbiologie, Hopital de Verdun | Smith Laboratory, Centre de Recherche CHU Sainte-Justine |
| EPI_ISL_444502 | Laboratoire de microbiologie, Hopital de Verdun | Smith Laboratory, Centre de Recherche CHU Sainte-Justine |
| EPI_ISL_444503 | Laboratoire de microbiologie, Hopital de Verdun | Smith Laboratory, Centre de Recherche CHU Sainte-Justine |
| EPI_ISL_444504 | Laboratoire de microbiologie, Hopital de Verdun | Smith Laboratory, Centre de Recherche CHU Sainte-Justine |
| EPI_ISL_444505 | Laboratoire de microbiologie, Hopital de Verdun | Smith Laboratory, Centre de Recherche CHU Sainte-Justine |
| EPI_ISL_444506 | Laboratoire de microbiologie, Hopital de Verdun | Smith Laboratory, Centre de Recherche CHU Sainte-Justine |
| EPI_ISL_444507 | Laboratoire de microbiologie, Hopital de Verdun | Smith Laboratory, Centre de Recherche CHU Sainte-Justine |
| EPI_ISL_444508 | Laboratoire de microbiologie, Hopital de Verdun | Smith Laboratory, Centre de Recherche CHU Sainte-Justine |
| EPI_ISL_444509 | Laboratoire de microbiologie, Hopital de Verdun | Smith Laboratory, Centre de Recherche CHU Sainte-Justine |
| EPI_ISL_444510 | Laboratoire de microbiologie, Hopital de Verdun | Smith Laboratory, Centre de Recherche CHU Sainte-Justine |
| EPI_ISL_444511 | Laboratoire de microbiologie, Hopital de Verdun | Smith Laboratory, Centre de Recherche CHU Sainte-Justine |
| EPI_ISL_444512 | Laboratoire de microbiologie, Hopital de Verdun | Smith Laboratory, Centre de Recherche CHU Sainte-Justine |
| EPI_ISL_444513 | Laboratoire de microbiologie, Hopital de Verdun | Smith Laboratory, Centre de Recherche CHU Sainte-Justine |
| EPI_ISL_444514 | Laboratoire de microbiologie, Hopital de Verdun | Smith Laboratory, Centre de Recherche CHU Sainte-Justine |
| EPI_ISL_444515 | Laboratoire de microbiologie, Hopital de Verdun | Smith Laboratory, Centre de Recherche CHU Sainte-Justine |
| EPI_ISL_444516 | Laboratoire de microbiologie, Hopital de Verdun | Smith Laboratory, Centre de Recherche CHU Sainte-Justine |
| EPI_ISL_444518 | unknown | Molecular Infectious Disease |

| EPI_Accession_ID | Originating Laboratory | Submitting Laboratory |
|---|---|---|
| EPI_ISL_444519 | unknown | Molecular Infectious Disease |
| EPI_ISL_444520 | Northwestern Memorial Hospital | Ozer Lab |
| EPI_ISL_444521 | Northwestern Memorial Hospital | Ozer Lab |
| EPI_ISL_444522 | Northwestern Memorial Hospital | Ozer Lab |
| EPI_ISL_444523 | Northwestern Memorial Hospital | Ozer Lab |
| EPI_ISL_444524 | Northwestern Memorial Hospital | Ozer Lab |
| EPI_ISL_444525 | Northwestern Memorial Hospital | Ozer Lab |
| EPI_ISL_444526 | Northwestern Memorial Hospital | Ozer Lab |
| EPI_ISL_444527 | Northwestern Memorial Hospital | Ozer Lab |
| EPI_ISL_444528 | Northwestern Memorial Hospital | Ozer Lab |
| EPI_ISL_444529 | Northwestern Memorial Hospital | Ozer Lab |
| EPI_ISL_444530 | Northwestern Memorial Hospital | Ozer Lab |
| EPI_ISL_444531 | Northwestern Memorial Hospital | Ozer Lab |
| EPI_ISL_444532 | Northwestern Memorial Hospital | Ozer Lab |
| EPI_ISL_444533 | Northwestern Memorial Hospital | Ozer Lab |
| EPI_ISL_444534 | Northwestern Memorial Hospital | Ozer Lab |
| EPI_ISL_444535 | Northwestern Memorial Hospital | Ozer Lab |
| EPI_ISL_444536 | Northwestern Memorial Hospital | Ozer Lab |
| EPI_ISL_444537 | Northwestern Memorial Hospital | Ozer Lab |
| EPI_ISL_444538 | Northwestern Memorial Hospital | Ozer Lab |
| EPI_ISL_444539 | Northwestern Memorial Hospital | Ozer Lab |
| EPI_ISL_444540 | Northwestern Memorial Hospital | Ozer Lab |
| EPI_ISL_444541 | Northwestern Memorial Hospital | Ozer Lab |
| EPI_ISL_444542 | Northwestern Memorial Hospital | Ozer Lab |
| EPI_ISL_444543 | Northwestern Memorial Hospital | Ozer Lab |
| EPI_ISL_444544 | Northwestern Memorial Hospital | Ozer Lab |
| EPI_ISL_444545 | Northwestern Memorial Hospital | Ozer Lab |
| EPI_ISL_444546 | Northwestern Memorial Hospital | Ozer Lab |
| EPI_ISL_444547 | Northwestern Memorial Hospital | Ozer Lab |
| EPI_ISL_444548 | Northwestern Memorial Hospital | Ozer Lab |
| EPI_ISL_444549 | Northwestern Memorial Hospital | Ozer Lab |
| EPI_ISL_444550 | Northwestern Memorial Hospital | Ozer Lab |
| EPI_ISL_444551 | Northwestern Memorial Hospital | Ozer Lab |

| EPI_Accession_ID | Originating Laboratory | Submitting Laboratory |
|---|---|---|
| EPI_ISL_444552 | Northwestern Memorial Hospital | Ozer Lab |
| EPI_ISL_444553 | Northwestern Memorial Hospital | Ozer Lab |
| EPI_ISL_444554 | Northwestern Memorial Hospital | Ozer Lab |
| EPI_ISL_444555 | Northwestern Memorial Hospital | Ozer Lab |
| EPI_ISL_444556 | Northwestern Memorial Hospital | Ozer Lab |
| EPI_ISL_444557 | Northwestern Memorial Hospital | Ozer Lab |
| EPI_ISL_444558 | Northwestern Memorial Hospital | Ozer Lab |
| EPI_ISL_444559 | Northwestern Memorial Hospital | Ozer Lab |
| EPI_ISL_444560 | Northwestern Memorial Hospital | Ozer Lab |
| EPI_ISL_444561 | Northwestern Memorial Hospital | Ozer Lab |
| EPI_ISL_444562 | Northwestern Memorial Hospital | Ozer Lab |
| EPI_ISL_444563 | Northwestern Memorial Hospital | Ozer Lab |
| EPI_ISL_444564 | Northwestern Memorial Hospital | Ozer Lab |
| EPI_ISL_444565 | Northwestern Memorial Hospital | Ozer Lab |
| EPI_ISL_444566 | Northwestern Memorial Hospital | Ozer Lab |
| EPI_ISL_444567 | Northwestern Memorial Hospital | Ozer Lab |
| EPI_ISL_444568 | Northwestern Memorial Hospital | Ozer Lab |
| EPI_ISL_444569 | Northwestern Memorial Hospital | Ozer Lab |
| EPI_ISL_444570 | Northwestern Memorial Hospital | Ozer Lab |
| EPI_ISL_444571 | Northwestern Memorial Hospital | Ozer Lab |
| EPI_ISL_444572 | Northwestern Memorial Hospital | Ozer Lab |
| EPI_ISL_444573 | Northwestern Memorial Hospital | Ozer Lab |
| EPI_ISL_444574 | Northwestern Memorial Hospital | Ozer Lab |
| EPI_ISL_444575 | Northwestern Memorial Hospital | Ozer Lab |
| EPI_ISL_444576 | Northwestern Memorial Hospital | Ozer Lab |
| EPI_ISL_444577 | Northwestern Memorial Hospital | Ozer Lab |
| EPI_ISL_444578 | Northwestern Memorial Hospital | Ozer Lab |
| EPI_ISL_444579 | Northwestern Memorial Hospital | Ozer Lab |
| EPI_ISL_444580 | Northwestern Memorial Hospital | Ozer Lab |
| EPI_ISL_444581 | Northwestern Memorial Hospital | Ozer Lab |
| EPI_ISL_444582 | Northwestern Memorial Hospital | Ozer Lab |
| EPI_ISL_444583 | Northwestern Memorial Hospital | Ozer Lab |
| EPI_ISL_444584 | Northwestern Memorial Hospital | Ozer Lab |

| EPI_Accession_ID | Originating Laboratory | Submitting Laboratory |
|---|---|---|
| EPI_ISL_444585 | Northwestern Memorial Hospital | Ozer Lab |
| EPI_ISL_444586 | Northwestern Memorial Hospital | Ozer Lab |
| EPI_ISL_444587 | Northwestern Memorial Hospital | Ozer Lab |
| EPI_ISL_444588 | Northwestern Memorial Hospital | Ozer Lab |
| EPI_ISL_444589 | Northwestern Memorial Hospital | Ozer Lab |
| EPI_ISL_444590 | Northwestern Memorial Hospital | Ozer Lab |
| EPI_ISL_444591 | Northwestern Memorial Hospital | Ozer Lab |
| EPI_ISL_444592 | Northwestern Memorial Hospital | Ozer Lab |
| EPI_ISL_444593 | Northwestern Memorial Hospital | Ozer Lab |
| EPI_ISL_444594 | Northwestern Memorial Hospital | Ozer Lab |
| EPI_ISL_444595 | Northwestern Memorial Hospital | Ozer Lab |
| EPI_ISL_444596 | Northwestern Memorial Hospital | Ozer Lab |
| EPI_ISL_444597 | Northwestern Memorial Hospital | Ozer Lab |
| EPI_ISL_444598 | Northwestern Memorial Hospital | Ozer Lab |
| EPI_ISL_444599 | Northwestern Memorial Hospital | Ozer Lab |
| EPI_ISL_444600 | Northwestern Memorial Hospital | Ozer Lab |
| EPI_ISL_444601 | Northwestern Memorial Hospital | Ozer Lab |
| EPI_ISL_444602 | Northwestern Memorial Hospital | Ozer Lab |
| EPI_ISL_444603 | Northwestern Memorial Hospital | Ozer Lab |
| EPI_ISL_444604 | Northwestern Memorial Hospital | Ozer Lab |
| EPI_ISL_444605 | Northwestern Memorial Hospital | Ozer Lab |
| EPI_ISL_444606 | Northwestern Memorial Hospital | Ozer Lab |
| EPI_ISL_444607 | Northwestern Memorial Hospital | Ozer Lab |
| EPI_ISL_444608 | Northwestern Memorial Hospital | Ozer Lab |
| EPI_ISL_444609 | Northwestern Memorial Hospital | Ozer Lab |
| EPI_ISL_444611 | Pathology Queensland | Public Health Virology Laboratory |
| EPI_ISL_444612 | QML Pathology | Public Health Virology Laboratory |
| EPI_ISL_444613 | NYU Langone Health | Departments of Pathology and Medicine, New York University School of Medicine |
| EPI_ISL_444614 | NYU Langone Health | Departments of Pathology and Medicine, New York University School of Medicine |
| EPI_ISL_444615 | NYU Langone Health | Departments of Pathology and Medicine, New York University School of Medicine |

| EPI_Accession_ID | Originating Laboratory | Submitting Laboratory |
|---|---|---|
| EPI_ISL_444616 | NYU Langone Health | Departments of Pathology and Medicine, New York University School of Medicine |
| EPI_ISL_444617 | NYU Langone Health | Departments of Pathology and Medicine, New York University School of Medicine |
| EPI_ISL_444618 | NYU Langone Health | Departments of Pathology and Medicine, New York University School of Medicine |
| EPI_ISL_444619 | NYU Langone Health | Departments of Pathology and Medicine, New York University School of Medicine |
| EPI_ISL_444620 | NYU Langone Health | Departments of Pathology and Medicine, New York University School of Medicine |
| EPI_ISL_444621 | NYU Langone Health | Departments of Pathology and Medicine, New York University School of Medicine |
| EPI_ISL_444622 | NYU Langone Health | Departments of Pathology and Medicine, New York University School of Medicine |
| EPI_ISL_444623 | NYU Langone Health | Departments of Pathology and Medicine, New York University School of Medicine |
| EPI_ISL_444624 | NYU Langone Health | Departments of Pathology and Medicine, New York University School of Medicine |
| EPI_ISL_444625 | NYU Langone Health | Departments of Pathology and Medicine, New York University School of Medicine |
| EPI_ISL_444626 | NYU Langone Health | Departments of Pathology and Medicine, New York University School of Medicine |
| EPI_ISL_444627 | NYU Langone Health | Departments of Pathology and Medicine, New York University School of Medicine |
| EPI_ISL_444628 | NYU Langone Health | Departments of Pathology and Medicine, New York University School of Medicine |
| EPI_ISL_444629 | NYU Langone Health | Departments of Pathology and Medicine, New York University School of Medicine |
| EPI_ISL_444630 | NYU Langone Health | Departments of Pathology and Medicine, New York University School of Medicine |
| EPI_ISL_444631 | NYU Langone Health | Departments of Pathology and Medicine, New York University School of Medicine |

| EPI_Accession_ID | Originating Laboratory | Submitting Laboratory |
|---|---|---|
| EPI_ISL_444632 | NYU Langone Health | Departments of Pathology and Medicine, New York University School of Medicine |
| EPI_ISL_444633 | NYU Langone Health | Departments of Pathology and Medicine, New York University School of Medicine |
| EPI_ISL_444634 | NYU Langone Health | Departments of Pathology and Medicine, New York University School of Medicine |
| EPI_ISL_444635 | NYU Langone Health | Departments of Pathology and Medicine, New York University School of Medicine |
| EPI_ISL_444636 | NYU Langone Health | Departments of Pathology and Medicine, New York University School of Medicine |
| EPI_ISL_444637 | NYU Langone Health | Departments of Pathology and Medicine, New York University School of Medicine |
| EPI_ISL_444638 | NYU Langone Health | Departments of Pathology and Medicine, New York University School of Medicine |
| EPI_ISL_444639 | NYU Langone Health | Departments of Pathology and Medicine, New York University School of Medicine |
| EPI_ISL_444640 | NYU Langone Health | Departments of Pathology and Medicine, New York University School of Medicine |
| EPI_ISL_444641 | NYU Langone Health | Departments of Pathology and Medicine, New York University School of Medicine |
| EPI_ISL_444642 | NYU Langone Health | Departments of Pathology and Medicine, New York University School of Medicine |
| EPI_ISL_444643 | NYU Langone Health | Departments of Pathology and Medicine, New York University School of Medicine |
| EPI_ISL_444644 | NYU Langone Health | Departments of Pathology and Medicine, New York University School of Medicine |
| EPI_ISL_444645 | NYU Langone Health | Departments of Pathology and Medicine, New York University School of Medicine |
| EPI_ISL_444646 | NYU Langone Health | Departments of Pathology and Medicine, New York University School of Medicine |
| EPI_ISL_444647 | NYU Langone Health | Departments of Pathology and Medicine, New York University School of Medicine |

| EPI_Accession_ID | Originating Laboratory | Submitting Laboratory |
|---|---|---|
| EPI_ISL_444648 | NYU Langone Health | Departments of Pathology and Medicine, New York University School of Medicine |
| EPI_ISL_444650 | NYU Langone Health | Departments of Pathology and Medicine, New York University School of Medicine |
| EPI_ISL_444651 | NYU Langone Health | Departments of Pathology and Medicine, New York University School of Medicine |
| EPI_ISL_444652 | NYU Langone Health | Departments of Pathology and Medicine, New York University School of Medicine |
| EPI_ISL_444653 | NYU Langone Health | Departments of Pathology and Medicine, New York University School of Medicine |
| EPI_ISL_444654 | NYU Langone Health | Departments of Pathology and Medicine, New York University School of Medicine |
| EPI_ISL_444655 | NYU Langone Health | Departments of Pathology and Medicine, New York University School of Medicine |
| EPI_ISL_444656 | NYU Langone Health | Departments of Pathology and Medicine, New York University School of Medicine |
| EPI_ISL_444657 | NYU Langone Health | Departments of Pathology and Medicine, New York University School of Medicine |
| EPI_ISL_444658 | NYU Langone Health | Departments of Pathology and Medicine, New York University School of Medicine |
| EPI_ISL_444659 | NYU Langone Health | Departments of Pathology and Medicine, New York University School of Medicine |
| EPI_ISL_444660 | NYU Langone Health | Departments of Pathology and Medicine, New York University School of Medicine |
| EPI_ISL_444661 | NYU Langone Health | Departments of Pathology and Medicine, New York University School of Medicine |
| EPI_ISL_444662 | NYU Langone Health | Departments of Pathology and Medicine, New York University School of Medicine |
| EPI_ISL_444663 | NYU Langone Health | Departments of Pathology and Medicine, New York University School of Medicine |
| EPI_ISL_444664 | NYU Langone Health | Departments of Pathology and Medicine, New York University School of Medicine |

| EPI_Accession_ID | Originating Laboratory | Submitting Laboratory |
|---|---|---|
| EPI_ISL_444665 | NYU Langone Health | Departments of Pathology and Medicine, New York University School of Medicine |
| EPI_ISL_444666 | NYU Langone Health | Departments of Pathology and Medicine, New York University School of Medicine |
| EPI_ISL_444667 | NYU Langone Health | Departments of Pathology and Medicine, New York University School of Medicine |
| EPI_ISL_444668 | NYU Langone Health | Departments of Pathology and Medicine, New York University School of Medicine |
| EPI_ISL_444669 | NYU Langone Health | Departments of Pathology and Medicine, New York University School of Medicine |
| EPI_ISL_444670 | NYU Langone Health | Departments of Pathology and Medicine, New York University School of Medicine |
| EPI_ISL_444671 | NYU Langone Health | Departments of Pathology and Medicine, New York University School of Medicine |
| EPI_ISL_444672 | NYU Langone Health | Departments of Pathology and Medicine, New York University School of Medicine |
| EPI_ISL_444673 | NYU Langone Health | Departments of Pathology and Medicine, New York University School of Medicine |
| EPI_ISL_444674 | NYU Langone Health | Departments of Pathology and Medicine, New York University School of Medicine |
| EPI_ISL_444675 | NYU Langone Health | Departments of Pathology and Medicine, New York University School of Medicine |
| EPI_ISL_444676 | NYU Langone Health | Departments of Pathology and Medicine, New York University School of Medicine |
| EPI_ISL_444677 | NYU Langone Health | Departments of Pathology and Medicine, New York University School of Medicine |
| EPI_ISL_444678 | NYU Langone Health | Departments of Pathology and Medicine, New York University School of Medicine |
| EPI_ISL_444679 | NYU Langone Health | Departments of Pathology and Medicine, New York University School of Medicine |
| EPI_ISL_444680 | NYU Langone Health | Departments of Pathology and Medicine, New York University School of Medicine |

| EPI_Accession_ID | Originating Laboratory | Submitting Laboratory |
|---|---|---|
| EPI_ISL_444681 | NYU Langone Health | Departments of Pathology and Medicine, New York University School of Medicine |
| EPI_ISL_444682 | NYU Langone Health | Departments of Pathology and Medicine, New York University School of Medicine |
| EPI_ISL_444683 | NYU Langone Health | Departments of Pathology and Medicine, New York University School of Medicine |
| EPI_ISL_444684 | NYU Langone Health | Departments of Pathology and Medicine, New York University School of Medicine |
| EPI_ISL_444685 | NYU Langone Health | Departments of Pathology and Medicine, New York University School of Medicine |
| EPI_ISL_444686 | NYU Langone Health | Departments of Pathology and Medicine, New York University School of Medicine |
| EPI_ISL_444687 | NYU Langone Health | Departments of Pathology and Medicine, New York University School of Medicine |
| EPI_ISL_444688 | NYU Langone Health | Departments of Pathology and Medicine, New York University School of Medicine |
| EPI_ISL_444689 | NYU Langone Health | Departments of Pathology and Medicine, New York University School of Medicine |
| EPI_ISL_444690 | NYU Langone Health | Departments of Pathology and Medicine, New York University School of Medicine |
| EPI_ISL_444691 | NYU Langone Health | Departments of Pathology and Medicine, New York University School of Medicine |
| EPI_ISL_444692 | NYU Langone Health | Departments of Pathology and Medicine, New York University School of Medicine |
| EPI_ISL_444693 | NYU Langone Health | Departments of Pathology and Medicine, New York University School of Medicine |
| EPI_ISL_444694 | NYU Langone Health | Departments of Pathology and Medicine, New York University School of Medicine |
| EPI_ISL_444695 | NYU Langone Health | Departments of Pathology and Medicine, New York University School of Medicine |
| EPI_ISL_444696 | NYU Langone Health | Departments of Pathology and Medicine, New York University School of Medicine |

| EPI_Accession_ID | Originating Laboratory | Submitting Laboratory |
|---|---|---|
| EPI_ISL_444697 | NYU Langone Health | Departments of Pathology and Medicine, New York University School of Medicine |
| EPI_ISL_444698 | NYU Langone Health | Departments of Pathology and Medicine, New York University School of Medicine |
| EPI_ISL_444699 | NYU Langone Health | Departments of Pathology and Medicine, New York University School of Medicine |
| EPI_ISL_444700 | NYU Langone Health | Departments of Pathology and Medicine, New York University School of Medicine |
| EPI_ISL_444701 | NYU Langone Health | Departments of Pathology and Medicine, New York University School of Medicine |
| EPI_ISL_444702 | NYU Langone Health | Departments of Pathology and Medicine, New York University School of Medicine |
| EPI_ISL_444703 | NYU Langone Health | Departments of Pathology and Medicine, New York University School of Medicine |
| EPI_ISL_444704 | NYU Langone Health | Departments of Pathology and Medicine, New York University School of Medicine |
| EPI_ISL_444705 | NYU Langone Health | Departments of Pathology and Medicine, New York University School of Medicine |
| EPI_ISL_444706 | NYU Langone Health | Departments of Pathology and Medicine, New York University School of Medicine |
| EPI_ISL_444707 | NYU Langone Health | Departments of Pathology and Medicine, New York University School of Medicine |
| EPI_ISL_444708 | NYU Langone Health | Departments of Pathology and Medicine, New York University School of Medicine |
| EPI_ISL_444709 | NYU Langone Health | Departments of Pathology and Medicine, New York University School of Medicine |
| EPI_ISL_444710 | NYU Langone Health | Departments of Pathology and Medicine, New York University School of Medicine |
| EPI_ISL_444711 | NYU Langone Health | Departments of Pathology and Medicine, New York University School of Medicine |
| EPI_ISL_444712 | NYU Langone Health | Departments of Pathology and Medicine, New York University School of Medicine |

| EPI_Accession_ID | Originating Laboratory | Submitting Laboratory |
|---|---|---|
| EPI_ISL_444713 | NYU Langone Health | Departments of Pathology and Medicine, New York University School of Medicine |
| EPI_ISL_444714 | NYU Langone Health | Departments of Pathology and Medicine, New York University School of Medicine |
| EPI_ISL_444715 | NYU Langone Health | Departments of Pathology and Medicine, New York University School of Medicine |
| EPI_ISL_444716 | NYU Langone Health | Departments of Pathology and Medicine, New York University School of Medicine |
| EPI_ISL_444717 | NYU Langone Health | Departments of Pathology and Medicine, New York University School of Medicine |
| EPI_ISL_444718 | NYU Langone Health | Departments of Pathology and Medicine, New York University School of Medicine |
| EPI_ISL_444719 | NYU Langone Health | Departments of Pathology and Medicine, New York University School of Medicine |
| EPI_ISL_444720 | NYU Langone Health | Departments of Pathology and Medicine, New York University School of Medicine |
| EPI_ISL_444721 | NYU Langone Health | Departments of Pathology and Medicine, New York University School of Medicine |
| EPI_ISL_444722 | NYU Langone Health | Departments of Pathology and Medicine, New York University School of Medicine |
| EPI_ISL_444723 | NYU Langone Health | Departments of Pathology and Medicine, New York University School of Medicine |
| EPI_ISL_444724 | NYU Langone Health | Departments of Pathology and Medicine, New York University School of Medicine |
| EPI_ISL_444725 | NYU Langone Health | Departments of Pathology and Medicine, New York University School of Medicine |
| EPI_ISL_444726 | NYU Langone Health | Departments of Pathology and Medicine, New York University School of Medicine |
| EPI_ISL_444727 | NYU Langone Health | Departments of Pathology and Medicine, New York University School of Medicine |
| EPI_ISL_444728 | NYU Langone Health | Departments of Pathology and Medicine, New York University School of Medicine |

| EPI_Accession_ID | Originating Laboratory | Submitting Laboratory |
|---|---|---|
| EPI_ISL_444729 | NYU Langone Health | Departments of Pathology and Medicine, New York University School of Medicine |
| EPI_ISL_444730 | NYU Langone Health | Departments of Pathology and Medicine, New York University School of Medicine |
| EPI_ISL_444731 | NYU Langone Health | Departments of Pathology and Medicine, New York University School of Medicine |
| EPI_ISL_444732 | NYU Langone Health | Departments of Pathology and Medicine, New York University School of Medicine |
| EPI_ISL_444733 | NYU Langone Health | Departments of Pathology and Medicine, New York University School of Medicine |
| EPI_ISL_444734 | NYU Langone Health | Departments of Pathology and Medicine, New York University School of Medicine |
| EPI_ISL_444735 | NYU Langone Health | Departments of Pathology and Medicine, New York University School of Medicine |
| EPI_ISL_444736 | NYU Langone Health | Departments of Pathology and Medicine, New York University School of Medicine |
| EPI_ISL_444737 | NYU Langone Health | Departments of Pathology and Medicine, New York University School of Medicine |
| EPI_ISL_444738 | NYU Langone Health | Departments of Pathology and Medicine, New York University School of Medicine |
| EPI_ISL_444739 | NYU Langone Health | Departments of Pathology and Medicine, New York University School of Medicine |
| EPI_ISL_444740 | NYU Langone Health | Departments of Pathology and Medicine, New York University School of Medicine |
| EPI_ISL_444741 | NYU Langone Health | Departments of Pathology and Medicine, New York University School of Medicine |
| EPI_ISL_444742 | NYU Langone Health | Departments of Pathology and Medicine, New York University School of Medicine |
| EPI_ISL_444743 | NYU Langone Health | Departments of Pathology and Medicine, New York University School of Medicine |
| EPI_ISL_444744 | NYU Langone Health | Departments of Pathology and Medicine, New York University School of Medicine |

| EPI_Accession_ID | Originating Laboratory | Submitting Laboratory |
|---|---|---|
| EPI_ISL_444745 | NYU Langone Health | Departments of Pathology and Medicine, New York University School of Medicine |
| EPI_ISL_444746 | NYU Langone Health | Departments of Pathology and Medicine, New York University School of Medicine |
| EPI_ISL_444747 | NYU Langone Health | Departments of Pathology and Medicine, New York University School of Medicine |
| EPI_ISL_444748 | NYU Langone Health | Departments of Pathology and Medicine, New York University School of Medicine |
| EPI_ISL_444749 | NYU Langone Health | Departments of Pathology and Medicine, New York University School of Medicine |
| EPI_ISL_444750 | NYU Langone Health | Departments of Pathology and Medicine, New York University School of Medicine |
| EPI_ISL_444751 | NYU Langone Health | Departments of Pathology and Medicine, New York University School of Medicine |
| EPI_ISL_444752 | NYU Langone Health | Departments of Pathology and Medicine, New York University School of Medicine |
| EPI_ISL_444753 | NYU Langone Health | Departments of Pathology and Medicine, New York University School of Medicine |
| EPI_ISL_444754 | NYU Langone Health | Departments of Pathology and Medicine, New York University School of Medicine |
| EPI_ISL_444755 | NYU Langone Health | Departments of Pathology and Medicine, New York University School of Medicine |
| EPI_ISL_444756 | NYU Langone Health | Departments of Pathology and Medicine, New York University School of Medicine |
| EPI_ISL_444757 | NYU Langone Health | Departments of Pathology and Medicine, New York University School of Medicine |
| EPI_ISL_444758 | NYU Langone Health | Departments of Pathology and Medicine, New York University School of Medicine |
| EPI_ISL_444759 | NYU Langone Health | Departments of Pathology and Medicine, New York University School of Medicine |
| EPI_ISL_444760 | NYU Langone Health | Departments of Pathology and Medicine, New York University School of Medicine |

| EPI_Accession_ID | Originating Laboratory | Submitting Laboratory |
|---|---|---|
| EPI_ISL_444761 | NYU Langone Health | Departments of Pathology and Medicine, New York University School of Medicine |
| EPI_ISL_444762 | NYU Langone Health | Departments of Pathology and Medicine, New York University School of Medicine |
| EPI_ISL_444763 | NYU Langone Health | Departments of Pathology and Medicine, New York University School of Medicine |
| EPI_ISL_444764 | NYU Langone Health | Departments of Pathology and Medicine, New York University School of Medicine |
| EPI_ISL_444765 | NYU Langone Health | Departments of Pathology and Medicine, New York University School of Medicine |
| EPI_ISL_444766 | NYU Langone Health | Departments of Pathology and Medicine, New York University School of Medicine |
| EPI_ISL_444767 | NYU Langone Health | Departments of Pathology and Medicine, New York University School of Medicine |
| EPI_ISL_444768 | NYU Langone Health | Departments of Pathology and Medicine, New York University School of Medicine |
| EPI_ISL_444769 | NYU Langone Health | Departments of Pathology and Medicine, New York University School of Medicine |
| EPI_ISL_444770 | NYU Langone Health | Departments of Pathology and Medicine, New York University School of Medicine |
| EPI_ISL_444771 | NYU Langone Health | Departments of Pathology and Medicine, New York University School of Medicine |
| EPI_ISL_444772 | NYU Langone Health | Departments of Pathology and Medicine, New York University School of Medicine |
| EPI_ISL_444773 | NYU Langone Health | Departments of Pathology and Medicine, New York University School of Medicine |
| EPI_ISL_444774 | NYU Langone Health | Departments of Pathology and Medicine, New York University School of Medicine |
| EPI_ISL_444775 | NYU Langone Health | Departments of Pathology and Medicine, New York University School of Medicine |
| EPI_ISL_444776 | NYU Langone Health | Departments of Pathology and Medicine, New York University School of Medicine |

| EPI_Accession_ID | Originating Laboratory | Submitting Laboratory |
|---|---|---|
| EPI_ISL_444777 | NYU Langone Health | Departments of Pathology and Medicine, New York University School of Medicine |
| EPI_ISL_444778 | NYU Langone Health | Departments of Pathology and Medicine, New York University School of Medicine |
| EPI_ISL_444779 | NYU Langone Health | Departments of Pathology and Medicine, New York University School of Medicine |
| EPI_ISL_444780 | NYU Langone Health | Departments of Pathology and Medicine, New York University School of Medicine |
| EPI_ISL_444781 | NYU Langone Health | Departments of Pathology and Medicine, New York University School of Medicine |
| EPI_ISL_444782 | NYU Langone Health | Departments of Pathology and Medicine, New York University School of Medicine |
| EPI_ISL_444783 | NYU Langone Health | Departments of Pathology and Medicine, New York University School of Medicine |
| EPI_ISL_444784 | NYU Langone Health | Departments of Pathology and Medicine, New York University School of Medicine |
| EPI_ISL_444785 | NYU Langone Health | Departments of Pathology and Medicine, New York University School of Medicine |
| EPI_ISL_444786 | NYU Langone Health | Departments of Pathology and Medicine, New York University School of Medicine |
| EPI_ISL_444787 | NYU Langone Health | Departments of Pathology and Medicine, New York University School of Medicine |
| EPI_ISL_444788 | NYU Langone Health | Departments of Pathology and Medicine, New York University School of Medicine |
| EPI_ISL_444789 | NYU Langone Health | Departments of Pathology and Medicine, New York University School of Medicine |
| EPI_ISL_444790 | NYU Langone Health | Departments of Pathology and Medicine, New York University School of Medicine |
| EPI_ISL_444791 | NYU Langone Health | Departments of Pathology and Medicine, New York University School of Medicine |
| EPI_ISL_444792 | NYU Langone Health | Departments of Pathology and Medicine, New York University School of Medicine |
| EPI_ISL_444793 | Pathology Queensland | Public Health Virology Laboratory |

| EPI_Accession_ID | Originating Laboratory | Submitting Laboratory |
|---|---|---|
| EPI_ISL_444794 | Cairns Hospital | Public Health Virology Laboratory |
| EPI_ISL_444817 | Department of Virus and Microbiological Special Diagnostics, Statens Serum Institut, Copenhagen, Denmark, Artillerivej 5, 2300 Copenahgen S | Albertsen lab, Department of Chemistry and Bioscience, Aalborg University, Denmark |
| EPI_ISL_444818 | Department of Virus and Microbiological Special Diagnostics, Statens Serum Institut, Copenhagen, Denmark, Artillerivej 5, 2300 Copenahgen S | Albertsen lab, Department of Chemistry and Bioscience, Aalborg University, Denmark |
| EPI_ISL_444819 | Department of Virus and Microbiological Special Diagnostics, Statens Serum Institut, Copenhagen, Denmark, Artillerivej 5, 2300 Copenahgen S | Albertsen lab, Department of Chemistry and Bioscience, Aalborg University, Denmark |
| EPI_ISL_444820 | Department of Virus and Microbiological Special Diagnostics, Statens Serum Institut, Copenhagen, Denmark, Artillerivej 5, 2300 Copenahgen S | Albertsen lab, Department of Chemistry and Bioscience, Aalborg University, Denmark |
| EPI_ISL_444821 | Department of Virus and Microbiological Special Diagnostics, Statens Serum Institut, Copenhagen, Denmark, Artillerivej 5, 2300 Copenahgen S | Albertsen lab, Department of Chemistry and Bioscience, Aalborg University, Denmark |
| EPI_ISL_444822 | Department of Virus and Microbiological Special Diagnostics, Statens Serum Institut, Copenhagen, Denmark, Artillerivej 5, 2300 Copenahgen S | Albertsen lab, Department of Chemistry and Bioscience, Aalborg University, Denmark |
| EPI_ISL_444823 | Department of Virus and Microbiological Special Diagnostics, Statens Serum Institut, Copenhagen, Denmark, Artillerivej 5, 2300 Copenahgen S | Albertsen lab, Department of Chemistry and Bioscience, Aalborg University, Denmark |
| EPI_ISL_444824 | Department of Virus and Microbiological Special Diagnostics, Statens Serum Institut, Copenhagen, Denmark, Artillerivej 5, 2300 Copenahgen S | Albertsen lab, Department of Chemistry and Bioscience, Aalborg University, Denmark |
| EPI_ISL_444825 | Department of Virus and Microbiological Special Diagnostics, Statens Serum Institut, Copenhagen, Denmark, Artillerivej 5, 2300 Copenahgen S | Albertsen lab, Department of Chemistry and Bioscience, Aalborg University, Denmark |
| EPI_ISL_444826 | Department of Virus and Microbiological Special Diagnostics, Statens Serum Institut, Copenhagen, Denmark, Artillerivej 5, 2300 Copenahgen S | Albertsen lab, Department of Chemistry and Bioscience, Aalborg University, Denmark |

| EPI_Accession_ID | Originating Laboratory | Submitting Laboratory |
|---|---|---|
| EPI_ISL_444827 | Department of Virus and Microbiological Special Diagnostics, Statens Serum Institut, Copenhagen, Denmark, Artillerivej 5, 2300 Copenahgen S | Albertsen lab, Department of Chemistry and Bioscience, Aalborg University, Denmark |
| EPI_ISL_444828 | Department of Virus and Microbiological Special Diagnostics, Statens Serum Institut, Copenhagen, Denmark, Artillerivej 5, 2300 Copenahgen S | Albertsen lab, Department of Chemistry and Bioscience, Aalborg University, Denmark |
| EPI_ISL_444829 | Department of Virus and Microbiological Special Diagnostics, Statens Serum Institut, Copenhagen, Denmark, Artillerivej 5, 2300 Copenahgen S | Albertsen lab, Department of Chemistry and Bioscience, Aalborg University, Denmark |
| EPI_ISL_444830 | Department of Virus and Microbiological Special Diagnostics, Statens Serum Institut, Copenhagen, Denmark, Artillerivej 5, 2300 Copenahgen S | Albertsen lab, Department of Chemistry and Bioscience, Aalborg University, Denmark |
| EPI_ISL_444831 | Department of Virus and Microbiological Special Diagnostics, Statens Serum Institut, Copenhagen, Denmark, Artillerivej 5, 2300 Copenahgen S | Albertsen lab, Department of Chemistry and Bioscience, Aalborg University, Denmark |
| EPI_ISL_444832 | Department of Virus and Microbiological Special Diagnostics, Statens Serum Institut, Copenhagen, Denmark, Artillerivej 5, 2300 Copenahgen S | Albertsen lab, Department of Chemistry and Bioscience, Aalborg University, Denmark |
| EPI_ISL_444833 | Department of Virus and Microbiological Special Diagnostics, Statens Serum Institut, Copenhagen, Denmark, Artillerivej 5, 2300 Copenahgen S | Albertsen lab, Department of Chemistry and Bioscience, Aalborg University, Denmark |
| EPI_ISL_444834 | Department of Virus and Microbiological Special Diagnostics, Statens Serum Institut, Copenhagen, Denmark, Artillerivej 5, 2300 Copenahgen S | Albertsen lab, Department of Chemistry and Bioscience, Aalborg University, Denmark |
| EPI_ISL_444835 | Department of Virus and Microbiological Special Diagnostics, Statens Serum Institut, Copenhagen, Denmark, Artillerivej 5, 2300 Copenahgen S | Albertsen lab, Department of Chemistry and Bioscience, Aalborg University, Denmark |
| EPI_ISL_444836 | Department of Virus and Microbiological Special Diagnostics, Statens Serum Institut, Copenhagen, Denmark, Artillerivej 5, 2300 Copenahgen S | Albertsen lab, Department of Chemistry and Bioscience, Aalborg University, Denmark |
| EPI_ISL_444837 | Department of Virus and Microbiological Special Diagnostics, Statens Serum Institut, Copenhagen, Denmark, Artillerivej 5, 2300 Copenahgen S | Albertsen lab, Department of Chemistry and Bioscience, Aalborg University, Denmark |

| EPI_Accession_ID | Originating Laboratory | Submitting Laboratory |
|---|---|---|
| EPI_ISL_444838 | Department of Virus and Microbiological Special Diagnostics, Statens Serum Institut, Copenhagen, Denmark, Artillerivej 5, 2300 Copenahgen S | Albertsen lab, Department of Chemistry and Bioscience, Aalborg University, Denmark |
| EPI_ISL_444839 | Department of Virus and Microbiological Special Diagnostics, Statens Serum Institut, Copenhagen, Denmark, Artillerivej 5, 2300 Copenahgen S | Albertsen lab, Department of Chemistry and Bioscience, Aalborg University, Denmark |
| EPI_ISL_444840 | Department of Virus and Microbiological Special Diagnostics, Statens Serum Institut, Copenhagen, Denmark, Artillerivej 5, 2300 Copenahgen S | Albertsen lab, Department of Chemistry and Bioscience, Aalborg University, Denmark |
| EPI_ISL_444841 | Department of Virus and Microbiological Special Diagnostics, Statens Serum Institut, Copenhagen, Denmark, Artillerivej 5, 2300 Copenahgen S | Albertsen lab, Department of Chemistry and Bioscience, Aalborg University, Denmark |
| EPI_ISL_444842 | Department of Virus and Microbiological Special Diagnostics, Statens Serum Institut, Copenhagen, Denmark, Artillerivej 5, 2300 Copenahgen S | Albertsen lab, Department of Chemistry and Bioscience, Aalborg University, Denmark |
| EPI_ISL_444843 | Department of Virus and Microbiological Special Diagnostics, Statens Serum Institut, Copenhagen, Denmark, Artillerivej 5, 2300 Copenahgen S | Albertsen lab, Department of Chemistry and Bioscience, Aalborg University, Denmark |
| EPI_ISL_444844 | Department of Virus and Microbiological Special Diagnostics, Statens Serum Institut, Copenhagen, Denmark, Artillerivej 5, 2300 Copenahgen S | Albertsen lab, Department of Chemistry and Bioscience, Aalborg University, Denmark |
| EPI_ISL_444845 | Department of Virus and Microbiological Special Diagnostics, Statens Serum Institut, Copenhagen, Denmark, Artillerivej 5, 2300 Copenahgen S | Albertsen lab, Department of Chemistry and Bioscience, Aalborg University, Denmark |
| EPI_ISL_444846 | Department of Virus and Microbiological Special Diagnostics, Statens Serum Institut, Copenhagen, Denmark, Artillerivej 5, 2300 Copenahgen S | Albertsen lab, Department of Chemistry and Bioscience, Aalborg University, Denmark |
| EPI_ISL_444847 | Department of Virus and Microbiological Special Diagnostics, Statens Serum Institut, Copenhagen, Denmark, Artillerivej 5, 2300 Copenahgen S | Albertsen lab, Department of Chemistry and Bioscience, Aalborg University, Denmark |
| EPI_ISL_444848 | Department of Virus and Microbiological Special Diagnostics, Statens Serum Institut, Copenhagen, Denmark, Artillerivej 5, 2300 Copenahgen S | Albertsen lab, Department of Chemistry and Bioscience, Aalborg University, Denmark |

| EPI_Accession_ID | Originating Laboratory | Submitting Laboratory |
|---|---|---|
| EPI_ISL_444849 | Department of Virus and Microbiological Special Diagnostics, Statens Serum Institut, Copenhagen, Denmark, Artillerivej 5, 2300 Copenahgen S | Albertsen lab, Department of Chemistry and Bioscience, Aalborg University, Denmark |
| EPI_ISL_444850 | Department of Virus and Microbiological Special Diagnostics, Statens Serum Institut, Copenhagen, Denmark, Artillerivej 5, 2300 Copenahgen S | Albertsen lab, Department of Chemistry and Bioscience, Aalborg University, Denmark |
| EPI_ISL_444851 | Department of Virus and Microbiological Special Diagnostics, Statens Serum Institut, Copenhagen, Denmark, Artillerivej 5, 2300 Copenahgen S | Albertsen lab, Department of Chemistry and Bioscience, Aalborg University, Denmark |
| EPI_ISL_444852 | Department of Virus and Microbiological Special Diagnostics, Statens Serum Institut, Copenhagen, Denmark, Artillerivej 5, 2300 Copenahgen S | Albertsen lab, Department of Chemistry and Bioscience, Aalborg University, Denmark |
| EPI_ISL_444853 | Department of Virus and Microbiological Special Diagnostics, Statens Serum Institut, Copenhagen, Denmark, Artillerivej 5, 2300 Copenahgen S | Albertsen lab, Department of Chemistry and Bioscience, Aalborg University, Denmark |
| EPI_ISL_444854 | Department of Virus and Microbiological Special Diagnostics, Statens Serum Institut, Copenhagen, Denmark, Artillerivej 5, 2300 Copenahgen S | Albertsen lab, Department of Chemistry and Bioscience, Aalborg University, Denmark |
| EPI_ISL_444855 | Department of Virus and Microbiological Special Diagnostics, Statens Serum Institut, Copenhagen, Denmark, Artillerivej 5, 2300 Copenahgen S | Albertsen lab, Department of Chemistry and Bioscience, Aalborg University, Denmark |
| EPI_ISL_444856 | Department of Virus and Microbiological Special Diagnostics, Statens Serum Institut, Copenhagen, Denmark, Artillerivej 5, 2300 Copenahgen S | Albertsen lab, Department of Chemistry and Bioscience, Aalborg University, Denmark |
| EPI_ISL_444857 | Department of Virus and Microbiological Special Diagnostics, Statens Serum Institut, Copenhagen, Denmark, Artillerivej 5, 2300 Copenahgen S | Albertsen lab, Department of Chemistry and Bioscience, Aalborg University, Denmark |
| EPI_ISL_444858 | Department of Virus and Microbiological Special Diagnostics, Statens Serum Institut, Copenhagen, Denmark, Artillerivej 5, 2300 Copenahgen S | Albertsen lab, Department of Chemistry and Bioscience, Aalborg University, Denmark |
| EPI_ISL_444859 | Department of Virus and Microbiological Special Diagnostics, Statens Serum Institut, Copenhagen, Denmark, Artillerivej 5, 2300 Copenahgen S | Albertsen lab, Department of Chemistry and Bioscience, Aalborg University, Denmark |

| EPI_Accession_ID | Originating Laboratory | Submitting Laboratory |
|---|---|---|
| EPI_ISL_444860 | Department of Virus and Microbiological Special Diagnostics, Statens Serum Institut, Copenhagen, Denmark, Artillerivej 5, 2300 Copenahgen S | Albertsen lab, Department of Chemistry and Bioscience, Aalborg University, Denmark |
| EPI_ISL_444861 | Department of Virus and Microbiological Special Diagnostics, Statens Serum Institut, Copenhagen, Denmark, Artillerivej 5, 2300 Copenahgen S | Albertsen lab, Department of Chemistry and Bioscience, Aalborg University, Denmark |
| EPI_ISL_444862 | Department of Virus and Microbiological Special Diagnostics, Statens Serum Institut, Copenhagen, Denmark, Artillerivej 5, 2300 Copenahgen S | Albertsen lab, Department of Chemistry and Bioscience, Aalborg University, Denmark |
| EPI_ISL_444863 | Department of Virus and Microbiological Special Diagnostics, Statens Serum Institut, Copenhagen, Denmark, Artillerivej 5, 2300 Copenahgen S | Albertsen lab, Department of Chemistry and Bioscience, Aalborg University, Denmark |
| EPI_ISL_444864 | Department of Virus and Microbiological Special Diagnostics, Statens Serum Institut, Copenhagen, Denmark, Artillerivej 5, 2300 Copenahgen S | Albertsen lab, Department of Chemistry and Bioscience, Aalborg University, Denmark |
| EPI_ISL_444865 | Department of Virus and Microbiological Special Diagnostics, Statens Serum Institut, Copenhagen, Denmark, Artillerivej 5, 2300 Copenahgen S | Albertsen lab, Department of Chemistry and Bioscience, Aalborg University, Denmark |
| EPI_ISL_444866 | Department of Virus and Microbiological Special Diagnostics, Statens Serum Institut, Copenhagen, Denmark, Artillerivej 5, 2300 Copenahgen S | Albertsen lab, Department of Chemistry and Bioscience, Aalborg University, Denmark |
| EPI_ISL_444867 | Department of Virus and Microbiological Special Diagnostics, Statens Serum Institut, Copenhagen, Denmark, Artillerivej 5, 2300 Copenahgen S | Albertsen lab, Department of Chemistry and Bioscience, Aalborg University, Denmark |
| EPI_ISL_444868 | Department of Virus and Microbiological Special Diagnostics, Statens Serum Institut, Copenhagen, Denmark, Artillerivej 5, 2300 Copenahgen S | Albertsen lab, Department of Chemistry and Bioscience, Aalborg University, Denmark |
| EPI_ISL_444869 | Department of Virus and Microbiological Special Diagnostics, Statens Serum Institut, Copenhagen, Denmark, Artillerivej 5, 2300 Copenahgen S | Albertsen lab, Department of Chemistry and Bioscience, Aalborg University, Denmark |
| EPI_ISL_444870 | Department of Virus and Microbiological Special Diagnostics, Statens Serum Institut, Copenhagen, Denmark, Artillerivej 5, 2300 Copenahgen S | Albertsen lab, Department of Chemistry and Bioscience, Aalborg University, Denmark |

| EPI_Accession_ID | Originating Laboratory | Submitting Laboratory |
|---|---|---|
| EPI_ISL_444871 | Department of Virus and Microbiological Special Diagnostics, Statens Serum Institut, Copenhagen, Denmark, Artillerivej 5, 2300 Copenahgen S | Albertsen lab, Department of Chemistry and Bioscience, Aalborg University, Denmark |
| EPI_ISL_444872 | Department of Virus and Microbiological Special Diagnostics, Statens Serum Institut, Copenhagen, Denmark, Artillerivej 5, 2300 Copenahgen S | Albertsen lab, Department of Chemistry and Bioscience, Aalborg University, Denmark |
| EPI_ISL_444873 | Department of Virus and Microbiological Special Diagnostics, Statens Serum Institut, Copenhagen, Denmark, Artillerivej 5, 2300 Copenahgen S | Albertsen lab, Department of Chemistry and Bioscience, Aalborg University, Denmark |
| EPI_ISL_444874 | Department of Virus and Microbiological Special Diagnostics, Statens Serum Institut, Copenhagen, Denmark, Artillerivej 5, 2300 Copenahgen S | Albertsen lab, Department of Chemistry and Bioscience, Aalborg University, Denmark |
| EPI_ISL_444875 | Department of Virus and Microbiological Special Diagnostics, Statens Serum Institut, Copenhagen, Denmark, Artillerivej 5, 2300 Copenahgen S | Albertsen lab, Department of Chemistry and Bioscience, Aalborg University, Denmark |
| EPI_ISL_444876 | Department of Virus and Microbiological Special Diagnostics, Statens Serum Institut, Copenhagen, Denmark, Artillerivej 5, 2300 Copenahgen S | Albertsen lab, Department of Chemistry and Bioscience, Aalborg University, Denmark |
| EPI_ISL_444877 | Department of Virus and Microbiological Special Diagnostics, Statens Serum Institut, Copenhagen, Denmark, Artillerivej 5, 2300 Copenahgen S | Albertsen lab, Department of Chemistry and Bioscience, Aalborg University, Denmark |
| EPI_ISL_444878 | Department of Virus and Microbiological Special Diagnostics, Statens Serum Institut, Copenhagen, Denmark, Artillerivej 5, 2300 Copenahgen S | Albertsen lab, Department of Chemistry and Bioscience, Aalborg University, Denmark |
| EPI_ISL_444879 | Department of Virus and Microbiological Special Diagnostics, Statens Serum Institut, Copenhagen, Denmark, Artillerivej 5, 2300 Copenahgen S | Albertsen lab, Department of Chemistry and Bioscience, Aalborg University, Denmark |
| EPI_ISL_444880 | Department of Virus and Microbiological Special Diagnostics, Statens Serum Institut, Copenhagen, Denmark, Artillerivej 5, 2300 Copenahgen S | Albertsen lab, Department of Chemistry and Bioscience, Aalborg University, Denmark |
| EPI_ISL_444881 | Department of Virus and Microbiological Special Diagnostics, Statens Serum Institut, Copenhagen, Denmark, Artillerivej 5, 2300 Copenahgen S | Albertsen lab, Department of Chemistry and Bioscience, Aalborg University, Denmark |

| EPI_Accession_ID | Originating Laboratory | Submitting Laboratory |
|---|---|---|
| EPI_ISL_444882 | Department of Virus and Microbiological Special Diagnostics, Statens Serum Institut, Copenhagen, Denmark, Artillerivej 5, 2300 Copenahgen S | Albertsen lab, Department of Chemistry and Bioscience, Aalborg University, Denmark |
| EPI_ISL_444883 | Department of Virus and Microbiological Special Diagnostics, Statens Serum Institut, Copenhagen, Denmark, Artillerivej 5, 2300 Copenahgen S | Albertsen lab, Department of Chemistry and Bioscience, Aalborg University, Denmark |
| EPI_ISL_444884 | Department of Virus and Microbiological Special Diagnostics, Statens Serum Institut, Copenhagen, Denmark, Artillerivej 5, 2300 Copenahgen S | Albertsen lab, Department of Chemistry and Bioscience, Aalborg University, Denmark |
| EPI_ISL_444885 | Department of Virus and Microbiological Special Diagnostics, Statens Serum Institut, Copenhagen, Denmark, Artillerivej 5, 2300 Copenahgen S | Albertsen lab, Department of Chemistry and Bioscience, Aalborg University, Denmark |
| EPI_ISL_444886 | Department of Virus and Microbiological Special Diagnostics, Statens Serum Institut, Copenhagen, Denmark, Artillerivej 5, 2300 Copenahgen S | Albertsen lab, Department of Chemistry and Bioscience, Aalborg University, Denmark |
| EPI_ISL_444887 | Department of Virus and Microbiological Special Diagnostics, Statens Serum Institut, Copenhagen, Denmark, Artillerivej 5, 2300 Copenahgen S | Albertsen lab, Department of Chemistry and Bioscience, Aalborg University, Denmark |
| EPI_ISL_444888 | Department of Virus and Microbiological Special Diagnostics, Statens Serum Institut, Copenhagen, Denmark, Artillerivej 5, 2300 Copenahgen S | Albertsen lab, Department of Chemistry and Bioscience, Aalborg University, Denmark |
| EPI_ISL_444889 | Department of Virus and Microbiological Special Diagnostics, Statens Serum Institut, Copenhagen, Denmark, Artillerivej 5, 2300 Copenahgen S | Albertsen lab, Department of Chemistry and Bioscience, Aalborg University, Denmark |
| EPI_ISL_444890 | Department of Virus and Microbiological Special Diagnostics, Statens Serum Institut, Copenhagen, Denmark, Artillerivej 5, 2300 Copenahgen S | Albertsen lab, Department of Chemistry and Bioscience, Aalborg University, Denmark |
| EPI_ISL_444891 | Department of Virus and Microbiological Special Diagnostics, Statens Serum Institut, Copenhagen, Denmark, Artillerivej 5, 2300 Copenahgen S | Albertsen lab, Department of Chemistry and Bioscience, Aalborg University, Denmark |
| EPI_ISL_444892 | Department of Virus and Microbiological Special Diagnostics, Statens Serum Institut, Copenhagen, Denmark, Artillerivej 5, 2300 Copenahgen S | Albertsen lab, Department of Chemistry and Bioscience, Aalborg University, Denmark |

| EPI_Accession_ID | Originating Laboratory | Submitting Laboratory |
|---|---|---|
| EPI_ISL_444893 | Department of Virus and Microbiological Special Diagnostics, Statens Serum Institut, Copenhagen, Denmark, Artillerivej 5, 2300 Copenahgen S | Albertsen lab, Department of Chemistry and Bioscience, Aalborg University, Denmark |
| EPI_ISL_444894 | Department of Virus and Microbiological Special Diagnostics, Statens Serum Institut, Copenhagen, Denmark, Artillerivej 5, 2300 Copenahgen S | Albertsen lab, Department of Chemistry and Bioscience, Aalborg University, Denmark |
| EPI_ISL_444895 | Department of Virus and Microbiological Special Diagnostics, Statens Serum Institut, Copenhagen, Denmark, Artillerivej 5, 2300 Copenahgen S | Albertsen lab, Department of Chemistry and Bioscience, Aalborg University, Denmark |
| EPI_ISL_444896 | Department of Virus and Microbiological Special Diagnostics, Statens Serum Institut, Copenhagen, Denmark, Artillerivej 5, 2300 Copenahgen S | Albertsen lab, Department of Chemistry and Bioscience, Aalborg University, Denmark |
| EPI_ISL_444897 | Department of Virus and Microbiological Special Diagnostics, Statens Serum Institut, Copenhagen, Denmark, Artillerivej 5, 2300 Copenahgen S | Albertsen lab, Department of Chemistry and Bioscience, Aalborg University, Denmark |
| EPI_ISL_444898 | Department of Virus and Microbiological Special Diagnostics, Statens Serum Institut, Copenhagen, Denmark, Artillerivej 5, 2300 Copenahgen S | Albertsen lab, Department of Chemistry and Bioscience, Aalborg University, Denmark |
| EPI_ISL_444899 | Department of Virus and Microbiological Special Diagnostics, Statens Serum Institut, Copenhagen, Denmark, Artillerivej 5, 2300 Copenahgen S | Albertsen lab, Department of Chemistry and Bioscience, Aalborg University, Denmark |
| EPI_ISL_444900 | Department of Virus and Microbiological Special Diagnostics, Statens Serum Institut, Copenhagen, Denmark, Artillerivej 5, 2300 Copenahgen S | Albertsen lab, Department of Chemistry and Bioscience, Aalborg University, Denmark |
| EPI_ISL_444901 | Department of Virus and Microbiological Special Diagnostics, Statens Serum Institut, Copenhagen, Denmark, Artillerivej 5, 2300 Copenahgen S | Albertsen lab, Department of Chemistry and Bioscience, Aalborg University, Denmark |
| EPI_ISL_444902 | Department of Virus and Microbiological Special Diagnostics, Statens Serum Institut, Copenhagen, Denmark, Artillerivej 5, 2300 Copenahgen S | Albertsen lab, Department of Chemistry and Bioscience, Aalborg University, Denmark |
| EPI_ISL_444903 | Department of Virus and Microbiological Special Diagnostics, Statens Serum Institut, Copenhagen, Denmark, Artillerivej 5, 2300 Copenahgen S | Albertsen lab, Department of Chemistry and Bioscience, Aalborg University, Denmark |

| EPI_Accession_ID | Originating Laboratory | Submitting Laboratory |
|---|---|---|
| EPI_ISL_444904 | Department of Virus and Microbiological Special Diagnostics, Statens Serum Institut, Copenhagen, Denmark, Artillerivej 5, 2300 Copenahgen S | Albertsen lab, Department of Chemistry and Bioscience, Aalborg University, Denmark |
| EPI_ISL_444905 | Department of Virus and Microbiological Special Diagnostics, Statens Serum Institut, Copenhagen, Denmark, Artillerivej 5, 2300 Copenahgen S | Albertsen lab, Department of Chemistry and Bioscience, Aalborg University, Denmark |
| EPI_ISL_444906 | Department of Virus and Microbiological Special Diagnostics, Statens Serum Institut, Copenhagen, Denmark, Artillerivej 5, 2300 Copenahgen S | Albertsen lab, Department of Chemistry and Bioscience, Aalborg University, Denmark |
| EPI_ISL_444907 | Department of Virus and Microbiological Special Diagnostics, Statens Serum Institut, Copenhagen, Denmark, Artillerivej 5, 2300 Copenahgen S | Albertsen lab, Department of Chemistry and Bioscience, Aalborg University, Denmark |
| EPI_ISL_444908 | Department of Virus and Microbiological Special Diagnostics, Statens Serum Institut, Copenhagen, Denmark, Artillerivej 5, 2300 Copenahgen S | Albertsen lab, Department of Chemistry and Bioscience, Aalborg University, Denmark |
| EPI_ISL_444909 | Department of Virus and Microbiological Special Diagnostics, Statens Serum Institut, Copenhagen, Denmark, Artillerivej 5, 2300 Copenahgen S | Albertsen lab, Department of Chemistry and Bioscience, Aalborg University, Denmark |
| EPI_ISL_444910 | Department of Virus and Microbiological Special Diagnostics, Statens Serum Institut, Copenhagen, Denmark, Artillerivej 5, 2300 Copenahgen S | Albertsen lab, Department of Chemistry and Bioscience, Aalborg University, Denmark |
| EPI_ISL_444911 | Department of Virus and Microbiological Special Diagnostics, Statens Serum Institut, Copenhagen, Denmark, Artillerivej 5, 2300 Copenahgen S | Albertsen lab, Department of Chemistry and Bioscience, Aalborg University, Denmark |
| EPI_ISL_444912 | Department of Virus and Microbiological Special Diagnostics, Statens Serum Institut, Copenhagen, Denmark, Artillerivej 5, 2300 Copenahgen S | Albertsen lab, Department of Chemistry and Bioscience, Aalborg University, Denmark |
| EPI_ISL_444913 | Department of Virus and Microbiological Special Diagnostics, Statens Serum Institut, Copenhagen, Denmark, Artillerivej 5, 2300 Copenahgen S | Albertsen lab, Department of Chemistry and Bioscience, Aalborg University, Denmark |
| EPI_ISL_444914 | Department of Virus and Microbiological Special Diagnostics, Statens Serum Institut, Copenhagen, Denmark, Artillerivej 5, 2300 Copenahgen S | Albertsen lab, Department of Chemistry and Bioscience, Aalborg University, Denmark |

| EPI_Accession_ID | Originating Laboratory | Submitting Laboratory |
|---|---|---|
| EPI_ISL_444915 | Department of Virus and Microbiological Special Diagnostics, Statens Serum Institut, Copenhagen, Denmark, Artillerivej 5, 2300 Copenahgen S | Albertsen lab, Department of Chemistry and Bioscience, Aalborg University, Denmark |
| EPI_ISL_444916 | Department of Virus and Microbiological Special Diagnostics, Statens Serum Institut, Copenhagen, Denmark, Artillerivej 5, 2300 Copenahgen S | Albertsen lab, Department of Chemistry and Bioscience, Aalborg University, Denmark |
| EPI_ISL_444917 | Department of Virus and Microbiological Special Diagnostics, Statens Serum Institut, Copenhagen, Denmark, Artillerivej 5, 2300 Copenahgen S | Albertsen lab, Department of Chemistry and Bioscience, Aalborg University, Denmark |
| EPI_ISL_444918 | Department of Virus and Microbiological Special Diagnostics, Statens Serum Institut, Copenhagen, Denmark, Artillerivej 5, 2300 Copenahgen S | Albertsen lab, Department of Chemistry and Bioscience, Aalborg University, Denmark |
| EPI_ISL_444919 | Department of Virus and Microbiological Special Diagnostics, Statens Serum Institut, Copenhagen, Denmark, Artillerivej 5, 2300 Copenahgen S | Albertsen lab, Department of Chemistry and Bioscience, Aalborg University, Denmark |
| EPI_ISL_444920 | Department of Virus and Microbiological Special Diagnostics, Statens Serum Institut, Copenhagen, Denmark, Artillerivej 5, 2300 Copenahgen S | Albertsen lab, Department of Chemistry and Bioscience, Aalborg University, Denmark |
| EPI_ISL_444921 | Department of Virus and Microbiological Special Diagnostics, Statens Serum Institut, Copenhagen, Denmark, Artillerivej 5, 2300 Copenahgen S | Albertsen lab, Department of Chemistry and Bioscience, Aalborg University, Denmark |
| EPI_ISL_444922 | Department of Virus and Microbiological Special Diagnostics, Statens Serum Institut, Copenhagen, Denmark, Artillerivej 5, 2300 Copenahgen S | Albertsen lab, Department of Chemistry and Bioscience, Aalborg University, Denmark |
| EPI_ISL_444923 | Department of Virus and Microbiological Special Diagnostics, Statens Serum Institut, Copenhagen, Denmark, Artillerivej 5, 2300 Copenahgen S | Albertsen lab, Department of Chemistry and Bioscience, Aalborg University, Denmark |
| EPI_ISL_444924 | Department of Virus and Microbiological Special Diagnostics, Statens Serum Institut, Copenhagen, Denmark, Artillerivej 5, 2300 Copenahgen S | Albertsen lab, Department of Chemistry and Bioscience, Aalborg University, Denmark |
| EPI_ISL_444925 | Department of Virus and Microbiological Special Diagnostics, Statens Serum Institut, Copenhagen, Denmark, Artillerivej 5, 2300 Copenahgen S | Albertsen lab, Department of Chemistry and Bioscience, Aalborg University, Denmark |

| EPI_Accession_ID | Originating Laboratory | Submitting Laboratory |
|---|---|---|
| EPI_ISL_444926 | Department of Virus and Microbiological Special Diagnostics, Statens Serum Institut, Copenhagen, Denmark, Artillerivej 5, 2300 Copenahgen S | Albertsen lab, Department of Chemistry and Bioscience, Aalborg University, Denmark |
| EPI_ISL_444927 | Department of Virus and Microbiological Special Diagnostics, Statens Serum Institut, Copenhagen, Denmark, Artillerivej 5, 2300 Copenahgen S | Albertsen lab, Department of Chemistry and Bioscience, Aalborg University, Denmark |
| EPI_ISL_444928 | Department of Virus and Microbiological Special Diagnostics, Statens Serum Institut, Copenhagen, Denmark, Artillerivej 5, 2300 Copenahgen S | Albertsen lab, Department of Chemistry and Bioscience, Aalborg University, Denmark |
| EPI_ISL_444929 | Department of Virus and Microbiological Special Diagnostics, Statens Serum Institut, Copenhagen, Denmark, Artillerivej 5, 2300 Copenahgen S | Albertsen lab, Department of Chemistry and Bioscience, Aalborg University, Denmark |
| EPI_ISL_444930 | Department of Virus and Microbiological Special Diagnostics, Statens Serum Institut, Copenhagen, Denmark, Artillerivej 5, 2300 Copenahgen S | Albertsen lab, Department of Chemistry and Bioscience, Aalborg University, Denmark |
| EPI_ISL_444931 | Department of Virus and Microbiological Special Diagnostics, Statens Serum Institut, Copenhagen, Denmark, Artillerivej 5, 2300 Copenahgen S | Albertsen lab, Department of Chemistry and Bioscience, Aalborg University, Denmark |
| EPI_ISL_444932 | Department of Virus and Microbiological Special Diagnostics, Statens Serum Institut, Copenhagen, Denmark, Artillerivej 5, 2300 Copenahgen S | Albertsen lab, Department of Chemistry and Bioscience, Aalborg University, Denmark |
| EPI_ISL_444933 | Department of Virus and Microbiological Special Diagnostics, Statens Serum Institut, Copenhagen, Denmark, Artillerivej 5, 2300 Copenahgen S | Albertsen lab, Department of Chemistry and Bioscience, Aalborg University, Denmark |
| EPI_ISL_444934 | Department of Virus and Microbiological Special Diagnostics, Statens Serum Institut, Copenhagen, Denmark, Artillerivej 5, 2300 Copenahgen S | Albertsen lab, Department of Chemistry and Bioscience, Aalborg University, Denmark |
| EPI_ISL_444935 | Department of Virus and Microbiological Special Diagnostics, Statens Serum Institut, Copenhagen, Denmark, Artillerivej 5, 2300 Copenahgen S | Albertsen lab, Department of Chemistry and Bioscience, Aalborg University, Denmark |
| EPI_ISL_444936 | Department of Virus and Microbiological Special Diagnostics, Statens Serum Institut, Copenhagen, Denmark, Artillerivej 5, 2300 Copenahgen S | Albertsen lab, Department of Chemistry and Bioscience, Aalborg University, Denmark |

| EPI_Accession_ID | Originating Laboratory | Submitting Laboratory |
|---|---|---|
| EPI_ISL_444937 | Department of Virus and Microbiological Special Diagnostics, Statens Serum Institut, Copenhagen, Denmark, Artillerivej 5, 2300 Copenahgen S | Albertsen lab, Department of Chemistry and Bioscience, Aalborg University, Denmark |
| EPI_ISL_444938 | Department of Virus and Microbiological Special Diagnostics, Statens Serum Institut, Copenhagen, Denmark, Artillerivej 5, 2300 Copenahgen S | Albertsen lab, Department of Chemistry and Bioscience, Aalborg University, Denmark |
| EPI_ISL_444939 | Department of Virus and Microbiological Special Diagnostics, Statens Serum Institut, Copenhagen, Denmark, Artillerivej 5, 2300 Copenahgen S | Albertsen lab, Department of Chemistry and Bioscience, Aalborg University, Denmark |
| EPI_ISL_444940 | Department of Virus and Microbiological Special Diagnostics, Statens Serum Institut, Copenhagen, Denmark, Artillerivej 5, 2300 Copenahgen S | Albertsen lab, Department of Chemistry and Bioscience, Aalborg University, Denmark |
| EPI_ISL_444941 | Department of Virus and Microbiological Special Diagnostics, Statens Serum Institut, Copenhagen, Denmark, Artillerivej 5, 2300 Copenahgen S | Albertsen lab, Department of Chemistry and Bioscience, Aalborg University, Denmark |
| EPI_ISL_444942 | Department of Virus and Microbiological Special Diagnostics, Statens Serum Institut, Copenhagen, Denmark, Artillerivej 5, 2300 Copenahgen S | Albertsen lab, Department of Chemistry and Bioscience, Aalborg University, Denmark |
| EPI_ISL_444943 | Department of Virus and Microbiological Special Diagnostics, Statens Serum Institut, Copenhagen, Denmark, Artillerivej 5, 2300 Copenahgen S | Albertsen lab, Department of Chemistry and Bioscience, Aalborg University, Denmark |
| EPI_ISL_444944 | Department of Virus and Microbiological Special Diagnostics, Statens Serum Institut, Copenhagen, Denmark, Artillerivej 5, 2300 Copenahgen S | Albertsen lab, Department of Chemistry and Bioscience, Aalborg University, Denmark |
| EPI_ISL_444945 | Department of Virus and Microbiological Special Diagnostics, Statens Serum Institut, Copenhagen, Denmark, Artillerivej 5, 2300 Copenahgen S | Albertsen lab, Department of Chemistry and Bioscience, Aalborg University, Denmark |
| EPI_ISL_444946 | Department of Virus and Microbiological Special Diagnostics, Statens Serum Institut, Copenhagen, Denmark, Artillerivej 5, 2300 Copenahgen S | Albertsen lab, Department of Chemistry and Bioscience, Aalborg University, Denmark |
| EPI_ISL_444947 | Department of Virus and Microbiological Special Diagnostics, Statens Serum Institut, Copenhagen, Denmark, Artillerivej 5, 2300 Copenahgen S | Albertsen lab, Department of Chemistry and Bioscience, Aalborg University, Denmark |

| EPI_Accession_ID | Originating Laboratory | Submitting Laboratory |
|---|---|---|
| EPI_ISL_444948 | Department of Virus and Microbiological Special Diagnostics, Statens Serum Institut, Copenhagen, Denmark, Artillerivej 5, 2300 Copenahgen S | Albertsen lab, Department of Chemistry and Bioscience, Aalborg University, Denmark |
| EPI_ISL_444949 | Department of Virus and Microbiological Special Diagnostics, Statens Serum Institut, Copenhagen, Denmark, Artillerivej 5, 2300 Copenahgen S | Albertsen lab, Department of Chemistry and Bioscience, Aalborg University, Denmark |
| EPI_ISL_444950 | Department of Virus and Microbiological Special Diagnostics, Statens Serum Institut, Copenhagen, Denmark, Artillerivej 5, 2300 Copenahgen S | Albertsen lab, Department of Chemistry and Bioscience, Aalborg University, Denmark |
| EPI_ISL_444951 | Department of Virus and Microbiological Special Diagnostics, Statens Serum Institut, Copenhagen, Denmark, Artillerivej 5, 2300 Copenahgen S | Albertsen lab, Department of Chemistry and Bioscience, Aalborg University, Denmark |
| EPI_ISL_444952 | Department of Virus and Microbiological Special Diagnostics, Statens Serum Institut, Copenhagen, Denmark, Artillerivej 5, 2300 Copenahgen S | Albertsen lab, Department of Chemistry and Bioscience, Aalborg University, Denmark |
| EPI_ISL_444953 | Department of Virus and Microbiological Special Diagnostics, Statens Serum Institut, Copenhagen, Denmark, Artillerivej 5, 2300 Copenahgen S | Albertsen lab, Department of Chemistry and Bioscience, Aalborg University, Denmark |
| EPI_ISL_444954 | Department of Virus and Microbiological Special Diagnostics, Statens Serum Institut, Copenhagen, Denmark, Artillerivej 5, 2300 Copenahgen S | Albertsen lab, Department of Chemistry and Bioscience, Aalborg University, Denmark |
| EPI_ISL_444955 | Department of Virus and Microbiological Special Diagnostics, Statens Serum Institut, Copenhagen, Denmark, Artillerivej 5, 2300 Copenahgen S | Albertsen lab, Department of Chemistry and Bioscience, Aalborg University, Denmark |
| EPI_ISL_444956 | Department of Virus and Microbiological Special Diagnostics, Statens Serum Institut, Copenhagen, Denmark, Artillerivej 5, 2300 Copenahgen S | Albertsen lab, Department of Chemistry and Bioscience, Aalborg University, Denmark |
| EPI_ISL_444957 | Department of Virus and Microbiological Special Diagnostics, Statens Serum Institut, Copenhagen, Denmark, Artillerivej 5, 2300 Copenahgen S | Albertsen lab, Department of Chemistry and Bioscience, Aalborg University, Denmark |
| EPI_ISL_444958 | Department of Virus and Microbiological Special Diagnostics, Statens Serum Institut, Copenhagen, Denmark, Artillerivej 5, 2300 Copenahgen S | Albertsen lab, Department of Chemistry and Bioscience, Aalborg University, Denmark |

| EPI_Accession_ID | Originating Laboratory | Submitting Laboratory |
|---|---|---|
| EPI_ISL_444959 | Department of Virus and Microbiological Special Diagnostics, Statens Serum Institut, Copenhagen, Denmark, Artillerivej 5, 2300 Copenahgen S | Albertsen lab, Department of Chemistry and Bioscience, Aalborg University, Denmark |
| EPI_ISL_444960 | Department of Virus and Microbiological Special Diagnostics, Statens Serum Institut, Copenhagen, Denmark, Artillerivej 5, 2300 Copenahgen S | Albertsen lab, Department of Chemistry and Bioscience, Aalborg University, Denmark |
| EPI_ISL_444961 | Department of Virus and Microbiological Special Diagnostics, Statens Serum Institut, Copenhagen, Denmark, Artillerivej 5, 2300 Copenahgen S | Albertsen lab, Department of Chemistry and Bioscience, Aalborg University, Denmark |
| EPI_ISL_444962 | Department of Virus and Microbiological Special Diagnostics, Statens Serum Institut, Copenhagen, Denmark, Artillerivej 5, 2300 Copenahgen S | Albertsen lab, Department of Chemistry and Bioscience, Aalborg University, Denmark |
| EPI_ISL_444963 | Department of Virus and Microbiological Special Diagnostics, Statens Serum Institut, Copenhagen, Denmark, Artillerivej 5, 2300 Copenahgen S | Albertsen lab, Department of Chemistry and Bioscience, Aalborg University, Denmark |
| EPI_ISL_444964 | Department of Virus and Microbiological Special Diagnostics, Statens Serum Institut, Copenhagen, Denmark, Artillerivej 5, 2300 Copenahgen S | Albertsen lab, Department of Chemistry and Bioscience, Aalborg University, Denmark |
| EPI_ISL_444965 | Department of Virus and Microbiological Special Diagnostics, Statens Serum Institut, Copenhagen, Denmark, Artillerivej 5, 2300 Copenahgen S | Albertsen lab, Department of Chemistry and Bioscience, Aalborg University, Denmark |
| EPI_ISL_444966 | Department of Virus and Microbiological Special Diagnostics, Statens Serum Institut, Copenhagen, Denmark, Artillerivej 5, 2300 Copenahgen S | Albertsen lab, Department of Chemistry and Bioscience, Aalborg University, Denmark |
| EPI_ISL_444967 | Department of Virus and Microbiological Special Diagnostics, Statens Serum Institut, Copenhagen, Denmark, Artillerivej 5, 2300 Copenahgen S | Albertsen lab, Department of Chemistry and Bioscience, Aalborg University, Denmark |
| EPI_ISL_444968 | Department of Virus and Microbiological Special Diagnostics, Statens Serum Institut, Copenhagen, Denmark, Artillerivej 5, 2300 Copenahgen S | Albertsen lab, Department of Chemistry and Bioscience, Aalborg University, Denmark |
| EPI_ISL_444969 | Guangzhou Eighth People's Hospital (Jiahe Sector) | Institute of Human Virology, Zhongshan School of Medicine, Sun Yat-sen University |

| EPI_Accession_ID | Originating Laboratory | Submitting Laboratory |
| --- | --- | --- |
| EPI_ISL_444971 | Hospital Universitari Vall d'Hebron - Vall d'Hebron Institut de Recerca | Hospital Universitari Vall d'Hebron |
| EPI_ISL_444972 | Hospital Universitari Vall d'Hebron - Vall d'Hebron Institut de Recerca | Hospital Universitari Vall d'Hebron |
| EPI_ISL_444973 | Hospital Universitari Vall d`Hebron - Vall d'Hebron Institut de Recerca | Hospital Universitari Vall d'Hebron |
| EPI_ISL_444974 | Hospital Universitari Vall d'Hebron - Vall d'Hebron Institut de Recerca | Hospital Universitari Vall d'Hebron |
| EPI_ISL_444975 | Hospital Universitari Vall d'Hebron - Vall d'Hebron Institut de Recerca | Hospital Universitari Vall d'Hebron |
| EPI_ISL_444976 | Hospital Universitari Vall d'Hebron - Vall d'Hebron Institut de Recerca | Hospital Universitari Vall d'Hebron |
| EPI_ISL_444977 | Hospital Universitari Vall d'Hebron - Vall d'Hebron Institut de Recerca | Hospital Universitari Vall d'Hebron |
| EPI_ISL_444978 | Hospital Universitari Vall d'Hebron - Vall d'Hebron Institut de Recerca | Hospital Universitari Vall d'Hebron |
| EPI_ISL_444979 | Hospital Universitari Vall d'Hebron - Vall d'Hebron Institut de Recerca | Hospital Universitari Vall d'Hebron |
| EPI_ISL_444980 | Hospital Universitari Vall d'Hebron - Vall d"hebron Institut de Recerca | Hospital Universitari Vall d'Hebron |
| EPI_ISL_444981 | Hospital Universitari Vall d'Hebron - Vall d"hebron Institut de Recerca | Hospital Universitari Vall d'Hebron |
| EPI_ISL_444982 | Hospital Universitari Vall d'Hebron - Vall d"hebron Institut de Recerca | Hospital Universitari Vall d'Hebron |
| EPI_ISL_444983 | Hospital Universitari Vall d'Hebron - Vall d"hebron Institut de Recerca | Hospital Universitari Vall d'Hebron |
| EPI_ISL_444984 | Hospital Universitari Vall d'Hebron - Vall d'Hebron Institut de Recerca | Hospital Universitari Vall d'Hebron |
| EPI_ISL_444985 | Hospital Universitari Vall d'Hebron - Vall d'Hebron Institut de Recerca | Hospital Universitari Vall d'Hebron |
| EPI_ISL_444986 | Hospital Universitari Vall d'Hebron - Vall d'Hebron Institut de Recerca | Hospital Universitari Vall d'Hebron |

| EPI_Accession_ID | Originating Laboratory | Submitting Laboratory |
|---|---|---|
| EPI_ISL_444987 | Hospital Universitari Vall d'Hebron - Vall d'Hebron Institut de Recerca | Hospital Universitari Vall d'Hebron |
| EPI_ISL_444988 | Hospital Universitari Vall d'Hebron - Vall d'Hebron Institut de Recerca | Hospital Universitari Vall d'Hebron |
| EPI_ISL_444989 | Hospital Universitari Vall d'Hebron - Vall d'Hebron Institut de Recerca | Hospital Universitari Vall d'Hebron |
| EPI_ISL_444990 | Hospital Universitari Vall d'Hebron - Vall d'Hebron Institut de Recerca | Hospital Universitari Vall d'Hebron |
| EPI_ISL_444994 | Naval Health Research Center | Naval Medical Research Center Biological Defense Research Directorate |
| EPI_ISL_444995 | Naval Health Research Center | Naval Medical Research Center Biological Defense Research Directorate |
| EPI_ISL_444996 | Naval Health Research Center | Naval Medical Research Center Biological Defense Research Directorate |
| EPI_ISL_444997 | Naval Health Research Center | Naval Medical Research Center Biological Defense Research Directorate |
| EPI_ISL_444998 | Naval Health Research Center | Naval Medical Research Center Biological Defense Research Directorate |
| EPI_ISL_444999 | Naval Health Research Center | Naval Medical Research Center Biological Defense Research Directorate |
| EPI_ISL_445000 | Naval Health Research Center | Naval Medical Research Center Biological Defense Research Directorate |
| EPI_ISL_445004 | Florida Bureau of Public Health Laboratories | Florida Bureau of Public Health Laboratories |
| EPI_ISL_445005 | Florida Bureau of Public Health Laboratories | Florida Bureau of Public Health Laboratories |
| EPI_ISL_445006 | Florida Bureau of Public Health Laboratories | Florida Bureau of Public Health Laboratories |
| EPI_ISL_445008 | Florida Bureau of Public Health Laboratories | Florida Bureau of Public Health Laboratories |
| EPI_ISL_445009 | Florida Bureau of Public Health Laboratories | Florida Bureau of Public Health Laboratories |
| EPI_ISL_445011 | Florida Bureau of Public Health Laboratories | Florida Bureau of Public Health Laboratories |
| EPI_ISL_445012 | Florida Bureau of Public Health Laboratories | Florida Bureau of Public Health Laboratories |
| EPI_ISL_445013 | Florida Bureau of Public Health Laboratories | Florida Bureau of Public Health Laboratories |
| EPI_ISL_445014 | Florida Bureau of Public Health Laboratories | Florida Bureau of Public Health Laboratories |
| EPI_ISL_445015 | Florida Bureau of Public Health Laboratories | Florida Bureau of Public Health Laboratories |
| EPI_ISL_445016 | Florida Bureau of Public Health Laboratories | Florida Bureau of Public Health Laboratories |

| EPI_Accession_ID | Originating Laboratory | Submitting Laboratory |
|---|---|---|
| EPI_ISL_445017 | Florida Bureau of Public Health Laboratories | Florida Bureau of Public Health Laboratories |
| EPI_ISL_445019 | Florida Bureau of Public Health Laboratories | Florida Bureau of Public Health Laboratories |
| EPI_ISL_445021 | Florida Bureau of Public Health Laboratories | Florida Bureau of Public Health Laboratories |
| EPI_ISL_445022 | Florida Bureau of Public Health Laboratories | Florida Bureau of Public Health Laboratories |
| EPI_ISL_445023 | Florida Bureau of Public Health Laboratories | Florida Bureau of Public Health Laboratories |
| EPI_ISL_445024 | Florida Bureau of Public Health Laboratories | Florida Bureau of Public Health Laboratories |
| EPI_ISL_445025 | Florida Bureau of Public Health Laboratories | Florida Bureau of Public Health Laboratories |
| EPI_ISL_445026 | Florida Bureau of Public Health Laboratories | Florida Bureau of Public Health Laboratories |
| EPI_ISL_445027 | Florida Bureau of Public Health Laboratories | Florida Bureau of Public Health Laboratories |
| EPI_ISL_445029 | Florida Bureau of Public Health Laboratories | Florida Bureau of Public Health Laboratories |
| EPI_ISL_445030 | Florida Bureau of Public Health Laboratories | Florida Bureau of Public Health Laboratories |
| EPI_ISL_445032 | Florida Bureau of Public Health Laboratories | Florida Bureau of Public Health Laboratories |
| EPI_ISL_445033 | Florida Bureau of Public Health Laboratories | Florida Bureau of Public Health Laboratories |
| EPI_ISL_445034 | Florida Bureau of Public Health Laboratories | Florida Bureau of Public Health Laboratories |
| EPI_ISL_445035 | Florida Bureau of Public Health Laboratories | Florida Bureau of Public Health Laboratories |
| EPI_ISL_445036 | Florida Bureau of Public Health Laboratories | Florida Bureau of Public Health Laboratories |
| EPI_ISL_445037 | Florida Bureau of Public Health Laboratories | Florida Bureau of Public Health Laboratories |
| EPI_ISL_445038 | Florida Bureau of Public Health Laboratories | Florida Bureau of Public Health Laboratories |
| EPI_ISL_445039 | Florida Bureau of Public Health Laboratories | Florida Bureau of Public Health Laboratories |
| EPI_ISL_445040 | Florida Bureau of Public Health Laboratories | Florida Bureau of Public Health Laboratories |
| EPI_ISL_445041 | Florida Bureau of Public Health Laboratories | Florida Bureau of Public Health Laboratories |
| EPI_ISL_445042 | Florida Bureau of Public Health Laboratories | Florida Bureau of Public Health Laboratories |
| EPI_ISL_445043 | Florida Bureau of Public Health Laboratories | Florida Bureau of Public Health Laboratories |
| EPI_ISL_445044 | Florida Bureau of Public Health Laboratories | Florida Bureau of Public Health Laboratories |
| EPI_ISL_445045 | Florida Bureau of Public Health Laboratories | Florida Bureau of Public Health Laboratories |
| EPI_ISL_445046 | Florida Bureau of Public Health Laboratories | Florida Bureau of Public Health Laboratories |
| EPI_ISL_445047 | Florida Bureau of Public Health Laboratories | Florida Bureau of Public Health Laboratories |
| EPI_ISL_445048 | Florida Bureau of Public Health Laboratories | Florida Bureau of Public Health Laboratories |
| EPI_ISL_445049 | Florida Bureau of Public Health Laboratories | Florida Bureau of Public Health Laboratories |
| EPI_ISL_445050 | Florida Bureau of Public Health Laboratories | Florida Bureau of Public Health Laboratories |
| EPI_ISL_445053 | Laboratoire National de Sante, Microbiology, Virology | Laboratoire National de Sante, Microbiology, Epidemiology and Microbial Genomics |

| EPI_Accession_ID | Originating Laboratory | Submitting Laboratory |
|---|---|---|
| EPI_ISL_445054 | Laboratoire National de Sante, Microbiology, Virology | Laboratoire National de Sante, Microbiology, Epidemiology and Microbial Genomics |
| EPI_ISL_445055 | Laboratoire National de Sante, Microbiology, Virology | Laboratoire National de Sante, Microbiology, Epidemiology and Microbial Genomics |
| EPI_ISL_445056 | Laboratoire National de Sante, Microbiology, Virology | Laboratoire National de Sante, Microbiology, Epidemiology and Microbial Genomics |
| EPI_ISL_445057 | Laboratoire National de Sante, Microbiology, Virology | Laboratoire National de Sante, Microbiology, Epidemiology and Microbial Genomics |
| EPI_ISL_445058 | Laboratoire National de Sante, Microbiology, Virology | Laboratoire National de Sante, Microbiology, Epidemiology and Microbial Genomics |
| EPI_ISL_445059 | Laboratoire National de Sante, Microbiology, Virology | Laboratoire National de Sante, Microbiology, Epidemiology and Microbial Genomics |
| EPI_ISL_445060 | Laboratoire National de Sante, Microbiology, Virology | Laboratoire National de Sante, Microbiology, Epidemiology and Microbial Genomics |
| EPI_ISL_445061 | Laboratoire National de Sante, Microbiology, Virology | Laboratoire National de Sante, Microbiology, Epidemiology and Microbial Genomics |
| EPI_ISL_445062 | Laboratoire National de Sante, Microbiology, Virology | Laboratoire National de Sante, Microbiology, Epidemiology and Microbial Genomics |
| EPI_ISL_445063 | Laboratoire National de Sante, Microbiology, Virology | Laboratoire National de Sante, Microbiology, Epidemiology and Microbial Genomics |
| EPI_ISL_445064 | Laboratoire National de Sante, Microbiology, Virology | Laboratoire National de Sante, Microbiology, Epidemiology and Microbial Genomics |
| EPI_ISL_445065 | Laboratoire National de Sante, Microbiology, Virology | Laboratoire National de Sante, Microbiology, Epidemiology and Microbial Genomics |
| EPI_ISL_445066 | Laboratoire National de Sante, Microbiology, Virology | Laboratoire National de Sante, Microbiology, Epidemiology and Microbial Genomics |
| EPI_ISL_445067 | Laboratoire National de Sante, Microbiology, Virology | Laboratoire National de Sante, Microbiology, Epidemiology and Microbial Genomics |
| EPI_ISL_445068 | Laboratoire National de Sante, Microbiology, Virology | Laboratoire National de Sante, Microbiology, Epidemiology and Microbial Genomics |
| EPI_ISL_445069 | Laboratoire National de Sante, Microbiology, Virology | Laboratoire National de Sante, Microbiology, Epidemiology and Microbial Genomics |

| EPI_Accession_ID | Originating Laboratory | Submitting Laboratory |
|---|---|---|
| EPI_ISL_445070 | Laboratoire National de Sante, Microbiology, Virology | Laboratoire National de Sante, Microbiology, Epidemiology and Microbial Genomics |
| EPI_ISL_445071 | Laboratoire National de Sante, Microbiology, Virology | Laboratoire National de Sante, Microbiology, Epidemiology and Microbial Genomics |
| EPI_ISL_445072 | Laboratoire National de Sante, Microbiology, Virology | Laboratoire National de Sante, Microbiology, Epidemiology and Microbial Genomics |
| EPI_ISL_445073 | Laboratoire National de Sante, Microbiology, Virology | Laboratoire National de Sante, Microbiology, Epidemiology and Microbial Genomics |
| EPI_ISL_445074 | Laboratoire National de Sante, Microbiology, Virology | Laboratoire National de Sante, Microbiology, Epidemiology and Microbial Genomics |
| EPI_ISL_445075 | Laboratoire National de Sante, Microbiology, Virology | Laboratoire National de Sante, Microbiology, Epidemiology and Microbial Genomics |
| EPI_ISL_445076 | Laboratoire National de Sante, Microbiology, Virology | Laboratoire National de Sante, Microbiology, Epidemiology and Microbial Genomics |
| EPI_ISL_445077 | M Health Fairview | University of Minnesota Genomics Center |
| EPI_ISL_445078 | Baylor College of Medicine | Baylor College of Medicine: HGSC |
| EPI_ISL_445079 | Baylor College of Medicine | Baylor College of Medicine: HGSC |
| EPI_ISL_445080 | Baylor College of Medicine | Baylor College of Medicine: HGSC |
| EPI_ISL_445081 | Baylor College of Medicine | Baylor College of Medicine: HGSC |
| EPI_ISL_445082 | Baylor College of Medicine | Baylor College of Medicine: HGSC |
| EPI_ISL_445083 | Baylor College of Medicine | Baylor College of Medicine: HGSC |
| EPI_ISL_445084 | Baylor College of Medicine | Baylor College of Medicine: HGSC |
| EPI_ISL_445085 | unknown | Virology Unit |
| EPI_ISL_445086 | unknown | Laboratory Diagnostic |
| EPI_ISL_445087 | unknown | Laboratory Diagnostic |
| EPI_ISL_445088 | unknown | Human Genetic Research Center |
| EPI_ISL_445094 | UC San Diego Center for Advanced Laboratory Medicine | Andersen lab at Scripps Research |
| EPI_ISL_445095 | UC San Diego Center for Advanced Laboratory Medicine | Andersen lab at Scripps Research |
| EPI_ISL_445096 | UC San Diego Center for Advanced Laboratory Medicine | Andersen lab at Scripps Research |

| EPI_Accession_ID | Originating Laboratory | Submitting Laboratory |
|---|---|---|
| EPI_ISL_445097 | UC San Diego Center for Advanced Laboratory Medicine | Andersen lab at Scripps Research |
| EPI_ISL_445098 | UC San Diego Center for Advanced Laboratory Medicine | Andersen lab at Scripps Research |
| EPI_ISL_445099 | UC San Diego Center for Advanced Laboratory Medicine | Andersen lab at Scripps Research |
| EPI_ISL_445100 | UC San Diego Center for Advanced Laboratory Medicine | Andersen lab at Scripps Research |
| EPI_ISL_445101 | UC San Diego Center for Advanced Laboratory Medicine | Andersen lab at Scripps Research |
| EPI_ISL_445102 | UC San Diego Center for Advanced Laboratory Medicine | Andersen lab at Scripps Research |
| EPI_ISL_445103 | UC San Diego Center for Advanced Laboratory Medicine | Andersen lab at Scripps Research |
| EPI_ISL_445104 | UC San Diego Center for Advanced Laboratory Medicine | Andersen lab at Scripps Research |
| EPI_ISL_445105 | UC San Diego Center for Advanced Laboratory Medicine | Andersen lab at Scripps Research |
| EPI_ISL_445106 | UC San Diego Center for Advanced Laboratory Medicine | Andersen lab at Scripps Research |
| EPI_ISL_445107 | UC San Diego Center for Advanced Laboratory Medicine | Andersen lab at Scripps Research |
| EPI_ISL_445108 | UC San Diego Center for Advanced Laboratory Medicine | Andersen lab at Scripps Research |
| EPI_ISL_445109 | UC San Diego Center for Advanced Laboratory Medicine | Andersen lab at Scripps Research |
| EPI_ISL_445110 | UC San Diego Center for Advanced Laboratory Medicine | Andersen lab at Scripps Research |
| EPI_ISL_445111 | UC San Diego Center for Advanced Laboratory Medicine | Andersen lab at Scripps Research |
| EPI_ISL_445112 | UC San Diego Center for Advanced Laboratory Medicine | Andersen lab at Scripps Research |

| EPI_Accession_ID | Originating Laboratory | Submitting Laboratory |
|---|---|---|
| EPI_ISL_445113 | UC San Diego Center for Advanced Laboratory Medicine | Andersen lab at Scripps Research |
| EPI_ISL_445114 | UC San Diego Center for Advanced Laboratory Medicine | Andersen lab at Scripps Research |
| EPI_ISL_445115 | UC San Diego Center for Advanced Laboratory Medicine | Andersen lab at Scripps Research |
| EPI_ISL_445116 | UC San Diego Center for Advanced Laboratory Medicine | Andersen lab at Scripps Research |
| EPI_ISL_445117 | UC San Diego Center for Advanced Laboratory Medicine | Andersen lab at Scripps Research |
| EPI_ISL_445118 | Rady's Childrens Hospital | Andersen lab at Scripps Research |
| EPI_ISL_445119 | Robert Garry lab | Andersen lab at Scripps Research |
| EPI_ISL_445120 | Robert Garry lab | Andersen lab at Scripps Research |
| EPI_ISL_445121 | Robert Garry lab | Andersen lab at Scripps Research |
| EPI_ISL_445122 | Robert Garry lab | Andersen lab at Scripps Research |
| EPI_ISL_445123 | Robert Garry lab | Andersen lab at Scripps Research |
| EPI_ISL_445124 | Robert Garry lab | Andersen lab at Scripps Research |
| EPI_ISL_445125 | Robert Garry lab | Andersen lab at Scripps Research |
| EPI_ISL_445126 | Robert Garry lab | Andersen lab at Scripps Research |
| EPI_ISL_445127 | Robert Garry lab | Andersen lab at Scripps Research |
| EPI_ISL_445128 | Robert Garry lab | Andersen lab at Scripps Research |
| EPI_ISL_445129 | Robert Garry lab | Andersen lab at Scripps Research |
| EPI_ISL_445130 | Robert Garry lab | Andersen lab at Scripps Research |
| EPI_ISL_445131 | Robert Garry lab | Andersen lab at Scripps Research |
| EPI_ISL_445132 | Robert Garry lab | Andersen lab at Scripps Research |
| EPI_ISL_445133 | Robert Garry lab | Andersen lab at Scripps Research |
| EPI_ISL_445134 | Robert Garry lab | Andersen lab at Scripps Research |
| EPI_ISL_445135 | Robert Garry lab | Andersen lab at Scripps Research |
| EPI_ISL_445136 | Robert Garry lab | Andersen lab at Scripps Research |
| EPI_ISL_445137 | Robert Garry lab | Andersen lab at Scripps Research |
| EPI_ISL_445138 | Robert Garry lab | Andersen lab at Scripps Research |
| EPI_ISL_445139 | Robert Garry lab | Andersen lab at Scripps Research |
| EPI_ISL_445140 | Robert Garry lab | Andersen lab at Scripps Research |

| EPI_Accession_ID | Originating Laboratory | Submitting Laboratory |
|---|---|---|
| EPI_ISL_445141 | Robert Garry lab | Andersen lab at Scripps Research |
| EPI_ISL_445142 | Robert Garry lab | Andersen lab at Scripps Research |
| EPI_ISL_445143 | Robert Garry lab | Andersen lab at Scripps Research |
| EPI_ISL_445144 | Robert Garry lab | Andersen lab at Scripps Research |
| EPI_ISL_445145 | Robert Garry lab | Andersen lab at Scripps Research |
| EPI_ISL_445146 | Robert Garry lab | Andersen lab at Scripps Research |
| EPI_ISL_445147 | Robert Garry lab | Andersen lab at Scripps Research |
| EPI_ISL_445148 | Robert Garry lab | Andersen lab at Scripps Research |
| EPI_ISL_445149 | Robert Garry lab | Andersen lab at Scripps Research |
| EPI_ISL_445150 | Robert Garry lab | Andersen lab at Scripps Research |
| EPI_ISL_445151 | Robert Garry lab | Andersen lab at Scripps Research |
| EPI_ISL_445152 | Robert Garry lab | Andersen lab at Scripps Research |
| EPI_ISL_445153 | Robert Garry lab | Andersen lab at Scripps Research |
| EPI_ISL_445154 | Robert Garry lab | Andersen lab at Scripps Research |
| EPI_ISL_445155 | Robert Garry lab | Andersen lab at Scripps Research |
| EPI_ISL_445156 | Robert Garry lab | Andersen lab at Scripps Research |
| EPI_ISL_445157 | Robert Garry lab | Andersen lab at Scripps Research |
| EPI_ISL_445158 | Robert Garry lab | Andersen lab at Scripps Research |
| EPI_ISL_445159 | Robert Garry lab | Andersen lab at Scripps Research |
| EPI_ISL_445160 | Robert Garry lab | Andersen lab at Scripps Research |
| EPI_ISL_445161 | Robert Garry lab | Andersen lab at Scripps Research |
| EPI_ISL_445162 | Robert Garry lab | Andersen lab at Scripps Research |
| EPI_ISL_445163 | Robert Garry lab | Andersen lab at Scripps Research |
| EPI_ISL_445164 | Scripps Medical Laboratory | Andersen lab at Scripps Research |
| EPI_ISL_445165 | Scripps Medical Laboratory | Andersen lab at Scripps Research |
| EPI_ISL_445166 | Scripps Medical Laboratory | Andersen lab at Scripps Research |
| EPI_ISL_445167 | Scripps Medical Laboratory | Andersen lab at Scripps Research |
| EPI_ISL_445168 | Scripps Medical Laboratory | Andersen lab at Scripps Research |
| EPI_ISL_445169 | UCSF Clinical Microbiology Laboratory | Chan-Zuckerberg Biohub |
| EPI_ISL_445170 | UCSF Clinical Microbiology Laboratory | Chan-Zuckerberg Biohub |
| EPI_ISL_445171 | UCSF Clinical Microbiology Laboratory | Chan-Zuckerberg Biohub |
| EPI_ISL_445172 | UCSF Clinical Microbiology Laboratory | Chan-Zuckerberg Biohub |
| EPI_ISL_445173 | UCSF Clinical Microbiology Laboratory | Chan-Zuckerberg Biohub |

| EPI_Accession_ID | Originating Laboratory | Submitting Laboratory |
|---|---|---|
| EPI_ISL_445174 | UCSF Clinical Microbiology Laboratory | Chan-Zuckerberg Biohub |
| EPI_ISL_445175 | UCSF Clinical Microbiology Laboratory | Chan-Zuckerberg Biohub |
| EPI_ISL_445176 | UCSF Clinical Microbiology Laboratory | Chan-Zuckerberg Biohub |
| EPI_ISL_445177 | UCSF Clinical Microbiology Laboratory | Chan-Zuckerberg Biohub |
| EPI_ISL_445178 | UCSF Clinical Microbiology Laboratory | Chan-Zuckerberg Biohub |
| EPI_ISL_445179 | UCSF Clinical Microbiology Laboratory | Chan-Zuckerberg Biohub |
| EPI_ISL_445180 | UCSF Clinical Microbiology Laboratory | Chan-Zuckerberg Biohub |
| EPI_ISL_445181 | UCSF Clinical Microbiology Laboratory | Chan-Zuckerberg Biohub |
| EPI_ISL_445182 | UCSF Clinical Microbiology Laboratory | Chan-Zuckerberg Biohub |
| EPI_ISL_445183 | unknown | Takayuki Hishiki Kanagawa Prefectural Institute of Public Health |
| EPI_ISL_445214 | DNA Solution Ltd. | DNA Solution Ltd. |
| EPI_ISL_445215 | DNA Solution Ltd. | DNA Solution Ltd. |
| EPI_ISL_445216 | DNA Solution Ltd. | DNA Solution Ltd. |
| EPI_ISL_445219 | Universidad del Valle, Laboratorio de Microbiologia, VIREM | Universidad del Valle, Universidad Nacional de Colombia-Sede Palmira, International Center for Tropical Agriculture |
| EPI_ISL_445220 | Laborarory for Respiratory Viruses, "Cantacuzino" National Military-Medical Institute for Resararch and Development | Cantacuzino Institute |
| EPI_ISL_445221 | Wasterlakarna | The Public Health Agency of Sweden |
| EPI_ISL_445222 | Saroledens Familjelakare | The Public Health Agency of Sweden |
| EPI_ISL_445223 | Victoria Vard och Halsa | The Public Health Agency of Sweden |
| EPI_ISL_445224 | Narhalsan Olskroken VC | The Public Health Agency of Sweden |
| EPI_ISL_445225 | Surbrunns VC | The Public Health Agency of Sweden |
| EPI_ISL_445226 | Saroledens Familjelakare | The Public Health Agency of Sweden |
| EPI_ISL_445227 | Uppsala Narakut Aleris | The Public Health Agency of Sweden |
| EPI_ISL_445228 | Ulltuna Vardcentral | The Public Health Agency of Sweden |
| EPI_ISL_445229 | Narhalsan Backa vardcentral | The Public Health Agency of Sweden |
| EPI_ISL_445230 | Uppsala Narakut Aleris | The Public Health Agency of Sweden |
| EPI_ISL_445231 | Uppsala Narakut Aleris | The Public Health Agency of Sweden |
| EPI_ISL_445232 | Kungsors VC | The Public Health Agency of Sweden |
| EPI_ISL_445233 | Vardcentralen Brinken | The Public Health Agency of Sweden |

| EPI_Accession_ID | Originating Laboratory | Submitting Laboratory |
|---|---|---|
| EPI_ISL_445234 | Wasterlakarna | The Public Health Agency of Sweden |
| EPI_ISL_445235 | Wasterlakarna | The Public Health Agency of Sweden |
| EPI_ISL_445236 | Narhalsan Backa vardcentral | The Public Health Agency of Sweden |
| EPI_ISL_445237 | Narhalsan Molnlycke, Barn och ungdomsmedicin | The Public Health Agency of Sweden |
| EPI_ISL_445238 | Ãƒâ€“resundslakarna | The Public Health Agency of Sweden |
| EPI_ISL_445239 | Uppsala Narakut Aleris | The Public Health Agency of Sweden |
| EPI_ISL_445240 | Ulltuna Vardcentral | The Public Health Agency of Sweden |
| EPI_ISL_445241 | Ãƒâ€“restadsklinikens VC | The Public Health Agency of Sweden |
| EPI_ISL_445242 | Jokkmokks Halsocentral | The Public Health Agency of Sweden |
| EPI_ISL_445243 | Laboratory for Respiratory Viruses, Cantacuzino National Military-Medical Institute for Research and Development | Cantacuzino Institute |
| EPI_ISL_445244 | Akbiomed lab | Tejgaon College bmb lab |
| EPI_ISL_445245 | CLINICA ALEMANA DE SANTIAGO S.A. | Instituto de Salud Publica de Chile |
| EPI_ISL_445246 | HOSPITAL PUERTO MONTT | Instituto de Salud Publica de Chile |
| EPI_ISL_445247 | UNIVERSIDAD DE LOS ANDES | Instituto de Salud Publica de Chile |
| EPI_ISL_445248 | CLINICA ALEMANA DE SANTIAGO S.A. | Instituto de Salud Publica de Chile |
| EPI_ISL_445249 | CLINICA SANTA MARIA S.A. | Instituto de Salud Publica de Chile |
| EPI_ISL_445250 | CLINICA ALEMANA DE SANTIAGO S.A. | Instituto de Salud Publica de Chile |
| EPI_ISL_445251 | HOSPITAL DE CARABINEROS | Instituto de Salud Publica de Chile |
| EPI_ISL_445252 | PONTIFICIA U. CATOLICA FAC. MEDICINA | Instituto de Salud Publica de Chile |
| EPI_ISL_445253 | CLINICA ALEMANA DE SANTIAGO S.A. | Instituto de Salud Publica de Chile |
| EPI_ISL_445254 | CLINICA ALEMANA DE SANTIAGO S.A. | Instituto de Salud Publica de Chile |
| EPI_ISL_445255 | CLINICA ALEMANA DE SANTIAGO S.A. | Instituto de Salud Publica de Chile |
| EPI_ISL_445256 | CLINICA LAS CONDES S.A. | Instituto de Salud Publica de Chile |
| EPI_ISL_445257 | CLINICA TABANCURA | Instituto de Salud Publica de Chile |
| EPI_ISL_445258 | CLINICA ALEMANA DE SANTIAGO S.A. | Instituto de Salud Publica de Chile |
| EPI_ISL_445259 | CLINICA LAS CONDES S.A. | Instituto de Salud Publica de Chile |
| EPI_ISL_445260 | CLINICA ALEMANA DE SANTIAGO S.A. | Instituto de Salud Publica de Chile |
| EPI_ISL_445261 | INTEGRAMEDICA LAB. CLINICO LTDA. | Instituto de Salud Publica de Chile |
| EPI_ISL_445262 | CLINICA TABANCURA | Instituto de Salud Publica de Chile |
| EPI_ISL_445263 | MEGASALUD SPA. | Instituto de Salud Publica de Chile |
| EPI_ISL_445264 | CLINICA REDSALUD VITACURA. | Instituto de Salud Publica de Chile |

| EPI_Accession_ID | Originating Laboratory | Submitting Laboratory |
|---|---|---|
| EPI_ISL_445265 | PONTIFICIA UNIVERSIDAD CATOLICA DE CHILE | Instituto de Salud Publica de Chile |
| EPI_ISL_445266 | CENTRO ONCOLOGICO DEL NORTE | Instituto de Salud Publica de Chile |
| EPI_ISL_445267 | CENTRO ONCOLOGICO DEL NORTE | Instituto de Salud Publica de Chile |
| EPI_ISL_445268 | HOSPITAL REG.LAUTARO NAVARRO AVARIA | Instituto de Salud Publica de Chile |
| EPI_ISL_445269 | HOSPITAL REG.LAUTARO NAVARRO AVARIA | Instituto de Salud Publica de Chile |
| EPI_ISL_445270 | HOSPITAL DR.HERNAN HENRIQUEZ ARAVENA | Instituto de Salud Publica de Chile |
| EPI_ISL_445271 | LABORATORIO CLINICA CHILLAN | Instituto de Salud Publica de Chile |
| EPI_ISL_445272 | CLINICA CIUDAD DEL MAR | Instituto de Salud Publica de Chile |
| EPI_ISL_445273 | LABORATORIO TORRE MEDICA LTDA. | Instituto de Salud Publica de Chile |
| EPI_ISL_445274 | LABORATORIO TORRE MEDICA LTDA. | Instituto de Salud Publica de Chile |
| EPI_ISL_445275 | HOSPITAL CLINICO FUSAT | Instituto de Salud Publica de Chile |
| EPI_ISL_445276 | HOSPITAL CLINICO FUSAT | Instituto de Salud Publica de Chile |
| EPI_ISL_445277 | FUNDACION DE SALUD EL TENIENTE | Instituto de Salud Publica de Chile |
| EPI_ISL_445278 | LABORATORIO TORRE MEDICA LTDA. | Instituto de Salud Publica de Chile |
| EPI_ISL_445279 | LABORATORIO INMUNOLAB SPA | Instituto de Salud Publica de Chile |
| EPI_ISL_445280 | HOSPITAL REG.LAUTARO NAVARRO AVARIA | Instituto de Salud Publica de Chile |
| EPI_ISL_445281 | HOSPITAL CLINICO DEL SUR | Instituto de Salud Publica de Chile |
| EPI_ISL_445282 | CLINICA MAGALLANES S.A. | Instituto de Salud Publica de Chile |
| EPI_ISL_445283 | CLINICA MAGALLANES S.A. | Instituto de Salud Publica de Chile |
| EPI_ISL_445284 | HOSPITAL REG.LAUTARO NAVARRO AVARIA | Instituto de Salud Publica de Chile |
| EPI_ISL_445285 | CLINICA CIUDAD DEL MAR | Instituto de Salud Publica de Chile |
| EPI_ISL_445286 | HOSPITAL HANGA ROA | Instituto de Salud Publica de Chile |
| EPI_ISL_445287 | CLINICA CIUDAD DEL MAR | Instituto de Salud Publica de Chile |
| EPI_ISL_445288 | HOSPITAL REG.LAUTARO NAVARRO AVARIA | Instituto de Salud Publica de Chile |
| EPI_ISL_445289 | HOSPITAL NAVAL PUERTO WILLIAMS | Instituto de Salud Publica de Chile |
| EPI_ISL_445290 | CLINICA MAGALLANES S.A. | Instituto de Salud Publica de Chile |
| EPI_ISL_445291 | CLINICA MAGALLANES S.A. | Instituto de Salud Publica de Chile |
| EPI_ISL_445292 | CLINICA MAGALLANES S.A. | Instituto de Salud Publica de Chile |
| EPI_ISL_445293 | HOSPITAL REG.LAUTARO NAVARRO AVARIA | Instituto de Salud Publica de Chile |
| EPI_ISL_445294 | HOSPITAL REG.LAUTARO NAVARRO AVARIA | Instituto de Salud Publica de Chile |
| EPI_ISL_445295 | HOSPITAL REG.LAUTARO NAVARRO AVARIA | Instituto de Salud Publica de Chile |
| EPI_ISL_445296 | HOSPITAL REGIONAL DE COYHAIQUE | Instituto de Salud Publica de Chile |
| EPI_ISL_445297 | CLINICA INTEGRAL S.A. | Instituto de Salud Publica de Chile |

| EPI_Accession_ID | Originating Laboratory | Submitting Laboratory |
|---|---|---|
| EPI_ISL_445298 | HOSPITAL DE SAN FERNANDO | Instituto de Salud Publica de Chile |
| EPI_ISL_445299 | CLINICA MAGALLANES S.A. | Instituto de Salud Publica de Chile |
| EPI_ISL_445300 | HOSPITAL DE RANCAGUA | Instituto de Salud Publica de Chile |
| EPI_ISL_445301 | HOSPITAL NAVAL PUERTO WILLIAMS | Instituto de Salud Publica de Chile |
| EPI_ISL_445302 | INSTITUTO MEDICO LEGAL | Instituto de Salud Publica de Chile |
| EPI_ISL_445303 | CTRO.DE SALUD FAMILIAR DR. RAUL YAZIGI | Instituto de Salud Publica de Chile |
| EPI_ISL_445304 | HOSPITAL SAN JUAN DE DIOS | Instituto de Salud Publica de Chile |
| EPI_ISL_445305 | HOSPITAL DE CARABINEROS | Instituto de Salud Publica de Chile |
| EPI_ISL_445306 | CLINICA UC SAN CARLOS DE APOQUINDO | Instituto de Salud Publica de Chile |
| EPI_ISL_445307 | HOSPITAL SAN JOSE DE MAIPO | Instituto de Salud Publica de Chile |
| EPI_ISL_445308 | HOSPITAL EL CARMEN DR.LUIS VALENTIN F. | Instituto de Salud Publica de Chile |
| EPI_ISL_445309 | MUTUAL DE SEGURIDAD C.CH.C. | Instituto de Salud Publica de Chile |
| EPI_ISL_445310 | HOSPITAL DR.SOTERO DEL RIO | Instituto de Salud Publica de Chile |
| EPI_ISL_445311 | MEGASALUD S.A. | Instituto de Salud Publica de Chile |
| EPI_ISL_445312 | CLINICA UC SAN CARLOS DE APOQUINDO | Instituto de Salud Publica de Chile |
| EPI_ISL_445313 | HOSPITAL EL CARMEN DR.LUIS VALENTIN F. | Instituto de Salud Publica de Chile |
| EPI_ISL_445314 | HOSPITAL EL CARMEN DR.LUIS VALENTIN F. | Instituto de Salud Publica de Chile |
| EPI_ISL_445315 | CLINICA UC SAN CARLOS DE APOQUINDO | Instituto de Salud Publica de Chile |
| EPI_ISL_445316 | CESFAM BALMACEDA DE RENCA | Instituto de Salud Publica de Chile |
| EPI_ISL_445317 | UNIV.DE CHILE HOSP.CLINICO | Instituto de Salud Publica de Chile |
| EPI_ISL_445318 | C.DE SALUD FAMILIAR PABLO NERUDA | Instituto de Salud Publica de Chile |
| EPI_ISL_445319 | HOSPITAL FELIX BULNES | Instituto de Salud Publica de Chile |
| EPI_ISL_445320 | C.C.SALUD FAMILIAR PADRE FELIX DONOSO G. | Instituto de Salud Publica de Chile |
| EPI_ISL_445321 | HOSPITAL FELIX BULNES | Instituto de Salud Publica de Chile |
| EPI_ISL_445322 | UNIV.DE CHILE HOSP.CLINICO | Instituto de Salud Publica de Chile |
| EPI_ISL_445323 | HOSP.ENFERMEDADES INFECCIOSAS | Instituto de Salud Publica de Chile |
| EPI_ISL_445324 | INTEGRAMEDICA S.A | Instituto de Salud Publica de Chile |
| EPI_ISL_445325 | HOSPITAL DR.SOTERO DEL RIO | Instituto de Salud Publica de Chile |
| EPI_ISL_445326 | ASISTENCIA PUBLICA DR.ALEJANDRO DEL RIO | Instituto de Salud Publica de Chile |
| EPI_ISL_445327 | HOSPITAL PADRE HURTADO | Instituto de Salud Publica de Chile |
| EPI_ISL_445328 | MEGASALUD S.A. | Instituto de Salud Publica de Chile |
| EPI_ISL_445329 | HOSPITAL DR.SOTERO DEL RIO | Instituto de Salud Publica de Chile |
| EPI_ISL_445330 | CLINICA VESPUCIO S. A. | Instituto de Salud Publica de Chile |

| EPI_Accession_ID | Originating Laboratory | Submitting Laboratory |
|---|---|---|
| EPI_ISL_445331 | HOSPITAL HERMINDA MARTIN CHILLAN | Instituto de Salud Publica de Chile |
| EPI_ISL_445332 | HOSPITAL HERMINDA MARTIN CHILLAN | Instituto de Salud Publica de Chile |
| EPI_ISL_445333 | LABORATORIO CLINICA UNIVERSITARIA DE CONCEPCION | Instituto de Salud Publica de Chile |
| EPI_ISL_445334 | CLINICA UNIVERSITARIA DE PUERTO MONTT S.A. | Instituto de Salud Publica de Chile |
| EPI_ISL_445335 | HOSPITAL DE CALBUCO | Instituto de Salud Publica de Chile |
| EPI_ISL_445336 | HOSPITAL LAS HIGUERAS DE TALCAHUANO | Instituto de Salud Publica de Chile |
| EPI_ISL_445337 | HOSPITAL HANGA ROA | Instituto de Salud Publica de Chile |
| EPI_ISL_445338 | HOSPITAL DR.HERNAN HENRIQUEZ ARAVENA | Instituto de Salud Publica de Chile |
| EPI_ISL_445339 | HOSPITAL DR.HERNAN HENRIQUEZ ARAVENA | Instituto de Salud Publica de Chile |
| EPI_ISL_445340 | HOSPITAL DR.HERNAN HENRIQUEZ ARAVENA | Instituto de Salud Publica de Chile |
| EPI_ISL_445341 | HOSPITAL DR.HERNAN HENRIQUEZ ARAVENA | Instituto de Salud Publica de Chile |
| EPI_ISL_445342 | HOSPITAL DR.HERNAN HENRIQUEZ ARAVENA | Instituto de Salud Publica de Chile |
| EPI_ISL_445343 | HOSPITAL DR.HERNAN HENRIQUEZ ARAVENA | Instituto de Salud Publica de Chile |
| EPI_ISL_445344 | HOSPITAL DR.HERNAN HENRIQUEZ ARAVENA | Instituto de Salud Publica de Chile |
| EPI_ISL_445345 | HOSPITAL DR.HERNAN HENRIQUEZ ARAVENA | Instituto de Salud Publica de Chile |
| EPI_ISL_445346 | HOSPITAL DR.HERNAN HENRIQUEZ ARAVENA | Instituto de Salud Publica de Chile |
| EPI_ISL_445347 | HOSPITAL DR.HERNAN HENRIQUEZ ARAVENA | Instituto de Salud Publica de Chile |
| EPI_ISL_445348 | HOSPITAL DR.HERNAN HENRIQUEZ ARAVENA | Instituto de Salud Publica de Chile |
| EPI_ISL_445349 | HOSPITAL SAN JUAN DE DIOS | Instituto de Salud Publica de Chile |
| EPI_ISL_445350 | HOSPITAL SAN JUAN DE DIOS | Instituto de Salud Publica de Chile |
| EPI_ISL_445351 | HOSPITAL SAN JUAN DE DIOS | Instituto de Salud Publica de Chile |
| EPI_ISL_445352 | HOSPITAL DEL PROFESOR | Instituto de Salud Publica de Chile |
| EPI_ISL_445353 | HOSPITAL PADRE HURTADO | Instituto de Salud Publica de Chile |
| EPI_ISL_445354 | HOSPITAL DE CARABINEROS | Instituto de Salud Publica de Chile |
| EPI_ISL_445355 | MUTUAL DE SEGURIDAD C.CH.C. | Instituto de Salud Publica de Chile |
| EPI_ISL_445356 | PONTIFICIA U. CATOLICA SERV. LABORATORIO | Instituto de Salud Publica de Chile |
| EPI_ISL_445357 | INTEGRAMEDICA CENTROS MEDICOS S.A. | Instituto de Salud Publica de Chile |
| EPI_ISL_445358 | MEGASALUD SPA. | Instituto de Salud Publica de Chile |
| EPI_ISL_445359 | HOSP. SANTIAGO ORIENTE DR. LUIS TISNE B. | Instituto de Salud Publica de Chile |
| EPI_ISL_445360 | HOSPITAL DEL PROFESOR | Instituto de Salud Publica de Chile |
| EPI_ISL_445361 | CLINICA UC SAN CARLOS DE APOQUINDO | Instituto de Salud Publica de Chile |
| EPI_ISL_445362 | BUPA SERVICIOS CLINICOS S.A | Instituto de Salud Publica de Chile |

| EPI_Accession_ID | Originating Laboratory | Submitting Laboratory |
|---|---|---|
| EPI_ISL_445363 | ASISTENCIA PUBLICA DR.ALEJANDRO DEL RIO | Instituto de Salud Publica de Chile |
| EPI_ISL_445364 | HOSPITAL EL CARMEN DR.LUIS VALENTIN F. | Instituto de Salud Publica de Chile |
| EPI_ISL_445365 | HOSPITAL DR.SOTERO DEL RIO | Instituto de Salud Publica de Chile |
| EPI_ISL_445366 | HOSPITAL DR.SOTERO DEL RIO | Instituto de Salud Publica de Chile |
| EPI_ISL_445367 | ASISTENCIA PUBLICA DR.ALEJANDRO DEL RIO | Instituto de Salud Publica de Chile |
| EPI_ISL_445368 | HOSPITAL DEL PROFESOR | Instituto de Salud Publica de Chile |
| EPI_ISL_445369 | HOSPITAL DE CARABINEROS | Instituto de Salud Publica de Chile |
| EPI_ISL_445370 | HOSPITAL DE CARABINEROS | Instituto de Salud Publica de Chile |
| EPI_ISL_445371 | HOSPITAL DR.SOTERO DEL RIO | Instituto de Salud Publica de Chile |
| EPI_ISL_445372 | HOSPITAL FF.AA. "CIRUJANO C. GUZMAN | Instituto de Salud Publica de Chile |
| EPI_ISL_445373 | HOSPITAL SAN JUAN DE DIOS | Instituto de Salud Publica de Chile |
| EPI_ISL_445374 | HOSPITAL SAN JUAN DE DIOS | Instituto de Salud Publica de Chile |
| EPI_ISL_445375 | HOSPITAL SAN JUAN DE DIOS | Instituto de Salud Publica de Chile |
| EPI_ISL_445376 | HOSPITAL SAN JUAN DE DIOS | Instituto de Salud Publica de Chile |
| EPI_ISL_445377 | HOSPITAL SAN JUAN DE DIOS | Instituto de Salud Publica de Chile |
| EPI_ISL_445378 | HOSPITAL DE BULNES | Instituto de Salud Publica de Chile |
| EPI_ISL_445379 | IMALAB- HOSPITAL FACH | Instituto de Salud Publica de Chile |
| EPI_ISL_445380 | Ramathibodi Hospital | COVID-19 Network Investigations (CONI) Alliance |
| EPI_ISL_445381 | Wales Specialist Virology Centre | Public Health Wales Microbiology Cardiff |
| EPI_ISL_445382 | Wales Specialist Virology Centre | Public Health Wales Microbiology Cardiff |
| EPI_ISL_445383 | Wales Specialist Virology Centre | Public Health Wales Microbiology Cardiff |
| EPI_ISL_445384 | Wales Specialist Virology Centre | Public Health Wales Microbiology Cardiff |
| EPI_ISL_445385 | Wales Specialist Virology Centre | Public Health Wales Microbiology Cardiff |
| EPI_ISL_445386 | Wales Specialist Virology Centre | Public Health Wales Microbiology Cardiff |
| EPI_ISL_445387 | Wales Specialist Virology Centre | Public Health Wales Microbiology Cardiff |
| EPI_ISL_445388 | Wales Specialist Virology Centre | Public Health Wales Microbiology Cardiff |
| EPI_ISL_445389 | Wales Specialist Virology Centre | Public Health Wales Microbiology Cardiff |
| EPI_ISL_445390 | Wales Specialist Virology Centre | Public Health Wales Microbiology Cardiff |
| EPI_ISL_445391 | Wales Specialist Virology Centre | Public Health Wales Microbiology Cardiff |
| EPI_ISL_445392 | Wales Specialist Virology Centre | Public Health Wales Microbiology Cardiff |
| EPI_ISL_445393 | Wales Specialist Virology Centre | Public Health Wales Microbiology Cardiff |
| EPI_ISL_445394 | Wales Specialist Virology Centre | Public Health Wales Microbiology Cardiff |
| EPI_ISL_445395 | Wales Specialist Virology Centre | Public Health Wales Microbiology Cardiff |

| EPI_Accession_ID | Originating Laboratory | Submitting Laboratory |
|---|---|---|
| EPI_ISL_445396 | Wales Specialist Virology Centre | Public Health Wales Microbiology Cardiff |
| EPI_ISL_445397 | Wales Specialist Virology Centre | Public Health Wales Microbiology Cardiff |
| EPI_ISL_445398 | Wales Specialist Virology Centre | Public Health Wales Microbiology Cardiff |
| EPI_ISL_445399 | Wales Specialist Virology Centre | Public Health Wales Microbiology Cardiff |
| EPI_ISL_445400 | Wales Specialist Virology Centre | Public Health Wales Microbiology Cardiff |
| EPI_ISL_445401 | Wales Specialist Virology Centre | Public Health Wales Microbiology Cardiff |
| EPI_ISL_445402 | Wales Specialist Virology Centre | Public Health Wales Microbiology Cardiff |
| EPI_ISL_445403 | Wales Specialist Virology Centre | Public Health Wales Microbiology Cardiff |
| EPI_ISL_445404 | Wales Specialist Virology Centre | Public Health Wales Microbiology Cardiff |
| EPI_ISL_445405 | Wales Specialist Virology Centre | Public Health Wales Microbiology Cardiff |
| EPI_ISL_445406 | Wales Specialist Virology Centre | Public Health Wales Microbiology Cardiff |
| EPI_ISL_445407 | Wales Specialist Virology Centre | Public Health Wales Microbiology Cardiff |
| EPI_ISL_445408 | Wales Specialist Virology Centre | Public Health Wales Microbiology Cardiff |
| EPI_ISL_445409 | Wales Specialist Virology Centre | Public Health Wales Microbiology Cardiff |
| EPI_ISL_445410 | Wales Specialist Virology Centre | Public Health Wales Microbiology Cardiff |
| EPI_ISL_445411 | Wales Specialist Virology Centre | Public Health Wales Microbiology Cardiff |
| EPI_ISL_445412 | Wales Specialist Virology Centre | Public Health Wales Microbiology Cardiff |
| EPI_ISL_445413 | Wales Specialist Virology Centre | Public Health Wales Microbiology Cardiff |
| EPI_ISL_445414 | Wales Specialist Virology Centre | Public Health Wales Microbiology Cardiff |
| EPI_ISL_445415 | Wales Specialist Virology Centre | Public Health Wales Microbiology Cardiff |
| EPI_ISL_445416 | Wales Specialist Virology Centre | Public Health Wales Microbiology Cardiff |
| EPI_ISL_445417 | Wales Specialist Virology Centre | Public Health Wales Microbiology Cardiff |
| EPI_ISL_445418 | Wales Specialist Virology Centre | Public Health Wales Microbiology Cardiff |
| EPI_ISL_445419 | Wales Specialist Virology Centre | Public Health Wales Microbiology Cardiff |
| EPI_ISL_445420 | Wales Specialist Virology Centre | Public Health Wales Microbiology Cardiff |
| EPI_ISL_445421 | Wales Specialist Virology Centre | Public Health Wales Microbiology Cardiff |
| EPI_ISL_445422 | Wales Specialist Virology Centre | Public Health Wales Microbiology Cardiff |
| EPI_ISL_445423 | Wales Specialist Virology Centre | Public Health Wales Microbiology Cardiff |
| EPI_ISL_445424 | Wales Specialist Virology Centre | Public Health Wales Microbiology Cardiff |
| EPI_ISL_445425 | Wales Specialist Virology Centre | Public Health Wales Microbiology Cardiff |
| EPI_ISL_445426 | Wales Specialist Virology Centre | Public Health Wales Microbiology Cardiff |
| EPI_ISL_445427 | Wales Specialist Virology Centre | Public Health Wales Microbiology Cardiff |
| EPI_ISL_445428 | Wales Specialist Virology Centre | Public Health Wales Microbiology Cardiff |

| EPI_Accession_ID | Originating Laboratory | Submitting Laboratory |
|---|---|---|
| EPI_ISL_445429 | Wales Specialist Virology Centre | Public Health Wales Microbiology Cardiff |
| EPI_ISL_445430 | Wales Specialist Virology Centre | Public Health Wales Microbiology Cardiff |
| EPI_ISL_445431 | Wales Specialist Virology Centre | Public Health Wales Microbiology Cardiff |
| EPI_ISL_445432 | Wales Specialist Virology Centre | Public Health Wales Microbiology Cardiff |
| EPI_ISL_445433 | Wales Specialist Virology Centre | Public Health Wales Microbiology Cardiff |
| EPI_ISL_445434 | Wales Specialist Virology Centre | Public Health Wales Microbiology Cardiff |
| EPI_ISL_445435 | Wales Specialist Virology Centre | Public Health Wales Microbiology Cardiff |
| EPI_ISL_445436 | Wales Specialist Virology Centre | Public Health Wales Microbiology Cardiff |
| EPI_ISL_445437 | Wales Specialist Virology Centre | Public Health Wales Microbiology Cardiff |
| EPI_ISL_445438 | Wales Specialist Virology Centre | Public Health Wales Microbiology Cardiff |
| EPI_ISL_445439 | Wales Specialist Virology Centre | Public Health Wales Microbiology Cardiff |
| EPI_ISL_445440 | Wales Specialist Virology Centre | Public Health Wales Microbiology Cardiff |
| EPI_ISL_445441 | Wales Specialist Virology Centre | Public Health Wales Microbiology Cardiff |
| EPI_ISL_445442 | Wales Specialist Virology Centre | Public Health Wales Microbiology Cardiff |
| EPI_ISL_445443 | Wales Specialist Virology Centre | Public Health Wales Microbiology Cardiff |
| EPI_ISL_445444 | Wales Specialist Virology Centre | Public Health Wales Microbiology Cardiff |
| EPI_ISL_445445 | Wales Specialist Virology Centre | Public Health Wales Microbiology Cardiff |
| EPI_ISL_445446 | Wales Specialist Virology Centre | Public Health Wales Microbiology Cardiff |
| EPI_ISL_445447 | Wales Specialist Virology Centre | Public Health Wales Microbiology Cardiff |
| EPI_ISL_445448 | Wales Specialist Virology Centre | Public Health Wales Microbiology Cardiff |
| EPI_ISL_445449 | Wales Specialist Virology Centre | Public Health Wales Microbiology Cardiff |
| EPI_ISL_445450 | Wales Specialist Virology Centre | Public Health Wales Microbiology Cardiff |
| EPI_ISL_445451 | Wales Specialist Virology Centre | Public Health Wales Microbiology Cardiff |
| EPI_ISL_445452 | Wales Specialist Virology Centre | Public Health Wales Microbiology Cardiff |
| EPI_ISL_445453 | Wales Specialist Virology Centre | Public Health Wales Microbiology Cardiff |
| EPI_ISL_445454 | Wales Specialist Virology Centre | Public Health Wales Microbiology Cardiff |
| EPI_ISL_445455 | Wales Specialist Virology Centre | Public Health Wales Microbiology Cardiff |
| EPI_ISL_445456 | Wales Specialist Virology Centre | Public Health Wales Microbiology Cardiff |
| EPI_ISL_445457 | Wales Specialist Virology Centre | Public Health Wales Microbiology Cardiff |
| EPI_ISL_445458 | Wales Specialist Virology Centre | Public Health Wales Microbiology Cardiff |
| EPI_ISL_445459 | Wales Specialist Virology Centre | Public Health Wales Microbiology Cardiff |
| EPI_ISL_445460 | Wales Specialist Virology Centre | Public Health Wales Microbiology Cardiff |
| EPI_ISL_445461 | Wales Specialist Virology Centre | Public Health Wales Microbiology Cardiff |

| EPI_Accession_ID | Originating Laboratory | Submitting Laboratory |
|---|---|---|
| EPI_ISL_445462 | Wales Specialist Virology Centre | Public Health Wales Microbiology Cardiff |
| EPI_ISL_445463 | Wales Specialist Virology Centre | Public Health Wales Microbiology Cardiff |
| EPI_ISL_445464 | Wales Specialist Virology Centre | Public Health Wales Microbiology Cardiff |
| EPI_ISL_445465 | Wales Specialist Virology Centre | Public Health Wales Microbiology Cardiff |
| EPI_ISL_445466 | Wales Specialist Virology Centre | Public Health Wales Microbiology Cardiff |
| EPI_ISL_445467 | Wales Specialist Virology Centre | Public Health Wales Microbiology Cardiff |
| EPI_ISL_445468 | Wales Specialist Virology Centre | Public Health Wales Microbiology Cardiff |
| EPI_ISL_445469 | Wales Specialist Virology Centre | Public Health Wales Microbiology Cardiff |
| EPI_ISL_445470 | Wales Specialist Virology Centre | Public Health Wales Microbiology Cardiff |
| EPI_ISL_445471 | Wales Specialist Virology Centre | Public Health Wales Microbiology Cardiff |
| EPI_ISL_445472 | Wales Specialist Virology Centre | Public Health Wales Microbiology Cardiff |
| EPI_ISL_445473 | Wales Specialist Virology Centre | Public Health Wales Microbiology Cardiff |
| EPI_ISL_445474 | Wales Specialist Virology Centre | Public Health Wales Microbiology Cardiff |
| EPI_ISL_445475 | Wales Specialist Virology Centre | Public Health Wales Microbiology Cardiff |
| EPI_ISL_445476 | Wales Specialist Virology Centre | Public Health Wales Microbiology Cardiff |
| EPI_ISL_445477 | Wales Specialist Virology Centre | Public Health Wales Microbiology Cardiff |
| EPI_ISL_445478 | Wales Specialist Virology Centre | Public Health Wales Microbiology Cardiff |
| EPI_ISL_445479 | Wales Specialist Virology Centre | Public Health Wales Microbiology Cardiff |
| EPI_ISL_445480 | Wales Specialist Virology Centre | Public Health Wales Microbiology Cardiff |
| EPI_ISL_445481 | Wales Specialist Virology Centre | Public Health Wales Microbiology Cardiff |
| EPI_ISL_445482 | Wales Specialist Virology Centre | Public Health Wales Microbiology Cardiff |
| EPI_ISL_445483 | Wales Specialist Virology Centre | Public Health Wales Microbiology Cardiff |
| EPI_ISL_445484 | Wales Specialist Virology Centre | Public Health Wales Microbiology Cardiff |
| EPI_ISL_445485 | Wales Specialist Virology Centre | Public Health Wales Microbiology Cardiff |
| EPI_ISL_445486 | Wales Specialist Virology Centre | Public Health Wales Microbiology Cardiff |
| EPI_ISL_445487 | Wales Specialist Virology Centre | Public Health Wales Microbiology Cardiff |
| EPI_ISL_445488 | Wales Specialist Virology Centre | Public Health Wales Microbiology Cardiff |
| EPI_ISL_445489 | Wales Specialist Virology Centre | Public Health Wales Microbiology Cardiff |
| EPI_ISL_445490 | Wales Specialist Virology Centre | Public Health Wales Microbiology Cardiff |
| EPI_ISL_445491 | Wales Specialist Virology Centre | Public Health Wales Microbiology Cardiff |
| EPI_ISL_445492 | Wales Specialist Virology Centre | Public Health Wales Microbiology Cardiff |
| EPI_ISL_445493 | Wales Specialist Virology Centre | Public Health Wales Microbiology Cardiff |
| EPI_ISL_445494 | Wales Specialist Virology Centre | Public Health Wales Microbiology Cardiff |

| EPI_Accession_ID | Originating Laboratory | Submitting Laboratory |
|---|---|---|
| EPI_ISL_445495 | Wales Specialist Virology Centre | Public Health Wales Microbiology Cardiff |
| EPI_ISL_445496 | Wales Specialist Virology Centre | Public Health Wales Microbiology Cardiff |
| EPI_ISL_445497 | Wales Specialist Virology Centre | Public Health Wales Microbiology Cardiff |
| EPI_ISL_445498 | Wales Specialist Virology Centre | Public Health Wales Microbiology Cardiff |
| EPI_ISL_445499 | Wales Specialist Virology Centre | Public Health Wales Microbiology Cardiff |
| EPI_ISL_445500 | Wales Specialist Virology Centre | Public Health Wales Microbiology Cardiff |
| EPI_ISL_445501 | Wales Specialist Virology Centre | Public Health Wales Microbiology Cardiff |
| EPI_ISL_445502 | Wales Specialist Virology Centre | Public Health Wales Microbiology Cardiff |
| EPI_ISL_445503 | Wales Specialist Virology Centre | Public Health Wales Microbiology Cardiff |
| EPI_ISL_445504 | Wales Specialist Virology Centre | Public Health Wales Microbiology Cardiff |
| EPI_ISL_445505 | Wales Specialist Virology Centre | Public Health Wales Microbiology Cardiff |
| EPI_ISL_445506 | Wales Specialist Virology Centre | Public Health Wales Microbiology Cardiff |
| EPI_ISL_445507 | Wales Specialist Virology Centre | Public Health Wales Microbiology Cardiff |
| EPI_ISL_445508 | Wales Specialist Virology Centre | Public Health Wales Microbiology Cardiff |
| EPI_ISL_445509 | Wales Specialist Virology Centre | Public Health Wales Microbiology Cardiff |
| EPI_ISL_445510 | Wales Specialist Virology Centre | Public Health Wales Microbiology Cardiff |
| EPI_ISL_445511 | Wales Specialist Virology Centre | Public Health Wales Microbiology Cardiff |
| EPI_ISL_445512 | Wales Specialist Virology Centre | Public Health Wales Microbiology Cardiff |
| EPI_ISL_445513 | Wales Specialist Virology Centre | Public Health Wales Microbiology Cardiff |
| EPI_ISL_445514 | Wales Specialist Virology Centre | Public Health Wales Microbiology Cardiff |
| EPI_ISL_445515 | Wales Specialist Virology Centre | Public Health Wales Microbiology Cardiff |
| EPI_ISL_445516 | Wales Specialist Virology Centre | Public Health Wales Microbiology Cardiff |
| EPI_ISL_445517 | Wales Specialist Virology Centre | Public Health Wales Microbiology Cardiff |
| EPI_ISL_445518 | Wales Specialist Virology Centre | Public Health Wales Microbiology Cardiff |
| EPI_ISL_445519 | Wales Specialist Virology Centre | Public Health Wales Microbiology Cardiff |
| EPI_ISL_445520 | Wales Specialist Virology Centre | Public Health Wales Microbiology Cardiff |
| EPI_ISL_445521 | Wales Specialist Virology Centre | Public Health Wales Microbiology Cardiff |
| EPI_ISL_445522 | Wales Specialist Virology Centre | Public Health Wales Microbiology Cardiff |
| EPI_ISL_445523 | Wales Specialist Virology Centre | Public Health Wales Microbiology Cardiff |
| EPI_ISL_445524 | Wales Specialist Virology Centre | Public Health Wales Microbiology Cardiff |
| EPI_ISL_445525 | Wales Specialist Virology Centre | Public Health Wales Microbiology Cardiff |
| EPI_ISL_445526 | Wales Specialist Virology Centre | Public Health Wales Microbiology Cardiff |
| EPI_ISL_445527 | Wales Specialist Virology Centre | Public Health Wales Microbiology Cardiff |

| EPI_Accession_ID | Originating Laboratory | Submitting Laboratory |
|---|---|---|
| EPI_ISL_445528 | Wales Specialist Virology Centre | Public Health Wales Microbiology Cardiff |
| EPI_ISL_445529 | Wales Specialist Virology Centre | Public Health Wales Microbiology Cardiff |
| EPI_ISL_445530 | Wales Specialist Virology Centre | Public Health Wales Microbiology Cardiff |
| EPI_ISL_445531 | Wales Specialist Virology Centre | Public Health Wales Microbiology Cardiff |
| EPI_ISL_445532 | Wales Specialist Virology Centre | Public Health Wales Microbiology Cardiff |
| EPI_ISL_445533 | Wales Specialist Virology Centre | Public Health Wales Microbiology Cardiff |
| EPI_ISL_445534 | Wales Specialist Virology Centre | Public Health Wales Microbiology Cardiff |
| EPI_ISL_445535 | Wales Specialist Virology Centre | Public Health Wales Microbiology Cardiff |
| EPI_ISL_445536 | Wales Specialist Virology Centre | Public Health Wales Microbiology Cardiff |
| EPI_ISL_445537 | Wales Specialist Virology Centre | Public Health Wales Microbiology Cardiff |
| EPI_ISL_445538 | Wales Specialist Virology Centre | Public Health Wales Microbiology Cardiff |
| EPI_ISL_445539 | Wales Specialist Virology Centre | Public Health Wales Microbiology Cardiff |
| EPI_ISL_445540 | Wales Specialist Virology Centre | Public Health Wales Microbiology Cardiff |
| EPI_ISL_445541 | Wales Specialist Virology Centre | Public Health Wales Microbiology Cardiff |
| EPI_ISL_445542 | Wales Specialist Virology Centre | Public Health Wales Microbiology Cardiff |
| EPI_ISL_445543 | Wales Specialist Virology Centre | Public Health Wales Microbiology Cardiff |
| EPI_ISL_445544 | Wales Specialist Virology Centre | Public Health Wales Microbiology Cardiff |
| EPI_ISL_445545 | Wales Specialist Virology Centre | Public Health Wales Microbiology Cardiff |
| EPI_ISL_445546 | Wales Specialist Virology Centre | Public Health Wales Microbiology Cardiff |
| EPI_ISL_445547 | Wales Specialist Virology Centre | Public Health Wales Microbiology Cardiff |
| EPI_ISL_445548 | Wales Specialist Virology Centre | Public Health Wales Microbiology Cardiff |
| EPI_ISL_445549 | Wales Specialist Virology Centre | Public Health Wales Microbiology Cardiff |
| EPI_ISL_445550 | Wales Specialist Virology Centre | Public Health Wales Microbiology Cardiff |
| EPI_ISL_445551 | Wales Specialist Virology Centre | Public Health Wales Microbiology Cardiff |
| EPI_ISL_445552 | Wales Specialist Virology Centre | Public Health Wales Microbiology Cardiff |
| EPI_ISL_445553 | Wales Specialist Virology Centre | Public Health Wales Microbiology Cardiff |
| EPI_ISL_445554 | Wales Specialist Virology Centre | Public Health Wales Microbiology Cardiff |
| EPI_ISL_445555 | Wales Specialist Virology Centre | Public Health Wales Microbiology Cardiff |
| EPI_ISL_445556 | Wales Specialist Virology Centre | Public Health Wales Microbiology Cardiff |
| EPI_ISL_445557 | Wales Specialist Virology Centre | Public Health Wales Microbiology Cardiff |
| EPI_ISL_445558 | Wales Specialist Virology Centre | Public Health Wales Microbiology Cardiff |
| EPI_ISL_445559 | Wales Specialist Virology Centre | Public Health Wales Microbiology Cardiff |
| EPI_ISL_445560 | Wales Specialist Virology Centre | Public Health Wales Microbiology Cardiff |

| EPI_Accession_ID | Originating Laboratory | Submitting Laboratory |
|---|---|---|
| EPI_ISL_445561 | Wales Specialist Virology Centre | Public Health Wales Microbiology Cardiff |
| EPI_ISL_445562 | Wales Specialist Virology Centre | Public Health Wales Microbiology Cardiff |
| EPI_ISL_445563 | Wales Specialist Virology Centre | Public Health Wales Microbiology Cardiff |
| EPI_ISL_445564 | Wales Specialist Virology Centre | Public Health Wales Microbiology Cardiff |
| EPI_ISL_445565 | Wales Specialist Virology Centre | Public Health Wales Microbiology Cardiff |
| EPI_ISL_445566 | Wales Specialist Virology Centre | Public Health Wales Microbiology Cardiff |
| EPI_ISL_445567 | Wales Specialist Virology Centre | Public Health Wales Microbiology Cardiff |
| EPI_ISL_445568 | Wales Specialist Virology Centre | Public Health Wales Microbiology Cardiff |
| EPI_ISL_445569 | Wales Specialist Virology Centre | Public Health Wales Microbiology Cardiff |
| EPI_ISL_445570 | Wales Specialist Virology Centre | Public Health Wales Microbiology Cardiff |
| EPI_ISL_445571 | Wales Specialist Virology Centre | Public Health Wales Microbiology Cardiff |
| EPI_ISL_445572 | Wales Specialist Virology Centre | Public Health Wales Microbiology Cardiff |
| EPI_ISL_445573 | Wales Specialist Virology Centre | Public Health Wales Microbiology Cardiff |
| EPI_ISL_445574 | Wales Specialist Virology Centre | Public Health Wales Microbiology Cardiff |
| EPI_ISL_445575 | Wales Specialist Virology Centre | Public Health Wales Microbiology Cardiff |
| EPI_ISL_445576 | Wales Specialist Virology Centre | Public Health Wales Microbiology Cardiff |
| EPI_ISL_445577 | Wales Specialist Virology Centre | Public Health Wales Microbiology Cardiff |
| EPI_ISL_445578 | Wales Specialist Virology Centre | Public Health Wales Microbiology Cardiff |
| EPI_ISL_445579 | Wales Specialist Virology Centre | Public Health Wales Microbiology Cardiff |
| EPI_ISL_445580 | Wales Specialist Virology Centre | Public Health Wales Microbiology Cardiff |
| EPI_ISL_445581 | Wales Specialist Virology Centre | Public Health Wales Microbiology Cardiff |
| EPI_ISL_445582 | Wales Specialist Virology Centre | Public Health Wales Microbiology Cardiff |
| EPI_ISL_445583 | Wales Specialist Virology Centre | Public Health Wales Microbiology Cardiff |
| EPI_ISL_445584 | Wales Specialist Virology Centre | Public Health Wales Microbiology Cardiff |
| EPI_ISL_445585 | Wales Specialist Virology Centre | Public Health Wales Microbiology Cardiff |
| EPI_ISL_445586 | Wales Specialist Virology Centre | Public Health Wales Microbiology Cardiff |
| EPI_ISL_445587 | Wales Specialist Virology Centre | Public Health Wales Microbiology Cardiff |
| EPI_ISL_445588 | Wales Specialist Virology Centre | Public Health Wales Microbiology Cardiff |
| EPI_ISL_445589 | Wales Specialist Virology Centre | Public Health Wales Microbiology Cardiff |
| EPI_ISL_445590 | Wales Specialist Virology Centre | Public Health Wales Microbiology Cardiff |
| EPI_ISL_445591 | Wales Specialist Virology Centre | Public Health Wales Microbiology Cardiff |
| EPI_ISL_445592 | Wales Specialist Virology Centre | Public Health Wales Microbiology Cardiff |
| EPI_ISL_445593 | Wales Specialist Virology Centre | Public Health Wales Microbiology Cardiff |

| EPI_Accession_ID | Originating Laboratory | Submitting Laboratory |
|---|---|---|
| EPI_ISL_445594 | Wales Specialist Virology Centre | Public Health Wales Microbiology Cardiff |
| EPI_ISL_445595 | Wales Specialist Virology Centre | Public Health Wales Microbiology Cardiff |
| EPI_ISL_445596 | Wales Specialist Virology Centre | Public Health Wales Microbiology Cardiff |
| EPI_ISL_445597 | Wales Specialist Virology Centre | Public Health Wales Microbiology Cardiff |
| EPI_ISL_445598 | Wales Specialist Virology Centre | Public Health Wales Microbiology Cardiff |
| EPI_ISL_445599 | Wales Specialist Virology Centre | Public Health Wales Microbiology Cardiff |
| EPI_ISL_445600 | Wales Specialist Virology Centre | Public Health Wales Microbiology Cardiff |
| EPI_ISL_445601 | Wales Specialist Virology Centre | Public Health Wales Microbiology Cardiff |
| EPI_ISL_445602 | Wales Specialist Virology Centre | Public Health Wales Microbiology Cardiff |
| EPI_ISL_445603 | Wales Specialist Virology Centre | Public Health Wales Microbiology Cardiff |
| EPI_ISL_445604 | Wales Specialist Virology Centre | Public Health Wales Microbiology Cardiff |
| EPI_ISL_445605 | Wales Specialist Virology Centre | Public Health Wales Microbiology Cardiff |
| EPI_ISL_445606 | Wales Specialist Virology Centre | Public Health Wales Microbiology Cardiff |
| EPI_ISL_445607 | Wales Specialist Virology Centre | Public Health Wales Microbiology Cardiff |
| EPI_ISL_445608 | Wales Specialist Virology Centre | Public Health Wales Microbiology Cardiff |
| EPI_ISL_445609 | Wales Specialist Virology Centre | Public Health Wales Microbiology Cardiff |
| EPI_ISL_445610 | Wales Specialist Virology Centre | Public Health Wales Microbiology Cardiff |
| EPI_ISL_445611 | Wales Specialist Virology Centre | Public Health Wales Microbiology Cardiff |
| EPI_ISL_445612 | Wales Specialist Virology Centre | Public Health Wales Microbiology Cardiff |
| EPI_ISL_445613 | Wales Specialist Virology Centre | Public Health Wales Microbiology Cardiff |
| EPI_ISL_445614 | Wales Specialist Virology Centre | Public Health Wales Microbiology Cardiff |
| EPI_ISL_445615 | Wales Specialist Virology Centre | Public Health Wales Microbiology Cardiff |
| EPI_ISL_445616 | Wales Specialist Virology Centre | Public Health Wales Microbiology Cardiff |
| EPI_ISL_445617 | Wales Specialist Virology Centre | Public Health Wales Microbiology Cardiff |
| EPI_ISL_445618 | Wales Specialist Virology Centre | Public Health Wales Microbiology Cardiff |
| EPI_ISL_445619 | Wales Specialist Virology Centre | Public Health Wales Microbiology Cardiff |
| EPI_ISL_445620 | Wales Specialist Virology Centre | Public Health Wales Microbiology Cardiff |
| EPI_ISL_445621 | Wales Specialist Virology Centre | Public Health Wales Microbiology Cardiff |
| EPI_ISL_445622 | Wales Specialist Virology Centre | Public Health Wales Microbiology Cardiff |
| EPI_ISL_445623 | Wales Specialist Virology Centre | Public Health Wales Microbiology Cardiff |
| EPI_ISL_445624 | Wales Specialist Virology Centre | Public Health Wales Microbiology Cardiff |
| EPI_ISL_445625 | Wales Specialist Virology Centre | Public Health Wales Microbiology Cardiff |
| EPI_ISL_445626 | Wales Specialist Virology Centre | Public Health Wales Microbiology Cardiff |

| EPI_Accession_ID | Originating Laboratory | Submitting Laboratory |
|---|---|---|
| EPI_ISL_445627 | Wales Specialist Virology Centre | Public Health Wales Microbiology Cardiff |
| EPI_ISL_445628 | Wales Specialist Virology Centre | Public Health Wales Microbiology Cardiff |
| EPI_ISL_445629 | Wales Specialist Virology Centre | Public Health Wales Microbiology Cardiff |
| EPI_ISL_445630 | Wales Specialist Virology Centre | Public Health Wales Microbiology Cardiff |
| EPI_ISL_445631 | Wales Specialist Virology Centre | Public Health Wales Microbiology Cardiff |
| EPI_ISL_445632 | Wales Specialist Virology Centre | Public Health Wales Microbiology Cardiff |
| EPI_ISL_445633 | Wales Specialist Virology Centre | Public Health Wales Microbiology Cardiff |
| EPI_ISL_445634 | Wales Specialist Virology Centre | Public Health Wales Microbiology Cardiff |
| EPI_ISL_445635 | Wales Specialist Virology Centre | Public Health Wales Microbiology Cardiff |
| EPI_ISL_445636 | Wales Specialist Virology Centre | Public Health Wales Microbiology Cardiff |
| EPI_ISL_445637 | Wales Specialist Virology Centre | Public Health Wales Microbiology Cardiff |
| EPI_ISL_445638 | Wales Specialist Virology Centre | Public Health Wales Microbiology Cardiff |
| EPI_ISL_445639 | Wales Specialist Virology Centre | Public Health Wales Microbiology Cardiff |
| EPI_ISL_445640 | Wales Specialist Virology Centre | Public Health Wales Microbiology Cardiff |
| EPI_ISL_445641 | Wales Specialist Virology Centre | Public Health Wales Microbiology Cardiff |
| EPI_ISL_445642 | Wales Specialist Virology Centre | Public Health Wales Microbiology Cardiff |
| EPI_ISL_445643 | Wales Specialist Virology Centre | Public Health Wales Microbiology Cardiff |
| EPI_ISL_445644 | Wales Specialist Virology Centre | Public Health Wales Microbiology Cardiff |
| EPI_ISL_445645 | Wales Specialist Virology Centre | Public Health Wales Microbiology Cardiff |
| EPI_ISL_445646 | Wales Specialist Virology Centre | Public Health Wales Microbiology Cardiff |
| EPI_ISL_445647 | Wales Specialist Virology Centre | Public Health Wales Microbiology Cardiff |
| EPI_ISL_445648 | Wales Specialist Virology Centre | Public Health Wales Microbiology Cardiff |
| EPI_ISL_445649 | Wales Specialist Virology Centre | Public Health Wales Microbiology Cardiff |
| EPI_ISL_445650 | Wales Specialist Virology Centre | Public Health Wales Microbiology Cardiff |
| EPI_ISL_445651 | Wales Specialist Virology Centre | Public Health Wales Microbiology Cardiff |
| EPI_ISL_445652 | Wales Specialist Virology Centre | Public Health Wales Microbiology Cardiff |
| EPI_ISL_445653 | Wales Specialist Virology Centre | Public Health Wales Microbiology Cardiff |
| EPI_ISL_445654 | Wales Specialist Virology Centre | Public Health Wales Microbiology Cardiff |
| EPI_ISL_445655 | Wales Specialist Virology Centre | Public Health Wales Microbiology Cardiff |
| EPI_ISL_445656 | Wales Specialist Virology Centre | Public Health Wales Microbiology Cardiff |
| EPI_ISL_445657 | Wales Specialist Virology Centre | Public Health Wales Microbiology Cardiff |
| EPI_ISL_445658 | Wales Specialist Virology Centre | Public Health Wales Microbiology Cardiff |
| EPI_ISL_445659 | Wales Specialist Virology Centre | Public Health Wales Microbiology Cardiff |

| EPI_Accession_ID | Originating Laboratory | Submitting Laboratory |
|---|---|---|
| EPI_ISL_445660 | Wales Specialist Virology Centre | Public Health Wales Microbiology Cardiff |
| EPI_ISL_445661 | Wales Specialist Virology Centre | Public Health Wales Microbiology Cardiff |
| EPI_ISL_445662 | Wales Specialist Virology Centre | Public Health Wales Microbiology Cardiff |
| EPI_ISL_445663 | Wales Specialist Virology Centre | Public Health Wales Microbiology Cardiff |
| EPI_ISL_445664 | Wales Specialist Virology Centre | Public Health Wales Microbiology Cardiff |
| EPI_ISL_445665 | Wales Specialist Virology Centre | Public Health Wales Microbiology Cardiff |
| EPI_ISL_445666 | Wales Specialist Virology Centre | Public Health Wales Microbiology Cardiff |
| EPI_ISL_445667 | Wales Specialist Virology Centre | Public Health Wales Microbiology Cardiff |
| EPI_ISL_445668 | Wales Specialist Virology Centre | Public Health Wales Microbiology Cardiff |
| EPI_ISL_445669 | Wales Specialist Virology Centre | Public Health Wales Microbiology Cardiff |
| EPI_ISL_445670 | Wales Specialist Virology Centre | Public Health Wales Microbiology Cardiff |
| EPI_ISL_445671 | Wales Specialist Virology Centre | Public Health Wales Microbiology Cardiff |
| EPI_ISL_445672 | Wales Specialist Virology Centre | Public Health Wales Microbiology Cardiff |
| EPI_ISL_445673 | Wales Specialist Virology Centre | Public Health Wales Microbiology Cardiff |
| EPI_ISL_445674 | Wales Specialist Virology Centre | Public Health Wales Microbiology Cardiff |
| EPI_ISL_445675 | Wales Specialist Virology Centre | Public Health Wales Microbiology Cardiff |
| EPI_ISL_445676 | Wales Specialist Virology Centre | Public Health Wales Microbiology Cardiff |
| EPI_ISL_445677 | Wales Specialist Virology Centre | Public Health Wales Microbiology Cardiff |
| EPI_ISL_445678 | Wales Specialist Virology Centre | Public Health Wales Microbiology Cardiff |
| EPI_ISL_445679 | Wales Specialist Virology Centre | Public Health Wales Microbiology Cardiff |
| EPI_ISL_445680 | Wales Specialist Virology Centre | Public Health Wales Microbiology Cardiff |
| EPI_ISL_445681 | Wales Specialist Virology Centre | Public Health Wales Microbiology Cardiff |
| EPI_ISL_445682 | Wales Specialist Virology Centre | Public Health Wales Microbiology Cardiff |
| EPI_ISL_445683 | Wales Specialist Virology Centre | Public Health Wales Microbiology Cardiff |
| EPI_ISL_445684 | Wales Specialist Virology Centre | Public Health Wales Microbiology Cardiff |
| EPI_ISL_445685 | Wales Specialist Virology Centre | Public Health Wales Microbiology Cardiff |
| EPI_ISL_445686 | Wales Specialist Virology Centre | Public Health Wales Microbiology Cardiff |
| EPI_ISL_445687 | Wales Specialist Virology Centre | Public Health Wales Microbiology Cardiff |
| EPI_ISL_445688 | Wales Specialist Virology Centre | Public Health Wales Microbiology Cardiff |
| EPI_ISL_445689 | Wales Specialist Virology Centre | Public Health Wales Microbiology Cardiff |
| EPI_ISL_445690 | Wales Specialist Virology Centre | Public Health Wales Microbiology Cardiff |
| EPI_ISL_445691 | Wales Specialist Virology Centre | Public Health Wales Microbiology Cardiff |
| EPI_ISL_445692 | Wales Specialist Virology Centre | Public Health Wales Microbiology Cardiff |

| EPI_Accession_ID | Originating Laboratory | Submitting Laboratory |
|---|---|---|
| EPI_ISL_445693 | Wales Specialist Virology Centre | Public Health Wales Microbiology Cardiff |
| EPI_ISL_445694 | Wales Specialist Virology Centre | Public Health Wales Microbiology Cardiff |
| EPI_ISL_445695 | Wales Specialist Virology Centre | Public Health Wales Microbiology Cardiff |
| EPI_ISL_445696 | Wales Specialist Virology Centre | Public Health Wales Microbiology Cardiff |
| EPI_ISL_445697 | Wales Specialist Virology Centre | Public Health Wales Microbiology Cardiff |
| EPI_ISL_445698 | Wales Specialist Virology Centre | Public Health Wales Microbiology Cardiff |
| EPI_ISL_445699 | Wales Specialist Virology Centre | Public Health Wales Microbiology Cardiff |
| EPI_ISL_445700 | Wales Specialist Virology Centre | Public Health Wales Microbiology Cardiff |
| EPI_ISL_445701 | Wales Specialist Virology Centre | Public Health Wales Microbiology Cardiff |
| EPI_ISL_445702 | Wales Specialist Virology Centre | Public Health Wales Microbiology Cardiff |
| EPI_ISL_445703 | Wales Specialist Virology Centre | Public Health Wales Microbiology Cardiff |
| EPI_ISL_445704 | Wales Specialist Virology Centre | Public Health Wales Microbiology Cardiff |
| EPI_ISL_445705 | Wales Specialist Virology Centre | Public Health Wales Microbiology Cardiff |
| EPI_ISL_445706 | Wales Specialist Virology Centre | Public Health Wales Microbiology Cardiff |
| EPI_ISL_445707 | Wales Specialist Virology Centre | Public Health Wales Microbiology Cardiff |
| EPI_ISL_445708 | Wales Specialist Virology Centre | Public Health Wales Microbiology Cardiff |
| EPI_ISL_445709 | Wales Specialist Virology Centre | Public Health Wales Microbiology Cardiff |
| EPI_ISL_445710 | Wales Specialist Virology Centre | Public Health Wales Microbiology Cardiff |
| EPI_ISL_445711 | Wales Specialist Virology Centre | Public Health Wales Microbiology Cardiff |
| EPI_ISL_445712 | Wales Specialist Virology Centre | Public Health Wales Microbiology Cardiff |
| EPI_ISL_445713 | Wales Specialist Virology Centre | Public Health Wales Microbiology Cardiff |
| EPI_ISL_445714 | Wales Specialist Virology Centre | Public Health Wales Microbiology Cardiff |
| EPI_ISL_445715 | Wales Specialist Virology Centre | Public Health Wales Microbiology Cardiff |
| EPI_ISL_445716 | Wales Specialist Virology Centre | Public Health Wales Microbiology Cardiff |
| EPI_ISL_445717 | Wales Specialist Virology Centre | Public Health Wales Microbiology Cardiff |
| EPI_ISL_445718 | Wales Specialist Virology Centre | Public Health Wales Microbiology Cardiff |
| EPI_ISL_445719 | Wales Specialist Virology Centre | Public Health Wales Microbiology Cardiff |
| EPI_ISL_445720 | Wales Specialist Virology Centre | Public Health Wales Microbiology Cardiff |
| EPI_ISL_445721 | Wales Specialist Virology Centre | Public Health Wales Microbiology Cardiff |
| EPI_ISL_445722 | Wales Specialist Virology Centre | Public Health Wales Microbiology Cardiff |
| EPI_ISL_445723 | Wales Specialist Virology Centre | Public Health Wales Microbiology Cardiff |
| EPI_ISL_445724 | Wales Specialist Virology Centre | Public Health Wales Microbiology Cardiff |
| EPI_ISL_445725 | Wales Specialist Virology Centre | Public Health Wales Microbiology Cardiff |

| EPI_Accession_ID | Originating Laboratory | Submitting Laboratory |
|---|---|---|
| EPI_ISL_445726 | Wales Specialist Virology Centre | Public Health Wales Microbiology Cardiff |
| EPI_ISL_445727 | Wales Specialist Virology Centre | Public Health Wales Microbiology Cardiff |
| EPI_ISL_445728 | Wales Specialist Virology Centre | Public Health Wales Microbiology Cardiff |
| EPI_ISL_445729 | Wales Specialist Virology Centre | Public Health Wales Microbiology Cardiff |
| EPI_ISL_445730 | Wales Specialist Virology Centre | Public Health Wales Microbiology Cardiff |
| EPI_ISL_445731 | Wales Specialist Virology Centre | Public Health Wales Microbiology Cardiff |
| EPI_ISL_445732 | Wales Specialist Virology Centre | Public Health Wales Microbiology Cardiff |
| EPI_ISL_445733 | Wales Specialist Virology Centre | Public Health Wales Microbiology Cardiff |
| EPI_ISL_445734 | Wales Specialist Virology Centre | Public Health Wales Microbiology Cardiff |
| EPI_ISL_445735 | Wales Specialist Virology Centre | Public Health Wales Microbiology Cardiff |
| EPI_ISL_445736 | Wales Specialist Virology Centre | Public Health Wales Microbiology Cardiff |
| EPI_ISL_445737 | Wales Specialist Virology Centre | Public Health Wales Microbiology Cardiff |
| EPI_ISL_445738 | Wales Specialist Virology Centre | Public Health Wales Microbiology Cardiff |
| EPI_ISL_445739 | Wales Specialist Virology Centre | Public Health Wales Microbiology Cardiff |
| EPI_ISL_445740 | Wales Specialist Virology Centre | Public Health Wales Microbiology Cardiff |
| EPI_ISL_445741 | Wales Specialist Virology Centre | Public Health Wales Microbiology Cardiff |
| EPI_ISL_445742 | Wales Specialist Virology Centre | Public Health Wales Microbiology Cardiff |
| EPI_ISL_445743 | Wales Specialist Virology Centre | Public Health Wales Microbiology Cardiff |
| EPI_ISL_445744 | Wales Specialist Virology Centre | Public Health Wales Microbiology Cardiff |
| EPI_ISL_445745 | Wales Specialist Virology Centre | Public Health Wales Microbiology Cardiff |
| EPI_ISL_445746 | Wales Specialist Virology Centre | Public Health Wales Microbiology Cardiff |
| EPI_ISL_445747 | Wales Specialist Virology Centre | Public Health Wales Microbiology Cardiff |
| EPI_ISL_445749 | Wales Specialist Virology Centre | Public Health Wales Microbiology Cardiff |
| EPI_ISL_445750 | Wales Specialist Virology Centre | Public Health Wales Microbiology Cardiff |
| EPI_ISL_445751 | Wales Specialist Virology Centre | Public Health Wales Microbiology Cardiff |
| EPI_ISL_445752 | Wales Specialist Virology Centre | Public Health Wales Microbiology Cardiff |
| EPI_ISL_445753 | Wales Specialist Virology Centre | Public Health Wales Microbiology Cardiff |
| EPI_ISL_445754 | Wales Specialist Virology Centre | Public Health Wales Microbiology Cardiff |
| EPI_ISL_445755 | Wales Specialist Virology Centre | Public Health Wales Microbiology Cardiff |
| EPI_ISL_445756 | Wales Specialist Virology Centre | Public Health Wales Microbiology Cardiff |
| EPI_ISL_445757 | Wales Specialist Virology Centre | Public Health Wales Microbiology Cardiff |
| EPI_ISL_445758 | Wales Specialist Virology Centre | Public Health Wales Microbiology Cardiff |
| EPI_ISL_445759 | Wales Specialist Virology Centre | Public Health Wales Microbiology Cardiff |

| EPI_Accession_ID | Originating Laboratory | Submitting Laboratory |
|---|---|---|
| EPI_ISL_445760 | Wales Specialist Virology Centre | Public Health Wales Microbiology Cardiff |
| EPI_ISL_445761 | Wales Specialist Virology Centre | Public Health Wales Microbiology Cardiff |
| EPI_ISL_445762 | Wales Specialist Virology Centre | Public Health Wales Microbiology Cardiff |
| EPI_ISL_445763 | Wales Specialist Virology Centre | Public Health Wales Microbiology Cardiff |
| EPI_ISL_445764 | Wales Specialist Virology Centre | Public Health Wales Microbiology Cardiff |
| EPI_ISL_445765 | Wales Specialist Virology Centre | Public Health Wales Microbiology Cardiff |
| EPI_ISL_445766 | Wales Specialist Virology Centre | Public Health Wales Microbiology Cardiff |
| EPI_ISL_445767 | Wales Specialist Virology Centre | Public Health Wales Microbiology Cardiff |
| EPI_ISL_445768 | Wales Specialist Virology Centre | Public Health Wales Microbiology Cardiff |
| EPI_ISL_445769 | Wales Specialist Virology Centre | Public Health Wales Microbiology Cardiff |
| EPI_ISL_445770 | Wales Specialist Virology Centre | Public Health Wales Microbiology Cardiff |
| EPI_ISL_445771 | Wales Specialist Virology Centre | Public Health Wales Microbiology Cardiff |
| EPI_ISL_445772 | Wales Specialist Virology Centre | Public Health Wales Microbiology Cardiff |
| EPI_ISL_445773 | Wales Specialist Virology Centre | Public Health Wales Microbiology Cardiff |
| EPI_ISL_445774 | Wales Specialist Virology Centre | Public Health Wales Microbiology Cardiff |
| EPI_ISL_445775 | Wales Specialist Virology Centre | Public Health Wales Microbiology Cardiff |
| EPI_ISL_445776 | Wales Specialist Virology Centre | Public Health Wales Microbiology Cardiff |
| EPI_ISL_445777 | Wales Specialist Virology Centre | Public Health Wales Microbiology Cardiff |
| EPI_ISL_445778 | Wales Specialist Virology Centre | Public Health Wales Microbiology Cardiff |
| EPI_ISL_445779 | Wales Specialist Virology Centre | Public Health Wales Microbiology Cardiff |
| EPI_ISL_445780 | Wales Specialist Virology Centre | Public Health Wales Microbiology Cardiff |
| EPI_ISL_445781 | Wales Specialist Virology Centre | Public Health Wales Microbiology Cardiff |
| EPI_ISL_445783 | Wales Specialist Virology Centre | Public Health Wales Microbiology Cardiff |
| EPI_ISL_445784 | Wales Specialist Virology Centre | Public Health Wales Microbiology Cardiff |
| EPI_ISL_445785 | Wales Specialist Virology Centre | Public Health Wales Microbiology Cardiff |
| EPI_ISL_445786 | Wales Specialist Virology Centre | Public Health Wales Microbiology Cardiff |
| EPI_ISL_445787 | Wales Specialist Virology Centre | Public Health Wales Microbiology Cardiff |
| EPI_ISL_445788 | Wales Specialist Virology Centre | Public Health Wales Microbiology Cardiff |
| EPI_ISL_445789 | Wales Specialist Virology Centre | Public Health Wales Microbiology Cardiff |
| EPI_ISL_445790 | Wales Specialist Virology Centre | Public Health Wales Microbiology Cardiff |
| EPI_ISL_445791 | Wales Specialist Virology Centre | Public Health Wales Microbiology Cardiff |
| EPI_ISL_445792 | Wales Specialist Virology Centre | Public Health Wales Microbiology Cardiff |
| EPI_ISL_445793 | Wales Specialist Virology Centre | Public Health Wales Microbiology Cardiff |

| EPI_Accession_ID | Originating Laboratory | Submitting Laboratory |
|---|---|---|
| EPI_ISL_445794 | Wales Specialist Virology Centre | Public Health Wales Microbiology Cardiff |
| EPI_ISL_445795 | Wales Specialist Virology Centre | Public Health Wales Microbiology Cardiff |
| EPI_ISL_445796 | Wales Specialist Virology Centre | Public Health Wales Microbiology Cardiff |
| EPI_ISL_445797 | Wales Specialist Virology Centre | Public Health Wales Microbiology Cardiff |
| EPI_ISL_445798 | Wales Specialist Virology Centre | Public Health Wales Microbiology Cardiff |
| EPI_ISL_445799 | Wales Specialist Virology Centre | Public Health Wales Microbiology Cardiff |
| EPI_ISL_445800 | Wales Specialist Virology Centre | Public Health Wales Microbiology Cardiff |
| EPI_ISL_445801 | Wales Specialist Virology Centre | Public Health Wales Microbiology Cardiff |
| EPI_ISL_445802 | Wales Specialist Virology Centre | Public Health Wales Microbiology Cardiff |
| EPI_ISL_445803 | Wales Specialist Virology Centre | Public Health Wales Microbiology Cardiff |
| EPI_ISL_445804 | Wales Specialist Virology Centre | Public Health Wales Microbiology Cardiff |
| EPI_ISL_445805 | Wales Specialist Virology Centre | Public Health Wales Microbiology Cardiff |
| EPI_ISL_445806 | Wales Specialist Virology Centre | Public Health Wales Microbiology Cardiff |
| EPI_ISL_445807 | Wales Specialist Virology Centre | Public Health Wales Microbiology Cardiff |
| EPI_ISL_445808 | Wales Specialist Virology Centre | Public Health Wales Microbiology Cardiff |
| EPI_ISL_445809 | Wales Specialist Virology Centre | Public Health Wales Microbiology Cardiff |
| EPI_ISL_445810 | Wales Specialist Virology Centre | Public Health Wales Microbiology Cardiff |
| EPI_ISL_445811 | Wales Specialist Virology Centre | Public Health Wales Microbiology Cardiff |
| EPI_ISL_445812 | Wales Specialist Virology Centre | Public Health Wales Microbiology Cardiff |
| EPI_ISL_445813 | Wales Specialist Virology Centre | Public Health Wales Microbiology Cardiff |
| EPI_ISL_445814 | Wales Specialist Virology Centre | Public Health Wales Microbiology Cardiff |
| EPI_ISL_445815 | Wales Specialist Virology Centre | Public Health Wales Microbiology Cardiff |
| EPI_ISL_445816 | Wales Specialist Virology Centre | Public Health Wales Microbiology Cardiff |
| EPI_ISL_445817 | Wales Specialist Virology Centre | Public Health Wales Microbiology Cardiff |
| EPI_ISL_445818 | Wales Specialist Virology Centre | Public Health Wales Microbiology Cardiff |
| EPI_ISL_445819 | Wales Specialist Virology Centre | Public Health Wales Microbiology Cardiff |
| EPI_ISL_445820 | Wales Specialist Virology Centre | Public Health Wales Microbiology Cardiff |
| EPI_ISL_445821 | Wales Specialist Virology Centre | Public Health Wales Microbiology Cardiff |
| EPI_ISL_445822 | Wales Specialist Virology Centre | Public Health Wales Microbiology Cardiff |
| EPI_ISL_445823 | Wales Specialist Virology Centre | Public Health Wales Microbiology Cardiff |
| EPI_ISL_445824 | Wales Specialist Virology Centre | Public Health Wales Microbiology Cardiff |
| EPI_ISL_445825 | Wales Specialist Virology Centre | Public Health Wales Microbiology Cardiff |
| EPI_ISL_445826 | Wales Specialist Virology Centre | Public Health Wales Microbiology Cardiff |

| EPI_Accession_ID | Originating Laboratory | Submitting Laboratory |
|---|---|---|
| EPI_ISL_445827 | Wales Specialist Virology Centre | Public Health Wales Microbiology Cardiff |
| EPI_ISL_445828 | Wales Specialist Virology Centre | Public Health Wales Microbiology Cardiff |
| EPI_ISL_445829 | Wales Specialist Virology Centre | Public Health Wales Microbiology Cardiff |
| EPI_ISL_445830 | Wales Specialist Virology Centre | Public Health Wales Microbiology Cardiff |
| EPI_ISL_445831 | Wales Specialist Virology Centre | Public Health Wales Microbiology Cardiff |
| EPI_ISL_445832 | Wales Specialist Virology Centre | Public Health Wales Microbiology Cardiff |
| EPI_ISL_445833 | Wales Specialist Virology Centre | Public Health Wales Microbiology Cardiff |
| EPI_ISL_445834 | Wales Specialist Virology Centre | Public Health Wales Microbiology Cardiff |
| EPI_ISL_445835 | Wales Specialist Virology Centre | Public Health Wales Microbiology Cardiff |
| EPI_ISL_445836 | Wales Specialist Virology Centre | Public Health Wales Microbiology Cardiff |
| EPI_ISL_445837 | Wales Specialist Virology Centre | Public Health Wales Microbiology Cardiff |
| EPI_ISL_445838 | Wales Specialist Virology Centre | Public Health Wales Microbiology Cardiff |
| EPI_ISL_445839 | Wales Specialist Virology Centre | Public Health Wales Microbiology Cardiff |
| EPI_ISL_445840 | Wales Specialist Virology Centre | Public Health Wales Microbiology Cardiff |
| EPI_ISL_445841 | Wales Specialist Virology Centre | Public Health Wales Microbiology Cardiff |
| EPI_ISL_445842 | Wales Specialist Virology Centre | Public Health Wales Microbiology Cardiff |
| EPI_ISL_445843 | Wales Specialist Virology Centre | Public Health Wales Microbiology Cardiff |
| EPI_ISL_445844 | Wales Specialist Virology Centre | Public Health Wales Microbiology Cardiff |
| EPI_ISL_445845 | Wales Specialist Virology Centre | Public Health Wales Microbiology Cardiff |
| EPI_ISL_445846 | Wales Specialist Virology Centre | Public Health Wales Microbiology Cardiff |
| EPI_ISL_445847 | Wales Specialist Virology Centre | Public Health Wales Microbiology Cardiff |
| EPI_ISL_445848 | Wales Specialist Virology Centre | Public Health Wales Microbiology Cardiff |
| EPI_ISL_445849 | Wales Specialist Virology Centre | Public Health Wales Microbiology Cardiff |
| EPI_ISL_445850 | Wales Specialist Virology Centre | Public Health Wales Microbiology Cardiff |
| EPI_ISL_445851 | Wales Specialist Virology Centre | Public Health Wales Microbiology Cardiff |
| EPI_ISL_445852 | Wales Specialist Virology Centre | Public Health Wales Microbiology Cardiff |
| EPI_ISL_445853 | Wales Specialist Virology Centre | Public Health Wales Microbiology Cardiff |
| EPI_ISL_445854 | Wales Specialist Virology Centre | Public Health Wales Microbiology Cardiff |
| EPI_ISL_445855 | Wales Specialist Virology Centre | Public Health Wales Microbiology Cardiff |
| EPI_ISL_445856 | Wales Specialist Virology Centre | Public Health Wales Microbiology Cardiff |
| EPI_ISL_445857 | Wales Specialist Virology Centre | Public Health Wales Microbiology Cardiff |
| EPI_ISL_445858 | Wales Specialist Virology Centre | Public Health Wales Microbiology Cardiff |
| EPI_ISL_445859 | Wales Specialist Virology Centre | Public Health Wales Microbiology Cardiff |

| EPI_Accession_ID | Originating Laboratory | Submitting Laboratory |
|---|---|---|
| EPI_ISL_445860 | Wales Specialist Virology Centre | Public Health Wales Microbiology Cardiff |
| EPI_ISL_445861 | Wales Specialist Virology Centre | Public Health Wales Microbiology Cardiff |
| EPI_ISL_445862 | Wales Specialist Virology Centre | Public Health Wales Microbiology Cardiff |
| EPI_ISL_445863 | Wales Specialist Virology Centre | Public Health Wales Microbiology Cardiff |
| EPI_ISL_445864 | Wales Specialist Virology Centre | Public Health Wales Microbiology Cardiff |
| EPI_ISL_445865 | Wales Specialist Virology Centre | Public Health Wales Microbiology Cardiff |
| EPI_ISL_445866 | Wales Specialist Virology Centre | Public Health Wales Microbiology Cardiff |
| EPI_ISL_445867 | Wales Specialist Virology Centre | Public Health Wales Microbiology Cardiff |
| EPI_ISL_445868 | Wales Specialist Virology Centre | Public Health Wales Microbiology Cardiff |
| EPI_ISL_445869 | Wales Specialist Virology Centre | Public Health Wales Microbiology Cardiff |
| EPI_ISL_445870 | Wales Specialist Virology Centre | Public Health Wales Microbiology Cardiff |
| EPI_ISL_445871 | Wales Specialist Virology Centre | Public Health Wales Microbiology Cardiff |
| EPI_ISL_445872 | Wales Specialist Virology Centre | Public Health Wales Microbiology Cardiff |
| EPI_ISL_445873 | Wales Specialist Virology Centre | Public Health Wales Microbiology Cardiff |
| EPI_ISL_445874 | Wales Specialist Virology Centre | Public Health Wales Microbiology Cardiff |
| EPI_ISL_445875 | Wales Specialist Virology Centre | Public Health Wales Microbiology Cardiff |
| EPI_ISL_445876 | Wales Specialist Virology Centre | Public Health Wales Microbiology Cardiff |
| EPI_ISL_445877 | Wales Specialist Virology Centre | Public Health Wales Microbiology Cardiff |
| EPI_ISL_445878 | Wales Specialist Virology Centre | Public Health Wales Microbiology Cardiff |
| EPI_ISL_445879 | Wales Specialist Virology Centre | Public Health Wales Microbiology Cardiff |
| EPI_ISL_445880 | Wales Specialist Virology Centre | Public Health Wales Microbiology Cardiff |
| EPI_ISL_445881 | Wales Specialist Virology Centre | Public Health Wales Microbiology Cardiff |
| EPI_ISL_445882 | Wales Specialist Virology Centre | Public Health Wales Microbiology Cardiff |
| EPI_ISL_445883 | Wales Specialist Virology Centre | Public Health Wales Microbiology Cardiff |
| EPI_ISL_445884 | Wales Specialist Virology Centre | Public Health Wales Microbiology Cardiff |
| EPI_ISL_445885 | Wales Specialist Virology Centre | Public Health Wales Microbiology Cardiff |
| EPI_ISL_445886 | Wales Specialist Virology Centre | Public Health Wales Microbiology Cardiff |
| EPI_ISL_445887 | Wales Specialist Virology Centre | Public Health Wales Microbiology Cardiff |
| EPI_ISL_445888 | Wales Specialist Virology Centre | Public Health Wales Microbiology Cardiff |
| EPI_ISL_445889 | Wales Specialist Virology Centre | Public Health Wales Microbiology Cardiff |
| EPI_ISL_445890 | Wales Specialist Virology Centre | Public Health Wales Microbiology Cardiff |
| EPI_ISL_445891 | Wales Specialist Virology Centre | Public Health Wales Microbiology Cardiff |
| EPI_ISL_445892 | Wales Specialist Virology Centre | Public Health Wales Microbiology Cardiff |

| EPI_Accession_ID | Originating Laboratory | Submitting Laboratory |
|---|---|---|
| EPI_ISL_445893 | Wales Specialist Virology Centre | Public Health Wales Microbiology Cardiff |
| EPI_ISL_445894 | Wales Specialist Virology Centre | Public Health Wales Microbiology Cardiff |
| EPI_ISL_445895 | Wales Specialist Virology Centre | Public Health Wales Microbiology Cardiff |
| EPI_ISL_445896 | Wales Specialist Virology Centre | Public Health Wales Microbiology Cardiff |
| EPI_ISL_445897 | Wales Specialist Virology Centre | Public Health Wales Microbiology Cardiff |
| EPI_ISL_445898 | Wales Specialist Virology Centre | Public Health Wales Microbiology Cardiff |
| EPI_ISL_445899 | Wales Specialist Virology Centre | Public Health Wales Microbiology Cardiff |
| EPI_ISL_445900 | Wales Specialist Virology Centre | Public Health Wales Microbiology Cardiff |
| EPI_ISL_445901 | Wales Specialist Virology Centre | Public Health Wales Microbiology Cardiff |
| EPI_ISL_445902 | Wales Specialist Virology Centre | Public Health Wales Microbiology Cardiff |
| EPI_ISL_445903 | Wales Specialist Virology Centre | Public Health Wales Microbiology Cardiff |
| EPI_ISL_445904 | Wales Specialist Virology Centre | Public Health Wales Microbiology Cardiff |
| EPI_ISL_445905 | Wales Specialist Virology Centre | Public Health Wales Microbiology Cardiff |
| EPI_ISL_445906 | Wales Specialist Virology Centre | Public Health Wales Microbiology Cardiff |
| EPI_ISL_445907 | Wales Specialist Virology Centre | Public Health Wales Microbiology Cardiff |
| EPI_ISL_445908 | Wales Specialist Virology Centre | Public Health Wales Microbiology Cardiff |
| EPI_ISL_445909 | Wales Specialist Virology Centre | Public Health Wales Microbiology Cardiff |
| EPI_ISL_445910 | Wales Specialist Virology Centre | Public Health Wales Microbiology Cardiff |
| EPI_ISL_445911 | Wales Specialist Virology Centre | Public Health Wales Microbiology Cardiff |
| EPI_ISL_445912 | Wales Specialist Virology Centre | Public Health Wales Microbiology Cardiff |
| EPI_ISL_445913 | Wales Specialist Virology Centre | Public Health Wales Microbiology Cardiff |
| EPI_ISL_445914 | Wales Specialist Virology Centre | Public Health Wales Microbiology Cardiff |
| EPI_ISL_445915 | Wales Specialist Virology Centre | Public Health Wales Microbiology Cardiff |
| EPI_ISL_445916 | Wales Specialist Virology Centre | Public Health Wales Microbiology Cardiff |
| EPI_ISL_445917 | Wales Specialist Virology Centre | Public Health Wales Microbiology Cardiff |
| EPI_ISL_445918 | Wales Specialist Virology Centre | Public Health Wales Microbiology Cardiff |
| EPI_ISL_445919 | Wales Specialist Virology Centre | Public Health Wales Microbiology Cardiff |
| EPI_ISL_445920 | Wales Specialist Virology Centre | Public Health Wales Microbiology Cardiff |
| EPI_ISL_445921 | Wales Specialist Virology Centre | Public Health Wales Microbiology Cardiff |
| EPI_ISL_445922 | Wales Specialist Virology Centre | Public Health Wales Microbiology Cardiff |
| EPI_ISL_445923 | Wales Specialist Virology Centre | Public Health Wales Microbiology Cardiff |
| EPI_ISL_445924 | Wales Specialist Virology Centre | Public Health Wales Microbiology Cardiff |
| EPI_ISL_445925 | Wales Specialist Virology Centre | Public Health Wales Microbiology Cardiff |

| EPI_Accession_ID | Originating Laboratory | Submitting Laboratory |
|---|---|---|
| EPI_ISL_445926 | Wales Specialist Virology Centre | Public Health Wales Microbiology Cardiff |
| EPI_ISL_445927 | Wales Specialist Virology Centre | Public Health Wales Microbiology Cardiff |
| EPI_ISL_445928 | Wales Specialist Virology Centre | Public Health Wales Microbiology Cardiff |
| EPI_ISL_445929 | Wales Specialist Virology Centre | Public Health Wales Microbiology Cardiff |
| EPI_ISL_445930 | Wales Specialist Virology Centre | Public Health Wales Microbiology Cardiff |
| EPI_ISL_445931 | Wales Specialist Virology Centre | Public Health Wales Microbiology Cardiff |
| EPI_ISL_445932 | Wales Specialist Virology Centre | Public Health Wales Microbiology Cardiff |
| EPI_ISL_445933 | Wales Specialist Virology Centre | Public Health Wales Microbiology Cardiff |
| EPI_ISL_445934 | Wales Specialist Virology Centre | Public Health Wales Microbiology Cardiff |
| EPI_ISL_445935 | Wales Specialist Virology Centre | Public Health Wales Microbiology Cardiff |
| EPI_ISL_445936 | Wales Specialist Virology Centre | Public Health Wales Microbiology Cardiff |
| EPI_ISL_445937 | Wales Specialist Virology Centre | Public Health Wales Microbiology Cardiff |
| EPI_ISL_445938 | Wales Specialist Virology Centre | Public Health Wales Microbiology Cardiff |
| EPI_ISL_445939 | Wales Specialist Virology Centre | Public Health Wales Microbiology Cardiff |
| EPI_ISL_445940 | Wales Specialist Virology Centre | Public Health Wales Microbiology Cardiff |
| EPI_ISL_445941 | Wales Specialist Virology Centre | Public Health Wales Microbiology Cardiff |
| EPI_ISL_445942 | Wales Specialist Virology Centre | Public Health Wales Microbiology Cardiff |
| EPI_ISL_445943 | Wales Specialist Virology Centre | Public Health Wales Microbiology Cardiff |
| EPI_ISL_445944 | Wales Specialist Virology Centre | Public Health Wales Microbiology Cardiff |
| EPI_ISL_445945 | Wales Specialist Virology Centre | Public Health Wales Microbiology Cardiff |
| EPI_ISL_445946 | Wales Specialist Virology Centre | Public Health Wales Microbiology Cardiff |
| EPI_ISL_445947 | Wales Specialist Virology Centre | Public Health Wales Microbiology Cardiff |
| EPI_ISL_445948 | Wales Specialist Virology Centre | Public Health Wales Microbiology Cardiff |
| EPI_ISL_445949 | Wales Specialist Virology Centre | Public Health Wales Microbiology Cardiff |
| EPI_ISL_445950 | Wales Specialist Virology Centre | Public Health Wales Microbiology Cardiff |
| EPI_ISL_445951 | Wales Specialist Virology Centre | Public Health Wales Microbiology Cardiff |
| EPI_ISL_445952 | Wales Specialist Virology Centre | Public Health Wales Microbiology Cardiff |
| EPI_ISL_445953 | Wales Specialist Virology Centre | Public Health Wales Microbiology Cardiff |
| EPI_ISL_445954 | Wales Specialist Virology Centre | Public Health Wales Microbiology Cardiff |
| EPI_ISL_445955 | Wales Specialist Virology Centre | Public Health Wales Microbiology Cardiff |
| EPI_ISL_445956 | Wales Specialist Virology Centre | Public Health Wales Microbiology Cardiff |
| EPI_ISL_445957 | Wales Specialist Virology Centre | Public Health Wales Microbiology Cardiff |
| EPI_ISL_445958 | Wales Specialist Virology Centre | Public Health Wales Microbiology Cardiff |

| EPI_Accession_ID | Originating Laboratory | Submitting Laboratory |
|---|---|---|
| EPI_ISL_445959 | Wales Specialist Virology Centre | Public Health Wales Microbiology Cardiff |
| EPI_ISL_445960 | Wales Specialist Virology Centre | Public Health Wales Microbiology Cardiff |
| EPI_ISL_445961 | Wales Specialist Virology Centre | Public Health Wales Microbiology Cardiff |
| EPI_ISL_445962 | Wales Specialist Virology Centre | Public Health Wales Microbiology Cardiff |
| EPI_ISL_445963 | Wales Specialist Virology Centre | Public Health Wales Microbiology Cardiff |
| EPI_ISL_445964 | Wales Specialist Virology Centre | Public Health Wales Microbiology Cardiff |
| EPI_ISL_445965 | Wales Specialist Virology Centre | Public Health Wales Microbiology Cardiff |
| EPI_ISL_445966 | Wales Specialist Virology Centre | Public Health Wales Microbiology Cardiff |
| EPI_ISL_445967 | Wales Specialist Virology Centre | Public Health Wales Microbiology Cardiff |
| EPI_ISL_445968 | Wales Specialist Virology Centre | Public Health Wales Microbiology Cardiff |
| EPI_ISL_445969 | Wales Specialist Virology Centre | Public Health Wales Microbiology Cardiff |
| EPI_ISL_445970 | Wales Specialist Virology Centre | Public Health Wales Microbiology Cardiff |
| EPI_ISL_445971 | Wales Specialist Virology Centre | Public Health Wales Microbiology Cardiff |
| EPI_ISL_445972 | Wales Specialist Virology Centre | Public Health Wales Microbiology Cardiff |
| EPI_ISL_445973 | Wales Specialist Virology Centre | Public Health Wales Microbiology Cardiff |
| EPI_ISL_445974 | Wales Specialist Virology Centre | Public Health Wales Microbiology Cardiff |
| EPI_ISL_445975 | Wales Specialist Virology Centre | Public Health Wales Microbiology Cardiff |
| EPI_ISL_445976 | Wales Specialist Virology Centre | Public Health Wales Microbiology Cardiff |
| EPI_ISL_445977 | Wales Specialist Virology Centre | Public Health Wales Microbiology Cardiff |
| EPI_ISL_445978 | Wales Specialist Virology Centre | Public Health Wales Microbiology Cardiff |
| EPI_ISL_445979 | Wales Specialist Virology Centre | Public Health Wales Microbiology Cardiff |
| EPI_ISL_445980 | Wales Specialist Virology Centre | Public Health Wales Microbiology Cardiff |
| EPI_ISL_445981 | Wales Specialist Virology Centre | Public Health Wales Microbiology Cardiff |
| EPI_ISL_445982 | Wales Specialist Virology Centre | Public Health Wales Microbiology Cardiff |
| EPI_ISL_445983 | Wales Specialist Virology Centre | Public Health Wales Microbiology Cardiff |
| EPI_ISL_445984 | Wales Specialist Virology Centre | Public Health Wales Microbiology Cardiff |
| EPI_ISL_445985 | Wales Specialist Virology Centre | Public Health Wales Microbiology Cardiff |
| EPI_ISL_445986 | Wales Specialist Virology Centre | Public Health Wales Microbiology Cardiff |
| EPI_ISL_445987 | Wales Specialist Virology Centre | Public Health Wales Microbiology Cardiff |
| EPI_ISL_445988 | Wales Specialist Virology Centre | Public Health Wales Microbiology Cardiff |
| EPI_ISL_445989 | Wales Specialist Virology Centre | Public Health Wales Microbiology Cardiff |
| EPI_ISL_445990 | Wales Specialist Virology Centre | Public Health Wales Microbiology Cardiff |
| EPI_ISL_445991 | Wales Specialist Virology Centre | Public Health Wales Microbiology Cardiff |

| EPI_Accession_ID | Originating Laboratory | Submitting Laboratory |
|---|---|---|
| EPI_ISL_445992 | Wales Specialist Virology Centre | Public Health Wales Microbiology Cardiff |
| EPI_ISL_445993 | Wales Specialist Virology Centre | Public Health Wales Microbiology Cardiff |
| EPI_ISL_445994 | Wales Specialist Virology Centre | Public Health Wales Microbiology Cardiff |
| EPI_ISL_445995 | Wales Specialist Virology Centre | Public Health Wales Microbiology Cardiff |
| EPI_ISL_445996 | Wales Specialist Virology Centre | Public Health Wales Microbiology Cardiff |
| EPI_ISL_445997 | Wales Specialist Virology Centre | Public Health Wales Microbiology Cardiff |
| EPI_ISL_445998 | Wales Specialist Virology Centre | Public Health Wales Microbiology Cardiff |
| EPI_ISL_445999 | Wales Specialist Virology Centre | Public Health Wales Microbiology Cardiff |
| EPI_ISL_446000 | Wales Specialist Virology Centre | Public Health Wales Microbiology Cardiff |
| EPI_ISL_446001 | Wales Specialist Virology Centre | Public Health Wales Microbiology Cardiff |
| EPI_ISL_446002 | Wales Specialist Virology Centre | Public Health Wales Microbiology Cardiff |
| EPI_ISL_446003 | Wales Specialist Virology Centre | Public Health Wales Microbiology Cardiff |
| EPI_ISL_446004 | Wales Specialist Virology Centre | Public Health Wales Microbiology Cardiff |
| EPI_ISL_446005 | Wales Specialist Virology Centre | Public Health Wales Microbiology Cardiff |
| EPI_ISL_446006 | Wales Specialist Virology Centre | Public Health Wales Microbiology Cardiff |
| EPI_ISL_446007 | Wales Specialist Virology Centre | Public Health Wales Microbiology Cardiff |
| EPI_ISL_446008 | Wales Specialist Virology Centre | Public Health Wales Microbiology Cardiff |
| EPI_ISL_446009 | Wales Specialist Virology Centre | Public Health Wales Microbiology Cardiff |
| EPI_ISL_446010 | Wales Specialist Virology Centre | Public Health Wales Microbiology Cardiff |
| EPI_ISL_446011 | Wales Specialist Virology Centre | Public Health Wales Microbiology Cardiff |
| EPI_ISL_446012 | Wales Specialist Virology Centre | Public Health Wales Microbiology Cardiff |
| EPI_ISL_446013 | Wales Specialist Virology Centre | Public Health Wales Microbiology Cardiff |
| EPI_ISL_446014 | Wales Specialist Virology Centre | Public Health Wales Microbiology Cardiff |
| EPI_ISL_446015 | Wales Specialist Virology Centre | Public Health Wales Microbiology Cardiff |
| EPI_ISL_446016 | Wales Specialist Virology Centre | Public Health Wales Microbiology Cardiff |
| EPI_ISL_446017 | Wales Specialist Virology Centre | Public Health Wales Microbiology Cardiff |
| EPI_ISL_446018 | Wales Specialist Virology Centre | Public Health Wales Microbiology Cardiff |
| EPI_ISL_446019 | Wales Specialist Virology Centre | Public Health Wales Microbiology Cardiff |
| EPI_ISL_446020 | Wales Specialist Virology Centre | Public Health Wales Microbiology Cardiff |
| EPI_ISL_446021 | Wales Specialist Virology Centre | Public Health Wales Microbiology Cardiff |
| EPI_ISL_446022 | Wales Specialist Virology Centre | Public Health Wales Microbiology Cardiff |
| EPI_ISL_446023 | Wales Specialist Virology Centre | Public Health Wales Microbiology Cardiff |
| EPI_ISL_446024 | Wales Specialist Virology Centre | Public Health Wales Microbiology Cardiff |

| EPI_Accession_ID | Originating Laboratory | Submitting Laboratory |
| --- | --- | --- |
| EPI_ISL_446025 | Wales Specialist Virology Centre | Public Health Wales Microbiology Cardiff |
| EPI_ISL_446026 | Wales Specialist Virology Centre | Public Health Wales Microbiology Cardiff |
| EPI_ISL_446027 | Wales Specialist Virology Centre | Public Health Wales Microbiology Cardiff |
| EPI_ISL_446028 | Wales Specialist Virology Centre | Public Health Wales Microbiology Cardiff |
| EPI_ISL_446029 | Wales Specialist Virology Centre | Public Health Wales Microbiology Cardiff |
| EPI_ISL_446030 | Wales Specialist Virology Centre | Public Health Wales Microbiology Cardiff |
| EPI_ISL_446031 | Wales Specialist Virology Centre | Public Health Wales Microbiology Cardiff |
| EPI_ISL_446032 | Wales Specialist Virology Centre | Public Health Wales Microbiology Cardiff |
| EPI_ISL_446033 | Wales Specialist Virology Centre | Public Health Wales Microbiology Cardiff |
| EPI_ISL_446034 | Wales Specialist Virology Centre | Public Health Wales Microbiology Cardiff |
| EPI_ISL_446035 | Wales Specialist Virology Centre | Public Health Wales Microbiology Cardiff |
| EPI_ISL_446036 | Wales Specialist Virology Centre | Public Health Wales Microbiology Cardiff |
| EPI_ISL_446037 | Wales Specialist Virology Centre | Public Health Wales Microbiology Cardiff |
| EPI_ISL_446038 | Wales Specialist Virology Centre | Public Health Wales Microbiology Cardiff |
| EPI_ISL_446039 | Wales Specialist Virology Centre | Public Health Wales Microbiology Cardiff |
| EPI_ISL_446040 | Wales Specialist Virology Centre | Public Health Wales Microbiology Cardiff |
| EPI_ISL_446041 | Wales Specialist Virology Centre | Public Health Wales Microbiology Cardiff |
| EPI_ISL_446042 | Wales Specialist Virology Centre | Public Health Wales Microbiology Cardiff |
| EPI_ISL_446043 | Wales Specialist Virology Centre | Public Health Wales Microbiology Cardiff |
| EPI_ISL_446044 | Wales Specialist Virology Centre | Public Health Wales Microbiology Cardiff |
| EPI_ISL_446045 | Wales Specialist Virology Centre | Public Health Wales Microbiology Cardiff |
| EPI_ISL_446046 | Wales Specialist Virology Centre | Public Health Wales Microbiology Cardiff |
| EPI_ISL_446047 | Wales Specialist Virology Centre | Public Health Wales Microbiology Cardiff |
| EPI_ISL_446048 | Wales Specialist Virology Centre | Public Health Wales Microbiology Cardiff |
| EPI_ISL_446049 | Wales Specialist Virology Centre | Public Health Wales Microbiology Cardiff |
| EPI_ISL_446050 | Wales Specialist Virology Centre | Public Health Wales Microbiology Cardiff |
| EPI_ISL_446051 | Wales Specialist Virology Centre | Public Health Wales Microbiology Cardiff |
| EPI_ISL_446052 | Wales Specialist Virology Centre | Public Health Wales Microbiology Cardiff |
| EPI_ISL_446053 | Wales Specialist Virology Centre | Public Health Wales Microbiology Cardiff |
| EPI_ISL_446054 | Wales Specialist Virology Centre | Public Health Wales Microbiology Cardiff |
| EPI_ISL_446055 | Wales Specialist Virology Centre | Public Health Wales Microbiology Cardiff |
| EPI_ISL_446056 | Wales Specialist Virology Centre | Public Health Wales Microbiology Cardiff |
| EPI_ISL_446057 | Wales Specialist Virology Centre | Public Health Wales Microbiology Cardiff |

| EPI_Accession_ID | Originating Laboratory | Submitting Laboratory |
|---|---|---|
| EPI_ISL_446058 | Wales Specialist Virology Centre | Public Health Wales Microbiology Cardiff |
| EPI_ISL_446059 | Wales Specialist Virology Centre | Public Health Wales Microbiology Cardiff |
| EPI_ISL_446060 | Wales Specialist Virology Centre | Public Health Wales Microbiology Cardiff |
| EPI_ISL_446061 | Wales Specialist Virology Centre | Public Health Wales Microbiology Cardiff |
| EPI_ISL_446062 | Wales Specialist Virology Centre | Public Health Wales Microbiology Cardiff |
| EPI_ISL_446063 | Wales Specialist Virology Centre | Public Health Wales Microbiology Cardiff |
| EPI_ISL_446064 | Wales Specialist Virology Centre | Public Health Wales Microbiology Cardiff |
| EPI_ISL_446065 | Wales Specialist Virology Centre | Public Health Wales Microbiology Cardiff |
| EPI_ISL_446066 | Wales Specialist Virology Centre | Public Health Wales Microbiology Cardiff |
| EPI_ISL_446067 | Wales Specialist Virology Centre | Public Health Wales Microbiology Cardiff |
| EPI_ISL_446068 | Wales Specialist Virology Centre | Public Health Wales Microbiology Cardiff |
| EPI_ISL_446069 | Wales Specialist Virology Centre | Public Health Wales Microbiology Cardiff |
| EPI_ISL_446070 | Wales Specialist Virology Centre | Public Health Wales Microbiology Cardiff |
| EPI_ISL_446071 | Wales Specialist Virology Centre | Public Health Wales Microbiology Cardiff |
| EPI_ISL_446072 | Wales Specialist Virology Centre | Public Health Wales Microbiology Cardiff |
| EPI_ISL_446073 | Wales Specialist Virology Centre | Public Health Wales Microbiology Cardiff |
| EPI_ISL_446074 | Wales Specialist Virology Centre | Public Health Wales Microbiology Cardiff |
| EPI_ISL_446075 | Wales Specialist Virology Centre | Public Health Wales Microbiology Cardiff |
| EPI_ISL_446076 | Wales Specialist Virology Centre | Public Health Wales Microbiology Cardiff |
| EPI_ISL_446077 | Wales Specialist Virology Centre | Public Health Wales Microbiology Cardiff |
| EPI_ISL_446078 | Wales Specialist Virology Centre | Public Health Wales Microbiology Cardiff |
| EPI_ISL_446079 | Wales Specialist Virology Centre | Public Health Wales Microbiology Cardiff |
| EPI_ISL_446080 | Wales Specialist Virology Centre | Public Health Wales Microbiology Cardiff |
| EPI_ISL_446081 | Wales Specialist Virology Centre | Public Health Wales Microbiology Cardiff |
| EPI_ISL_446082 | Wales Specialist Virology Centre | Public Health Wales Microbiology Cardiff |
| EPI_ISL_446083 | Wales Specialist Virology Centre | Public Health Wales Microbiology Cardiff |
| EPI_ISL_446084 | Wales Specialist Virology Centre | Public Health Wales Microbiology Cardiff |
| EPI_ISL_446085 | Wales Specialist Virology Centre | Public Health Wales Microbiology Cardiff |
| EPI_ISL_446086 | Wales Specialist Virology Centre | Public Health Wales Microbiology Cardiff |
| EPI_ISL_446087 | Wales Specialist Virology Centre | Public Health Wales Microbiology Cardiff |
| EPI_ISL_446088 | Wales Specialist Virology Centre | Public Health Wales Microbiology Cardiff |
| EPI_ISL_446089 | Wales Specialist Virology Centre | Public Health Wales Microbiology Cardiff |
| EPI_ISL_446090 | Wales Specialist Virology Centre | Public Health Wales Microbiology Cardiff |

| EPI_Accession_ID | Originating Laboratory | Submitting Laboratory |
|---|---|---|
| EPI_ISL_446091 | Wales Specialist Virology Centre | Public Health Wales Microbiology Cardiff |
| EPI_ISL_446092 | Wales Specialist Virology Centre | Public Health Wales Microbiology Cardiff |
| EPI_ISL_446093 | Wales Specialist Virology Centre | Public Health Wales Microbiology Cardiff |
| EPI_ISL_446094 | Wales Specialist Virology Centre | Public Health Wales Microbiology Cardiff |
| EPI_ISL_446095 | Wales Specialist Virology Centre | Public Health Wales Microbiology Cardiff |
| EPI_ISL_446096 | Wales Specialist Virology Centre | Public Health Wales Microbiology Cardiff |
| EPI_ISL_446097 | Wales Specialist Virology Centre | Public Health Wales Microbiology Cardiff |
| EPI_ISL_446098 | Wales Specialist Virology Centre | Public Health Wales Microbiology Cardiff |
| EPI_ISL_446099 | Wales Specialist Virology Centre | Public Health Wales Microbiology Cardiff |
| EPI_ISL_446100 | Wales Specialist Virology Centre | Public Health Wales Microbiology Cardiff |
| EPI_ISL_446101 | Wales Specialist Virology Centre | Public Health Wales Microbiology Cardiff |
| EPI_ISL_446102 | Wales Specialist Virology Centre | Public Health Wales Microbiology Cardiff |
| EPI_ISL_446103 | Wales Specialist Virology Centre | Public Health Wales Microbiology Cardiff |
| EPI_ISL_446104 | Wales Specialist Virology Centre | Public Health Wales Microbiology Cardiff |
| EPI_ISL_446105 | Wales Specialist Virology Centre | Public Health Wales Microbiology Cardiff |
| EPI_ISL_446106 | Wales Specialist Virology Centre | Public Health Wales Microbiology Cardiff |
| EPI_ISL_446107 | Wales Specialist Virology Centre | Public Health Wales Microbiology Cardiff |
| EPI_ISL_446108 | Wales Specialist Virology Centre | Public Health Wales Microbiology Cardiff |
| EPI_ISL_446109 | Wales Specialist Virology Centre | Public Health Wales Microbiology Cardiff |
| EPI_ISL_446110 | Wales Specialist Virology Centre | Public Health Wales Microbiology Cardiff |
| EPI_ISL_446111 | Wales Specialist Virology Centre | Public Health Wales Microbiology Cardiff |
| EPI_ISL_446112 | Wales Specialist Virology Centre | Public Health Wales Microbiology Cardiff |
| EPI_ISL_446113 | Wales Specialist Virology Centre | Public Health Wales Microbiology Cardiff |
| EPI_ISL_446114 | Wales Specialist Virology Centre | Public Health Wales Microbiology Cardiff |
| EPI_ISL_446115 | Wales Specialist Virology Centre | Public Health Wales Microbiology Cardiff |
| EPI_ISL_446116 | Wales Specialist Virology Centre | Public Health Wales Microbiology Cardiff |
| EPI_ISL_446117 | Wales Specialist Virology Centre | Public Health Wales Microbiology Cardiff |
| EPI_ISL_446118 | Wales Specialist Virology Centre | Public Health Wales Microbiology Cardiff |
| EPI_ISL_446119 | Wales Specialist Virology Centre | Public Health Wales Microbiology Cardiff |
| EPI_ISL_446120 | Wales Specialist Virology Centre | Public Health Wales Microbiology Cardiff |
| EPI_ISL_446121 | Wales Specialist Virology Centre | Public Health Wales Microbiology Cardiff |
| EPI_ISL_446122 | Wales Specialist Virology Centre | Public Health Wales Microbiology Cardiff |
| EPI_ISL_446123 | Wales Specialist Virology Centre | Public Health Wales Microbiology Cardiff |

| EPI_Accession_ID | Originating Laboratory | Submitting Laboratory |
|---|---|---|
| EPI_ISL_446124 | Wales Specialist Virology Centre | Public Health Wales Microbiology Cardiff |
| EPI_ISL_446125 | Wales Specialist Virology Centre | Public Health Wales Microbiology Cardiff |
| EPI_ISL_446126 | Wales Specialist Virology Centre | Public Health Wales Microbiology Cardiff |
| EPI_ISL_446127 | Wales Specialist Virology Centre | Public Health Wales Microbiology Cardiff |
| EPI_ISL_446128 | Wales Specialist Virology Centre | Public Health Wales Microbiology Cardiff |
| EPI_ISL_446129 | Wales Specialist Virology Centre | Public Health Wales Microbiology Cardiff |
| EPI_ISL_446130 | Wales Specialist Virology Centre | Public Health Wales Microbiology Cardiff |
| EPI_ISL_446131 | Wales Specialist Virology Centre | Public Health Wales Microbiology Cardiff |
| EPI_ISL_446132 | Wales Specialist Virology Centre | Public Health Wales Microbiology Cardiff |
| EPI_ISL_446133 | Wales Specialist Virology Centre | Public Health Wales Microbiology Cardiff |
| EPI_ISL_446134 | Wales Specialist Virology Centre | Public Health Wales Microbiology Cardiff |
| EPI_ISL_446135 | Wales Specialist Virology Centre | Public Health Wales Microbiology Cardiff |
| EPI_ISL_446136 | Wales Specialist Virology Centre | Public Health Wales Microbiology Cardiff |
| EPI_ISL_446137 | Wales Specialist Virology Centre | Public Health Wales Microbiology Cardiff |
| EPI_ISL_446138 | Wales Specialist Virology Centre | Public Health Wales Microbiology Cardiff |
| EPI_ISL_446139 | Wales Specialist Virology Centre | Public Health Wales Microbiology Cardiff |
| EPI_ISL_446140 | Wales Specialist Virology Centre | Public Health Wales Microbiology Cardiff |
| EPI_ISL_446141 | Wales Specialist Virology Centre | Public Health Wales Microbiology Cardiff |
| EPI_ISL_446142 | Wales Specialist Virology Centre | Public Health Wales Microbiology Cardiff |
| EPI_ISL_446143 | Wales Specialist Virology Centre | Public Health Wales Microbiology Cardiff |
| EPI_ISL_446144 | Wales Specialist Virology Centre | Public Health Wales Microbiology Cardiff |
| EPI_ISL_446145 | Wales Specialist Virology Centre | Public Health Wales Microbiology Cardiff |
| EPI_ISL_446146 | Wales Specialist Virology Centre | Public Health Wales Microbiology Cardiff |
| EPI_ISL_446147 | Wales Specialist Virology Centre | Public Health Wales Microbiology Cardiff |
| EPI_ISL_446148 | Wales Specialist Virology Centre | Public Health Wales Microbiology Cardiff |
| EPI_ISL_446149 | Wales Specialist Virology Centre | Public Health Wales Microbiology Cardiff |
| EPI_ISL_446150 | Wales Specialist Virology Centre | Public Health Wales Microbiology Cardiff |
| EPI_ISL_446151 | Wales Specialist Virology Centre | Public Health Wales Microbiology Cardiff |
| EPI_ISL_446152 | Wales Specialist Virology Centre | Public Health Wales Microbiology Cardiff |
| EPI_ISL_446153 | Wales Specialist Virology Centre | Public Health Wales Microbiology Cardiff |
| EPI_ISL_446154 | Wales Specialist Virology Centre | Public Health Wales Microbiology Cardiff |
| EPI_ISL_446155 | Wales Specialist Virology Centre | Public Health Wales Microbiology Cardiff |
| EPI_ISL_446156 | Wales Specialist Virology Centre | Public Health Wales Microbiology Cardiff |

| EPI_Accession_ID | Originating Laboratory | Submitting Laboratory |
|---|---|---|
| EPI_ISL_446157 | Wales Specialist Virology Centre | Public Health Wales Microbiology Cardiff |
| EPI_ISL_446158 | Wales Specialist Virology Centre | Public Health Wales Microbiology Cardiff |
| EPI_ISL_446159 | Wales Specialist Virology Centre | Public Health Wales Microbiology Cardiff |
| EPI_ISL_446160 | Wales Specialist Virology Centre | Public Health Wales Microbiology Cardiff |
| EPI_ISL_446161 | Wales Specialist Virology Centre | Public Health Wales Microbiology Cardiff |
| EPI_ISL_446162 | Wales Specialist Virology Centre | Public Health Wales Microbiology Cardiff |
| EPI_ISL_446163 | Wales Specialist Virology Centre | Public Health Wales Microbiology Cardiff |
| EPI_ISL_446164 | Wales Specialist Virology Centre | Public Health Wales Microbiology Cardiff |
| EPI_ISL_446165 | Wales Specialist Virology Centre | Public Health Wales Microbiology Cardiff |
| EPI_ISL_446166 | Wales Specialist Virology Centre | Public Health Wales Microbiology Cardiff |
| EPI_ISL_446167 | Wales Specialist Virology Centre | Public Health Wales Microbiology Cardiff |
| EPI_ISL_446168 | Wales Specialist Virology Centre | Public Health Wales Microbiology Cardiff |
| EPI_ISL_446169 | Wales Specialist Virology Centre | Public Health Wales Microbiology Cardiff |
| EPI_ISL_446170 | Wales Specialist Virology Centre | Public Health Wales Microbiology Cardiff |
| EPI_ISL_446171 | Wales Specialist Virology Centre | Public Health Wales Microbiology Cardiff |
| EPI_ISL_446172 | Wales Specialist Virology Centre | Public Health Wales Microbiology Cardiff |
| EPI_ISL_446173 | Wales Specialist Virology Centre | Public Health Wales Microbiology Cardiff |
| EPI_ISL_446174 | Wales Specialist Virology Centre | Public Health Wales Microbiology Cardiff |
| EPI_ISL_446175 | Wales Specialist Virology Centre | Public Health Wales Microbiology Cardiff |
| EPI_ISL_446176 | Wales Specialist Virology Centre | Public Health Wales Microbiology Cardiff |
| EPI_ISL_446177 | Wales Specialist Virology Centre | Public Health Wales Microbiology Cardiff |
| EPI_ISL_446178 | Wales Specialist Virology Centre | Public Health Wales Microbiology Cardiff |
| EPI_ISL_446179 | Wales Specialist Virology Centre | Public Health Wales Microbiology Cardiff |
| EPI_ISL_446180 | Wales Specialist Virology Centre | Public Health Wales Microbiology Cardiff |
| EPI_ISL_446181 | Wales Specialist Virology Centre | Public Health Wales Microbiology Cardiff |
| EPI_ISL_446182 | Wales Specialist Virology Centre | Public Health Wales Microbiology Cardiff |
| EPI_ISL_446183 | Wales Specialist Virology Centre | Public Health Wales Microbiology Cardiff |
| EPI_ISL_446184 | Wales Specialist Virology Centre | Public Health Wales Microbiology Cardiff |
| EPI_ISL_446185 | Wales Specialist Virology Centre | Public Health Wales Microbiology Cardiff |
| EPI_ISL_446186 | Wales Specialist Virology Centre | Public Health Wales Microbiology Cardiff |
| EPI_ISL_446187 | Wales Specialist Virology Centre | Public Health Wales Microbiology Cardiff |
| EPI_ISL_446188 | Wales Specialist Virology Centre | Public Health Wales Microbiology Cardiff |
| EPI_ISL_446189 | Wales Specialist Virology Centre | Public Health Wales Microbiology Cardiff |

| EPI_Accession_ID | Originating Laboratory | Submitting Laboratory |
|---|---|---|
| EPI_ISL_446190 | Wales Specialist Virology Centre | Public Health Wales Microbiology Cardiff |
| EPI_ISL_446191 | Wales Specialist Virology Centre | Public Health Wales Microbiology Cardiff |
| EPI_ISL_446192 | Wales Specialist Virology Centre | Public Health Wales Microbiology Cardiff |
| EPI_ISL_446193 | Wales Specialist Virology Centre | Public Health Wales Microbiology Cardiff |
| EPI_ISL_446194 | Wales Specialist Virology Centre | Public Health Wales Microbiology Cardiff |
| EPI_ISL_446195 | Wales Specialist Virology Centre | Public Health Wales Microbiology Cardiff |
| EPI_ISL_446196 | Wales Specialist Virology Centre | Public Health Wales Microbiology Cardiff |
| EPI_ISL_446197 | Wales Specialist Virology Centre | Public Health Wales Microbiology Cardiff |
| EPI_ISL_446198 | Wales Specialist Virology Centre | Public Health Wales Microbiology Cardiff |
| EPI_ISL_446199 | Wales Specialist Virology Centre | Public Health Wales Microbiology Cardiff |
| EPI_ISL_446200 | Wales Specialist Virology Centre | Public Health Wales Microbiology Cardiff |
| EPI_ISL_446201 | Wales Specialist Virology Centre | Public Health Wales Microbiology Cardiff |
| EPI_ISL_446202 | Wales Specialist Virology Centre | Public Health Wales Microbiology Cardiff |
| EPI_ISL_446203 | Wales Specialist Virology Centre | Public Health Wales Microbiology Cardiff |
| EPI_ISL_446204 | Wales Specialist Virology Centre | Public Health Wales Microbiology Cardiff |
| EPI_ISL_446205 | Wales Specialist Virology Centre | Public Health Wales Microbiology Cardiff |
| EPI_ISL_446206 | Wales Specialist Virology Centre | Public Health Wales Microbiology Cardiff |
| EPI_ISL_446207 | Wales Specialist Virology Centre | Public Health Wales Microbiology Cardiff |
| EPI_ISL_446208 | Wales Specialist Virology Centre | Public Health Wales Microbiology Cardiff |
| EPI_ISL_446209 | Wales Specialist Virology Centre | Public Health Wales Microbiology Cardiff |
| EPI_ISL_446210 | Wales Specialist Virology Centre | Public Health Wales Microbiology Cardiff |
| EPI_ISL_446211 | Wales Specialist Virology Centre | Public Health Wales Microbiology Cardiff |
| EPI_ISL_446212 | Wales Specialist Virology Centre | Public Health Wales Microbiology Cardiff |
| EPI_ISL_446213 | Wales Specialist Virology Centre | Public Health Wales Microbiology Cardiff |
| EPI_ISL_446214 | Wales Specialist Virology Centre | Public Health Wales Microbiology Cardiff |
| EPI_ISL_446215 | Wales Specialist Virology Centre | Public Health Wales Microbiology Cardiff |
| EPI_ISL_446216 | Wales Specialist Virology Centre | Public Health Wales Microbiology Cardiff |
| EPI_ISL_446217 | Wales Specialist Virology Centre | Public Health Wales Microbiology Cardiff |
| EPI_ISL_446218 | Wales Specialist Virology Centre | Public Health Wales Microbiology Cardiff |
| EPI_ISL_446219 | Wales Specialist Virology Centre | Public Health Wales Microbiology Cardiff |
| EPI_ISL_446220 | Wales Specialist Virology Centre | Public Health Wales Microbiology Cardiff |
| EPI_ISL_446221 | Wales Specialist Virology Centre | Public Health Wales Microbiology Cardiff |
| EPI_ISL_446222 | Wales Specialist Virology Centre | Public Health Wales Microbiology Cardiff |

| EPI_Accession_ID | Originating Laboratory | Submitting Laboratory |
|---|---|---|
| EPI_ISL_446223 | Wales Specialist Virology Centre | Public Health Wales Microbiology Cardiff |
| EPI_ISL_446224 | Wales Specialist Virology Centre | Public Health Wales Microbiology Cardiff |
| EPI_ISL_446225 | Wales Specialist Virology Centre | Public Health Wales Microbiology Cardiff |
| EPI_ISL_446226 | Wales Specialist Virology Centre | Public Health Wales Microbiology Cardiff |
| EPI_ISL_446227 | Wales Specialist Virology Centre | Public Health Wales Microbiology Cardiff |
| EPI_ISL_446228 | Wales Specialist Virology Centre | Public Health Wales Microbiology Cardiff |
| EPI_ISL_446229 | Wales Specialist Virology Centre | Public Health Wales Microbiology Cardiff |
| EPI_ISL_446230 | Wales Specialist Virology Centre | Public Health Wales Microbiology Cardiff |
| EPI_ISL_446231 | Wales Specialist Virology Centre | Public Health Wales Microbiology Cardiff |
| EPI_ISL_446232 | Wales Specialist Virology Centre | Public Health Wales Microbiology Cardiff |
| EPI_ISL_446233 | Wales Specialist Virology Centre | Public Health Wales Microbiology Cardiff |
| EPI_ISL_446234 | Wales Specialist Virology Centre | Public Health Wales Microbiology Cardiff |
| EPI_ISL_446235 | Wales Specialist Virology Centre | Public Health Wales Microbiology Cardiff |
| EPI_ISL_446236 | Wales Specialist Virology Centre | Public Health Wales Microbiology Cardiff |
| EPI_ISL_446237 | Wales Specialist Virology Centre | Public Health Wales Microbiology Cardiff |
| EPI_ISL_446238 | Wales Specialist Virology Centre | Public Health Wales Microbiology Cardiff |
| EPI_ISL_446239 | Wales Specialist Virology Centre | Public Health Wales Microbiology Cardiff |
| EPI_ISL_446240 | Wales Specialist Virology Centre | Public Health Wales Microbiology Cardiff |
| EPI_ISL_446241 | Wales Specialist Virology Centre | Public Health Wales Microbiology Cardiff |
| EPI_ISL_446242 | Wales Specialist Virology Centre | Public Health Wales Microbiology Cardiff |
| EPI_ISL_446243 | Wales Specialist Virology Centre | Public Health Wales Microbiology Cardiff |
| EPI_ISL_446244 | Wales Specialist Virology Centre | Public Health Wales Microbiology Cardiff |
| EPI_ISL_446245 | Wales Specialist Virology Centre | Public Health Wales Microbiology Cardiff |
| EPI_ISL_446246 | Wales Specialist Virology Centre | Public Health Wales Microbiology Cardiff |
| EPI_ISL_446247 | Wales Specialist Virology Centre | Public Health Wales Microbiology Cardiff |
| EPI_ISL_446248 | Wales Specialist Virology Centre | Public Health Wales Microbiology Cardiff |
| EPI_ISL_446249 | Wales Specialist Virology Centre | Public Health Wales Microbiology Cardiff |
| EPI_ISL_446250 | Wales Specialist Virology Centre | Public Health Wales Microbiology Cardiff |
| EPI_ISL_446251 | Wales Specialist Virology Centre | Public Health Wales Microbiology Cardiff |
| EPI_ISL_446252 | Wales Specialist Virology Centre | Public Health Wales Microbiology Cardiff |
| EPI_ISL_446253 | Wales Specialist Virology Centre | Public Health Wales Microbiology Cardiff |
| EPI_ISL_446254 | Wales Specialist Virology Centre | Public Health Wales Microbiology Cardiff |
| EPI_ISL_446255 | Wales Specialist Virology Centre | Public Health Wales Microbiology Cardiff |

| EPI_Accession_ID | Originating Laboratory | Submitting Laboratory |
|---|---|---|
| EPI_ISL_446256 | Wales Specialist Virology Centre | Public Health Wales Microbiology Cardiff |
| EPI_ISL_446257 | Wales Specialist Virology Centre | Public Health Wales Microbiology Cardiff |
| EPI_ISL_446258 | Wales Specialist Virology Centre | Public Health Wales Microbiology Cardiff |
| EPI_ISL_446259 | Wales Specialist Virology Centre | Public Health Wales Microbiology Cardiff |
| EPI_ISL_446260 | Wales Specialist Virology Centre | Public Health Wales Microbiology Cardiff |
| EPI_ISL_446261 | Wales Specialist Virology Centre | Public Health Wales Microbiology Cardiff |
| EPI_ISL_446262 | Wales Specialist Virology Centre | Public Health Wales Microbiology Cardiff |
| EPI_ISL_446263 | Wales Specialist Virology Centre | Public Health Wales Microbiology Cardiff |
| EPI_ISL_446264 | Wales Specialist Virology Centre | Public Health Wales Microbiology Cardiff |
| EPI_ISL_446265 | Wales Specialist Virology Centre | Public Health Wales Microbiology Cardiff |
| EPI_ISL_446266 | Wales Specialist Virology Centre | Public Health Wales Microbiology Cardiff |
| EPI_ISL_446267 | Wales Specialist Virology Centre | Public Health Wales Microbiology Cardiff |
| EPI_ISL_446268 | Wales Specialist Virology Centre | Public Health Wales Microbiology Cardiff |
| EPI_ISL_446269 | Wales Specialist Virology Centre | Public Health Wales Microbiology Cardiff |
| EPI_ISL_446270 | Wales Specialist Virology Centre | Public Health Wales Microbiology Cardiff |
| EPI_ISL_446271 | Wales Specialist Virology Centre | Public Health Wales Microbiology Cardiff |
| EPI_ISL_446272 | Wales Specialist Virology Centre | Public Health Wales Microbiology Cardiff |
| EPI_ISL_446273 | Wales Specialist Virology Centre | Public Health Wales Microbiology Cardiff |
| EPI_ISL_446274 | Wales Specialist Virology Centre | Public Health Wales Microbiology Cardiff |
| EPI_ISL_446275 | Wales Specialist Virology Centre | Public Health Wales Microbiology Cardiff |
| EPI_ISL_446276 | Wales Specialist Virology Centre | Public Health Wales Microbiology Cardiff |
| EPI_ISL_446277 | Wales Specialist Virology Centre | Public Health Wales Microbiology Cardiff |
| EPI_ISL_446278 | Wales Specialist Virology Centre | Public Health Wales Microbiology Cardiff |
| EPI_ISL_446279 | Wales Specialist Virology Centre | Public Health Wales Microbiology Cardiff |
| EPI_ISL_446280 | Wales Specialist Virology Centre | Public Health Wales Microbiology Cardiff |
| EPI_ISL_446281 | Wales Specialist Virology Centre | Public Health Wales Microbiology Cardiff |
| EPI_ISL_446282 | Wales Specialist Virology Centre | Public Health Wales Microbiology Cardiff |
| EPI_ISL_446283 | Wales Specialist Virology Centre | Public Health Wales Microbiology Cardiff |
| EPI_ISL_446284 | Wales Specialist Virology Centre | Public Health Wales Microbiology Cardiff |
| EPI_ISL_446285 | Wales Specialist Virology Centre | Public Health Wales Microbiology Cardiff |
| EPI_ISL_446286 | Wales Specialist Virology Centre | Public Health Wales Microbiology Cardiff |
| EPI_ISL_446287 | Wales Specialist Virology Centre | Public Health Wales Microbiology Cardiff |
| EPI_ISL_446288 | Wales Specialist Virology Centre | Public Health Wales Microbiology Cardiff |

| EPI_Accession_ID | Originating Laboratory | Submitting Laboratory |
|---|---|---|
| EPI_ISL_446289 | Wales Specialist Virology Centre | Public Health Wales Microbiology Cardiff |
| EPI_ISL_446290 | Wales Specialist Virology Centre | Public Health Wales Microbiology Cardiff |
| EPI_ISL_446291 | Wales Specialist Virology Centre | Public Health Wales Microbiology Cardiff |
| EPI_ISL_446292 | Wales Specialist Virology Centre | Public Health Wales Microbiology Cardiff |
| EPI_ISL_446293 | Wales Specialist Virology Centre | Public Health Wales Microbiology Cardiff |
| EPI_ISL_446294 | Wales Specialist Virology Centre | Public Health Wales Microbiology Cardiff |
| EPI_ISL_446295 | Wales Specialist Virology Centre | Public Health Wales Microbiology Cardiff |
| EPI_ISL_446296 | Wales Specialist Virology Centre | Public Health Wales Microbiology Cardiff |
| EPI_ISL_446297 | Wales Specialist Virology Centre | Public Health Wales Microbiology Cardiff |
| EPI_ISL_446298 | Wales Specialist Virology Centre | Public Health Wales Microbiology Cardiff |
| EPI_ISL_446299 | Wales Specialist Virology Centre | Public Health Wales Microbiology Cardiff |
| EPI_ISL_446300 | Wales Specialist Virology Centre | Public Health Wales Microbiology Cardiff |
| EPI_ISL_446301 | Wales Specialist Virology Centre | Public Health Wales Microbiology Cardiff |
| EPI_ISL_446302 | Wales Specialist Virology Centre | Public Health Wales Microbiology Cardiff |
| EPI_ISL_446303 | Wales Specialist Virology Centre | Public Health Wales Microbiology Cardiff |
| EPI_ISL_446304 | Wales Specialist Virology Centre | Public Health Wales Microbiology Cardiff |
| EPI_ISL_446305 | Wales Specialist Virology Centre | Public Health Wales Microbiology Cardiff |
| EPI_ISL_446306 | Wales Specialist Virology Centre | Public Health Wales Microbiology Cardiff |
| EPI_ISL_446307 | Wales Specialist Virology Centre | Public Health Wales Microbiology Cardiff |
| EPI_ISL_446308 | Wales Specialist Virology Centre | Public Health Wales Microbiology Cardiff |
| EPI_ISL_446309 | Wales Specialist Virology Centre | Public Health Wales Microbiology Cardiff |
| EPI_ISL_446310 | Wales Specialist Virology Centre | Public Health Wales Microbiology Cardiff |
| EPI_ISL_446311 | Wales Specialist Virology Centre | Public Health Wales Microbiology Cardiff |
| EPI_ISL_446312 | Wales Specialist Virology Centre | Public Health Wales Microbiology Cardiff |
| EPI_ISL_446313 | Wales Specialist Virology Centre | Public Health Wales Microbiology Cardiff |
| EPI_ISL_446314 | Wales Specialist Virology Centre | Public Health Wales Microbiology Cardiff |
| EPI_ISL_446315 | Wales Specialist Virology Centre | Public Health Wales Microbiology Cardiff |
| EPI_ISL_446316 | Wales Specialist Virology Centre | Public Health Wales Microbiology Cardiff |
| EPI_ISL_446317 | Wales Specialist Virology Centre | Public Health Wales Microbiology Cardiff |
| EPI_ISL_446318 | Wales Specialist Virology Centre | Public Health Wales Microbiology Cardiff |
| EPI_ISL_446319 | Wales Specialist Virology Centre | Public Health Wales Microbiology Cardiff |
| EPI_ISL_446320 | Wales Specialist Virology Centre | Public Health Wales Microbiology Cardiff |
| EPI_ISL_446321 | Wales Specialist Virology Centre | Public Health Wales Microbiology Cardiff |

| EPI_Accession_ID | Originating Laboratory | Submitting Laboratory |
|---|---|---|
| EPI_ISL_446322 | Wales Specialist Virology Centre | Public Health Wales Microbiology Cardiff |
| EPI_ISL_446323 | Wales Specialist Virology Centre | Public Health Wales Microbiology Cardiff |
| EPI_ISL_446324 | Wales Specialist Virology Centre | Public Health Wales Microbiology Cardiff |
| EPI_ISL_446325 | Wales Specialist Virology Centre | Public Health Wales Microbiology Cardiff |
| EPI_ISL_446326 | Wales Specialist Virology Centre | Public Health Wales Microbiology Cardiff |
| EPI_ISL_446327 | Wales Specialist Virology Centre | Public Health Wales Microbiology Cardiff |
| EPI_ISL_446328 | Wales Specialist Virology Centre | Public Health Wales Microbiology Cardiff |
| EPI_ISL_446329 | Wales Specialist Virology Centre | Public Health Wales Microbiology Cardiff |
| EPI_ISL_446330 | Wales Specialist Virology Centre | Public Health Wales Microbiology Cardiff |
| EPI_ISL_446331 | Wales Specialist Virology Centre | Public Health Wales Microbiology Cardiff |
| EPI_ISL_446332 | Wales Specialist Virology Centre | Public Health Wales Microbiology Cardiff |
| EPI_ISL_446333 | Wales Specialist Virology Centre | Public Health Wales Microbiology Cardiff |
| EPI_ISL_446334 | Wales Specialist Virology Centre | Public Health Wales Microbiology Cardiff |
| EPI_ISL_446335 | Wales Specialist Virology Centre | Public Health Wales Microbiology Cardiff |
| EPI_ISL_446336 | Wales Specialist Virology Centre | Public Health Wales Microbiology Cardiff |
| EPI_ISL_446337 | Wales Specialist Virology Centre | Public Health Wales Microbiology Cardiff |
| EPI_ISL_446338 | Wales Specialist Virology Centre | Public Health Wales Microbiology Cardiff |
| EPI_ISL_446339 | Wales Specialist Virology Centre | Public Health Wales Microbiology Cardiff |
| EPI_ISL_446340 | Wales Specialist Virology Centre | Public Health Wales Microbiology Cardiff |
| EPI_ISL_446341 | Wales Specialist Virology Centre | Public Health Wales Microbiology Cardiff |
| EPI_ISL_446342 | Wales Specialist Virology Centre | Public Health Wales Microbiology Cardiff |
| EPI_ISL_446343 | Wales Specialist Virology Centre | Public Health Wales Microbiology Cardiff |
| EPI_ISL_446344 | Wales Specialist Virology Centre | Public Health Wales Microbiology Cardiff |
| EPI_ISL_446345 | Wales Specialist Virology Centre | Public Health Wales Microbiology Cardiff |
| EPI_ISL_446346 | Wales Specialist Virology Centre | Public Health Wales Microbiology Cardiff |
| EPI_ISL_446347 | Wales Specialist Virology Centre | Public Health Wales Microbiology Cardiff |
| EPI_ISL_446348 | Wales Specialist Virology Centre | Public Health Wales Microbiology Cardiff |
| EPI_ISL_446349 | Wales Specialist Virology Centre | Public Health Wales Microbiology Cardiff |
| EPI_ISL_446350 | Wales Specialist Virology Centre | Public Health Wales Microbiology Cardiff |
| EPI_ISL_446351 | Wales Specialist Virology Centre | Public Health Wales Microbiology Cardiff |
| EPI_ISL_446352 | Wales Specialist Virology Centre | Public Health Wales Microbiology Cardiff |
| EPI_ISL_446353 | Wales Specialist Virology Centre | Public Health Wales Microbiology Cardiff |
| EPI_ISL_446354 | Wales Specialist Virology Centre | Public Health Wales Microbiology Cardiff |

| EPI_Accession_ID | Originating Laboratory | Submitting Laboratory |
| --- | --- | --- |
| EPI_ISL_446355 | Wales Specialist Virology Centre | Public Health Wales Microbiology Cardiff |
| EPI_ISL_446356 | Wales Specialist Virology Centre | Public Health Wales Microbiology Cardiff |
| EPI_ISL_446357 | Wales Specialist Virology Centre | Public Health Wales Microbiology Cardiff |
| EPI_ISL_446358 | Wales Specialist Virology Centre | Public Health Wales Microbiology Cardiff |
| EPI_ISL_446359 | Wales Specialist Virology Centre | Public Health Wales Microbiology Cardiff |
| EPI_ISL_446360 | Wales Specialist Virology Centre | Public Health Wales Microbiology Cardiff |
| EPI_ISL_446361 | Wales Specialist Virology Centre | Public Health Wales Microbiology Cardiff |
| EPI_ISL_446362 | Wales Specialist Virology Centre | Public Health Wales Microbiology Cardiff |
| EPI_ISL_446363 | Wales Specialist Virology Centre | Public Health Wales Microbiology Cardiff |
| EPI_ISL_446364 | Wales Specialist Virology Centre | Public Health Wales Microbiology Cardiff |
| EPI_ISL_446365 | Wales Specialist Virology Centre | Public Health Wales Microbiology Cardiff |
| EPI_ISL_446366 | Wales Specialist Virology Centre | Public Health Wales Microbiology Cardiff |
| EPI_ISL_446367 | Wales Specialist Virology Centre | Public Health Wales Microbiology Cardiff |
| EPI_ISL_446368 | Wales Specialist Virology Centre | Public Health Wales Microbiology Cardiff |
| EPI_ISL_446369 | Wales Specialist Virology Centre | Public Health Wales Microbiology Cardiff |
| EPI_ISL_446370 | Wales Specialist Virology Centre | Public Health Wales Microbiology Cardiff |
| EPI_ISL_446371 | Wales Specialist Virology Centre | Public Health Wales Microbiology Cardiff |
| EPI_ISL_446372 | Wales Specialist Virology Centre | Public Health Wales Microbiology Cardiff |
| EPI_ISL_446373 | Wales Specialist Virology Centre | Public Health Wales Microbiology Cardiff |
| EPI_ISL_446374 | Wales Specialist Virology Centre | Public Health Wales Microbiology Cardiff |
| EPI_ISL_446375 | Wales Specialist Virology Centre | Public Health Wales Microbiology Cardiff |
| EPI_ISL_446376 | Wales Specialist Virology Centre | Public Health Wales Microbiology Cardiff |
| EPI_ISL_446377 | Wales Specialist Virology Centre | Public Health Wales Microbiology Cardiff |
| EPI_ISL_446378 | Wales Specialist Virology Centre | Public Health Wales Microbiology Cardiff |
| EPI_ISL_446379 | Wales Specialist Virology Centre | Public Health Wales Microbiology Cardiff |
| EPI_ISL_446380 | Wales Specialist Virology Centre | Public Health Wales Microbiology Cardiff |
| EPI_ISL_446381 | Wales Specialist Virology Centre | Public Health Wales Microbiology Cardiff |
| EPI_ISL_446382 | Wales Specialist Virology Centre | Public Health Wales Microbiology Cardiff |
| EPI_ISL_446383 | Wales Specialist Virology Centre | Public Health Wales Microbiology Cardiff |
| EPI_ISL_446384 | Wales Specialist Virology Centre | Public Health Wales Microbiology Cardiff |
| EPI_ISL_446385 | Wales Specialist Virology Centre | Public Health Wales Microbiology Cardiff |
| EPI_ISL_446386 | Wales Specialist Virology Centre | Public Health Wales Microbiology Cardiff |
| EPI_ISL_446387 | Wales Specialist Virology Centre | Public Health Wales Microbiology Cardiff |

| EPI_Accession_ID | Originating Laboratory | Submitting Laboratory |
|---|---|---|
| EPI_ISL_446388 | Wales Specialist Virology Centre | Public Health Wales Microbiology Cardiff |
| EPI_ISL_446389 | Wales Specialist Virology Centre | Public Health Wales Microbiology Cardiff |
| EPI_ISL_446390 | Wales Specialist Virology Centre | Public Health Wales Microbiology Cardiff |
| EPI_ISL_446391 | Wales Specialist Virology Centre | Public Health Wales Microbiology Cardiff |
| EPI_ISL_446392 | Wales Specialist Virology Centre | Public Health Wales Microbiology Cardiff |
| EPI_ISL_446393 | Wales Specialist Virology Centre | Public Health Wales Microbiology Cardiff |
| EPI_ISL_446394 | Wales Specialist Virology Centre | Public Health Wales Microbiology Cardiff |
| EPI_ISL_446395 | Wales Specialist Virology Centre | Public Health Wales Microbiology Cardiff |
| EPI_ISL_446396 | Wales Specialist Virology Centre | Public Health Wales Microbiology Cardiff |
| EPI_ISL_446397 | Wales Specialist Virology Centre | Public Health Wales Microbiology Cardiff |
| EPI_ISL_446398 | Wales Specialist Virology Centre | Public Health Wales Microbiology Cardiff |
| EPI_ISL_446399 | Wales Specialist Virology Centre | Public Health Wales Microbiology Cardiff |
| EPI_ISL_446400 | Wales Specialist Virology Centre | Public Health Wales Microbiology Cardiff |
| EPI_ISL_446401 | Wales Specialist Virology Centre | Public Health Wales Microbiology Cardiff |
| EPI_ISL_446402 | Wales Specialist Virology Centre | Public Health Wales Microbiology Cardiff |
| EPI_ISL_446403 | Wales Specialist Virology Centre | Public Health Wales Microbiology Cardiff |
| EPI_ISL_446404 | Wales Specialist Virology Centre | Public Health Wales Microbiology Cardiff |
| EPI_ISL_446405 | Wales Specialist Virology Centre | Public Health Wales Microbiology Cardiff |
| EPI_ISL_446406 | Wales Specialist Virology Centre | Public Health Wales Microbiology Cardiff |
| EPI_ISL_446407 | Wales Specialist Virology Centre | Public Health Wales Microbiology Cardiff |
| EPI_ISL_446408 | Wales Specialist Virology Centre | Public Health Wales Microbiology Cardiff |
| EPI_ISL_446409 | Wales Specialist Virology Centre | Public Health Wales Microbiology Cardiff |
| EPI_ISL_446410 | Wales Specialist Virology Centre | Public Health Wales Microbiology Cardiff |
| EPI_ISL_446411 | Wales Specialist Virology Centre | Public Health Wales Microbiology Cardiff |
| EPI_ISL_446412 | Wales Specialist Virology Centre | Public Health Wales Microbiology Cardiff |
| EPI_ISL_446413 | Wales Specialist Virology Centre | Public Health Wales Microbiology Cardiff |
| EPI_ISL_446414 | Wales Specialist Virology Centre | Public Health Wales Microbiology Cardiff |
| EPI_ISL_446415 | Wales Specialist Virology Centre | Public Health Wales Microbiology Cardiff |
| EPI_ISL_446416 | Wales Specialist Virology Centre | Public Health Wales Microbiology Cardiff |
| EPI_ISL_446417 | Wales Specialist Virology Centre | Public Health Wales Microbiology Cardiff |
| EPI_ISL_446418 | Wales Specialist Virology Centre | Public Health Wales Microbiology Cardiff |
| EPI_ISL_446419 | Wales Specialist Virology Centre | Public Health Wales Microbiology Cardiff |
| EPI_ISL_446420 | Wales Specialist Virology Centre | Public Health Wales Microbiology Cardiff |

| EPI_Accession_ID | Originating Laboratory | Submitting Laboratory |
| --- | --- | --- |
| EPI_ISL_446421 | Wales Specialist Virology Centre | Public Health Wales Microbiology Cardiff |
| EPI_ISL_446422 | Wales Specialist Virology Centre | Public Health Wales Microbiology Cardiff |
| EPI_ISL_446423 | Wales Specialist Virology Centre | Public Health Wales Microbiology Cardiff |
| EPI_ISL_446424 | Wales Specialist Virology Centre | Public Health Wales Microbiology Cardiff |
| EPI_ISL_446425 | Wales Specialist Virology Centre | Public Health Wales Microbiology Cardiff |
| EPI_ISL_446426 | Wales Specialist Virology Centre | Public Health Wales Microbiology Cardiff |
| EPI_ISL_446427 | Wales Specialist Virology Centre | Public Health Wales Microbiology Cardiff |
| EPI_ISL_446428 | Wales Specialist Virology Centre | Public Health Wales Microbiology Cardiff |
| EPI_ISL_446429 | Wales Specialist Virology Centre | Public Health Wales Microbiology Cardiff |
| EPI_ISL_446430 | Wales Specialist Virology Centre | Public Health Wales Microbiology Cardiff |
| EPI_ISL_446431 | Wales Specialist Virology Centre | Public Health Wales Microbiology Cardiff |
| EPI_ISL_446432 | Wales Specialist Virology Centre | Public Health Wales Microbiology Cardiff |
| EPI_ISL_446433 | Wales Specialist Virology Centre | Public Health Wales Microbiology Cardiff |
| EPI_ISL_446434 | Wales Specialist Virology Centre | Public Health Wales Microbiology Cardiff |
| EPI_ISL_446435 | Wales Specialist Virology Centre | Public Health Wales Microbiology Cardiff |
| EPI_ISL_446436 | Wales Specialist Virology Centre | Public Health Wales Microbiology Cardiff |
| EPI_ISL_446437 | Wales Specialist Virology Centre | Public Health Wales Microbiology Cardiff |
| EPI_ISL_446438 | Wales Specialist Virology Centre | Public Health Wales Microbiology Cardiff |
| EPI_ISL_446439 | Wales Specialist Virology Centre | Public Health Wales Microbiology Cardiff |
| EPI_ISL_446440 | Wales Specialist Virology Centre | Public Health Wales Microbiology Cardiff |
| EPI_ISL_446441 | Wales Specialist Virology Centre | Public Health Wales Microbiology Cardiff |
| EPI_ISL_446442 | Wales Specialist Virology Centre | Public Health Wales Microbiology Cardiff |
| EPI_ISL_446443 | Wales Specialist Virology Centre | Public Health Wales Microbiology Cardiff |
| EPI_ISL_446444 | Wales Specialist Virology Centre | Public Health Wales Microbiology Cardiff |
| EPI_ISL_446445 | Wales Specialist Virology Centre | Public Health Wales Microbiology Cardiff |
| EPI_ISL_446446 | Wales Specialist Virology Centre | Public Health Wales Microbiology Cardiff |
| EPI_ISL_446447 | Wales Specialist Virology Centre | Public Health Wales Microbiology Cardiff |
| EPI_ISL_446448 | Wales Specialist Virology Centre | Public Health Wales Microbiology Cardiff |
| EPI_ISL_446449 | Wales Specialist Virology Centre | Public Health Wales Microbiology Cardiff |
| EPI_ISL_446450 | Wales Specialist Virology Centre | Public Health Wales Microbiology Cardiff |
| EPI_ISL_446451 | Wales Specialist Virology Centre | Public Health Wales Microbiology Cardiff |
| EPI_ISL_446452 | Wales Specialist Virology Centre | Public Health Wales Microbiology Cardiff |
| EPI_ISL_446453 | Wales Specialist Virology Centre | Public Health Wales Microbiology Cardiff |

| EPI_Accession_ID | Originating Laboratory | Submitting Laboratory |
|---|---|---|
| EPI_ISL_446454 | Wales Specialist Virology Centre | Public Health Wales Microbiology Cardiff |
| EPI_ISL_446455 | Wales Specialist Virology Centre | Public Health Wales Microbiology Cardiff |
| EPI_ISL_446456 | Wales Specialist Virology Centre | Public Health Wales Microbiology Cardiff |
| EPI_ISL_446457 | Wales Specialist Virology Centre | Public Health Wales Microbiology Cardiff |
| EPI_ISL_446458 | Wales Specialist Virology Centre | Public Health Wales Microbiology Cardiff |
| EPI_ISL_446459 | Wales Specialist Virology Centre | Public Health Wales Microbiology Cardiff |
| EPI_ISL_446460 | Wales Specialist Virology Centre | Public Health Wales Microbiology Cardiff |
| EPI_ISL_446461 | Wales Specialist Virology Centre | Public Health Wales Microbiology Cardiff |
| EPI_ISL_446462 | Wales Specialist Virology Centre | Public Health Wales Microbiology Cardiff |
| EPI_ISL_446463 | Wales Specialist Virology Centre | Public Health Wales Microbiology Cardiff |
| EPI_ISL_446464 | Wales Specialist Virology Centre | Public Health Wales Microbiology Cardiff |
| EPI_ISL_446465 | Wales Specialist Virology Centre | Public Health Wales Microbiology Cardiff |
| EPI_ISL_446466 | Wales Specialist Virology Centre | Public Health Wales Microbiology Cardiff |
| EPI_ISL_446467 | Wales Specialist Virology Centre | Public Health Wales Microbiology Cardiff |
| EPI_ISL_446468 | Wales Specialist Virology Centre | Public Health Wales Microbiology Cardiff |
| EPI_ISL_446469 | Wales Specialist Virology Centre | Public Health Wales Microbiology Cardiff |
| EPI_ISL_446470 | Wales Specialist Virology Centre | Public Health Wales Microbiology Cardiff |
| EPI_ISL_446471 | Wales Specialist Virology Centre | Public Health Wales Microbiology Cardiff |
| EPI_ISL_446472 | Wales Specialist Virology Centre | Public Health Wales Microbiology Cardiff |
| EPI_ISL_446473 | Wales Specialist Virology Centre | Public Health Wales Microbiology Cardiff |
| EPI_ISL_446474 | Wales Specialist Virology Centre | Public Health Wales Microbiology Cardiff |
| EPI_ISL_446475 | Wales Specialist Virology Centre | Public Health Wales Microbiology Cardiff |
| EPI_ISL_446476 | Wales Specialist Virology Centre | Public Health Wales Microbiology Cardiff |
| EPI_ISL_446477 | Wales Specialist Virology Centre | Public Health Wales Microbiology Cardiff |
| EPI_ISL_446478 | Wales Specialist Virology Centre | Public Health Wales Microbiology Cardiff |
| EPI_ISL_446479 | Wales Specialist Virology Centre | Public Health Wales Microbiology Cardiff |
| EPI_ISL_446480 | Wales Specialist Virology Centre | Public Health Wales Microbiology Cardiff |
| EPI_ISL_446481 | Wales Specialist Virology Centre | Public Health Wales Microbiology Cardiff |
| EPI_ISL_446482 | Wales Specialist Virology Centre | Public Health Wales Microbiology Cardiff |
| EPI_ISL_446483 | Wales Specialist Virology Centre | Public Health Wales Microbiology Cardiff |
| EPI_ISL_446484 | Wales Specialist Virology Centre | Public Health Wales Microbiology Cardiff |
| EPI_ISL_446485 | Wales Specialist Virology Centre | Public Health Wales Microbiology Cardiff |
| EPI_ISL_446486 | Wales Specialist Virology Centre | Public Health Wales Microbiology Cardiff |

| EPI_Accession_ID | Originating Laboratory | Submitting Laboratory |
|---|---|---|
| EPI_ISL_446487 | Wales Specialist Virology Centre | Public Health Wales Microbiology Cardiff |
| EPI_ISL_446488 | Wales Specialist Virology Centre | Public Health Wales Microbiology Cardiff |
| EPI_ISL_446489 | Wales Specialist Virology Centre | Public Health Wales Microbiology Cardiff |
| EPI_ISL_446490 | Wales Specialist Virology Centre | Public Health Wales Microbiology Cardiff |
| EPI_ISL_446491 | Wales Specialist Virology Centre | Public Health Wales Microbiology Cardiff |
| EPI_ISL_446492 | Wales Specialist Virology Centre | Public Health Wales Microbiology Cardiff |
| EPI_ISL_446493 | Wales Specialist Virology Centre | Public Health Wales Microbiology Cardiff |
| EPI_ISL_446494 | Wales Specialist Virology Centre | Public Health Wales Microbiology Cardiff |
| EPI_ISL_446495 | Wales Specialist Virology Centre | Public Health Wales Microbiology Cardiff |
| EPI_ISL_446496 | Wales Specialist Virology Centre | Public Health Wales Microbiology Cardiff |
| EPI_ISL_446497 | Wales Specialist Virology Centre | Public Health Wales Microbiology Cardiff |
| EPI_ISL_446498 | Wales Specialist Virology Centre | Public Health Wales Microbiology Cardiff |
| EPI_ISL_446499 | Wales Specialist Virology Centre | Public Health Wales Microbiology Cardiff |
| EPI_ISL_446500 | Wales Specialist Virology Centre | Public Health Wales Microbiology Cardiff |
| EPI_ISL_446501 | Wales Specialist Virology Centre | Public Health Wales Microbiology Cardiff |
| EPI_ISL_446502 | Wales Specialist Virology Centre | Public Health Wales Microbiology Cardiff |
| EPI_ISL_446503 | Wales Specialist Virology Centre | Public Health Wales Microbiology Cardiff |
| EPI_ISL_446504 | Wales Specialist Virology Centre | Public Health Wales Microbiology Cardiff |
| EPI_ISL_446505 | Wales Specialist Virology Centre | Public Health Wales Microbiology Cardiff |
| EPI_ISL_446506 | Wales Specialist Virology Centre | Public Health Wales Microbiology Cardiff |
| EPI_ISL_446507 | Wales Specialist Virology Centre | Public Health Wales Microbiology Cardiff |
| EPI_ISL_446508 | Wales Specialist Virology Centre | Public Health Wales Microbiology Cardiff |
| EPI_ISL_446509 | Wales Specialist Virology Centre | Public Health Wales Microbiology Cardiff |
| EPI_ISL_446510 | Wales Specialist Virology Centre | Public Health Wales Microbiology Cardiff |
| EPI_ISL_446511 | Wales Specialist Virology Centre | Public Health Wales Microbiology Cardiff |
| EPI_ISL_446512 | Wales Specialist Virology Centre | Public Health Wales Microbiology Cardiff |
| EPI_ISL_446513 | Wales Specialist Virology Centre | Public Health Wales Microbiology Cardiff |
| EPI_ISL_446514 | Wales Specialist Virology Centre | Public Health Wales Microbiology Cardiff |
| EPI_ISL_446515 | Wales Specialist Virology Centre | Public Health Wales Microbiology Cardiff |
| EPI_ISL_446516 | Wales Specialist Virology Centre | Public Health Wales Microbiology Cardiff |
| EPI_ISL_446517 | Wales Specialist Virology Centre | Public Health Wales Microbiology Cardiff |
| EPI_ISL_446518 | Wales Specialist Virology Centre | Public Health Wales Microbiology Cardiff |
| EPI_ISL_446519 | Wales Specialist Virology Centre | Public Health Wales Microbiology Cardiff |

| EPI_Accession_ID | Originating Laboratory | Submitting Laboratory |
|---|---|---|
| EPI_ISL_446520 | Wales Specialist Virology Centre | Public Health Wales Microbiology Cardiff |
| EPI_ISL_446521 | Wales Specialist Virology Centre | Public Health Wales Microbiology Cardiff |
| EPI_ISL_446522 | Wales Specialist Virology Centre | Public Health Wales Microbiology Cardiff |
| EPI_ISL_446523 | Wales Specialist Virology Centre | Public Health Wales Microbiology Cardiff |
| EPI_ISL_446524 | Wales Specialist Virology Centre | Public Health Wales Microbiology Cardiff |
| EPI_ISL_446525 | Wales Specialist Virology Centre | Public Health Wales Microbiology Cardiff |
| EPI_ISL_446526 | Wales Specialist Virology Centre | Public Health Wales Microbiology Cardiff |
| EPI_ISL_446527 | Wales Specialist Virology Centre | Public Health Wales Microbiology Cardiff |
| EPI_ISL_446528 | Wales Specialist Virology Centre | Public Health Wales Microbiology Cardiff |
| EPI_ISL_446529 | Wales Specialist Virology Centre | Public Health Wales Microbiology Cardiff |
| EPI_ISL_446530 | Wales Specialist Virology Centre | Public Health Wales Microbiology Cardiff |
| EPI_ISL_446531 | Wales Specialist Virology Centre | Public Health Wales Microbiology Cardiff |
| EPI_ISL_446532 | Wales Specialist Virology Centre | Public Health Wales Microbiology Cardiff |
| EPI_ISL_446533 | Wales Specialist Virology Centre | Public Health Wales Microbiology Cardiff |
| EPI_ISL_446534 | Wales Specialist Virology Centre | Public Health Wales Microbiology Cardiff |
| EPI_ISL_446535 | Wales Specialist Virology Centre | Public Health Wales Microbiology Cardiff |
| EPI_ISL_446536 | Wales Specialist Virology Centre | Public Health Wales Microbiology Cardiff |
| EPI_ISL_446537 | Wales Specialist Virology Centre | Public Health Wales Microbiology Cardiff |
| EPI_ISL_446538 | Wales Specialist Virology Centre | Public Health Wales Microbiology Cardiff |
| EPI_ISL_446539 | Wales Specialist Virology Centre | Public Health Wales Microbiology Cardiff |
| EPI_ISL_446540 | Wales Specialist Virology Centre | Public Health Wales Microbiology Cardiff |
| EPI_ISL_446541 | Wales Specialist Virology Centre | Public Health Wales Microbiology Cardiff |
| EPI_ISL_446542 | Wales Specialist Virology Centre | Public Health Wales Microbiology Cardiff |
| EPI_ISL_446543 | Wales Specialist Virology Centre | Public Health Wales Microbiology Cardiff |
| EPI_ISL_446544 | Wales Specialist Virology Centre | Public Health Wales Microbiology Cardiff |
| EPI_ISL_446545 | Wales Specialist Virology Centre | Public Health Wales Microbiology Cardiff |
| EPI_ISL_446546 | Wales Specialist Virology Centre | Public Health Wales Microbiology Cardiff |
| EPI_ISL_446547 | Wales Specialist Virology Centre | Public Health Wales Microbiology Cardiff |
| EPI_ISL_446548 | Wales Specialist Virology Centre | Public Health Wales Microbiology Cardiff |
| EPI_ISL_446549 | Wales Specialist Virology Centre | Public Health Wales Microbiology Cardiff |
| EPI_ISL_446550 | Wales Specialist Virology Centre | Public Health Wales Microbiology Cardiff |
| EPI_ISL_446551 | Wales Specialist Virology Centre | Public Health Wales Microbiology Cardiff |
| EPI_ISL_446552 | Wales Specialist Virology Centre | Public Health Wales Microbiology Cardiff |

| EPI_Accession_ID | Originating Laboratory | Submitting Laboratory |
|---|---|---|
| EPI_ISL_446553 | Wales Specialist Virology Centre | Public Health Wales Microbiology Cardiff |
| EPI_ISL_446554 | Wales Specialist Virology Centre | Public Health Wales Microbiology Cardiff |
| EPI_ISL_446555 | Wales Specialist Virology Centre | Public Health Wales Microbiology Cardiff |
| EPI_ISL_446556 | Wales Specialist Virology Centre | Public Health Wales Microbiology Cardiff |
| EPI_ISL_446557 | Wales Specialist Virology Centre | Public Health Wales Microbiology Cardiff |
| EPI_ISL_446558 | Wales Specialist Virology Centre | Public Health Wales Microbiology Cardiff |
| EPI_ISL_446559 | Wales Specialist Virology Centre | Public Health Wales Microbiology Cardiff |
| EPI_ISL_446560 | Wales Specialist Virology Centre | Public Health Wales Microbiology Cardiff |
| EPI_ISL_446561 | Wales Specialist Virology Centre | Public Health Wales Microbiology Cardiff |
| EPI_ISL_446562 | Wales Specialist Virology Centre | Public Health Wales Microbiology Cardiff |
| EPI_ISL_446563 | Wales Specialist Virology Centre | Public Health Wales Microbiology Cardiff |
| EPI_ISL_446564 | Wales Specialist Virology Centre | Public Health Wales Microbiology Cardiff |
| EPI_ISL_446565 | Wales Specialist Virology Centre | Public Health Wales Microbiology Cardiff |
| EPI_ISL_446566 | Wales Specialist Virology Centre | Public Health Wales Microbiology Cardiff |
| EPI_ISL_446567 | Wales Specialist Virology Centre | Public Health Wales Microbiology Cardiff |
| EPI_ISL_446568 | Wales Specialist Virology Centre | Public Health Wales Microbiology Cardiff |
| EPI_ISL_446569 | Wales Specialist Virology Centre | Public Health Wales Microbiology Cardiff |
| EPI_ISL_446570 | Wales Specialist Virology Centre | Public Health Wales Microbiology Cardiff |
| EPI_ISL_446571 | Wales Specialist Virology Centre | Public Health Wales Microbiology Cardiff |
| EPI_ISL_446572 | Wales Specialist Virology Centre | Public Health Wales Microbiology Cardiff |
| EPI_ISL_446573 | Wales Specialist Virology Centre | Public Health Wales Microbiology Cardiff |
| EPI_ISL_446574 | Wales Specialist Virology Centre | Public Health Wales Microbiology Cardiff |
| EPI_ISL_446575 | Wales Specialist Virology Centre | Public Health Wales Microbiology Cardiff |
| EPI_ISL_446576 | Wales Specialist Virology Centre | Public Health Wales Microbiology Cardiff |
| EPI_ISL_446577 | Wales Specialist Virology Centre | Public Health Wales Microbiology Cardiff |
| EPI_ISL_446578 | Wales Specialist Virology Centre | Public Health Wales Microbiology Cardiff |
| EPI_ISL_446579 | Wales Specialist Virology Centre | Public Health Wales Microbiology Cardiff |
| EPI_ISL_446580 | Wales Specialist Virology Centre | Public Health Wales Microbiology Cardiff |
| EPI_ISL_446581 | Wales Specialist Virology Centre | Public Health Wales Microbiology Cardiff |
| EPI_ISL_446582 | Wales Specialist Virology Centre | Public Health Wales Microbiology Cardiff |
| EPI_ISL_446583 | Wales Specialist Virology Centre | Public Health Wales Microbiology Cardiff |
| EPI_ISL_446584 | Wales Specialist Virology Centre | Public Health Wales Microbiology Cardiff |
| EPI_ISL_446585 | Wales Specialist Virology Centre | Public Health Wales Microbiology Cardiff |

| EPI_Accession_ID | Originating Laboratory | Submitting Laboratory |
|---|---|---|
| EPI_ISL_446586 | Wales Specialist Virology Centre | Public Health Wales Microbiology Cardiff |
| EPI_ISL_446587 | Wales Specialist Virology Centre | Public Health Wales Microbiology Cardiff |
| EPI_ISL_446588 | Wales Specialist Virology Centre | Public Health Wales Microbiology Cardiff |
| EPI_ISL_446589 | Wales Specialist Virology Centre | Public Health Wales Microbiology Cardiff |
| EPI_ISL_446590 | Wales Specialist Virology Centre | Public Health Wales Microbiology Cardiff |
| EPI_ISL_446591 | Wales Specialist Virology Centre | Public Health Wales Microbiology Cardiff |
| EPI_ISL_446592 | Wales Specialist Virology Centre | Public Health Wales Microbiology Cardiff |
| EPI_ISL_446593 | Wales Specialist Virology Centre | Public Health Wales Microbiology Cardiff |
| EPI_ISL_446594 | Wales Specialist Virology Centre | Public Health Wales Microbiology Cardiff |
| EPI_ISL_446595 | Wales Specialist Virology Centre | Public Health Wales Microbiology Cardiff |
| EPI_ISL_446596 | Wales Specialist Virology Centre | Public Health Wales Microbiology Cardiff |
| EPI_ISL_446597 | Wales Specialist Virology Centre | Public Health Wales Microbiology Cardiff |
| EPI_ISL_446598 | Wales Specialist Virology Centre | Public Health Wales Microbiology Cardiff |
| EPI_ISL_446599 | Wales Specialist Virology Centre | Public Health Wales Microbiology Cardiff |
| EPI_ISL_446600 | Wales Specialist Virology Centre | Public Health Wales Microbiology Cardiff |
| EPI_ISL_446601 | Wales Specialist Virology Centre | Public Health Wales Microbiology Cardiff |
| EPI_ISL_446602 | Wales Specialist Virology Centre | Public Health Wales Microbiology Cardiff |
| EPI_ISL_446603 | Wales Specialist Virology Centre | Public Health Wales Microbiology Cardiff |
| EPI_ISL_446604 | Wales Specialist Virology Centre | Public Health Wales Microbiology Cardiff |
| EPI_ISL_446605 | Wales Specialist Virology Centre | Public Health Wales Microbiology Cardiff |
| EPI_ISL_446606 | Wales Specialist Virology Centre | Public Health Wales Microbiology Cardiff |
| EPI_ISL_446607 | Wales Specialist Virology Centre | Public Health Wales Microbiology Cardiff |
| EPI_ISL_446608 | Wales Specialist Virology Centre | Public Health Wales Microbiology Cardiff |
| EPI_ISL_446609 | Wales Specialist Virology Centre | Public Health Wales Microbiology Cardiff |
| EPI_ISL_446610 | Wales Specialist Virology Centre | Public Health Wales Microbiology Cardiff |
| EPI_ISL_446611 | Wales Specialist Virology Centre | Public Health Wales Microbiology Cardiff |
| EPI_ISL_446612 | Wales Specialist Virology Centre | Public Health Wales Microbiology Cardiff |
| EPI_ISL_446613 | Wales Specialist Virology Centre | Public Health Wales Microbiology Cardiff |
| EPI_ISL_446614 | Wales Specialist Virology Centre | Public Health Wales Microbiology Cardiff |
| EPI_ISL_446615 | Wales Specialist Virology Centre | Public Health Wales Microbiology Cardiff |
| EPI_ISL_446616 | Wales Specialist Virology Centre | Public Health Wales Microbiology Cardiff |
| EPI_ISL_446617 | Wales Specialist Virology Centre | Public Health Wales Microbiology Cardiff |
| EPI_ISL_446618 | Wales Specialist Virology Centre | Public Health Wales Microbiology Cardiff |

| EPI_Accession_ID | Originating Laboratory | Submitting Laboratory |
|---|---|---|
| EPI_ISL_446619 | Wales Specialist Virology Centre | Public Health Wales Microbiology Cardiff |
| EPI_ISL_446620 | Wales Specialist Virology Centre | Public Health Wales Microbiology Cardiff |
| EPI_ISL_446621 | Wales Specialist Virology Centre | Public Health Wales Microbiology Cardiff |
| EPI_ISL_446622 | Wales Specialist Virology Centre | Public Health Wales Microbiology Cardiff |
| EPI_ISL_446623 | Wales Specialist Virology Centre | Public Health Wales Microbiology Cardiff |
| EPI_ISL_446624 | Wales Specialist Virology Centre | Public Health Wales Microbiology Cardiff |
| EPI_ISL_446625 | Wales Specialist Virology Centre | Public Health Wales Microbiology Cardiff |
| EPI_ISL_446626 | Wales Specialist Virology Centre | Public Health Wales Microbiology Cardiff |
| EPI_ISL_446627 | Wales Specialist Virology Centre | Public Health Wales Microbiology Cardiff |
| EPI_ISL_446628 | Wales Specialist Virology Centre | Public Health Wales Microbiology Cardiff |
| EPI_ISL_446629 | Wales Specialist Virology Centre | Public Health Wales Microbiology Cardiff |
| EPI_ISL_446630 | Wales Specialist Virology Centre | Public Health Wales Microbiology Cardiff |
| EPI_ISL_446631 | Wales Specialist Virology Centre | Public Health Wales Microbiology Cardiff |
| EPI_ISL_446632 | Wales Specialist Virology Centre | Public Health Wales Microbiology Cardiff |
| EPI_ISL_446633 | Wales Specialist Virology Centre | Public Health Wales Microbiology Cardiff |
| EPI_ISL_446634 | Wales Specialist Virology Centre | Public Health Wales Microbiology Cardiff |
| EPI_ISL_446635 | Wales Specialist Virology Centre | Public Health Wales Microbiology Cardiff |
| EPI_ISL_446636 | Wales Specialist Virology Centre | Public Health Wales Microbiology Cardiff |
| EPI_ISL_446637 | Wales Specialist Virology Centre | Public Health Wales Microbiology Cardiff |
| EPI_ISL_446638 | Wales Specialist Virology Centre | Public Health Wales Microbiology Cardiff |
| EPI_ISL_446639 | Wales Specialist Virology Centre | Public Health Wales Microbiology Cardiff |
| EPI_ISL_446640 | Wales Specialist Virology Centre | Public Health Wales Microbiology Cardiff |
| EPI_ISL_446641 | Wales Specialist Virology Centre | Public Health Wales Microbiology Cardiff |
| EPI_ISL_446642 | Wales Specialist Virology Centre | Public Health Wales Microbiology Cardiff |
| EPI_ISL_446643 | Wales Specialist Virology Centre | Public Health Wales Microbiology Cardiff |
| EPI_ISL_446644 | Wales Specialist Virology Centre | Public Health Wales Microbiology Cardiff |
| EPI_ISL_446645 | Wales Specialist Virology Centre | Public Health Wales Microbiology Cardiff |
| EPI_ISL_446646 | Wales Specialist Virology Centre | Public Health Wales Microbiology Cardiff |
| EPI_ISL_446647 | Wales Specialist Virology Centre | Public Health Wales Microbiology Cardiff |
| EPI_ISL_446648 | Wales Specialist Virology Centre | Public Health Wales Microbiology Cardiff |
| EPI_ISL_446649 | Wales Specialist Virology Centre | Public Health Wales Microbiology Cardiff |
| EPI_ISL_446650 | Wales Specialist Virology Centre | Public Health Wales Microbiology Cardiff |
| EPI_ISL_446651 | Wales Specialist Virology Centre | Public Health Wales Microbiology Cardiff |

| EPI_Accession_ID | Originating Laboratory | Submitting Laboratory |
|---|---|---|
| EPI_ISL_446652 | Wales Specialist Virology Centre | Public Health Wales Microbiology Cardiff |
| EPI_ISL_446653 | Wales Specialist Virology Centre | Public Health Wales Microbiology Cardiff |
| EPI_ISL_446654 | Wales Specialist Virology Centre | Public Health Wales Microbiology Cardiff |
| EPI_ISL_446655 | Wales Specialist Virology Centre | Public Health Wales Microbiology Cardiff |
| EPI_ISL_446656 | Wales Specialist Virology Centre | Public Health Wales Microbiology Cardiff |
| EPI_ISL_446657 | Wales Specialist Virology Centre | Public Health Wales Microbiology Cardiff |
| EPI_ISL_446658 | Wales Specialist Virology Centre | Public Health Wales Microbiology Cardiff |
| EPI_ISL_446659 | Wales Specialist Virology Centre | Public Health Wales Microbiology Cardiff |
| EPI_ISL_446660 | Wales Specialist Virology Centre | Public Health Wales Microbiology Cardiff |
| EPI_ISL_446661 | Wales Specialist Virology Centre | Public Health Wales Microbiology Cardiff |
| EPI_ISL_446662 | Wales Specialist Virology Centre | Public Health Wales Microbiology Cardiff |
| EPI_ISL_446663 | Wales Specialist Virology Centre | Public Health Wales Microbiology Cardiff |
| EPI_ISL_446664 | Wales Specialist Virology Centre | Public Health Wales Microbiology Cardiff |
| EPI_ISL_446665 | Wales Specialist Virology Centre | Public Health Wales Microbiology Cardiff |
| EPI_ISL_446666 | Wales Specialist Virology Centre | Public Health Wales Microbiology Cardiff |
| EPI_ISL_446667 | Wales Specialist Virology Centre | Public Health Wales Microbiology Cardiff |
| EPI_ISL_446668 | Wales Specialist Virology Centre | Public Health Wales Microbiology Cardiff |
| EPI_ISL_446670 | Wales Specialist Virology Centre | Public Health Wales Microbiology Cardiff |
| EPI_ISL_446671 | Wales Specialist Virology Centre | Public Health Wales Microbiology Cardiff |
| EPI_ISL_446672 | Wales Specialist Virology Centre | Public Health Wales Microbiology Cardiff |
| EPI_ISL_446673 | Wales Specialist Virology Centre | Public Health Wales Microbiology Cardiff |
| EPI_ISL_446675 | Wales Specialist Virology Centre | Public Health Wales Microbiology Cardiff |
| EPI_ISL_446676 | Wales Specialist Virology Centre | Public Health Wales Microbiology Cardiff |
| EPI_ISL_446677 | Wales Specialist Virology Centre | Public Health Wales Microbiology Cardiff |
| EPI_ISL_446678 | Wales Specialist Virology Centre | Public Health Wales Microbiology Cardiff |
| EPI_ISL_446679 | Wales Specialist Virology Centre | Public Health Wales Microbiology Cardiff |
| EPI_ISL_446680 | Wales Specialist Virology Centre | Public Health Wales Microbiology Cardiff |
| EPI_ISL_446681 | Wales Specialist Virology Centre | Public Health Wales Microbiology Cardiff |
| EPI_ISL_446682 | Wales Specialist Virology Centre | Public Health Wales Microbiology Cardiff |
| EPI_ISL_446683 | Wales Specialist Virology Centre | Public Health Wales Microbiology Cardiff |
| EPI_ISL_446684 | Wales Specialist Virology Centre | Public Health Wales Microbiology Cardiff |
| EPI_ISL_446686 | Wales Specialist Virology Centre | Public Health Wales Microbiology Cardiff |
| EPI_ISL_446687 | Wales Specialist Virology Centre | Public Health Wales Microbiology Cardiff |

| EPI_Accession_ID | Originating Laboratory | Submitting Laboratory |
|---|---|---|
| EPI_ISL_446688 | Wales Specialist Virology Centre | Public Health Wales Microbiology Cardiff |
| EPI_ISL_446689 | Wales Specialist Virology Centre | Public Health Wales Microbiology Cardiff |
| EPI_ISL_446690 | Wales Specialist Virology Centre | Public Health Wales Microbiology Cardiff |
| EPI_ISL_446691 | Wales Specialist Virology Centre | Public Health Wales Microbiology Cardiff |
| EPI_ISL_446692 | Wales Specialist Virology Centre | Public Health Wales Microbiology Cardiff |
| EPI_ISL_446693 | Wales Specialist Virology Centre | Public Health Wales Microbiology Cardiff |
| EPI_ISL_446694 | Wales Specialist Virology Centre | Public Health Wales Microbiology Cardiff |
| EPI_ISL_446695 | Wales Specialist Virology Centre | Public Health Wales Microbiology Cardiff |
| EPI_ISL_446696 | Wales Specialist Virology Centre | Public Health Wales Microbiology Cardiff |
| EPI_ISL_446697 | Wales Specialist Virology Centre | Public Health Wales Microbiology Cardiff |
| EPI_ISL_446698 | Wales Specialist Virology Centre | Public Health Wales Microbiology Cardiff |
| EPI_ISL_446699 | Wales Specialist Virology Centre | Public Health Wales Microbiology Cardiff |
| EPI_ISL_446700 | Wales Specialist Virology Centre | Public Health Wales Microbiology Cardiff |
| EPI_ISL_446701 | Wales Specialist Virology Centre | Public Health Wales Microbiology Cardiff |
| EPI_ISL_446702 | Wales Specialist Virology Centre | Public Health Wales Microbiology Cardiff |
| EPI_ISL_446703 | Wales Specialist Virology Centre | Public Health Wales Microbiology Cardiff |
| EPI_ISL_446704 | Wales Specialist Virology Centre | Public Health Wales Microbiology Cardiff |
| EPI_ISL_446705 | Wales Specialist Virology Centre | Public Health Wales Microbiology Cardiff |
| EPI_ISL_446706 | Wales Specialist Virology Centre | Public Health Wales Microbiology Cardiff |
| EPI_ISL_446707 | Wales Specialist Virology Centre | Public Health Wales Microbiology Cardiff |
| EPI_ISL_446708 | Wales Specialist Virology Centre | Public Health Wales Microbiology Cardiff |
| EPI_ISL_446709 | Wales Specialist Virology Centre | Public Health Wales Microbiology Cardiff |
| EPI_ISL_446710 | Wales Specialist Virology Centre | Public Health Wales Microbiology Cardiff |
| EPI_ISL_446711 | Wales Specialist Virology Centre | Public Health Wales Microbiology Cardiff |
| EPI_ISL_446712 | Wales Specialist Virology Centre | Public Health Wales Microbiology Cardiff |
| EPI_ISL_446713 | Wales Specialist Virology Centre | Public Health Wales Microbiology Cardiff |
| EPI_ISL_446714 | Wales Specialist Virology Centre | Public Health Wales Microbiology Cardiff |
| EPI_ISL_446715 | Wales Specialist Virology Centre | Public Health Wales Microbiology Cardiff |
| EPI_ISL_446716 | Wales Specialist Virology Centre | Public Health Wales Microbiology Cardiff |
| EPI_ISL_446717 | Wales Specialist Virology Centre | Public Health Wales Microbiology Cardiff |
| EPI_ISL_446718 | Wales Specialist Virology Centre | Public Health Wales Microbiology Cardiff |
| EPI_ISL_446719 | Wales Specialist Virology Centre | Public Health Wales Microbiology Cardiff |
| EPI_ISL_446720 | Wales Specialist Virology Centre | Public Health Wales Microbiology Cardiff |

| EPI_Accession_ID | Originating Laboratory | Submitting Laboratory |
|---|---|---|
| EPI_ISL_446721 | Wales Specialist Virology Centre | Public Health Wales Microbiology Cardiff |
| EPI_ISL_446722 | Wales Specialist Virology Centre | Public Health Wales Microbiology Cardiff |
| EPI_ISL_446723 | Wales Specialist Virology Centre | Public Health Wales Microbiology Cardiff |
| EPI_ISL_446724 | Wales Specialist Virology Centre | Public Health Wales Microbiology Cardiff |
| EPI_ISL_446725 | Wales Specialist Virology Centre | Public Health Wales Microbiology Cardiff |
| EPI_ISL_446726 | Wales Specialist Virology Centre | Public Health Wales Microbiology Cardiff |
| EPI_ISL_446727 | Wales Specialist Virology Centre | Public Health Wales Microbiology Cardiff |
| EPI_ISL_446728 | Wales Specialist Virology Centre | Public Health Wales Microbiology Cardiff |
| EPI_ISL_446729 | Wales Specialist Virology Centre | Public Health Wales Microbiology Cardiff |
| EPI_ISL_446730 | Wales Specialist Virology Centre | Public Health Wales Microbiology Cardiff |
| EPI_ISL_446731 | Wales Specialist Virology Centre | Public Health Wales Microbiology Cardiff |
| EPI_ISL_446732 | Wales Specialist Virology Centre | Public Health Wales Microbiology Cardiff |
| EPI_ISL_446733 | Wales Specialist Virology Centre | Public Health Wales Microbiology Cardiff |
| EPI_ISL_446734 | Wales Specialist Virology Centre | Public Health Wales Microbiology Cardiff |
| EPI_ISL_446735 | Wales Specialist Virology Centre | Public Health Wales Microbiology Cardiff |
| EPI_ISL_446736 | Wales Specialist Virology Centre | Public Health Wales Microbiology Cardiff |
| EPI_ISL_446737 | Wales Specialist Virology Centre | Public Health Wales Microbiology Cardiff |
| EPI_ISL_446738 | Wales Specialist Virology Centre | Public Health Wales Microbiology Cardiff |
| EPI_ISL_446739 | Wales Specialist Virology Centre | Public Health Wales Microbiology Cardiff |
| EPI_ISL_446740 | Wales Specialist Virology Centre | Public Health Wales Microbiology Cardiff |
| EPI_ISL_446741 | Wales Specialist Virology Centre | Public Health Wales Microbiology Cardiff |
| EPI_ISL_446742 | Wales Specialist Virology Centre | Public Health Wales Microbiology Cardiff |
| EPI_ISL_446743 | Wales Specialist Virology Centre | Public Health Wales Microbiology Cardiff |
| EPI_ISL_446744 | Wales Specialist Virology Centre | Public Health Wales Microbiology Cardiff |
| EPI_ISL_446745 | Wales Specialist Virology Centre | Public Health Wales Microbiology Cardiff |
| EPI_ISL_446746 | Wales Specialist Virology Centre | Public Health Wales Microbiology Cardiff |
| EPI_ISL_446747 | Wales Specialist Virology Centre | Public Health Wales Microbiology Cardiff |
| EPI_ISL_446748 | Wales Specialist Virology Centre | Public Health Wales Microbiology Cardiff |
| EPI_ISL_446749 | Wales Specialist Virology Centre | Public Health Wales Microbiology Cardiff |
| EPI_ISL_446750 | Wales Specialist Virology Centre | Public Health Wales Microbiology Cardiff |
| EPI_ISL_446751 | Wales Specialist Virology Centre | Public Health Wales Microbiology Cardiff |
| EPI_ISL_446752 | Wales Specialist Virology Centre | Public Health Wales Microbiology Cardiff |
| EPI_ISL_446753 | Wales Specialist Virology Centre | Public Health Wales Microbiology Cardiff |

| EPI_Accession_ID | Originating Laboratory | Submitting Laboratory |
|---|---|---|
| EPI_ISL_446754 | Wales Specialist Virology Centre | Public Health Wales Microbiology Cardiff |
| EPI_ISL_446755 | Wales Specialist Virology Centre | Public Health Wales Microbiology Cardiff |
| EPI_ISL_446756 | Wales Specialist Virology Centre | Public Health Wales Microbiology Cardiff |
| EPI_ISL_446757 | Wales Specialist Virology Centre | Public Health Wales Microbiology Cardiff |
| EPI_ISL_446758 | Wales Specialist Virology Centre | Public Health Wales Microbiology Cardiff |
| EPI_ISL_446759 | Wales Specialist Virology Centre | Public Health Wales Microbiology Cardiff |
| EPI_ISL_446760 | Wales Specialist Virology Centre | Public Health Wales Microbiology Cardiff |
| EPI_ISL_446761 | Wales Specialist Virology Centre | Public Health Wales Microbiology Cardiff |
| EPI_ISL_446762 | Wales Specialist Virology Centre | Public Health Wales Microbiology Cardiff |
| EPI_ISL_446763 | Wales Specialist Virology Centre | Public Health Wales Microbiology Cardiff |
| EPI_ISL_446764 | Wales Specialist Virology Centre | Public Health Wales Microbiology Cardiff |
| EPI_ISL_446765 | Wales Specialist Virology Centre | Public Health Wales Microbiology Cardiff |
| EPI_ISL_446766 | Wales Specialist Virology Centre | Public Health Wales Microbiology Cardiff |
| EPI_ISL_446767 | Wales Specialist Virology Centre | Public Health Wales Microbiology Cardiff |
| EPI_ISL_446768 | Wales Specialist Virology Centre | Public Health Wales Microbiology Cardiff |
| EPI_ISL_446769 | Wales Specialist Virology Centre | Public Health Wales Microbiology Cardiff |
| EPI_ISL_446770 | Wales Specialist Virology Centre | Public Health Wales Microbiology Cardiff |
| EPI_ISL_446771 | Wales Specialist Virology Centre | Public Health Wales Microbiology Cardiff |
| EPI_ISL_446772 | Wales Specialist Virology Centre | Public Health Wales Microbiology Cardiff |
| EPI_ISL_446773 | Wales Specialist Virology Centre | Public Health Wales Microbiology Cardiff |
| EPI_ISL_446774 | Wales Specialist Virology Centre | Public Health Wales Microbiology Cardiff |
| EPI_ISL_446775 | Wales Specialist Virology Centre | Public Health Wales Microbiology Cardiff |
| EPI_ISL_446776 | Wales Specialist Virology Centre | Public Health Wales Microbiology Cardiff |
| EPI_ISL_446777 | Wales Specialist Virology Centre | Public Health Wales Microbiology Cardiff |
| EPI_ISL_446778 | Wales Specialist Virology Centre | Public Health Wales Microbiology Cardiff |
| EPI_ISL_446779 | Wales Specialist Virology Centre | Public Health Wales Microbiology Cardiff |
| EPI_ISL_446780 | Wales Specialist Virology Centre | Public Health Wales Microbiology Cardiff |
| EPI_ISL_446781 | Wales Specialist Virology Centre | Public Health Wales Microbiology Cardiff |
| EPI_ISL_446782 | Wales Specialist Virology Centre | Public Health Wales Microbiology Cardiff |
| EPI_ISL_446783 | Wales Specialist Virology Centre | Public Health Wales Microbiology Cardiff |
| EPI_ISL_446784 | Wales Specialist Virology Centre | Public Health Wales Microbiology Cardiff |
| EPI_ISL_446785 | Wales Specialist Virology Centre | Public Health Wales Microbiology Cardiff |
| EPI_ISL_446786 | Wales Specialist Virology Centre | Public Health Wales Microbiology Cardiff |

| EPI_Accession_ID | Originating Laboratory | Submitting Laboratory |
|---|---|---|
| EPI_ISL_446787 | Wales Specialist Virology Centre | Public Health Wales Microbiology Cardiff |
| EPI_ISL_446788 | Wales Specialist Virology Centre | Public Health Wales Microbiology Cardiff |
| EPI_ISL_446789 | Wales Specialist Virology Centre | Public Health Wales Microbiology Cardiff |
| EPI_ISL_446790 | Wales Specialist Virology Centre | Public Health Wales Microbiology Cardiff |
| EPI_ISL_446791 | Wales Specialist Virology Centre | Public Health Wales Microbiology Cardiff |
| EPI_ISL_446792 | Wales Specialist Virology Centre | Public Health Wales Microbiology Cardiff |
| EPI_ISL_446793 | Wales Specialist Virology Centre | Public Health Wales Microbiology Cardiff |
| EPI_ISL_446794 | Wales Specialist Virology Centre | Public Health Wales Microbiology Cardiff |
| EPI_ISL_446795 | Wales Specialist Virology Centre | Public Health Wales Microbiology Cardiff |
| EPI_ISL_446796 | Wales Specialist Virology Centre | Public Health Wales Microbiology Cardiff |
| EPI_ISL_446797 | Wales Specialist Virology Centre | Public Health Wales Microbiology Cardiff |
| EPI_ISL_446798 | Wales Specialist Virology Centre | Public Health Wales Microbiology Cardiff |
| EPI_ISL_446799 | Wales Specialist Virology Centre | Public Health Wales Microbiology Cardiff |
| EPI_ISL_446800 | Wales Specialist Virology Centre | Public Health Wales Microbiology Cardiff |
| EPI_ISL_446801 | Wales Specialist Virology Centre | Public Health Wales Microbiology Cardiff |
| EPI_ISL_446802 | Wales Specialist Virology Centre | Public Health Wales Microbiology Cardiff |
| EPI_ISL_446803 | Wales Specialist Virology Centre | Public Health Wales Microbiology Cardiff |
| EPI_ISL_446804 | Wales Specialist Virology Centre | Public Health Wales Microbiology Cardiff |
| EPI_ISL_446805 | Wales Specialist Virology Centre | Public Health Wales Microbiology Cardiff |
| EPI_ISL_446806 | Wales Specialist Virology Centre | Public Health Wales Microbiology Cardiff |
| EPI_ISL_446807 | Wales Specialist Virology Centre | Public Health Wales Microbiology Cardiff |
| EPI_ISL_446808 | Wales Specialist Virology Centre | Public Health Wales Microbiology Cardiff |
| EPI_ISL_446809 | Wales Specialist Virology Centre | Public Health Wales Microbiology Cardiff |
| EPI_ISL_446810 | Wales Specialist Virology Centre | Public Health Wales Microbiology Cardiff |
| EPI_ISL_446811 | Wales Specialist Virology Centre | Public Health Wales Microbiology Cardiff |
| EPI_ISL_446812 | Wales Specialist Virology Centre | Public Health Wales Microbiology Cardiff |
| EPI_ISL_446813 | Wales Specialist Virology Centre | Public Health Wales Microbiology Cardiff |
| EPI_ISL_446814 | Wales Specialist Virology Centre | Public Health Wales Microbiology Cardiff |
| EPI_ISL_446815 | Wales Specialist Virology Centre | Public Health Wales Microbiology Cardiff |
| EPI_ISL_446816 | Wales Specialist Virology Centre | Public Health Wales Microbiology Cardiff |
| EPI_ISL_446817 | Wales Specialist Virology Centre | Public Health Wales Microbiology Cardiff |
| EPI_ISL_446818 | Wales Specialist Virology Centre | Public Health Wales Microbiology Cardiff |
| EPI_ISL_446819 | Wales Specialist Virology Centre | Public Health Wales Microbiology Cardiff |

| EPI_Accession_ID | Originating Laboratory | Submitting Laboratory |
|---|---|---|
| EPI_ISL_446820 | Wales Specialist Virology Centre | Public Health Wales Microbiology Cardiff |
| EPI_ISL_446821 | Wales Specialist Virology Centre | Public Health Wales Microbiology Cardiff |
| EPI_ISL_446822 | Wales Specialist Virology Centre | Public Health Wales Microbiology Cardiff |
| EPI_ISL_446823 | Wales Specialist Virology Centre | Public Health Wales Microbiology Cardiff |
| EPI_ISL_446824 | Wales Specialist Virology Centre | Public Health Wales Microbiology Cardiff |
| EPI_ISL_446825 | Wales Specialist Virology Centre | Public Health Wales Microbiology Cardiff |
| EPI_ISL_446826 | Wales Specialist Virology Centre | Public Health Wales Microbiology Cardiff |
| EPI_ISL_446827 | Wales Specialist Virology Centre | Public Health Wales Microbiology Cardiff |
| EPI_ISL_446828 | Wales Specialist Virology Centre | Public Health Wales Microbiology Cardiff |
| EPI_ISL_446829 | Wales Specialist Virology Centre | Public Health Wales Microbiology Cardiff |
| EPI_ISL_446830 | Wales Specialist Virology Centre | Public Health Wales Microbiology Cardiff |
| EPI_ISL_446831 | Wales Specialist Virology Centre | Public Health Wales Microbiology Cardiff |
| EPI_ISL_446832 | Wales Specialist Virology Centre | Public Health Wales Microbiology Cardiff |
| EPI_ISL_446833 | Wales Specialist Virology Centre | Public Health Wales Microbiology Cardiff |
| EPI_ISL_446834 | Wales Specialist Virology Centre | Public Health Wales Microbiology Cardiff |
| EPI_ISL_446835 | Wales Specialist Virology Centre | Public Health Wales Microbiology Cardiff |
| EPI_ISL_446836 | Wales Specialist Virology Centre | Public Health Wales Microbiology Cardiff |
| EPI_ISL_446837 | Wales Specialist Virology Centre | Public Health Wales Microbiology Cardiff |
| EPI_ISL_446838 | Wales Specialist Virology Centre | Public Health Wales Microbiology Cardiff |
| EPI_ISL_446839 | Wales Specialist Virology Centre | Public Health Wales Microbiology Cardiff |
| EPI_ISL_446840 | Wales Specialist Virology Centre | Public Health Wales Microbiology Cardiff |
| EPI_ISL_446841 | Wales Specialist Virology Centre | Public Health Wales Microbiology Cardiff |
| EPI_ISL_446842 | Wales Specialist Virology Centre | Public Health Wales Microbiology Cardiff |
| EPI_ISL_446843 | Wales Specialist Virology Centre | Public Health Wales Microbiology Cardiff |
| EPI_ISL_446844 | Wales Specialist Virology Centre | Public Health Wales Microbiology Cardiff |
| EPI_ISL_446845 | Wales Specialist Virology Centre | Public Health Wales Microbiology Cardiff |
| EPI_ISL_446846 | Wales Specialist Virology Centre | Public Health Wales Microbiology Cardiff |
| EPI_ISL_446847 | Wales Specialist Virology Centre | Public Health Wales Microbiology Cardiff |
| EPI_ISL_446848 | Wales Specialist Virology Centre | Public Health Wales Microbiology Cardiff |
| EPI_ISL_446849 | Wales Specialist Virology Centre | Public Health Wales Microbiology Cardiff |
| EPI_ISL_446850 | Wales Specialist Virology Centre | Public Health Wales Microbiology Cardiff |
| EPI_ISL_446851 | Wales Specialist Virology Centre | Public Health Wales Microbiology Cardiff |
| EPI_ISL_446852 | Wales Specialist Virology Centre | Public Health Wales Microbiology Cardiff |

| EPI_Accession_ID | Originating Laboratory | Submitting Laboratory |
|---|---|---|
| EPI_ISL_446853 | Wales Specialist Virology Centre | Public Health Wales Microbiology Cardiff |
| EPI_ISL_446854 | Wales Specialist Virology Centre | Public Health Wales Microbiology Cardiff |
| EPI_ISL_446855 | Wales Specialist Virology Centre | Public Health Wales Microbiology Cardiff |
| EPI_ISL_446856 | Wales Specialist Virology Centre | Public Health Wales Microbiology Cardiff |
| EPI_ISL_446857 | Wales Specialist Virology Centre | Public Health Wales Microbiology Cardiff |
| EPI_ISL_446858 | Wales Specialist Virology Centre | Public Health Wales Microbiology Cardiff |
| EPI_ISL_446859 | Wales Specialist Virology Centre | Public Health Wales Microbiology Cardiff |
| EPI_ISL_446860 | Wales Specialist Virology Centre | Public Health Wales Microbiology Cardiff |
| EPI_ISL_446861 | Wales Specialist Virology Centre | Public Health Wales Microbiology Cardiff |
| EPI_ISL_446862 | Wales Specialist Virology Centre | Public Health Wales Microbiology Cardiff |
| EPI_ISL_446863 | Wales Specialist Virology Centre | Public Health Wales Microbiology Cardiff |
| EPI_ISL_446864 | Wales Specialist Virology Centre | Public Health Wales Microbiology Cardiff |
| EPI_ISL_446865 | Wales Specialist Virology Centre | Public Health Wales Microbiology Cardiff |
| EPI_ISL_446866 | Wales Specialist Virology Centre | Public Health Wales Microbiology Cardiff |
| EPI_ISL_446867 | Wales Specialist Virology Centre | Public Health Wales Microbiology Cardiff |
| EPI_ISL_446868 | Wales Specialist Virology Centre | Public Health Wales Microbiology Cardiff |
| EPI_ISL_446869 | Wales Specialist Virology Centre | Public Health Wales Microbiology Cardiff |
| EPI_ISL_446870 | Wales Specialist Virology Centre | Public Health Wales Microbiology Cardiff |
| EPI_ISL_446871 | Wales Specialist Virology Centre | Public Health Wales Microbiology Cardiff |
| EPI_ISL_446872 | Wales Specialist Virology Centre | Public Health Wales Microbiology Cardiff |
| EPI_ISL_446873 | Wales Specialist Virology Centre | Public Health Wales Microbiology Cardiff |
| EPI_ISL_446874 | Wales Specialist Virology Centre | Public Health Wales Microbiology Cardiff |
| EPI_ISL_446875 | Wales Specialist Virology Centre | Public Health Wales Microbiology Cardiff |
| EPI_ISL_446876 | Wales Specialist Virology Centre | Public Health Wales Microbiology Cardiff |
| EPI_ISL_446877 | Wales Specialist Virology Centre | Public Health Wales Microbiology Cardiff |
| EPI_ISL_446878 | Wales Specialist Virology Centre | Public Health Wales Microbiology Cardiff |
| EPI_ISL_446879 | Wales Specialist Virology Centre | Public Health Wales Microbiology Cardiff |
| EPI_ISL_446880 | Wales Specialist Virology Centre | Public Health Wales Microbiology Cardiff |
| EPI_ISL_446881 | Wales Specialist Virology Centre | Public Health Wales Microbiology Cardiff |
| EPI_ISL_446882 | Wales Specialist Virology Centre | Public Health Wales Microbiology Cardiff |
| EPI_ISL_446883 | Wales Specialist Virology Centre | Public Health Wales Microbiology Cardiff |
| EPI_ISL_446884 | Wales Specialist Virology Centre | Public Health Wales Microbiology Cardiff |
| EPI_ISL_446885 | Wales Specialist Virology Centre | Public Health Wales Microbiology Cardiff |

| EPI_Accession_ID | Originating Laboratory | Submitting Laboratory |
|---|---|---|
| EPI_ISL_446886 | Wales Specialist Virology Centre | Public Health Wales Microbiology Cardiff |
| EPI_ISL_446887 | Wales Specialist Virology Centre | Public Health Wales Microbiology Cardiff |
| EPI_ISL_446888 | Wales Specialist Virology Centre | Public Health Wales Microbiology Cardiff |
| EPI_ISL_446889 | Wales Specialist Virology Centre | Public Health Wales Microbiology Cardiff |
| EPI_ISL_446890 | Wales Specialist Virology Centre | Public Health Wales Microbiology Cardiff |
| EPI_ISL_446891 | Wales Specialist Virology Centre | Public Health Wales Microbiology Cardiff |
| EPI_ISL_446892 | Wales Specialist Virology Centre | Public Health Wales Microbiology Cardiff |
| EPI_ISL_446893 | Wales Specialist Virology Centre | Public Health Wales Microbiology Cardiff |
| EPI_ISL_446894 | Wales Specialist Virology Centre | Public Health Wales Microbiology Cardiff |
| EPI_ISL_446895 | Wales Specialist Virology Centre | Public Health Wales Microbiology Cardiff |
| EPI_ISL_446896 | Wales Specialist Virology Centre | Public Health Wales Microbiology Cardiff |
| EPI_ISL_446897 | Wales Specialist Virology Centre | Public Health Wales Microbiology Cardiff |
| EPI_ISL_446898 | Wales Specialist Virology Centre | Public Health Wales Microbiology Cardiff |
| EPI_ISL_446899 | Wales Specialist Virology Centre | Public Health Wales Microbiology Cardiff |
| EPI_ISL_446900 | Wales Specialist Virology Centre | Public Health Wales Microbiology Cardiff |
| EPI_ISL_446901 | Wales Specialist Virology Centre | Public Health Wales Microbiology Cardiff |
| EPI_ISL_446902 | Wales Specialist Virology Centre | Public Health Wales Microbiology Cardiff |
| EPI_ISL_446903 | Wales Specialist Virology Centre | Public Health Wales Microbiology Cardiff |
| EPI_ISL_446904 | Wales Specialist Virology Centre | Public Health Wales Microbiology Cardiff |
| EPI_ISL_446905 | Wales Specialist Virology Centre | Public Health Wales Microbiology Cardiff |
| EPI_ISL_446906 | Wales Specialist Virology Centre | Public Health Wales Microbiology Cardiff |
| EPI_ISL_446907 | Wales Specialist Virology Centre | Public Health Wales Microbiology Cardiff |
| EPI_ISL_446908 | Wales Specialist Virology Centre | Public Health Wales Microbiology Cardiff |
| EPI_ISL_446909 | Wales Specialist Virology Centre | Public Health Wales Microbiology Cardiff |
| EPI_ISL_446910 | Wales Specialist Virology Centre | Public Health Wales Microbiology Cardiff |
| EPI_ISL_446911 | Wales Specialist Virology Centre | Public Health Wales Microbiology Cardiff |
| EPI_ISL_446912 | Wales Specialist Virology Centre | Public Health Wales Microbiology Cardiff |
| EPI_ISL_446913 | Wales Specialist Virology Centre | Public Health Wales Microbiology Cardiff |
| EPI_ISL_446914 | Wales Specialist Virology Centre | Public Health Wales Microbiology Cardiff |
| EPI_ISL_446915 | Wales Specialist Virology Centre | Public Health Wales Microbiology Cardiff |
| EPI_ISL_446916 | Wales Specialist Virology Centre | Public Health Wales Microbiology Cardiff |
| EPI_ISL_446917 | Wales Specialist Virology Centre | Public Health Wales Microbiology Cardiff |
| EPI_ISL_446918 | Wales Specialist Virology Centre | Public Health Wales Microbiology Cardiff |

| EPI_Accession_ID | Originating Laboratory | Submitting Laboratory |
|---|---|---|
| EPI_ISL_446919 | Wales Specialist Virology Centre | Public Health Wales Microbiology Cardiff |
| EPI_ISL_446920 | Wales Specialist Virology Centre | Public Health Wales Microbiology Cardiff |
| EPI_ISL_446921 | Wales Specialist Virology Centre | Public Health Wales Microbiology Cardiff |
| EPI_ISL_446922 | Wales Specialist Virology Centre | Public Health Wales Microbiology Cardiff |
| EPI_ISL_446923 | Wales Specialist Virology Centre | Public Health Wales Microbiology Cardiff |
| EPI_ISL_446924 | Wales Specialist Virology Centre | Public Health Wales Microbiology Cardiff |
| EPI_ISL_446925 | Wales Specialist Virology Centre | Public Health Wales Microbiology Cardiff |
| EPI_ISL_446926 | Wales Specialist Virology Centre | Public Health Wales Microbiology Cardiff |
| EPI_ISL_446927 | Wales Specialist Virology Centre | Public Health Wales Microbiology Cardiff |
| EPI_ISL_446928 | Wales Specialist Virology Centre | Public Health Wales Microbiology Cardiff |
| EPI_ISL_446929 | Wales Specialist Virology Centre | Public Health Wales Microbiology Cardiff |
| EPI_ISL_446930 | Wales Specialist Virology Centre | Public Health Wales Microbiology Cardiff |
| EPI_ISL_446931 | Wales Specialist Virology Centre | Public Health Wales Microbiology Cardiff |
| EPI_ISL_446932 | Wales Specialist Virology Centre | Public Health Wales Microbiology Cardiff |
| EPI_ISL_446933 | Wales Specialist Virology Centre | Public Health Wales Microbiology Cardiff |
| EPI_ISL_446934 | Wales Specialist Virology Centre | Public Health Wales Microbiology Cardiff |
| EPI_ISL_446935 | Wales Specialist Virology Centre | Public Health Wales Microbiology Cardiff |
| EPI_ISL_446936 | Wales Specialist Virology Centre | Public Health Wales Microbiology Cardiff |
| EPI_ISL_446937 | Wales Specialist Virology Centre | Public Health Wales Microbiology Cardiff |
| EPI_ISL_446938 | Wales Specialist Virology Centre | Public Health Wales Microbiology Cardiff |
| EPI_ISL_446939 | Wales Specialist Virology Centre | Public Health Wales Microbiology Cardiff |
| EPI_ISL_446940 | Wales Specialist Virology Centre | Public Health Wales Microbiology Cardiff |
| EPI_ISL_446941 | Wales Specialist Virology Centre | Public Health Wales Microbiology Cardiff |
| EPI_ISL_446942 | Wales Specialist Virology Centre | Public Health Wales Microbiology Cardiff |
| EPI_ISL_446943 | Wales Specialist Virology Centre | Public Health Wales Microbiology Cardiff |
| EPI_ISL_446944 | Wales Specialist Virology Centre | Public Health Wales Microbiology Cardiff |
| EPI_ISL_446946 | Wales Specialist Virology Centre | Public Health Wales Microbiology Cardiff |
| EPI_ISL_446947 | Wales Specialist Virology Centre | Public Health Wales Microbiology Cardiff |
| EPI_ISL_446948 | Wales Specialist Virology Centre | Public Health Wales Microbiology Cardiff |
| EPI_ISL_446949 | Wales Specialist Virology Centre | Public Health Wales Microbiology Cardiff |
| EPI_ISL_446950 | Wales Specialist Virology Centre | Public Health Wales Microbiology Cardiff |
| EPI_ISL_446951 | Wales Specialist Virology Centre | Public Health Wales Microbiology Cardiff |
| EPI_ISL_446952 | Wales Specialist Virology Centre | Public Health Wales Microbiology Cardiff |

| EPI_Accession_ID | Originating Laboratory | Submitting Laboratory |
| --- | --- | --- |
| EPI_ISL_446953 | Wales Specialist Virology Centre | Public Health Wales Microbiology Cardiff |
| EPI_ISL_446954 | Wales Specialist Virology Centre | Public Health Wales Microbiology Cardiff |
| EPI_ISL_446955 | Wales Specialist Virology Centre | Public Health Wales Microbiology Cardiff |
| EPI_ISL_446956 | Wales Specialist Virology Centre | Public Health Wales Microbiology Cardiff |
| EPI_ISL_446957 | Wales Specialist Virology Centre | Public Health Wales Microbiology Cardiff |
| EPI_ISL_446958 | Wales Specialist Virology Centre | Public Health Wales Microbiology Cardiff |
| EPI_ISL_446959 | Wales Specialist Virology Centre | Public Health Wales Microbiology Cardiff |
| EPI_ISL_446960 | Wales Specialist Virology Centre | Public Health Wales Microbiology Cardiff |
| EPI_ISL_446961 | Wales Specialist Virology Centre | Public Health Wales Microbiology Cardiff |
| EPI_ISL_446962 | Wales Specialist Virology Centre | Public Health Wales Microbiology Cardiff |
| EPI_ISL_446963 | Wales Specialist Virology Centre | Public Health Wales Microbiology Cardiff |
| EPI_ISL_446964 | Wales Specialist Virology Centre | Public Health Wales Microbiology Cardiff |
| EPI_ISL_446965 | Wales Specialist Virology Centre | Public Health Wales Microbiology Cardiff |
| EPI_ISL_446966 | Wales Specialist Virology Centre | Public Health Wales Microbiology Cardiff |
| EPI_ISL_446967 | Wales Specialist Virology Centre | Public Health Wales Microbiology Cardiff |
| EPI_ISL_446968 | Wales Specialist Virology Centre | Public Health Wales Microbiology Cardiff |
| EPI_ISL_446969 | Wales Specialist Virology Centre | Public Health Wales Microbiology Cardiff |
| EPI_ISL_446970 | Wales Specialist Virology Centre | Public Health Wales Microbiology Cardiff |
| EPI_ISL_446971 | Wales Specialist Virology Centre | Public Health Wales Microbiology Cardiff |
| EPI_ISL_446972 | Wales Specialist Virology Centre | Public Health Wales Microbiology Cardiff |
| EPI_ISL_446973 | Wales Specialist Virology Centre | Public Health Wales Microbiology Cardiff |
| EPI_ISL_446974 | Wales Specialist Virology Centre | Public Health Wales Microbiology Cardiff |
| EPI_ISL_446975 | Wales Specialist Virology Centre | Public Health Wales Microbiology Cardiff |
| EPI_ISL_446976 | Wales Specialist Virology Centre | Public Health Wales Microbiology Cardiff |
| EPI_ISL_446977 | Wales Specialist Virology Centre | Public Health Wales Microbiology Cardiff |
| EPI_ISL_446978 | Wales Specialist Virology Centre | Public Health Wales Microbiology Cardiff |
| EPI_ISL_446979 | Wales Specialist Virology Centre | Public Health Wales Microbiology Cardiff |
| EPI_ISL_446980 | Wales Specialist Virology Centre | Public Health Wales Microbiology Cardiff |
| EPI_ISL_446981 | Wales Specialist Virology Centre | Public Health Wales Microbiology Cardiff |
| EPI_ISL_446982 | Wales Specialist Virology Centre | Public Health Wales Microbiology Cardiff |
| EPI_ISL_446983 | Wales Specialist Virology Centre | Public Health Wales Microbiology Cardiff |
| EPI_ISL_446984 | Wales Specialist Virology Centre | Public Health Wales Microbiology Cardiff |
| EPI_ISL_446985 | Wales Specialist Virology Centre | Public Health Wales Microbiology Cardiff |

| EPI_Accession_ID | Originating Laboratory | Submitting Laboratory |
|---|---|---|
| EPI_ISL_446986 | Wales Specialist Virology Centre | Public Health Wales Microbiology Cardiff |
| EPI_ISL_446987 | Wales Specialist Virology Centre | Public Health Wales Microbiology Cardiff |
| EPI_ISL_446988 | Wales Specialist Virology Centre | Public Health Wales Microbiology Cardiff |
| EPI_ISL_446989 | Wales Specialist Virology Centre | Public Health Wales Microbiology Cardiff |
| EPI_ISL_446990 | Wales Specialist Virology Centre | Public Health Wales Microbiology Cardiff |
| EPI_ISL_446991 | Wales Specialist Virology Centre | Public Health Wales Microbiology Cardiff |
| EPI_ISL_446992 | Wales Specialist Virology Centre | Public Health Wales Microbiology Cardiff |
| EPI_ISL_446993 | Wales Specialist Virology Centre | Public Health Wales Microbiology Cardiff |
| EPI_ISL_446994 | Wales Specialist Virology Centre | Public Health Wales Microbiology Cardiff |
| EPI_ISL_446995 | Wales Specialist Virology Centre | Public Health Wales Microbiology Cardiff |
| EPI_ISL_446996 | Ramathibodi Hospital | COVID-19 Network Investigations (CONI) Alliance |
| EPI_ISL_446997 | Ramathibodi Hospital | COVID-19 Network Investigations (CONI) Alliance |
| EPI_ISL_446998 | Ramathibodi Hospital | COVID-19 Network Investigations (CONI) Alliance |
| EPI_ISL_446999 | Ramathibodi Hospital | COVID-19 Network Investigations (CONI) Alliance |
| EPI_ISL_447000 | Ramathibodi Hospital | COVID-19 Network Investigations (CONI) Alliance |
| EPI_ISL_447001 | Ramathibodi Hospital | COVID-19 Network Investigations (CONI) Alliance |
| EPI_ISL_447002 | Ramathibodi Hospital | COVID-19 Network Investigations (CONI) Alliance |
| EPI_ISL_447003 | Ramathibodi Hospital | COVID-19 Network Investigations (CONI) Alliance |
| EPI_ISL_447004 | Ramathibodi Hospital | COVID-19 Network Investigations (CONI) Alliance |
| EPI_ISL_447005 | Ramathibodi Hospital | COVID-19 Network Investigations (CONI) Alliance |
| EPI_ISL_447006 | Ramathibodi Hospital | COVID-19 Network Investigations (CONI) Alliance |
| EPI_ISL_447007 | Ramathibodi Hospital | COVID-19 Network Investigations (CONI) Alliance |
| EPI_ISL_447008 | Ramathibodi Hospital | COVID-19 Network Investigations (CONI) Alliance |
| EPI_ISL_447009 | Ramathibodi Hospital | COVID-19 Network Investigations (CONI) Alliance |
| EPI_ISL_447010 | Ramathibodi Hospital | COVID-19 Network Investigations (CONI) Alliance |
| EPI_ISL_447011 | Ramathibodi Hospital | COVID-19 Network Investigations (CONI) Alliance |
| EPI_ISL_447012 | Ramathibodi Hospital | COVID-19 Network Investigations (CONI) Alliance |
| EPI_ISL_447013 | Ramathibodi Hospital | COVID-19 Network Investigations (CONI) Alliance |
| EPI_ISL_447014 | Ramathibodi Hospital | COVID-19 Network Investigations (CONI) Alliance |
| EPI_ISL_447015 | Ramathibodi Hospital | COVID-19 Network Investigations (CONI) Alliance |
| EPI_ISL_447016 | Ramathibodi Hospital | COVID-19 Network Investigations (CONI) Alliance |
| EPI_ISL_447017 | Ramathibodi Hospital | COVID-19 Network Investigations (CONI) Alliance |
| EPI_ISL_447018 | Ramathibodi Hospital | COVID-19 Network Investigations (CONI) Alliance |

| EPI_Accession_ID | Originating Laboratory | Submitting Laboratory |
|---|---|---|
| EPI_ISL_447019 | Ramathibodi Hospital | COVID-19 Network Investigations (CONI) Alliance |
| EPI_ISL_447020 | Ramathibodi Hospital | COVID-19 Network Investigations (CONI) Alliance |
| EPI_ISL_447021 | Ramathibodi Hospital | COVID-19 Network Investigations (CONI) Alliance |
| EPI_ISL_447022 | Ramathibodi Hospital | COVID-19 Network Investigations (CONI) Alliance |
| EPI_ISL_447023 | Ramathibodi Hospital | COVID-19 Network Investigations (CONI) Alliance |
| EPI_ISL_447024 | Ramathibodi Hospital | COVID-19 Network Investigations (CONI) Alliance |
| EPI_ISL_447025 | Ramathibodi Hospital | COVID-19 Network Investigations (CONI) Alliance |
| EPI_ISL_447026 | Ramathibodi Hospital | COVID-19 Network Investigations (CONI) Alliance |
| EPI_ISL_447027 | Ramathibodi Hospital | COVID-19 Network Investigations (CONI) Alliance |
| EPI_ISL_447028 | Ramathibodi Hospital | COVID-19 Network Investigations (CONI) Alliance |
| EPI_ISL_447029 | Ramathibodi Hospital | COVID-19 Network Investigations (CONI) Alliance |
| EPI_ISL_447030 | B.J. Medical College and Civil hospital | Gujarat Biotechnology Research Centre |
| EPI_ISL_447031 | B.J. Medical College and Civil hospital | Gujarat Biotechnology Research Centre |
| EPI_ISL_447032 | B.J. Medical College and Civil hospital | Gujarat Biotechnology Research Centre |
| EPI_ISL_447033 | B.J. Medical College and Civil hospital | Gujarat Biotechnology Research Centre |
| EPI_ISL_447034 | B.J. Medical College and Civil hospital | Gujarat Biotechnology Research Centre |
| EPI_ISL_447035 | B.J. Medical College and Civil hospital | Gujarat Biotechnology Research Centre |
| EPI_ISL_447036 | B.J. Medical College and Civil hospital | Gujarat Biotechnology Research Centre |
| EPI_ISL_447037 | B.J. Medical College and Civil hospital | Gujarat Biotechnology Research Centre |
| EPI_ISL_447038 | B.J. Medical College and Civil hospital | Gujarat Biotechnology Research Centre |
| EPI_ISL_447039 | B.J. Medical College and Civil hospital | Gujarat Biotechnology Research Centre |
| EPI_ISL_447040 | B.J. Medical College and Civil hospital | Gujarat Biotechnology Research Centre |
| EPI_ISL_447041 | B.J. Medical College and Civil hospital | Gujarat Biotechnology Research Centre |
| EPI_ISL_447042 | B.J. Medical College and Civil hospital | Gujarat Biotechnology Research Centre |
| EPI_ISL_447043 | B.J. Medical College and Civil hospital | Gujarat Biotechnology Research Centre |
| EPI_ISL_447044 | B.J. Medical College and Civil hospital | Gujarat Biotechnology Research Centre |
| EPI_ISL_447045 | B.J. Medical College and Civil hospital | Gujarat Biotechnology Research Centre |
| EPI_ISL_447046 | B.J. Medical College and Civil hospital | Gujarat Biotechnology Research Centre |
| EPI_ISL_447047 | GMERS Medical College and Hospital, Gandhinagar | Gujarat Biotechnology Research Centre |
| EPI_ISL_447048 | GMERS Medical College and Hospital, Gandhinagar | Gujarat Biotechnology Research Centre |
| EPI_ISL_447049 | GMERS Medical College and Hospital, Gandhinagar | Gujarat Biotechnology Research Centre |
| EPI_ISL_447050 | GMERS Medical College and Hospital, Gandhinagar | Gujarat Biotechnology Research Centre |
| EPI_ISL_447051 | GMERS Medical College and Hospital, Gandhinagar | Gujarat Biotechnology Research Centre |

| EPI_Accession_ID | Originating Laboratory | Submitting Laboratory |
| --- | --- | --- |
| EPI_ISL_447052 | GMERS Medical College and Hospital, Gandhinagar | Gujarat Biotechnology Research Centre |
| EPI_ISL_447053 | GMERS Medical College and Hospital, Gandhinagar | Gujarat Biotechnology Research Centre |
| EPI_ISL_447054 | Cantacuzino National Military-Medical Institute for Research and Development | Cantacuzino Institute |
| EPI_ISL_447055 | Department for Virology, Molecular Biology and Genome Research, R. G. Lugar Center for Public Health Research, National Center for Disease Control and Public Health (NCDC) of Georgia. | Department for Virology, Molecular Biology and Genome Research, R. G. Lugar Center for Public Health Research, National Center for Disease Control and Public Health (NCDC) of Georgia. |
| EPI_ISL_447056 | Department for Virology, Molecular Biology and Genome Research, R. G. Lugar Center for Public Health Research, National Center for Disease Control and Public Health (NCDC) of Georgia. | Department for Virology, Molecular Biology and Genome Research, R. G. Lugar Center for Public Health Research, National Center for Disease Control and Public Health (NCDC) of Georgia. |
| EPI_ISL_447057 | Michigan Department of Health and Human Services, Bureau of Laboratories | Michigan Department of Health and Human Services, Bureau of Laboratories |
| EPI_ISL_447058 | Michigan Department of Health and Human Services, Bureau of Laboratories | Michigan Department of Health and Human Services, Bureau of Laboratories |
| EPI_ISL_447059 | Michigan Department of Health and Human Services, Bureau of Laboratories | Michigan Department of Health and Human Services, Bureau of Laboratories |
| EPI_ISL_447060 | Michigan Department of Health and Human Services, Bureau of Laboratories | Michigan Department of Health and Human Services, Bureau of Laboratories |
| EPI_ISL_447061 | Michigan Department of Health and Human Services, Bureau of Laboratories | Michigan Department of Health and Human Services, Bureau of Laboratories |
| EPI_ISL_447062 | Michigan Department of Health and Human Services, Bureau of Laboratories | Michigan Department of Health and Human Services, Bureau of Laboratories |
| EPI_ISL_447063 | Michigan Department of Health and Human Services, Bureau of Laboratories | Michigan Department of Health and Human Services, Bureau of Laboratories |
| EPI_ISL_447064 | Michigan Department of Health and Human Services, Bureau of Laboratories | Michigan Department of Health and Human Services, Bureau of Laboratories |
| EPI_ISL_447065 | Michigan Department of Health and Human Services, Bureau of Laboratories | Michigan Department of Health and Human Services, Bureau of Laboratories |
| EPI_ISL_447066 | Michigan Department of Health and Human Services, Bureau of Laboratories | Michigan Department of Health and Human Services, Bureau of Laboratories |

| EPI_Accession_ID | Originating Laboratory | Submitting Laboratory |
|---|---|---|
| EPI_ISL_447067 | Michigan Department of Health and Human Services, Bureau of Laboratories | Michigan Department of Health and Human Services, Bureau of Laboratories |
| EPI_ISL_447068 | Michigan Department of Health and Human Services, Bureau of Laboratories | Michigan Department of Health and Human Services, Bureau of Laboratories |
| EPI_ISL_447069 | Michigan Department of Health and Human Services, Bureau of Laboratories | Michigan Department of Health and Human Services, Bureau of Laboratories |
| EPI_ISL_447070 | Michigan Department of Health and Human Services, Bureau of Laboratories | Michigan Department of Health and Human Services, Bureau of Laboratories |
| EPI_ISL_447071 | Michigan Department of Health and Human Services, Bureau of Laboratories | Michigan Department of Health and Human Services, Bureau of Laboratories |
| EPI_ISL_447072 | Michigan Department of Health and Human Services, Bureau of Laboratories | Michigan Department of Health and Human Services, Bureau of Laboratories |
| EPI_ISL_447073 | Michigan Department of Health and Human Services, Bureau of Laboratories | Michigan Department of Health and Human Services, Bureau of Laboratories |
| EPI_ISL_447074 | Michigan Department of Health and Human Services, Bureau of Laboratories | Michigan Department of Health and Human Services, Bureau of Laboratories |
| EPI_ISL_447075 | Michigan Department of Health and Human Services, Bureau of Laboratories | Michigan Department of Health and Human Services, Bureau of Laboratories |
| EPI_ISL_447076 | Michigan Department of Health and Human Services, Bureau of Laboratories | Michigan Department of Health and Human Services, Bureau of Laboratories |
| EPI_ISL_447077 | Michigan Department of Health and Human Services, Bureau of Laboratories | Michigan Department of Health and Human Services, Bureau of Laboratories |
| EPI_ISL_447078 | Michigan Department of Health and Human Services, Bureau of Laboratories | Michigan Department of Health and Human Services, Bureau of Laboratories |
| EPI_ISL_447079 | Michigan Department of Health and Human Services, Bureau of Laboratories | Michigan Department of Health and Human Services, Bureau of Laboratories |
| EPI_ISL_447080 | Michigan Department of Health and Human Services, Bureau of Laboratories | Michigan Department of Health and Human Services, Bureau of Laboratories |
| EPI_ISL_447081 | Michigan Department of Health and Human Services, Bureau of Laboratories | Michigan Department of Health and Human Services, Bureau of Laboratories |
| EPI_ISL_447082 | Michigan Department of Health and Human Services, Bureau of Laboratories | Michigan Department of Health and Human Services, Bureau of Laboratories |

| EPI_Accession_ID | Originating Laboratory | Submitting Laboratory |
|---|---|---|
| EPI_ISL_447083 | Michigan Department of Health and Human Services, Bureau of Laboratories | Michigan Department of Health and Human Services, Bureau of Laboratories |
| EPI_ISL_447084 | Michigan Department of Health and Human Services, Bureau of Laboratories | Michigan Department of Health and Human Services, Bureau of Laboratories |
| EPI_ISL_447085 | Michigan Department of Health and Human Services, Bureau of Laboratories | Michigan Department of Health and Human Services, Bureau of Laboratories |
| EPI_ISL_447086 | Michigan Department of Health and Human Services, Bureau of Laboratories | Michigan Department of Health and Human Services, Bureau of Laboratories |
| EPI_ISL_447087 | Michigan Department of Health and Human Services, Bureau of Laboratories | Michigan Department of Health and Human Services, Bureau of Laboratories |
| EPI_ISL_447088 | Michigan Department of Health and Human Services, Bureau of Laboratories | Michigan Department of Health and Human Services, Bureau of Laboratories |
| EPI_ISL_447089 | Michigan Department of Health and Human Services, Bureau of Laboratories | Michigan Department of Health and Human Services, Bureau of Laboratories |
| EPI_ISL_447090 | Michigan Department of Health and Human Services, Bureau of Laboratories | Michigan Department of Health and Human Services, Bureau of Laboratories |
| EPI_ISL_447091 | Michigan Department of Health and Human Services, Bureau of Laboratories | Michigan Department of Health and Human Services, Bureau of Laboratories |
| EPI_ISL_447092 | Michigan Department of Health and Human Services, Bureau of Laboratories | Michigan Department of Health and Human Services, Bureau of Laboratories |
| EPI_ISL_447094 | Michigan Department of Health and Human Services, Bureau of Laboratories | Michigan Department of Health and Human Services, Bureau of Laboratories |
| EPI_ISL_447095 | Michigan Department of Health and Human Services, Bureau of Laboratories | Michigan Department of Health and Human Services, Bureau of Laboratories |
| EPI_ISL_447096 | Michigan Department of Health and Human Services, Bureau of Laboratories | Michigan Department of Health and Human Services, Bureau of Laboratories |
| EPI_ISL_447097 | Michigan Department of Health and Human Services, Bureau of Laboratories | Michigan Department of Health and Human Services, Bureau of Laboratories |
| EPI_ISL_447098 | Michigan Department of Health and Human Services, Bureau of Laboratories | Michigan Department of Health and Human Services, Bureau of Laboratories |
| EPI_ISL_447099 | Michigan Department of Health and Human Services, Bureau of Laboratories | Michigan Department of Health and Human Services, Bureau of Laboratories |

| EPI_Accession_ID | Originating Laboratory | Submitting Laboratory |
|---|---|---|
| EPI_ISL_447100 | Michigan Department of Health and Human Services, Bureau of Laboratories | Michigan Department of Health and Human Services, Bureau of Laboratories |
| EPI_ISL_447101 | Michigan Department of Health and Human Services, Bureau of Laboratories | Michigan Department of Health and Human Services, Bureau of Laboratories |
| EPI_ISL_447102 | Michigan Department of Health and Human Services, Bureau of Laboratories | Michigan Department of Health and Human Services, Bureau of Laboratories |
| EPI_ISL_447103 | Michigan Department of Health and Human Services, Bureau of Laboratories | Michigan Department of Health and Human Services, Bureau of Laboratories |
| EPI_ISL_447104 | Michigan Department of Health and Human Services, Bureau of Laboratories | Michigan Department of Health and Human Services, Bureau of Laboratories |
| EPI_ISL_447105 | Michigan Department of Health and Human Services, Bureau of Laboratories | Michigan Department of Health and Human Services, Bureau of Laboratories |
| EPI_ISL_447106 | Michigan Department of Health and Human Services, Bureau of Laboratories | Michigan Department of Health and Human Services, Bureau of Laboratories |
| EPI_ISL_447108 | Michigan Department of Health and Human Services, Bureau of Laboratories | Michigan Department of Health and Human Services, Bureau of Laboratories |
| EPI_ISL_447109 | Michigan Department of Health and Human Services, Bureau of Laboratories | Michigan Department of Health and Human Services, Bureau of Laboratories |
| EPI_ISL_447110 | Michigan Department of Health and Human Services, Bureau of Laboratories | Michigan Department of Health and Human Services, Bureau of Laboratories |
| EPI_ISL_447111 | Michigan Department of Health and Human Services, Bureau of Laboratories | Michigan Department of Health and Human Services, Bureau of Laboratories |
| EPI_ISL_447112 | Michigan Department of Health and Human Services, Bureau of Laboratories | Michigan Department of Health and Human Services, Bureau of Laboratories |
| EPI_ISL_447113 | Michigan Department of Health and Human Services, Bureau of Laboratories | Michigan Department of Health and Human Services, Bureau of Laboratories |
| EPI_ISL_447114 | Michigan Department of Health and Human Services, Bureau of Laboratories | Michigan Department of Health and Human Services, Bureau of Laboratories |
| EPI_ISL_447115 | Michigan Department of Health and Human Services, Bureau of Laboratories | Michigan Department of Health and Human Services, Bureau of Laboratories |
| EPI_ISL_447116 | Michigan Department of Health and Human Services, Bureau of Laboratories | Michigan Department of Health and Human Services, Bureau of Laboratories |

| EPI_Accession_ID | Originating Laboratory | Submitting Laboratory |
|---|---|---|
| EPI_ISL_447117 | Michigan Department of Health and Human Services, Bureau of Laboratories | Michigan Department of Health and Human Services, Bureau of Laboratories |
| EPI_ISL_447118 | Michigan Department of Health and Human Services, Bureau of Laboratories | Michigan Department of Health and Human Services, Bureau of Laboratories |
| EPI_ISL_447119 | HOSPITAL DR.HERNAN HENRIQUEZ ARAVENA | Instituto de Salud Publica de Chile |
| EPI_ISL_447120 | Department of Clinical Microbiology | GIGA Medical Genomics |
| EPI_ISL_447121 | Department of Clinical Microbiology | GIGA Medical Genomics |
| EPI_ISL_447122 | Department of Clinical Microbiology | GIGA Medical Genomics |
| EPI_ISL_447123 | Department of Clinical Microbiology | GIGA Medical Genomics |
| EPI_ISL_447124 | Department of Clinical Microbiology | GIGA Medical Genomics |
| EPI_ISL_447125 | Department of Clinical Microbiology | GIGA Medical Genomics |
| EPI_ISL_447126 | Department of Clinical Microbiology | GIGA Medical Genomics |
| EPI_ISL_447127 | Department of Clinical Microbiology | GIGA Medical Genomics |
| EPI_ISL_447128 | Department of Clinical Microbiology | GIGA Medical Genomics |
| EPI_ISL_447129 | Department of Clinical Microbiology | GIGA Medical Genomics |
| EPI_ISL_447130 | Department of Clinical Microbiology | GIGA Medical Genomics |
| EPI_ISL_447131 | Department of Clinical Microbiology | GIGA Medical Genomics |
| EPI_ISL_447132 | Department of Clinical Microbiology | GIGA Medical Genomics |
| EPI_ISL_447133 | Department of Clinical Microbiology | GIGA Medical Genomics |
| EPI_ISL_447134 | Department of Clinical Microbiology | GIGA Medical Genomics |
| EPI_ISL_447135 | Department of Clinical Microbiology | GIGA Medical Genomics |
| EPI_ISL_447136 | Department of Clinical Microbiology | GIGA Medical Genomics |
| EPI_ISL_447137 | Department of Clinical Microbiology | GIGA Medical Genomics |
| EPI_ISL_447138 | Department of Clinical Microbiology | GIGA Medical Genomics |
| EPI_ISL_447139 | Department of Clinical Microbiology | GIGA Medical Genomics |
| EPI_ISL_447140 | Department of Clinical Microbiology | GIGA Medical Genomics |
| EPI_ISL_447141 | Department of Clinical Microbiology | GIGA Medical Genomics |
| EPI_ISL_447142 | Department of Clinical Microbiology | GIGA Medical Genomics |
| EPI_ISL_447143 | Department of Clinical Microbiology | GIGA Medical Genomics |
| EPI_ISL_447144 | Department of Clinical Microbiology | GIGA Medical Genomics |
| EPI_ISL_447145 | Department of Clinical Microbiology | GIGA Medical Genomics |
| EPI_ISL_447146 | Department of Clinical Microbiology | GIGA Medical Genomics |
| EPI_ISL_447147 | Department of Clinical Microbiology | GIGA Medical Genomics |

| EPI_Accession_ID | Originating Laboratory | Submitting Laboratory |
|---|---|---|
| EPI_ISL_447148 | Department of Clinical Microbiology | GIGA Medical Genomics |
| EPI_ISL_447149 | Department of Clinical Microbiology | GIGA Medical Genomics |
| EPI_ISL_447150 | Department of Clinical Microbiology | GIGA Medical Genomics |
| EPI_ISL_447151 | Department of Clinical Microbiology | GIGA Medical Genomics |
| EPI_ISL_447152 | Department of Clinical Microbiology | GIGA Medical Genomics |
| EPI_ISL_447153 | Department of Clinical Microbiology | GIGA Medical Genomics |
| EPI_ISL_447154 | Department of Clinical Microbiology | GIGA Medical Genomics |
| EPI_ISL_447155 | Department of Clinical Microbiology | GIGA Medical Genomics |
| EPI_ISL_447156 | Department of Clinical Microbiology | GIGA Medical Genomics |
| EPI_ISL_447157 | Department of Clinical Microbiology | GIGA Medical Genomics |
| EPI_ISL_447158 | Department of Clinical Microbiology | GIGA Medical Genomics |
| EPI_ISL_447159 | Department of Clinical Microbiology | GIGA Medical Genomics |
| EPI_ISL_447160 | Department of Clinical Microbiology | GIGA Medical Genomics |
| EPI_ISL_447161 | Department of Clinical Microbiology | GIGA Medical Genomics |
| EPI_ISL_447162 | Department of Clinical Microbiology | GIGA Medical Genomics |
| EPI_ISL_447163 | Michigan Department of Health and Human Services, Bureau of Laboratories | Michigan Department of Health and Human Services, Bureau of Laboratories |
| EPI_ISL_447164 | Michigan Department of Health and Human Services, Bureau of Laboratories | Michigan Department of Health and Human Services, Bureau of Laboratories |
| EPI_ISL_447165 | Michigan Department of Health and Human Services, Bureau of Laboratories | Michigan Department of Health and Human Services, Bureau of Laboratories |
| EPI_ISL_447166 | Michigan Department of Health and Human Services, Bureau of Laboratories | Michigan Department of Health and Human Services, Bureau of Laboratories |
| EPI_ISL_447167 | Michigan Department of Health and Human Services, Bureau of Laboratories | Michigan Department of Health and Human Services, Bureau of Laboratories |
| EPI_ISL_447168 | Michigan Department of Health and Human Services, Bureau of Laboratories | Michigan Department of Health and Human Services, Bureau of Laboratories |
| EPI_ISL_447169 | Michigan Department of Health and Human Services, Bureau of Laboratories | Michigan Department of Health and Human Services, Bureau of Laboratories |
| EPI_ISL_447170 | Michigan Department of Health and Human Services, Bureau of Laboratories | Michigan Department of Health and Human Services, Bureau of Laboratories |
| EPI_ISL_447171 | Michigan Department of Health and Human Services, Bureau of Laboratories | Michigan Department of Health and Human Services, Bureau of Laboratories |

| EPI_Accession_ID | Originating Laboratory | Submitting Laboratory |
|---|---|---|
| EPI_ISL_447172 | Michigan Department of Health and Human Services, Bureau of Laboratories | Michigan Department of Health and Human Services, Bureau of Laboratories |
| EPI_ISL_447173 | Michigan Department of Health and Human Services, Bureau of Laboratories | Michigan Department of Health and Human Services, Bureau of Laboratories |
| EPI_ISL_447174 | Michigan Department of Health and Human Services, Bureau of Laboratories | Michigan Department of Health and Human Services, Bureau of Laboratories |
| EPI_ISL_447175 | Michigan Department of Health and Human Services, Bureau of Laboratories | Michigan Department of Health and Human Services, Bureau of Laboratories |
| EPI_ISL_447176 | Michigan Department of Health and Human Services, Bureau of Laboratories | Michigan Department of Health and Human Services, Bureau of Laboratories |
| EPI_ISL_447177 | Michigan Department of Health and Human Services, Bureau of Laboratories | Michigan Department of Health and Human Services, Bureau of Laboratories |
| EPI_ISL_447178 | Michigan Department of Health and Human Services, Bureau of Laboratories | Michigan Department of Health and Human Services, Bureau of Laboratories |
| EPI_ISL_447179 | Michigan Department of Health and Human Services, Bureau of Laboratories | Michigan Department of Health and Human Services, Bureau of Laboratories |
| EPI_ISL_447180 | Michigan Department of Health and Human Services, Bureau of Laboratories | Michigan Department of Health and Human Services, Bureau of Laboratories |
| EPI_ISL_447181 | Michigan Department of Health and Human Services, Bureau of Laboratories | Michigan Department of Health and Human Services, Bureau of Laboratories |
| EPI_ISL_447182 | Michigan Department of Health and Human Services, Bureau of Laboratories | Michigan Department of Health and Human Services, Bureau of Laboratories |
| EPI_ISL_447183 | Michigan Department of Health and Human Services, Bureau of Laboratories | Michigan Department of Health and Human Services, Bureau of Laboratories |
| EPI_ISL_447184 | Michigan Department of Health and Human Services, Bureau of Laboratories | Michigan Department of Health and Human Services, Bureau of Laboratories |
| EPI_ISL_447185 | Michigan Department of Health and Human Services, Bureau of Laboratories | Michigan Department of Health and Human Services, Bureau of Laboratories |
| EPI_ISL_447186 | Michigan Department of Health and Human Services, Bureau of Laboratories | Michigan Department of Health and Human Services, Bureau of Laboratories |
| EPI_ISL_447187 | Michigan Department of Health and Human Services, Bureau of Laboratories | Michigan Department of Health and Human Services, Bureau of Laboratories |

| EPI_Accession_ID | Originating Laboratory | Submitting Laboratory |
|---|---|---|
| EPI_ISL_447188 | Michigan Department of Health and Human Services, Bureau of Laboratories | Michigan Department of Health and Human Services, Bureau of Laboratories |
| EPI_ISL_447189 | Michigan Department of Health and Human Services, Bureau of Laboratories | Michigan Department of Health and Human Services, Bureau of Laboratories |
| EPI_ISL_447190 | Michigan Department of Health and Human Services, Bureau of Laboratories | Michigan Department of Health and Human Services, Bureau of Laboratories |
| EPI_ISL_447191 | Michigan Department of Health and Human Services, Bureau of Laboratories | Michigan Department of Health and Human Services, Bureau of Laboratories |
| EPI_ISL_447192 | Michigan Department of Health and Human Services, Bureau of Laboratories | Michigan Department of Health and Human Services, Bureau of Laboratories |
| EPI_ISL_447193 | Michigan Department of Health and Human Services, Bureau of Laboratories | Michigan Department of Health and Human Services, Bureau of Laboratories |
| EPI_ISL_447194 | Michigan Department of Health and Human Services, Bureau of Laboratories | Michigan Department of Health and Human Services, Bureau of Laboratories |
| EPI_ISL_447195 | Michigan Department of Health and Human Services, Bureau of Laboratories | Michigan Department of Health and Human Services, Bureau of Laboratories |
| EPI_ISL_447196 | Michigan Department of Health and Human Services, Bureau of Laboratories | Michigan Department of Health and Human Services, Bureau of Laboratories |
| EPI_ISL_447197 | Michigan Department of Health and Human Services, Bureau of Laboratories | Michigan Department of Health and Human Services, Bureau of Laboratories |
| EPI_ISL_447198 | Michigan Department of Health and Human Services, Bureau of Laboratories | Michigan Department of Health and Human Services, Bureau of Laboratories |
| EPI_ISL_447199 | Michigan Department of Health and Human Services, Bureau of Laboratories | Michigan Department of Health and Human Services, Bureau of Laboratories |
| EPI_ISL_447200 | Michigan Department of Health and Human Services, Bureau of Laboratories | Michigan Department of Health and Human Services, Bureau of Laboratories |
| EPI_ISL_447201 | Michigan Department of Health and Human Services, Bureau of Laboratories | Michigan Department of Health and Human Services, Bureau of Laboratories |
| EPI_ISL_447202 | Michigan Department of Health and Human Services, Bureau of Laboratories | Michigan Department of Health and Human Services, Bureau of Laboratories |
| EPI_ISL_447203 | Michigan Department of Health and Human Services, Bureau of Laboratories | Michigan Department of Health and Human Services, Bureau of Laboratories |

| EPI_Accession_ID | Originating Laboratory | Submitting Laboratory |
|---|---|---|
| EPI_ISL_447204 | Michigan Department of Health and Human Services, Bureau of Laboratories | Michigan Department of Health and Human Services, Bureau of Laboratories |
| EPI_ISL_447205 | Michigan Department of Health and Human Services, Bureau of Laboratories | Michigan Department of Health and Human Services, Bureau of Laboratories |
| EPI_ISL_447206 | Michigan Department of Health and Human Services, Bureau of Laboratories | Michigan Department of Health and Human Services, Bureau of Laboratories |
| EPI_ISL_447207 | Michigan Department of Health and Human Services, Bureau of Laboratories | Michigan Department of Health and Human Services, Bureau of Laboratories |
| EPI_ISL_447208 | Michigan Department of Health and Human Services, Bureau of Laboratories | Michigan Department of Health and Human Services, Bureau of Laboratories |
| EPI_ISL_447209 | Michigan Department of Health and Human Services, Bureau of Laboratories | Michigan Department of Health and Human Services, Bureau of Laboratories |
| EPI_ISL_447210 | Michigan Department of Health and Human Services, Bureau of Laboratories | Michigan Department of Health and Human Services, Bureau of Laboratories |
| EPI_ISL_447211 | Michigan Department of Health and Human Services, Bureau of Laboratories | Michigan Department of Health and Human Services, Bureau of Laboratories |
| EPI_ISL_447212 | Michigan Department of Health and Human Services, Bureau of Laboratories | Michigan Department of Health and Human Services, Bureau of Laboratories |
| EPI_ISL_447213 | Michigan Department of Health and Human Services, Bureau of Laboratories | Michigan Department of Health and Human Services, Bureau of Laboratories |
| EPI_ISL_447214 | Michigan Department of Health and Human Services, Bureau of Laboratories | Michigan Department of Health and Human Services, Bureau of Laboratories |
| EPI_ISL_447215 | Michigan Department of Health and Human Services, Bureau of Laboratories | Michigan Department of Health and Human Services, Bureau of Laboratories |
| EPI_ISL_447216 | Michigan Department of Health and Human Services, Bureau of Laboratories | Michigan Department of Health and Human Services, Bureau of Laboratories |
| EPI_ISL_447217 | Michigan Department of Health and Human Services, Bureau of Laboratories | Michigan Department of Health and Human Services, Bureau of Laboratories |
| EPI_ISL_447218 | Michigan Department of Health and Human Services, Bureau of Laboratories | Michigan Department of Health and Human Services, Bureau of Laboratories |
| EPI_ISL_447219 | Michigan Department of Health and Human Services, Bureau of Laboratories | Michigan Department of Health and Human Services, Bureau of Laboratories |

| EPI_Accession_ID | Originating Laboratory | Submitting Laboratory |
|---|---|---|
| EPI_ISL_447220 | Michigan Department of Health and Human Services, Bureau of Laboratories | Michigan Department of Health and Human Services, Bureau of Laboratories |
| EPI_ISL_447221 | Michigan Department of Health and Human Services, Bureau of Laboratories | Michigan Department of Health and Human Services, Bureau of Laboratories |
| EPI_ISL_447222 | Michigan Department of Health and Human Services, Bureau of Laboratories | Michigan Department of Health and Human Services, Bureau of Laboratories |
| EPI_ISL_447223 | Michigan Department of Health and Human Services, Bureau of Laboratories | Michigan Department of Health and Human Services, Bureau of Laboratories |
| EPI_ISL_447224 | Michigan Department of Health and Human Services, Bureau of Laboratories | Michigan Department of Health and Human Services, Bureau of Laboratories |
| EPI_ISL_447225 | Michigan Department of Health and Human Services, Bureau of Laboratories | Michigan Department of Health and Human Services, Bureau of Laboratories |
| EPI_ISL_447226 | Michigan Department of Health and Human Services, Bureau of Laboratories | Michigan Department of Health and Human Services, Bureau of Laboratories |
| EPI_ISL_447227 | Michigan Department of Health and Human Services, Bureau of Laboratories | Michigan Department of Health and Human Services, Bureau of Laboratories |
| EPI_ISL_447228 | Michigan Department of Health and Human Services, Bureau of Laboratories | Michigan Department of Health and Human Services, Bureau of Laboratories |
| EPI_ISL_447229 | Michigan Department of Health and Human Services, Bureau of Laboratories | Michigan Department of Health and Human Services, Bureau of Laboratories |
| EPI_ISL_447230 | Viral Respiratory Lab, National Institute for Biomedical Research (INRB) | Pathogen Sequencing Lab, National Institute for Biomedical Research (INRB) |
| EPI_ISL_447231 | Viral Respiratory Lab, National Institute for Biomedical Research (INRB) | Pathogen Sequencing Lab, National Institute for Biomedical Research (INRB) |
| EPI_ISL_447232 | Viral Respiratory Lab, National Institute for Biomedical Research (INRB) | Pathogen Sequencing Lab, National Institute for Biomedical Research (INRB) |
| EPI_ISL_447233 | Viral Respiratory Lab, National Institute for Biomedical Research (INRB) | Pathogen Sequencing Lab, National Institute for Biomedical Research (INRB) |
| EPI_ISL_447234 | Viral Respiratory Lab, National Institute for Biomedical Research (INRB) | Pathogen Sequencing Lab, National Institute for Biomedical Research (INRB) |
| EPI_ISL_447235 | Viral Respiratory Lab, National Institute for Biomedical Research (INRB) | Pathogen Sequencing Lab, National Institute for Biomedical Research (INRB) |

| EPI_Accession_ID | Originating Laboratory | Submitting Laboratory |
|---|---|---|
| EPI_ISL_447236 | Viral Respiratory Lab, National Institute for Biomedical Research (INRB) | Pathogen Sequencing Lab, National Institute for Biomedical Research (INRB) |
| EPI_ISL_447237 | Viral Respiratory Lab, National Institute for Biomedical Research (INRB) | Pathogen Sequencing Lab, National Institute for Biomedical Research (INRB) |
| EPI_ISL_447238 | Viral Respiratory Lab, National Institute for Biomedical Research (INRB) | Pathogen Sequencing Lab, National Institute for Biomedical Research (INRB) |
| EPI_ISL_447239 | Viral Respiratory Lab, National Institute for Biomedical Research (INRB) | Pathogen Sequencing Lab, National Institute for Biomedical Research (INRB) |
| EPI_ISL_447240 | Viral Respiratory Lab, National Institute for Biomedical Research (INRB) | Pathogen Sequencing Lab, National Institute for Biomedical Research (INRB) |
| EPI_ISL_447241 | Viral Respiratory Lab, National Institute for Biomedical Research (INRB) | Pathogen Sequencing Lab, National Institute for Biomedical Research (INRB) |
| EPI_ISL_447242 | Viral Respiratory Lab, National Institute for Biomedical Research (INRB) | Pathogen Sequencing Lab, National Institute for Biomedical Research (INRB) |
| EPI_ISL_447243 | Viral Respiratory Lab, National Institute for Biomedical Research (INRB) | Pathogen Sequencing Lab, National Institute for Biomedical Research (INRB) |
| EPI_ISL_447244 | Viral Respiratory Lab, National Institute for Biomedical Research (INRB) | Pathogen Sequencing Lab, National Institute for Biomedical Research (INRB) |
| EPI_ISL_447245 | Viral Respiratory Lab, National Institute for Biomedical Research (INRB) | Pathogen Sequencing Lab, National Institute for Biomedical Research (INRB) |
| EPI_ISL_447246 | Viral Respiratory Lab, National Institute for Biomedical Research (INRB) | Pathogen Sequencing Lab, National Institute for Biomedical Research (INRB) |
| EPI_ISL_447247 | Viral Respiratory Lab, National Institute for Biomedical Research (INRB) | Pathogen Sequencing Lab, National Institute for Biomedical Research (INRB) |
| EPI_ISL_447248 | Viral Respiratory Lab, National Institute for Biomedical Research (INRB) | Pathogen Sequencing Lab, National Institute for Biomedical Research (INRB) |
| EPI_ISL_447249 | Viral Respiratory Lab, National Institute for Biomedical Research (INRB) | Pathogen Sequencing Lab, National Institute for Biomedical Research (INRB) |
| EPI_ISL_447250 | Central Virology Laboratory | Central Virology Laboratory |
| EPI_ISL_447251 | Central Virology Laboratory | Central Virology Laboratory |
| EPI_ISL_447252 | TSGH-CP molecular lab | TSGH-CP molecular lab |
| EPI_ISL_447253 | TSGH-CP molecular lab | TSGH-CP molecular lab |
| EPI_ISL_447255 | TSGH-CP molecular lab | TSGH-CP molecular lab |

| EPI_Accession_ID | Originating Laboratory | Submitting Laboratory |
|---|---|---|
| EPI_ISL_447256 | TSGH-CP molecular lab | TSGH-CP molecular lab |
| EPI_ISL_447257 | TSGH-CP molecular lab | TSGH-CP molecular lab |
| EPI_ISL_447258 | Microbiology laboratory, Assuta Ashdod University-Affiliated Hospital | Stern Lab |
| EPI_ISL_447259 | Microbiology laboratory, Assuta Ashdod University-Affiliated Hospital | Stern Lab |
| EPI_ISL_447260 | Microbiology laboratory, Assuta Ashdod University-Affiliated Hospital | Stern Lab |
| EPI_ISL_447261 | Microbiology laboratory, Assuta Ashdod University-Affiliated Hospital | Stern Lab |
| EPI_ISL_447262 | Microbiology laboratory, Assuta Ashdod University-Affiliated Hospital | Stern Lab |
| EPI_ISL_447263 | Microbiology laboratory, Assuta Ashdod University-Affiliated Hospital | Stern Lab |
| EPI_ISL_447264 | Microbiology laboratory, Assuta Ashdod University-Affiliated Hospital | Stern Lab |
| EPI_ISL_447265 | Microbiology laboratory, Assuta Ashdod University-Affiliated Hospital | Stern Lab |
| EPI_ISL_447266 | Microbiology laboratory, Assuta Ashdod University-Affiliated Hospital | Stern Lab |
| EPI_ISL_447267 | Microbiology laboratory, Assuta Ashdod University-Affiliated Hospital | Stern Lab |
| EPI_ISL_447268 | Microbiology laboratory, Assuta Ashdod University-Affiliated Hospital | Stern Lab |
| EPI_ISL_447269 | Microbiology laboratory, Assuta Ashdod University-Affiliated Hospital | Stern Lab |
| EPI_ISL_447270 | Microbiology laboratory, Assuta Ashdod University-Affiliated Hospital | Stern Lab |
| EPI_ISL_447271 | Microbiology laboratory, Assuta Ashdod University-Affiliated Hospital | Stern Lab |
| EPI_ISL_447272 | Microbiology laboratory, Assuta Ashdod University-Affiliated Hospital | Stern Lab |

| EPI_Accession_ID | Originating Laboratory | Submitting Laboratory |
|---|---|---|
| EPI_ISL_447273 | Microbiology laboratory, Assuta Ashdod University-Affiliated Hospital | Stern Lab |
| EPI_ISL_447274 | Microbiology laboratory, Assuta Ashdod University-Affiliated Hospital | Stern Lab |
| EPI_ISL_447275 | Microbiology laboratory, Assuta Ashdod University-Affiliated Hospital | Stern Lab |
| EPI_ISL_447276 | Microbiology laboratory, Assuta Ashdod University-Affiliated Hospital | Stern Lab |
| EPI_ISL_447277 | Microbiology laboratory, Assuta Ashdod University-Affiliated Hospital | Stern Lab |
| EPI_ISL_447278 | Microbiology laboratory, Assuta Ashdod University-Affiliated Hospital | Stern Lab |
| EPI_ISL_447279 | Microbiology laboratory, Assuta Ashdod University-Affiliated Hospital | Stern Lab |
| EPI_ISL_447280 | Microbiology laboratory, Assuta Ashdod University-Affiliated Hospital | Stern Lab |
| EPI_ISL_447281 | Microbiology Division, Barzilai University Medical Center | Stern Lab |
| EPI_ISL_447282 | Microbiology Division, Barzilai University Medical Center | Stern Lab |
| EPI_ISL_447283 | Microbiology Division, Barzilai University Medical Center | Stern Lab |
| EPI_ISL_447284 | Microbiology Division, Barzilai University Medical Center | Stern Lab |
| EPI_ISL_447285 | Microbiology Division, Barzilai University Medical Center | Stern Lab |
| EPI_ISL_447286 | Microbiology Division, Barzilai University Medical Center | Stern Lab |
| EPI_ISL_447287 | Microbiology Division, Barzilai University Medical Center | Stern Lab |
| EPI_ISL_447288 | Microbiology Division, Barzilai University Medical Center | Stern Lab |

| EPI_Accession_ID | Originating Laboratory | Submitting Laboratory |
|---|---|---|
| EPI_ISL_447289 | Microbiology Division, Barzilai University Medical Center | Stern Lab |
| EPI_ISL_447290 | Microbiology Division, Barzilai University Medical Center | Stern Lab |
| EPI_ISL_447291 | Microbiology Division, Barzilai University Medical Center | Stern Lab |
| EPI_ISL_447292 | Microbiology Division, Barzilai University Medical Center | Stern Lab |
| EPI_ISL_447293 | Microbiology Division, Barzilai University Medical Center | Stern Lab |
| EPI_ISL_447294 | Microbiology Division, Barzilai University Medical Center | Stern Lab |
| EPI_ISL_447295 | Microbiology Division, Barzilai University Medical Center | Stern Lab |
| EPI_ISL_447296 | Microbiology Division, Barzilai University Medical Center | Stern Lab |
| EPI_ISL_447297 | Microbiology Division, Barzilai University Medical Center | Stern Lab |
| EPI_ISL_447298 | Microbiology Division, Barzilai University Medical Center | Stern Lab |
| EPI_ISL_447299 | Microbiology Division, Barzilai University Medical Center | Stern Lab |
| EPI_ISL_447300 | Microbiology Division, Barzilai University Medical Center | Stern Lab |
| EPI_ISL_447301 | Microbiology Division, Barzilai University Medical Center | Stern Lab |
| EPI_ISL_447302 | Microbiology Division, Barzilai University Medical Center | Stern Lab |
| EPI_ISL_447303 | Microbiology Division, Barzilai University Medical Center | Stern Lab |
| EPI_ISL_447304 | Microbiology Division, Barzilai University Medical Center | Stern Lab |

| EPI_Accession_ID | Originating Laboratory | Submitting Laboratory |
|---|---|---|
| EPI_ISL_447305 | Microbiology Division, Barzilai University Medical Center | Stern Lab |
| EPI_ISL_447306 | Microbiology Division, Barzilai University Medical Center | Stern Lab |
| EPI_ISL_447307 | Microbiology Division, Barzilai University Medical Center | Stern Lab |
| EPI_ISL_447308 | Microbiology Division, Barzilai University Medical Center | Stern Lab |
| EPI_ISL_447309 | Microbiology Division, Barzilai University Medical Center | Stern Lab |
| EPI_ISL_447310 | Microbiology Division, Barzilai University Medical Center | Stern Lab |
| EPI_ISL_447311 | Clinical Virology Laboratory, Soroka Medical Center and the Faculty of Health Sciences, Ben-Gurion University of the Negev | Stern Lab |
| EPI_ISL_447312 | Clinical Virology Laboratory, Soroka Medical Center and the Faculty of Health Sciences, Ben-Gurion University of the Negev | Stern Lab |
| EPI_ISL_447313 | Clinical Virology Laboratory, Soroka Medical Center and the Faculty of Health Sciences, Ben-Gurion University of the Negev | Stern Lab |
| EPI_ISL_447314 | Clinical Virology Laboratory, Soroka Medical Center and the Faculty of Health Sciences, Ben-Gurion University of the Negev | Stern Lab |
| EPI_ISL_447315 | Clinical Virology Laboratory, Soroka Medical Center and the Faculty of Health Sciences, Ben-Gurion University of the Negev | Stern Lab |
| EPI_ISL_447316 | Clinical Virology Laboratory, Soroka Medical Center and the Faculty of Health Sciences, Ben-Gurion University of the Negev | Stern Lab |
| EPI_ISL_447317 | Clinical Virology Laboratory, Soroka Medical Center and the Faculty of Health Sciences, Ben-Gurion University of the Negev | Stern Lab |

| EPI_Accession_ID | Originating Laboratory | Submitting Laboratory |
|---|---|---|
| EPI_ISL_447318 | Clinical Virology Laboratory, Soroka Medical Center and the Faculty of Health Sciences, Ben-Gurion University of the Negev | Stern Lab |
| EPI_ISL_447319 | Clinical Virology Laboratory, Soroka Medical Center and the Faculty of Health Sciences, Ben-Gurion University of the Negev | Stern Lab |
| EPI_ISL_447320 | Clinical Virology Laboratory, Soroka Medical Center and the Faculty of Health Sciences, Ben-Gurion University of the Negev | Stern Lab |
| EPI_ISL_447321 | Clinical Virology Laboratory, Soroka Medical Center and the Faculty of Health Sciences, Ben-Gurion University of the Negev | Stern Lab |
| EPI_ISL_447322 | Clinical Virology Laboratory, Soroka Medical Center and the Faculty of Health Sciences, Ben-Gurion University of the Negev | Stern Lab |
| EPI_ISL_447323 | Clinical Virology Laboratory, Soroka Medical Center and the Faculty of Health Sciences, Ben-Gurion University of the Negev | Stern Lab |
| EPI_ISL_447324 | Clinical Virology Laboratory, Soroka Medical Center and the Faculty of Health Sciences, Ben-Gurion University of the Negev | Stern Lab |
| EPI_ISL_447325 | Clinical Virology Laboratory, Soroka Medical Center and the Faculty of Health Sciences, Ben-Gurion University of the Negev | Stern Lab |
| EPI_ISL_447326 | Clinical Virology Laboratory, Soroka Medical Center and the Faculty of Health Sciences, Ben-Gurion University of the Negev | Stern Lab |
| EPI_ISL_447327 | Clinical Virology Laboratory, Soroka Medical Center and the Faculty of Health Sciences, Ben-Gurion University of the Negev | Stern Lab |
| EPI_ISL_447328 | Clinical Virology Laboratory, Soroka Medical Center and the Faculty of Health Sciences, Ben-Gurion University of the Negev | Stern Lab |

| EPI_Accession_ID | Originating Laboratory | Submitting Laboratory |
|---|---|---|
| EPI_ISL_447329 | Clinical Virology Laboratory, Soroka Medical Center and the Faculty of Health Sciences, Ben-Gurion University of the Negev | Stern Lab |
| EPI_ISL_447330 | Clinical Virology Laboratory, Soroka Medical Center and the Faculty of Health Sciences, Ben-Gurion University of the Negev | Stern Lab |
| EPI_ISL_447331 | Clinical Virology Unit, Hadassah Hebrew University Medical Center | Stern Lab |
| EPI_ISL_447332 | Clinical Virology Unit, Hadassah Hebrew University Medical Center | Stern Lab |
| EPI_ISL_447333 | Clinical Virology Unit, Hadassah Hebrew University Medical Center | Stern Lab |
| EPI_ISL_447334 | Clinical Virology Unit, Hadassah Hebrew University Medical Center | Stern Lab |
| EPI_ISL_447335 | Clinical Virology Unit, Hadassah Hebrew University Medical Center | Stern Lab |
| EPI_ISL_447336 | Clinical Virology Unit, Hadassah Hebrew University Medical Center | Stern Lab |
| EPI_ISL_447337 | Clinical Virology Unit, Hadassah Hebrew University Medical Center | Stern Lab |
| EPI_ISL_447338 | Clinical Virology Unit, Hadassah Hebrew University Medical Center | Stern Lab |
| EPI_ISL_447339 | Clinical Virology Unit, Hadassah Hebrew University Medical Center | Stern Lab |
| EPI_ISL_447340 | Clinical Virology Unit, Hadassah Hebrew University Medical Center | Stern Lab |
| EPI_ISL_447341 | Clinical Virology Unit, Hadassah Hebrew University Medical Center | Stern Lab |
| EPI_ISL_447342 | Clinical Virology Unit, Hadassah Hebrew University Medical Center | Stern Lab |
| EPI_ISL_447343 | Clinical Virology Unit, Hadassah Hebrew University Medical Center | Stern Lab |

| EPI_Accession_ID | Originating Laboratory | Submitting Laboratory |
|---|---|---|
| EPI_ISL_447344 | Clinical Virology Unit, Hadassah Hebrew University Medical Center | Stern Lab |
| EPI_ISL_447345 | Clinical Virology Unit, Hadassah Hebrew University Medical Center | Stern Lab |
| EPI_ISL_447346 | Clinical Virology Unit, Hadassah Hebrew University Medical Center | Stern Lab |
| EPI_ISL_447347 | Clinical Virology Unit, Hadassah Hebrew University Medical Center | Stern Lab |
| EPI_ISL_447348 | Clinical Virology Unit, Hadassah Hebrew University Medical Center | Stern Lab |
| EPI_ISL_447349 | Clinical Virology Unit, Hadassah Hebrew University Medical Center | Stern Lab |
| EPI_ISL_447350 | Clinical Virology Unit, Hadassah Hebrew University Medical Center | Stern Lab |
| EPI_ISL_447351 | Clinical Virology Unit, Hadassah Hebrew University Medical Center | Stern Lab |
| EPI_ISL_447352 | Clinical Virology Unit, Hadassah Hebrew University Medical Center | Stern Lab |
| EPI_ISL_447353 | Clinical Virology Unit, Hadassah Hebrew University Medical Center | Stern Lab |
| EPI_ISL_447354 | Clinical Virology Unit, Hadassah Hebrew University Medical Center | Stern Lab |
| EPI_ISL_447355 | Clinical Virology Unit, Hadassah Hebrew University Medical Center | Stern Lab |
| EPI_ISL_447356 | Clinical Virology Unit, Hadassah Hebrew University Medical Center | Stern Lab |
| EPI_ISL_447357 | Clinical Virology Unit, Hadassah Hebrew University Medical Center | Stern Lab |
| EPI_ISL_447358 | Clinical Virology Unit, Hadassah Hebrew University Medical Center | Stern Lab |
| EPI_ISL_447359 | Clinical Virology Unit, Hadassah Hebrew University Medical Center | Stern Lab |

| EPI_Accession_ID | Originating Laboratory | Submitting Laboratory |
| --- | --- | --- |
| EPI_ISL_447360 | Clinical Virology Unit, Hadassah Hebrew University Medical Center | Stern Lab |
| EPI_ISL_447361 | Clinical Virology Unit, Hadassah Hebrew University Medical Center | Stern Lab |
| EPI_ISL_447362 | Clinical Virology Unit, Hadassah Hebrew University Medical Center | Stern Lab |
| EPI_ISL_447363 | Clinical Virology Unit, Hadassah Hebrew University Medical Center | Stern Lab |
| EPI_ISL_447364 | Clinical Virology Unit, Hadassah Hebrew University Medical Center | Stern Lab |
| EPI_ISL_447365 | Clinical Virology Unit, Hadassah Hebrew University Medical Center | Stern Lab |
| EPI_ISL_447366 | Clinical Virology Unit, Hadassah Hebrew University Medical Center | Stern Lab |
| EPI_ISL_447367 | Clinical Virology Unit, Hadassah Hebrew University Medical Center | Stern Lab |
| EPI_ISL_447368 | Clinical Virology Unit, Hadassah Hebrew University Medical Center | Stern Lab |
| EPI_ISL_447369 | Clinical Virology Unit, Hadassah Hebrew University Medical Center | Stern Lab |
| EPI_ISL_447370 | Clinical Virology Unit, Hadassah Hebrew University Medical Center | Stern Lab |
| EPI_ISL_447371 | Clinical Virology Unit, Hadassah Hebrew University Medical Center | Stern Lab |
| EPI_ISL_447372 | Clinical Virology Unit, Hadassah Hebrew University Medical Center | Stern Lab |
| EPI_ISL_447373 | Clinical Virology Unit, Hadassah Hebrew University Medical Center | Stern Lab |
| EPI_ISL_447374 | Clinical Virology Unit, Hadassah Hebrew University Medical Center | Stern Lab |
| EPI_ISL_447375 | Clinical Virology Unit, Hadassah Hebrew University Medical Center | Stern Lab |

| EPI_Accession_ID | Originating Laboratory | Submitting Laboratory |
|---|---|---|
| EPI_ISL_447376 | Clinical Virology Unit, Hadassah Hebrew University Medical Center | Stern Lab |
| EPI_ISL_447377 | Clinical Virology Unit, Hadassah Hebrew University Medical Center | Stern Lab |
| EPI_ISL_447378 | Clinical Virology Unit, Hadassah Hebrew University Medical Center | Stern Lab |
| EPI_ISL_447379 | Clinical Virology Unit, Hadassah Hebrew University Medical Center | Stern Lab |
| EPI_ISL_447380 | Clinical Virology Unit, Hadassah Hebrew University Medical Center | Stern Lab |
| EPI_ISL_447381 | Clinical Virology Unit, Hadassah Hebrew University Medical Center | Stern Lab |
| EPI_ISL_447382 | Clinical Virology Unit, Hadassah Hebrew University Medical Center | Stern Lab |
| EPI_ISL_447383 | Clinical Microbiology Laboratory, The Baruch Padeh Medical Center, Poriya | Stern Lab |
| EPI_ISL_447384 | Clinical Microbiology Laboratory, The Baruch Padeh Medical Center, Poriya | Stern Lab |
| EPI_ISL_447385 | Clinical Microbiology Laboratory, The Baruch Padeh Medical Center, Poriya | Stern Lab |
| EPI_ISL_447386 | Clinical Microbiology Laboratory, The Baruch Padeh Medical Center, Poriya | Stern Lab |
| EPI_ISL_447387 | Clinical Microbiology Laboratory, The Baruch Padeh Medical Center, Poriya | Stern Lab |
| EPI_ISL_447388 | Clinical Microbiology Laboratory, The Baruch Padeh Medical Center, Poriya | Stern Lab |
| EPI_ISL_447389 | Clinical Microbiology Laboratory, The Baruch Padeh Medical Center, Poriya | Stern Lab |
| EPI_ISL_447390 | Clinical Microbiology Laboratory, The Baruch Padeh Medical Center, Poriya | Stern Lab |
| EPI_ISL_447391 | Clinical Microbiology Laboratory, The Baruch Padeh Medical Center, Poriya | Stern Lab |

| EPI_Accession_ID | Originating Laboratory | Submitting Laboratory |
|---|---|---|
| EPI_ISL_447392 | Clinical Microbiology Laboratory, The Baruch Padeh Medical Center, Poriya | Stern Lab |
| EPI_ISL_447393 | Clinical Microbiology Laboratory, The Baruch Padeh Medical Center, Poriya | Stern Lab |
| EPI_ISL_447394 | Clinical Microbiology Laboratory, The Baruch Padeh Medical Center, Poriya | Stern Lab |
| EPI_ISL_447395 | Clinical Microbiology Laboratory, The Baruch Padeh Medical Center, Poriya | Stern Lab |
| EPI_ISL_447396 | Clinical Microbiology Laboratory, The Baruch Padeh Medical Center, Poriya | Stern Lab |
| EPI_ISL_447397 | Clinical Microbiology Laboratory, The Baruch Padeh Medical Center, Poriya | Stern Lab |
| EPI_ISL_447398 | Clinical Microbiology Laboratory, The Baruch Padeh Medical Center, Poriya | Stern Lab |
| EPI_ISL_447399 | Clinical Microbiology Laboratory, The Baruch Padeh Medical Center, Poriya | Stern Lab |
| EPI_ISL_447400 | Clinical Microbiology Laboratory, The Baruch Padeh Medical Center, Poriya | Stern Lab |
| EPI_ISL_447401 | Clinical Microbiology Laboratory, The Baruch Padeh Medical Center, Poriya | Stern Lab |
| EPI_ISL_447402 | Clinical Microbiology Laboratory, The Baruch Padeh Medical Center, Poriya | Stern Lab |
| EPI_ISL_447403 | Clinical Microbiology Laboratory, The Baruch Padeh Medical Center, Poriya | Stern Lab |
| EPI_ISL_447404 | Clinical Microbiology Laboratory, The Baruch Padeh Medical Center, Poriya | Stern Lab |
| EPI_ISL_447405 | Clinical Microbiology Laboratory, The Baruch Padeh Medical Center, Poriya | Stern Lab |
| EPI_ISL_447406 | Clinical Microbiology Laboratory, The Baruch Padeh Medical Center, Poriya | Stern Lab |
| EPI_ISL_447407 | Clinical Virology Unit, Hadassah Hebrew University Medical Center | Stern Lab |

| EPI_Accession_ID | Originating Laboratory | Submitting Laboratory |
|---|---|---|
| EPI_ISL_447408 | Clinical Virology Unit, Hadassah Hebrew University Medical Center | Stern Lab |
| EPI_ISL_447409 | Clinical Virology Unit, Hadassah Hebrew University Medical Center | Stern Lab |
| EPI_ISL_447410 | Clinical Virology Unit, Hadassah Hebrew University Medical Center | Stern Lab |
| EPI_ISL_447411 | Clinical Virology Unit, Hadassah Hebrew University Medical Center | Stern Lab |
| EPI_ISL_447412 | Clinical Virology Unit, Hadassah Hebrew University Medical Center | Stern Lab |
| EPI_ISL_447413 | Clinical Virology Unit, Hadassah Hebrew University Medical Center | Stern Lab |
| EPI_ISL_447414 | Clinical Virology Unit, Hadassah Hebrew University Medical Center | Stern Lab |
| EPI_ISL_447415 | Clinical Virology Unit, Hadassah Hebrew University Medical Center | Stern Lab |
| EPI_ISL_447416 | Clinical Virology Unit, Hadassah Hebrew University Medical Center | Stern Lab |
| EPI_ISL_447417 | Clinical Microbiology Laboratory, The Baruch Padeh Medical Center, Poriya | Stern Lab |
| EPI_ISL_447418 | Clinical Microbiology Laboratory, The Baruch Padeh Medical Center, Poriya | Stern Lab |
| EPI_ISL_447419 | Clinical Microbiology Laboratory, Sheba Medical Center | Stern Lab |
| EPI_ISL_447420 | Clinical Microbiology Laboratory, Sheba Medical Center | Stern Lab |
| EPI_ISL_447421 | Clinical Microbiology Laboratory, Sheba Medical Center | Stern Lab |
| EPI_ISL_447422 | Clinical Microbiology Laboratory, Sheba Medical Center | Stern Lab |
| EPI_ISL_447423 | Clinical Microbiology Laboratory, Sheba Medical Center | Stern Lab |

| EPI_Accession_ID | Originating Laboratory | Submitting Laboratory |
|---|---|---|
| EPI_ISL_447424 | Clinical Microbiology Laboratory, Sheba Medical Center | Stern Lab |
| EPI_ISL_447425 | Clinical Microbiology Laboratory, Sheba Medical Center | Stern Lab |
| EPI_ISL_447426 | Clinical Microbiology Laboratory, Sheba Medical Center | Stern Lab |
| EPI_ISL_447427 | Clinical Microbiology Laboratory, Sheba Medical Center | Stern Lab |
| EPI_ISL_447428 | Clinical Microbiology Laboratory, Sheba Medical Center | Stern Lab |
| EPI_ISL_447429 | Clinical Microbiology Laboratory, Sheba Medical Center | Stern Lab |
| EPI_ISL_447430 | Clinical Microbiology Laboratory, Sheba Medical Center | Stern Lab |
| EPI_ISL_447431 | Clinical Microbiology Laboratory, Sheba Medical Center | Stern Lab |
| EPI_ISL_447432 | Clinical Microbiology Laboratory, Sheba Medical Center | Stern Lab |
| EPI_ISL_447433 | Clinical Microbiology Laboratory, Sheba Medical Center | Stern Lab |
| EPI_ISL_447434 | Clinical Microbiology Laboratory, Sheba Medical Center | Stern Lab |
| EPI_ISL_447435 | Clinical Microbiology Laboratory, Sheba Medical Center | Stern Lab |
| EPI_ISL_447436 | Clinical Microbiology Laboratory, Sheba Medical Center | Stern Lab |
| EPI_ISL_447437 | Clinical Microbiology Laboratory, Sheba Medical Center | Stern Lab |
| EPI_ISL_447438 | Clinical Microbiology Laboratory, Sheba Medical Center | Stern Lab |
| EPI_ISL_447439 | Clinical Microbiology Laboratory, Sheba Medical Center | Stern Lab |

| EPI_Accession_ID | Originating Laboratory | Submitting Laboratory |
|---|---|---|
| EPI_ISL_447440 | Clinical Microbiology Laboratory, Sheba Medical Center | Stern Lab |
| EPI_ISL_447441 | Clinical Microbiology Laboratory, Sheba Medical Center | Stern Lab |
| EPI_ISL_447442 | Clinical Microbiology Laboratory, Sheba Medical Center | Stern Lab |
| EPI_ISL_447443 | Clinical Microbiology Laboratory, Sheba Medical Center | Stern Lab |
| EPI_ISL_447444 | Clinical Microbiology Laboratory, Sheba Medical Center | Stern Lab |
| EPI_ISL_447445 | Clinical Microbiology Laboratory, Sheba Medical Center | Stern Lab |
| EPI_ISL_447446 | Clinical Microbiology Laboratory, Sheba Medical Center | Stern Lab |
| EPI_ISL_447447 | Clinical Microbiology Laboratory, Sheba Medical Center | Stern Lab |
| EPI_ISL_447448 | Clinical Microbiology Laboratory, Sheba Medical Center | Stern Lab |
| EPI_ISL_447449 | Clinical Microbiology Laboratory, Sheba Medical Center | Stern Lab |
| EPI_ISL_447450 | Clinical Microbiology Laboratory, Sheba Medical Center | Stern Lab |
| EPI_ISL_447451 | Clinical Microbiology Laboratory, Sheba Medical Center | Stern Lab |
| EPI_ISL_447452 | Clinical Microbiology Laboratory, Sheba Medical Center | Stern Lab |
| EPI_ISL_447453 | Clinical Microbiology Laboratory, Sheba Medical Center | Stern Lab |
| EPI_ISL_447454 | Clinical Microbiology Laboratory, Sheba Medical Center | Stern Lab |
| EPI_ISL_447455 | Clinical Microbiology Laboratory, Sheba Medical Center | Stern Lab |

| EPI_Accession_ID | Originating Laboratory | Submitting Laboratory |
|---|---|---|
| EPI_ISL_447456 | Clinical Microbiology Laboratory, Sheba Medical Center | Stern Lab |
| EPI_ISL_447457 | Clinical Microbiology Laboratory, Sheba Medical Center | Stern Lab |
| EPI_ISL_447458 | Clinical Microbiology Laboratory, Sheba Medical Center | Stern Lab |
| EPI_ISL_447459 | Clinical Microbiology Laboratory, Sheba Medical Center | Stern Lab |
| EPI_ISL_447460 | Clinical Microbiology Laboratory, Sheba Medical Center | Stern Lab |
| EPI_ISL_447461 | Clinical Microbiology Laboratory, Sheba Medical Center | Stern Lab |
| EPI_ISL_447462 | Clinical Microbiology Laboratory, Sheba Medical Center | Stern Lab |
| EPI_ISL_447463 | Clinical Microbiology Laboratory, Sheba Medical Center | Stern Lab |
| EPI_ISL_447464 | Clinical Microbiology Laboratory, Sheba Medical Center | Stern Lab |
| EPI_ISL_447465 | Clinical Microbiology Laboratory, Sheba Medical Center | Stern Lab |
| EPI_ISL_447466 | Clinical Microbiology Laboratory, Sheba Medical Center | Stern Lab |
| EPI_ISL_447467 | Clinical Microbiology Laboratory, Sheba Medical Center | Stern Lab |
| EPI_ISL_447468 | Clinical Microbiology Laboratory, Sheba Medical Center | Stern Lab |
| EPI_ISL_447469 | Clinical Microbiology Laboratory, Sheba Medical Center | Stern Lab |
| EPI_ISL_447470 | Servicio de Microbiologia. Hospital Clinico Universitario de Valencia | Sequencing and Bioinformatics Service and Molecular Epidemiology Research Group. FISABIO-Public Health |
| EPI_ISL_447471 | Servicio de Microbiologia. Hospital Clinico Universitario de Valencia | Sequencing and Bioinformatics Service and Molecular Epidemiology Research Group. FISABIO-Public Health |

| EPI_Accession_ID | Originating Laboratory | Submitting Laboratory |
|---|---|---|
| EPI_ISL_447472 | Servicio de Microbiologia. Hospital Clinico Universitario de Valencia | Sequencing and Bioinformatics Service and Molecular Epidemiology Research Group. FISABIO-Public Health |
| EPI_ISL_447473 | Servicio de Microbiologia. Hospital Clinico Universitario de Valencia | Sequencing and Bioinformatics Service and Molecular Epidemiology Research Group. FISABIO-Public Health |
| EPI_ISL_447474 | Servicio de Microbiologia. Hospital Clinico Universitario de Valencia | Sequencing and Bioinformatics Service and Molecular Epidemiology Research Group. FISABIO-Public Health |
| EPI_ISL_447475 | Servicio de Microbiologia. Hospital Clinico Universitario de Valencia | Sequencing and Bioinformatics Service and Molecular Epidemiology Research Group. FISABIO-Public Health |
| EPI_ISL_447476 | Servicio de Microbiologia. Hospital Clinico Universitario de Valencia | Sequencing and Bioinformatics Service and Molecular Epidemiology Research Group. FISABIO-Public Health |
| EPI_ISL_447477 | Servicio de Microbiologia. Hospital Clinico Universitario de Valencia | Sequencing and Bioinformatics Service and Molecular Epidemiology Research Group. FISABIO-Public Health |
| EPI_ISL_447478 | Servicio de Microbiologia. Hospital Clinico Universitario de Valencia | Sequencing and Bioinformatics Service and Molecular Epidemiology Research Group. FISABIO-Public Health |
| EPI_ISL_447479 | Servicio de Microbiologia. Hospital Clinico Universitario de Valencia | Sequencing and Bioinformatics Service and Molecular Epidemiology Research Group. FISABIO-Public Health |
| EPI_ISL_447480 | Servicio de Microbiologia. Hospital Clinico Universitario de Valencia | Sequencing and Bioinformatics Service and Molecular Epidemiology Research Group. FISABIO-Public Health |
| EPI_ISL_447481 | Servicio de Microbiologia. Hospital Clinico Universitario de Valencia | Sequencing and Bioinformatics Service and Molecular Epidemiology Research Group. FISABIO-Public Health |
| EPI_ISL_447482 | Servicio de Microbiologia. Hospital Clinico Universitario de Valencia | Sequencing and Bioinformatics Service and Molecular Epidemiology Research Group. FISABIO-Public Health |
| EPI_ISL_447483 | Servicio de Microbiologia. Hospital Clinico Universitario de Valencia | Sequencing and Bioinformatics Service and Molecular Epidemiology Research Group. FISABIO-Public Health |
| EPI_ISL_447484 | Servicio de Microbiologia. Hospital Clinico Universitario de Valencia | Sequencing and Bioinformatics Service and Molecular Epidemiology Research Group. FISABIO-Public Health |
| EPI_ISL_447485 | Servicio de Microbiologia. Hospital Clinico Universitario de Valencia | Sequencing and Bioinformatics Service and Molecular Epidemiology Research Group. FISABIO-Public Health |
| EPI_ISL_447486 | Servicio de Microbiologia. Hospital Clinico Universitario de Valencia | Sequencing and Bioinformatics Service and Molecular Epidemiology Research Group. FISABIO-Public Health |
| EPI_ISL_447487 | Servicio de Microbiologia. Hospital Clinico Universitario de Valencia | Sequencing and Bioinformatics Service and Molecular Epidemiology Research Group. FISABIO-Public Health |

| EPI_Accession_ID | Originating Laboratory | Submitting Laboratory |
|---|---|---|
| EPI_ISL_447488 | Servicio de Microbiologia. Hospital Clinico Universitario de Valencia | Sequencing and Bioinformatics Service and Molecular Epidemiology Research Group. FISABIO-Public Health |
| EPI_ISL_447489 | Servicio de Microbiologia. Hospital Clinico Universitario de Valencia | Sequencing and Bioinformatics Service and Molecular Epidemiology Research Group. FISABIO-Public Health |
| EPI_ISL_447490 | Servicio de Microbiologia. Hospital Clinico Universitario de Valencia | Sequencing and Bioinformatics Service and Molecular Epidemiology Research Group. FISABIO-Public Health |
| EPI_ISL_447491 | Servicio de Microbiologia. Hospital Clinico Universitario de Valencia | Sequencing and Bioinformatics Service and Molecular Epidemiology Research Group. FISABIO-Public Health |
| EPI_ISL_447492 | Servicio de Microbiologia. Hospital Clinico Universitario de Valencia | Sequencing and Bioinformatics Service and Molecular Epidemiology Research Group. FISABIO-Public Health |
| EPI_ISL_447493 | Servicio de Microbiologia. Hospital Clinico Universitario de Valencia | Sequencing and Bioinformatics Service and Molecular Epidemiology Research Group. FISABIO-Public Health |
| EPI_ISL_447494 | Servicio de Microbiologia. Hospital Clinico Universitario de Valencia | Sequencing and Bioinformatics Service and Molecular Epidemiology Research Group. FISABIO-Public Health |
| EPI_ISL_447495 | Servicio de Microbiologia. Hospital Clinico Universitario de Valencia | Sequencing and Bioinformatics Service and Molecular Epidemiology Research Group. FISABIO-Public Health |
| EPI_ISL_447496 | Servicio de Microbiologia. Hospital Clinico Universitario de Valencia | Sequencing and Bioinformatics Service and Molecular Epidemiology Research Group. FISABIO-Public Health |
| EPI_ISL_447497 | Servicio de Microbiologia. Hospital Clinico Universitario de Valencia | Sequencing and Bioinformatics Service and Molecular Epidemiology Research Group. FISABIO-Public Health |
| EPI_ISL_447498 | Servicio de Microbiologia. Hospital Clinico Universitario de Valencia | Sequencing and Bioinformatics Service and Molecular Epidemiology Research Group. FISABIO-Public Health |
| EPI_ISL_447499 | Servicio de Microbiologia. Hospital Clinico Universitario de Valencia | Sequencing and Bioinformatics Service and Molecular Epidemiology Research Group. FISABIO-Public Health |
| EPI_ISL_447500 | Servicio de Microbiologia. Hospital Clinico Universitario de Valencia | Sequencing and Bioinformatics Service and Molecular Epidemiology Research Group. FISABIO-Public Health |
| EPI_ISL_447501 | Servicio de Microbiologia. Hospital Clinico Universitario de Valencia | Sequencing and Bioinformatics Service and Molecular Epidemiology Research Group. FISABIO-Public Health |
| EPI_ISL_447502 | Servicio de Microbiologia. Hospital Clinico Universitario de Valencia | Sequencing and Bioinformatics Service and Molecular Epidemiology Research Group. FISABIO-Public Health |
| EPI_ISL_447503 | Servicio de Microbiologia. Hospital Clinico Universitario de Valencia | Sequencing and Bioinformatics Service and Molecular Epidemiology Research Group. FISABIO-Public Health |

| EPI_Accession_ID | Originating Laboratory | Submitting Laboratory |
|---|---|---|
| EPI_ISL_447504 | Servicio de Microbiologia. Hospital Clinico Universitario de Valencia | Sequencing and Bioinformatics Service and Molecular Epidemiology Research Group. FISABIO-Public Health |
| EPI_ISL_447505 | Servicio de Microbiologia. Hospital Clinico Universitario de Valencia | Sequencing and Bioinformatics Service and Molecular Epidemiology Research Group. FISABIO-Public Health |
| EPI_ISL_447506 | Servicio de Microbiologia. Hospital Clinico Universitario de Valencia | Sequencing and Bioinformatics Service and Molecular Epidemiology Research Group. FISABIO-Public Health |
| EPI_ISL_447507 | Servicio de Microbiologia. Hospital Clinico Universitario de Valencia | Sequencing and Bioinformatics Service and Molecular Epidemiology Research Group. FISABIO-Public Health |
| EPI_ISL_447508 | Servicio de Microbiologia. Hospital Clinico Universitario de Valencia | Sequencing and Bioinformatics Service and Molecular Epidemiology Research Group. FISABIO-Public Health |
| EPI_ISL_447509 | Servicio de Microbiologia. Hospital Clinico Universitario de Valencia | Sequencing and Bioinformatics Service and Molecular Epidemiology Research Group. FISABIO-Public Health |
| EPI_ISL_447510 | Servicio de Microbiologia. Hospital Clinico Universitario de Valencia | Sequencing and Bioinformatics Service and Molecular Epidemiology Research Group. FISABIO-Public Health |
| EPI_ISL_447511 | Servicio de Microbiologia. Hospital Clinico Universitario de Valencia | Sequencing and Bioinformatics Service and Molecular Epidemiology Research Group. FISABIO-Public Health |
| EPI_ISL_447512 | Servicio de Microbiologia. Hospital Clinico Universitario de Valencia | Sequencing and Bioinformatics Service and Molecular Epidemiology Research Group. FISABIO-Public Health |
| EPI_ISL_447513 | Servicio de Microbiologia. Hospital Clinico Universitario de Valencia | Sequencing and Bioinformatics Service and Molecular Epidemiology Research Group. FISABIO-Public Health |
| EPI_ISL_447514 | Servicio de Microbiologia. Hospital Clinico Universitario de Valencia | Sequencing and Bioinformatics Service and Molecular Epidemiology Research Group. FISABIO-Public Health |
| EPI_ISL_447515 | Servicio de Microbiologia. Hospital Clinico Universitario de Valencia | Sequencing and Bioinformatics Service and Molecular Epidemiology Research Group. FISABIO-Public Health |
| EPI_ISL_447516 | Servicio de Microbiologia. Hospital Clinico Universitario de Valencia | Sequencing and Bioinformatics Service and Molecular Epidemiology Research Group. FISABIO-Public Health |
| EPI_ISL_447517 | Servicio de Microbiologia. Hospital Clinico Universitario de Valencia | Sequencing and Bioinformatics Service and Molecular Epidemiology Research Group. FISABIO-Public Health |
| EPI_ISL_447518 | Servicio de Microbiologia. Hospital Clinico Universitario de Valencia | Sequencing and Bioinformatics Service and Molecular Epidemiology Research Group. FISABIO-Public Health |
| EPI_ISL_447519 | Servicio de Microbiologia. Hospital Clinico Universitario de Valencia | Sequencing and Bioinformatics Service and Molecular Epidemiology Research Group. FISABIO-Public Health |

| EPI_Accession_ID | Originating Laboratory | Submitting Laboratory |
| --- | --- | --- |
| EPI_ISL_447520 | Servicio de Microbiologia. Hospital Clinico Universitario de Valencia | Sequencing and Bioinformatics Service and Molecular Epidemiology Research Group. FISABIO-Public Health |
| EPI_ISL_447521 | Servicio de Microbiologia. Hospital Clinico Universitario de Valencia | Sequencing and Bioinformatics Service and Molecular Epidemiology Research Group. FISABIO-Public Health |
| EPI_ISL_447522 | Servicio de Microbiologia. Hospital Clinico Universitario de Valencia | Sequencing and Bioinformatics Service and Molecular Epidemiology Research Group. FISABIO-Public Health |
| EPI_ISL_447523 | Servicio de Microbiologia. Hospital Clinico Universitario de Valencia | Sequencing and Bioinformatics Service and Molecular Epidemiology Research Group. FISABIO-Public Health |
| EPI_ISL_447524 | Servicio de Microbiologia. Hospital Clinico Universitario de Valencia | Sequencing and Bioinformatics Service and Molecular Epidemiology Research Group. FISABIO-Public Health |
| EPI_ISL_447525 | Servicio de Microbiologia. Hospital Clinico Universitario de Valencia | Sequencing and Bioinformatics Service and Molecular Epidemiology Research Group. FISABIO-Public Health |
| EPI_ISL_447526 | Servicio de Microbiologia. Hospital Clinico Universitario de Valencia | Sequencing and Bioinformatics Service and Molecular Epidemiology Research Group. FISABIO-Public Health |
| EPI_ISL_447527 | Servicio de Microbiologia. Hospital Clinico Universitario de Valencia | Sequencing and Bioinformatics Service and Molecular Epidemiology Research Group. FISABIO-Public Health |
| EPI_ISL_447528 | Servicio de Microbiologia. Hospital Clinico Universitario de Valencia | Sequencing and Bioinformatics Service and Molecular Epidemiology Research Group. FISABIO-Public Health |
| EPI_ISL_447529 | Servicio de Microbiologia. Hospital Clinico Universitario de Valencia | Sequencing and Bioinformatics Service and Molecular Epidemiology Research Group. FISABIO-Public Health |
| EPI_ISL_447530 | Servicio de Microbiologia. Hospital Clinico Universitario de Valencia | Sequencing and Bioinformatics Service and Molecular Epidemiology Research Group. FISABIO-Public Health |
| EPI_ISL_447531 | Servicio de Microbiologia. Hospital Clinico Universitario de Valencia | Sequencing and Bioinformatics Service and Molecular Epidemiology Research Group. FISABIO-Public Health |
| EPI_ISL_447532 | Hospital Universitari Vall d'Hebron - Vall d'Hebron Institut de Recerca | Hospital Universitari Vall d'Hebron |
| EPI_ISL_447533 | Hospital Universitari Vall d'Hebron - Vall d'Hebron Institut de Recerca | Hospital Universitari Vall d'Hebron |
| EPI_ISL_447534 | Gujarat Biotechnology Research Centre | Gujarat Biotechnology Research Centre |
| EPI_ISL_447535 | Gujarat Biotechnology Research Centre | Gujarat Biotechnology Research Centre |
| EPI_ISL_447536 | Gujarat Biotechnology Research Centre | Gujarat Biotechnology Research Centre |
| EPI_ISL_447537 | Gujarat Biotechnology Research Centre | Gujarat Biotechnology Research Centre |
| EPI_ISL_447538 | Gujarat Biotechnology Research Centre | Gujarat Biotechnology Research Centre |

| EPI_Accession_ID | Originating Laboratory | Submitting Laboratory |
|---|---|---|
| EPI_ISL_447539 | Gujarat Biotechnology Research Centre | Gujarat Biotechnology Research Centre |
| EPI_ISL_447540 | Gujarat Biotechnology Research Centre | Gujarat Biotechnology Research Centre |
| EPI_ISL_447541 | Gujarat Biotechnology Research Centre | Gujarat Biotechnology Research Centre |
| EPI_ISL_447542 | Gujarat Biotechnology Research Centre | Gujarat Biotechnology Research Centre |
| EPI_ISL_447543 | Gujarat Biotechnology Research Centre | Gujarat Biotechnology Research Centre |
| EPI_ISL_447544 | Gujarat Biotechnology Research Centre | Gujarat Biotechnology Research Centre |
| EPI_ISL_447545 | Gujarat Biotechnology Research Centre | Gujarat Biotechnology Research Centre |
| EPI_ISL_447546 | Gujarat Biotechnology Research Centre | Gujarat Biotechnology Research Centre |
| EPI_ISL_447547 | GMERS Medical College and Hospital, Gandhinagar | Gujarat Biotechnology Research Centre |
| EPI_ISL_447548 | GMERS Medical College and Hospital, Gandhinagar | Gujarat Biotechnology Research Centre |
| EPI_ISL_447549 | GMERS Medical College and Hospital, Gandhinagar | Gujarat Biotechnology Research Centre |
| EPI_ISL_447550 | GMERS Medical College and Hospital, Gandhinagar | Gujarat Biotechnology Research Centre |
| EPI_ISL_447551 | GMERS Medical College and Hospital, Gandhinagar | Gujarat Biotechnology Research Centre |
| EPI_ISL_447552 | GMERS Medical College and Hospital, Gandhinagar | Gujarat Biotechnology Research Centre |
| EPI_ISL_447553 | GMERS Medical College and Hospital, Gandhinagar | Gujarat Biotechnology Research Centre |
| EPI_ISL_447554 | GMERS Medical College and Hospital, Gandhinagar | Gujarat Biotechnology Research Centre |
| EPI_ISL_447555 | GMERS Medical College and Hospital, Gandhinagar | Gujarat Biotechnology Research Centre |
| EPI_ISL_447556 | CSIR-Centre for Cellular and Molecular Biology | CSIR-Centre for Cellular and Molecular Biology |
| EPI_ISL_447557 | CSIR-Centre for Cellular and Molecular Biology | CSIR-Centre for Cellular and Molecular Biology |
| EPI_ISL_447558 | CSIR-Centre for Cellular and Molecular Biology | CSIR-Centre for Cellular and Molecular Biology |
| EPI_ISL_447559 | CSIR-Centre for Cellular and Molecular Biology | CSIR-Centre for Cellular and Molecular Biology |
| EPI_ISL_447560 | CSIR-Centre for Cellular and Molecular Biology | CSIR-Centre for Cellular and Molecular Biology |
| EPI_ISL_447561 | CSIR-Centre for Cellular and Molecular Biology | CSIR-Centre for Cellular and Molecular Biology |
| EPI_ISL_447562 | CSIR-Centre for Cellular and Molecular Biology | CSIR-Centre for Cellular and Molecular Biology |
| EPI_ISL_447563 | CSIR-Centre for Cellular and Molecular Biology | CSIR-Centre for Cellular and Molecular Biology |
| EPI_ISL_447564 | CSIR-Centre for Cellular and Molecular Biology | CSIR-Centre for Cellular and Molecular Biology |
| EPI_ISL_447565 | CSIR-Centre for Cellular and Molecular Biology | CSIR-Centre for Cellular and Molecular Biology |
| EPI_ISL_447566 | CSIR-Centre for Cellular and Molecular Biology | CSIR-Centre for Cellular and Molecular Biology |
| EPI_ISL_447567 | CSIR-Centre for Cellular and Molecular Biology | CSIR-Centre for Cellular and Molecular Biology |
| EPI_ISL_447568 | CSIR-Centre for Cellular and Molecular Biology | CSIR-Centre for Cellular and Molecular Biology |
| EPI_ISL_447569 | CSIR-Centre for Cellular and Molecular Biology | CSIR-Centre for Cellular and Molecular Biology |
| EPI_ISL_447570 | CSIR-Centre for Cellular and Molecular Biology | CSIR-Centre for Cellular and Molecular Biology |
| EPI_ISL_447571 | CSIR-Centre for Cellular and Molecular Biology | CSIR-Centre for Cellular and Molecular Biology |

| EPI_Accession_ID | Originating Laboratory | Submitting Laboratory |
|---|---|---|
| EPI_ISL_447572 | CSIR-Centre for Cellular and Molecular Biology | CSIR-Centre for Cellular and Molecular Biology |
| EPI_ISL_447573 | CSIR-Centre for Cellular and Molecular Biology | CSIR-Centre for Cellular and Molecular Biology |
| EPI_ISL_447574 | CSIR-Centre for Cellular and Molecular Biology | CSIR-Centre for Cellular and Molecular Biology |
| EPI_ISL_447575 | CSIR-Centre for Cellular and Molecular Biology | CSIR-Centre for Cellular and Molecular Biology |
| EPI_ISL_447576 | CSIR-Centre for Cellular and Molecular Biology | CSIR-Centre for Cellular and Molecular Biology |
| EPI_ISL_447577 | CSIR-Centre for Cellular and Molecular Biology | CSIR-Centre for Cellular and Molecular Biology |
| EPI_ISL_447578 | CSIR-Centre for Cellular and Molecular Biology | CSIR-Centre for Cellular and Molecular Biology |
| EPI_ISL_447579 | CSIR-Centre for Cellular and Molecular Biology | CSIR-Centre for Cellular and Molecular Biology |
| EPI_ISL_447580 | CSIR-Centre for Cellular and Molecular Biology | CSIR-Centre for Cellular and Molecular Biology |
| EPI_ISL_447581 | CSIR-Centre for Cellular and Molecular Biology | CSIR-Centre for Cellular and Molecular Biology |
| EPI_ISL_447582 | CSIR-Centre for Cellular and Molecular Biology | CSIR-Centre for Cellular and Molecular Biology |
| EPI_ISL_447583 | CSIR-Centre for Cellular and Molecular Biology | CSIR-Centre for Cellular and Molecular Biology |
| EPI_ISL_447584 | Tamil Nadu Veterinary and Animal Sciences University | CSIR-Centre for Cellular and Molecular Biology |
| EPI_ISL_447585 | Tamil Nadu Veterinary and Animal Sciences University | CSIR-Centre for Cellular and Molecular Biology |
| EPI_ISL_447586 | Tamil Nadu Veterinary and Animal Sciences University | CSIR-Centre for Cellular and Molecular Biology |
| EPI_ISL_447587 | Tamil Nadu Veterinary and Animal Sciences University | CSIR-Centre for Cellular and Molecular Biology |
| EPI_ISL_447588 | Lednicky Lab | Lednicky lab |
| EPI_ISL_447589 | Lednicky Lab | Lednicky Lab |
| EPI_ISL_447592 | TSGH-CP molecular lab | TSGH-CP molecular lab |
| EPI_ISL_447593 | TSGH-CP molecular lab | TSGH-CP molecular lab |
| EPI_ISL_447594 | Caloundra Hospital | Public Health Virology Laboratory |
| EPI_ISL_447595 | Pathology Queensland, Sunshine Coast University Hospital | Public Health Virology Laboratory |
| EPI_ISL_447596 | Viral Respiratory Lab, National Institute for Biomedical Research (INRB) | Pathogen Sequencing Lab, National Institute for Biomedical Research (INRB) |
| EPI_ISL_447597 | Viral Respiratory Lab, National Institute for Biomedical Research (INRB) | Pathogen Sequencing Lab, National Institute for Biomedical Research (INRB) |
| EPI_ISL_447599 | Viral Respiratory Lab, National Institute for Biomedical Research (INRB) | Pathogen Sequencing Lab, National Institute for Biomedical Research (INRB) |
| EPI_ISL_447600 | Viral Respiratory Lab, National Institute for Biomedical Research (INRB) | Pathogen Sequencing Lab, National Institute for Biomedical Research (INRB) |
| EPI_ISL_447601 | Viral Respiratory Lab, National Institute for Biomedical Research (INRB) | Pathogen Sequencing Lab, National Institute for Biomedical Research (INRB) |

| EPI_Accession_ID | Originating Laboratory | Submitting Laboratory |
|---|---|---|
| EPI_ISL_447602 | Viral Respiratory Lab, National Institute for Biomedical Research (INRB) | Pathogen Sequencing Lab, National Institute for Biomedical Research (INRB) |
| EPI_ISL_447603 | Viral Respiratory Lab, National Institute for Biomedical Research (INRB) | Pathogen Sequencing Lab, National Institute for Biomedical Research (INRB) |
| EPI_ISL_447604 | Viral Respiratory Lab, National Institute for Biomedical Research (INRB) | Pathogen Sequencing Lab, National Institute for Biomedical Research (INRB) |
| EPI_ISL_447605 | Viral Respiratory Lab, National Institute for Biomedical Research (INRB) | Pathogen Sequencing Lab, National Institute for Biomedical Research (INRB) |
| EPI_ISL_447606 | Viral Respiratory Lab, National Institute for Biomedical Research (INRB) | Pathogen Sequencing Lab, National Institute for Biomedical Research (INRB) |
| EPI_ISL_447607 | Viral Respiratory Lab, National Institute for Biomedical Research (INRB) | Pathogen Sequencing Lab, National Institute for Biomedical Research (INRB) |
| EPI_ISL_447614 | Department of Laboratory Medicine, National Taiwan University Hospital | Microbial Genomics Core Lab, National Taiwan University Centers of Genomic and Precision Medicine |
| EPI_ISL_447615 | Department of Laboratory Medicine, National Taiwan University Hospital | Microbial Genomics Core Lab, National Taiwan University Centers of Genomic and Precision Medicine |
| EPI_ISL_447616 | Department of Laboratory Medicine, National Taiwan University Hospital | Microbial Genomics Core Lab, National Taiwan University Centers of Genomic and Precision Medicine |
| EPI_ISL_447617 | Department of Laboratory Medicine, National Taiwan University Hospital | Microbial Genomics Core Lab, National Taiwan University Centers of Genomic and Precision Medicine |
| EPI_ISL_447618 | Department of Laboratory Medicine, National Taiwan University Hospital | Microbial Genomics Core Lab, National Taiwan University Centers of Genomic and Precision Medicine |
| EPI_ISL_447619 | Department of Laboratory Medicine, National Taiwan University Hospital | Microbial Genomics Core Lab, National Taiwan University Centers of Genomic and Precision Medicine |
| EPI_ISL_447620 | Department of Laboratory Medicine, National Taiwan University Hospital | Microbial Genomics Core Lab, National Taiwan University Centers of Genomic and Precision Medicine |
| EPI_ISL_447621 | Department of Laboratory Medicine, National Taiwan University Hospital | Microbial Genomics Core Lab, National Taiwan University Centers of Genomic and Precision Medicine |
| EPI_ISL_447622 | Department of Laboratory Medicine, National Taiwan University Hospital | Microbial Genomics Core Lab, National Taiwan University Centers of Genomic and Precision Medicine |
| EPI_ISL_447623 | unknown | Virology |
| EPI_ISL_447624 | unknown | Virology |
| EPI_ISL_447625 | unknown | Virology |

| EPI_Accession_ID | Originating Laboratory | Submitting Laboratory |
|---|---|---|
| EPI_ISL_447626 | unknown | Virology |
| EPI_ISL_447627 | unknown | Virology |
| EPI_ISL_447628 | unknown | Virology |
| EPI_ISL_447629 | unknown | Virology |
| EPI_ISL_447630 | unknown | Virology |
| EPI_ISL_447631 | unknown | Virology |
| EPI_ISL_447632 | unknown | Virology |
| EPI_ISL_447633 | unknown | Virology |
| EPI_ISL_447634 | unknown | Virology |
| EPI_ISL_447635 | unknown | Department of Medicine |
| EPI_ISL_447636 | unknown | Department of Medicine |
| EPI_ISL_447637 | unknown | Department of Medicine |
| EPI_ISL_447638 | unknown | Department of Medicine |
| EPI_ISL_447639 | unknown | Department of Medicine |
| EPI_ISL_447640 | unknown | Department of Medicine |
| EPI_ISL_447641 | unknown | Department of Medicine |
| EPI_ISL_447642 | unknown | Department of Medicine |
| EPI_ISL_447643 | unknown | Department of Medicine |
| EPI_ISL_447644 | unknown | Department of Medicine |
| EPI_ISL_447645 | unknown | Department of Medicine |
| EPI_ISL_447646 | unknown | Department of Medicine |
| EPI_ISL_447647 | unknown | Department of Medicine |
| EPI_ISL_447648 | unknown | Department of Medicine |
| EPI_ISL_447649 | unknown | Department of Medicine |
| EPI_ISL_447650 | unknown | Department of Medicine |
| EPI_ISL_447651 | unknown | Department of Medicine |
| EPI_ISL_447652 | unknown | Department of Medicine |
| EPI_ISL_447653 | unknown | Department of Medicine |

| EPI_Accession_ID | Originating Laboratory | Submitting Laboratory |
|---|---|---|
| EPI_ISL_447734 | Grupo de Investigaciones MicrobiolÃ³gicas-UR (GIMUR), Departamento de BiologÃ-a, Facultad de Ciencias Naturales, Universidad del Rosario, BogotÃ¡, Colombia | Grupo de Investigaciones MicrobiolÃ³gicas-UR (GIMUR), Departamento de BiologÃ-a, Facultad de Ciencias Naturales, Universidad del Rosario, BogotÃ¡, Colombia Instituto Nacional de Salud, BogotÃ¡, Colombia Icahn School of Medicine at Mount Sinai, New York, USA |
| EPI_ISL_447735 | Grupo de Investigaciones MicrobiolÃ³gicas-UR (GIMUR), Departamento de BiologÃ-a, Facultad de Ciencias Naturales, Universidad del Rosario, BogotÃ¡, Colombia | Grupo de Investigaciones MicrobiolÃ³gicas-UR (GIMUR), Departamento de BiologÃ-a, Facultad de Ciencias Naturales, Universidad del Rosario, BogotÃ¡, Colombia Instituto Nacional de Salud, BogotÃ¡, Colombia Icahn School of Medicine at Mount Sinai, New York, USA |
| EPI_ISL_447736 | Grupo de Investigaciones MicrobiolÃ³gicas-UR (GIMUR), Departamento de BiologÃ-a, Facultad de Ciencias Naturales, Universidad del Rosario, BogotÃ¡, Colombia | Grupo de Investigaciones MicrobiolÃ³gicas-UR (GIMUR), Departamento de BiologÃ-a, Facultad de Ciencias Naturales, Universidad del Rosario, BogotÃ¡, Colombia Instituto Nacional de Salud, BogotÃ¡, Colombia Icahn School of Medicine at Mount Sinai, New York, USA |
| EPI_ISL_447737 | Grupo de Investigaciones MicrobiolÃ³gicas-UR (GIMUR), Departamento de BiologÃ-a, Facultad de Ciencias Naturales, Universidad del Rosario, BogotÃ¡, Colombia | Grupo de Investigaciones MicrobiolÃ³gicas-UR (GIMUR), Departamento de BiologÃ-a, Facultad de Ciencias Naturales, Universidad del Rosario, BogotÃ¡, Colombia Instituto Nacional de Salud, BogotÃ¡, Colombia Icahn School of Medicine at Mount Sinai, New York, USA |
| EPI_ISL_447738 | Grupo de Investigaciones MicrobiolÃ³gicas-UR (GIMUR), Departamento de BiologÃ-a, Facultad de Ciencias Naturales, Universidad del Rosario, BogotÃ¡, Colombia | Grupo de Investigaciones MicrobiolÃ³gicas-UR (GIMUR), Departamento de BiologÃ-a, Facultad de Ciencias Naturales, Universidad del Rosario, BogotÃ¡, Colombia Instituto Nacional de Salud, BogotÃ¡, Colombia Icahn School of Medicine at Mount Sinai, New York, USA |

| EPI_Accession_ID | Originating Laboratory | Submitting Laboratory |
|---|---|---|
| EPI_ISL_447739 | Grupo de Investigaciones MicrobiolÃ³gicas-UR (GIMUR), Departamento de BiologÃ-a, Facultad de Ciencias Naturales, Universidad del Rosario, BogotÃ¡, Colombia | Grupo de Investigaciones MicrobiolÃ³gicas-UR (GIMUR), Departamento de BiologÃ-a, Facultad de Ciencias Naturales, Universidad del Rosario, BogotÃ¡, Colombia Instituto Nacional de Salud, BogotÃ¡, Colombia Icahn School of Medicine at Mount Sinai, New York, USA |
| EPI_ISL_447740 | Grupo de Investigaciones MicrobiolÃ³gicas-UR (GIMUR), Departamento de BiologÃ-a, Facultad de Ciencias Naturales, Universidad del Rosario, BogotÃ¡, Colombia | Grupo de Investigaciones MicrobiolÃ³gicas-UR (GIMUR), Departamento de BiologÃ-a, Facultad de Ciencias Naturales, Universidad del Rosario, BogotÃ¡, Colombia Instituto Nacional de Salud, BogotÃ¡, Colombia Icahn School of Medicine at Mount Sinai, New York, USA |
| EPI_ISL_447741 | Grupo de Investigaciones MicrobiolÃ³gicas-UR (GIMUR), Departamento de BiologÃ-a, Facultad de Ciencias Naturales, Universidad del Rosario, BogotÃ¡, Colombia | Grupo de Investigaciones MicrobiolÃ³gicas-UR (GIMUR), Departamento de BiologÃ-a, Facultad de Ciencias Naturales, Universidad del Rosario, BogotÃ¡, Colombia Instituto Nacional de Salud, BogotÃ¡, Colombia Icahn School of Medicine at Mount Sinai, New York, USA |
| EPI_ISL_447742 | Grupo de Investigaciones MicrobiolÃ³gicas-UR (GIMUR), Departamento de BiologÃ-a, Facultad de Ciencias Naturales, Universidad del Rosario, BogotÃ¡, Colombia | Grupo de Investigaciones MicrobiolÃ³gicas-UR (GIMUR), Departamento de BiologÃ-a, Facultad de Ciencias Naturales, Universidad del Rosario, BogotÃ¡, Colombia Instituto Nacional de Salud, BogotÃ¡, Colombia Icahn School of Medicine at Mount Sinai, New York, USA |
| EPI_ISL_447743 | Grupo de Investigaciones MicrobiolÃ³gicas-UR (GIMUR), Departamento de BiologÃ-a, Facultad de Ciencias Naturales, Universidad del Rosario, BogotÃ¡, Colombia | Grupo de Investigaciones MicrobiolÃ³gicas-UR (GIMUR), Departamento de BiologÃ-a, Facultad de Ciencias Naturales, Universidad del Rosario, BogotÃ¡, Colombia Instituto Nacional de Salud, BogotÃ¡, Colombia Icahn School of Medicine at Mount Sinai, New York, USA |

| EPI_Accession_ID | Originating Laboratory | Submitting Laboratory |
| --- | --- | --- |
| EPI_ISL_447744 | Grupo de Investigaciones MicrobiolÃ³gicas-UR (GIMUR), Departamento de BiologÃ-a, Facultad de Ciencias Naturales, Universidad del Rosario, BogotÃ¡, Colombia | Grupo de Investigaciones MicrobiolÃ³gicas-UR (GIMUR), Departamento de BiologÃ-a, Facultad de Ciencias Naturales, Universidad del Rosario, BogotÃ¡, Colombia Instituto Nacional de Salud, BogotÃ¡, Colombia Icahn School of Medicine at Mount Sinai, New York, USA |
| EPI_ISL_447745 | Grupo de Investigaciones MicrobiolÃ³gicas-UR (GIMUR), Departamento de BiologÃ-a, Facultad de Ciencias Naturales, Universidad del Rosario, BogotÃ¡, Colombia | Grupo de Investigaciones MicrobiolÃ³gicas-UR (GIMUR), Departamento de BiologÃ-a, Facultad de Ciencias Naturales, Universidad del Rosario, BogotÃ¡, Colombia Instituto Nacional de Salud, BogotÃ¡, Colombia Icahn School of Medicine at Mount Sinai, New York, USA |
| EPI_ISL_447746 | Grupo de Investigaciones MicrobiolÃ³gicas-UR (GIMUR), Departamento de BiologÃ-a, Facultad de Ciencias Naturales, Universidad del Rosario, BogotÃ¡, Colombia | Grupo de Investigaciones MicrobiolÃ³gicas-UR (GIMUR), Departamento de BiologÃ-a, Facultad de Ciencias Naturales, Universidad del Rosario, BogotÃ¡, Colombia Instituto Nacional de Salud, BogotÃ¡, Colombia Icahn School of Medicine at Mount Sinai, New York, USA |
| EPI_ISL_447747 | Grupo de Investigaciones MicrobiolÃ³gicas-UR (GIMUR), Departamento de BiologÃ-a, Facultad de Ciencias Naturales, Universidad del Rosario, BogotÃ¡, Colombia | Grupo de Investigaciones MicrobiolÃ³gicas-UR (GIMUR), Departamento de BiologÃ-a, Facultad de Ciencias Naturales, Universidad del Rosario, BogotÃ¡, Colombia Instituto Nacional de Salud, BogotÃ¡, Colombia Icahn School of Medicine at Mount Sinai, New York, USA |
| EPI_ISL_447748 | Grupo de Investigaciones MicrobiolÃ³gicas-UR (GIMUR), Departamento de BiologÃ-a, Facultad de Ciencias Naturales, Universidad del Rosario, BogotÃ¡, Colombia | Grupo de Investigaciones MicrobiolÃ³gicas-UR (GIMUR), Departamento de BiologÃ-a, Facultad de Ciencias Naturales, Universidad del Rosario, BogotÃ¡, Colombia Instituto Nacional de Salud, BogotÃ¡, Colombia Icahn School of Medicine at Mount Sinai, New York, USA |

| EPI_Accession_ID | Originating Laboratory | Submitting Laboratory |
|---|---|---|
| EPI_ISL_447749 | Grupo de Investigaciones MicrobiolÃ³gicas-UR (GIMUR), Departamento de BiologÃ-a, Facultad de Ciencias Naturales, Universidad del Rosario, BogotÃ¡, Colombia | Grupo de Investigaciones MicrobiolÃ³gicas-UR (GIMUR), Departamento de BiologÃ-a, Facultad de Ciencias Naturales, Universidad del Rosario, BogotÃ¡, Colombia Instituto Nacional de Salud, BogotÃ¡, Colombia Icahn School of Medicine at Mount Sinai, New York, USA |
| EPI_ISL_447750 | Grupo de Investigaciones MicrobiolÃ³gicas-UR (GIMUR), Departamento de BiologÃ-a, Facultad de Ciencias Naturales, Universidad del Rosario, BogotÃ¡, Colombia | Grupo de Investigaciones MicrobiolÃ³gicas-UR (GIMUR), Departamento de BiologÃ-a, Facultad de Ciencias Naturales, Universidad del Rosario, BogotÃ¡, Colombia Instituto Nacional de Salud, BogotÃ¡, Colombia Icahn School of Medicine at Mount Sinai, New York, USA |
| EPI_ISL_447751 | Grupo de Investigaciones MicrobiolÃ³gicas-UR (GIMUR), Departamento de BiologÃ-a, Facultad de Ciencias Naturales, Universidad del Rosario, BogotÃ¡, Colombia | Grupo de Investigaciones MicrobiolÃ³gicas-UR (GIMUR), Departamento de BiologÃ-a, Facultad de Ciencias Naturales, Universidad del Rosario, BogotÃ¡, Colombia Instituto Nacional de Salud, BogotÃ¡, Colombia Icahn School of Medicine at Mount Sinai, New York, USA |
| EPI_ISL_447752 | Grupo de Investigaciones MicrobiolÃ³gicas-UR (GIMUR), Departamento de BiologÃ-a, Facultad de Ciencias Naturales, Universidad del Rosario, BogotÃ¡, Colombia | Grupo de Investigaciones MicrobiolÃ³gicas-UR (GIMUR), Departamento de BiologÃ-a, Facultad de Ciencias Naturales, Universidad del Rosario, BogotÃ¡, Colombia Instituto Nacional de Salud, BogotÃ¡, Colombia Icahn School of Medicine at Mount Sinai, New York, USA |
| EPI_ISL_447753 | Grupo de Investigaciones MicrobiolÃ³gicas-UR (GIMUR), Departamento de BiologÃ-a, Facultad de Ciencias Naturales, Universidad del Rosario, BogotÃ¡, Colombia | Grupo de Investigaciones MicrobiolÃ³gicas-UR (GIMUR), Departamento de BiologÃ-a, Facultad de Ciencias Naturales, Universidad del Rosario, BogotÃ¡, Colombia Instituto Nacional de Salud, BogotÃ¡, Colombia Icahn School of Medicine at Mount Sinai, New York, USA |

| EPI_Accession_ID | Originating Laboratory | Submitting Laboratory |
|---|---|---|
| EPI_ISL_447754 | Grupo de Investigaciones MicrobiolÃ³gicas-UR (GIMUR), Departamento de BiologÃ-a, Facultad de Ciencias Naturales, Universidad del Rosario, BogotÃ¡, Colombia | Grupo de Investigaciones MicrobiolÃ³gicas-UR (GIMUR), Departamento de BiologÃ-a, Facultad de Ciencias Naturales, Universidad del Rosario, BogotÃ¡, Colombia Instituto Nacional de Salud, BogotÃ¡, Colombia Icahn School of Medicine at Mount Sinai, New York, USA |
| EPI_ISL_447755 | Instituto Nacional de Salud, BogotÃ¡, Colombia | Grupo de Investigaciones MicrobiolÃ³gicas-UR (GIMUR), Departamento de BiologÃ-a, Facultad de Ciencias Naturales, Universidad del Rosario, BogotÃ¡, Colombia Instituto Nacional de Salud, BogotÃ¡, Colombia Icahn School of Medicine at Mount Sinai, New York, USA |
| EPI_ISL_447756 | Instituto Nacional de Salud, BogotÃ¡, Colombia | Grupo de Investigaciones MicrobiolÃ³gicas-UR (GIMUR), Departamento de BiologÃ-a, Facultad de Ciencias Naturales, Universidad del Rosario, BogotÃ¡, Colombia Instituto Nacional de Salud, BogotÃ¡, Colombia Icahn School of Medicine at Mount Sinai, New York, USA |
| EPI_ISL_447757 | Instituto Nacional de Salud, BogotÃ¡, Colombia | Grupo de Investigaciones MicrobiolÃ³gicas-UR (GIMUR), Departamento de BiologÃ-a, Facultad de Ciencias Naturales, Universidad del Rosario, BogotÃ¡, Colombia Instituto Nacional de Salud, BogotÃ¡, Colombia Icahn School of Medicine at Mount Sinai, New York, USA |
| EPI_ISL_447758 | Instituto Nacional de Salud, BogotÃ¡, Colombia | Grupo de Investigaciones MicrobiolÃ³gicas-UR (GIMUR), Departamento de BiologÃ-a, Facultad de Ciencias Naturales, Universidad del Rosario, BogotÃ¡, Colombia Instituto Nacional de Salud, BogotÃ¡, Colombia Icahn School of Medicine at Mount Sinai, New York, USA |

| EPI_Accession_ID | Originating Laboratory | Submitting Laboratory |
|---|---|---|
| EPI_ISL_447759 | Instituto Nacional de Salud, BogotÃ¡, Colombia | Grupo de Investigaciones MicrobiolÃ³gicas-UR (GIMUR), Departamento de BiologÃ-a, Facultad de Ciencias Naturales, Universidad del Rosario, BogotÃ¡, Colombia Instituto Nacional de Salud, BogotÃ¡, Colombia Icahn School of Medicine at Mount Sinai, New York, USA |
| EPI_ISL_447760 | Instituto Nacional de Salud, BogotÃ¡, Colombia | Grupo de Investigaciones MicrobiolÃ³gicas-UR (GIMUR), Departamento de BiologÃ-a, Facultad de Ciencias Naturales, Universidad del Rosario, BogotÃ¡, Colombia Instituto Nacional de Salud, BogotÃ¡, Colombia Icahn School of Medicine at Mount Sinai, New York, USA |
| EPI_ISL_447761 | Instituto Nacional de Salud, BogotÃ¡, Colombia | Grupo de Investigaciones MicrobiolÃ³gicas-UR (GIMUR), Departamento de BiologÃ-a, Facultad de Ciencias Naturales, Universidad del Rosario, BogotÃ¡, Colombia Instituto Nacional de Salud, BogotÃ¡, Colombia Icahn School of Medicine at Mount Sinai, New York, USA |
| EPI_ISL_447762 | Instituto Nacional de Salud, BogotÃ¡, Colombia | Grupo de Investigaciones MicrobiolÃ³gicas-UR (GIMUR), Departamento de BiologÃ-a, Facultad de Ciencias Naturales, Universidad del Rosario, BogotÃ¡, Colombia Instituto Nacional de Salud, BogotÃ¡, Colombia Icahn School of Medicine at Mount Sinai, New York, USA |
| EPI_ISL_447763 | Instituto Nacional de Salud, BogotÃ¡, Colombia | Grupo de Investigaciones MicrobiolÃ³gicas-UR (GIMUR), Departamento de BiologÃ-a, Facultad de Ciencias Naturales, Universidad del Rosario, BogotÃ¡, Colombia Instituto Nacional de Salud, BogotÃ¡, Colombia Icahn School of Medicine at Mount Sinai, New York, USA |

| EPI_Accession_ID | Originating Laboratory | Submitting Laboratory |
|---|---|---|
| EPI_ISL_447765 | Instituto Nacional de Salud, BogotÃ¡, Colombia | Grupo de Investigaciones MicrobiolÃ³gicas-UR (GIMUR), Departamento de BiologÃ-a, Facultad de Ciencias Naturales, Universidad del Rosario, BogotÃ¡, Colombia Instituto Nacional de Salud, BogotÃ¡, Colombia Icahn School of Medicine at Mount Sinai, New York, USA |
| EPI_ISL_447766 | Instituto Nacional de Salud, BogotÃ¡, Colombia | Grupo de Investigaciones MicrobiolÃ³gicas-UR (GIMUR), Departamento de BiologÃ-a, Facultad de Ciencias Naturales, Universidad del Rosario, BogotÃ¡, Colombia Instituto Nacional de Salud, BogotÃ¡, Colombia Icahn School of Medicine at Mount Sinai, New York, USA |
| EPI_ISL_447767 | Instituto Nacional de Salud, BogotÃ¡, Colombia | Grupo de Investigaciones MicrobiolÃ³gicas-UR (GIMUR), Departamento de BiologÃ-a, Facultad de Ciencias Naturales, Universidad del Rosario, BogotÃ¡, Colombia Instituto Nacional de Salud, BogotÃ¡, Colombia Icahn School of Medicine at Mount Sinai, New York, USA |
| EPI_ISL_447768 | Instituto Nacional de Salud, BogotÃ¡, Colombia | Grupo de Investigaciones MicrobiolÃ³gicas-UR (GIMUR), Departamento de BiologÃ-a, Facultad de Ciencias Naturales, Universidad del Rosario, BogotÃ¡, Colombia Instituto Nacional de Salud, BogotÃ¡, Colombia Icahn School of Medicine at Mount Sinai, New York, USA |
| EPI_ISL_447769 | Instituto Nacional de Salud, BogotÃ¡, Colombia | Grupo de Investigaciones MicrobiolÃ³gicas-UR (GIMUR), Departamento de BiologÃ-a, Facultad de Ciencias Naturales, Universidad del Rosario, BogotÃ¡, Colombia Instituto Nacional de Salud, BogotÃ¡, Colombia Icahn School of Medicine at Mount Sinai, New York, USA |

| EPI_Accession_ID | Originating Laboratory | Submitting Laboratory |
|---|---|---|
| EPI_ISL_447771 | Instituto Nacional de Salud, BogotÃ¡, Colombia | Grupo de Investigaciones MicrobiolÃ³gicas-UR (GIMUR), Departamento de BiologÃ-a, Facultad de Ciencias Naturales, Universidad del Rosario, BogotÃ¡, Colombia Instituto Nacional de Salud, BogotÃ¡, Colombia Icahn School of Medicine at Mount Sinai, New York, USA |
| EPI_ISL_447774 | Instituto Nacional de Salud, BogotÃ¡, Colombia | Grupo de Investigaciones MicrobiolÃ³gicas-UR (GIMUR), Departamento de BiologÃ-a, Facultad de Ciencias Naturales, Universidad del Rosario, BogotÃ¡, Colombia Instituto Nacional de Salud, BogotÃ¡, Colombia Icahn School of Medicine at Mount Sinai, New York, USA |
| EPI_ISL_447775 | Instituto Nacional de Salud, BogotÃ¡, Colombia | Grupo de Investigaciones MicrobiolÃ³gicas-UR (GIMUR), Departamento de BiologÃ-a, Facultad de Ciencias Naturales, Universidad del Rosario, BogotÃ¡, Colombia Instituto Nacional de Salud, BogotÃ¡, Colombia Icahn School of Medicine at Mount Sinai, New York, USA |
| EPI_ISL_447777 | Instituto Nacional de Salud, BogotÃ¡, Colombia | Grupo de Investigaciones MicrobiolÃ³gicas-UR (GIMUR), Departamento de BiologÃ-a, Facultad de Ciencias Naturales, Universidad del Rosario, BogotÃ¡, Colombia Instituto Nacional de Salud, BogotÃ¡, Colombia Icahn School of Medicine at Mount Sinai, New York, USA |
| EPI_ISL_447778 | Instituto Nacional de Salud, BogotÃ¡, Colombia | Grupo de Investigaciones MicrobiolÃ³gicas-UR (GIMUR), Departamento de BiologÃ-a, Facultad de Ciencias Naturales, Universidad del Rosario, BogotÃ¡, Colombia Instituto Nacional de Salud, BogotÃ¡, Colombia Icahn School of Medicine at Mount Sinai, New York, USA |

| EPI_Accession_ID | Originating Laboratory | Submitting Laboratory |
|---|---|---|
| EPI_ISL_447779 | Instituto Nacional de Salud, BogotÃ¡, Colombia | Grupo de Investigaciones MicrobiolÃ³gicas-UR (GIMUR), Departamento de BiologÃ-a, Facultad de Ciencias Naturales, Universidad del Rosario, BogotÃ¡, Colombia Instituto Nacional de Salud, BogotÃ¡, Colombia Icahn School of Medicine at Mount Sinai, New York, USA |
| EPI_ISL_447780 | Instituto Nacional de Salud, BogotÃ¡, Colombia | Grupo de Investigaciones MicrobiolÃ³gicas-UR (GIMUR), Departamento de BiologÃ-a, Facultad de Ciencias Naturales, Universidad del Rosario, BogotÃ¡, Colombia Instituto Nacional de Salud, BogotÃ¡, Colombia Icahn School of Medicine at Mount Sinai, New York, USA |
| EPI_ISL_447781 | Instituto Nacional de Salud, BogotÃ¡, Colombia | Grupo de Investigaciones MicrobiolÃ³gicas-UR (GIMUR), Departamento de BiologÃ-a, Facultad de Ciencias Naturales, Universidad del Rosario, BogotÃ¡, Colombia Instituto Nacional de Salud, BogotÃ¡, Colombia Icahn School of Medicine at Mount Sinai, New York, USA |
| EPI_ISL_447782 | Instituto Nacional de Salud, BogotÃ¡, Colombia | Grupo de Investigaciones MicrobiolÃ³gicas-UR (GIMUR), Departamento de BiologÃ-a, Facultad de Ciencias Naturales, Universidad del Rosario, BogotÃ¡, Colombia Instituto Nacional de Salud, BogotÃ¡, Colombia Icahn School of Medicine at Mount Sinai, New York, USA |
| EPI_ISL_447783 | Instituto Nacional de Salud, BogotÃ¡, Colombia | Grupo de Investigaciones MicrobiolÃ³gicas-UR (GIMUR), Departamento de BiologÃ-a, Facultad de Ciencias Naturales, Universidad del Rosario, BogotÃ¡, Colombia Instituto Nacional de Salud, BogotÃ¡, Colombia Icahn School of Medicine at Mount Sinai, New York, USA |

| EPI_Accession_ID | Originating Laboratory | Submitting Laboratory |
|---|---|---|
| EPI_ISL_447784 | Instituto Nacional de Salud, BogotÃ¡, Colombia | Grupo de Investigaciones MicrobiolÃ³gicas-UR (GIMUR), Departamento de BiologÃ-a, Facultad de Ciencias Naturales, Universidad del Rosario, BogotÃ¡, Colombia Instituto Nacional de Salud, BogotÃ¡, Colombia Icahn School of Medicine at Mount Sinai, New York, USA |
| EPI_ISL_447785 | Instituto Nacional de Salud, BogotÃ¡, Colombia | Grupo de Investigaciones MicrobiolÃ³gicas-UR (GIMUR), Departamento de BiologÃ-a, Facultad de Ciencias Naturales, Universidad del Rosario, BogotÃ¡, Colombia Instituto Nacional de Salud, BogotÃ¡, Colombia Icahn School of Medicine at Mount Sinai, New York, USA |
| EPI_ISL_447786 | Instituto Nacional de Salud, BogotÃ¡, Colombia | Grupo de Investigaciones MicrobiolÃ³gicas-UR (GIMUR), Departamento de BiologÃ-a, Facultad de Ciencias Naturales, Universidad del Rosario, BogotÃ¡, Colombia Instituto Nacional de Salud, BogotÃ¡, Colombia Icahn School of Medicine at Mount Sinai, New York, USA |
| EPI_ISL_447787 | Instituto Nacional de Salud, BogotÃ¡, Colombia | Grupo de Investigaciones MicrobiolÃ³gicas-UR (GIMUR), Departamento de BiologÃ-a, Facultad de Ciencias Naturales, Universidad del Rosario, BogotÃ¡, Colombia Instituto Nacional de Salud, BogotÃ¡, Colombia Icahn School of Medicine at Mount Sinai, New York, USA |
| EPI_ISL_447789 | Instituto Nacional de Salud, BogotÃ¡, Colombia | Grupo de Investigaciones MicrobiolÃ³gicas-UR (GIMUR), Departamento de BiologÃ-a, Facultad de Ciencias Naturales, Universidad del Rosario, BogotÃ¡, Colombia Instituto Nacional de Salud, BogotÃ¡, Colombia Icahn School of Medicine at Mount Sinai, New York, USA |

| EPI_Accession_ID | Originating Laboratory | Submitting Laboratory |
|---|---|---|
| EPI_ISL_447790 | Instituto Nacional de Salud, BogotÃ¡, Colombia | Grupo de Investigaciones MicrobiolÃ³gicas-UR (GIMUR), Departamento de BiologÃa, Facultad de Ciencias Naturales, Universidad del Rosario, BogotÃ¡, Colombia Instituto Nacional de Salud, BogotÃ¡, Colombia Icahn School of Medicine at Mount Sinai, New York, USA |
| EPI_ISL_447791 | Instituto Nacional de Salud, BogotÃ¡, Colombia | Grupo de Investigaciones MicrobiolÃ³gicas-UR (GIMUR), Departamento de BiologÃa, Facultad de Ciencias Naturales, Universidad del Rosario, BogotÃ¡, Colombia Instituto Nacional de Salud, BogotÃ¡, Colombia Icahn School of Medicine at Mount Sinai, New York, USA |
| EPI_ISL_447793 | Instituto Nacional de Salud, BogotÃ¡, Colombia | Grupo de Investigaciones MicrobiolÃ³gicas-UR (GIMUR), Departamento de BiologÃa, Facultad de Ciencias Naturales, Universidad del Rosario, BogotÃ¡, Colombia Instituto Nacional de Salud, BogotÃ¡, Colombia Icahn School of Medicine at Mount Sinai, New York, USA |
| EPI_ISL_447794 | Instituto Nacional de Salud, BogotÃ¡, Colombia | Grupo de Investigaciones MicrobiolÃ³gicas-UR (GIMUR), Departamento de BiologÃa, Facultad de Ciencias Naturales, Universidad del Rosario, BogotÃ¡, Colombia Instituto Nacional de Salud, BogotÃ¡, Colombia Icahn School of Medicine at Mount Sinai, New York, USA |
| EPI_ISL_447795 | Instituto Nacional de Salud, BogotÃ¡, Colombia | Grupo de Investigaciones MicrobiolÃ³gicas-UR (GIMUR), Departamento de BiologÃa, Facultad de Ciencias Naturales, Universidad del Rosario, BogotÃ¡, Colombia Instituto Nacional de Salud, BogotÃ¡, Colombia Icahn School of Medicine at Mount Sinai, New York, USA |

| EPI_Accession_ID | Originating Laboratory | Submitting Laboratory |
|---|---|---|
| EPI_ISL_447796 | Instituto Nacional de Salud, BogotÃ¡, Colombia | Grupo de Investigaciones MicrobiolÃ³gicas-UR (GIMUR), Departamento de BiologÃ-a, Facultad de Ciencias Naturales, Universidad del Rosario, BogotÃ¡, Colombia Instituto Nacional de Salud, BogotÃ¡, Colombia Icahn School of Medicine at Mount Sinai, New York, USA |
| EPI_ISL_447797 | Instituto Nacional de Salud, BogotÃ¡, Colombia | Grupo de Investigaciones MicrobiolÃ³gicas-UR (GIMUR), Departamento de BiologÃ-a, Facultad de Ciencias Naturales, Universidad del Rosario, BogotÃ¡, Colombia Instituto Nacional de Salud, BogotÃ¡, Colombia Icahn School of Medicine at Mount Sinai, New York, USA |
| EPI_ISL_447798 | Instituto Nacional de Salud, BogotÃ¡, Colombia | Grupo de Investigaciones MicrobiolÃ³gicas-UR (GIMUR), Departamento de BiologÃ-a, Facultad de Ciencias Naturales, Universidad del Rosario, BogotÃ¡, Colombia Instituto Nacional de Salud, BogotÃ¡, Colombia Icahn School of Medicine at Mount Sinai, New York, USA |
| EPI_ISL_447799 | Instituto Nacional de Salud, BogotÃ¡, Colombia | Grupo de Investigaciones MicrobiolÃ³gicas-UR (GIMUR), Departamento de BiologÃ-a, Facultad de Ciencias Naturales, Universidad del Rosario, BogotÃ¡, Colombia Instituto Nacional de Salud, BogotÃ¡, Colombia Icahn School of Medicine at Mount Sinai, New York, USA |
| EPI_ISL_447800 | Instituto Nacional de Salud, BogotÃ¡, Colombia | Grupo de Investigaciones MicrobiolÃ³gicas-UR (GIMUR), Departamento de BiologÃ-a, Facultad de Ciencias Naturales, Universidad del Rosario, BogotÃ¡, Colombia Instituto Nacional de Salud, BogotÃ¡, Colombia Icahn School of Medicine at Mount Sinai, New York, USA |

| EPI_Accession_ID | Originating Laboratory | Submitting Laboratory |
| --- | --- | --- |
| EPI_ISL_447801 | Instituto Nacional de Salud, BogotÃ¡, Colombia | Grupo de Investigaciones MicrobiolÃ³gicas-UR (GIMUR), Departamento de BiologÃa, Facultad de Ciencias Naturales, Universidad del Rosario, BogotÃ¡, Colombia Instituto Nacional de Salud, BogotÃ¡, Colombia Icahn School of Medicine at Mount Sinai, New York, USA |
| EPI_ISL_447802 | Instituto Nacional de Salud, BogotÃ¡, Colombia | Grupo de Investigaciones MicrobiolÃ³gicas-UR (GIMUR), Departamento de BiologÃa, Facultad de Ciencias Naturales, Universidad del Rosario, BogotÃ¡, Colombia Instituto Nacional de Salud, BogotÃ¡, Colombia Icahn School of Medicine at Mount Sinai, New York, USA |
| EPI_ISL_447803 | Instituto Nacional de Salud, BogotÃ¡, Colombia | Grupo de Investigaciones MicrobiolÃ³gicas-UR (GIMUR), Departamento de BiologÃa, Facultad de Ciencias Naturales, Universidad del Rosario, BogotÃ¡, Colombia Instituto Nacional de Salud, BogotÃ¡, Colombia Icahn School of Medicine at Mount Sinai, New York, USA |
| EPI_ISL_447804 | Instituto Nacional de Salud, BogotÃ¡, Colombia | Grupo de Investigaciones MicrobiolÃ³gicas-UR (GIMUR), Departamento de BiologÃa, Facultad de Ciencias Naturales, Universidad del Rosario, BogotÃ¡, Colombia Instituto Nacional de Salud, BogotÃ¡, Colombia Icahn School of Medicine at Mount Sinai, New York, USA |
| EPI_ISL_447805 | Instituto Nacional de Salud, BogotÃ¡, Colombia | Grupo de Investigaciones MicrobiolÃ³gicas-UR (GIMUR), Departamento de BiologÃa, Facultad de Ciencias Naturales, Universidad del Rosario, BogotÃ¡, Colombia Instituto Nacional de Salud, BogotÃ¡, Colombia Icahn School of Medicine at Mount Sinai, New York, USA |

| EPI_Accession_ID | Originating Laboratory | Submitting Laboratory |
| --- | --- | --- |
| EPI_ISL_447806 | Instituto Nacional de Salud, BogotÃ¡, Colombia | Grupo de Investigaciones MicrobiolÃ³gicas-UR (GIMUR), Departamento de BiologÃ-a, Facultad de Ciencias Naturales, Universidad del Rosario, BogotÃ¡, Colombia Instituto Nacional de Salud, BogotÃ¡, Colombia Icahn School of Medicine at Mount Sinai, New York, USA |
| EPI_ISL_447807 | Instituto Nacional de Salud, BogotÃ¡, Colombia | Grupo de Investigaciones MicrobiolÃ³gicas-UR (GIMUR), Departamento de BiologÃ-a, Facultad de Ciencias Naturales, Universidad del Rosario, BogotÃ¡, Colombia Instituto Nacional de Salud, BogotÃ¡, Colombia Icahn School of Medicine at Mount Sinai, New York, USA |
| EPI_ISL_447808 | Instituto Nacional de Salud, BogotÃ¡, Colombia | Grupo de Investigaciones MicrobiolÃ³gicas-UR (GIMUR), Departamento de BiologÃ-a, Facultad de Ciencias Naturales, Universidad del Rosario, BogotÃ¡, Colombia Instituto Nacional de Salud, BogotÃ¡, Colombia Icahn School of Medicine at Mount Sinai, New York, USA |
| EPI_ISL_447809 | Instituto Nacional de Salud, BogotÃ¡, Colombia | Grupo de Investigaciones MicrobiolÃ³gicas-UR (GIMUR), Departamento de BiologÃ-a, Facultad de Ciencias Naturales, Universidad del Rosario, BogotÃ¡, Colombia Instituto Nacional de Salud, BogotÃ¡, Colombia Icahn School of Medicine at Mount Sinai, New York, USA |
| EPI_ISL_447810 | Instituto Nacional de Salud, BogotÃ¡, Colombia | Grupo de Investigaciones MicrobiolÃ³gicas-UR (GIMUR), Departamento de BiologÃ-a, Facultad de Ciencias Naturales, Universidad del Rosario, BogotÃ¡, Colombia Instituto Nacional de Salud, BogotÃ¡, Colombia Icahn School of Medicine at Mount Sinai, New York, USA |

| EPI_Accession_ID | Originating Laboratory | Submitting Laboratory |
| --- | --- | --- |
| EPI_ISL_447811 | Instituto Nacional de Salud, BogotÃ¡, Colombia | Grupo de Investigaciones MicrobiolÃ³gicas-UR (GIMUR), Departamento de BiologÃ-a, Facultad de Ciencias Naturales, Universidad del Rosario, BogotÃ¡, Colombia Instituto Nacional de Salud, BogotÃ¡, Colombia Icahn School of Medicine at Mount Sinai, New York, USA |
| EPI_ISL_447812 | Instituto Nacional de Salud, BogotÃ¡, Colombia | Grupo de Investigaciones MicrobiolÃ³gicas-UR (GIMUR), Departamento de BiologÃ-a, Facultad de Ciencias Naturales, Universidad del Rosario, BogotÃ¡, Colombia Instituto Nacional de Salud, BogotÃ¡, Colombia Icahn School of Medicine at Mount Sinai, New York, USA |
| EPI_ISL_447813 | Instituto Nacional de Salud, BogotÃ¡, Colombia | Grupo de Investigaciones MicrobiolÃ³gicas-UR (GIMUR), Departamento de BiologÃ-a, Facultad de Ciencias Naturales, Universidad del Rosario, BogotÃ¡, Colombia Instituto Nacional de Salud, BogotÃ¡, Colombia Icahn School of Medicine at Mount Sinai, New York, USA |
| EPI_ISL_447814 | Instituto Nacional de Salud, BogotÃ¡, Colombia | Grupo de Investigaciones MicrobiolÃ³gicas-UR (GIMUR), Departamento de BiologÃ-a, Facultad de Ciencias Naturales, Universidad del Rosario, BogotÃ¡, Colombia Instituto Nacional de Salud, BogotÃ¡, Colombia Icahn School of Medicine at Mount Sinai, New York, USA |
| EPI_ISL_447815 | Instituto Nacional de Salud, BogotÃ¡, Colombia | Grupo de Investigaciones MicrobiolÃ³gicas-UR (GIMUR), Departamento de BiologÃ-a, Facultad de Ciencias Naturales, Universidad del Rosario, BogotÃ¡, Colombia Instituto Nacional de Salud, BogotÃ¡, Colombia Icahn School of Medicine at Mount Sinai, New York, USA |

| EPI_Accession_ID | Originating Laboratory | Submitting Laboratory |
|---|---|---|
| EPI_ISL_447816 | Instituto Nacional de Salud, BogotÃ¡, Colombia | Grupo de Investigaciones MicrobiolÃ³gicas-UR (GIMUR), Departamento de BiologÃ-a, Facultad de Ciencias Naturales, Universidad del Rosario, BogotÃ¡, Colombia Instituto Nacional de Salud, BogotÃ¡, Colombia Icahn School of Medicine at Mount Sinai, New York, USA |
| EPI_ISL_447817 | Instituto Nacional de Salud, BogotÃ¡, Colombia | Grupo de Investigaciones MicrobiolÃ³gicas-UR (GIMUR), Departamento de BiologÃ-a, Facultad de Ciencias Naturales, Universidad del Rosario, BogotÃ¡, Colombia Instituto Nacional de Salud, BogotÃ¡, Colombia Icahn School of Medicine at Mount Sinai, New York, USA |
| EPI_ISL_447832 | unknown | Department of Medicine |
| EPI_ISL_447833 | unknown | Department of Medicine |
| EPI_ISL_447834 | unknown | Department of Medicine |
| EPI_ISL_447835 | unknown | Department of Medicine |
| EPI_ISL_447836 | unknown | Department of Medicine |
| EPI_ISL_447837 | Dept. of Medical Microbiology, Stavanger University Hospital, Helse Stavanger HF, | Norwegian Institute of Public Health, Department of Virology |
| EPI_ISL_447838 | Medical Microbiology Unit, Department for Laboratory Medicine, Drammen Hospital, Vestre Viken Health Trust, | Norwegian Institute of Public Health, Department of Virology |
| EPI_ISL_447839 | Medical Microbiology Unit, Department for Laboratory Medicine, Drammen Hospital, Vestre Viken Health Trust, | Norwegian Institute of Public Health, Department of Virology |
| EPI_ISL_447840 | DC Public Health Lab/ Dept. of Forensic Sciences | Pathogen Discovery, Respiratory Viruses Branch, Division of Viral Diseases, Centers for Disease Control and Prevention |
| EPI_ISL_447841 | FL Bureau of Public Health Laboratories-Tampa | Pathogen Discovery, Respiratory Viruses Branch, Division of Viral Diseases, Centers for Disease Control and Prevention |

| EPI_Accession_ID | Originating Laboratory | Submitting Laboratory |
|---|---|---|
| EPI_ISL_447842 | IA State Hygienic Laboratory | Pathogen Discovery, Respiratory Viruses Branch, Division of Viral Diseases, Centers for Disease Control and Prevention |
| EPI_ISL_447843 | MD DOH Laboratories Administration | Pathogen Discovery, Respiratory Viruses Branch, Division of Viral Diseases, Centers for Disease Control and Prevention |
| EPI_ISL_447844 | PA Department of Health, Bureau of Laboratories | Pathogen Discovery, Respiratory Viruses Branch, Division of Viral Diseases, Centers for Disease Control and Prevention |
| EPI_ISL_447845 | PR - Biological and Chemical Emergencies Lab Office of Public Health Preparedness and Response | Pathogen Discovery, Respiratory Viruses Branch, Division of Viral Diseases, Centers for Disease Control and Prevention |
| EPI_ISL_447846 | VT Dept. of Health Laboratory | Pathogen Discovery, Respiratory Viruses Branch, Division of Viral Diseases, Centers for Disease Control and Prevention |
| EPI_ISL_447847 | CSIR-Centre for Cellular and Molecular Biology | CSIR-Centre for Cellular and Molecular Biology |
| EPI_ISL_447848 | CSIR-Centre for Cellular and Molecular Biology | CSIR-Centre for Cellular and Molecular Biology |
| EPI_ISL_447849 | CSIR-Centre for Cellular and Molecular Biology | CSIR-Centre for Cellular and Molecular Biology |
| EPI_ISL_447850 | CSIR-Centre for Cellular and Molecular Biology | CSIR-Centre for Cellular and Molecular Biology |
| EPI_ISL_447851 | CSIR-Centre for Cellular and Molecular Biology | CSIR-Centre for Cellular and Molecular Biology |
| EPI_ISL_447852 | CSIR-Centre for Cellular and Molecular Biology | CSIR-Centre for Cellular and Molecular Biology |
| EPI_ISL_447853 | CSIR-Centre for Cellular and Molecular Biology | CSIR-Centre for Cellular and Molecular Biology |
| EPI_ISL_447854 | CSIR-Centre for Cellular and Molecular Biology | CSIR-Centre for Cellular and Molecular Biology |
| EPI_ISL_447855 | CSIR-Centre for Cellular and Molecular Biology | CSIR-Centre for Cellular and Molecular Biology |
| EPI_ISL_447856 | CSIR-Centre for Cellular and Molecular Biology | CSIR-Centre for Cellular and Molecular Biology |
| EPI_ISL_447857 | CSIR-Centre for Cellular and Molecular Biology | CSIR-Centre for Cellular and Molecular Biology |
| EPI_ISL_447858 | CSIR-Centre for Cellular and Molecular Biology | CSIR-Centre for Cellular and Molecular Biology |
| EPI_ISL_447859 | CSIR-Centre for Cellular and Molecular Biology | CSIR-Centre for Cellular and Molecular Biology |
| EPI_ISL_447860 | CSIR-Centre for Cellular and Molecular Biology | CSIR-Centre for Cellular and Molecular Biology |
| EPI_ISL_447861 | CSIR-Centre for Cellular and Molecular Biology | CSIR-Centre for Cellular and Molecular Biology |
| EPI_ISL_447862 | CSIR-Centre for Cellular and Molecular Biology | CSIR-Centre for Cellular and Molecular Biology |
| EPI_ISL_447863 | CSIR-Centre for Cellular and Molecular Biology | CSIR-Centre for Cellular and Molecular Biology |
| EPI_ISL_447864 | CSIR-Centre for Cellular and Molecular Biology | CSIR-Centre for Cellular and Molecular Biology |

| EPI_Accession_ID | Originating Laboratory | Submitting Laboratory |
|---|---|---|
| EPI_ISL_447865 | CSIR-Centre for Cellular and Molecular Biology | CSIR-Centre for Cellular and Molecular Biology |
| EPI_ISL_447866 | CSIR-Centre for Cellular and Molecular Biology | CSIR-Centre for Cellular and Molecular Biology |
| EPI_ISL_447886 | unknown | Pathogen Discovery |
| EPI_ISL_447887 | University of California, Davis | Chan-Zuckerberg Biohub |
| EPI_ISL_447888 | University of California, Davis | Chan-Zuckerberg Biohub |
| EPI_ISL_447889 | University of California, Davis | Chan-Zuckerberg Biohub |
| EPI_ISL_447890 | University of California, Davis | Chan-Zuckerberg Biohub |
| EPI_ISL_447891 | University of California, Davis | Chan-Zuckerberg Biohub |
| EPI_ISL_447892 | University of California, Davis | Chan-Zuckerberg Biohub |
| EPI_ISL_447893 | University of California, Davis | Chan-Zuckerberg Biohub |
| EPI_ISL_447894 | University of California, Davis | Chan-Zuckerberg Biohub |
| EPI_ISL_447895 | University of California, Davis | Chan-Zuckerberg Biohub |
| EPI_ISL_447896 | University of California, Davis | Chan-Zuckerberg Biohub |
| EPI_ISL_447900 | Lednicky Laboratory at Emerging Pathogens Institute | Environmental and Global Health |
| EPI_ISL_447903 | unknown | Environmental and Global Health |
| EPI_ISL_447904 | National Institute of Biotechnology | National Institute of Biotechnology |
| EPI_ISL_447905 | unknown | Environmental and Global Health |
| EPI_ISL_447909 | n/a | National Institute of Health. Department of medical Sciences, Ministry of Public Health, Thailand |
| EPI_ISL_447910 | n/a | National Institute of Health. Department of medical Sciences, Ministry of Public Health, Thailand |
| EPI_ISL_447911 | n/a | National Institute of Health. Department of medical Sciences, Ministry of Public Health, Thailand |
| EPI_ISL_447912 | n/a | National Institute of Health. Department of medical Sciences, Ministry of Public Health, Thailand |
| EPI_ISL_447913 | n/a | National Institute of Health. Department of medical Sciences, Ministry of Public Health, Thailand |
| EPI_ISL_447914 | n/a | National Institute of Health. Department of medical Sciences, Ministry of Public Health, Thailand |
| EPI_ISL_447915 | n/a | National Institute of Health. Department of medical Sciences, Ministry of Public Health, Thailand |
| EPI_ISL_447916 | n/a | National Institute of Health. Department of medical Sciences, Ministry of Public Health, Thailand |

| EPI_Accession_ID | Originating Laboratory | Submitting Laboratory |
|---|---|---|
| EPI_ISL_447917 | n/a | National Institute of Health. Department of medical Sciences, Ministry of Public Health, Thailand |
| EPI_ISL_447918 | n/a | National Institute of Health. Department of medical Sciences, Ministry of Public Health, Thailand |
| EPI_ISL_447919 | n/a | National Institute of Health. Department of medical Sciences, Ministry of Public Health, Thailand |
| EPI_ISL_447920 | n/a | National Institute of Health. Department of medical Sciences, Ministry of Public Health, Thailand |
| EPI_ISL_447921 | n/a | National Institute of Health. Department of medical Sciences, Ministry of Public Health, Thailand |
| EPI_ISL_447922 | University of Birmingham | COVID-19 Genomics UK (COG-UK) Consortium |
| EPI_ISL_447923 | University of Birmingham | COVID-19 Genomics UK (COG-UK) Consortium |
| EPI_ISL_447924 | University of Birmingham | COVID-19 Genomics UK (COG-UK) Consortium |
| EPI_ISL_447925 | University of Birmingham | COVID-19 Genomics UK (COG-UK) Consortium |
| EPI_ISL_447926 | University of Birmingham | COVID-19 Genomics UK (COG-UK) Consortium |
| EPI_ISL_447927 | University of Birmingham | COVID-19 Genomics UK (COG-UK) Consortium |
| EPI_ISL_447928 | University of Birmingham | COVID-19 Genomics UK (COG-UK) Consortium |
| EPI_ISL_447929 | University of Birmingham | COVID-19 Genomics UK (COG-UK) Consortium |
| EPI_ISL_447930 | University of Birmingham | COVID-19 Genomics UK (COG-UK) Consortium |
| EPI_ISL_447931 | University of Birmingham | COVID-19 Genomics UK (COG-UK) Consortium |
| EPI_ISL_447932 | University of Birmingham | COVID-19 Genomics UK (COG-UK) Consortium |
| EPI_ISL_447933 | University of Birmingham | COVID-19 Genomics UK (COG-UK) Consortium |
| EPI_ISL_447934 | University of Birmingham | COVID-19 Genomics UK (COG-UK) Consortium |
| EPI_ISL_447935 | University of Birmingham | COVID-19 Genomics UK (COG-UK) Consortium |
| EPI_ISL_447936 | University of Birmingham | COVID-19 Genomics UK (COG-UK) Consortium |
| EPI_ISL_447937 | University of Birmingham | COVID-19 Genomics UK (COG-UK) Consortium |
| EPI_ISL_447938 | University of Birmingham | COVID-19 Genomics UK (COG-UK) Consortium |
| EPI_ISL_447939 | University of Birmingham | COVID-19 Genomics UK (COG-UK) Consortium |
| EPI_ISL_447940 | University of Birmingham | COVID-19 Genomics UK (COG-UK) Consortium |
| EPI_ISL_447941 | University of Birmingham | COVID-19 Genomics UK (COG-UK) Consortium |
| EPI_ISL_447942 | University of Birmingham | COVID-19 Genomics UK (COG-UK) Consortium |
| EPI_ISL_447943 | University of Birmingham | COVID-19 Genomics UK (COG-UK) Consortium |
| EPI_ISL_447944 | University of Birmingham | COVID-19 Genomics UK (COG-UK) Consortium |

| EPI_Accession_ID | Originating Laboratory | Submitting Laboratory |
|---|---|---|
| EPI_ISL_447945 | Department of Pathology, University of Cambridge | COVID-19 Genomics UK (COG-UK) Consortium |
| EPI_ISL_447946 | Department of Pathology, University of Cambridge | COVID-19 Genomics UK (COG-UK) Consortium |
| EPI_ISL_447947 | Department of Pathology, University of Cambridge | COVID-19 Genomics UK (COG-UK) Consortium |
| EPI_ISL_447948 | Department of Pathology, University of Cambridge | COVID-19 Genomics UK (COG-UK) Consortium |
| EPI_ISL_447949 | Department of Pathology, University of Cambridge | COVID-19 Genomics UK (COG-UK) Consortium |
| EPI_ISL_447950 | Department of Pathology, University of Cambridge | COVID-19 Genomics UK (COG-UK) Consortium |
| EPI_ISL_447951 | Department of Pathology, University of Cambridge | COVID-19 Genomics UK (COG-UK) Consortium |
| EPI_ISL_447952 | Department of Pathology, University of Cambridge | COVID-19 Genomics UK (COG-UK) Consortium |
| EPI_ISL_447953 | Department of Pathology, University of Cambridge | COVID-19 Genomics UK (COG-UK) Consortium |
| EPI_ISL_447954 | Department of Pathology, University of Cambridge | COVID-19 Genomics UK (COG-UK) Consortium |
| EPI_ISL_447955 | Department of Pathology, University of Cambridge | COVID-19 Genomics UK (COG-UK) Consortium |
| EPI_ISL_447956 | Department of Pathology, University of Cambridge | COVID-19 Genomics UK (COG-UK) Consortium |
| EPI_ISL_447957 | Department of Pathology, University of Cambridge | COVID-19 Genomics UK (COG-UK) Consortium |
| EPI_ISL_447958 | Department of Pathology, University of Cambridge | COVID-19 Genomics UK (COG-UK) Consortium |
| EPI_ISL_447959 | Department of Pathology, University of Cambridge | COVID-19 Genomics UK (COG-UK) Consortium |
| EPI_ISL_447960 | Department of Pathology, University of Cambridge | COVID-19 Genomics UK (COG-UK) Consortium |
| EPI_ISL_447961 | Department of Pathology, University of Cambridge | COVID-19 Genomics UK (COG-UK) Consortium |
| EPI_ISL_447962 | Department of Pathology, University of Cambridge | COVID-19 Genomics UK (COG-UK) Consortium |
| EPI_ISL_447963 | Department of Pathology, University of Cambridge | COVID-19 Genomics UK (COG-UK) Consortium |
| EPI_ISL_447964 | Department of Pathology, University of Cambridge | COVID-19 Genomics UK (COG-UK) Consortium |
| EPI_ISL_447965 | Department of Pathology, University of Cambridge | COVID-19 Genomics UK (COG-UK) Consortium |
| EPI_ISL_447966 | Department of Pathology, University of Cambridge | COVID-19 Genomics UK (COG-UK) Consortium |
| EPI_ISL_447967 | Department of Pathology, University of Cambridge | COVID-19 Genomics UK (COG-UK) Consortium |
| EPI_ISL_447968 | Department of Pathology, University of Cambridge | COVID-19 Genomics UK (COG-UK) Consortium |
| EPI_ISL_447969 | Department of Pathology, University of Cambridge | COVID-19 Genomics UK (COG-UK) Consortium |
| EPI_ISL_447970 | Department of Pathology, University of Cambridge | COVID-19 Genomics UK (COG-UK) Consortium |
| EPI_ISL_447971 | Department of Pathology, University of Cambridge | COVID-19 Genomics UK (COG-UK) Consortium |
| EPI_ISL_447972 | Department of Pathology, University of Cambridge | COVID-19 Genomics UK (COG-UK) Consortium |
| EPI_ISL_447973 | Department of Pathology, University of Cambridge | COVID-19 Genomics UK (COG-UK) Consortium |
| EPI_ISL_447974 | Department of Pathology, University of Cambridge | COVID-19 Genomics UK (COG-UK) Consortium |
| EPI_ISL_447975 | Department of Pathology, University of Cambridge | COVID-19 Genomics UK (COG-UK) Consortium |
| EPI_ISL_447976 | Department of Pathology, University of Cambridge | COVID-19 Genomics UK (COG-UK) Consortium |
| EPI_ISL_447977 | Department of Pathology, University of Cambridge | COVID-19 Genomics UK (COG-UK) Consortium |

| EPI_Accession_ID | Originating Laboratory | Submitting Laboratory |
|---|---|---|
| EPI_ISL_447978 | Department of Pathology, University of Cambridge | COVID-19 Genomics UK (COG-UK) Consortium |
| EPI_ISL_447979 | Department of Pathology, University of Cambridge | COVID-19 Genomics UK (COG-UK) Consortium |
| EPI_ISL_447980 | Department of Pathology, University of Cambridge | COVID-19 Genomics UK (COG-UK) Consortium |
| EPI_ISL_447981 | Department of Pathology, University of Cambridge | COVID-19 Genomics UK (COG-UK) Consortium |
| EPI_ISL_447982 | Department of Pathology, University of Cambridge | COVID-19 Genomics UK (COG-UK) Consortium |
| EPI_ISL_447983 | Department of Pathology, University of Cambridge | COVID-19 Genomics UK (COG-UK) Consortium |
| EPI_ISL_447987 | Department of Pathology, University of Cambridge | COVID-19 Genomics UK (COG-UK) Consortium |
| EPI_ISL_447988 | Department of Pathology, University of Cambridge | COVID-19 Genomics UK (COG-UK) Consortium |
| EPI_ISL_447989 | Department of Pathology, University of Cambridge | COVID-19 Genomics UK (COG-UK) Consortium |
| EPI_ISL_447990 | Department of Pathology, University of Cambridge | COVID-19 Genomics UK (COG-UK) Consortium |
| EPI_ISL_447991 | Department of Pathology, University of Cambridge | COVID-19 Genomics UK (COG-UK) Consortium |
| EPI_ISL_447992 | Department of Pathology, University of Cambridge | COVID-19 Genomics UK (COG-UK) Consortium |
| EPI_ISL_447993 | Department of Pathology, University of Cambridge | COVID-19 Genomics UK (COG-UK) Consortium |
| EPI_ISL_447994 | Department of Pathology, University of Cambridge | COVID-19 Genomics UK (COG-UK) Consortium |
| EPI_ISL_447995 | Department of Pathology, University of Cambridge | COVID-19 Genomics UK (COG-UK) Consortium |
| EPI_ISL_447996 | Department of Pathology, University of Cambridge | COVID-19 Genomics UK (COG-UK) Consortium |
| EPI_ISL_447997 | Department of Pathology, University of Cambridge | COVID-19 Genomics UK (COG-UK) Consortium |
| EPI_ISL_447998 | Department of Pathology, University of Cambridge | COVID-19 Genomics UK (COG-UK) Consortium |
| EPI_ISL_447999 | Department of Pathology, University of Cambridge | COVID-19 Genomics UK (COG-UK) Consortium |
| EPI_ISL_448000 | Department of Pathology, University of Cambridge | COVID-19 Genomics UK (COG-UK) Consortium |
| EPI_ISL_448001 | Department of Pathology, University of Cambridge | COVID-19 Genomics UK (COG-UK) Consortium |
| EPI_ISL_448002 | Department of Pathology, University of Cambridge | COVID-19 Genomics UK (COG-UK) Consortium |
| EPI_ISL_448003 | Department of Pathology, University of Cambridge | COVID-19 Genomics UK (COG-UK) Consortium |
| EPI_ISL_448004 | Department of Pathology, University of Cambridge | COVID-19 Genomics UK (COG-UK) Consortium |
| EPI_ISL_448005 | Department of Pathology, University of Cambridge | COVID-19 Genomics UK (COG-UK) Consortium |
| EPI_ISL_448006 | Department of Pathology, University of Cambridge | COVID-19 Genomics UK (COG-UK) Consortium |
| EPI_ISL_448007 | Department of Pathology, University of Cambridge | COVID-19 Genomics UK (COG-UK) Consortium |
| EPI_ISL_448008 | Department of Pathology, University of Cambridge | COVID-19 Genomics UK (COG-UK) Consortium |
| EPI_ISL_448009 | Department of Pathology, University of Cambridge | COVID-19 Genomics UK (COG-UK) Consortium |
| EPI_ISL_448010 | Department of Pathology, University of Cambridge | COVID-19 Genomics UK (COG-UK) Consortium |
| EPI_ISL_448011 | Department of Pathology, University of Cambridge | COVID-19 Genomics UK (COG-UK) Consortium |
| EPI_ISL_448012 | Department of Pathology, University of Cambridge | COVID-19 Genomics UK (COG-UK) Consortium |
| EPI_ISL_448013 | Department of Pathology, University of Cambridge | COVID-19 Genomics UK (COG-UK) Consortium |

| EPI_Accession_ID | Originating Laboratory | Submitting Laboratory |
|---|---|---|
| EPI_ISL_448015 | Department of Pathology, University of Cambridge | COVID-19 Genomics UK (COG-UK) Consortium |
| EPI_ISL_448016 | Department of Pathology, University of Cambridge | COVID-19 Genomics UK (COG-UK) Consortium |
| EPI_ISL_448017 | Department of Pathology, University of Cambridge | COVID-19 Genomics UK (COG-UK) Consortium |
| EPI_ISL_448018 | Department of Pathology, University of Cambridge | COVID-19 Genomics UK (COG-UK) Consortium |
| EPI_ISL_448019 | Department of Pathology, University of Cambridge | COVID-19 Genomics UK (COG-UK) Consortium |
| EPI_ISL_448020 | Department of Pathology, University of Cambridge | COVID-19 Genomics UK (COG-UK) Consortium |
| EPI_ISL_448021 | Department of Pathology, University of Cambridge | COVID-19 Genomics UK (COG-UK) Consortium |
| EPI_ISL_448022 | Department of Pathology, University of Cambridge | COVID-19 Genomics UK (COG-UK) Consortium |
| EPI_ISL_448023 | Department of Pathology, University of Cambridge | COVID-19 Genomics UK (COG-UK) Consortium |
| EPI_ISL_448024 | Department of Pathology, University of Cambridge | COVID-19 Genomics UK (COG-UK) Consortium |
| EPI_ISL_448025 | Department of Pathology, University of Cambridge | COVID-19 Genomics UK (COG-UK) Consortium |
| EPI_ISL_448026 | Department of Pathology, University of Cambridge | COVID-19 Genomics UK (COG-UK) Consortium |
| EPI_ISL_448027 | Department of Pathology, University of Cambridge | COVID-19 Genomics UK (COG-UK) Consortium |
| EPI_ISL_448028 | Department of Pathology, University of Cambridge | COVID-19 Genomics UK (COG-UK) Consortium |
| EPI_ISL_448029 | Department of Pathology, University of Cambridge | COVID-19 Genomics UK (COG-UK) Consortium |
| EPI_ISL_448031 | Department of Pathology, University of Cambridge | COVID-19 Genomics UK (COG-UK) Consortium |
| EPI_ISL_448032 | Department of Pathology, University of Cambridge | COVID-19 Genomics UK (COG-UK) Consortium |
| EPI_ISL_448033 | Department of Pathology, University of Cambridge | COVID-19 Genomics UK (COG-UK) Consortium |
| EPI_ISL_448034 | Department of Pathology, University of Cambridge | COVID-19 Genomics UK (COG-UK) Consortium |
| EPI_ISL_448035 | Department of Pathology, University of Cambridge | COVID-19 Genomics UK (COG-UK) Consortium |
| EPI_ISL_448036 | Department of Pathology, University of Cambridge | COVID-19 Genomics UK (COG-UK) Consortium |
| EPI_ISL_448037 | Department of Pathology, University of Cambridge | COVID-19 Genomics UK (COG-UK) Consortium |
| EPI_ISL_448039 | Department of Pathology, University of Cambridge | COVID-19 Genomics UK (COG-UK) Consortium |
| EPI_ISL_448040 | Department of Pathology, University of Cambridge | COVID-19 Genomics UK (COG-UK) Consortium |
| EPI_ISL_448041 | Department of Pathology, University of Cambridge | COVID-19 Genomics UK (COG-UK) Consortium |
| EPI_ISL_448042 | Department of Pathology, University of Cambridge | COVID-19 Genomics UK (COG-UK) Consortium |
| EPI_ISL_448043 | Department of Pathology, University of Cambridge | COVID-19 Genomics UK (COG-UK) Consortium |
| EPI_ISL_448044 | Department of Pathology, University of Cambridge | COVID-19 Genomics UK (COG-UK) Consortium |
| EPI_ISL_448045 | Department of Pathology, University of Cambridge | COVID-19 Genomics UK (COG-UK) Consortium |
| EPI_ISL_448046 | Department of Pathology, University of Cambridge | COVID-19 Genomics UK (COG-UK) Consortium |
| EPI_ISL_448047 | Department of Pathology, University of Cambridge | COVID-19 Genomics UK (COG-UK) Consortium |
| EPI_ISL_448048 | Department of Pathology, University of Cambridge | COVID-19 Genomics UK (COG-UK) Consortium |
| EPI_ISL_448049 | Department of Pathology, University of Cambridge | COVID-19 Genomics UK (COG-UK) Consortium |

| EPI_Accession_ID | Originating Laboratory | Submitting Laboratory |
|---|---|---|
| EPI_ISL_448050 | Department of Pathology, University of Cambridge | COVID-19 Genomics UK (COG-UK) Consortium |
| EPI_ISL_448051 | Department of Pathology, University of Cambridge | COVID-19 Genomics UK (COG-UK) Consortium |
| EPI_ISL_448052 | Department of Pathology, University of Cambridge | COVID-19 Genomics UK (COG-UK) Consortium |
| EPI_ISL_448053 | Department of Pathology, University of Cambridge | COVID-19 Genomics UK (COG-UK) Consortium |
| EPI_ISL_448054 | Department of Pathology, University of Cambridge | COVID-19 Genomics UK (COG-UK) Consortium |
| EPI_ISL_448055 | Department of Pathology, University of Cambridge | COVID-19 Genomics UK (COG-UK) Consortium |
| EPI_ISL_448056 | Department of Pathology, University of Cambridge | COVID-19 Genomics UK (COG-UK) Consortium |
| EPI_ISL_448057 | Department of Pathology, University of Cambridge | COVID-19 Genomics UK (COG-UK) Consortium |
| EPI_ISL_448058 | Department of Pathology, University of Cambridge | COVID-19 Genomics UK (COG-UK) Consortium |
| EPI_ISL_448059 | Department of Pathology, University of Cambridge | COVID-19 Genomics UK (COG-UK) Consortium |
| EPI_ISL_448060 | Department of Pathology, University of Cambridge | COVID-19 Genomics UK (COG-UK) Consortium |
| EPI_ISL_448061 | Department of Pathology, University of Cambridge | COVID-19 Genomics UK (COG-UK) Consortium |
| EPI_ISL_448062 | Department of Pathology, University of Cambridge | COVID-19 Genomics UK (COG-UK) Consortium |
| EPI_ISL_448063 | Department of Pathology, University of Cambridge | COVID-19 Genomics UK (COG-UK) Consortium |
| EPI_ISL_448064 | Department of Pathology, University of Cambridge | COVID-19 Genomics UK (COG-UK) Consortium |
| EPI_ISL_448065 | Department of Pathology, University of Cambridge | COVID-19 Genomics UK (COG-UK) Consortium |
| EPI_ISL_448066 | Department of Pathology, University of Cambridge | COVID-19 Genomics UK (COG-UK) Consortium |
| EPI_ISL_448067 | Department of Pathology, University of Cambridge | COVID-19 Genomics UK (COG-UK) Consortium |
| EPI_ISL_448068 | Department of Pathology, University of Cambridge | COVID-19 Genomics UK (COG-UK) Consortium |
| EPI_ISL_448069 | Department of Pathology, University of Cambridge | COVID-19 Genomics UK (COG-UK) Consortium |
| EPI_ISL_448070 | Department of Pathology, University of Cambridge | COVID-19 Genomics UK (COG-UK) Consortium |
| EPI_ISL_448071 | Department of Pathology, University of Cambridge | COVID-19 Genomics UK (COG-UK) Consortium |
| EPI_ISL_448074 | Department of Pathology, University of Cambridge | COVID-19 Genomics UK (COG-UK) Consortium |
| EPI_ISL_448075 | Department of Pathology, University of Cambridge | COVID-19 Genomics UK (COG-UK) Consortium |
| EPI_ISL_448076 | Department of Pathology, University of Cambridge | COVID-19 Genomics UK (COG-UK) Consortium |
| EPI_ISL_448077 | Department of Pathology, University of Cambridge | COVID-19 Genomics UK (COG-UK) Consortium |
| EPI_ISL_448078 | Department of Pathology, University of Cambridge | COVID-19 Genomics UK (COG-UK) Consortium |
| EPI_ISL_448079 | Department of Pathology, University of Cambridge | COVID-19 Genomics UK (COG-UK) Consortium |
| EPI_ISL_448080 | Department of Pathology, University of Cambridge | COVID-19 Genomics UK (COG-UK) Consortium |
| EPI_ISL_448081 | Department of Pathology, University of Cambridge | COVID-19 Genomics UK (COG-UK) Consortium |
| EPI_ISL_448082 | Department of Pathology, University of Cambridge | COVID-19 Genomics UK (COG-UK) Consortium |
| EPI_ISL_448083 | Department of Pathology, University of Cambridge | COVID-19 Genomics UK (COG-UK) Consortium |
| EPI_ISL_448084 | Department of Pathology, University of Cambridge | COVID-19 Genomics UK (COG-UK) Consortium |

| EPI_Accession_ID | Originating Laboratory | Submitting Laboratory |
|---|---|---|
| EPI_ISL_448086 | Department of Pathology, University of Cambridge | COVID-19 Genomics UK (COG-UK) Consortium |
| EPI_ISL_448087 | Department of Pathology, University of Cambridge | COVID-19 Genomics UK (COG-UK) Consortium |
| EPI_ISL_448088 | Department of Pathology, University of Cambridge | COVID-19 Genomics UK (COG-UK) Consortium |
| EPI_ISL_448089 | Department of Pathology, University of Cambridge | COVID-19 Genomics UK (COG-UK) Consortium |
| EPI_ISL_448090 | Department of Pathology, University of Cambridge | COVID-19 Genomics UK (COG-UK) Consortium |
| EPI_ISL_448092 | Department of Pathology, University of Cambridge | COVID-19 Genomics UK (COG-UK) Consortium |
| EPI_ISL_448096 | Department of Pathology, University of Cambridge | COVID-19 Genomics UK (COG-UK) Consortium |
| EPI_ISL_448097 | Department of Pathology, University of Cambridge | COVID-19 Genomics UK (COG-UK) Consortium |
| EPI_ISL_448098 | Department of Pathology, University of Cambridge | COVID-19 Genomics UK (COG-UK) Consortium |
| EPI_ISL_448099 | Department of Pathology, University of Cambridge | COVID-19 Genomics UK (COG-UK) Consortium |
| EPI_ISL_448100 | Department of Pathology, University of Cambridge | COVID-19 Genomics UK (COG-UK) Consortium |
| EPI_ISL_448101 | Department of Pathology, University of Cambridge | COVID-19 Genomics UK (COG-UK) Consortium |
| EPI_ISL_448102 | Department of Pathology, University of Cambridge | COVID-19 Genomics UK (COG-UK) Consortium |
| EPI_ISL_448103 | Department of Pathology, University of Cambridge | COVID-19 Genomics UK (COG-UK) Consortium |
| EPI_ISL_448104 | Department of Pathology, University of Cambridge | COVID-19 Genomics UK (COG-UK) Consortium |
| EPI_ISL_448106 | Department of Pathology, University of Cambridge | COVID-19 Genomics UK (COG-UK) Consortium |
| EPI_ISL_448107 | Department of Pathology, University of Cambridge | COVID-19 Genomics UK (COG-UK) Consortium |
| EPI_ISL_448108 | Department of Pathology, University of Cambridge | COVID-19 Genomics UK (COG-UK) Consortium |
| EPI_ISL_448109 | Department of Pathology, University of Cambridge | COVID-19 Genomics UK (COG-UK) Consortium |
| EPI_ISL_448110 | Department of Pathology, University of Cambridge | COVID-19 Genomics UK (COG-UK) Consortium |
| EPI_ISL_448111 | Department of Pathology, University of Cambridge | COVID-19 Genomics UK (COG-UK) Consortium |
| EPI_ISL_448112 | Department of Pathology, University of Cambridge | COVID-19 Genomics UK (COG-UK) Consortium |
| EPI_ISL_448113 | Department of Pathology, University of Cambridge | COVID-19 Genomics UK (COG-UK) Consortium |
| EPI_ISL_448114 | Department of Pathology, University of Cambridge | COVID-19 Genomics UK (COG-UK) Consortium |
| EPI_ISL_448115 | Department of Pathology, University of Cambridge | COVID-19 Genomics UK (COG-UK) Consortium |
| EPI_ISL_448116 | West of Scotland Specialist Virology Centre, NHSGGC / MRC-University of Glasgow Centre for Virus Research | COVID-19 Genomics UK (COG-UK) Consortium |
| EPI_ISL_448117 | West of Scotland Specialist Virology Centre, NHSGGC / MRC-University of Glasgow Centre for Virus Research | COVID-19 Genomics UK (COG-UK) Consortium |
| EPI_ISL_448118 | West of Scotland Specialist Virology Centre, NHSGGC / MRC-University of Glasgow Centre for Virus Research | COVID-19 Genomics UK (COG-UK) Consortium |
| EPI_ISL_448119 | West of Scotland Specialist Virology Centre, NHSGGC / MRC-University of Glasgow Centre for Virus Research | COVID-19 Genomics UK (COG-UK) Consortium |

| EPI_Accession_ID | Originating Laboratory | Submitting Laboratory |
|---|---|---|
| EPI_ISL_448120 | West of Scotland Specialist Virology Centre, NHSGGC / MRC-University of Glasgow Centre for Virus Research | COVID-19 Genomics UK (COG-UK) Consortium |
| EPI_ISL_448121 | West of Scotland Specialist Virology Centre, NHSGGC / MRC-University of Glasgow Centre for Virus Research | COVID-19 Genomics UK (COG-UK) Consortium |
| EPI_ISL_448122 | West of Scotland Specialist Virology Centre, NHSGGC / MRC-University of Glasgow Centre for Virus Research | COVID-19 Genomics UK (COG-UK) Consortium |
| EPI_ISL_448123 | West of Scotland Specialist Virology Centre, NHSGGC / MRC-University of Glasgow Centre for Virus Research | COVID-19 Genomics UK (COG-UK) Consortium |
| EPI_ISL_448124 | West of Scotland Specialist Virology Centre, NHSGGC / MRC-University of Glasgow Centre for Virus Research | COVID-19 Genomics UK (COG-UK) Consortium |
| EPI_ISL_448125 | West of Scotland Specialist Virology Centre, NHSGGC / MRC-University of Glasgow Centre for Virus Research | COVID-19 Genomics UK (COG-UK) Consortium |
| EPI_ISL_448126 | West of Scotland Specialist Virology Centre, NHSGGC / MRC-University of Glasgow Centre for Virus Research | COVID-19 Genomics UK (COG-UK) Consortium |
| EPI_ISL_448127 | West of Scotland Specialist Virology Centre, NHSGGC / MRC-University of Glasgow Centre for Virus Research | COVID-19 Genomics UK (COG-UK) Consortium |
| EPI_ISL_448128 | West of Scotland Specialist Virology Centre, NHSGGC / MRC-University of Glasgow Centre for Virus Research | COVID-19 Genomics UK (COG-UK) Consortium |
| EPI_ISL_448129 | West of Scotland Specialist Virology Centre, NHSGGC / MRC-University of Glasgow Centre for Virus Research | COVID-19 Genomics UK (COG-UK) Consortium |
| EPI_ISL_448130 | West of Scotland Specialist Virology Centre, NHSGGC / MRC-University of Glasgow Centre for Virus Research | COVID-19 Genomics UK (COG-UK) Consortium |
| EPI_ISL_448131 | West of Scotland Specialist Virology Centre, NHSGGC / MRC-University of Glasgow Centre for Virus Research | COVID-19 Genomics UK (COG-UK) Consortium |
| EPI_ISL_448132 | West of Scotland Specialist Virology Centre, NHSGGC / MRC-University of Glasgow Centre for Virus Research | COVID-19 Genomics UK (COG-UK) Consortium |
| EPI_ISL_448133 | West of Scotland Specialist Virology Centre, NHSGGC / MRC-University of Glasgow Centre for Virus Research | COVID-19 Genomics UK (COG-UK) Consortium |
| EPI_ISL_448134 | West of Scotland Specialist Virology Centre, NHSGGC / MRC-University of Glasgow Centre for Virus Research | COVID-19 Genomics UK (COG-UK) Consortium |
| EPI_ISL_448135 | West of Scotland Specialist Virology Centre, NHSGGC / MRC-University of Glasgow Centre for Virus Research | COVID-19 Genomics UK (COG-UK) Consortium |

| EPI_Accession_ID | Originating Laboratory | Submitting Laboratory |
|---|---|---|
| EPI_ISL_448136 | West of Scotland Specialist Virology Centre, NHSGGC / MRC-University of Glasgow Centre for Virus Research | COVID-19 Genomics UK (COG-UK) Consortium |
| EPI_ISL_448137 | West of Scotland Specialist Virology Centre, NHSGGC / MRC-University of Glasgow Centre for Virus Research | COVID-19 Genomics UK (COG-UK) Consortium |
| EPI_ISL_448138 | West of Scotland Specialist Virology Centre, NHSGGC / MRC-University of Glasgow Centre for Virus Research | COVID-19 Genomics UK (COG-UK) Consortium |
| EPI_ISL_448139 | West of Scotland Specialist Virology Centre, NHSGGC / MRC-University of Glasgow Centre for Virus Research | COVID-19 Genomics UK (COG-UK) Consortium |
| EPI_ISL_448140 | West of Scotland Specialist Virology Centre, NHSGGC / MRC-University of Glasgow Centre for Virus Research | COVID-19 Genomics UK (COG-UK) Consortium |
| EPI_ISL_448141 | West of Scotland Specialist Virology Centre, NHSGGC / MRC-University of Glasgow Centre for Virus Research | COVID-19 Genomics UK (COG-UK) Consortium |
| EPI_ISL_448142 | West of Scotland Specialist Virology Centre, NHSGGC / MRC-University of Glasgow Centre for Virus Research | COVID-19 Genomics UK (COG-UK) Consortium |
| EPI_ISL_448143 | West of Scotland Specialist Virology Centre, NHSGGC / MRC-University of Glasgow Centre for Virus Research | COVID-19 Genomics UK (COG-UK) Consortium |
| EPI_ISL_448144 | West of Scotland Specialist Virology Centre, NHSGGC / MRC-University of Glasgow Centre for Virus Research | COVID-19 Genomics UK (COG-UK) Consortium |
| EPI_ISL_448145 | West of Scotland Specialist Virology Centre, NHSGGC / MRC-University of Glasgow Centre for Virus Research | COVID-19 Genomics UK (COG-UK) Consortium |
| EPI_ISL_448146 | West of Scotland Specialist Virology Centre, NHSGGC / MRC-University of Glasgow Centre for Virus Research | COVID-19 Genomics UK (COG-UK) Consortium |
| EPI_ISL_448147 | West of Scotland Specialist Virology Centre, NHSGGC / MRC-University of Glasgow Centre for Virus Research | COVID-19 Genomics UK (COG-UK) Consortium |
| EPI_ISL_448148 | West of Scotland Specialist Virology Centre, NHSGGC / MRC-University of Glasgow Centre for Virus Research | COVID-19 Genomics UK (COG-UK) Consortium |
| EPI_ISL_448149 | West of Scotland Specialist Virology Centre, NHSGGC / MRC-University of Glasgow Centre for Virus Research | COVID-19 Genomics UK (COG-UK) Consortium |
| EPI_ISL_448150 | West of Scotland Specialist Virology Centre, NHSGGC / MRC-University of Glasgow Centre for Virus Research | COVID-19 Genomics UK (COG-UK) Consortium |
| EPI_ISL_448151 | West of Scotland Specialist Virology Centre, NHSGGC / MRC-University of Glasgow Centre for Virus Research | COVID-19 Genomics UK (COG-UK) Consortium |

| EPI_Accession_ID | Originating Laboratory | Submitting Laboratory |
|---|---|---|
| EPI_ISL_448152 | West of Scotland Specialist Virology Centre, NHSGGC / MRC-University of Glasgow Centre for Virus Research | COVID-19 Genomics UK (COG-UK) Consortium |
| EPI_ISL_448153 | West of Scotland Specialist Virology Centre, NHSGGC / MRC-University of Glasgow Centre for Virus Research | COVID-19 Genomics UK (COG-UK) Consortium |
| EPI_ISL_448154 | West of Scotland Specialist Virology Centre, NHSGGC / MRC-University of Glasgow Centre for Virus Research | COVID-19 Genomics UK (COG-UK) Consortium |
| EPI_ISL_448155 | West of Scotland Specialist Virology Centre, NHSGGC / MRC-University of Glasgow Centre for Virus Research | COVID-19 Genomics UK (COG-UK) Consortium |
| EPI_ISL_448156 | West of Scotland Specialist Virology Centre, NHSGGC / MRC-University of Glasgow Centre for Virus Research | COVID-19 Genomics UK (COG-UK) Consortium |
| EPI_ISL_448157 | West of Scotland Specialist Virology Centre, NHSGGC / MRC-University of Glasgow Centre for Virus Research | COVID-19 Genomics UK (COG-UK) Consortium |
| EPI_ISL_448158 | West of Scotland Specialist Virology Centre, NHSGGC / MRC-University of Glasgow Centre for Virus Research | COVID-19 Genomics UK (COG-UK) Consortium |
| EPI_ISL_448159 | West of Scotland Specialist Virology Centre, NHSGGC / MRC-University of Glasgow Centre for Virus Research | COVID-19 Genomics UK (COG-UK) Consortium |
| EPI_ISL_448160 | West of Scotland Specialist Virology Centre, NHSGGC / MRC-University of Glasgow Centre for Virus Research | COVID-19 Genomics UK (COG-UK) Consortium |
| EPI_ISL_448161 | West of Scotland Specialist Virology Centre, NHSGGC / MRC-University of Glasgow Centre for Virus Research | COVID-19 Genomics UK (COG-UK) Consortium |
| EPI_ISL_448162 | West of Scotland Specialist Virology Centre, NHSGGC / MRC-University of Glasgow Centre for Virus Research | COVID-19 Genomics UK (COG-UK) Consortium |
| EPI_ISL_448163 | West of Scotland Specialist Virology Centre, NHSGGC / MRC-University of Glasgow Centre for Virus Research | COVID-19 Genomics UK (COG-UK) Consortium |
| EPI_ISL_448164 | West of Scotland Specialist Virology Centre, NHSGGC / MRC-University of Glasgow Centre for Virus Research | COVID-19 Genomics UK (COG-UK) Consortium |
| EPI_ISL_448165 | West of Scotland Specialist Virology Centre, NHSGGC / MRC-University of Glasgow Centre for Virus Research | COVID-19 Genomics UK (COG-UK) Consortium |
| EPI_ISL_448166 | West of Scotland Specialist Virology Centre, NHSGGC / MRC-University of Glasgow Centre for Virus Research | COVID-19 Genomics UK (COG-UK) Consortium |
| EPI_ISL_448167 | West of Scotland Specialist Virology Centre, NHSGGC / MRC-University of Glasgow Centre for Virus Research | COVID-19 Genomics UK (COG-UK) Consortium |

| EPI_Accession_ID | Originating Laboratory | Submitting Laboratory |
|---|---|---|
| EPI_ISL_448168 | West of Scotland Specialist Virology Centre, NHSGGC / MRC-University of Glasgow Centre for Virus Research | COVID-19 Genomics UK (COG-UK) Consortium |
| EPI_ISL_448169 | West of Scotland Specialist Virology Centre, NHSGGC / MRC-University of Glasgow Centre for Virus Research | COVID-19 Genomics UK (COG-UK) Consortium |
| EPI_ISL_448170 | West of Scotland Specialist Virology Centre, NHSGGC / MRC-University of Glasgow Centre for Virus Research | COVID-19 Genomics UK (COG-UK) Consortium |
| EPI_ISL_448171 | West of Scotland Specialist Virology Centre, NHSGGC / MRC-University of Glasgow Centre for Virus Research | COVID-19 Genomics UK (COG-UK) Consortium |
| EPI_ISL_448172 | West of Scotland Specialist Virology Centre, NHSGGC / MRC-University of Glasgow Centre for Virus Research | COVID-19 Genomics UK (COG-UK) Consortium |
| EPI_ISL_448173 | West of Scotland Specialist Virology Centre, NHSGGC / MRC-University of Glasgow Centre for Virus Research | COVID-19 Genomics UK (COG-UK) Consortium |
| EPI_ISL_448174 | West of Scotland Specialist Virology Centre, NHSGGC / MRC-University of Glasgow Centre for Virus Research | COVID-19 Genomics UK (COG-UK) Consortium |
| EPI_ISL_448175 | West of Scotland Specialist Virology Centre, NHSGGC / MRC-University of Glasgow Centre for Virus Research | COVID-19 Genomics UK (COG-UK) Consortium |
| EPI_ISL_448176 | West of Scotland Specialist Virology Centre, NHSGGC / MRC-University of Glasgow Centre for Virus Research | COVID-19 Genomics UK (COG-UK) Consortium |
| EPI_ISL_448177 | West of Scotland Specialist Virology Centre, NHSGGC / MRC-University of Glasgow Centre for Virus Research | COVID-19 Genomics UK (COG-UK) Consortium |
| EPI_ISL_448178 | West of Scotland Specialist Virology Centre, NHSGGC / MRC-University of Glasgow Centre for Virus Research | COVID-19 Genomics UK (COG-UK) Consortium |
| EPI_ISL_448179 | West of Scotland Specialist Virology Centre, NHSGGC / MRC-University of Glasgow Centre for Virus Research | COVID-19 Genomics UK (COG-UK) Consortium |
| EPI_ISL_448180 | West of Scotland Specialist Virology Centre, NHSGGC / MRC-University of Glasgow Centre for Virus Research | COVID-19 Genomics UK (COG-UK) Consortium |
| EPI_ISL_448181 | West of Scotland Specialist Virology Centre, NHSGGC / MRC-University of Glasgow Centre for Virus Research | COVID-19 Genomics UK (COG-UK) Consortium |
| EPI_ISL_448182 | West of Scotland Specialist Virology Centre, NHSGGC / MRC-University of Glasgow Centre for Virus Research | COVID-19 Genomics UK (COG-UK) Consortium |
| EPI_ISL_448183 | West of Scotland Specialist Virology Centre, NHSGGC / MRC-University of Glasgow Centre for Virus Research | COVID-19 Genomics UK (COG-UK) Consortium |

| EPI_Accession_ID | Originating Laboratory | Submitting Laboratory |
|---|---|---|
| EPI_ISL_448184 | West of Scotland Specialist Virology Centre, NHSGGC / MRC-University of Glasgow Centre for Virus Research | COVID-19 Genomics UK (COG-UK) Consortium |
| EPI_ISL_448185 | West of Scotland Specialist Virology Centre, NHSGGC / MRC-University of Glasgow Centre for Virus Research | COVID-19 Genomics UK (COG-UK) Consortium |
| EPI_ISL_448186 | West of Scotland Specialist Virology Centre, NHSGGC / MRC-University of Glasgow Centre for Virus Research | COVID-19 Genomics UK (COG-UK) Consortium |
| EPI_ISL_448187 | West of Scotland Specialist Virology Centre, NHSGGC / MRC-University of Glasgow Centre for Virus Research | COVID-19 Genomics UK (COG-UK) Consortium |
| EPI_ISL_448188 | West of Scotland Specialist Virology Centre, NHSGGC / MRC-University of Glasgow Centre for Virus Research | COVID-19 Genomics UK (COG-UK) Consortium |
| EPI_ISL_448189 | West of Scotland Specialist Virology Centre, NHSGGC / MRC-University of Glasgow Centre for Virus Research | COVID-19 Genomics UK (COG-UK) Consortium |
| EPI_ISL_448190 | West of Scotland Specialist Virology Centre, NHSGGC / MRC-University of Glasgow Centre for Virus Research | COVID-19 Genomics UK (COG-UK) Consortium |
| EPI_ISL_448191 | West of Scotland Specialist Virology Centre, NHSGGC / MRC-University of Glasgow Centre for Virus Research | COVID-19 Genomics UK (COG-UK) Consortium |
| EPI_ISL_448192 | West of Scotland Specialist Virology Centre, NHSGGC / MRC-University of Glasgow Centre for Virus Research | COVID-19 Genomics UK (COG-UK) Consortium |
| EPI_ISL_448193 | West of Scotland Specialist Virology Centre, NHSGGC / MRC-University of Glasgow Centre for Virus Research | COVID-19 Genomics UK (COG-UK) Consortium |
| EPI_ISL_448194 | West of Scotland Specialist Virology Centre, NHSGGC / MRC-University of Glasgow Centre for Virus Research | COVID-19 Genomics UK (COG-UK) Consortium |
| EPI_ISL_448195 | West of Scotland Specialist Virology Centre, NHSGGC / MRC-University of Glasgow Centre for Virus Research | COVID-19 Genomics UK (COG-UK) Consortium |
| EPI_ISL_448196 | West of Scotland Specialist Virology Centre, NHSGGC / MRC-University of Glasgow Centre for Virus Research | COVID-19 Genomics UK (COG-UK) Consortium |
| EPI_ISL_448197 | West of Scotland Specialist Virology Centre, NHSGGC / MRC-University of Glasgow Centre for Virus Research | COVID-19 Genomics UK (COG-UK) Consortium |
| EPI_ISL_448198 | West of Scotland Specialist Virology Centre, NHSGGC / MRC-University of Glasgow Centre for Virus Research | COVID-19 Genomics UK (COG-UK) Consortium |
| EPI_ISL_448199 | West of Scotland Specialist Virology Centre, NHSGGC / MRC-University of Glasgow Centre for Virus Research | COVID-19 Genomics UK (COG-UK) Consortium |

| EPI_Accession_ID | Originating Laboratory | Submitting Laboratory |
|---|---|---|
| EPI_ISL_448200 | West of Scotland Specialist Virology Centre, NHSGGC / MRC-University of Glasgow Centre for Virus Research | COVID-19 Genomics UK (COG-UK) Consortium |
| EPI_ISL_448201 | West of Scotland Specialist Virology Centre, NHSGGC / MRC-University of Glasgow Centre for Virus Research | COVID-19 Genomics UK (COG-UK) Consortium |
| EPI_ISL_448202 | West of Scotland Specialist Virology Centre, NHSGGC / MRC-University of Glasgow Centre for Virus Research | COVID-19 Genomics UK (COG-UK) Consortium |
| EPI_ISL_448203 | West of Scotland Specialist Virology Centre, NHSGGC / MRC-University of Glasgow Centre for Virus Research | COVID-19 Genomics UK (COG-UK) Consortium |
| EPI_ISL_448204 | West of Scotland Specialist Virology Centre, NHSGGC / MRC-University of Glasgow Centre for Virus Research | COVID-19 Genomics UK (COG-UK) Consortium |
| EPI_ISL_448205 | West of Scotland Specialist Virology Centre, NHSGGC / MRC-University of Glasgow Centre for Virus Research | COVID-19 Genomics UK (COG-UK) Consortium |
| EPI_ISL_448206 | West of Scotland Specialist Virology Centre, NHSGGC / MRC-University of Glasgow Centre for Virus Research | COVID-19 Genomics UK (COG-UK) Consortium |
| EPI_ISL_448207 | West of Scotland Specialist Virology Centre, NHSGGC / MRC-University of Glasgow Centre for Virus Research | COVID-19 Genomics UK (COG-UK) Consortium |
| EPI_ISL_448208 | West of Scotland Specialist Virology Centre, NHSGGC / MRC-University of Glasgow Centre for Virus Research | COVID-19 Genomics UK (COG-UK) Consortium |
| EPI_ISL_448209 | West of Scotland Specialist Virology Centre, NHSGGC / MRC-University of Glasgow Centre for Virus Research | COVID-19 Genomics UK (COG-UK) Consortium |
| EPI_ISL_448210 | West of Scotland Specialist Virology Centre, NHSGGC / MRC-University of Glasgow Centre for Virus Research | COVID-19 Genomics UK (COG-UK) Consortium |
| EPI_ISL_448211 | West of Scotland Specialist Virology Centre, NHSGGC / MRC-University of Glasgow Centre for Virus Research | COVID-19 Genomics UK (COG-UK) Consortium |
| EPI_ISL_448212 | West of Scotland Specialist Virology Centre, NHSGGC / MRC-University of Glasgow Centre for Virus Research | COVID-19 Genomics UK (COG-UK) Consortium |
| EPI_ISL_448213 | West of Scotland Specialist Virology Centre, NHSGGC / MRC-University of Glasgow Centre for Virus Research | COVID-19 Genomics UK (COG-UK) Consortium |
| EPI_ISL_448214 | West of Scotland Specialist Virology Centre, NHSGGC / MRC-University of Glasgow Centre for Virus Research | COVID-19 Genomics UK (COG-UK) Consortium |
| EPI_ISL_448215 | West of Scotland Specialist Virology Centre, NHSGGC / MRC-University of Glasgow Centre for Virus Research | COVID-19 Genomics UK (COG-UK) Consortium |

| EPI_Accession_ID | Originating Laboratory | Submitting Laboratory |
|---|---|---|
| EPI_ISL_448216 | West of Scotland Specialist Virology Centre, NHSGGC / MRC-University of Glasgow Centre for Virus Research | COVID-19 Genomics UK (COG-UK) Consortium |
| EPI_ISL_448217 | West of Scotland Specialist Virology Centre, NHSGGC / MRC-University of Glasgow Centre for Virus Research | COVID-19 Genomics UK (COG-UK) Consortium |
| EPI_ISL_448218 | West of Scotland Specialist Virology Centre, NHSGGC / MRC-University of Glasgow Centre for Virus Research | COVID-19 Genomics UK (COG-UK) Consortium |
| EPI_ISL_448219 | West of Scotland Specialist Virology Centre, NHSGGC / MRC-University of Glasgow Centre for Virus Research | COVID-19 Genomics UK (COG-UK) Consortium |
| EPI_ISL_448220 | West of Scotland Specialist Virology Centre, NHSGGC / MRC-University of Glasgow Centre for Virus Research | COVID-19 Genomics UK (COG-UK) Consortium |
| EPI_ISL_448221 | West of Scotland Specialist Virology Centre, NHSGGC / MRC-University of Glasgow Centre for Virus Research | COVID-19 Genomics UK (COG-UK) Consortium |
| EPI_ISL_448222 | Pasteur Insitute Ho Chi Minh City | National Key Laboratory of Gene Technology, Institute of Biotechnology, Vietnam Academy of Science and Technology |
| EPI_ISL_448223 | Quadram Institute Bioscience | COVID-19 Genomics UK (COG-UK) Consortium |
| EPI_ISL_448224 | Quadram Institute Bioscience | COVID-19 Genomics UK (COG-UK) Consortium |
| EPI_ISL_448225 | Quadram Institute Bioscience | COVID-19 Genomics UK (COG-UK) Consortium |
| EPI_ISL_448226 | Quadram Institute Bioscience | COVID-19 Genomics UK (COG-UK) Consortium |
| EPI_ISL_448227 | Quadram Institute Bioscience | COVID-19 Genomics UK (COG-UK) Consortium |
| EPI_ISL_448229 | Quadram Institute Bioscience | COVID-19 Genomics UK (COG-UK) Consortium |
| EPI_ISL_448230 | Quadram Institute Bioscience | COVID-19 Genomics UK (COG-UK) Consortium |
| EPI_ISL_448231 | Quadram Institute Bioscience | COVID-19 Genomics UK (COG-UK) Consortium |
| EPI_ISL_448232 | Quadram Institute Bioscience | COVID-19 Genomics UK (COG-UK) Consortium |
| EPI_ISL_448233 | Quadram Institute Bioscience | COVID-19 Genomics UK (COG-UK) Consortium |
| EPI_ISL_448234 | Quadram Institute Bioscience | COVID-19 Genomics UK (COG-UK) Consortium |
| EPI_ISL_448235 | Quadram Institute Bioscience | COVID-19 Genomics UK (COG-UK) Consortium |
| EPI_ISL_448237 | Quadram Institute Bioscience | COVID-19 Genomics UK (COG-UK) Consortium |
| EPI_ISL_448238 | Quadram Institute Bioscience | COVID-19 Genomics UK (COG-UK) Consortium |
| EPI_ISL_448239 | Quadram Institute Bioscience | COVID-19 Genomics UK (COG-UK) Consortium |
| EPI_ISL_448242 | Quadram Institute Bioscience | COVID-19 Genomics UK (COG-UK) Consortium |
| EPI_ISL_448243 | Quadram Institute Bioscience | COVID-19 Genomics UK (COG-UK) Consortium |
| EPI_ISL_448244 | Quadram Institute Bioscience | COVID-19 Genomics UK (COG-UK) Consortium |

| EPI_Accession_ID | Originating Laboratory | Submitting Laboratory |
|---|---|---|
| EPI_ISL_448245 | Quadram Institute Bioscience | COVID-19 Genomics UK (COG-UK) Consortium |
| EPI_ISL_448246 | Quadram Institute Bioscience | COVID-19 Genomics UK (COG-UK) Consortium |
| EPI_ISL_448248 | Quadram Institute Bioscience | COVID-19 Genomics UK (COG-UK) Consortium |
| EPI_ISL_448249 | Quadram Institute Bioscience | COVID-19 Genomics UK (COG-UK) Consortium |
| EPI_ISL_448251 | Quadram Institute Bioscience | COVID-19 Genomics UK (COG-UK) Consortium |
| EPI_ISL_448252 | Quadram Institute Bioscience | COVID-19 Genomics UK (COG-UK) Consortium |
| EPI_ISL_448254 | Quadram Institute Bioscience | COVID-19 Genomics UK (COG-UK) Consortium |
| EPI_ISL_448255 | Quadram Institute Bioscience | COVID-19 Genomics UK (COG-UK) Consortium |
| EPI_ISL_448256 | Quadram Institute Bioscience | COVID-19 Genomics UK (COG-UK) Consortium |
| EPI_ISL_448257 | Quadram Institute Bioscience | COVID-19 Genomics UK (COG-UK) Consortium |
| EPI_ISL_448258 | Quadram Institute Bioscience | COVID-19 Genomics UK (COG-UK) Consortium |
| EPI_ISL_448259 | Quadram Institute Bioscience | COVID-19 Genomics UK (COG-UK) Consortium |
| EPI_ISL_448262 | Quadram Institute Bioscience | COVID-19 Genomics UK (COG-UK) Consortium |
| EPI_ISL_448265 | Quadram Institute Bioscience | COVID-19 Genomics UK (COG-UK) Consortium |
| EPI_ISL_448266 | Quadram Institute Bioscience | COVID-19 Genomics UK (COG-UK) Consortium |
| EPI_ISL_448267 | Quadram Institute Bioscience | COVID-19 Genomics UK (COG-UK) Consortium |
| EPI_ISL_448268 | Quadram Institute Bioscience | COVID-19 Genomics UK (COG-UK) Consortium |
| EPI_ISL_448269 | Quadram Institute Bioscience | COVID-19 Genomics UK (COG-UK) Consortium |
| EPI_ISL_448270 | Quadram Institute Bioscience | COVID-19 Genomics UK (COG-UK) Consortium |
| EPI_ISL_448272 | Quadram Institute Bioscience | COVID-19 Genomics UK (COG-UK) Consortium |
| EPI_ISL_448273 | Quadram Institute Bioscience | COVID-19 Genomics UK (COG-UK) Consortium |
| EPI_ISL_448274 | Quadram Institute Bioscience | COVID-19 Genomics UK (COG-UK) Consortium |
| EPI_ISL_448276 | Quadram Institute Bioscience | COVID-19 Genomics UK (COG-UK) Consortium |
| EPI_ISL_448277 | Quadram Institute Bioscience | COVID-19 Genomics UK (COG-UK) Consortium |
| EPI_ISL_448278 | Quadram Institute Bioscience | COVID-19 Genomics UK (COG-UK) Consortium |
| EPI_ISL_448279 | Quadram Institute Bioscience | COVID-19 Genomics UK (COG-UK) Consortium |
| EPI_ISL_448280 | Quadram Institute Bioscience | COVID-19 Genomics UK (COG-UK) Consortium |
| EPI_ISL_448281 | Quadram Institute Bioscience | COVID-19 Genomics UK (COG-UK) Consortium |
| EPI_ISL_448282 | Quadram Institute Bioscience | COVID-19 Genomics UK (COG-UK) Consortium |
| EPI_ISL_448283 | Quadram Institute Bioscience | COVID-19 Genomics UK (COG-UK) Consortium |
| EPI_ISL_448284 | Quadram Institute Bioscience | COVID-19 Genomics UK (COG-UK) Consortium |
| EPI_ISL_448285 | Quadram Institute Bioscience | COVID-19 Genomics UK (COG-UK) Consortium |
| EPI_ISL_448286 | Quadram Institute Bioscience | COVID-19 Genomics UK (COG-UK) Consortium |

| EPI_Accession_ID | Originating Laboratory | Submitting Laboratory |
| --- | --- | --- |
| EPI_ISL_448287 | Quadram Institute Bioscience | COVID-19 Genomics UK (COG-UK) Consortium |
| EPI_ISL_448288 | Quadram Institute Bioscience | COVID-19 Genomics UK (COG-UK) Consortium |
| EPI_ISL_448289 | Quadram Institute Bioscience | COVID-19 Genomics UK (COG-UK) Consortium |
| EPI_ISL_448290 | Quadram Institute Bioscience | COVID-19 Genomics UK (COG-UK) Consortium |
| EPI_ISL_448291 | Quadram Institute Bioscience | COVID-19 Genomics UK (COG-UK) Consortium |
| EPI_ISL_448292 | Quadram Institute Bioscience | COVID-19 Genomics UK (COG-UK) Consortium |
| EPI_ISL_448293 | Quadram Institute Bioscience | COVID-19 Genomics UK (COG-UK) Consortium |
| EPI_ISL_448294 | Quadram Institute Bioscience | COVID-19 Genomics UK (COG-UK) Consortium |
| EPI_ISL_448295 | Quadram Institute Bioscience | COVID-19 Genomics UK (COG-UK) Consortium |
| EPI_ISL_448296 | Quadram Institute Bioscience | COVID-19 Genomics UK (COG-UK) Consortium |
| EPI_ISL_448297 | Quadram Institute Bioscience | COVID-19 Genomics UK (COG-UK) Consortium |
| EPI_ISL_448298 | Quadram Institute Bioscience | COVID-19 Genomics UK (COG-UK) Consortium |
| EPI_ISL_448299 | Quadram Institute Bioscience | COVID-19 Genomics UK (COG-UK) Consortium |
| EPI_ISL_448300 | Quadram Institute Bioscience | COVID-19 Genomics UK (COG-UK) Consortium |
| EPI_ISL_448301 | Quadram Institute Bioscience | COVID-19 Genomics UK (COG-UK) Consortium |
| EPI_ISL_448303 | Quadram Institute Bioscience | COVID-19 Genomics UK (COG-UK) Consortium |
| EPI_ISL_448304 | Quadram Institute Bioscience | COVID-19 Genomics UK (COG-UK) Consortium |
| EPI_ISL_448305 | Quadram Institute Bioscience | COVID-19 Genomics UK (COG-UK) Consortium |
| EPI_ISL_448307 | Quadram Institute Bioscience | COVID-19 Genomics UK (COG-UK) Consortium |
| EPI_ISL_448309 | Quadram Institute Bioscience | COVID-19 Genomics UK (COG-UK) Consortium |
| EPI_ISL_448310 | Quadram Institute Bioscience | COVID-19 Genomics UK (COG-UK) Consortium |
| EPI_ISL_448311 | Quadram Institute Bioscience | COVID-19 Genomics UK (COG-UK) Consortium |
| EPI_ISL_448312 | Quadram Institute Bioscience | COVID-19 Genomics UK (COG-UK) Consortium |
| EPI_ISL_448313 | Quadram Institute Bioscience | COVID-19 Genomics UK (COG-UK) Consortium |
| EPI_ISL_448314 | Quadram Institute Bioscience | COVID-19 Genomics UK (COG-UK) Consortium |
| EPI_ISL_448315 | Quadram Institute Bioscience | COVID-19 Genomics UK (COG-UK) Consortium |
| EPI_ISL_448316 | Quadram Institute Bioscience | COVID-19 Genomics UK (COG-UK) Consortium |
| EPI_ISL_448317 | Quadram Institute Bioscience | COVID-19 Genomics UK (COG-UK) Consortium |
| EPI_ISL_448318 | Quadram Institute Bioscience | COVID-19 Genomics UK (COG-UK) Consortium |
| EPI_ISL_448319 | Quadram Institute Bioscience | COVID-19 Genomics UK (COG-UK) Consortium |
| EPI_ISL_448320 | Quadram Institute Bioscience | COVID-19 Genomics UK (COG-UK) Consortium |
| EPI_ISL_448321 | Quadram Institute Bioscience | COVID-19 Genomics UK (COG-UK) Consortium |
| EPI_ISL_448322 | Quadram Institute Bioscience | COVID-19 Genomics UK (COG-UK) Consortium |

| EPI_Accession_ID | Originating Laboratory | Submitting Laboratory |
|---|---|---|
| EPI_ISL_448323 | Quadram Institute Bioscience | COVID-19 Genomics UK (COG-UK) Consortium |
| EPI_ISL_448324 | Quadram Institute Bioscience | COVID-19 Genomics UK (COG-UK) Consortium |
| EPI_ISL_448325 | Quadram Institute Bioscience | COVID-19 Genomics UK (COG-UK) Consortium |
| EPI_ISL_448326 | Quadram Institute Bioscience | COVID-19 Genomics UK (COG-UK) Consortium |
| EPI_ISL_448327 | Quadram Institute Bioscience | COVID-19 Genomics UK (COG-UK) Consortium |
| EPI_ISL_448328 | Quadram Institute Bioscience | COVID-19 Genomics UK (COG-UK) Consortium |
| EPI_ISL_448329 | Quadram Institute Bioscience | COVID-19 Genomics UK (COG-UK) Consortium |
| EPI_ISL_448330 | Quadram Institute Bioscience | COVID-19 Genomics UK (COG-UK) Consortium |
| EPI_ISL_448331 | Quadram Institute Bioscience | COVID-19 Genomics UK (COG-UK) Consortium |
| EPI_ISL_448332 | Quadram Institute Bioscience | COVID-19 Genomics UK (COG-UK) Consortium |
| EPI_ISL_448333 | Quadram Institute Bioscience | COVID-19 Genomics UK (COG-UK) Consortium |
| EPI_ISL_448334 | Quadram Institute Bioscience | COVID-19 Genomics UK (COG-UK) Consortium |
| EPI_ISL_448335 | Quadram Institute Bioscience | COVID-19 Genomics UK (COG-UK) Consortium |
| EPI_ISL_448336 | Quadram Institute Bioscience | COVID-19 Genomics UK (COG-UK) Consortium |
| EPI_ISL_448337 | Quadram Institute Bioscience | COVID-19 Genomics UK (COG-UK) Consortium |
| EPI_ISL_448338 | Quadram Institute Bioscience | COVID-19 Genomics UK (COG-UK) Consortium |
| EPI_ISL_448339 | Quadram Institute Bioscience | COVID-19 Genomics UK (COG-UK) Consortium |
| EPI_ISL_448340 | Quadram Institute Bioscience | COVID-19 Genomics UK (COG-UK) Consortium |
| EPI_ISL_448341 | Quadram Institute Bioscience | COVID-19 Genomics UK (COG-UK) Consortium |
| EPI_ISL_448342 | Quadram Institute Bioscience | COVID-19 Genomics UK (COG-UK) Consortium |
| EPI_ISL_448343 | Quadram Institute Bioscience | COVID-19 Genomics UK (COG-UK) Consortium |
| EPI_ISL_448344 | Quadram Institute Bioscience | COVID-19 Genomics UK (COG-UK) Consortium |
| EPI_ISL_448345 | Quadram Institute Bioscience | COVID-19 Genomics UK (COG-UK) Consortium |
| EPI_ISL_448346 | Quadram Institute Bioscience | COVID-19 Genomics UK (COG-UK) Consortium |
| EPI_ISL_448347 | Quadram Institute Bioscience | COVID-19 Genomics UK (COG-UK) Consortium |
| EPI_ISL_448348 | Quadram Institute Bioscience | COVID-19 Genomics UK (COG-UK) Consortium |
| EPI_ISL_448349 | Quadram Institute Bioscience | COVID-19 Genomics UK (COG-UK) Consortium |
| EPI_ISL_448350 | Quadram Institute Bioscience | COVID-19 Genomics UK (COG-UK) Consortium |
| EPI_ISL_448351 | Quadram Institute Bioscience | COVID-19 Genomics UK (COG-UK) Consortium |
| EPI_ISL_448352 | Quadram Institute Bioscience | COVID-19 Genomics UK (COG-UK) Consortium |
| EPI_ISL_448353 | Quadram Institute Bioscience | COVID-19 Genomics UK (COG-UK) Consortium |
| EPI_ISL_448354 | Quadram Institute Bioscience | COVID-19 Genomics UK (COG-UK) Consortium |
| EPI_ISL_448355 | Quadram Institute Bioscience | COVID-19 Genomics UK (COG-UK) Consortium |

| EPI_Accession_ID | Originating Laboratory | Submitting Laboratory |
|---|---|---|
| EPI_ISL_448356 | Quadram Institute Bioscience | COVID-19 Genomics UK (COG-UK) Consortium |
| EPI_ISL_448357 | Quadram Institute Bioscience | COVID-19 Genomics UK (COG-UK) Consortium |
| EPI_ISL_448358 | Quadram Institute Bioscience | COVID-19 Genomics UK (COG-UK) Consortium |
| EPI_ISL_448359 | Quadram Institute Bioscience | COVID-19 Genomics UK (COG-UK) Consortium |
| EPI_ISL_448360 | Quadram Institute Bioscience | COVID-19 Genomics UK (COG-UK) Consortium |
| EPI_ISL_448361 | Quadram Institute Bioscience | COVID-19 Genomics UK (COG-UK) Consortium |
| EPI_ISL_448362 | Quadram Institute Bioscience | COVID-19 Genomics UK (COG-UK) Consortium |
| EPI_ISL_448363 | Quadram Institute Bioscience | COVID-19 Genomics UK (COG-UK) Consortium |
| EPI_ISL_448364 | Quadram Institute Bioscience | COVID-19 Genomics UK (COG-UK) Consortium |
| EPI_ISL_448365 | Quadram Institute Bioscience | COVID-19 Genomics UK (COG-UK) Consortium |
| EPI_ISL_448366 | Quadram Institute Bioscience | COVID-19 Genomics UK (COG-UK) Consortium |
| EPI_ISL_448367 | Quadram Institute Bioscience | COVID-19 Genomics UK (COG-UK) Consortium |
| EPI_ISL_448368 | Quadram Institute Bioscience | COVID-19 Genomics UK (COG-UK) Consortium |
| EPI_ISL_448369 | Quadram Institute Bioscience | COVID-19 Genomics UK (COG-UK) Consortium |
| EPI_ISL_448370 | Quadram Institute Bioscience | COVID-19 Genomics UK (COG-UK) Consortium |
| EPI_ISL_448371 | Quadram Institute Bioscience | COVID-19 Genomics UK (COG-UK) Consortium |
| EPI_ISL_448372 | Quadram Institute Bioscience | COVID-19 Genomics UK (COG-UK) Consortium |
| EPI_ISL_448373 | Quadram Institute Bioscience | COVID-19 Genomics UK (COG-UK) Consortium |
| EPI_ISL_448374 | Quadram Institute Bioscience | COVID-19 Genomics UK (COG-UK) Consortium |
| EPI_ISL_448375 | Quadram Institute Bioscience | COVID-19 Genomics UK (COG-UK) Consortium |
| EPI_ISL_448376 | Quadram Institute Bioscience | COVID-19 Genomics UK (COG-UK) Consortium |
| EPI_ISL_448377 | Quadram Institute Bioscience | COVID-19 Genomics UK (COG-UK) Consortium |
| EPI_ISL_448378 | Quadram Institute Bioscience | COVID-19 Genomics UK (COG-UK) Consortium |
| EPI_ISL_448379 | Quadram Institute Bioscience | COVID-19 Genomics UK (COG-UK) Consortium |
| EPI_ISL_448380 | Quadram Institute Bioscience | COVID-19 Genomics UK (COG-UK) Consortium |
| EPI_ISL_448381 | Quadram Institute Bioscience | COVID-19 Genomics UK (COG-UK) Consortium |
| EPI_ISL_448382 | Quadram Institute Bioscience | COVID-19 Genomics UK (COG-UK) Consortium |
| EPI_ISL_448383 | Quadram Institute Bioscience | COVID-19 Genomics UK (COG-UK) Consortium |
| EPI_ISL_448384 | Quadram Institute Bioscience | COVID-19 Genomics UK (COG-UK) Consortium |
| EPI_ISL_448385 | Quadram Institute Bioscience | COVID-19 Genomics UK (COG-UK) Consortium |
| EPI_ISL_448386 | Quadram Institute Bioscience | COVID-19 Genomics UK (COG-UK) Consortium |
| EPI_ISL_448387 | Quadram Institute Bioscience | COVID-19 Genomics UK (COG-UK) Consortium |
| EPI_ISL_448388 | Quadram Institute Bioscience | COVID-19 Genomics UK (COG-UK) Consortium |

| EPI_Accession_ID | Originating Laboratory | Submitting Laboratory |
|---|---|---|
| EPI_ISL_448389 | Queens Medical Centre, Clinical Microbiology Department / DeepSeq Nottingham | COVID-19 Genomics UK (COG-UK) Consortium |
| EPI_ISL_448390 | Queens Medical Centre, Clinical Microbiology Department / DeepSeq Nottingham | COVID-19 Genomics UK (COG-UK) Consortium |
| EPI_ISL_448391 | Queens Medical Centre, Clinical Microbiology Department / DeepSeq Nottingham | COVID-19 Genomics UK (COG-UK) Consortium |
| EPI_ISL_448392 | Queens Medical Centre, Clinical Microbiology Department / DeepSeq Nottingham | COVID-19 Genomics UK (COG-UK) Consortium |
| EPI_ISL_448393 | Queens Medical Centre, Clinical Microbiology Department / DeepSeq Nottingham | COVID-19 Genomics UK (COG-UK) Consortium |
| EPI_ISL_448394 | Queens Medical Centre, Clinical Microbiology Department / DeepSeq Nottingham | COVID-19 Genomics UK (COG-UK) Consortium |
| EPI_ISL_448395 | Queens Medical Centre, Clinical Microbiology Department / DeepSeq Nottingham | COVID-19 Genomics UK (COG-UK) Consortium |
| EPI_ISL_448396 | Queens Medical Centre, Clinical Microbiology Department / DeepSeq Nottingham | COVID-19 Genomics UK (COG-UK) Consortium |
| EPI_ISL_448397 | Queens Medical Centre, Clinical Microbiology Department / DeepSeq Nottingham | COVID-19 Genomics UK (COG-UK) Consortium |
| EPI_ISL_448398 | Queens Medical Centre, Clinical Microbiology Department / DeepSeq Nottingham | COVID-19 Genomics UK (COG-UK) Consortium |
| EPI_ISL_448399 | Queens Medical Centre, Clinical Microbiology Department / DeepSeq Nottingham | COVID-19 Genomics UK (COG-UK) Consortium |
| EPI_ISL_448400 | Queens Medical Centre, Clinical Microbiology Department / DeepSeq Nottingham | COVID-19 Genomics UK (COG-UK) Consortium |
| EPI_ISL_448401 | Queens Medical Centre, Clinical Microbiology Department / DeepSeq Nottingham | COVID-19 Genomics UK (COG-UK) Consortium |
| EPI_ISL_448402 | Queens Medical Centre, Clinical Microbiology Department / DeepSeq Nottingham | COVID-19 Genomics UK (COG-UK) Consortium |
| EPI_ISL_448403 | Queens Medical Centre, Clinical Microbiology Department / DeepSeq Nottingham | COVID-19 Genomics UK (COG-UK) Consortium |
| EPI_ISL_448404 | Queens Medical Centre, Clinical Microbiology Department / DeepSeq Nottingham | COVID-19 Genomics UK (COG-UK) Consortium |

| EPI_Accession_ID | Originating Laboratory | Submitting Laboratory |
| --- | --- | --- |
| EPI_ISL_448405 | Queens Medical Centre, Clinical Microbiology Department / DeepSeq Nottingham | COVID-19 Genomics UK (COG-UK) Consortium |
| EPI_ISL_448406 | Queens Medical Centre, Clinical Microbiology Department / DeepSeq Nottingham | COVID-19 Genomics UK (COG-UK) Consortium |
| EPI_ISL_448407 | Queens Medical Centre, Clinical Microbiology Department / DeepSeq Nottingham | COVID-19 Genomics UK (COG-UK) Consortium |
| EPI_ISL_448408 | Queens Medical Centre, Clinical Microbiology Department / DeepSeq Nottingham | COVID-19 Genomics UK (COG-UK) Consortium |
| EPI_ISL_448409 | Queens Medical Centre, Clinical Microbiology Department / DeepSeq Nottingham | COVID-19 Genomics UK (COG-UK) Consortium |
| EPI_ISL_448410 | Queens Medical Centre, Clinical Microbiology Department / DeepSeq Nottingham | COVID-19 Genomics UK (COG-UK) Consortium |
| EPI_ISL_448411 | Queens Medical Centre, Clinical Microbiology Department / DeepSeq Nottingham | COVID-19 Genomics UK (COG-UK) Consortium |
| EPI_ISL_448412 | Queens Medical Centre, Clinical Microbiology Department / DeepSeq Nottingham | COVID-19 Genomics UK (COG-UK) Consortium |
| EPI_ISL_448413 | Queens Medical Centre, Clinical Microbiology Department / DeepSeq Nottingham | COVID-19 Genomics UK (COG-UK) Consortium |
| EPI_ISL_448414 | Queens Medical Centre, Clinical Microbiology Department / DeepSeq Nottingham | COVID-19 Genomics UK (COG-UK) Consortium |
| EPI_ISL_448415 | Queens Medical Centre, Clinical Microbiology Department / DeepSeq Nottingham | COVID-19 Genomics UK (COG-UK) Consortium |
| EPI_ISL_448416 | Queens Medical Centre, Clinical Microbiology Department / DeepSeq Nottingham | COVID-19 Genomics UK (COG-UK) Consortium |
| EPI_ISL_448417 | Queens Medical Centre, Clinical Microbiology Department / DeepSeq Nottingham | COVID-19 Genomics UK (COG-UK) Consortium |
| EPI_ISL_448418 | Queens Medical Centre, Clinical Microbiology Department / DeepSeq Nottingham | COVID-19 Genomics UK (COG-UK) Consortium |
| EPI_ISL_448419 | Queens Medical Centre, Clinical Microbiology Department / DeepSeq Nottingham | COVID-19 Genomics UK (COG-UK) Consortium |
| EPI_ISL_448420 | Queens Medical Centre, Clinical Microbiology Department / DeepSeq Nottingham | COVID-19 Genomics UK (COG-UK) Consortium |

| EPI_Accession_ID | Originating Laboratory | Submitting Laboratory |
|---|---|---|
| EPI_ISL_448421 | Queens Medical Centre, Clinical Microbiology Department / DeepSeq Nottingham | COVID-19 Genomics UK (COG-UK) Consortium |
| EPI_ISL_448422 | Queens Medical Centre, Clinical Microbiology Department / DeepSeq Nottingham | COVID-19 Genomics UK (COG-UK) Consortium |
| EPI_ISL_448423 | Queens Medical Centre, Clinical Microbiology Department / DeepSeq Nottingham | COVID-19 Genomics UK (COG-UK) Consortium |
| EPI_ISL_448424 | Queens Medical Centre, Clinical Microbiology Department / DeepSeq Nottingham | COVID-19 Genomics UK (COG-UK) Consortium |
| EPI_ISL_448425 | Queens Medical Centre, Clinical Microbiology Department / DeepSeq Nottingham | COVID-19 Genomics UK (COG-UK) Consortium |
| EPI_ISL_448426 | Queens Medical Centre, Clinical Microbiology Department / DeepSeq Nottingham | COVID-19 Genomics UK (COG-UK) Consortium |
| EPI_ISL_448427 | Queens Medical Centre, Clinical Microbiology Department / DeepSeq Nottingham | COVID-19 Genomics UK (COG-UK) Consortium |
| EPI_ISL_448428 | Queens Medical Centre, Clinical Microbiology Department / DeepSeq Nottingham | COVID-19 Genomics UK (COG-UK) Consortium |
| EPI_ISL_448429 | Queens Medical Centre, Clinical Microbiology Department / DeepSeq Nottingham | COVID-19 Genomics UK (COG-UK) Consortium |
| EPI_ISL_448430 | Queens Medical Centre, Clinical Microbiology Department / DeepSeq Nottingham | COVID-19 Genomics UK (COG-UK) Consortium |
| EPI_ISL_448431 | Queens Medical Centre, Clinical Microbiology Department / DeepSeq Nottingham | COVID-19 Genomics UK (COG-UK) Consortium |
| EPI_ISL_448432 | Queens Medical Centre, Clinical Microbiology Department / DeepSeq Nottingham | COVID-19 Genomics UK (COG-UK) Consortium |
| EPI_ISL_448433 | Queens Medical Centre, Clinical Microbiology Department / DeepSeq Nottingham | COVID-19 Genomics UK (COG-UK) Consortium |
| EPI_ISL_448434 | Queens Medical Centre, Clinical Microbiology Department / DeepSeq Nottingham | COVID-19 Genomics UK (COG-UK) Consortium |
| EPI_ISL_448435 | Queens Medical Centre, Clinical Microbiology Department / DeepSeq Nottingham | COVID-19 Genomics UK (COG-UK) Consortium |
| EPI_ISL_448436 | Queens Medical Centre, Clinical Microbiology Department / DeepSeq Nottingham | COVID-19 Genomics UK (COG-UK) Consortium |

| EPI_Accession_ID | Originating Laboratory | Submitting Laboratory |
|---|---|---|
| EPI_ISL_448437 | Queens Medical Centre, Clinical Microbiology Department / DeepSeq Nottingham | COVID-19 Genomics UK (COG-UK) Consortium |
| EPI_ISL_448438 | Queens Medical Centre, Clinical Microbiology Department / DeepSeq Nottingham | COVID-19 Genomics UK (COG-UK) Consortium |
| EPI_ISL_448439 | Queens Medical Centre, Clinical Microbiology Department / DeepSeq Nottingham | COVID-19 Genomics UK (COG-UK) Consortium |
| EPI_ISL_448440 | Queens Medical Centre, Clinical Microbiology Department / DeepSeq Nottingham | COVID-19 Genomics UK (COG-UK) Consortium |
| EPI_ISL_448441 | Queens Medical Centre, Clinical Microbiology Department / DeepSeq Nottingham | COVID-19 Genomics UK (COG-UK) Consortium |
| EPI_ISL_448442 | Queens Medical Centre, Clinical Microbiology Department / DeepSeq Nottingham | COVID-19 Genomics UK (COG-UK) Consortium |
| EPI_ISL_448443 | Queens Medical Centre, Clinical Microbiology Department / DeepSeq Nottingham | COVID-19 Genomics UK (COG-UK) Consortium |
| EPI_ISL_448444 | Queens Medical Centre, Clinical Microbiology Department / DeepSeq Nottingham | COVID-19 Genomics UK (COG-UK) Consortium |
| EPI_ISL_448445 | Queens Medical Centre, Clinical Microbiology Department / DeepSeq Nottingham | COVID-19 Genomics UK (COG-UK) Consortium |
| EPI_ISL_448446 | Queens Medical Centre, Clinical Microbiology Department / DeepSeq Nottingham | COVID-19 Genomics UK (COG-UK) Consortium |
| EPI_ISL_448447 | Queens Medical Centre, Clinical Microbiology Department / DeepSeq Nottingham | COVID-19 Genomics UK (COG-UK) Consortium |
| EPI_ISL_448448 | Queens Medical Centre, Clinical Microbiology Department / DeepSeq Nottingham | COVID-19 Genomics UK (COG-UK) Consortium |
| EPI_ISL_448449 | Oxford Viromics, NDM, University of Oxford; Oxford University Hospitals; Basingstoke and North Hampshire Hospital | COVID-19 Genomics UK (COG-UK) Consortium |
| EPI_ISL_448450 | Oxford Viromics, NDM, University of Oxford; Oxford University Hospitals; Basingstoke and North Hampshire Hospital | COVID-19 Genomics UK (COG-UK) Consortium |
| EPI_ISL_448451 | Oxford Viromics, NDM, University of Oxford; Oxford University Hospitals; Basingstoke and North Hampshire Hospital | COVID-19 Genomics UK (COG-UK) Consortium |

| EPI_Accession_ID | Originating Laboratory | Submitting Laboratory |
|---|---|---|
| EPI_ISL_448452 | Oxford Viromics, NDM, University of Oxford; Oxford University Hospitals; Basingstoke and North Hampshire Hospital | COVID-19 Genomics UK (COG-UK) Consortium |
| EPI_ISL_448453 | Oxford Viromics, NDM, University of Oxford; Oxford University Hospitals; Basingstoke and North Hampshire Hospital | COVID-19 Genomics UK (COG-UK) Consortium |
| EPI_ISL_448454 | Oxford Viromics, NDM, University of Oxford; Oxford University Hospitals; Basingstoke and North Hampshire Hospital | COVID-19 Genomics UK (COG-UK) Consortium |
| EPI_ISL_448455 | Oxford Viromics, NDM, University of Oxford; Oxford University Hospitals; Basingstoke and North Hampshire Hospital | COVID-19 Genomics UK (COG-UK) Consortium |
| EPI_ISL_448456 | Oxford Viromics, NDM, University of Oxford; Oxford University Hospitals; Basingstoke and North Hampshire Hospital | COVID-19 Genomics UK (COG-UK) Consortium |
| EPI_ISL_448457 | Oxford Viromics, NDM, University of Oxford; Oxford University Hospitals; Basingstoke and North Hampshire Hospital | COVID-19 Genomics UK (COG-UK) Consortium |
| EPI_ISL_448458 | Oxford Viromics, NDM, University of Oxford; Oxford University Hospitals; Basingstoke and North Hampshire Hospital | COVID-19 Genomics UK (COG-UK) Consortium |
| EPI_ISL_448459 | Oxford Viromics, NDM, University of Oxford; Oxford University Hospitals; Basingstoke and North Hampshire Hospital | COVID-19 Genomics UK (COG-UK) Consortium |
| EPI_ISL_448460 | Oxford Viromics, NDM, University of Oxford; Oxford University Hospitals; Basingstoke and North Hampshire Hospital | COVID-19 Genomics UK (COG-UK) Consortium |
| EPI_ISL_448461 | Oxford Viromics, NDM, University of Oxford; Oxford University Hospitals; Basingstoke and North Hampshire Hospital | COVID-19 Genomics UK (COG-UK) Consortium |
| EPI_ISL_448462 | Oxford Viromics, NDM, University of Oxford; Oxford University Hospitals; Basingstoke and North Hampshire Hospital | COVID-19 Genomics UK (COG-UK) Consortium |

| EPI_Accession_ID | Originating Laboratory | Submitting Laboratory |
|---|---|---|
| EPI_ISL_448463 | Oxford Viromics, NDM, University of Oxford; Oxford University Hospitals; Basingstoke and North Hampshire Hospital | COVID-19 Genomics UK (COG-UK) Consortium |
| EPI_ISL_448464 | Oxford Viromics, NDM, University of Oxford; Oxford University Hospitals; Basingstoke and North Hampshire Hospital | COVID-19 Genomics UK (COG-UK) Consortium |
| EPI_ISL_448465 | Oxford Viromics, NDM, University of Oxford; Oxford University Hospitals; Basingstoke and North Hampshire Hospital | COVID-19 Genomics UK (COG-UK) Consortium |
| EPI_ISL_448466 | Oxford Viromics, NDM, University of Oxford; Oxford University Hospitals; Basingstoke and North Hampshire Hospital | COVID-19 Genomics UK (COG-UK) Consortium |
| EPI_ISL_448467 | Oxford Viromics, NDM, University of Oxford; Oxford University Hospitals; Basingstoke and North Hampshire Hospital | COVID-19 Genomics UK (COG-UK) Consortium |
| EPI_ISL_448468 | Oxford Viromics, NDM, University of Oxford; Oxford University Hospitals; Basingstoke and North Hampshire Hospital | COVID-19 Genomics UK (COG-UK) Consortium |
| EPI_ISL_448469 | Oxford Viromics, NDM, University of Oxford; Oxford University Hospitals; Basingstoke and North Hampshire Hospital | COVID-19 Genomics UK (COG-UK) Consortium |
| EPI_ISL_448470 | Oxford Viromics, NDM, University of Oxford; Oxford University Hospitals; Basingstoke and North Hampshire Hospital | COVID-19 Genomics UK (COG-UK) Consortium |
| EPI_ISL_448471 | Oxford Viromics, NDM, University of Oxford; Oxford University Hospitals; Basingstoke and North Hampshire Hospital | COVID-19 Genomics UK (COG-UK) Consortium |
| EPI_ISL_448472 | Oxford Viromics, NDM, University of Oxford; Oxford University Hospitals; Basingstoke and North Hampshire Hospital | COVID-19 Genomics UK (COG-UK) Consortium |
| EPI_ISL_448473 | Oxford Viromics, NDM, University of Oxford; Oxford University Hospitals; Basingstoke and North Hampshire Hospital | COVID-19 Genomics UK (COG-UK) Consortium |

| EPI_Accession_ID | Originating Laboratory | Submitting Laboratory |
|---|---|---|
| EPI_ISL_448474 | Oxford Viromics, NDM, University of Oxford; Oxford University Hospitals; Basingstoke and North Hampshire Hospital | COVID-19 Genomics UK (COG-UK) Consortium |
| EPI_ISL_448475 | Oxford Viromics, NDM, University of Oxford; Oxford University Hospitals; Basingstoke and North Hampshire Hospital | COVID-19 Genomics UK (COG-UK) Consortium |
| EPI_ISL_448476 | Oxford Viromics, NDM, University of Oxford; Oxford University Hospitals; Basingstoke and North Hampshire Hospital | COVID-19 Genomics UK (COG-UK) Consortium |
| EPI_ISL_448477 | Oxford Viromics, NDM, University of Oxford; Oxford University Hospitals; Basingstoke and North Hampshire Hospital | COVID-19 Genomics UK (COG-UK) Consortium |
| EPI_ISL_448478 | Oxford Viromics, NDM, University of Oxford; Oxford University Hospitals; Basingstoke and North Hampshire Hospital | COVID-19 Genomics UK (COG-UK) Consortium |
| EPI_ISL_448479 | Oxford Viromics, NDM, University of Oxford; Oxford University Hospitals; Basingstoke and North Hampshire Hospital | COVID-19 Genomics UK (COG-UK) Consortium |
| EPI_ISL_448480 | Oxford Viromics, NDM, University of Oxford; Oxford University Hospitals; Basingstoke and North Hampshire Hospital | COVID-19 Genomics UK (COG-UK) Consortium |
| EPI_ISL_448481 | Oxford Viromics, NDM, University of Oxford; Oxford University Hospitals; Basingstoke and North Hampshire Hospital | COVID-19 Genomics UK (COG-UK) Consortium |
| EPI_ISL_448482 | Oxford Viromics, NDM, University of Oxford; Oxford University Hospitals; Basingstoke and North Hampshire Hospital | COVID-19 Genomics UK (COG-UK) Consortium |
| EPI_ISL_448483 | Oxford Viromics, NDM, University of Oxford; Oxford University Hospitals; Basingstoke and North Hampshire Hospital | COVID-19 Genomics UK (COG-UK) Consortium |
| EPI_ISL_448484 | Oxford Viromics, NDM, University of Oxford; Oxford University Hospitals; Basingstoke and North Hampshire Hospital | COVID-19 Genomics UK (COG-UK) Consortium |

| EPI_Accession_ID | Originating Laboratory | Submitting Laboratory |
| --- | --- | --- |
| EPI_ISL_448485 | Oxford Viromics, NDM, University of Oxford; Oxford University Hospitals; Basingstoke and North Hampshire Hospital | COVID-19 Genomics UK (COG-UK) Consortium |
| EPI_ISL_448486 | Oxford Viromics, NDM, University of Oxford; Oxford University Hospitals; Basingstoke and North Hampshire Hospital | COVID-19 Genomics UK (COG-UK) Consortium |
| EPI_ISL_448487 | Oxford Viromics, NDM, University of Oxford; Oxford University Hospitals; Basingstoke and North Hampshire Hospital | COVID-19 Genomics UK (COG-UK) Consortium |
| EPI_ISL_448488 | Oxford Viromics, NDM, University of Oxford; Oxford University Hospitals; Basingstoke and North Hampshire Hospital | COVID-19 Genomics UK (COG-UK) Consortium |
| EPI_ISL_448489 | Oxford Viromics, NDM, University of Oxford; Oxford University Hospitals; Basingstoke and North Hampshire Hospital | COVID-19 Genomics UK (COG-UK) Consortium |
| EPI_ISL_448490 | Oxford Viromics, NDM, University of Oxford; Oxford University Hospitals; Basingstoke and North Hampshire Hospital | COVID-19 Genomics UK (COG-UK) Consortium |
| EPI_ISL_448491 | Oxford Viromics, NDM, University of Oxford; Oxford University Hospitals; Basingstoke and North Hampshire Hospital | COVID-19 Genomics UK (COG-UK) Consortium |
| EPI_ISL_448492 | Oxford Viromics, NDM, University of Oxford; Oxford University Hospitals; Basingstoke and North Hampshire Hospital | COVID-19 Genomics UK (COG-UK) Consortium |
| EPI_ISL_448493 | Oxford Viromics, NDM, University of Oxford; Oxford University Hospitals; Basingstoke and North Hampshire Hospital | COVID-19 Genomics UK (COG-UK) Consortium |
| EPI_ISL_448494 | Oxford Viromics, NDM, University of Oxford; Oxford University Hospitals; Basingstoke and North Hampshire Hospital | COVID-19 Genomics UK (COG-UK) Consortium |
| EPI_ISL_448495 | Oxford Viromics, NDM, University of Oxford; Oxford University Hospitals; Basingstoke and North Hampshire Hospital | COVID-19 Genomics UK (COG-UK) Consortium |

| EPI_Accession_ID | Originating Laboratory | Submitting Laboratory |
|---|---|---|
| EPI_ISL_448496 | Oxford Viromics, NDM, University of Oxford; Oxford University Hospitals; Basingstoke and North Hampshire Hospital | COVID-19 Genomics UK (COG-UK) Consortium |
| EPI_ISL_448497 | Oxford Viromics, NDM, University of Oxford; Oxford University Hospitals; Basingstoke and North Hampshire Hospital | COVID-19 Genomics UK (COG-UK) Consortium |
| EPI_ISL_448498 | Oxford Viromics, NDM, University of Oxford; Oxford University Hospitals; Basingstoke and North Hampshire Hospital | COVID-19 Genomics UK (COG-UK) Consortium |
| EPI_ISL_448499 | Oxford Viromics, NDM, University of Oxford; Oxford University Hospitals; Basingstoke and North Hampshire Hospital | COVID-19 Genomics UK (COG-UK) Consortium |
| EPI_ISL_448500 | Oxford Viromics, NDM, University of Oxford; Oxford University Hospitals; Basingstoke and North Hampshire Hospital | COVID-19 Genomics UK (COG-UK) Consortium |
| EPI_ISL_448501 | Oxford Viromics, NDM, University of Oxford; Oxford University Hospitals; Basingstoke and North Hampshire Hospital | COVID-19 Genomics UK (COG-UK) Consortium |
| EPI_ISL_448502 | Oxford Viromics, NDM, University of Oxford; Oxford University Hospitals; Basingstoke and North Hampshire Hospital | COVID-19 Genomics UK (COG-UK) Consortium |
| EPI_ISL_448503 | Oxford Viromics, NDM, University of Oxford; Oxford University Hospitals; Basingstoke and North Hampshire Hospital | COVID-19 Genomics UK (COG-UK) Consortium |
| EPI_ISL_448504 | Oxford Viromics, NDM, University of Oxford; Oxford University Hospitals; Basingstoke and North Hampshire Hospital | COVID-19 Genomics UK (COG-UK) Consortium |
| EPI_ISL_448505 | Oxford Viromics, NDM, University of Oxford; Oxford University Hospitals; Basingstoke and North Hampshire Hospital | COVID-19 Genomics UK (COG-UK) Consortium |
| EPI_ISL_448506 | Oxford Viromics, NDM, University of Oxford; Oxford University Hospitals; Basingstoke and North Hampshire Hospital | COVID-19 Genomics UK (COG-UK) Consortium |

| EPI_Accession_ID | Originating Laboratory | Submitting Laboratory |
|---|---|---|
| EPI_ISL_448507 | Oxford Viromics, NDM, University of Oxford; Oxford University Hospitals; Basingstoke and North Hampshire Hospital | COVID-19 Genomics UK (COG-UK) Consortium |
| EPI_ISL_448508 | Oxford Viromics, NDM, University of Oxford; Oxford University Hospitals; Basingstoke and North Hampshire Hospital | COVID-19 Genomics UK (COG-UK) Consortium |
| EPI_ISL_448509 | Oxford Viromics, NDM, University of Oxford; Oxford University Hospitals; Basingstoke and North Hampshire Hospital | COVID-19 Genomics UK (COG-UK) Consortium |
| EPI_ISL_448510 | Oxford Viromics, NDM, University of Oxford; Oxford University Hospitals; Basingstoke and North Hampshire Hospital | COVID-19 Genomics UK (COG-UK) Consortium |
| EPI_ISL_448511 | Oxford Viromics, NDM, University of Oxford; Oxford University Hospitals; Basingstoke and North Hampshire Hospital | COVID-19 Genomics UK (COG-UK) Consortium |
| EPI_ISL_448512 | Oxford Viromics, NDM, University of Oxford; Oxford University Hospitals; Basingstoke and North Hampshire Hospital | COVID-19 Genomics UK (COG-UK) Consortium |
| EPI_ISL_448513 | Oxford Viromics, NDM, University of Oxford; Oxford University Hospitals; Basingstoke and North Hampshire Hospital | COVID-19 Genomics UK (COG-UK) Consortium |
| EPI_ISL_448514 | Oxford Viromics, NDM, University of Oxford; Oxford University Hospitals; Basingstoke and North Hampshire Hospital | COVID-19 Genomics UK (COG-UK) Consortium |
| EPI_ISL_448515 | Oxford Viromics, NDM, University of Oxford; Oxford University Hospitals; Basingstoke and North Hampshire Hospital | COVID-19 Genomics UK (COG-UK) Consortium |
| EPI_ISL_448516 | Oxford Viromics, NDM, University of Oxford; Oxford University Hospitals; Basingstoke and North Hampshire Hospital | COVID-19 Genomics UK (COG-UK) Consortium |
| EPI_ISL_448517 | Oxford Viromics, NDM, University of Oxford; Oxford University Hospitals; Basingstoke and North Hampshire Hospital | COVID-19 Genomics UK (COG-UK) Consortium |

| EPI_Accession_ID | Originating Laboratory | Submitting Laboratory |
| --- | --- | --- |
| EPI_ISL_448518 | Oxford Viromics, NDM, University of Oxford; Oxford University Hospitals; Basingstoke and North Hampshire Hospital | COVID-19 Genomics UK (COG-UK) Consortium |
| EPI_ISL_448519 | Oxford Viromics, NDM, University of Oxford; Oxford University Hospitals; Basingstoke and North Hampshire Hospital | COVID-19 Genomics UK (COG-UK) Consortium |
| EPI_ISL_448520 | Oxford Viromics, NDM, University of Oxford; Oxford University Hospitals; Basingstoke and North Hampshire Hospital | COVID-19 Genomics UK (COG-UK) Consortium |
| EPI_ISL_448521 | Oxford Viromics, NDM, University of Oxford; Oxford University Hospitals; Basingstoke and North Hampshire Hospital | COVID-19 Genomics UK (COG-UK) Consortium |
| EPI_ISL_448522 | Oxford Viromics, NDM, University of Oxford; Oxford University Hospitals; Basingstoke and North Hampshire Hospital | COVID-19 Genomics UK (COG-UK) Consortium |
| EPI_ISL_448523 | Oxford Viromics, NDM, University of Oxford; Oxford University Hospitals; Basingstoke and North Hampshire Hospital | COVID-19 Genomics UK (COG-UK) Consortium |
| EPI_ISL_448524 | Oxford Viromics, NDM, University of Oxford; Oxford University Hospitals; Basingstoke and North Hampshire Hospital | COVID-19 Genomics UK (COG-UK) Consortium |
| EPI_ISL_448525 | Oxford Viromics, NDM, University of Oxford; Oxford University Hospitals; Basingstoke and North Hampshire Hospital | COVID-19 Genomics UK (COG-UK) Consortium |
| EPI_ISL_448526 | Oxford Viromics, NDM, University of Oxford; Oxford University Hospitals; Basingstoke and North Hampshire Hospital | COVID-19 Genomics UK (COG-UK) Consortium |
| EPI_ISL_448527 | Oxford Viromics, NDM, University of Oxford; Oxford University Hospitals; Basingstoke and North Hampshire Hospital | COVID-19 Genomics UK (COG-UK) Consortium |
| EPI_ISL_448528 | Oxford Viromics, NDM, University of Oxford; Oxford University Hospitals; Basingstoke and North Hampshire Hospital | COVID-19 Genomics UK (COG-UK) Consortium |

| EPI_Accession_ID | Originating Laboratory | Submitting Laboratory |
|---|---|---|
| EPI_ISL_448529 | Oxford Viromics, NDM, University of Oxford; Oxford University Hospitals; Basingstoke and North Hampshire Hospital | COVID-19 Genomics UK (COG-UK) Consortium |
| EPI_ISL_448530 | Oxford Viromics, NDM, University of Oxford; Oxford University Hospitals; Basingstoke and North Hampshire Hospital | COVID-19 Genomics UK (COG-UK) Consortium |
| EPI_ISL_448531 | Oxford Viromics, NDM, University of Oxford; Oxford University Hospitals; Basingstoke and North Hampshire Hospital | COVID-19 Genomics UK (COG-UK) Consortium |
| EPI_ISL_448532 | Oxford Viromics, NDM, University of Oxford; Oxford University Hospitals; Basingstoke and North Hampshire Hospital | COVID-19 Genomics UK (COG-UK) Consortium |
| EPI_ISL_448533 | Oxford Viromics, NDM, University of Oxford; Oxford University Hospitals; Basingstoke and North Hampshire Hospital | COVID-19 Genomics UK (COG-UK) Consortium |
| EPI_ISL_448534 | Oxford Viromics, NDM, University of Oxford; Oxford University Hospitals; Basingstoke and North Hampshire Hospital | COVID-19 Genomics UK (COG-UK) Consortium |
| EPI_ISL_448535 | Oxford Viromics, NDM, University of Oxford; Oxford University Hospitals; Basingstoke and North Hampshire Hospital | COVID-19 Genomics UK (COG-UK) Consortium |
| EPI_ISL_448536 | Oxford Viromics, NDM, University of Oxford; Oxford University Hospitals; Basingstoke and North Hampshire Hospital | COVID-19 Genomics UK (COG-UK) Consortium |
| EPI_ISL_448537 | Oxford Viromics, NDM, University of Oxford; Oxford University Hospitals; Basingstoke and North Hampshire Hospital | COVID-19 Genomics UK (COG-UK) Consortium |
| EPI_ISL_448538 | Oxford Viromics, NDM, University of Oxford; Oxford University Hospitals; Basingstoke and North Hampshire Hospital | COVID-19 Genomics UK (COG-UK) Consortium |
| EPI_ISL_448539 | Oxford Viromics, NDM, University of Oxford; Oxford University Hospitals; Basingstoke and North Hampshire Hospital | COVID-19 Genomics UK (COG-UK) Consortium |

| EPI_Accession_ID | Originating Laboratory | Submitting Laboratory |
|---|---|---|
| EPI_ISL_448540 | Oxford Viromics, NDM, University of Oxford; Oxford University Hospitals; Basingstoke and North Hampshire Hospital | COVID-19 Genomics UK (COG-UK) Consortium |
| EPI_ISL_448541 | Oxford Viromics, NDM, University of Oxford; Oxford University Hospitals; Basingstoke and North Hampshire Hospital | COVID-19 Genomics UK (COG-UK) Consortium |
| EPI_ISL_448542 | Oxford Viromics, NDM, University of Oxford; Oxford University Hospitals; Basingstoke and North Hampshire Hospital | COVID-19 Genomics UK (COG-UK) Consortium |
| EPI_ISL_448543 | Oxford Viromics, NDM, University of Oxford; Oxford University Hospitals; Basingstoke and North Hampshire Hospital | COVID-19 Genomics UK (COG-UK) Consortium |
| EPI_ISL_448544 | Oxford Viromics, NDM, University of Oxford; Oxford University Hospitals; Basingstoke and North Hampshire Hospital | COVID-19 Genomics UK (COG-UK) Consortium |
| EPI_ISL_448545 | Oxford Viromics, NDM, University of Oxford; Oxford University Hospitals; Basingstoke and North Hampshire Hospital | COVID-19 Genomics UK (COG-UK) Consortium |
| EPI_ISL_448546 | Oxford Viromics, NDM, University of Oxford; Oxford University Hospitals; Basingstoke and North Hampshire Hospital | COVID-19 Genomics UK (COG-UK) Consortium |
| EPI_ISL_448547 | Oxford Viromics, NDM, University of Oxford; Oxford University Hospitals; Basingstoke and North Hampshire Hospital | COVID-19 Genomics UK (COG-UK) Consortium |
| EPI_ISL_448549 | Oxford Viromics, NDM, University of Oxford; Oxford University Hospitals; Basingstoke and North Hampshire Hospital | COVID-19 Genomics UK (COG-UK) Consortium |
| EPI_ISL_448551 | Oxford Viromics, NDM, University of Oxford; Oxford University Hospitals; Basingstoke and North Hampshire Hospital | COVID-19 Genomics UK (COG-UK) Consortium |
| EPI_ISL_448553 | Oxford Viromics, NDM, University of Oxford; Oxford University Hospitals; Basingstoke and North Hampshire Hospital | COVID-19 Genomics UK (COG-UK) Consortium |

| EPI_Accession_ID | Originating Laboratory | Submitting Laboratory |
|---|---|---|
| EPI_ISL_448555 | Oxford Viromics, NDM, University of Oxford; Oxford University Hospitals; Basingstoke and North Hampshire Hospital | COVID-19 Genomics UK (COG-UK) Consortium |
| EPI_ISL_448557 | Oxford Viromics, NDM, University of Oxford; Oxford University Hospitals; Basingstoke and North Hampshire Hospital | COVID-19 Genomics UK (COG-UK) Consortium |
| EPI_ISL_448559 | Oxford Viromics, NDM, University of Oxford; Oxford University Hospitals; Basingstoke and North Hampshire Hospital | COVID-19 Genomics UK (COG-UK) Consortium |
| EPI_ISL_448561 | Oxford Viromics, NDM, University of Oxford; Oxford University Hospitals; Basingstoke and North Hampshire Hospital | COVID-19 Genomics UK (COG-UK) Consortium |
| EPI_ISL_448564 | Oxford Viromics, NDM, University of Oxford; Oxford University Hospitals; Basingstoke and North Hampshire Hospital | COVID-19 Genomics UK (COG-UK) Consortium |
| EPI_ISL_448565 | Oxford Viromics, NDM, University of Oxford; Oxford University Hospitals; Basingstoke and North Hampshire Hospital | COVID-19 Genomics UK (COG-UK) Consortium |
| EPI_ISL_448774 | Oxford Viromics, NDM, University of Oxford; Oxford University Hospitals; Basingstoke and North Hampshire Hospital | COVID-19 Genomics UK (COG-UK) Consortium |
| EPI_ISL_448775 | Oxford Viromics, NDM, University of Oxford; Oxford University Hospitals; Basingstoke and North Hampshire Hospital | COVID-19 Genomics UK (COG-UK) Consortium |
| EPI_ISL_448776 | Oxford Viromics, NDM, University of Oxford; Oxford University Hospitals; Basingstoke and North Hampshire Hospital | COVID-19 Genomics UK (COG-UK) Consortium |
| EPI_ISL_448777 | Oxford Viromics, NDM, University of Oxford; Oxford University Hospitals; Basingstoke and North Hampshire Hospital | COVID-19 Genomics UK (COG-UK) Consortium |
| EPI_ISL_448778 | Oxford Viromics, NDM, University of Oxford; Oxford University Hospitals; Basingstoke and North Hampshire Hospital | COVID-19 Genomics UK (COG-UK) Consortium |

| EPI_Accession_ID | Originating Laboratory | Submitting Laboratory |
|---|---|---|
| EPI_ISL_448779 | Oxford Viromics, NDM, University of Oxford; Oxford University Hospitals; Basingstoke and North Hampshire Hospital | COVID-19 Genomics UK (COG-UK) Consortium |
| EPI_ISL_448780 | Oxford Viromics, NDM, University of Oxford; Oxford University Hospitals; Basingstoke and North Hampshire Hospital | COVID-19 Genomics UK (COG-UK) Consortium |
| EPI_ISL_448781 | Oxford Viromics, NDM, University of Oxford; Oxford University Hospitals; Basingstoke and North Hampshire Hospital | COVID-19 Genomics UK (COG-UK) Consortium |
| EPI_ISL_448782 | Oxford Viromics, NDM, University of Oxford; Oxford University Hospitals; Basingstoke and North Hampshire Hospital | COVID-19 Genomics UK (COG-UK) Consortium |
| EPI_ISL_448783 | Oxford Viromics, NDM, University of Oxford; Oxford University Hospitals; Basingstoke and North Hampshire Hospital | COVID-19 Genomics UK (COG-UK) Consortium |
| EPI_ISL_448784 | Oxford Viromics, NDM, University of Oxford; Oxford University Hospitals; Basingstoke and North Hampshire Hospital | COVID-19 Genomics UK (COG-UK) Consortium |
| EPI_ISL_448785 | Oxford Viromics, NDM, University of Oxford; Oxford University Hospitals; Basingstoke and North Hampshire Hospital | COVID-19 Genomics UK (COG-UK) Consortium |
| EPI_ISL_448786 | Oxford Viromics, NDM, University of Oxford; Oxford University Hospitals; Basingstoke and North Hampshire Hospital | COVID-19 Genomics UK (COG-UK) Consortium |
| EPI_ISL_448787 | Oxford Viromics, NDM, University of Oxford; Oxford University Hospitals; Basingstoke and North Hampshire Hospital | COVID-19 Genomics UK (COG-UK) Consortium |
| EPI_ISL_448788 | Oxford Viromics, NDM, University of Oxford; Oxford University Hospitals; Basingstoke and North Hampshire Hospital | COVID-19 Genomics UK (COG-UK) Consortium |
| EPI_ISL_448789 | Oxford Viromics, NDM, University of Oxford; Oxford University Hospitals; Basingstoke and North Hampshire Hospital | COVID-19 Genomics UK (COG-UK) Consortium |

| EPI_Accession_ID | Originating Laboratory | Submitting Laboratory |
|---|---|---|
| EPI_ISL_448790 | Oxford Viromics, NDM, University of Oxford; Oxford University Hospitals; Basingstoke and North Hampshire Hospital | COVID-19 Genomics UK (COG-UK) Consortium |
| EPI_ISL_448791 | Oxford Viromics, NDM, University of Oxford; Oxford University Hospitals; Basingstoke and North Hampshire Hospital | COVID-19 Genomics UK (COG-UK) Consortium |
| EPI_ISL_448792 | Oxford Viromics, NDM, University of Oxford; Oxford University Hospitals; Basingstoke and North Hampshire Hospital | COVID-19 Genomics UK (COG-UK) Consortium |
| EPI_ISL_448793 | Oxford Viromics, NDM, University of Oxford; Oxford University Hospitals; Basingstoke and North Hampshire Hospital | COVID-19 Genomics UK (COG-UK) Consortium |
| EPI_ISL_448794 | Oxford Viromics, NDM, University of Oxford; Oxford University Hospitals; Basingstoke and North Hampshire Hospital | COVID-19 Genomics UK (COG-UK) Consortium |
| EPI_ISL_448795 | Oxford Viromics, NDM, University of Oxford; Oxford University Hospitals; Basingstoke and North Hampshire Hospital | COVID-19 Genomics UK (COG-UK) Consortium |
| EPI_ISL_448796 | Oxford Viromics, NDM, University of Oxford; Oxford University Hospitals; Basingstoke and North Hampshire Hospital | COVID-19 Genomics UK (COG-UK) Consortium |
| EPI_ISL_448797 | Oxford Viromics, NDM, University of Oxford; Oxford University Hospitals; Basingstoke and North Hampshire Hospital | COVID-19 Genomics UK (COG-UK) Consortium |
| EPI_ISL_448798 | Oxford Viromics, NDM, University of Oxford; Oxford University Hospitals; Basingstoke and North Hampshire Hospital | COVID-19 Genomics UK (COG-UK) Consortium |
| EPI_ISL_448799 | Oxford Viromics, NDM, University of Oxford; Oxford University Hospitals; Basingstoke and North Hampshire Hospital | COVID-19 Genomics UK (COG-UK) Consortium |
| EPI_ISL_448800 | Oxford Viromics, NDM, University of Oxford; Oxford University Hospitals; Basingstoke and North Hampshire Hospital | COVID-19 Genomics UK (COG-UK) Consortium |

| EPI_Accession_ID | Originating Laboratory | Submitting Laboratory |
|---|---|---|
| EPI_ISL_448801 | Oxford Viromics, NDM, University of Oxford; Oxford University Hospitals; Basingstoke and North Hampshire Hospital | COVID-19 Genomics UK (COG-UK) Consortium |
| EPI_ISL_448802 | Oxford Viromics, NDM, University of Oxford; Oxford University Hospitals; Basingstoke and North Hampshire Hospital | COVID-19 Genomics UK (COG-UK) Consortium |
| EPI_ISL_448803 | Oxford Viromics, NDM, University of Oxford; Oxford University Hospitals; Basingstoke and North Hampshire Hospital | COVID-19 Genomics UK (COG-UK) Consortium |
| EPI_ISL_448804 | Oxford Viromics, NDM, University of Oxford; Oxford University Hospitals; Basingstoke and North Hampshire Hospital | COVID-19 Genomics UK (COG-UK) Consortium |
| EPI_ISL_448805 | Oxford Viromics, NDM, University of Oxford; Oxford University Hospitals; Basingstoke and North Hampshire Hospital | COVID-19 Genomics UK (COG-UK) Consortium |
| EPI_ISL_448806 | Oxford Viromics, NDM, University of Oxford; Oxford University Hospitals; Basingstoke and North Hampshire Hospital | COVID-19 Genomics UK (COG-UK) Consortium |
| EPI_ISL_448807 | Oxford Viromics, NDM, University of Oxford; Oxford University Hospitals; Basingstoke and North Hampshire Hospital | COVID-19 Genomics UK (COG-UK) Consortium |
| EPI_ISL_448808 | Oxford Viromics, NDM, University of Oxford; Oxford University Hospitals; Basingstoke and North Hampshire Hospital | COVID-19 Genomics UK (COG-UK) Consortium |
| EPI_ISL_448809 | Oxford Viromics, NDM, University of Oxford; Oxford University Hospitals; Basingstoke and North Hampshire Hospital | COVID-19 Genomics UK (COG-UK) Consortium |
| EPI_ISL_448810 | Oxford Viromics, NDM, University of Oxford; Oxford University Hospitals; Basingstoke and North Hampshire Hospital | COVID-19 Genomics UK (COG-UK) Consortium |
| EPI_ISL_448811 | Oxford Viromics, NDM, University of Oxford; Oxford University Hospitals; Basingstoke and North Hampshire Hospital | COVID-19 Genomics UK (COG-UK) Consortium |

| EPI_Accession_ID | Originating Laboratory | Submitting Laboratory |
|---|---|---|
| EPI_ISL_448812 | Oxford Viromics, NDM, University of Oxford; Oxford University Hospitals; Basingstoke and North Hampshire Hospital | COVID-19 Genomics UK (COG-UK) Consortium |
| EPI_ISL_448813 | Oxford Viromics, NDM, University of Oxford; Oxford University Hospitals; Basingstoke and North Hampshire Hospital | COVID-19 Genomics UK (COG-UK) Consortium |
| EPI_ISL_448814 | Oxford Viromics, NDM, University of Oxford; Oxford University Hospitals; Basingstoke and North Hampshire Hospital | COVID-19 Genomics UK (COG-UK) Consortium |
| EPI_ISL_448815 | Oxford Viromics, NDM, University of Oxford; Oxford University Hospitals; Basingstoke and North Hampshire Hospital | COVID-19 Genomics UK (COG-UK) Consortium |
| EPI_ISL_448816 | Oxford Viromics, NDM, University of Oxford; Oxford University Hospitals; Basingstoke and North Hampshire Hospital | COVID-19 Genomics UK (COG-UK) Consortium |
| EPI_ISL_448817 | Oxford Viromics, NDM, University of Oxford; Oxford University Hospitals; Basingstoke and North Hampshire Hospital | COVID-19 Genomics UK (COG-UK) Consortium |
| EPI_ISL_448818 | Oxford Viromics, NDM, University of Oxford; Oxford University Hospitals; Basingstoke and North Hampshire Hospital | COVID-19 Genomics UK (COG-UK) Consortium |
| EPI_ISL_448819 | Oxford Viromics, NDM, University of Oxford; Oxford University Hospitals; Basingstoke and North Hampshire Hospital | COVID-19 Genomics UK (COG-UK) Consortium |
| EPI_ISL_448820 | Oxford Viromics, NDM, University of Oxford; Oxford University Hospitals; Basingstoke and North Hampshire Hospital | COVID-19 Genomics UK (COG-UK) Consortium |
| EPI_ISL_448821 | Oxford Viromics, NDM, University of Oxford; Oxford University Hospitals; Basingstoke and North Hampshire Hospital | COVID-19 Genomics UK (COG-UK) Consortium |
| EPI_ISL_448822 | Oxford Viromics, NDM, University of Oxford; Oxford University Hospitals; Basingstoke and North Hampshire Hospital | COVID-19 Genomics UK (COG-UK) Consortium |

| EPI_Accession_ID | Originating Laboratory | Submitting Laboratory |
|---|---|---|
| EPI_ISL_448823 | Oxford Viromics, NDM, University of Oxford; Oxford University Hospitals; Basingstoke and North Hampshire Hospital | COVID-19 Genomics UK (COG-UK) Consortium |
| EPI_ISL_448824 | Oxford Viromics, NDM, University of Oxford; Oxford University Hospitals; Basingstoke and North Hampshire Hospital | COVID-19 Genomics UK (COG-UK) Consortium |
| EPI_ISL_448825 | Oxford Viromics, NDM, University of Oxford; Oxford University Hospitals; Basingstoke and North Hampshire Hospital | COVID-19 Genomics UK (COG-UK) Consortium |
| EPI_ISL_448826 | Virology Department, Sheffield Teaching Hospitals NHS Foundation Trust/Department of Infection, Immunity and Cardiovascular Disease, The Medical School, University of Sheffield | COVID-19 Genomics UK (COG-UK) Consortium |
| EPI_ISL_448827 | Virology Department, Sheffield Teaching Hospitals NHS Foundation Trust/Department of Infection, Immunity and Cardiovascular Disease, The Medical School, University of Sheffield | COVID-19 Genomics UK (COG-UK) Consortium |
| EPI_ISL_448828 | Virology Department, Sheffield Teaching Hospitals NHS Foundation Trust/Department of Infection, Immunity and Cardiovascular Disease, The Medical School, University of Sheffield | COVID-19 Genomics UK (COG-UK) Consortium |
| EPI_ISL_448829 | Virology Department, Sheffield Teaching Hospitals NHS Foundation Trust/Department of Infection, Immunity and Cardiovascular Disease, The Medical School, University of Sheffield | COVID-19 Genomics UK (COG-UK) Consortium |
| EPI_ISL_448830 | Virology Department, Sheffield Teaching Hospitals NHS Foundation Trust/Department of Infection, Immunity and Cardiovascular Disease, The Medical School, University of Sheffield | COVID-19 Genomics UK (COG-UK) Consortium |
| EPI_ISL_448831 | Virology Department, Sheffield Teaching Hospitals NHS Foundation Trust/Department of Infection, Immunity and Cardiovascular Disease, The Medical School, University of Sheffield | COVID-19 Genomics UK (COG-UK) Consortium |

| EPI_Accession_ID | Originating Laboratory | Submitting Laboratory |
|---|---|---|
| EPI_ISL_448832 | Virology Department, Sheffield Teaching Hospitals NHS Foundation Trust/Department of Infection, Immunity and Cardiovascular Disease, The Medical School, University of Sheffield | COVID-19 Genomics UK (COG-UK) Consortium |
| EPI_ISL_448833 | Virology Department, Sheffield Teaching Hospitals NHS Foundation Trust/Department of Infection, Immunity and Cardiovascular Disease, The Medical School, University of Sheffield | COVID-19 Genomics UK (COG-UK) Consortium |
| EPI_ISL_448834 | Virology Department, Sheffield Teaching Hospitals NHS Foundation Trust/Department of Infection, Immunity and Cardiovascular Disease, The Medical School, University of Sheffield | COVID-19 Genomics UK (COG-UK) Consortium |
| EPI_ISL_448835 | Virology Department, Sheffield Teaching Hospitals NHS Foundation Trust/Department of Infection, Immunity and Cardiovascular Disease, The Medical School, University of Sheffield | COVID-19 Genomics UK (COG-UK) Consortium |
| EPI_ISL_448836 | Virology Department, Sheffield Teaching Hospitals NHS Foundation Trust/Department of Infection, Immunity and Cardiovascular Disease, The Medical School, University of Sheffield | COVID-19 Genomics UK (COG-UK) Consortium |
| EPI_ISL_448837 | Virology Department, Sheffield Teaching Hospitals NHS Foundation Trust/Department of Infection, Immunity and Cardiovascular Disease, The Medical School, University of Sheffield | COVID-19 Genomics UK (COG-UK) Consortium |
| EPI_ISL_448838 | Virology Department, Sheffield Teaching Hospitals NHS Foundation Trust/Department of Infection, Immunity and Cardiovascular Disease, The Medical School, University of Sheffield | COVID-19 Genomics UK (COG-UK) Consortium |
| EPI_ISL_448839 | Virology Department, Sheffield Teaching Hospitals NHS Foundation Trust/Department of Infection, Immunity and Cardiovascular Disease, The Medical School, University of Sheffield | COVID-19 Genomics UK (COG-UK) Consortium |

| EPI_Accession_ID | Originating Laboratory | Submitting Laboratory |
|---|---|---|
| EPI_ISL_448840 | Virology Department, Sheffield Teaching Hospitals NHS Foundation Trust/Department of Infection, Immunity and Cardiovascular Disease, The Medical School, University of Sheffield | COVID-19 Genomics UK (COG-UK) Consortium |
| EPI_ISL_448841 | Virology Department, Sheffield Teaching Hospitals NHS Foundation Trust/Department of Infection, Immunity and Cardiovascular Disease, The Medical School, University of Sheffield | COVID-19 Genomics UK (COG-UK) Consortium |
| EPI_ISL_448842 | Virology Department, Sheffield Teaching Hospitals NHS Foundation Trust/Department of Infection, Immunity and Cardiovascular Disease, The Medical School, University of Sheffield | COVID-19 Genomics UK (COG-UK) Consortium |
| EPI_ISL_448843 | Virology Department, Sheffield Teaching Hospitals NHS Foundation Trust/Department of Infection, Immunity and Cardiovascular Disease, The Medical School, University of Sheffield | COVID-19 Genomics UK (COG-UK) Consortium |
| EPI_ISL_448844 | Virology Laboratory, Castle Hill Hospital, Hull University Teaching Hospitals NHS Trust/Department of Infection, Immunity and Cardiovascular Disease, The Medical School, University of Sheffield | COVID-19 Genomics UK (COG-UK) Consortium |
| EPI_ISL_448845 | Virology Department, Sheffield Teaching Hospitals NHS Foundation Trust/Department of Infection, Immunity and Cardiovascular Disease, The Medical School, University of Sheffield | COVID-19 Genomics UK (COG-UK) Consortium |
| EPI_ISL_448846 | Virology Department, Sheffield Teaching Hospitals NHS Foundation Trust/Department of Infection, Immunity and Cardiovascular Disease, The Medical School, University of Sheffield | COVID-19 Genomics UK (COG-UK) Consortium |
| EPI_ISL_448847 | Virology Department, Sheffield Teaching Hospitals NHS Foundation Trust/Department of Infection, Immunity and Cardiovascular Disease, The Medical School, University of Sheffield | COVID-19 Genomics UK (COG-UK) Consortium |

| EPI_Accession_ID | Originating Laboratory | Submitting Laboratory |
|---|---|---|
| EPI_ISL_448848 | Virology Department, Sheffield Teaching Hospitals NHS Foundation Trust/Department of Infection, Immunity and Cardiovascular Disease, The Medical School, University of Sheffield | COVID-19 Genomics UK (COG-UK) Consortium |
| EPI_ISL_448849 | Virology Department, Sheffield Teaching Hospitals NHS Foundation Trust/Department of Infection, Immunity and Cardiovascular Disease, The Medical School, University of Sheffield | COVID-19 Genomics UK (COG-UK) Consortium |
| EPI_ISL_448850 | Virology Laboratory, Castle Hill Hospital, Hull University Teaching Hospitals NHS Trust/Department of Infection, Immunity and Cardiovascular Disease, The Medical School, University of Sheffield | COVID-19 Genomics UK (COG-UK) Consortium |
| EPI_ISL_448851 | Virology Department, Sheffield Teaching Hospitals NHS Foundation Trust/Department of Infection, Immunity and Cardiovascular Disease, The Medical School, University of Sheffield | COVID-19 Genomics UK (COG-UK) Consortium |
| EPI_ISL_448852 | Virology Department, Sheffield Teaching Hospitals NHS Foundation Trust/Department of Infection, Immunity and Cardiovascular Disease, The Medical School, University of Sheffield | COVID-19 Genomics UK (COG-UK) Consortium |
| EPI_ISL_448853 | Virology Department, Sheffield Teaching Hospitals NHS Foundation Trust/Department of Infection, Immunity and Cardiovascular Disease, The Medical School, University of Sheffield | COVID-19 Genomics UK (COG-UK) Consortium |
| EPI_ISL_448854 | Virology Department, Sheffield Teaching Hospitals NHS Foundation Trust/Department of Infection, Immunity and Cardiovascular Disease, The Medical School, University of Sheffield | COVID-19 Genomics UK (COG-UK) Consortium |
| EPI_ISL_448855 | Virology Department, Sheffield Teaching Hospitals NHS Foundation Trust/Department of Infection, Immunity and Cardiovascular Disease, The Medical School, University of Sheffield | COVID-19 Genomics UK (COG-UK) Consortium |

| EPI_Accession_ID | Originating Laboratory | Submitting Laboratory |
|---|---|---|
| EPI_ISL_448856 | Virology Department, Sheffield Teaching Hospitals NHS Foundation Trust/Department of Infection, Immunity and Cardiovascular Disease, The Medical School, University of Sheffield | COVID-19 Genomics UK (COG-UK) Consortium |
| EPI_ISL_448857 | Virology Department, Sheffield Teaching Hospitals NHS Foundation Trust/Department of Infection, Immunity and Cardiovascular Disease, The Medical School, University of Sheffield | COVID-19 Genomics UK (COG-UK) Consortium |
| EPI_ISL_448858 | Virology Department, Sheffield Teaching Hospitals NHS Foundation Trust/Department of Infection, Immunity and Cardiovascular Disease, The Medical School, University of Sheffield | COVID-19 Genomics UK (COG-UK) Consortium |
| EPI_ISL_448859 | Virology Department, Sheffield Teaching Hospitals NHS Foundation Trust/Department of Infection, Immunity and Cardiovascular Disease, The Medical School, University of Sheffield | COVID-19 Genomics UK (COG-UK) Consortium |
| EPI_ISL_448860 | Virology Department, Sheffield Teaching Hospitals NHS Foundation Trust/Department of Infection, Immunity and Cardiovascular Disease, The Medical School, University of Sheffield | COVID-19 Genomics UK (COG-UK) Consortium |
| EPI_ISL_448861 | Virology Department, Sheffield Teaching Hospitals NHS Foundation Trust/Department of Infection, Immunity and Cardiovascular Disease, The Medical School, University of Sheffield | COVID-19 Genomics UK (COG-UK) Consortium |
| EPI_ISL_448862 | Virology Department, Sheffield Teaching Hospitals NHS Foundation Trust/Department of Infection, Immunity and Cardiovascular Disease, The Medical School, University of Sheffield | COVID-19 Genomics UK (COG-UK) Consortium |
| EPI_ISL_448863 | Virology Department, Sheffield Teaching Hospitals NHS Foundation Trust/Department of Infection, Immunity and Cardiovascular Disease, The Medical School, University of Sheffield | COVID-19 Genomics UK (COG-UK) Consortium |

| EPI_Accession_ID | Originating Laboratory | Submitting Laboratory |
|---|---|---|
| EPI_ISL_448864 | Virology Laboratory, Castle Hill Hospital, Hull University Teaching Hospitals NHS Trust/Department of Infection, Immunity and Cardiovascular Disease, The Medical School, University of Sheffield | COVID-19 Genomics UK (COG-UK) Consortium |
| EPI_ISL_448865 | Virology Department, Sheffield Teaching Hospitals NHS Foundation Trust/Department of Infection, Immunity and Cardiovascular Disease, The Medical School, University of Sheffield | COVID-19 Genomics UK (COG-UK) Consortium |
| EPI_ISL_448866 | Virology Laboratory, Castle Hill Hospital, Hull University Teaching Hospitals NHS Trust/Department of Infection, Immunity and Cardiovascular Disease, The Medical School, University of Sheffield | COVID-19 Genomics UK (COG-UK) Consortium |
| EPI_ISL_448867 | Virology Department, Sheffield Teaching Hospitals NHS Foundation Trust/Department of Infection, Immunity and Cardiovascular Disease, The Medical School, University of Sheffield | COVID-19 Genomics UK (COG-UK) Consortium |
| EPI_ISL_448868 | Virology Department, Sheffield Teaching Hospitals NHS Foundation Trust/Department of Infection, Immunity and Cardiovascular Disease, The Medical School, University of Sheffield | COVID-19 Genomics UK (COG-UK) Consortium |
| EPI_ISL_448869 | Virology Department, Sheffield Teaching Hospitals NHS Foundation Trust/Department of Infection, Immunity and Cardiovascular Disease, The Medical School, University of Sheffield | COVID-19 Genomics UK (COG-UK) Consortium |
| EPI_ISL_448870 | Virology Department, Sheffield Teaching Hospitals NHS Foundation Trust/Department of Infection, Immunity and Cardiovascular Disease, The Medical School, University of Sheffield | COVID-19 Genomics UK (COG-UK) Consortium |
| EPI_ISL_448871 | Virology Department, Sheffield Teaching Hospitals NHS Foundation Trust/Department of Infection, Immunity and Cardiovascular Disease, The Medical School, University of Sheffield | COVID-19 Genomics UK (COG-UK) Consortium |

| EPI_Accession_ID | Originating Laboratory | Submitting Laboratory |
|---|---|---|
| EPI_ISL_448872 | Virology Department, Sheffield Teaching Hospitals NHS Foundation Trust/Department of Infection, Immunity and Cardiovascular Disease, The Medical School, University of Sheffield | COVID-19 Genomics UK (COG-UK) Consortium |
| EPI_ISL_448873 | Virology Department, Sheffield Teaching Hospitals NHS Foundation Trust/Department of Infection, Immunity and Cardiovascular Disease, The Medical School, University of Sheffield | COVID-19 Genomics UK (COG-UK) Consortium |
| EPI_ISL_448874 | Virology Department, Sheffield Teaching Hospitals NHS Foundation Trust/Department of Infection, Immunity and Cardiovascular Disease, The Medical School, University of Sheffield | COVID-19 Genomics UK (COG-UK) Consortium |
| EPI_ISL_448875 | Virology Department, Sheffield Teaching Hospitals NHS Foundation Trust/Department of Infection, Immunity and Cardiovascular Disease, The Medical School, University of Sheffield | COVID-19 Genomics UK (COG-UK) Consortium |
| EPI_ISL_448876 | Virology Department, Sheffield Teaching Hospitals NHS Foundation Trust/Department of Infection, Immunity and Cardiovascular Disease, The Medical School, University of Sheffield | COVID-19 Genomics UK (COG-UK) Consortium |
| EPI_ISL_448877 | Virology Department, Sheffield Teaching Hospitals NHS Foundation Trust/Department of Infection, Immunity and Cardiovascular Disease, The Medical School, University of Sheffield | COVID-19 Genomics UK (COG-UK) Consortium |
| EPI_ISL_448878 | Virology Department, Sheffield Teaching Hospitals NHS Foundation Trust/Department of Infection, Immunity and Cardiovascular Disease, The Medical School, University of Sheffield | COVID-19 Genomics UK (COG-UK) Consortium |
| EPI_ISL_448879 | Virology Department, Sheffield Teaching Hospitals NHS Foundation Trust/Department of Infection, Immunity and Cardiovascular Disease, The Medical School, University of Sheffield | COVID-19 Genomics UK (COG-UK) Consortium |

| EPI_Accession_ID | Originating Laboratory | Submitting Laboratory |
|---|---|---|
| EPI_ISL_448880 | Virology Department, Sheffield Teaching Hospitals NHS Foundation Trust/Department of Infection, Immunity and Cardiovascular Disease, The Medical School, University of Sheffield | COVID-19 Genomics UK (COG-UK) Consortium |
| EPI_ISL_448881 | Virology Department, Sheffield Teaching Hospitals NHS Foundation Trust/Department of Infection, Immunity and Cardiovascular Disease, The Medical School, University of Sheffield | COVID-19 Genomics UK (COG-UK) Consortium |
| EPI_ISL_448882 | Virology Department, Sheffield Teaching Hospitals NHS Foundation Trust/Department of Infection, Immunity and Cardiovascular Disease, The Medical School, University of Sheffield | COVID-19 Genomics UK (COG-UK) Consortium |
| EPI_ISL_448883 | Virology Department, Sheffield Teaching Hospitals NHS Foundation Trust/Department of Infection, Immunity and Cardiovascular Disease, The Medical School, University of Sheffield | COVID-19 Genomics UK (COG-UK) Consortium |
| EPI_ISL_448884 | Virology Department, Sheffield Teaching Hospitals NHS Foundation Trust/Department of Infection, Immunity and Cardiovascular Disease, The Medical School, University of Sheffield | COVID-19 Genomics UK (COG-UK) Consortium |
| EPI_ISL_448885 | Virology Department, Sheffield Teaching Hospitals NHS Foundation Trust/Department of Infection, Immunity and Cardiovascular Disease, The Medical School, University of Sheffield | COVID-19 Genomics UK (COG-UK) Consortium |
| EPI_ISL_448886 | Virology Department, Sheffield Teaching Hospitals NHS Foundation Trust/Department of Infection, Immunity and Cardiovascular Disease, The Medical School, University of Sheffield | COVID-19 Genomics UK (COG-UK) Consortium |
| EPI_ISL_448887 | Virology Department, Sheffield Teaching Hospitals NHS Foundation Trust/Department of Infection, Immunity and Cardiovascular Disease, The Medical School, University of Sheffield | COVID-19 Genomics UK (COG-UK) Consortium |

| EPI_Accession_ID | Originating Laboratory | Submitting Laboratory |
|---|---|---|
| EPI_ISL_448888 | Virology Department, Sheffield Teaching Hospitals NHS Foundation Trust/Department of Infection, Immunity and Cardiovascular Disease, The Medical School, University of Sheffield | COVID-19 Genomics UK (COG-UK) Consortium |
| EPI_ISL_448889 | Virology Department, Sheffield Teaching Hospitals NHS Foundation Trust/Department of Infection, Immunity and Cardiovascular Disease, The Medical School, University of Sheffield | COVID-19 Genomics UK (COG-UK) Consortium |
| EPI_ISL_448890 | Virology Department, Sheffield Teaching Hospitals NHS Foundation Trust/Department of Infection, Immunity and Cardiovascular Disease, The Medical School, University of Sheffield | COVID-19 Genomics UK (COG-UK) Consortium |
| EPI_ISL_448891 | Virology Department, Sheffield Teaching Hospitals NHS Foundation Trust/Department of Infection, Immunity and Cardiovascular Disease, The Medical School, University of Sheffield | COVID-19 Genomics UK (COG-UK) Consortium |
| EPI_ISL_448892 | Virology Department, Sheffield Teaching Hospitals NHS Foundation Trust/Department of Infection, Immunity and Cardiovascular Disease, The Medical School, University of Sheffield | COVID-19 Genomics UK (COG-UK) Consortium |
| EPI_ISL_448893 | Virology Department, Sheffield Teaching Hospitals NHS Foundation Trust/Department of Infection, Immunity and Cardiovascular Disease, The Medical School, University of Sheffield | COVID-19 Genomics UK (COG-UK) Consortium |
| EPI_ISL_448894 | Virology Laboratory, Castle Hill Hospital, Hull University Teaching Hospitals NHS Trust/Department of Infection, Immunity and Cardiovascular Disease, The Medical School, University of Sheffield | COVID-19 Genomics UK (COG-UK) Consortium |
| EPI_ISL_448895 | Virology Department, Sheffield Teaching Hospitals NHS Foundation Trust/Department of Infection, Immunity and Cardiovascular Disease, The Medical School, University of Sheffield | COVID-19 Genomics UK (COG-UK) Consortium |

| EPI_Accession_ID | Originating Laboratory | Submitting Laboratory |
| --- | --- | --- |
| EPI_ISL_448896 | Virology Laboratory, Castle Hill Hospital, Hull University Teaching Hospitals NHS Trust/Department of Infection, Immunity and Cardiovascular Disease, The Medical School, University of Sheffield | COVID-19 Genomics UK (COG-UK) Consortium |
| EPI_ISL_448897 | Virology Department, Sheffield Teaching Hospitals NHS Foundation Trust/Department of Infection, Immunity and Cardiovascular Disease, The Medical School, University of Sheffield | COVID-19 Genomics UK (COG-UK) Consortium |
| EPI_ISL_448898 | Virology Department, Sheffield Teaching Hospitals NHS Foundation Trust/Department of Infection, Immunity and Cardiovascular Disease, The Medical School, University of Sheffield | COVID-19 Genomics UK (COG-UK) Consortium |
| EPI_ISL_448899 | Virology Department, Sheffield Teaching Hospitals NHS Foundation Trust/Department of Infection, Immunity and Cardiovascular Disease, The Medical School, University of Sheffield | COVID-19 Genomics UK (COG-UK) Consortium |
| EPI_ISL_448900 | Virology Department, Sheffield Teaching Hospitals NHS Foundation Trust/Department of Infection, Immunity and Cardiovascular Disease, The Medical School, University of Sheffield | COVID-19 Genomics UK (COG-UK) Consortium |
| EPI_ISL_448901 | Virology Department, Sheffield Teaching Hospitals NHS Foundation Trust/Department of Infection, Immunity and Cardiovascular Disease, The Medical School, University of Sheffield | COVID-19 Genomics UK (COG-UK) Consortium |
| EPI_ISL_448902 | Virology Laboratory, Castle Hill Hospital, Hull University Teaching Hospitals NHS Trust/Department of Infection, Immunity and Cardiovascular Disease, The Medical School, University of Sheffield | COVID-19 Genomics UK (COG-UK) Consortium |
| EPI_ISL_448903 | Virology Department, Sheffield Teaching Hospitals NHS Foundation Trust/Department of Infection, Immunity and Cardiovascular Disease, The Medical School, University of Sheffield | COVID-19 Genomics UK (COG-UK) Consortium |

| EPI_Accession_ID | Originating Laboratory | Submitting Laboratory |
|---|---|---|
| EPI_ISL_448904 | Virology Department, Sheffield Teaching Hospitals NHS Foundation Trust/Department of Infection, Immunity and Cardiovascular Disease, The Medical School, University of Sheffield | COVID-19 Genomics UK (COG-UK) Consortium |
| EPI_ISL_448905 | Virology Department, Sheffield Teaching Hospitals NHS Foundation Trust/Department of Infection, Immunity and Cardiovascular Disease, The Medical School, University of Sheffield | COVID-19 Genomics UK (COG-UK) Consortium |
| EPI_ISL_448906 | Virology Laboratory, Castle Hill Hospital, Hull University Teaching Hospitals NHS Trust/Department of Infection, Immunity and Cardiovascular Disease, The Medical School, University of Sheffield | COVID-19 Genomics UK (COG-UK) Consortium |
| EPI_ISL_448907 | Virology Department, Sheffield Teaching Hospitals NHS Foundation Trust/Department of Infection, Immunity and Cardiovascular Disease, The Medical School, University of Sheffield | COVID-19 Genomics UK (COG-UK) Consortium |
| EPI_ISL_448908 | Virology Department, Sheffield Teaching Hospitals NHS Foundation Trust/Department of Infection, Immunity and Cardiovascular Disease, The Medical School, University of Sheffield | COVID-19 Genomics UK (COG-UK) Consortium |
| EPI_ISL_448909 | Virology Department, Sheffield Teaching Hospitals NHS Foundation Trust/Department of Infection, Immunity and Cardiovascular Disease, The Medical School, University of Sheffield | COVID-19 Genomics UK (COG-UK) Consortium |
| EPI_ISL_448910 | Virology Department, Sheffield Teaching Hospitals NHS Foundation Trust/Department of Infection, Immunity and Cardiovascular Disease, The Medical School, University of Sheffield | COVID-19 Genomics UK (COG-UK) Consortium |
| EPI_ISL_448911 | Virology Department, Sheffield Teaching Hospitals NHS Foundation Trust/Department of Infection, Immunity and Cardiovascular Disease, The Medical School, University of Sheffield | COVID-19 Genomics UK (COG-UK) Consortium |

| EPI_Accession_ID | Originating Laboratory | Submitting Laboratory |
| --- | --- | --- |
| EPI_ISL_448912 | Virology Department, Sheffield Teaching Hospitals NHS Foundation Trust/Department of Infection, Immunity and Cardiovascular Disease, The Medical School, University of Sheffield | COVID-19 Genomics UK (COG-UK) Consortium |
| EPI_ISL_448913 | Virology Department, Sheffield Teaching Hospitals NHS Foundation Trust/Department of Infection, Immunity and Cardiovascular Disease, The Medical School, University of Sheffield | COVID-19 Genomics UK (COG-UK) Consortium |
| EPI_ISL_448914 | Virology Department, Sheffield Teaching Hospitals NHS Foundation Trust/Department of Infection, Immunity and Cardiovascular Disease, The Medical School, University of Sheffield | COVID-19 Genomics UK (COG-UK) Consortium |
| EPI_ISL_448915 | Virology Department, Sheffield Teaching Hospitals NHS Foundation Trust/Department of Infection, Immunity and Cardiovascular Disease, The Medical School, University of Sheffield | COVID-19 Genomics UK (COG-UK) Consortium |
| EPI_ISL_448916 | Virology Department, Sheffield Teaching Hospitals NHS Foundation Trust/Department of Infection, Immunity and Cardiovascular Disease, The Medical School, University of Sheffield | COVID-19 Genomics UK (COG-UK) Consortium |
| EPI_ISL_448917 | Virology Department, Sheffield Teaching Hospitals NHS Foundation Trust/Department of Infection, Immunity and Cardiovascular Disease, The Medical School, University of Sheffield | COVID-19 Genomics UK (COG-UK) Consortium |
| EPI_ISL_448918 | Regional Virus Laboratory, Belfast Health and Social Care Trust | COVID-19 Genomics UK (COG-UK) Consortium |
| EPI_ISL_448919 | Regional Virus Laboratory, Belfast Health and Social Care Trust | COVID-19 Genomics UK (COG-UK) Consortium |
| EPI_ISL_448920 | Regional Virus Laboratory, Belfast Health and Social Care Trust | COVID-19 Genomics UK (COG-UK) Consortium |
| EPI_ISL_448921 | Regional Virus Laboratory, Belfast Health and Social Care Trust | COVID-19 Genomics UK (COG-UK) Consortium |

| EPI_Accession_ID | Originating Laboratory | Submitting Laboratory |
|---|---|---|
| EPI_ISL_448922 | Regional Virus Laboratory, Belfast Health and Social Care Trust | COVID-19 Genomics UK (COG-UK) Consortium |
| EPI_ISL_448923 | Regional Virus Laboratory, Belfast Health and Social Care Trust | COVID-19 Genomics UK (COG-UK) Consortium |
| EPI_ISL_448924 | Regional Virus Laboratory, Belfast Health and Social Care Trust | COVID-19 Genomics UK (COG-UK) Consortium |
| EPI_ISL_448925 | Regional Virus Laboratory, Belfast Health and Social Care Trust | COVID-19 Genomics UK (COG-UK) Consortium |
| EPI_ISL_448926 | Regional Virus Laboratory, Belfast Health and Social Care Trust | COVID-19 Genomics UK (COG-UK) Consortium |
| EPI_ISL_448927 | Regional Virus Laboratory, Belfast Health and Social Care Trust | COVID-19 Genomics UK (COG-UK) Consortium |
| EPI_ISL_448928 | Regional Virus Laboratory, Belfast Health and Social Care Trust | COVID-19 Genomics UK (COG-UK) Consortium |
| EPI_ISL_448929 | Regional Virus Laboratory, Belfast Health and Social Care Trust | COVID-19 Genomics UK (COG-UK) Consortium |
| EPI_ISL_448930 | Regional Virus Laboratory, Belfast Health and Social Care Trust | COVID-19 Genomics UK (COG-UK) Consortium |
| EPI_ISL_448931 | Regional Virus Laboratory, Belfast Health and Social Care Trust | COVID-19 Genomics UK (COG-UK) Consortium |
| EPI_ISL_448932 | Regional Virus Laboratory, Belfast Health and Social Care Trust | COVID-19 Genomics UK (COG-UK) Consortium |
| EPI_ISL_448933 | Regional Virus Laboratory, Belfast Health and Social Care Trust | COVID-19 Genomics UK (COG-UK) Consortium |
| EPI_ISL_448934 | Regional Virus Laboratory, Belfast Health and Social Care Trust | COVID-19 Genomics UK (COG-UK) Consortium |
| EPI_ISL_448935 | Regional Virus Laboratory, Belfast Health and Social Care Trust | COVID-19 Genomics UK (COG-UK) Consortium |
| EPI_ISL_448936 | Regional Virus Laboratory, Belfast Health and Social Care Trust | COVID-19 Genomics UK (COG-UK) Consortium |
| EPI_ISL_448937 | Regional Virus Laboratory, Belfast Health and Social Care Trust | COVID-19 Genomics UK (COG-UK) Consortium |

| EPI_Accession_ID | Originating Laboratory | Submitting Laboratory |
|---|---|---|
| EPI_ISL_448938 | Regional Virus Laboratory, Belfast Health and Social Care Trust | COVID-19 Genomics UK (COG-UK) Consortium |
| EPI_ISL_448939 | Regional Virus Laboratory, Belfast Health and Social Care Trust | COVID-19 Genomics UK (COG-UK) Consortium |
| EPI_ISL_448940 | Regional Virus Laboratory, Belfast Health and Social Care Trust | COVID-19 Genomics UK (COG-UK) Consortium |
| EPI_ISL_448941 | Regional Virus Laboratory, Belfast Health and Social Care Trust | COVID-19 Genomics UK (COG-UK) Consortium |
| EPI_ISL_448942 | Regional Virus Laboratory, Belfast Health and Social Care Trust | COVID-19 Genomics UK (COG-UK) Consortium |
| EPI_ISL_448943 | Regional Virus Laboratory, Belfast Health and Social Care Trust | COVID-19 Genomics UK (COG-UK) Consortium |
| EPI_ISL_448944 | Regional Virus Laboratory, Belfast Health and Social Care Trust | COVID-19 Genomics UK (COG-UK) Consortium |
| EPI_ISL_448945 | Regional Virus Laboratory, Belfast Health and Social Care Trust | COVID-19 Genomics UK (COG-UK) Consortium |
| EPI_ISL_448946 | Regional Virus Laboratory, Belfast Health and Social Care Trust | COVID-19 Genomics UK (COG-UK) Consortium |
| EPI_ISL_448947 | Regional Virus Laboratory, Belfast Health and Social Care Trust | COVID-19 Genomics UK (COG-UK) Consortium |
| EPI_ISL_448948 | Regional Virus Laboratory, Belfast Health and Social Care Trust | COVID-19 Genomics UK (COG-UK) Consortium |
| EPI_ISL_448949 | Regional Virus Laboratory, Belfast Health and Social Care Trust | COVID-19 Genomics UK (COG-UK) Consortium |
| EPI_ISL_448950 | Regional Virus Laboratory, Belfast Health and Social Care Trust | COVID-19 Genomics UK (COG-UK) Consortium |
| EPI_ISL_448951 | Regional Virus Laboratory, Belfast Health and Social Care Trust | COVID-19 Genomics UK (COG-UK) Consortium |
| EPI_ISL_448952 | Regional Virus Laboratory, Belfast Health and Social Care Trust | COVID-19 Genomics UK (COG-UK) Consortium |
| EPI_ISL_448953 | Regional Virus Laboratory, Belfast Health and Social Care Trust | COVID-19 Genomics UK (COG-UK) Consortium |

| EPI_Accession_ID | Originating Laboratory | Submitting Laboratory |
|---|---|---|
| EPI_ISL_448954 | Regional Virus Laboratory, Belfast Health and Social Care Trust | COVID-19 Genomics UK (COG-UK) Consortium |
| EPI_ISL_448955 | Regional Virus Laboratory, Belfast Health and Social Care Trust | COVID-19 Genomics UK (COG-UK) Consortium |
| EPI_ISL_448956 | Regional Virus Laboratory, Belfast Health and Social Care Trust | COVID-19 Genomics UK (COG-UK) Consortium |
| EPI_ISL_448957 | Regional Virus Laboratory, Belfast Health and Social Care Trust | COVID-19 Genomics UK (COG-UK) Consortium |
| EPI_ISL_448958 | Regional Virus Laboratory, Belfast Health and Social Care Trust | COVID-19 Genomics UK (COG-UK) Consortium |
| EPI_ISL_448959 | Regional Virus Laboratory, Belfast Health and Social Care Trust | COVID-19 Genomics UK (COG-UK) Consortium |
| EPI_ISL_448960 | Regional Virus Laboratory, Belfast Health and Social Care Trust | COVID-19 Genomics UK (COG-UK) Consortium |
| EPI_ISL_448961 | Regional Virus Laboratory, Belfast Health and Social Care Trust | COVID-19 Genomics UK (COG-UK) Consortium |
| EPI_ISL_448962 | Regional Virus Laboratory, Belfast Health and Social Care Trust | COVID-19 Genomics UK (COG-UK) Consortium |
| EPI_ISL_448963 | Regional Virus Laboratory, Belfast Health and Social Care Trust | COVID-19 Genomics UK (COG-UK) Consortium |
| EPI_ISL_448964 | Regional Virus Laboratory, Belfast Health and Social Care Trust | COVID-19 Genomics UK (COG-UK) Consortium |
| EPI_ISL_448965 | Regional Virus Laboratory, Belfast Health and Social Care Trust | COVID-19 Genomics UK (COG-UK) Consortium |
| EPI_ISL_448966 | Regional Virus Laboratory, Belfast Health and Social Care Trust | COVID-19 Genomics UK (COG-UK) Consortium |
| EPI_ISL_448967 | Regional Virus Laboratory, Belfast Health and Social Care Trust | COVID-19 Genomics UK (COG-UK) Consortium |
| EPI_ISL_448968 | Regional Virus Laboratory, Belfast Health and Social Care Trust | COVID-19 Genomics UK (COG-UK) Consortium |
| EPI_ISL_448969 | Regional Virus Laboratory, Belfast Health and Social Care Trust | COVID-19 Genomics UK (COG-UK) Consortium |

| EPI_Accession_ID | Originating Laboratory | Submitting Laboratory |
| --- | --- | --- |
| EPI_ISL_448970 | Regional Virus Laboratory, Belfast Health and Social Care Trust | COVID-19 Genomics UK (COG-UK) Consortium |
| EPI_ISL_448971 | Regional Virus Laboratory, Belfast Health and Social Care Trust | COVID-19 Genomics UK (COG-UK) Consortium |
| EPI_ISL_448972 | Regional Virus Laboratory, Belfast Health and Social Care Trust | COVID-19 Genomics UK (COG-UK) Consortium |
| EPI_ISL_448973 | Regional Virus Laboratory, Belfast Health and Social Care Trust | COVID-19 Genomics UK (COG-UK) Consortium |
| EPI_ISL_448974 | Regional Virus Laboratory, Belfast Health and Social Care Trust | COVID-19 Genomics UK (COG-UK) Consortium |
| EPI_ISL_448975 | Regional Virus Laboratory, Belfast Health and Social Care Trust | COVID-19 Genomics UK (COG-UK) Consortium |
| EPI_ISL_448976 | Regional Virus Laboratory, Belfast Health and Social Care Trust | COVID-19 Genomics UK (COG-UK) Consortium |
| EPI_ISL_448977 | Regional Virus Laboratory, Belfast Health and Social Care Trust | COVID-19 Genomics UK (COG-UK) Consortium |
| EPI_ISL_448978 | Quadram Institute Bioscience | COVID-19 Genomics UK (COG-UK) Consortium |
| EPI_ISL_448979 | Quadram Institute Bioscience | COVID-19 Genomics UK (COG-UK) Consortium |
| EPI_ISL_448980 | Quadram Institute Bioscience | COVID-19 Genomics UK (COG-UK) Consortium |
| EPI_ISL_448981 | Quadram Institute Bioscience | COVID-19 Genomics UK (COG-UK) Consortium |
| EPI_ISL_448982 | Quadram Institute Bioscience | COVID-19 Genomics UK (COG-UK) Consortium |
| EPI_ISL_448983 | Quadram Institute Bioscience | COVID-19 Genomics UK (COG-UK) Consortium |
| EPI_ISL_448984 | Quadram Institute Bioscience | COVID-19 Genomics UK (COG-UK) Consortium |
| EPI_ISL_448985 | Quadram Institute Bioscience | COVID-19 Genomics UK (COG-UK) Consortium |
| EPI_ISL_448986 | Quadram Institute Bioscience | COVID-19 Genomics UK (COG-UK) Consortium |
| EPI_ISL_448988 | Quadram Institute Bioscience | COVID-19 Genomics UK (COG-UK) Consortium |
| EPI_ISL_448989 | Quadram Institute Bioscience | COVID-19 Genomics UK (COG-UK) Consortium |
| EPI_ISL_448990 | Quadram Institute Bioscience | COVID-19 Genomics UK (COG-UK) Consortium |
| EPI_ISL_448991 | Quadram Institute Bioscience | COVID-19 Genomics UK (COG-UK) Consortium |
| EPI_ISL_448992 | Quadram Institute Bioscience | COVID-19 Genomics UK (COG-UK) Consortium |
| EPI_ISL_448994 | Quadram Institute Bioscience | COVID-19 Genomics UK (COG-UK) Consortium |
| EPI_ISL_448995 | Quadram Institute Bioscience | COVID-19 Genomics UK (COG-UK) Consortium |
| EPI_ISL_448996 | Quadram Institute Bioscience | COVID-19 Genomics UK (COG-UK) Consortium |

| EPI_Accession_ID | Originating Laboratory | Submitting Laboratory |
|---|---|---|
| EPI_ISL_448997 | Quadram Institute Bioscience | COVID-19 Genomics UK (COG-UK) Consortium |
| EPI_ISL_448998 | Quadram Institute Bioscience | COVID-19 Genomics UK (COG-UK) Consortium |
| EPI_ISL_448999 | Quadram Institute Bioscience | COVID-19 Genomics UK (COG-UK) Consortium |
| EPI_ISL_449000 | Quadram Institute Bioscience | COVID-19 Genomics UK (COG-UK) Consortium |
| EPI_ISL_449001 | Quadram Institute Bioscience | COVID-19 Genomics UK (COG-UK) Consortium |
| EPI_ISL_449002 | Quadram Institute Bioscience | COVID-19 Genomics UK (COG-UK) Consortium |
| EPI_ISL_449003 | Quadram Institute Bioscience | COVID-19 Genomics UK (COG-UK) Consortium |
| EPI_ISL_449004 | Quadram Institute Bioscience | COVID-19 Genomics UK (COG-UK) Consortium |
| EPI_ISL_449005 | Quadram Institute Bioscience | COVID-19 Genomics UK (COG-UK) Consortium |
| EPI_ISL_449006 | Quadram Institute Bioscience | COVID-19 Genomics UK (COG-UK) Consortium |
| EPI_ISL_449007 | Quadram Institute Bioscience | COVID-19 Genomics UK (COG-UK) Consortium |
| EPI_ISL_449008 | Quadram Institute Bioscience | COVID-19 Genomics UK (COG-UK) Consortium |
| EPI_ISL_449009 | Quadram Institute Bioscience | COVID-19 Genomics UK (COG-UK) Consortium |
| EPI_ISL_449010 | Quadram Institute Bioscience | COVID-19 Genomics UK (COG-UK) Consortium |
| EPI_ISL_449011 | Quadram Institute Bioscience | COVID-19 Genomics UK (COG-UK) Consortium |
| EPI_ISL_449012 | Quadram Institute Bioscience | COVID-19 Genomics UK (COG-UK) Consortium |
| EPI_ISL_449013 | Quadram Institute Bioscience | COVID-19 Genomics UK (COG-UK) Consortium |
| EPI_ISL_449014 | Quadram Institute Bioscience | COVID-19 Genomics UK (COG-UK) Consortium |
| EPI_ISL_449015 | Quadram Institute Bioscience | COVID-19 Genomics UK (COG-UK) Consortium |
| EPI_ISL_449016 | Quadram Institute Bioscience | COVID-19 Genomics UK (COG-UK) Consortium |
| EPI_ISL_449017 | Quadram Institute Bioscience | COVID-19 Genomics UK (COG-UK) Consortium |
| EPI_ISL_449018 | Quadram Institute Bioscience | COVID-19 Genomics UK (COG-UK) Consortium |
| EPI_ISL_449019 | Quadram Institute Bioscience | COVID-19 Genomics UK (COG-UK) Consortium |
| EPI_ISL_449020 | Quadram Institute Bioscience | COVID-19 Genomics UK (COG-UK) Consortium |
| EPI_ISL_449021 | Quadram Institute Bioscience | COVID-19 Genomics UK (COG-UK) Consortium |
| EPI_ISL_449022 | Quadram Institute Bioscience | COVID-19 Genomics UK (COG-UK) Consortium |
| EPI_ISL_449023 | Quadram Institute Bioscience | COVID-19 Genomics UK (COG-UK) Consortium |
| EPI_ISL_449024 | Quadram Institute Bioscience | COVID-19 Genomics UK (COG-UK) Consortium |
| EPI_ISL_449025 | Quadram Institute Bioscience | COVID-19 Genomics UK (COG-UK) Consortium |
| EPI_ISL_449026 | Quadram Institute Bioscience | COVID-19 Genomics UK (COG-UK) Consortium |
| EPI_ISL_449027 | Quadram Institute Bioscience | COVID-19 Genomics UK (COG-UK) Consortium |
| EPI_ISL_449028 | Quadram Institute Bioscience | COVID-19 Genomics UK (COG-UK) Consortium |
| EPI_ISL_449029 | Quadram Institute Bioscience | COVID-19 Genomics UK (COG-UK) Consortium |

| EPI_Accession_ID | Originating Laboratory | Submitting Laboratory |
|---|---|---|
| EPI_ISL_449030 | Quadram Institute Bioscience | COVID-19 Genomics UK (COG-UK) Consortium |
| EPI_ISL_449031 | Quadram Institute Bioscience | COVID-19 Genomics UK (COG-UK) Consortium |
| EPI_ISL_449032 | Quadram Institute Bioscience | COVID-19 Genomics UK (COG-UK) Consortium |
| EPI_ISL_449033 | Quadram Institute Bioscience | COVID-19 Genomics UK (COG-UK) Consortium |
| EPI_ISL_449034 | Quadram Institute Bioscience | COVID-19 Genomics UK (COG-UK) Consortium |
| EPI_ISL_449035 | Quadram Institute Bioscience | COVID-19 Genomics UK (COG-UK) Consortium |
| EPI_ISL_449036 | Quadram Institute Bioscience | COVID-19 Genomics UK (COG-UK) Consortium |
| EPI_ISL_449037 | Quadram Institute Bioscience | COVID-19 Genomics UK (COG-UK) Consortium |
| EPI_ISL_449038 | Quadram Institute Bioscience | COVID-19 Genomics UK (COG-UK) Consortium |
| EPI_ISL_449039 | Quadram Institute Bioscience | COVID-19 Genomics UK (COG-UK) Consortium |
| EPI_ISL_449040 | Quadram Institute Bioscience | COVID-19 Genomics UK (COG-UK) Consortium |
| EPI_ISL_449041 | Quadram Institute Bioscience | COVID-19 Genomics UK (COG-UK) Consortium |
| EPI_ISL_449042 | Quadram Institute Bioscience | COVID-19 Genomics UK (COG-UK) Consortium |
| EPI_ISL_449043 | Quadram Institute Bioscience | COVID-19 Genomics UK (COG-UK) Consortium |
| EPI_ISL_449044 | Quadram Institute Bioscience | COVID-19 Genomics UK (COG-UK) Consortium |
| EPI_ISL_449045 | Quadram Institute Bioscience | COVID-19 Genomics UK (COG-UK) Consortium |
| EPI_ISL_449046 | Quadram Institute Bioscience | COVID-19 Genomics UK (COG-UK) Consortium |
| EPI_ISL_449047 | Quadram Institute Bioscience | COVID-19 Genomics UK (COG-UK) Consortium |
| EPI_ISL_449048 | Quadram Institute Bioscience | COVID-19 Genomics UK (COG-UK) Consortium |
| EPI_ISL_449049 | Quadram Institute Bioscience | COVID-19 Genomics UK (COG-UK) Consortium |
| EPI_ISL_449050 | Quadram Institute Bioscience | COVID-19 Genomics UK (COG-UK) Consortium |
| EPI_ISL_449051 | Quadram Institute Bioscience | COVID-19 Genomics UK (COG-UK) Consortium |
| EPI_ISL_449052 | Quadram Institute Bioscience | COVID-19 Genomics UK (COG-UK) Consortium |
| EPI_ISL_449053 | Quadram Institute Bioscience | COVID-19 Genomics UK (COG-UK) Consortium |
| EPI_ISL_449054 | Quadram Institute Bioscience | COVID-19 Genomics UK (COG-UK) Consortium |
| EPI_ISL_449055 | Quadram Institute Bioscience | COVID-19 Genomics UK (COG-UK) Consortium |
| EPI_ISL_449056 | Quadram Institute Bioscience | COVID-19 Genomics UK (COG-UK) Consortium |
| EPI_ISL_449057 | Quadram Institute Bioscience | COVID-19 Genomics UK (COG-UK) Consortium |
| EPI_ISL_449058 | Quadram Institute Bioscience | COVID-19 Genomics UK (COG-UK) Consortium |
| EPI_ISL_449059 | Quadram Institute Bioscience | COVID-19 Genomics UK (COG-UK) Consortium |
| EPI_ISL_449060 | Quadram Institute Bioscience | COVID-19 Genomics UK (COG-UK) Consortium |
| EPI_ISL_449061 | Quadram Institute Bioscience | COVID-19 Genomics UK (COG-UK) Consortium |
| EPI_ISL_449062 | Quadram Institute Bioscience | COVID-19 Genomics UK (COG-UK) Consortium |

| EPI_Accession_ID | Originating Laboratory | Submitting Laboratory |
|---|---|---|
| EPI_ISL_449063 | Quadram Institute Bioscience | COVID-19 Genomics UK (COG-UK) Consortium |
| EPI_ISL_449064 | Quadram Institute Bioscience | COVID-19 Genomics UK (COG-UK) Consortium |
| EPI_ISL_449065 | Quadram Institute Bioscience | COVID-19 Genomics UK (COG-UK) Consortium |
| EPI_ISL_449066 | Quadram Institute Bioscience | COVID-19 Genomics UK (COG-UK) Consortium |
| EPI_ISL_449067 | Quadram Institute Bioscience | COVID-19 Genomics UK (COG-UK) Consortium |
| EPI_ISL_449068 | Quadram Institute Bioscience | COVID-19 Genomics UK (COG-UK) Consortium |
| EPI_ISL_449069 | Quadram Institute Bioscience | COVID-19 Genomics UK (COG-UK) Consortium |
| EPI_ISL_449070 | Quadram Institute Bioscience | COVID-19 Genomics UK (COG-UK) Consortium |
| EPI_ISL_449071 | Quadram Institute Bioscience | COVID-19 Genomics UK (COG-UK) Consortium |
| EPI_ISL_449072 | Quadram Institute Bioscience | COVID-19 Genomics UK (COG-UK) Consortium |
| EPI_ISL_449073 | Quadram Institute Bioscience | COVID-19 Genomics UK (COG-UK) Consortium |
| EPI_ISL_449074 | Quadram Institute Bioscience | COVID-19 Genomics UK (COG-UK) Consortium |
| EPI_ISL_449075 | Quadram Institute Bioscience | COVID-19 Genomics UK (COG-UK) Consortium |
| EPI_ISL_449076 | Quadram Institute Bioscience | COVID-19 Genomics UK (COG-UK) Consortium |
| EPI_ISL_449077 | Quadram Institute Bioscience | COVID-19 Genomics UK (COG-UK) Consortium |
| EPI_ISL_449078 | Quadram Institute Bioscience | COVID-19 Genomics UK (COG-UK) Consortium |
| EPI_ISL_449079 | Quadram Institute Bioscience | COVID-19 Genomics UK (COG-UK) Consortium |
| EPI_ISL_449080 | Quadram Institute Bioscience | COVID-19 Genomics UK (COG-UK) Consortium |
| EPI_ISL_449081 | Quadram Institute Bioscience | COVID-19 Genomics UK (COG-UK) Consortium |
| EPI_ISL_449082 | Quadram Institute Bioscience | COVID-19 Genomics UK (COG-UK) Consortium |
| EPI_ISL_449083 | Quadram Institute Bioscience | COVID-19 Genomics UK (COG-UK) Consortium |
| EPI_ISL_449084 | Quadram Institute Bioscience | COVID-19 Genomics UK (COG-UK) Consortium |
| EPI_ISL_449085 | Quadram Institute Bioscience | COVID-19 Genomics UK (COG-UK) Consortium |
| EPI_ISL_449086 | Quadram Institute Bioscience | COVID-19 Genomics UK (COG-UK) Consortium |
| EPI_ISL_449087 | Quadram Institute Bioscience | COVID-19 Genomics UK (COG-UK) Consortium |
| EPI_ISL_449088 | Quadram Institute Bioscience | COVID-19 Genomics UK (COG-UK) Consortium |
| EPI_ISL_449089 | Quadram Institute Bioscience | COVID-19 Genomics UK (COG-UK) Consortium |
| EPI_ISL_449090 | Quadram Institute Bioscience | COVID-19 Genomics UK (COG-UK) Consortium |
| EPI_ISL_449091 | Quadram Institute Bioscience | COVID-19 Genomics UK (COG-UK) Consortium |
| EPI_ISL_449092 | Quadram Institute Bioscience | COVID-19 Genomics UK (COG-UK) Consortium |
| EPI_ISL_449093 | Quadram Institute Bioscience | COVID-19 Genomics UK (COG-UK) Consortium |
| EPI_ISL_449094 | Quadram Institute Bioscience | COVID-19 Genomics UK (COG-UK) Consortium |
| EPI_ISL_449095 | Quadram Institute Bioscience | COVID-19 Genomics UK (COG-UK) Consortium |

| EPI_Accession_ID | Originating Laboratory | Submitting Laboratory |
|---|---|---|
| EPI_ISL_449096 | Quadram Institute Bioscience | COVID-19 Genomics UK (COG-UK) Consortium |
| EPI_ISL_449097 | Quadram Institute Bioscience | COVID-19 Genomics UK (COG-UK) Consortium |
| EPI_ISL_449098 | Quadram Institute Bioscience | COVID-19 Genomics UK (COG-UK) Consortium |
| EPI_ISL_449100 | Quadram Institute Bioscience | COVID-19 Genomics UK (COG-UK) Consortium |
| EPI_ISL_449101 | Quadram Institute Bioscience | COVID-19 Genomics UK (COG-UK) Consortium |
| EPI_ISL_449102 | Quadram Institute Bioscience | COVID-19 Genomics UK (COG-UK) Consortium |
| EPI_ISL_449103 | Quadram Institute Bioscience | COVID-19 Genomics UK (COG-UK) Consortium |
| EPI_ISL_449104 | Quadram Institute Bioscience | COVID-19 Genomics UK (COG-UK) Consortium |
| EPI_ISL_449105 | Quadram Institute Bioscience | COVID-19 Genomics UK (COG-UK) Consortium |
| EPI_ISL_449107 | Quadram Institute Bioscience | COVID-19 Genomics UK (COG-UK) Consortium |
| EPI_ISL_449108 | Quadram Institute Bioscience | COVID-19 Genomics UK (COG-UK) Consortium |
| EPI_ISL_449109 | Quadram Institute Bioscience | COVID-19 Genomics UK (COG-UK) Consortium |
| EPI_ISL_449111 | Quadram Institute Bioscience | COVID-19 Genomics UK (COG-UK) Consortium |
| EPI_ISL_449112 | Quadram Institute Bioscience | COVID-19 Genomics UK (COG-UK) Consortium |
| EPI_ISL_449113 | Quadram Institute Bioscience | COVID-19 Genomics UK (COG-UK) Consortium |
| EPI_ISL_449114 | Quadram Institute Bioscience | COVID-19 Genomics UK (COG-UK) Consortium |
| EPI_ISL_449116 | Quadram Institute Bioscience | COVID-19 Genomics UK (COG-UK) Consortium |
| EPI_ISL_449117 | Quadram Institute Bioscience | COVID-19 Genomics UK (COG-UK) Consortium |
| EPI_ISL_449118 | Quadram Institute Bioscience | COVID-19 Genomics UK (COG-UK) Consortium |
| EPI_ISL_449119 | Quadram Institute Bioscience | COVID-19 Genomics UK (COG-UK) Consortium |
| EPI_ISL_449120 | Quadram Institute Bioscience | COVID-19 Genomics UK (COG-UK) Consortium |
| EPI_ISL_449121 | Quadram Institute Bioscience | COVID-19 Genomics UK (COG-UK) Consortium |
| EPI_ISL_449122 | Quadram Institute Bioscience | COVID-19 Genomics UK (COG-UK) Consortium |
| EPI_ISL_449123 | Quadram Institute Bioscience | COVID-19 Genomics UK (COG-UK) Consortium |
| EPI_ISL_449124 | Quadram Institute Bioscience | COVID-19 Genomics UK (COG-UK) Consortium |
| EPI_ISL_449125 | Quadram Institute Bioscience | COVID-19 Genomics UK (COG-UK) Consortium |
| EPI_ISL_449127 | Quadram Institute Bioscience | COVID-19 Genomics UK (COG-UK) Consortium |
| EPI_ISL_449128 | Quadram Institute Bioscience | COVID-19 Genomics UK (COG-UK) Consortium |
| EPI_ISL_449129 | Quadram Institute Bioscience | COVID-19 Genomics UK (COG-UK) Consortium |
| EPI_ISL_449130 | Quadram Institute Bioscience | COVID-19 Genomics UK (COG-UK) Consortium |
| EPI_ISL_449131 | Quadram Institute Bioscience | COVID-19 Genomics UK (COG-UK) Consortium |
| EPI_ISL_449132 | Quadram Institute Bioscience | COVID-19 Genomics UK (COG-UK) Consortium |
| EPI_ISL_449133 | Quadram Institute Bioscience | COVID-19 Genomics UK (COG-UK) Consortium |

| EPI_Accession_ID | Originating Laboratory | Submitting Laboratory |
|---|---|---|
| EPI_ISL_449135 | Quadram Institute Bioscience | COVID-19 Genomics UK (COG-UK) Consortium |
| EPI_ISL_449136 | Quadram Institute Bioscience | COVID-19 Genomics UK (COG-UK) Consortium |
| EPI_ISL_449139 | Quadram Institute Bioscience | COVID-19 Genomics UK (COG-UK) Consortium |
| EPI_ISL_449140 | Quadram Institute Bioscience | COVID-19 Genomics UK (COG-UK) Consortium |
| EPI_ISL_449141 | Quadram Institute Bioscience | COVID-19 Genomics UK (COG-UK) Consortium |
| EPI_ISL_449143 | Quadram Institute Bioscience | COVID-19 Genomics UK (COG-UK) Consortium |
| EPI_ISL_449145 | Quadram Institute Bioscience | COVID-19 Genomics UK (COG-UK) Consortium |
| EPI_ISL_449146 | Quadram Institute Bioscience | COVID-19 Genomics UK (COG-UK) Consortium |
| EPI_ISL_449147 | Quadram Institute Bioscience | COVID-19 Genomics UK (COG-UK) Consortium |
| EPI_ISL_449148 | Quadram Institute Bioscience | COVID-19 Genomics UK (COG-UK) Consortium |
| EPI_ISL_449149 | Quadram Institute Bioscience | COVID-19 Genomics UK (COG-UK) Consortium |
| EPI_ISL_449150 | Quadram Institute Bioscience | COVID-19 Genomics UK (COG-UK) Consortium |
| EPI_ISL_449151 | Quadram Institute Bioscience | COVID-19 Genomics UK (COG-UK) Consortium |
| EPI_ISL_449152 | Quadram Institute Bioscience | COVID-19 Genomics UK (COG-UK) Consortium |
| EPI_ISL_449153 | Quadram Institute Bioscience | COVID-19 Genomics UK (COG-UK) Consortium |
| EPI_ISL_449154 | Quadram Institute Bioscience | COVID-19 Genomics UK (COG-UK) Consortium |
| EPI_ISL_449155 | Quadram Institute Bioscience | COVID-19 Genomics UK (COG-UK) Consortium |
| EPI_ISL_449156 | Quadram Institute Bioscience | COVID-19 Genomics UK (COG-UK) Consortium |
| EPI_ISL_449157 | Quadram Institute Bioscience | COVID-19 Genomics UK (COG-UK) Consortium |
| EPI_ISL_449158 | Quadram Institute Bioscience | COVID-19 Genomics UK (COG-UK) Consortium |
| EPI_ISL_449159 | Quadram Institute Bioscience | COVID-19 Genomics UK (COG-UK) Consortium |
| EPI_ISL_449160 | Quadram Institute Bioscience | COVID-19 Genomics UK (COG-UK) Consortium |
| EPI_ISL_449161 | Quadram Institute Bioscience | COVID-19 Genomics UK (COG-UK) Consortium |
| EPI_ISL_449162 | Quadram Institute Bioscience | COVID-19 Genomics UK (COG-UK) Consortium |
| EPI_ISL_449163 | Quadram Institute Bioscience | COVID-19 Genomics UK (COG-UK) Consortium |
| EPI_ISL_449164 | Quadram Institute Bioscience | COVID-19 Genomics UK (COG-UK) Consortium |
| EPI_ISL_449165 | Quadram Institute Bioscience | COVID-19 Genomics UK (COG-UK) Consortium |
| EPI_ISL_449166 | Quadram Institute Bioscience | COVID-19 Genomics UK (COG-UK) Consortium |
| EPI_ISL_449167 | Quadram Institute Bioscience | COVID-19 Genomics UK (COG-UK) Consortium |
| EPI_ISL_449168 | Quadram Institute Bioscience | COVID-19 Genomics UK (COG-UK) Consortium |
| EPI_ISL_449169 | Quadram Institute Bioscience | COVID-19 Genomics UK (COG-UK) Consortium |
| EPI_ISL_449170 | Quadram Institute Bioscience | COVID-19 Genomics UK (COG-UK) Consortium |
| EPI_ISL_449171 | Quadram Institute Bioscience | COVID-19 Genomics UK (COG-UK) Consortium |

| EPI_Accession_ID | Originating Laboratory | Submitting Laboratory |
|---|---|---|
| EPI_ISL_449172 | Quadram Institute Bioscience | COVID-19 Genomics UK (COG-UK) Consortium |
| EPI_ISL_449173 | Quadram Institute Bioscience | COVID-19 Genomics UK (COG-UK) Consortium |
| EPI_ISL_449174 | Quadram Institute Bioscience | COVID-19 Genomics UK (COG-UK) Consortium |
| EPI_ISL_449175 | Quadram Institute Bioscience | COVID-19 Genomics UK (COG-UK) Consortium |
| EPI_ISL_449176 | West of Scotland Specialist Virology Centre, NHSGGC / MRC-University of Glasgow Centre for Virus Research | COVID-19 Genomics UK (COG-UK) Consortium |
| EPI_ISL_449177 | West of Scotland Specialist Virology Centre, NHSGGC / MRC-University of Glasgow Centre for Virus Research | COVID-19 Genomics UK (COG-UK) Consortium |
| EPI_ISL_449178 | West of Scotland Specialist Virology Centre, NHSGGC / MRC-University of Glasgow Centre for Virus Research | COVID-19 Genomics UK (COG-UK) Consortium |
| EPI_ISL_449179 | West of Scotland Specialist Virology Centre, NHSGGC / MRC-University of Glasgow Centre for Virus Research | COVID-19 Genomics UK (COG-UK) Consortium |
| EPI_ISL_449180 | West of Scotland Specialist Virology Centre, NHSGGC / MRC-University of Glasgow Centre for Virus Research | COVID-19 Genomics UK (COG-UK) Consortium |
| EPI_ISL_449181 | West of Scotland Specialist Virology Centre, NHSGGC / MRC-University of Glasgow Centre for Virus Research | COVID-19 Genomics UK (COG-UK) Consortium |
| EPI_ISL_449182 | West of Scotland Specialist Virology Centre, NHSGGC / MRC-University of Glasgow Centre for Virus Research | COVID-19 Genomics UK (COG-UK) Consortium |
| EPI_ISL_449183 | West of Scotland Specialist Virology Centre, NHSGGC / MRC-University of Glasgow Centre for Virus Research | COVID-19 Genomics UK (COG-UK) Consortium |
| EPI_ISL_449184 | West of Scotland Specialist Virology Centre, NHSGGC / MRC-University of Glasgow Centre for Virus Research | COVID-19 Genomics UK (COG-UK) Consortium |
| EPI_ISL_449185 | West of Scotland Specialist Virology Centre, NHSGGC / MRC-University of Glasgow Centre for Virus Research | COVID-19 Genomics UK (COG-UK) Consortium |
| EPI_ISL_449186 | West of Scotland Specialist Virology Centre, NHSGGC / MRC-University of Glasgow Centre for Virus Research | COVID-19 Genomics UK (COG-UK) Consortium |
| EPI_ISL_449187 | West of Scotland Specialist Virology Centre, NHSGGC / MRC-University of Glasgow Centre for Virus Research | COVID-19 Genomics UK (COG-UK) Consortium |
| EPI_ISL_449188 | West of Scotland Specialist Virology Centre, NHSGGC / MRC-University of Glasgow Centre for Virus Research | COVID-19 Genomics UK (COG-UK) Consortium |
| EPI_ISL_449189 | West of Scotland Specialist Virology Centre, NHSGGC / MRC-University of Glasgow Centre for Virus Research | COVID-19 Genomics UK (COG-UK) Consortium |

| EPI_Accession_ID | Originating Laboratory | Submitting Laboratory |
|---|---|---|
| EPI_ISL_449190 | West of Scotland Specialist Virology Centre, NHSGGC / MRC-University of Glasgow Centre for Virus Research | COVID-19 Genomics UK (COG-UK) Consortium |
| EPI_ISL_449191 | West of Scotland Specialist Virology Centre, NHSGGC / MRC-University of Glasgow Centre for Virus Research | COVID-19 Genomics UK (COG-UK) Consortium |
| EPI_ISL_449192 | West of Scotland Specialist Virology Centre, NHSGGC / MRC-University of Glasgow Centre for Virus Research | COVID-19 Genomics UK (COG-UK) Consortium |
| EPI_ISL_449193 | West of Scotland Specialist Virology Centre, NHSGGC / MRC-University of Glasgow Centre for Virus Research | COVID-19 Genomics UK (COG-UK) Consortium |
| EPI_ISL_449194 | West of Scotland Specialist Virology Centre, NHSGGC / MRC-University of Glasgow Centre for Virus Research | COVID-19 Genomics UK (COG-UK) Consortium |
| EPI_ISL_449195 | West of Scotland Specialist Virology Centre, NHSGGC / MRC-University of Glasgow Centre for Virus Research | COVID-19 Genomics UK (COG-UK) Consortium |
| EPI_ISL_449196 | West of Scotland Specialist Virology Centre, NHSGGC / MRC-University of Glasgow Centre for Virus Research | COVID-19 Genomics UK (COG-UK) Consortium |
| EPI_ISL_449197 | West of Scotland Specialist Virology Centre, NHSGGC / MRC-University of Glasgow Centre for Virus Research | COVID-19 Genomics UK (COG-UK) Consortium |
| EPI_ISL_449198 | West of Scotland Specialist Virology Centre, NHSGGC / MRC-University of Glasgow Centre for Virus Research | COVID-19 Genomics UK (COG-UK) Consortium |
| EPI_ISL_449199 | West of Scotland Specialist Virology Centre, NHSGGC / MRC-University of Glasgow Centre for Virus Research | COVID-19 Genomics UK (COG-UK) Consortium |
| EPI_ISL_449200 | West of Scotland Specialist Virology Centre, NHSGGC / MRC-University of Glasgow Centre for Virus Research | COVID-19 Genomics UK (COG-UK) Consortium |
| EPI_ISL_449201 | West of Scotland Specialist Virology Centre, NHSGGC / MRC-University of Glasgow Centre for Virus Research | COVID-19 Genomics UK (COG-UK) Consortium |
| EPI_ISL_449202 | West of Scotland Specialist Virology Centre, NHSGGC / MRC-University of Glasgow Centre for Virus Research | COVID-19 Genomics UK (COG-UK) Consortium |
| EPI_ISL_449203 | West of Scotland Specialist Virology Centre, NHSGGC / MRC-University of Glasgow Centre for Virus Research | COVID-19 Genomics UK (COG-UK) Consortium |
| EPI_ISL_449204 | West of Scotland Specialist Virology Centre, NHSGGC / MRC-University of Glasgow Centre for Virus Research | COVID-19 Genomics UK (COG-UK) Consortium |
| EPI_ISL_449205 | West of Scotland Specialist Virology Centre, NHSGGC / MRC-University of Glasgow Centre for Virus Research | COVID-19 Genomics UK (COG-UK) Consortium |

| EPI_Accession_ID | Originating Laboratory | Submitting Laboratory |
|---|---|---|
| EPI_ISL_449206 | West of Scotland Specialist Virology Centre, NHSGGC / MRC-University of Glasgow Centre for Virus Research | COVID-19 Genomics UK (COG-UK) Consortium |
| EPI_ISL_449207 | West of Scotland Specialist Virology Centre, NHSGGC / MRC-University of Glasgow Centre for Virus Research | COVID-19 Genomics UK (COG-UK) Consortium |
| EPI_ISL_449208 | West of Scotland Specialist Virology Centre, NHSGGC / MRC-University of Glasgow Centre for Virus Research | COVID-19 Genomics UK (COG-UK) Consortium |
| EPI_ISL_449209 | West of Scotland Specialist Virology Centre, NHSGGC / MRC-University of Glasgow Centre for Virus Research | COVID-19 Genomics UK (COG-UK) Consortium |
| EPI_ISL_449210 | West of Scotland Specialist Virology Centre, NHSGGC / MRC-University of Glasgow Centre for Virus Research | COVID-19 Genomics UK (COG-UK) Consortium |
| EPI_ISL_449211 | West of Scotland Specialist Virology Centre, NHSGGC / MRC-University of Glasgow Centre for Virus Research | COVID-19 Genomics UK (COG-UK) Consortium |
| EPI_ISL_449212 | West of Scotland Specialist Virology Centre, NHSGGC / MRC-University of Glasgow Centre for Virus Research | COVID-19 Genomics UK (COG-UK) Consortium |
| EPI_ISL_449213 | West of Scotland Specialist Virology Centre, NHSGGC / MRC-University of Glasgow Centre for Virus Research | COVID-19 Genomics UK (COG-UK) Consortium |
| EPI_ISL_449214 | West of Scotland Specialist Virology Centre, NHSGGC / MRC-University of Glasgow Centre for Virus Research | COVID-19 Genomics UK (COG-UK) Consortium |
| EPI_ISL_449215 | West of Scotland Specialist Virology Centre, NHSGGC / MRC-University of Glasgow Centre for Virus Research | COVID-19 Genomics UK (COG-UK) Consortium |
| EPI_ISL_449216 | West of Scotland Specialist Virology Centre, NHSGGC / MRC-University of Glasgow Centre for Virus Research | COVID-19 Genomics UK (COG-UK) Consortium |
| EPI_ISL_449217 | West of Scotland Specialist Virology Centre, NHSGGC / MRC-University of Glasgow Centre for Virus Research | COVID-19 Genomics UK (COG-UK) Consortium |
| EPI_ISL_449218 | West of Scotland Specialist Virology Centre, NHSGGC / MRC-University of Glasgow Centre for Virus Research | COVID-19 Genomics UK (COG-UK) Consortium |
| EPI_ISL_449219 | West of Scotland Specialist Virology Centre, NHSGGC / MRC-University of Glasgow Centre for Virus Research | COVID-19 Genomics UK (COG-UK) Consortium |
| EPI_ISL_449220 | West of Scotland Specialist Virology Centre, NHSGGC / MRC-University of Glasgow Centre for Virus Research | COVID-19 Genomics UK (COG-UK) Consortium |
| EPI_ISL_449221 | West of Scotland Specialist Virology Centre, NHSGGC / MRC-University of Glasgow Centre for Virus Research | COVID-19 Genomics UK (COG-UK) Consortium |

| EPI_Accession_ID | Originating Laboratory | Submitting Laboratory |
|---|---|---|
| EPI_ISL_449222 | West of Scotland Specialist Virology Centre, NHSGGC / MRC-University of Glasgow Centre for Virus Research | COVID-19 Genomics UK (COG-UK) Consortium |
| EPI_ISL_449223 | West of Scotland Specialist Virology Centre, NHSGGC / MRC-University of Glasgow Centre for Virus Research | COVID-19 Genomics UK (COG-UK) Consortium |
| EPI_ISL_449224 | West of Scotland Specialist Virology Centre, NHSGGC / MRC-University of Glasgow Centre for Virus Research | COVID-19 Genomics UK (COG-UK) Consortium |
| EPI_ISL_449225 | West of Scotland Specialist Virology Centre, NHSGGC / MRC-University of Glasgow Centre for Virus Research | COVID-19 Genomics UK (COG-UK) Consortium |
| EPI_ISL_449226 | West of Scotland Specialist Virology Centre, NHSGGC / MRC-University of Glasgow Centre for Virus Research | COVID-19 Genomics UK (COG-UK) Consortium |
| EPI_ISL_449227 | West of Scotland Specialist Virology Centre, NHSGGC / MRC-University of Glasgow Centre for Virus Research | COVID-19 Genomics UK (COG-UK) Consortium |
| EPI_ISL_449228 | West of Scotland Specialist Virology Centre, NHSGGC / MRC-University of Glasgow Centre for Virus Research | COVID-19 Genomics UK (COG-UK) Consortium |
| EPI_ISL_449229 | West of Scotland Specialist Virology Centre, NHSGGC / MRC-University of Glasgow Centre for Virus Research | COVID-19 Genomics UK (COG-UK) Consortium |
| EPI_ISL_449230 | West of Scotland Specialist Virology Centre, NHSGGC / MRC-University of Glasgow Centre for Virus Research | COVID-19 Genomics UK (COG-UK) Consortium |
| EPI_ISL_449231 | West of Scotland Specialist Virology Centre, NHSGGC / MRC-University of Glasgow Centre for Virus Research | COVID-19 Genomics UK (COG-UK) Consortium |
| EPI_ISL_449232 | West of Scotland Specialist Virology Centre, NHSGGC / MRC-University of Glasgow Centre for Virus Research | COVID-19 Genomics UK (COG-UK) Consortium |
| EPI_ISL_449233 | West of Scotland Specialist Virology Centre, NHSGGC / MRC-University of Glasgow Centre for Virus Research | COVID-19 Genomics UK (COG-UK) Consortium |
| EPI_ISL_449234 | West of Scotland Specialist Virology Centre, NHSGGC / MRC-University of Glasgow Centre for Virus Research | COVID-19 Genomics UK (COG-UK) Consortium |
| EPI_ISL_449235 | West of Scotland Specialist Virology Centre, NHSGGC / MRC-University of Glasgow Centre for Virus Research | COVID-19 Genomics UK (COG-UK) Consortium |
| EPI_ISL_449236 | West of Scotland Specialist Virology Centre, NHSGGC / MRC-University of Glasgow Centre for Virus Research | COVID-19 Genomics UK (COG-UK) Consortium |
| EPI_ISL_449237 | West of Scotland Specialist Virology Centre, NHSGGC / MRC-University of Glasgow Centre for Virus Research | COVID-19 Genomics UK (COG-UK) Consortium |

| EPI_Accession_ID | Originating Laboratory | Submitting Laboratory |
|---|---|---|
| EPI_ISL_449238 | West of Scotland Specialist Virology Centre, NHSGGC / MRC-University of Glasgow Centre for Virus Research | COVID-19 Genomics UK (COG-UK) Consortium |
| EPI_ISL_449239 | West of Scotland Specialist Virology Centre, NHSGGC / MRC-University of Glasgow Centre for Virus Research | COVID-19 Genomics UK (COG-UK) Consortium |
| EPI_ISL_449240 | West of Scotland Specialist Virology Centre, NHSGGC / MRC-University of Glasgow Centre for Virus Research | COVID-19 Genomics UK (COG-UK) Consortium |
| EPI_ISL_449241 | West of Scotland Specialist Virology Centre, NHSGGC / MRC-University of Glasgow Centre for Virus Research | COVID-19 Genomics UK (COG-UK) Consortium |
| EPI_ISL_449242 | West of Scotland Specialist Virology Centre, NHSGGC / MRC-University of Glasgow Centre for Virus Research | COVID-19 Genomics UK (COG-UK) Consortium |
| EPI_ISL_449243 | West of Scotland Specialist Virology Centre, NHSGGC / MRC-University of Glasgow Centre for Virus Research | COVID-19 Genomics UK (COG-UK) Consortium |
| EPI_ISL_449244 | West of Scotland Specialist Virology Centre, NHSGGC / MRC-University of Glasgow Centre for Virus Research | COVID-19 Genomics UK (COG-UK) Consortium |
| EPI_ISL_449245 | West of Scotland Specialist Virology Centre, NHSGGC / MRC-University of Glasgow Centre for Virus Research | COVID-19 Genomics UK (COG-UK) Consortium |
| EPI_ISL_449246 | West of Scotland Specialist Virology Centre, NHSGGC / MRC-University of Glasgow Centre for Virus Research | COVID-19 Genomics UK (COG-UK) Consortium |
| EPI_ISL_449247 | West of Scotland Specialist Virology Centre, NHSGGC / MRC-University of Glasgow Centre for Virus Research | COVID-19 Genomics UK (COG-UK) Consortium |
| EPI_ISL_449248 | West of Scotland Specialist Virology Centre, NHSGGC / MRC-University of Glasgow Centre for Virus Research | COVID-19 Genomics UK (COG-UK) Consortium |
| EPI_ISL_449249 | West of Scotland Specialist Virology Centre, NHSGGC / MRC-University of Glasgow Centre for Virus Research | COVID-19 Genomics UK (COG-UK) Consortium |
| EPI_ISL_449250 | West of Scotland Specialist Virology Centre, NHSGGC / MRC-University of Glasgow Centre for Virus Research | COVID-19 Genomics UK (COG-UK) Consortium |
| EPI_ISL_449251 | West of Scotland Specialist Virology Centre, NHSGGC / MRC-University of Glasgow Centre for Virus Research | COVID-19 Genomics UK (COG-UK) Consortium |
| EPI_ISL_449252 | West of Scotland Specialist Virology Centre, NHSGGC / MRC-University of Glasgow Centre for Virus Research | COVID-19 Genomics UK (COG-UK) Consortium |
| EPI_ISL_449253 | West of Scotland Specialist Virology Centre, NHSGGC / MRC-University of Glasgow Centre for Virus Research | COVID-19 Genomics UK (COG-UK) Consortium |

| EPI_Accession_ID | Originating Laboratory | Submitting Laboratory |
|---|---|---|
| EPI_ISL_449254 | West of Scotland Specialist Virology Centre, NHSGGC / MRC-University of Glasgow Centre for Virus Research | COVID-19 Genomics UK (COG-UK) Consortium |
| EPI_ISL_449255 | West of Scotland Specialist Virology Centre, NHSGGC / MRC-University of Glasgow Centre for Virus Research | COVID-19 Genomics UK (COG-UK) Consortium |
| EPI_ISL_449256 | West of Scotland Specialist Virology Centre, NHSGGC / MRC-University of Glasgow Centre for Virus Research | COVID-19 Genomics UK (COG-UK) Consortium |
| EPI_ISL_449257 | West of Scotland Specialist Virology Centre, NHSGGC / MRC-University of Glasgow Centre for Virus Research | COVID-19 Genomics UK (COG-UK) Consortium |
| EPI_ISL_449258 | West of Scotland Specialist Virology Centre, NHSGGC / MRC-University of Glasgow Centre for Virus Research | COVID-19 Genomics UK (COG-UK) Consortium |
| EPI_ISL_449259 | Virology Department, Royal Infirmary of Edinburgh, NHS Lothian / School of Biological Sciences, University of Edinburgh / Institute of Genetics and Molecular Medicine, University of Edinburgh | COVID-19 Genomics UK (COG-UK) Consortium |
| EPI_ISL_449260 | Virology Department, Royal Infirmary of Edinburgh, NHS Lothian / School of Biological Sciences, University of Edinburgh / Institute of Genetics and Molecular Medicine, University of Edinburgh | COVID-19 Genomics UK (COG-UK) Consortium |
| EPI_ISL_449261 | Virology Department, Royal Infirmary of Edinburgh, NHS Lothian / School of Biological Sciences, University of Edinburgh / Institute of Genetics and Molecular Medicine, University of Edinburgh | COVID-19 Genomics UK (COG-UK) Consortium |
| EPI_ISL_449262 | Virology Department, Royal Infirmary of Edinburgh, NHS Lothian / School of Biological Sciences, University of Edinburgh / Institute of Genetics and Molecular Medicine, University of Edinburgh | COVID-19 Genomics UK (COG-UK) Consortium |
| EPI_ISL_449263 | Virology Department, Royal Infirmary of Edinburgh, NHS Lothian / School of Biological Sciences, University of Edinburgh / Institute of Genetics and Molecular Medicine, University of Edinburgh | COVID-19 Genomics UK (COG-UK) Consortium |

| EPI_Accession_ID | Originating Laboratory | Submitting Laboratory |
|---|---|---|
| EPI_ISL_449264 | Virology Department, Royal Infirmary of Edinburgh, NHS Lothian / School of Biological Sciences, University of Edinburgh / Institute of Genetics and Molecular Medicine, University of Edinburgh | COVID-19 Genomics UK (COG-UK) Consortium |
| EPI_ISL_449265 | Virology Department, Royal Infirmary of Edinburgh, NHS Lothian / School of Biological Sciences, University of Edinburgh / Institute of Genetics and Molecular Medicine, University of Edinburgh | COVID-19 Genomics UK (COG-UK) Consortium |
| EPI_ISL_449266 | Virology Department, Royal Infirmary of Edinburgh, NHS Lothian / School of Biological Sciences, University of Edinburgh / Institute of Genetics and Molecular Medicine, University of Edinburgh | COVID-19 Genomics UK (COG-UK) Consortium |
| EPI_ISL_449267 | Virology Department, Royal Infirmary of Edinburgh, NHS Lothian / School of Biological Sciences, University of Edinburgh / Institute of Genetics and Molecular Medicine, University of Edinburgh | COVID-19 Genomics UK (COG-UK) Consortium |
| EPI_ISL_449268 | Virology Department, Royal Infirmary of Edinburgh, NHS Lothian / School of Biological Sciences, University of Edinburgh / Institute of Genetics and Molecular Medicine, University of Edinburgh | COVID-19 Genomics UK (COG-UK) Consortium |
| EPI_ISL_449269 | Virology Department, Royal Infirmary of Edinburgh, NHS Lothian / School of Biological Sciences, University of Edinburgh / Institute of Genetics and Molecular Medicine, University of Edinburgh | COVID-19 Genomics UK (COG-UK) Consortium |
| EPI_ISL_449270 | Virology Department, Royal Infirmary of Edinburgh, NHS Lothian / School of Biological Sciences, University of Edinburgh / Institute of Genetics and Molecular Medicine, University of Edinburgh | COVID-19 Genomics UK (COG-UK) Consortium |
| EPI_ISL_449271 | Virology Department, Royal Infirmary of Edinburgh, NHS Lothian / School of Biological Sciences, University of Edinburgh / Institute of Genetics and Molecular Medicine, University of Edinburgh | COVID-19 Genomics UK (COG-UK) Consortium |

| EPI_Accession_ID | Originating Laboratory | Submitting Laboratory |
|---|---|---|
| EPI_ISL_449272 | Virology Department, Royal Infirmary of Edinburgh, NHS Lothian / School of Biological Sciences, University of Edinburgh / Institute of Genetics and Molecular Medicine, University of Edinburgh | COVID-19 Genomics UK (COG-UK) Consortium |
| EPI_ISL_449273 | Virology Department, Royal Infirmary of Edinburgh, NHS Lothian / School of Biological Sciences, University of Edinburgh / Institute of Genetics and Molecular Medicine, University of Edinburgh | COVID-19 Genomics UK (COG-UK) Consortium |
| EPI_ISL_449274 | Virology Department, Royal Infirmary of Edinburgh, NHS Lothian / School of Biological Sciences, University of Edinburgh / Institute of Genetics and Molecular Medicine, University of Edinburgh | COVID-19 Genomics UK (COG-UK) Consortium |
| EPI_ISL_449275 | Virology Department, Royal Infirmary of Edinburgh, NHS Lothian / School of Biological Sciences, University of Edinburgh / Institute of Genetics and Molecular Medicine, University of Edinburgh | COVID-19 Genomics UK (COG-UK) Consortium |
| EPI_ISL_449276 | Virology Department, Royal Infirmary of Edinburgh, NHS Lothian / School of Biological Sciences, University of Edinburgh / Institute of Genetics and Molecular Medicine, University of Edinburgh | COVID-19 Genomics UK (COG-UK) Consortium |
| EPI_ISL_449277 | Virology Department, Royal Infirmary of Edinburgh, NHS Lothian / School of Biological Sciences, University of Edinburgh / Institute of Genetics and Molecular Medicine, University of Edinburgh | COVID-19 Genomics UK (COG-UK) Consortium |
| EPI_ISL_449278 | Virology Department, Royal Infirmary of Edinburgh, NHS Lothian / School of Biological Sciences, University of Edinburgh / Institute of Genetics and Molecular Medicine, University of Edinburgh | COVID-19 Genomics UK (COG-UK) Consortium |
| EPI_ISL_449279 | Virology Department, Royal Infirmary of Edinburgh, NHS Lothian / School of Biological Sciences, University of Edinburgh / Institute of Genetics and Molecular Medicine, University of Edinburgh | COVID-19 Genomics UK (COG-UK) Consortium |

| EPI_Accession_ID | Originating Laboratory | Submitting Laboratory |
|---|---|---|
| EPI_ISL_449280 | Virology Department, Royal Infirmary of Edinburgh, NHS Lothian / School of Biological Sciences, University of Edinburgh / Institute of Genetics and Molecular Medicine, University of Edinburgh | COVID-19 Genomics UK (COG-UK) Consortium |
| EPI_ISL_449281 | Virology Department, Royal Infirmary of Edinburgh, NHS Lothian / School of Biological Sciences, University of Edinburgh / Institute of Genetics and Molecular Medicine, University of Edinburgh | COVID-19 Genomics UK (COG-UK) Consortium |
| EPI_ISL_449282 | Virology Department, Royal Infirmary of Edinburgh, NHS Lothian / School of Biological Sciences, University of Edinburgh / Institute of Genetics and Molecular Medicine, University of Edinburgh | COVID-19 Genomics UK (COG-UK) Consortium |
| EPI_ISL_449283 | Virology Department, Royal Infirmary of Edinburgh, NHS Lothian / School of Biological Sciences, University of Edinburgh / Institute of Genetics and Molecular Medicine, University of Edinburgh | COVID-19 Genomics UK (COG-UK) Consortium |
| EPI_ISL_449284 | Virology Department, Royal Infirmary of Edinburgh, NHS Lothian / School of Biological Sciences, University of Edinburgh / Institute of Genetics and Molecular Medicine, University of Edinburgh | COVID-19 Genomics UK (COG-UK) Consortium |
| EPI_ISL_449285 | Virology Department, Royal Infirmary of Edinburgh, NHS Lothian / School of Biological Sciences, University of Edinburgh / Institute of Genetics and Molecular Medicine, University of Edinburgh | COVID-19 Genomics UK (COG-UK) Consortium |
| EPI_ISL_449286 | Virology Department, Royal Infirmary of Edinburgh, NHS Lothian / School of Biological Sciences, University of Edinburgh / Institute of Genetics and Molecular Medicine, University of Edinburgh | COVID-19 Genomics UK (COG-UK) Consortium |
| EPI_ISL_449287 | Virology Department, Royal Infirmary of Edinburgh, NHS Lothian / School of Biological Sciences, University of Edinburgh / Institute of Genetics and Molecular Medicine, University of Edinburgh | COVID-19 Genomics UK (COG-UK) Consortium |

| EPI_Accession_ID | Originating Laboratory | Submitting Laboratory |
|---|---|---|
| EPI_ISL_449288 | Virology Department, Royal Infirmary of Edinburgh, NHS Lothian / School of Biological Sciences, University of Edinburgh / Institute of Genetics and Molecular Medicine, University of Edinburgh | COVID-19 Genomics UK (COG-UK) Consortium |
| EPI_ISL_449289 | Virology Department, Royal Infirmary of Edinburgh, NHS Lothian / School of Biological Sciences, University of Edinburgh / Institute of Genetics and Molecular Medicine, University of Edinburgh | COVID-19 Genomics UK (COG-UK) Consortium |
| EPI_ISL_449290 | Virology Department, Royal Infirmary of Edinburgh, NHS Lothian / School of Biological Sciences, University of Edinburgh / Institute of Genetics and Molecular Medicine, University of Edinburgh | COVID-19 Genomics UK (COG-UK) Consortium |
| EPI_ISL_449291 | Virology Department, Royal Infirmary of Edinburgh, NHS Lothian / School of Biological Sciences, University of Edinburgh / Institute of Genetics and Molecular Medicine, University of Edinburgh | COVID-19 Genomics UK (COG-UK) Consortium |
| EPI_ISL_449292 | Virology Department, Royal Infirmary of Edinburgh, NHS Lothian / School of Biological Sciences, University of Edinburgh / Institute of Genetics and Molecular Medicine, University of Edinburgh | COVID-19 Genomics UK (COG-UK) Consortium |
| EPI_ISL_449293 | Virology Department, Royal Infirmary of Edinburgh, NHS Lothian / School of Biological Sciences, University of Edinburgh / Institute of Genetics and Molecular Medicine, University of Edinburgh | COVID-19 Genomics UK (COG-UK) Consortium |
| EPI_ISL_449294 | Virology Department, Royal Infirmary of Edinburgh, NHS Lothian / School of Biological Sciences, University of Edinburgh / Institute of Genetics and Molecular Medicine, University of Edinburgh | COVID-19 Genomics UK (COG-UK) Consortium |
| EPI_ISL_449295 | Virology Department, Royal Infirmary of Edinburgh, NHS Lothian / School of Biological Sciences, University of Edinburgh / Institute of Genetics and Molecular Medicine, University of Edinburgh | COVID-19 Genomics UK (COG-UK) Consortium |

| EPI_Accession_ID | Originating Laboratory | Submitting Laboratory |
| --- | --- | --- |
| EPI_ISL_449296 | Virology Department, Royal Infirmary of Edinburgh, NHS Lothian / School of Biological Sciences, University of Edinburgh / Institute of Genetics and Molecular Medicine, University of Edinburgh | COVID-19 Genomics UK (COG-UK) Consortium |
| EPI_ISL_449297 | Virology Department, Royal Infirmary of Edinburgh, NHS Lothian / School of Biological Sciences, University of Edinburgh / Institute of Genetics and Molecular Medicine, University of Edinburgh | COVID-19 Genomics UK (COG-UK) Consortium |
| EPI_ISL_449298 | Virology Department, Royal Infirmary of Edinburgh, NHS Lothian / School of Biological Sciences, University of Edinburgh / Institute of Genetics and Molecular Medicine, University of Edinburgh | COVID-19 Genomics UK (COG-UK) Consortium |
| EPI_ISL_449299 | Virology Department, Royal Infirmary of Edinburgh, NHS Lothian / School of Biological Sciences, University of Edinburgh / Institute of Genetics and Molecular Medicine, University of Edinburgh | COVID-19 Genomics UK (COG-UK) Consortium |
| EPI_ISL_449300 | Virology Department, Royal Infirmary of Edinburgh, NHS Lothian / School of Biological Sciences, University of Edinburgh / Institute of Genetics and Molecular Medicine, University of Edinburgh | COVID-19 Genomics UK (COG-UK) Consortium |
| EPI_ISL_449301 | Virology Department, Royal Infirmary of Edinburgh, NHS Lothian / School of Biological Sciences, University of Edinburgh / Institute of Genetics and Molecular Medicine, University of Edinburgh | COVID-19 Genomics UK (COG-UK) Consortium |
| EPI_ISL_449302 | Virology Department, Royal Infirmary of Edinburgh, NHS Lothian / School of Biological Sciences, University of Edinburgh / Institute of Genetics and Molecular Medicine, University of Edinburgh | COVID-19 Genomics UK (COG-UK) Consortium |
| EPI_ISL_449303 | Virology Department, Royal Infirmary of Edinburgh, NHS Lothian / School of Biological Sciences, University of Edinburgh / Institute of Genetics and Molecular Medicine, University of Edinburgh | COVID-19 Genomics UK (COG-UK) Consortium |

| EPI_Accession_ID | Originating Laboratory | Submitting Laboratory |
|---|---|---|
| EPI_ISL_449304 | Virology Department, Royal Infirmary of Edinburgh, NHS Lothian / School of Biological Sciences, University of Edinburgh / Institute of Genetics and Molecular Medicine, University of Edinburgh | COVID-19 Genomics UK (COG-UK) Consortium |
| EPI_ISL_449305 | Virology Department, Royal Infirmary of Edinburgh, NHS Lothian / School of Biological Sciences, University of Edinburgh / Institute of Genetics and Molecular Medicine, University of Edinburgh | COVID-19 Genomics UK (COG-UK) Consortium |
| EPI_ISL_449306 | Virology Department, Royal Infirmary of Edinburgh, NHS Lothian / School of Biological Sciences, University of Edinburgh / Institute of Genetics and Molecular Medicine, University of Edinburgh | COVID-19 Genomics UK (COG-UK) Consortium |
| EPI_ISL_449307 | Virology Department, Royal Infirmary of Edinburgh, NHS Lothian / School of Biological Sciences, University of Edinburgh / Institute of Genetics and Molecular Medicine, University of Edinburgh | COVID-19 Genomics UK (COG-UK) Consortium |
| EPI_ISL_449308 | Virology Department, Royal Infirmary of Edinburgh, NHS Lothian / School of Biological Sciences, University of Edinburgh / Institute of Genetics and Molecular Medicine, University of Edinburgh | COVID-19 Genomics UK (COG-UK) Consortium |
| EPI_ISL_449309 | Virology Department, Royal Infirmary of Edinburgh, NHS Lothian / School of Biological Sciences, University of Edinburgh / Institute of Genetics and Molecular Medicine, University of Edinburgh | COVID-19 Genomics UK (COG-UK) Consortium |
| EPI_ISL_449310 | Virology Department, Royal Infirmary of Edinburgh, NHS Lothian / School of Biological Sciences, University of Edinburgh / Institute of Genetics and Molecular Medicine, University of Edinburgh | COVID-19 Genomics UK (COG-UK) Consortium |
| EPI_ISL_449311 | Virology Department, Royal Infirmary of Edinburgh, NHS Lothian / School of Biological Sciences, University of Edinburgh / Institute of Genetics and Molecular Medicine, University of Edinburgh | COVID-19 Genomics UK (COG-UK) Consortium |

| EPI_Accession_ID | Originating Laboratory | Submitting Laboratory |
|---|---|---|
| EPI_ISL_449312 | Virology Department, Royal Infirmary of Edinburgh, NHS Lothian / School of Biological Sciences, University of Edinburgh / Institute of Genetics and Molecular Medicine, University of Edinburgh | COVID-19 Genomics UK (COG-UK) Consortium |
| EPI_ISL_449313 | Virology Department, Royal Infirmary of Edinburgh, NHS Lothian / School of Biological Sciences, University of Edinburgh / Institute of Genetics and Molecular Medicine, University of Edinburgh | COVID-19 Genomics UK (COG-UK) Consortium |
| EPI_ISL_449314 | Virology Department, Royal Infirmary of Edinburgh, NHS Lothian / School of Biological Sciences, University of Edinburgh / Institute of Genetics and Molecular Medicine, University of Edinburgh | COVID-19 Genomics UK (COG-UK) Consortium |
| EPI_ISL_449315 | Virology Department, Royal Infirmary of Edinburgh, NHS Lothian / School of Biological Sciences, University of Edinburgh / Institute of Genetics and Molecular Medicine, University of Edinburgh | COVID-19 Genomics UK (COG-UK) Consortium |
| EPI_ISL_449316 | Virology Department, Royal Infirmary of Edinburgh, NHS Lothian / School of Biological Sciences, University of Edinburgh / Institute of Genetics and Molecular Medicine, University of Edinburgh | COVID-19 Genomics UK (COG-UK) Consortium |
| EPI_ISL_449317 | Virology Department, Royal Infirmary of Edinburgh, NHS Lothian / School of Biological Sciences, University of Edinburgh / Institute of Genetics and Molecular Medicine, University of Edinburgh | COVID-19 Genomics UK (COG-UK) Consortium |
| EPI_ISL_449318 | Virology Department, Royal Infirmary of Edinburgh, NHS Lothian / School of Biological Sciences, University of Edinburgh / Institute of Genetics and Molecular Medicine, University of Edinburgh | COVID-19 Genomics UK (COG-UK) Consortium |
| EPI_ISL_449319 | Virology Department, Royal Infirmary of Edinburgh, NHS Lothian / School of Biological Sciences, University of Edinburgh / Institute of Genetics and Molecular Medicine, University of Edinburgh | COVID-19 Genomics UK (COG-UK) Consortium |

| EPI_Accession_ID | Originating Laboratory | Submitting Laboratory |
|---|---|---|
| EPI_ISL_449320 | Virology Department, Royal Infirmary of Edinburgh, NHS Lothian / School of Biological Sciences, University of Edinburgh / Institute of Genetics and Molecular Medicine, University of Edinburgh | COVID-19 Genomics UK (COG-UK) Consortium |
| EPI_ISL_449321 | Virology Department, Royal Infirmary of Edinburgh, NHS Lothian / School of Biological Sciences, University of Edinburgh / Institute of Genetics and Molecular Medicine, University of Edinburgh | COVID-19 Genomics UK (COG-UK) Consortium |
| EPI_ISL_449322 | Virology Department, Royal Infirmary of Edinburgh, NHS Lothian / School of Biological Sciences, University of Edinburgh / Institute of Genetics and Molecular Medicine, University of Edinburgh | COVID-19 Genomics UK (COG-UK) Consortium |
| EPI_ISL_449323 | Virology Department, Royal Infirmary of Edinburgh, NHS Lothian / School of Biological Sciences, University of Edinburgh / Institute of Genetics and Molecular Medicine, University of Edinburgh | COVID-19 Genomics UK (COG-UK) Consortium |
| EPI_ISL_449324 | Virology Department, Royal Infirmary of Edinburgh, NHS Lothian / School of Biological Sciences, University of Edinburgh / Institute of Genetics and Molecular Medicine, University of Edinburgh | COVID-19 Genomics UK (COG-UK) Consortium |
| EPI_ISL_449325 | Virology Department, Royal Infirmary of Edinburgh, NHS Lothian / School of Biological Sciences, University of Edinburgh / Institute of Genetics and Molecular Medicine, University of Edinburgh | COVID-19 Genomics UK (COG-UK) Consortium |
| EPI_ISL_449326 | Virology Department, Royal Infirmary of Edinburgh, NHS Lothian / School of Biological Sciences, University of Edinburgh / Institute of Genetics and Molecular Medicine, University of Edinburgh | COVID-19 Genomics UK (COG-UK) Consortium |
| EPI_ISL_449327 | Virology Department, Royal Infirmary of Edinburgh, NHS Lothian / School of Biological Sciences, University of Edinburgh / Institute of Genetics and Molecular Medicine, University of Edinburgh | COVID-19 Genomics UK (COG-UK) Consortium |

| EPI_Accession_ID | Originating Laboratory | Submitting Laboratory |
|---|---|---|
| EPI_ISL_449328 | Virology Department, Royal Infirmary of Edinburgh, NHS Lothian / School of Biological Sciences, University of Edinburgh / Institute of Genetics and Molecular Medicine, University of Edinburgh | COVID-19 Genomics UK (COG-UK) Consortium |
| EPI_ISL_449329 | Virology Department, Royal Infirmary of Edinburgh, NHS Lothian / School of Biological Sciences, University of Edinburgh / Institute of Genetics and Molecular Medicine, University of Edinburgh | COVID-19 Genomics UK (COG-UK) Consortium |
| EPI_ISL_449330 | Liverpool Clinical Laboratories | COVID-19 Genomics UK (COG-UK) Consortium |
| EPI_ISL_449331 | Liverpool Clinical Laboratories | COVID-19 Genomics UK (COG-UK) Consortium |
| EPI_ISL_449332 | Liverpool Clinical Laboratories | COVID-19 Genomics UK (COG-UK) Consortium |
| EPI_ISL_449333 | Liverpool Clinical Laboratories | COVID-19 Genomics UK (COG-UK) Consortium |
| EPI_ISL_449334 | Liverpool Clinical Laboratories | COVID-19 Genomics UK (COG-UK) Consortium |
| EPI_ISL_449335 | Liverpool Clinical Laboratories | COVID-19 Genomics UK (COG-UK) Consortium |
| EPI_ISL_449336 | Liverpool Clinical Laboratories | COVID-19 Genomics UK (COG-UK) Consortium |
| EPI_ISL_449337 | Liverpool Clinical Laboratories | COVID-19 Genomics UK (COG-UK) Consortium |
| EPI_ISL_449338 | Liverpool Clinical Laboratories | COVID-19 Genomics UK (COG-UK) Consortium |
| EPI_ISL_449339 | Liverpool Clinical Laboratories | COVID-19 Genomics UK (COG-UK) Consortium |
| EPI_ISL_449340 | Liverpool Clinical Laboratories | COVID-19 Genomics UK (COG-UK) Consortium |
| EPI_ISL_449341 | Liverpool Clinical Laboratories | COVID-19 Genomics UK (COG-UK) Consortium |
| EPI_ISL_449342 | Liverpool Clinical Laboratories | COVID-19 Genomics UK (COG-UK) Consortium |
| EPI_ISL_449343 | Liverpool Clinical Laboratories | COVID-19 Genomics UK (COG-UK) Consortium |
| EPI_ISL_449344 | Liverpool Clinical Laboratories | COVID-19 Genomics UK (COG-UK) Consortium |
| EPI_ISL_449345 | Liverpool Clinical Laboratories | COVID-19 Genomics UK (COG-UK) Consortium |
| EPI_ISL_449346 | Liverpool Clinical Laboratories | COVID-19 Genomics UK (COG-UK) Consortium |
| EPI_ISL_449347 | Liverpool Clinical Laboratories | COVID-19 Genomics UK (COG-UK) Consortium |
| EPI_ISL_449348 | Liverpool Clinical Laboratories | COVID-19 Genomics UK (COG-UK) Consortium |
| EPI_ISL_449349 | Liverpool Clinical Laboratories | COVID-19 Genomics UK (COG-UK) Consortium |
| EPI_ISL_449350 | Liverpool Clinical Laboratories | COVID-19 Genomics UK (COG-UK) Consortium |
| EPI_ISL_449351 | Liverpool Clinical Laboratories | COVID-19 Genomics UK (COG-UK) Consortium |
| EPI_ISL_449352 | Liverpool Clinical Laboratories | COVID-19 Genomics UK (COG-UK) Consortium |
| EPI_ISL_449353 | Liverpool Clinical Laboratories | COVID-19 Genomics UK (COG-UK) Consortium |
| EPI_ISL_449354 | Liverpool Clinical Laboratories | COVID-19 Genomics UK (COG-UK) Consortium |

| EPI_Accession_ID | Originating Laboratory | Submitting Laboratory |
|---|---|---|
| EPI_ISL_449355 | Liverpool Clinical Laboratories | COVID-19 Genomics UK (COG-UK) Consortium |
| EPI_ISL_449356 | Liverpool Clinical Laboratories | COVID-19 Genomics UK (COG-UK) Consortium |
| EPI_ISL_449357 | Liverpool Clinical Laboratories | COVID-19 Genomics UK (COG-UK) Consortium |
| EPI_ISL_449358 | Liverpool Clinical Laboratories | COVID-19 Genomics UK (COG-UK) Consortium |
| EPI_ISL_449359 | Liverpool Clinical Laboratories | COVID-19 Genomics UK (COG-UK) Consortium |
| EPI_ISL_449360 | Liverpool Clinical Laboratories | COVID-19 Genomics UK (COG-UK) Consortium |
| EPI_ISL_449361 | Liverpool Clinical Laboratories | COVID-19 Genomics UK (COG-UK) Consortium |
| EPI_ISL_449362 | Liverpool Clinical Laboratories | COVID-19 Genomics UK (COG-UK) Consortium |
| EPI_ISL_449363 | Liverpool Clinical Laboratories | COVID-19 Genomics UK (COG-UK) Consortium |
| EPI_ISL_449364 | Liverpool Clinical Laboratories | COVID-19 Genomics UK (COG-UK) Consortium |
| EPI_ISL_449365 | Liverpool Clinical Laboratories | COVID-19 Genomics UK (COG-UK) Consortium |
| EPI_ISL_449366 | Liverpool Clinical Laboratories | COVID-19 Genomics UK (COG-UK) Consortium |
| EPI_ISL_449367 | Liverpool Clinical Laboratories | COVID-19 Genomics UK (COG-UK) Consortium |
| EPI_ISL_449368 | Liverpool Clinical Laboratories | COVID-19 Genomics UK (COG-UK) Consortium |
| EPI_ISL_449369 | Liverpool Clinical Laboratories | COVID-19 Genomics UK (COG-UK) Consortium |
| EPI_ISL_449370 | Liverpool Clinical Laboratories | COVID-19 Genomics UK (COG-UK) Consortium |
| EPI_ISL_449371 | Liverpool Clinical Laboratories | COVID-19 Genomics UK (COG-UK) Consortium |
| EPI_ISL_449372 | Liverpool Clinical Laboratories | COVID-19 Genomics UK (COG-UK) Consortium |
| EPI_ISL_449373 | Liverpool Clinical Laboratories | COVID-19 Genomics UK (COG-UK) Consortium |
| EPI_ISL_449374 | Liverpool Clinical Laboratories | COVID-19 Genomics UK (COG-UK) Consortium |
| EPI_ISL_449375 | Liverpool Clinical Laboratories | COVID-19 Genomics UK (COG-UK) Consortium |
| EPI_ISL_449376 | Liverpool Clinical Laboratories | COVID-19 Genomics UK (COG-UK) Consortium |
| EPI_ISL_449377 | Liverpool Clinical Laboratories | COVID-19 Genomics UK (COG-UK) Consortium |
| EPI_ISL_449378 | Liverpool Clinical Laboratories | COVID-19 Genomics UK (COG-UK) Consortium |
| EPI_ISL_449379 | Liverpool Clinical Laboratories | COVID-19 Genomics UK (COG-UK) Consortium |
| EPI_ISL_449380 | Liverpool Clinical Laboratories | COVID-19 Genomics UK (COG-UK) Consortium |
| EPI_ISL_449381 | Liverpool Clinical Laboratories | COVID-19 Genomics UK (COG-UK) Consortium |
| EPI_ISL_449382 | Liverpool Clinical Laboratories | COVID-19 Genomics UK (COG-UK) Consortium |
| EPI_ISL_449383 | Liverpool Clinical Laboratories | COVID-19 Genomics UK (COG-UK) Consortium |
| EPI_ISL_449384 | Liverpool Clinical Laboratories | COVID-19 Genomics UK (COG-UK) Consortium |
| EPI_ISL_449385 | Liverpool Clinical Laboratories | COVID-19 Genomics UK (COG-UK) Consortium |
| EPI_ISL_449386 | Liverpool Clinical Laboratories | COVID-19 Genomics UK (COG-UK) Consortium |
| EPI_ISL_449387 | Liverpool Clinical Laboratories | COVID-19 Genomics UK (COG-UK) Consortium |

| EPI_Accession_ID | Originating Laboratory | Submitting Laboratory |
|---|---|---|
| EPI_ISL_449388 | Liverpool Clinical Laboratories | COVID-19 Genomics UK (COG-UK) Consortium |
| EPI_ISL_449389 | Liverpool Clinical Laboratories | COVID-19 Genomics UK (COG-UK) Consortium |
| EPI_ISL_449390 | Liverpool Clinical Laboratories | COVID-19 Genomics UK (COG-UK) Consortium |
| EPI_ISL_449391 | Liverpool Clinical Laboratories | COVID-19 Genomics UK (COG-UK) Consortium |
| EPI_ISL_449392 | Liverpool Clinical Laboratories | COVID-19 Genomics UK (COG-UK) Consortium |
| EPI_ISL_449393 | Liverpool Clinical Laboratories | COVID-19 Genomics UK (COG-UK) Consortium |
| EPI_ISL_449394 | Liverpool Clinical Laboratories | COVID-19 Genomics UK (COG-UK) Consortium |
| EPI_ISL_449395 | Liverpool Clinical Laboratories | COVID-19 Genomics UK (COG-UK) Consortium |
| EPI_ISL_449396 | Liverpool Clinical Laboratories | COVID-19 Genomics UK (COG-UK) Consortium |
| EPI_ISL_449397 | Liverpool Clinical Laboratories | COVID-19 Genomics UK (COG-UK) Consortium |
| EPI_ISL_449398 | Liverpool Clinical Laboratories | COVID-19 Genomics UK (COG-UK) Consortium |
| EPI_ISL_449399 | Liverpool Clinical Laboratories | COVID-19 Genomics UK (COG-UK) Consortium |
| EPI_ISL_449400 | Liverpool Clinical Laboratories | COVID-19 Genomics UK (COG-UK) Consortium |
| EPI_ISL_449401 | Liverpool Clinical Laboratories | COVID-19 Genomics UK (COG-UK) Consortium |
| EPI_ISL_449402 | Liverpool Clinical Laboratories | COVID-19 Genomics UK (COG-UK) Consortium |
| EPI_ISL_449403 | Liverpool Clinical Laboratories | COVID-19 Genomics UK (COG-UK) Consortium |
| EPI_ISL_449404 | Liverpool Clinical Laboratories | COVID-19 Genomics UK (COG-UK) Consortium |
| EPI_ISL_449405 | Liverpool Clinical Laboratories | COVID-19 Genomics UK (COG-UK) Consortium |
| EPI_ISL_449406 | Liverpool Clinical Laboratories | COVID-19 Genomics UK (COG-UK) Consortium |
| EPI_ISL_449407 | Liverpool Clinical Laboratories | COVID-19 Genomics UK (COG-UK) Consortium |
| EPI_ISL_449408 | Liverpool Clinical Laboratories | COVID-19 Genomics UK (COG-UK) Consortium |
| EPI_ISL_449409 | Liverpool Clinical Laboratories | COVID-19 Genomics UK (COG-UK) Consortium |
| EPI_ISL_449410 | Liverpool Clinical Laboratories | COVID-19 Genomics UK (COG-UK) Consortium |
| EPI_ISL_449411 | Liverpool Clinical Laboratories | COVID-19 Genomics UK (COG-UK) Consortium |
| EPI_ISL_449412 | Liverpool Clinical Laboratories | COVID-19 Genomics UK (COG-UK) Consortium |
| EPI_ISL_449413 | Liverpool Clinical Laboratories | COVID-19 Genomics UK (COG-UK) Consortium |
| EPI_ISL_449414 | Liverpool Clinical Laboratories | COVID-19 Genomics UK (COG-UK) Consortium |
| EPI_ISL_449415 | Liverpool Clinical Laboratories | COVID-19 Genomics UK (COG-UK) Consortium |
| EPI_ISL_449416 | Liverpool Clinical Laboratories | COVID-19 Genomics UK (COG-UK) Consortium |
| EPI_ISL_449417 | Liverpool Clinical Laboratories | COVID-19 Genomics UK (COG-UK) Consortium |
| EPI_ISL_449418 | Liverpool Clinical Laboratories | COVID-19 Genomics UK (COG-UK) Consortium |
| EPI_ISL_449419 | Liverpool Clinical Laboratories | COVID-19 Genomics UK (COG-UK) Consortium |
| EPI_ISL_449420 | Liverpool Clinical Laboratories | COVID-19 Genomics UK (COG-UK) Consortium |

| EPI_Accession_ID | Originating Laboratory | Submitting Laboratory |
|---|---|---|
| EPI_ISL_449421 | Liverpool Clinical Laboratories | COVID-19 Genomics UK (COG-UK) Consortium |
| EPI_ISL_449422 | Liverpool Clinical Laboratories | COVID-19 Genomics UK (COG-UK) Consortium |
| EPI_ISL_449423 | Liverpool Clinical Laboratories | COVID-19 Genomics UK (COG-UK) Consortium |
| EPI_ISL_449424 | Liverpool Clinical Laboratories | COVID-19 Genomics UK (COG-UK) Consortium |
| EPI_ISL_449425 | Liverpool Clinical Laboratories | COVID-19 Genomics UK (COG-UK) Consortium |
| EPI_ISL_449426 | Liverpool Clinical Laboratories | COVID-19 Genomics UK (COG-UK) Consortium |
| EPI_ISL_449427 | Liverpool Clinical Laboratories | COVID-19 Genomics UK (COG-UK) Consortium |
| EPI_ISL_449428 | Liverpool Clinical Laboratories | COVID-19 Genomics UK (COG-UK) Consortium |
| EPI_ISL_449429 | Liverpool Clinical Laboratories | COVID-19 Genomics UK (COG-UK) Consortium |
| EPI_ISL_449430 | Liverpool Clinical Laboratories | COVID-19 Genomics UK (COG-UK) Consortium |
| EPI_ISL_449431 | Liverpool Clinical Laboratories | COVID-19 Genomics UK (COG-UK) Consortium |
| EPI_ISL_449432 | Liverpool Clinical Laboratories | COVID-19 Genomics UK (COG-UK) Consortium |
| EPI_ISL_449433 | Liverpool Clinical Laboratories | COVID-19 Genomics UK (COG-UK) Consortium |
| EPI_ISL_449434 | Liverpool Clinical Laboratories | COVID-19 Genomics UK (COG-UK) Consortium |
| EPI_ISL_449435 | Liverpool Clinical Laboratories | COVID-19 Genomics UK (COG-UK) Consortium |
| EPI_ISL_449436 | Liverpool Clinical Laboratories | COVID-19 Genomics UK (COG-UK) Consortium |
| EPI_ISL_449437 | Liverpool Clinical Laboratories | COVID-19 Genomics UK (COG-UK) Consortium |
| EPI_ISL_449438 | Liverpool Clinical Laboratories | COVID-19 Genomics UK (COG-UK) Consortium |
| EPI_ISL_449439 | Liverpool Clinical Laboratories | COVID-19 Genomics UK (COG-UK) Consortium |
| EPI_ISL_449440 | Liverpool Clinical Laboratories | COVID-19 Genomics UK (COG-UK) Consortium |
| EPI_ISL_449441 | Liverpool Clinical Laboratories | COVID-19 Genomics UK (COG-UK) Consortium |
| EPI_ISL_449442 | Liverpool Clinical Laboratories | COVID-19 Genomics UK (COG-UK) Consortium |
| EPI_ISL_449443 | Liverpool Clinical Laboratories | COVID-19 Genomics UK (COG-UK) Consortium |
| EPI_ISL_449444 | Liverpool Clinical Laboratories | COVID-19 Genomics UK (COG-UK) Consortium |
| EPI_ISL_449445 | Liverpool Clinical Laboratories | COVID-19 Genomics UK (COG-UK) Consortium |
| EPI_ISL_449446 | Liverpool Clinical Laboratories | COVID-19 Genomics UK (COG-UK) Consortium |
| EPI_ISL_449447 | Liverpool Clinical Laboratories | COVID-19 Genomics UK (COG-UK) Consortium |
| EPI_ISL_449448 | Liverpool Clinical Laboratories | COVID-19 Genomics UK (COG-UK) Consortium |
| EPI_ISL_449449 | Liverpool Clinical Laboratories | COVID-19 Genomics UK (COG-UK) Consortium |
| EPI_ISL_449450 | Liverpool Clinical Laboratories | COVID-19 Genomics UK (COG-UK) Consortium |
| EPI_ISL_449451 | Liverpool Clinical Laboratories | COVID-19 Genomics UK (COG-UK) Consortium |
| EPI_ISL_449452 | Liverpool Clinical Laboratories | COVID-19 Genomics UK (COG-UK) Consortium |
| EPI_ISL_449453 | Liverpool Clinical Laboratories | COVID-19 Genomics UK (COG-UK) Consortium |

| EPI_Accession_ID | Originating Laboratory | Submitting Laboratory |
|---|---|---|
| EPI_ISL_449454 | Liverpool Clinical Laboratories | COVID-19 Genomics UK (COG-UK) Consortium |
| EPI_ISL_449455 | Liverpool Clinical Laboratories | COVID-19 Genomics UK (COG-UK) Consortium |
| EPI_ISL_449456 | Liverpool Clinical Laboratories | COVID-19 Genomics UK (COG-UK) Consortium |
| EPI_ISL_449457 | Liverpool Clinical Laboratories | COVID-19 Genomics UK (COG-UK) Consortium |
| EPI_ISL_449458 | Liverpool Clinical Laboratories | COVID-19 Genomics UK (COG-UK) Consortium |
| EPI_ISL_449459 | Liverpool Clinical Laboratories | COVID-19 Genomics UK (COG-UK) Consortium |
| EPI_ISL_449460 | Liverpool Clinical Laboratories | COVID-19 Genomics UK (COG-UK) Consortium |
| EPI_ISL_449461 | Liverpool Clinical Laboratories | COVID-19 Genomics UK (COG-UK) Consortium |
| EPI_ISL_449462 | Liverpool Clinical Laboratories | COVID-19 Genomics UK (COG-UK) Consortium |
| EPI_ISL_449463 | Liverpool Clinical Laboratories | COVID-19 Genomics UK (COG-UK) Consortium |
| EPI_ISL_449464 | Liverpool Clinical Laboratories | COVID-19 Genomics UK (COG-UK) Consortium |
| EPI_ISL_449465 | Liverpool Clinical Laboratories | COVID-19 Genomics UK (COG-UK) Consortium |
| EPI_ISL_449466 | Liverpool Clinical Laboratories | COVID-19 Genomics UK (COG-UK) Consortium |
| EPI_ISL_449467 | Liverpool Clinical Laboratories | COVID-19 Genomics UK (COG-UK) Consortium |
| EPI_ISL_449468 | Liverpool Clinical Laboratories | COVID-19 Genomics UK (COG-UK) Consortium |
| EPI_ISL_449469 | Liverpool Clinical Laboratories | COVID-19 Genomics UK (COG-UK) Consortium |
| EPI_ISL_449470 | Liverpool Clinical Laboratories | COVID-19 Genomics UK (COG-UK) Consortium |
| EPI_ISL_449471 | Liverpool Clinical Laboratories | COVID-19 Genomics UK (COG-UK) Consortium |
| EPI_ISL_449472 | Liverpool Clinical Laboratories | COVID-19 Genomics UK (COG-UK) Consortium |
| EPI_ISL_449473 | Liverpool Clinical Laboratories | COVID-19 Genomics UK (COG-UK) Consortium |
| EPI_ISL_449474 | Liverpool Clinical Laboratories | COVID-19 Genomics UK (COG-UK) Consortium |
| EPI_ISL_449475 | Liverpool Clinical Laboratories | COVID-19 Genomics UK (COG-UK) Consortium |
| EPI_ISL_449476 | unknown | Department of Respiratory and Critical Care |
| EPI_ISL_449477 | unknown | Department of Respiratory and Critical Care |
| EPI_ISL_449478 | unknown | Department of Respiratory and Critical Care |
| EPI_ISL_449479 | unknown | Department of Respiratory and Critical Care |
| EPI_ISL_449480 | unknown | Department of Respiratory and Critical Care |
| EPI_ISL_449481 | unknown | Department of Respiratory and Critical Care |
| EPI_ISL_449482 | unknown | Department of Respiratory and Critical Care |
| EPI_ISL_449483 | unknown | Department of Respiratory and Critical Care |
| EPI_ISL_449484 | unknown | Department of Respiratory and Critical Care |
| EPI_ISL_449485 | unknown | Department of Respiratory and Critical Care |
| EPI_ISL_449486 | unknown | Department of Respiratory and Critical Care |

| EPI_Accession_ID | Originating Laboratory | Submitting Laboratory |
|---|---|---|
| EPI_ISL_449487 | unknown | Department of Respiratory and Critical Care |
| EPI_ISL_449488 | Liverpool Clinical Laboratories | COVID-19 Genomics UK (COG-UK) Consortium |
| EPI_ISL_449489 | Liverpool Clinical Laboratories | COVID-19 Genomics UK (COG-UK) Consortium |
| EPI_ISL_449490 | Liverpool Clinical Laboratories | COVID-19 Genomics UK (COG-UK) Consortium |
| EPI_ISL_449491 | Liverpool Clinical Laboratories | COVID-19 Genomics UK (COG-UK) Consortium |
| EPI_ISL_449492 | Liverpool Clinical Laboratories | COVID-19 Genomics UK (COG-UK) Consortium |
| EPI_ISL_449493 | Liverpool Clinical Laboratories | COVID-19 Genomics UK (COG-UK) Consortium |
| EPI_ISL_449494 | Liverpool Clinical Laboratories | COVID-19 Genomics UK (COG-UK) Consortium |
| EPI_ISL_449495 | Liverpool Clinical Laboratories | COVID-19 Genomics UK (COG-UK) Consortium |
| EPI_ISL_449496 | Liverpool Clinical Laboratories | COVID-19 Genomics UK (COG-UK) Consortium |
| EPI_ISL_449497 | Liverpool Clinical Laboratories | COVID-19 Genomics UK (COG-UK) Consortium |
| EPI_ISL_449498 | Liverpool Clinical Laboratories | COVID-19 Genomics UK (COG-UK) Consortium |
| EPI_ISL_449499 | Liverpool Clinical Laboratories | COVID-19 Genomics UK (COG-UK) Consortium |
| EPI_ISL_449500 | Liverpool Clinical Laboratories | COVID-19 Genomics UK (COG-UK) Consortium |
| EPI_ISL_449501 | Liverpool Clinical Laboratories | COVID-19 Genomics UK (COG-UK) Consortium |
| EPI_ISL_449502 | Liverpool Clinical Laboratories | COVID-19 Genomics UK (COG-UK) Consortium |
| EPI_ISL_449503 | Liverpool Clinical Laboratories | COVID-19 Genomics UK (COG-UK) Consortium |
| EPI_ISL_449504 | Liverpool Clinical Laboratories | COVID-19 Genomics UK (COG-UK) Consortium |
| EPI_ISL_449505 | Liverpool Clinical Laboratories | COVID-19 Genomics UK (COG-UK) Consortium |
| EPI_ISL_449506 | Liverpool Clinical Laboratories | COVID-19 Genomics UK (COG-UK) Consortium |
| EPI_ISL_449507 | Liverpool Clinical Laboratories | COVID-19 Genomics UK (COG-UK) Consortium |
| EPI_ISL_449508 | Liverpool Clinical Laboratories | COVID-19 Genomics UK (COG-UK) Consortium |
| EPI_ISL_449509 | Liverpool Clinical Laboratories | COVID-19 Genomics UK (COG-UK) Consortium |
| EPI_ISL_449510 | Liverpool Clinical Laboratories | COVID-19 Genomics UK (COG-UK) Consortium |
| EPI_ISL_449511 | Liverpool Clinical Laboratories | COVID-19 Genomics UK (COG-UK) Consortium |
| EPI_ISL_449512 | Liverpool Clinical Laboratories | COVID-19 Genomics UK (COG-UK) Consortium |
| EPI_ISL_449513 | Liverpool Clinical Laboratories | COVID-19 Genomics UK (COG-UK) Consortium |
| EPI_ISL_449514 | Liverpool Clinical Laboratories | COVID-19 Genomics UK (COG-UK) Consortium |
| EPI_ISL_449515 | Liverpool Clinical Laboratories | COVID-19 Genomics UK (COG-UK) Consortium |
| EPI_ISL_449516 | Liverpool Clinical Laboratories | COVID-19 Genomics UK (COG-UK) Consortium |
| EPI_ISL_449517 | Liverpool Clinical Laboratories | COVID-19 Genomics UK (COG-UK) Consortium |
| EPI_ISL_449518 | Liverpool Clinical Laboratories | COVID-19 Genomics UK (COG-UK) Consortium |
| EPI_ISL_449519 | Liverpool Clinical Laboratories | COVID-19 Genomics UK (COG-UK) Consortium |

| EPI_Accession_ID | Originating Laboratory | Submitting Laboratory |
|---|---|---|
| EPI_ISL_449520 | Liverpool Clinical Laboratories | COVID-19 Genomics UK (COG-UK) Consortium |
| EPI_ISL_449521 | Liverpool Clinical Laboratories | COVID-19 Genomics UK (COG-UK) Consortium |
| EPI_ISL_449522 | Liverpool Clinical Laboratories | COVID-19 Genomics UK (COG-UK) Consortium |
| EPI_ISL_449523 | Liverpool Clinical Laboratories | COVID-19 Genomics UK (COG-UK) Consortium |
| EPI_ISL_449524 | Liverpool Clinical Laboratories | COVID-19 Genomics UK (COG-UK) Consortium |
| EPI_ISL_449525 | Liverpool Clinical Laboratories | COVID-19 Genomics UK (COG-UK) Consortium |
| EPI_ISL_449526 | Liverpool Clinical Laboratories | COVID-19 Genomics UK (COG-UK) Consortium |
| EPI_ISL_449527 | Liverpool Clinical Laboratories | COVID-19 Genomics UK (COG-UK) Consortium |
| EPI_ISL_449528 | Liverpool Clinical Laboratories | COVID-19 Genomics UK (COG-UK) Consortium |
| EPI_ISL_449529 | Liverpool Clinical Laboratories | COVID-19 Genomics UK (COG-UK) Consortium |
| EPI_ISL_449530 | Liverpool Clinical Laboratories | COVID-19 Genomics UK (COG-UK) Consortium |
| EPI_ISL_449531 | Liverpool Clinical Laboratories | COVID-19 Genomics UK (COG-UK) Consortium |
| EPI_ISL_449532 | Liverpool Clinical Laboratories | COVID-19 Genomics UK (COG-UK) Consortium |
| EPI_ISL_449533 | Liverpool Clinical Laboratories | COVID-19 Genomics UK (COG-UK) Consortium |
| EPI_ISL_449534 | Liverpool Clinical Laboratories | COVID-19 Genomics UK (COG-UK) Consortium |
| EPI_ISL_449535 | Liverpool Clinical Laboratories | COVID-19 Genomics UK (COG-UK) Consortium |
| EPI_ISL_449536 | Liverpool Clinical Laboratories | COVID-19 Genomics UK (COG-UK) Consortium |
| EPI_ISL_449537 | Liverpool Clinical Laboratories | COVID-19 Genomics UK (COG-UK) Consortium |
| EPI_ISL_449538 | Liverpool Clinical Laboratories | COVID-19 Genomics UK (COG-UK) Consortium |
| EPI_ISL_449539 | Liverpool Clinical Laboratories | COVID-19 Genomics UK (COG-UK) Consortium |
| EPI_ISL_449540 | Liverpool Clinical Laboratories | COVID-19 Genomics UK (COG-UK) Consortium |
| EPI_ISL_449541 | Liverpool Clinical Laboratories | COVID-19 Genomics UK (COG-UK) Consortium |
| EPI_ISL_449542 | Liverpool Clinical Laboratories | COVID-19 Genomics UK (COG-UK) Consortium |
| EPI_ISL_449543 | Liverpool Clinical Laboratories | COVID-19 Genomics UK (COG-UK) Consortium |
| EPI_ISL_449544 | Liverpool Clinical Laboratories | COVID-19 Genomics UK (COG-UK) Consortium |
| EPI_ISL_449545 | Liverpool Clinical Laboratories | COVID-19 Genomics UK (COG-UK) Consortium |
| EPI_ISL_449546 | Liverpool Clinical Laboratories | COVID-19 Genomics UK (COG-UK) Consortium |
| EPI_ISL_449547 | Liverpool Clinical Laboratories | COVID-19 Genomics UK (COG-UK) Consortium |
| EPI_ISL_449548 | Liverpool Clinical Laboratories | COVID-19 Genomics UK (COG-UK) Consortium |
| EPI_ISL_449549 | Liverpool Clinical Laboratories | COVID-19 Genomics UK (COG-UK) Consortium |
| EPI_ISL_449550 | Liverpool Clinical Laboratories | COVID-19 Genomics UK (COG-UK) Consortium |
| EPI_ISL_449551 | Liverpool Clinical Laboratories | COVID-19 Genomics UK (COG-UK) Consortium |
| EPI_ISL_449552 | Liverpool Clinical Laboratories | COVID-19 Genomics UK (COG-UK) Consortium |

| EPI_Accession_ID | Originating Laboratory | Submitting Laboratory |
|---|---|---|
| EPI_ISL_449553 | Liverpool Clinical Laboratories | COVID-19 Genomics UK (COG-UK) Consortium |
| EPI_ISL_449554 | Liverpool Clinical Laboratories | COVID-19 Genomics UK (COG-UK) Consortium |
| EPI_ISL_449555 | Liverpool Clinical Laboratories | COVID-19 Genomics UK (COG-UK) Consortium |
| EPI_ISL_449556 | Liverpool Clinical Laboratories | COVID-19 Genomics UK (COG-UK) Consortium |
| EPI_ISL_449557 | Liverpool Clinical Laboratories | COVID-19 Genomics UK (COG-UK) Consortium |
| EPI_ISL_449558 | Liverpool Clinical Laboratories | COVID-19 Genomics UK (COG-UK) Consortium |
| EPI_ISL_449559 | Liverpool Clinical Laboratories | COVID-19 Genomics UK (COG-UK) Consortium |
| EPI_ISL_449560 | Liverpool Clinical Laboratories | COVID-19 Genomics UK (COG-UK) Consortium |
| EPI_ISL_449561 | Liverpool Clinical Laboratories | COVID-19 Genomics UK (COG-UK) Consortium |
| EPI_ISL_449562 | Liverpool Clinical Laboratories | COVID-19 Genomics UK (COG-UK) Consortium |
| EPI_ISL_449563 | Liverpool Clinical Laboratories | COVID-19 Genomics UK (COG-UK) Consortium |
| EPI_ISL_449564 | Liverpool Clinical Laboratories | COVID-19 Genomics UK (COG-UK) Consortium |
| EPI_ISL_449565 | Liverpool Clinical Laboratories | COVID-19 Genomics UK (COG-UK) Consortium |
| EPI_ISL_449566 | Liverpool Clinical Laboratories | COVID-19 Genomics UK (COG-UK) Consortium |
| EPI_ISL_449567 | Liverpool Clinical Laboratories | COVID-19 Genomics UK (COG-UK) Consortium |
| EPI_ISL_449568 | Liverpool Clinical Laboratories | COVID-19 Genomics UK (COG-UK) Consortium |
| EPI_ISL_449569 | Liverpool Clinical Laboratories | COVID-19 Genomics UK (COG-UK) Consortium |
| EPI_ISL_449570 | Liverpool Clinical Laboratories | COVID-19 Genomics UK (COG-UK) Consortium |
| EPI_ISL_449571 | Liverpool Clinical Laboratories | COVID-19 Genomics UK (COG-UK) Consortium |
| EPI_ISL_449572 | Liverpool Clinical Laboratories | COVID-19 Genomics UK (COG-UK) Consortium |
| EPI_ISL_449573 | Liverpool Clinical Laboratories | COVID-19 Genomics UK (COG-UK) Consortium |
| EPI_ISL_449574 | Liverpool Clinical Laboratories | COVID-19 Genomics UK (COG-UK) Consortium |
| EPI_ISL_449575 | Liverpool Clinical Laboratories | COVID-19 Genomics UK (COG-UK) Consortium |
| EPI_ISL_449576 | Liverpool Clinical Laboratories | COVID-19 Genomics UK (COG-UK) Consortium |
| EPI_ISL_449577 | Liverpool Clinical Laboratories | COVID-19 Genomics UK (COG-UK) Consortium |
| EPI_ISL_449578 | Liverpool Clinical Laboratories | COVID-19 Genomics UK (COG-UK) Consortium |
| EPI_ISL_449579 | Liverpool Clinical Laboratories | COVID-19 Genomics UK (COG-UK) Consortium |
| EPI_ISL_449580 | Liverpool Clinical Laboratories | COVID-19 Genomics UK (COG-UK) Consortium |
| EPI_ISL_449581 | Liverpool Clinical Laboratories | COVID-19 Genomics UK (COG-UK) Consortium |
| EPI_ISL_449582 | Liverpool Clinical Laboratories | COVID-19 Genomics UK (COG-UK) Consortium |
| EPI_ISL_449583 | Liverpool Clinical Laboratories | COVID-19 Genomics UK (COG-UK) Consortium |
| EPI_ISL_449584 | Liverpool Clinical Laboratories | COVID-19 Genomics UK (COG-UK) Consortium |
| EPI_ISL_449585 | Liverpool Clinical Laboratories | COVID-19 Genomics UK (COG-UK) Consortium |

| EPI_Accession_ID | Originating Laboratory | Submitting Laboratory |
|---|---|---|
| EPI_ISL_449586 | Liverpool Clinical Laboratories | COVID-19 Genomics UK (COG-UK) Consortium |
| EPI_ISL_449587 | Liverpool Clinical Laboratories | COVID-19 Genomics UK (COG-UK) Consortium |
| EPI_ISL_449588 | Liverpool Clinical Laboratories | COVID-19 Genomics UK (COG-UK) Consortium |
| EPI_ISL_449589 | Liverpool Clinical Laboratories | COVID-19 Genomics UK (COG-UK) Consortium |
| EPI_ISL_449590 | Liverpool Clinical Laboratories | COVID-19 Genomics UK (COG-UK) Consortium |
| EPI_ISL_449591 | Liverpool Clinical Laboratories | COVID-19 Genomics UK (COG-UK) Consortium |
| EPI_ISL_449592 | Liverpool Clinical Laboratories | COVID-19 Genomics UK (COG-UK) Consortium |
| EPI_ISL_449593 | Liverpool Clinical Laboratories | COVID-19 Genomics UK (COG-UK) Consortium |
| EPI_ISL_449594 | Liverpool Clinical Laboratories | COVID-19 Genomics UK (COG-UK) Consortium |
| EPI_ISL_449595 | Liverpool Clinical Laboratories | COVID-19 Genomics UK (COG-UK) Consortium |
| EPI_ISL_449596 | Liverpool Clinical Laboratories | COVID-19 Genomics UK (COG-UK) Consortium |
| EPI_ISL_449597 | Liverpool Clinical Laboratories | COVID-19 Genomics UK (COG-UK) Consortium |
| EPI_ISL_449598 | Liverpool Clinical Laboratories | COVID-19 Genomics UK (COG-UK) Consortium |
| EPI_ISL_449599 | Liverpool Clinical Laboratories | COVID-19 Genomics UK (COG-UK) Consortium |
| EPI_ISL_449600 | Liverpool Clinical Laboratories | COVID-19 Genomics UK (COG-UK) Consortium |
| EPI_ISL_449601 | Liverpool Clinical Laboratories | COVID-19 Genomics UK (COG-UK) Consortium |
| EPI_ISL_449602 | Liverpool Clinical Laboratories | COVID-19 Genomics UK (COG-UK) Consortium |
| EPI_ISL_449603 | Liverpool Clinical Laboratories | COVID-19 Genomics UK (COG-UK) Consortium |
| EPI_ISL_449604 | Liverpool Clinical Laboratories | COVID-19 Genomics UK (COG-UK) Consortium |
| EPI_ISL_449605 | Liverpool Clinical Laboratories | COVID-19 Genomics UK (COG-UK) Consortium |
| EPI_ISL_449606 | Liverpool Clinical Laboratories | COVID-19 Genomics UK (COG-UK) Consortium |
| EPI_ISL_449607 | Liverpool Clinical Laboratories | COVID-19 Genomics UK (COG-UK) Consortium |
| EPI_ISL_449608 | Liverpool Clinical Laboratories | COVID-19 Genomics UK (COG-UK) Consortium |
| EPI_ISL_449609 | Liverpool Clinical Laboratories | COVID-19 Genomics UK (COG-UK) Consortium |
| EPI_ISL_449610 | Liverpool Clinical Laboratories | COVID-19 Genomics UK (COG-UK) Consortium |
| EPI_ISL_449611 | Liverpool Clinical Laboratories | COVID-19 Genomics UK (COG-UK) Consortium |
| EPI_ISL_449612 | Liverpool Clinical Laboratories | COVID-19 Genomics UK (COG-UK) Consortium |
| EPI_ISL_449613 | Liverpool Clinical Laboratories | COVID-19 Genomics UK (COG-UK) Consortium |
| EPI_ISL_449614 | Liverpool Clinical Laboratories | COVID-19 Genomics UK (COG-UK) Consortium |
| EPI_ISL_449615 | Liverpool Clinical Laboratories | COVID-19 Genomics UK (COG-UK) Consortium |
| EPI_ISL_449616 | Liverpool Clinical Laboratories | COVID-19 Genomics UK (COG-UK) Consortium |
| EPI_ISL_449617 | Liverpool Clinical Laboratories | COVID-19 Genomics UK (COG-UK) Consortium |
| EPI_ISL_449618 | Liverpool Clinical Laboratories | COVID-19 Genomics UK (COG-UK) Consortium |

| EPI_Accession_ID | Originating Laboratory | Submitting Laboratory |
|---|---|---|
| EPI_ISL_449619 | Liverpool Clinical Laboratories | COVID-19 Genomics UK (COG-UK) Consortium |
| EPI_ISL_449620 | Liverpool Clinical Laboratories | COVID-19 Genomics UK (COG-UK) Consortium |
| EPI_ISL_449621 | Liverpool Clinical Laboratories | COVID-19 Genomics UK (COG-UK) Consortium |
| EPI_ISL_449622 | Liverpool Clinical Laboratories | COVID-19 Genomics UK (COG-UK) Consortium |
| EPI_ISL_449623 | Liverpool Clinical Laboratories | COVID-19 Genomics UK (COG-UK) Consortium |
| EPI_ISL_449624 | Liverpool Clinical Laboratories | COVID-19 Genomics UK (COG-UK) Consortium |
| EPI_ISL_449625 | Liverpool Clinical Laboratories | COVID-19 Genomics UK (COG-UK) Consortium |
| EPI_ISL_449626 | University College London, Great Ormond Street Hospital for Children NHS Foundation Trust, Imperial College Healthcare NHS Trust | COVID-19 Genomics UK (COG-UK) Consortium |
| EPI_ISL_449627 | University College London, Great Ormond Street Hospital for Children NHS Foundation Trust, Imperial College Healthcare NHS Trust | COVID-19 Genomics UK (COG-UK) Consortium |
| EPI_ISL_449628 | University College London, Great Ormond Street Hospital for Children NHS Foundation Trust, Imperial College Healthcare NHS Trust | COVID-19 Genomics UK (COG-UK) Consortium |
| EPI_ISL_449629 | University College London, Great Ormond Street Hospital for Children NHS Foundation Trust, Imperial College Healthcare NHS Trust | COVID-19 Genomics UK (COG-UK) Consortium |
| EPI_ISL_449630 | University College London, Great Ormond Street Hospital for Children NHS Foundation Trust, Imperial College Healthcare NHS Trust | COVID-19 Genomics UK (COG-UK) Consortium |
| EPI_ISL_449631 | University College London, Great Ormond Street Hospital for Children NHS Foundation Trust, Imperial College Healthcare NHS Trust | COVID-19 Genomics UK (COG-UK) Consortium |
| EPI_ISL_449632 | University College London, Great Ormond Street Hospital for Children NHS Foundation Trust, Imperial College Healthcare NHS Trust | COVID-19 Genomics UK (COG-UK) Consortium |
| EPI_ISL_449633 | University College London, Great Ormond Street Hospital for Children NHS Foundation Trust, Imperial College Healthcare NHS Trust | COVID-19 Genomics UK (COG-UK) Consortium |

| EPI_Accession_ID | Originating Laboratory | Submitting Laboratory |
|---|---|---|
| EPI_ISL_449634 | University College London, Great Ormond Street Hospital for Children NHS Foundation Trust, Imperial College Healthcare NHS Trust | COVID-19 Genomics UK (COG-UK) Consortium |
| EPI_ISL_449635 | University College London, Great Ormond Street Hospital for Children NHS Foundation Trust, Imperial College Healthcare NHS Trust | COVID-19 Genomics UK (COG-UK) Consortium |
| EPI_ISL_449636 | University College London, Great Ormond Street Hospital for Children NHS Foundation Trust, Imperial College Healthcare NHS Trust | COVID-19 Genomics UK (COG-UK) Consortium |
| EPI_ISL_449637 | University College London, Great Ormond Street Hospital for Children NHS Foundation Trust, Imperial College Healthcare NHS Trust | COVID-19 Genomics UK (COG-UK) Consortium |
| EPI_ISL_449638 | University College London, Great Ormond Street Hospital for Children NHS Foundation Trust, Imperial College Healthcare NHS Trust | COVID-19 Genomics UK (COG-UK) Consortium |
| EPI_ISL_449639 | University College London, Great Ormond Street Hospital for Children NHS Foundation Trust, Imperial College Healthcare NHS Trust | COVID-19 Genomics UK (COG-UK) Consortium |
| EPI_ISL_449640 | University College London, Great Ormond Street Hospital for Children NHS Foundation Trust, Imperial College Healthcare NHS Trust | COVID-19 Genomics UK (COG-UK) Consortium |
| EPI_ISL_449641 | University College London, Great Ormond Street Hospital for Children NHS Foundation Trust, Imperial College Healthcare NHS Trust | COVID-19 Genomics UK (COG-UK) Consortium |
| EPI_ISL_449642 | University College London, Great Ormond Street Hospital for Children NHS Foundation Trust, Imperial College Healthcare NHS Trust | COVID-19 Genomics UK (COG-UK) Consortium |
| EPI_ISL_449643 | University College London, Great Ormond Street Hospital for Children NHS Foundation Trust, Imperial College Healthcare NHS Trust | COVID-19 Genomics UK (COG-UK) Consortium |
| EPI_ISL_449644 | University College London, Great Ormond Street Hospital for Children NHS Foundation Trust, Imperial College Healthcare NHS Trust | COVID-19 Genomics UK (COG-UK) Consortium |

| EPI_Accession_ID | Originating Laboratory | Submitting Laboratory |
|---|---|---|
| EPI_ISL_449645 | University College London, Great Ormond Street Hospital for Children NHS Foundation Trust, Imperial College Healthcare NHS Trust | COVID-19 Genomics UK (COG-UK) Consortium |
| EPI_ISL_449646 | University College London, Great Ormond Street Hospital for Children NHS Foundation Trust, Imperial College Healthcare NHS Trust | COVID-19 Genomics UK (COG-UK) Consortium |
| EPI_ISL_449647 | University College London, Great Ormond Street Hospital for Children NHS Foundation Trust, Imperial College Healthcare NHS Trust | COVID-19 Genomics UK (COG-UK) Consortium |
| EPI_ISL_449648 | University College London, Great Ormond Street Hospital for Children NHS Foundation Trust, Imperial College Healthcare NHS Trust | COVID-19 Genomics UK (COG-UK) Consortium |
| EPI_ISL_449649 | University College London, Great Ormond Street Hospital for Children NHS Foundation Trust, Imperial College Healthcare NHS Trust | COVID-19 Genomics UK (COG-UK) Consortium |
| EPI_ISL_449650 | University College London, Great Ormond Street Hospital for Children NHS Foundation Trust, Imperial College Healthcare NHS Trust | COVID-19 Genomics UK (COG-UK) Consortium |
| EPI_ISL_449651 | University College London, Great Ormond Street Hospital for Children NHS Foundation Trust, Imperial College Healthcare NHS Trust | COVID-19 Genomics UK (COG-UK) Consortium |
| EPI_ISL_449652 | University College London, Great Ormond Street Hospital for Children NHS Foundation Trust, Imperial College Healthcare NHS Trust | COVID-19 Genomics UK (COG-UK) Consortium |
| EPI_ISL_449653 | University College London, Great Ormond Street Hospital for Children NHS Foundation Trust, Imperial College Healthcare NHS Trust | COVID-19 Genomics UK (COG-UK) Consortium |
| EPI_ISL_449654 | University College London, Great Ormond Street Hospital for Children NHS Foundation Trust, Imperial College Healthcare NHS Trust | COVID-19 Genomics UK (COG-UK) Consortium |
| EPI_ISL_449655 | University College London, Great Ormond Street Hospital for Children NHS Foundation Trust, Imperial College Healthcare NHS Trust | COVID-19 Genomics UK (COG-UK) Consortium |

| EPI_Accession_ID | Originating Laboratory | Submitting Laboratory |
|---|---|---|
| EPI_ISL_449656 | University College London, Great Ormond Street Hospital for Children NHS Foundation Trust, Imperial College Healthcare NHS Trust | COVID-19 Genomics UK (COG-UK) Consortium |
| EPI_ISL_449657 | University College London, Great Ormond Street Hospital for Children NHS Foundation Trust, Imperial College Healthcare NHS Trust | COVID-19 Genomics UK (COG-UK) Consortium |
| EPI_ISL_449658 | University College London, Great Ormond Street Hospital for Children NHS Foundation Trust, Imperial College Healthcare NHS Trust | COVID-19 Genomics UK (COG-UK) Consortium |
| EPI_ISL_449659 | University College London, Great Ormond Street Hospital for Children NHS Foundation Trust, Imperial College Healthcare NHS Trust | COVID-19 Genomics UK (COG-UK) Consortium |
| EPI_ISL_449660 | University College London, Great Ormond Street Hospital for Children NHS Foundation Trust, Imperial College Healthcare NHS Trust | COVID-19 Genomics UK (COG-UK) Consortium |
| EPI_ISL_449661 | University College London, Great Ormond Street Hospital for Children NHS Foundation Trust, Imperial College Healthcare NHS Trust | COVID-19 Genomics UK (COG-UK) Consortium |
| EPI_ISL_449662 | University College London, Great Ormond Street Hospital for Children NHS Foundation Trust, Imperial College Healthcare NHS Trust | COVID-19 Genomics UK (COG-UK) Consortium |
| EPI_ISL_449663 | University College London, Great Ormond Street Hospital for Children NHS Foundation Trust, Imperial College Healthcare NHS Trust | COVID-19 Genomics UK (COG-UK) Consortium |
| EPI_ISL_449664 | University College London, Great Ormond Street Hospital for Children NHS Foundation Trust, Imperial College Healthcare NHS Trust | COVID-19 Genomics UK (COG-UK) Consortium |
| EPI_ISL_449665 | University College London, Great Ormond Street Hospital for Children NHS Foundation Trust, Imperial College Healthcare NHS Trust | COVID-19 Genomics UK (COG-UK) Consortium |
| EPI_ISL_449666 | University College London, Great Ormond Street Hospital for Children NHS Foundation Trust, Imperial College Healthcare NHS Trust | COVID-19 Genomics UK (COG-UK) Consortium |

| EPI_Accession_ID | Originating Laboratory | Submitting Laboratory |
|---|---|---|
| EPI_ISL_449667 | University College London, Great Ormond Street Hospital for Children NHS Foundation Trust, Imperial College Healthcare NHS Trust | COVID-19 Genomics UK (COG-UK) Consortium |
| EPI_ISL_449668 | University College London, Great Ormond Street Hospital for Children NHS Foundation Trust, Imperial College Healthcare NHS Trust | COVID-19 Genomics UK (COG-UK) Consortium |
| EPI_ISL_449669 | University College London, Great Ormond Street Hospital for Children NHS Foundation Trust, Imperial College Healthcare NHS Trust | COVID-19 Genomics UK (COG-UK) Consortium |
| EPI_ISL_449670 | University College London, Great Ormond Street Hospital for Children NHS Foundation Trust, Imperial College Healthcare NHS Trust | COVID-19 Genomics UK (COG-UK) Consortium |
| EPI_ISL_449671 | University College London, Great Ormond Street Hospital for Children NHS Foundation Trust, Imperial College Healthcare NHS Trust | COVID-19 Genomics UK (COG-UK) Consortium |
| EPI_ISL_449672 | University College London, Great Ormond Street Hospital for Children NHS Foundation Trust, Imperial College Healthcare NHS Trust | COVID-19 Genomics UK (COG-UK) Consortium |
| EPI_ISL_449673 | University College London, Great Ormond Street Hospital for Children NHS Foundation Trust, Imperial College Healthcare NHS Trust | COVID-19 Genomics UK (COG-UK) Consortium |
| EPI_ISL_449674 | University College London, Great Ormond Street Hospital for Children NHS Foundation Trust, Imperial College Healthcare NHS Trust | COVID-19 Genomics UK (COG-UK) Consortium |
| EPI_ISL_449675 | University College London, Great Ormond Street Hospital for Children NHS Foundation Trust, Imperial College Healthcare NHS Trust | COVID-19 Genomics UK (COG-UK) Consortium |
| EPI_ISL_449676 | University College London, Great Ormond Street Hospital for Children NHS Foundation Trust, Imperial College Healthcare NHS Trust | COVID-19 Genomics UK (COG-UK) Consortium |
| EPI_ISL_449677 | University College London, Great Ormond Street Hospital for Children NHS Foundation Trust, Imperial College Healthcare NHS Trust | COVID-19 Genomics UK (COG-UK) Consortium |

| EPI_Accession_ID | Originating Laboratory | Submitting Laboratory |
|---|---|---|
| EPI_ISL_449678 | University College London, Great Ormond Street Hospital for Children NHS Foundation Trust, Imperial College Healthcare NHS Trust | COVID-19 Genomics UK (COG-UK) Consortium |
| EPI_ISL_449679 | University College London, Great Ormond Street Hospital for Children NHS Foundation Trust, Imperial College Healthcare NHS Trust | COVID-19 Genomics UK (COG-UK) Consortium |
| EPI_ISL_449680 | University College London, Great Ormond Street Hospital for Children NHS Foundation Trust, Imperial College Healthcare NHS Trust | COVID-19 Genomics UK (COG-UK) Consortium |
| EPI_ISL_449681 | University College London, Great Ormond Street Hospital for Children NHS Foundation Trust, Imperial College Healthcare NHS Trust | COVID-19 Genomics UK (COG-UK) Consortium |
| EPI_ISL_449682 | University College London, Great Ormond Street Hospital for Children NHS Foundation Trust, Imperial College Healthcare NHS Trust | COVID-19 Genomics UK (COG-UK) Consortium |
| EPI_ISL_449683 | University College London, Great Ormond Street Hospital for Children NHS Foundation Trust, Imperial College Healthcare NHS Trust | COVID-19 Genomics UK (COG-UK) Consortium |
| EPI_ISL_449684 | University College London, Great Ormond Street Hospital for Children NHS Foundation Trust, Imperial College Healthcare NHS Trust | COVID-19 Genomics UK (COG-UK) Consortium |
| EPI_ISL_449685 | University College London, Great Ormond Street Hospital for Children NHS Foundation Trust, Imperial College Healthcare NHS Trust | COVID-19 Genomics UK (COG-UK) Consortium |
| EPI_ISL_449686 | University College London, Great Ormond Street Hospital for Children NHS Foundation Trust, Imperial College Healthcare NHS Trust | COVID-19 Genomics UK (COG-UK) Consortium |
| EPI_ISL_449687 | University College London, Great Ormond Street Hospital for Children NHS Foundation Trust, Imperial College Healthcare NHS Trust | COVID-19 Genomics UK (COG-UK) Consortium |
| EPI_ISL_449688 | University College London, Great Ormond Street Hospital for Children NHS Foundation Trust, Imperial College Healthcare NHS Trust | COVID-19 Genomics UK (COG-UK) Consortium |

| EPI_Accession_ID | Originating Laboratory | Submitting Laboratory |
|---|---|---|
| EPI_ISL_449689 | University College London, Great Ormond Street Hospital for Children NHS Foundation Trust, Imperial College Healthcare NHS Trust | COVID-19 Genomics UK (COG-UK) Consortium |
| EPI_ISL_449690 | University College London, Great Ormond Street Hospital for Children NHS Foundation Trust, Imperial College Healthcare NHS Trust | COVID-19 Genomics UK (COG-UK) Consortium |
| EPI_ISL_449691 | University College London, Great Ormond Street Hospital for Children NHS Foundation Trust, Imperial College Healthcare NHS Trust | COVID-19 Genomics UK (COG-UK) Consortium |
| EPI_ISL_449692 | University College London, Great Ormond Street Hospital for Children NHS Foundation Trust, Imperial College Healthcare NHS Trust | COVID-19 Genomics UK (COG-UK) Consortium |
| EPI_ISL_449693 | University College London, Great Ormond Street Hospital for Children NHS Foundation Trust, Imperial College Healthcare NHS Trust | COVID-19 Genomics UK (COG-UK) Consortium |
| EPI_ISL_449694 | University College London, Great Ormond Street Hospital for Children NHS Foundation Trust, Imperial College Healthcare NHS Trust | COVID-19 Genomics UK (COG-UK) Consortium |
| EPI_ISL_449695 | University College London, Great Ormond Street Hospital for Children NHS Foundation Trust, Imperial College Healthcare NHS Trust | COVID-19 Genomics UK (COG-UK) Consortium |
| EPI_ISL_449696 | University College London, Great Ormond Street Hospital for Children NHS Foundation Trust, Imperial College Healthcare NHS Trust | COVID-19 Genomics UK (COG-UK) Consortium |
| EPI_ISL_449697 | University College London, Great Ormond Street Hospital for Children NHS Foundation Trust, Imperial College Healthcare NHS Trust | COVID-19 Genomics UK (COG-UK) Consortium |
| EPI_ISL_449698 | University College London, Great Ormond Street Hospital for Children NHS Foundation Trust, Imperial College Healthcare NHS Trust | COVID-19 Genomics UK (COG-UK) Consortium |
| EPI_ISL_449699 | University College London, Great Ormond Street Hospital for Children NHS Foundation Trust, Imperial College Healthcare NHS Trust | COVID-19 Genomics UK (COG-UK) Consortium |

| EPI_Accession_ID | Originating Laboratory | Submitting Laboratory |
| --- | --- | --- |
| EPI_ISL_449700 | University College London, Great Ormond Street Hospital for Children NHS Foundation Trust, Imperial College Healthcare NHS Trust | COVID-19 Genomics UK (COG-UK) Consortium |
| EPI_ISL_449701 | University College London, Great Ormond Street Hospital for Children NHS Foundation Trust, Imperial College Healthcare NHS Trust | COVID-19 Genomics UK (COG-UK) Consortium |
| EPI_ISL_449702 | University College London, Great Ormond Street Hospital for Children NHS Foundation Trust, Imperial College Healthcare NHS Trust | COVID-19 Genomics UK (COG-UK) Consortium |
| EPI_ISL_449703 | University College London, Great Ormond Street Hospital for Children NHS Foundation Trust, Imperial College Healthcare NHS Trust | COVID-19 Genomics UK (COG-UK) Consortium |
| EPI_ISL_449704 | University College London, Great Ormond Street Hospital for Children NHS Foundation Trust, Imperial College Healthcare NHS Trust | COVID-19 Genomics UK (COG-UK) Consortium |
| EPI_ISL_449705 | University College London, Great Ormond Street Hospital for Children NHS Foundation Trust, Imperial College Healthcare NHS Trust | COVID-19 Genomics UK (COG-UK) Consortium |
| EPI_ISL_449706 | University College London, Great Ormond Street Hospital for Children NHS Foundation Trust, Imperial College Healthcare NHS Trust | COVID-19 Genomics UK (COG-UK) Consortium |
| EPI_ISL_449707 | University College London, Great Ormond Street Hospital for Children NHS Foundation Trust, Imperial College Healthcare NHS Trust | COVID-19 Genomics UK (COG-UK) Consortium |
| EPI_ISL_449708 | University College London, Great Ormond Street Hospital for Children NHS Foundation Trust, Imperial College Healthcare NHS Trust | COVID-19 Genomics UK (COG-UK) Consortium |
| EPI_ISL_449709 | University College London, Great Ormond Street Hospital for Children NHS Foundation Trust, Imperial College Healthcare NHS Trust | COVID-19 Genomics UK (COG-UK) Consortium |
| EPI_ISL_449710 | University College London, Great Ormond Street Hospital for Children NHS Foundation Trust, Imperial College Healthcare NHS Trust | COVID-19 Genomics UK (COG-UK) Consortium |

| EPI_Accession_ID | Originating Laboratory | Submitting Laboratory |
|---|---|---|
| EPI_ISL_449711 | University College London, Great Ormond Street Hospital for Children NHS Foundation Trust, Imperial College Healthcare NHS Trust | COVID-19 Genomics UK (COG-UK) Consortium |
| EPI_ISL_449712 | University College London, Great Ormond Street Hospital for Children NHS Foundation Trust, Imperial College Healthcare NHS Trust | COVID-19 Genomics UK (COG-UK) Consortium |
| EPI_ISL_449713 | University College London, Great Ormond Street Hospital for Children NHS Foundation Trust, Imperial College Healthcare NHS Trust | COVID-19 Genomics UK (COG-UK) Consortium |
| EPI_ISL_449714 | University College London, Great Ormond Street Hospital for Children NHS Foundation Trust, Imperial College Healthcare NHS Trust | COVID-19 Genomics UK (COG-UK) Consortium |
| EPI_ISL_449715 | University College London, Great Ormond Street Hospital for Children NHS Foundation Trust, Imperial College Healthcare NHS Trust | COVID-19 Genomics UK (COG-UK) Consortium |
| EPI_ISL_449716 | University College London, Great Ormond Street Hospital for Children NHS Foundation Trust, Imperial College Healthcare NHS Trust | COVID-19 Genomics UK (COG-UK) Consortium |
| EPI_ISL_449717 | University College London, Great Ormond Street Hospital for Children NHS Foundation Trust, Imperial College Healthcare NHS Trust | COVID-19 Genomics UK (COG-UK) Consortium |
| EPI_ISL_449718 | University College London, Great Ormond Street Hospital for Children NHS Foundation Trust, Imperial College Healthcare NHS Trust | COVID-19 Genomics UK (COG-UK) Consortium |
| EPI_ISL_449719 | University College London, Great Ormond Street Hospital for Children NHS Foundation Trust, Imperial College Healthcare NHS Trust | COVID-19 Genomics UK (COG-UK) Consortium |
| EPI_ISL_449720 | University College London, Great Ormond Street Hospital for Children NHS Foundation Trust, Imperial College Healthcare NHS Trust | COVID-19 Genomics UK (COG-UK) Consortium |
| EPI_ISL_449721 | University College London, Great Ormond Street Hospital for Children NHS Foundation Trust, Imperial College Healthcare NHS Trust | COVID-19 Genomics UK (COG-UK) Consortium |

| EPI_Accession_ID | Originating Laboratory | Submitting Laboratory |
|---|---|---|
| EPI_ISL_449722 | University College London, Great Ormond Street Hospital for Children NHS Foundation Trust, Imperial College Healthcare NHS Trust | COVID-19 Genomics UK (COG-UK) Consortium |
| EPI_ISL_449723 | University College London, Great Ormond Street Hospital for Children NHS Foundation Trust, Imperial College Healthcare NHS Trust | COVID-19 Genomics UK (COG-UK) Consortium |
| EPI_ISL_449724 | University College London, Great Ormond Street Hospital for Children NHS Foundation Trust, Imperial College Healthcare NHS Trust | COVID-19 Genomics UK (COG-UK) Consortium |
| EPI_ISL_449725 | University College London, Great Ormond Street Hospital for Children NHS Foundation Trust, Imperial College Healthcare NHS Trust | COVID-19 Genomics UK (COG-UK) Consortium |
| EPI_ISL_449726 | University College London, Great Ormond Street Hospital for Children NHS Foundation Trust, Imperial College Healthcare NHS Trust | COVID-19 Genomics UK (COG-UK) Consortium |
| EPI_ISL_449727 | University College London, Great Ormond Street Hospital for Children NHS Foundation Trust, Imperial College Healthcare NHS Trust | COVID-19 Genomics UK (COG-UK) Consortium |
| EPI_ISL_449728 | University College London, Great Ormond Street Hospital for Children NHS Foundation Trust, Imperial College Healthcare NHS Trust | COVID-19 Genomics UK (COG-UK) Consortium |
| EPI_ISL_449729 | University College London, Great Ormond Street Hospital for Children NHS Foundation Trust, Imperial College Healthcare NHS Trust | COVID-19 Genomics UK (COG-UK) Consortium |
| EPI_ISL_449786 | Ostfold Hospital Trust - Kalnes, Centre for Laboratory Medicine, Section for gene technology and infection serology | Norwegian Institute of Public Health, Department of Virology |
| EPI_ISL_449787 | Furst Medical Laboratory | Norwegian Institute of Public Health, Department of Virology |
| EPI_ISL_449788 | Furst Medical Laboratory | Norwegian Institute of Public Health, Department of Virology |
| EPI_ISL_449789 | Dept. of Medical Microbiology, Stavanger University Hospital, Helse Stavanger HF | Norwegian Institute of Public Health, Department of Virology |

| EPI_Accession_ID | Originating Laboratory | Submitting Laboratory |
|---|---|---|
| EPI_ISL_449790 | Dept. of Medical Microbiology, Stavanger University Hospital, Helse Stavanger HF | Norwegian Institute of Public Health, Department of Virology |
| EPI_ISL_449791 | Dept. of Medical Microbiology, Stavanger University Hospital, Helse Stavanger HF | Norwegian Institute of Public Health, Department of Virology |
| EPI_ISL_449792 | Dept. of Medical Microbiology, Stavanger University Hospital, Helse Stavanger HF | Norwegian Institute of Public Health, Department of Virology |
| EPI_ISL_449793 | Dept. of Medical Microbiology, Stavanger University Hospital, Helse Stavanger HF | Norwegian Institute of Public Health, Department of Virology |
| EPI_ISL_449794 | Dept. of Medical Microbiology, Stavanger University Hospital, Helse Stavanger HF | Norwegian Institute of Public Health, Department of Virology |
| EPI_ISL_449799 | National Laboratory for Health, Environment and Food | National Laboratory for Health, Environment and Food |
| EPI_ISL_449800 | HOSPITAL SAN JUAN DE DIOS | Instituto de Salud Publica de Chile |
| EPI_ISL_449801 | Utah Public Health Laboratory | Utah Public Health Laboratory |
| EPI_ISL_449802 | Utah Public Health Laboratory | Utah Public Health Laboratory |
| EPI_ISL_449803 | Utah Public Health Laboratory | Utah Public Health Laboratory |
| EPI_ISL_449804 | Utah Public Health Laboratory | Utah Public Health Laboratory |
| EPI_ISL_449805 | Utah Public Health Laboratory | Utah Public Health Laboratory |
| EPI_ISL_449806 | Utah Public Health Laboratory | Utah Public Health Laboratory |
| EPI_ISL_449807 | Utah Public Health Laboratory | Utah Public Health Laboratory |
| EPI_ISL_449808 | Utah Public Health Laboratory | Utah Public Health Laboratory |
| EPI_ISL_449809 | Utah Public Health Laboratory | Utah Public Health Laboratory |
| EPI_ISL_449810 | Utah Public Health Laboratory | Utah Public Health Laboratory |
| EPI_ISL_449811 | Utah Public Health Laboratory | Utah Public Health Laboratory |
| EPI_ISL_449812 | Utah Public Health Laboratory | Utah Public Health Laboratory |
| EPI_ISL_449813 | Utah Public Health Laboratory | Utah Public Health Laboratory |
| EPI_ISL_449814 | Utah Public Health Laboratory | Utah Public Health Laboratory |
| EPI_ISL_449815 | Utah Public Health Laboratory | Utah Public Health Laboratory |
| EPI_ISL_449816 | Utah Public Health Laboratory | Utah Public Health Laboratory |
| EPI_ISL_449817 | Utah Public Health Laboratory | Utah Public Health Laboratory |
| EPI_ISL_449818 | Utah Public Health Laboratory | Utah Public Health Laboratory |
| EPI_ISL_449819 | Utah Public Health Laboratory | Utah Public Health Laboratory |
| EPI_ISL_449820 | Utah Public Health Laboratory | Utah Public Health Laboratory |
| EPI_ISL_449821 | Utah Public Health Laboratory | Utah Public Health Laboratory |

| EPI_Accession_ID | Originating Laboratory | Submitting Laboratory |
|---|---|---|
| EPI_ISL_449822 | Utah Public Health Laboratory | Utah Public Health Laboratory |
| EPI_ISL_449823 | Utah Public Health Laboratory | Utah Public Health Laboratory |
| EPI_ISL_449824 | Utah Public Health Laboratory | Utah Public Health Laboratory |
| EPI_ISL_449825 | Utah Public Health Laboratory | Utah Public Health Laboratory |
| EPI_ISL_449826 | Utah Public Health Laboratory | Utah Public Health Laboratory |
| EPI_ISL_449827 | Utah Public Health Laboratory | Utah Public Health Laboratory |
| EPI_ISL_449828 | Utah Public Health Laboratory | Utah Public Health Laboratory |
| EPI_ISL_449829 | Utah Public Health Laboratory | Utah Public Health Laboratory |
| EPI_ISL_449830 | Utah Public Health Laboratory | Utah Public Health Laboratory |
| EPI_ISL_449831 | Utah Public Health Laboratory | Utah Public Health Laboratory |
| EPI_ISL_449832 | Utah Public Health Laboratory | Utah Public Health Laboratory |
| EPI_ISL_449833 | Utah Public Health Laboratory | Utah Public Health Laboratory |
| EPI_ISL_449834 | Utah Public Health Laboratory | Utah Public Health Laboratory |
| EPI_ISL_449835 | Utah Public Health Laboratory | Utah Public Health Laboratory |
| EPI_ISL_449836 | Utah Public Health Laboratory | Utah Public Health Laboratory |
| EPI_ISL_449837 | Utah Public Health Laboratory | Utah Public Health Laboratory |
| EPI_ISL_449838 | Utah Public Health Laboratory | Utah Public Health Laboratory |
| EPI_ISL_449839 | Washington State Department of Health | Seattle Flu Study |
| EPI_ISL_449840 | Washington State Department of Health | Seattle Flu Study |
| EPI_ISL_449841 | Washington State Department of Health | Seattle Flu Study |
| EPI_ISL_449842 | Washington State Department of Health | Seattle Flu Study |
| EPI_ISL_449843 | Washington State Department of Health | Seattle Flu Study |
| EPI_ISL_449844 | Washington State Department of Health | Seattle Flu Study |
| EPI_ISL_449845 | Washington State Department of Health | Seattle Flu Study |
| EPI_ISL_449846 | Washington State Department of Health | Seattle Flu Study |
| EPI_ISL_449847 | Washington State Department of Health | Seattle Flu Study |
| EPI_ISL_449848 | Washington State Department of Health | Seattle Flu Study |
| EPI_ISL_449849 | Washington State Department of Health | Seattle Flu Study |
| EPI_ISL_449850 | Washington State Department of Health | Seattle Flu Study |
| EPI_ISL_449851 | Washington State Department of Health | Seattle Flu Study |
| EPI_ISL_449852 | Washington State Department of Health | Seattle Flu Study |
| EPI_ISL_449853 | Washington State Department of Health | Seattle Flu Study |
| EPI_ISL_449854 | Washington State Department of Health | Seattle Flu Study |

| EPI_Accession_ID | Originating Laboratory | Submitting Laboratory |
|---|---|---|
| EPI_ISL_449855 | Washington State Department of Health | Seattle Flu Study |
| EPI_ISL_449856 | Washington State Department of Health | Seattle Flu Study |
| EPI_ISL_449857 | Washington State Department of Health | Seattle Flu Study |
| EPI_ISL_449858 | Washington State Department of Health | Seattle Flu Study |
| EPI_ISL_449859 | Washington State Department of Health | Seattle Flu Study |
| EPI_ISL_449860 | Washington State Department of Health | Seattle Flu Study |
| EPI_ISL_449861 | Washington State Department of Health | Seattle Flu Study |
| EPI_ISL_449862 | Washington State Department of Health | Seattle Flu Study |
| EPI_ISL_449863 | Washington State Department of Health | Seattle Flu Study |
| EPI_ISL_449864 | Washington State Department of Health | Seattle Flu Study |
| EPI_ISL_449865 | Washington State Department of Health | Seattle Flu Study |
| EPI_ISL_449866 | Washington State Department of Health | Seattle Flu Study |
| EPI_ISL_449867 | Washington State Department of Health | Seattle Flu Study |
| EPI_ISL_449868 | Washington State Department of Health | Seattle Flu Study |
| EPI_ISL_449869 | Washington State Department of Health | Seattle Flu Study |
| EPI_ISL_449870 | Washington State Department of Health | Seattle Flu Study |
| EPI_ISL_449871 | Washington State Department of Health | Seattle Flu Study |
| EPI_ISL_449872 | Washington State Department of Health | Seattle Flu Study |
| EPI_ISL_449873 | Washington State Department of Health | Seattle Flu Study |
| EPI_ISL_449874 | Washington State Department of Health | Seattle Flu Study |
| EPI_ISL_449875 | Washington State Department of Health | Seattle Flu Study |
| EPI_ISL_449876 | Washington State Department of Health | Seattle Flu Study |
| EPI_ISL_449877 | Washington State Department of Health | Seattle Flu Study |
| EPI_ISL_449878 | Washington State Department of Health | Seattle Flu Study |
| EPI_ISL_449879 | Washington State Department of Health | Seattle Flu Study |
| EPI_ISL_449880 | Washington State Department of Health | Seattle Flu Study |
| EPI_ISL_449881 | Washington State Department of Health | Seattle Flu Study |
| EPI_ISL_449882 | Washington State Department of Health | Seattle Flu Study |
| EPI_ISL_449883 | Washington State Department of Health | Seattle Flu Study |
| EPI_ISL_449884 | Washington State Department of Health | Seattle Flu Study |
| EPI_ISL_449885 | Washington State Department of Health | Seattle Flu Study |
| EPI_ISL_449886 | Washington State Department of Health | Seattle Flu Study |
| EPI_ISL_449887 | Washington State Department of Health | Seattle Flu Study |

| EPI_Accession_ID | Originating Laboratory | Submitting Laboratory |
| --- | --- | --- |
| EPI_ISL_449888 | Washington State Department of Health | Seattle Flu Study |
| EPI_ISL_449889 | Washington State Department of Health | Seattle Flu Study |
| EPI_ISL_449890 | Washington State Department of Health | Seattle Flu Study |
| EPI_ISL_449891 | Washington State Department of Health | Seattle Flu Study |
| EPI_ISL_449892 | Washington State Department of Health | Seattle Flu Study |
| EPI_ISL_449893 | Washington State Department of Health | Seattle Flu Study |
| EPI_ISL_449894 | Washington State Department of Health | Seattle Flu Study |
| EPI_ISL_449895 | Washington State Department of Health | Seattle Flu Study |
| EPI_ISL_449896 | Washington State Department of Health | Seattle Flu Study |
| EPI_ISL_449897 | Washington State Department of Health | Seattle Flu Study |
| EPI_ISL_449898 | Washington State Department of Health | Seattle Flu Study |
| EPI_ISL_449899 | Washington State Department of Health | Seattle Flu Study |
| EPI_ISL_449900 | Washington State Department of Health | Seattle Flu Study |
| EPI_ISL_449901 | Washington State Department of Health | Seattle Flu Study |
| EPI_ISL_449902 | Washington State Department of Health | Seattle Flu Study |
| EPI_ISL_449903 | Washington State Department of Health | Seattle Flu Study |
| EPI_ISL_449904 | Washington State Department of Health | Seattle Flu Study |
| EPI_ISL_449905 | Washington State Department of Health | Seattle Flu Study |
| EPI_ISL_449906 | Washington State Department of Health | Seattle Flu Study |
| EPI_ISL_449907 | Washington State Department of Health | Seattle Flu Study |
| EPI_ISL_449908 | Washington State Department of Health | Seattle Flu Study |
| EPI_ISL_449909 | Washington State Department of Health | Seattle Flu Study |
| EPI_ISL_449910 | Washington State Department of Health | Seattle Flu Study |
| EPI_ISL_449911 | Washington State Department of Health | Seattle Flu Study |
| EPI_ISL_449912 | Washington State Department of Health | Seattle Flu Study |
| EPI_ISL_449913 | Washington State Department of Health | Seattle Flu Study |
| EPI_ISL_449914 | Washington State Department of Health | Seattle Flu Study |
| EPI_ISL_449915 | Washington State Department of Health | Seattle Flu Study |
| EPI_ISL_449916 | Washington State Department of Health | Seattle Flu Study |
| EPI_ISL_449917 | Washington State Department of Health | Seattle Flu Study |
| EPI_ISL_449918 | Washington State Department of Health | Seattle Flu Study |
| EPI_ISL_449919 | Washington State Department of Health | Seattle Flu Study |
| EPI_ISL_449920 | Washington State Department of Health | Seattle Flu Study |

| EPI_Accession_ID | Originating Laboratory | Submitting Laboratory |
|---|---|---|
| EPI_ISL_449921 | Washington State Department of Health | Seattle Flu Study |
| EPI_ISL_449922 | Washington State Department of Health | Seattle Flu Study |
| EPI_ISL_449923 | Washington State Department of Health | Seattle Flu Study |
| EPI_ISL_449924 | Washington State Department of Health | Seattle Flu Study |
| EPI_ISL_449925 | Washington State Department of Health | Seattle Flu Study |
| EPI_ISL_449926 | Washington State Department of Health | Seattle Flu Study |
| EPI_ISL_449927 | Washington State Department of Health | Seattle Flu Study |
| EPI_ISL_449928 | Washington State Department of Health | Seattle Flu Study |
| EPI_ISL_449929 | Washington State Department of Health | Seattle Flu Study |
| EPI_ISL_449930 | Washington State Department of Health | Seattle Flu Study |
| EPI_ISL_449931 | Washington State Department of Health | Seattle Flu Study |
| EPI_ISL_449932 | Washington State Department of Health | Seattle Flu Study |
| EPI_ISL_449933 | Washington State Department of Health | Seattle Flu Study |
| EPI_ISL_449934 | Washington State Department of Health | Seattle Flu Study |
| EPI_ISL_449935 | Washington State Department of Health | Seattle Flu Study |
| EPI_ISL_449936 | Washington State Department of Health | Seattle Flu Study |
| EPI_ISL_449937 | Washington State Department of Health | Seattle Flu Study |
| EPI_ISL_449938 | Washington State Department of Health | Seattle Flu Study |
| EPI_ISL_449939 | Washington State Department of Health | Seattle Flu Study |
| EPI_ISL_449940 | Washington State Department of Health | Seattle Flu Study |
| EPI_ISL_449941 | Washington State Department of Health | Seattle Flu Study |
| EPI_ISL_449942 | Washington State Department of Health | Seattle Flu Study |
| EPI_ISL_449943 | Washington State Department of Health | Seattle Flu Study |
| EPI_ISL_449944 | Washington State Department of Health | Seattle Flu Study |
| EPI_ISL_449945 | Washington State Department of Health | Seattle Flu Study |
| EPI_ISL_449946 | Washington State Department of Health | Seattle Flu Study |
| EPI_ISL_449947 | Washington State Department of Health | Seattle Flu Study |
| EPI_ISL_449948 | Washington State Department of Health | Seattle Flu Study |
| EPI_ISL_449949 | Washington State Department of Health | Seattle Flu Study |
| EPI_ISL_449950 | Washington State Department of Health | Seattle Flu Study |
| EPI_ISL_449951 | Washington State Department of Health | Seattle Flu Study |
| EPI_ISL_449952 | Washington State Department of Health | Seattle Flu Study |
| EPI_ISL_449953 | Washington State Department of Health | Seattle Flu Study |

| EPI_Accession_ID | Originating Laboratory | Submitting Laboratory |
|---|---|---|
| EPI_ISL_449954 | Washington State Department of Health | Seattle Flu Study |
| EPI_ISL_449955 | Washington State Department of Health | Seattle Flu Study |
| EPI_ISL_449956 | Washington State Department of Health | Seattle Flu Study |
| EPI_ISL_449957 | Washington State Department of Health | Seattle Flu Study |
| EPI_ISL_449958 | Washington State Department of Health | Seattle Flu Study |
| EPI_ISL_449959 | Washington State Department of Health | Seattle Flu Study |
| EPI_ISL_449960 | Washington State Department of Health | Seattle Flu Study |
| EPI_ISL_449961 | Washington State Department of Health | Seattle Flu Study |
| EPI_ISL_449962 | Washington State Department of Health | Seattle Flu Study |
| EPI_ISL_449963 | Washington State Department of Health | Seattle Flu Study |
| EPI_ISL_449964 | Washington State Department of Health | Seattle Flu Study |
| EPI_ISL_449965 | Washington State Department of Health | Seattle Flu Study |
| EPI_ISL_449966 | Washington State Department of Health | Seattle Flu Study |
| EPI_ISL_449967 | Washington State Department of Health | Seattle Flu Study |
| EPI_ISL_449968 | Washington State Department of Health | Seattle Flu Study |
| EPI_ISL_449969 | Washington State Department of Health | Seattle Flu Study |
| EPI_ISL_449970 | Washington State Department of Health | Seattle Flu Study |
| EPI_ISL_449971 | Washington State Department of Health | Seattle Flu Study |
| EPI_ISL_449972 | Washington State Department of Health | Seattle Flu Study |
| EPI_ISL_449973 | Washington State Department of Health | Seattle Flu Study |
| EPI_ISL_449974 | Washington State Department of Health | Seattle Flu Study |
| EPI_ISL_449975 | Washington State Department of Health | Seattle Flu Study |
| EPI_ISL_449976 | Washington State Department of Health | Seattle Flu Study |
| EPI_ISL_449977 | Washington State Department of Health | Seattle Flu Study |
| EPI_ISL_449978 | Washington State Department of Health | Seattle Flu Study |
| EPI_ISL_449979 | Washington State Department of Health | Seattle Flu Study |
| EPI_ISL_449980 | Washington State Department of Health | Seattle Flu Study |
| EPI_ISL_449981 | Washington State Department of Health | Seattle Flu Study |
| EPI_ISL_449982 | Washington State Department of Health | Seattle Flu Study |
| EPI_ISL_449983 | Washington State Department of Health | Seattle Flu Study |
| EPI_ISL_449984 | Washington State Department of Health | Seattle Flu Study |
| EPI_ISL_449985 | Washington State Department of Health | Seattle Flu Study |
| EPI_ISL_449986 | Washington State Department of Health | Seattle Flu Study |

| EPI_Accession_ID | Originating Laboratory | Submitting Laboratory |
| --- | --- | --- |
| EPI_ISL_449987 | Washington State Department of Health | Seattle Flu Study |
| EPI_ISL_449988 | Washington State Department of Health | Seattle Flu Study |
| EPI_ISL_449989 | Washington State Department of Health | Seattle Flu Study |
| EPI_ISL_449990 | Washington State Department of Health | Seattle Flu Study |
| EPI_ISL_449991 | Washington State Department of Health | Seattle Flu Study |
| EPI_ISL_449992 | Washington State Department of Health | Seattle Flu Study |
| EPI_ISL_449993 | Washington State Department of Health | Seattle Flu Study |
| EPI_ISL_449994 | Washington State Department of Health | Seattle Flu Study |
| EPI_ISL_449995 | Washington State Department of Health | Seattle Flu Study |
| EPI_ISL_449996 | Washington State Department of Health | Seattle Flu Study |
| EPI_ISL_449997 | Washington State Department of Health | Seattle Flu Study |
| EPI_ISL_449998 | Washington State Department of Health | Seattle Flu Study |
| EPI_ISL_449999 | Washington State Department of Health | Seattle Flu Study |
| EPI_ISL_450000 | Washington State Department of Health | Seattle Flu Study |
| EPI_ISL_450001 | Washington State Department of Health | Seattle Flu Study |
| EPI_ISL_450002 | Washington State Department of Health | Seattle Flu Study |
| EPI_ISL_450003 | Washington State Department of Health | Seattle Flu Study |
| EPI_ISL_450004 | MSHS Clinical Microbiology Laboratories | MSHS Pathogen Surveillance Program |
| EPI_ISL_450005 | MSHS Clinical Microbiology Laboratories | MSHS Pathogen Surveillance Program |
| EPI_ISL_450006 | MSHS Clinical Microbiology Laboratories | MSHS Pathogen Surveillance Program |
| EPI_ISL_450007 | MSHS Clinical Microbiology Laboratories | MSHS Pathogen Surveillance Program |
| EPI_ISL_450008 | MSHS Clinical Microbiology Laboratories | MSHS Pathogen Surveillance Program |
| EPI_ISL_450009 | MSHS Clinical Microbiology Laboratories | MSHS Pathogen Surveillance Program |
| EPI_ISL_450010 | MSHS Clinical Microbiology Laboratories | MSHS Pathogen Surveillance Program |
| EPI_ISL_450011 | MSHS Clinical Microbiology Laboratories | MSHS Pathogen Surveillance Program |
| EPI_ISL_450012 | MSHS Clinical Microbiology Laboratories | MSHS Pathogen Surveillance Program |
| EPI_ISL_450013 | MSHS Clinical Microbiology Laboratories | MSHS Pathogen Surveillance Program |
| EPI_ISL_450014 | MSHS Clinical Microbiology Laboratories | MSHS Pathogen Surveillance Program |
| EPI_ISL_450015 | MSHS Clinical Microbiology Laboratories | MSHS Pathogen Surveillance Program |
| EPI_ISL_450016 | MSHS Clinical Microbiology Laboratories | MSHS Pathogen Surveillance Program |
| EPI_ISL_450017 | MSHS Clinical Microbiology Laboratories | MSHS Pathogen Surveillance Program |
| EPI_ISL_450018 | MSHS Clinical Microbiology Laboratories | MSHS Pathogen Surveillance Program |
| EPI_ISL_450019 | MSHS Clinical Microbiology Laboratories | MSHS Pathogen Surveillance Program |

| EPI_Accession_ID | Originating Laboratory | Submitting Laboratory |
|---|---|---|
| EPI_ISL_450020 | MSHS Clinical Microbiology Laboratories | MSHS Pathogen Surveillance Program |
| EPI_ISL_450021 | MSHS Clinical Microbiology Laboratories | MSHS Pathogen Surveillance Program |
| EPI_ISL_450022 | MSHS Clinical Microbiology Laboratories | MSHS Pathogen Surveillance Program |
| EPI_ISL_450023 | MSHS Clinical Microbiology Laboratories | MSHS Pathogen Surveillance Program |
| EPI_ISL_450024 | MSHS Clinical Microbiology Laboratories | MSHS Pathogen Surveillance Program |
| EPI_ISL_450025 | MSHS Clinical Microbiology Laboratories | MSHS Pathogen Surveillance Program |
| EPI_ISL_450026 | MSHS Clinical Microbiology Laboratories | MSHS Pathogen Surveillance Program |
| EPI_ISL_450027 | MSHS Clinical Microbiology Laboratories | MSHS Pathogen Surveillance Program |
| EPI_ISL_450028 | MSHS Clinical Microbiology Laboratories | MSHS Pathogen Surveillance Program |
| EPI_ISL_450029 | MSHS Clinical Microbiology Laboratories | MSHS Pathogen Surveillance Program |
| EPI_ISL_450030 | MSHS Clinical Microbiology Laboratories | MSHS Pathogen Surveillance Program |
| EPI_ISL_450031 | MSHS Clinical Microbiology Laboratories | MSHS Pathogen Surveillance Program |
| EPI_ISL_450032 | MSHS Clinical Microbiology Laboratories | MSHS Pathogen Surveillance Program |
| EPI_ISL_450033 | MSHS Clinical Microbiology Laboratories | MSHS Pathogen Surveillance Program |
| EPI_ISL_450034 | MSHS Clinical Microbiology Laboratories | MSHS Pathogen Surveillance Program |
| EPI_ISL_450035 | MSHS Clinical Microbiology Laboratories | MSHS Pathogen Surveillance Program |
| EPI_ISL_450036 | MSHS Clinical Microbiology Laboratories | MSHS Pathogen Surveillance Program |
| EPI_ISL_450037 | MSHS Clinical Microbiology Laboratories | MSHS Pathogen Surveillance Program |
| EPI_ISL_450038 | MSHS Clinical Microbiology Laboratories | MSHS Pathogen Surveillance Program |
| EPI_ISL_450039 | MSHS Clinical Microbiology Laboratories | MSHS Pathogen Surveillance Program |
| EPI_ISL_450040 | MSHS Clinical Microbiology Laboratories | MSHS Pathogen Surveillance Program |
| EPI_ISL_450041 | MSHS Clinical Microbiology Laboratories | MSHS Pathogen Surveillance Program |
| EPI_ISL_450042 | MSHS Clinical Microbiology Laboratories | MSHS Pathogen Surveillance Program |
| EPI_ISL_450043 | MSHS Clinical Microbiology Laboratories | MSHS Pathogen Surveillance Program |
| EPI_ISL_450044 | MSHS Clinical Microbiology Laboratories | MSHS Pathogen Surveillance Program |
| EPI_ISL_450045 | MSHS Clinical Microbiology Laboratories | MSHS Pathogen Surveillance Program |
| EPI_ISL_450046 | MSHS Clinical Microbiology Laboratories | MSHS Pathogen Surveillance Program |
| EPI_ISL_450047 | MSHS Clinical Microbiology Laboratories | MSHS Pathogen Surveillance Program |
| EPI_ISL_450048 | MSHS Clinical Microbiology Laboratories | MSHS Pathogen Surveillance Program |
| EPI_ISL_450049 | MSHS Clinical Microbiology Laboratories | MSHS Pathogen Surveillance Program |
| EPI_ISL_450050 | MSHS Clinical Microbiology Laboratories | MSHS Pathogen Surveillance Program |
| EPI_ISL_450051 | MSHS Clinical Microbiology Laboratories | MSHS Pathogen Surveillance Program |
| EPI_ISL_450052 | MSHS Clinical Microbiology Laboratories | MSHS Pathogen Surveillance Program |

| EPI_Accession_ID | Originating Laboratory | Submitting Laboratory |
|---|---|---|
| EPI_ISL_450053 | MSHS Clinical Microbiology Laboratories | MSHS Pathogen Surveillance Program |
| EPI_ISL_450054 | MSHS Clinical Microbiology Laboratories | MSHS Pathogen Surveillance Program |
| EPI_ISL_450055 | MSHS Clinical Microbiology Laboratories | MSHS Pathogen Surveillance Program |
| EPI_ISL_450056 | MSHS Clinical Microbiology Laboratories | MSHS Pathogen Surveillance Program |
| EPI_ISL_450057 | MSHS Clinical Microbiology Laboratories | MSHS Pathogen Surveillance Program |
| EPI_ISL_450058 | MSHS Clinical Microbiology Laboratories | MSHS Pathogen Surveillance Program |
| EPI_ISL_450059 | MSHS Clinical Microbiology Laboratories | MSHS Pathogen Surveillance Program |
| EPI_ISL_450060 | MSHS Clinical Microbiology Laboratories | MSHS Pathogen Surveillance Program |
| EPI_ISL_450061 | MSHS Clinical Microbiology Laboratories | MSHS Pathogen Surveillance Program |
| EPI_ISL_450062 | MSHS Clinical Microbiology Laboratories | MSHS Pathogen Surveillance Program |
| EPI_ISL_450063 | MSHS Clinical Microbiology Laboratories | MSHS Pathogen Surveillance Program |
| EPI_ISL_450064 | MSHS Clinical Microbiology Laboratories | MSHS Pathogen Surveillance Program |
| EPI_ISL_450065 | MSHS Clinical Microbiology Laboratories | MSHS Pathogen Surveillance Program |
| EPI_ISL_450066 | MSHS Clinical Microbiology Laboratories | MSHS Pathogen Surveillance Program |
| EPI_ISL_450067 | MSHS Clinical Microbiology Laboratories | MSHS Pathogen Surveillance Program |
| EPI_ISL_450068 | MSHS Clinical Microbiology Laboratories | MSHS Pathogen Surveillance Program |
| EPI_ISL_450069 | MSHS Clinical Microbiology Laboratories | MSHS Pathogen Surveillance Program |
| EPI_ISL_450070 | MSHS Clinical Microbiology Laboratories | MSHS Pathogen Surveillance Program |
| EPI_ISL_450071 | MSHS Clinical Microbiology Laboratories | MSHS Pathogen Surveillance Program |
| EPI_ISL_450072 | MSHS Clinical Microbiology Laboratories | MSHS Pathogen Surveillance Program |
| EPI_ISL_450073 | MSHS Clinical Microbiology Laboratories | MSHS Pathogen Surveillance Program |
| EPI_ISL_450074 | MSHS Clinical Microbiology Laboratories | MSHS Pathogen Surveillance Program |
| EPI_ISL_450075 | MSHS Clinical Microbiology Laboratories | MSHS Pathogen Surveillance Program |
| EPI_ISL_450076 | MSHS Clinical Microbiology Laboratories | MSHS Pathogen Surveillance Program |
| EPI_ISL_450077 | MSHS Clinical Microbiology Laboratories | MSHS Pathogen Surveillance Program |
| EPI_ISL_450078 | MSHS Clinical Microbiology Laboratories | MSHS Pathogen Surveillance Program |
| EPI_ISL_450079 | MSHS Clinical Microbiology Laboratories | MSHS Pathogen Surveillance Program |
| EPI_ISL_450080 | MSHS Clinical Microbiology Laboratories | MSHS Pathogen Surveillance Program |
| EPI_ISL_450081 | MSHS Clinical Microbiology Laboratories | MSHS Pathogen Surveillance Program |
| EPI_ISL_450082 | MSHS Clinical Microbiology Laboratories | MSHS Pathogen Surveillance Program |
| EPI_ISL_450083 | MSHS Clinical Microbiology Laboratories | MSHS Pathogen Surveillance Program |
| EPI_ISL_450084 | MSHS Clinical Microbiology Laboratories | MSHS Pathogen Surveillance Program |
| EPI_ISL_450085 | MSHS Clinical Microbiology Laboratories | MSHS Pathogen Surveillance Program |

| EPI_Accession_ID | Originating Laboratory | Submitting Laboratory |
|---|---|---|
| EPI_ISL_450086 | MSHS Clinical Microbiology Laboratories | MSHS Pathogen Surveillance Program |
| EPI_ISL_450087 | MSHS Clinical Microbiology Laboratories | MSHS Pathogen Surveillance Program |
| EPI_ISL_450088 | MSHS Clinical Microbiology Laboratories | MSHS Pathogen Surveillance Program |
| EPI_ISL_450089 | MSHS Clinical Microbiology Laboratories | MSHS Pathogen Surveillance Program |
| EPI_ISL_450090 | MSHS Clinical Microbiology Laboratories | MSHS Pathogen Surveillance Program |
| EPI_ISL_450091 | MSHS Clinical Microbiology Laboratories | MSHS Pathogen Surveillance Program |
| EPI_ISL_450092 | MSHS Clinical Microbiology Laboratories | MSHS Pathogen Surveillance Program |
| EPI_ISL_450093 | MSHS Clinical Microbiology Laboratories | MSHS Pathogen Surveillance Program |
| EPI_ISL_450094 | MSHS Clinical Microbiology Laboratories | MSHS Pathogen Surveillance Program |
| EPI_ISL_450095 | MSHS Clinical Microbiology Laboratories | MSHS Pathogen Surveillance Program |
| EPI_ISL_450096 | MSHS Clinical Microbiology Laboratories | MSHS Pathogen Surveillance Program |
| EPI_ISL_450097 | MSHS Clinical Microbiology Laboratories | MSHS Pathogen Surveillance Program |
| EPI_ISL_450098 | MSHS Clinical Microbiology Laboratories | MSHS Pathogen Surveillance Program |
| EPI_ISL_450099 | MSHS Clinical Microbiology Laboratories | MSHS Pathogen Surveillance Program |
| EPI_ISL_450100 | MSHS Clinical Microbiology Laboratories | MSHS Pathogen Surveillance Program |
| EPI_ISL_450101 | MSHS Clinical Microbiology Laboratories | MSHS Pathogen Surveillance Program |
| EPI_ISL_450102 | MSHS Clinical Microbiology Laboratories | MSHS Pathogen Surveillance Program |
| EPI_ISL_450103 | MSHS Clinical Microbiology Laboratories | MSHS Pathogen Surveillance Program |
| EPI_ISL_450104 | MSHS Clinical Microbiology Laboratories | MSHS Pathogen Surveillance Program |
| EPI_ISL_450105 | MSHS Clinical Microbiology Laboratories | MSHS Pathogen Surveillance Program |
| EPI_ISL_450106 | MSHS Clinical Microbiology Laboratories | MSHS Pathogen Surveillance Program |
| EPI_ISL_450107 | MSHS Clinical Microbiology Laboratories | MSHS Pathogen Surveillance Program |
| EPI_ISL_450108 | MSHS Clinical Microbiology Laboratories | MSHS Pathogen Surveillance Program |
| EPI_ISL_450109 | MSHS Clinical Microbiology Laboratories | MSHS Pathogen Surveillance Program |
| EPI_ISL_450110 | MSHS Clinical Microbiology Laboratories | MSHS Pathogen Surveillance Program |
| EPI_ISL_450111 | MSHS Clinical Microbiology Laboratories | MSHS Pathogen Surveillance Program |
| EPI_ISL_450112 | MSHS Clinical Microbiology Laboratories | MSHS Pathogen Surveillance Program |
| EPI_ISL_450113 | MSHS Clinical Microbiology Laboratories | MSHS Pathogen Surveillance Program |
| EPI_ISL_450114 | MSHS Clinical Microbiology Laboratories | MSHS Pathogen Surveillance Program |
| EPI_ISL_450115 | MSHS Clinical Microbiology Laboratories | MSHS Pathogen Surveillance Program |
| EPI_ISL_450116 | MSHS Clinical Microbiology Laboratories | MSHS Pathogen Surveillance Program |
| EPI_ISL_450117 | MSHS Clinical Microbiology Laboratories | MSHS Pathogen Surveillance Program |
| EPI_ISL_450118 | MSHS Clinical Microbiology Laboratories | MSHS Pathogen Surveillance Program |

| EPI_Accession_ID | Originating Laboratory | Submitting Laboratory |
|---|---|---|
| EPI_ISL_450119 | MSHS Clinical Microbiology Laboratories | MSHS Pathogen Surveillance Program |
| EPI_ISL_450120 | MSHS Clinical Microbiology Laboratories | MSHS Pathogen Surveillance Program |
| EPI_ISL_450121 | MSHS Clinical Microbiology Laboratories | MSHS Pathogen Surveillance Program |
| EPI_ISL_450122 | MSHS Clinical Microbiology Laboratories | MSHS Pathogen Surveillance Program |
| EPI_ISL_450123 | MSHS Clinical Microbiology Laboratories | MSHS Pathogen Surveillance Program |
| EPI_ISL_450124 | MSHS Clinical Microbiology Laboratories | MSHS Pathogen Surveillance Program |
| EPI_ISL_450125 | MSHS Clinical Microbiology Laboratories | MSHS Pathogen Surveillance Program |
| EPI_ISL_450126 | MSHS Clinical Microbiology Laboratories | MSHS Pathogen Surveillance Program |
| EPI_ISL_450127 | MSHS Clinical Microbiology Laboratories | MSHS Pathogen Surveillance Program |
| EPI_ISL_450128 | MSHS Clinical Microbiology Laboratories | MSHS Pathogen Surveillance Program |
| EPI_ISL_450129 | MSHS Clinical Microbiology Laboratories | MSHS Pathogen Surveillance Program |
| EPI_ISL_450130 | MSHS Clinical Microbiology Laboratories | MSHS Pathogen Surveillance Program |
| EPI_ISL_450131 | MSHS Clinical Microbiology Laboratories | MSHS Pathogen Surveillance Program |
| EPI_ISL_450132 | MSHS Clinical Microbiology Laboratories | MSHS Pathogen Surveillance Program |
| EPI_ISL_450133 | MSHS Clinical Microbiology Laboratories | MSHS Pathogen Surveillance Program |
| EPI_ISL_450134 | MSHS Clinical Microbiology Laboratories | MSHS Pathogen Surveillance Program |
| EPI_ISL_450135 | MSHS Clinical Microbiology Laboratories | MSHS Pathogen Surveillance Program |
| EPI_ISL_450136 | MSHS Clinical Microbiology Laboratories | MSHS Pathogen Surveillance Program |
| EPI_ISL_450137 | MSHS Clinical Microbiology Laboratories | MSHS Pathogen Surveillance Program |
| EPI_ISL_450138 | MSHS Clinical Microbiology Laboratories | MSHS Pathogen Surveillance Program |
| EPI_ISL_450139 | MSHS Clinical Microbiology Laboratories | MSHS Pathogen Surveillance Program |
| EPI_ISL_450140 | MSHS Clinical Microbiology Laboratories | MSHS Pathogen Surveillance Program |
| EPI_ISL_450141 | MSHS Clinical Microbiology Laboratories | MSHS Pathogen Surveillance Program |
| EPI_ISL_450142 | MSHS Clinical Microbiology Laboratories | MSHS Pathogen Surveillance Program |
| EPI_ISL_450143 | MSHS Clinical Microbiology Laboratories | MSHS Pathogen Surveillance Program |
| EPI_ISL_450144 | MSHS Clinical Microbiology Laboratories | MSHS Pathogen Surveillance Program |
| EPI_ISL_450145 | MSHS Clinical Microbiology Laboratories | MSHS Pathogen Surveillance Program |
| EPI_ISL_450146 | MSHS Clinical Microbiology Laboratories | MSHS Pathogen Surveillance Program |
| EPI_ISL_450147 | MSHS Clinical Microbiology Laboratories | MSHS Pathogen Surveillance Program |
| EPI_ISL_450148 | MSHS Clinical Microbiology Laboratories | MSHS Pathogen Surveillance Program |
| EPI_ISL_450149 | MSHS Clinical Microbiology Laboratories | MSHS Pathogen Surveillance Program |
| EPI_ISL_450150 | MSHS Clinical Microbiology Laboratories | MSHS Pathogen Surveillance Program |
| EPI_ISL_450151 | MSHS Clinical Microbiology Laboratories | MSHS Pathogen Surveillance Program |

| EPI_Accession_ID | Originating Laboratory | Submitting Laboratory |
|---|---|---|
| EPI_ISL_450152 | MSHS Clinical Microbiology Laboratories | MSHS Pathogen Surveillance Program |
| EPI_ISL_450153 | MSHS Clinical Microbiology Laboratories | MSHS Pathogen Surveillance Program |
| EPI_ISL_450154 | MSHS Clinical Microbiology Laboratories | MSHS Pathogen Surveillance Program |
| EPI_ISL_450155 | MSHS Clinical Microbiology Laboratories | MSHS Pathogen Surveillance Program |
| EPI_ISL_450156 | MSHS Clinical Microbiology Laboratories | MSHS Pathogen Surveillance Program |
| EPI_ISL_450157 | MSHS Clinical Microbiology Laboratories | MSHS Pathogen Surveillance Program |
| EPI_ISL_450158 | MSHS Clinical Microbiology Laboratories | MSHS Pathogen Surveillance Program |
| EPI_ISL_450159 | MSHS Clinical Microbiology Laboratories | MSHS Pathogen Surveillance Program |
| EPI_ISL_450160 | MSHS Clinical Microbiology Laboratories | MSHS Pathogen Surveillance Program |
| EPI_ISL_450161 | MSHS Clinical Microbiology Laboratories | MSHS Pathogen Surveillance Program |
| EPI_ISL_450162 | MSHS Clinical Microbiology Laboratories | MSHS Pathogen Surveillance Program |
| EPI_ISL_450163 | MSHS Clinical Microbiology Laboratories | MSHS Pathogen Surveillance Program |
| EPI_ISL_450164 | MSHS Clinical Microbiology Laboratories | MSHS Pathogen Surveillance Program |
| EPI_ISL_450165 | MSHS Clinical Microbiology Laboratories | MSHS Pathogen Surveillance Program |
| EPI_ISL_450166 | MSHS Clinical Microbiology Laboratories | MSHS Pathogen Surveillance Program |
| EPI_ISL_450167 | MSHS Clinical Microbiology Laboratories | MSHS Pathogen Surveillance Program |
| EPI_ISL_450168 | MSHS Clinical Microbiology Laboratories | MSHS Pathogen Surveillance Program |
| EPI_ISL_450169 | MSHS Clinical Microbiology Laboratories | MSHS Pathogen Surveillance Program |
| EPI_ISL_450170 | MSHS Clinical Microbiology Laboratories | MSHS Pathogen Surveillance Program |
| EPI_ISL_450171 | National Influenza centre, Sri Lanka | The University of Hong Kong |
| EPI_ISL_450172 | National Influenza centre, Sri Lanka | The University of Hong Kong |
| EPI_ISL_450173 | Robert Garry lab | Andersen lab at Scripps Research |
| EPI_ISL_450174 | Robert Garry lab | Andersen lab at Scripps Research |
| EPI_ISL_450175 | Robert Garry lab | Andersen lab at Scripps Research |
| EPI_ISL_450176 | Robert Garry lab | Andersen lab at Scripps Research |
| EPI_ISL_450177 | Robert Garry lab | Andersen lab at Scripps Research |
| EPI_ISL_450178 | Robert Garry lab | Andersen lab at Scripps Research |
| EPI_ISL_450179 | Robert Garry lab | Andersen lab at Scripps Research |
| EPI_ISL_450180 | Robert Garry lab | Andersen lab at Scripps Research |
| EPI_ISL_450181 | Robert Garry lab | Andersen lab at Scripps Research |
| EPI_ISL_450182 | Robert Garry lab | Andersen lab at Scripps Research |
| EPI_ISL_450183 | Robert Garry lab | Andersen lab at Scripps Research |
| EPI_ISL_450184 | Robert Garry lab | Andersen lab at Scripps Research |

| EPI_Accession_ID | Originating Laboratory | Submitting Laboratory |
|---|---|---|
| EPI_ISL_450185 | Robert Garry lab | Andersen lab at Scripps Research |
| EPI_ISL_450186 | Biolab Diagnostic Laboratories | Andersen lab at Scripps Research |
| EPI_ISL_450187 | Biolab Diagnostic Laboratories | Andersen lab at Scripps Research |
| EPI_ISL_450188 | Biolab Diagnostic Laboratories | Andersen lab at Scripps Research |
| EPI_ISL_450189 | Biolab Diagnostic Laboratories | Andersen lab at Scripps Research |
| EPI_ISL_450190 | Rady's Childrens Hospital | Andersen lab at Scripps Research |
| EPI_ISL_450191 | Scripps Medical Laboratory | Andersen lab at Scripps Research |
| EPI_ISL_450192 | Scripps Medical Laboratory | Andersen lab at Scripps Research |
| EPI_ISL_450193 | SA Pathology | SA Pathology |
| EPI_ISL_450194 | SA Pathology | SA Pathology |
| EPI_ISL_450195 | SA Pathology | SA Pathology |
| EPI_ISL_450196 | bumrungrad international hospital | National Institute of Health. Department of medical Sciences, Ministry of Public Health, Thailand |
| EPI_ISL_450197 | - | National Institute of Health. Department of medical Sciences, Ministry of Public Health, Thailand |
| EPI_ISL_450198 | unknown | Department of Virology |
| EPI_ISL_450199 | unknown | Department of Virology |
| EPI_ISL_450200 | unknown | Department of Virology |
| EPI_ISL_450203 | unknown | Department of Virology |
| EPI_ISL_450204 | unknown | Department of Virology |
| EPI_ISL_450207 | unknown | Department of Virology |
| EPI_ISL_450208 | unknown | Department of Virology |
| EPI_ISL_450209 | unknown | Department of Virology |
| EPI_ISL_450212 | unknown | Microbiological Diagnostic Unit Public Health Laboratory (MDU-PHL) and Victorian Infectious Disease Reference Laboratory (VIDRL) |
| EPI_ISL_450213 | unknown | Microbiological Diagnostic Unit Public Health Laboratory (MDU-PHL) and Victorian Infectious Disease Reference Laboratory (VIDRL) |
| EPI_ISL_450214 | unknown | Microbiological Diagnostic Unit Public Health Laboratory (MDU-PHL) and Victorian Infectious Disease Reference Laboratory (VIDRL) |

| EPI_Accession_ID | Originating Laboratory | Submitting Laboratory |
|---|---|---|
| EPI_ISL_450215 | unknown | Microbiological Diagnostic Unit Public Health Laboratory (MDU-PHL) and Victorian Infectious Disease Reference Laboratory (VIDRL) |
| EPI_ISL_450216 | unknown | Microbiological Diagnostic Unit Public Health Laboratory (MDU-PHL) and Victorian Infectious Disease Reference Laboratory (VIDRL) |
| EPI_ISL_450231 | Robert Garry lab | Andersen lab at Scripps Research |
| EPI_ISL_450232 | UCSF Clinical Microbiology Laboratory | Chiu Laboratory, University of California, San Francisco |
| EPI_ISL_450233 | UCSF Clinical Microbiology Laboratory | Chiu Laboratory, University of California, San Francisco |
| EPI_ISL_450234 | UCSF Clinical Microbiology Laboratory | Chiu Laboratory, University of California, San Francisco |
| EPI_ISL_450235 | UCSF Clinical Microbiology Laboratory | Chiu Laboratory, University of California, San Francisco |
| EPI_ISL_450236 | UCSF Clinical Microbiology Laboratory | Chiu Laboratory, University of California, San Francisco |
| EPI_ISL_450237 | UCSF Clinical Microbiology Laboratory | Chiu Laboratory, University of California, San Francisco |
| EPI_ISL_450238 | UCSF Clinical Microbiology Laboratory | Chiu Laboratory, University of California, San Francisco |
| EPI_ISL_450239 | UCSF Clinical Microbiology Laboratory | Chiu Laboratory, University of California, San Francisco |
| EPI_ISL_450240 | UCSF Clinical Microbiology Laboratory | Chiu Laboratory, University of California, San Francisco |
| EPI_ISL_450241 | WHO National Influenza Centre Russian Federation | WHO National Influenza Centre Russian Federation |
| EPI_ISL_450242 | WHO National Influenza Centre Russian Federation | WHO National Influenza Centre Russian Federation |
| EPI_ISL_450243 | WHO National Influenza Centre Russian Federation | WHO National Influenza Centre Russian Federation |
| EPI_ISL_450244 | WHO National Influenza Centre Russian Federation | WHO National Influenza Centre Russian Federation |
| EPI_ISL_450245 | WHO National Influenza Centre Russian Federation | WHO National Influenza Centre Russian Federation |
| EPI_ISL_450246 | WHO National Influenza Centre Russian Federation | WHO National Influenza Centre Russian Federation |
| EPI_ISL_450247 | WHO National Influenza Centre Russian Federation | WHO National Influenza Centre Russian Federation |
| EPI_ISL_450248 | WHO National Influenza Centre Russian Federation | WHO National Influenza Centre Russian Federation |
| EPI_ISL_450249 | WHO National Influenza Centre Russian Federation | WHO National Influenza Centre Russian Federation |
| EPI_ISL_450250 | WHO National Influenza Centre Russian Federation | WHO National Influenza Centre Russian Federation |
| EPI_ISL_450251 | WHO National Influenza Centre Russian Federation | WHO National Influenza Centre Russian Federation |
| EPI_ISL_450252 | WHO National Influenza Centre Russian Federation | WHO National Influenza Centre Russian Federation |
| EPI_ISL_450253 | WHO National Influenza Centre Russian Federation | WHO National Influenza Centre Russian Federation |
| EPI_ISL_450254 | WHO National Influenza Centre Russian Federation | WHO National Influenza Centre Russian Federation |
| EPI_ISL_450255 | WHO National Influenza Centre Russian Federation | WHO National Influenza Centre Russian Federation |
| EPI_ISL_450256 | WHO National Influenza Centre Russian Federation | WHO National Influenza Centre Russian Federation |
| EPI_ISL_450257 | WHO National Influenza Centre Russian Federation | WHO National Influenza Centre Russian Federation |

| EPI_Accession_ID | Originating Laboratory | Submitting Laboratory |
|---|---|---|
| EPI_ISL_450258 | WHO National Influenza Centre Russian Federation | WHO National Influenza Centre Russian Federation |
| EPI_ISL_450259 | WHO National Influenza Centre Russian Federation | WHO National Influenza Centre Russian Federation |
| EPI_ISL_450260 | WHO National Influenza Centre Russian Federation | WHO National Influenza Centre Russian Federation |
| EPI_ISL_450261 | WHO National Influenza Centre Russian Federation | WHO National Influenza Centre Russian Federation |
| EPI_ISL_450262 | WHO National Influenza Centre Russian Federation | WHO National Influenza Centre Russian Federation |
| EPI_ISL_450263 | WHO National Influenza Centre Russian Federation | WHO National Influenza Centre Russian Federation |
| EPI_ISL_450264 | WHO National Influenza Centre Russian Federation | WHO National Influenza Centre Russian Federation |
| EPI_ISL_450265 | WHO National Influenza Centre Russian Federation | WHO National Influenza Centre Russian Federation |
| EPI_ISL_450266 | WHO National Influenza Centre Russian Federation | WHO National Influenza Centre Russian Federation |
| EPI_ISL_450267 | WHO National Influenza Centre Russian Federation | WHO National Influenza Centre Russian Federation |
| EPI_ISL_450268 | WHO National Influenza Centre Russian Federation | WHO National Influenza Centre Russian Federation |
| EPI_ISL_450269 | WHO National Influenza Centre Russian Federation | WHO National Influenza Centre Russian Federation |
| EPI_ISL_450270 | WHO National Influenza Centre Russian Federation | WHO National Influenza Centre Russian Federation |
| EPI_ISL_450271 | WHO National Influenza Centre Russian Federation | WHO National Influenza Centre Russian Federation |
| EPI_ISL_450272 | WHO National Influenza Centre Russian Federation | WHO National Influenza Centre Russian Federation |
| EPI_ISL_450273 | WHO National Influenza Centre Russian Federation | WHO National Influenza Centre Russian Federation |
| EPI_ISL_450274 | WHO National Influenza Centre Russian Federation | WHO National Influenza Centre Russian Federation |
| EPI_ISL_450275 | WHO National Influenza Centre Russian Federation | WHO National Influenza Centre Russian Federation |
| EPI_ISL_450276 | WHO National Influenza Centre Russian Federation | WHO National Influenza Centre Russian Federation |
| EPI_ISL_450277 | WHO National Influenza Centre Russian Federation | WHO National Influenza Centre Russian Federation |
| EPI_ISL_450278 | WHO National Influenza Centre Russian Federation | WHO National Influenza Centre Russian Federation |
| EPI_ISL_450279 | WHO National Influenza Centre Russian Federation | WHO National Influenza Centre Russian Federation |
| EPI_ISL_450280 | WHO National Influenza Centre Russian Federation | WHO National Influenza Centre Russian Federation |
| EPI_ISL_450281 | WHO National Influenza Centre Russian Federation | WHO National Influenza Centre Russian Federation |
| EPI_ISL_450282 | WHO National Influenza Centre Russian Federation | WHO National Influenza Centre Russian Federation |
| EPI_ISL_450283 | WHO National Influenza Centre Russian Federation | WHO National Influenza Centre Russian Federation |
| EPI_ISL_450284 | WHO National Influenza Centre Russian Federation | WHO National Influenza Centre Russian Federation |
| EPI_ISL_450285 | WHO National Influenza Centre Russian Federation | WHO National Influenza Centre Russian Federation |
| EPI_ISL_450286 | WHO National Influenza Centre Russian Federation | WHO National Influenza Centre Russian Federation |
| EPI_ISL_450287 | WHO National Influenza Centre Russian Federation | WHO National Influenza Centre Russian Federation |
| EPI_ISL_450288 | WHO National Influenza Centre Russian Federation | WHO National Influenza Centre Russian Federation |
| EPI_ISL_450289 | WHO National Influenza Centre Russian Federation | WHO National Influenza Centre Russian Federation |
| EPI_ISL_450290 | WHO National Influenza Centre Russian Federation | WHO National Influenza Centre Russian Federation |

| EPI_Accession_ID | Originating Laboratory | Submitting Laboratory |
|---|---|---|
| EPI_ISL_450291 | WHO National Influenza Centre Russian Federation | WHO National Influenza Centre Russian Federation |
| EPI_ISL_450292 | WHO National Influenza Centre Russian Federation | WHO National Influenza Centre Russian Federation |
| EPI_ISL_450293 | WHO National Influenza Centre Russian Federation | WHO National Influenza Centre Russian Federation |
| EPI_ISL_450294 | Institute of Human Genetics, Polish Academy of Sciences Sanitary and Epidemiological Station in PoznaÅ„ | Institute of Human Genetics, Polish Academy of Sciences |
| EPI_ISL_450295 | Institute of Human Genetics, Polish Academy of Sciences; Sanitary and Epidemiological Station in PoznaÅ„ | Institute of Human Genetics, Polish Academy of Sciences |
| EPI_ISL_450296 | National Institute for Communicable Diseases of the National Health Laboratory Service | National Institute for Communicable Diseases of the National Health Laboratory Service |
| EPI_ISL_450297 | National Institute for Communicable Diseases of the National Health Laboratory Service | National Institute for Communicable Diseases of the National Health Laboratory Service |
| EPI_ISL_450298 | National Institute for Communicable Diseases of the National Health Laboratory Service | National Institute for Communicable Diseases of the National Health Laboratory Service |
| EPI_ISL_450299 | National Institute for Communicable Diseases of the National Health Laboratory Service | National Institute for Communicable Diseases of the National Health Laboratory Service |
| EPI_ISL_450300 | National Institute for Communicable Diseases of the National Health Laboratory Service | National Institute for Communicable Diseases of the National Health Laboratory Service |
| EPI_ISL_450301 | National Institute for Communicable Diseases of the National Health Laboratory Service | National Institute for Communicable Diseases of the National Health Laboratory Service |
| EPI_ISL_450302 | Centre hospitalier Anna-Laberge | Laboratoire de santÃ© publique du QuÃ©bec |
| EPI_ISL_450303 | Centre hospitalier Anna-Laberge | Laboratoire de santÃ© publique du QuÃ©bec |
| EPI_ISL_450304 | HÃ´pital Charles-LeMoyne | Laboratoire de santÃ© publique du QuÃ©bec |
| EPI_ISL_450305 | HÃ´pital Charles-LeMoyne | Laboratoire de santÃ© publique du QuÃ©bec |
| EPI_ISL_450306 | CSSS Haut-Richelieu/Rouville (HÃ´pital) | Laboratoire de santÃ© publique du QuÃ©bec |
| EPI_ISL_450307 | HÃ´pital du SuroÃ®t | Laboratoire de santÃ© publique du QuÃ©bec |
| EPI_ISL_450308 | HÃ´pital Charles-LeMoyne | Laboratoire de santÃ© publique du QuÃ©bec |
| EPI_ISL_450309 | HÃ´pital Pierre-Boucher | Laboratoire de santÃ© publique du QuÃ©bec |
| EPI_ISL_450310 | HÃ´pital Charles-LeMoyne | Laboratoire de santÃ© publique du QuÃ©bec |
| EPI_ISL_450311 | HÃ´pital du SuroÃ®t | Laboratoire de santÃ© publique du QuÃ©bec |
| EPI_ISL_450312 | HÃ´pital du SuroÃ®t | Laboratoire de santÃ© publique du QuÃ©bec |
| EPI_ISL_450313 | HÃ´pital du SuroÃ®t | Laboratoire de santÃ© publique du QuÃ©bec |

| EPI_Accession_ID | Originating Laboratory | Submitting Laboratory |
|---|---|---|
| EPI_ISL_450314 | HÃ´pital Pierre-Boucher | Laboratoire de santÃ© publique du QuÃ©bec |
| EPI_ISL_450315 | HÃ´pital Pierre-Boucher | Laboratoire de santÃ© publique du QuÃ©bec |
| EPI_ISL_450316 | HÃ´pital Charles-LeMoyne | Laboratoire de santÃ© publique du QuÃ©bec |
| EPI_ISL_450317 | HÃ´pital Pierre-Boucher | Laboratoire de santÃ© publique du QuÃ©bec |
| EPI_ISL_450318 | HÃ´pital Pierre-Boucher | Laboratoire de santÃ© publique du QuÃ©bec |
| EPI_ISL_450321 | NIV Pune | CSIR-Centre for Cellular and Molecular Biology |
| EPI_ISL_450322 | NIV Pune | CSIR-Centre for Cellular and Molecular Biology |
| EPI_ISL_450323 | NIV Pune | CSIR-Centre for Cellular and Molecular Biology |
| EPI_ISL_450324 | NIV Pune | CSIR-Centre for Cellular and Molecular Biology |
| EPI_ISL_450325 | NIV Pune | CSIR-Centre for Cellular and Molecular Biology |
| EPI_ISL_450326 | CSIR-Centre for Cellular and Molecular Biology | CSIR-Centre for Cellular and Molecular Biology |
| EPI_ISL_450327 | CSIR-Centre for Cellular and Molecular Biology | CSIR-Centre for Cellular and Molecular Biology |
| EPI_ISL_450328 | CSIR-Centre for Cellular and Molecular Biology | CSIR-Centre for Cellular and Molecular Biology |
| EPI_ISL_450329 | CSIR-Centre for Cellular and Molecular Biology | CSIR-Centre for Cellular and Molecular Biology |
| EPI_ISL_450330 | CSIR-Centre for Cellular and Molecular Biology | CSIR-Centre for Cellular and Molecular Biology |
| EPI_ISL_450331 | CSIR-Centre for Cellular and Molecular Biology | CSIR-Centre for Cellular and Molecular Biology |
| EPI_ISL_450332 | CSIR-Centre for Cellular and Molecular Biology | CSIR-Centre for Cellular and Molecular Biology |
| EPI_ISL_450337 | Hospital Universitari Vall d'Hebron - Vall d'Hebron Institut de Recerca | Hospital Universitari Vall d'Hebron |
| EPI_ISL_450338 | Institute of Human Genetics, Polish Academy of Sciences; Sanitary and Epidemiological Station in PoznaÅ„ | Institute of Human Genetics, Polish Academy of Sciences |
| EPI_ISL_450339 | Corona (COVID-19) Testing Laboratory and Bangladesh Institute of Tropical & Infectious Diseases | Basic and Applied Research on Jute Project |
| EPI_ISL_450340 | Corona (COVID-19) Testing Laboratory and Bangladesh Institute of Tropical & Infectious Diseases | Basic and Applied Research on Jute Project |
| EPI_ISL_450341 | unknown | Basic and Applied Research on Jute Project |
| EPI_ISL_450342 | Corona (COVID-19) Testing Laboratory and Bangladesh Institute of Tropical & Infectious Diseases | Basic and Applied Research on Jute Project |
| EPI_ISL_450343 | Corona (COVID-19) Testing Laboratory and Bangladesh Institute of Tropical & Infectious Diseases | Basic and Applied Research on Jute Project |
| EPI_ISL_450344 | Corona (COVID-19) Testing Laboratory and Bangladesh Institute of Tropical & Infectious Diseases | Basic and Applied Research on Jute Project |

| EPI_Accession_ID | Originating Laboratory | Submitting Laboratory |
|---|---|---|
| EPI_ISL_450345 | Corona (COVID-19) Testing Laboratory and Bangladesh Institute of Tropical & Infectious Diseases | Basic and Applied Research on Jute Project |
| EPI_ISL_450346 | St.Olavs hospital/NTNU | Institute of Genomics Core Facility, University of Tartu |
| EPI_ISL_450347 | St.Olavs hospital/NTNU | Institute of Genomics Core Facility, University of Tartu |
| EPI_ISL_450348 | St.Olavs hospital/NTNU | Institute of Genomics Core Facility, University of Tartu |
| EPI_ISL_450349 | St.Olavs hospital/NTNU | Institute of Genomics Core Facility, University of Tartu |
| EPI_ISL_450350 | St.Olavs hospital/NTNU | Institute of Genomics Core Facility, University of Tartu |
| EPI_ISL_450351 | St.Olavs hospital/NTNU | Institute of Genomics Core Facility, University of Tartu |
| EPI_ISL_450352 | St.Olavs hospital/NTNU | Institute of Genomics Core Facility, University of Tartu |
| EPI_ISL_450393 | NYU Langone Health | Departments of Pathology and Medicine, New York University School of Medicine |
| EPI_ISL_450394 | NYU Langone Health | Departments of Pathology and Medicine, New York University School of Medicine |
| EPI_ISL_450395 | NYU Langone Health | Departments of Pathology and Medicine, New York University School of Medicine |
| EPI_ISL_450396 | NYU Langone Health | Departments of Pathology and Medicine, New York University School of Medicine |
| EPI_ISL_450397 | NYU Langone Health | Departments of Pathology and Medicine, New York University School of Medicine |
| EPI_ISL_450398 | NYU Langone Health | Departments of Pathology and Medicine, New York University School of Medicine |
| EPI_ISL_450399 | NYU Langone Health | Departments of Pathology and Medicine, New York University School of Medicine |
| EPI_ISL_450400 | NYU Langone Health | Departments of Pathology and Medicine, New York University School of Medicine |
| EPI_ISL_450401 | NYU Langone Health | Departments of Pathology and Medicine, New York University School of Medicine |
| EPI_ISL_450402 | NYU Langone Health | Departments of Pathology and Medicine, New York University School of Medicine |
| EPI_ISL_450403 | unknown | School of Public Health |
| EPI_ISL_450404 | unknown | School of Public Health |
| EPI_ISL_450405 | unknown | School of Public Health |
| EPI_ISL_450408 | unknown | Microbiology |

| EPI_Accession_ID | Originating Laboratory | Submitting Laboratory |
|---|---|---|
| EPI_ISL_450409 | unknown | Microbiology |
| EPI_ISL_450410 | unknown | Microbiology |
| EPI_ISL_450411 | unknown | Microbiology |
| EPI_ISL_450412 | unknown | Microbiology |
| EPI_ISL_450413 | unknown | Center for Diagnostics |
| EPI_ISL_450442 | unknown | The Department of Infectious Disease Prevention and Control |
| EPI_ISL_450444 | unknown | Dept. OPA |
| EPI_ISL_450445 | Stanford clinical virology lab | Chan-Zuckerberg Biohub |
| EPI_ISL_450446 | Stanford clinical virology lab | Chan-Zuckerberg Biohub |
| EPI_ISL_450447 | Stanford clinical virology lab | Chan-Zuckerberg Biohub |
| EPI_ISL_450448 | Stanford clinical virology lab | Chan-Zuckerberg Biohub |
| EPI_ISL_450449 | Stanford clinical virology lab | Chan-Zuckerberg Biohub |
| EPI_ISL_450450 | Stanford clinical virology lab | Chan-Zuckerberg Biohub |
| EPI_ISL_450451 | Stanford clinical virology lab | Chan-Zuckerberg Biohub |
| EPI_ISL_450452 | Stanford clinical virology lab | Chan-Zuckerberg Biohub |
| EPI_ISL_450453 | Stanford clinical virology lab | Chan-Zuckerberg Biohub |
| EPI_ISL_450454 | Stanford clinical virology lab | Chan-Zuckerberg Biohub |
| EPI_ISL_450455 | Stanford clinical virology lab | Chan-Zuckerberg Biohub |
| EPI_ISL_450456 | Stanford clinical virology lab | Chan-Zuckerberg Biohub |
| EPI_ISL_450457 | Stanford clinical virology lab | Chan-Zuckerberg Biohub |
| EPI_ISL_450458 | Stanford clinical virology lab | Chan-Zuckerberg Biohub |
| EPI_ISL_450459 | Stanford clinical virology lab | Chan-Zuckerberg Biohub |
| EPI_ISL_450460 | Stanford clinical virology lab | Chan-Zuckerberg Biohub |
| EPI_ISL_450461 | Stanford clinical virology lab | Chan-Zuckerberg Biohub |
| EPI_ISL_450462 | Stanford clinical virology lab | Chan-Zuckerberg Biohub |
| EPI_ISL_450463 | Stanford clinical virology lab | Chan-Zuckerberg Biohub |
| EPI_ISL_450464 | Stanford clinical virology lab | Chan-Zuckerberg Biohub |
| EPI_ISL_450465 | Stanford clinical virology lab | Chan-Zuckerberg Biohub |
| EPI_ISL_450466 | Stanford clinical virology lab | Chan-Zuckerberg Biohub |
| EPI_ISL_450467 | Stanford clinical virology lab | Chan-Zuckerberg Biohub |
| EPI_ISL_450468 | Stanford clinical virology lab | Chan-Zuckerberg Biohub |
| EPI_ISL_450469 | Stanford clinical virology lab | Chan-Zuckerberg Biohub |

| EPI_Accession_ID | Originating Laboratory | Submitting Laboratory |
|---|---|---|
| EPI_ISL_450470 | Stanford clinical virology lab | Chan-Zuckerberg Biohub |
| EPI_ISL_450471 | Stanford clinical virology lab | Chan-Zuckerberg Biohub |
| EPI_ISL_450472 | Stanford clinical virology lab | Chan-Zuckerberg Biohub |
| EPI_ISL_450473 | Stanford clinical virology lab | Chan-Zuckerberg Biohub |
| EPI_ISL_450474 | Stanford clinical virology lab | Chan-Zuckerberg Biohub |
| EPI_ISL_450475 | Stanford clinical virology lab | Chan-Zuckerberg Biohub |
| EPI_ISL_450476 | Stanford clinical virology lab | Chan-Zuckerberg Biohub |
| EPI_ISL_450477 | Stanford clinical virology lab | Chan-Zuckerberg Biohub |
| EPI_ISL_450478 | Stanford clinical virology lab | Chan-Zuckerberg Biohub |
| EPI_ISL_450479 | Stanford clinical virology lab | Chan-Zuckerberg Biohub |
| EPI_ISL_450480 | Stanford clinical virology lab | Chan-Zuckerberg Biohub |
| EPI_ISL_450481 | Stanford clinical virology lab | Chan-Zuckerberg Biohub |
| EPI_ISL_450484 | unknown | Data Science |
| EPI_ISL_450485 | unknown | Data Science |
| EPI_ISL_450486 | unknown | Data Science |
| EPI_ISL_450487 | unknown | Data Science |
| EPI_ISL_450489 | unknown | CAS Key Laboratory of Special Pathogens and Biosafety and Center for Emerging Infectious Diseases |
| EPI_ISL_450495 | National Institute for Communicable Diseases of the National Health Laboratory Service | National Institute for Communicable Diseases of the National Health Laboratory Service |
| EPI_ISL_450500 | unknown | CAS Key Laboratory of Special Pathogens and Biosafety and Center for Emerging Infectious Diseases |
| EPI_ISL_450501 | unknown | CAS Key Laboratory of Special Pathogens and Biosafety and Center for Emerging Infectious Diseases |
| EPI_ISL_450502 | unknown | CAS Key Laboratory of Special Pathogens and Biosafety and Center for Emerging Infectious Diseases |
| EPI_ISL_450503 | unknown | CAS Key Laboratory of Special Pathogens and Biosafety and Center for Emerging Infectious Diseases |
| EPI_ISL_450504 | unknown | CAS Key Laboratory of Special Pathogens and Biosafety and Center for Emerging Infectious Diseases |
| EPI_ISL_450506 | unknown | Clinical Laboratory |
| EPI_ISL_450508 | Rafik Hariri University Hospital | Rafik Hariri University Hospital |
| EPI_ISL_450509 | Rafik Hariri University Hospital | Rafik Hariri University Hospital |

| EPI_Accession_ID | Originating Laboratory | Submitting Laboratory |
|---|---|---|
| EPI_ISL_450511 | Rafik Hariri University Hospital | Rafik Hariri University Hospital |
| EPI_ISL_450512 | Rafik Hariri University Hospital | Rafik Hariri University Hospital |
| EPI_ISL_450514 | Rafik Hariri University Hospital | Rafik Hariri University Hospital |
| EPI_ISL_450515 | Rafik Hariri University Hospital | Rafik Hariri University Hospital |
| EPI_ISL_450516 | Rafik Hariri University Hospital | Rafik Hariri University Hospital |
| EPI_ISL_450518 | E. Gulbja Laboratorija | Latvian Biomedical Research and Study Centre |
| EPI_ISL_450519 | E. Gulbja Laboratorija | Latvian Biomedical Research and Study Centre |
| EPI_ISL_450520 | CentrÄ�Ä�Laboratorija | Latvian Biomedical Research and Study Centre |
| EPI_ISL_450521 | CentrÄ�Ä�Laboratorija | Latvian Biomedical Research and Study Centre |
| EPI_ISL_450522 | CentrÄ�Ä�Laboratorija | Latvian Biomedical Research and Study Centre |
| EPI_ISL_450523 | CentrÄ�Ä�Laboratorija | Latvian Biomedical Research and Study Centre |
| EPI_ISL_450524 | CentrÄ�Ä�Laboratorija | Latvian Biomedical Research and Study Centre |
| EPI_ISL_450525 | Hematology Laboratory, Section of Molecular Diagnostics, University Clinical Centre, Medical University of Gdansk | Department of Virology, Faculty of Medicine, University of Helsinki, Helsinki, Finland |
| EPI_ISL_450526 | Hematology Laboratory, Section of Molecular Diagnostics, University Clinical Centre, Medical University of Gdansk | Department of Virology, Faculty of Medicine, University of Helsinki, Helsinki, Finland |
| EPI_ISL_450527 | Hematology Laboratory, Section of Molecular Diagnostics, University Clinical Centre, Medical University of Gdansk | Department of Virology, Faculty of Medicine, University of Helsinki, Helsinki, Finland |
| EPI_ISL_450528 | Hematology Laboratory, Section of Molecular Diagnostics, University Clinical Centre, Medical University of Gdansk | Department of Virology, Faculty of Medicine, University of Helsinki, Helsinki, Finland |
| EPI_ISL_450529 | Hematology Laboratory, Section of Molecular Diagnostics, University Clinical Centre, Medical University of Gdansk | Department of Virology, Faculty of Medicine, University of Helsinki, Helsinki, Finland |
| EPI_ISL_450530 | Hematology Laboratory, Section of Molecular Diagnostics, University Clinical Centre, Medical University of Gdansk | Department of Virology, Faculty of Medicine, University of Helsinki, Helsinki, Finland |
| EPI_ISL_450538 | Utah Public Health Laboratory | Utah Public Health Laboratory |
| EPI_ISL_450539 | Utah Public Health Laboratory | Utah Public Health Laboratory |
| EPI_ISL_450540 | Utah Public Health Laboratory | Utah Public Health Laboratory |

| EPI_Accession_ID | Originating Laboratory | Submitting Laboratory |
|---|---|---|
| EPI_ISL_450541 | Utah Public Health Laboratory | Utah Public Health Laboratory |
| EPI_ISL_450542 | Utah Public Health Laboratory | Utah Public Health Laboratory |
| EPI_ISL_450543 | Utah Public Health Laboratory | Utah Public Health Laboratory |
| EPI_ISL_450544 | Utah Public Health Laboratory | Utah Public Health Laboratory |
| EPI_ISL_450545 | Utah Public Health Laboratory | Utah Public Health Laboratory |
| EPI_ISL_450546 | Utah Public Health Laboratory | Utah Public Health Laboratory |
| EPI_ISL_450547 | Utah Public Health Laboratory | Utah Public Health Laboratory |
| EPI_ISL_450548 | Utah Public Health Laboratory | Utah Public Health Laboratory |
| EPI_ISL_450549 | Utah Public Health Laboratory | Utah Public Health Laboratory |
| EPI_ISL_450550 | Utah Public Health Laboratory | Utah Public Health Laboratory |
| EPI_ISL_450551 | Utah Public Health Laboratory | Utah Public Health Laboratory |
| EPI_ISL_450552 | Utah Public Health Laboratory | Utah Public Health Laboratory |
| EPI_ISL_450553 | Utah Public Health Laboratory | Utah Public Health Laboratory |
| EPI_ISL_450554 | Utah Public Health Laboratory | Utah Public Health Laboratory |
| EPI_ISL_450555 | Utah Public Health Laboratory | Utah Public Health Laboratory |
| EPI_ISL_450556 | Utah Public Health Laboratory | Utah Public Health Laboratory |
| EPI_ISL_450557 | Utah Public Health Laboratory | Utah Public Health Laboratory |
| EPI_ISL_450559 | Utah Public Health Laboratory | Utah Public Health Laboratory |
| EPI_ISL_450560 | Utah Public Health Laboratory | Utah Public Health Laboratory |
| EPI_ISL_450561 | Utah Public Health Laboratory | Utah Public Health Laboratory |
| EPI_ISL_450562 | Utah Public Health Laboratory | Utah Public Health Laboratory |
| EPI_ISL_450563 | Utah Public Health Laboratory | Utah Public Health Laboratory |
| EPI_ISL_450564 | Utah Public Health Laboratory | Utah Public Health Laboratory |
| EPI_ISL_450565 | Utah Public Health Laboratory | Utah Public Health Laboratory |
| EPI_ISL_450566 | Utah Public Health Laboratory | Utah Public Health Laboratory |
| EPI_ISL_450567 | Utah Public Health Laboratory | Utah Public Health Laboratory |
| EPI_ISL_450568 | Utah Public Health Laboratory | Utah Public Health Laboratory |
| EPI_ISL_450570 | Michigan Department of Health and Human Services, Bureau of Laboratories | Michigan Department of Health and Human Services, Bureau of Laboratories |
| EPI_ISL_450571 | Michigan Department of Health and Human Services, Bureau of Laboratories | Michigan Department of Health and Human Services, Bureau of Laboratories |
| EPI_ISL_450572 | Michigan Department of Health and Human Services, Bureau of Laboratories | Michigan Department of Health and Human Services, Bureau of Laboratories |

| EPI_Accession_ID | Originating Laboratory | Submitting Laboratory |
|---|---|---|
| EPI_ISL_450573 | Michigan Department of Health and Human Services, Bureau of Laboratories | Michigan Department of Health and Human Services, Bureau of Laboratories |
| EPI_ISL_450574 | Michigan Department of Health and Human Services, Bureau of Laboratories | Michigan Department of Health and Human Services, Bureau of Laboratories |
| EPI_ISL_450575 | Michigan Department of Health and Human Services, Bureau of Laboratories | Michigan Department of Health and Human Services, Bureau of Laboratories |
| EPI_ISL_450576 | Michigan Department of Health and Human Services, Bureau of Laboratories | Michigan Department of Health and Human Services, Bureau of Laboratories |
| EPI_ISL_450577 | Michigan Department of Health and Human Services, Bureau of Laboratories | Michigan Department of Health and Human Services, Bureau of Laboratories |
| EPI_ISL_450578 | Michigan Department of Health and Human Services, Bureau of Laboratories | Michigan Department of Health and Human Services, Bureau of Laboratories |
| EPI_ISL_450579 | Michigan Department of Health and Human Services, Bureau of Laboratories | Michigan Department of Health and Human Services, Bureau of Laboratories |
| EPI_ISL_450580 | Michigan Department of Health and Human Services, Bureau of Laboratories | Michigan Department of Health and Human Services, Bureau of Laboratories |
| EPI_ISL_450581 | Michigan Department of Health and Human Services, Bureau of Laboratories | Michigan Department of Health and Human Services, Bureau of Laboratories |
| EPI_ISL_450582 | Michigan Department of Health and Human Services, Bureau of Laboratories | Michigan Department of Health and Human Services, Bureau of Laboratories |
| EPI_ISL_450583 | Michigan Department of Health and Human Services, Bureau of Laboratories | Michigan Department of Health and Human Services, Bureau of Laboratories |
| EPI_ISL_450584 | Michigan Department of Health and Human Services, Bureau of Laboratories | Michigan Department of Health and Human Services, Bureau of Laboratories |
| EPI_ISL_450585 | Michigan Department of Health and Human Services, Bureau of Laboratories | Michigan Department of Health and Human Services, Bureau of Laboratories |
| EPI_ISL_450586 | Michigan Department of Health and Human Services, Bureau of Laboratories | Michigan Department of Health and Human Services, Bureau of Laboratories |
| EPI_ISL_450587 | Michigan Department of Health and Human Services, Bureau of Laboratories | Michigan Department of Health and Human Services, Bureau of Laboratories |
| EPI_ISL_450588 | Michigan Department of Health and Human Services, Bureau of Laboratories | Michigan Department of Health and Human Services, Bureau of Laboratories |

| EPI_Accession_ID | Originating Laboratory | Submitting Laboratory |
|---|---|---|
| EPI_ISL_450590 | Michigan Department of Health and Human Services, Bureau of Laboratories | Michigan Department of Health and Human Services, Bureau of Laboratories |
| EPI_ISL_450591 | Michigan Department of Health and Human Services, Bureau of Laboratories | Michigan Department of Health and Human Services, Bureau of Laboratories |
| EPI_ISL_450592 | Michigan Department of Health and Human Services, Bureau of Laboratories | Michigan Department of Health and Human Services, Bureau of Laboratories |
| EPI_ISL_450593 | Michigan Department of Health and Human Services, Bureau of Laboratories | Michigan Department of Health and Human Services, Bureau of Laboratories |
| EPI_ISL_450594 | Michigan Department of Health and Human Services, Bureau of Laboratories | Michigan Department of Health and Human Services, Bureau of Laboratories |
| EPI_ISL_450595 | Michigan Department of Health and Human Services, Bureau of Laboratories | Michigan Department of Health and Human Services, Bureau of Laboratories |
| EPI_ISL_450596 | Michigan Department of Health and Human Services, Bureau of Laboratories | Michigan Department of Health and Human Services, Bureau of Laboratories |
| EPI_ISL_450597 | Michigan Department of Health and Human Services, Bureau of Laboratories | Michigan Department of Health and Human Services, Bureau of Laboratories |
| EPI_ISL_450598 | Michigan Department of Health and Human Services, Bureau of Laboratories | Michigan Department of Health and Human Services, Bureau of Laboratories |
| EPI_ISL_450599 | Michigan Department of Health and Human Services, Bureau of Laboratories | Michigan Department of Health and Human Services, Bureau of Laboratories |
| EPI_ISL_450600 | Michigan Department of Health and Human Services, Bureau of Laboratories | Michigan Department of Health and Human Services, Bureau of Laboratories |
| EPI_ISL_450601 | Michigan Department of Health and Human Services, Bureau of Laboratories | Michigan Department of Health and Human Services, Bureau of Laboratories |
| EPI_ISL_450602 | Michigan Department of Health and Human Services, Bureau of Laboratories | Michigan Department of Health and Human Services, Bureau of Laboratories |
| EPI_ISL_450603 | Michigan Department of Health and Human Services, Bureau of Laboratories | Michigan Department of Health and Human Services, Bureau of Laboratories |
| EPI_ISL_450604 | Michigan Department of Health and Human Services, Bureau of Laboratories | Michigan Department of Health and Human Services, Bureau of Laboratories |
| EPI_ISL_450605 | Michigan Department of Health and Human Services, Bureau of Laboratories | Michigan Department of Health and Human Services, Bureau of Laboratories |

| EPI_Accession_ID | Originating Laboratory | Submitting Laboratory |
|---|---|---|
| EPI_ISL_450606 | Michigan Department of Health and Human Services, Bureau of Laboratories | Michigan Department of Health and Human Services, Bureau of Laboratories |
| EPI_ISL_450607 | Michigan Department of Health and Human Services, Bureau of Laboratories | Michigan Department of Health and Human Services, Bureau of Laboratories |
| EPI_ISL_450608 | Michigan Department of Health and Human Services, Bureau of Laboratories | Michigan Department of Health and Human Services, Bureau of Laboratories |
| EPI_ISL_450609 | Michigan Department of Health and Human Services, Bureau of Laboratories | Michigan Department of Health and Human Services, Bureau of Laboratories |
| EPI_ISL_450610 | Michigan Department of Health and Human Services, Bureau of Laboratories | Michigan Department of Health and Human Services, Bureau of Laboratories |
| EPI_ISL_450611 | Michigan Department of Health and Human Services, Bureau of Laboratories | Michigan Department of Health and Human Services, Bureau of Laboratories |
| EPI_ISL_450612 | Michigan Department of Health and Human Services, Bureau of Laboratories | Michigan Department of Health and Human Services, Bureau of Laboratories |
| EPI_ISL_450613 | Michigan Department of Health and Human Services, Bureau of Laboratories | Michigan Department of Health and Human Services, Bureau of Laboratories |
| EPI_ISL_450614 | Michigan Department of Health and Human Services, Bureau of Laboratories | Michigan Department of Health and Human Services, Bureau of Laboratories |
| EPI_ISL_450615 | Michigan Department of Health and Human Services, Bureau of Laboratories | Michigan Department of Health and Human Services, Bureau of Laboratories |
| EPI_ISL_450616 | Michigan Department of Health and Human Services, Bureau of Laboratories | Michigan Department of Health and Human Services, Bureau of Laboratories |
| EPI_ISL_450617 | Michigan Department of Health and Human Services, Bureau of Laboratories | Michigan Department of Health and Human Services, Bureau of Laboratories |
| EPI_ISL_450618 | Michigan Department of Health and Human Services, Bureau of Laboratories | Michigan Department of Health and Human Services, Bureau of Laboratories |
| EPI_ISL_450619 | Michigan Department of Health and Human Services, Bureau of Laboratories | Michigan Department of Health and Human Services, Bureau of Laboratories |
| EPI_ISL_450620 | Michigan Department of Health and Human Services, Bureau of Laboratories | Michigan Department of Health and Human Services, Bureau of Laboratories |
| EPI_ISL_450621 | Michigan Department of Health and Human Services, Bureau of Laboratories | Michigan Department of Health and Human Services, Bureau of Laboratories |

| EPI_Accession_ID | Originating Laboratory | Submitting Laboratory |
|---|---|---|
| EPI_ISL_450622 | Michigan Department of Health and Human Services, Bureau of Laboratories | Michigan Department of Health and Human Services, Bureau of Laboratories |
| EPI_ISL_450623 | Michigan Department of Health and Human Services, Bureau of Laboratories | Michigan Department of Health and Human Services, Bureau of Laboratories |
| EPI_ISL_450624 | Michigan Department of Health and Human Services, Bureau of Laboratories | Michigan Department of Health and Human Services, Bureau of Laboratories |
| EPI_ISL_450625 | Michigan Department of Health and Human Services, Bureau of Laboratories | Michigan Department of Health and Human Services, Bureau of Laboratories |
| EPI_ISL_450626 | Michigan Department of Health and Human Services, Bureau of Laboratories | Michigan Department of Health and Human Services, Bureau of Laboratories |
| EPI_ISL_450627 | Michigan Department of Health and Human Services, Bureau of Laboratories | Michigan Department of Health and Human Services, Bureau of Laboratories |
| EPI_ISL_450628 | Michigan Department of Health and Human Services, Bureau of Laboratories | Michigan Department of Health and Human Services, Bureau of Laboratories |
| EPI_ISL_450629 | Michigan Department of Health and Human Services, Bureau of Laboratories | Michigan Department of Health and Human Services, Bureau of Laboratories |
| EPI_ISL_450630 | Michigan Department of Health and Human Services, Bureau of Laboratories | Michigan Department of Health and Human Services, Bureau of Laboratories |
| EPI_ISL_450631 | Michigan Department of Health and Human Services, Bureau of Laboratories | Michigan Department of Health and Human Services, Bureau of Laboratories |
| EPI_ISL_450632 | Michigan Department of Health and Human Services, Bureau of Laboratories | Michigan Department of Health and Human Services, Bureau of Laboratories |
| EPI_ISL_450633 | Michigan Department of Health and Human Services, Bureau of Laboratories | Michigan Department of Health and Human Services, Bureau of Laboratories |
| EPI_ISL_450634 | Michigan Department of Health and Human Services, Bureau of Laboratories | Michigan Department of Health and Human Services, Bureau of Laboratories |
| EPI_ISL_450635 | Michigan Department of Health and Human Services, Bureau of Laboratories | Michigan Department of Health and Human Services, Bureau of Laboratories |
| EPI_ISL_450636 | Michigan Department of Health and Human Services, Bureau of Laboratories | Michigan Department of Health and Human Services, Bureau of Laboratories |
| EPI_ISL_450637 | Michigan Department of Health and Human Services, Bureau of Laboratories | Michigan Department of Health and Human Services, Bureau of Laboratories |

| EPI_Accession_ID | Originating Laboratory | Submitting Laboratory |
|---|---|---|
| EPI_ISL_450638 | Laboratoire de microbiologie, Hopital de Verdun | Smith Laboratory, Centre de Recherche CHU Sainte-Justine |
| EPI_ISL_450639 | Laboratoire de microbiologie, Hopital de Verdun | Smith Laboratory, Centre de Recherche CHU Sainte-Justine |
| EPI_ISL_450640 | Laboratoire de microbiologie, Hopital de Verdun | Smith Laboratory, Centre de Recherche CHU Sainte-Justine |
| EPI_ISL_450641 | Laboratoire de microbiologie, Hopital de Verdun | Smith Laboratory, Centre de Recherche CHU Sainte-Justine |
| EPI_ISL_450642 | Laboratoire de microbiologie, Hopital de Verdun | Smith Laboratory, Centre de Recherche CHU Sainte-Justine |
| EPI_ISL_450643 | Laboratoire de microbiologie, Hopital de Verdun | Smith Laboratory, Centre de Recherche CHU Sainte-Justine |
| EPI_ISL_450644 | Laboratoire de microbiologie, Hopital de Verdun | Smith Laboratory, Centre de Recherche CHU Sainte-Justine |
| EPI_ISL_450645 | Laboratoire de microbiologie, Hopital de Verdun | Smith Laboratory, Centre de Recherche CHU Sainte-Justine |
| EPI_ISL_450646 | Laboratoire de microbiologie, Hopital de Verdun | Smith Laboratory, Centre de Recherche CHU Sainte-Justine |
| EPI_ISL_450647 | Laboratoire de microbiologie, Hopital de Verdun | Smith Laboratory, Centre de Recherche CHU Sainte-Justine |
| EPI_ISL_450648 | Laboratoire de microbiologie, Hopital de Verdun | Smith Laboratory, Centre de Recherche CHU Sainte-Justine |
| EPI_ISL_450649 | Laboratoire de microbiologie, Hopital de Verdun | Smith Laboratory, Centre de Recherche CHU Sainte-Justine |
| EPI_ISL_450650 | Laboratoire de microbiologie, Hopital de Verdun | Smith Laboratory, Centre de Recherche CHU Sainte-Justine |
| EPI_ISL_450651 | Laboratoire de microbiologie, Hopital de Verdun | Smith Laboratory, Centre de Recherche CHU Sainte-Justine |
| EPI_ISL_450652 | Laboratoire de microbiologie, Hopital de Verdun | Smith Laboratory, Centre de Recherche CHU Sainte-Justine |
| EPI_ISL_450653 | Laboratoire de microbiologie, Hopital de Verdun | Smith Laboratory, Centre de Recherche CHU Sainte-Justine |

| EPI_Accession_ID | Originating Laboratory | Submitting Laboratory |
|---|---|---|
| EPI_ISL_450700 | University of Wisconsin-Madison AIDS Vaccine Research Laboratories | University of Wisconsin-Madison AIDS Vaccine Research Laboratories |
| EPI_ISL_450701 | University of Wisconsin-Madison AIDS Vaccine Research Laboratories | University of Wisconsin-Madison AIDS Vaccine Research Laboratories |
| EPI_ISL_450702 | University of Wisconsin-Madison AIDS Vaccine Research Laboratories | University of Wisconsin-Madison AIDS Vaccine Research Laboratories |
| EPI_ISL_450703 | University of Wisconsin-Madison AIDS Vaccine Research Laboratories | University of Wisconsin-Madison AIDS Vaccine Research Laboratories |
| EPI_ISL_450704 | University of Wisconsin-Madison AIDS Vaccine Research Laboratories | University of Wisconsin-Madison AIDS Vaccine Research Laboratories |
| EPI_ISL_450705 | University of Wisconsin-Madison AIDS Vaccine Research Laboratories | University of Wisconsin-Madison AIDS Vaccine Research Laboratories |
| EPI_ISL_450706 | University of Wisconsin-Madison AIDS Vaccine Research Laboratories | University of Wisconsin-Madison AIDS Vaccine Research Laboratories |
| EPI_ISL_450707 | University of Wisconsin-Madison AIDS Vaccine Research Laboratories | University of Wisconsin-Madison AIDS Vaccine Research Laboratories |
| EPI_ISL_450708 | University of Wisconsin-Madison AIDS Vaccine Research Laboratories | University of Wisconsin-Madison AIDS Vaccine Research Laboratories |
| EPI_ISL_450709 | University of Wisconsin-Madison AIDS Vaccine Research Laboratories | University of Wisconsin-Madison AIDS Vaccine Research Laboratories |
| EPI_ISL_450710 | University of Wisconsin-Madison AIDS Vaccine Research Laboratories | University of Wisconsin-Madison AIDS Vaccine Research Laboratories |
| EPI_ISL_450711 | University of Wisconsin-Madison AIDS Vaccine Research Laboratories | University of Wisconsin-Madison AIDS Vaccine Research Laboratories |
| EPI_ISL_450712 | University of Wisconsin-Madison AIDS Vaccine Research Laboratories | University of Wisconsin-Madison AIDS Vaccine Research Laboratories |
| EPI_ISL_450713 | University of Wisconsin-Madison AIDS Vaccine Research Laboratories | University of Wisconsin-Madison AIDS Vaccine Research Laboratories |
| EPI_ISL_450714 | University of Wisconsin-Madison AIDS Vaccine Research Laboratories | University of Wisconsin-Madison AIDS Vaccine Research Laboratories |
| EPI_ISL_450715 | University of Wisconsin-Madison AIDS Vaccine Research Laboratories | University of Wisconsin-Madison AIDS Vaccine Research Laboratories |

| EPI_Accession_ID | Originating Laboratory | Submitting Laboratory |
|---|---|---|
| EPI_ISL_450716 | University of Wisconsin-Madison AIDS Vaccine Research Laboratories | University of Wisconsin-Madison AIDS Vaccine Research Laboratories |
| EPI_ISL_450717 | University of Wisconsin-Madison AIDS Vaccine Research Laboratories | University of Wisconsin-Madison AIDS Vaccine Research Laboratories |
| EPI_ISL_450718 | University of Wisconsin-Madison AIDS Vaccine Research Laboratories | University of Wisconsin-Madison AIDS Vaccine Research Laboratories |
| EPI_ISL_450719 | University of Wisconsin-Madison AIDS Vaccine Research Laboratories | University of Wisconsin-Madison AIDS Vaccine Research Laboratories |
| EPI_ISL_450720 | University of Wisconsin-Madison AIDS Vaccine Research Laboratories | University of Wisconsin-Madison AIDS Vaccine Research Laboratories |
| EPI_ISL_450721 | University of Wisconsin-Madison AIDS Vaccine Research Laboratories | University of Wisconsin-Madison AIDS Vaccine Research Laboratories |
| EPI_ISL_450722 | University of Wisconsin-Madison AIDS Vaccine Research Laboratories | University of Wisconsin-Madison AIDS Vaccine Research Laboratories |
| EPI_ISL_450723 | Ramathibodi Hospital | COVID-19 Network Investigations (CONI) Alliance |
| EPI_ISL_450724 | Hospital AZ Rivierenland | Institute of Tropical Medicine |
| EPI_ISL_450725 | Hospital AZ Rivierenland | Institute of Tropical Medicine |
| EPI_ISL_450726 | Hospital AZ Rivierenland | Institute of Tropical Medicine |
| EPI_ISL_450727 | Hospital AZ Rivierenland | Institute of Tropical Medicine |
| EPI_ISL_450728 | Hospital AZ Rivierenland | Institute of Tropical Medicine |
| EPI_ISL_450729 | Hospital AZ Rivierenland | Institute of Tropical Medicine |
| EPI_ISL_450730 | Hospital AZ Rivierenland | Institute of Tropical Medicine |
| EPI_ISL_450731 | Hospital AZ Rivierenland | Institute of Tropical Medicine |
| EPI_ISL_450732 | Hospital AZ Rivierenland | Institute of Tropical Medicine |
| EPI_ISL_450734 | Hospital AZ Rivierenland | Institute of Tropical Medicine |
| EPI_ISL_450735 | Hospital AZ Rivierenland | Institute of Tropical Medicine |
| EPI_ISL_450736 | Hospital AZ Rivierenland | Institute of Tropical Medicine |
| EPI_ISL_450737 | Hospital AZ Rivierenland | Institute of Tropical Medicine |
| EPI_ISL_450738 | OUCRU/HTD | OUCRU/HTD |
| EPI_ISL_450739 | OUCRU/HTD | OUCRU/HTD |
| EPI_ISL_450740 | OUCRU/HTD | OUCRU/HTD |
| EPI_ISL_450741 | OUCRU/HTD | OUCRU/HTD |
| EPI_ISL_450742 | OUCRU/HTD | OUCRU/HTD |

| EPI_Accession_ID | Originating Laboratory | Submitting Laboratory |
|---|---|---|
| EPI_ISL_450743 | OUCRU/HTD | OUCRU/HTD |
| EPI_ISL_450744 | OUCRU/HTD | OUCRU/HTD |
| EPI_ISL_450745 | OUCRU/HTD | OUCRU/HTD |
| EPI_ISL_450746 | Laboratory of Molecular Biology, Diagnostyka sp. z o.o. | Laboratory of Recombinant Vaccines |
| EPI_ISL_450747 | Sunnybrook Health Sciences Centre | Department of Laboratory Medicine and Molecular Diagnostics, Sunnybrook Health Sciences Centre |
| EPI_ISL_450748 | Minnesota Department of Health, Public Health Laboratory | Minnesota Department of Health, Public Health Laboratory |
| EPI_ISL_450749 | Minnesota Department of Health, Public Health Laboratory | Minnesota Department of Health, Public Health Laboratory |
| EPI_ISL_450750 | Minnesota Department of Health, Public Health Laboratory | Minnesota Department of Health, Public Health Laboratory |
| EPI_ISL_450751 | Minnesota Department of Health, Public Health Laboratory | Minnesota Department of Health, Public Health Laboratory |
| EPI_ISL_450752 | Minnesota Department of Health, Public Health Laboratory | Minnesota Department of Health, Public Health Laboratory |
| EPI_ISL_450753 | Minnesota Department of Health, Public Health Laboratory | Minnesota Department of Health, Public Health Laboratory |
| EPI_ISL_450754 | Minnesota Department of Health, Public Health Laboratory | Minnesota Department of Health, Public Health Laboratory |
| EPI_ISL_450755 | Minnesota Department of Health, Public Health Laboratory | Minnesota Department of Health, Public Health Laboratory |
| EPI_ISL_450756 | Minnesota Department of Health, Public Health Laboratory | Minnesota Department of Health, Public Health Laboratory |
| EPI_ISL_450757 | Minnesota Department of Health, Public Health Laboratory | Minnesota Department of Health, Public Health Laboratory |
| EPI_ISL_450758 | Minnesota Department of Health, Public Health Laboratory | Minnesota Department of Health, Public Health Laboratory |
| EPI_ISL_450759 | Minnesota Department of Health, Public Health Laboratory | Minnesota Department of Health, Public Health Laboratory |
| EPI_ISL_450760 | Minnesota Department of Health, Public Health Laboratory | Minnesota Department of Health, Public Health Laboratory |

| EPI_Accession_ID | Originating Laboratory | Submitting Laboratory |
|---|---|---|
| EPI_ISL_450761 | Minnesota Department of Health, Public Health Laboratory | Minnesota Department of Health, Public Health Laboratory |
| EPI_ISL_450762 | Minnesota Department of Health, Public Health Laboratory | Minnesota Department of Health, Public Health Laboratory |
| EPI_ISL_450763 | Minnesota Department of Health, Public Health Laboratory | Minnesota Department of Health, Public Health Laboratory |
| EPI_ISL_450764 | Minnesota Department of Health, Public Health Laboratory | Minnesota Department of Health, Public Health Laboratory |
| EPI_ISL_450765 | Minnesota Department of Health, Public Health Laboratory | Minnesota Department of Health, Public Health Laboratory |
| EPI_ISL_450766 | Minnesota Department of Health, Public Health Laboratory | Minnesota Department of Health, Public Health Laboratory |
| EPI_ISL_450767 | Minnesota Department of Health, Public Health Laboratory | Minnesota Department of Health, Public Health Laboratory |
| EPI_ISL_450768 | Minnesota Department of Health, Public Health Laboratory | Minnesota Department of Health, Public Health Laboratory |
| EPI_ISL_450769 | Minnesota Department of Health, Public Health Laboratory | Minnesota Department of Health, Public Health Laboratory |
| EPI_ISL_450770 | Minnesota Department of Health, Public Health Laboratory | Minnesota Department of Health, Public Health Laboratory |
| EPI_ISL_450771 | Minnesota Department of Health, Public Health Laboratory | Minnesota Department of Health, Public Health Laboratory |
| EPI_ISL_450772 | Minnesota Department of Health, Public Health Laboratory | Minnesota Department of Health, Public Health Laboratory |
| EPI_ISL_450773 | Minnesota Department of Health, Public Health Laboratory | Minnesota Department of Health, Public Health Laboratory |
| EPI_ISL_450774 | Minnesota Department of Health, Public Health Laboratory | Minnesota Department of Health, Public Health Laboratory |
| EPI_ISL_450775 | Minnesota Department of Health, Public Health Laboratory | Minnesota Department of Health, Public Health Laboratory |
| EPI_ISL_450776 | Minnesota Department of Health, Public Health Laboratory | Minnesota Department of Health, Public Health Laboratory |

| EPI_Accession_ID | Originating Laboratory | Submitting Laboratory |
| --- | --- | --- |
| EPI_ISL_450777 | Minnesota Department of Health, Public Health Laboratory | Minnesota Department of Health, Public Health Laboratory |
| EPI_ISL_450778 | Minnesota Department of Health, Public Health Laboratory | Minnesota Department of Health, Public Health Laboratory |
| EPI_ISL_450779 | Minnesota Department of Health, Public Health Laboratory | Minnesota Department of Health, Public Health Laboratory |
| EPI_ISL_450780 | Minnesota Department of Health, Public Health Laboratory | Minnesota Department of Health, Public Health Laboratory |
| EPI_ISL_450781 | Government Medical College-Bhavnagar | Gujarat Biotechnology Research Centre |
| EPI_ISL_450782 | Government Medical College-Bhavnagar | Gujarat Biotechnology Research Centre |
| EPI_ISL_450783 | Government Medical College-Bhavnagar | Gujarat Biotechnology Research Centre |
| EPI_ISL_450784 | Government Medical College-Bhavnagar | Gujarat Biotechnology Research Centre |
| EPI_ISL_450785 | Pandit Deendayal Upadhyay Government Medical College, Rajkot | Gujarat Biotechnology Research Centre |
| EPI_ISL_450786 | Pandit Deendayal Upadhyay Government Medical College, Rajkot | Gujarat Biotechnology Research Centre |
| EPI_ISL_450787 | Pandit Deendayal Upadhyay Government Medical College, Rajkot | Gujarat Biotechnology Research Centre |
| EPI_ISL_450788 | Pandit Deendayal Upadhyay Government Medical College, Rajkot | Gujarat Biotechnology Research Centre |
| EPI_ISL_450789 | Pandit Deendayal Upadhyay Government Medical College, Rajkot | Gujarat Biotechnology Research Centre |
| EPI_ISL_450790 | Pandit Deendayal Upadhyay Government Medical College, Rajkot | Gujarat Biotechnology Research Centre |
| EPI_ISL_450791 | Pandit Deendayal Upadhyay Government Medical College, Rajkot | Gujarat Biotechnology Research Centre |
| EPI_ISL_450792 | Jamaica Ministry of Health and Wellness | Pathogen Discovery, Respiratory Viruses Branch, Division of Viral Diseases, Centers for Disease Control and Prevention |
| EPI_ISL_450793 | Jamaica Ministry of Health and Wellness | Pathogen Discovery, Respiratory Viruses Branch, Division of Viral Diseases, Centers for Disease Control and Prevention |

| EPI_Accession_ID | Originating Laboratory | Submitting Laboratory |
| --- | --- | --- |
| EPI_ISL_450794 | Jamaica Ministry of Health and Wellness | Pathogen Discovery, Respiratory Viruses Branch, Division of Viral Diseases, Centers for Disease Control and Prevention |
| EPI_ISL_450795 | Jamaica Ministry of Health and Wellness | Pathogen Discovery, Respiratory Viruses Branch, Division of Viral Diseases, Centers for Disease Control and Prevention |
| EPI_ISL_450796 | Jamaica Ministry of Health and Wellness | Pathogen Discovery, Respiratory Viruses Branch, Division of Viral Diseases, Centers for Disease Control and Prevention |
| EPI_ISL_450797 | Jamaica Ministry of Health and Wellness | Pathogen Discovery, Respiratory Viruses Branch, Division of Viral Diseases, Centers for Disease Control and Prevention |
| EPI_ISL_450798 | Jamaica Ministry of Health and Wellness | Pathogen Discovery, Respiratory Viruses Branch, Division of Viral Diseases, Centers for Disease Control and Prevention |
| EPI_ISL_450799 | Jamaica Ministry of Health and Wellness | Pathogen Discovery, Respiratory Viruses Branch, Division of Viral Diseases, Centers for Disease Control and Prevention |
| EPI_ISL_450800 | AR Dept. of Health-Public Health Lab | Pathogen Discovery, Respiratory Viruses Branch, Division of Viral Diseases, Centers for Disease Control and Prevention |
| EPI_ISL_450801 | Georgia Department of Health | Pathogen Discovery, Respiratory Viruses Branch, Division of Viral Diseases, Centers for Disease Control and Prevention |
| EPI_ISL_450802 | PA Department of Health, Bureau of Laboratories | Pathogen Discovery, Respiratory Viruses Branch, Division of Viral Diseases, Centers for Disease Control and Prevention |
| EPI_ISL_450803 | PA Department of Health, Bureau of Laboratories | Pathogen Discovery, Respiratory Viruses Branch, Division of Viral Diseases, Centers for Disease Control and Prevention |
| EPI_ISL_450804 | VI-US Virgin Islands Department of Health | Pathogen Discovery, Respiratory Viruses Branch, Division of Viral Diseases, Centers for Disease Control and Prevention |

| EPI_Accession_ID | Originating Laboratory | Submitting Laboratory |
| --- | --- | --- |
| EPI_ISL_450805 | VI-US Virgin Islands Department of Health | Pathogen Discovery, Respiratory Viruses Branch, Division of Viral Diseases, Centers for Disease Control and Prevention |
| EPI_ISL_450806 | VI-US Virgin Islands Department of Health | Pathogen Discovery, Respiratory Viruses Branch, Division of Viral Diseases, Centers for Disease Control and Prevention |
| EPI_ISL_450807 | Victoria Vard och Halsa | The Public Health Agency of Sweden |
| EPI_ISL_450808 | Saroledens Familjelakare | The Public Health Agency of Sweden |
| EPI_ISL_450809 | Kungsholmsdoktorn | The Public Health Agency of Sweden |
| EPI_ISL_450810 | Saroledens Familjelakare | The Public Health Agency of Sweden |
| EPI_ISL_450811 | Knivsta VC | The Public Health Agency of Sweden |
| EPI_ISL_450812 | Uppsala Narakut Aleris | The Public Health Agency of Sweden |
| EPI_ISL_450813 | Bla Kustens halsocentral | The Public Health Agency of Sweden |
| EPI_ISL_450814 | Huslakarna Varmbadhuset Varberg | The Public Health Agency of Sweden |
| EPI_ISL_450815 | Narhalsan Molnlycke, Barn och ungdomsmedicin | The Public Health Agency of Sweden |
| EPI_ISL_450816 | orestadsklinikens VC | The Public Health Agency of Sweden |
| EPI_ISL_450817 | Narhalsan Backa vardcentral | The Public Health Agency of Sweden |
| EPI_ISL_450818 | Aneby VC | The Public Health Agency of Sweden |
| EPI_ISL_450819 | Hovas Askim Familjelakare och BVC | The Public Health Agency of Sweden |
| EPI_ISL_450820 | Narhalsan Molnlycke, Barn och ungdomsmedicin | The Public Health Agency of Sweden |
| EPI_ISL_450821 | Jarpens HC | The Public Health Agency of Sweden |
| EPI_ISL_450822 | Jarpens HC | The Public Health Agency of Sweden |
| EPI_ISL_450823 | Narhalsan Backa vardcentral | The Public Health Agency of Sweden |
| EPI_ISL_450824 | Ulltuna Vardcentral | The Public Health Agency of Sweden |
| EPI_ISL_450825 | Narhalsan Backa vardcentral | The Public Health Agency of Sweden |
| EPI_ISL_450826 | Uppsala Narakut Aleris | The Public Health Agency of Sweden |
| EPI_ISL_450827 | Uppsala Narakut Aleris | The Public Health Agency of Sweden |
| EPI_ISL_450828 | Uppsala Narakut Aleris | The Public Health Agency of Sweden |
| EPI_ISL_450829 | Narhalsan Sjobo vardcentral | The Public Health Agency of Sweden |
| EPI_ISL_450830 | Narhalsan Olskroken VC | The Public Health Agency of Sweden |
| EPI_ISL_450831 | Wetterhalsan | The Public Health Agency of Sweden |
| EPI_ISL_450832 | Byjorden vardcentral | The Public Health Agency of Sweden |
| EPI_ISL_450833 | Wetterhalsan | The Public Health Agency of Sweden |

| EPI_Accession_ID | Originating Laboratory | Submitting Laboratory |
|---|---|---|
| EPI_ISL_450834 | Klinisk mikrobiologi och vardhygien Halmstad | The Public Health Agency of Sweden |
| EPI_ISL_450835 | Unilabs Skovde | The Public Health Agency of Sweden |
| EPI_ISL_450836 | Laboratoriemedicin | The Public Health Agency of Sweden |
| EPI_ISL_450837 | Laboratoriemedicin | The Public Health Agency of Sweden |
| EPI_ISL_450838 | Utah Public Health Laboratory | Utah Public Health Laboratory |
| EPI_ISL_450839 | COVID-19 Laboratory Centre for Advanced Research in Sciences (CARS), University of Dhaka, Dhaka-1000, Bangladesh | DNA Solution Ltd |
| EPI_ISL_450840 | COVID-19 Laboratory | DNA Solution Ltd. L-5 |
| EPI_ISL_450841 | COVID-19 Laboratory | DNA Solution Ltd |
| EPI_ISL_450843 | COVID-19 Laboratory | DNA Solution Ltd. |
| EPI_ISL_450844 | Florida Bureau of Public Health Laboratories | Florida Bureau of Public Health Laboratories |
| EPI_ISL_450845 | Florida Bureau of Public Health Laboratories | Florida Bureau of Public Health Laboratories |
| EPI_ISL_450846 | Florida Bureau of Public Health Laboratories | Florida Bureau of Public Health Laboratories |
| EPI_ISL_450847 | Florida Bureau of Public Health Laboratories | Florida Bureau of Public Health Laboratories |
| EPI_ISL_450848 | Florida Bureau of Public Health Laboratories | Florida Bureau of Public Health Laboratories |
| EPI_ISL_450849 | Florida Bureau of Public Health Laboratories | Florida Bureau of Public Health Laboratories |
| EPI_ISL_450850 | Florida Bureau of Public Health Laboratories | Florida Bureau of Public Health Laboratories |
| EPI_ISL_450851 | Florida Bureau of Public Health Laboratories | Florida Bureau of Public Health Laboratories |
| EPI_ISL_450852 | Florida Bureau of Public Health Laboratories | Florida Bureau of Public Health Laboratories |
| EPI_ISL_450854 | Florida Bureau of Public Health Laboratories | Florida Bureau of Public Health Laboratories |
| EPI_ISL_450855 | Florida Bureau of Public Health Laboratories | Florida Bureau of Public Health Laboratories |
| EPI_ISL_450856 | Florida Bureau of Public Health Laboratories | Florida Bureau of Public Health Laboratories |
| EPI_ISL_450857 | Florida Bureau of Public Health Laboratories | Florida Bureau of Public Health Laboratories |
| EPI_ISL_450858 | Florida Bureau of Public Health Laboratories | Florida Bureau of Public Health Laboratories |
| EPI_ISL_450859 | Florida Bureau of Public Health Laboratories | Florida Bureau of Public Health Laboratories |
| EPI_ISL_450861 | Florida Bureau of Public Health Laboratories | Florida Bureau of Public Health Laboratories |
| EPI_ISL_450862 | Florida Bureau of Public Health Laboratories | Florida Bureau of Public Health Laboratories |
| EPI_ISL_450863 | Florida Bureau of Public Health Laboratories | Florida Bureau of Public Health Laboratories |
| EPI_ISL_450864 | Florida Bureau of Public Health Laboratories | Florida Bureau of Public Health Laboratories |
| EPI_ISL_450865 | Florida Bureau of Public Health Laboratories | Florida Bureau of Public Health Laboratories |
| EPI_ISL_450866 | Florida Bureau of Public Health Laboratories | Florida Bureau of Public Health Laboratories |
| EPI_ISL_450867 | Florida Bureau of Public Health Laboratories | Florida Bureau of Public Health Laboratories |

| EPI_Accession_ID | Originating Laboratory | Submitting Laboratory |
|---|---|---|
| EPI_ISL_450868 | Florida Bureau of Public Health Laboratories | Florida Bureau of Public Health Laboratories |
| EPI_ISL_450869 | Florida Bureau of Public Health Laboratories | Florida Bureau of Public Health Laboratories |
| EPI_ISL_450870 | Florida Bureau of Public Health Laboratories | Florida Bureau of Public Health Laboratories |
| EPI_ISL_450871 | Florida Bureau of Public Health Laboratories | Florida Bureau of Public Health Laboratories |
| EPI_ISL_450872 | Florida Bureau of Public Health Laboratories | Florida Bureau of Public Health Laboratories |
| EPI_ISL_450873 | Evandro Chagas Institute | Evandro Chagas Institute |
| EPI_ISL_450874 | Evandro Chagas Institute | Evandro Chagas Institute |
| EPI_ISL_451076 | West China Hospital of Sichuan University | State Key Laboratory of Biotherapy of Sichuan University |
| EPI_ISL_451077 | SA Pathology | SA Pathology |
| EPI_ISL_451078 | SA Pathology | SA Pathology |
| EPI_ISL_451079 | SA Pathology | SA Pathology |
| EPI_ISL_451080 | SA Pathology | SA Pathology |
| EPI_ISL_451081 | SA Pathology | SA Pathology |
| EPI_ISL_451082 | SA Pathology | SA Pathology |
| EPI_ISL_451083 | SA Pathology | SA Pathology |
| EPI_ISL_451084 | SA Pathology | SA Pathology |
| EPI_ISL_451085 | SA Pathology | SA Pathology |
| EPI_ISL_451086 | SA Pathology | SA Pathology |
| EPI_ISL_451087 | SA Pathology | SA Pathology |
| EPI_ISL_451088 | SA Pathology | SA Pathology |
| EPI_ISL_451089 | SA Pathology | SA Pathology |
| EPI_ISL_451090 | SA Pathology | SA Pathology |
| EPI_ISL_451091 | SA Pathology | SA Pathology |
| EPI_ISL_451092 | SA Pathology | SA Pathology |
| EPI_ISL_451093 | SA Pathology | SA Pathology |
| EPI_ISL_451094 | SA Pathology | SA Pathology |
| EPI_ISL_451095 | SA Pathology | SA Pathology |
| EPI_ISL_451096 | SA Pathology | SA Pathology |
| EPI_ISL_451097 | SA Pathology | SA Pathology |
| EPI_ISL_451098 | SA Pathology | SA Pathology |
| EPI_ISL_451099 | SA Pathology | SA Pathology |
| EPI_ISL_451100 | SA Pathology | SA Pathology |

| EPI_Accession_ID | Originating Laboratory | Submitting Laboratory |
|---|---|---|
| EPI_ISL_451101 | SA Pathology | SA Pathology |
| EPI_ISL_451102 | SA Pathology | SA Pathology |
| EPI_ISL_451103 | SA Pathology | SA Pathology |
| EPI_ISL_451104 | SA Pathology | SA Pathology |
| EPI_ISL_451105 | SA Pathology | SA Pathology |
| EPI_ISL_451106 | SA Pathology | SA Pathology |
| EPI_ISL_451107 | SA Pathology | SA Pathology |
| EPI_ISL_451108 | SA Pathology | SA Pathology |
| EPI_ISL_451109 | SA Pathology | SA Pathology |
| EPI_ISL_451110 | SA Pathology | SA Pathology |
| EPI_ISL_451111 | SA Pathology | SA Pathology |
| EPI_ISL_451112 | SA Pathology | SA Pathology |
| EPI_ISL_451113 | SA Pathology | SA Pathology |
| EPI_ISL_451114 | SA Pathology | SA Pathology |
| EPI_ISL_451115 | SA Pathology | SA Pathology |
| EPI_ISL_451116 | SA Pathology | SA Pathology |
| EPI_ISL_451117 | SA Pathology | SA Pathology |
| EPI_ISL_451118 | SA Pathology | SA Pathology |
| EPI_ISL_451119 | SA Pathology | SA Pathology |
| EPI_ISL_451120 | SA Pathology | SA Pathology |
| EPI_ISL_451121 | SA Pathology | SA Pathology |
| EPI_ISL_451122 | SA Pathology | SA Pathology |
| EPI_ISL_451123 | SA Pathology | SA Pathology |
| EPI_ISL_451124 | SA Pathology | SA Pathology |
| EPI_ISL_451125 | SA Pathology | SA Pathology |
| EPI_ISL_451126 | SA Pathology | SA Pathology |
| EPI_ISL_451127 | SA Pathology | SA Pathology |
| EPI_ISL_451128 | SA Pathology | SA Pathology |
| EPI_ISL_451129 | SA Pathology | SA Pathology |
| EPI_ISL_451130 | SA Pathology | SA Pathology |
| EPI_ISL_451131 | SA Pathology | SA Pathology |
| EPI_ISL_451132 | SA Pathology | SA Pathology |
| EPI_ISL_451133 | SA Pathology | SA Pathology |

| EPI_Accession_ID | Originating Laboratory | Submitting Laboratory |
|---|---|---|
| EPI_ISL_451134 | SA Pathology | SA Pathology |
| EPI_ISL_451135 | SA Pathology | SA Pathology |
| EPI_ISL_451136 | SA Pathology | SA Pathology |
| EPI_ISL_451137 | SA Pathology | SA Pathology |
| EPI_ISL_451138 | SA Pathology | SA Pathology |
| EPI_ISL_451139 | SA Pathology | SA Pathology |
| EPI_ISL_451140 | SA Pathology | SA Pathology |
| EPI_ISL_451141 | SA Pathology | SA Pathology |
| EPI_ISL_451142 | SA Pathology | SA Pathology |
| EPI_ISL_451143 | SA Pathology | SA Pathology |
| EPI_ISL_451144 | SA Pathology | SA Pathology |
| EPI_ISL_451145 | SA Pathology | SA Pathology |
| EPI_ISL_451146 | SA Pathology | SA Pathology |
| EPI_ISL_451147 | SA Pathology | SA Pathology |
| EPI_ISL_451148 | SA Pathology | SA Pathology |
| EPI_ISL_451149 | M.P Shah Government Medocal college Jamnagar | Gujarat Biotechnology Research Centre |
| EPI_ISL_451150 | M.P Shah Government Medocal college Jamnagar | Gujarat Biotechnology Research Centre |
| EPI_ISL_451151 | M.P Shah Government Medocal college Jamnagar | Gujarat Biotechnology Research Centre |
| EPI_ISL_451152 | M.P Shah Government Medocal college Jamnagar | Gujarat Biotechnology Research Centre |
| EPI_ISL_451153 | M.P Shah Government Medocal college Jamnagar | Gujarat Biotechnology Research Centre |
| EPI_ISL_451154 | Government Medical College, Vadodara | Gujarat Biotechnology Research Centre |
| EPI_ISL_451155 | Government Medical College, Vadodara | Gujarat Biotechnology Research Centre |
| EPI_ISL_451156 | Government Medical College, Vadodara | Gujarat Biotechnology Research Centre |
| EPI_ISL_451157 | Government Medical College, Vadodara | Gujarat Biotechnology Research Centre |
| EPI_ISL_451158 | Government Medical College, Vadodara | Gujarat Biotechnology Research Centre |
| EPI_ISL_451159 | Government Medical College, Vadodara | Gujarat Biotechnology Research Centre |
| EPI_ISL_451160 | Government Medical College, Vadodara | Gujarat Biotechnology Research Centre |
| EPI_ISL_451161 | Government Medical College, Vadodara | Gujarat Biotechnology Research Centre |
| EPI_ISL_451162 | Government Medical College, Vadodara | Gujarat Biotechnology Research Centre |
| EPI_ISL_451163 | Government Medical College, Vadodara | Gujarat Biotechnology Research Centre |
| EPI_ISL_451164 | Lab voor klinische biologie | Onderzoeksgroep Virologie |
| EPI_ISL_451165 | Lab voor klinische biologie | Onderzoeksgroep Virologie |
| EPI_ISL_451166 | Lab voor klinische biologie | Onderzoeksgroep Virologie |

| EPI_Accession_ID | Originating Laboratory | Submitting Laboratory |
|---|---|---|
| EPI_ISL_451167 | Lab voor klinische biologie | Onderzoeksgroep Virologie |
| EPI_ISL_451168 | Lab voor klinische biologie | Onderzoeksgroep Virologie |
| EPI_ISL_451169 | Lab voor klinische biologie | Onderzoeksgroep Virologie |
| EPI_ISL_451170 | Lab voor klinische biologie | Onderzoeksgroep Virologie |
| EPI_ISL_451171 | Lab voor klinische biologie | Onderzoeksgroep Virologie |
| EPI_ISL_451172 | Lab voor klinische biologie | Onderzoeksgroep Virologie |
| EPI_ISL_451173 | Lab voor klinische biologie | Onderzoeksgroep Virologie |
| EPI_ISL_451174 | Lab voor klinische biologie | Onderzoeksgroep Virologie |
| EPI_ISL_451175 | Lab voor klinische biologie | Onderzoeksgroep Virologie |
| EPI_ISL_451176 | Lab voor klinische biologie | Onderzoeksgroep Virologie |
| EPI_ISL_451177 | Lab voor klinische biologie | Onderzoeksgroep Virologie |
| EPI_ISL_451178 | Lab voor klinische biologie | Onderzoeksgroep Virologie |
| EPI_ISL_451179 | Lab voor klinische biologie | Onderzoeksgroep Virologie |
| EPI_ISL_451180 | Lab voor klinische biologie | Onderzoeksgroep Virologie |
| EPI_ISL_451181 | Lab voor klinische biologie | Onderzoeksgroep Virologie |
| EPI_ISL_451182 | Lab voor klinische biologie | Onderzoeksgroep Virologie |
| EPI_ISL_451183 | Uganda Virus Research Institute | MRC/UVRI & LSHTM Uganda Research Unit |
| EPI_ISL_451184 | Uganda Virus Research Institute | MRC/UVRI & LSHTM Uganda Research Unit |
| EPI_ISL_451185 | Uganda Virus Research Institute | MRC/UVRI & LSHTM Uganda Research Unit |
| EPI_ISL_451186 | Uganda Virus Research Institute | MRC/UVRI & LSHTM Uganda Research Unit |
| EPI_ISL_451187 | Uganda Virus Research Institute | MRC/UVRI & LSHTM Uganda Research Unit |
| EPI_ISL_451188 | Uganda Virus Research Institute | MRC/UVRI & LSHTM Uganda Research Unit |
| EPI_ISL_451189 | Uganda Virus Research Institute | MRC/UVRI & LSHTM Uganda Research Unit |
| EPI_ISL_451190 | Uganda Virus Research Institute | MRC/UVRI & LSHTM Uganda Research Unit |
| EPI_ISL_451191 | Uganda Virus Research Institute | MRC/UVRI & LSHTM Uganda Research Unit |
| EPI_ISL_451192 | Uganda Virus Research Institute | MRC/UVRI & LSHTM Uganda Research Unit |
| EPI_ISL_451193 | Uganda Virus Research Institute | MRC/UVRI & LSHTM Uganda Research Unit |
| EPI_ISL_451194 | Uganda Virus Research Institute | MRC/UVRI & LSHTM Uganda Research Unit |
| EPI_ISL_451195 | Uganda Virus Research Institute | MRC/UVRI & LSHTM Uganda Research Unit |
| EPI_ISL_451196 | Uganda Virus Research Institute | MRC/UVRI & LSHTM Uganda Research Unit |
| EPI_ISL_451197 | Uganda Virus Research Institute | MRC/UVRI & LSHTM Uganda Research Unit |
| EPI_ISL_451198 | Uganda Virus Research Institute | MRC/UVRI & LSHTM Uganda Research Unit |
| EPI_ISL_451199 | Uganda Virus Research Institute | MRC/UVRI & LSHTM Uganda Research Unit |

| EPI_Accession_ID | Originating Laboratory | Submitting Laboratory |
|---|---|---|
| EPI_ISL_451200 | Uganda Virus Research Institute | MRC/UVRI & LSHTM Uganda Research Unit |
| EPI_ISL_451201 | Uganda Virus Research Institute | MRC/UVRI & LSHTM Uganda Research Unit |
| EPI_ISL_451202 | Uganda Virus Research Institute | MRC/UVRI & LSHTM Uganda Research Unit |
| EPI_ISL_451203 | LSUHS Emerging Viral Threat Laboratory | Microbial Genome Sequencing Center |
| EPI_ISL_451204 | LSUHS Emerging Viral Threat Laboratory | Microbial Genome Sequencing Center |
| EPI_ISL_451205 | LSUHS Emerging Viral Threat Laboratory | Microbial Genome Sequencing Center |
| EPI_ISL_451206 | LSUHS Emerging Viral Threat Laboratory | Microbial Genome Sequencing Center |
| EPI_ISL_451207 | LSUHS Emerging Viral Threat Laboratory | Microbial Genome Sequencing Center |
| EPI_ISL_451208 | LSUHS Emerging Viral Threat Laboratory | Microbial Genome Sequencing Center |
| EPI_ISL_451209 | LSUHS Emerging Viral Threat Laboratory | Microbial Genome Sequencing Center |
| EPI_ISL_451210 | LSUHS Emerging Viral Threat Laboratory | Microbial Genome Sequencing Center |
| EPI_ISL_451211 | LSUHS Emerging Viral Threat Laboratory | Microbial Genome Sequencing Center |
| EPI_ISL_451212 | LSUHS Emerging Viral Threat Laboratory | Microbial Genome Sequencing Center |
| EPI_ISL_451213 | LSUHS Emerging Viral Threat Laboratory | Microbial Genome Sequencing Center |
| EPI_ISL_451214 | LSUHS Emerging Viral Threat Laboratory | Microbial Genome Sequencing Center |
| EPI_ISL_451215 | LSUHS Emerging Viral Threat Laboratory | Microbial Genome Sequencing Center |
| EPI_ISL_451216 | LSUHS Emerging Viral Threat Laboratory | Microbial Genome Sequencing Center |
| EPI_ISL_451217 | LSUHS Emerging Viral Threat Laboratory | Microbial Genome Sequencing Center |
| EPI_ISL_451218 | LSUHS Emerging Viral Threat Laboratory | Microbial Genome Sequencing Center |
| EPI_ISL_451219 | LSUHS Emerging Viral Threat Laboratory | Microbial Genome Sequencing Center |
| EPI_ISL_451220 | LSUHS Emerging Viral Threat Laboratory | Microbial Genome Sequencing Center |
| EPI_ISL_451221 | LSUHS Emerging Viral Threat Laboratory | Microbial Genome Sequencing Center |
| EPI_ISL_451222 | LSUHS Emerging Viral Threat Laboratory | Microbial Genome Sequencing Center |
| EPI_ISL_451223 | LSUHS Emerging Viral Threat Laboratory | Microbial Genome Sequencing Center |
| EPI_ISL_451224 | LSUHS Emerging Viral Threat Laboratory | Microbial Genome Sequencing Center |
| EPI_ISL_451225 | LSUHS Emerging Viral Threat Laboratory | Microbial Genome Sequencing Center |
| EPI_ISL_451226 | LSUHS Emerging Viral Threat Laboratory | Microbial Genome Sequencing Center |
| EPI_ISL_451227 | LSUHS Emerging Viral Threat Laboratory | Microbial Genome Sequencing Center |
| EPI_ISL_451228 | LSUHS Emerging Viral Threat Laboratory | Microbial Genome Sequencing Center |
| EPI_ISL_451229 | LSUHS Emerging Viral Threat Laboratory | Microbial Genome Sequencing Center |
| EPI_ISL_451230 | LSUHS Emerging Viral Threat Laboratory | Microbial Genome Sequencing Center |
| EPI_ISL_451231 | LSUHS Emerging Viral Threat Laboratory | Microbial Genome Sequencing Center |
| EPI_ISL_451232 | LSUHS Emerging Viral Threat Laboratory | Microbial Genome Sequencing Center |

| EPI_Accession_ID | Originating Laboratory | Submitting Laboratory |
|---|---|---|
| EPI_ISL_451233 | LSUHS Emerging Viral Threat Laboratory | Microbial Genome Sequencing Center |
| EPI_ISL_451234 | LSUHS Emerging Viral Threat Laboratory | Microbial Genome Sequencing Center |
| EPI_ISL_451235 | LSUHS Emerging Viral Threat Laboratory | Microbial Genome Sequencing Center |
| EPI_ISL_451236 | LSUHS Emerging Viral Threat Laboratory | Microbial Genome Sequencing Center |
| EPI_ISL_451237 | LSUHS Emerging Viral Threat Laboratory | Microbial Genome Sequencing Center |
| EPI_ISL_451238 | LSUHS Emerging Viral Threat Laboratory | Microbial Genome Sequencing Center |
| EPI_ISL_451239 | LSUHS Emerging Viral Threat Laboratory | Microbial Genome Sequencing Center |
| EPI_ISL_451240 | LSUHS Emerging Viral Threat Laboratory | Microbial Genome Sequencing Center |
| EPI_ISL_451241 | LSUHS Emerging Viral Threat Laboratory | Microbial Genome Sequencing Center |
| EPI_ISL_451242 | LSUHS Emerging Viral Threat Laboratory | Microbial Genome Sequencing Center |
| EPI_ISL_451243 | LSUHS Emerging Viral Threat Laboratory | Microbial Genome Sequencing Center |
| EPI_ISL_451244 | LSUHS Emerging Viral Threat Laboratory | Microbial Genome Sequencing Center |
| EPI_ISL_451245 | LSUHS Emerging Viral Threat Laboratory | Microbial Genome Sequencing Center |
| EPI_ISL_451246 | LSUHS Emerging Viral Threat Laboratory | Microbial Genome Sequencing Center |
| EPI_ISL_451247 | LSUHS Emerging Viral Threat Laboratory | Microbial Genome Sequencing Center |
| EPI_ISL_451248 | LSUHS Emerging Viral Threat Laboratory | Microbial Genome Sequencing Center |
| EPI_ISL_451249 | LSUHS Emerging Viral Threat Laboratory | Microbial Genome Sequencing Center |
| EPI_ISL_451250 | LSUHS Emerging Viral Threat Laboratory | Microbial Genome Sequencing Center |
| EPI_ISL_451251 | LSUHS Emerging Viral Threat Laboratory | Microbial Genome Sequencing Center |
| EPI_ISL_451252 | LSUHS Emerging Viral Threat Laboratory | Microbial Genome Sequencing Center |
| EPI_ISL_451253 | LSUHS Emerging Viral Threat Laboratory | Microbial Genome Sequencing Center |
| EPI_ISL_451254 | LSUHS Emerging Viral Threat Laboratory | Microbial Genome Sequencing Center |
| EPI_ISL_451255 | LSUHS Emerging Viral Threat Laboratory | Microbial Genome Sequencing Center |
| EPI_ISL_451256 | LSUHS Emerging Viral Threat Laboratory | Microbial Genome Sequencing Center |
| EPI_ISL_451257 | LSUHS Emerging Viral Threat Laboratory | Microbial Genome Sequencing Center |
| EPI_ISL_451258 | LSUHS Emerging Viral Threat Laboratory | Microbial Genome Sequencing Center |
| EPI_ISL_451259 | LSUHS Emerging Viral Threat Laboratory | Microbial Genome Sequencing Center |
| EPI_ISL_451260 | LSUHS Emerging Viral Threat Laboratory | Microbial Genome Sequencing Center |
| EPI_ISL_451261 | LSUHS Emerging Viral Threat Laboratory | Microbial Genome Sequencing Center |
| EPI_ISL_451262 | LSUHS Emerging Viral Threat Laboratory | Microbial Genome Sequencing Center |
| EPI_ISL_451263 | LSUHS Emerging Viral Threat Laboratory | Microbial Genome Sequencing Center |
| EPI_ISL_451264 | LSUHS Emerging Viral Threat Laboratory | Microbial Genome Sequencing Center |
| EPI_ISL_451265 | LSUHS Emerging Viral Threat Laboratory | Microbial Genome Sequencing Center |

| EPI_Accession_ID | Originating Laboratory | Submitting Laboratory |
|---|---|---|
| EPI_ISL_451266 | LSUHS Emerging Viral Threat Laboratory | Microbial Genome Sequencing Center |
| EPI_ISL_451267 | LSUHS Emerging Viral Threat Laboratory | Microbial Genome Sequencing Center |
| EPI_ISL_451268 | LSUHS Emerging Viral Threat Laboratory | Microbial Genome Sequencing Center |
| EPI_ISL_451269 | LSUHS Emerging Viral Threat Laboratory | Microbial Genome Sequencing Center |
| EPI_ISL_451270 | LSUHS Emerging Viral Threat Laboratory | Microbial Genome Sequencing Center |
| EPI_ISL_451271 | LSUHS Emerging Viral Threat Laboratory | Microbial Genome Sequencing Center |
| EPI_ISL_451272 | LSUHS Emerging Viral Threat Laboratory | Microbial Genome Sequencing Center |
| EPI_ISL_451273 | LSUHS Emerging Viral Threat Laboratory | Microbial Genome Sequencing Center |
| EPI_ISL_451274 | LSUHS Emerging Viral Threat Laboratory | Microbial Genome Sequencing Center |
| EPI_ISL_451275 | LSUHS Emerging Viral Threat Laboratory | Microbial Genome Sequencing Center |
| EPI_ISL_451276 | LSUHS Emerging Viral Threat Laboratory | Microbial Genome Sequencing Center |
| EPI_ISL_451277 | LSUHS Emerging Viral Threat Laboratory | Microbial Genome Sequencing Center |
| EPI_ISL_451278 | LSUHS Emerging Viral Threat Laboratory | Microbial Genome Sequencing Center |
| EPI_ISL_451279 | LSUHS Emerging Viral Threat Laboratory | Microbial Genome Sequencing Center |
| EPI_ISL_451280 | LSUHS Emerging Viral Threat Laboratory | Microbial Genome Sequencing Center |
| EPI_ISL_451281 | LSUHS Emerging Viral Threat Laboratory | Microbial Genome Sequencing Center |
| EPI_ISL_451282 | LSUHS Emerging Viral Threat Laboratory | Microbial Genome Sequencing Center |
| EPI_ISL_451283 | LSUHS Emerging Viral Threat Laboratory | Microbial Genome Sequencing Center |
| EPI_ISL_451284 | LSUHS Emerging Viral Threat Laboratory | Microbial Genome Sequencing Center |
| EPI_ISL_451285 | LSUHS Emerging Viral Threat Laboratory | Microbial Genome Sequencing Center |
| EPI_ISL_451286 | LSUHS Emerging Viral Threat Laboratory | Microbial Genome Sequencing Center |
| EPI_ISL_451287 | LSUHS Emerging Viral Threat Laboratory | Microbial Genome Sequencing Center |
| EPI_ISL_451288 | LSUHS Emerging Viral Threat Laboratory | Microbial Genome Sequencing Center |
| EPI_ISL_451289 | LSUHS Emerging Viral Threat Laboratory | Microbial Genome Sequencing Center |
| EPI_ISL_451290 | LSUHS Emerging Viral Threat Laboratory | Microbial Genome Sequencing Center |
| EPI_ISL_451291 | LSUHS Emerging Viral Threat Laboratory | Microbial Genome Sequencing Center |
| EPI_ISL_451292 | LSUHS Emerging Viral Threat Laboratory | Microbial Genome Sequencing Center |
| EPI_ISL_451293 | LSUHS Emerging Viral Threat Laboratory | Microbial Genome Sequencing Center |
| EPI_ISL_451294 | LSUHS Emerging Viral Threat Laboratory | Microbial Genome Sequencing Center |
| EPI_ISL_451295 | LSUHS Emerging Viral Threat Laboratory | Microbial Genome Sequencing Center |
| EPI_ISL_451296 | LSUHS Emerging Viral Threat Laboratory | Microbial Genome Sequencing Center |
| EPI_ISL_451297 | LSUHS Emerging Viral Threat Laboratory | Microbial Genome Sequencing Center |
| EPI_ISL_451298 | Laboratory of Virology, INMI Lazzaro Spallanzani IRCCS | Laboratory of Virology, INMI Lazzaro Spallanzani IRCCS |

| EPI_Accession_ID | Originating Laboratory | Submitting Laboratory |
|---|---|---|
| EPI_ISL_451299 | Laboratory of Virology, INMI Lazzaro Spallanzani IRCCS | Laboratory of Virology, INMI Lazzaro Spallanzani IRCCS |
| EPI_ISL_451300 | Laboratory of Virology, INMI Lazzaro Spallanzani IRCCS | Laboratory of Virology, INMI Lazzaro Spallanzani IRCCS |
| EPI_ISL_451301 | Laboratory of Virology, INMI Lazzaro Spallanzani IRCCS | Laboratory of Virology, INMI Lazzaro Spallanzani IRCCS |
| EPI_ISL_451302 | Laboratory of Virology, INMI Lazzaro Spallanzani IRCCS | Laboratory of Virology, INMI Lazzaro Spallanzani IRCCS |
| EPI_ISL_451303 | Laboratory of Virology, INMI Lazzaro Spallanzani IRCCS | Laboratory of Virology, INMI Lazzaro Spallanzani IRCCS |
| EPI_ISL_451304 | Laboratory of Virology, INMI Lazzaro Spallanzani IRCCS | Laboratory of Virology, INMI Lazzaro Spallanzani IRCCS |
| EPI_ISL_451305 | Laboratory of Virology, INMI Lazzaro Spallanzani IRCCS | Laboratory of Virology, INMI Lazzaro Spallanzani IRCCS |
| EPI_ISL_451306 | Molecular Virology Unit, Fondazione IRCCS Policlinico San Matteo , Pavia | Laboratory of Virology, INMI Lazzaro Spallanzani IRCCS |
| EPI_ISL_451307 | Molecular Virology Unit, Fondazione IRCCS Policlinico San Matteo , Pavia | Laboratory of Virology, INMI Lazzaro Spallanzani IRCCS |
| EPI_ISL_451308 | Molecular Virology Unit, Fondazione IRCCS Policlinico San Matteo , Pavia | Laboratory of Virology, INMI Lazzaro Spallanzani IRCCS |
| EPI_ISL_451309 | Molecular Virology Unit, Fondazione IRCCS Policlinico San Matteo , Pavia | Laboratory of Virology, INMI Lazzaro Spallanzani IRCCS |
| EPI_ISL_451310 | Hellenic Pasteur Institute, National Influenza Reference laboratory of Southern Greece & Unit of Bioinformatics and Applied Genomics | Hellenic Pasteur Institute, National Influenza Reference laboratory of Southern Greece & Unit of Bioinformatics and Applied Genomics |
| EPI_ISL_451311 | Hellenic Pasteur Institute, National Influenza Reference laboratory of Southern Greece & Unit of Bioinformatics and Applied Genomics | Hellenic Pasteur Institute, National Influenza Reference laboratory of Southern Greece & Unit of Bioinformatics and Applied Genomics |
| EPI_ISL_451312 | West China Hospital of Sichuan University | State Key Laboratory of Biotherapy of Sichuan University |
| EPI_ISL_451313 | West China Hospital of Sichuan University | State Key Laboratory of Biotherapy of Sichuan University |
| EPI_ISL_451314 | West China Hospital of Sichuan University | State Key Laboratory of Biotherapy of Sichuan University |
| EPI_ISL_451315 | West China Hospital of Sichuan University | State Key Laboratory of Biotherapy of Sichuan University |
| EPI_ISL_451316 | West China Hospital of Sichuan University | State Key Laboratory of Biotherapy of Sichuan University |
| EPI_ISL_451317 | West China Hospital of Sichuan University | State Key Laboratory of Biotherapy of Sichuan University |

| EPI_Accession_ID | Originating Laboratory | Submitting Laboratory |
| --- | --- | --- |
| EPI_ISL_451318 | West China Hospital of Sichuan University | State Key Laboratory of Biotherapy of Sichuan University |
| EPI_ISL_451319 | West China Hospital of Sichuan University | State Key Laboratory of Biotherapy of Sichuan University |
| EPI_ISL_451320 | West China Hospital of Sichuan University | State Key Laboratory of Biotherapy of Sichuan University |
| EPI_ISL_451321 | West China Hospital of Sichuan University | State Key Laboratory of Biotherapy of Sichuan University |
| EPI_ISL_451322 | West China Hospital of Sichuan University | State Key Laboratory of Biotherapy of Sichuan University |
| EPI_ISL_451323 | West China Hospital of Sichuan University | State Key Laboratory of Biotherapy of Sichuan University |
| EPI_ISL_451324 | West China Hospital of Sichuan University | State Key Laboratory of Biotherapy of Sichuan University |
| EPI_ISL_451325 | West China Hospital of Sichuan University | State Key Laboratory of Biotherapy of Sichuan University |
| EPI_ISL_451326 | West China Hospital of Sichuan University | State Key Laboratory of Biotherapy of Sichuan University |
| EPI_ISL_451327 | West China Hospital of Sichuan University | State Key Laboratory of Biotherapy of Sichuan University |
| EPI_ISL_451328 | West China Hospital of Sichuan University | State Key Laboratory of Biotherapy of Sichuan University |
| EPI_ISL_451329 | West China Hospital of Sichuan University | State Key Laboratory of Biotherapy of Sichuan University |
| EPI_ISL_451330 | West China Hospital of Sichuan University | State Key Laboratory of Biotherapy of Sichuan University |
| EPI_ISL_451331 | West China Hospital of Sichuan University | State Key Laboratory of Biotherapy of Sichuan University |
| EPI_ISL_451332 | West China Hospital of Sichuan University | State Key Laboratory of Biotherapy of Sichuan University |
| EPI_ISL_451333 | West China Hospital of Sichuan University | State Key Laboratory of Biotherapy of Sichuan University |

| EPI_Accession_ID | Originating Laboratory | Submitting Laboratory |
|---|---|---|
| EPI_ISL_451334 | West China Hospital of Sichuan University | State Key Laboratory of Biotherapy of Sichuan University |
| EPI_ISL_451335 | West China Hospital of Sichuan University | State Key Laboratory of Biotherapy of Sichuan University |
| EPI_ISL_451336 | West China Hospital of Sichuan University | State Key Laboratory of Biotherapy of Sichuan University |
| EPI_ISL_451337 | West China Hospital of Sichuan University | State Key Laboratory of Biotherapy of Sichuan University |
| EPI_ISL_451338 | West China Hospital of Sichuan University | State Key Laboratory of Biotherapy of Sichuan University |
| EPI_ISL_451339 | West China Hospital of Sichuan University | State Key Laboratory of Biotherapy of Sichuan University |
| EPI_ISL_451340 | West China Hospital of Sichuan University | State Key Laboratory of Biotherapy of Sichuan University |
| EPI_ISL_451341 | West China Hospital of Sichuan University | State Key Laboratory of Biotherapy of Sichuan University |
| EPI_ISL_451342 | West China Hospital of Sichuan University | State Key Laboratory of Biotherapy of Sichuan University |
| EPI_ISL_451343 | West China Hospital of Sichuan University | State Key Laboratory of Biotherapy of Sichuan University |
| EPI_ISL_451344 | West China Hospital of Sichuan University | State Key Laboratory of Biotherapy of Sichuan University |
| EPI_ISL_451345 | West China Hospital of Sichuan University | State Key Laboratory of Biotherapy of Sichuan University |
| EPI_ISL_451346 | West China Hospital of Sichuan University | State Key Laboratory of Biotherapy of Sichuan University |
| EPI_ISL_451347 | West China Hospital of Sichuan University | State Key Laboratory of Biotherapy of Sichuan University |
| EPI_ISL_451348 | West China Hospital of Sichuan University | State Key Laboratory of Biotherapy of Sichuan University |
| EPI_ISL_451349 | West China Hospital of Sichuan University | State Key Laboratory of Biotherapy of Sichuan University |

| EPI_Accession_ID | Originating Laboratory | Submitting Laboratory |
|---|---|---|
| EPI_ISL_451350 | West China Hospital of Sichuan University | State Key Laboratory of Biotherapy of Sichuan University |
| EPI_ISL_451351 | West China Hospital of Sichuan University | State Key Laboratory of Biotherapy of Sichuan University |
| EPI_ISL_451352 | West China Hospital of Sichuan University | State Key Laboratory of Biotherapy of Sichuan University |
| EPI_ISL_451353 | West China Hospital of Sichuan University | State Key Laboratory of Biotherapy of Sichuan University |
| EPI_ISL_451354 | West China Hospital of Sichuan University | State Key Laboratory of Biotherapy of Sichuan University |
| EPI_ISL_451355 | West China Hospital of Sichuan University | State Key Laboratory of Biotherapy of Sichuan University |
| EPI_ISL_451356 | West China Hospital of Sichuan University | State Key Laboratory of Biotherapy of Sichuan University |
| EPI_ISL_451357 | West China Hospital of Sichuan University | State Key Laboratory of Biotherapy of Sichuan University |
| EPI_ISL_451358 | West China Hospital of Sichuan University | State Key Laboratory of Biotherapy of Sichuan University |
| EPI_ISL_451359 | West China Hospital of Sichuan University | State Key Laboratory of Biotherapy of Sichuan University |
| EPI_ISL_451360 | West China Hospital of Sichuan University | State Key Laboratory of Biotherapy of Sichuan University |
| EPI_ISL_451361 | West China Hospital of Sichuan University | State Key Laboratory of Biotherapy of Sichuan University |
| EPI_ISL_451362 | West China Hospital of Sichuan University | State Key Laboratory of Biotherapy of Sichuan University |
| EPI_ISL_451363 | West China Hospital of Sichuan University | State Key Laboratory of Biotherapy of Sichuan University |
| EPI_ISL_451364 | West China Hospital of Sichuan University | State Key Laboratory of Biotherapy of Sichuan University |
| EPI_ISL_451365 | West China Hospital of Sichuan University | State Key Laboratory of Biotherapy of Sichuan University |

| EPI_Accession_ID | Originating Laboratory | Submitting Laboratory |
|---|---|---|
| EPI_ISL_451366 | West China Hospital of Sichuan University | State Key Laboratory of Biotherapy of Sichuan University |
| EPI_ISL_451367 | West China Hospital of Sichuan University | State Key Laboratory of Biotherapy of Sichuan University |
| EPI_ISL_451368 | West China Hospital of Sichuan University | State Key Laboratory of Biotherapy of Sichuan University |
| EPI_ISL_451369 | West China Hospital of Sichuan University | State Key Laboratory of Biotherapy of Sichuan University |
| EPI_ISL_451370 | West China Hospital of Sichuan University | State Key Laboratory of Biotherapy of Sichuan University |
| EPI_ISL_451371 | West China Hospital of Sichuan University | State Key Laboratory of Biotherapy of Sichuan University |
| EPI_ISL_451372 | West China Hospital of Sichuan University | State Key Laboratory of Biotherapy of Sichuan University |
| EPI_ISL_451373 | West China Hospital of Sichuan University | State Key Laboratory of Biotherapy of Sichuan University |
| EPI_ISL_451374 | West China Hospital of Sichuan University | State Key Laboratory of Biotherapy of Sichuan University |
| EPI_ISL_451375 | West China Hospital of Sichuan University | State Key Laboratory of Biotherapy of Sichuan University |
| EPI_ISL_451376 | West China Hospital of Sichuan University | State Key Laboratory of Biotherapy of Sichuan University |
| EPI_ISL_451378 | West China Hospital of Sichuan University | State Key Laboratory of Biotherapy of Sichuan University |
| EPI_ISL_451379 | West China Hospital of Sichuan University | State Key Laboratory of Biotherapy of Sichuan University |
| EPI_ISL_451380 | West China Hospital of Sichuan University | State Key Laboratory of Biotherapy of Sichuan University |
| EPI_ISL_451381 | West China Hospital of Sichuan University | State Key Laboratory of Biotherapy of Sichuan University |
| EPI_ISL_451382 | West China Hospital of Sichuan University | State Key Laboratory of Biotherapy of Sichuan University |

| EPI_Accession_ID | Originating Laboratory | Submitting Laboratory |
|---|---|---|
| EPI_ISL_451383 | West China Hospital of Sichuan University | State Key Laboratory of Biotherapy of Sichuan University |
| EPI_ISL_451384 | West China Hospital of Sichuan University | State Key Laboratory of Biotherapy of Sichuan University |
| EPI_ISL_451385 | West China Hospital of Sichuan University | State Key Laboratory of Biotherapy of Sichuan University |
| EPI_ISL_451386 | West China Hospital of Sichuan University | State Key Laboratory of Biotherapy of Sichuan University |
| EPI_ISL_451387 | West China Hospital of Sichuan University | State Key Laboratory of Biotherapy of Sichuan University |
| EPI_ISL_451388 | West China Hospital of Sichuan University | State Key Laboratory of Biotherapy of Sichuan University |
| EPI_ISL_451389 | West China Hospital of Sichuan University | State Key Laboratory of Biotherapy of Sichuan University |
| EPI_ISL_451390 | West China Hospital of Sichuan University | State Key Laboratory of Biotherapy of Sichuan University |
| EPI_ISL_451391 | West China Hospital of Sichuan University | State Key Laboratory of Biotherapy of Sichuan University |
| EPI_ISL_451392 | West China Hospital of Sichuan University | State Key Laboratory of Biotherapy of Sichuan University |
| EPI_ISL_451393 | West China Hospital of Sichuan University | State Key Laboratory of Biotherapy of Sichuan University |
| EPI_ISL_451394 | West China Hospital of Sichuan University | State Key Laboratory of Biotherapy of Sichuan University |
| EPI_ISL_451395 | West China Hospital of Sichuan University | State Key Laboratory of Biotherapy of Sichuan University |
| EPI_ISL_451396 | West China Hospital of Sichuan University | State Key Laboratory of Biotherapy of Sichuan University |
| EPI_ISL_451397 | West China Hospital of Sichuan University | State Key Laboratory of Biotherapy of Sichuan University |
| EPI_ISL_451398 | West China Hospital of Sichuan University | State Key Laboratory of Biotherapy of Sichuan University |

| EPI_Accession_ID | Originating Laboratory | Submitting Laboratory |
|---|---|---|
| EPI_ISL_451399 | West China Hospital of Sichuan University | State Key Laboratory of Biotherapy of Sichuan University |
| EPI_ISL_451400 | Laboratoire de Recherche et d'Analyse MÃ©dicale de la Gendarmerie Royale | Laboratoire de Recherche et d'Analyse MÃ©dicale de la Gendarmerie Royale |
| EPI_ISL_451401 | NYU Langone Health | Departments of Pathology and Medicine, New York University School of Medicine |
| EPI_ISL_451402 | NYU Langone Health | Departments of Pathology and Medicine, New York University School of Medicine |
| EPI_ISL_451403 | NYU Langone Health | Departments of Pathology and Medicine, New York University School of Medicine |
| EPI_ISL_451404 | NYU Langone Health | Departments of Pathology and Medicine, New York University School of Medicine |
| EPI_ISL_451405 | NYU Langone Health | Departments of Pathology and Medicine, New York University School of Medicine |
| EPI_ISL_451406 | NYU Langone Health | Departments of Pathology and Medicine, New York University School of Medicine |
| EPI_ISL_451407 | NYU Langone Health | Departments of Pathology and Medicine, New York University School of Medicine |
| EPI_ISL_451408 | NYU Langone Health | Departments of Pathology and Medicine, New York University School of Medicine |
| EPI_ISL_451409 | NYU Langone Health | Departments of Pathology and Medicine, New York University School of Medicine |
| EPI_ISL_451410 | NYU Langone Health | Departments of Pathology and Medicine, New York University School of Medicine |
| EPI_ISL_451411 | NYU Langone Health | Departments of Pathology and Medicine, New York University School of Medicine |
| EPI_ISL_451412 | NYU Langone Health | Departments of Pathology and Medicine, New York University School of Medicine |
| EPI_ISL_451413 | NYU Langone Health | Departments of Pathology and Medicine, New York University School of Medicine |
| EPI_ISL_451414 | NYU Langone Health | Departments of Pathology and Medicine, New York University School of Medicine |

| EPI_Accession_ID | Originating Laboratory | Submitting Laboratory |
|---|---|---|
| EPI_ISL_451415 | NYU Langone Health | Departments of Pathology and Medicine, New York University School of Medicine |
| EPI_ISL_451416 | NYU Langone Health | Departments of Pathology and Medicine, New York University School of Medicine |
| EPI_ISL_451417 | NYU Langone Health | Departments of Pathology and Medicine, New York University School of Medicine |
| EPI_ISL_451418 | NYU Langone Health | Departments of Pathology and Medicine, New York University School of Medicine |
| EPI_ISL_451419 | NYU Langone Health | Departments of Pathology and Medicine, New York University School of Medicine |
| EPI_ISL_451420 | NYU Langone Health | Departments of Pathology and Medicine, New York University School of Medicine |
| EPI_ISL_451421 | NYU Langone Health | Departments of Pathology and Medicine, New York University School of Medicine |
| EPI_ISL_451422 | NYU Langone Health | Departments of Pathology and Medicine, New York University School of Medicine |
| EPI_ISL_451423 | NYU Langone Health | Departments of Pathology and Medicine, New York University School of Medicine |
| EPI_ISL_451424 | NYU Langone Health | Departments of Pathology and Medicine, New York University School of Medicine |
| EPI_ISL_451425 | NYU Langone Health | Departments of Pathology and Medicine, New York University School of Medicine |
| EPI_ISL_451426 | NYU Langone Health | Departments of Pathology and Medicine, New York University School of Medicine |
| EPI_ISL_451427 | NYU Langone Health | Departments of Pathology and Medicine, New York University School of Medicine |
| EPI_ISL_451428 | NYU Langone Health | Departments of Pathology and Medicine, New York University School of Medicine |
| EPI_ISL_451429 | NYU Langone Health | Departments of Pathology and Medicine, New York University School of Medicine |
| EPI_ISL_451430 | NYU Langone Health | Departments of Pathology and Medicine, New York University School of Medicine |

| EPI_Accession_ID | Originating Laboratory | Submitting Laboratory |
|---|---|---|
| EPI_ISL_451431 | NYU Langone Health | Departments of Pathology and Medicine, New York University School of Medicine |
| EPI_ISL_451432 | NYU Langone Health | Departments of Pathology and Medicine, New York University School of Medicine |
| EPI_ISL_451433 | NYU Langone Health | Departments of Pathology and Medicine, New York University School of Medicine |
| EPI_ISL_451434 | NYU Langone Health | Departments of Pathology and Medicine, New York University School of Medicine |
| EPI_ISL_451435 | NYU Langone Health | Departments of Pathology and Medicine, New York University School of Medicine |
| EPI_ISL_451436 | NYU Langone Health | Departments of Pathology and Medicine, New York University School of Medicine |
| EPI_ISL_451437 | NYU Langone Health | Departments of Pathology and Medicine, New York University School of Medicine |
| EPI_ISL_451438 | NYU Langone Health | Departments of Pathology and Medicine, New York University School of Medicine |
| EPI_ISL_451439 | NYU Langone Health | Departments of Pathology and Medicine, New York University School of Medicine |
| EPI_ISL_451440 | NYU Langone Health | Departments of Pathology and Medicine, New York University School of Medicine |
| EPI_ISL_451441 | NYU Langone Health | Departments of Pathology and Medicine, New York University School of Medicine |
| EPI_ISL_451442 | NYU Langone Health | Departments of Pathology and Medicine, New York University School of Medicine |
| EPI_ISL_451443 | NYU Langone Health | Departments of Pathology and Medicine, New York University School of Medicine |
| EPI_ISL_451444 | NYU Langone Health | Departments of Pathology and Medicine, New York University School of Medicine |
| EPI_ISL_451445 | NYU Langone Health | Departments of Pathology and Medicine, New York University School of Medicine |
| EPI_ISL_451446 | NYU Langone Health | Departments of Pathology and Medicine, New York University School of Medicine |

| EPI_Accession_ID | Originating Laboratory | Submitting Laboratory |
|---|---|---|
| EPI_ISL_451447 | NYU Langone Health | Departments of Pathology and Medicine, New York University School of Medicine |
| EPI_ISL_451448 | NYU Langone Health | Departments of Pathology and Medicine, New York University School of Medicine |
| EPI_ISL_451449 | NYU Langone Health | Departments of Pathology and Medicine, New York University School of Medicine |
| EPI_ISL_451450 | NYU Langone Health | Departments of Pathology and Medicine, New York University School of Medicine |
| EPI_ISL_451451 | NYU Langone Health | Departments of Pathology and Medicine, New York University School of Medicine |
| EPI_ISL_451452 | NYU Langone Health | Departments of Pathology and Medicine, New York University School of Medicine |
| EPI_ISL_451453 | NYU Langone Health | Departments of Pathology and Medicine, New York University School of Medicine |
| EPI_ISL_451454 | NYU Langone Health | Departments of Pathology and Medicine, New York University School of Medicine |
| EPI_ISL_451455 | NYU Langone Health | Departments of Pathology and Medicine, New York University School of Medicine |
| EPI_ISL_451456 | NYU Langone Health | Departments of Pathology and Medicine, New York University School of Medicine |
| EPI_ISL_451457 | NYU Langone Health | Departments of Pathology and Medicine, New York University School of Medicine |
| EPI_ISL_451458 | NYU Langone Health | Departments of Pathology and Medicine, New York University School of Medicine |
| EPI_ISL_451459 | NYU Langone Health | Departments of Pathology and Medicine, New York University School of Medicine |
| EPI_ISL_451460 | NYU Langone Health | Departments of Pathology and Medicine, New York University School of Medicine |
| EPI_ISL_451461 | NYU Langone Health | Departments of Pathology and Medicine, New York University School of Medicine |
| EPI_ISL_451462 | NYU Langone Health | Departments of Pathology and Medicine, New York University School of Medicine |

| EPI_Accession_ID | Originating Laboratory | Submitting Laboratory |
|---|---|---|
| EPI_ISL_451463 | NYU Langone Health | Departments of Pathology and Medicine, New York University School of Medicine |
| EPI_ISL_451464 | NYU Langone Health | Departments of Pathology and Medicine, New York University School of Medicine |
| EPI_ISL_451465 | NYU Langone Health | Departments of Pathology and Medicine, New York University School of Medicine |
| EPI_ISL_451466 | NYU Langone Health | Departments of Pathology and Medicine, New York University School of Medicine |
| EPI_ISL_451467 | NYU Langone Health | Departments of Pathology and Medicine, New York University School of Medicine |
| EPI_ISL_451468 | NYU Langone Health | Departments of Pathology and Medicine, New York University School of Medicine |
| EPI_ISL_451469 | NYU Langone Health | Departments of Pathology and Medicine, New York University School of Medicine |
| EPI_ISL_451470 | NYU Langone Health | Departments of Pathology and Medicine, New York University School of Medicine |
| EPI_ISL_451471 | NYU Langone Health | Departments of Pathology and Medicine, New York University School of Medicine |
| EPI_ISL_451472 | NYU Langone Health | Departments of Pathology and Medicine, New York University School of Medicine |
| EPI_ISL_451473 | NYU Langone Health | Departments of Pathology and Medicine, New York University School of Medicine |
| EPI_ISL_451474 | NYU Langone Health | Departments of Pathology and Medicine, New York University School of Medicine |
| EPI_ISL_451475 | NYU Langone Health | Departments of Pathology and Medicine, New York University School of Medicine |
| EPI_ISL_451476 | NYU Langone Health | Departments of Pathology and Medicine, New York University School of Medicine |
| EPI_ISL_451477 | NYU Langone Health | Departments of Pathology and Medicine, New York University School of Medicine |
| EPI_ISL_451478 | NYU Langone Health | Departments of Pathology and Medicine, New York University School of Medicine |

| EPI_Accession_ID | Originating Laboratory | Submitting Laboratory |
|---|---|---|
| EPI_ISL_451479 | NYU Langone Health | Departments of Pathology and Medicine, New York University School of Medicine |
| EPI_ISL_451486 | Australian Clinical Labs | NSW Health Pathology - Institute of Clinical Pathology and Medical Research; Westmead Hospital; University of Sydney |
| EPI_ISL_451487 | Pathology North - NSW Health Pathology | NSW Health Pathology - Institute of Clinical Pathology and Medical Research; Westmead Hospital; University of Sydney |
| EPI_ISL_451488 | Pathology North - NSW Health Pathology | NSW Health Pathology - Institute of Clinical Pathology and Medical Research; Westmead Hospital; University of Sydney |
| EPI_ISL_451489 | Laverty Pathology | NSW Health Pathology - Institute of Clinical Pathology and Medical Research; Westmead Hospital; University of Sydney |
| EPI_ISL_451490 | Pathology West - NSW Health Pathology | NSW Health Pathology - Institute of Clinical Pathology and Medical Research; Westmead Hospital; University of Sydney |
| EPI_ISL_451491 | Pathology West - NSW Health Pathology | NSW Health Pathology - Institute of Clinical Pathology and Medical Research; Westmead Hospital; University of Sydney |
| EPI_ISL_451492 | Pathology West - NSW Health Pathology | NSW Health Pathology - Institute of Clinical Pathology and Medical Research; Westmead Hospital; University of Sydney |
| EPI_ISL_451493 | Pathology West - NSW Health Pathology | NSW Health Pathology - Institute of Clinical Pathology and Medical Research; Westmead Hospital; University of Sydney |
| EPI_ISL_451494 | Pathology West - NSW Health Pathology | NSW Health Pathology - Institute of Clinical Pathology and Medical Research; Westmead Hospital; University of Sydney |
| EPI_ISL_451495 | Pathology West - NSW Health Pathology | NSW Health Pathology - Institute of Clinical Pathology and Medical Research; Westmead Hospital; University of Sydney |

| EPI_Accession_ID | Originating Laboratory | Submitting Laboratory |
|---|---|---|
| EPI_ISL_451496 | Pathology West - NSW Health Pathology | NSW Health Pathology - Institute of Clinical Pathology and Medical Research; Westmead Hospital; University of Sydney |
| EPI_ISL_451497 | Pathology West - NSW Health Pathology | NSW Health Pathology - Institute of Clinical Pathology and Medical Research; Westmead Hospital; University of Sydney |
| EPI_ISL_451498 | Pathology West - NSW Health Pathology | NSW Health Pathology - Institute of Clinical Pathology and Medical Research; Westmead Hospital; University of Sydney |
| EPI_ISL_451499 | Pathology West - NSW Health Pathology | NSW Health Pathology - Institute of Clinical Pathology and Medical Research; Westmead Hospital; University of Sydney |
| EPI_ISL_451500 | Pathology West - NSW Health Pathology | NSW Health Pathology - Institute of Clinical Pathology and Medical Research; Westmead Hospital; University of Sydney |
| EPI_ISL_451501 | Pathology West - NSW Health Pathology | NSW Health Pathology - Institute of Clinical Pathology and Medical Research; Westmead Hospital; University of Sydney |
| EPI_ISL_451502 | Pathology West - NSW Health Pathology | NSW Health Pathology - Institute of Clinical Pathology and Medical Research; Westmead Hospital; University of Sydney |
| EPI_ISL_451503 | Pathology West - NSW Health Pathology | NSW Health Pathology - Institute of Clinical Pathology and Medical Research; Westmead Hospital; University of Sydney |
| EPI_ISL_451504 | Pathology West - NSW Health Pathology | NSW Health Pathology - Institute of Clinical Pathology and Medical Research; Westmead Hospital; University of Sydney |
| EPI_ISL_451505 | Pathology West - NSW Health Pathology | NSW Health Pathology - Institute of Clinical Pathology and Medical Research; Westmead Hospital; University of Sydney |
| EPI_ISL_451506 | Pathology West - NSW Health Pathology | NSW Health Pathology - Institute of Clinical Pathology and Medical Research; Westmead Hospital; University of Sydney |

| EPI_Accession_ID | Originating Laboratory | Submitting Laboratory |
|---|---|---|
| EPI_ISL_451507 | Pathology West - NSW Health Pathology | NSW Health Pathology - Institute of Clinical Pathology and Medical Research; Westmead Hospital; University of Sydney |
| EPI_ISL_451508 | Pathology West - NSW Health Pathology | NSW Health Pathology - Institute of Clinical Pathology and Medical Research; Westmead Hospital; University of Sydney |
| EPI_ISL_451509 | Pathology West - NSW Health Pathology | NSW Health Pathology - Institute of Clinical Pathology and Medical Research; Westmead Hospital; University of Sydney |
| EPI_ISL_451510 | Pathology West - NSW Health Pathology | NSW Health Pathology - Institute of Clinical Pathology and Medical Research; Westmead Hospital; University of Sydney |
| EPI_ISL_451511 | Pathology West - NSW Health Pathology | NSW Health Pathology - Institute of Clinical Pathology and Medical Research; Westmead Hospital; University of Sydney |
| EPI_ISL_451512 | Pathology West - NSW Health Pathology | NSW Health Pathology - Institute of Clinical Pathology and Medical Research; Westmead Hospital; University of Sydney |
| EPI_ISL_451513 | Pathology West - NSW Health Pathology | NSW Health Pathology - Institute of Clinical Pathology and Medical Research; Westmead Hospital; University of Sydney |
| EPI_ISL_451514 | Pathology West - NSW Health Pathology | NSW Health Pathology - Institute of Clinical Pathology and Medical Research; Westmead Hospital; University of Sydney |
| EPI_ISL_451515 | Pathology West - NSW Health Pathology | NSW Health Pathology - Institute of Clinical Pathology and Medical Research; Westmead Hospital; University of Sydney |
| EPI_ISL_451516 | Pathology West - NSW Health Pathology | NSW Health Pathology - Institute of Clinical Pathology and Medical Research; Westmead Hospital; University of Sydney |
| EPI_ISL_451517 | South Eastern Area Laboratory Services | NSW Health Pathology - Institute of Clinical Pathology and Medical Research; Westmead Hospital; University of Sydney |

| EPI_Accession_ID | Originating Laboratory | Submitting Laboratory |
|---|---|---|
| EPI_ISL_451518 | South Eastern Area Laboratory Services | NSW Health Pathology - Institute of Clinical Pathology and Medical Research; Westmead Hospital; University of Sydney |
| EPI_ISL_451519 | South Eastern Area Laboratory Services | NSW Health Pathology - Institute of Clinical Pathology and Medical Research; Westmead Hospital; University of Sydney |
| EPI_ISL_451520 | South Eastern Area Laboratory Services | NSW Health Pathology - Institute of Clinical Pathology and Medical Research; Westmead Hospital; University of Sydney |
| EPI_ISL_451521 | South Eastern Area Laboratory Services | NSW Health Pathology - Institute of Clinical Pathology and Medical Research; Westmead Hospital; University of Sydney |
| EPI_ISL_451522 | Pathology West - NSW Health Pathology | NSW Health Pathology - Institute of Clinical Pathology and Medical Research; Westmead Hospital; University of Sydney |
| EPI_ISL_451523 | Pathology West - NSW Health Pathology | NSW Health Pathology - Institute of Clinical Pathology and Medical Research; Westmead Hospital; University of Sydney |
| EPI_ISL_451524 | Pathology West - NSW Health Pathology | NSW Health Pathology - Institute of Clinical Pathology and Medical Research; Westmead Hospital; University of Sydney |
| EPI_ISL_451525 | Pathology West - NSW Health Pathology | NSW Health Pathology - Institute of Clinical Pathology and Medical Research; Westmead Hospital; University of Sydney |
| EPI_ISL_451526 | Pathology West - NSW Health Pathology | NSW Health Pathology - Institute of Clinical Pathology and Medical Research; Westmead Hospital; University of Sydney |
| EPI_ISL_451527 | Pathology West - NSW Health Pathology | NSW Health Pathology - Institute of Clinical Pathology and Medical Research; Westmead Hospital; University of Sydney |
| EPI_ISL_451528 | Pathology West - NSW Health Pathology | NSW Health Pathology - Institute of Clinical Pathology and Medical Research; Westmead Hospital; University of Sydney |

| EPI_Accession_ID | Originating Laboratory | Submitting Laboratory |
|---|---|---|
| EPI_ISL_451529 | Pathology West - NSW Health Pathology | NSW Health Pathology - Institute of Clinical Pathology and Medical Research; Westmead Hospital; University of Sydney |
| EPI_ISL_451530 | Pathology Sydney South West - NSW Health Pathology | NSW Health Pathology - Institute of Clinical Pathology and Medical Research; Westmead Hospital; University of Sydney |
| EPI_ISL_451531 | Pathology Sydney South West - NSW Health Pathology | NSW Health Pathology - Institute of Clinical Pathology and Medical Research; Westmead Hospital; University of Sydney |
| EPI_ISL_451532 | Pathology West - NSW Health Pathology | NSW Health Pathology - Institute of Clinical Pathology and Medical Research; Westmead Hospital; University of Sydney |
| EPI_ISL_451533 | Pathology Sydney South West - NSW Health Pathology | NSW Health Pathology - Institute of Clinical Pathology and Medical Research; Westmead Hospital; University of Sydney |
| EPI_ISL_451534 | Medlab Pathology | NSW Health Pathology - Institute of Clinical Pathology and Medical Research; Westmead Hospital; University of Sydney |
| EPI_ISL_451535 | Pathology West - NSW Health Pathology | NSW Health Pathology - Institute of Clinical Pathology and Medical Research; Westmead Hospital; University of Sydney |
| EPI_ISL_451536 | Pathology Sydney South West - NSW Health Pathology | NSW Health Pathology - Institute of Clinical Pathology and Medical Research; Westmead Hospital; University of Sydney |
| EPI_ISL_451537 | Pathology West - NSW Health Pathology | NSW Health Pathology - Institute of Clinical Pathology and Medical Research; Westmead Hospital; University of Sydney |
| EPI_ISL_451538 | Pathology West - NSW Health Pathology | NSW Health Pathology - Institute of Clinical Pathology and Medical Research; Westmead Hospital; University of Sydney |
| EPI_ISL_451539 | Pathology West - NSW Health Pathology | NSW Health Pathology - Institute of Clinical Pathology and Medical Research; Westmead Hospital; University of Sydney |

| EPI_Accession_ID | Originating Laboratory | Submitting Laboratory |
|---|---|---|
| EPI_ISL_451540 | ACT pathology | NSW Health Pathology - Institute of Clinical Pathology and Medical Research; Westmead Hospital; University of Sydney |
| EPI_ISL_451541 | Pathology West - NSW Health Pathology | NSW Health Pathology - Institute of Clinical Pathology and Medical Research; Westmead Hospital; University of Sydney |
| EPI_ISL_451542 | Pathology Sydney South West - NSW Health Pathology | NSW Health Pathology - Institute of Clinical Pathology and Medical Research; Westmead Hospital; University of Sydney |
| EPI_ISL_451543 | Pathology Sydney South West - NSW Health Pathology | NSW Health Pathology - Institute of Clinical Pathology and Medical Research; Westmead Hospital; University of Sydney |
| EPI_ISL_451544 | Pathology Sydney South West - NSW Health Pathology | NSW Health Pathology - Institute of Clinical Pathology and Medical Research; Westmead Hospital; University of Sydney |
| EPI_ISL_451545 | Laverty Pathology | NSW Health Pathology - Institute of Clinical Pathology and Medical Research; Westmead Hospital; University of Sydney |
| EPI_ISL_451546 | Pathology West - NSW Health Pathology | NSW Health Pathology - Institute of Clinical Pathology and Medical Research; Westmead Hospital; University of Sydney |
| EPI_ISL_451547 | Pathology West - NSW Health Pathology | NSW Health Pathology - Institute of Clinical Pathology and Medical Research; Westmead Hospital; University of Sydney |
| EPI_ISL_451548 | Childrens Hospital Westmead | NSW Health Pathology - Institute of Clinical Pathology and Medical Research; Westmead Hospital; University of Sydney |
| EPI_ISL_451549 | Pathology Sydney South West - NSW Health Pathology | NSW Health Pathology - Institute of Clinical Pathology and Medical Research; Westmead Hospital; University of Sydney |
| EPI_ISL_451550 | Pathology West - NSW Health Pathology | NSW Health Pathology - Institute of Clinical Pathology and Medical Research; Westmead Hospital; University of Sydney |

| EPI_Accession_ID | Originating Laboratory | Submitting Laboratory |
|---|---|---|
| EPI_ISL_451551 | Pathology West - NSW Health Pathology | NSW Health Pathology - Institute of Clinical Pathology and Medical Research; Westmead Hospital; University of Sydney |
| EPI_ISL_451552 | Pathology North - NSW Health Pathology | NSW Health Pathology - Institute of Clinical Pathology and Medical Research; Westmead Hospital; University of Sydney |
| EPI_ISL_451553 | Medlab Pathology | NSW Health Pathology - Institute of Clinical Pathology and Medical Research; Westmead Hospital; University of Sydney |
| EPI_ISL_451554 | Medlab Pathology | NSW Health Pathology - Institute of Clinical Pathology and Medical Research; Westmead Hospital; University of Sydney |
| EPI_ISL_451555 | Medlab Pathology | NSW Health Pathology - Institute of Clinical Pathology and Medical Research; Westmead Hospital; University of Sydney |
| EPI_ISL_451556 | Pathology West - NSW Health Pathology | NSW Health Pathology - Institute of Clinical Pathology and Medical Research; Westmead Hospital; University of Sydney |
| EPI_ISL_451557 | Pathology West - NSW Health Pathology | NSW Health Pathology - Institute of Clinical Pathology and Medical Research; Westmead Hospital; University of Sydney |
| EPI_ISL_451558 | Medlab Pathology | NSW Health Pathology - Institute of Clinical Pathology and Medical Research; Westmead Hospital; University of Sydney |
| EPI_ISL_451559 | Pathology Sydney South West - NSW Health Pathology | NSW Health Pathology - Institute of Clinical Pathology and Medical Research; Westmead Hospital; University of Sydney |
| EPI_ISL_451560 | Pathology Sydney South West - NSW Health Pathology | NSW Health Pathology - Institute of Clinical Pathology and Medical Research; Westmead Hospital; University of Sydney |
| EPI_ISL_451561 | Pathology Sydney South West - NSW Health Pathology | NSW Health Pathology - Institute of Clinical Pathology and Medical Research; Westmead Hospital; University of Sydney |

| EPI_Accession_ID | Originating Laboratory | Submitting Laboratory |
|---|---|---|
| EPI_ISL_451562 | Pathology Sydney South West - NSW Health Pathology | NSW Health Pathology - Institute of Clinical Pathology and Medical Research; Westmead Hospital; University of Sydney |
| EPI_ISL_451563 | Pathology Sydney South West - NSW Health Pathology | NSW Health Pathology - Institute of Clinical Pathology and Medical Research; Westmead Hospital; University of Sydney |
| EPI_ISL_451564 | Pathology Sydney South West - NSW Health Pathology | NSW Health Pathology - Institute of Clinical Pathology and Medical Research; Westmead Hospital; University of Sydney |
| EPI_ISL_451565 | Pathology Sydney South West - NSW Health Pathology | NSW Health Pathology - Institute of Clinical Pathology and Medical Research; Westmead Hospital; University of Sydney |
| EPI_ISL_451566 | Pathology West - NSW Health Pathology | NSW Health Pathology - Institute of Clinical Pathology and Medical Research; Westmead Hospital; University of Sydney |
| EPI_ISL_451567 | Medlab Pathology | NSW Health Pathology - Institute of Clinical Pathology and Medical Research; Westmead Hospital; University of Sydney |
| EPI_ISL_451568 | Pathology West - NSW Health Pathology | NSW Health Pathology - Institute of Clinical Pathology and Medical Research; Westmead Hospital; University of Sydney |
| EPI_ISL_451569 | Pathology West - NSW Health Pathology | NSW Health Pathology - Institute of Clinical Pathology and Medical Research; Westmead Hospital; University of Sydney |
| EPI_ISL_451570 | Pathology West - NSW Health Pathology | NSW Health Pathology - Institute of Clinical Pathology and Medical Research; Westmead Hospital; University of Sydney |
| EPI_ISL_451571 | Pathology West - NSW Health Pathology | NSW Health Pathology - Institute of Clinical Pathology and Medical Research; Westmead Hospital; University of Sydney |
| EPI_ISL_451572 | Laverty Pathology | NSW Health Pathology - Institute of Clinical Pathology and Medical Research; Westmead Hospital; University of Sydney |

| EPI_Accession_ID | Originating Laboratory | Submitting Laboratory |
|---|---|---|
| EPI_ISL_451573 | Pathology West - NSW Health Pathology | NSW Health Pathology - Institute of Clinical Pathology and Medical Research; Westmead Hospital; University of Sydney |
| EPI_ISL_451574 | Australian Clinical Labs | NSW Health Pathology - Institute of Clinical Pathology and Medical Research; Westmead Hospital; University of Sydney |
| EPI_ISL_451575 | Pathology West - NSW Health Pathology | NSW Health Pathology - Institute of Clinical Pathology and Medical Research; Westmead Hospital; University of Sydney |
| EPI_ISL_451576 | Pathology West - NSW Health Pathology | NSW Health Pathology - Institute of Clinical Pathology and Medical Research; Westmead Hospital; University of Sydney |
| EPI_ISL_451577 | Pathology West - NSW Health Pathology | NSW Health Pathology - Institute of Clinical Pathology and Medical Research; Westmead Hospital; University of Sydney |
| EPI_ISL_451578 | Pathology Sydney South West - NSW Health Pathology | NSW Health Pathology - Institute of Clinical Pathology and Medical Research; Westmead Hospital; University of Sydney |
| EPI_ISL_451579 | Pathology West - NSW Health Pathology | NSW Health Pathology - Institute of Clinical Pathology and Medical Research; Westmead Hospital; University of Sydney |
| EPI_ISL_451580 | Pathology West - NSW Health Pathology | NSW Health Pathology - Institute of Clinical Pathology and Medical Research; Westmead Hospital; University of Sydney |
| EPI_ISL_451581 | Pathology Sydney South West - NSW Health Pathology | NSW Health Pathology - Institute of Clinical Pathology and Medical Research; Westmead Hospital; University of Sydney |
| EPI_ISL_451582 | Pathology West - NSW Health Pathology | NSW Health Pathology - Institute of Clinical Pathology and Medical Research; Westmead Hospital; University of Sydney |
| EPI_ISL_451583 | Medlab Pathology | NSW Health Pathology - Institute of Clinical Pathology and Medical Research; Westmead Hospital; University of Sydney |

| EPI_Accession_ID | Originating Laboratory | Submitting Laboratory |
|---|---|---|
| EPI_ISL_451584 | Pathology West - NSW Health Pathology | NSW Health Pathology - Institute of Clinical Pathology and Medical Research; Westmead Hospital; University of Sydney |
| EPI_ISL_451585 | Pathology West - NSW Health Pathology | NSW Health Pathology - Institute of Clinical Pathology and Medical Research; Westmead Hospital; University of Sydney |
| EPI_ISL_451586 | Pathology West - NSW Health Pathology | NSW Health Pathology - Institute of Clinical Pathology and Medical Research; Westmead Hospital; University of Sydney |
| EPI_ISL_451587 | Pathology North Hunter- NSW Health Pathology | NSW Health Pathology - Institute of Clinical Pathology and Medical Research; Westmead Hospital; University of Sydney |
| EPI_ISL_451588 | ACT pathology | NSW Health Pathology - Institute of Clinical Pathology and Medical Research; Westmead Hospital; University of Sydney |
| EPI_ISL_451589 | Pathology North Hunter- NSW Health Pathology | NSW Health Pathology - Institute of Clinical Pathology and Medical Research; Westmead Hospital; University of Sydney |
| EPI_ISL_451590 | ACT pathology | NSW Health Pathology - Institute of Clinical Pathology and Medical Research; Westmead Hospital; University of Sydney |
| EPI_ISL_451591 | ACT pathology | NSW Health Pathology - Institute of Clinical Pathology and Medical Research; Westmead Hospital; University of Sydney |
| EPI_ISL_451592 | ACT pathology | NSW Health Pathology - Institute of Clinical Pathology and Medical Research; Westmead Hospital; University of Sydney |
| EPI_ISL_451593 | ACT pathology | NSW Health Pathology - Institute of Clinical Pathology and Medical Research; Westmead Hospital; University of Sydney |
| EPI_ISL_451594 | Childrens Hospital Westmead | NSW Health Pathology - Institute of Clinical Pathology and Medical Research; Westmead Hospital; University of Sydney |

| EPI_Accession_ID | Originating Laboratory | Submitting Laboratory |
|---|---|---|
| EPI_ISL_451595 | ACT pathology | NSW Health Pathology - Institute of Clinical Pathology and Medical Research; Westmead Hospital; University of Sydney |
| EPI_ISL_451596 | ACT pathology | NSW Health Pathology - Institute of Clinical Pathology and Medical Research; Westmead Hospital; University of Sydney |
| EPI_ISL_451597 | Medlab Pathology | NSW Health Pathology - Institute of Clinical Pathology and Medical Research; Westmead Hospital; University of Sydney |
| EPI_ISL_451598 | ACT pathology | NSW Health Pathology - Institute of Clinical Pathology and Medical Research; Westmead Hospital; University of Sydney |
| EPI_ISL_451599 | Australian Clinical Labs | NSW Health Pathology - Institute of Clinical Pathology and Medical Research; Westmead Hospital; University of Sydney |
| EPI_ISL_451600 | Pathology North Hunter- NSW Health Pathology | NSW Health Pathology - Institute of Clinical Pathology and Medical Research; Westmead Hospital; University of Sydney |
| EPI_ISL_451601 | Douglas Hanly Moir Pathology | NSW Health Pathology - Institute of Clinical Pathology and Medical Research; Westmead Hospital; University of Sydney |
| EPI_ISL_451602 | Laverty Pathology | NSW Health Pathology - Institute of Clinical Pathology and Medical Research; Westmead Hospital; University of Sydney |
| EPI_ISL_451603 | Pathology Sydney South West - NSW Health Pathology | NSW Health Pathology - Institute of Clinical Pathology and Medical Research; Westmead Hospital; University of Sydney |
| EPI_ISL_451604 | Pathology North - NSW Health Pathology | NSW Health Pathology - Institute of Clinical Pathology and Medical Research; Westmead Hospital; University of Sydney |
| EPI_ISL_451605 | Childrens Hospital Westmead | NSW Health Pathology - Institute of Clinical Pathology and Medical Research; Westmead Hospital; University of Sydney |

| EPI_Accession_ID | Originating Laboratory | Submitting Laboratory |
|---|---|---|
| EPI_ISL_451606 | Pathology West - NSW Health Pathology | NSW Health Pathology - Institute of Clinical Pathology and Medical Research; Westmead Hospital; University of Sydney |
| EPI_ISL_451607 | Pathology West - NSW Health Pathology | NSW Health Pathology - Institute of Clinical Pathology and Medical Research; Westmead Hospital; University of Sydney |
| EPI_ISL_451608 | Pathology Sydney South West - NSW Health Pathology | NSW Health Pathology - Institute of Clinical Pathology and Medical Research; Westmead Hospital; University of Sydney |
| EPI_ISL_451609 | Laverty Pathology | NSW Health Pathology - Institute of Clinical Pathology and Medical Research; Westmead Hospital; University of Sydney |
| EPI_ISL_451610 | Medlab Pathology | NSW Health Pathology - Institute of Clinical Pathology and Medical Research; Westmead Hospital; University of Sydney |
| EPI_ISL_451611 | Laverty Pathology | NSW Health Pathology - Institute of Clinical Pathology and Medical Research; Westmead Hospital; University of Sydney |
| EPI_ISL_451612 | ACT pathology | NSW Health Pathology - Institute of Clinical Pathology and Medical Research; Westmead Hospital; University of Sydney |
| EPI_ISL_451613 | Pathology West - NSW Health Pathology | NSW Health Pathology - Institute of Clinical Pathology and Medical Research; Westmead Hospital; University of Sydney |
| EPI_ISL_451614 | Pathology West - NSW Health Pathology | NSW Health Pathology - Institute of Clinical Pathology and Medical Research; Westmead Hospital; University of Sydney |
| EPI_ISL_451615 | Pathology West - NSW Health Pathology | NSW Health Pathology - Institute of Clinical Pathology and Medical Research; Westmead Hospital; University of Sydney |
| EPI_ISL_451616 | Pathology West - NSW Health Pathology | NSW Health Pathology - Institute of Clinical Pathology and Medical Research; Westmead Hospital; University of Sydney |

| EPI_Accession_ID | Originating Laboratory | Submitting Laboratory |
|---|---|---|
| EPI_ISL_451617 | Pathology West - NSW Health Pathology | NSW Health Pathology - Institute of Clinical Pathology and Medical Research; Westmead Hospital; University of Sydney |
| EPI_ISL_451618 | Pathology West - NSW Health Pathology | NSW Health Pathology - Institute of Clinical Pathology and Medical Research; Westmead Hospital; University of Sydney |
| EPI_ISL_451619 | Pathology West - NSW Health Pathology | NSW Health Pathology - Institute of Clinical Pathology and Medical Research; Westmead Hospital; University of Sydney |
| EPI_ISL_451620 | Pathology West - NSW Health Pathology | NSW Health Pathology - Institute of Clinical Pathology and Medical Research; Westmead Hospital; University of Sydney |
| EPI_ISL_451621 | Pathology West - NSW Health Pathology | NSW Health Pathology - Institute of Clinical Pathology and Medical Research; Westmead Hospital; University of Sydney |
| EPI_ISL_451622 | Pathology West - NSW Health Pathology | NSW Health Pathology - Institute of Clinical Pathology and Medical Research; Westmead Hospital; University of Sydney |
| EPI_ISL_451623 | Pathology West - NSW Health Pathology | NSW Health Pathology - Institute of Clinical Pathology and Medical Research; Westmead Hospital; University of Sydney |
| EPI_ISL_451624 | Pathology West - NSW Health Pathology | NSW Health Pathology - Institute of Clinical Pathology and Medical Research; Westmead Hospital; University of Sydney |
| EPI_ISL_451625 | Pathology West - NSW Health Pathology | NSW Health Pathology - Institute of Clinical Pathology and Medical Research; Westmead Hospital; University of Sydney |
| EPI_ISL_451626 | Pathology West - NSW Health Pathology | NSW Health Pathology - Institute of Clinical Pathology and Medical Research; Westmead Hospital; University of Sydney |
| EPI_ISL_451627 | Pathology West - NSW Health Pathology | NSW Health Pathology - Institute of Clinical Pathology and Medical Research; Westmead Hospital; University of Sydney |

| EPI_Accession_ID | Originating Laboratory | Submitting Laboratory |
|---|---|---|
| EPI_ISL_451628 | Pathology West - NSW Health Pathology | NSW Health Pathology - Institute of Clinical Pathology and Medical Research; Westmead Hospital; University of Sydney |
| EPI_ISL_451629 | Pathology West - NSW Health Pathology | NSW Health Pathology - Institute of Clinical Pathology and Medical Research; Westmead Hospital; University of Sydney |
| EPI_ISL_451630 | Pathology West - NSW Health Pathology | NSW Health Pathology - Institute of Clinical Pathology and Medical Research; Westmead Hospital; University of Sydney |
| EPI_ISL_451631 | Laverty Pathology | NSW Health Pathology - Institute of Clinical Pathology and Medical Research; Westmead Hospital; University of Sydney |
| EPI_ISL_451632 | South Eastern Area Laboratory Services | NSW Health Pathology - Institute of Clinical Pathology and Medical Research; Westmead Hospital; University of Sydney |
| EPI_ISL_451633 | South Eastern Area Laboratory Services | NSW Health Pathology - Institute of Clinical Pathology and Medical Research; Westmead Hospital; University of Sydney |
| EPI_ISL_451634 | South Eastern Area Laboratory Services | NSW Health Pathology - Institute of Clinical Pathology and Medical Research; Westmead Hospital; University of Sydney |
| EPI_ISL_451635 | South Eastern Area Laboratory Services | NSW Health Pathology - Institute of Clinical Pathology and Medical Research; Westmead Hospital; University of Sydney |
| EPI_ISL_451636 | South Eastern Area Laboratory Services | NSW Health Pathology - Institute of Clinical Pathology and Medical Research; Westmead Hospital; University of Sydney |
| EPI_ISL_451637 | Pathology West - NSW Health Pathology | NSW Health Pathology - Institute of Clinical Pathology and Medical Research; Westmead Hospital; University of Sydney |
| EPI_ISL_451638 | Pathology West - NSW Health Pathology | NSW Health Pathology - Institute of Clinical Pathology and Medical Research; Westmead Hospital; University of Sydney |

| EPI_Accession_ID | Originating Laboratory | Submitting Laboratory |
|---|---|---|
| EPI_ISL_451639 | Pathology West - NSW Health Pathology | NSW Health Pathology - Institute of Clinical Pathology and Medical Research; Westmead Hospital; University of Sydney |
| EPI_ISL_451640 | Pathology West - NSW Health Pathology | NSW Health Pathology - Institute of Clinical Pathology and Medical Research; Westmead Hospital; University of Sydney |
| EPI_ISL_451641 | Laverty Pathology | NSW Health Pathology - Institute of Clinical Pathology and Medical Research; Westmead Hospital; University of Sydney |
| EPI_ISL_451642 | Pathology Sydney South West - NSW Health Pathology | NSW Health Pathology - Institute of Clinical Pathology and Medical Research; Westmead Hospital; University of Sydney |
| EPI_ISL_451643 | Pathology Sydney South West - NSW Health Pathology | NSW Health Pathology - Institute of Clinical Pathology and Medical Research; Westmead Hospital; University of Sydney |
| EPI_ISL_451644 | Laboratory of Molecular Biology, Diagnostyka sp. z o.o. | Laboratory of Recombinant Vaccines |
| EPI_ISL_451645 | Laboratory of Molecular Biology, Diagnostyka sp. z o.o. | Laboratory of Recombinant Vaccines |
| EPI_ISL_451646 | Laboratory of Molecular Biology, Diagnostyka sp. z o.o. | Laboratory of Recombinant Vaccines |
| EPI_ISL_451647 | Laboratory of Molecular Biology, Diagnostyka sp. z o.o. | Laboratory of Recombinant Vaccines |
| EPI_ISL_451648 | Hematology Laboratory, Section of Molecular Diagnostics, University Clinical Centre, Medical University of Gdansk | Laboratory of Recombinant Vaccines |
| EPI_ISL_451649 | Hematology Laboratory, Section of Molecular Diagnostics, University Clinical Centre, Medical University of Gdansk | Laboratory of Recombinant Vaccines |
| EPI_ISL_451650 | Hematology Laboratory, Section of Molecular Diagnostics, University Clinical Centre, Medical University of Gdansk | Laboratory of Recombinant Vaccines |
| EPI_ISL_451651 | Hematology Laboratory, Section of Molecular Diagnostics, University Clinical Centre, Medical University of Gdansk | Laboratory of Recombinant Vaccines |

| EPI_Accession_ID | Originating Laboratory | Submitting Laboratory |
|---|---|---|
| EPI_ISL_451652 | Hematology Laboratory, Section of Molecular Diagnostics, University Clinical Centre, Medical University of Gdansk | Laboratory of Recombinant Vaccines |
| EPI_ISL_451653 | Hematology Laboratory, Section of Molecular Diagnostics, University Clinical Centre, Medical University of Gdansk | Laboratory of Recombinant Vaccines |
| EPI_ISL_451654 | Hematology Laboratory, Section of Molecular Diagnostics, University Clinical Centre, Medical University of Gdansk | Laboratory of Recombinant Vaccines |
| EPI_ISL_451655 | State Sanitary Inspectorate | Laboratory of Recombinant Vaccines |
| EPI_ISL_451656 | State Sanitary Inspectorate | Laboratory of Recombinant Vaccines |
| EPI_ISL_451657 | State Sanitary Inspectorate | Laboratory of Recombinant Vaccines |
| EPI_ISL_451658 | State Sanitary Inspectorate | Laboratory of Recombinant Vaccines |
| EPI_ISL_451659 | State Sanitary Inspectorate | Laboratory of Recombinant Vaccines |
| EPI_ISL_451660 | State Sanitary Inspectorate | Laboratory of Recombinant Vaccines |
| EPI_ISL_451661 | State Sanitary Inspectorate | Laboratory of Recombinant Vaccines |
| EPI_ISL_451662 | State Sanitary Inspectorate | Laboratory of Recombinant Vaccines |
| EPI_ISL_451663 | State Sanitary Inspectorate | Laboratory of Recombinant Vaccines |
| EPI_ISL_451664 | State Sanitary Inspectorate | Laboratory of Recombinant Vaccines |
| EPI_ISL_451665 | NYU Langone Health | Departments of Pathology and Medicine, New York University School of Medicine |
| EPI_ISL_451666 | M.P Shah Government Medocal college Jamnagar | Gujarat Biotechnology Research Centre |
| EPI_ISL_451668 | Viollier AG | Department of Biosystems Science and Engineering, ETH ZÃ¼rich |
| EPI_ISL_451669 | Viollier AG | Department of Biosystems Science and Engineering, ETH ZÃ¼rich |
| EPI_ISL_451670 | Viollier AG | Department of Biosystems Science and Engineering, ETH ZÃ¼rich |
| EPI_ISL_451671 | Viollier AG | Department of Biosystems Science and Engineering, ETH ZÃ¼rich |
| EPI_ISL_451672 | Viollier AG | Department of Biosystems Science and Engineering, ETH ZÃ¼rich |

| EPI_Accession_ID | Originating Laboratory | Submitting Laboratory |
|---|---|---|
| EPI_ISL_451673 | Viollier AG | Department of Biosystems Science and Engineering, ETH ZÃ¼rich |
| EPI_ISL_451674 | Viollier AG | Department of Biosystems Science and Engineering, ETH ZÃ¼rich |
| EPI_ISL_451675 | Viollier AG | Department of Biosystems Science and Engineering, ETH ZÃ¼rich |
| EPI_ISL_451676 | Viollier AG | Department of Biosystems Science and Engineering, ETH ZÃ¼rich |
| EPI_ISL_451677 | Viollier AG | Department of Biosystems Science and Engineering, ETH ZÃ¼rich |
| EPI_ISL_451678 | Viollier AG | Department of Biosystems Science and Engineering, ETH ZÃ¼rich |
| EPI_ISL_451679 | Viollier AG | Department of Biosystems Science and Engineering, ETH ZÃ¼rich |
| EPI_ISL_451680 | Viollier AG | Department of Biosystems Science and Engineering, ETH ZÃ¼rich |
| EPI_ISL_451681 | Viollier AG | Department of Biosystems Science and Engineering, ETH ZÃ¼rich |
| EPI_ISL_451682 | Viollier AG | Department of Biosystems Science and Engineering, ETH ZÃ¼rich |
| EPI_ISL_451683 | Viollier AG | Department of Biosystems Science and Engineering, ETH ZÃ¼rich |
| EPI_ISL_451684 | Viollier AG | Department of Biosystems Science and Engineering, ETH ZÃ¼rich |
| EPI_ISL_451685 | Viollier AG | Department of Biosystems Science and Engineering, ETH ZÃ¼rich |
| EPI_ISL_451686 | Viollier AG | Department of Biosystems Science and Engineering, ETH ZÃ¼rich |
| EPI_ISL_451687 | Viollier AG | Department of Biosystems Science and Engineering, ETH ZÃ¼rich |
| EPI_ISL_451688 | Viollier AG | Department of Biosystems Science and Engineering, ETH ZÃ¼rich |

| EPI_Accession_ID | Originating Laboratory | Submitting Laboratory |
|---|---|---|
| EPI_ISL_451689 | Viollier AG | Department of Biosystems Science and Engineering, ETH ZÃ¼rich |
| EPI_ISL_451690 | Viollier AG | Department of Biosystems Science and Engineering, ETH ZÃ¼rich |
| EPI_ISL_451691 | Viollier AG | Department of Biosystems Science and Engineering, ETH ZÃ¼rich |
| EPI_ISL_451692 | Viollier AG | Department of Biosystems Science and Engineering, ETH ZÃ¼rich |
| EPI_ISL_451693 | Viollier AG | Department of Biosystems Science and Engineering, ETH ZÃ¼rich |
| EPI_ISL_451694 | Viollier AG | Department of Biosystems Science and Engineering, ETH ZÃ¼rich |
| EPI_ISL_451695 | Viollier AG | Department of Biosystems Science and Engineering, ETH ZÃ¼rich |
| EPI_ISL_451696 | Viollier AG | Department of Biosystems Science and Engineering, ETH ZÃ¼rich |
| EPI_ISL_451697 | Viollier AG | Department of Biosystems Science and Engineering, ETH ZÃ¼rich |
| EPI_ISL_451698 | Viollier AG | Department of Biosystems Science and Engineering, ETH ZÃ¼rich |
| EPI_ISL_451699 | Viollier AG | Department of Biosystems Science and Engineering, ETH ZÃ¼rich |
| EPI_ISL_451700 | Viollier AG | Department of Biosystems Science and Engineering, ETH ZÃ¼rich |
| EPI_ISL_451701 | Viollier AG | Department of Biosystems Science and Engineering, ETH ZÃ¼rich |
| EPI_ISL_451702 | Viollier AG | Department of Biosystems Science and Engineering, ETH ZÃ¼rich |
| EPI_ISL_451703 | Viollier AG | Department of Biosystems Science and Engineering, ETH ZÃ¼rich |
| EPI_ISL_451704 | Viollier AG | Department of Biosystems Science and Engineering, ETH ZÃ¼rich |

| EPI_Accession_ID | Originating Laboratory | Submitting Laboratory |
|---|---|---|
| EPI_ISL_451705 | Viollier AG | Department of Biosystems Science and Engineering, ETH ZÃ¼rich |
| EPI_ISL_451706 | Viollier AG | Department of Biosystems Science and Engineering, ETH ZÃ¼rich |
| EPI_ISL_451707 | Viollier AG | Department of Biosystems Science and Engineering, ETH ZÃ¼rich |
| EPI_ISL_451708 | Viollier AG | Department of Biosystems Science and Engineering, ETH ZÃ¼rich |
| EPI_ISL_451709 | Viollier AG | Department of Biosystems Science and Engineering, ETH ZÃ¼rich |
| EPI_ISL_451710 | Viollier AG | Department of Biosystems Science and Engineering, ETH ZÃ¼rich |
| EPI_ISL_451711 | Viollier AG | Department of Biosystems Science and Engineering, ETH ZÃ¼rich |
| EPI_ISL_451712 | Viollier AG | Department of Biosystems Science and Engineering, ETH ZÃ¼rich |
| EPI_ISL_451713 | Viollier AG | Department of Biosystems Science and Engineering, ETH ZÃ¼rich |
| EPI_ISL_451714 | Viollier AG | Department of Biosystems Science and Engineering, ETH ZÃ¼rich |
| EPI_ISL_451716 | Viollier AG | Department of Biosystems Science and Engineering, ETH ZÃ¼rich |
| EPI_ISL_451717 | Viollier AG | Department of Biosystems Science and Engineering, ETH ZÃ¼rich |
| EPI_ISL_451718 | Viollier AG | Department of Biosystems Science and Engineering, ETH ZÃ¼rich |
| EPI_ISL_451719 | Viollier AG | Department of Biosystems Science and Engineering, ETH ZÃ¼rich |
| EPI_ISL_451720 | Viollier AG | Department of Biosystems Science and Engineering, ETH ZÃ¼rich |
| EPI_ISL_451721 | Viollier AG | Department of Biosystems Science and Engineering, ETH ZÃ¼rich |

| EPI_Accession_ID | Originating Laboratory | Submitting Laboratory |
|---|---|---|
| EPI_ISL_451722 | Viollier AG | Department of Biosystems Science and Engineering, ETH ZÃ¼rich |
| EPI_ISL_451723 | Viollier AG | Department of Biosystems Science and Engineering, ETH ZÃ¼rich |
| EPI_ISL_451724 | Viollier AG | Department of Biosystems Science and Engineering, ETH ZÃ¼rich |
| EPI_ISL_451725 | Viollier AG | Department of Biosystems Science and Engineering, ETH ZÃ¼rich |
| EPI_ISL_451726 | Viollier AG | Department of Biosystems Science and Engineering, ETH ZÃ¼rich |
| EPI_ISL_451727 | Viollier AG | Department of Biosystems Science and Engineering, ETH ZÃ¼rich |
| EPI_ISL_451728 | Viollier AG | Department of Biosystems Science and Engineering, ETH ZÃ¼rich |
| EPI_ISL_451729 | Viollier AG | Department of Biosystems Science and Engineering, ETH ZÃ¼rich |
| EPI_ISL_451730 | Viollier AG | Department of Biosystems Science and Engineering, ETH ZÃ¼rich |
| EPI_ISL_451731 | Viollier AG | Department of Biosystems Science and Engineering, ETH ZÃ¼rich |
| EPI_ISL_451732 | Viollier AG | Department of Biosystems Science and Engineering, ETH ZÃ¼rich |
| EPI_ISL_451734 | Viollier AG | Department of Biosystems Science and Engineering, ETH ZÃ¼rich |
| EPI_ISL_451736 | Viollier AG | Department of Biosystems Science and Engineering, ETH ZÃ¼rich |
| EPI_ISL_451737 | Viollier AG | Department of Biosystems Science and Engineering, ETH ZÃ¼rich |
| EPI_ISL_451738 | Viollier AG | Department of Biosystems Science and Engineering, ETH ZÃ¼rich |
| EPI_ISL_451739 | Viollier AG | Department of Biosystems Science and Engineering, ETH ZÃ¼rich |

| EPI_Accession_ID | Originating Laboratory | Submitting Laboratory |
|---|---|---|
| EPI_ISL_451740 | Viollier AG | Department of Biosystems Science and Engineering, ETH ZÃ¼rich |
| EPI_ISL_451741 | Viollier AG | Department of Biosystems Science and Engineering, ETH ZÃ¼rich |
| EPI_ISL_451742 | Viollier AG | Department of Biosystems Science and Engineering, ETH ZÃ¼rich |
| EPI_ISL_451744 | Viollier AG | Department of Biosystems Science and Engineering, ETH ZÃ¼rich |
| EPI_ISL_451745 | Viollier AG | Department of Biosystems Science and Engineering, ETH ZÃ¼rich |
| EPI_ISL_451746 | Viollier AG | Department of Biosystems Science and Engineering, ETH ZÃ¼rich |
| EPI_ISL_451747 | Viollier AG | Department of Biosystems Science and Engineering, ETH ZÃ¼rich |
| EPI_ISL_451748 | Viollier AG | Department of Biosystems Science and Engineering, ETH ZÃ¼rich |
| EPI_ISL_451749 | Viollier AG | Department of Biosystems Science and Engineering, ETH ZÃ¼rich |
| EPI_ISL_451750 | Viollier AG | Department of Biosystems Science and Engineering, ETH ZÃ¼rich |
| EPI_ISL_451751 | Viollier AG | Department of Biosystems Science and Engineering, ETH ZÃ¼rich |
| EPI_ISL_451752 | Viollier AG | Department of Biosystems Science and Engineering, ETH ZÃ¼rich |
| EPI_ISL_451753 | Viollier AG | Department of Biosystems Science and Engineering, ETH ZÃ¼rich |
| EPI_ISL_451754 | Viollier AG | Department of Biosystems Science and Engineering, ETH ZÃ¼rich |
| EPI_ISL_451755 | Viollier AG | Department of Biosystems Science and Engineering, ETH ZÃ¼rich |
| EPI_ISL_451756 | Viollier AG | Department of Biosystems Science and Engineering, ETH ZÃ¼rich |

| EPI_Accession_ID | Originating Laboratory | Submitting Laboratory |
|---|---|---|
| EPI_ISL_451757 | Viollier AG | Department of Biosystems Science and Engineering, ETH ZÃ¼rich |
| EPI_ISL_451758 | Viollier AG | Department of Biosystems Science and Engineering, ETH ZÃ¼rich |
| EPI_ISL_451759 | Viollier AG | Department of Biosystems Science and Engineering, ETH ZÃ¼rich |
| EPI_ISL_451760 | Viollier AG | Department of Biosystems Science and Engineering, ETH ZÃ¼rich |
| EPI_ISL_451761 | Viollier AG | Department of Biosystems Science and Engineering, ETH ZÃ¼rich |
| EPI_ISL_451762 | Viollier AG | Department of Biosystems Science and Engineering, ETH ZÃ¼rich |
| EPI_ISL_451763 | Viollier AG | Department of Biosystems Science and Engineering, ETH ZÃ¼rich |
| EPI_ISL_451764 | Viollier AG | Department of Biosystems Science and Engineering, ETH ZÃ¼rich |
| EPI_ISL_451765 | Viollier AG | Department of Biosystems Science and Engineering, ETH ZÃ¼rich |
| EPI_ISL_451766 | Viollier AG | Department of Biosystems Science and Engineering, ETH ZÃ¼rich |
| EPI_ISL_451767 | Viollier AG | Department of Biosystems Science and Engineering, ETH ZÃ¼rich |
| EPI_ISL_451769 | Viollier AG | Department of Biosystems Science and Engineering, ETH ZÃ¼rich |
| EPI_ISL_451770 | Viollier AG | Department of Biosystems Science and Engineering, ETH ZÃ¼rich |
| EPI_ISL_451771 | Viollier AG | Department of Biosystems Science and Engineering, ETH ZÃ¼rich |
| EPI_ISL_451772 | Viollier AG | Department of Biosystems Science and Engineering, ETH ZÃ¼rich |
| EPI_ISL_451773 | Viollier AG | Department of Biosystems Science and Engineering, ETH ZÃ¼rich |

| EPI_Accession_ID | Originating Laboratory | Submitting Laboratory |
|---|---|---|
| EPI_ISL_451774 | Viollier AG | Department of Biosystems Science and Engineering, ETH ZÃ¼rich |
| EPI_ISL_451775 | Viollier AG | Department of Biosystems Science and Engineering, ETH ZÃ¼rich |
| EPI_ISL_451776 | Viollier AG | Department of Biosystems Science and Engineering, ETH ZÃ¼rich |
| EPI_ISL_451777 | Viollier AG | Department of Biosystems Science and Engineering, ETH ZÃ¼rich |
| EPI_ISL_451778 | Viollier AG | Department of Biosystems Science and Engineering, ETH ZÃ¼rich |
| EPI_ISL_451779 | Viollier AG | Department of Biosystems Science and Engineering, ETH ZÃ¼rich |
| EPI_ISL_451780 | Viollier AG | Department of Biosystems Science and Engineering, ETH ZÃ¼rich |
| EPI_ISL_451781 | Viollier AG | Department of Biosystems Science and Engineering, ETH ZÃ¼rich |
| EPI_ISL_451782 | Viollier AG | Department of Biosystems Science and Engineering, ETH ZÃ¼rich |
| EPI_ISL_451783 | Viollier AG | Department of Biosystems Science and Engineering, ETH ZÃ¼rich |
| EPI_ISL_451784 | Viollier AG | Department of Biosystems Science and Engineering, ETH ZÃ¼rich |
| EPI_ISL_451785 | Viollier AG | Department of Biosystems Science and Engineering, ETH ZÃ¼rich |
| EPI_ISL_451786 | Viollier AG | Department of Biosystems Science and Engineering, ETH ZÃ¼rich |
| EPI_ISL_451787 | Viollier AG | Department of Biosystems Science and Engineering, ETH ZÃ¼rich |
| EPI_ISL_451788 | Viollier AG | Department of Biosystems Science and Engineering, ETH ZÃ¼rich |
| EPI_ISL_451789 | Viollier AG | Department of Biosystems Science and Engineering, ETH ZÃ¼rich |

| EPI_Accession_ID | Originating Laboratory | Submitting Laboratory |
|---|---|---|
| EPI_ISL_451790 | Viollier AG | Department of Biosystems Science and Engineering, ETH ZÃ¼rich |
| EPI_ISL_451791 | Viollier AG | Department of Biosystems Science and Engineering, ETH ZÃ¼rich |
| EPI_ISL_451792 | Viollier AG | Department of Biosystems Science and Engineering, ETH ZÃ¼rich |
| EPI_ISL_451793 | Viollier AG | Department of Biosystems Science and Engineering, ETH ZÃ¼rich |
| EPI_ISL_451794 | Viollier AG | Department of Biosystems Science and Engineering, ETH ZÃ¼rich |
| EPI_ISL_451795 | Viollier AG | Department of Biosystems Science and Engineering, ETH ZÃ¼rich |
| EPI_ISL_451796 | Viollier AG | Department of Biosystems Science and Engineering, ETH ZÃ¼rich |
| EPI_ISL_451797 | Viollier AG | Department of Biosystems Science and Engineering, ETH ZÃ¼rich |
| EPI_ISL_451798 | Viollier AG | Department of Biosystems Science and Engineering, ETH ZÃ¼rich |
| EPI_ISL_451799 | Viollier AG | Department of Biosystems Science and Engineering, ETH ZÃ¼rich |
| EPI_ISL_451800 | Viollier AG | Department of Biosystems Science and Engineering, ETH ZÃ¼rich |
| EPI_ISL_451801 | Viollier AG | Department of Biosystems Science and Engineering, ETH ZÃ¼rich |
| EPI_ISL_451802 | Viollier AG | Department of Biosystems Science and Engineering, ETH ZÃ¼rich |
| EPI_ISL_451803 | Viollier AG | Department of Biosystems Science and Engineering, ETH ZÃ¼rich |
| EPI_ISL_451804 | Viollier AG | Department of Biosystems Science and Engineering, ETH ZÃ¼rich |
| EPI_ISL_451805 | Viollier AG | Department of Biosystems Science and Engineering, ETH ZÃ¼rich |

| EPI_Accession_ID | Originating Laboratory | Submitting Laboratory |
|---|---|---|
| EPI_ISL_451806 | Viollier AG | Department of Biosystems Science and Engineering, ETH ZÃ¼rich |
| EPI_ISL_451807 | Viollier AG | Department of Biosystems Science and Engineering, ETH ZÃ¼rich |
| EPI_ISL_451808 | Viollier AG | Department of Biosystems Science and Engineering, ETH ZÃ¼rich |
| EPI_ISL_451809 | Viollier AG | Department of Biosystems Science and Engineering, ETH ZÃ¼rich |
| EPI_ISL_451810 | Viollier AG | Department of Biosystems Science and Engineering, ETH ZÃ¼rich |
| EPI_ISL_451811 | Viollier AG | Department of Biosystems Science and Engineering, ETH ZÃ¼rich |
| EPI_ISL_451812 | Viollier AG | Department of Biosystems Science and Engineering, ETH ZÃ¼rich |
| EPI_ISL_451813 | Viollier AG | Department of Biosystems Science and Engineering, ETH ZÃ¼rich |
| EPI_ISL_451814 | Viollier AG | Department of Biosystems Science and Engineering, ETH ZÃ¼rich |
| EPI_ISL_451815 | Viollier AG | Department of Biosystems Science and Engineering, ETH ZÃ¼rich |
| EPI_ISL_451816 | Viollier AG | Department of Biosystems Science and Engineering, ETH ZÃ¼rich |
| EPI_ISL_451817 | Viollier AG | Department of Biosystems Science and Engineering, ETH ZÃ¼rich |
| EPI_ISL_451818 | Viollier AG | Department of Biosystems Science and Engineering, ETH ZÃ¼rich |
| EPI_ISL_451819 | Viollier AG | Department of Biosystems Science and Engineering, ETH ZÃ¼rich |
| EPI_ISL_451820 | Viollier AG | Department of Biosystems Science and Engineering, ETH ZÃ¼rich |
| EPI_ISL_451821 | Viollier AG | Department of Biosystems Science and Engineering, ETH ZÃ¼rich |

| EPI_Accession_ID | Originating Laboratory | Submitting Laboratory |
|---|---|---|
| EPI_ISL_451822 | Viollier AG | Department of Biosystems Science and Engineering, ETH ZÃ¼rich |
| EPI_ISL_451823 | Viollier AG | Department of Biosystems Science and Engineering, ETH ZÃ¼rich |
| EPI_ISL_451824 | Viollier AG | Department of Biosystems Science and Engineering, ETH ZÃ¼rich |
| EPI_ISL_451826 | Viollier AG | Department of Biosystems Science and Engineering, ETH ZÃ¼rich |
| EPI_ISL_451827 | Viollier AG | Department of Biosystems Science and Engineering, ETH ZÃ¼rich |
| EPI_ISL_451829 | Viollier AG | Department of Biosystems Science and Engineering, ETH ZÃ¼rich |
| EPI_ISL_451830 | Viollier AG | Department of Biosystems Science and Engineering, ETH ZÃ¼rich |
| EPI_ISL_451831 | Viollier AG | Department of Biosystems Science and Engineering, ETH ZÃ¼rich |
| EPI_ISL_451832 | Viollier AG | Department of Biosystems Science and Engineering, ETH ZÃ¼rich |
| EPI_ISL_451833 | Viollier AG | Department of Biosystems Science and Engineering, ETH ZÃ¼rich |
| EPI_ISL_451834 | Viollier AG | Department of Biosystems Science and Engineering, ETH ZÃ¼rich |
| EPI_ISL_451835 | Viollier AG | Department of Biosystems Science and Engineering, ETH ZÃ¼rich |
| EPI_ISL_451836 | Viollier AG | Department of Biosystems Science and Engineering, ETH ZÃ¼rich |
| EPI_ISL_451837 | Viollier AG | Department of Biosystems Science and Engineering, ETH ZÃ¼rich |
| EPI_ISL_451838 | Viollier AG | Department of Biosystems Science and Engineering, ETH ZÃ¼rich |
| EPI_ISL_451839 | Viollier AG | Department of Biosystems Science and Engineering, ETH ZÃ¼rich |

| EPI_Accession_ID | Originating Laboratory | Submitting Laboratory |
|---|---|---|
| EPI_ISL_451840 | Viollier AG | Department of Biosystems Science and Engineering, ETH ZÃ¼rich |
| EPI_ISL_451841 | Viollier AG | Department of Biosystems Science and Engineering, ETH ZÃ¼rich |
| EPI_ISL_451842 | Viollier AG | Department of Biosystems Science and Engineering, ETH ZÃ¼rich |
| EPI_ISL_451843 | Viollier AG | Department of Biosystems Science and Engineering, ETH ZÃ¼rich |
| EPI_ISL_451844 | Viollier AG | Department of Biosystems Science and Engineering, ETH ZÃ¼rich |
| EPI_ISL_451845 | Viollier AG | Department of Biosystems Science and Engineering, ETH ZÃ¼rich |
| EPI_ISL_451846 | Viollier AG | Department of Biosystems Science and Engineering, ETH ZÃ¼rich |
| EPI_ISL_451847 | Viollier AG | Department of Biosystems Science and Engineering, ETH ZÃ¼rich |
| EPI_ISL_451848 | Viollier AG | Department of Biosystems Science and Engineering, ETH ZÃ¼rich |
| EPI_ISL_451849 | Viollier AG | Department of Biosystems Science and Engineering, ETH ZÃ¼rich |
| EPI_ISL_451850 | Viollier AG | Department of Biosystems Science and Engineering, ETH ZÃ¼rich |
| EPI_ISL_451851 | Viollier AG | Department of Biosystems Science and Engineering, ETH ZÃ¼rich |
| EPI_ISL_451852 | Viollier AG | Department of Biosystems Science and Engineering, ETH ZÃ¼rich |
| EPI_ISL_451853 | Viollier AG | Department of Biosystems Science and Engineering, ETH ZÃ¼rich |
| EPI_ISL_451854 | Viollier AG | Department of Biosystems Science and Engineering, ETH ZÃ¼rich |
| EPI_ISL_451855 | Viollier AG | Department of Biosystems Science and Engineering, ETH ZÃ¼rich |

| EPI_Accession_ID | Originating Laboratory | Submitting Laboratory |
|---|---|---|
| EPI_ISL_451856 | Viollier AG | Department of Biosystems Science and Engineering, ETH ZÃ¼rich |
| EPI_ISL_451857 | Viollier AG | Department of Biosystems Science and Engineering, ETH ZÃ¼rich |
| EPI_ISL_451858 | Viollier AG | Department of Biosystems Science and Engineering, ETH ZÃ¼rich |
| EPI_ISL_451859 | Viollier AG | Department of Biosystems Science and Engineering, ETH ZÃ¼rich |
| EPI_ISL_451860 | Viollier AG | Department of Biosystems Science and Engineering, ETH ZÃ¼rich |
| EPI_ISL_451861 | Viollier AG | Department of Biosystems Science and Engineering, ETH ZÃ¼rich |
| EPI_ISL_451862 | Viollier AG | Department of Biosystems Science and Engineering, ETH ZÃ¼rich |
| EPI_ISL_451863 | Viollier AG | Department of Biosystems Science and Engineering, ETH ZÃ¼rich |
| EPI_ISL_451864 | Viollier AG | Department of Biosystems Science and Engineering, ETH ZÃ¼rich |
| EPI_ISL_451865 | Viollier AG | Department of Biosystems Science and Engineering, ETH ZÃ¼rich |
| EPI_ISL_451866 | Viollier AG | Department of Biosystems Science and Engineering, ETH ZÃ¼rich |
| EPI_ISL_451867 | Viollier AG | Department of Biosystems Science and Engineering, ETH ZÃ¼rich |
| EPI_ISL_451868 | Viollier AG | Department of Biosystems Science and Engineering, ETH ZÃ¼rich |
| EPI_ISL_451869 | Viollier AG | Department of Biosystems Science and Engineering, ETH ZÃ¼rich |
| EPI_ISL_451870 | Viollier AG | Department of Biosystems Science and Engineering, ETH ZÃ¼rich |
| EPI_ISL_451871 | Viollier AG | Department of Biosystems Science and Engineering, ETH ZÃ¼rich |

| EPI_Accession_ID | Originating Laboratory | Submitting Laboratory |
|---|---|---|
| EPI_ISL_451872 | Viollier AG | Department of Biosystems Science and Engineering, ETH ZÃ¼rich |
| EPI_ISL_451873 | Viollier AG | Department of Biosystems Science and Engineering, ETH ZÃ¼rich |
| EPI_ISL_451874 | Viollier AG | Department of Biosystems Science and Engineering, ETH ZÃ¼rich |
| EPI_ISL_451875 | Viollier AG | Department of Biosystems Science and Engineering, ETH ZÃ¼rich |
| EPI_ISL_451876 | Viollier AG | Department of Biosystems Science and Engineering, ETH ZÃ¼rich |
| EPI_ISL_451877 | Viollier AG | Department of Biosystems Science and Engineering, ETH ZÃ¼rich |
| EPI_ISL_451878 | Viollier AG | Department of Biosystems Science and Engineering, ETH ZÃ¼rich |
| EPI_ISL_451879 | Viollier AG | Department of Biosystems Science and Engineering, ETH ZÃ¼rich |
| EPI_ISL_451880 | Viollier AG | Department of Biosystems Science and Engineering, ETH ZÃ¼rich |
| EPI_ISL_451882 | Viollier AG | Department of Biosystems Science and Engineering, ETH ZÃ¼rich |
| EPI_ISL_451883 | Viollier AG | Department of Biosystems Science and Engineering, ETH ZÃ¼rich |
| EPI_ISL_451884 | Viollier AG | Department of Biosystems Science and Engineering, ETH ZÃ¼rich |
| EPI_ISL_451885 | Viollier AG | Department of Biosystems Science and Engineering, ETH ZÃ¼rich |
| EPI_ISL_451886 | Viollier AG | Department of Biosystems Science and Engineering, ETH ZÃ¼rich |
| EPI_ISL_451887 | Viollier AG | Department of Biosystems Science and Engineering, ETH ZÃ¼rich |
| EPI_ISL_451888 | Viollier AG | Department of Biosystems Science and Engineering, ETH ZÃ¼rich |

| EPI_Accession_ID | Originating Laboratory | Submitting Laboratory |
|---|---|---|
| EPI_ISL_451889 | Viollier AG | Department of Biosystems Science and Engineering, ETH ZÃ¼rich |
| EPI_ISL_451890 | Viollier AG | Department of Biosystems Science and Engineering, ETH ZÃ¼rich |
| EPI_ISL_451891 | Viollier AG | Department of Biosystems Science and Engineering, ETH ZÃ¼rich |
| EPI_ISL_451892 | Viollier AG | Department of Biosystems Science and Engineering, ETH ZÃ¼rich |
| EPI_ISL_451893 | Viollier AG | Department of Biosystems Science and Engineering, ETH ZÃ¼rich |
| EPI_ISL_451894 | Viollier AG | Department of Biosystems Science and Engineering, ETH ZÃ¼rich |
| EPI_ISL_451896 | Viollier AG | Department of Biosystems Science and Engineering, ETH ZÃ¼rich |
| EPI_ISL_451898 | Viollier AG | Department of Biosystems Science and Engineering, ETH ZÃ¼rich |
| EPI_ISL_451899 | Viollier AG | Department of Biosystems Science and Engineering, ETH ZÃ¼rich |
| EPI_ISL_451900 | Viollier AG | Department of Biosystems Science and Engineering, ETH ZÃ¼rich |
| EPI_ISL_451902 | Viollier AG | Department of Biosystems Science and Engineering, ETH ZÃ¼rich |
| EPI_ISL_451903 | Viollier AG | Department of Biosystems Science and Engineering, ETH ZÃ¼rich |
| EPI_ISL_451905 | Viollier AG | Department of Biosystems Science and Engineering, ETH ZÃ¼rich |
| EPI_ISL_451906 | Viollier AG | Department of Biosystems Science and Engineering, ETH ZÃ¼rich |
| EPI_ISL_451907 | Viollier AG | Department of Biosystems Science and Engineering, ETH ZÃ¼rich |
| EPI_ISL_451908 | Viollier AG | Department of Biosystems Science and Engineering, ETH ZÃ¼rich |

| EPI_Accession_ID | Originating Laboratory | Submitting Laboratory |
|---|---|---|
| EPI_ISL_451909 | Viollier AG | Department of Biosystems Science and Engineering, ETH ZÃ¼rich |
| EPI_ISL_451910 | Viollier AG | Department of Biosystems Science and Engineering, ETH ZÃ¼rich |
| EPI_ISL_451911 | Viollier AG | Department of Biosystems Science and Engineering, ETH ZÃ¼rich |
| EPI_ISL_451912 | Viollier AG | Department of Biosystems Science and Engineering, ETH ZÃ¼rich |
| EPI_ISL_451913 | Viollier AG | Department of Biosystems Science and Engineering, ETH ZÃ¼rich |
| EPI_ISL_451914 | Viollier AG | Department of Biosystems Science and Engineering, ETH ZÃ¼rich |
| EPI_ISL_451915 | Viollier AG | Department of Biosystems Science and Engineering, ETH ZÃ¼rich |
| EPI_ISL_451916 | Viollier AG | Department of Biosystems Science and Engineering, ETH ZÃ¼rich |
| EPI_ISL_451934 | Research Unit, University Hospital for Infectious Diseases "Dr. Fran MihaljeviÄ‡" | Cicin Sain lab, Helmholtz Centre for Infection Research |
| EPI_ISL_451935 | CUB Hopital Erasme Laboratoire d'Anatomie Pathologique | CUB Hopital Erasme Laboratoire d'Anatomie Pathologique |
| EPI_ISL_451936 | Max von Pettenkofer Institute, Virology, National Reference Center for Retroviruses, LMU MÃ¼nchen | Laboratory for Functional Genome Analysis, Dept. Genomics, Gene Center of the LMU Munich |
| EPI_ISL_451937 | Max von Pettenkofer Institute, Virology, National Reference Center for Retroviruses, LMU MÃ¼nchen | Laboratory for Functional Genome Analysis, Dept. Genomics, Gene Center of the LMU Munich |
| EPI_ISL_451938 | Max von Pettenkofer Institute, Virology, National Reference Center for Retroviruses, LMU MÃ¼nchen | Laboratory for Functional Genome Analysis, Dept. Genomics, Gene Center of the LMU Munich |
| EPI_ISL_451939 | Max von Pettenkofer Institute, Virology, National Reference Center for Retroviruses, LMU MÃ¼nchen | Laboratory for Functional Genome Analysis, Dept. Genomics, Gene Center of the LMU Munich |
| EPI_ISL_451940 | Max von Pettenkofer Institute, Virology, National Reference Center for Retroviruses, LMU MÃ¼nchen | Laboratory for Functional Genome Analysis, Dept. Genomics, Gene Center of the LMU Munich |
| EPI_ISL_451941 | Max von Pettenkofer Institute, Virology, National Reference Center for Retroviruses, LMU MÃ¼nchen | Laboratory for Functional Genome Analysis, Dept. Genomics, Gene Center of the LMU Munich |

| EPI_Accession_ID | Originating Laboratory | Submitting Laboratory |
|---|---|---|
| EPI_ISL_451942 | Max von Pettenkofer Institute, Virology, National Reference Center for Retroviruses, LMU MÃ¼nchen | Laboratory for Functional Genome Analysis, Dept. Genomics, Gene Center of the LMU Munich |
| EPI_ISL_451943 | Max von Pettenkofer Institute, Virology, National Reference Center for Retroviruses, LMU MÃ¼nchen | Laboratory for Functional Genome Analysis, Dept. Genomics, Gene Center of the LMU Munich |
| EPI_ISL_451944 | Max von Pettenkofer Institute, Virology, National Reference Center for Retroviruses, LMU MÃ¼nchen | Laboratory for Functional Genome Analysis, Dept. Genomics, Gene Center of the LMU Munich |
| EPI_ISL_451945 | Max von Pettenkofer Institute, Virology, National Reference Center for Retroviruses, LMU MÃ¼nchen | Laboratory for Functional Genome Analysis, Dept. Genomics, Gene Center of the LMU Munich |
| EPI_ISL_451946 | Max von Pettenkofer Institute, Virology, National Reference Center for Retroviruses, LMU MÃ¼nchen | Laboratory for Functional Genome Analysis, Dept. Genomics, Gene Center of the LMU Munich |
| EPI_ISL_451957 | Molecular Pathology Division, Department of Pathology, Hong Kong Sanatorium & Hospital | Molecular Pathology Division, Department of Pathology, Hong Kong Sanatorium & Hospital |
| EPI_ISL_451958 | unknown | Jamil-ur-Rahman Center for Genome Research |
| EPI_ISL_451960 | unknown | Respiratory Viruses Branch |
| EPI_ISL_451961 | Istituto Zooprofilattico Sperimentale Puglia e Basilicata; Dipartimento di Bioscienze, Biotecnologie e Biofarmaceutica dellâ€™UniversitÃ degli Studi di Bari â€œA.Moroâ€�Istituto di Biomembrane. Bioenergetica e Biotecnologie Molecolari del Consiglio Nazionale delle Ricerche di Bari | Beaconlab (Bioinformatics Evolution and Comparative Genomics lab), Dept of Biosciences, University of Milan |
| EPI_ISL_451962 | Istituto Zooprofilattico Sperimentale Puglia e Basilicata; Dipartimento di Bioscienze, Biotecnologie e Biofarmaceutica dellâ€™UniversitÃ degli Studi di Bari â€œA.Moroâ€�Istituto di Biomembrane. Bioenergetica e Biotecnologie Molecolari del Consiglio Nazionale delle Ricerche di Bari | Beaconlab (Bioinformatics, Evolution and Comparative Genomics lab), Dept of Biosciences, University on Milan |
| EPI_ISL_451963 | Federal Budget Institution of Science, State Research Center for Applied Microbiology & Biotechnology | Federal Budget Institution of Science, State Research Center for Applied Microbiology & Biotechnology |
| EPI_ISL_451965 | Federal Budget Institution of Science, State Research Center for Applied Microbiology & Biotechnology | Federal Budget Institution of Science, State Research Center for Applied Microbiology & Biotechnology |
| EPI_ISL_451968 | Federal Budget Institution of Science, State Research Center for Applied Microbiology & Biotechnology | Federal Budget Institution of Science, State Research Center for Applied Microbiology & Biotechnology |

| EPI_Accession_ID | Originating Laboratory | Submitting Laboratory |
|---|---|---|
| EPI_ISL_451969 | Federal Budget Institution of Science, State Research Center for Applied Microbiology & Biotechnology | Federal Budget Institution of Science, State Research Center for Applied Microbiology & Biotechnology |
| EPI_ISL_451970 | Federal Budget Institution of Science, State Research Center for Applied Microbiology & Biotechnology | Federal Budget Institution of Science, State Research Center for Applied Microbiology & Biotechnology |
| EPI_ISL_451971 | 1. ViroGenetics - BSL3 Laboratory of Virology, MaÅ‚opolska Centre of Biotechnology, Jagiellonian University; 2. II Department of Internal Medicine, Faculty of Medicine, Jagiellonian University Medical College; 3. DIAGNOSTYKA Ltd. | 1. ViroGenetics - BSL3 Laboratory of Virology, MaÅ‚opolska Centre of Biotechnology, Jagiellonian University; 2. II Department of Internal Medicine, Faculty of Medicine, Jagiellonian University Medical College. |
| EPI_ISL_451972 | 1. ViroGenetics - BSL3 Laboratory of Virology, MaÅ‚opolska Centre of Biotechnology, Jagiellonian University; 2. II Department of Internal Medicine, Faculty of Medicine, Jagiellonian University Medical College; 3. DIAGNOSTYKA Ltd. | 1. ViroGenetics - BSL3 Laboratory of Virology, MaÅ‚opolska Centre of Biotechnology, Jagiellonian University; 2. II Department of Internal Medicine, Faculty of Medicine, Jagiellonian University Medical College. |
| EPI_ISL_451973 | 1. ViroGenetics - BSL3 Laboratory of Virology, MaÅ‚opolska Centre of Biotechnology, Jagiellonian University; 2. II Department of Internal Medicine, Faculty of Medicine, Jagiellonian University Medical College; 3. DIAGNOSTYKA Ltd. | 1. ViroGenetics - BSL3 Laboratory of Virology, MaÅ‚opolska Centre of Biotechnology, Jagiellonian University; 2. II Department of Internal Medicine, Faculty of Medicine, Jagiellonian University Medical College. |
| EPI_ISL_451974 | 1. ViroGenetics - BSL3 Laboratory of Virology, MaÅ‚opolska Centre of Biotechnology, Jagiellonian University; 2. II Department of Internal Medicine, Faculty of Medicine, Jagiellonian University Medical College; 3. DIAGNOSTYKA Ltd. | 1. ViroGenetics - BSL3 Laboratory of Virology, MaÅ‚opolska Centre of Biotechnology, Jagiellonian University; 2. II Department of Internal Medicine, Faculty of Medicine, Jagiellonian University Medical College. |
| EPI_ISL_451975 | 1. ViroGenetics - BSL3 Laboratory of Virology, MaÅ‚opolska Centre of Biotechnology, Jagiellonian University; 2. II Department of Internal Medicine, Faculty of Medicine, Jagiellonian University Medical College; 3. DIAGNOSTYKA Ltd. | 1. ViroGenetics - BSL3 Laboratory of Virology, MaÅ‚opolska Centre of Biotechnology, Jagiellonian University; 2. II Department of Internal Medicine, Faculty of Medicine, Jagiellonian University Medical College. |

| EPI_Accession_ID | Originating Laboratory | Submitting Laboratory |
|---|---|---|
| EPI_ISL_451976 | 1. ViroGenetics - BSL3 Laboratory of Virology, MaÅ‚opolska Centre of Biotechnology, Jagiellonian University; 2. II Department of Internal Medicine, Faculty of Medicine, Jagiellonian University Medical College; 3. DIAGNOSTYKA Ltd. | 1. ViroGenetics - BSL3 Laboratory of Virology, MaÅ‚opolska Centre of Biotechnology, Jagiellonian University; 2. II Department of Internal Medicine, Faculty of Medicine, Jagiellonian University Medical College. |
| EPI_ISL_451977 | 1. ViroGenetics - BSL3 Laboratory of Virology, MaÅ‚opolska Centre of Biotechnology, Jagiellonian University; 2. II Department of Internal Medicine, Faculty of Medicine, Jagiellonian University Medical College; 3. DIAGNOSTYKA Ltd. | 1. ViroGenetics - BSL3 Laboratory of Virology, MaÅ‚opolska Centre of Biotechnology, Jagiellonian University; 2. II Department of Internal Medicine, Faculty of Medicine, Jagiellonian University Medical College. |
| EPI_ISL_451978 | 1. ViroGenetics - BSL3 Laboratory of Virology, MaÅ‚opolska Centre of Biotechnology, Jagiellonian University; 2. II Department of Internal Medicine, Faculty of Medicine, Jagiellonian University Medical College; 3. DIAGNOSTYKA Ltd. | 1. ViroGenetics - BSL3 Laboratory of Virology, MaÅ‚opolska Centre of Biotechnology, Jagiellonian University; 2. II Department of Internal Medicine, Faculty of Medicine, Jagiellonian University Medical College. |
| EPI_ISL_451979 | 1. ViroGenetics - BSL3 Laboratory of Virology, MaÅ‚opolska Centre of Biotechnology, Jagiellonian University; 2. II Department of Internal Medicine, Faculty of Medicine, Jagiellonian University Medical College; 3. DIAGNOSTYKA Ltd. | 1. ViroGenetics - BSL3 Laboratory of Virology, MaÅ‚opolska Centre of Biotechnology, Jagiellonian University; 2. II Department of Internal Medicine, Faculty of Medicine, Jagiellonian University Medical College. |
| EPI_ISL_451980 | 1. ViroGenetics - BSL3 Laboratory of Virology, MaÅ‚opolska Centre of Biotechnology, Jagiellonian University; 2. II Department of Internal Medicine, Faculty of Medicine, Jagiellonian University Medical College; 3. DIAGNOSTYKA Ltd. | 1. ViroGenetics - BSL3 Laboratory of Virology, MaÅ‚opolska Centre of Biotechnology, Jagiellonian University; 2. II Department of Internal Medicine, Faculty of Medicine, Jagiellonian University Medical College. |
| EPI_ISL_451981 | 1. ViroGenetics - BSL3 Laboratory of Virology, MaÅ‚opolska Centre of Biotechnology, Jagiellonian University; 2. II Department of Internal Medicine, Faculty of Medicine, Jagiellonian University Medical College; 3. DIAGNOSTYKA Ltd. | 1. ViroGenetics - BSL3 Laboratory of Virology, MaÅ‚opolska Centre of Biotechnology, Jagiellonian University; 2. II Department of Internal Medicine, Faculty of Medicine, Jagiellonian University Medical College. |

| EPI_Accession_ID | Originating Laboratory | Submitting Laboratory |
|---|---|---|
| EPI_ISL_451982 | 1. ViroGenetics - BSL3 Laboratory of Virology, MaÅ‚opolska Centre of Biotechnology, Jagiellonian University; 2. II Department of Internal Medicine, Faculty of Medicine, Jagiellonian University Medical College; 3. DIAGNOSTYKA Ltd. | 1. ViroGenetics - BSL3 Laboratory of Virology, MaÅ‚opolska Centre of Biotechnology, Jagiellonian University; 2. II Department of Internal Medicine, Faculty of Medicine, Jagiellonian University Medical College. |
| EPI_ISL_451983 | 1. ViroGenetics - BSL3 Laboratory of Virology, MaÅ‚opolska Centre of Biotechnology, Jagiellonian University; 2. II Department of Internal Medicine, Faculty of Medicine, Jagiellonian University Medical College; 3. DIAGNOSTYKA Ltd. | 1. ViroGenetics - BSL3 Laboratory of Virology, MaÅ‚opolska Centre of Biotechnology, Jagiellonian University; 2. II Department of Internal Medicine, Faculty of Medicine, Jagiellonian University Medical College. |
| EPI_ISL_451984 | 1. ViroGenetics - BSL3 Laboratory of Virology, MaÅ‚opolska Centre of Biotechnology, Jagiellonian University; 2. II Department of Internal Medicine, Faculty of Medicine, Jagiellonian University Medical College; 3. DIAGNOSTYKA Ltd. | 1. ViroGenetics - BSL3 Laboratory of Virology, MaÅ‚opolska Centre of Biotechnology, Jagiellonian University; 2. II Department of Internal Medicine, Faculty of Medicine, Jagiellonian University Medical College. |
| EPI_ISL_451985 | 1. ViroGenetics - BSL3 Laboratory of Virology, MaÅ‚opolska Centre of Biotechnology, Jagiellonian University; 2. II Department of Internal Medicine, Faculty of Medicine, Jagiellonian University Medical College; 3. DIAGNOSTYKA Ltd. | 1. ViroGenetics - BSL3 Laboratory of Virology, MaÅ‚opolska Centre of Biotechnology, Jagiellonian University; 2. II Department of Internal Medicine, Faculty of Medicine, Jagiellonian University Medical College. |
| EPI_ISL_451986 | 1. ViroGenetics - BSL3 Laboratory of Virology, MaÅ‚opolska Centre of Biotechnology, Jagiellonian University; 2. II Department of Internal Medicine, Faculty of Medicine, Jagiellonian University Medical College; 3. DIAGNOSTYKA Ltd. | 1. ViroGenetics - BSL3 Laboratory of Virology, MaÅ‚opolska Centre of Biotechnology, Jagiellonian University; 2. II Department of Internal Medicine, Faculty of Medicine, Jagiellonian University Medical College. |
| EPI_ISL_451987 | 1. ViroGenetics - BSL3 Laboratory of Virology, MaÅ‚opolska Centre of Biotechnology, Jagiellonian University; 2. II Department of Internal Medicine, Faculty of Medicine, Jagiellonian University Medical College; 3. DIAGNOSTYKA Ltd. | 1. ViroGenetics - BSL3 Laboratory of Virology, MaÅ‚opolska Centre of Biotechnology, Jagiellonian University; 2. II Department of Internal Medicine, Faculty of Medicine, Jagiellonian University Medical College. |
| EPI_ISL_451988 | Department of Clinical Microbiology, Copenhagen University Hospital, Hvidovre, Kettegaard Alle 30, 2650 Hvidovre. | Albertsen lab, Department of Chemistry and Bioscience, Aalborg University, Denmark |

| EPI_Accession_ID | Originating Laboratory | Submitting Laboratory |
|---|---|---|
| EPI_ISL_451989 | Department of Clinical Microbiology, Copenhagen University Hospital, Hvidovre, Kettegaard Alle 30, 2650 Hvidovre. | Albertsen lab, Department of Chemistry and Bioscience, Aalborg University, Denmark |
| EPI_ISL_451990 | Department of Clinical Microbiology, Copenhagen University Hospital, Hvidovre, Kettegaard Alle 30, 2650 Hvidovre. | Albertsen lab, Department of Chemistry and Bioscience, Aalborg University, Denmark |
| EPI_ISL_451991 | Department of Clinical Microbiology, Copenhagen University Hospital, Hvidovre, Kettegaard Alle 30, 2650 Hvidovre. | Albertsen lab, Department of Chemistry and Bioscience, Aalborg University, Denmark |
| EPI_ISL_451992 | Department of Clinical Microbiology, Copenhagen University Hospital, Hvidovre, Kettegaard Alle 30, 2650 Hvidovre. | Albertsen lab, Department of Chemistry and Bioscience, Aalborg University, Denmark |
| EPI_ISL_451993 | Department of Clinical Microbiology, Copenhagen University Hospital, Hvidovre, Kettegaard Alle 30, 2650 Hvidovre. | Albertsen lab, Department of Chemistry and Bioscience, Aalborg University, Denmark |
| EPI_ISL_451994 | Department of Clinical Microbiology, Copenhagen University Hospital, Hvidovre, Kettegaard Alle 30, 2650 Hvidovre. | Albertsen lab, Department of Chemistry and Bioscience, Aalborg University, Denmark |
| EPI_ISL_451995 | Department of Clinical Microbiology, Copenhagen University Hospital, Hvidovre, Kettegaard Alle 30, 2650 Hvidovre. | Albertsen lab, Department of Chemistry and Bioscience, Aalborg University, Denmark |
| EPI_ISL_451996 | Department of Clinical Microbiology, Copenhagen University Hospital, Hvidovre, Kettegaard Alle 30, 2650 Hvidovre. | Albertsen lab, Department of Chemistry and Bioscience, Aalborg University, Denmark |
| EPI_ISL_451997 | Department of Clinical Microbiology, Copenhagen University Hospital, Hvidovre, Kettegaard Alle 30, 2650 Hvidovre. | Albertsen lab, Department of Chemistry and Bioscience, Aalborg University, Denmark |
| EPI_ISL_451998 | Department of Clinical Microbiology, Copenhagen University Hospital, Hvidovre, Kettegaard Alle 30, 2650 Hvidovre. | Albertsen lab, Department of Chemistry and Bioscience, Aalborg University, Denmark |
| EPI_ISL_451999 | Department of Clinical Microbiology, Copenhagen University Hospital, Hvidovre, Kettegaard Alle 30, 2650 Hvidovre. | Albertsen lab, Department of Chemistry and Bioscience, Aalborg University, Denmark |

| EPI_Accession_ID | Originating Laboratory | Submitting Laboratory |
|---|---|---|
| EPI_ISL_452000 | Department of Clinical Microbiology, Copenhagen University Hospital, Hvidovre, Kettegaard Alle 30, 2650 Hvidovre. | Albertsen lab, Department of Chemistry and Bioscience, Aalborg University, Denmark |
| EPI_ISL_452001 | Department of Clinical Microbiology, Copenhagen University Hospital, Hvidovre, Kettegaard Alle 30, 2650 Hvidovre. | Albertsen lab, Department of Chemistry and Bioscience, Aalborg University, Denmark |
| EPI_ISL_452002 | Department of Clinical Microbiology, Copenhagen University Hospital, Hvidovre, Kettegaard Alle 30, 2650 Hvidovre. | Albertsen lab, Department of Chemistry and Bioscience, Aalborg University, Denmark |
| EPI_ISL_452003 | Department of Clinical Microbiology, Copenhagen University Hospital, Hvidovre, Kettegaard Alle 30, 2650 Hvidovre. | Albertsen lab, Department of Chemistry and Bioscience, Aalborg University, Denmark |
| EPI_ISL_452004 | Department of Clinical Microbiology, Copenhagen University Hospital, Hvidovre, Kettegaard Alle 30, 2650 Hvidovre. | Albertsen lab, Department of Chemistry and Bioscience, Aalborg University, Denmark |
| EPI_ISL_452005 | Department of Clinical Microbiology, Copenhagen University Hospital, Hvidovre, Kettegaard Alle 30, 2650 Hvidovre. | Albertsen lab, Department of Chemistry and Bioscience, Aalborg University, Denmark |
| EPI_ISL_452006 | Department of Clinical Microbiology, Copenhagen University Hospital, Hvidovre, Kettegaard Alle 30, 2650 Hvidovre. | Albertsen lab, Department of Chemistry and Bioscience, Aalborg University, Denmark |
| EPI_ISL_452007 | Department of Clinical Microbiology, Copenhagen University Hospital, Hvidovre, Kettegaard Alle 30, 2650 Hvidovre. | Albertsen lab, Department of Chemistry and Bioscience, Aalborg University, Denmark |
| EPI_ISL_452008 | Department of Clinical Microbiology, Copenhagen University Hospital, Hvidovre, Kettegaard Alle 30, 2650 Hvidovre. | Albertsen lab, Department of Chemistry and Bioscience, Aalborg University, Denmark |
| EPI_ISL_452009 | Department of Clinical Microbiology, Copenhagen University Hospital, Hvidovre, Kettegaard Alle 30, 2650 Hvidovre. | Albertsen lab, Department of Chemistry and Bioscience, Aalborg University, Denmark |
| EPI_ISL_452010 | Department of Clinical Microbiology, Copenhagen University Hospital, Hvidovre, Kettegaard Alle 30, 2650 Hvidovre. | Albertsen lab, Department of Chemistry and Bioscience, Aalborg University, Denmark |

| EPI_Accession_ID | Originating Laboratory | Submitting Laboratory |
|---|---|---|
| EPI_ISL_452011 | Department of Clinical Microbiology, Copenhagen University Hospital, Hvidovre, Kettegaard Alle 30, 2650 Hvidovre. | Albertsen lab, Department of Chemistry and Bioscience, Aalborg University, Denmark |
| EPI_ISL_452012 | Department of Clinical Microbiology, Copenhagen University Hospital, Hvidovre, Kettegaard Alle 30, 2650 Hvidovre. | Albertsen lab, Department of Chemistry and Bioscience, Aalborg University, Denmark |
| EPI_ISL_452013 | Department of Clinical Microbiology, Copenhagen University Hospital, Hvidovre, Kettegaard Alle 30, 2650 Hvidovre. | Albertsen lab, Department of Chemistry and Bioscience, Aalborg University, Denmark |
| EPI_ISL_452014 | Department of Clinical Microbiology, Copenhagen University Hospital, Hvidovre, Kettegaard Alle 30, 2650 Hvidovre. | Albertsen lab, Department of Chemistry and Bioscience, Aalborg University, Denmark |
| EPI_ISL_452015 | Department of Clinical Microbiology, Copenhagen University Hospital, Hvidovre, Kettegaard Alle 30, 2650 Hvidovre. | Albertsen lab, Department of Chemistry and Bioscience, Aalborg University, Denmark |
| EPI_ISL_452016 | Department of Clinical Microbiology, Copenhagen University Hospital, Hvidovre, Kettegaard Alle 30, 2650 Hvidovre. | Albertsen lab, Department of Chemistry and Bioscience, Aalborg University, Denmark |
| EPI_ISL_452017 | Department of Clinical Microbiology, Copenhagen University Hospital, Hvidovre, Kettegaard Alle 30, 2650 Hvidovre. | Albertsen lab, Department of Chemistry and Bioscience, Aalborg University, Denmark |
| EPI_ISL_452018 | Department of Clinical Microbiology, Copenhagen University Hospital, Hvidovre, Kettegaard Alle 30, 2650 Hvidovre. | Albertsen lab, Department of Chemistry and Bioscience, Aalborg University, Denmark |
| EPI_ISL_452019 | Department of Clinical Microbiology, Copenhagen University Hospital, Hvidovre, Kettegaard Alle 30, 2650 Hvidovre. | Albertsen lab, Department of Chemistry and Bioscience, Aalborg University, Denmark |
| EPI_ISL_452020 | Department of Clinical Microbiology, Copenhagen University Hospital, Hvidovre, Kettegaard Alle 30, 2650 Hvidovre. | Albertsen lab, Department of Chemistry and Bioscience, Aalborg University, Denmark |
| EPI_ISL_452021 | Department of Clinical Microbiology, Copenhagen University Hospital, Hvidovre, Kettegaard Alle 30, 2650 Hvidovre. | Albertsen lab, Department of Chemistry and Bioscience, Aalborg University, Denmark |

| EPI_Accession_ID | Originating Laboratory | Submitting Laboratory |
|---|---|---|
| EPI_ISL_452022 | Department of Clinical Microbiology, Copenhagen University Hospital, Hvidovre, Kettegaard Alle 30, 2650 Hvidovre. | Albertsen lab, Department of Chemistry and Bioscience, Aalborg University, Denmark |
| EPI_ISL_452023 | Department of Clinical Microbiology, Copenhagen University Hospital, Hvidovre, Kettegaard Alle 30, 2650 Hvidovre. | Albertsen lab, Department of Chemistry and Bioscience, Aalborg University, Denmark |
| EPI_ISL_452024 | Department of Clinical Microbiology, Copenhagen University Hospital, Hvidovre, Kettegaard Alle 30, 2650 Hvidovre. | Albertsen lab, Department of Chemistry and Bioscience, Aalborg University, Denmark |
| EPI_ISL_452025 | Department of Clinical Microbiology, Copenhagen University Hospital, Hvidovre, Kettegaard Alle 30, 2650 Hvidovre. | Albertsen lab, Department of Chemistry and Bioscience, Aalborg University, Denmark |
| EPI_ISL_452026 | Department of Clinical Microbiology, Copenhagen University Hospital, Hvidovre, Kettegaard Alle 30, 2650 Hvidovre. | Albertsen lab, Department of Chemistry and Bioscience, Aalborg University, Denmark |
| EPI_ISL_452027 | Department of Clinical Microbiology, Copenhagen University Hospital, Hvidovre, Kettegaard Alle 30, 2650 Hvidovre. | Albertsen lab, Department of Chemistry and Bioscience, Aalborg University, Denmark |
| EPI_ISL_452028 | Department of Clinical Microbiology, Copenhagen University Hospital, Hvidovre, Kettegaard Alle 30, 2650 Hvidovre. | Albertsen lab, Department of Chemistry and Bioscience, Aalborg University, Denmark |
| EPI_ISL_452029 | Department of Clinical Microbiology, Copenhagen University Hospital, Hvidovre, Kettegaard Alle 30, 2650 Hvidovre. | Albertsen lab, Department of Chemistry and Bioscience, Aalborg University, Denmark |
| EPI_ISL_452030 | Department of Clinical Microbiology, Copenhagen University Hospital, Hvidovre, Kettegaard Alle 30, 2650 Hvidovre. | Albertsen lab, Department of Chemistry and Bioscience, Aalborg University, Denmark |
| EPI_ISL_452031 | Department of Clinical Microbiology, Copenhagen University Hospital, Hvidovre, Kettegaard Alle 30, 2650 Hvidovre. | Albertsen lab, Department of Chemistry and Bioscience, Aalborg University, Denmark |
| EPI_ISL_452032 | Department of Clinical Microbiology, Copenhagen University Hospital, Hvidovre, Kettegaard Alle 30, 2650 Hvidovre. | Albertsen lab, Department of Chemistry and Bioscience, Aalborg University, Denmark |

| EPI_Accession_ID | Originating Laboratory | Submitting Laboratory |
|---|---|---|
| EPI_ISL_452033 | Department of Clinical Microbiology, Copenhagen University Hospital, Hvidovre, Kettegaard Alle 30, 2650 Hvidovre. | Albertsen lab, Department of Chemistry and Bioscience, Aalborg University, Denmark |
| EPI_ISL_452034 | Department of Clinical Microbiology, Copenhagen University Hospital, Hvidovre, Kettegaard Alle 30, 2650 Hvidovre. | Albertsen lab, Department of Chemistry and Bioscience, Aalborg University, Denmark |
| EPI_ISL_452035 | Department of Clinical Microbiology, Copenhagen University Hospital, Hvidovre, Kettegaard Alle 30, 2650 Hvidovre. | Albertsen lab, Department of Chemistry and Bioscience, Aalborg University, Denmark |
| EPI_ISL_452036 | Department of Clinical Microbiology, Copenhagen University Hospital, Hvidovre, Kettegaard Alle 30, 2650 Hvidovre. | Albertsen lab, Department of Chemistry and Bioscience, Aalborg University, Denmark |
| EPI_ISL_452037 | Department of Clinical Microbiology, Copenhagen University Hospital, Hvidovre, Kettegaard Alle 30, 2650 Hvidovre. | Albertsen lab, Department of Chemistry and Bioscience, Aalborg University, Denmark |
| EPI_ISL_452038 | Department of Clinical Microbiology, Copenhagen University Hospital, Hvidovre, Kettegaard Alle 30, 2650 Hvidovre. | Albertsen lab, Department of Chemistry and Bioscience, Aalborg University, Denmark |
| EPI_ISL_452039 | Department of Clinical Microbiology, Copenhagen University Hospital, Hvidovre, Kettegaard Alle 30, 2650 Hvidovre. | Albertsen lab, Department of Chemistry and Bioscience, Aalborg University, Denmark |
| EPI_ISL_452040 | Department of Clinical Microbiology, Copenhagen University Hospital, Hvidovre, Kettegaard Alle 30, 2650 Hvidovre. | Albertsen lab, Department of Chemistry and Bioscience, Aalborg University, Denmark |
| EPI_ISL_452041 | Department of Clinical Microbiology, Copenhagen University Hospital, Hvidovre, Kettegaard Alle 30, 2650 Hvidovre. | Albertsen lab, Department of Chemistry and Bioscience, Aalborg University, Denmark |
| EPI_ISL_452042 | Department of Clinical Microbiology, Copenhagen University Hospital, Hvidovre, Kettegaard Alle 30, 2650 Hvidovre. | Albertsen lab, Department of Chemistry and Bioscience, Aalborg University, Denmark |
| EPI_ISL_452043 | Department of Clinical Microbiology, Copenhagen University Hospital, Hvidovre, Kettegaard Alle 30, 2650 Hvidovre. | Albertsen lab, Department of Chemistry and Bioscience, Aalborg University, Denmark |

| EPI_Accession_ID | Originating Laboratory | Submitting Laboratory |
|---|---|---|
| EPI_ISL_452044 | Department of Clinical Microbiology, Copenhagen University Hospital, Hvidovre, Kettegaard Alle 30, 2650 Hvidovre. | Albertsen lab, Department of Chemistry and Bioscience, Aalborg University, Denmark |
| EPI_ISL_452045 | Department of Clinical Microbiology, Copenhagen University Hospital, Hvidovre, Kettegaard Alle 30, 2650 Hvidovre. | Albertsen lab, Department of Chemistry and Bioscience, Aalborg University, Denmark |
| EPI_ISL_452046 | Department of Clinical Microbiology, Copenhagen University Hospital, Hvidovre, Kettegaard Alle 30, 2650 Hvidovre. | Albertsen lab, Department of Chemistry and Bioscience, Aalborg University, Denmark |
| EPI_ISL_452047 | Department of Clinical Microbiology, Copenhagen University Hospital, Hvidovre, Kettegaard Alle 30, 2650 Hvidovre. | Albertsen lab, Department of Chemistry and Bioscience, Aalborg University, Denmark |
| EPI_ISL_452048 | Department of Clinical Microbiology, Copenhagen University Hospital, Hvidovre, Kettegaard Alle 30, 2650 Hvidovre. | Albertsen lab, Department of Chemistry and Bioscience, Aalborg University, Denmark |
| EPI_ISL_452049 | Department of Clinical Microbiology, Copenhagen University Hospital, Hvidovre, Kettegaard Alle 30, 2650 Hvidovre. | Albertsen lab, Department of Chemistry and Bioscience, Aalborg University, Denmark |
| EPI_ISL_452050 | Department of Clinical Microbiology, Copenhagen University Hospital, Hvidovre, Kettegaard Alle 30, 2650 Hvidovre. | Albertsen lab, Department of Chemistry and Bioscience, Aalborg University, Denmark |
| EPI_ISL_452051 | Department of Clinical Microbiology, Copenhagen University Hospital, Hvidovre, Kettegaard Alle 30, 2650 Hvidovre. | Albertsen lab, Department of Chemistry and Bioscience, Aalborg University, Denmark |
| EPI_ISL_452052 | Department of Clinical Microbiology, Copenhagen University Hospital, Hvidovre, Kettegaard Alle 30, 2650 Hvidovre. | Albertsen lab, Department of Chemistry and Bioscience, Aalborg University, Denmark |
| EPI_ISL_452053 | Department of Clinical Microbiology, Copenhagen University Hospital, Hvidovre, Kettegaard Alle 30, 2650 Hvidovre. | Albertsen lab, Department of Chemistry and Bioscience, Aalborg University, Denmark |
| EPI_ISL_452054 | Department of Clinical Microbiology, Copenhagen University Hospital, Hvidovre, Kettegaard Alle 30, 2650 Hvidovre. | Albertsen lab, Department of Chemistry and Bioscience, Aalborg University, Denmark |

| EPI_Accession_ID | Originating Laboratory | Submitting Laboratory |
|---|---|---|
| EPI_ISL_452055 | Department of Clinical Microbiology, Copenhagen University Hospital, Hvidovre, Kettegaard Alle 30, 2650 Hvidovre. | Albertsen lab, Department of Chemistry and Bioscience, Aalborg University, Denmark |
| EPI_ISL_452056 | Department of Clinical Microbiology, Copenhagen University Hospital, Hvidovre, Kettegaard Alle 30, 2650 Hvidovre. | Albertsen lab, Department of Chemistry and Bioscience, Aalborg University, Denmark |
| EPI_ISL_452057 | Department of Clinical Microbiology, Copenhagen University Hospital, Hvidovre, Kettegaard Alle 30, 2650 Hvidovre. | Albertsen lab, Department of Chemistry and Bioscience, Aalborg University, Denmark |
| EPI_ISL_452058 | Department of Clinical Microbiology, Copenhagen University Hospital, Hvidovre, Kettegaard Alle 30, 2650 Hvidovre. | Albertsen lab, Department of Chemistry and Bioscience, Aalborg University, Denmark |
| EPI_ISL_452059 | Department of Clinical Microbiology, Copenhagen University Hospital, Hvidovre, Kettegaard Alle 30, 2650 Hvidovre. | Albertsen lab, Department of Chemistry and Bioscience, Aalborg University, Denmark |
| EPI_ISL_452060 | Department of Clinical Microbiology, Copenhagen University Hospital, Hvidovre, Kettegaard Alle 30, 2650 Hvidovre. | Albertsen lab, Department of Chemistry and Bioscience, Aalborg University, Denmark |
| EPI_ISL_452061 | Department of Clinical Microbiology, Copenhagen University Hospital, Hvidovre, Kettegaard Alle 30, 2650 Hvidovre. | Albertsen lab, Department of Chemistry and Bioscience, Aalborg University, Denmark |
| EPI_ISL_452062 | Department of Clinical Microbiology, Copenhagen University Hospital, Hvidovre, Kettegaard Alle 30, 2650 Hvidovre. | Albertsen lab, Department of Chemistry and Bioscience, Aalborg University, Denmark |
| EPI_ISL_452063 | Department of Clinical Microbiology, Copenhagen University Hospital, Hvidovre, Kettegaard Alle 30, 2650 Hvidovre. | Albertsen lab, Department of Chemistry and Bioscience, Aalborg University, Denmark |
| EPI_ISL_452064 | Department of Clinical Microbiology, Copenhagen University Hospital, Hvidovre, Kettegaard Alle 30, 2650 Hvidovre. | Albertsen lab, Department of Chemistry and Bioscience, Aalborg University, Denmark |
| EPI_ISL_452065 | Department of Clinical Microbiology, Copenhagen University Hospital, Hvidovre, Kettegaard Alle 30, 2650 Hvidovre. | Albertsen lab, Department of Chemistry and Bioscience, Aalborg University, Denmark |

| EPI_Accession_ID | Originating Laboratory | Submitting Laboratory |
|---|---|---|
| EPI_ISL_452066 | Department of Clinical Microbiology, Copenhagen University Hospital, Hvidovre, Kettegaard Alle 30, 2650 Hvidovre. | Albertsen lab, Department of Chemistry and Bioscience, Aalborg University, Denmark |
| EPI_ISL_452067 | Department of Clinical Microbiology, Copenhagen University Hospital, Hvidovre, Kettegaard Alle 30, 2650 Hvidovre. | Albertsen lab, Department of Chemistry and Bioscience, Aalborg University, Denmark |
| EPI_ISL_452068 | Department of Clinical Microbiology, Copenhagen University Hospital, Hvidovre, Kettegaard Alle 30, 2650 Hvidovre. | Albertsen lab, Department of Chemistry and Bioscience, Aalborg University, Denmark |
| EPI_ISL_452069 | Department of Clinical Microbiology, Copenhagen University Hospital, Hvidovre, Kettegaard Alle 30, 2650 Hvidovre. | Albertsen lab, Department of Chemistry and Bioscience, Aalborg University, Denmark |
| EPI_ISL_452070 | Department of Clinical Microbiology, Copenhagen University Hospital, Hvidovre, Kettegaard Alle 30, 2650 Hvidovre. | Albertsen lab, Department of Chemistry and Bioscience, Aalborg University, Denmark |
| EPI_ISL_452071 | Department of Clinical Microbiology, Copenhagen University Hospital, Hvidovre, Kettegaard Alle 30, 2650 Hvidovre. | Albertsen lab, Department of Chemistry and Bioscience, Aalborg University, Denmark |
| EPI_ISL_452072 | Department of Clinical Microbiology, Copenhagen University Hospital, Hvidovre, Kettegaard Alle 30, 2650 Hvidovre. | Albertsen lab, Department of Chemistry and Bioscience, Aalborg University, Denmark |
| EPI_ISL_452073 | Department of Clinical Microbiology, Copenhagen University Hospital, Hvidovre, Kettegaard Alle 30, 2650 Hvidovre. | Albertsen lab, Department of Chemistry and Bioscience, Aalborg University, Denmark |
| EPI_ISL_452074 | Department of Clinical Microbiology, Copenhagen University Hospital, Hvidovre, Kettegaard Alle 30, 2650 Hvidovre. | Albertsen lab, Department of Chemistry and Bioscience, Aalborg University, Denmark |
| EPI_ISL_452075 | Department of Clinical Microbiology, Copenhagen University Hospital, Hvidovre, Kettegaard Alle 30, 2650 Hvidovre. | Albertsen lab, Department of Chemistry and Bioscience, Aalborg University, Denmark |
| EPI_ISL_452076 | Department of Clinical Microbiology, Copenhagen University Hospital, Hvidovre, Kettegaard Alle 30, 2650 Hvidovre. | Albertsen lab, Department of Chemistry and Bioscience, Aalborg University, Denmark |

| EPI_Accession_ID | Originating Laboratory | Submitting Laboratory |
| --- | --- | --- |
| EPI_ISL_452077 | Department of Clinical Microbiology, Copenhagen University Hospital, Hvidovre, Kettegaard Alle 30, 2650 Hvidovre. | Albertsen lab, Department of Chemistry and Bioscience, Aalborg University, Denmark |
| EPI_ISL_452078 | Department of Clinical Microbiology, Copenhagen University Hospital, Hvidovre, Kettegaard Alle 30, 2650 Hvidovre. | Albertsen lab, Department of Chemistry and Bioscience, Aalborg University, Denmark |
| EPI_ISL_452079 | Department of Clinical Microbiology, Copenhagen University Hospital, Hvidovre, Kettegaard Alle 30, 2650 Hvidovre. | Albertsen lab, Department of Chemistry and Bioscience, Aalborg University, Denmark |
| EPI_ISL_452080 | Department of Clinical Microbiology, Copenhagen University Hospital, Hvidovre, Kettegaard Alle 30, 2650 Hvidovre. | Albertsen lab, Department of Chemistry and Bioscience, Aalborg University, Denmark |
| EPI_ISL_452081 | Department of Clinical Microbiology, Copenhagen University Hospital, Hvidovre, Kettegaard Alle 30, 2650 Hvidovre. | Albertsen lab, Department of Chemistry and Bioscience, Aalborg University, Denmark |
| EPI_ISL_452082 | Department of Clinical Microbiology, Copenhagen University Hospital, Hvidovre, Kettegaard Alle 30, 2650 Hvidovre. | Albertsen lab, Department of Chemistry and Bioscience, Aalborg University, Denmark |
| EPI_ISL_452083 | Department of Clinical Microbiology, Copenhagen University Hospital, Hvidovre, Kettegaard Alle 30, 2650 Hvidovre. | Albertsen lab, Department of Chemistry and Bioscience, Aalborg University, Denmark |
| EPI_ISL_452084 | Department of Clinical Microbiology, Copenhagen University Hospital, Hvidovre, Kettegaard Alle 30, 2650 Hvidovre. | Albertsen lab, Department of Chemistry and Bioscience, Aalborg University, Denmark |
| EPI_ISL_452085 | Department of Clinical Microbiology, Copenhagen University Hospital, Hvidovre, Kettegaard Alle 30, 2650 Hvidovre. | Albertsen lab, Department of Chemistry and Bioscience, Aalborg University, Denmark |
| EPI_ISL_452086 | Department of Clinical Microbiology, Copenhagen University Hospital, Hvidovre, Kettegaard Alle 30, 2650 Hvidovre. | Albertsen lab, Department of Chemistry and Bioscience, Aalborg University, Denmark |
| EPI_ISL_452087 | Department of Clinical Microbiology, Copenhagen University Hospital, Hvidovre, Kettegaard Alle 30, 2650 Hvidovre. | Albertsen lab, Department of Chemistry and Bioscience, Aalborg University, Denmark |

| EPI_Accession_ID | Originating Laboratory | Submitting Laboratory |
|---|---|---|
| EPI_ISL_452088 | Department of Clinical Microbiology, Copenhagen University Hospital, Hvidovre, Kettegaard Alle 30, 2650 Hvidovre. | Albertsen lab, Department of Chemistry and Bioscience, Aalborg University, Denmark |
| EPI_ISL_452089 | Department of Clinical Microbiology, Copenhagen University Hospital, Hvidovre, Kettegaard Alle 30, 2650 Hvidovre. | Albertsen lab, Department of Chemistry and Bioscience, Aalborg University, Denmark |
| EPI_ISL_452090 | Department of Clinical Microbiology, Copenhagen University Hospital, Hvidovre, Kettegaard Alle 30, 2650 Hvidovre. | Albertsen lab, Department of Chemistry and Bioscience, Aalborg University, Denmark |
| EPI_ISL_452091 | Department of Clinical Microbiology, Copenhagen University Hospital, Hvidovre, Kettegaard Alle 30, 2650 Hvidovre. | Albertsen lab, Department of Chemistry and Bioscience, Aalborg University, Denmark |
| EPI_ISL_452092 | Department of Clinical Microbiology, Copenhagen University Hospital, Hvidovre, Kettegaard Alle 30, 2650 Hvidovre. | Albertsen lab, Department of Chemistry and Bioscience, Aalborg University, Denmark |
| EPI_ISL_452093 | Department of Clinical Microbiology, Copenhagen University Hospital, Hvidovre, Kettegaard Alle 30, 2650 Hvidovre. | Albertsen lab, Department of Chemistry and Bioscience, Aalborg University, Denmark |
| EPI_ISL_452094 | Department of Clinical Microbiology, Copenhagen University Hospital, Hvidovre, Kettegaard Alle 30, 2650 Hvidovre. | Albertsen lab, Department of Chemistry and Bioscience, Aalborg University, Denmark |
| EPI_ISL_452095 | Department of Clinical Microbiology, Copenhagen University Hospital, Hvidovre, Kettegaard Alle 30, 2650 Hvidovre. | Albertsen lab, Department of Chemistry and Bioscience, Aalborg University, Denmark |
| EPI_ISL_452096 | Department of Clinical Microbiology, Copenhagen University Hospital, Hvidovre, Kettegaard Alle 30, 2650 Hvidovre. | Albertsen lab, Department of Chemistry and Bioscience, Aalborg University, Denmark |
| EPI_ISL_452097 | Department of Clinical Microbiology, Copenhagen University Hospital, Hvidovre, Kettegaard Alle 30, 2650 Hvidovre. | Albertsen lab, Department of Chemistry and Bioscience, Aalborg University, Denmark |
| EPI_ISL_452098 | Department of Clinical Microbiology, Copenhagen University Hospital, Hvidovre, Kettegaard Alle 30, 2650 Hvidovre. | Albertsen lab, Department of Chemistry and Bioscience, Aalborg University, Denmark |

| EPI_Accession_ID | Originating Laboratory | Submitting Laboratory |
|---|---|---|
| EPI_ISL_452099 | Department of Clinical Microbiology, Copenhagen University Hospital, Hvidovre, Kettegaard Alle 30, 2650 Hvidovre. | Albertsen lab, Department of Chemistry and Bioscience, Aalborg University, Denmark |
| EPI_ISL_452100 | Department of Clinical Microbiology, Copenhagen University Hospital, Hvidovre, Kettegaard Alle 30, 2650 Hvidovre. | Albertsen lab, Department of Chemistry and Bioscience, Aalborg University, Denmark |
| EPI_ISL_452101 | Department of Virus and Microbiological Special Diagnostics, Statens Serum Institut, Copenhagen, Denmark, Artillerivej 5, 2300 Copenahgen S | Albertsen lab, Department of Chemistry and Bioscience, Aalborg University, Denmark |
| EPI_ISL_452102 | Department of Virus and Microbiological Special Diagnostics, Statens Serum Institut, Copenhagen, Denmark, Artillerivej 5, 2300 Copenahgen S | Albertsen lab, Department of Chemistry and Bioscience, Aalborg University, Denmark |
| EPI_ISL_452103 | Max von Pettenkofer Institute, Virology, National Reference Center for Retroviruses, LMU MÃ¼nchen | Laboratory for Functional Genome Analysis, Dept. Genomics, Gene Center of the LMU Munich |
| EPI_ISL_452104 | Max von Pettenkofer Institute, Virology, National Reference Center for Retroviruses, LMU MÃ¼nchen | Laboratory for Functional Genome Analysis, Dept. Genomics, Gene Center of the LMU Munich |
| EPI_ISL_452105 | Texas DSHS Lab Services | Pathogen Discovery, Respiratory Viruses Branch, Division of Viral Diseases, Centers for Disease Control and Prevention |
| EPI_ISL_452106 | LA Office of Public Health Laboratories | Pathogen Discovery, Respiratory Viruses Branch, Division of Viral Diseases, Centers for Disease Control and Prevention |
| EPI_ISL_452107 | LA Office of Public Health Laboratories | Pathogen Discovery, Respiratory Viruses Branch, Division of Viral Diseases, Centers for Disease Control and Prevention |
| EPI_ISL_452108 | MN Department of Health | Pathogen Discovery, Respiratory Viruses Branch, Division of Viral Diseases, Centers for Disease Control and Prevention |
| EPI_ISL_452109 | NM Department of Health | Pathogen Discovery, Respiratory Viruses Branch, Division of Viral Diseases, Centers for Disease Control and Prevention |

| EPI_Accession_ID | Originating Laboratory | Submitting Laboratory |
|---|---|---|
| EPI_ISL_452110 | FL Bureau of Public Health Laboratories | Pathogen Discovery, Respiratory Viruses Branch, Division of Viral Diseases, Centers for Disease Control and Prevention |
| EPI_ISL_452111 | FL Bureau of Public Health Laboratories | Pathogen Discovery, Respiratory Viruses Branch, Division of Viral Diseases, Centers for Disease Control and Prevention |
| EPI_ISL_452112 | Texas DSHS Lab Services | Pathogen Discovery, Respiratory Viruses Branch, Division of Viral Diseases, Centers for Disease Control and Prevention |
| EPI_ISL_452113 | MN Department of Health | Pathogen Discovery, Respiratory Viruses Branch, Division of Viral Diseases, Centers for Disease Control and Prevention |
| EPI_ISL_452114 | KS Health and Environmental Laboratories | Pathogen Discovery, Respiratory Viruses Branch, Division of Viral Diseases, Centers for Disease Control and Prevention |
| EPI_ISL_452115 | Texas DSHS Lab Services | Pathogen Discovery, Respiratory Viruses Branch, Division of Viral Diseases, Centers for Disease Control and Prevention |
| EPI_ISL_452116 | NJ Public Health and Environmental Laboratories | Pathogen Discovery, Respiratory Viruses Branch, Division of Viral Diseases, Centers for Disease Control and Prevention |
| EPI_ISL_452117 | NJ Public Health and Environmental Laboratories | Pathogen Discovery, Respiratory Viruses Branch, Division of Viral Diseases, Centers for Disease Control and Prevention |
| EPI_ISL_452118 | AZ SPHL, Arizona Department of Health Services | Pathogen Discovery, Respiratory Viruses Branch, Division of Viral Diseases, Centers for Disease Control and Prevention |
| EPI_ISL_452119 | VI-US Virgin Islands Department of Health | Pathogen Discovery, Respiratory Viruses Branch, Division of Viral Diseases, Centers for Disease Control and Prevention |
| EPI_ISL_452120 | Georgia Department of Health | Pathogen Discovery, Respiratory Viruses Branch, Division of Viral Diseases, Centers for Disease Control and Prevention |

| EPI_Accession_ID | Originating Laboratory | Submitting Laboratory |
|---|---|---|
| EPI_ISL_452121 | VI-US Virgin Islands Department of Health | Pathogen Discovery, Respiratory Viruses Branch, Division of Viral Diseases, Centers for Disease Control and Prevention |
| EPI_ISL_452122 | VI-US Virgin Islands Department of Health | Pathogen Discovery, Respiratory Viruses Branch, Division of Viral Diseases, Centers for Disease Control and Prevention |
| EPI_ISL_452123 | VI-US Virgin Islands Department of Health | Pathogen Discovery, Respiratory Viruses Branch, Division of Viral Diseases, Centers for Disease Control and Prevention |
| EPI_ISL_452124 | NC State Laboratory of Public Health | Pathogen Discovery, Respiratory Viruses Branch, Division of Viral Diseases, Centers for Disease Control and Prevention |
| EPI_ISL_452125 | Georgia Department of Health | Pathogen Discovery, Respiratory Viruses Branch, Division of Viral Diseases, Centers for Disease Control and Prevention |
| EPI_ISL_452126 | NC State Laboratory of Public Health | Pathogen Discovery, Respiratory Viruses Branch, Division of Viral Diseases, Centers for Disease Control and Prevention |
| EPI_ISL_452127 | CO Department of Public Health and Environment | Pathogen Discovery, Respiratory Viruses Branch, Division of Viral Diseases, Centers for Disease Control and Prevention |
| EPI_ISL_452128 | CO Department of Public Health and Environment | Pathogen Discovery, Respiratory Viruses Branch, Division of Viral Diseases, Centers for Disease Control and Prevention |
| EPI_ISL_452129 | CO Department of Public Health and Environment | Pathogen Discovery, Respiratory Viruses Branch, Division of Viral Diseases, Centers for Disease Control and Prevention |
| EPI_ISL_452130 | CO Department of Public Health and Environment | Pathogen Discovery, Respiratory Viruses Branch, Division of Viral Diseases, Centers for Disease Control and Prevention |
| EPI_ISL_452131 | IN State Department of Health Laboratory Services | Pathogen Discovery, Respiratory Viruses Branch, Division of Viral Diseases, Centers for Disease Control and Prevention |

| EPI_Accession_ID | Originating Laboratory | Submitting Laboratory |
|---|---|---|
| EPI_ISL_452132 | FL Bureau of Public Health Laboratories | Pathogen Discovery, Respiratory Viruses Branch, Division of Viral Diseases, Centers for Disease Control and Prevention |
| EPI_ISL_452133 | MN Department of Health | Pathogen Discovery, Respiratory Viruses Branch, Division of Viral Diseases, Centers for Disease Control and Prevention |
| EPI_ISL_452134 | IL Department of Public Health Chicago Laboratory | Pathogen Discovery, Respiratory Viruses Branch, Division of Viral Diseases, Centers for Disease Control and Prevention |
| EPI_ISL_452135 | IN State Department of Health Laboratory Services | Pathogen Discovery, Respiratory Viruses Branch, Division of Viral Diseases, Centers for Disease Control and Prevention |
| EPI_ISL_452136 | IN State Department of Health Laboratory Services | Pathogen Discovery, Respiratory Viruses Branch, Division of Viral Diseases, Centers for Disease Control and Prevention |
| EPI_ISL_452137 | VI-US Virgin Islands Department of Health | Pathogen Discovery, Respiratory Viruses Branch, Division of Viral Diseases, Centers for Disease Control and Prevention |
| EPI_ISL_452138 | VI-US Virgin Islands Department of Health | Pathogen Discovery, Respiratory Viruses Branch, Division of Viral Diseases, Centers for Disease Control and Prevention |
| EPI_ISL_452139 | Instituto de Diagnostico y Referencia Epidemiologicos (INDRE) | Instituto de diagnÃ³stico y Referencia Epidemiologicos (INDRE) |
| EPI_ISL_452140 | CUB Hopital Erasme Laboratoire d'Anatomie Pathologique | CUB Hopital Erasme Laboratoire d'Anatomie Pathologique |
| EPI_ISL_452141 | Instituto de Diagnostico y Referencia Epidemiologicos (INDRE) | Instituto de diagnÃ³stico y Referencia Epidemiologicos (INDRE) |
| EPI_ISL_452142 | CUB Hopital Erasme Laboratoire d'Anatomie Pathologique | CUB Hopital Erasme Laboratoire d'Anatomie Pathologique |
| EPI_ISL_452144 | Yale COVID-19 Biorepository | Grubaugh Lab - Yale School of Public Health |
| EPI_ISL_452145 | Yale COVID-19 Biorepository | Grubaugh Lab - Yale School of Public Health |
| EPI_ISL_452146 | Yale COVID-19 Biorepository | Grubaugh Lab - Yale School of Public Health |
| EPI_ISL_452147 | Yale COVID-19 Biorepository | Grubaugh Lab - Yale School of Public Health |

| EPI_Accession_ID | Originating Laboratory | Submitting Laboratory |
|---|---|---|
| EPI_ISL_452148 | CUB Hopital Erasme Laboratoire d'Anatomie Pathologique | CUB Hopital Erasme Laboratoire d'Anatomie Pathologique |
| EPI_ISL_452149 | CUB Hopital Erasme Laboratoire d'Anatomie Pathologique | CUB Hopital Erasme Laboratoire d'Anatomie Pathologique |
| EPI_ISL_452150 | CUB Hopital Erasme Laboratoire d'Anatomie Pathologique | CUB Hopital Erasme Laboratoire d'Anatomie Pathologique |
| EPI_ISL_452151 | CUB Hopital Erasme Laboratoire d'Anatomie Pathologique | CUB Hopital Erasme Laboratoire d'Anatomie Pathologique |
| EPI_ISL_452152 | CUB Hopital Erasme Laboratoire d'Anatomie Pathologique | CUB Hopital Erasme Laboratoire d'Anatomie Pathologique |
| EPI_ISL_452153 | Utah Public Health Laboratory | Utah Public Health Laboratory |
| EPI_ISL_452154 | Utah Public Health Laboratory | Utah Public Health Laboratory |
| EPI_ISL_452155 | Utah Public Health Laboratory | Utah Public Health Laboratory |
| EPI_ISL_452156 | Utah Public Health Laboratory | Utah Public Health Laboratory |
| EPI_ISL_452157 | Utah Public Health Laboratory | Utah Public Health Laboratory |
| EPI_ISL_452158 | Utah Public Health Laboratory | Utah Public Health Laboratory |
| EPI_ISL_452159 | Utah Public Health Laboratory | Utah Public Health Laboratory |
| EPI_ISL_452160 | Utah Public Health Laboratory | Utah Public Health Laboratory |
| EPI_ISL_452161 | Utah Public Health Laboratory | Utah Public Health Laboratory |
| EPI_ISL_452162 | Utah Public Health Laboratory | Utah Public Health Laboratory |
| EPI_ISL_452163 | Utah Public Health Laboratory | Utah Public Health Laboratory |
| EPI_ISL_452164 | Utah Public Health Laboratory | Utah Public Health Laboratory |
| EPI_ISL_452165 | Utah Public Health Laboratory | Utah Public Health Laboratory |
| EPI_ISL_452166 | Utah Public Health Laboratory | Utah Public Health Laboratory |
| EPI_ISL_452167 | Utah Public Health Laboratory | Utah Public Health Laboratory |
| EPI_ISL_452168 | Utah Public Health Laboratory | Utah Public Health Laboratory |
| EPI_ISL_452169 | Utah Public Health Laboratory | Utah Public Health Laboratory |
| EPI_ISL_452170 | Utah Public Health Laboratory | Utah Public Health Laboratory |
| EPI_ISL_452171 | Utah Public Health Laboratory | Utah Public Health Laboratory |
| EPI_ISL_452172 | Utah Public Health Laboratory | Utah Public Health Laboratory |
| EPI_ISL_452173 | Utah Public Health Laboratory | Utah Public Health Laboratory |
| EPI_ISL_452174 | Utah Public Health Laboratory | Utah Public Health Laboratory |
| EPI_ISL_452175 | Utah Public Health Laboratory | Utah Public Health Laboratory |

| EPI_Accession_ID | Originating Laboratory | Submitting Laboratory |
|---|---|---|
| EPI_ISL_452176 | Utah Public Health Laboratory | Utah Public Health Laboratory |
| EPI_ISL_452177 | Utah Public Health Laboratory | Utah Public Health Laboratory |
| EPI_ISL_452178 | Laboratory Medicine | Department of Laboratory Medicine, Lin-Kou Chang Gung Memorial Hospital, Taoyuan, Taiwan |
| EPI_ISL_452179 | Laboratory Medicine | Department of Laboratory Medicine, Lin-Kou Chang Gung Memorial Hospital, Taoyuan, Taiwan |
| EPI_ISL_452180 | Center of Medical Microbiology, Virology, and Hospital Hygiene, University of Duesseldorf | Center of Medical Microbiology, Virology, and Hospital Hygiene, University of Duesseldorf |
| EPI_ISL_452181 | ULSS9 Distretto di Bussolengo | Istituto Zooprofilattico Sperimentale delle Venezie |
| EPI_ISL_452182 | ULSS9 Distretto di Bussolengo | Istituto Zooprofilattico Sperimentale delle Venezie |
| EPI_ISL_452183 | ULSS9 Distretto di Bussolengo | Istituto Zooprofilattico Sperimentale delle Venezie |
| EPI_ISL_452184 | ULSS9 Distretto di Bussolengo | Istituto Zooprofilattico Sperimentale delle Venezie |
| EPI_ISL_452185 | ULSS9 Distretto di Bussolengo | Istituto Zooprofilattico Sperimentale delle Venezie |
| EPI_ISL_452186 | ULSS9 Distretto di Bussolengo | Istituto Zooprofilattico Sperimentale delle Venezie |
| EPI_ISL_452187 | ULSS9 Distretto di Bussolengo | Istituto Zooprofilattico Sperimentale delle Venezie |
| EPI_ISL_452188 | ULSS9 Distretto di Bussolengo | Istituto Zooprofilattico Sperimentale delle Venezie |
| EPI_ISL_452189 | ULSS9 Distretto di Bussolengo | Istituto Zooprofilattico Sperimentale delle Venezie |
| EPI_ISL_452190 | ULSS9 Distretto di San Bonifacio | Istituto Zooprofilattico Sperimentale delle Venezie |
| EPI_ISL_452191 | ULSS9 Distretto di San Bonifacio | Istituto Zooprofilattico Sperimentale delle Venezie |
| EPI_ISL_452192 | NIV Influenza | NIV Influenza |
| EPI_ISL_452193 | NIV Influenza | NIV Influenza |
| EPI_ISL_452194 | NIV Influenza | NIV Influenza |
| EPI_ISL_452195 | NIV Influenza | NIV Influenza |
| EPI_ISL_452196 | NIV Influenza | NIV Influenza |
| EPI_ISL_452197 | NIV Influenza | NIV Influenza |
| EPI_ISL_452198 | NIV Influenza | NIV Influenza |
| EPI_ISL_452199 | NIV Influenza | NIV Influenza |
| EPI_ISL_452200 | NIV Influenza | NIV Influenza |
| EPI_ISL_452201 | NIV Influenza | NIV Influenza |
| EPI_ISL_452202 | NIV Influenza | NIV Influenza |
| EPI_ISL_452203 | NIV Influenza | NIV Influenza |
| EPI_ISL_452204 | NIV Influenza | NIV Influenza |
| EPI_ISL_452205 | NIV Influenza | NIV Influenza |

| EPI_Accession_ID | Originating Laboratory | Submitting Laboratory |
|---|---|---|
| EPI_ISL_452206 | NIV Influenza | NIV Influenza |
| EPI_ISL_452207 | NIV Influenza | NIV Influenza |
| EPI_ISL_452208 | NIV Influenza | NIV Influenza |
| EPI_ISL_452209 | NIV Influenza | NIV Influenza |
| EPI_ISL_452210 | NIV Influenza | NIV Influenza |
| EPI_ISL_452211 | NIV Influenza | NIV Influenza |
| EPI_ISL_452212 | NIV Influenza | NIV Influenza |
| EPI_ISL_452213 | NIV Influenza | NIV Influenza |
| EPI_ISL_452214 | NIV Influenza | NIV Influenza |
| EPI_ISL_452215 | NIV Influenza | NIV Influenza |
| EPI_ISL_452216 | NIV Influenza | NIV Influenza |
| EPI_ISL_452217 | NIV Influenza | NIV Influenza |
| EPI_ISL_452218 | University Hospital Frankfurt | Widera/Toptan |
| EPI_ISL_452219 | University Hospital Frankfurt | Widera/Toptan |
| EPI_ISL_452220 | University Hospital Frankfurt | Widera/Toptan |
| EPI_ISL_452221 | University Hospital Frankfurt | Widera/Toptan |
| EPI_ISL_452222 | University Hospital Frankfurt | Widera/Toptan |
| EPI_ISL_452223 | University Hospital Frankfurt | Widera/Toptan |
| EPI_ISL_452229 | Narhalsan Backa vardcentral | The Public Health Agency of Sweden |
| EPI_ISL_452230 | Narhalsan Backa vardcentral | The Public Health Agency of Sweden |
| EPI_ISL_452231 | Saroledens Familjelakare | The Public Health Agency of Sweden |
| EPI_ISL_452232 | Saroledens Familjelakare | The Public Health Agency of Sweden |
| EPI_ISL_452233 | Victoria Vard och Halsa | The Public Health Agency of Sweden |
| EPI_ISL_452234 | Din Klinik | The Public Health Agency of Sweden |
| EPI_ISL_452235 | Narhalsan Backa vardcentral | The Public Health Agency of Sweden |
| EPI_ISL_452236 | VC Sorgenfrimottagningen | The Public Health Agency of Sweden |
| EPI_ISL_452237 | Huddinge VC | The Public Health Agency of Sweden |
| EPI_ISL_452238 | Narhalsan Sjobo vardcentral | The Public Health Agency of Sweden |
| EPI_ISL_452239 | Narhalsan Backa vardcentral | The Public Health Agency of Sweden |
| EPI_ISL_452240 | Huddinge VC | The Public Health Agency of Sweden |
| EPI_ISL_452241 | Huslakarna Varmbadhuset Varberg | The Public Health Agency of Sweden |
| EPI_ISL_452242 | Wernstedt Medical AB | The Public Health Agency of Sweden |

| EPI_Accession_ID | Originating Laboratory | Submitting Laboratory |
|---|---|---|
| EPI_ISL_452249 | Respiratory Virus Unit, Microbiology Services Colindale, Public Health England | Respiratory Virus Unit, Microbiology Services Colindale, Public Health England |
| EPI_ISL_452250 | Respiratory Virus Unit, Microbiology Services Colindale, Public Health England | Respiratory Virus Unit, Microbiology Services Colindale, Public Health England |
| EPI_ISL_452251 | Respiratory Virus Unit, Microbiology Services Colindale, Public Health England | Respiratory Virus Unit, Microbiology Services Colindale, Public Health England |
| EPI_ISL_452252 | Respiratory Virus Unit, Microbiology Services Colindale, Public Health England | Respiratory Virus Unit, Microbiology Services Colindale, Public Health England |
| EPI_ISL_452253 | Respiratory Virus Unit, Microbiology Services Colindale, Public Health England | Respiratory Virus Unit, Microbiology Services Colindale, Public Health England |
| EPI_ISL_452254 | Respiratory Virus Unit, Microbiology Services Colindale, Public Health England | Respiratory Virus Unit, Microbiology Services Colindale, Public Health England |
| EPI_ISL_452255 | Respiratory Virus Unit, Microbiology Services Colindale, Public Health England | Respiratory Virus Unit, Microbiology Services Colindale, Public Health England |
| EPI_ISL_452256 | Respiratory Virus Unit, Microbiology Services Colindale, Public Health England | Respiratory Virus Unit, Microbiology Services Colindale, Public Health England |
| EPI_ISL_452257 | Respiratory Virus Unit, Microbiology Services Colindale, Public Health England | Respiratory Virus Unit, Microbiology Services Colindale, Public Health England |
| EPI_ISL_452258 | Respiratory Virus Unit, Microbiology Services Colindale, Public Health England | Respiratory Virus Unit, Microbiology Services Colindale, Public Health England |
| EPI_ISL_452262 | Michigan Department of Health and Human Services, Bureau of Laboratories | Michigan Department of Health and Human Services, Bureau of Laboratories |
| EPI_ISL_452263 | Michigan Department of Health and Human Services, Bureau of Laboratories | Michigan Department of Health and Human Services, Bureau of Laboratories |
| EPI_ISL_452264 | Michigan Department of Health and Human Services, Bureau of Laboratories | Michigan Department of Health and Human Services, Bureau of Laboratories |
| EPI_ISL_452265 | Michigan Department of Health and Human Services, Bureau of Laboratories | Michigan Department of Health and Human Services, Bureau of Laboratories |
| EPI_ISL_452266 | Michigan Department of Health and Human Services, Bureau of Laboratories | Michigan Department of Health and Human Services, Bureau of Laboratories |
| EPI_ISL_452267 | Michigan Department of Health and Human Services, Bureau of Laboratories | Michigan Department of Health and Human Services, Bureau of Laboratories |

| EPI_Accession_ID | Originating Laboratory | Submitting Laboratory |
|---|---|---|
| EPI_ISL_452268 | Michigan Department of Health and Human Services, Bureau of Laboratories | Michigan Department of Health and Human Services, Bureau of Laboratories |
| EPI_ISL_452269 | Michigan Department of Health and Human Services, Bureau of Laboratories | Michigan Department of Health and Human Services, Bureau of Laboratories |
| EPI_ISL_452270 | Michigan Department of Health and Human Services, Bureau of Laboratories | Michigan Department of Health and Human Services, Bureau of Laboratories |
| EPI_ISL_452271 | Michigan Department of Health and Human Services, Bureau of Laboratories | Michigan Department of Health and Human Services, Bureau of Laboratories |
| EPI_ISL_452272 | Michigan Department of Health and Human Services, Bureau of Laboratories | Michigan Department of Health and Human Services, Bureau of Laboratories |
| EPI_ISL_452273 | Michigan Department of Health and Human Services, Bureau of Laboratories | Michigan Department of Health and Human Services, Bureau of Laboratories |
| EPI_ISL_452274 | Michigan Department of Health and Human Services, Bureau of Laboratories | Michigan Department of Health and Human Services, Bureau of Laboratories |
| EPI_ISL_452275 | Michigan Department of Health and Human Services, Bureau of Laboratories | Michigan Department of Health and Human Services, Bureau of Laboratories |
| EPI_ISL_452276 | Michigan Department of Health and Human Services, Bureau of Laboratories | Michigan Department of Health and Human Services, Bureau of Laboratories |
| EPI_ISL_452277 | Michigan Department of Health and Human Services, Bureau of Laboratories | Michigan Department of Health and Human Services, Bureau of Laboratories |
| EPI_ISL_452278 | Michigan Department of Health and Human Services, Bureau of Laboratories | Michigan Department of Health and Human Services, Bureau of Laboratories |
| EPI_ISL_452279 | Michigan Department of Health and Human Services, Bureau of Laboratories | Michigan Department of Health and Human Services, Bureau of Laboratories |
| EPI_ISL_452280 | Michigan Department of Health and Human Services, Bureau of Laboratories | Michigan Department of Health and Human Services, Bureau of Laboratories |
| EPI_ISL_452281 | Michigan Department of Health and Human Services, Bureau of Laboratories | Michigan Department of Health and Human Services, Bureau of Laboratories |
| EPI_ISL_452282 | Michigan Department of Health and Human Services, Bureau of Laboratories | Michigan Department of Health and Human Services, Bureau of Laboratories |
| EPI_ISL_452283 | Michigan Department of Health and Human Services, Bureau of Laboratories | Michigan Department of Health and Human Services, Bureau of Laboratories |

| EPI_Accession_ID | Originating Laboratory | Submitting Laboratory |
|---|---|---|
| EPI_ISL_452284 | Michigan Department of Health and Human Services, Bureau of Laboratories | Michigan Department of Health and Human Services, Bureau of Laboratories |
| EPI_ISL_452285 | Michigan Department of Health and Human Services, Bureau of Laboratories | Michigan Department of Health and Human Services, Bureau of Laboratories |
| EPI_ISL_452286 | Michigan Department of Health and Human Services, Bureau of Laboratories | Michigan Department of Health and Human Services, Bureau of Laboratories |
| EPI_ISL_452287 | Michigan Department of Health and Human Services, Bureau of Laboratories | Michigan Department of Health and Human Services, Bureau of Laboratories |
| EPI_ISL_452288 | Michigan Department of Health and Human Services, Bureau of Laboratories | Michigan Department of Health and Human Services, Bureau of Laboratories |
| EPI_ISL_452289 | Michigan Department of Health and Human Services, Bureau of Laboratories | Michigan Department of Health and Human Services, Bureau of Laboratories |
| EPI_ISL_452290 | Michigan Department of Health and Human Services, Bureau of Laboratories | Michigan Department of Health and Human Services, Bureau of Laboratories |
| EPI_ISL_452291 | Michigan Department of Health and Human Services, Bureau of Laboratories | Michigan Department of Health and Human Services, Bureau of Laboratories |
| EPI_ISL_452292 | Michigan Department of Health and Human Services, Bureau of Laboratories | Michigan Department of Health and Human Services, Bureau of Laboratories |
| EPI_ISL_452293 | Michigan Department of Health and Human Services, Bureau of Laboratories | Michigan Department of Health and Human Services, Bureau of Laboratories |
| EPI_ISL_452294 | Michigan Department of Health and Human Services, Bureau of Laboratories | Michigan Department of Health and Human Services, Bureau of Laboratories |
| EPI_ISL_452295 | Michigan Department of Health and Human Services, Bureau of Laboratories | Michigan Department of Health and Human Services, Bureau of Laboratories |
| EPI_ISL_452296 | Michigan Department of Health and Human Services, Bureau of Laboratories | Michigan Department of Health and Human Services, Bureau of Laboratories |
| EPI_ISL_452297 | Michigan Department of Health and Human Services, Bureau of Laboratories | Michigan Department of Health and Human Services, Bureau of Laboratories |
| EPI_ISL_452298 | Michigan Department of Health and Human Services, Bureau of Laboratories | Michigan Department of Health and Human Services, Bureau of Laboratories |
| EPI_ISL_452299 | Michigan Department of Health and Human Services, Bureau of Laboratories | Michigan Department of Health and Human Services, Bureau of Laboratories |

| EPI_Accession_ID | Originating Laboratory | Submitting Laboratory |
|---|---|---|
| EPI_ISL_452300 | Michigan Department of Health and Human Services, Bureau of Laboratories | Michigan Department of Health and Human Services, Bureau of Laboratories |
| EPI_ISL_452301 | Michigan Department of Health and Human Services, Bureau of Laboratories | Michigan Department of Health and Human Services, Bureau of Laboratories |
| EPI_ISL_452302 | Michigan Department of Health and Human Services, Bureau of Laboratories | Michigan Department of Health and Human Services, Bureau of Laboratories |
| EPI_ISL_452303 | Michigan Department of Health and Human Services, Bureau of Laboratories | Michigan Department of Health and Human Services, Bureau of Laboratories |
| EPI_ISL_452304 | Michigan Department of Health and Human Services, Bureau of Laboratories | Michigan Department of Health and Human Services, Bureau of Laboratories |
| EPI_ISL_452305 | Michigan Department of Health and Human Services, Bureau of Laboratories | Michigan Department of Health and Human Services, Bureau of Laboratories |
| EPI_ISL_452306 | Michigan Department of Health and Human Services, Bureau of Laboratories | Michigan Department of Health and Human Services, Bureau of Laboratories |
| EPI_ISL_452307 | Michigan Department of Health and Human Services, Bureau of Laboratories | Michigan Department of Health and Human Services, Bureau of Laboratories |
| EPI_ISL_452308 | Michigan Department of Health and Human Services, Bureau of Laboratories | Michigan Department of Health and Human Services, Bureau of Laboratories |
| EPI_ISL_452309 | Michigan Department of Health and Human Services, Bureau of Laboratories | Michigan Department of Health and Human Services, Bureau of Laboratories |
| EPI_ISL_452310 | Michigan Department of Health and Human Services, Bureau of Laboratories | Michigan Department of Health and Human Services, Bureau of Laboratories |
| EPI_ISL_452311 | Michigan Department of Health and Human Services, Bureau of Laboratories | Michigan Department of Health and Human Services, Bureau of Laboratories |
| EPI_ISL_452312 | Michigan Department of Health and Human Services, Bureau of Laboratories | Michigan Department of Health and Human Services, Bureau of Laboratories |
| EPI_ISL_452313 | Michigan Department of Health and Human Services, Bureau of Laboratories | Michigan Department of Health and Human Services, Bureau of Laboratories |
| EPI_ISL_452314 | Michigan Department of Health and Human Services, Bureau of Laboratories | Michigan Department of Health and Human Services, Bureau of Laboratories |
| EPI_ISL_452315 | Michigan Department of Health and Human Services, Bureau of Laboratories | Michigan Department of Health and Human Services, Bureau of Laboratories |

| EPI_Accession_ID | Originating Laboratory | Submitting Laboratory |
|---|---|---|
| EPI_ISL_452316 | Michigan Department of Health and Human Services, Bureau of Laboratories | Michigan Department of Health and Human Services, Bureau of Laboratories |
| EPI_ISL_452317 | Michigan Department of Health and Human Services, Bureau of Laboratories | Michigan Department of Health and Human Services, Bureau of Laboratories |
| EPI_ISL_452318 | Michigan Department of Health and Human Services, Bureau of Laboratories | Michigan Department of Health and Human Services, Bureau of Laboratories |
| EPI_ISL_452319 | Michigan Department of Health and Human Services, Bureau of Laboratories | Michigan Department of Health and Human Services, Bureau of Laboratories |
| EPI_ISL_452320 | Michigan Department of Health and Human Services, Bureau of Laboratories | Michigan Department of Health and Human Services, Bureau of Laboratories |
| EPI_ISL_452321 | Michigan Department of Health and Human Services, Bureau of Laboratories | Michigan Department of Health and Human Services, Bureau of Laboratories |
| EPI_ISL_452322 | Michigan Department of Health and Human Services, Bureau of Laboratories | Michigan Department of Health and Human Services, Bureau of Laboratories |
| EPI_ISL_452323 | Michigan Department of Health and Human Services, Bureau of Laboratories | Michigan Department of Health and Human Services, Bureau of Laboratories |
| EPI_ISL_452324 | Michigan Department of Health and Human Services, Bureau of Laboratories | Michigan Department of Health and Human Services, Bureau of Laboratories |
| EPI_ISL_452325 | Michigan Department of Health and Human Services, Bureau of Laboratories | Michigan Department of Health and Human Services, Bureau of Laboratories |
| EPI_ISL_452326 | Michigan Department of Health and Human Services, Bureau of Laboratories | Michigan Department of Health and Human Services, Bureau of Laboratories |
| EPI_ISL_452327 | Laboratory of Infectious Diseases Center of Beijing Ditan Hospital | Laboratory of Infectious Diseases Center of Beijing Ditan Hospital |
| EPI_ISL_452328 | Laboratory of Infectious Diseases Center of Beijing Ditan Hospital | Laboratory of Infectious Diseases Center of Beijing Ditan Hospital |
| EPI_ISL_452329 | Laboratory of Infectious Diseases Center of Beijing Ditan Hospital | Laboratory of Infectious Diseases Center of Beijing Ditan Hospital |
| EPI_ISL_452330 | Laboratory of Infectious Diseases Center of Beijing Ditan Hospital | Laboratory of Infectious Diseases Center of Beijing Ditan Hospital |
| EPI_ISL_452331 | Laboratory of Infectious Diseases Center of Beijing Ditan Hospital | Laboratory of Infectious Diseases Center of Beijing Ditan Hospital |

| EPI_Accession_ID | Originating Laboratory | Submitting Laboratory |
|---|---|---|
| EPI_ISL_452332 | Laboratory of Infectious Diseases Center of Beijing Ditan Hospital | Laboratory of Infectious Diseases Center of Beijing Ditan Hospital |
| EPI_ISL_452333 | Laboratory of Infectious Diseases Center of Beijing Ditan Hospital | Laboratory of Infectious Diseases Center of Beijing Ditan Hospital |
| EPI_ISL_452334 | Laboratory of Infectious Diseases Center of Beijing Ditan Hospital | Laboratory of Infectious Diseases Center of Beijing Ditan Hospital |
| EPI_ISL_452335 | Laboratory of Infectious Diseases Center of Beijing Ditan Hospital | Laboratory of Infectious Diseases Center of Beijing Ditan Hospital |
| EPI_ISL_452336 | Laboratory of Infectious Diseases Center of Beijing Ditan Hospital | Laboratory of Infectious Diseases Center of Beijing Ditan Hospital |
| EPI_ISL_452337 | Laboratory of Infectious Diseases Center of Beijing Ditan Hospital | Laboratory of Infectious Diseases Center of Beijing Ditan Hospital |
| EPI_ISL_452338 | Laboratory of Infectious Diseases Center of Beijing Ditan Hospital | Laboratory of Infectious Diseases Center of Beijing Ditan Hospital |
| EPI_ISL_452339 | Laboratory of Infectious Diseases Center of Beijing Ditan Hospital | Laboratory of Infectious Diseases Center of Beijing Ditan Hospital |
| EPI_ISL_452340 | Laboratory of Infectious Diseases Center of Beijing Ditan Hospital | Laboratory of Infectious Diseases Center of Beijing Ditan Hospital |
| EPI_ISL_452341 | Laboratory of Infectious Diseases Center of Beijing Ditan Hospital | Laboratory of Infectious Diseases Center of Beijing Ditan Hospital |
| EPI_ISL_452342 | Laboratory of Infectious Diseases Center of Beijing Ditan Hospital | Laboratory of Infectious Diseases Center of Beijing Ditan Hospital |
| EPI_ISL_452343 | Laboratory of Infectious Diseases Center of Beijing Ditan Hospital | Laboratory of Infectious Diseases Center of Beijing Ditan Hospital |
| EPI_ISL_452344 | Laboratory of Infectious Diseases Center of Beijing Ditan Hospital | Laboratory of Infectious Diseases Center of Beijing Ditan Hospital |
| EPI_ISL_452345 | Laboratory of Infectious Diseases Center of Beijing Ditan Hospital | Laboratory of Infectious Diseases Center of Beijing Ditan Hospital |
| EPI_ISL_452346 | Laboratory of Infectious Diseases Center of Beijing Ditan Hospital | Laboratory of Infectious Diseases Center of Beijing Ditan Hospital |
| EPI_ISL_452347 | Laboratory of Infectious Diseases Center of Beijing Ditan Hospital | Laboratory of Infectious Diseases Center of Beijing Ditan Hospital |

| EPI_Accession_ID | Originating Laboratory | Submitting Laboratory |
|---|---|---|
| EPI_ISL_452348 | Laboratory of Infectious Diseases Center of Beijing Ditan Hospital | Laboratory of Infectious Diseases Center of Beijing Ditan Hospital |
| EPI_ISL_452349 | Laboratory of Infectious Diseases Center of Beijing Ditan Hospital | Laboratory of Infectious Diseases Center of Beijing Ditan Hospital |
| EPI_ISL_452350 | Laboratory of Infectious Diseases Center of Beijing Ditan Hospital | Laboratory of Infectious Diseases Center of Beijing Ditan Hospital |
| EPI_ISL_452351 | Laboratory of Infectious Diseases Center of Beijing Ditan Hospital | Laboratory of Infectious Diseases Center of Beijing Ditan Hospital |
| EPI_ISL_452352 | Laboratory of Infectious Diseases Center of Beijing Ditan Hospital | Laboratory of Infectious Diseases Center of Beijing Ditan Hospital |
| EPI_ISL_452353 | Laboratory of Infectious Diseases Center of Beijing Ditan Hospital | Laboratory of Infectious Diseases Center of Beijing Ditan Hospital |
| EPI_ISL_452354 | Laboratory of Infectious Diseases Center of Beijing Ditan Hospital | Laboratory of Infectious Diseases Center of Beijing Ditan Hospital |
| EPI_ISL_452355 | Laboratory of Infectious Diseases Center of Beijing Ditan Hospital | Laboratory of Infectious Diseases Center of Beijing Ditan Hospital |
| EPI_ISL_452356 | Laboratory of Infectious Diseases Center of Beijing Ditan Hospital | Laboratory of Infectious Diseases Center of Beijing Ditan Hospital |
| EPI_ISL_452357 | Laboratory of Infectious Diseases Center of Beijing Ditan Hospital | Laboratory of Infectious Diseases Center of Beijing Ditan Hospital |
| EPI_ISL_452358 | Laboratory of Infectious Diseases Center of Beijing Ditan Hospital | Laboratory of Infectious Diseases Center of Beijing Ditan Hospital |
| EPI_ISL_452359 | Laboratory of Infectious Diseases Center of Beijing Ditan Hospital | Laboratory of Infectious Diseases Center of Beijing Ditan Hospital |
| EPI_ISL_452360 | Laboratory of Infectious Diseases Center of Beijing Ditan Hospital | Laboratory of Infectious Diseases Center of Beijing Ditan Hospital |
| EPI_ISL_452361 | Laboratory of Infectious Diseases Center of Beijing Ditan Hospital | Laboratory of Infectious Diseases Center of Beijing Ditan Hospital |
| EPI_ISL_452362 | Laboratory of Infectious Diseases Center of Beijing Ditan Hospital | Laboratory of Infectious Diseases Center of Beijing Ditan Hospital |
| EPI_ISL_452363 | Laboratory of Infectious Diseases Center of Beijing Ditan Hospital | Laboratory of Infectious Diseases Center of Beijing Ditan Hospital |

| EPI_Accession_ID | Originating Laboratory | Submitting Laboratory |
|---|---|---|
| EPI_ISL_452364 | Laboratory of Infectious Diseases Center of Beijing Ditan Hospital | Laboratory of Infectious Diseases Center of Beijing Ditan Hospital |
| EPI_ISL_452365 | Servicio de MicrobiologÃ-a. HRU de MÃ¡laga. Servicio Andaluz de Salud | SeqCOVID-SPAIN consortium/IBV(CSIC) |
| EPI_ISL_452366 | Servicio de MicrobiologÃ-a. HRU de MÃ¡laga. Servicio Andaluz de Salud | SeqCOVID-SPAIN consortium/IBV(CSIC) |
| EPI_ISL_452367 | Servicio de MicrobiologÃ-a. HRU de MÃ¡laga. Servicio Andaluz de Salud | SeqCOVID-SPAIN consortium/IBV(CSIC) |
| EPI_ISL_452368 | Servicio de MicrobiologÃ-a. HRU de MÃ¡laga. Servicio Andaluz de Salud | SeqCOVID-SPAIN consortium/IBV(CSIC) |
| EPI_ISL_452369 | Servicio de MicrobiologÃ-a. HRU de MÃ¡laga. Servicio Andaluz de Salud | SeqCOVID-SPAIN consortium/IBV(CSIC) |
| EPI_ISL_452370 | Servicio de MicrobiologÃ-a. HRU de MÃ¡laga. Servicio Andaluz de Salud | SeqCOVID-SPAIN consortium/IBV(CSIC) |
| EPI_ISL_452371 | Servicio de MicrobiologÃ-a. HRU de MÃ¡laga. Servicio Andaluz de Salud | SeqCOVID-SPAIN consortium/IBV(CSIC) |
| EPI_ISL_452372 | Servicio de MicrobiologÃ-a. HRU de MÃ¡laga. Servicio Andaluz de Salud | SeqCOVID-SPAIN consortium/IBV(CSIC) |
| EPI_ISL_452373 | Servicio de MicrobiologÃ-a. HRU de MÃ¡laga. Servicio Andaluz de Salud | SeqCOVID-SPAIN consortium/IBV(CSIC) |
| EPI_ISL_452374 | Servicio de MicrobiologÃ-a. HRU de MÃ¡laga. Servicio Andaluz de Salud | SeqCOVID-SPAIN consortium/IBV(CSIC) |
| EPI_ISL_452375 | Servicio de MicrobiologÃ-a. HRU de MÃ¡laga. Servicio Andaluz de Salud | SeqCOVID-SPAIN consortium/IBV(CSIC) |
| EPI_ISL_452376 | Servicio de MicrobiologÃ-a. HRU de MÃ¡laga. Servicio Andaluz de Salud | SeqCOVID-SPAIN consortium/IBV(CSIC) |
| EPI_ISL_452377 | Servicio de MicrobiologÃ-a. HRU de MÃ¡laga. Servicio Andaluz de Salud | SeqCOVID-SPAIN consortium/IBV(CSIC) |
| EPI_ISL_452378 | Servicio de MicrobiologÃ-a. HRU de MÃ¡laga. Servicio Andaluz de Salud | SeqCOVID-SPAIN consortium/IBV(CSIC) |
| EPI_ISL_452379 | Servicio de MicrobiologÃ-a. HRU de MÃ¡laga. Servicio Andaluz de Salud | SeqCOVID-SPAIN consortium/IBV(CSIC) |

| EPI_Accession_ID | Originating Laboratory | Submitting Laboratory |
|---|---|---|
| EPI_ISL_452380 | Servicio de MicrobiologÃ-a. HRU de MÃ¡laga. Servicio Andaluz de Salud | SeqCOVID-SPAIN consortium/IBV(CSIC) |
| EPI_ISL_452381 | Servicio de MicrobiologÃ-a. HRU de MÃ¡laga. Servicio Andaluz de Salud | SeqCOVID-SPAIN consortium/IBV(CSIC) |
| EPI_ISL_452382 | Servicio de MicrobiologÃ-a. HRU de MÃ¡laga. Servicio Andaluz de Salud | SeqCOVID-SPAIN consortium/IBV(CSIC) |
| EPI_ISL_452383 | Servicio de MicrobiologÃ-a. HRU de MÃ¡laga. Servicio Andaluz de Salud | SeqCOVID-SPAIN consortium/IBV(CSIC) |
| EPI_ISL_452384 | Servicio de MicrobiologÃ-a. HRU de MÃ¡laga. Servicio Andaluz de Salud | SeqCOVID-SPAIN consortium/IBV(CSIC) |
| EPI_ISL_452385 | Servicio de MicrobiologÃ-a. HRU de MÃ¡laga. Servicio Andaluz de Salud | SeqCOVID-SPAIN consortium/IBV(CSIC) |
| EPI_ISL_452386 | Servicio de MicrobiologÃ-a. HRU de MÃ¡laga. Servicio Andaluz de Salud | SeqCOVID-SPAIN consortium/IBV(CSIC) |
| EPI_ISL_452387 | Servicio de MicrobiologÃ-a. HRU de MÃ¡laga. Servicio Andaluz de Salud | SeqCOVID-SPAIN consortium/IBV(CSIC) |
| EPI_ISL_452388 | Servicio de MicrobiologÃ-a. HRU de MÃ¡laga. Servicio Andaluz de Salud | SeqCOVID-SPAIN consortium/IBV(CSIC) |
| EPI_ISL_452389 | Servicio de MicrobiologÃ-a. HRU de MÃ¡laga. Servicio Andaluz de Salud | SeqCOVID-SPAIN consortium/IBV(CSIC) |
| EPI_ISL_452390 | Servicio de MicrobiologÃ-a. HRU de MÃ¡laga. Servicio Andaluz de Salud | SeqCOVID-SPAIN consortium/IBV(CSIC) |
| EPI_ISL_452391 | Servicio de MicrobiologÃ-a. HRU de MÃ¡laga. Servicio Andaluz de Salud | SeqCOVID-SPAIN consortium/IBV(CSIC) |
| EPI_ISL_452392 | Servicio de MicrobiologÃ-a. HRU de MÃ¡laga. Servicio Andaluz de Salud | SeqCOVID-SPAIN consortium/IBV(CSIC) |
| EPI_ISL_452393 | Servicio de MicrobiologÃ-a. HRU de MÃ¡laga. Servicio Andaluz de Salud | SeqCOVID-SPAIN consortium/IBV(CSIC) |
| EPI_ISL_452394 | Servicio de MicrobiologÃ-a. HRU de MÃ¡laga. Servicio Andaluz de Salud | SeqCOVID-SPAIN consortium/IBV(CSIC) |
| EPI_ISL_452395 | Servicio de MicrobiologÃ-a. HRU de MÃ¡laga. Servicio Andaluz de Salud | SeqCOVID-SPAIN consortium/IBV(CSIC) |

| EPI_Accession_ID | Originating Laboratory | Submitting Laboratory |
|---|---|---|
| EPI_ISL_452396 | Servicio de MicrobiologÃ-a. HRU de MÃ¡laga. Servicio Andaluz de Salud | SeqCOVID-SPAIN consortium/IBV(CSIC) |
| EPI_ISL_452397 | Servicio de MicrobiologÃ-a. HRU de MÃ¡laga. Servicio Andaluz de Salud | SeqCOVID-SPAIN consortium/IBV(CSIC) |
| EPI_ISL_452398 | Servicio de MicrobiologÃ-a. HRU de MÃ¡laga. Servicio Andaluz de Salud | SeqCOVID-SPAIN consortium/IBV(CSIC) |
| EPI_ISL_452399 | Servicio de MicrobiologÃ-a. HRU de MÃ¡laga. Servicio Andaluz de Salud | SeqCOVID-SPAIN consortium/IBV(CSIC) |
| EPI_ISL_452400 | Servicio de MicrobiologÃ-a. HRU de MÃ¡laga. Servicio Andaluz de Salud | SeqCOVID-SPAIN consortium/IBV(CSIC) |
| EPI_ISL_452401 | Servicio de MicrobiologÃ-a. HRU de MÃ¡laga. Servicio Andaluz de Salud | SeqCOVID-SPAIN consortium/IBV(CSIC) |
| EPI_ISL_452402 | Servicio de MicrobiologÃ-a. HRU de MÃ¡laga. Servicio Andaluz de Salud | SeqCOVID-SPAIN consortium/IBV(CSIC) |
| EPI_ISL_452403 | Servicio de MicrobiologÃ-a. HRU de MÃ¡laga. Servicio Andaluz de Salud | SeqCOVID-SPAIN consortium/IBV(CSIC) |
| EPI_ISL_452404 | Servicio de MicrobiologÃ-a. HRU de MÃ¡laga. Servicio Andaluz de Salud | SeqCOVID-SPAIN consortium/IBV(CSIC) |
| EPI_ISL_452405 | Servicio de MicrobiologÃ-a. HRU de MÃ¡laga. Servicio Andaluz de Salud | SeqCOVID-SPAIN consortium/IBV(CSIC) |
| EPI_ISL_452406 | Servicio de MicrobiologÃ-a. HRU de MÃ¡laga. Servicio Andaluz de Salud | SeqCOVID-SPAIN consortium/IBV(CSIC) |
| EPI_ISL_452407 | Servicio de MicrobiologÃ-a. HRU de MÃ¡laga. Servicio Andaluz de Salud | SeqCOVID-SPAIN consortium/IBV(CSIC) |
| EPI_ISL_452408 | Servicio de MicrobiologÃ-a. HRU de MÃ¡laga. Servicio Andaluz de Salud | SeqCOVID-SPAIN consortium/IBV(CSIC) |
| EPI_ISL_452409 | Servicio de MicrobiologÃ-a. HRU de MÃ¡laga. Servicio Andaluz de Salud | SeqCOVID-SPAIN consortium/IBV(CSIC) |
| EPI_ISL_452410 | Servicio de MicrobiologÃ-a. HRU de MÃ¡laga. Servicio Andaluz de Salud | SeqCOVID-SPAIN consortium/IBV(CSIC) |
| EPI_ISL_452411 | Servicio de MicrobiologÃ-a. HRU de MÃ¡laga. Servicio Andaluz de Salud | SeqCOVID-SPAIN consortium/IBV(CSIC) |

| EPI_Accession_ID | Originating Laboratory | Submitting Laboratory |
|---|---|---|
| EPI_ISL_452412 | Servicio de MicrobiologÃ-a. HRU de MÃ¡laga. Servicio Andaluz de Salud | SeqCOVID-SPAIN consortium/IBV(CSIC) |
| EPI_ISL_452413 | Servicio de MicrobiologÃ-a. HRU de MÃ¡laga. Servicio Andaluz de Salud | SeqCOVID-SPAIN consortium/IBV(CSIC) |
| EPI_ISL_452414 | Servicio de MicrobiologÃ-a. HRU de MÃ¡laga. Servicio Andaluz de Salud | SeqCOVID-SPAIN consortium/IBV(CSIC) |
| EPI_ISL_452415 | Servicio de MicrobiologÃ-a. HRU de MÃ¡laga. Servicio Andaluz de Salud | SeqCOVID-SPAIN consortium/IBV(CSIC) |
| EPI_ISL_452416 | Servicio de MicrobiologÃ-a. HRU de MÃ¡laga. Servicio Andaluz de Salud | SeqCOVID-SPAIN consortium/IBV(CSIC) |
| EPI_ISL_452417 | Servicio de MicrobiologÃ-a. HRU de MÃ¡laga. Servicio Andaluz de Salud | SeqCOVID-SPAIN consortium/IBV(CSIC) |
| EPI_ISL_452418 | Servicio de MicrobiologÃ-a. HRU de MÃ¡laga. Servicio Andaluz de Salud | SeqCOVID-SPAIN consortium/IBV(CSIC) |
| EPI_ISL_452419 | Servicio de MicrobiologÃ-a. HRU de MÃ¡laga. Servicio Andaluz de Salud | SeqCOVID-SPAIN consortium/IBV(CSIC) |
| EPI_ISL_452420 | Servicio de MicrobiologÃ-a. HRU de MÃ¡laga. Servicio Andaluz de Salud | SeqCOVID-SPAIN consortium/IBV(CSIC) |
| EPI_ISL_452421 | Servicio de MicrobiologÃ-a. HRU de MÃ¡laga. Servicio Andaluz de Salud | SeqCOVID-SPAIN consortium/IBV(CSIC) |
| EPI_ISL_452422 | Servicio de MicrobiologÃ-a. HRU de MÃ¡laga. Servicio Andaluz de Salud | SeqCOVID-SPAIN consortium/IBV(CSIC) |
| EPI_ISL_452423 | Servicio de MicrobiologÃ-a. HRU de MÃ¡laga. Servicio Andaluz de Salud | SeqCOVID-SPAIN consortium/IBV(CSIC) |
| EPI_ISL_452424 | Servicio de MicrobiologÃ-a. HRU de MÃ¡laga. Servicio Andaluz de Salud | SeqCOVID-SPAIN consortium/IBV(CSIC) |
| EPI_ISL_452425 | Servicio de MicrobiologÃ-a. HRU de MÃ¡laga. Servicio Andaluz de Salud | SeqCOVID-SPAIN consortium/IBV(CSIC) |
| EPI_ISL_452426 | Servicio de MicrobiologÃ-a. HRU de MÃ¡laga. Servicio Andaluz de Salud | SeqCOVID-SPAIN consortium/IBV(CSIC) |
| EPI_ISL_452427 | Servicio de MicrobiologÃ-a. HRU de MÃ¡laga. Servicio Andaluz de Salud | SeqCOVID-SPAIN consortium/IBV(CSIC) |

| EPI_Accession_ID | Originating Laboratory | Submitting Laboratory |
|---|---|---|
| EPI_ISL_452428 | Servicio de MicrobiologÃ-a. HRU de MÃ¡laga. Servicio Andaluz de Salud | SeqCOVID-SPAIN consortium/IBV(CSIC) |
| EPI_ISL_452429 | Servicio de MicrobiologÃ-a. HRU de MÃ¡laga. Servicio Andaluz de Salud | SeqCOVID-SPAIN consortium/IBV(CSIC) |
| EPI_ISL_452430 | Servicio de MicrobiologÃ-a. HRU de MÃ¡laga. Servicio Andaluz de Salud | SeqCOVID-SPAIN consortium/IBV(CSIC) |
| EPI_ISL_452431 | Servicio de MicrobiologÃ-a. HRU de MÃ¡laga. Servicio Andaluz de Salud | SeqCOVID-SPAIN consortium/IBV(CSIC) |
| EPI_ISL_452432 | Servicio de MicrobiologÃ-a. HRU de MÃ¡laga. Servicio Andaluz de Salud | SeqCOVID-SPAIN consortium/IBV(CSIC) |
| EPI_ISL_452433 | Servicio de MicrobiologÃ-a. HRU de MÃ¡laga. Servicio Andaluz de Salud | SeqCOVID-SPAIN consortium/IBV(CSIC) |
| EPI_ISL_452434 | Servicio de MicrobiologÃ-a. HRU de MÃ¡laga. Servicio Andaluz de Salud | SeqCOVID-SPAIN consortium/IBV(CSIC) |
| EPI_ISL_452435 | Servicio de MicrobiologÃ-a. HRU de MÃ¡laga. Servicio Andaluz de Salud | SeqCOVID-SPAIN consortium/IBV(CSIC) |
| EPI_ISL_452436 | Servicio de MicrobiologÃ-a. HRU de MÃ¡laga. Servicio Andaluz de Salud | SeqCOVID-SPAIN consortium/IBV(CSIC) |
| EPI_ISL_452437 | Servicio de MicrobiologÃ-a. HRU de MÃ¡laga. Servicio Andaluz de Salud | SeqCOVID-SPAIN consortium/IBV(CSIC) |
| EPI_ISL_452438 | Servicio de MicrobiologÃ-a. HRU de MÃ¡laga. Servicio Andaluz de Salud | SeqCOVID-SPAIN consortium/IBV(CSIC) |
| EPI_ISL_452439 | Servicio de MicrobiologÃ-a. HRU de MÃ¡laga. Servicio Andaluz de Salud | SeqCOVID-SPAIN consortium/IBV(CSIC) |
| EPI_ISL_452440 | Servicio de MicrobiologÃ-a. HRU de MÃ¡laga. Servicio Andaluz de Salud | SeqCOVID-SPAIN consortium/IBV(CSIC) |
| EPI_ISL_452441 | Servicio de MicrobiologÃ-a. HRU de MÃ¡laga. Servicio Andaluz de Salud | SeqCOVID-SPAIN consortium/IBV(CSIC) |
| EPI_ISL_452442 | Servicio de MicrobiologÃ-a. HRU de MÃ¡laga. Servicio Andaluz de Salud | SeqCOVID-SPAIN consortium/IBV(CSIC) |
| EPI_ISL_452443 | Servicio de MicrobiologÃ-a. HRU de MÃ¡laga. Servicio Andaluz de Salud | SeqCOVID-SPAIN consortium/IBV(CSIC) |

| EPI_Accession_ID | Originating Laboratory | Submitting Laboratory |
|---|---|---|
| EPI_ISL_452444 | Servicio de MicrobiologÃ-a. HRU de MÃ¡laga. Servicio Andaluz de Salud | SeqCOVID-SPAIN consortium/IBV(CSIC) |
| EPI_ISL_452445 | Servicio de MicrobiologÃ-a. HRU de MÃ¡laga. Servicio Andaluz de Salud | SeqCOVID-SPAIN consortium/IBV(CSIC) |
| EPI_ISL_452446 | Servicio de MicrobiologÃ-a. HRU de MÃ¡laga. Servicio Andaluz de Salud | SeqCOVID-SPAIN consortium/IBV(CSIC) |
| EPI_ISL_452447 | Servicio de MicrobiologÃ-a. HRU de MÃ¡laga. Servicio Andaluz de Salud | SeqCOVID-SPAIN consortium/IBV(CSIC) |
| EPI_ISL_452448 | Servicio de MicrobiologÃ-a. HRU de MÃ¡laga. Servicio Andaluz de Salud | SeqCOVID-SPAIN consortium/IBV(CSIC) |
| EPI_ISL_452449 | Servicio de MicrobiologÃ-a. HRU de MÃ¡laga. Servicio Andaluz de Salud | SeqCOVID-SPAIN consortium/IBV(CSIC) |
| EPI_ISL_452450 | Servicio de MicrobiologÃ-a. HRU de MÃ¡laga. Servicio Andaluz de Salud | SeqCOVID-SPAIN consortium/IBV(CSIC) |
| EPI_ISL_452451 | Servicio de MicrobiologÃ-a. HRU de MÃ¡laga. Servicio Andaluz de Salud | SeqCOVID-SPAIN consortium/IBV(CSIC) |
| EPI_ISL_452452 | Servicio de MicrobiologÃ-a. HRU de MÃ¡laga. Servicio Andaluz de Salud | SeqCOVID-SPAIN consortium/IBV(CSIC) |
| EPI_ISL_452453 | Hospital Universitario Puerta del Mar de CÃ¡diz - INIBICA | SeqCOVID-SPAIN consortium/IBV(CSIC) |
| EPI_ISL_452454 | Hospital Universitario Puerta del Mar de CÃ¡diz - INIBICA | SeqCOVID-SPAIN consortium/IBV(CSIC) |
| EPI_ISL_452455 | Hospital Universitario Puerta del Mar de CÃ¡diz - INIBICA | SeqCOVID-SPAIN consortium/IBV(CSIC) |
| EPI_ISL_452456 | Hospital Universitario Puerta del Mar de CÃ¡diz - INIBICA | SeqCOVID-SPAIN consortium/IBV(CSIC) |
| EPI_ISL_452457 | Hospital Universitario Puerta del Mar de CÃ¡diz - INIBICA | SeqCOVID-SPAIN consortium/IBV(CSIC) |
| EPI_ISL_452458 | Hospital Universitario Puerta del Mar de CÃ¡diz - INIBICA | SeqCOVID-SPAIN consortium/IBV(CSIC) |
| EPI_ISL_452459 | Hospital Universitario Puerta del Mar de CÃ¡diz - INIBICA | SeqCOVID-SPAIN consortium/IBV(CSIC) |

| EPI_Accession_ID | Originating Laboratory | Submitting Laboratory |
|---|---|---|
| EPI_ISL_452460 | Hospital Universitario Puerta del Mar de CÃ¡diz - INIBICA | SeqCOVID-SPAIN consortium/IBV(CSIC) |
| EPI_ISL_452461 | Hospital Universitario Puerta del Mar de CÃ¡diz - INIBICA | SeqCOVID-SPAIN consortium/IBV(CSIC) |
| EPI_ISL_452462 | Hospital Universitario Puerta del Mar de CÃ¡diz - INIBICA | SeqCOVID-SPAIN consortium/IBV(CSIC) |
| EPI_ISL_452463 | Hospital Universitario Puerta del Mar de CÃ¡diz - INIBICA | SeqCOVID-SPAIN consortium/IBV(CSIC) |
| EPI_ISL_452464 | Hospital Universitario Puerta del Mar de CÃ¡diz - INIBICA | SeqCOVID-SPAIN consortium/IBV(CSIC) |
| EPI_ISL_452465 | Hospital Universitario Puerta del Mar de CÃ¡diz - INIBICA | SeqCOVID-SPAIN consortium/IBV(CSIC) |
| EPI_ISL_452466 | Hospital Universitario Puerta del Mar de CÃ¡diz - INIBICA | SeqCOVID-SPAIN consortium/IBV(CSIC) |
| EPI_ISL_452467 | Hospital Universitario Puerta del Mar de CÃ¡diz - INIBICA | SeqCOVID-SPAIN consortium/IBV(CSIC) |
| EPI_ISL_452468 | Hospital Universitario Puerta del Mar de CÃ¡diz - INIBICA | SeqCOVID-SPAIN consortium/IBV(CSIC) |
| EPI_ISL_452469 | Hospital Universitario Puerta del Mar de CÃ¡diz - INIBICA | SeqCOVID-SPAIN consortium/IBV(CSIC) |
| EPI_ISL_452470 | Hospital Universitario Puerta del Mar de CÃ¡diz - INIBICA | SeqCOVID-SPAIN consortium/IBV(CSIC) |
| EPI_ISL_452471 | Hospital Universitario Puerta del Mar de CÃ¡diz - INIBICA | SeqCOVID-SPAIN consortium/IBV(CSIC) |
| EPI_ISL_452472 | ClÃ-nica Universidad de Navarra. Servicio de Enfermedades Infecciosas y MicrobiologÃ-a clÃ-nica | SeqCOVID-SPAIN consortium/IBV(CSIC) |
| EPI_ISL_452473 | ClÃ-nica Universidad de Navarra. Servicio de Enfermedades Infecciosas y MicrobiologÃ-a clÃ-nica | SeqCOVID-SPAIN consortium/IBV(CSIC) |
| EPI_ISL_452474 | ClÃ-nica Universidad de Navarra. Servicio de Enfermedades Infecciosas y MicrobiologÃ-a clÃ-nica | SeqCOVID-SPAIN consortium/IBV(CSIC) |
| EPI_ISL_452475 | ClÃ-nica Universidad de Navarra. Servicio de Enfermedades Infecciosas y MicrobiologÃ-a clÃ-nica | SeqCOVID-SPAIN consortium/IBV(CSIC) |

| EPI_Accession_ID | Originating Laboratory | Submitting Laboratory |
|---|---|---|
| EPI_ISL_452476 | ClÃ-nica Universidad de Navarra. Servicio de Enfermedades Infecciosas y MicrobiologÃ-a clÃ-nica | SeqCOVID-SPAIN consortium/IBV(CSIC) |
| EPI_ISL_452477 | ClÃ-nica Universidad de Navarra. Servicio de Enfermedades Infecciosas y MicrobiologÃ-a clÃ-nica | SeqCOVID-SPAIN consortium/IBV(CSIC) |
| EPI_ISL_452478 | ClÃ-nica Universidad de Navarra. Servicio de Enfermedades Infecciosas y MicrobiologÃ-a clÃ-nica | SeqCOVID-SPAIN consortium/IBV(CSIC) |
| EPI_ISL_452479 | ClÃ-nica Universidad de Navarra. Servicio de Enfermedades Infecciosas y MicrobiologÃ-a clÃ-nica | SeqCOVID-SPAIN consortium/IBV(CSIC) |
| EPI_ISL_452480 | ClÃ-nica Universidad de Navarra. Servicio de Enfermedades Infecciosas y MicrobiologÃ-a clÃ-nica | SeqCOVID-SPAIN consortium/IBV(CSIC) |
| EPI_ISL_452481 | ClÃ-nica Universidad de Navarra. Servicio de Enfermedades Infecciosas y MicrobiologÃ-a clÃ-nica | SeqCOVID-SPAIN consortium/IBV(CSIC) |
| EPI_ISL_452482 | ClÃ-nica Universidad de Navarra. Servicio de Enfermedades Infecciosas y MicrobiologÃ-a clÃ-nica | SeqCOVID-SPAIN consortium/IBV(CSIC) |
| EPI_ISL_452483 | ClÃ-nica Universidad de Navarra. Servicio de Enfermedades Infecciosas y MicrobiologÃ-a clÃ-nica | SeqCOVID-SPAIN consortium/IBV(CSIC) |
| EPI_ISL_452484 | ClÃ-nica Universidad de Navarra. Servicio de Enfermedades Infecciosas y MicrobiologÃ-a clÃ-nica | SeqCOVID-SPAIN consortium/IBV(CSIC) |
| EPI_ISL_452485 | ClÃ-nica Universidad de Navarra. Servicio de Enfermedades Infecciosas y MicrobiologÃ-a clÃ-nica | SeqCOVID-SPAIN consortium/IBV(CSIC) |
| EPI_ISL_452486 | ClÃ-nica Universidad de Navarra. Servicio de Enfermedades Infecciosas y MicrobiologÃ-a clÃ-nica | SeqCOVID-SPAIN consortium/IBV(CSIC) |
| EPI_ISL_452487 | ClÃ-nica Universidad de Navarra. Servicio de Enfermedades Infecciosas y MicrobiologÃ-a clÃ-nica | SeqCOVID-SPAIN consortium/IBV(CSIC) |
| EPI_ISL_452488 | ClÃ-nica Universidad de Navarra. Servicio de Enfermedades Infecciosas y MicrobiologÃ-a clÃ-nica | SeqCOVID-SPAIN consortium/IBV(CSIC) |
| EPI_ISL_452489 | ClÃ-nica Universidad de Navarra. Servicio de Enfermedades Infecciosas y MicrobiologÃ-a clÃ-nica | SeqCOVID-SPAIN consortium/IBV(CSIC) |
| EPI_ISL_452490 | ClÃ-nica Universidad de Navarra. Servicio de Enfermedades Infecciosas y MicrobiologÃ-a clÃ-nica | SeqCOVID-SPAIN consortium/IBV(CSIC) |
| EPI_ISL_452491 | ClÃ-nica Universidad de Navarra. Servicio de Enfermedades Infecciosas y MicrobiologÃ-a clÃ-nica | SeqCOVID-SPAIN consortium/IBV(CSIC) |

| EPI_Accession_ID | Originating Laboratory | Submitting Laboratory |
|---|---|---|
| EPI_ISL_452492 | ClÃ-nica Universidad de Navarra. Servicio de Enfermedades Infecciosas y MicrobiologÃ-a clÃ-nica | SeqCOVID-SPAIN consortium/IBV(CSIC) |
| EPI_ISL_452493 | ClÃ-nica Universidad de Navarra. Servicio de Enfermedades Infecciosas y MicrobiologÃ-a clÃ-nica | SeqCOVID-SPAIN consortium/IBV(CSIC) |
| EPI_ISL_452494 | ClÃ-nica Universidad de Navarra. Servicio de Enfermedades Infecciosas y MicrobiologÃ-a clÃ-nica | SeqCOVID-SPAIN consortium/IBV(CSIC) |
| EPI_ISL_452495 | ClÃ-nica Universidad de Navarra. Servicio de Enfermedades Infecciosas y MicrobiologÃ-a clÃ-nica | SeqCOVID-SPAIN consortium/IBV(CSIC) |
| EPI_ISL_452496 | ClÃ-nica Universidad de Navarra. Servicio de Enfermedades Infecciosas y MicrobiologÃ-a clÃ-nica | SeqCOVID-SPAIN consortium/IBV(CSIC) |
| EPI_ISL_452497 | ClÃ-nica Universidad de Navarra. Servicio de Enfermedades Infecciosas y MicrobiologÃ-a clÃ-nica | SeqCOVID-SPAIN consortium/IBV(CSIC) |
| EPI_ISL_452498 | ClÃ-nica Universidad de Navarra. Servicio de Enfermedades Infecciosas y MicrobiologÃ-a clÃ-nica | SeqCOVID-SPAIN consortium/IBV(CSIC) |
| EPI_ISL_452499 | ClÃ-nica Universidad de Navarra. Servicio de Enfermedades Infecciosas y MicrobiologÃ-a clÃ-nica | SeqCOVID-SPAIN consortium/IBV(CSIC) |
| EPI_ISL_452500 | ClÃ-nica Universidad de Navarra. Servicio de Enfermedades Infecciosas y MicrobiologÃ-a clÃ-nica | SeqCOVID-SPAIN consortium/IBV(CSIC) |
| EPI_ISL_452501 | ClÃ-nica Universidad de Navarra. Servicio de Enfermedades Infecciosas y MicrobiologÃ-a clÃ-nica | SeqCOVID-SPAIN consortium/IBV(CSIC) |
| EPI_ISL_452502 | ClÃ-nica Universidad de Navarra. Servicio de Enfermedades Infecciosas y MicrobiologÃ-a clÃ-nica | SeqCOVID-SPAIN consortium/IBV(CSIC) |
| EPI_ISL_452503 | ClÃ-nica Universidad de Navarra. Servicio de Enfermedades Infecciosas y MicrobiologÃ-a clÃ-nica | SeqCOVID-SPAIN consortium/IBV(CSIC) |
| EPI_ISL_452504 | ClÃ-nica Universidad de Navarra. Servicio de Enfermedades Infecciosas y MicrobiologÃ-a clÃ-nica | SeqCOVID-SPAIN consortium/IBV(CSIC) |
| EPI_ISL_452505 | ClÃ-nica Universidad de Navarra. Servicio de Enfermedades Infecciosas y MicrobiologÃ-a clÃ-nica | SeqCOVID-SPAIN consortium/IBV(CSIC) |
| EPI_ISL_452506 | ClÃ-nica Universidad de Navarra. Servicio de Enfermedades Infecciosas y MicrobiologÃ-a clÃ-nica | SeqCOVID-SPAIN consortium/IBV(CSIC) |
| EPI_ISL_452507 | ClÃ-nica Universidad de Navarra. Servicio de Enfermedades Infecciosas y MicrobiologÃ-a clÃ-nica | SeqCOVID-SPAIN consortium/IBV(CSIC) |

| EPI_Accession_ID | Originating Laboratory | Submitting Laboratory |
|---|---|---|
| EPI_ISL_452508 | ClÃ-nica Universidad de Navarra. Servicio de Enfermedades Infecciosas y MicrobiologÃ-a clÃ-nica | SeqCOVID-SPAIN consortium/IBV(CSIC) |
| EPI_ISL_452509 | ClÃ-nica Universidad de Navarra. Servicio de Enfermedades Infecciosas y MicrobiologÃ-a clÃ-nica | SeqCOVID-SPAIN consortium/IBV(CSIC) |
| EPI_ISL_452510 | ClÃ-nica Universidad de Navarra. Servicio de Enfermedades Infecciosas y MicrobiologÃ-a clÃ-nica | SeqCOVID-SPAIN consortium/IBV(CSIC) |
| EPI_ISL_452511 | ClÃ-nica Universidad de Navarra. Servicio de Enfermedades Infecciosas y MicrobiologÃ-a clÃ-nica | SeqCOVID-SPAIN consortium/IBV(CSIC) |
| EPI_ISL_452512 | ClÃ-nica Universidad de Navarra. Servicio de Enfermedades Infecciosas y MicrobiologÃ-a clÃ-nica | SeqCOVID-SPAIN consortium/IBV(CSIC) |
| EPI_ISL_452513 | ClÃ-nica Universidad de Navarra. Servicio de Enfermedades Infecciosas y MicrobiologÃ-a clÃ-nica | SeqCOVID-SPAIN consortium/IBV(CSIC) |
| EPI_ISL_452514 | ClÃ-nica Universidad de Navarra. Servicio de Enfermedades Infecciosas y MicrobiologÃ-a clÃ-nica | SeqCOVID-SPAIN consortium/IBV(CSIC) |
| EPI_ISL_452515 | ClÃ-nica Universidad de Navarra. Servicio de Enfermedades Infecciosas y MicrobiologÃ-a clÃ-nica | SeqCOVID-SPAIN consortium/IBV(CSIC) |
| EPI_ISL_452516 | ClÃ-nica Universidad de Navarra. Servicio de Enfermedades Infecciosas y MicrobiologÃ-a clÃ-nica | SeqCOVID-SPAIN consortium/IBV(CSIC) |
| EPI_ISL_452517 | ClÃ-nica Universidad de Navarra. Servicio de Enfermedades Infecciosas y MicrobiologÃ-a clÃ-nica | SeqCOVID-SPAIN consortium/IBV(CSIC) |
| EPI_ISL_452518 | ClÃ-nica Universidad de Navarra. Servicio de Enfermedades Infecciosas y MicrobiologÃ-a clÃ-nica | SeqCOVID-SPAIN consortium/IBV(CSIC) |
| EPI_ISL_452519 | ClÃ-nica Universidad de Navarra. Servicio de Enfermedades Infecciosas y MicrobiologÃ-a clÃ-nica | SeqCOVID-SPAIN consortium/IBV(CSIC) |
| EPI_ISL_452520 | ClÃ-nica Universidad de Navarra. Servicio de Enfermedades Infecciosas y MicrobiologÃ-a clÃ-nica | SeqCOVID-SPAIN consortium/IBV(CSIC) |
| EPI_ISL_452521 | ClÃ-nica Universidad de Navarra. Servicio de Enfermedades Infecciosas y MicrobiologÃ-a clÃ-nica | SeqCOVID-SPAIN consortium/IBV(CSIC) |
| EPI_ISL_452522 | ClÃ-nica Universidad de Navarra. Servicio de Enfermedades Infecciosas y MicrobiologÃ-a clÃ-nica | SeqCOVID-SPAIN consortium/IBV(CSIC) |
| EPI_ISL_452523 | ClÃ-nica Universidad de Navarra. Servicio de Enfermedades Infecciosas y MicrobiologÃ-a clÃ-nica | SeqCOVID-SPAIN consortium/IBV(CSIC) |

| EPI_Accession_ID | Originating Laboratory | Submitting Laboratory |
|---|---|---|
| EPI_ISL_452524 | ClÃ-nica Universidad de Navarra. Servicio de Enfermedades Infecciosas y MicrobiologÃ-a clÃ-nica | SeqCOVID-SPAIN consortium/IBV(CSIC) |
| EPI_ISL_452525 | ClÃ-nica Universidad de Navarra. Servicio de Enfermedades Infecciosas y MicrobiologÃ-a clÃ-nica | SeqCOVID-SPAIN consortium/IBV(CSIC) |
| EPI_ISL_452526 | ClÃ-nica Universidad de Navarra. Servicio de Enfermedades Infecciosas y MicrobiologÃ-a clÃ-nica | SeqCOVID-SPAIN consortium/IBV(CSIC) |
| EPI_ISL_452527 | ClÃ-nica Universidad de Navarra. Servicio de Enfermedades Infecciosas y MicrobiologÃ-a clÃ-nica | SeqCOVID-SPAIN consortium/IBV(CSIC) |
| EPI_ISL_452528 | ClÃ-nica Universidad de Navarra. Servicio de Enfermedades Infecciosas y MicrobiologÃ-a clÃ-nica | SeqCOVID-SPAIN consortium/IBV(CSIC) |
| EPI_ISL_452529 | ClÃ-nica Universidad de Navarra. Servicio de Enfermedades Infecciosas y MicrobiologÃ-a clÃ-nica | SeqCOVID-SPAIN consortium/IBV(CSIC) |
| EPI_ISL_452530 | ClÃ-nica Universidad de Navarra. Servicio de Enfermedades Infecciosas y MicrobiologÃ-a clÃ-nica | SeqCOVID-SPAIN consortium/IBV(CSIC) |
| EPI_ISL_452531 | ClÃ-nica Universidad de Navarra. Servicio de Enfermedades Infecciosas y MicrobiologÃ-a clÃ-nica | SeqCOVID-SPAIN consortium/IBV(CSIC) |
| EPI_ISL_452532 | ClÃ-nica Universidad de Navarra. Servicio de Enfermedades Infecciosas y MicrobiologÃ-a clÃ-nica | SeqCOVID-SPAIN consortium/IBV(CSIC) |
| EPI_ISL_452533 | ClÃ-nica Universidad de Navarra. Servicio de Enfermedades Infecciosas y MicrobiologÃ-a clÃ-nica | SeqCOVID-SPAIN consortium/IBV(CSIC) |
| EPI_ISL_452534 | ClÃ-nica Universidad de Navarra. Servicio de Enfermedades Infecciosas y MicrobiologÃ-a clÃ-nica | SeqCOVID-SPAIN consortium/IBV(CSIC) |
| EPI_ISL_452535 | ClÃ-nica Universidad de Navarra. Servicio de Enfermedades Infecciosas y MicrobiologÃ-a clÃ-nica | SeqCOVID-SPAIN consortium/IBV(CSIC) |
| EPI_ISL_452536 | ClÃ-nica Universidad de Navarra. Servicio de Enfermedades Infecciosas y MicrobiologÃ-a clÃ-nica | SeqCOVID-SPAIN consortium/IBV(CSIC) |
| EPI_ISL_452537 | ClÃ-nica Universidad de Navarra. Servicio de Enfermedades Infecciosas y MicrobiologÃ-a clÃ-nica | SeqCOVID-SPAIN consortium/IBV(CSIC) |
| EPI_ISL_452538 | ClÃ-nica Universidad de Navarra. Servicio de Enfermedades Infecciosas y MicrobiologÃ-a clÃ-nica | SeqCOVID-SPAIN consortium/IBV(CSIC) |
| EPI_ISL_452539 | ClÃ-nica Universidad de Navarra. Servicio de Enfermedades Infecciosas y MicrobiologÃ-a clÃ-nica | SeqCOVID-SPAIN consortium/IBV(CSIC) |

| EPI_Accession_ID | Originating Laboratory | Submitting Laboratory |
|---|---|---|
| EPI_ISL_452540 | ClÃ-nica Universidad de Navarra. Servicio de Enfermedades Infecciosas y MicrobiologÃ-a clÃ-nica | SeqCOVID-SPAIN consortium/IBV(CSIC) |
| EPI_ISL_452541 | ClÃ-nica Universidad de Navarra. Servicio de Enfermedades Infecciosas y MicrobiologÃ-a clÃ-nica | SeqCOVID-SPAIN consortium/IBV(CSIC) |
| EPI_ISL_452542 | ClÃ-nica Universidad de Navarra. Servicio de Enfermedades Infecciosas y MicrobiologÃ-a clÃ-nica | SeqCOVID-SPAIN consortium/IBV(CSIC) |
| EPI_ISL_452543 | ClÃ-nica Universidad de Navarra. Servicio de Enfermedades Infecciosas y MicrobiologÃ-a clÃ-nica | SeqCOVID-SPAIN consortium/IBV(CSIC) |
| EPI_ISL_452544 | Servicio de MicrobiologÃ-a y ParasitologÃ-a clÃ-nica. UCEIMP. Hospital Universitario Virgen del RocÃ-o/IBIS/CSIC/US. | SeqCOVID-SPAIN consortium/IBV(CSIC) |
| EPI_ISL_452545 | Servicio de MicrobiologÃ-a y ParasitologÃ-a clÃ-nica. UCEIMP. Hospital Universitario Virgen del RocÃ-o/IBIS/CSIC/US. | SeqCOVID-SPAIN consortium/IBV(CSIC) |
| EPI_ISL_452546 | Servicio de MicrobiologÃ-a y ParasitologÃ-a clÃ-nica. UCEIMP. Hospital Universitario Virgen del RocÃ-o/IBIS/CSIC/US. | SeqCOVID-SPAIN consortium/IBV(CSIC) |
| EPI_ISL_452547 | Servicio de MicrobiologÃ-a y ParasitologÃ-a clÃ-nica. UCEIMP. Hospital Universitario Virgen del RocÃ-o/IBIS/CSIC/US. | SeqCOVID-SPAIN consortium/IBV(CSIC) |
| EPI_ISL_452548 | Servicio de MicrobiologÃ-a y ParasitologÃ-a clÃ-nica. UCEIMP. Hospital Universitario Virgen del RocÃ-o/IBIS/CSIC/US. | SeqCOVID-SPAIN consortium/IBV(CSIC) |
| EPI_ISL_452549 | Servicio de MicrobiologÃ-a y ParasitologÃ-a clÃ-nica. UCEIMP. Hospital Universitario Virgen del RocÃ-o/IBIS/CSIC/US. | SeqCOVID-SPAIN consortium/IBV(CSIC) |
| EPI_ISL_452550 | Servicio de MicrobiologÃ-a y ParasitologÃ-a clÃ-nica. UCEIMP. Hospital Universitario Virgen del RocÃ-o/IBIS/CSIC/US. | SeqCOVID-SPAIN consortium/IBV(CSIC) |
| EPI_ISL_452551 | Servicio de MicrobiologÃ-a y ParasitologÃ-a clÃ-nica. UCEIMP. Hospital Universitario Virgen del RocÃ-o/IBIS/CSIC/US. | SeqCOVID-SPAIN consortium/IBV(CSIC) |

| EPI_Accession_ID | Originating Laboratory | Submitting Laboratory |
|---|---|---|
| EPI_ISL_452552 | Servicio de MicrobiologÃ-a y ParasitologÃ-a clÃ-nica. UCEIMP. Hospital Universitario Virgen del RocÃ-o/IBIS/CSIC/US. | SeqCOVID-SPAIN consortium/IBV(CSIC) |
| EPI_ISL_452553 | Servicio de MicrobiologÃ-a y ParasitologÃ-a clÃ-nica. UCEIMP. Hospital Universitario Virgen del RocÃ-o/IBIS/CSIC/US. | SeqCOVID-SPAIN consortium/IBV(CSIC) |
| EPI_ISL_452554 | Servicio de MicrobiologÃ-a y ParasitologÃ-a clÃ-nica. UCEIMP. Hospital Universitario Virgen del RocÃ-o/IBIS/CSIC/US. | SeqCOVID-SPAIN consortium/IBV(CSIC) |
| EPI_ISL_452555 | Servicio de MicrobiologÃ-a y ParasitologÃ-a clÃ-nica. UCEIMP. Hospital Universitario Virgen del RocÃ-o/IBIS/CSIC/US. | SeqCOVID-SPAIN consortium/IBV(CSIC) |
| EPI_ISL_452556 | Servicio de MicrobiologÃ-a y ParasitologÃ-a clÃ-nica. UCEIMP. Hospital Universitario Virgen del RocÃ-o/IBIS/CSIC/US. | SeqCOVID-SPAIN consortium/IBV(CSIC) |
| EPI_ISL_452557 | Servicio de MicrobiologÃ-a y ParasitologÃ-a clÃ-nica. UCEIMP. Hospital Universitario Virgen del RocÃ-o/IBIS/CSIC/US. | SeqCOVID-SPAIN consortium/IBV(CSIC) |
| EPI_ISL_452558 | Servicio de MicrobiologÃ-a y ParasitologÃ-a clÃ-nica. UCEIMP. Hospital Universitario Virgen del RocÃ-o/IBIS/CSIC/US. | SeqCOVID-SPAIN consortium/IBV(CSIC) |
| EPI_ISL_452559 | Servicio de MicrobiologÃ-a y ParasitologÃ-a clÃ-nica. UCEIMP. Hospital Universitario Virgen del RocÃ-o/IBIS/CSIC/US. | SeqCOVID-SPAIN consortium/IBV(CSIC) |
| EPI_ISL_452560 | Servicio de MicrobiologÃ-a y ParasitologÃ-a clÃ-nica. UCEIMP. Hospital Universitario Virgen del RocÃ-o/IBIS/CSIC/US. | SeqCOVID-SPAIN consortium/IBV(CSIC) |
| EPI_ISL_452561 | Servicio de MicrobiologÃ-a y ParasitologÃ-a clÃ-nica. UCEIMP. Hospital Universitario Virgen del RocÃ-o/IBIS/CSIC/US. | SeqCOVID-SPAIN consortium/IBV(CSIC) |
| EPI_ISL_452562 | Servicio de MicrobiologÃ-a y ParasitologÃ-a clÃ-nica. UCEIMP. Hospital Universitario Virgen del RocÃ-o/IBIS/CSIC/US. | SeqCOVID-SPAIN consortium/IBV(CSIC) |

| EPI_Accession_ID | Originating Laboratory | Submitting Laboratory |
|---|---|---|
| EPI_ISL_452563 | Servicio de MicrobiologÃ-a y ParasitologÃ-a clÃ-nica. UCEIMP. Hospital Universitario Virgen del RocÃ-o/IBIS/CSIC/US. | SeqCOVID-SPAIN consortium/IBV(CSIC) |
| EPI_ISL_452564 | Servicio de MicrobiologÃ-a y ParasitologÃ-a clÃ-nica. UCEIMP. Hospital Universitario Virgen del RocÃ-o/IBIS/CSIC/US. | SeqCOVID-SPAIN consortium/IBV(CSIC) |
| EPI_ISL_452565 | Servicio de MicrobiologÃ-a y ParasitologÃ-a clÃ-nica. UCEIMP. Hospital Universitario Virgen del RocÃ-o/IBIS/CSIC/US. | SeqCOVID-SPAIN consortium/IBV(CSIC) |
| EPI_ISL_452566 | Servicio de MicrobiologÃ-a y ParasitologÃ-a clÃ-nica. UCEIMP. Hospital Universitario Virgen del RocÃ-o/IBIS/CSIC/US. | SeqCOVID-SPAIN consortium/IBV(CSIC) |
| EPI_ISL_452567 | Servicio de MicrobiologÃ-a y ParasitologÃ-a clÃ-nica. UCEIMP. Hospital Universitario Virgen del RocÃ-o/IBIS/CSIC/US. | SeqCOVID-SPAIN consortium/IBV(CSIC) |
| EPI_ISL_452568 | Servicio de MicrobiologÃ-a y ParasitologÃ-a clÃ-nica. UCEIMP. Hospital Universitario Virgen del RocÃ-o/IBIS/CSIC/US. | SeqCOVID-SPAIN consortium/IBV(CSIC) |
| EPI_ISL_452569 | Servicio de MicrobiologÃ-a y ParasitologÃ-a clÃ-nica. UCEIMP. Hospital Universitario Virgen del RocÃ-o/IBIS/CSIC/US. | SeqCOVID-SPAIN consortium/IBV(CSIC) |
| EPI_ISL_452570 | Servicio de MicrobiologÃ-a y ParasitologÃ-a clÃ-nica. UCEIMP. Hospital Universitario Virgen del RocÃ-o/IBIS/CSIC/US. | SeqCOVID-SPAIN consortium/IBV(CSIC) |
| EPI_ISL_452571 | Servicio de MicrobiologÃ-a y ParasitologÃ-a clÃ-nica. UCEIMP. Hospital Universitario Virgen del RocÃ-o/IBIS/CSIC/US. | SeqCOVID-SPAIN consortium/IBV(CSIC) |
| EPI_ISL_452572 | Servicio de MicrobiologÃ-a y ParasitologÃ-a clÃ-nica. UCEIMP. Hospital Universitario Virgen del RocÃ-o/IBIS/CSIC/US. | SeqCOVID-SPAIN consortium/IBV(CSIC) |
| EPI_ISL_452573 | Servicio de MicrobiologÃ-a y ParasitologÃ-a clÃ-nica. UCEIMP. Hospital Universitario Virgen del RocÃ-o/IBIS/CSIC/US. | SeqCOVID-SPAIN consortium/IBV(CSIC) |

| EPI_Accession_ID | Originating Laboratory | Submitting Laboratory |
|---|---|---|
| EPI_ISL_452574 | Servicio de MicrobiologÃ-a y ParasitologÃ-a clÃ-nica. UCEIMP. Hospital Universitario Virgen del RocÃ-o/IBIS/CSIC/US. | SeqCOVID-SPAIN consortium/IBV(CSIC) |
| EPI_ISL_452575 | Servicio de MicrobiologÃ-a y ParasitologÃ-a clÃ-nica. UCEIMP. Hospital Universitario Virgen del RocÃ-o/IBIS/CSIC/US. | SeqCOVID-SPAIN consortium/IBV(CSIC) |
| EPI_ISL_452576 | Servicio de MicrobiologÃ-a y ParasitologÃ-a clÃ-nica. UCEIMP. Hospital Universitario Virgen del RocÃ-o/IBIS/CSIC/US. | SeqCOVID-SPAIN consortium/IBV(CSIC) |
| EPI_ISL_452577 | Servicio de MicrobiologÃ-a y ParasitologÃ-a clÃ-nica. UCEIMP. Hospital Universitario Virgen del RocÃ-o/IBIS/CSIC/US. | SeqCOVID-SPAIN consortium/IBV(CSIC) |
| EPI_ISL_452578 | Servicio de MicrobiologÃ-a y ParasitologÃ-a clÃ-nica. UCEIMP. Hospital Universitario Virgen del RocÃ-o/IBIS/CSIC/US. | SeqCOVID-SPAIN consortium/IBV(CSIC) |
| EPI_ISL_452579 | Servicio de MicrobiologÃ-a y ParasitologÃ-a clÃ-nica. UCEIMP. Hospital Universitario Virgen del RocÃ-o/IBIS/CSIC/US. | SeqCOVID-SPAIN consortium/IBV(CSIC) |
| EPI_ISL_452580 | Servicio de MicrobiologÃ-a y ParasitologÃ-a clÃ-nica. UCEIMP. Hospital Universitario Virgen del RocÃ-o/IBIS/CSIC/US. | SeqCOVID-SPAIN consortium/IBV(CSIC) |
| EPI_ISL_452581 | Servicio de MicrobiologÃ-a y ParasitologÃ-a clÃ-nica. UCEIMP. Hospital Universitario Virgen del RocÃ-o/IBIS/CSIC/US. | SeqCOVID-SPAIN consortium/IBV(CSIC) |
| EPI_ISL_452582 | Servicio de MicrobiologÃ-a y ParasitologÃ-a clÃ-nica. UCEIMP. Hospital Universitario Virgen del RocÃ-o/IBIS/CSIC/US. | SeqCOVID-SPAIN consortium/IBV(CSIC) |
| EPI_ISL_452583 | Servicio de MicrobiologÃ-a y ParasitologÃ-a clÃ-nica. UCEIMP. Hospital Universitario Virgen del RocÃ-o/IBIS/CSIC/US. | SeqCOVID-SPAIN consortium/IBV(CSIC) |
| EPI_ISL_452584 | Servicio de MicrobiologÃ-a y ParasitologÃ-a clÃ-nica. UCEIMP. Hospital Universitario Virgen del RocÃ-o/IBIS/CSIC/US. | SeqCOVID-SPAIN consortium/IBV(CSIC) |

| EPI_Accession_ID | Originating Laboratory | Submitting Laboratory |
| --- | --- | --- |
| EPI_ISL_452585 | Servicio de MicrobiologÃ-a y ParasitologÃ-a clÃ-nica. UCEIMP. Hospital Universitario Virgen del RocÃ-o/IBIS/CSIC/US. | SeqCOVID-SPAIN consortium/IBV(CSIC) |
| EPI_ISL_452586 | Servicio de MicrobiologÃ-a y ParasitologÃ-a clÃ-nica. UCEIMP. Hospital Universitario Virgen del RocÃ-o/IBIS/CSIC/US. | SeqCOVID-SPAIN consortium/IBV(CSIC) |
| EPI_ISL_452587 | Servicio de MicrobiologÃ-a y ParasitologÃ-a clÃ-nica. UCEIMP. Hospital Universitario Virgen del RocÃ-o/IBIS/CSIC/US. | SeqCOVID-SPAIN consortium/IBV(CSIC) |
| EPI_ISL_452588 | Servicio de MicrobiologÃ-a y ParasitologÃ-a clÃ-nica. UCEIMP. Hospital Universitario Virgen del RocÃ-o/IBIS/CSIC/US. | SeqCOVID-SPAIN consortium/IBV(CSIC) |
| EPI_ISL_452589 | Servicio de MicrobiologÃ-a y ParasitologÃ-a clÃ-nica. UCEIMP. Hospital Universitario Virgen del RocÃ-o/IBIS/CSIC/US. | SeqCOVID-SPAIN consortium/IBV(CSIC) |
| EPI_ISL_452590 | Servicio de MicrobiologÃ-a y ParasitologÃ-a clÃ-nica. UCEIMP. Hospital Universitario Virgen del RocÃ-o/IBIS/CSIC/US. | SeqCOVID-SPAIN consortium/IBV(CSIC) |
| EPI_ISL_452591 | Servicio de MicrobiologÃ-a y ParasitologÃ-a clÃ-nica. UCEIMP. Hospital Universitario Virgen del RocÃ-o/IBIS/CSIC/US. | SeqCOVID-SPAIN consortium/IBV(CSIC) |
| EPI_ISL_452592 | Servicio de MicrobiologÃ-a y ParasitologÃ-a clÃ-nica. UCEIMP. Hospital Universitario Virgen del RocÃ-o/IBIS/CSIC/US. | SeqCOVID-SPAIN consortium/IBV(CSIC) |
| EPI_ISL_452593 | Servicio de MicrobiologÃ-a y ParasitologÃ-a clÃ-nica. UCEIMP. Hospital Universitario Virgen del RocÃ-o/IBIS/CSIC/US. | SeqCOVID-SPAIN consortium/IBV(CSIC) |
| EPI_ISL_452594 | Servicio de MicrobiologÃ-a y ParasitologÃ-a clÃ-nica. UCEIMP. Hospital Universitario Virgen del RocÃ-o/IBIS/CSIC/US. | SeqCOVID-SPAIN consortium/IBV(CSIC) |
| EPI_ISL_452595 | Servicio de MicrobiologÃ-a y ParasitologÃ-a clÃ-nica. UCEIMP. Hospital Universitario Virgen del RocÃ-o/IBIS/CSIC/US. | SeqCOVID-SPAIN consortium/IBV(CSIC) |

| EPI_Accession_ID | Originating Laboratory | Submitting Laboratory |
|---|---|---|
| EPI_ISL_452596 | Servicio de MicrobiologÃ-a y ParasitologÃ-a clÃ-nica. UCEIMP. Hospital Universitario Virgen del RocÃ-o/IBIS/CSIC/US. | SeqCOVID-SPAIN consortium/IBV(CSIC) |
| EPI_ISL_452597 | Servicio de MicrobiologÃ-a y ParasitologÃ-a clÃ-nica. UCEIMP. Hospital Universitario Virgen del RocÃ-o/IBIS/CSIC/US. | SeqCOVID-SPAIN consortium/IBV(CSIC) |
| EPI_ISL_452598 | Servicio de MicrobiologÃ-a y ParasitologÃ-a clÃ-nica. UCEIMP. Hospital Universitario Virgen del RocÃ-o/IBIS/CSIC/US. | SeqCOVID-SPAIN consortium/IBV(CSIC) |
| EPI_ISL_452599 | Servicio de MicrobiologÃ-a y ParasitologÃ-a clÃ-nica. UCEIMP. Hospital Universitario Virgen del RocÃ-o/IBIS/CSIC/US. | SeqCOVID-SPAIN consortium/IBV(CSIC) |
| EPI_ISL_452600 | Servicio de MicrobiologÃ-a y ParasitologÃ-a clÃ-nica. UCEIMP. Hospital Universitario Virgen del RocÃ-o/IBIS/CSIC/US. | SeqCOVID-SPAIN consortium/IBV(CSIC) |
| EPI_ISL_452601 | Servicio de MicrobiologÃ-a y ParasitologÃ-a clÃ-nica. UCEIMP. Hospital Universitario Virgen del RocÃ-o/IBIS/CSIC/US. | SeqCOVID-SPAIN consortium/IBV(CSIC) |
| EPI_ISL_452602 | Servicio de MicrobiologÃ-a y ParasitologÃ-a clÃ-nica. UCEIMP. Hospital Universitario Virgen del RocÃ-o/IBIS/CSIC/US. | SeqCOVID-SPAIN consortium/IBV(CSIC) |
| EPI_ISL_452603 | Servicio de MicrobiologÃ-a y ParasitologÃ-a clÃ-nica. UCEIMP. Hospital Universitario Virgen del RocÃ-o/IBIS/CSIC/US. | SeqCOVID-SPAIN consortium/IBV(CSIC) |
| EPI_ISL_452604 | Servicio de MicrobiologÃ-a y ParasitologÃ-a clÃ-nica. UCEIMP. Hospital Universitario Virgen del RocÃ-o/IBIS/CSIC/US. | SeqCOVID-SPAIN consortium/IBV(CSIC) |
| EPI_ISL_452605 | Servicio de MicrobiologÃ-a y ParasitologÃ-a clÃ-nica. UCEIMP. Hospital Universitario Virgen del RocÃ-o/IBIS/CSIC/US. | SeqCOVID-SPAIN consortium/IBV(CSIC) |
| EPI_ISL_452606 | Servicio de MicrobiologÃ-a y ParasitologÃ-a clÃ-nica. UCEIMP. Hospital Universitario Virgen del RocÃ-o/IBIS/CSIC/US. | SeqCOVID-SPAIN consortium/IBV(CSIC) |

| EPI_Accession_ID | Originating Laboratory | Submitting Laboratory |
|---|---|---|
| EPI_ISL_452607 | Servicio de MicrobiologÃ-a y ParasitologÃ-a clÃ-nica. UCEIMP. Hospital Universitario Virgen del RocÃ-o/IBIS/CSIC/US. | SeqCOVID-SPAIN consortium/IBV(CSIC) |
| EPI_ISL_452608 | Servicio de MicrobiologÃ-a y ParasitologÃ-a clÃ-nica. UCEIMP. Hospital Universitario Virgen del RocÃ-o/IBIS/CSIC/US. | SeqCOVID-SPAIN consortium/IBV(CSIC) |
| EPI_ISL_452609 | Servicio de MicrobiologÃ-a y ParasitologÃ-a clÃ-nica. UCEIMP. Hospital Universitario Virgen del RocÃ-o/IBIS/CSIC/US. | SeqCOVID-SPAIN consortium/IBV(CSIC) |
| EPI_ISL_452610 | Servicio de MicrobiologÃ-a y ParasitologÃ-a clÃ-nica. UCEIMP. Hospital Universitario Virgen del RocÃ-o/IBIS/CSIC/US. | SeqCOVID-SPAIN consortium/IBV(CSIC) |
| EPI_ISL_452611 | Servicio de MicrobiologÃ-a y ParasitologÃ-a clÃ-nica. UCEIMP. Hospital Universitario Virgen del RocÃ-o/IBIS/CSIC/US. | SeqCOVID-SPAIN consortium/IBV(CSIC) |
| EPI_ISL_452612 | Servicio de MicrobiologÃ-a y ParasitologÃ-a clÃ-nica. UCEIMP. Hospital Universitario Virgen del RocÃ-o/IBIS/CSIC/US. | SeqCOVID-SPAIN consortium/IBV(CSIC) |
| EPI_ISL_452613 | Servicio de MicrobiologÃ-a y ParasitologÃ-a clÃ-nica. UCEIMP. Hospital Universitario Virgen del RocÃ-o/IBIS/CSIC/US. | SeqCOVID-SPAIN consortium/IBV(CSIC) |
| EPI_ISL_452614 | Servicio de MicrobiologÃ-a y ParasitologÃ-a clÃ-nica. UCEIMP. Hospital Universitario Virgen del RocÃ-o/IBIS/CSIC/US. | SeqCOVID-SPAIN consortium/IBV(CSIC) |
| EPI_ISL_452615 | Servicio de MicrobiologÃ-a y ParasitologÃ-a clÃ-nica. UCEIMP. Hospital Universitario Virgen del RocÃ-o/IBIS/CSIC/US. | SeqCOVID-SPAIN consortium/IBV(CSIC) |
| EPI_ISL_452616 | Servicio de MicrobiologÃ-a y ParasitologÃ-a clÃ-nica. UCEIMP. Hospital Universitario Virgen del RocÃ-o/IBIS/CSIC/US. | SeqCOVID-SPAIN consortium/IBV(CSIC) |

| EPI_Accession_ID | Originating Laboratory | Submitting Laboratory |
|---|---|---|
| EPI_ISL_452617 | Servicio de MicrobiologÃ-a. Hospital Universitario Donostia. OSI Donostialdea. Ã�ea de Enfermedades Infecciosas, Grupo de InfecciÃ³n Respiratoria y Resistencia Antimicrobiana. Instituto de InvestigaciÃ³n Sanitaria Biodonostia. | SeqCOVID-SPAIN consortium/IBV(CSIC) |
| EPI_ISL_452618 | Servicio de MicrobiologÃ-a. Hospital Universitario Donostia. OSI Donostialdea. Ã�ea de Enfermedades Infecciosas, Grupo de InfecciÃ³n Respiratoria y Resistencia Antimicrobiana. Instituto de InvestigaciÃ³n Sanitaria Biodonostia. | SeqCOVID-SPAIN consortium/IBV(CSIC) |
| EPI_ISL_452619 | Servicio de MicrobiologÃ-a. Hospital Universitario Donostia. OSI Donostialdea. Ã�ea de Enfermedades Infecciosas, Grupo de InfecciÃ³n Respiratoria y Resistencia Antimicrobiana. Instituto de InvestigaciÃ³n Sanitaria Biodonostia. | SeqCOVID-SPAIN consortium/IBV(CSIC) |
| EPI_ISL_452620 | Servicio de MicrobiologÃ-a. Hospital Universitario Donostia. OSI Donostialdea. Ã�ea de Enfermedades Infecciosas, Grupo de InfecciÃ³n Respiratoria y Resistencia Antimicrobiana. Instituto de InvestigaciÃ³n Sanitaria Biodonostia. | SeqCOVID-SPAIN consortium/IBV(CSIC) |
| EPI_ISL_452621 | Servicio de MicrobiologÃ-a. Hospital Universitario Donostia. OSI Donostialdea. Ã�ea de Enfermedades Infecciosas, Grupo de InfecciÃ³n Respiratoria y Resistencia Antimicrobiana. Instituto de InvestigaciÃ³n Sanitaria Biodonostia. | SeqCOVID-SPAIN consortium/IBV(CSIC) |
| EPI_ISL_452622 | Servicio de MicrobiologÃ-a. Hospital Universitario Donostia. OSI Donostialdea. Ã�ea de Enfermedades Infecciosas, Grupo de InfecciÃ³n Respiratoria y Resistencia Antimicrobiana. Instituto de InvestigaciÃ³n Sanitaria Biodonostia. | SeqCOVID-SPAIN consortium/IBV(CSIC) |

| EPI_Accession_ID | Originating Laboratory | Submitting Laboratory |
|---|---|---|
| EPI_ISL_452623 | Servicio de MicrobiologÃ-a. Hospital Universitario Donostia. OSI Donostialdea. Ã�ea de Enfermedades Infecciosas, Grupo de InfecciÃ³n Respiratoria y Resistencia Antimicrobiana. Instituto de InvestigaciÃ³n Sanitaria Biodonostia. | SeqCOVID-SPAIN consortium/IBV(CSIC) |
| EPI_ISL_452624 | Servicio de MicrobiologÃ-a. Hospital Universitario Donostia. OSI Donostialdea. Ã�ea de Enfermedades Infecciosas, Grupo de InfecciÃ³n Respiratoria y Resistencia Antimicrobiana. Instituto de InvestigaciÃ³n Sanitaria Biodonostia. | SeqCOVID-SPAIN consortium/IBV(CSIC) |
| EPI_ISL_452625 | Servicio de MicrobiologÃ-a. Hospital Universitario Donostia. OSI Donostialdea. Ã�ea de Enfermedades Infecciosas, Grupo de InfecciÃ³n Respiratoria y Resistencia Antimicrobiana. Instituto de InvestigaciÃ³n Sanitaria Biodonostia. | SeqCOVID-SPAIN consortium/IBV(CSIC) |
| EPI_ISL_452626 | Servicio de MicrobiologÃ-a. Hospital Universitario Donostia. OSI Donostialdea. Ã�ea de Enfermedades Infecciosas, Grupo de InfecciÃ³n Respiratoria y Resistencia Antimicrobiana. Instituto de InvestigaciÃ³n Sanitaria Biodonostia. | SeqCOVID-SPAIN consortium/IBV(CSIC) |
| EPI_ISL_452627 | Servicio de MicrobiologÃ-a. Hospital Universitario Donostia. OSI Donostialdea. Ã�ea de Enfermedades Infecciosas, Grupo de InfecciÃ³n Respiratoria y Resistencia Antimicrobiana. Instituto de InvestigaciÃ³n Sanitaria Biodonostia. | SeqCOVID-SPAIN consortium/IBV(CSIC) |
| EPI_ISL_452628 | Servicio de MicrobiologÃ-a. Hospital Universitario Donostia. OSI Donostialdea. Ã�ea de Enfermedades Infecciosas, Grupo de InfecciÃ³n Respiratoria y Resistencia Antimicrobiana. Instituto de InvestigaciÃ³n Sanitaria Biodonostia. | SeqCOVID-SPAIN consortium/IBV(CSIC) |

| EPI_Accession_ID | Originating Laboratory | Submitting Laboratory |
|---|---|---|
| EPI_ISL_452629 | Servicio de MicrobiologÃ-a. Hospital Universitario Donostia. OSI Donostialdea. Ã�ea de Enfermedades Infecciosas, Grupo de InfecciÃ³n Respiratoria y Resistencia Antimicrobiana. Instituto de InvestigaciÃ³n Sanitaria Biodonostia. | SeqCOVID-SPAIN consortium/IBV(CSIC) |
| EPI_ISL_452630 | Servicio de MicrobiologÃ-a. Hospital Universitario Donostia. OSI Donostialdea. Ã�ea de Enfermedades Infecciosas, Grupo de InfecciÃ³n Respiratoria y Resistencia Antimicrobiana. Instituto de InvestigaciÃ³n Sanitaria Biodonostia. | SeqCOVID-SPAIN consortium/IBV(CSIC) |
| EPI_ISL_452631 | Servicio de MicrobiologÃ-a. Hospital Universitario Donostia. OSI Donostialdea. Ã�ea de Enfermedades Infecciosas, Grupo de InfecciÃ³n Respiratoria y Resistencia Antimicrobiana. Instituto de InvestigaciÃ³n Sanitaria Biodonostia. | SeqCOVID-SPAIN consortium/IBV(CSIC) |
| EPI_ISL_452632 | Servicio de MicrobiologÃ-a. Hospital Universitario Donostia. OSI Donostialdea. Ã�ea de Enfermedades Infecciosas, Grupo de InfecciÃ³n Respiratoria y Resistencia Antimicrobiana. Instituto de InvestigaciÃ³n Sanitaria Biodonostia. | SeqCOVID-SPAIN consortium/IBV(CSIC) |
| EPI_ISL_452633 | Servicio de MicrobiologÃ-a. Hospital Universitario Donostia. OSI Donostialdea. Ã�ea de Enfermedades Infecciosas, Grupo de InfecciÃ³n Respiratoria y Resistencia Antimicrobiana. Instituto de InvestigaciÃ³n Sanitaria Biodonostia. | SeqCOVID-SPAIN consortium/IBV(CSIC) |
| EPI_ISL_452634 | Servicio de MicrobiologÃ-a. Hospital Universitario Donostia. OSI Donostialdea. Ã�ea de Enfermedades Infecciosas, Grupo de InfecciÃ³n Respiratoria y Resistencia Antimicrobiana. Instituto de InvestigaciÃ³n Sanitaria Biodonostia. | SeqCOVID-SPAIN consortium/IBV(CSIC) |

| EPI_Accession_ID | Originating Laboratory | Submitting Laboratory |
| --- | --- | --- |
| EPI_ISL_452635 | Servicio de MicrobiologÃ-a. Hospital Universitario Donostia. OSI Donostialdea. Ã�ea de Enfermedades Infecciosas, Grupo de InfecciÃ³n Respiratoria y Resistencia Antimicrobiana. Instituto de InvestigaciÃ³n Sanitaria Biodonostia. | SeqCOVID-SPAIN consortium/IBV(CSIC) |
| EPI_ISL_452636 | Servicio de MicrobiologÃ-a. Hospital Universitario Donostia. OSI Donostialdea. Ã�ea de Enfermedades Infecciosas, Grupo de InfecciÃ³n Respiratoria y Resistencia Antimicrobiana. Instituto de InvestigaciÃ³n Sanitaria Biodonostia. | SeqCOVID-SPAIN consortium/IBV(CSIC) |
| EPI_ISL_452637 | Servicio de MicrobiologÃ-a. Hospital Universitario Donostia. OSI Donostialdea. Ã�ea de Enfermedades Infecciosas, Grupo de InfecciÃ³n Respiratoria y Resistencia Antimicrobiana. Instituto de InvestigaciÃ³n Sanitaria Biodonostia. | SeqCOVID-SPAIN consortium/IBV(CSIC) |
| EPI_ISL_452638 | Servicio de MicrobiologÃ-a. Hospital Universitario Donostia. OSI Donostialdea. Ã�ea de Enfermedades Infecciosas, Grupo de InfecciÃ³n Respiratoria y Resistencia Antimicrobiana. Instituto de InvestigaciÃ³n Sanitaria Biodonostia. | SeqCOVID-SPAIN consortium/IBV(CSIC) |
| EPI_ISL_452639 | Servicio de MicrobiologÃ-a. Hospital Universitario Donostia. OSI Donostialdea. Ã�ea de Enfermedades Infecciosas, Grupo de InfecciÃ³n Respiratoria y Resistencia Antimicrobiana. Instituto de InvestigaciÃ³n Sanitaria Biodonostia. | SeqCOVID-SPAIN consortium/IBV(CSIC) |
| EPI_ISL_452640 | Servicio de MicrobiologÃ-a. Hospital Universitario Donostia. OSI Donostialdea. Ã�ea de Enfermedades Infecciosas, Grupo de InfecciÃ³n Respiratoria y Resistencia Antimicrobiana. Instituto de InvestigaciÃ³n Sanitaria Biodonostia. | SeqCOVID-SPAIN consortium/IBV(CSIC) |

| EPI_Accession_ID | Originating Laboratory | Submitting Laboratory |
|---|---|---|
| EPI_ISL_452641 | Servicio de MicrobiologÃ-a. Hospital Universitario Donostia. OSI Donostialdea. Ã�ea de Enfermedades Infecciosas, Grupo de InfecciÃ³n Respiratoria y Resistencia Antimicrobiana. Instituto de InvestigaciÃ³n Sanitaria Biodonostia. | SeqCOVID-SPAIN consortium/IBV(CSIC) |
| EPI_ISL_452642 | Servicio de MicrobiologÃ-a. Hospital Universitario Donostia. OSI Donostialdea. Ã�ea de Enfermedades Infecciosas, Grupo de InfecciÃ³n Respiratoria y Resistencia Antimicrobiana. Instituto de InvestigaciÃ³n Sanitaria Biodonostia. | SeqCOVID-SPAIN consortium/IBV(CSIC) |
| EPI_ISL_452643 | Servicio de MicrobiologÃ-a. Hospital Universitario Donostia. OSI Donostialdea. Ã�ea de Enfermedades Infecciosas, Grupo de InfecciÃ³n Respiratoria y Resistencia Antimicrobiana. Instituto de InvestigaciÃ³n Sanitaria Biodonostia. | SeqCOVID-SPAIN consortium/IBV(CSIC) |
| EPI_ISL_452644 | Servicio de MicrobiologÃ-a. Hospital Universitario Donostia. OSI Donostialdea. Ã�ea de Enfermedades Infecciosas, Grupo de InfecciÃ³n Respiratoria y Resistencia Antimicrobiana. Instituto de InvestigaciÃ³n Sanitaria Biodonostia. | SeqCOVID-SPAIN consortium/IBV(CSIC) |
| EPI_ISL_452645 | Servicio de MicrobiologÃ-a. Hospital Universitario Donostia. OSI Donostialdea. Ã�ea de Enfermedades Infecciosas, Grupo de InfecciÃ³n Respiratoria y Resistencia Antimicrobiana. Instituto de InvestigaciÃ³n Sanitaria Biodonostia. | SeqCOVID-SPAIN consortium/IBV(CSIC) |
| EPI_ISL_452646 | Servicio de MicrobiologÃ-a. Hospital Universitario Donostia. OSI Donostialdea. Ã�ea de Enfermedades Infecciosas, Grupo de InfecciÃ³n Respiratoria y Resistencia Antimicrobiana. Instituto de InvestigaciÃ³n Sanitaria Biodonostia. | SeqCOVID-SPAIN consortium/IBV(CSIC) |

| EPI_Accession_ID | Originating Laboratory | Submitting Laboratory |
|---|---|---|
| EPI_ISL_452647 | Servicio de MicrobiologÃ-a. Hospital Universitario Donostia. OSI Donostialdea. Ã�ea de Enfermedades Infecciosas, Grupo de InfecciÃ³n Respiratoria y Resistencia Antimicrobiana. Instituto de InvestigaciÃ³n Sanitaria Biodonostia. | SeqCOVID-SPAIN consortium/IBV(CSIC) |
| EPI_ISL_452648 | Servicio de MicrobiologÃ-a. Hospital Universitario Donostia. OSI Donostialdea. Ã�ea de Enfermedades Infecciosas, Grupo de InfecciÃ³n Respiratoria y Resistencia Antimicrobiana. Instituto de InvestigaciÃ³n Sanitaria Biodonostia. | SeqCOVID-SPAIN consortium/IBV(CSIC) |
| EPI_ISL_452649 | Servicio de MicrobiologÃ-a. Hospital Universitario Donostia. OSI Donostialdea. Ã�ea de Enfermedades Infecciosas, Grupo de InfecciÃ³n Respiratoria y Resistencia Antimicrobiana. Instituto de InvestigaciÃ³n Sanitaria Biodonostia. | SeqCOVID-SPAIN consortium/IBV(CSIC) |
| EPI_ISL_452650 | Servicio de MicrobiologÃ-a. Hospital Universitario Donostia. OSI Donostialdea. Ã�ea de Enfermedades Infecciosas, Grupo de InfecciÃ³n Respiratoria y Resistencia Antimicrobiana. Instituto de InvestigaciÃ³n Sanitaria Biodonostia. | SeqCOVID-SPAIN consortium/IBV(CSIC) |
| EPI_ISL_452651 | Servicio de MicrobiologÃ-a. Hospital Universitario Donostia. OSI Donostialdea. Ã�ea de Enfermedades Infecciosas, Grupo de InfecciÃ³n Respiratoria y Resistencia Antimicrobiana. Instituto de InvestigaciÃ³n Sanitaria Biodonostia. | SeqCOVID-SPAIN consortium/IBV(CSIC) |
| EPI_ISL_452652 | Servicio de MicrobiologÃ-a. Hospital Universitario Donostia. OSI Donostialdea. Ã�ea de Enfermedades Infecciosas, Grupo de InfecciÃ³n Respiratoria y Resistencia Antimicrobiana. Instituto de InvestigaciÃ³n Sanitaria Biodonostia. | SeqCOVID-SPAIN consortium/IBV(CSIC) |

| EPI_Accession_ID | Originating Laboratory | Submitting Laboratory |
|---|---|---|
| EPI_ISL_452653 | Servicio de MicrobiologÃ-a. Hospital Universitario Donostia. OSI Donostialdea. Ã�ea de Enfermedades Infecciosas, Grupo de InfecciÃ³n Respiratoria y Resistencia Antimicrobiana. Instituto de InvestigaciÃ³n Sanitaria Biodonostia. | SeqCOVID-SPAIN consortium/IBV(CSIC) |
| EPI_ISL_452654 | Servicio de MicrobiologÃ-a. Hospital Universitario Donostia. OSI Donostialdea. Ã�ea de Enfermedades Infecciosas, Grupo de InfecciÃ³n Respiratoria y Resistencia Antimicrobiana. Instituto de InvestigaciÃ³n Sanitaria Biodonostia. | SeqCOVID-SPAIN consortium/IBV(CSIC) |
| EPI_ISL_452655 | Servicio de MicrobiologÃ-a. Hospital Universitario Donostia. OSI Donostialdea. Ã�ea de Enfermedades Infecciosas, Grupo de InfecciÃ³n Respiratoria y Resistencia Antimicrobiana. Instituto de InvestigaciÃ³n Sanitaria Biodonostia. | SeqCOVID-SPAIN consortium/IBV(CSIC) |
| EPI_ISL_452656 | Servicio de MicrobiologÃ-a. Hospital Universitario Donostia. OSI Donostialdea. Ã�ea de Enfermedades Infecciosas, Grupo de InfecciÃ³n Respiratoria y Resistencia Antimicrobiana. Instituto de InvestigaciÃ³n Sanitaria Biodonostia. | SeqCOVID-SPAIN consortium/IBV(CSIC) |
| EPI_ISL_452657 | Servicio de MicrobiologÃ-a. Hospital Universitario Donostia. OSI Donostialdea. Ã�ea de Enfermedades Infecciosas, Grupo de InfecciÃ³n Respiratoria y Resistencia Antimicrobiana. Instituto de InvestigaciÃ³n Sanitaria Biodonostia. | SeqCOVID-SPAIN consortium/IBV(CSIC) |
| EPI_ISL_452658 | Servicio de MicrobiologÃ-a. Hospital Universitario Donostia. OSI Donostialdea. Ã�ea de Enfermedades Infecciosas, Grupo de InfecciÃ³n Respiratoria y Resistencia Antimicrobiana. Instituto de InvestigaciÃ³n Sanitaria Biodonostia. | SeqCOVID-SPAIN consortium/IBV(CSIC) |

| EPI_Accession_ID | Originating Laboratory | Submitting Laboratory |
|---|---|---|
| EPI_ISL_452659 | Servicio de MicrobiologÃ-a. Hospital Universitario Donostia. OSI Donostialdea. Ã�ea de Enfermedades Infecciosas, Grupo de InfecciÃ³n Respiratoria y Resistencia Antimicrobiana. Instituto de InvestigaciÃ³n Sanitaria Biodonostia. | SeqCOVID-SPAIN consortium/IBV(CSIC) |
| EPI_ISL_452660 | Servicio de MicrobiologÃ-a. Hospital Universitario Donostia. OSI Donostialdea. Ã�ea de Enfermedades Infecciosas, Grupo de InfecciÃ³n Respiratoria y Resistencia Antimicrobiana. Instituto de InvestigaciÃ³n Sanitaria Biodonostia. | SeqCOVID-SPAIN consortium/IBV(CSIC) |
| EPI_ISL_452661 | Servicio de MicrobiologÃ-a. Hospital Universitario Donostia. OSI Donostialdea. Ã�ea de Enfermedades Infecciosas, Grupo de InfecciÃ³n Respiratoria y Resistencia Antimicrobiana. Instituto de InvestigaciÃ³n Sanitaria Biodonostia. | SeqCOVID-SPAIN consortium/IBV(CSIC) |
| EPI_ISL_452662 | Servicio de MicrobiologÃ-a. Hospital Universitario Donostia. OSI Donostialdea. Ã�ea de Enfermedades Infecciosas, Grupo de InfecciÃ³n Respiratoria y Resistencia Antimicrobiana. Instituto de InvestigaciÃ³n Sanitaria Biodonostia. | SeqCOVID-SPAIN consortium/IBV(CSIC) |
| EPI_ISL_452663 | Servicio de MicrobiologÃ-a. Hospital Universitario Donostia. OSI Donostialdea. Ã�ea de Enfermedades Infecciosas, Grupo de InfecciÃ³n Respiratoria y Resistencia Antimicrobiana. Instituto de InvestigaciÃ³n Sanitaria Biodonostia. | SeqCOVID-SPAIN consortium/IBV(CSIC) |
| EPI_ISL_452664 | Servicio de MicrobiologÃ-a. Hospital Universitario Donostia. OSI Donostialdea. Ã�ea de Enfermedades Infecciosas, Grupo de InfecciÃ³n Respiratoria y Resistencia Antimicrobiana. Instituto de InvestigaciÃ³n Sanitaria Biodonostia. | SeqCOVID-SPAIN consortium/IBV(CSIC) |

| EPI_Accession_ID | Originating Laboratory | Submitting Laboratory |
|---|---|---|
| EPI_ISL_452665 | Servicio de MicrobiologÃ-a. Hospital Universitario Donostia. OSI Donostialdea. Ã�ea de Enfermedades Infecciosas, Grupo de InfecciÃ³n Respiratoria y Resistencia Antimicrobiana. Instituto de InvestigaciÃ³n Sanitaria Biodonostia. | SeqCOVID-SPAIN consortium/IBV(CSIC) |
| EPI_ISL_452666 | Servicio de MicrobiologÃ-a. Hospital Universitario Donostia. OSI Donostialdea. Ã�ea de Enfermedades Infecciosas, Grupo de InfecciÃ³n Respiratoria y Resistencia Antimicrobiana. Instituto de InvestigaciÃ³n Sanitaria Biodonostia. | SeqCOVID-SPAIN consortium/IBV(CSIC) |
| EPI_ISL_452667 | Servicio de MicrobiologÃ-a. Hospital Universitario Donostia. OSI Donostialdea. Ã�ea de Enfermedades Infecciosas, Grupo de InfecciÃ³n Respiratoria y Resistencia Antimicrobiana. Instituto de InvestigaciÃ³n Sanitaria Biodonostia. | SeqCOVID-SPAIN consortium/IBV(CSIC) |
| EPI_ISL_452668 | Servicio de MicrobiologÃ-a. Hospital Universitario Donostia. OSI Donostialdea. Ã�ea de Enfermedades Infecciosas, Grupo de InfecciÃ³n Respiratoria y Resistencia Antimicrobiana. Instituto de InvestigaciÃ³n Sanitaria Biodonostia. | SeqCOVID-SPAIN consortium/IBV(CSIC) |
| EPI_ISL_452669 | Servicio de MicrobiologÃ-a. Hospital Universitario Donostia. OSI Donostialdea. Ã�ea de Enfermedades Infecciosas, Grupo de InfecciÃ³n Respiratoria y Resistencia Antimicrobiana. Instituto de InvestigaciÃ³n Sanitaria Biodonostia. | SeqCOVID-SPAIN consortium/IBV(CSIC) |
| EPI_ISL_452670 | Servicio de MicrobiologÃ-a. Hospital Universitario Donostia. OSI Donostialdea. Ã�ea de Enfermedades Infecciosas, Grupo de InfecciÃ³n Respiratoria y Resistencia Antimicrobiana. Instituto de InvestigaciÃ³n Sanitaria Biodonostia. | SeqCOVID-SPAIN consortium/IBV(CSIC) |

| EPI_Accession_ID | Originating Laboratory | Submitting Laboratory |
|---|---|---|
| EPI_ISL_452671 | Servicio de MicrobiologÃ-a. Hospital Universitario Donostia. OSI Donostialdea. Ã�ea de Enfermedades Infecciosas, Grupo de InfecciÃ³n Respiratoria y Resistencia Antimicrobiana. Instituto de InvestigaciÃ³n Sanitaria Biodonostia. | SeqCOVID-SPAIN consortium/IBV(CSIC) |
| EPI_ISL_452672 | Servicio de MicrobiologÃ-a. Hospital Universitario Donostia. OSI Donostialdea. Ã�ea de Enfermedades Infecciosas, Grupo de InfecciÃ³n Respiratoria y Resistencia Antimicrobiana. Instituto de InvestigaciÃ³n Sanitaria Biodonostia. | SeqCOVID-SPAIN consortium/IBV(CSIC) |
| EPI_ISL_452673 | Servicio de MicrobiologÃ-a. Hospital Universitario Donostia. OSI Donostialdea. Ã�ea de Enfermedades Infecciosas, Grupo de InfecciÃ³n Respiratoria y Resistencia Antimicrobiana. Instituto de InvestigaciÃ³n Sanitaria Biodonostia. | SeqCOVID-SPAIN consortium/IBV(CSIC) |
| EPI_ISL_452674 | Servicio de MicrobiologÃ-a. Hospital Universitario Donostia. OSI Donostialdea. Ã�ea de Enfermedades Infecciosas, Grupo de InfecciÃ³n Respiratoria y Resistencia Antimicrobiana. Instituto de InvestigaciÃ³n Sanitaria Biodonostia. | SeqCOVID-SPAIN consortium/IBV(CSIC) |
| EPI_ISL_452675 | Servicio de MicrobiologÃ-a. Hospital Universitario Donostia. OSI Donostialdea. Ã�ea de Enfermedades Infecciosas, Grupo de InfecciÃ³n Respiratoria y Resistencia Antimicrobiana. Instituto de InvestigaciÃ³n Sanitaria Biodonostia. | SeqCOVID-SPAIN consortium/IBV(CSIC) |
| EPI_ISL_452676 | Servicio de MicrobiologÃ-a. Hospital Universitario Donostia. OSI Donostialdea. Ã�ea de Enfermedades Infecciosas, Grupo de InfecciÃ³n Respiratoria y Resistencia Antimicrobiana. Instituto de InvestigaciÃ³n Sanitaria Biodonostia. | SeqCOVID-SPAIN consortium/IBV(CSIC) |

| EPI_Accession_ID | Originating Laboratory | Submitting Laboratory |
|---|---|---|
| EPI_ISL_452677 | Servicio de MicrobiologÃ-a. Hospital Universitario Donostia. OSI Donostialdea. Ã�ea de Enfermedades Infecciosas, Grupo de InfecciÃ³n Respiratoria y Resistencia Antimicrobiana. Instituto de InvestigaciÃ³n Sanitaria Biodonostia. | SeqCOVID-SPAIN consortium/IBV(CSIC) |
| EPI_ISL_452678 | Servicio de MicrobiologÃ-a. Hospital Universitario Donostia. OSI Donostialdea. Ã�ea de Enfermedades Infecciosas, Grupo de InfecciÃ³n Respiratoria y Resistencia Antimicrobiana. Instituto de InvestigaciÃ³n Sanitaria Biodonostia. | SeqCOVID-SPAIN consortium/IBV(CSIC) |
| EPI_ISL_452679 | Servicio de MicrobiologÃ-a. Hospital Universitario Donostia. OSI Donostialdea. Ã�ea de Enfermedades Infecciosas, Grupo de InfecciÃ³n Respiratoria y Resistencia Antimicrobiana. Instituto de InvestigaciÃ³n Sanitaria Biodonostia. | SeqCOVID-SPAIN consortium/IBV(CSIC) |
| EPI_ISL_452680 | Servicio de MicrobiologÃ-a. Hospital Universitario Donostia. OSI Donostialdea. Ã�ea de Enfermedades Infecciosas, Grupo de InfecciÃ³n Respiratoria y Resistencia Antimicrobiana. Instituto de InvestigaciÃ³n Sanitaria Biodonostia. | SeqCOVID-SPAIN consortium/IBV(CSIC) |
| EPI_ISL_452681 | Servicio de MicrobiologÃ-a. Hospital Universitario Donostia. OSI Donostialdea. Ã�ea de Enfermedades Infecciosas, Grupo de InfecciÃ³n Respiratoria y Resistencia Antimicrobiana. Instituto de InvestigaciÃ³n Sanitaria Biodonostia. | SeqCOVID-SPAIN consortium/IBV(CSIC) |
| EPI_ISL_452682 | Servicio de MicrobiologÃ-a. Hospital Universitario Donostia. OSI Donostialdea. Ã�ea de Enfermedades Infecciosas, Grupo de InfecciÃ³n Respiratoria y Resistencia Antimicrobiana. Instituto de InvestigaciÃ³n Sanitaria Biodonostia. | SeqCOVID-SPAIN consortium/IBV(CSIC) |

| EPI_Accession_ID | Originating Laboratory | Submitting Laboratory |
|---|---|---|
| EPI_ISL_452683 | Servicio de MicrobiologÃ-a. Hospital Universitario Donostia. OSI Donostialdea. Ã�ea de Enfermedades Infecciosas, Grupo de InfecciÃ³n Respiratoria y Resistencia Antimicrobiana. Instituto de InvestigaciÃ³n Sanitaria Biodonostia. | SeqCOVID-SPAIN consortium/IBV(CSIC) |
| EPI_ISL_452684 | Servicio de MicrobiologÃ-a. Hospital Universitario Donostia. OSI Donostialdea. Ã�ea de Enfermedades Infecciosas, Grupo de InfecciÃ³n Respiratoria y Resistencia Antimicrobiana. Instituto de InvestigaciÃ³n Sanitaria Biodonostia. | SeqCOVID-SPAIN consortium/IBV(CSIC) |
| EPI_ISL_452685 | Servicio de MicrobiologÃ-a. Hospital Universitario Donostia. OSI Donostialdea. Ã�ea de Enfermedades Infecciosas, Grupo de InfecciÃ³n Respiratoria y Resistencia Antimicrobiana. Instituto de InvestigaciÃ³n Sanitaria Biodonostia. | SeqCOVID-SPAIN consortium/IBV(CSIC) |
| EPI_ISL_452686 | Servicio de MicrobiologÃ-a. Hospital Universitario Donostia. OSI Donostialdea. Ã�ea de Enfermedades Infecciosas, Grupo de InfecciÃ³n Respiratoria y Resistencia Antimicrobiana. Instituto de InvestigaciÃ³n Sanitaria Biodonostia. | SeqCOVID-SPAIN consortium/IBV(CSIC) |
| EPI_ISL_452687 | Servicio de MicrobiologÃ-a. Hospital Universitario Donostia. OSI Donostialdea. Ã�ea de Enfermedades Infecciosas, Grupo de InfecciÃ³n Respiratoria y Resistencia Antimicrobiana. Instituto de InvestigaciÃ³n Sanitaria Biodonostia. | SeqCOVID-SPAIN consortium/IBV(CSIC) |
| EPI_ISL_452688 | Servicio de MicrobiologÃ-a. Hospital Universitario Donostia. OSI Donostialdea. Ã�ea de Enfermedades Infecciosas, Grupo de InfecciÃ³n Respiratoria y Resistencia Antimicrobiana. Instituto de InvestigaciÃ³n Sanitaria Biodonostia. | SeqCOVID-SPAIN consortium/IBV(CSIC) |

| EPI_Accession_ID | Originating Laboratory | Submitting Laboratory |
|---|---|---|
| EPI_ISL_452689 | Servicio de MicrobiologÃ-a. Hospital Universitario Donostia. OSI Donostialdea. Ã�ea de Enfermedades Infecciosas, Grupo de InfecciÃ³n Respiratoria y Resistencia Antimicrobiana. Instituto de InvestigaciÃ³n Sanitaria Biodonostia. | SeqCOVID-SPAIN consortium/IBV(CSIC) |
| EPI_ISL_452690 | Servicio de MicrobiologÃ-a. Hospital Universitario Donostia. OSI Donostialdea. Ã�ea de Enfermedades Infecciosas, Grupo de InfecciÃ³n Respiratoria y Resistencia Antimicrobiana. Instituto de InvestigaciÃ³n Sanitaria Biodonostia. | SeqCOVID-SPAIN consortium/IBV(CSIC) |
| EPI_ISL_452691 | Servicio de MicrobiologÃ-a. Hospital Universitario Donostia. OSI Donostialdea. Ã�ea de Enfermedades Infecciosas, Grupo de InfecciÃ³n Respiratoria y Resistencia Antimicrobiana. Instituto de InvestigaciÃ³n Sanitaria Biodonostia. | SeqCOVID-SPAIN consortium/IBV(CSIC) |
| EPI_ISL_452692 | Hospital Universitario Araba. Vitoria-Gasteiz, | SeqCOVID-SPAIN consortium/IBV(CSIC) |
| EPI_ISL_452693 | Hospital Universitario Araba. Vitoria-Gasteiz, | SeqCOVID-SPAIN consortium/IBV(CSIC) |
| EPI_ISL_452694 | Hospital Universitario Araba. Vitoria-Gasteiz, | SeqCOVID-SPAIN consortium/IBV(CSIC) |
| EPI_ISL_452695 | Hospital Universitario Araba. Vitoria-Gasteiz, | SeqCOVID-SPAIN consortium/IBV(CSIC) |
| EPI_ISL_452696 | Hospital Universitario Araba. Vitoria-Gasteiz, | SeqCOVID-SPAIN consortium/IBV(CSIC) |
| EPI_ISL_452697 | Hospital Universitario Araba. Vitoria-Gasteiz, | SeqCOVID-SPAIN consortium/IBV(CSIC) |
| EPI_ISL_452698 | Hospital Universitario Araba. Vitoria-Gasteiz, | SeqCOVID-SPAIN consortium/IBV(CSIC) |
| EPI_ISL_452699 | Hospital Universitario Araba. Vitoria-Gasteiz, | SeqCOVID-SPAIN consortium/IBV(CSIC) |
| EPI_ISL_452700 | Hospital Universitario Araba. Vitoria-Gasteiz, | SeqCOVID-SPAIN consortium/IBV(CSIC) |
| EPI_ISL_452701 | Hospital Universitario Araba. Vitoria-Gasteiz, | SeqCOVID-SPAIN consortium/IBV(CSIC) |
| EPI_ISL_452702 | Hospital Universitario Araba. Vitoria-Gasteiz, | SeqCOVID-SPAIN consortium/IBV(CSIC) |
| EPI_ISL_452703 | Hospital Universitario Araba. Vitoria-Gasteiz, | SeqCOVID-SPAIN consortium/IBV(CSIC) |
| EPI_ISL_452704 | Hospital Universitario Araba. Vitoria-Gasteiz, | SeqCOVID-SPAIN consortium/IBV(CSIC) |
| EPI_ISL_452705 | Hospital Universitario Araba. Vitoria-Gasteiz, | SeqCOVID-SPAIN consortium/IBV(CSIC) |
| EPI_ISL_452706 | Hospital Universitario Araba. Vitoria-Gasteiz, | SeqCOVID-SPAIN consortium/IBV(CSIC) |
| EPI_ISL_452707 | Hospital Universitario Araba. Vitoria-Gasteiz, | SeqCOVID-SPAIN consortium/IBV(CSIC) |
| EPI_ISL_452708 | Hospital Universitario Araba. Vitoria-Gasteiz, | SeqCOVID-SPAIN consortium/IBV(CSIC) |
| EPI_ISL_452709 | Hospital Universitario Araba. Vitoria-Gasteiz, | SeqCOVID-SPAIN consortium/IBV(CSIC) |

| EPI_Accession_ID | Originating Laboratory | Submitting Laboratory |
|---|---|---|
| EPI_ISL_452710 | Hospital Universitario Araba. Vitoria-Gasteiz, | SeqCOVID-SPAIN consortium/IBV(CSIC) |
| EPI_ISL_452711 | Hospital Universitario Araba. Vitoria-Gasteiz, | SeqCOVID-SPAIN consortium/IBV(CSIC) |
| EPI_ISL_452712 | Hospital Universitario Araba. Vitoria-Gasteiz, | SeqCOVID-SPAIN consortium/IBV(CSIC) |
| EPI_ISL_452713 | Hospital Universitario Araba. Vitoria-Gasteiz, | SeqCOVID-SPAIN consortium/IBV(CSIC) |
| EPI_ISL_452714 | Hospital Universitario Araba. Vitoria-Gasteiz, | SeqCOVID-SPAIN consortium/IBV(CSIC) |
| EPI_ISL_452715 | Hospital Universitario Araba. Vitoria-Gasteiz, | SeqCOVID-SPAIN consortium/IBV(CSIC) |
| EPI_ISL_452716 | Hospital Universitario Araba. Vitoria-Gasteiz, | SeqCOVID-SPAIN consortium/IBV(CSIC) |
| EPI_ISL_452717 | Hospital Universitario Araba. Vitoria-Gasteiz, | SeqCOVID-SPAIN consortium/IBV(CSIC) |
| EPI_ISL_452718 | Hospital Universitario Araba. Vitoria-Gasteiz, | SeqCOVID-SPAIN consortium/IBV(CSIC) |
| EPI_ISL_452719 | Hospital Universitario Araba. Vitoria-Gasteiz, | SeqCOVID-SPAIN consortium/IBV(CSIC) |
| EPI_ISL_452720 | Hospital Universitario Araba. Vitoria-Gasteiz, | SeqCOVID-SPAIN consortium/IBV(CSIC) |
| EPI_ISL_452721 | Hospital Universitario Araba. Vitoria-Gasteiz, | SeqCOVID-SPAIN consortium/IBV(CSIC) |
| EPI_ISL_452722 | Hospital Universitario Araba. Vitoria-Gasteiz, | SeqCOVID-SPAIN consortium/IBV(CSIC) |
| EPI_ISL_452723 | Hospital Universitario Araba. Vitoria-Gasteiz, | SeqCOVID-SPAIN consortium/IBV(CSIC) |
| EPI_ISL_452724 | Hospital Universitario Araba. Vitoria-Gasteiz, | SeqCOVID-SPAIN consortium/IBV(CSIC) |
| EPI_ISL_452725 | Hospital Universitario Araba. Vitoria-Gasteiz, | SeqCOVID-SPAIN consortium/IBV(CSIC) |
| EPI_ISL_452726 | Hospital Universitario Araba. Vitoria-Gasteiz, | SeqCOVID-SPAIN consortium/IBV(CSIC) |
| EPI_ISL_452727 | Hospital Universitario Araba. Vitoria-Gasteiz, | SeqCOVID-SPAIN consortium/IBV(CSIC) |
| EPI_ISL_452728 | Hospital Universitario Araba. Vitoria-Gasteiz, | SeqCOVID-SPAIN consortium/IBV(CSIC) |
| EPI_ISL_452729 | Hospital Universitario Araba. Vitoria-Gasteiz, | SeqCOVID-SPAIN consortium/IBV(CSIC) |
| EPI_ISL_452730 | Hospital Universitario Araba. Vitoria-Gasteiz, | SeqCOVID-SPAIN consortium/IBV(CSIC) |
| EPI_ISL_452731 | Hospital Universitario Araba. Vitoria-Gasteiz, | SeqCOVID-SPAIN consortium/IBV(CSIC) |
| EPI_ISL_452732 | Hospital Universitario Araba. Vitoria-Gasteiz, | SeqCOVID-SPAIN consortium/IBV(CSIC) |
| EPI_ISL_452733 | Hospital Universitario Araba. Vitoria-Gasteiz, | SeqCOVID-SPAIN consortium/IBV(CSIC) |
| EPI_ISL_452734 | Hospital Universitario Araba. Vitoria-Gasteiz, | SeqCOVID-SPAIN consortium/IBV(CSIC) |
| EPI_ISL_452735 | Hospital Universitario Araba. Vitoria-Gasteiz, | SeqCOVID-SPAIN consortium/IBV(CSIC) |
| EPI_ISL_452736 | Hospital Universitario Araba. Vitoria-Gasteiz, | SeqCOVID-SPAIN consortium/IBV(CSIC) |
| EPI_ISL_452737 | Hospital Universitario Araba. Vitoria-Gasteiz, | SeqCOVID-SPAIN consortium/IBV(CSIC) |
| EPI_ISL_452738 | Hospital Universitario Araba. Vitoria-Gasteiz, | SeqCOVID-SPAIN consortium/IBV(CSIC) |
| EPI_ISL_452739 | Hospital Universitario Araba. Vitoria-Gasteiz, | SeqCOVID-SPAIN consortium/IBV(CSIC) |
| EPI_ISL_452740 | Hospital Universitario Araba. Vitoria-Gasteiz, | SeqCOVID-SPAIN consortium/IBV(CSIC) |
| EPI_ISL_452741 | Hospital Universitario Araba. Vitoria-Gasteiz, | SeqCOVID-SPAIN consortium/IBV(CSIC) |
| EPI_ISL_452742 | Hospital Universitario Araba. Vitoria-Gasteiz, | SeqCOVID-SPAIN consortium/IBV(CSIC) |

| EPI_Accession_ID | Originating Laboratory | Submitting Laboratory |
|---|---|---|
| EPI_ISL_452743 | Hospital Universitario Araba. Vitoria-Gasteiz, | SeqCOVID-SPAIN consortium/IBV(CSIC) |
| EPI_ISL_452744 | Hospital Universitario Araba. Vitoria-Gasteiz, | SeqCOVID-SPAIN consortium/IBV(CSIC) |
| EPI_ISL_452745 | Hospital Universitario Araba. Vitoria-Gasteiz, | SeqCOVID-SPAIN consortium/IBV(CSIC) |
| EPI_ISL_452746 | Hospital Universitario Araba. Vitoria-Gasteiz, | SeqCOVID-SPAIN consortium/IBV(CSIC) |
| EPI_ISL_452747 | Hospital Universitario Araba. Vitoria-Gasteiz, | SeqCOVID-SPAIN consortium/IBV(CSIC) |
| EPI_ISL_452748 | Hospital Universitario Araba. Vitoria-Gasteiz, | SeqCOVID-SPAIN consortium/IBV(CSIC) |
| EPI_ISL_452749 | Hospital Universitario Araba. Vitoria-Gasteiz, | SeqCOVID-SPAIN consortium/IBV(CSIC) |
| EPI_ISL_452750 | Hospital Universitario Araba. Vitoria-Gasteiz, | SeqCOVID-SPAIN consortium/IBV(CSIC) |
| EPI_ISL_452751 | Hospital Universitario Araba. Vitoria-Gasteiz, | SeqCOVID-SPAIN consortium/IBV(CSIC) |
| EPI_ISL_452752 | Hospital Universitario Araba. Vitoria-Gasteiz, | SeqCOVID-SPAIN consortium/IBV(CSIC) |
| EPI_ISL_452753 | Hospital Universitario Araba. Vitoria-Gasteiz, | SeqCOVID-SPAIN consortium/IBV(CSIC) |
| EPI_ISL_452754 | Hospital Universitario Araba. Vitoria-Gasteiz, | SeqCOVID-SPAIN consortium/IBV(CSIC) |
| EPI_ISL_452755 | Hospital Universitario Araba. Vitoria-Gasteiz, | SeqCOVID-SPAIN consortium/IBV(CSIC) |
| EPI_ISL_452756 | Hospital Universitario Araba. Vitoria-Gasteiz, | SeqCOVID-SPAIN consortium/IBV(CSIC) |
| EPI_ISL_452757 | Hospital Universitario Araba. Vitoria-Gasteiz, | SeqCOVID-SPAIN consortium/IBV(CSIC) |
| EPI_ISL_452758 | Hospital Universitario Araba. Vitoria-Gasteiz, | SeqCOVID-SPAIN consortium/IBV(CSIC) |
| EPI_ISL_452759 | Hospital Universitario Araba. Vitoria-Gasteiz, | SeqCOVID-SPAIN consortium/IBV(CSIC) |
| EPI_ISL_452760 | Hospital Universitario Araba. Vitoria-Gasteiz, | SeqCOVID-SPAIN consortium/IBV(CSIC) |
| EPI_ISL_452761 | Hospital Universitario Araba. Vitoria-Gasteiz, | SeqCOVID-SPAIN consortium/IBV(CSIC) |
| EPI_ISL_452762 | Hospital Universitario Araba. Vitoria-Gasteiz, | SeqCOVID-SPAIN consortium/IBV(CSIC) |
| EPI_ISL_452763 | Hospital Universitario Araba. Vitoria-Gasteiz, | SeqCOVID-SPAIN consortium/IBV(CSIC) |
| EPI_ISL_452764 | Hospital Universitario Araba. Vitoria-Gasteiz, | SeqCOVID-SPAIN consortium/IBV(CSIC) |
| EPI_ISL_452765 | Hospital Universitario Araba. Vitoria-Gasteiz, | SeqCOVID-SPAIN consortium/IBV(CSIC) |
| EPI_ISL_452766 | Hospital Universitario Araba. Vitoria-Gasteiz, | SeqCOVID-SPAIN consortium/IBV(CSIC) |
| EPI_ISL_452767 | Hospital Universitario Araba. Vitoria-Gasteiz, | SeqCOVID-SPAIN consortium/IBV(CSIC) |
| EPI_ISL_452768 | Hospital Universitario Araba. Vitoria-Gasteiz, | SeqCOVID-SPAIN consortium/IBV(CSIC) |
| EPI_ISL_452769 | Hospital Universitario Araba. Vitoria-Gasteiz, | SeqCOVID-SPAIN consortium/IBV(CSIC) |
| EPI_ISL_452770 | Hospital Universitario Araba. Vitoria-Gasteiz, | SeqCOVID-SPAIN consortium/IBV(CSIC) |
| EPI_ISL_452771 | Hospital Universitario Araba. Vitoria-Gasteiz, | SeqCOVID-SPAIN consortium/IBV(CSIC) |
| EPI_ISL_452772 | Hospital Universitario Araba. Vitoria-Gasteiz, | SeqCOVID-SPAIN consortium/IBV(CSIC) |
| EPI_ISL_452773 | Hospital Universitario Araba. Vitoria-Gasteiz, | SeqCOVID-SPAIN consortium/IBV(CSIC) |
| EPI_ISL_452774 | Hospital Universitario Araba. Vitoria-Gasteiz, | SeqCOVID-SPAIN consortium/IBV(CSIC) |
| EPI_ISL_452775 | Hospital Universitario Araba. Vitoria-Gasteiz, | SeqCOVID-SPAIN consortium/IBV(CSIC) |

| EPI_Accession_ID | Originating Laboratory | Submitting Laboratory |
|---|---|---|
| EPI_ISL_452776 | Hospital Universitario Araba. Vitoria-Gasteiz, | SeqCOVID-SPAIN consortium/IBV(CSIC) |
| EPI_ISL_452777 | Hospital Universitario Araba. Vitoria-Gasteiz, | SeqCOVID-SPAIN consortium/IBV(CSIC) |
| EPI_ISL_452778 | Hospital Universitario Araba. Vitoria-Gasteiz, | SeqCOVID-SPAIN consortium/IBV(CSIC) |
| EPI_ISL_452779 | Hospital Universitario Araba. Vitoria-Gasteiz, | SeqCOVID-SPAIN consortium/IBV(CSIC) |
| EPI_ISL_452780 | Hospital Universitario Araba. Vitoria-Gasteiz, | SeqCOVID-SPAIN consortium/IBV(CSIC) |
| EPI_ISL_452781 | Hospital Universitario Araba. Vitoria-Gasteiz, | SeqCOVID-SPAIN consortium/IBV(CSIC) |
| EPI_ISL_452782 | Hospital Universitario Araba. Vitoria-Gasteiz, | SeqCOVID-SPAIN consortium/IBV(CSIC) |
| EPI_ISL_452783 | Hospital Universitario Araba. Vitoria-Gasteiz, | SeqCOVID-SPAIN consortium/IBV(CSIC) |
| EPI_ISL_452784 | Hospital Universitario Araba. Vitoria-Gasteiz, | SeqCOVID-SPAIN consortium/IBV(CSIC) |
| EPI_ISL_452785 | Hospital Universitario Araba. Vitoria-Gasteiz, | SeqCOVID-SPAIN consortium/IBV(CSIC) |
| EPI_ISL_452786 | Hospital Universitario Araba. Vitoria-Gasteiz, | SeqCOVID-SPAIN consortium/IBV(CSIC) |
| EPI_ISL_452787 | ICAR-National Institute of High Security Animal Diseases | ICAR-National Institute of High Security Animal Diseases |
| EPI_ISL_452788 | ICAR-National Institute of High Security Animal Diseases | ICAR-National Institute of High Security Animal Diseases |
| EPI_ISL_452789 | ICAR-National Institute of High Security Animal Diseases | ICAR-National Institute of High Security Animal Diseases |
| EPI_ISL_452790 | ICAR-National Institute of High Security Animal Diseases | ICAR-National Institute of High Security Animal Diseases |
| EPI_ISL_452791 | ICAR-National Institute of High Security Animal Diseases | ICAR-National Institute of High Security Animal Diseases |
| EPI_ISL_452792 | ICAR-National Institute of High Security Animal Diseases | ICAR-National Institute of High Security Animal Diseases |
| EPI_ISL_452793 | ICAR-National Institute of High Security Animal Diseases | ICAR-National Institute of High Security Animal Diseases |
| EPI_ISL_452794 | ICAR-National Institute of High Security Animal Diseases | ICAR-National Institute of High Security Animal Diseases |
| EPI_ISL_452795 | ICAR-National Institute of High Security Animal Diseases | ICAR-National Institute of High Security Animal Diseases |
| EPI_ISL_452796 | Virginia DCLS | Virginia DCLS |
| EPI_ISL_452797 | Virginia DCLS | Virginia DCLS |
| EPI_ISL_452798 | Virginia DCLS | Virginia DCLS |
| EPI_ISL_452799 | Virginia DCLS | Virginia DCLS |

| EPI_Accession_ID | Originating Laboratory | Submitting Laboratory |
|---|---|---|
| EPI_ISL_452800 | Virginia DCLS | Virginia DCLS |
| EPI_ISL_452801 | Virginia DCLS | Virginia DCLS |
| EPI_ISL_452802 | Virginia DCLS | Virginia DCLS |
| EPI_ISL_452803 | Virginia DCLS | Virginia DCLS |
| EPI_ISL_452804 | Virginia DCLS | Virginia DCLS |
| EPI_ISL_452805 | Virginia DCLS | Virginia DCLS |
| EPI_ISL_452806 | Virginia DCLS | Virginia DCLS |
| EPI_ISL_452807 | Virginia DCLS | Virginia DCLS |
| EPI_ISL_452808 | Virginia DCLS | Virginia DCLS |
| EPI_ISL_452809 | Virginia DCLS | Virginia DCLS |
| EPI_ISL_452810 | Virginia DCLS | Virginia DCLS |
| EPI_ISL_452811 | Virginia DCLS | Virginia DCLS |
| EPI_ISL_452812 | Virginia DCLS | Virginia DCLS |
| EPI_ISL_452813 | Virginia DCLS | Virginia DCLS |
| EPI_ISL_452814 | Virginia DCLS | Virginia DCLS |
| EPI_ISL_452815 | Virginia DCLS | Virginia DCLS |
| EPI_ISL_452816 | Virginia DCLS | Virginia DCLS |
| EPI_ISL_452817 | Virginia DCLS | Virginia DCLS |
| EPI_ISL_452818 | Virginia DCLS | Virginia DCLS |
| EPI_ISL_452819 | Virginia DCLS | Virginia DCLS |
| EPI_ISL_452820 | Virginia DCLS | Virginia DCLS |
| EPI_ISL_452821 | Virginia DCLS | Virginia DCLS |
| EPI_ISL_452822 | Virginia DCLS | Virginia DCLS |
| EPI_ISL_452823 | Virginia DCLS | Virginia DCLS |
| EPI_ISL_452824 | Virginia DCLS | Virginia DCLS |
| EPI_ISL_452825 | Virginia DCLS | Virginia DCLS |
| EPI_ISL_452826 | Virginia DCLS | Virginia DCLS |
| EPI_ISL_452827 | Virginia DCLS | Virginia DCLS |
| EPI_ISL_452828 | Virginia DCLS | Virginia DCLS |
| EPI_ISL_452829 | Virginia DCLS | Virginia DCLS |
| EPI_ISL_452830 | Virginia DCLS | Virginia DCLS |
| EPI_ISL_452831 | Virginia DCLS | Virginia DCLS |
| EPI_ISL_452832 | Virginia DCLS | Virginia DCLS |

| EPI_Accession_ID | Originating Laboratory | Submitting Laboratory |
|---|---|---|
| EPI_ISL_452833 | Virginia DCLS | Virginia DCLS |
| EPI_ISL_452834 | Virginia DCLS | Virginia DCLS |
| EPI_ISL_452835 | Virginia DCLS | Virginia DCLS |
| EPI_ISL_452836 | Virginia DCLS | Virginia DCLS |
| EPI_ISL_452837 | Virginia DCLS | Virginia DCLS |
| EPI_ISL_452838 | Virginia DCLS | Virginia DCLS |
| EPI_ISL_452839 | Virginia DCLS | Virginia DCLS |
| EPI_ISL_452840 | Virginia DCLS | Virginia DCLS |
| EPI_ISL_452841 | Virginia DCLS | Virginia DCLS |
| EPI_ISL_452842 | Virginia DCLS | Virginia DCLS |
| EPI_ISL_452843 | Virginia DCLS | Virginia DCLS |
| EPI_ISL_452844 | Virginia DCLS | Virginia DCLS |
| EPI_ISL_452845 | Virginia DCLS | Virginia DCLS |
| EPI_ISL_452846 | Virginia DCLS | Virginia DCLS |
| EPI_ISL_452847 | Virginia DCLS | Virginia DCLS |
| EPI_ISL_452848 | Virginia DCLS | Virginia DCLS |
| EPI_ISL_452849 | Virginia DCLS | Virginia DCLS |
| EPI_ISL_452850 | Virginia DCLS | Virginia DCLS |
| EPI_ISL_452851 | Virginia DCLS | Virginia DCLS |
| EPI_ISL_452852 | Virginia DCLS | Virginia DCLS |
| EPI_ISL_452853 | Virginia DCLS | Virginia DCLS |
| EPI_ISL_452854 | Virginia DCLS | Virginia DCLS |
| EPI_ISL_452855 | Virginia DCLS | Virginia DCLS |
| EPI_ISL_452856 | Virginia DCLS | Virginia DCLS |
| EPI_ISL_452857 | Department of Pathology, University of Cambridge | COVID-19 Genomics UK (COG-UK) Consortium |
| EPI_ISL_452858 | Department of Pathology, University of Cambridge | COVID-19 Genomics UK (COG-UK) Consortium |
| EPI_ISL_452859 | Department of Pathology, University of Cambridge | COVID-19 Genomics UK (COG-UK) Consortium |
| EPI_ISL_452860 | Department of Pathology, University of Cambridge | COVID-19 Genomics UK (COG-UK) Consortium |
| EPI_ISL_452861 | Department of Pathology, University of Cambridge | COVID-19 Genomics UK (COG-UK) Consortium |
| EPI_ISL_452862 | Department of Pathology, University of Cambridge | COVID-19 Genomics UK (COG-UK) Consortium |
| EPI_ISL_452863 | Department of Pathology, University of Cambridge | COVID-19 Genomics UK (COG-UK) Consortium |
| EPI_ISL_452864 | Department of Pathology, University of Cambridge | COVID-19 Genomics UK (COG-UK) Consortium |
| EPI_ISL_452865 | Department of Pathology, University of Cambridge | COVID-19 Genomics UK (COG-UK) Consortium |

| EPI_Accession_ID | Originating Laboratory | Submitting Laboratory |
| --- | --- | --- |
| EPI_ISL_452866 | Department of Pathology, University of Cambridge | COVID-19 Genomics UK (COG-UK) Consortium |
| EPI_ISL_452867 | Department of Pathology, University of Cambridge | COVID-19 Genomics UK (COG-UK) Consortium |
| EPI_ISL_452868 | Department of Pathology, University of Cambridge | COVID-19 Genomics UK (COG-UK) Consortium |
| EPI_ISL_452869 | Department of Pathology, University of Cambridge | COVID-19 Genomics UK (COG-UK) Consortium |
| EPI_ISL_452870 | Department of Pathology, University of Cambridge | COVID-19 Genomics UK (COG-UK) Consortium |
| EPI_ISL_452871 | Department of Pathology, University of Cambridge | COVID-19 Genomics UK (COG-UK) Consortium |
| EPI_ISL_452872 | Department of Pathology, University of Cambridge | COVID-19 Genomics UK (COG-UK) Consortium |
| EPI_ISL_452873 | Department of Pathology, University of Cambridge | COVID-19 Genomics UK (COG-UK) Consortium |
| EPI_ISL_452874 | Department of Pathology, University of Cambridge | COVID-19 Genomics UK (COG-UK) Consortium |
| EPI_ISL_452875 | Department of Pathology, University of Cambridge | COVID-19 Genomics UK (COG-UK) Consortium |
| EPI_ISL_452876 | Department of Pathology, University of Cambridge | COVID-19 Genomics UK (COG-UK) Consortium |
| EPI_ISL_452877 | Department of Pathology, University of Cambridge | COVID-19 Genomics UK (COG-UK) Consortium |
| EPI_ISL_452878 | Department of Pathology, University of Cambridge | COVID-19 Genomics UK (COG-UK) Consortium |
| EPI_ISL_452879 | Department of Pathology, University of Cambridge | COVID-19 Genomics UK (COG-UK) Consortium |
| EPI_ISL_452880 | Department of Pathology, University of Cambridge | COVID-19 Genomics UK (COG-UK) Consortium |
| EPI_ISL_452881 | Department of Pathology, University of Cambridge | COVID-19 Genomics UK (COG-UK) Consortium |
| EPI_ISL_452882 | Department of Pathology, University of Cambridge | COVID-19 Genomics UK (COG-UK) Consortium |
| EPI_ISL_452883 | Department of Pathology, University of Cambridge | COVID-19 Genomics UK (COG-UK) Consortium |
| EPI_ISL_452884 | Department of Pathology, University of Cambridge | COVID-19 Genomics UK (COG-UK) Consortium |
| EPI_ISL_452885 | Department of Pathology, University of Cambridge | COVID-19 Genomics UK (COG-UK) Consortium |
| EPI_ISL_452886 | Department of Pathology, University of Cambridge | COVID-19 Genomics UK (COG-UK) Consortium |
| EPI_ISL_452887 | Department of Pathology, University of Cambridge | COVID-19 Genomics UK (COG-UK) Consortium |
| EPI_ISL_452888 | Department of Pathology, University of Cambridge | COVID-19 Genomics UK (COG-UK) Consortium |
| EPI_ISL_452889 | Department of Pathology, University of Cambridge | COVID-19 Genomics UK (COG-UK) Consortium |
| EPI_ISL_452890 | Department of Pathology, University of Cambridge | COVID-19 Genomics UK (COG-UK) Consortium |
| EPI_ISL_452891 | Department of Pathology, University of Cambridge | COVID-19 Genomics UK (COG-UK) Consortium |
| EPI_ISL_452892 | Department of Pathology, University of Cambridge | COVID-19 Genomics UK (COG-UK) Consortium |
| EPI_ISL_452893 | Department of Pathology, University of Cambridge | COVID-19 Genomics UK (COG-UK) Consortium |
| EPI_ISL_452894 | Department of Pathology, University of Cambridge | COVID-19 Genomics UK (COG-UK) Consortium |
| EPI_ISL_452895 | Department of Pathology, University of Cambridge | COVID-19 Genomics UK (COG-UK) Consortium |
| EPI_ISL_452896 | Department of Pathology, University of Cambridge | COVID-19 Genomics UK (COG-UK) Consortium |
| EPI_ISL_452897 | Department of Pathology, University of Cambridge | COVID-19 Genomics UK (COG-UK) Consortium |
| EPI_ISL_452898 | Department of Pathology, University of Cambridge | COVID-19 Genomics UK (COG-UK) Consortium |

| EPI_Accession_ID | Originating Laboratory | Submitting Laboratory |
| --- | --- | --- |
| EPI_ISL_452899 | Department of Pathology, University of Cambridge | COVID-19 Genomics UK (COG-UK) Consortium |
| EPI_ISL_452900 | Department of Pathology, University of Cambridge | COVID-19 Genomics UK (COG-UK) Consortium |
| EPI_ISL_452901 | Department of Pathology, University of Cambridge | COVID-19 Genomics UK (COG-UK) Consortium |
| EPI_ISL_452903 | Department of Pathology, University of Cambridge | COVID-19 Genomics UK (COG-UK) Consortium |
| EPI_ISL_452904 | Department of Pathology, University of Cambridge | COVID-19 Genomics UK (COG-UK) Consortium |
| EPI_ISL_452905 | Department of Pathology, University of Cambridge | COVID-19 Genomics UK (COG-UK) Consortium |
| EPI_ISL_452906 | Department of Pathology, University of Cambridge | COVID-19 Genomics UK (COG-UK) Consortium |
| EPI_ISL_452910 | Department of Pathology, University of Cambridge | COVID-19 Genomics UK (COG-UK) Consortium |
| EPI_ISL_452911 | Department of Pathology, University of Cambridge | COVID-19 Genomics UK (COG-UK) Consortium |
| EPI_ISL_452912 | Department of Pathology, University of Cambridge | COVID-19 Genomics UK (COG-UK) Consortium |
| EPI_ISL_452913 | Department of Pathology, University of Cambridge | COVID-19 Genomics UK (COG-UK) Consortium |
| EPI_ISL_452914 | Department of Pathology, University of Cambridge | COVID-19 Genomics UK (COG-UK) Consortium |
| EPI_ISL_452915 | Department of Pathology, University of Cambridge | COVID-19 Genomics UK (COG-UK) Consortium |
| EPI_ISL_452916 | Department of Pathology, University of Cambridge | COVID-19 Genomics UK (COG-UK) Consortium |
| EPI_ISL_452917 | Department of Pathology, University of Cambridge | COVID-19 Genomics UK (COG-UK) Consortium |
| EPI_ISL_452918 | Department of Pathology, University of Cambridge | COVID-19 Genomics UK (COG-UK) Consortium |
| EPI_ISL_452919 | Department of Pathology, University of Cambridge | COVID-19 Genomics UK (COG-UK) Consortium |
| EPI_ISL_452920 | Department of Pathology, University of Cambridge | COVID-19 Genomics UK (COG-UK) Consortium |
| EPI_ISL_452921 | Department of Pathology, University of Cambridge | COVID-19 Genomics UK (COG-UK) Consortium |
| EPI_ISL_452922 | Department of Pathology, University of Cambridge | COVID-19 Genomics UK (COG-UK) Consortium |
| EPI_ISL_452923 | Department of Pathology, University of Cambridge | COVID-19 Genomics UK (COG-UK) Consortium |
| EPI_ISL_452924 | Department of Pathology, University of Cambridge | COVID-19 Genomics UK (COG-UK) Consortium |
| EPI_ISL_452925 | Department of Pathology, University of Cambridge | COVID-19 Genomics UK (COG-UK) Consortium |
| EPI_ISL_452926 | Department of Pathology, University of Cambridge | COVID-19 Genomics UK (COG-UK) Consortium |
| EPI_ISL_452927 | Department of Pathology, University of Cambridge | COVID-19 Genomics UK (COG-UK) Consortium |
| EPI_ISL_452928 | Department of Pathology, University of Cambridge | COVID-19 Genomics UK (COG-UK) Consortium |
| EPI_ISL_452929 | Department of Pathology, University of Cambridge | COVID-19 Genomics UK (COG-UK) Consortium |
| EPI_ISL_452930 | Department of Pathology, University of Cambridge | COVID-19 Genomics UK (COG-UK) Consortium |
| EPI_ISL_452932 | Department of Pathology, University of Cambridge | COVID-19 Genomics UK (COG-UK) Consortium |
| EPI_ISL_452933 | Department of Pathology, University of Cambridge | COVID-19 Genomics UK (COG-UK) Consortium |
| EPI_ISL_452934 | Department of Pathology, University of Cambridge | COVID-19 Genomics UK (COG-UK) Consortium |
| EPI_ISL_452935 | Department of Pathology, University of Cambridge | COVID-19 Genomics UK (COG-UK) Consortium |
| EPI_ISL_452936 | Department of Pathology, University of Cambridge | COVID-19 Genomics UK (COG-UK) Consortium |

| EPI_Accession_ID | Originating Laboratory | Submitting Laboratory |
|---|---|---|
| EPI_ISL_452937 | Department of Pathology, University of Cambridge | COVID-19 Genomics UK (COG-UK) Consortium |
| EPI_ISL_452938 | Department of Pathology, University of Cambridge | COVID-19 Genomics UK (COG-UK) Consortium |
| EPI_ISL_452939 | Department of Pathology, University of Cambridge | COVID-19 Genomics UK (COG-UK) Consortium |
| EPI_ISL_452940 | Department of Pathology, University of Cambridge | COVID-19 Genomics UK (COG-UK) Consortium |
| EPI_ISL_452941 | Department of Pathology, University of Cambridge | COVID-19 Genomics UK (COG-UK) Consortium |
| EPI_ISL_452942 | Department of Pathology, University of Cambridge | COVID-19 Genomics UK (COG-UK) Consortium |
| EPI_ISL_452943 | Department of Pathology, University of Cambridge | COVID-19 Genomics UK (COG-UK) Consortium |
| EPI_ISL_452944 | Department of Pathology, University of Cambridge | COVID-19 Genomics UK (COG-UK) Consortium |
| EPI_ISL_452945 | Department of Pathology, University of Cambridge | COVID-19 Genomics UK (COG-UK) Consortium |
| EPI_ISL_452946 | Department of Pathology, University of Cambridge | COVID-19 Genomics UK (COG-UK) Consortium |
| EPI_ISL_452947 | Department of Pathology, University of Cambridge | COVID-19 Genomics UK (COG-UK) Consortium |
| EPI_ISL_452948 | Department of Pathology, University of Cambridge | COVID-19 Genomics UK (COG-UK) Consortium |
| EPI_ISL_452949 | Department of Pathology, University of Cambridge | COVID-19 Genomics UK (COG-UK) Consortium |
| EPI_ISL_452951 | Department of Pathology, University of Cambridge | COVID-19 Genomics UK (COG-UK) Consortium |
| EPI_ISL_452952 | Department of Pathology, University of Cambridge | COVID-19 Genomics UK (COG-UK) Consortium |
| EPI_ISL_452953 | Department of Pathology, University of Cambridge | COVID-19 Genomics UK (COG-UK) Consortium |
| EPI_ISL_452954 | Department of Pathology, University of Cambridge | COVID-19 Genomics UK (COG-UK) Consortium |
| EPI_ISL_452956 | Department of Pathology, University of Cambridge | COVID-19 Genomics UK (COG-UK) Consortium |
| EPI_ISL_452957 | Department of Pathology, University of Cambridge | COVID-19 Genomics UK (COG-UK) Consortium |
| EPI_ISL_452958 | Department of Pathology, University of Cambridge | COVID-19 Genomics UK (COG-UK) Consortium |
| EPI_ISL_452959 | Department of Pathology, University of Cambridge | COVID-19 Genomics UK (COG-UK) Consortium |
| EPI_ISL_452960 | Department of Pathology, University of Cambridge | COVID-19 Genomics UK (COG-UK) Consortium |
| EPI_ISL_452961 | Department of Pathology, University of Cambridge | COVID-19 Genomics UK (COG-UK) Consortium |
| EPI_ISL_452962 | Department of Pathology, University of Cambridge | COVID-19 Genomics UK (COG-UK) Consortium |
| EPI_ISL_452963 | Department of Pathology, University of Cambridge | COVID-19 Genomics UK (COG-UK) Consortium |
| EPI_ISL_452964 | Department of Pathology, University of Cambridge | COVID-19 Genomics UK (COG-UK) Consortium |
| EPI_ISL_452965 | Department of Pathology, University of Cambridge | COVID-19 Genomics UK (COG-UK) Consortium |
| EPI_ISL_452966 | Department of Pathology, University of Cambridge | COVID-19 Genomics UK (COG-UK) Consortium |
| EPI_ISL_452967 | Department of Pathology, University of Cambridge | COVID-19 Genomics UK (COG-UK) Consortium |
| EPI_ISL_452968 | Department of Pathology, University of Cambridge | COVID-19 Genomics UK (COG-UK) Consortium |
| EPI_ISL_452969 | Department of Pathology, University of Cambridge | COVID-19 Genomics UK (COG-UK) Consortium |
| EPI_ISL_452970 | Department of Pathology, University of Cambridge | COVID-19 Genomics UK (COG-UK) Consortium |
| EPI_ISL_452971 | Department of Pathology, University of Cambridge | COVID-19 Genomics UK (COG-UK) Consortium |

| EPI_Accession_ID | Originating Laboratory | Submitting Laboratory |
|---|---|---|
| EPI_ISL_452972 | Department of Pathology, University of Cambridge | COVID-19 Genomics UK (COG-UK) Consortium |
| EPI_ISL_452973 | Department of Pathology, University of Cambridge | COVID-19 Genomics UK (COG-UK) Consortium |
| EPI_ISL_452974 | Department of Pathology, University of Cambridge | COVID-19 Genomics UK (COG-UK) Consortium |
| EPI_ISL_452975 | Department of Pathology, University of Cambridge | COVID-19 Genomics UK (COG-UK) Consortium |
| EPI_ISL_452976 | Department of Pathology, University of Cambridge | COVID-19 Genomics UK (COG-UK) Consortium |
| EPI_ISL_452977 | Department of Pathology, University of Cambridge | COVID-19 Genomics UK (COG-UK) Consortium |
| EPI_ISL_452978 | Department of Pathology, University of Cambridge | COVID-19 Genomics UK (COG-UK) Consortium |
| EPI_ISL_452979 | Department of Pathology, University of Cambridge | COVID-19 Genomics UK (COG-UK) Consortium |
| EPI_ISL_452980 | Department of Pathology, University of Cambridge | COVID-19 Genomics UK (COG-UK) Consortium |
| EPI_ISL_452981 | Department of Pathology, University of Cambridge | COVID-19 Genomics UK (COG-UK) Consortium |
| EPI_ISL_452982 | Department of Pathology, University of Cambridge | COVID-19 Genomics UK (COG-UK) Consortium |
| EPI_ISL_452983 | Department of Pathology, University of Cambridge | COVID-19 Genomics UK (COG-UK) Consortium |
| EPI_ISL_452984 | Department of Pathology, University of Cambridge | COVID-19 Genomics UK (COG-UK) Consortium |
| EPI_ISL_452985 | Department of Pathology, University of Cambridge | COVID-19 Genomics UK (COG-UK) Consortium |
| EPI_ISL_452986 | Department of Pathology, University of Cambridge | COVID-19 Genomics UK (COG-UK) Consortium |
| EPI_ISL_452987 | Department of Pathology, University of Cambridge | COVID-19 Genomics UK (COG-UK) Consortium |
| EPI_ISL_452988 | Department of Pathology, University of Cambridge | COVID-19 Genomics UK (COG-UK) Consortium |
| EPI_ISL_452989 | Department of Pathology, University of Cambridge | COVID-19 Genomics UK (COG-UK) Consortium |
| EPI_ISL_452990 | Department of Pathology, University of Cambridge | COVID-19 Genomics UK (COG-UK) Consortium |
| EPI_ISL_452991 | Department of Pathology, University of Cambridge | COVID-19 Genomics UK (COG-UK) Consortium |
| EPI_ISL_452992 | Department of Pathology, University of Cambridge | COVID-19 Genomics UK (COG-UK) Consortium |
| EPI_ISL_452993 | Department of Pathology, University of Cambridge | COVID-19 Genomics UK (COG-UK) Consortium |
| EPI_ISL_452994 | Department of Pathology, University of Cambridge | COVID-19 Genomics UK (COG-UK) Consortium |
| EPI_ISL_452995 | Department of Pathology, University of Cambridge | COVID-19 Genomics UK (COG-UK) Consortium |
| EPI_ISL_452996 | Department of Pathology, University of Cambridge | COVID-19 Genomics UK (COG-UK) Consortium |
| EPI_ISL_452997 | Department of Pathology, University of Cambridge | COVID-19 Genomics UK (COG-UK) Consortium |
| EPI_ISL_452998 | Department of Pathology, University of Cambridge | COVID-19 Genomics UK (COG-UK) Consortium |
| EPI_ISL_452999 | Department of Pathology, University of Cambridge | COVID-19 Genomics UK (COG-UK) Consortium |
| EPI_ISL_453000 | Department of Pathology, University of Cambridge | COVID-19 Genomics UK (COG-UK) Consortium |
| EPI_ISL_453001 | Department of Pathology, University of Cambridge | COVID-19 Genomics UK (COG-UK) Consortium |
| EPI_ISL_453002 | Department of Pathology, University of Cambridge | COVID-19 Genomics UK (COG-UK) Consortium |
| EPI_ISL_453003 | Department of Pathology, University of Cambridge | COVID-19 Genomics UK (COG-UK) Consortium |
| EPI_ISL_453004 | Department of Pathology, University of Cambridge | COVID-19 Genomics UK (COG-UK) Consortium |

| EPI_Accession_ID | Originating Laboratory | Submitting Laboratory |
|---|---|---|
| EPI_ISL_453005 | Department of Pathology, University of Cambridge | COVID-19 Genomics UK (COG-UK) Consortium |
| EPI_ISL_453006 | West of Scotland Specialist Virology Centre, NHSGGC / MRC-University of Glasgow Centre for Virus Research | COVID-19 Genomics UK (COG-UK) Consortium |
| EPI_ISL_453007 | West of Scotland Specialist Virology Centre, NHSGGC / MRC-University of Glasgow Centre for Virus Research | COVID-19 Genomics UK (COG-UK) Consortium |
| EPI_ISL_453008 | West of Scotland Specialist Virology Centre, NHSGGC / MRC-University of Glasgow Centre for Virus Research | COVID-19 Genomics UK (COG-UK) Consortium |
| EPI_ISL_453009 | West of Scotland Specialist Virology Centre, NHSGGC / MRC-University of Glasgow Centre for Virus Research | COVID-19 Genomics UK (COG-UK) Consortium |
| EPI_ISL_453010 | West of Scotland Specialist Virology Centre, NHSGGC / MRC-University of Glasgow Centre for Virus Research | COVID-19 Genomics UK (COG-UK) Consortium |
| EPI_ISL_453011 | West of Scotland Specialist Virology Centre, NHSGGC / MRC-University of Glasgow Centre for Virus Research | COVID-19 Genomics UK (COG-UK) Consortium |
| EPI_ISL_453012 | West of Scotland Specialist Virology Centre, NHSGGC / MRC-University of Glasgow Centre for Virus Research | COVID-19 Genomics UK (COG-UK) Consortium |
| EPI_ISL_453013 | West of Scotland Specialist Virology Centre, NHSGGC / MRC-University of Glasgow Centre for Virus Research | COVID-19 Genomics UK (COG-UK) Consortium |
| EPI_ISL_453014 | West of Scotland Specialist Virology Centre, NHSGGC / MRC-University of Glasgow Centre for Virus Research | COVID-19 Genomics UK (COG-UK) Consortium |
| EPI_ISL_453015 | West of Scotland Specialist Virology Centre, NHSGGC / MRC-University of Glasgow Centre for Virus Research | COVID-19 Genomics UK (COG-UK) Consortium |
| EPI_ISL_453016 | West of Scotland Specialist Virology Centre, NHSGGC / MRC-University of Glasgow Centre for Virus Research | COVID-19 Genomics UK (COG-UK) Consortium |
| EPI_ISL_453017 | West of Scotland Specialist Virology Centre, NHSGGC / MRC-University of Glasgow Centre for Virus Research | COVID-19 Genomics UK (COG-UK) Consortium |
| EPI_ISL_453018 | West of Scotland Specialist Virology Centre, NHSGGC / MRC-University of Glasgow Centre for Virus Research | COVID-19 Genomics UK (COG-UK) Consortium |
| EPI_ISL_453019 | West of Scotland Specialist Virology Centre, NHSGGC / MRC-University of Glasgow Centre for Virus Research | COVID-19 Genomics UK (COG-UK) Consortium |
| EPI_ISL_453020 | West of Scotland Specialist Virology Centre, NHSGGC / MRC-University of Glasgow Centre for Virus Research | COVID-19 Genomics UK (COG-UK) Consortium |
| EPI_ISL_453021 | West of Scotland Specialist Virology Centre, NHSGGC / MRC-University of Glasgow Centre for Virus Research | COVID-19 Genomics UK (COG-UK) Consortium |

| EPI_Accession_ID | Originating Laboratory | Submitting Laboratory |
|---|---|---|
| EPI_ISL_453022 | West of Scotland Specialist Virology Centre, NHSGGC / MRC-University of Glasgow Centre for Virus Research | COVID-19 Genomics UK (COG-UK) Consortium |
| EPI_ISL_453023 | West of Scotland Specialist Virology Centre, NHSGGC / MRC-University of Glasgow Centre for Virus Research | COVID-19 Genomics UK (COG-UK) Consortium |
| EPI_ISL_453024 | West of Scotland Specialist Virology Centre, NHSGGC / MRC-University of Glasgow Centre for Virus Research | COVID-19 Genomics UK (COG-UK) Consortium |
| EPI_ISL_453025 | West of Scotland Specialist Virology Centre, NHSGGC / MRC-University of Glasgow Centre for Virus Research | COVID-19 Genomics UK (COG-UK) Consortium |
| EPI_ISL_453026 | West of Scotland Specialist Virology Centre, NHSGGC / MRC-University of Glasgow Centre for Virus Research | COVID-19 Genomics UK (COG-UK) Consortium |
| EPI_ISL_453027 | West of Scotland Specialist Virology Centre, NHSGGC / MRC-University of Glasgow Centre for Virus Research | COVID-19 Genomics UK (COG-UK) Consortium |
| EPI_ISL_453028 | West of Scotland Specialist Virology Centre, NHSGGC / MRC-University of Glasgow Centre for Virus Research | COVID-19 Genomics UK (COG-UK) Consortium |
| EPI_ISL_453029 | West of Scotland Specialist Virology Centre, NHSGGC / MRC-University of Glasgow Centre for Virus Research | COVID-19 Genomics UK (COG-UK) Consortium |
| EPI_ISL_453030 | West of Scotland Specialist Virology Centre, NHSGGC / MRC-University of Glasgow Centre for Virus Research | COVID-19 Genomics UK (COG-UK) Consortium |
| EPI_ISL_453031 | West of Scotland Specialist Virology Centre, NHSGGC / MRC-University of Glasgow Centre for Virus Research | COVID-19 Genomics UK (COG-UK) Consortium |
| EPI_ISL_453032 | West of Scotland Specialist Virology Centre, NHSGGC / MRC-University of Glasgow Centre for Virus Research | COVID-19 Genomics UK (COG-UK) Consortium |
| EPI_ISL_453033 | West of Scotland Specialist Virology Centre, NHSGGC / MRC-University of Glasgow Centre for Virus Research | COVID-19 Genomics UK (COG-UK) Consortium |
| EPI_ISL_453034 | West of Scotland Specialist Virology Centre, NHSGGC / MRC-University of Glasgow Centre for Virus Research | COVID-19 Genomics UK (COG-UK) Consortium |
| EPI_ISL_453035 | West of Scotland Specialist Virology Centre, NHSGGC / MRC-University of Glasgow Centre for Virus Research | COVID-19 Genomics UK (COG-UK) Consortium |
| EPI_ISL_453036 | West of Scotland Specialist Virology Centre, NHSGGC / MRC-University of Glasgow Centre for Virus Research | COVID-19 Genomics UK (COG-UK) Consortium |
| EPI_ISL_453037 | West of Scotland Specialist Virology Centre, NHSGGC / MRC-University of Glasgow Centre for Virus Research | COVID-19 Genomics UK (COG-UK) Consortium |

| EPI_Accession_ID | Originating Laboratory | Submitting Laboratory |
|---|---|---|
| EPI_ISL_453038 | West of Scotland Specialist Virology Centre, NHSGGC / MRC-University of Glasgow Centre for Virus Research | COVID-19 Genomics UK (COG-UK) Consortium |
| EPI_ISL_453039 | West of Scotland Specialist Virology Centre, NHSGGC / MRC-University of Glasgow Centre for Virus Research | COVID-19 Genomics UK (COG-UK) Consortium |
| EPI_ISL_453040 | West of Scotland Specialist Virology Centre, NHSGGC / MRC-University of Glasgow Centre for Virus Research | COVID-19 Genomics UK (COG-UK) Consortium |
| EPI_ISL_453041 | West of Scotland Specialist Virology Centre, NHSGGC / MRC-University of Glasgow Centre for Virus Research | COVID-19 Genomics UK (COG-UK) Consortium |
| EPI_ISL_453042 | West of Scotland Specialist Virology Centre, NHSGGC / MRC-University of Glasgow Centre for Virus Research | COVID-19 Genomics UK (COG-UK) Consortium |
| EPI_ISL_453043 | West of Scotland Specialist Virology Centre, NHSGGC / MRC-University of Glasgow Centre for Virus Research | COVID-19 Genomics UK (COG-UK) Consortium |
| EPI_ISL_453044 | West of Scotland Specialist Virology Centre, NHSGGC / MRC-University of Glasgow Centre for Virus Research | COVID-19 Genomics UK (COG-UK) Consortium |
| EPI_ISL_453045 | West of Scotland Specialist Virology Centre, NHSGGC / MRC-University of Glasgow Centre for Virus Research | COVID-19 Genomics UK (COG-UK) Consortium |
| EPI_ISL_453046 | West of Scotland Specialist Virology Centre, NHSGGC / MRC-University of Glasgow Centre for Virus Research | COVID-19 Genomics UK (COG-UK) Consortium |
| EPI_ISL_453047 | West of Scotland Specialist Virology Centre, NHSGGC / MRC-University of Glasgow Centre for Virus Research | COVID-19 Genomics UK (COG-UK) Consortium |
| EPI_ISL_453048 | West of Scotland Specialist Virology Centre, NHSGGC / MRC-University of Glasgow Centre for Virus Research | COVID-19 Genomics UK (COG-UK) Consortium |
| EPI_ISL_453049 | West of Scotland Specialist Virology Centre, NHSGGC / MRC-University of Glasgow Centre for Virus Research | COVID-19 Genomics UK (COG-UK) Consortium |
| EPI_ISL_453050 | West of Scotland Specialist Virology Centre, NHSGGC / MRC-University of Glasgow Centre for Virus Research | COVID-19 Genomics UK (COG-UK) Consortium |
| EPI_ISL_453051 | West of Scotland Specialist Virology Centre, NHSGGC / MRC-University of Glasgow Centre for Virus Research | COVID-19 Genomics UK (COG-UK) Consortium |
| EPI_ISL_453052 | West of Scotland Specialist Virology Centre, NHSGGC / MRC-University of Glasgow Centre for Virus Research | COVID-19 Genomics UK (COG-UK) Consortium |
| EPI_ISL_453053 | West of Scotland Specialist Virology Centre, NHSGGC / MRC-University of Glasgow Centre for Virus Research | COVID-19 Genomics UK (COG-UK) Consortium |

| EPI_Accession_ID | Originating Laboratory | Submitting Laboratory |
|---|---|---|
| EPI_ISL_453054 | West of Scotland Specialist Virology Centre, NHSGGC / MRC-University of Glasgow Centre for Virus Research | COVID-19 Genomics UK (COG-UK) Consortium |
| EPI_ISL_453055 | West of Scotland Specialist Virology Centre, NHSGGC / MRC-University of Glasgow Centre for Virus Research | COVID-19 Genomics UK (COG-UK) Consortium |
| EPI_ISL_453056 | West of Scotland Specialist Virology Centre, NHSGGC / MRC-University of Glasgow Centre for Virus Research | COVID-19 Genomics UK (COG-UK) Consortium |
| EPI_ISL_453057 | West of Scotland Specialist Virology Centre, NHSGGC / MRC-University of Glasgow Centre for Virus Research | COVID-19 Genomics UK (COG-UK) Consortium |
| EPI_ISL_453058 | West of Scotland Specialist Virology Centre, NHSGGC / MRC-University of Glasgow Centre for Virus Research | COVID-19 Genomics UK (COG-UK) Consortium |
| EPI_ISL_453059 | West of Scotland Specialist Virology Centre, NHSGGC / MRC-University of Glasgow Centre for Virus Research | COVID-19 Genomics UK (COG-UK) Consortium |
| EPI_ISL_453060 | West of Scotland Specialist Virology Centre, NHSGGC / MRC-University of Glasgow Centre for Virus Research | COVID-19 Genomics UK (COG-UK) Consortium |
| EPI_ISL_453061 | West of Scotland Specialist Virology Centre, NHSGGC / MRC-University of Glasgow Centre for Virus Research | COVID-19 Genomics UK (COG-UK) Consortium |
| EPI_ISL_453062 | West of Scotland Specialist Virology Centre, NHSGGC / MRC-University of Glasgow Centre for Virus Research | COVID-19 Genomics UK (COG-UK) Consortium |
| EPI_ISL_453063 | West of Scotland Specialist Virology Centre, NHSGGC / MRC-University of Glasgow Centre for Virus Research | COVID-19 Genomics UK (COG-UK) Consortium |
| EPI_ISL_453064 | West of Scotland Specialist Virology Centre, NHSGGC / MRC-University of Glasgow Centre for Virus Research | COVID-19 Genomics UK (COG-UK) Consortium |
| EPI_ISL_453065 | West of Scotland Specialist Virology Centre, NHSGGC / MRC-University of Glasgow Centre for Virus Research | COVID-19 Genomics UK (COG-UK) Consortium |
| EPI_ISL_453066 | West of Scotland Specialist Virology Centre, NHSGGC / MRC-University of Glasgow Centre for Virus Research | COVID-19 Genomics UK (COG-UK) Consortium |
| EPI_ISL_453067 | West of Scotland Specialist Virology Centre, NHSGGC / MRC-University of Glasgow Centre for Virus Research | COVID-19 Genomics UK (COG-UK) Consortium |
| EPI_ISL_453068 | West of Scotland Specialist Virology Centre, NHSGGC / MRC-University of Glasgow Centre for Virus Research | COVID-19 Genomics UK (COG-UK) Consortium |
| EPI_ISL_453069 | West of Scotland Specialist Virology Centre, NHSGGC / MRC-University of Glasgow Centre for Virus Research | COVID-19 Genomics UK (COG-UK) Consortium |

| EPI_Accession_ID | Originating Laboratory | Submitting Laboratory |
|---|---|---|
| EPI_ISL_453070 | West of Scotland Specialist Virology Centre, NHSGGC / MRC-University of Glasgow Centre for Virus Research | COVID-19 Genomics UK (COG-UK) Consortium |
| EPI_ISL_453071 | West of Scotland Specialist Virology Centre, NHSGGC / MRC-University of Glasgow Centre for Virus Research | COVID-19 Genomics UK (COG-UK) Consortium |
| EPI_ISL_453072 | West of Scotland Specialist Virology Centre, NHSGGC / MRC-University of Glasgow Centre for Virus Research | COVID-19 Genomics UK (COG-UK) Consortium |
| EPI_ISL_453073 | West of Scotland Specialist Virology Centre, NHSGGC / MRC-University of Glasgow Centre for Virus Research | COVID-19 Genomics UK (COG-UK) Consortium |
| EPI_ISL_453074 | West of Scotland Specialist Virology Centre, NHSGGC / MRC-University of Glasgow Centre for Virus Research | COVID-19 Genomics UK (COG-UK) Consortium |
| EPI_ISL_453075 | West of Scotland Specialist Virology Centre, NHSGGC / MRC-University of Glasgow Centre for Virus Research | COVID-19 Genomics UK (COG-UK) Consortium |
| EPI_ISL_453076 | West of Scotland Specialist Virology Centre, NHSGGC / MRC-University of Glasgow Centre for Virus Research | COVID-19 Genomics UK (COG-UK) Consortium |
| EPI_ISL_453077 | West of Scotland Specialist Virology Centre, NHSGGC / MRC-University of Glasgow Centre for Virus Research | COVID-19 Genomics UK (COG-UK) Consortium |
| EPI_ISL_453078 | West of Scotland Specialist Virology Centre, NHSGGC / MRC-University of Glasgow Centre for Virus Research | COVID-19 Genomics UK (COG-UK) Consortium |
| EPI_ISL_453079 | West of Scotland Specialist Virology Centre, NHSGGC / MRC-University of Glasgow Centre for Virus Research | COVID-19 Genomics UK (COG-UK) Consortium |
| EPI_ISL_453080 | West of Scotland Specialist Virology Centre, NHSGGC / MRC-University of Glasgow Centre for Virus Research | COVID-19 Genomics UK (COG-UK) Consortium |
| EPI_ISL_453081 | West of Scotland Specialist Virology Centre, NHSGGC / MRC-University of Glasgow Centre for Virus Research | COVID-19 Genomics UK (COG-UK) Consortium |
| EPI_ISL_453082 | West of Scotland Specialist Virology Centre, NHSGGC / MRC-University of Glasgow Centre for Virus Research | COVID-19 Genomics UK (COG-UK) Consortium |
| EPI_ISL_453083 | West of Scotland Specialist Virology Centre, NHSGGC / MRC-University of Glasgow Centre for Virus Research | COVID-19 Genomics UK (COG-UK) Consortium |
| EPI_ISL_453084 | West of Scotland Specialist Virology Centre, NHSGGC / MRC-University of Glasgow Centre for Virus Research | COVID-19 Genomics UK (COG-UK) Consortium |
| EPI_ISL_453085 | West of Scotland Specialist Virology Centre, NHSGGC / MRC-University of Glasgow Centre for Virus Research | COVID-19 Genomics UK (COG-UK) Consortium |

| EPI_Accession_ID | Originating Laboratory | Submitting Laboratory |
|---|---|---|
| EPI_ISL_453086 | West of Scotland Specialist Virology Centre, NHSGGC / MRC-University of Glasgow Centre for Virus Research | COVID-19 Genomics UK (COG-UK) Consortium |
| EPI_ISL_453087 | West of Scotland Specialist Virology Centre, NHSGGC / MRC-University of Glasgow Centre for Virus Research | COVID-19 Genomics UK (COG-UK) Consortium |
| EPI_ISL_453088 | West of Scotland Specialist Virology Centre, NHSGGC / MRC-University of Glasgow Centre for Virus Research | COVID-19 Genomics UK (COG-UK) Consortium |
| EPI_ISL_453089 | West of Scotland Specialist Virology Centre, NHSGGC / MRC-University of Glasgow Centre for Virus Research | COVID-19 Genomics UK (COG-UK) Consortium |
| EPI_ISL_453090 | West of Scotland Specialist Virology Centre, NHSGGC / MRC-University of Glasgow Centre for Virus Research | COVID-19 Genomics UK (COG-UK) Consortium |
| EPI_ISL_453091 | West of Scotland Specialist Virology Centre, NHSGGC / MRC-University of Glasgow Centre for Virus Research | COVID-19 Genomics UK (COG-UK) Consortium |
| EPI_ISL_453092 | West of Scotland Specialist Virology Centre, NHSGGC / MRC-University of Glasgow Centre for Virus Research | COVID-19 Genomics UK (COG-UK) Consortium |
| EPI_ISL_453093 | West of Scotland Specialist Virology Centre, NHSGGC / MRC-University of Glasgow Centre for Virus Research | COVID-19 Genomics UK (COG-UK) Consortium |
| EPI_ISL_453094 | West of Scotland Specialist Virology Centre, NHSGGC / MRC-University of Glasgow Centre for Virus Research | COVID-19 Genomics UK (COG-UK) Consortium |
| EPI_ISL_453095 | West of Scotland Specialist Virology Centre, NHSGGC / MRC-University of Glasgow Centre for Virus Research | COVID-19 Genomics UK (COG-UK) Consortium |
| EPI_ISL_453096 | West of Scotland Specialist Virology Centre, NHSGGC / MRC-University of Glasgow Centre for Virus Research | COVID-19 Genomics UK (COG-UK) Consortium |
| EPI_ISL_453097 | West of Scotland Specialist Virology Centre, NHSGGC / MRC-University of Glasgow Centre for Virus Research | COVID-19 Genomics UK (COG-UK) Consortium |
| EPI_ISL_453098 | West of Scotland Specialist Virology Centre, NHSGGC / MRC-University of Glasgow Centre for Virus Research | COVID-19 Genomics UK (COG-UK) Consortium |
| EPI_ISL_453099 | Virology Department, Royal Infirmary of Edinburgh, NHS Lothian / School of Biological Sciences, University of Edinburgh / Institute of Genetics and Molecular Medicine, University of Edinburgh | COVID-19 Genomics UK (COG-UK) Consortium |

| EPI_Accession_ID | Originating Laboratory | Submitting Laboratory |
|---|---|---|
| EPI_ISL_453100 | Virology Department, Royal Infirmary of Edinburgh, NHS Lothian / School of Biological Sciences, University of Edinburgh / Institute of Genetics and Molecular Medicine, University of Edinburgh | COVID-19 Genomics UK (COG-UK) Consortium |
| EPI_ISL_453101 | Virology Department, Royal Infirmary of Edinburgh, NHS Lothian / School of Biological Sciences, University of Edinburgh / Institute of Genetics and Molecular Medicine, University of Edinburgh | COVID-19 Genomics UK (COG-UK) Consortium |
| EPI_ISL_453102 | Virology Department, Royal Infirmary of Edinburgh, NHS Lothian / School of Biological Sciences, University of Edinburgh / Institute of Genetics and Molecular Medicine, University of Edinburgh | COVID-19 Genomics UK (COG-UK) Consortium |
| EPI_ISL_453103 | Virology Department, Royal Infirmary of Edinburgh, NHS Lothian / School of Biological Sciences, University of Edinburgh / Institute of Genetics and Molecular Medicine, University of Edinburgh | COVID-19 Genomics UK (COG-UK) Consortium |
| EPI_ISL_453104 | Virology Department, Royal Infirmary of Edinburgh, NHS Lothian / School of Biological Sciences, University of Edinburgh / Institute of Genetics and Molecular Medicine, University of Edinburgh | COVID-19 Genomics UK (COG-UK) Consortium |
| EPI_ISL_453105 | Virology Department, Royal Infirmary of Edinburgh, NHS Lothian / School of Biological Sciences, University of Edinburgh / Institute of Genetics and Molecular Medicine, University of Edinburgh | COVID-19 Genomics UK (COG-UK) Consortium |
| EPI_ISL_453106 | Virology Department, Royal Infirmary of Edinburgh, NHS Lothian / School of Biological Sciences, University of Edinburgh / Institute of Genetics and Molecular Medicine, University of Edinburgh | COVID-19 Genomics UK (COG-UK) Consortium |
| EPI_ISL_453107 | Virology Department, Royal Infirmary of Edinburgh, NHS Lothian / School of Biological Sciences, University of Edinburgh / Institute of Genetics and Molecular Medicine, University of Edinburgh | COVID-19 Genomics UK (COG-UK) Consortium |

| EPI_Accession_ID | Originating Laboratory | Submitting Laboratory |
|---|---|---|
| EPI_ISL_453108 | Virology Department, Royal Infirmary of Edinburgh, NHS Lothian / School of Biological Sciences, University of Edinburgh / Institute of Genetics and Molecular Medicine, University of Edinburgh | COVID-19 Genomics UK (COG-UK) Consortium |
| EPI_ISL_453109 | Virology Department, Royal Infirmary of Edinburgh, NHS Lothian / School of Biological Sciences, University of Edinburgh / Institute of Genetics and Molecular Medicine, University of Edinburgh | COVID-19 Genomics UK (COG-UK) Consortium |
| EPI_ISL_453110 | Virology Department, Royal Infirmary of Edinburgh, NHS Lothian / School of Biological Sciences, University of Edinburgh / Institute of Genetics and Molecular Medicine, University of Edinburgh | COVID-19 Genomics UK (COG-UK) Consortium |
| EPI_ISL_453111 | Virology Department, Royal Infirmary of Edinburgh, NHS Lothian / School of Biological Sciences, University of Edinburgh / Institute of Genetics and Molecular Medicine, University of Edinburgh | COVID-19 Genomics UK (COG-UK) Consortium |
| EPI_ISL_453112 | Virology Department, Royal Infirmary of Edinburgh, NHS Lothian / School of Biological Sciences, University of Edinburgh / Institute of Genetics and Molecular Medicine, University of Edinburgh | COVID-19 Genomics UK (COG-UK) Consortium |
| EPI_ISL_453113 | Virology Department, Royal Infirmary of Edinburgh, NHS Lothian / School of Biological Sciences, University of Edinburgh / Institute of Genetics and Molecular Medicine, University of Edinburgh | COVID-19 Genomics UK (COG-UK) Consortium |
| EPI_ISL_453114 | Virology Department, Royal Infirmary of Edinburgh, NHS Lothian / School of Biological Sciences, University of Edinburgh / Institute of Genetics and Molecular Medicine, University of Edinburgh | COVID-19 Genomics UK (COG-UK) Consortium |
| EPI_ISL_453115 | Virology Department, Royal Infirmary of Edinburgh, NHS Lothian / School of Biological Sciences, University of Edinburgh / Institute of Genetics and Molecular Medicine, University of Edinburgh | COVID-19 Genomics UK (COG-UK) Consortium |

| EPI_Accession_ID | Originating Laboratory | Submitting Laboratory |
| --- | --- | --- |
| EPI_ISL_453116 | Virology Department, Royal Infirmary of Edinburgh, NHS Lothian / School of Biological Sciences, University of Edinburgh / Institute of Genetics and Molecular Medicine, University of Edinburgh | COVID-19 Genomics UK (COG-UK) Consortium |
| EPI_ISL_453117 | Virology Department, Royal Infirmary of Edinburgh, NHS Lothian / School of Biological Sciences, University of Edinburgh / Institute of Genetics and Molecular Medicine, University of Edinburgh | COVID-19 Genomics UK (COG-UK) Consortium |
| EPI_ISL_453118 | Virology Department, Royal Infirmary of Edinburgh, NHS Lothian / School of Biological Sciences, University of Edinburgh / Institute of Genetics and Molecular Medicine, University of Edinburgh | COVID-19 Genomics UK (COG-UK) Consortium |
| EPI_ISL_453119 | Virology Department, Royal Infirmary of Edinburgh, NHS Lothian / School of Biological Sciences, University of Edinburgh / Institute of Genetics and Molecular Medicine, University of Edinburgh | COVID-19 Genomics UK (COG-UK) Consortium |
| EPI_ISL_453120 | Virology Department, Royal Infirmary of Edinburgh, NHS Lothian / School of Biological Sciences, University of Edinburgh / Institute of Genetics and Molecular Medicine, University of Edinburgh | COVID-19 Genomics UK (COG-UK) Consortium |
| EPI_ISL_453121 | Virology Department, Royal Infirmary of Edinburgh, NHS Lothian / School of Biological Sciences, University of Edinburgh / Institute of Genetics and Molecular Medicine, University of Edinburgh | COVID-19 Genomics UK (COG-UK) Consortium |
| EPI_ISL_453122 | Virology Department, Royal Infirmary of Edinburgh, NHS Lothian / School of Biological Sciences, University of Edinburgh / Institute of Genetics and Molecular Medicine, University of Edinburgh | COVID-19 Genomics UK (COG-UK) Consortium |
| EPI_ISL_453123 | Virology Department, Royal Infirmary of Edinburgh, NHS Lothian / School of Biological Sciences, University of Edinburgh / Institute of Genetics and Molecular Medicine, University of Edinburgh | COVID-19 Genomics UK (COG-UK) Consortium |

| EPI_Accession_ID | Originating Laboratory | Submitting Laboratory |
|---|---|---|
| EPI_ISL_453124 | Virology Department, Royal Infirmary of Edinburgh, NHS Lothian / School of Biological Sciences, University of Edinburgh / Institute of Genetics and Molecular Medicine, University of Edinburgh | COVID-19 Genomics UK (COG-UK) Consortium |
| EPI_ISL_453125 | Virology Department, Royal Infirmary of Edinburgh, NHS Lothian / School of Biological Sciences, University of Edinburgh / Institute of Genetics and Molecular Medicine, University of Edinburgh | COVID-19 Genomics UK (COG-UK) Consortium |
| EPI_ISL_453126 | Virology Department, Royal Infirmary of Edinburgh, NHS Lothian / School of Biological Sciences, University of Edinburgh / Institute of Genetics and Molecular Medicine, University of Edinburgh | COVID-19 Genomics UK (COG-UK) Consortium |
| EPI_ISL_453127 | Virology Department, Royal Infirmary of Edinburgh, NHS Lothian / School of Biological Sciences, University of Edinburgh / Institute of Genetics and Molecular Medicine, University of Edinburgh | COVID-19 Genomics UK (COG-UK) Consortium |
| EPI_ISL_453128 | Virology Department, Royal Infirmary of Edinburgh, NHS Lothian / School of Biological Sciences, University of Edinburgh / Institute of Genetics and Molecular Medicine, University of Edinburgh | COVID-19 Genomics UK (COG-UK) Consortium |
| EPI_ISL_453129 | Virology Department, Royal Infirmary of Edinburgh, NHS Lothian / School of Biological Sciences, University of Edinburgh / Institute of Genetics and Molecular Medicine, University of Edinburgh | COVID-19 Genomics UK (COG-UK) Consortium |
| EPI_ISL_453130 | Virology Department, Royal Infirmary of Edinburgh, NHS Lothian / School of Biological Sciences, University of Edinburgh / Institute of Genetics and Molecular Medicine, University of Edinburgh | COVID-19 Genomics UK (COG-UK) Consortium |
| EPI_ISL_453131 | Virology Department, Royal Infirmary of Edinburgh, NHS Lothian / School of Biological Sciences, University of Edinburgh / Institute of Genetics and Molecular Medicine, University of Edinburgh | COVID-19 Genomics UK (COG-UK) Consortium |

| EPI_Accession_ID | Originating Laboratory | Submitting Laboratory |
| --- | --- | --- |
| EPI_ISL_453132 | Virology Department, Royal Infirmary of Edinburgh, NHS Lothian / School of Biological Sciences, University of Edinburgh / Institute of Genetics and Molecular Medicine, University of Edinburgh | COVID-19 Genomics UK (COG-UK) Consortium |
| EPI_ISL_453133 | Virology Department, Royal Infirmary of Edinburgh, NHS Lothian / School of Biological Sciences, University of Edinburgh / Institute of Genetics and Molecular Medicine, University of Edinburgh | COVID-19 Genomics UK (COG-UK) Consortium |
| EPI_ISL_453134 | Virology Department, Royal Infirmary of Edinburgh, NHS Lothian / School of Biological Sciences, University of Edinburgh / Institute of Genetics and Molecular Medicine, University of Edinburgh | COVID-19 Genomics UK (COG-UK) Consortium |
| EPI_ISL_453135 | Virology Department, Royal Infirmary of Edinburgh, NHS Lothian / School of Biological Sciences, University of Edinburgh / Institute of Genetics and Molecular Medicine, University of Edinburgh | COVID-19 Genomics UK (COG-UK) Consortium |
| EPI_ISL_453136 | Virology Department, Royal Infirmary of Edinburgh, NHS Lothian / School of Biological Sciences, University of Edinburgh / Institute of Genetics and Molecular Medicine, University of Edinburgh | COVID-19 Genomics UK (COG-UK) Consortium |
| EPI_ISL_453137 | Virology Department, Royal Infirmary of Edinburgh, NHS Lothian / School of Biological Sciences, University of Edinburgh / Institute of Genetics and Molecular Medicine, University of Edinburgh | COVID-19 Genomics UK (COG-UK) Consortium |
| EPI_ISL_453138 | Virology Department, Royal Infirmary of Edinburgh, NHS Lothian / School of Biological Sciences, University of Edinburgh / Institute of Genetics and Molecular Medicine, University of Edinburgh | COVID-19 Genomics UK (COG-UK) Consortium |
| EPI_ISL_453139 | Virology Department, Royal Infirmary of Edinburgh, NHS Lothian / School of Biological Sciences, University of Edinburgh / Institute of Genetics and Molecular Medicine, University of Edinburgh | COVID-19 Genomics UK (COG-UK) Consortium |

| EPI_Accession_ID | Originating Laboratory | Submitting Laboratory |
| --- | --- | --- |
| EPI_ISL_453140 | Virology Department, Royal Infirmary of Edinburgh, NHS Lothian / School of Biological Sciences, University of Edinburgh / Institute of Genetics and Molecular Medicine, University of Edinburgh | COVID-19 Genomics UK (COG-UK) Consortium |
| EPI_ISL_453141 | Virology Department, Royal Infirmary of Edinburgh, NHS Lothian / School of Biological Sciences, University of Edinburgh / Institute of Genetics and Molecular Medicine, University of Edinburgh | COVID-19 Genomics UK (COG-UK) Consortium |
| EPI_ISL_453142 | Virology Department, Royal Infirmary of Edinburgh, NHS Lothian / School of Biological Sciences, University of Edinburgh / Institute of Genetics and Molecular Medicine, University of Edinburgh | COVID-19 Genomics UK (COG-UK) Consortium |
| EPI_ISL_453143 | Virology Department, Royal Infirmary of Edinburgh, NHS Lothian / School of Biological Sciences, University of Edinburgh / Institute of Genetics and Molecular Medicine, University of Edinburgh | COVID-19 Genomics UK (COG-UK) Consortium |
| EPI_ISL_453144 | Virology Department, Royal Infirmary of Edinburgh, NHS Lothian / School of Biological Sciences, University of Edinburgh / Institute of Genetics and Molecular Medicine, University of Edinburgh | COVID-19 Genomics UK (COG-UK) Consortium |
| EPI_ISL_453145 | Virology Department, Royal Infirmary of Edinburgh, NHS Lothian / School of Biological Sciences, University of Edinburgh / Institute of Genetics and Molecular Medicine, University of Edinburgh | COVID-19 Genomics UK (COG-UK) Consortium |
| EPI_ISL_453146 | Virology Department, Royal Infirmary of Edinburgh, NHS Lothian / School of Biological Sciences, University of Edinburgh / Institute of Genetics and Molecular Medicine, University of Edinburgh | COVID-19 Genomics UK (COG-UK) Consortium |
| EPI_ISL_453147 | Virology Department, Royal Infirmary of Edinburgh, NHS Lothian / School of Biological Sciences, University of Edinburgh / Institute of Genetics and Molecular Medicine, University of Edinburgh | COVID-19 Genomics UK (COG-UK) Consortium |

| EPI_Accession_ID | Originating Laboratory | Submitting Laboratory |
|---|---|---|
| EPI_ISL_453148 | Virology Department, Royal Infirmary of Edinburgh, NHS Lothian / School of Biological Sciences, University of Edinburgh / Institute of Genetics and Molecular Medicine, University of Edinburgh | COVID-19 Genomics UK (COG-UK) Consortium |
| EPI_ISL_453149 | Virology Department, Royal Infirmary of Edinburgh, NHS Lothian / School of Biological Sciences, University of Edinburgh / Institute of Genetics and Molecular Medicine, University of Edinburgh | COVID-19 Genomics UK (COG-UK) Consortium |
| EPI_ISL_453150 | Virology Department, Royal Infirmary of Edinburgh, NHS Lothian / School of Biological Sciences, University of Edinburgh / Institute of Genetics and Molecular Medicine, University of Edinburgh | COVID-19 Genomics UK (COG-UK) Consortium |
| EPI_ISL_453151 | Virology Department, Royal Infirmary of Edinburgh, NHS Lothian / School of Biological Sciences, University of Edinburgh / Institute of Genetics and Molecular Medicine, University of Edinburgh | COVID-19 Genomics UK (COG-UK) Consortium |
| EPI_ISL_453152 | Virology Department, Royal Infirmary of Edinburgh, NHS Lothian / School of Biological Sciences, University of Edinburgh / Institute of Genetics and Molecular Medicine, University of Edinburgh | COVID-19 Genomics UK (COG-UK) Consortium |
| EPI_ISL_453153 | Virology Department, Royal Infirmary of Edinburgh, NHS Lothian / School of Biological Sciences, University of Edinburgh / Institute of Genetics and Molecular Medicine, University of Edinburgh | COVID-19 Genomics UK (COG-UK) Consortium |
| EPI_ISL_453154 | Virology Department, Royal Infirmary of Edinburgh, NHS Lothian / School of Biological Sciences, University of Edinburgh / Institute of Genetics and Molecular Medicine, University of Edinburgh | COVID-19 Genomics UK (COG-UK) Consortium |
| EPI_ISL_453155 | Virology Department, Royal Infirmary of Edinburgh, NHS Lothian / School of Biological Sciences, University of Edinburgh / Institute of Genetics and Molecular Medicine, University of Edinburgh | COVID-19 Genomics UK (COG-UK) Consortium |

| EPI_Accession_ID | Originating Laboratory | Submitting Laboratory |
|---|---|---|
| EPI_ISL_453156 | Virology Department, Royal Infirmary of Edinburgh, NHS Lothian / School of Biological Sciences, University of Edinburgh / Institute of Genetics and Molecular Medicine, University of Edinburgh | COVID-19 Genomics UK (COG-UK) Consortium |
| EPI_ISL_453157 | Virology Department, Royal Infirmary of Edinburgh, NHS Lothian / School of Biological Sciences, University of Edinburgh / Institute of Genetics and Molecular Medicine, University of Edinburgh | COVID-19 Genomics UK (COG-UK) Consortium |
| EPI_ISL_453158 | Virology Department, Royal Infirmary of Edinburgh, NHS Lothian / School of Biological Sciences, University of Edinburgh / Institute of Genetics and Molecular Medicine, University of Edinburgh | COVID-19 Genomics UK (COG-UK) Consortium |
| EPI_ISL_453159 | Virology Department, Royal Infirmary of Edinburgh, NHS Lothian / School of Biological Sciences, University of Edinburgh / Institute of Genetics and Molecular Medicine, University of Edinburgh | COVID-19 Genomics UK (COG-UK) Consortium |
| EPI_ISL_453160 | Virology Department, Royal Infirmary of Edinburgh, NHS Lothian / School of Biological Sciences, University of Edinburgh / Institute of Genetics and Molecular Medicine, University of Edinburgh | COVID-19 Genomics UK (COG-UK) Consortium |
| EPI_ISL_453161 | Virology Department, Royal Infirmary of Edinburgh, NHS Lothian / School of Biological Sciences, University of Edinburgh / Institute of Genetics and Molecular Medicine, University of Edinburgh | COVID-19 Genomics UK (COG-UK) Consortium |
| EPI_ISL_453162 | Virology Department, Royal Infirmary of Edinburgh, NHS Lothian / School of Biological Sciences, University of Edinburgh / Institute of Genetics and Molecular Medicine, University of Edinburgh | COVID-19 Genomics UK (COG-UK) Consortium |
| EPI_ISL_453163 | Virology Department, Royal Infirmary of Edinburgh, NHS Lothian / School of Biological Sciences, University of Edinburgh / Institute of Genetics and Molecular Medicine, University of Edinburgh | COVID-19 Genomics UK (COG-UK) Consortium |

| EPI_Accession_ID | Originating Laboratory | Submitting Laboratory |
| --- | --- | --- |
| EPI_ISL_453164 | Virology Department, Royal Infirmary of Edinburgh, NHS Lothian / School of Biological Sciences, University of Edinburgh / Institute of Genetics and Molecular Medicine, University of Edinburgh | COVID-19 Genomics UK (COG-UK) Consortium |
| EPI_ISL_453165 | Virology Department, Royal Infirmary of Edinburgh, NHS Lothian / School of Biological Sciences, University of Edinburgh / Institute of Genetics and Molecular Medicine, University of Edinburgh | COVID-19 Genomics UK (COG-UK) Consortium |
| EPI_ISL_453166 | Virology Department, Royal Infirmary of Edinburgh, NHS Lothian / School of Biological Sciences, University of Edinburgh / Institute of Genetics and Molecular Medicine, University of Edinburgh | COVID-19 Genomics UK (COG-UK) Consortium |
| EPI_ISL_453167 | Virology Department, Royal Infirmary of Edinburgh, NHS Lothian / School of Biological Sciences, University of Edinburgh / Institute of Genetics and Molecular Medicine, University of Edinburgh | COVID-19 Genomics UK (COG-UK) Consortium |
| EPI_ISL_453168 | Virology Department, Royal Infirmary of Edinburgh, NHS Lothian / School of Biological Sciences, University of Edinburgh / Institute of Genetics and Molecular Medicine, University of Edinburgh | COVID-19 Genomics UK (COG-UK) Consortium |
| EPI_ISL_453169 | Virology Department, Royal Infirmary of Edinburgh, NHS Lothian / School of Biological Sciences, University of Edinburgh / Institute of Genetics and Molecular Medicine, University of Edinburgh | COVID-19 Genomics UK (COG-UK) Consortium |
| EPI_ISL_453170 | Virology Department, Royal Infirmary of Edinburgh, NHS Lothian / School of Biological Sciences, University of Edinburgh / Institute of Genetics and Molecular Medicine, University of Edinburgh | COVID-19 Genomics UK (COG-UK) Consortium |
| EPI_ISL_453171 | Virology Department, Royal Infirmary of Edinburgh, NHS Lothian / School of Biological Sciences, University of Edinburgh / Institute of Genetics and Molecular Medicine, University of Edinburgh | COVID-19 Genomics UK (COG-UK) Consortium |

| EPI_Accession_ID | Originating Laboratory | Submitting Laboratory |
|---|---|---|
| EPI_ISL_453172 | Virology Department, Royal Infirmary of Edinburgh, NHS Lothian / School of Biological Sciences, University of Edinburgh / Institute of Genetics and Molecular Medicine, University of Edinburgh | COVID-19 Genomics UK (COG-UK) Consortium |
| EPI_ISL_453173 | Virology Department, Royal Infirmary of Edinburgh, NHS Lothian / School of Biological Sciences, University of Edinburgh / Institute of Genetics and Molecular Medicine, University of Edinburgh | COVID-19 Genomics UK (COG-UK) Consortium |
| EPI_ISL_453174 | Virology Department, Royal Infirmary of Edinburgh, NHS Lothian / School of Biological Sciences, University of Edinburgh / Institute of Genetics and Molecular Medicine, University of Edinburgh | COVID-19 Genomics UK (COG-UK) Consortium |
| EPI_ISL_453175 | Virology Department, Royal Infirmary of Edinburgh, NHS Lothian / School of Biological Sciences, University of Edinburgh / Institute of Genetics and Molecular Medicine, University of Edinburgh | COVID-19 Genomics UK (COG-UK) Consortium |
| EPI_ISL_453176 | Virology Department, Royal Infirmary of Edinburgh, NHS Lothian / School of Biological Sciences, University of Edinburgh / Institute of Genetics and Molecular Medicine, University of Edinburgh | COVID-19 Genomics UK (COG-UK) Consortium |
| EPI_ISL_453177 | Virology Department, Royal Infirmary of Edinburgh, NHS Lothian / School of Biological Sciences, University of Edinburgh / Institute of Genetics and Molecular Medicine, University of Edinburgh | COVID-19 Genomics UK (COG-UK) Consortium |
| EPI_ISL_453178 | Virology Department, Royal Infirmary of Edinburgh, NHS Lothian / School of Biological Sciences, University of Edinburgh / Institute of Genetics and Molecular Medicine, University of Edinburgh | COVID-19 Genomics UK (COG-UK) Consortium |
| EPI_ISL_453179 | Virology Department, Royal Infirmary of Edinburgh, NHS Lothian / School of Biological Sciences, University of Edinburgh / Institute of Genetics and Molecular Medicine, University of Edinburgh | COVID-19 Genomics UK (COG-UK) Consortium |

| EPI_Accession_ID | Originating Laboratory | Submitting Laboratory |
|---|---|---|
| EPI_ISL_453180 | Virology Department, Royal Infirmary of Edinburgh, NHS Lothian / School of Biological Sciences, University of Edinburgh / Institute of Genetics and Molecular Medicine, University of Edinburgh | COVID-19 Genomics UK (COG-UK) Consortium |
| EPI_ISL_453181 | Virology Department, Royal Infirmary of Edinburgh, NHS Lothian / School of Biological Sciences, University of Edinburgh / Institute of Genetics and Molecular Medicine, University of Edinburgh | COVID-19 Genomics UK (COG-UK) Consortium |
| EPI_ISL_453182 | Virology Department, Royal Infirmary of Edinburgh, NHS Lothian / School of Biological Sciences, University of Edinburgh / Institute of Genetics and Molecular Medicine, University of Edinburgh | COVID-19 Genomics UK (COG-UK) Consortium |
| EPI_ISL_453183 | Virology Department, Royal Infirmary of Edinburgh, NHS Lothian / School of Biological Sciences, University of Edinburgh / Institute of Genetics and Molecular Medicine, University of Edinburgh | COVID-19 Genomics UK (COG-UK) Consortium |
| EPI_ISL_453184 | Virology Department, Royal Infirmary of Edinburgh, NHS Lothian / School of Biological Sciences, University of Edinburgh / Institute of Genetics and Molecular Medicine, University of Edinburgh | COVID-19 Genomics UK (COG-UK) Consortium |
| EPI_ISL_453185 | Virology Department, Royal Infirmary of Edinburgh, NHS Lothian / School of Biological Sciences, University of Edinburgh / Institute of Genetics and Molecular Medicine, University of Edinburgh | COVID-19 Genomics UK (COG-UK) Consortium |
| EPI_ISL_453186 | Virology Department, Royal Infirmary of Edinburgh, NHS Lothian / School of Biological Sciences, University of Edinburgh / Institute of Genetics and Molecular Medicine, University of Edinburgh | COVID-19 Genomics UK (COG-UK) Consortium |
| EPI_ISL_453187 | Virology Department, Royal Infirmary of Edinburgh, NHS Lothian / School of Biological Sciences, University of Edinburgh / Institute of Genetics and Molecular Medicine, University of Edinburgh | COVID-19 Genomics UK (COG-UK) Consortium |

| EPI_Accession_ID | Originating Laboratory | Submitting Laboratory |
|---|---|---|
| EPI_ISL_453188 | Virology Department, Royal Infirmary of Edinburgh, NHS Lothian / School of Biological Sciences, University of Edinburgh / Institute of Genetics and Molecular Medicine, University of Edinburgh | COVID-19 Genomics UK (COG-UK) Consortium |
| EPI_ISL_453189 | Virology Department, Royal Infirmary of Edinburgh, NHS Lothian / School of Biological Sciences, University of Edinburgh / Institute of Genetics and Molecular Medicine, University of Edinburgh | COVID-19 Genomics UK (COG-UK) Consortium |
| EPI_ISL_453190 | Virology Department, Royal Infirmary of Edinburgh, NHS Lothian / School of Biological Sciences, University of Edinburgh / Institute of Genetics and Molecular Medicine, University of Edinburgh | COVID-19 Genomics UK (COG-UK) Consortium |
| EPI_ISL_453191 | Virology Department, Royal Infirmary of Edinburgh, NHS Lothian / School of Biological Sciences, University of Edinburgh / Institute of Genetics and Molecular Medicine, University of Edinburgh | COVID-19 Genomics UK (COG-UK) Consortium |
| EPI_ISL_453192 | Virology Department, Royal Infirmary of Edinburgh, NHS Lothian / School of Biological Sciences, University of Edinburgh / Institute of Genetics and Molecular Medicine, University of Edinburgh | COVID-19 Genomics UK (COG-UK) Consortium |
| EPI_ISL_453193 | Virology Department, Royal Infirmary of Edinburgh, NHS Lothian / School of Biological Sciences, University of Edinburgh / Institute of Genetics and Molecular Medicine, University of Edinburgh | COVID-19 Genomics UK (COG-UK) Consortium |
| EPI_ISL_453194 | Virology Department, Royal Infirmary of Edinburgh, NHS Lothian / School of Biological Sciences, University of Edinburgh / Institute of Genetics and Molecular Medicine, University of Edinburgh | COVID-19 Genomics UK (COG-UK) Consortium |
| EPI_ISL_453195 | Liverpool Clinical Laboratories | COVID-19 Genomics UK (COG-UK) Consortium |
| EPI_ISL_453196 | Liverpool Clinical Laboratories | COVID-19 Genomics UK (COG-UK) Consortium |
| EPI_ISL_453197 | Liverpool Clinical Laboratories | COVID-19 Genomics UK (COG-UK) Consortium |
| EPI_ISL_453198 | Liverpool Clinical Laboratories | COVID-19 Genomics UK (COG-UK) Consortium |
| EPI_ISL_453199 | Liverpool Clinical Laboratories | COVID-19 Genomics UK (COG-UK) Consortium |

| EPI_Accession_ID | Originating Laboratory | Submitting Laboratory |
|---|---|---|
| EPI_ISL_453200 | Liverpool Clinical Laboratories | COVID-19 Genomics UK (COG-UK) Consortium |
| EPI_ISL_453201 | Liverpool Clinical Laboratories | COVID-19 Genomics UK (COG-UK) Consortium |
| EPI_ISL_453202 | Liverpool Clinical Laboratories | COVID-19 Genomics UK (COG-UK) Consortium |
| EPI_ISL_453203 | Liverpool Clinical Laboratories | COVID-19 Genomics UK (COG-UK) Consortium |
| EPI_ISL_453204 | Liverpool Clinical Laboratories | COVID-19 Genomics UK (COG-UK) Consortium |
| EPI_ISL_453205 | Liverpool Clinical Laboratories | COVID-19 Genomics UK (COG-UK) Consortium |
| EPI_ISL_453206 | Liverpool Clinical Laboratories | COVID-19 Genomics UK (COG-UK) Consortium |
| EPI_ISL_453207 | Liverpool Clinical Laboratories | COVID-19 Genomics UK (COG-UK) Consortium |
| EPI_ISL_453208 | Liverpool Clinical Laboratories | COVID-19 Genomics UK (COG-UK) Consortium |
| EPI_ISL_453209 | Liverpool Clinical Laboratories | COVID-19 Genomics UK (COG-UK) Consortium |
| EPI_ISL_453210 | Liverpool Clinical Laboratories | COVID-19 Genomics UK (COG-UK) Consortium |
| EPI_ISL_453211 | Liverpool Clinical Laboratories | COVID-19 Genomics UK (COG-UK) Consortium |
| EPI_ISL_453212 | Liverpool Clinical Laboratories | COVID-19 Genomics UK (COG-UK) Consortium |
| EPI_ISL_453213 | Liverpool Clinical Laboratories | COVID-19 Genomics UK (COG-UK) Consortium |
| EPI_ISL_453214 | Liverpool Clinical Laboratories | COVID-19 Genomics UK (COG-UK) Consortium |
| EPI_ISL_453215 | Liverpool Clinical Laboratories | COVID-19 Genomics UK (COG-UK) Consortium |
| EPI_ISL_453216 | Liverpool Clinical Laboratories | COVID-19 Genomics UK (COG-UK) Consortium |
| EPI_ISL_453217 | Liverpool Clinical Laboratories | COVID-19 Genomics UK (COG-UK) Consortium |
| EPI_ISL_453218 | Liverpool Clinical Laboratories | COVID-19 Genomics UK (COG-UK) Consortium |
| EPI_ISL_453219 | Liverpool Clinical Laboratories | COVID-19 Genomics UK (COG-UK) Consortium |
| EPI_ISL_453220 | Liverpool Clinical Laboratories | COVID-19 Genomics UK (COG-UK) Consortium |
| EPI_ISL_453221 | Liverpool Clinical Laboratories | COVID-19 Genomics UK (COG-UK) Consortium |
| EPI_ISL_453222 | Liverpool Clinical Laboratories | COVID-19 Genomics UK (COG-UK) Consortium |
| EPI_ISL_453223 | Liverpool Clinical Laboratories | COVID-19 Genomics UK (COG-UK) Consortium |
| EPI_ISL_453224 | Liverpool Clinical Laboratories | COVID-19 Genomics UK (COG-UK) Consortium |
| EPI_ISL_453225 | Liverpool Clinical Laboratories | COVID-19 Genomics UK (COG-UK) Consortium |
| EPI_ISL_453226 | Liverpool Clinical Laboratories | COVID-19 Genomics UK (COG-UK) Consortium |
| EPI_ISL_453227 | Liverpool Clinical Laboratories | COVID-19 Genomics UK (COG-UK) Consortium |
| EPI_ISL_453228 | Liverpool Clinical Laboratories | COVID-19 Genomics UK (COG-UK) Consortium |
| EPI_ISL_453229 | Liverpool Clinical Laboratories | COVID-19 Genomics UK (COG-UK) Consortium |
| EPI_ISL_453230 | Liverpool Clinical Laboratories | COVID-19 Genomics UK (COG-UK) Consortium |
| EPI_ISL_453231 | Liverpool Clinical Laboratories | COVID-19 Genomics UK (COG-UK) Consortium |
| EPI_ISL_453232 | Liverpool Clinical Laboratories | COVID-19 Genomics UK (COG-UK) Consortium |

| EPI_Accession_ID | Originating Laboratory | Submitting Laboratory |
|---|---|---|
| EPI_ISL_453233 | Liverpool Clinical Laboratories | COVID-19 Genomics UK (COG-UK) Consortium |
| EPI_ISL_453234 | Liverpool Clinical Laboratories | COVID-19 Genomics UK (COG-UK) Consortium |
| EPI_ISL_453235 | Liverpool Clinical Laboratories | COVID-19 Genomics UK (COG-UK) Consortium |
| EPI_ISL_453236 | Liverpool Clinical Laboratories | COVID-19 Genomics UK (COG-UK) Consortium |
| EPI_ISL_453237 | Liverpool Clinical Laboratories | COVID-19 Genomics UK (COG-UK) Consortium |
| EPI_ISL_453238 | Liverpool Clinical Laboratories | COVID-19 Genomics UK (COG-UK) Consortium |
| EPI_ISL_453239 | Liverpool Clinical Laboratories | COVID-19 Genomics UK (COG-UK) Consortium |
| EPI_ISL_453240 | Liverpool Clinical Laboratories | COVID-19 Genomics UK (COG-UK) Consortium |
| EPI_ISL_453241 | Liverpool Clinical Laboratories | COVID-19 Genomics UK (COG-UK) Consortium |
| EPI_ISL_453242 | Liverpool Clinical Laboratories | COVID-19 Genomics UK (COG-UK) Consortium |
| EPI_ISL_453243 | Liverpool Clinical Laboratories | COVID-19 Genomics UK (COG-UK) Consortium |
| EPI_ISL_453244 | Liverpool Clinical Laboratories | COVID-19 Genomics UK (COG-UK) Consortium |
| EPI_ISL_453245 | Liverpool Clinical Laboratories | COVID-19 Genomics UK (COG-UK) Consortium |
| EPI_ISL_453246 | Liverpool Clinical Laboratories | COVID-19 Genomics UK (COG-UK) Consortium |
| EPI_ISL_453247 | Liverpool Clinical Laboratories | COVID-19 Genomics UK (COG-UK) Consortium |
| EPI_ISL_453248 | Liverpool Clinical Laboratories | COVID-19 Genomics UK (COG-UK) Consortium |
| EPI_ISL_453249 | Liverpool Clinical Laboratories | COVID-19 Genomics UK (COG-UK) Consortium |
| EPI_ISL_453250 | Liverpool Clinical Laboratories | COVID-19 Genomics UK (COG-UK) Consortium |
| EPI_ISL_453251 | Liverpool Clinical Laboratories | COVID-19 Genomics UK (COG-UK) Consortium |
| EPI_ISL_453252 | Liverpool Clinical Laboratories | COVID-19 Genomics UK (COG-UK) Consortium |
| EPI_ISL_453253 | Liverpool Clinical Laboratories | COVID-19 Genomics UK (COG-UK) Consortium |
| EPI_ISL_453254 | Liverpool Clinical Laboratories | COVID-19 Genomics UK (COG-UK) Consortium |
| EPI_ISL_453255 | Liverpool Clinical Laboratories | COVID-19 Genomics UK (COG-UK) Consortium |
| EPI_ISL_453256 | Liverpool Clinical Laboratories | COVID-19 Genomics UK (COG-UK) Consortium |
| EPI_ISL_453257 | Liverpool Clinical Laboratories | COVID-19 Genomics UK (COG-UK) Consortium |
| EPI_ISL_453258 | Liverpool Clinical Laboratories | COVID-19 Genomics UK (COG-UK) Consortium |
| EPI_ISL_453259 | Liverpool Clinical Laboratories | COVID-19 Genomics UK (COG-UK) Consortium |
| EPI_ISL_453260 | Liverpool Clinical Laboratories | COVID-19 Genomics UK (COG-UK) Consortium |
| EPI_ISL_453261 | Liverpool Clinical Laboratories | COVID-19 Genomics UK (COG-UK) Consortium |
| EPI_ISL_453262 | Liverpool Clinical Laboratories | COVID-19 Genomics UK (COG-UK) Consortium |
| EPI_ISL_453263 | Liverpool Clinical Laboratories | COVID-19 Genomics UK (COG-UK) Consortium |
| EPI_ISL_453264 | Liverpool Clinical Laboratories | COVID-19 Genomics UK (COG-UK) Consortium |
| EPI_ISL_453265 | Liverpool Clinical Laboratories | COVID-19 Genomics UK (COG-UK) Consortium |

| EPI_Accession_ID | Originating Laboratory | Submitting Laboratory |
|---|---|---|
| EPI_ISL_453266 | Liverpool Clinical Laboratories | COVID-19 Genomics UK (COG-UK) Consortium |
| EPI_ISL_453267 | Liverpool Clinical Laboratories | COVID-19 Genomics UK (COG-UK) Consortium |
| EPI_ISL_453268 | Liverpool Clinical Laboratories | COVID-19 Genomics UK (COG-UK) Consortium |
| EPI_ISL_453269 | Liverpool Clinical Laboratories | COVID-19 Genomics UK (COG-UK) Consortium |
| EPI_ISL_453270 | Liverpool Clinical Laboratories | COVID-19 Genomics UK (COG-UK) Consortium |
| EPI_ISL_453271 | Liverpool Clinical Laboratories | COVID-19 Genomics UK (COG-UK) Consortium |
| EPI_ISL_453272 | Liverpool Clinical Laboratories | COVID-19 Genomics UK (COG-UK) Consortium |
| EPI_ISL_453273 | Liverpool Clinical Laboratories | COVID-19 Genomics UK (COG-UK) Consortium |
| EPI_ISL_453274 | Liverpool Clinical Laboratories | COVID-19 Genomics UK (COG-UK) Consortium |
| EPI_ISL_453275 | Liverpool Clinical Laboratories | COVID-19 Genomics UK (COG-UK) Consortium |
| EPI_ISL_453276 | Liverpool Clinical Laboratories | COVID-19 Genomics UK (COG-UK) Consortium |
| EPI_ISL_453277 | Liverpool Clinical Laboratories | COVID-19 Genomics UK (COG-UK) Consortium |
| EPI_ISL_453278 | Liverpool Clinical Laboratories | COVID-19 Genomics UK (COG-UK) Consortium |
| EPI_ISL_453279 | Liverpool Clinical Laboratories | COVID-19 Genomics UK (COG-UK) Consortium |
| EPI_ISL_453280 | Liverpool Clinical Laboratories | COVID-19 Genomics UK (COG-UK) Consortium |
| EPI_ISL_453281 | Liverpool Clinical Laboratories | COVID-19 Genomics UK (COG-UK) Consortium |
| EPI_ISL_453282 | Liverpool Clinical Laboratories | COVID-19 Genomics UK (COG-UK) Consortium |
| EPI_ISL_453283 | Liverpool Clinical Laboratories | COVID-19 Genomics UK (COG-UK) Consortium |
| EPI_ISL_453284 | Liverpool Clinical Laboratories | COVID-19 Genomics UK (COG-UK) Consortium |
| EPI_ISL_453285 | Liverpool Clinical Laboratories | COVID-19 Genomics UK (COG-UK) Consortium |
| EPI_ISL_453286 | Liverpool Clinical Laboratories | COVID-19 Genomics UK (COG-UK) Consortium |
| EPI_ISL_453287 | Liverpool Clinical Laboratories | COVID-19 Genomics UK (COG-UK) Consortium |
| EPI_ISL_453288 | Liverpool Clinical Laboratories | COVID-19 Genomics UK (COG-UK) Consortium |
| EPI_ISL_453289 | Liverpool Clinical Laboratories | COVID-19 Genomics UK (COG-UK) Consortium |
| EPI_ISL_453290 | Liverpool Clinical Laboratories | COVID-19 Genomics UK (COG-UK) Consortium |
| EPI_ISL_453291 | Liverpool Clinical Laboratories | COVID-19 Genomics UK (COG-UK) Consortium |
| EPI_ISL_453292 | Liverpool Clinical Laboratories | COVID-19 Genomics UK (COG-UK) Consortium |
| EPI_ISL_453293 | Liverpool Clinical Laboratories | COVID-19 Genomics UK (COG-UK) Consortium |
| EPI_ISL_453294 | Liverpool Clinical Laboratories | COVID-19 Genomics UK (COG-UK) Consortium |
| EPI_ISL_453295 | Liverpool Clinical Laboratories | COVID-19 Genomics UK (COG-UK) Consortium |
| EPI_ISL_453296 | Liverpool Clinical Laboratories | COVID-19 Genomics UK (COG-UK) Consortium |
| EPI_ISL_453297 | Liverpool Clinical Laboratories | COVID-19 Genomics UK (COG-UK) Consortium |
| EPI_ISL_453298 | Liverpool Clinical Laboratories | COVID-19 Genomics UK (COG-UK) Consortium |

| EPI_Accession_ID | Originating Laboratory | Submitting Laboratory |
|---|---|---|
| EPI_ISL_453299 | Liverpool Clinical Laboratories | COVID-19 Genomics UK (COG-UK) Consortium |
| EPI_ISL_453300 | Liverpool Clinical Laboratories | COVID-19 Genomics UK (COG-UK) Consortium |
| EPI_ISL_453301 | Liverpool Clinical Laboratories | COVID-19 Genomics UK (COG-UK) Consortium |
| EPI_ISL_453302 | Liverpool Clinical Laboratories | COVID-19 Genomics UK (COG-UK) Consortium |
| EPI_ISL_453303 | Liverpool Clinical Laboratories | COVID-19 Genomics UK (COG-UK) Consortium |
| EPI_ISL_453304 | Liverpool Clinical Laboratories | COVID-19 Genomics UK (COG-UK) Consortium |
| EPI_ISL_453305 | Liverpool Clinical Laboratories | COVID-19 Genomics UK (COG-UK) Consortium |
| EPI_ISL_453306 | Liverpool Clinical Laboratories | COVID-19 Genomics UK (COG-UK) Consortium |
| EPI_ISL_453307 | Liverpool Clinical Laboratories | COVID-19 Genomics UK (COG-UK) Consortium |
| EPI_ISL_453308 | Liverpool Clinical Laboratories | COVID-19 Genomics UK (COG-UK) Consortium |
| EPI_ISL_453309 | Liverpool Clinical Laboratories | COVID-19 Genomics UK (COG-UK) Consortium |
| EPI_ISL_453310 | Liverpool Clinical Laboratories | COVID-19 Genomics UK (COG-UK) Consortium |
| EPI_ISL_453311 | Liverpool Clinical Laboratories | COVID-19 Genomics UK (COG-UK) Consortium |
| EPI_ISL_453312 | Liverpool Clinical Laboratories | COVID-19 Genomics UK (COG-UK) Consortium |
| EPI_ISL_453313 | Liverpool Clinical Laboratories | COVID-19 Genomics UK (COG-UK) Consortium |
| EPI_ISL_453314 | Liverpool Clinical Laboratories | COVID-19 Genomics UK (COG-UK) Consortium |
| EPI_ISL_453315 | Liverpool Clinical Laboratories | COVID-19 Genomics UK (COG-UK) Consortium |
| EPI_ISL_453316 | Liverpool Clinical Laboratories | COVID-19 Genomics UK (COG-UK) Consortium |
| EPI_ISL_453317 | Liverpool Clinical Laboratories | COVID-19 Genomics UK (COG-UK) Consortium |
| EPI_ISL_453318 | Liverpool Clinical Laboratories | COVID-19 Genomics UK (COG-UK) Consortium |
| EPI_ISL_453319 | Liverpool Clinical Laboratories | COVID-19 Genomics UK (COG-UK) Consortium |
| EPI_ISL_453320 | Liverpool Clinical Laboratories | COVID-19 Genomics UK (COG-UK) Consortium |
| EPI_ISL_453321 | Liverpool Clinical Laboratories | COVID-19 Genomics UK (COG-UK) Consortium |
| EPI_ISL_453322 | Liverpool Clinical Laboratories | COVID-19 Genomics UK (COG-UK) Consortium |
| EPI_ISL_453323 | Liverpool Clinical Laboratories | COVID-19 Genomics UK (COG-UK) Consortium |
| EPI_ISL_453324 | Liverpool Clinical Laboratories | COVID-19 Genomics UK (COG-UK) Consortium |
| EPI_ISL_453325 | Liverpool Clinical Laboratories | COVID-19 Genomics UK (COG-UK) Consortium |
| EPI_ISL_453326 | Liverpool Clinical Laboratories | COVID-19 Genomics UK (COG-UK) Consortium |
| EPI_ISL_453327 | Liverpool Clinical Laboratories | COVID-19 Genomics UK (COG-UK) Consortium |
| EPI_ISL_453328 | Liverpool Clinical Laboratories | COVID-19 Genomics UK (COG-UK) Consortium |
| EPI_ISL_453329 | Liverpool Clinical Laboratories | COVID-19 Genomics UK (COG-UK) Consortium |
| EPI_ISL_453330 | Liverpool Clinical Laboratories | COVID-19 Genomics UK (COG-UK) Consortium |
| EPI_ISL_453331 | Liverpool Clinical Laboratories | COVID-19 Genomics UK (COG-UK) Consortium |

| EPI_Accession_ID | Originating Laboratory | Submitting Laboratory |
|---|---|---|
| EPI_ISL_453332 | Liverpool Clinical Laboratories | COVID-19 Genomics UK (COG-UK) Consortium |
| EPI_ISL_453333 | Liverpool Clinical Laboratories | COVID-19 Genomics UK (COG-UK) Consortium |
| EPI_ISL_453334 | Liverpool Clinical Laboratories | COVID-19 Genomics UK (COG-UK) Consortium |
| EPI_ISL_453335 | Liverpool Clinical Laboratories | COVID-19 Genomics UK (COG-UK) Consortium |
| EPI_ISL_453336 | Liverpool Clinical Laboratories | COVID-19 Genomics UK (COG-UK) Consortium |
| EPI_ISL_453337 | Liverpool Clinical Laboratories | COVID-19 Genomics UK (COG-UK) Consortium |
| EPI_ISL_453338 | Liverpool Clinical Laboratories | COVID-19 Genomics UK (COG-UK) Consortium |
| EPI_ISL_453339 | Liverpool Clinical Laboratories | COVID-19 Genomics UK (COG-UK) Consortium |
| EPI_ISL_453340 | Liverpool Clinical Laboratories | COVID-19 Genomics UK (COG-UK) Consortium |
| EPI_ISL_453341 | Liverpool Clinical Laboratories | COVID-19 Genomics UK (COG-UK) Consortium |
| EPI_ISL_453342 | Liverpool Clinical Laboratories | COVID-19 Genomics UK (COG-UK) Consortium |
| EPI_ISL_453343 | Liverpool Clinical Laboratories | COVID-19 Genomics UK (COG-UK) Consortium |
| EPI_ISL_453344 | Liverpool Clinical Laboratories | COVID-19 Genomics UK (COG-UK) Consortium |
| EPI_ISL_453345 | Liverpool Clinical Laboratories | COVID-19 Genomics UK (COG-UK) Consortium |
| EPI_ISL_453346 | Liverpool Clinical Laboratories | COVID-19 Genomics UK (COG-UK) Consortium |
| EPI_ISL_453347 | Liverpool Clinical Laboratories | COVID-19 Genomics UK (COG-UK) Consortium |
| EPI_ISL_453348 | Liverpool Clinical Laboratories | COVID-19 Genomics UK (COG-UK) Consortium |
| EPI_ISL_453349 | Liverpool Clinical Laboratories | COVID-19 Genomics UK (COG-UK) Consortium |
| EPI_ISL_453350 | Liverpool Clinical Laboratories | COVID-19 Genomics UK (COG-UK) Consortium |
| EPI_ISL_453351 | Liverpool Clinical Laboratories | COVID-19 Genomics UK (COG-UK) Consortium |
| EPI_ISL_453352 | Liverpool Clinical Laboratories | COVID-19 Genomics UK (COG-UK) Consortium |
| EPI_ISL_453353 | Liverpool Clinical Laboratories | COVID-19 Genomics UK (COG-UK) Consortium |
| EPI_ISL_453354 | Liverpool Clinical Laboratories | COVID-19 Genomics UK (COG-UK) Consortium |
| EPI_ISL_453355 | Liverpool Clinical Laboratories | COVID-19 Genomics UK (COG-UK) Consortium |
| EPI_ISL_453356 | Liverpool Clinical Laboratories | COVID-19 Genomics UK (COG-UK) Consortium |
| EPI_ISL_453357 | Liverpool Clinical Laboratories | COVID-19 Genomics UK (COG-UK) Consortium |
| EPI_ISL_453358 | Liverpool Clinical Laboratories | COVID-19 Genomics UK (COG-UK) Consortium |
| EPI_ISL_453359 | Liverpool Clinical Laboratories | COVID-19 Genomics UK (COG-UK) Consortium |
| EPI_ISL_453360 | Liverpool Clinical Laboratories | COVID-19 Genomics UK (COG-UK) Consortium |
| EPI_ISL_453361 | Liverpool Clinical Laboratories | COVID-19 Genomics UK (COG-UK) Consortium |
| EPI_ISL_453362 | Liverpool Clinical Laboratories | COVID-19 Genomics UK (COG-UK) Consortium |
| EPI_ISL_453363 | Liverpool Clinical Laboratories | COVID-19 Genomics UK (COG-UK) Consortium |
| EPI_ISL_453364 | Liverpool Clinical Laboratories | COVID-19 Genomics UK (COG-UK) Consortium |

| EPI_Accession_ID | Originating Laboratory | Submitting Laboratory |
|---|---|---|
| EPI_ISL_453365 | Liverpool Clinical Laboratories | COVID-19 Genomics UK (COG-UK) Consortium |
| EPI_ISL_453366 | Liverpool Clinical Laboratories | COVID-19 Genomics UK (COG-UK) Consortium |
| EPI_ISL_453367 | Liverpool Clinical Laboratories | COVID-19 Genomics UK (COG-UK) Consortium |
| EPI_ISL_453368 | Liverpool Clinical Laboratories | COVID-19 Genomics UK (COG-UK) Consortium |
| EPI_ISL_453369 | Liverpool Clinical Laboratories | COVID-19 Genomics UK (COG-UK) Consortium |
| EPI_ISL_453370 | Liverpool Clinical Laboratories | COVID-19 Genomics UK (COG-UK) Consortium |
| EPI_ISL_453371 | Liverpool Clinical Laboratories | COVID-19 Genomics UK (COG-UK) Consortium |
| EPI_ISL_453372 | Liverpool Clinical Laboratories | COVID-19 Genomics UK (COG-UK) Consortium |
| EPI_ISL_453373 | Liverpool Clinical Laboratories | COVID-19 Genomics UK (COG-UK) Consortium |
| EPI_ISL_453374 | Liverpool Clinical Laboratories | COVID-19 Genomics UK (COG-UK) Consortium |
| EPI_ISL_453375 | Liverpool Clinical Laboratories | COVID-19 Genomics UK (COG-UK) Consortium |
| EPI_ISL_453376 | Liverpool Clinical Laboratories | COVID-19 Genomics UK (COG-UK) Consortium |
| EPI_ISL_453377 | Liverpool Clinical Laboratories | COVID-19 Genomics UK (COG-UK) Consortium |
| EPI_ISL_453378 | Liverpool Clinical Laboratories | COVID-19 Genomics UK (COG-UK) Consortium |
| EPI_ISL_453379 | Liverpool Clinical Laboratories | COVID-19 Genomics UK (COG-UK) Consortium |
| EPI_ISL_453380 | Liverpool Clinical Laboratories | COVID-19 Genomics UK (COG-UK) Consortium |
| EPI_ISL_453381 | Liverpool Clinical Laboratories | COVID-19 Genomics UK (COG-UK) Consortium |
| EPI_ISL_453382 | Liverpool Clinical Laboratories | COVID-19 Genomics UK (COG-UK) Consortium |
| EPI_ISL_453383 | Liverpool Clinical Laboratories | COVID-19 Genomics UK (COG-UK) Consortium |
| EPI_ISL_453384 | Liverpool Clinical Laboratories | COVID-19 Genomics UK (COG-UK) Consortium |
| EPI_ISL_453385 | Liverpool Clinical Laboratories | COVID-19 Genomics UK (COG-UK) Consortium |
| EPI_ISL_453386 | Liverpool Clinical Laboratories | COVID-19 Genomics UK (COG-UK) Consortium |
| EPI_ISL_453387 | Liverpool Clinical Laboratories | COVID-19 Genomics UK (COG-UK) Consortium |
| EPI_ISL_453388 | Liverpool Clinical Laboratories | COVID-19 Genomics UK (COG-UK) Consortium |
| EPI_ISL_453389 | Liverpool Clinical Laboratories | COVID-19 Genomics UK (COG-UK) Consortium |
| EPI_ISL_453390 | Liverpool Clinical Laboratories | COVID-19 Genomics UK (COG-UK) Consortium |
| EPI_ISL_453391 | Liverpool Clinical Laboratories | COVID-19 Genomics UK (COG-UK) Consortium |
| EPI_ISL_453392 | Liverpool Clinical Laboratories | COVID-19 Genomics UK (COG-UK) Consortium |
| EPI_ISL_453393 | Liverpool Clinical Laboratories | COVID-19 Genomics UK (COG-UK) Consortium |
| EPI_ISL_453394 | Liverpool Clinical Laboratories | COVID-19 Genomics UK (COG-UK) Consortium |
| EPI_ISL_453395 | Liverpool Clinical Laboratories | COVID-19 Genomics UK (COG-UK) Consortium |
| EPI_ISL_453396 | Liverpool Clinical Laboratories | COVID-19 Genomics UK (COG-UK) Consortium |
| EPI_ISL_453397 | Liverpool Clinical Laboratories | COVID-19 Genomics UK (COG-UK) Consortium |

| EPI_Accession_ID | Originating Laboratory | Submitting Laboratory |
|---|---|---|
| EPI_ISL_453398 | Liverpool Clinical Laboratories | COVID-19 Genomics UK (COG-UK) Consortium |
| EPI_ISL_453399 | Liverpool Clinical Laboratories | COVID-19 Genomics UK (COG-UK) Consortium |
| EPI_ISL_453400 | Liverpool Clinical Laboratories | COVID-19 Genomics UK (COG-UK) Consortium |
| EPI_ISL_453401 | Liverpool Clinical Laboratories | COVID-19 Genomics UK (COG-UK) Consortium |
| EPI_ISL_453402 | Liverpool Clinical Laboratories | COVID-19 Genomics UK (COG-UK) Consortium |
| EPI_ISL_453403 | Liverpool Clinical Laboratories | COVID-19 Genomics UK (COG-UK) Consortium |
| EPI_ISL_453404 | Liverpool Clinical Laboratories | COVID-19 Genomics UK (COG-UK) Consortium |
| EPI_ISL_453405 | Liverpool Clinical Laboratories | COVID-19 Genomics UK (COG-UK) Consortium |
| EPI_ISL_453406 | Liverpool Clinical Laboratories | COVID-19 Genomics UK (COG-UK) Consortium |
| EPI_ISL_453407 | Liverpool Clinical Laboratories | COVID-19 Genomics UK (COG-UK) Consortium |
| EPI_ISL_453408 | Liverpool Clinical Laboratories | COVID-19 Genomics UK (COG-UK) Consortium |
| EPI_ISL_453409 | Liverpool Clinical Laboratories | COVID-19 Genomics UK (COG-UK) Consortium |
| EPI_ISL_453410 | Liverpool Clinical Laboratories | COVID-19 Genomics UK (COG-UK) Consortium |
| EPI_ISL_453411 | Liverpool Clinical Laboratories | COVID-19 Genomics UK (COG-UK) Consortium |
| EPI_ISL_453412 | Liverpool Clinical Laboratories | COVID-19 Genomics UK (COG-UK) Consortium |
| EPI_ISL_453413 | Liverpool Clinical Laboratories | COVID-19 Genomics UK (COG-UK) Consortium |
| EPI_ISL_453414 | Liverpool Clinical Laboratories | COVID-19 Genomics UK (COG-UK) Consortium |
| EPI_ISL_453415 | Liverpool Clinical Laboratories | COVID-19 Genomics UK (COG-UK) Consortium |
| EPI_ISL_453416 | Liverpool Clinical Laboratories | COVID-19 Genomics UK (COG-UK) Consortium |
| EPI_ISL_453417 | Liverpool Clinical Laboratories | COVID-19 Genomics UK (COG-UK) Consortium |
| EPI_ISL_453418 | Liverpool Clinical Laboratories | COVID-19 Genomics UK (COG-UK) Consortium |
| EPI_ISL_453419 | Liverpool Clinical Laboratories | COVID-19 Genomics UK (COG-UK) Consortium |
| EPI_ISL_453420 | Liverpool Clinical Laboratories | COVID-19 Genomics UK (COG-UK) Consortium |
| EPI_ISL_453421 | Liverpool Clinical Laboratories | COVID-19 Genomics UK (COG-UK) Consortium |
| EPI_ISL_453422 | Liverpool Clinical Laboratories | COVID-19 Genomics UK (COG-UK) Consortium |
| EPI_ISL_453423 | Liverpool Clinical Laboratories | COVID-19 Genomics UK (COG-UK) Consortium |
| EPI_ISL_453424 | Liverpool Clinical Laboratories | COVID-19 Genomics UK (COG-UK) Consortium |
| EPI_ISL_453425 | Liverpool Clinical Laboratories | COVID-19 Genomics UK (COG-UK) Consortium |
| EPI_ISL_453426 | Liverpool Clinical Laboratories | COVID-19 Genomics UK (COG-UK) Consortium |
| EPI_ISL_453427 | Liverpool Clinical Laboratories | COVID-19 Genomics UK (COG-UK) Consortium |
| EPI_ISL_453428 | Liverpool Clinical Laboratories | COVID-19 Genomics UK (COG-UK) Consortium |
| EPI_ISL_453429 | Liverpool Clinical Laboratories | COVID-19 Genomics UK (COG-UK) Consortium |
| EPI_ISL_453430 | Liverpool Clinical Laboratories | COVID-19 Genomics UK (COG-UK) Consortium |

| EPI_Accession_ID | Originating Laboratory | Submitting Laboratory |
| --- | --- | --- |
| EPI_ISL_453431 | Liverpool Clinical Laboratories | COVID-19 Genomics UK (COG-UK) Consortium |
| EPI_ISL_453432 | Liverpool Clinical Laboratories | COVID-19 Genomics UK (COG-UK) Consortium |
| EPI_ISL_453433 | Liverpool Clinical Laboratories | COVID-19 Genomics UK (COG-UK) Consortium |
| EPI_ISL_453434 | Liverpool Clinical Laboratories | COVID-19 Genomics UK (COG-UK) Consortium |
| EPI_ISL_453435 | Liverpool Clinical Laboratories | COVID-19 Genomics UK (COG-UK) Consortium |
| EPI_ISL_453436 | Liverpool Clinical Laboratories | COVID-19 Genomics UK (COG-UK) Consortium |
| EPI_ISL_453437 | Liverpool Clinical Laboratories | COVID-19 Genomics UK (COG-UK) Consortium |
| EPI_ISL_453438 | Liverpool Clinical Laboratories | COVID-19 Genomics UK (COG-UK) Consortium |
| EPI_ISL_453439 | Liverpool Clinical Laboratories | COVID-19 Genomics UK (COG-UK) Consortium |
| EPI_ISL_453440 | Liverpool Clinical Laboratories | COVID-19 Genomics UK (COG-UK) Consortium |
| EPI_ISL_453441 | Liverpool Clinical Laboratories | COVID-19 Genomics UK (COG-UK) Consortium |
| EPI_ISL_453442 | Liverpool Clinical Laboratories | COVID-19 Genomics UK (COG-UK) Consortium |
| EPI_ISL_453443 | Liverpool Clinical Laboratories | COVID-19 Genomics UK (COG-UK) Consortium |
| EPI_ISL_453444 | Liverpool Clinical Laboratories | COVID-19 Genomics UK (COG-UK) Consortium |
| EPI_ISL_453445 | Liverpool Clinical Laboratories | COVID-19 Genomics UK (COG-UK) Consortium |
| EPI_ISL_453446 | Liverpool Clinical Laboratories | COVID-19 Genomics UK (COG-UK) Consortium |
| EPI_ISL_453447 | Liverpool Clinical Laboratories | COVID-19 Genomics UK (COG-UK) Consortium |
| EPI_ISL_453448 | Liverpool Clinical Laboratories | COVID-19 Genomics UK (COG-UK) Consortium |
| EPI_ISL_453449 | Liverpool Clinical Laboratories | COVID-19 Genomics UK (COG-UK) Consortium |
| EPI_ISL_453450 | Liverpool Clinical Laboratories | COVID-19 Genomics UK (COG-UK) Consortium |
| EPI_ISL_453451 | Liverpool Clinical Laboratories | COVID-19 Genomics UK (COG-UK) Consortium |
| EPI_ISL_453452 | Liverpool Clinical Laboratories | COVID-19 Genomics UK (COG-UK) Consortium |
| EPI_ISL_453453 | Liverpool Clinical Laboratories | COVID-19 Genomics UK (COG-UK) Consortium |
| EPI_ISL_453454 | Liverpool Clinical Laboratories | COVID-19 Genomics UK (COG-UK) Consortium |
| EPI_ISL_453455 | Liverpool Clinical Laboratories | COVID-19 Genomics UK (COG-UK) Consortium |
| EPI_ISL_453456 | Liverpool Clinical Laboratories | COVID-19 Genomics UK (COG-UK) Consortium |
| EPI_ISL_453457 | Liverpool Clinical Laboratories | COVID-19 Genomics UK (COG-UK) Consortium |
| EPI_ISL_453458 | Liverpool Clinical Laboratories | COVID-19 Genomics UK (COG-UK) Consortium |
| EPI_ISL_453459 | Liverpool Clinical Laboratories | COVID-19 Genomics UK (COG-UK) Consortium |
| EPI_ISL_453460 | University College London, Great Ormond Street Hospital for Children NHS Foundation Trust, Imperial College Healthcare NHS Trust | COVID-19 Genomics UK (COG-UK) Consortium |

| EPI_Accession_ID | Originating Laboratory | Submitting Laboratory |
|---|---|---|
| EPI_ISL_453461 | University College London, Great Ormond Street Hospital for Children NHS Foundation Trust, Imperial College Healthcare NHS Trust | COVID-19 Genomics UK (COG-UK) Consortium |
| EPI_ISL_453462 | University College London, Great Ormond Street Hospital for Children NHS Foundation Trust, Imperial College Healthcare NHS Trust | COVID-19 Genomics UK (COG-UK) Consortium |
| EPI_ISL_453463 | University College London, Great Ormond Street Hospital for Children NHS Foundation Trust, Imperial College Healthcare NHS Trust | COVID-19 Genomics UK (COG-UK) Consortium |
| EPI_ISL_453464 | University College London, Great Ormond Street Hospital for Children NHS Foundation Trust, Imperial College Healthcare NHS Trust | COVID-19 Genomics UK (COG-UK) Consortium |
| EPI_ISL_453465 | Regional Virus Laboratory, Belfast Health and Social Care Trust | COVID-19 Genomics UK (COG-UK) Consortium |
| EPI_ISL_453466 | Regional Virus Laboratory, Belfast Health and Social Care Trust | COVID-19 Genomics UK (COG-UK) Consortium |
| EPI_ISL_453467 | Regional Virus Laboratory, Belfast Health and Social Care Trust | COVID-19 Genomics UK (COG-UK) Consortium |
| EPI_ISL_453468 | Regional Virus Laboratory, Belfast Health and Social Care Trust | COVID-19 Genomics UK (COG-UK) Consortium |
| EPI_ISL_453469 | Regional Virus Laboratory, Belfast Health and Social Care Trust | COVID-19 Genomics UK (COG-UK) Consortium |
| EPI_ISL_453470 | Regional Virus Laboratory, Belfast Health and Social Care Trust | COVID-19 Genomics UK (COG-UK) Consortium |
| EPI_ISL_453471 | Regional Virus Laboratory, Belfast Health and Social Care Trust | COVID-19 Genomics UK (COG-UK) Consortium |
| EPI_ISL_453472 | Regional Virus Laboratory, Belfast Health and Social Care Trust | COVID-19 Genomics UK (COG-UK) Consortium |
| EPI_ISL_453473 | Regional Virus Laboratory, Belfast Health and Social Care Trust | COVID-19 Genomics UK (COG-UK) Consortium |
| EPI_ISL_453474 | Regional Virus Laboratory, Belfast Health and Social Care Trust | COVID-19 Genomics UK (COG-UK) Consortium |

| EPI_Accession_ID | Originating Laboratory | Submitting Laboratory |
|---|---|---|
| EPI_ISL_453475 | Regional Virus Laboratory, Belfast Health and Social Care Trust | COVID-19 Genomics UK (COG-UK) Consortium |
| EPI_ISL_453476 | Regional Virus Laboratory, Belfast Health and Social Care Trust | COVID-19 Genomics UK (COG-UK) Consortium |
| EPI_ISL_453477 | Regional Virus Laboratory, Belfast Health and Social Care Trust | COVID-19 Genomics UK (COG-UK) Consortium |
| EPI_ISL_453478 | Regional Virus Laboratory, Belfast Health and Social Care Trust | COVID-19 Genomics UK (COG-UK) Consortium |
| EPI_ISL_453479 | Regional Virus Laboratory, Belfast Health and Social Care Trust | COVID-19 Genomics UK (COG-UK) Consortium |
| EPI_ISL_453480 | Regional Virus Laboratory, Belfast Health and Social Care Trust | COVID-19 Genomics UK (COG-UK) Consortium |
| EPI_ISL_453481 | Regional Virus Laboratory, Belfast Health and Social Care Trust | COVID-19 Genomics UK (COG-UK) Consortium |
| EPI_ISL_453482 | Regional Virus Laboratory, Belfast Health and Social Care Trust | COVID-19 Genomics UK (COG-UK) Consortium |
| EPI_ISL_453483 | Regional Virus Laboratory, Belfast Health and Social Care Trust | COVID-19 Genomics UK (COG-UK) Consortium |
| EPI_ISL_453484 | Regional Virus Laboratory, Belfast Health and Social Care Trust | COVID-19 Genomics UK (COG-UK) Consortium |
| EPI_ISL_453485 | Regional Virus Laboratory, Belfast Health and Social Care Trust | COVID-19 Genomics UK (COG-UK) Consortium |
| EPI_ISL_453486 | Northumbria University / South Tees Hospitals NHS Foundation Trust / North Cumbria Integrated Care NHS Foundation Trust / North Tees and Hartlepool NHS Foundation Trust / Newcastle Hospitals NHS Foundation Trust | COVID-19 Genomics UK (COG-UK) Consortium |
| EPI_ISL_453487 | Northumbria University / South Tees Hospitals NHS Foundation Trust / North Cumbria Integrated Care NHS Foundation Trust / North Tees and Hartlepool NHS Foundation Trust / Newcastle Hospitals NHS Foundation Trust | COVID-19 Genomics UK (COG-UK) Consortium |

| EPI_Accession_ID | Originating Laboratory | Submitting Laboratory |
|---|---|---|
| EPI_ISL_453488 | Northumbria University / South Tees Hospitals NHS Foundation Trust / North Cumbria Integrated Care NHS Foundation Trust / North Tees and Hartlepool NHS Foundation Trust / Newcastle Hospitals NHS Foundation Trust | COVID-19 Genomics UK (COG-UK) Consortium |
| EPI_ISL_453489 | Northumbria University / South Tees Hospitals NHS Foundation Trust / North Cumbria Integrated Care NHS Foundation Trust / North Tees and Hartlepool NHS Foundation Trust / Newcastle Hospitals NHS Foundation Trust | COVID-19 Genomics UK (COG-UK) Consortium |
| EPI_ISL_453490 | Northumbria University / South Tees Hospitals NHS Foundation Trust / North Cumbria Integrated Care NHS Foundation Trust / North Tees and Hartlepool NHS Foundation Trust / Newcastle Hospitals NHS Foundation Trust | COVID-19 Genomics UK (COG-UK) Consortium |
| EPI_ISL_453491 | Northumbria University / South Tees Hospitals NHS Foundation Trust / North Cumbria Integrated Care NHS Foundation Trust / North Tees and Hartlepool NHS Foundation Trust / Newcastle Hospitals NHS Foundation Trust | COVID-19 Genomics UK (COG-UK) Consortium |
| EPI_ISL_453492 | Northumbria University / South Tees Hospitals NHS Foundation Trust / North Cumbria Integrated Care NHS Foundation Trust / North Tees and Hartlepool NHS Foundation Trust / Newcastle Hospitals NHS Foundation Trust | COVID-19 Genomics UK (COG-UK) Consortium |
| EPI_ISL_453493 | Northumbria University / South Tees Hospitals NHS Foundation Trust / North Cumbria Integrated Care NHS Foundation Trust / North Tees and Hartlepool NHS Foundation Trust / Newcastle Hospitals NHS Foundation Trust | COVID-19 Genomics UK (COG-UK) Consortium |

| EPI_Accession_ID | Originating Laboratory | Submitting Laboratory |
|---|---|---|
| EPI_ISL_453494 | Northumbria University / South Tees Hospitals NHS Foundation Trust / North Cumbria Integrated Care NHS Foundation Trust / North Tees and Hartlepool NHS Foundation Trust / Newcastle Hospitals NHS Foundation Trust | COVID-19 Genomics UK (COG-UK) Consortium |
| EPI_ISL_453495 | Northumbria University / South Tees Hospitals NHS Foundation Trust / North Cumbria Integrated Care NHS Foundation Trust / North Tees and Hartlepool NHS Foundation Trust / Newcastle Hospitals NHS Foundation Trust | COVID-19 Genomics UK (COG-UK) Consortium |
| EPI_ISL_453496 | Northumbria University / South Tees Hospitals NHS Foundation Trust / North Cumbria Integrated Care NHS Foundation Trust / North Tees and Hartlepool NHS Foundation Trust / Newcastle Hospitals NHS Foundation Trust | COVID-19 Genomics UK (COG-UK) Consortium |
| EPI_ISL_453497 | Northumbria University / South Tees Hospitals NHS Foundation Trust / North Cumbria Integrated Care NHS Foundation Trust / North Tees and Hartlepool NHS Foundation Trust / Newcastle Hospitals NHS Foundation Trust | COVID-19 Genomics UK (COG-UK) Consortium |
| EPI_ISL_453498 | Northumbria University / South Tees Hospitals NHS Foundation Trust / North Cumbria Integrated Care NHS Foundation Trust / North Tees and Hartlepool NHS Foundation Trust / Newcastle Hospitals NHS Foundation Trust | COVID-19 Genomics UK (COG-UK) Consortium |
| EPI_ISL_453499 | Northumbria University / South Tees Hospitals NHS Foundation Trust / North Cumbria Integrated Care NHS Foundation Trust / North Tees and Hartlepool NHS Foundation Trust / Newcastle Hospitals NHS Foundation Trust | COVID-19 Genomics UK (COG-UK) Consortium |

| EPI_Accession_ID | Originating Laboratory | Submitting Laboratory |
|---|---|---|
| EPI_ISL_453500 | Northumbria University / South Tees Hospitals NHS Foundation Trust / North Cumbria Integrated Care NHS Foundation Trust / North Tees and Hartlepool NHS Foundation Trust / Newcastle Hospitals NHS Foundation Trust | COVID-19 Genomics UK (COG-UK) Consortium |
| EPI_ISL_453501 | Northumbria University / South Tees Hospitals NHS Foundation Trust / North Cumbria Integrated Care NHS Foundation Trust / North Tees and Hartlepool NHS Foundation Trust / Newcastle Hospitals NHS Foundation Trust | COVID-19 Genomics UK (COG-UK) Consortium |
| EPI_ISL_453502 | Northumbria University / South Tees Hospitals NHS Foundation Trust / North Cumbria Integrated Care NHS Foundation Trust / North Tees and Hartlepool NHS Foundation Trust / Newcastle Hospitals NHS Foundation Trust | COVID-19 Genomics UK (COG-UK) Consortium |
| EPI_ISL_453503 | Northumbria University / South Tees Hospitals NHS Foundation Trust / North Cumbria Integrated Care NHS Foundation Trust / North Tees and Hartlepool NHS Foundation Trust / Newcastle Hospitals NHS Foundation Trust | COVID-19 Genomics UK (COG-UK) Consortium |
| EPI_ISL_453504 | Northumbria University / South Tees Hospitals NHS Foundation Trust / North Cumbria Integrated Care NHS Foundation Trust / North Tees and Hartlepool NHS Foundation Trust / Newcastle Hospitals NHS Foundation Trust | COVID-19 Genomics UK (COG-UK) Consortium |
| EPI_ISL_453505 | Northumbria University / South Tees Hospitals NHS Foundation Trust / North Cumbria Integrated Care NHS Foundation Trust / North Tees and Hartlepool NHS Foundation Trust / Newcastle Hospitals NHS Foundation Trust | COVID-19 Genomics UK (COG-UK) Consortium |

| EPI_Accession_ID | Originating Laboratory | Submitting Laboratory |
|---|---|---|
| EPI_ISL_453506 | Northumbria University / South Tees Hospitals NHS Foundation Trust / North Cumbria Integrated Care NHS Foundation Trust / North Tees and Hartlepool NHS Foundation Trust / Newcastle Hospitals NHS Foundation Trust | COVID-19 Genomics UK (COG-UK) Consortium |
| EPI_ISL_453507 | Northumbria University / South Tees Hospitals NHS Foundation Trust / North Cumbria Integrated Care NHS Foundation Trust / North Tees and Hartlepool NHS Foundation Trust / Newcastle Hospitals NHS Foundation Trust | COVID-19 Genomics UK (COG-UK) Consortium |
| EPI_ISL_453508 | Northumbria University / South Tees Hospitals NHS Foundation Trust / North Cumbria Integrated Care NHS Foundation Trust / North Tees and Hartlepool NHS Foundation Trust / Newcastle Hospitals NHS Foundation Trust | COVID-19 Genomics UK (COG-UK) Consortium |
| EPI_ISL_453509 | Northumbria University / South Tees Hospitals NHS Foundation Trust / North Cumbria Integrated Care NHS Foundation Trust / North Tees and Hartlepool NHS Foundation Trust / Newcastle Hospitals NHS Foundation Trust | COVID-19 Genomics UK (COG-UK) Consortium |
| EPI_ISL_453510 | Northumbria University / South Tees Hospitals NHS Foundation Trust / North Cumbria Integrated Care NHS Foundation Trust / North Tees and Hartlepool NHS Foundation Trust / Newcastle Hospitals NHS Foundation Trust | COVID-19 Genomics UK (COG-UK) Consortium |
| EPI_ISL_453511 | Northumbria University / South Tees Hospitals NHS Foundation Trust / North Cumbria Integrated Care NHS Foundation Trust / North Tees and Hartlepool NHS Foundation Trust / Newcastle Hospitals NHS Foundation Trust | COVID-19 Genomics UK (COG-UK) Consortium |

| EPI_Accession_ID | Originating Laboratory | Submitting Laboratory |
|---|---|---|
| EPI_ISL_453512 | Northumbria University / South Tees Hospitals NHS Foundation Trust / North Cumbria Integrated Care NHS Foundation Trust / North Tees and Hartlepool NHS Foundation Trust / Newcastle Hospitals NHS Foundation Trust | COVID-19 Genomics UK (COG-UK) Consortium |
| EPI_ISL_453513 | Northumbria University / South Tees Hospitals NHS Foundation Trust / North Cumbria Integrated Care NHS Foundation Trust / North Tees and Hartlepool NHS Foundation Trust / Newcastle Hospitals NHS Foundation Trust | COVID-19 Genomics UK (COG-UK) Consortium |
| EPI_ISL_453514 | Northumbria University / South Tees Hospitals NHS Foundation Trust / North Cumbria Integrated Care NHS Foundation Trust / North Tees and Hartlepool NHS Foundation Trust / Newcastle Hospitals NHS Foundation Trust | COVID-19 Genomics UK (COG-UK) Consortium |
| EPI_ISL_453515 | Northumbria University / South Tees Hospitals NHS Foundation Trust / North Cumbria Integrated Care NHS Foundation Trust / North Tees and Hartlepool NHS Foundation Trust / Newcastle Hospitals NHS Foundation Trust | COVID-19 Genomics UK (COG-UK) Consortium |
| EPI_ISL_453516 | Northumbria University / South Tees Hospitals NHS Foundation Trust / North Cumbria Integrated Care NHS Foundation Trust / North Tees and Hartlepool NHS Foundation Trust / Newcastle Hospitals NHS Foundation Trust | COVID-19 Genomics UK (COG-UK) Consortium |
| EPI_ISL_453517 | Northumbria University / South Tees Hospitals NHS Foundation Trust / North Cumbria Integrated Care NHS Foundation Trust / North Tees and Hartlepool NHS Foundation Trust / Newcastle Hospitals NHS Foundation Trust | COVID-19 Genomics UK (COG-UK) Consortium |

| EPI_Accession_ID | Originating Laboratory | Submitting Laboratory |
|---|---|---|
| EPI_ISL_453518 | Northumbria University / South Tees Hospitals NHS Foundation Trust / North Cumbria Integrated Care NHS Foundation Trust / North Tees and Hartlepool NHS Foundation Trust / Newcastle Hospitals NHS Foundation Trust | COVID-19 Genomics UK (COG-UK) Consortium |
| EPI_ISL_453519 | Northumbria University / South Tees Hospitals NHS Foundation Trust / North Cumbria Integrated Care NHS Foundation Trust / North Tees and Hartlepool NHS Foundation Trust / Newcastle Hospitals NHS Foundation Trust | COVID-19 Genomics UK (COG-UK) Consortium |
| EPI_ISL_453520 | Northumbria University / South Tees Hospitals NHS Foundation Trust / North Cumbria Integrated Care NHS Foundation Trust / North Tees and Hartlepool NHS Foundation Trust / Newcastle Hospitals NHS Foundation Trust | COVID-19 Genomics UK (COG-UK) Consortium |
| EPI_ISL_453521 | Northumbria University / South Tees Hospitals NHS Foundation Trust / North Cumbria Integrated Care NHS Foundation Trust / North Tees and Hartlepool NHS Foundation Trust / Newcastle Hospitals NHS Foundation Trust | COVID-19 Genomics UK (COG-UK) Consortium |
| EPI_ISL_453522 | Northumbria University / South Tees Hospitals NHS Foundation Trust / North Cumbria Integrated Care NHS Foundation Trust / North Tees and Hartlepool NHS Foundation Trust / Newcastle Hospitals NHS Foundation Trust | COVID-19 Genomics UK (COG-UK) Consortium |
| EPI_ISL_453523 | Northumbria University / South Tees Hospitals NHS Foundation Trust / North Cumbria Integrated Care NHS Foundation Trust / North Tees and Hartlepool NHS Foundation Trust / Newcastle Hospitals NHS Foundation Trust | COVID-19 Genomics UK (COG-UK) Consortium |

| EPI_Accession_ID | Originating Laboratory | Submitting Laboratory |
|---|---|---|
| EPI_ISL_453524 | Northumbria University / South Tees Hospitals NHS Foundation Trust / North Cumbria Integrated Care NHS Foundation Trust / North Tees and Hartlepool NHS Foundation Trust / Newcastle Hospitals NHS Foundation Trust | COVID-19 Genomics UK (COG-UK) Consortium |
| EPI_ISL_453525 | Northumbria University / South Tees Hospitals NHS Foundation Trust / North Cumbria Integrated Care NHS Foundation Trust / North Tees and Hartlepool NHS Foundation Trust / Newcastle Hospitals NHS Foundation Trust | COVID-19 Genomics UK (COG-UK) Consortium |
| EPI_ISL_453526 | Northumbria University / South Tees Hospitals NHS Foundation Trust / North Cumbria Integrated Care NHS Foundation Trust / North Tees and Hartlepool NHS Foundation Trust / Newcastle Hospitals NHS Foundation Trust | COVID-19 Genomics UK (COG-UK) Consortium |
| EPI_ISL_453527 | Northumbria University / South Tees Hospitals NHS Foundation Trust / North Cumbria Integrated Care NHS Foundation Trust / North Tees and Hartlepool NHS Foundation Trust / Newcastle Hospitals NHS Foundation Trust | COVID-19 Genomics UK (COG-UK) Consortium |
| EPI_ISL_453528 | Northumbria University / South Tees Hospitals NHS Foundation Trust / North Cumbria Integrated Care NHS Foundation Trust / North Tees and Hartlepool NHS Foundation Trust / Newcastle Hospitals NHS Foundation Trust | COVID-19 Genomics UK (COG-UK) Consortium |
| EPI_ISL_453529 | Northumbria University / South Tees Hospitals NHS Foundation Trust / North Cumbria Integrated Care NHS Foundation Trust / North Tees and Hartlepool NHS Foundation Trust / Newcastle Hospitals NHS Foundation Trust | COVID-19 Genomics UK (COG-UK) Consortium |

| EPI_Accession_ID | Originating Laboratory | Submitting Laboratory |
|---|---|---|
| EPI_ISL_453530 | Northumbria University / South Tees Hospitals NHS Foundation Trust / North Cumbria Integrated Care NHS Foundation Trust / North Tees and Hartlepool NHS Foundation Trust / Newcastle Hospitals NHS Foundation Trust | COVID-19 Genomics UK (COG-UK) Consortium |
| EPI_ISL_453531 | Northumbria University / South Tees Hospitals NHS Foundation Trust / North Cumbria Integrated Care NHS Foundation Trust / North Tees and Hartlepool NHS Foundation Trust / Newcastle Hospitals NHS Foundation Trust | COVID-19 Genomics UK (COG-UK) Consortium |
| EPI_ISL_453532 | Northumbria University / South Tees Hospitals NHS Foundation Trust / North Cumbria Integrated Care NHS Foundation Trust / North Tees and Hartlepool NHS Foundation Trust / Newcastle Hospitals NHS Foundation Trust | COVID-19 Genomics UK (COG-UK) Consortium |
| EPI_ISL_453533 | Northumbria University / South Tees Hospitals NHS Foundation Trust / North Cumbria Integrated Care NHS Foundation Trust / North Tees and Hartlepool NHS Foundation Trust / Newcastle Hospitals NHS Foundation Trust | COVID-19 Genomics UK (COG-UK) Consortium |
| EPI_ISL_453534 | Northumbria University / South Tees Hospitals NHS Foundation Trust / North Cumbria Integrated Care NHS Foundation Trust / North Tees and Hartlepool NHS Foundation Trust / Newcastle Hospitals NHS Foundation Trust | COVID-19 Genomics UK (COG-UK) Consortium |
| EPI_ISL_453535 | Northumbria University / South Tees Hospitals NHS Foundation Trust / North Cumbria Integrated Care NHS Foundation Trust / North Tees and Hartlepool NHS Foundation Trust / Newcastle Hospitals NHS Foundation Trust | COVID-19 Genomics UK (COG-UK) Consortium |

| EPI_Accession_ID | Originating Laboratory | Submitting Laboratory |
|---|---|---|
| EPI_ISL_453536 | Northumbria University / South Tees Hospitals NHS Foundation Trust / North Cumbria Integrated Care NHS Foundation Trust / North Tees and Hartlepool NHS Foundation Trust / Newcastle Hospitals NHS Foundation Trust | COVID-19 Genomics UK (COG-UK) Consortium |
| EPI_ISL_453537 | Northumbria University / South Tees Hospitals NHS Foundation Trust / North Cumbria Integrated Care NHS Foundation Trust / North Tees and Hartlepool NHS Foundation Trust / Newcastle Hospitals NHS Foundation Trust | COVID-19 Genomics UK (COG-UK) Consortium |
| EPI_ISL_453538 | Northumbria University / South Tees Hospitals NHS Foundation Trust / North Cumbria Integrated Care NHS Foundation Trust / North Tees and Hartlepool NHS Foundation Trust / Newcastle Hospitals NHS Foundation Trust | COVID-19 Genomics UK (COG-UK) Consortium |
| EPI_ISL_453539 | Northumbria University / South Tees Hospitals NHS Foundation Trust / North Cumbria Integrated Care NHS Foundation Trust / North Tees and Hartlepool NHS Foundation Trust / Newcastle Hospitals NHS Foundation Trust | COVID-19 Genomics UK (COG-UK) Consortium |
| EPI_ISL_453540 | Northumbria University / South Tees Hospitals NHS Foundation Trust / North Cumbria Integrated Care NHS Foundation Trust / North Tees and Hartlepool NHS Foundation Trust / Newcastle Hospitals NHS Foundation Trust | COVID-19 Genomics UK (COG-UK) Consortium |
| EPI_ISL_453541 | Northumbria University / South Tees Hospitals NHS Foundation Trust / North Cumbria Integrated Care NHS Foundation Trust / North Tees and Hartlepool NHS Foundation Trust / Newcastle Hospitals NHS Foundation Trust | COVID-19 Genomics UK (COG-UK) Consortium |

| EPI_Accession_ID | Originating Laboratory | Submitting Laboratory |
|---|---|---|
| EPI_ISL_453542 | Northumbria University / South Tees Hospitals NHS Foundation Trust / North Cumbria Integrated Care NHS Foundation Trust / North Tees and Hartlepool NHS Foundation Trust / Newcastle Hospitals NHS Foundation Trust | COVID-19 Genomics UK (COG-UK) Consortium |
| EPI_ISL_453543 | Northumbria University / South Tees Hospitals NHS Foundation Trust / North Cumbria Integrated Care NHS Foundation Trust / North Tees and Hartlepool NHS Foundation Trust / Newcastle Hospitals NHS Foundation Trust | Northumbria University |
| EPI_ISL_453544 | Northumbria University / South Tees Hospitals NHS Foundation Trust / North Cumbria Integrated Care NHS Foundation Trust / North Tees and Hartlepool NHS Foundation Trust / Newcastle Hospitals NHS Foundation Trust | COVID-19 Genomics UK (COG-UK) Consortium |
| EPI_ISL_453545 | Quadram Institute Bioscience | COVID-19 Genomics UK (COG-UK) Consortium |
| EPI_ISL_453546 | Quadram Institute Bioscience | COVID-19 Genomics UK (COG-UK) Consortium |
| EPI_ISL_453547 | Quadram Institute Bioscience | COVID-19 Genomics UK (COG-UK) Consortium |
| EPI_ISL_453548 | Quadram Institute Bioscience | COVID-19 Genomics UK (COG-UK) Consortium |
| EPI_ISL_453549 | Quadram Institute Bioscience | COVID-19 Genomics UK (COG-UK) Consortium |
| EPI_ISL_453550 | Quadram Institute Bioscience | COVID-19 Genomics UK (COG-UK) Consortium |
| EPI_ISL_453551 | Quadram Institute Bioscience | COVID-19 Genomics UK (COG-UK) Consortium |
| EPI_ISL_453552 | Quadram Institute Bioscience | COVID-19 Genomics UK (COG-UK) Consortium |
| EPI_ISL_453553 | Quadram Institute Bioscience | COVID-19 Genomics UK (COG-UK) Consortium |
| EPI_ISL_453554 | Quadram Institute Bioscience | COVID-19 Genomics UK (COG-UK) Consortium |
| EPI_ISL_453555 | Quadram Institute Bioscience | COVID-19 Genomics UK (COG-UK) Consortium |
| EPI_ISL_453556 | Quadram Institute Bioscience | COVID-19 Genomics UK (COG-UK) Consortium |
| EPI_ISL_453557 | Quadram Institute Bioscience | COVID-19 Genomics UK (COG-UK) Consortium |
| EPI_ISL_453558 | Quadram Institute Bioscience | COVID-19 Genomics UK (COG-UK) Consortium |
| EPI_ISL_453559 | Quadram Institute Bioscience | COVID-19 Genomics UK (COG-UK) Consortium |
| EPI_ISL_453560 | Quadram Institute Bioscience | COVID-19 Genomics UK (COG-UK) Consortium |
| EPI_ISL_453561 | Quadram Institute Bioscience | COVID-19 Genomics UK (COG-UK) Consortium |
| EPI_ISL_453562 | Quadram Institute Bioscience | COVID-19 Genomics UK (COG-UK) Consortium |

| EPI_Accession_ID | Originating Laboratory | Submitting Laboratory |
|---|---|---|
| EPI_ISL_453563 | Quadram Institute Bioscience | COVID-19 Genomics UK (COG-UK) Consortium |
| EPI_ISL_453564 | Quadram Institute Bioscience | COVID-19 Genomics UK (COG-UK) Consortium |
| EPI_ISL_453565 | Quadram Institute Bioscience | COVID-19 Genomics UK (COG-UK) Consortium |
| EPI_ISL_453566 | Quadram Institute Bioscience | COVID-19 Genomics UK (COG-UK) Consortium |
| EPI_ISL_453567 | Quadram Institute Bioscience | COVID-19 Genomics UK (COG-UK) Consortium |
| EPI_ISL_453568 | Quadram Institute Bioscience | COVID-19 Genomics UK (COG-UK) Consortium |
| EPI_ISL_453569 | Quadram Institute Bioscience | COVID-19 Genomics UK (COG-UK) Consortium |
| EPI_ISL_453570 | Quadram Institute Bioscience | COVID-19 Genomics UK (COG-UK) Consortium |
| EPI_ISL_453571 | Quadram Institute Bioscience | COVID-19 Genomics UK (COG-UK) Consortium |
| EPI_ISL_453572 | Quadram Institute Bioscience | COVID-19 Genomics UK (COG-UK) Consortium |
| EPI_ISL_453573 | Quadram Institute Bioscience | COVID-19 Genomics UK (COG-UK) Consortium |
| EPI_ISL_453574 | Quadram Institute Bioscience | COVID-19 Genomics UK (COG-UK) Consortium |
| EPI_ISL_453575 | Quadram Institute Bioscience | COVID-19 Genomics UK (COG-UK) Consortium |
| EPI_ISL_453576 | Quadram Institute Bioscience | COVID-19 Genomics UK (COG-UK) Consortium |
| EPI_ISL_453577 | Quadram Institute Bioscience | COVID-19 Genomics UK (COG-UK) Consortium |
| EPI_ISL_453578 | Quadram Institute Bioscience | COVID-19 Genomics UK (COG-UK) Consortium |
| EPI_ISL_453579 | Quadram Institute Bioscience | COVID-19 Genomics UK (COG-UK) Consortium |
| EPI_ISL_453580 | Quadram Institute Bioscience | COVID-19 Genomics UK (COG-UK) Consortium |
| EPI_ISL_453581 | Quadram Institute Bioscience | COVID-19 Genomics UK (COG-UK) Consortium |
| EPI_ISL_453582 | Quadram Institute Bioscience | COVID-19 Genomics UK (COG-UK) Consortium |
| EPI_ISL_453583 | Quadram Institute Bioscience | COVID-19 Genomics UK (COG-UK) Consortium |
| EPI_ISL_453584 | Quadram Institute Bioscience | COVID-19 Genomics UK (COG-UK) Consortium |
| EPI_ISL_453585 | Quadram Institute Bioscience | COVID-19 Genomics UK (COG-UK) Consortium |
| EPI_ISL_453586 | Quadram Institute Bioscience | COVID-19 Genomics UK (COG-UK) Consortium |
| EPI_ISL_453587 | Quadram Institute Bioscience | COVID-19 Genomics UK (COG-UK) Consortium |
| EPI_ISL_453588 | Quadram Institute Bioscience | COVID-19 Genomics UK (COG-UK) Consortium |
| EPI_ISL_453589 | Quadram Institute Bioscience | COVID-19 Genomics UK (COG-UK) Consortium |
| EPI_ISL_453590 | Quadram Institute Bioscience | COVID-19 Genomics UK (COG-UK) Consortium |
| EPI_ISL_453591 | Quadram Institute Bioscience | COVID-19 Genomics UK (COG-UK) Consortium |
| EPI_ISL_453592 | Quadram Institute Bioscience | COVID-19 Genomics UK (COG-UK) Consortium |
| EPI_ISL_453593 | Quadram Institute Bioscience | COVID-19 Genomics UK (COG-UK) Consortium |
| EPI_ISL_453594 | Quadram Institute Bioscience | COVID-19 Genomics UK (COG-UK) Consortium |
| EPI_ISL_453595 | Quadram Institute Bioscience | COVID-19 Genomics UK (COG-UK) Consortium |

| EPI_Accession_ID | Originating Laboratory | Submitting Laboratory |
|---|---|---|
| EPI_ISL_453596 | Quadram Institute Bioscience | COVID-19 Genomics UK (COG-UK) Consortium |
| EPI_ISL_453597 | Quadram Institute Bioscience | COVID-19 Genomics UK (COG-UK) Consortium |
| EPI_ISL_453598 | Quadram Institute Bioscience | COVID-19 Genomics UK (COG-UK) Consortium |
| EPI_ISL_453599 | Quadram Institute Bioscience | COVID-19 Genomics UK (COG-UK) Consortium |
| EPI_ISL_453600 | Quadram Institute Bioscience | COVID-19 Genomics UK (COG-UK) Consortium |
| EPI_ISL_453601 | Quadram Institute Bioscience | COVID-19 Genomics UK (COG-UK) Consortium |
| EPI_ISL_453602 | Quadram Institute Bioscience | COVID-19 Genomics UK (COG-UK) Consortium |
| EPI_ISL_453603 | Quadram Institute Bioscience | COVID-19 Genomics UK (COG-UK) Consortium |
| EPI_ISL_453604 | Quadram Institute Bioscience | COVID-19 Genomics UK (COG-UK) Consortium |
| EPI_ISL_453605 | Quadram Institute Bioscience | COVID-19 Genomics UK (COG-UK) Consortium |
| EPI_ISL_453606 | Quadram Institute Bioscience | COVID-19 Genomics UK (COG-UK) Consortium |
| EPI_ISL_453607 | Quadram Institute Bioscience | COVID-19 Genomics UK (COG-UK) Consortium |
| EPI_ISL_453608 | Quadram Institute Bioscience | COVID-19 Genomics UK (COG-UK) Consortium |
| EPI_ISL_453609 | Quadram Institute Bioscience | COVID-19 Genomics UK (COG-UK) Consortium |
| EPI_ISL_453610 | Quadram Institute Bioscience | COVID-19 Genomics UK (COG-UK) Consortium |
| EPI_ISL_453611 | Quadram Institute Bioscience | COVID-19 Genomics UK (COG-UK) Consortium |
| EPI_ISL_453612 | Quadram Institute Bioscience | COVID-19 Genomics UK (COG-UK) Consortium |
| EPI_ISL_453613 | Quadram Institute Bioscience | COVID-19 Genomics UK (COG-UK) Consortium |
| EPI_ISL_453614 | Queens Medical Centre, Clinical Microbiology Department / DeepSeq Nottingham | COVID-19 Genomics UK (COG-UK) Consortium |
| EPI_ISL_453615 | Queens Medical Centre, Clinical Microbiology Department / DeepSeq Nottingham | COVID-19 Genomics UK (COG-UK) Consortium |
| EPI_ISL_453616 | Queens Medical Centre, Clinical Microbiology Department / DeepSeq Nottingham | COVID-19 Genomics UK (COG-UK) Consortium |
| EPI_ISL_453617 | Queens Medical Centre, Clinical Microbiology Department / DeepSeq Nottingham | COVID-19 Genomics UK (COG-UK) Consortium |
| EPI_ISL_453618 | Queens Medical Centre, Clinical Microbiology Department / DeepSeq Nottingham | COVID-19 Genomics UK (COG-UK) Consortium |
| EPI_ISL_453619 | Queens Medical Centre, Clinical Microbiology Department / DeepSeq Nottingham | COVID-19 Genomics UK (COG-UK) Consortium |
| EPI_ISL_453620 | Queens Medical Centre, Clinical Microbiology Department / DeepSeq Nottingham | COVID-19 Genomics UK (COG-UK) Consortium |

| EPI_Accession_ID | Originating Laboratory | Submitting Laboratory |
|---|---|---|
| EPI_ISL_453621 | Queens Medical Centre, Clinical Microbiology Department / DeepSeq Nottingham | COVID-19 Genomics UK (COG-UK) Consortium |
| EPI_ISL_453622 | Queens Medical Centre, Clinical Microbiology Department / DeepSeq Nottingham | COVID-19 Genomics UK (COG-UK) Consortium |
| EPI_ISL_453623 | Queens Medical Centre, Clinical Microbiology Department / DeepSeq Nottingham | COVID-19 Genomics UK (COG-UK) Consortium |
| EPI_ISL_453624 | Queens Medical Centre, Clinical Microbiology Department / DeepSeq Nottingham | COVID-19 Genomics UK (COG-UK) Consortium |
| EPI_ISL_453625 | Queens Medical Centre, Clinical Microbiology Department / DeepSeq Nottingham | COVID-19 Genomics UK (COG-UK) Consortium |
| EPI_ISL_453626 | Queens Medical Centre, Clinical Microbiology Department / DeepSeq Nottingham | COVID-19 Genomics UK (COG-UK) Consortium |
| EPI_ISL_453627 | Queens Medical Centre, Clinical Microbiology Department / DeepSeq Nottingham | COVID-19 Genomics UK (COG-UK) Consortium |
| EPI_ISL_453628 | Queens Medical Centre, Clinical Microbiology Department / DeepSeq Nottingham | COVID-19 Genomics UK (COG-UK) Consortium |
| EPI_ISL_453629 | Queens Medical Centre, Clinical Microbiology Department / DeepSeq Nottingham | COVID-19 Genomics UK (COG-UK) Consortium |
| EPI_ISL_453630 | Queens Medical Centre, Clinical Microbiology Department / DeepSeq Nottingham | COVID-19 Genomics UK (COG-UK) Consortium |
| EPI_ISL_453631 | Queens Medical Centre, Clinical Microbiology Department / DeepSeq Nottingham | COVID-19 Genomics UK (COG-UK) Consortium |
| EPI_ISL_453632 | Queens Medical Centre, Clinical Microbiology Department / DeepSeq Nottingham | COVID-19 Genomics UK (COG-UK) Consortium |
| EPI_ISL_453633 | Queens Medical Centre, Clinical Microbiology Department / DeepSeq Nottingham | COVID-19 Genomics UK (COG-UK) Consortium |
| EPI_ISL_453634 | Queens Medical Centre, Clinical Microbiology Department / DeepSeq Nottingham | COVID-19 Genomics UK (COG-UK) Consortium |
| EPI_ISL_453635 | Queens Medical Centre, Clinical Microbiology Department / DeepSeq Nottingham | COVID-19 Genomics UK (COG-UK) Consortium |
| EPI_ISL_453636 | Queens Medical Centre, Clinical Microbiology Department / DeepSeq Nottingham | COVID-19 Genomics UK (COG-UK) Consortium |

| EPI_Accession_ID | Originating Laboratory | Submitting Laboratory |
|---|---|---|
| EPI_ISL_453637 | Queens Medical Centre, Clinical Microbiology Department / DeepSeq Nottingham | COVID-19 Genomics UK (COG-UK) Consortium |
| EPI_ISL_453638 | Queens Medical Centre, Clinical Microbiology Department / DeepSeq Nottingham | COVID-19 Genomics UK (COG-UK) Consortium |
| EPI_ISL_453639 | Queens Medical Centre, Clinical Microbiology Department / DeepSeq Nottingham | COVID-19 Genomics UK (COG-UK) Consortium |
| EPI_ISL_453640 | Queens Medical Centre, Clinical Microbiology Department / DeepSeq Nottingham | COVID-19 Genomics UK (COG-UK) Consortium |
| EPI_ISL_453641 | Queens Medical Centre, Clinical Microbiology Department / DeepSeq Nottingham | COVID-19 Genomics UK (COG-UK) Consortium |
| EPI_ISL_453642 | Queens Medical Centre, Clinical Microbiology Department / DeepSeq Nottingham | COVID-19 Genomics UK (COG-UK) Consortium |
| EPI_ISL_453643 | Queens Medical Centre, Clinical Microbiology Department / DeepSeq Nottingham | COVID-19 Genomics UK (COG-UK) Consortium |
| EPI_ISL_453644 | Queens Medical Centre, Clinical Microbiology Department / DeepSeq Nottingham | COVID-19 Genomics UK (COG-UK) Consortium |
| EPI_ISL_453645 | Queens Medical Centre, Clinical Microbiology Department / DeepSeq Nottingham | COVID-19 Genomics UK (COG-UK) Consortium |
| EPI_ISL_453646 | Queens Medical Centre, Clinical Microbiology Department / DeepSeq Nottingham | COVID-19 Genomics UK (COG-UK) Consortium |
| EPI_ISL_453647 | Queens Medical Centre, Clinical Microbiology Department / DeepSeq Nottingham | COVID-19 Genomics UK (COG-UK) Consortium |
| EPI_ISL_453648 | Queens Medical Centre, Clinical Microbiology Department / DeepSeq Nottingham | COVID-19 Genomics UK (COG-UK) Consortium |
| EPI_ISL_453649 | Queens Medical Centre, Clinical Microbiology Department / DeepSeq Nottingham | COVID-19 Genomics UK (COG-UK) Consortium |
| EPI_ISL_453650 | Queens Medical Centre, Clinical Microbiology Department / DeepSeq Nottingham | COVID-19 Genomics UK (COG-UK) Consortium |
| EPI_ISL_453651 | Queens Medical Centre, Clinical Microbiology Department / DeepSeq Nottingham | COVID-19 Genomics UK (COG-UK) Consortium |
| EPI_ISL_453652 | Queens Medical Centre, Clinical Microbiology Department / DeepSeq Nottingham | COVID-19 Genomics UK (COG-UK) Consortium |

| EPI_Accession_ID | Originating Laboratory | Submitting Laboratory |
|---|---|---|
| EPI_ISL_453653 | Queens Medical Centre, Clinical Microbiology Department / DeepSeq Nottingham | COVID-19 Genomics UK (COG-UK) Consortium |
| EPI_ISL_453654 | Queens Medical Centre, Clinical Microbiology Department / DeepSeq Nottingham | COVID-19 Genomics UK (COG-UK) Consortium |
| EPI_ISL_453655 | Queens Medical Centre, Clinical Microbiology Department / DeepSeq Nottingham | COVID-19 Genomics UK (COG-UK) Consortium |
| EPI_ISL_453656 | Queens Medical Centre, Clinical Microbiology Department / DeepSeq Nottingham | COVID-19 Genomics UK (COG-UK) Consortium |
| EPI_ISL_453657 | Queens Medical Centre, Clinical Microbiology Department / DeepSeq Nottingham | COVID-19 Genomics UK (COG-UK) Consortium |
| EPI_ISL_453658 | Queens Medical Centre, Clinical Microbiology Department / DeepSeq Nottingham | COVID-19 Genomics UK (COG-UK) Consortium |
| EPI_ISL_453659 | Queens Medical Centre, Clinical Microbiology Department / DeepSeq Nottingham | COVID-19 Genomics UK (COG-UK) Consortium |
| EPI_ISL_453660 | Queens Medical Centre, Clinical Microbiology Department / DeepSeq Nottingham | COVID-19 Genomics UK (COG-UK) Consortium |
| EPI_ISL_453661 | Queens Medical Centre, Clinical Microbiology Department / DeepSeq Nottingham | COVID-19 Genomics UK (COG-UK) Consortium |
| EPI_ISL_453662 | Queens Medical Centre, Clinical Microbiology Department / DeepSeq Nottingham | COVID-19 Genomics UK (COG-UK) Consortium |
| EPI_ISL_453663 | Queens Medical Centre, Clinical Microbiology Department / DeepSeq Nottingham | COVID-19 Genomics UK (COG-UK) Consortium |
| EPI_ISL_453664 | Queens Medical Centre, Clinical Microbiology Department / DeepSeq Nottingham | COVID-19 Genomics UK (COG-UK) Consortium |
| EPI_ISL_453665 | Centre for Enzyme Innovation, University of Portsmouth / Translational Research Laboratory, Portsmouth Hospitals NHS Trust | University of Portsmouth |
| EPI_ISL_453666 | Centre for Enzyme Innovation, University of Portsmouth / Translational Research Laboratory, Portsmouth Hospitals NHS Trust | COVID-19 Genomics UK (COG-UK) Consortium |
| EPI_ISL_453667 | Centre for Enzyme Innovation, University of Portsmouth / Translational Research Laboratory, Portsmouth Hospitals NHS Trust | COVID-19 Genomics UK (COG-UK) Consortium |

| EPI_Accession_ID | Originating Laboratory | Submitting Laboratory |
|---|---|---|
| EPI_ISL_453668 | Centre for Enzyme Innovation, University of Portsmouth / Translational Research Laboratory, Portsmouth Hospitals NHS Trust | COVID-19 Genomics UK (COG-UK) Consortium |
| EPI_ISL_453669 | Centre for Enzyme Innovation, University of Portsmouth / Translational Research Laboratory, Portsmouth Hospitals NHS Trust | COVID-19 Genomics UK (COG-UK) Consortium |
| EPI_ISL_453670 | Centre for Enzyme Innovation, University of Portsmouth / Translational Research Laboratory, Portsmouth Hospitals NHS Trust | COVID-19 Genomics UK (COG-UK) Consortium |
| EPI_ISL_453671 | Centre for Enzyme Innovation, University of Portsmouth / Translational Research Laboratory, Portsmouth Hospitals NHS Trust | COVID-19 Genomics UK (COG-UK) Consortium |
| EPI_ISL_453672 | Centre for Enzyme Innovation, University of Portsmouth / Translational Research Laboratory, Portsmouth Hospitals NHS Trust | COVID-19 Genomics UK (COG-UK) Consortium |
| EPI_ISL_453673 | Centre for Enzyme Innovation, University of Portsmouth / Translational Research Laboratory, Portsmouth Hospitals NHS Trust | COVID-19 Genomics UK (COG-UK) Consortium |
| EPI_ISL_453674 | Centre for Enzyme Innovation, University of Portsmouth / Translational Research Laboratory, Portsmouth Hospitals NHS Trust | COVID-19 Genomics UK (COG-UK) Consortium |
| EPI_ISL_453675 | Centre for Enzyme Innovation, University of Portsmouth / Translational Research Laboratory, Portsmouth Hospitals NHS Trust | COVID-19 Genomics UK (COG-UK) Consortium |
| EPI_ISL_453676 | Centre for Enzyme Innovation, University of Portsmouth / Translational Research Laboratory, Portsmouth Hospitals NHS Trust | COVID-19 Genomics UK (COG-UK) Consortium |
| EPI_ISL_453677 | Centre for Enzyme Innovation, University of Portsmouth / Translational Research Laboratory, Portsmouth Hospitals NHS Trust | COVID-19 Genomics UK (COG-UK) Consortium |
| EPI_ISL_453678 | Centre for Enzyme Innovation, University of Portsmouth / Translational Research Laboratory, Portsmouth Hospitals NHS Trust | COVID-19 Genomics UK (COG-UK) Consortium |

| EPI_Accession_ID | Originating Laboratory | Submitting Laboratory |
|---|---|---|
| EPI_ISL_453679 | Centre for Enzyme Innovation, University of Portsmouth / Translational Research Laboratory, Portsmouth Hospitals NHS Trust | COVID-19 Genomics UK (COG-UK) Consortium |
| EPI_ISL_453680 | Centre for Enzyme Innovation, University of Portsmouth / Translational Research Laboratory, Portsmouth Hospitals NHS Trust | COVID-19 Genomics UK (COG-UK) Consortium |
| EPI_ISL_453681 | Centre for Enzyme Innovation, University of Portsmouth / Translational Research Laboratory, Portsmouth Hospitals NHS Trust | COVID-19 Genomics UK (COG-UK) Consortium |
| EPI_ISL_453682 | Centre for Enzyme Innovation, University of Portsmouth / Translational Research Laboratory, Portsmouth Hospitals NHS Trust | COVID-19 Genomics UK (COG-UK) Consortium |
| EPI_ISL_453683 | Centre for Enzyme Innovation, University of Portsmouth / Translational Research Laboratory, Portsmouth Hospitals NHS Trust | COVID-19 Genomics UK (COG-UK) Consortium |
| EPI_ISL_453684 | Centre for Enzyme Innovation, University of Portsmouth / Translational Research Laboratory, Portsmouth Hospitals NHS Trust | COVID-19 Genomics UK (COG-UK) Consortium |
| EPI_ISL_453685 | Centre for Enzyme Innovation, University of Portsmouth / Translational Research Laboratory, Portsmouth Hospitals NHS Trust | COVID-19 Genomics UK (COG-UK) Consortium |
| EPI_ISL_453686 | Centre for Enzyme Innovation, University of Portsmouth / Translational Research Laboratory, Portsmouth Hospitals NHS Trust | COVID-19 Genomics UK (COG-UK) Consortium |
| EPI_ISL_453687 | Centre for Enzyme Innovation, University of Portsmouth / Translational Research Laboratory, Portsmouth Hospitals NHS Trust | COVID-19 Genomics UK (COG-UK) Consortium |
| EPI_ISL_453688 | Centre for Enzyme Innovation, University of Portsmouth / Translational Research Laboratory, Portsmouth Hospitals NHS Trust | COVID-19 Genomics UK (COG-UK) Consortium |
| EPI_ISL_453689 | Centre for Enzyme Innovation, University of Portsmouth / Translational Research Laboratory, Portsmouth Hospitals NHS Trust | COVID-19 Genomics UK (COG-UK) Consortium |

| EPI_Accession_ID | Originating Laboratory | Submitting Laboratory |
|---|---|---|
| EPI_ISL_453690 | Centre for Enzyme Innovation, University of Portsmouth / Translational Research Laboratory, Portsmouth Hospitals NHS Trust | COVID-19 Genomics UK (COG-UK) Consortium |
| EPI_ISL_453691 | Centre for Enzyme Innovation, University of Portsmouth / Translational Research Laboratory, Portsmouth Hospitals NHS Trust | COVID-19 Genomics UK (COG-UK) Consortium |
| EPI_ISL_453692 | Centre for Enzyme Innovation, University of Portsmouth / Translational Research Laboratory, Portsmouth Hospitals NHS Trust | COVID-19 Genomics UK (COG-UK) Consortium |
| EPI_ISL_453693 | Centre for Enzyme Innovation, University of Portsmouth / Translational Research Laboratory, Portsmouth Hospitals NHS Trust | COVID-19 Genomics UK (COG-UK) Consortium |
| EPI_ISL_453694 | Centre for Enzyme Innovation, University of Portsmouth / Translational Research Laboratory, Portsmouth Hospitals NHS Trust | COVID-19 Genomics UK (COG-UK) Consortium |
| EPI_ISL_453695 | Centre for Enzyme Innovation, University of Portsmouth / Translational Research Laboratory, Portsmouth Hospitals NHS Trust | COVID-19 Genomics UK (COG-UK) Consortium |
| EPI_ISL_453696 | Virology Department, Sheffield Teaching Hospitals NHS Foundation Trust/Department of Infection, Immunity and Cardiovascular Disease, The Medical School, University of Sheffield | COVID-19 Genomics UK (COG-UK) Consortium |
| EPI_ISL_453697 | Virology Department, Sheffield Teaching Hospitals NHS Foundation Trust/Department of Infection, Immunity and Cardiovascular Disease, The Medical School, University of Sheffield | COVID-19 Genomics UK (COG-UK) Consortium |
| EPI_ISL_453698 | Virology Department, Sheffield Teaching Hospitals NHS Foundation Trust/Department of Infection, Immunity and Cardiovascular Disease, The Medical School, University of Sheffield | COVID-19 Genomics UK (COG-UK) Consortium |

| EPI_Accession_ID | Originating Laboratory | Submitting Laboratory |
|---|---|---|
| EPI_ISL_453699 | Virology Department, Sheffield Teaching Hospitals NHS Foundation Trust/Department of Infection, Immunity and Cardiovascular Disease, The Medical School, University of Sheffield | COVID-19 Genomics UK (COG-UK) Consortium |
| EPI_ISL_453700 | Virology Department, Sheffield Teaching Hospitals NHS Foundation Trust/Department of Infection, Immunity and Cardiovascular Disease, The Medical School, University of Sheffield | COVID-19 Genomics UK (COG-UK) Consortium |
| EPI_ISL_453701 | Virology Department, Sheffield Teaching Hospitals NHS Foundation Trust/Department of Infection, Immunity and Cardiovascular Disease, The Medical School, University of Sheffield | COVID-19 Genomics UK (COG-UK) Consortium |
| EPI_ISL_453702 | Virology Department, Sheffield Teaching Hospitals NHS Foundation Trust/Department of Infection, Immunity and Cardiovascular Disease, The Medical School, University of Sheffield | COVID-19 Genomics UK (COG-UK) Consortium |
| EPI_ISL_453703 | Virology Department, Sheffield Teaching Hospitals NHS Foundation Trust/Department of Infection, Immunity and Cardiovascular Disease, The Medical School, University of Sheffield | COVID-19 Genomics UK (COG-UK) Consortium |
| EPI_ISL_453704 | Virology Department, Sheffield Teaching Hospitals NHS Foundation Trust/Department of Infection, Immunity and Cardiovascular Disease, The Medical School, University of Sheffield | COVID-19 Genomics UK (COG-UK) Consortium |
| EPI_ISL_453705 | Virology Department, Sheffield Teaching Hospitals NHS Foundation Trust/Department of Infection, Immunity and Cardiovascular Disease, The Medical School, University of Sheffield | COVID-19 Genomics UK (COG-UK) Consortium |
| EPI_ISL_453706 | Virology Department, Sheffield Teaching Hospitals NHS Foundation Trust/Department of Infection, Immunity and Cardiovascular Disease, The Medical School, University of Sheffield | COVID-19 Genomics UK (COG-UK) Consortium |

| EPI_Accession_ID | Originating Laboratory | Submitting Laboratory |
|---|---|---|
| EPI_ISL_453707 | Virology Department, Sheffield Teaching Hospitals NHS Foundation Trust/Department of Infection, Immunity and Cardiovascular Disease, The Medical School, University of Sheffield | COVID-19 Genomics UK (COG-UK) Consortium |
| EPI_ISL_453708 | Virology Department, Sheffield Teaching Hospitals NHS Foundation Trust/Department of Infection, Immunity and Cardiovascular Disease, The Medical School, University of Sheffield | COVID-19 Genomics UK (COG-UK) Consortium |
| EPI_ISL_453709 | Virology Department, Sheffield Teaching Hospitals NHS Foundation Trust/Department of Infection, Immunity and Cardiovascular Disease, The Medical School, University of Sheffield | COVID-19 Genomics UK (COG-UK) Consortium |
| EPI_ISL_453710 | Virology Department, Sheffield Teaching Hospitals NHS Foundation Trust/Department of Infection, Immunity and Cardiovascular Disease, The Medical School, University of Sheffield | COVID-19 Genomics UK (COG-UK) Consortium |
| EPI_ISL_453711 | Virology Department, Sheffield Teaching Hospitals NHS Foundation Trust/Department of Infection, Immunity and Cardiovascular Disease, The Medical School, University of Sheffield | COVID-19 Genomics UK (COG-UK) Consortium |
| EPI_ISL_453712 | Virology Department, Sheffield Teaching Hospitals NHS Foundation Trust/Department of Infection, Immunity and Cardiovascular Disease, The Medical School, University of Sheffield | COVID-19 Genomics UK (COG-UK) Consortium |
| EPI_ISL_453713 | Virology Department, Sheffield Teaching Hospitals NHS Foundation Trust/Department of Infection, Immunity and Cardiovascular Disease, The Medical School, University of Sheffield | COVID-19 Genomics UK (COG-UK) Consortium |
| EPI_ISL_453714 | Virology Department, Sheffield Teaching Hospitals NHS Foundation Trust/Department of Infection, Immunity and Cardiovascular Disease, The Medical School, University of Sheffield | COVID-19 Genomics UK (COG-UK) Consortium |

| EPI_Accession_ID | Originating Laboratory | Submitting Laboratory |
|---|---|---|
| EPI_ISL_453715 | Virology Department, Sheffield Teaching Hospitals NHS Foundation Trust/Department of Infection, Immunity and Cardiovascular Disease, The Medical School, University of Sheffield | COVID-19 Genomics UK (COG-UK) Consortium |
| EPI_ISL_453716 | Virology Department, Sheffield Teaching Hospitals NHS Foundation Trust/Department of Infection, Immunity and Cardiovascular Disease, The Medical School, University of Sheffield | COVID-19 Genomics UK (COG-UK) Consortium |
| EPI_ISL_453717 | Virology Department, Sheffield Teaching Hospitals NHS Foundation Trust/Department of Infection, Immunity and Cardiovascular Disease, The Medical School, University of Sheffield | COVID-19 Genomics UK (COG-UK) Consortium |
| EPI_ISL_453718 | Virology Department, Sheffield Teaching Hospitals NHS Foundation Trust/Department of Infection, Immunity and Cardiovascular Disease, The Medical School, University of Sheffield | COVID-19 Genomics UK (COG-UK) Consortium |
| EPI_ISL_453719 | Virology Department, Sheffield Teaching Hospitals NHS Foundation Trust/Department of Infection, Immunity and Cardiovascular Disease, The Medical School, University of Sheffield | COVID-19 Genomics UK (COG-UK) Consortium |
| EPI_ISL_453720 | Virology Department, Sheffield Teaching Hospitals NHS Foundation Trust/Department of Infection, Immunity and Cardiovascular Disease, The Medical School, University of Sheffield | COVID-19 Genomics UK (COG-UK) Consortium |
| EPI_ISL_453721 | Virology Department, Sheffield Teaching Hospitals NHS Foundation Trust/Department of Infection, Immunity and Cardiovascular Disease, The Medical School, University of Sheffield | COVID-19 Genomics UK (COG-UK) Consortium |
| EPI_ISL_453722 | Virology Department, Sheffield Teaching Hospitals NHS Foundation Trust/Department of Infection, Immunity and Cardiovascular Disease, The Medical School, University of Sheffield | COVID-19 Genomics UK (COG-UK) Consortium |

| EPI_Accession_ID | Originating Laboratory | Submitting Laboratory |
|---|---|---|
| EPI_ISL_453723 | Virology Department, Sheffield Teaching Hospitals NHS Foundation Trust/Department of Infection, Immunity and Cardiovascular Disease, The Medical School, University of Sheffield | COVID-19 Genomics UK (COG-UK) Consortium |
| EPI_ISL_453724 | Virology Department, Sheffield Teaching Hospitals NHS Foundation Trust/Department of Infection, Immunity and Cardiovascular Disease, The Medical School, University of Sheffield | COVID-19 Genomics UK (COG-UK) Consortium |
| EPI_ISL_453725 | Virology Department, Sheffield Teaching Hospitals NHS Foundation Trust/Department of Infection, Immunity and Cardiovascular Disease, The Medical School, University of Sheffield | COVID-19 Genomics UK (COG-UK) Consortium |
| EPI_ISL_453726 | Virology Department, Sheffield Teaching Hospitals NHS Foundation Trust/Department of Infection, Immunity and Cardiovascular Disease, The Medical School, University of Sheffield | COVID-19 Genomics UK (COG-UK) Consortium |
| EPI_ISL_453727 | Virology Department, Sheffield Teaching Hospitals NHS Foundation Trust/Department of Infection, Immunity and Cardiovascular Disease, The Medical School, University of Sheffield | COVID-19 Genomics UK (COG-UK) Consortium |
| EPI_ISL_453728 | Virology Department, Sheffield Teaching Hospitals NHS Foundation Trust/Department of Infection, Immunity and Cardiovascular Disease, The Medical School, University of Sheffield | COVID-19 Genomics UK (COG-UK) Consortium |
| EPI_ISL_453729 | Virology Department, Sheffield Teaching Hospitals NHS Foundation Trust/Department of Infection, Immunity and Cardiovascular Disease, The Medical School, University of Sheffield | COVID-19 Genomics UK (COG-UK) Consortium |
| EPI_ISL_453730 | Virology Department, Sheffield Teaching Hospitals NHS Foundation Trust/Department of Infection, Immunity and Cardiovascular Disease, The Medical School, University of Sheffield | COVID-19 Genomics UK (COG-UK) Consortium |

| EPI_Accession_ID | Originating Laboratory | Submitting Laboratory |
|---|---|---|
| EPI_ISL_453731 | Virology Department, Sheffield Teaching Hospitals NHS Foundation Trust/Department of Infection, Immunity and Cardiovascular Disease, The Medical School, University of Sheffield | COVID-19 Genomics UK (COG-UK) Consortium |
| EPI_ISL_453732 | Virology Department, Sheffield Teaching Hospitals NHS Foundation Trust/Department of Infection, Immunity and Cardiovascular Disease, The Medical School, University of Sheffield | COVID-19 Genomics UK (COG-UK) Consortium |
| EPI_ISL_453733 | Virology Department, Sheffield Teaching Hospitals NHS Foundation Trust/Department of Infection, Immunity and Cardiovascular Disease, The Medical School, University of Sheffield | COVID-19 Genomics UK (COG-UK) Consortium |
| EPI_ISL_453734 | Virology Department, Sheffield Teaching Hospitals NHS Foundation Trust/Department of Infection, Immunity and Cardiovascular Disease, The Medical School, University of Sheffield | COVID-19 Genomics UK (COG-UK) Consortium |
| EPI_ISL_453735 | Virology Department, Sheffield Teaching Hospitals NHS Foundation Trust/Department of Infection, Immunity and Cardiovascular Disease, The Medical School, University of Sheffield | COVID-19 Genomics UK (COG-UK) Consortium |
| EPI_ISL_453736 | Virology Department, Sheffield Teaching Hospitals NHS Foundation Trust/Department of Infection, Immunity and Cardiovascular Disease, The Medical School, University of Sheffield | COVID-19 Genomics UK (COG-UK) Consortium |
| EPI_ISL_453737 | Virology Department, Sheffield Teaching Hospitals NHS Foundation Trust/Department of Infection, Immunity and Cardiovascular Disease, The Medical School, University of Sheffield | COVID-19 Genomics UK (COG-UK) Consortium |
| EPI_ISL_453738 | Virology Department, Sheffield Teaching Hospitals NHS Foundation Trust/Department of Infection, Immunity and Cardiovascular Disease, The Medical School, University of Sheffield | COVID-19 Genomics UK (COG-UK) Consortium |

| EPI_Accession_ID | Originating Laboratory | Submitting Laboratory |
|---|---|---|
| EPI_ISL_453739 | Virology Department, Sheffield Teaching Hospitals NHS Foundation Trust/Department of Infection, Immunity and Cardiovascular Disease, The Medical School, University of Sheffield | COVID-19 Genomics UK (COG-UK) Consortium |
| EPI_ISL_453740 | Virology Department, Sheffield Teaching Hospitals NHS Foundation Trust/Department of Infection, Immunity and Cardiovascular Disease, The Medical School, University of Sheffield | COVID-19 Genomics UK (COG-UK) Consortium |
| EPI_ISL_453741 | Virology Department, Sheffield Teaching Hospitals NHS Foundation Trust/Department of Infection, Immunity and Cardiovascular Disease, The Medical School, University of Sheffield | COVID-19 Genomics UK (COG-UK) Consortium |
| EPI_ISL_453742 | Virology Department, Sheffield Teaching Hospitals NHS Foundation Trust/Department of Infection, Immunity and Cardiovascular Disease, The Medical School, University of Sheffield | COVID-19 Genomics UK (COG-UK) Consortium |
| EPI_ISL_453743 | Virology Department, Sheffield Teaching Hospitals NHS Foundation Trust/Department of Infection, Immunity and Cardiovascular Disease, The Medical School, University of Sheffield | COVID-19 Genomics UK (COG-UK) Consortium |
| EPI_ISL_453744 | Virology Department, Sheffield Teaching Hospitals NHS Foundation Trust/Department of Infection, Immunity and Cardiovascular Disease, The Medical School, University of Sheffield | COVID-19 Genomics UK (COG-UK) Consortium |
| EPI_ISL_453745 | Virology Department, Sheffield Teaching Hospitals NHS Foundation Trust/Department of Infection, Immunity and Cardiovascular Disease, The Medical School, University of Sheffield | COVID-19 Genomics UK (COG-UK) Consortium |
| EPI_ISL_453746 | Virology Department, Sheffield Teaching Hospitals NHS Foundation Trust/Department of Infection, Immunity and Cardiovascular Disease, The Medical School, University of Sheffield | COVID-19 Genomics UK (COG-UK) Consortium |

| EPI_Accession_ID | Originating Laboratory | Submitting Laboratory |
|---|---|---|
| EPI_ISL_453747 | Virology Department, Sheffield Teaching Hospitals NHS Foundation Trust/Department of Infection, Immunity and Cardiovascular Disease, The Medical School, University of Sheffield | COVID-19 Genomics UK (COG-UK) Consortium |
| EPI_ISL_453748 | Virology Department, Sheffield Teaching Hospitals NHS Foundation Trust/Department of Infection, Immunity and Cardiovascular Disease, The Medical School, University of Sheffield | COVID-19 Genomics UK (COG-UK) Consortium |
| EPI_ISL_453749 | Virology Department, Sheffield Teaching Hospitals NHS Foundation Trust/Department of Infection, Immunity and Cardiovascular Disease, The Medical School, University of Sheffield | COVID-19 Genomics UK (COG-UK) Consortium |
| EPI_ISL_453750 | Virology Department, Sheffield Teaching Hospitals NHS Foundation Trust/Department of Infection, Immunity and Cardiovascular Disease, The Medical School, University of Sheffield | COVID-19 Genomics UK (COG-UK) Consortium |
| EPI_ISL_453751 | Virology Department, Sheffield Teaching Hospitals NHS Foundation Trust/Department of Infection, Immunity and Cardiovascular Disease, The Medical School, University of Sheffield | COVID-19 Genomics UK (COG-UK) Consortium |
| EPI_ISL_453752 | Virology Department, Sheffield Teaching Hospitals NHS Foundation Trust/Department of Infection, Immunity and Cardiovascular Disease, The Medical School, University of Sheffield | COVID-19 Genomics UK (COG-UK) Consortium |
| EPI_ISL_453753 | Virology Department, Sheffield Teaching Hospitals NHS Foundation Trust/Department of Infection, Immunity and Cardiovascular Disease, The Medical School, University of Sheffield | COVID-19 Genomics UK (COG-UK) Consortium |
| EPI_ISL_453754 | Virology Department, Sheffield Teaching Hospitals NHS Foundation Trust/Department of Infection, Immunity and Cardiovascular Disease, The Medical School, University of Sheffield | COVID-19 Genomics UK (COG-UK) Consortium |

| EPI_Accession_ID | Originating Laboratory | Submitting Laboratory |
|---|---|---|
| EPI_ISL_453755 | Virology Department, Sheffield Teaching Hospitals NHS Foundation Trust/Department of Infection, Immunity and Cardiovascular Disease, The Medical School, University of Sheffield | COVID-19 Genomics UK (COG-UK) Consortium |
| EPI_ISL_453756 | Virology Department, Sheffield Teaching Hospitals NHS Foundation Trust/Department of Infection, Immunity and Cardiovascular Disease, The Medical School, University of Sheffield | COVID-19 Genomics UK (COG-UK) Consortium |
| EPI_ISL_453757 | Virology Department, Sheffield Teaching Hospitals NHS Foundation Trust/Department of Infection, Immunity and Cardiovascular Disease, The Medical School, University of Sheffield | COVID-19 Genomics UK (COG-UK) Consortium |
| EPI_ISL_453758 | Virology Department, Sheffield Teaching Hospitals NHS Foundation Trust/Department of Infection, Immunity and Cardiovascular Disease, The Medical School, University of Sheffield | COVID-19 Genomics UK (COG-UK) Consortium |
| EPI_ISL_453759 | Virology Department, Sheffield Teaching Hospitals NHS Foundation Trust/Department of Infection, Immunity and Cardiovascular Disease, The Medical School, University of Sheffield | COVID-19 Genomics UK (COG-UK) Consortium |
| EPI_ISL_453760 | Virology Department, Sheffield Teaching Hospitals NHS Foundation Trust/Department of Infection, Immunity and Cardiovascular Disease, The Medical School, University of Sheffield | COVID-19 Genomics UK (COG-UK) Consortium |
| EPI_ISL_453761 | Virology Department, Sheffield Teaching Hospitals NHS Foundation Trust/Department of Infection, Immunity and Cardiovascular Disease, The Medical School, University of Sheffield | COVID-19 Genomics UK (COG-UK) Consortium |
| EPI_ISL_453762 | Virology Department, Sheffield Teaching Hospitals NHS Foundation Trust/Department of Infection, Immunity and Cardiovascular Disease, The Medical School, University of Sheffield | COVID-19 Genomics UK (COG-UK) Consortium |

| EPI_Accession_ID | Originating Laboratory | Submitting Laboratory |
|---|---|---|
| EPI_ISL_453763 | Virology Department, Sheffield Teaching Hospitals NHS Foundation Trust/Department of Infection, Immunity and Cardiovascular Disease, The Medical School, University of Sheffield | COVID-19 Genomics UK (COG-UK) Consortium |
| EPI_ISL_453764 | Virology Department, Sheffield Teaching Hospitals NHS Foundation Trust/Department of Infection, Immunity and Cardiovascular Disease, The Medical School, University of Sheffield | COVID-19 Genomics UK (COG-UK) Consortium |
| EPI_ISL_453765 | Virology Department, Sheffield Teaching Hospitals NHS Foundation Trust/Department of Infection, Immunity and Cardiovascular Disease, The Medical School, University of Sheffield | COVID-19 Genomics UK (COG-UK) Consortium |
| EPI_ISL_453766 | Virology Department, Sheffield Teaching Hospitals NHS Foundation Trust/Department of Infection, Immunity and Cardiovascular Disease, The Medical School, University of Sheffield | COVID-19 Genomics UK (COG-UK) Consortium |
| EPI_ISL_453767 | Virology Department, Sheffield Teaching Hospitals NHS Foundation Trust/Department of Infection, Immunity and Cardiovascular Disease, The Medical School, University of Sheffield | COVID-19 Genomics UK (COG-UK) Consortium |
| EPI_ISL_453768 | Virology Department, Sheffield Teaching Hospitals NHS Foundation Trust/Department of Infection, Immunity and Cardiovascular Disease, The Medical School, University of Sheffield | COVID-19 Genomics UK (COG-UK) Consortium |
| EPI_ISL_453769 | Virology Department, Sheffield Teaching Hospitals NHS Foundation Trust/Department of Infection, Immunity and Cardiovascular Disease, The Medical School, University of Sheffield | COVID-19 Genomics UK (COG-UK) Consortium |
| EPI_ISL_453770 | Virology Department, Sheffield Teaching Hospitals NHS Foundation Trust/Department of Infection, Immunity and Cardiovascular Disease, The Medical School, University of Sheffield | COVID-19 Genomics UK (COG-UK) Consortium |

| EPI_Accession_ID | Originating Laboratory | Submitting Laboratory |
|---|---|---|
| EPI_ISL_453771 | Virology Department, Sheffield Teaching Hospitals NHS Foundation Trust/Department of Infection, Immunity and Cardiovascular Disease, The Medical School, University of Sheffield | COVID-19 Genomics UK (COG-UK) Consortium |
| EPI_ISL_453772 | Virology Department, Sheffield Teaching Hospitals NHS Foundation Trust/Department of Infection, Immunity and Cardiovascular Disease, The Medical School, University of Sheffield | COVID-19 Genomics UK (COG-UK) Consortium |
| EPI_ISL_453773 | Virology Department, Sheffield Teaching Hospitals NHS Foundation Trust/Department of Infection, Immunity and Cardiovascular Disease, The Medical School, University of Sheffield | COVID-19 Genomics UK (COG-UK) Consortium |
| EPI_ISL_453774 | Virology Department, Sheffield Teaching Hospitals NHS Foundation Trust/Department of Infection, Immunity and Cardiovascular Disease, The Medical School, University of Sheffield | COVID-19 Genomics UK (COG-UK) Consortium |
| EPI_ISL_453775 | Virology Department, Sheffield Teaching Hospitals NHS Foundation Trust/Department of Infection, Immunity and Cardiovascular Disease, The Medical School, University of Sheffield | COVID-19 Genomics UK (COG-UK) Consortium |
| EPI_ISL_453776 | Virology Department, Sheffield Teaching Hospitals NHS Foundation Trust/Department of Infection, Immunity and Cardiovascular Disease, The Medical School, University of Sheffield | COVID-19 Genomics UK (COG-UK) Consortium |
| EPI_ISL_453777 | Virology Department, Sheffield Teaching Hospitals NHS Foundation Trust/Department of Infection, Immunity and Cardiovascular Disease, The Medical School, University of Sheffield | COVID-19 Genomics UK (COG-UK) Consortium |
| EPI_ISL_453778 | Virology Department, Sheffield Teaching Hospitals NHS Foundation Trust/Department of Infection, Immunity and Cardiovascular Disease, The Medical School, University of Sheffield | COVID-19 Genomics UK (COG-UK) Consortium |
| EPI_ISL_453813 | unknown | Instituto Nacional de Saude (INSA) |

| EPI_Accession_ID | Originating Laboratory | Submitting Laboratory |
|---|---|---|
| EPI_ISL_453814 | unknown | Instituto Nacional de Saude (INSA) |
| EPI_ISL_453815 | unknown | Instituto Nacional de Saude (INSA) |
| EPI_ISL_453816 | unknown | Instituto Nacional de Saude (INSA) |
| EPI_ISL_453817 | unknown | Instituto Nacional de Saude (INSA) |
| EPI_ISL_453818 | unknown | Instituto Nacional de Saude (INSA) |
| EPI_ISL_453819 | unknown | Instituto Nacional de Saude (INSA) |
| EPI_ISL_453820 | unknown | Instituto Nacional de Saude (INSA) |
| EPI_ISL_453821 | unknown | Instituto Nacional de Saude (INSA) |
| EPI_ISL_453822 | unknown | Instituto Nacional de Saude (INSA) |
| EPI_ISL_453823 | unknown | Instituto Nacional de Saude (INSA) |
| EPI_ISL_453824 | unknown | Instituto Nacional de Saude (INSA) |
| EPI_ISL_453825 | unknown | Instituto Nacional de Saude (INSA) |
| EPI_ISL_453826 | unknown | Instituto Nacional de Saude (INSA) |
| EPI_ISL_453827 | unknown | Instituto Nacional de Saude (INSA) |
| EPI_ISL_453828 | unknown | Instituto Nacional de Saude (INSA) |
| EPI_ISL_453829 | unknown | Instituto Nacional de Saude (INSA) |
| EPI_ISL_453830 | unknown | Instituto Nacional de Saude (INSA) |
| EPI_ISL_453831 | unknown | Instituto Nacional de Saude (INSA) |
| EPI_ISL_453832 | unknown | Instituto Nacional de Saude (INSA) |
| EPI_ISL_453833 | unknown | Instituto Nacional de Saude (INSA) |
| EPI_ISL_453834 | unknown | Instituto Nacional de Saude (INSA) |
| EPI_ISL_453835 | unknown | Instituto Nacional de Saude (INSA) |
| EPI_ISL_453836 | unknown | Instituto Nacional de Saude (INSA) |
| EPI_ISL_453837 | unknown | Instituto Nacional de Saude (INSA) |
| EPI_ISL_453838 | unknown | Instituto Nacional de Saude (INSA) |
| EPI_ISL_453839 | unknown | Instituto Nacional de Saude (INSA) |
| EPI_ISL_453840 | unknown | Instituto Nacional de Saude (INSA) |
| EPI_ISL_453841 | unknown | Instituto Nacional de Saude (INSA) |
| EPI_ISL_453842 | unknown | Instituto Nacional de Saude (INSA) |
| EPI_ISL_453843 | unknown | Instituto Nacional de Saude (INSA) |
| EPI_ISL_453844 | unknown | Instituto Nacional de Saude (INSA) |
| EPI_ISL_453845 | unknown | Instituto Nacional de Saude (INSA) |
| EPI_ISL_453846 | unknown | Instituto Nacional de Saude (INSA) |

| EPI_Accession_ID | Originating Laboratory | Submitting Laboratory |
|---|---|---|
| EPI_ISL_453847 | unknown | Instituto Nacional de Saude (INSA) |
| EPI_ISL_453848 | unknown | Instituto Nacional de Saude (INSA) |
| EPI_ISL_453849 | unknown | Instituto Nacional de Saude (INSA) |
| EPI_ISL_453850 | unknown | Instituto Nacional de Saude (INSA) |
| EPI_ISL_453851 | unknown | Instituto Nacional de Saude (INSA) |
| EPI_ISL_453852 | unknown | Instituto Nacional de Saude (INSA) |
| EPI_ISL_453853 | unknown | Instituto Nacional de Saude (INSA) |
| EPI_ISL_453854 | unknown | Instituto Nacional de Saude (INSA) |
| EPI_ISL_453855 | unknown | Instituto Nacional de Saude (INSA) |
| EPI_ISL_453856 | unknown | Instituto Nacional de Saude (INSA) |
| EPI_ISL_453857 | unknown | Instituto Nacional de Saude (INSA) |
| EPI_ISL_453858 | unknown | Instituto Nacional de Saude (INSA) |
| EPI_ISL_453859 | unknown | Instituto Nacional de Saude (INSA) |
| EPI_ISL_453860 | unknown | Instituto Nacional de Saude (INSA) |
| EPI_ISL_453861 | unknown | Instituto Nacional de Saude (INSA) |
| EPI_ISL_453862 | unknown | Instituto Nacional de Saude (INSA) |
| EPI_ISL_453863 | unknown | Instituto Nacional de Saude (INSA) |
| EPI_ISL_453864 | unknown | Instituto Nacional de Saude (INSA) |
| EPI_ISL_453865 | unknown | Instituto Nacional de Saude (INSA) |
| EPI_ISL_453866 | unknown | Instituto Nacional de Saude (INSA) |
| EPI_ISL_453867 | unknown | Instituto Nacional de Saude (INSA) |
| EPI_ISL_453868 | unknown | Instituto Nacional de Saude (INSA) |
| EPI_ISL_453869 | unknown | Instituto Nacional de Saude (INSA) |
| EPI_ISL_453870 | unknown | Instituto Nacional de Saude (INSA) |
| EPI_ISL_453871 | unknown | Instituto Nacional de Saude (INSA) |
| EPI_ISL_453872 | unknown | Instituto Nacional de Saude (INSA) |
| EPI_ISL_453873 | unknown | Instituto Nacional de Saude (INSA) |
| EPI_ISL_453874 | unknown | Instituto Nacional de Saude (INSA) |
| EPI_ISL_453875 | unknown | Instituto Nacional de Saude (INSA) |
| EPI_ISL_453876 | unknown | Instituto Nacional de Saude (INSA) |
| EPI_ISL_453877 | unknown | Instituto Nacional de Saude (INSA) |
| EPI_ISL_453878 | unknown | Instituto Nacional de Saude (INSA) |
| EPI_ISL_453879 | unknown | Instituto Nacional de Saude (INSA) |

| EPI_Accession_ID | Originating Laboratory | Submitting Laboratory |
|---|---|---|
| EPI_ISL_453880 | unknown | Instituto Nacional de Saude (INSA) |
| EPI_ISL_453881 | unknown | Instituto Nacional de Saude (INSA) |
| EPI_ISL_453882 | unknown | Instituto Nacional de Saude (INSA) |
| EPI_ISL_453883 | unknown | Instituto Nacional de Saude (INSA) |
| EPI_ISL_453884 | unknown | Instituto Nacional de Saude (INSA) |
| EPI_ISL_453885 | unknown | Instituto Nacional de Saude (INSA) |
| EPI_ISL_453886 | unknown | Instituto Nacional de Saude (INSA) |
| EPI_ISL_453887 | unknown | Instituto Nacional de Saude (INSA) |
| EPI_ISL_453888 | unknown | Instituto Nacional de Saude (INSA) |
| EPI_ISL_453889 | unknown | Instituto Nacional de Saude (INSA) |
| EPI_ISL_453890 | unknown | Instituto Nacional de Saude (INSA) |
| EPI_ISL_453891 | unknown | Instituto Nacional de Saude (INSA) |
| EPI_ISL_453892 | unknown | Instituto Nacional de Saude (INSA) |
| EPI_ISL_453893 | unknown | Instituto Nacional de Saude (INSA) |
| EPI_ISL_453894 | unknown | Instituto Nacional de Saude (INSA) |
| EPI_ISL_453895 | unknown | Instituto Nacional de Saude (INSA) |
| EPI_ISL_453896 | unknown | Instituto Nacional de Saude (INSA) |
| EPI_ISL_453897 | unknown | Instituto Nacional de Saude (INSA) |
| EPI_ISL_453898 | unknown | Instituto Nacional de Saude (INSA) |
| EPI_ISL_453899 | unknown | Instituto Nacional de Saude (INSA) |
| EPI_ISL_453900 | unknown | Instituto Nacional de Saude (INSA) |
| EPI_ISL_453901 | unknown | Instituto Nacional de Saude (INSA) |
| EPI_ISL_453902 | unknown | Instituto Nacional de Saude (INSA) |
| EPI_ISL_453903 | unknown | Instituto Nacional de Saude (INSA) |
| EPI_ISL_453904 | unknown | Instituto Nacional de Saude (INSA) |
| EPI_ISL_453905 | unknown | Instituto Nacional de Saude (INSA) |
| EPI_ISL_453906 | unknown | Instituto Nacional de Saude (INSA) |
| EPI_ISL_453907 | unknown | Instituto Nacional de Saude (INSA) |
| EPI_ISL_453908 | unknown | Instituto Nacional de Saude (INSA) |
| EPI_ISL_453909 | unknown | Instituto Nacional de Saude (INSA) |
| EPI_ISL_453910 | unknown | Instituto Nacional de Saude (INSA) |
| EPI_ISL_453911 | unknown | Instituto Nacional de Saude (INSA) |
| EPI_ISL_453912 | unknown | Instituto Nacional de Saude (INSA) |

| EPI_Accession_ID | Originating Laboratory | Submitting Laboratory |
|---|---|---|
| EPI_ISL_453913 | unknown | Instituto Nacional de Saude (INSA) |
| EPI_ISL_453914 | unknown | Instituto Nacional de Saude (INSA) |
| EPI_ISL_453915 | unknown | Instituto Nacional de Saude (INSA) |
| EPI_ISL_453916 | unknown | Instituto Nacional de Saude (INSA) |
| EPI_ISL_453917 | unknown | Instituto Nacional de Saude (INSA) |
| EPI_ISL_453919 | unknown | Instituto Nacional de Saude (INSA) |
| EPI_ISL_453920 | unknown | Instituto Nacional de Saude (INSA) |
| EPI_ISL_453921 | unknown | Instituto Nacional de Saude (INSA) |
| EPI_ISL_453922 | unknown | Instituto Nacional de Saude (INSA) |
| EPI_ISL_453923 | unknown | Instituto Nacional de Saude (INSA) |
| EPI_ISL_453924 | unknown | Instituto Nacional de Saude (INSA) |
| EPI_ISL_453925 | unknown | Instituto Nacional de Saude (INSA) |
| EPI_ISL_453926 | unknown | Instituto Nacional de Saude (INSA) |
| EPI_ISL_453927 | unknown | Instituto Nacional de Saude (INSA) |
| EPI_ISL_453928 | unknown | Instituto Nacional de Saude (INSA) |
| EPI_ISL_453929 | unknown | Instituto Nacional de Saude (INSA) |
| EPI_ISL_453930 | unknown | Instituto Nacional de Saude (INSA) |
| EPI_ISL_453931 | unknown | Instituto Nacional de Saude (INSA) |
| EPI_ISL_453932 | unknown | Instituto Nacional de Saude (INSA) |
| EPI_ISL_453933 | unknown | Instituto Nacional de Saude (INSA) |
| EPI_ISL_453934 | unknown | Instituto Nacional de Saude (INSA) |
| EPI_ISL_453935 | unknown | Instituto Nacional de Saude (INSA) |
| EPI_ISL_453936 | unknown | Instituto Nacional de Saude (INSA) |
| EPI_ISL_453937 | unknown | Instituto Nacional de Saude (INSA) |
| EPI_ISL_453939 | unknown | Instituto Nacional de Saude (INSA) |
| EPI_ISL_453940 | unknown | Instituto Nacional de Saude (INSA) |
| EPI_ISL_453941 | unknown | Instituto Nacional de Saude (INSA) |
| EPI_ISL_453942 | unknown | Instituto Nacional de Saude (INSA) |
| EPI_ISL_453943 | unknown | Instituto Nacional de Saude (INSA) |
| EPI_ISL_453944 | unknown | Instituto Nacional de Saude (INSA) |
| EPI_ISL_453945 | unknown | Instituto Nacional de Saude (INSA) |
| EPI_ISL_453946 | unknown | Instituto Nacional de Saude (INSA) |
| EPI_ISL_453947 | unknown | Instituto Nacional de Saude (INSA) |

| EPI_Accession_ID | Originating Laboratory | Submitting Laboratory |
|---|---|---|
| EPI_ISL_453948 | unknown | Instituto Nacional de Saude (INSA) |
| EPI_ISL_453949 | unknown | Instituto Nacional de Saude (INSA) |
| EPI_ISL_453950 | unknown | Instituto Nacional de Saude (INSA) |
| EPI_ISL_453951 | unknown | Instituto Nacional de Saude (INSA) |
| EPI_ISL_453952 | unknown | Instituto Nacional de Saude (INSA) |
| EPI_ISL_453953 | unknown | Instituto Nacional de Saude (INSA) |
| EPI_ISL_453954 | unknown | Instituto Nacional de Saude (INSA) |
| EPI_ISL_453955 | unknown | Instituto Nacional de Saude (INSA) |
| EPI_ISL_453956 | unknown | Instituto Nacional de Saude (INSA) |
| EPI_ISL_453957 | unknown | Instituto Nacional de Saude (INSA) |
| EPI_ISL_453958 | unknown | Instituto Nacional de Saude (INSA) |
| EPI_ISL_453959 | unknown | Instituto Nacional de Saude (INSA) |
| EPI_ISL_453960 | unknown | Instituto Nacional de Saude (INSA) |
| EPI_ISL_453961 | unknown | Instituto Nacional de Saude (INSA) |
| EPI_ISL_453962 | unknown | Instituto Nacional de Saude (INSA) |
| EPI_ISL_453963 | unknown | Instituto Nacional de Saude (INSA) |
| EPI_ISL_453964 | unknown | Instituto Nacional de Saude (INSA) |
| EPI_ISL_453965 | unknown | Instituto Nacional de Saude (INSA) |
| EPI_ISL_453966 | unknown | Instituto Nacional de Saude (INSA) |
| EPI_ISL_453967 | unknown | Instituto Nacional de Saude (INSA) |
| EPI_ISL_453968 | unknown | Instituto Nacional de Saude (INSA) |
| EPI_ISL_453969 | unknown | Instituto Nacional de Saude (INSA) |
| EPI_ISL_453970 | unknown | Instituto Nacional de Saude (INSA) |
| EPI_ISL_453971 | unknown | Instituto Nacional de Saude (INSA) |
| EPI_ISL_453972 | unknown | Instituto Nacional de Saude (INSA) |
| EPI_ISL_453973 | unknown | Instituto Nacional de Saude (INSA) |
| EPI_ISL_453974 | unknown | Instituto Nacional de Saude (INSA) |
| EPI_ISL_453975 | unknown | Instituto Nacional de Saude (INSA) |
| EPI_ISL_453976 | unknown | Instituto Nacional de Saude (INSA) |
| EPI_ISL_453977 | unknown | Instituto Nacional de Saude (INSA) |
| EPI_ISL_453978 | unknown | Instituto Nacional de Saude (INSA) |
| EPI_ISL_453979 | unknown | Instituto Nacional de Saude (INSA) |
| EPI_ISL_453980 | unknown | Instituto Nacional de Saude (INSA) |

| EPI_Accession_ID | Originating Laboratory | Submitting Laboratory |
|---|---|---|
| EPI_ISL_453981 | unknown | Instituto Nacional de Saude (INSA) |
| EPI_ISL_453982 | unknown | Instituto Nacional de Saude (INSA) |
| EPI_ISL_453983 | unknown | Instituto Nacional de Saude (INSA) |
| EPI_ISL_453984 | unknown | Instituto Nacional de Saude (INSA) |
| EPI_ISL_453985 | unknown | Instituto Nacional de Saude (INSA) |
| EPI_ISL_453986 | unknown | Instituto Nacional de Saude (INSA) |
| EPI_ISL_453987 | unknown | Instituto Nacional de Saude (INSA) |
| EPI_ISL_453988 | unknown | Instituto Nacional de Saude (INSA) |
| EPI_ISL_453989 | unknown | Instituto Nacional de Saude (INSA) |
| EPI_ISL_453990 | unknown | Instituto Nacional de Saude (INSA) |
| EPI_ISL_453991 | unknown | Instituto Nacional de Saude (INSA) |
| EPI_ISL_453992 | unknown | Instituto Nacional de Saude (INSA) |
| EPI_ISL_453993 | unknown | Instituto Nacional de Saude (INSA) |
| EPI_ISL_453994 | unknown | Instituto Nacional de Saude (INSA) |
| EPI_ISL_453995 | unknown | Instituto Nacional de Saude (INSA) |
| EPI_ISL_453996 | unknown | Instituto Nacional de Saude (INSA) |
| EPI_ISL_453997 | unknown | Instituto Nacional de Saude (INSA) |
| EPI_ISL_453998 | unknown | Instituto Nacional de Saude (INSA) |
| EPI_ISL_453999 | unknown | Instituto Nacional de Saude (INSA) |
| EPI_ISL_454000 | unknown | Instituto Nacional de Saude (INSA) |
| EPI_ISL_454001 | unknown | Instituto Nacional de Saude (INSA) |
| EPI_ISL_454002 | unknown | Instituto Nacional de Saude (INSA) |
| EPI_ISL_454003 | unknown | Instituto Nacional de Saude (INSA) |
| EPI_ISL_454004 | unknown | Instituto Nacional de Saude (INSA) |
| EPI_ISL_454005 | unknown | Instituto Nacional de Saude (INSA) |
| EPI_ISL_454006 | unknown | Instituto Nacional de Saude (INSA) |
| EPI_ISL_454007 | unknown | Instituto Nacional de Saude (INSA) |
| EPI_ISL_454008 | unknown | Instituto Nacional de Saude (INSA) |
| EPI_ISL_454009 | unknown | Instituto Nacional de Saude (INSA) |
| EPI_ISL_454010 | unknown | Instituto Nacional de Saude (INSA) |
| EPI_ISL_454011 | unknown | Instituto Nacional de Saude (INSA) |
| EPI_ISL_454012 | unknown | Instituto Nacional de Saude (INSA) |
| EPI_ISL_454013 | unknown | Instituto Nacional de Saude (INSA) |

| EPI_Accession_ID | Originating Laboratory | Submitting Laboratory |
|---|---|---|
| EPI_ISL_454014 | unknown | Instituto Nacional de Saude (INSA) |
| EPI_ISL_454015 | unknown | Instituto Nacional de Saude (INSA) |
| EPI_ISL_454016 | unknown | Instituto Nacional de Saude (INSA) |
| EPI_ISL_454017 | unknown | Instituto Nacional de Saude (INSA) |
| EPI_ISL_454018 | unknown | Instituto Nacional de Saude (INSA) |
| EPI_ISL_454019 | unknown | Instituto Nacional de Saude (INSA) |
| EPI_ISL_454020 | unknown | Instituto Nacional de Saude (INSA) |
| EPI_ISL_454021 | unknown | Instituto Nacional de Saude (INSA) |
| EPI_ISL_454022 | unknown | Instituto Nacional de Saude (INSA) |
| EPI_ISL_454023 | unknown | Instituto Nacional de Saude (INSA) |
| EPI_ISL_454024 | unknown | Instituto Nacional de Saude (INSA) |
| EPI_ISL_454025 | unknown | Instituto Nacional de Saude (INSA) |
| EPI_ISL_454026 | unknown | Instituto Nacional de Saude (INSA) |
| EPI_ISL_454027 | unknown | Instituto Nacional de Saude (INSA) |
| EPI_ISL_454028 | unknown | Instituto Nacional de Saude (INSA) |
| EPI_ISL_454029 | unknown | Instituto Nacional de Saude (INSA) |
| EPI_ISL_454030 | unknown | Instituto Nacional de Saude (INSA) |
| EPI_ISL_454031 | unknown | Instituto Nacional de Saude (INSA) |
| EPI_ISL_454032 | unknown | Instituto Nacional de Saude (INSA) |
| EPI_ISL_454033 | unknown | Instituto Nacional de Saude (INSA) |
| EPI_ISL_454034 | unknown | Instituto Nacional de Saude (INSA) |
| EPI_ISL_454035 | unknown | Instituto Nacional de Saude (INSA) |
| EPI_ISL_454036 | unknown | Instituto Nacional de Saude (INSA) |
| EPI_ISL_454037 | unknown | Instituto Nacional de Saude (INSA) |
| EPI_ISL_454038 | unknown | Instituto Nacional de Saude (INSA) |
| EPI_ISL_454039 | unknown | Instituto Nacional de Saude (INSA) |
| EPI_ISL_454040 | unknown | Instituto Nacional de Saude (INSA) |
| EPI_ISL_454041 | unknown | Instituto Nacional de Saude (INSA) |
| EPI_ISL_454042 | unknown | Instituto Nacional de Saude (INSA) |
| EPI_ISL_454043 | unknown | Instituto Nacional de Saude (INSA) |
| EPI_ISL_454044 | unknown | Instituto Nacional de Saude (INSA) |
| EPI_ISL_454045 | unknown | Instituto Nacional de Saude (INSA) |
| EPI_ISL_454046 | unknown | Instituto Nacional de Saude (INSA) |

| EPI_Accession_ID | Originating Laboratory | Submitting Laboratory |
|---|---|---|
| EPI_ISL_454047 | unknown | Instituto Nacional de Saude (INSA) |
| EPI_ISL_454048 | unknown | Instituto Nacional de Saude (INSA) |
| EPI_ISL_454049 | unknown | Instituto Nacional de Saude (INSA) |
| EPI_ISL_454050 | unknown | Instituto Nacional de Saude (INSA) |
| EPI_ISL_454051 | unknown | Instituto Nacional de Saude (INSA) |
| EPI_ISL_454052 | unknown | Instituto Nacional de Saude (INSA) |
| EPI_ISL_454053 | unknown | Instituto Nacional de Saude (INSA) |
| EPI_ISL_454054 | unknown | Instituto Nacional de Saude (INSA) |
| EPI_ISL_454055 | unknown | Instituto Nacional de Saude (INSA) |
| EPI_ISL_454056 | unknown | Instituto Nacional de Saude (INSA) |
| EPI_ISL_454057 | unknown | Instituto Nacional de Saude (INSA) |
| EPI_ISL_454058 | unknown | Instituto Nacional de Saude (INSA) |
| EPI_ISL_454059 | unknown | Instituto Nacional de Saude (INSA) |
| EPI_ISL_454060 | unknown | Instituto Nacional de Saude (INSA) |
| EPI_ISL_454061 | unknown | Instituto Nacional de Saude (INSA) |
| EPI_ISL_454062 | unknown | Instituto Nacional de Saude (INSA) |
| EPI_ISL_454063 | unknown | Instituto Nacional de Saude (INSA) |
| EPI_ISL_454064 | unknown | Instituto Nacional de Saude (INSA) |
| EPI_ISL_454065 | unknown | Instituto Nacional de Saude (INSA) |
| EPI_ISL_454066 | unknown | Instituto Nacional de Saude (INSA) |
| EPI_ISL_454067 | unknown | Instituto Nacional de Saude (INSA) |
| EPI_ISL_454068 | unknown | Instituto Nacional de Saude (INSA) |
| EPI_ISL_454069 | unknown | Instituto Nacional de Saude (INSA) |
| EPI_ISL_454070 | unknown | Instituto Nacional de Saude (INSA) |
| EPI_ISL_454071 | unknown | Instituto Nacional de Saude (INSA) |
| EPI_ISL_454072 | unknown | Instituto Nacional de Saude (INSA) |
| EPI_ISL_454073 | unknown | Instituto Nacional de Saude (INSA) |
| EPI_ISL_454074 | unknown | Instituto Nacional de Saude (INSA) |
| EPI_ISL_454075 | unknown | Instituto Nacional de Saude (INSA) |
| EPI_ISL_454076 | unknown | Instituto Nacional de Saude (INSA) |
| EPI_ISL_454077 | unknown | Instituto Nacional de Saude (INSA) |
| EPI_ISL_454078 | unknown | Instituto Nacional de Saude (INSA) |
| EPI_ISL_454079 | unknown | Instituto Nacional de Saude (INSA) |

| EPI_Accession_ID | Originating Laboratory | Submitting Laboratory |
|---|---|---|
| EPI_ISL_454080 | unknown | Instituto Nacional de Saude (INSA) |
| EPI_ISL_454081 | unknown | Instituto Nacional de Saude (INSA) |
| EPI_ISL_454082 | unknown | Instituto Nacional de Saude (INSA) |
| EPI_ISL_454083 | unknown | Instituto Nacional de Saude (INSA) |
| EPI_ISL_454084 | unknown | Instituto Nacional de Saude (INSA) |
| EPI_ISL_454085 | unknown | Instituto Nacional de Saude (INSA) |
| EPI_ISL_454086 | unknown | Instituto Nacional de Saude (INSA) |
| EPI_ISL_454087 | unknown | Instituto Nacional de Saude (INSA) |
| EPI_ISL_454088 | unknown | Instituto Nacional de Saude (INSA) |
| EPI_ISL_454089 | unknown | Instituto Nacional de Saude (INSA) |
| EPI_ISL_454090 | unknown | Instituto Nacional de Saude (INSA) |
| EPI_ISL_454091 | unknown | Instituto Nacional de Saude (INSA) |
| EPI_ISL_454092 | unknown | Instituto Nacional de Saude (INSA) |
| EPI_ISL_454093 | unknown | Instituto Nacional de Saude (INSA) |
| EPI_ISL_454094 | unknown | Instituto Nacional de Saude (INSA) |
| EPI_ISL_454095 | unknown | Instituto Nacional de Saude (INSA) |
| EPI_ISL_454096 | unknown | Instituto Nacional de Saude (INSA) |
| EPI_ISL_454097 | unknown | Instituto Nacional de Saude (INSA) |
| EPI_ISL_454098 | unknown | Instituto Nacional de Saude (INSA) |
| EPI_ISL_454099 | unknown | Instituto Nacional de Saude (INSA) |
| EPI_ISL_454100 | unknown | Instituto Nacional de Saude (INSA) |
| EPI_ISL_454101 | unknown | Instituto Nacional de Saude (INSA) |
| EPI_ISL_454102 | unknown | Instituto Nacional de Saude (INSA) |
| EPI_ISL_454103 | unknown | Instituto Nacional de Saude (INSA) |
| EPI_ISL_454104 | unknown | Instituto Nacional de Saude (INSA) |
| EPI_ISL_454105 | unknown | Instituto Nacional de Saude (INSA) |
| EPI_ISL_454106 | unknown | Instituto Nacional de Saude (INSA) |
| EPI_ISL_454107 | unknown | Instituto Nacional de Saude (INSA) |
| EPI_ISL_454108 | unknown | Instituto Nacional de Saude (INSA) |
| EPI_ISL_454109 | unknown | Instituto Nacional de Saude (INSA) |
| EPI_ISL_454110 | unknown | Instituto Nacional de Saude (INSA) |
| EPI_ISL_454111 | unknown | Instituto Nacional de Saude (INSA) |
| EPI_ISL_454112 | unknown | Instituto Nacional de Saude (INSA) |

| EPI_Accession_ID | Originating Laboratory | Submitting Laboratory |
|---|---|---|
| EPI_ISL_454113 | unknown | Instituto Nacional de Saude (INSA) |
| EPI_ISL_454114 | unknown | Instituto Nacional de Saude (INSA) |
| EPI_ISL_454115 | unknown | Instituto Nacional de Saude (INSA) |
| EPI_ISL_454116 | unknown | Instituto Nacional de Saude (INSA) |
| EPI_ISL_454117 | unknown | Instituto Nacional de Saude (INSA) |
| EPI_ISL_454118 | unknown | Instituto Nacional de Saude (INSA) |
| EPI_ISL_454119 | unknown | Instituto Nacional de Saude (INSA) |
| EPI_ISL_454120 | unknown | Instituto Nacional de Saude (INSA) |
| EPI_ISL_454121 | unknown | Instituto Nacional de Saude (INSA) |
| EPI_ISL_454122 | unknown | Instituto Nacional de Saude (INSA) |
| EPI_ISL_454123 | unknown | Instituto Nacional de Saude (INSA) |
| EPI_ISL_454124 | unknown | Instituto Nacional de Saude (INSA) |
| EPI_ISL_454125 | unknown | Instituto Nacional de Saude (INSA) |
| EPI_ISL_454126 | unknown | Instituto Nacional de Saude (INSA) |
| EPI_ISL_454127 | unknown | Instituto Nacional de Saude (INSA) |
| EPI_ISL_454128 | unknown | Instituto Nacional de Saude (INSA) |
| EPI_ISL_454129 | unknown | Instituto Nacional de Saude (INSA) |
| EPI_ISL_454130 | unknown | Instituto Nacional de Saude (INSA) |
| EPI_ISL_454131 | unknown | Instituto Nacional de Saude (INSA) |
| EPI_ISL_454132 | unknown | Instituto Nacional de Saude (INSA) |
| EPI_ISL_454133 | unknown | Instituto Nacional de Saude (INSA) |
| EPI_ISL_454134 | unknown | Instituto Nacional de Saude (INSA) |
| EPI_ISL_454135 | unknown | Instituto Nacional de Saude (INSA) |
| EPI_ISL_454136 | unknown | Instituto Nacional de Saude (INSA) |
| EPI_ISL_454137 | unknown | Instituto Nacional de Saude (INSA) |
| EPI_ISL_454138 | unknown | Instituto Nacional de Saude (INSA) |
| EPI_ISL_454139 | unknown | Instituto Nacional de Saude (INSA) |
| EPI_ISL_454140 | unknown | Instituto Nacional de Saude (INSA) |
| EPI_ISL_454141 | unknown | Instituto Nacional de Saude (INSA) |
| EPI_ISL_454142 | unknown | Instituto Nacional de Saude (INSA) |
| EPI_ISL_454143 | unknown | Instituto Nacional de Saude (INSA) |
| EPI_ISL_454144 | unknown | Instituto Nacional de Saude (INSA) |
| EPI_ISL_454145 | unknown | Instituto Nacional de Saude (INSA) |

| EPI_Accession_ID | Originating Laboratory | Submitting Laboratory |
|---|---|---|
| EPI_ISL_454146 | unknown | Instituto Nacional de Saude (INSA) |
| EPI_ISL_454147 | unknown | Instituto Nacional de Saude (INSA) |
| EPI_ISL_454148 | unknown | Instituto Nacional de Saude (INSA) |
| EPI_ISL_454149 | unknown | Instituto Nacional de Saude (INSA) |
| EPI_ISL_454150 | unknown | Instituto Nacional de Saude (INSA) |
| EPI_ISL_454151 | unknown | Instituto Nacional de Saude (INSA) |
| EPI_ISL_454152 | unknown | Instituto Nacional de Saude (INSA) |
| EPI_ISL_454153 | unknown | Instituto Nacional de Saude (INSA) |
| EPI_ISL_454154 | unknown | Instituto Nacional de Saude (INSA) |
| EPI_ISL_454155 | unknown | Instituto Nacional de Saude (INSA) |
| EPI_ISL_454158 | unknown | Instituto Nacional de Saude (INSA) |
| EPI_ISL_454159 | unknown | Instituto Nacional de Saude (INSA) |
| EPI_ISL_454160 | unknown | Instituto Nacional de Saude (INSA) |
| EPI_ISL_454161 | unknown | Instituto Nacional de Saude (INSA) |
| EPI_ISL_454162 | unknown | Instituto Nacional de Saude (INSA) |
| EPI_ISL_454163 | unknown | Instituto Nacional de Saude (INSA) |
| EPI_ISL_454164 | unknown | Instituto Nacional de Saude (INSA) |
| EPI_ISL_454165 | unknown | Instituto Nacional de Saude (INSA) |
| EPI_ISL_454166 | unknown | Instituto Nacional de Saude (INSA) |
| EPI_ISL_454167 | unknown | Instituto Nacional de Saude (INSA) |
| EPI_ISL_454168 | unknown | Instituto Nacional de Saude (INSA) |
| EPI_ISL_454169 | unknown | Instituto Nacional de Saude (INSA) |
| EPI_ISL_454170 | unknown | Instituto Nacional de Saude (INSA) |
| EPI_ISL_454171 | unknown | Instituto Nacional de Saude (INSA) |
| EPI_ISL_454172 | unknown | Instituto Nacional de Saude (INSA) |
| EPI_ISL_454173 | unknown | Instituto Nacional de Saude (INSA) |
| EPI_ISL_454174 | unknown | Instituto Nacional de Saude (INSA) |
| EPI_ISL_454175 | unknown | Instituto Nacional de Saude (INSA) |
| EPI_ISL_454176 | unknown | Instituto Nacional de Saude (INSA) |
| EPI_ISL_454177 | unknown | Instituto Nacional de Saude (INSA) |
| EPI_ISL_454178 | unknown | Instituto Nacional de Saude (INSA) |
| EPI_ISL_454179 | unknown | Instituto Nacional de Saude (INSA) |
| EPI_ISL_454180 | unknown | Instituto Nacional de Saude (INSA) |

| EPI_Accession_ID | Originating Laboratory | Submitting Laboratory |
|---|---|---|
| EPI_ISL_454181 | unknown | Instituto Nacional de Saude (INSA) |
| EPI_ISL_454182 | unknown | Instituto Nacional de Saude (INSA) |
| EPI_ISL_454183 | unknown | Instituto Nacional de Saude (INSA) |
| EPI_ISL_454184 | unknown | Instituto Nacional de Saude (INSA) |
| EPI_ISL_454185 | unknown | Instituto Nacional de Saude (INSA) |
| EPI_ISL_454186 | unknown | Instituto Nacional de Saude (INSA) |
| EPI_ISL_454187 | unknown | Instituto Nacional de Saude (INSA) |
| EPI_ISL_454188 | unknown | Instituto Nacional de Saude (INSA) |
| EPI_ISL_454189 | unknown | Instituto Nacional de Saude (INSA) |
| EPI_ISL_454190 | unknown | Instituto Nacional de Saude (INSA) |
| EPI_ISL_454191 | unknown | Instituto Nacional de Saude (INSA) |
| EPI_ISL_454192 | unknown | Instituto Nacional de Saude (INSA) |
| EPI_ISL_454193 | unknown | Instituto Nacional de Saude (INSA) |
| EPI_ISL_454194 | unknown | Instituto Nacional de Saude (INSA) |
| EPI_ISL_454195 | unknown | Instituto Nacional de Saude (INSA) |
| EPI_ISL_454196 | unknown | Instituto Nacional de Saude (INSA) |
| EPI_ISL_454197 | unknown | Instituto Nacional de Saude (INSA) |
| EPI_ISL_454198 | unknown | Instituto Nacional de Saude (INSA) |
| EPI_ISL_454199 | unknown | Instituto Nacional de Saude (INSA) |
| EPI_ISL_454200 | unknown | Instituto Nacional de Saude (INSA) |
| EPI_ISL_454201 | unknown | Instituto Nacional de Saude (INSA) |
| EPI_ISL_454202 | unknown | Instituto Nacional de Saude (INSA) |
| EPI_ISL_454203 | unknown | Instituto Nacional de Saude (INSA) |
| EPI_ISL_454204 | unknown | Instituto Nacional de Saude (INSA) |
| EPI_ISL_454205 | unknown | Instituto Nacional de Saude (INSA) |
| EPI_ISL_454206 | unknown | Instituto Nacional de Saude (INSA) |
| EPI_ISL_454207 | unknown | Instituto Nacional de Saude (INSA) |
| EPI_ISL_454208 | unknown | Instituto Nacional de Saude (INSA) |
| EPI_ISL_454209 | unknown | Instituto Nacional de Saude (INSA) |
| EPI_ISL_454210 | unknown | Instituto Nacional de Saude (INSA) |
| EPI_ISL_454211 | unknown | Instituto Nacional de Saude (INSA) |
| EPI_ISL_454212 | unknown | Instituto Nacional de Saude (INSA) |
| EPI_ISL_454213 | unknown | Instituto Nacional de Saude (INSA) |

| EPI_Accession_ID | Originating Laboratory | Submitting Laboratory |
|---|---|---|
| EPI_ISL_454214 | unknown | Instituto Nacional de Saude (INSA) |
| EPI_ISL_454215 | unknown | Instituto Nacional de Saude (INSA) |
| EPI_ISL_454216 | unknown | Instituto Nacional de Saude (INSA) |
| EPI_ISL_454217 | unknown | Instituto Nacional de Saude (INSA) |
| EPI_ISL_454218 | unknown | Instituto Nacional de Saude (INSA) |
| EPI_ISL_454219 | unknown | Instituto Nacional de Saude (INSA) |
| EPI_ISL_454220 | unknown | Instituto Nacional de Saude (INSA) |
| EPI_ISL_454221 | unknown | Instituto Nacional de Saude (INSA) |
| EPI_ISL_454222 | unknown | Instituto Nacional de Saude (INSA) |
| EPI_ISL_454223 | unknown | Instituto Nacional de Saude (INSA) |
| EPI_ISL_454224 | unknown | Instituto Nacional de Saude (INSA) |
| EPI_ISL_454225 | unknown | Instituto Nacional de Saude (INSA) |
| EPI_ISL_454226 | unknown | Instituto Nacional de Saude (INSA) |
| EPI_ISL_454227 | unknown | Instituto Nacional de Saude (INSA) |
| EPI_ISL_454228 | unknown | Instituto Nacional de Saude (INSA) |
| EPI_ISL_454229 | unknown | Instituto Nacional de Saude (INSA) |
| EPI_ISL_454230 | unknown | Instituto Nacional de Saude (INSA) |
| EPI_ISL_454231 | unknown | Instituto Nacional de Saude (INSA) |
| EPI_ISL_454232 | unknown | Instituto Nacional de Saude (INSA) |
| EPI_ISL_454233 | unknown | Instituto Nacional de Saude (INSA) |
| EPI_ISL_454234 | unknown | Instituto Nacional de Saude (INSA) |
| EPI_ISL_454235 | unknown | Instituto Nacional de Saude (INSA) |
| EPI_ISL_454236 | unknown | Instituto Nacional de Saude (INSA) |
| EPI_ISL_454237 | unknown | Instituto Nacional de Saude (INSA) |
| EPI_ISL_454238 | unknown | Instituto Nacional de Saude (INSA) |
| EPI_ISL_454239 | unknown | Instituto Nacional de Saude (INSA) |
| EPI_ISL_454240 | unknown | Instituto Nacional de Saude (INSA) |
| EPI_ISL_454241 | unknown | Instituto Nacional de Saude (INSA) |
| EPI_ISL_454242 | unknown | Instituto Nacional de Saude (INSA) |
| EPI_ISL_454243 | unknown | Instituto Nacional de Saude (INSA) |
| EPI_ISL_454244 | unknown | Instituto Nacional de Saude (INSA) |
| EPI_ISL_454245 | unknown | Instituto Nacional de Saude (INSA) |
| EPI_ISL_454246 | unknown | Instituto Nacional de Saude (INSA) |

| EPI_Accession_ID | Originating Laboratory | Submitting Laboratory |
|---|---|---|
| EPI_ISL_454247 | unknown | Instituto Nacional de Saude (INSA) |
| EPI_ISL_454248 | unknown | Instituto Nacional de Saude (INSA) |
| EPI_ISL_454249 | unknown | Instituto Nacional de Saude (INSA) |
| EPI_ISL_454250 | unknown | Instituto Nacional de Saude (INSA) |
| EPI_ISL_454251 | unknown | Instituto Nacional de Saude (INSA) |
| EPI_ISL_454252 | unknown | Instituto Nacional de Saude (INSA) |
| EPI_ISL_454253 | unknown | Instituto Nacional de Saude (INSA) |
| EPI_ISL_454254 | unknown | Instituto Nacional de Saude (INSA) |
| EPI_ISL_454255 | unknown | Instituto Nacional de Saude (INSA) |
| EPI_ISL_454256 | unknown | Instituto Nacional de Saude (INSA) |
| EPI_ISL_454257 | unknown | Instituto Nacional de Saude (INSA) |
| EPI_ISL_454258 | unknown | Instituto Nacional de Saude (INSA) |
| EPI_ISL_454259 | unknown | Instituto Nacional de Saude (INSA) |
| EPI_ISL_454260 | unknown | Instituto Nacional de Saude (INSA) |
| EPI_ISL_454261 | unknown | Instituto Nacional de Saude (INSA) |
| EPI_ISL_454262 | unknown | Instituto Nacional de Saude (INSA) |
| EPI_ISL_454263 | unknown | Instituto Nacional de Saude (INSA) |
| EPI_ISL_454264 | unknown | Instituto Nacional de Saude (INSA) |
| EPI_ISL_454265 | unknown | Instituto Nacional de Saude (INSA) |
| EPI_ISL_454266 | unknown | Instituto Nacional de Saude (INSA) |
| EPI_ISL_454267 | unknown | Instituto Nacional de Saude (INSA) |
| EPI_ISL_454268 | unknown | Instituto Nacional de Saude (INSA) |
| EPI_ISL_454269 | unknown | Instituto Nacional de Saude (INSA) |
| EPI_ISL_454270 | unknown | Instituto Nacional de Saude (INSA) |
| EPI_ISL_454271 | unknown | Instituto Nacional de Saude (INSA) |
| EPI_ISL_454272 | unknown | Instituto Nacional de Saude (INSA) |
| EPI_ISL_454274 | unknown | Instituto Nacional de Saude (INSA) |
| EPI_ISL_454275 | unknown | Instituto Nacional de Saude (INSA) |
| EPI_ISL_454276 | unknown | Instituto Nacional de Saude (INSA) |
| EPI_ISL_454277 | unknown | Instituto Nacional de Saude (INSA) |
| EPI_ISL_454278 | unknown | Instituto Nacional de Saude (INSA) |
| EPI_ISL_454279 | unknown | Instituto Nacional de Saude (INSA) |
| EPI_ISL_454280 | unknown | Instituto Nacional de Saude (INSA) |

| EPI_Accession_ID | Originating Laboratory | Submitting Laboratory |
|---|---|---|
| EPI_ISL_454281 | unknown | Instituto Nacional de Saude (INSA) |
| EPI_ISL_454282 | unknown | Instituto Nacional de Saude (INSA) |
| EPI_ISL_454283 | unknown | Instituto Nacional de Saude (INSA) |
| EPI_ISL_454284 | unknown | Instituto Nacional de Saude (INSA) |
| EPI_ISL_454285 | unknown | Instituto Nacional de Saude (INSA) |
| EPI_ISL_454286 | unknown | Instituto Nacional de Saude (INSA) |
| EPI_ISL_454287 | unknown | Instituto Nacional de Saude (INSA) |
| EPI_ISL_454288 | unknown | Instituto Nacional de Saude (INSA) |
| EPI_ISL_454289 | unknown | Instituto Nacional de Saude (INSA) |
| EPI_ISL_454290 | unknown | Instituto Nacional de Saude (INSA) |
| EPI_ISL_454291 | unknown | Instituto Nacional de Saude (INSA) |
| EPI_ISL_454292 | unknown | Instituto Nacional de Saude (INSA) |
| EPI_ISL_454293 | unknown | Instituto Nacional de Saude (INSA) |
| EPI_ISL_454294 | unknown | Instituto Nacional de Saude (INSA) |
| EPI_ISL_454295 | unknown | Instituto Nacional de Saude (INSA) |
| EPI_ISL_454296 | unknown | Instituto Nacional de Saude (INSA) |
| EPI_ISL_454297 | unknown | Instituto Nacional de Saude (INSA) |
| EPI_ISL_454298 | unknown | Instituto Nacional de Saude (INSA) |
| EPI_ISL_454299 | unknown | Instituto Nacional de Saude (INSA) |
| EPI_ISL_454300 | unknown | Instituto Nacional de Saude (INSA) |
| EPI_ISL_454301 | unknown | Instituto Nacional de Saude (INSA) |
| EPI_ISL_454302 | unknown | Instituto Nacional de Saude (INSA) |
| EPI_ISL_454303 | unknown | Instituto Nacional de Saude (INSA) |
| EPI_ISL_454304 | unknown | Instituto Nacional de Saude (INSA) |
| EPI_ISL_454305 | unknown | Instituto Nacional de Saude (INSA) |
| EPI_ISL_454307 | unknown | Instituto Nacional de Saude (INSA) |
| EPI_ISL_454308 | unknown | Instituto Nacional de Saude (INSA) |
| EPI_ISL_454309 | unknown | Instituto Nacional de Saude (INSA) |
| EPI_ISL_454310 | unknown | Instituto Nacional de Saude (INSA) |
| EPI_ISL_454311 | unknown | Instituto Nacional de Saude (INSA) |
| EPI_ISL_454312 | unknown | Instituto Nacional de Saude (INSA) |
| EPI_ISL_454313 | unknown | Instituto Nacional de Saude (INSA) |
| EPI_ISL_454314 | unknown | Instituto Nacional de Saude (INSA) |

| EPI_Accession_ID | Originating Laboratory | Submitting Laboratory |
|---|---|---|
| EPI_ISL_454315 | unknown | Instituto Nacional de Saude (INSA) |
| EPI_ISL_454316 | unknown | Instituto Nacional de Saude (INSA) |
| EPI_ISL_454317 | unknown | Instituto Nacional de Saude (INSA) |
| EPI_ISL_454318 | unknown | Instituto Nacional de Saude (INSA) |
| EPI_ISL_454319 | unknown | Instituto Nacional de Saude (INSA) |
| EPI_ISL_454320 | unknown | Instituto Nacional de Saude (INSA) |
| EPI_ISL_454321 | unknown | Instituto Nacional de Saude (INSA) |
| EPI_ISL_454322 | unknown | Instituto Nacional de Saude (INSA) |
| EPI_ISL_454323 | unknown | Instituto Nacional de Saude (INSA) |
| EPI_ISL_454324 | unknown | Instituto Nacional de Saude (INSA) |
| EPI_ISL_454325 | unknown | Instituto Nacional de Saude (INSA) |
| EPI_ISL_454326 | unknown | Instituto Nacional de Saude (INSA) |
| EPI_ISL_454327 | unknown | Instituto Nacional de Saude (INSA) |
| EPI_ISL_454328 | unknown | Instituto Nacional de Saude (INSA) |
| EPI_ISL_454329 | unknown | Instituto Nacional de Saude (INSA) |
| EPI_ISL_454330 | unknown | Instituto Nacional de Saude (INSA) |
| EPI_ISL_454331 | unknown | Instituto Nacional de Saude (INSA) |
| EPI_ISL_454332 | unknown | Instituto Nacional de Saude (INSA) |
| EPI_ISL_454333 | unknown | Instituto Nacional de Saude (INSA) |
| EPI_ISL_454334 | unknown | Instituto Nacional de Saude (INSA) |
| EPI_ISL_454335 | unknown | Instituto Nacional de Saude (INSA) |
| EPI_ISL_454336 | unknown | Instituto Nacional de Saude (INSA) |
| EPI_ISL_454337 | unknown | Instituto Nacional de Saude (INSA) |
| EPI_ISL_454338 | unknown | Instituto Nacional de Saude (INSA) |
| EPI_ISL_454339 | unknown | Instituto Nacional de Saude (INSA) |
| EPI_ISL_454340 | unknown | Instituto Nacional de Saude (INSA) |
| EPI_ISL_454341 | unknown | Instituto Nacional de Saude (INSA) |
| EPI_ISL_454342 | unknown | Instituto Nacional de Saude (INSA) |
| EPI_ISL_454343 | unknown | Instituto Nacional de Saude (INSA) |
| EPI_ISL_454344 | unknown | Instituto Nacional de Saude (INSA) |
| EPI_ISL_454345 | unknown | Instituto Nacional de Saude (INSA) |
| EPI_ISL_454346 | unknown | Instituto Nacional de Saude (INSA) |
| EPI_ISL_454347 | unknown | Instituto Nacional de Saude (INSA) |

| EPI_Accession_ID | Originating Laboratory | Submitting Laboratory |
|---|---|---|
| EPI_ISL_454348 | unknown | Instituto Nacional de Saude (INSA) |
| EPI_ISL_454349 | unknown | Instituto Nacional de Saude (INSA) |
| EPI_ISL_454350 | unknown | Instituto Nacional de Saude (INSA) |
| EPI_ISL_454351 | unknown | Instituto Nacional de Saude (INSA) |
| EPI_ISL_454352 | unknown | Instituto Nacional de Saude (INSA) |
| EPI_ISL_454353 | Microbial Genome Sequencing Center; Microbial Genomic Epidemiology Laboratory | Microbial Genomic Epidemiology Laboratory, University of Pittsburgh |
| EPI_ISL_454354 | UPMC Clinical Microbiology Laboratory | Microbial Genome Sequencing Center; Microbial Genomic Epidemiology Laboratory, University of Pittsburgh |
| EPI_ISL_454355 | UPMC Clinical Microbiology Laboratory | Microbial Genome Sequencing Center, Microbial Genomic Epidemiological Laboratory |
| EPI_ISL_454356 | UPMC Clinical Microbiology Laboratory | Microbial Genome Sequencing Center, Microbial Genomic Epidemiological Laboratory |
| EPI_ISL_454357 | UPMC Clinical Microbiology Laboratory | Microbial Genome Sequencing Center, Microbial Genomic Epidemiological Laboratory |
| EPI_ISL_454358 | UPMC Clinical Microbiology Laboratory | Microbial Genome Sequencing Center, Microbial Genomic Epidemiological Laboratory |
| EPI_ISL_454359 | UPMC Clinical Microbiology Laboratory | Microbial Genome Sequencing Center, Microbial Genomic Epidemiological Laboratory |
| EPI_ISL_454360 | UPMC Clinical Microbiology Laboratory | Microbial Genome Sequencing Center, Microbial Genomic Epidemiological Laboratory |
| EPI_ISL_454361 | UPMC Clinical Microbiology Laboratory | Microbial Genome Sequencing Center, Microbial Genomic Epidemiological Laboratory |
| EPI_ISL_454362 | UPMC Clinical Microbiology Laboratory | Microbial Genome Sequencing Center, Microbial Genomic Epidemiological Laboratory |
| EPI_ISL_454363 | UPMC Clinical Microbiology Laboratory | Microbial Genome Sequencing Center, Microbial Genomic Epidemiological Laboratory |
| EPI_ISL_454364 | UPMC Clinical Microbiology Laboratory | Microbial Genome Sequencing Center, Microbial Genomic Epidemiological Laboratory |
| EPI_ISL_454365 | UPMC Clinical Microbiology Laboratory | Microbial Genome Sequencing Center, Microbial Genomic Epidemiological Laboratory |

| EPI_Accession_ID | Originating Laboratory | Submitting Laboratory |
|---|---|---|
| EPI_ISL_454366 | UPMC Clinical Microbiology Laboratory | Microbial Genome Sequencing Center, Microbial Genomic Epidemiological Laboratory |
| EPI_ISL_454367 | UPMC Clinical Microbiology Laboratory | Microbial Genome Sequencing Center, Microbial Genomic Epidemiological Laboratory |
| EPI_ISL_454368 | UPMC Clinical Microbiology Laboratory | Microbial Genome Sequencing Center, Microbial Genomic Epidemiological Laboratory |
| EPI_ISL_454370 | UPMC Clinical Microbiology Laboratory | Microbial Genome Sequencing Center, Microbial Genomic Epidemiological Laboratory |
| EPI_ISL_454371 | UPMC Clinical Microbiology Laboratory | Microbial Genome Sequencing Center, Microbial Genomic Epidemiological Laboratory |
| EPI_ISL_454372 | UPMC Clinical Microbiology Laboratory | Microbial Genome Sequencing Center, Microbial Genomic Epidemiological Laboratory |
| EPI_ISL_454373 | UPMC Clinical Microbiology Laboratory | Microbial Genome Sequencing Center, Microbial Genomic Epidemiological Laboratory |
| EPI_ISL_454374 | UPMC Clinical Microbiology Laboratory | Microbial Genome Sequencing Center, Microbial Genomic Epidemiological Laboratory |
| EPI_ISL_454375 | UPMC Clinical Microbiology Laboratory | Microbial Genome Sequencing Center, Microbial Genomic Epidemiological Laboratory |
| EPI_ISL_454376 | UPMC Clinical Microbiology Laboratory | Microbial Genome Sequencing Center, Microbial Genomic Epidemiological Laboratory |
| EPI_ISL_454378 | UPMC Clinical Microbiology Laboratory | Microbial Genome Sequencing Center, Microbial Genomic Epidemiological Laboratory |
| EPI_ISL_454379 | UPMC Clinical Microbiology Laboratory | Microbial Genome Sequencing Center, Microbial Genomic Epidemiological Laboratory |
| EPI_ISL_454380 | UPMC Clinical Microbiology Laboratory | Microbial Genome Sequencing Center, Microbial Genomic Epidemiological Laboratory |
| EPI_ISL_454381 | UPMC Clinical Microbiology Laboratory | Microbial Genome Sequencing Center, Microbial Genomic Epidemiological Laboratory |
| EPI_ISL_454382 | UPMC Clinical Microbiology Laboratory | Microbial Genome Sequencing Center, Microbial Genomic Epidemiological Laboratory |
| EPI_ISL_454383 | UPMC Clinical Microbiology Laboratory | Microbial Genome Sequencing Center, Microbial Genomic Epidemiological Laboratory |

| EPI_Accession_ID | Originating Laboratory | Submitting Laboratory |
|---|---|---|
| EPI_ISL_454384 | UPMC Clinical Microbiology Laboratory | Microbial Genome Sequencing Center, Microbial Genomic Epidemiological Laboratory |
| EPI_ISL_454387 | UPMC Clinical Microbiology Laboratory | Microbial Genome Sequencing Center, Microbial Genomic Epidemiological Laboratory |
| EPI_ISL_454399 | UPMC Clinical Microbiology Laboratory | Microbial Genome Sequencing Center, Microbial Genomic Epidemiological Laboratory |
| EPI_ISL_454401 | UPMC Clinical Microbiology Laboratory | Microbial Genome Sequencing Center, Microbial Genomic Epidemiological Laboratory |
| EPI_ISL_454402 | UPMC Clinical Microbiology Laboratory | Microbial Genome Sequencing Center, Microbial Genomic Epidemiological Laboratory |
| EPI_ISL_454403 | UPMC Clinical Microbiology Laboratory | Microbial Genome Sequencing Center, Microbial Genomic Epidemiological Laboratory |
| EPI_ISL_454404 | UPMC Clinical Microbiology Laboratory | Microbial Genome Sequencing Center, Microbial Genomic Epidemiological Laboratory |
| EPI_ISL_454406 | UPMC Clinical Microbiology Laboratory | Microbial Genome Sequencing Center, Microbial Genomic Epidemiological Laboratory |
| EPI_ISL_454407 | UPMC Clinical Microbiology Laboratory | Microbial Genome Sequencing Center, Microbial Genomic Epidemiological Laboratory |
| EPI_ISL_454408 | UPMC Clinical Microbiology Laboratory | Microbial Genome Sequencing Center, Microbial Genomic Epidemiological Laboratory |
| EPI_ISL_454409 | UPMC Clinical Microbiology Laboratory | Microbial Genome Sequencing Center, Microbial Genomic Epidemiological Laboratory |
| EPI_ISL_454410 | UPMC Clinical Microbiology Laboratory | Microbial Genome Sequencing Center, Microbial Genomic Epidemiological Laboratory |
| EPI_ISL_454411 | UPMC Clinical Microbiology Laboratory | Microbial Genome Sequencing Center, Microbial Genomic Epidemiological Laboratory |
| EPI_ISL_454416 | unknown | Department of Medical Microbiology |
| EPI_ISL_454417 | unknown | Research and Experiment Center |
| EPI_ISL_454418 | unknown | Research and Experiment Center |
| EPI_ISL_454420 | Rafik Hariri University Hospital | Rafik Hariri University Hospital |
| EPI_ISL_454421 | Maryland Department of Health Laboratories Administration | Maryland Department of Health Laboratories Administration |

| EPI_Accession_ID | Originating Laboratory | Submitting Laboratory |
|---|---|---|
| EPI_ISL_454422 | Maryland Department of Health Laboratories Administration | Maryland Department of Health Laboratories Administration |
| EPI_ISL_454423 | Maryland Department of Health Laboratories Administration | Maryland Department of Health Laboratories Administration |
| EPI_ISL_454424 | Maryland Department of Health Laboratories Administration | Maryland Department of Health Laboratories Administration |
| EPI_ISL_454425 | Maryland Department of Health Laboratories Administration | Maryland Department of Health Laboratories Administration |
| EPI_ISL_454426 | Maryland Department of Health Laboratories Administration | Maryland Department of Health Laboratories Administration |
| EPI_ISL_454427 | Maryland Department of Health Laboratories Administration | Maryland Department of Health Laboratories Administration |
| EPI_ISL_454428 | Maryland Department of Health Laboratories Administration | Maryland Department of Health Laboratories Administration |
| EPI_ISL_454432 | Maryland Department of Health Laboratories Administration | Maryland Department of Health Laboratories Administration |
| EPI_ISL_454433 | Maryland Department of Health Laboratories Administration | Maryland Department of Health Laboratories Administration |
| EPI_ISL_454434 | Maryland Department of Health Laboratories Administration | Maryland Department of Health Laboratories Administration |
| EPI_ISL_454436 | Halmstad klinisk mikrobiologi | The Public Health Agency of Sweden |
| EPI_ISL_454437 | Halmstad klinisk mikrobiologi | The Public Health Agency of Sweden |
| EPI_ISL_454438 | Halmstad klinisk mikrobiologi | The Public Health Agency of Sweden |
| EPI_ISL_454439 | Halmstad klinisk mikrobiologi | The Public Health Agency of Sweden |
| EPI_ISL_454440 | Halmstad klinisk mikrobiologi | The Public Health Agency of Sweden |
| EPI_ISL_454441 | Halmstad klinisk mikrobiologi | The Public Health Agency of Sweden |
| EPI_ISL_454442 | Halmstad klinisk mikrobiologi | The Public Health Agency of Sweden |
| EPI_ISL_454443 | Halmstad klinisk mikrobiologi | The Public Health Agency of Sweden |
| EPI_ISL_454444 | Halmstad klinisk mikrobiologi | The Public Health Agency of Sweden |
| EPI_ISL_454445 | Halmstad klinisk mikrobiologi | The Public Health Agency of Sweden |
| EPI_ISL_454446 | Halmstad klinisk mikrobiologi | The Public Health Agency of Sweden |
| EPI_ISL_454447 | Karolinska Universitetslaboratoriet | The Public Health Agency of Sweden |
| EPI_ISL_454448 | Karolinska Universitetslaboratoriet | The Public Health Agency of Sweden |

| EPI_Accession_ID | Originating Laboratory | Submitting Laboratory |
| --- | --- | --- |
| EPI_ISL_454449 | Karolinska Universitetslaboratoriet | The Public Health Agency of Sweden |
| EPI_ISL_454450 | Karolinska Universitetslaboratoriet | The Public Health Agency of Sweden |
| EPI_ISL_454451 | Karolinska Universitetslaboratoriet | The Public Health Agency of Sweden |
| EPI_ISL_454452 | Karolinska Universitetslaboratoriet | The Public Health Agency of Sweden |
| EPI_ISL_454453 | Karolinska Universitetslaboratoriet | The Public Health Agency of Sweden |
| EPI_ISL_454454 | Karolinska Universitetslaboratoriet | The Public Health Agency of Sweden |
| EPI_ISL_454455 | Karolinska Universitetslaboratoriet | The Public Health Agency of Sweden |
| EPI_ISL_454456 | Karolinska Universitetslaboratoriet | The Public Health Agency of Sweden |
| EPI_ISL_454457 | Karolinska Universitetslaboratoriet | The Public Health Agency of Sweden |
| EPI_ISL_454458 | Karolinska Universitetslaboratoriet | The Public Health Agency of Sweden |
| EPI_ISL_454459 | Karolinska Universitetslaboratoriet | The Public Health Agency of Sweden |
| EPI_ISL_454460 | Karolinska Universitetslaboratoriet | The Public Health Agency of Sweden |
| EPI_ISL_454461 | Karolinska Universitetslaboratoriet | The Public Health Agency of Sweden |
| EPI_ISL_454462 | Karolinska Universitetslaboratoriet | The Public Health Agency of Sweden |
| EPI_ISL_454463 | Karolinska Universitetslaboratoriet | The Public Health Agency of Sweden |
| EPI_ISL_454464 | Karolinska Universitetslaboratoriet | The Public Health Agency of Sweden |
| EPI_ISL_454465 | Karolinska Universitetslaboratoriet | The Public Health Agency of Sweden |
| EPI_ISL_454466 | Karolinska Universitetslaboratoriet | The Public Health Agency of Sweden |
| EPI_ISL_454467 | Karolinska Universitetslaboratoriet | The Public Health Agency of Sweden |
| EPI_ISL_454468 | Karolinska Universitetslaboratoriet | The Public Health Agency of Sweden |
| EPI_ISL_454469 | Karolinska Universitetslaboratoriet | The Public Health Agency of Sweden |
| EPI_ISL_454470 | Karolinska Universitetslaboratoriet | The Public Health Agency of Sweden |
| EPI_ISL_454471 | Karolinska Universitetslaboratoriet | The Public Health Agency of Sweden |
| EPI_ISL_454472 | Karolinska Universitetslaboratoriet | The Public Health Agency of Sweden |
| EPI_ISL_454473 | Karolinska Universitetslaboratoriet | The Public Health Agency of Sweden |
| EPI_ISL_454474 | Karolinska Universitetslaboratoriet | The Public Health Agency of Sweden |
| EPI_ISL_454475 | Karolinska Universitetslaboratoriet | The Public Health Agency of Sweden |
| EPI_ISL_454476 | Karolinska Universitetslaboratoriet | The Public Health Agency of Sweden |
| EPI_ISL_454477 | Karolinska Universitetslaboratoriet | The Public Health Agency of Sweden |
| EPI_ISL_454478 | Karolinska Universitetslaboratoriet | The Public Health Agency of Sweden |
| EPI_ISL_454479 | Karolinska Universitetslaboratoriet | The Public Health Agency of Sweden |
| EPI_ISL_454480 | Karolinska Universitetslaboratoriet | The Public Health Agency of Sweden |
| EPI_ISL_454481 | Karolinska Universitetslaboratoriet | The Public Health Agency of Sweden |

| EPI_Accession_ID | Originating Laboratory | Submitting Laboratory |
|---|---|---|
| EPI_ISL_454482 | Karolinska Universitetslaboratoriet | The Public Health Agency of Sweden |
| EPI_ISL_454483 | Karolinska Universitetslaboratoriet | The Public Health Agency of Sweden |
| EPI_ISL_454484 | Karolinska Universitetslaboratoriet | The Public Health Agency of Sweden |
| EPI_ISL_454485 | Karolinska Universitetslaboratoriet | The Public Health Agency of Sweden |
| EPI_ISL_454486 | Karolinska Universitetslaboratoriet | The Public Health Agency of Sweden |
| EPI_ISL_454487 | Karolinska Universitetslaboratoriet | The Public Health Agency of Sweden |
| EPI_ISL_454488 | Karolinska Universitetslaboratoriet | The Public Health Agency of Sweden |
| EPI_ISL_454489 | Karolinska Universitetslaboratoriet | The Public Health Agency of Sweden |
| EPI_ISL_454490 | Karolinska Universitetslaboratoriet | The Public Health Agency of Sweden |
| EPI_ISL_454491 | Karolinska Universitetslaboratoriet | The Public Health Agency of Sweden |
| EPI_ISL_454492 | Karolinska Universitetslaboratoriet | The Public Health Agency of Sweden |
| EPI_ISL_454493 | Karolinska Universitetslaboratoriet | The Public Health Agency of Sweden |
| EPI_ISL_454494 | Skovde/Unilabs | The Public Health Agency of Sweden |
| EPI_ISL_454495 | Skovde/Unilabs | The Public Health Agency of Sweden |
| EPI_ISL_454496 | Skovde/Unilabs | The Public Health Agency of Sweden |
| EPI_ISL_454497 | RSE "National Center for Biotechnology" | RSE "National Center for Biotechnology" |
| EPI_ISL_454498 | RSE "National Center for Biotechnology" | RSE "National Center for Biotechnology" |
| EPI_ISL_454499 | RSE "National Center for Biotechnology" | RSE "National Center for Biotechnology" |
| EPI_ISL_454500 | RSE "National Center for Biotechnology" | RSE "National Center for Biotechnology" |
| EPI_ISL_454501 | RSE "National Center for Biotechnology" | RSE "National Center for Biotechnology" |
| EPI_ISL_454502 | RSE "National Center for Biotechnology" | RSE "National Center for Biotechnology" |
| EPI_ISL_454503 | RSE "National Center for Biotechnology" | RSE "National Center for Biotechnology" |
| EPI_ISL_454504 | RSE "National Center for Biotechnology" | RSE "National Center for Biotechnology" |
| EPI_ISL_454505 | RSE "National Center for Biotechnology" | RSE "National Center for Biotechnology" |
| EPI_ISL_454506 | RSE "National Center for Biotechnology" | RSE "National Center for Biotechnology" |
| EPI_ISL_454507 | RSE "National Center for Biotechnology" | RSE "National Center for Biotechnology" |
| EPI_ISL_454508 | RSE "National Center for Biotechnology" | RSE "National Center for Biotechnology" |
| EPI_ISL_454509 | RSE "National Center for Biotechnology" | RSE "National Center for Biotechnology" |
| EPI_ISL_454510 | RSE "National Center for Biotechnology" | RSE "National Center for Biotechnology" |
| EPI_ISL_454511 | RSE "National Center for Biotechnology" | RSE "National Center for Biotechnology" |
| EPI_ISL_454512 | RSE "National Center for Biotechnology" | RSE "National Center for Biotechnology" |
| EPI_ISL_454513 | RSE "National Center for Biotechnology" | RSE "National Center for Biotechnology" |
| EPI_ISL_454514 | RSE "National Center for Biotechnology" | RSE "National Center for Biotechnology" |

| EPI_Accession_ID | Originating Laboratory | Submitting Laboratory |
|---|---|---|
| EPI_ISL_454515 | RSE "National Center for Biotechnology" | RSE "National Center for Biotechnology" |
| EPI_ISL_454516 | RSE "National Center for Biotechnology" | RSE "National Center for Biotechnology" |
| EPI_ISL_454517 | RSE "National Center for Biotechnology" | RSE "National Center for Biotechnology" |
| EPI_ISL_454518 | RSE "National Center for Biotechnology" | RSE "National Center for Biotechnology" |
| EPI_ISL_454519 | RSE "National Center for Biotechnology" | RSE "National Center for Biotechnology" |
| EPI_ISL_454520 | RSE "National Center for Biotechnology" | RSE "National Center for Biotechnology" |
| EPI_ISL_454521 | NIV Influenza | NIV Influenza |
| EPI_ISL_454522 | NIV Influenza | NIV Influenza |
| EPI_ISL_454524 | NIV Influenza | NIV Influenza |
| EPI_ISL_454525 | NIV Influenza | NIV Influenza |
| EPI_ISL_454526 | NIV Influenza | NIV Influenza |
| EPI_ISL_454527 | NIV Influenza | NIV Influenza |
| EPI_ISL_454528 | NIV Influenza | NIV Influenza |
| EPI_ISL_454529 | NIV Influenza | NIV Influenza |
| EPI_ISL_454530 | NIV Influenza | NIV Influenza |
| EPI_ISL_454531 | NIV Influenza | NIV Influenza |
| EPI_ISL_454532 | NIV Influenza | NIV Influenza |
| EPI_ISL_454533 | NIV Influenza | NIV Influenza |
| EPI_ISL_454534 | NIV Influenza | NIV Influenza |
| EPI_ISL_454535 | NIV Influenza | NIV Influenza |
| EPI_ISL_454536 | NIV Influenza | NIV Influenza |
| EPI_ISL_454537 | NIV Influenza | NIV Influenza |
| EPI_ISL_454540 | NIV Influenza | NIV Influenza |
| EPI_ISL_454541 | NIV Influenza | NIV Influenza |
| EPI_ISL_454542 | NIV Influenza | NIV Influenza |
| EPI_ISL_454543 | NIV Influenza | NIV Influenza |
| EPI_ISL_454544 | NIV Influenza | NIV Influenza |
| EPI_ISL_454546 | NIV Influenza | NIV Influenza |
| EPI_ISL_454547 | NIV Influenza | NIV Influenza |
| EPI_ISL_454548 | NIV Influenza | NIV Influenza |
| EPI_ISL_454549 | NIV Influenza | NIV Influenza |
| EPI_ISL_454550 | NIV Influenza | NIV Influenza |
| EPI_ISL_454551 | NIV Influenza | NIV Influenza |

| EPI_Accession_ID | Originating Laboratory | Submitting Laboratory |
|---|---|---|
| EPI_ISL_454552 | NIV Influenza | NIV Influenza |
| EPI_ISL_454555 | NIV Influenza | NIV Influenza |
| EPI_ISL_454556 | NIV Influenza | NIV Influenza |
| EPI_ISL_454557 | NIV Influenza | NIV Influenza |
| EPI_ISL_454558 | NIV Influenza | NIV Influenza |
| EPI_ISL_454560 | NIV Influenza | NIV Influenza |
| EPI_ISL_454561 | NIV Influenza | NIV Influenza |
| EPI_ISL_454562 | NIV Influenza | NIV Influenza |
| EPI_ISL_454563 | NIV Influenza | NIV Influenza |
| EPI_ISL_454564 | NIV Influenza | NIV Influenza |
| EPI_ISL_454565 | NIV Influenza | NIV Influenza |
| EPI_ISL_454566 | NIV Influenza | NIV Influenza |
| EPI_ISL_454567 | NIV Influenza | NIV Influenza |
| EPI_ISL_454568 | NIV Influenza | NIV Influenza |
| EPI_ISL_454569 | NIV Influenza | NIV Influenza |
| EPI_ISL_454570 | NIV Influenza | NIV Influenza |
| EPI_ISL_454571 | National Center of Expertise | National Center for Expertise, National Center for Biotechnology, Kazakhstan |
| EPI_ISL_454572 | National Center of Expertise | National Center for Expertise, Kazakhstan National Center for Biotechnology, Kazakhstan |
| EPI_ISL_454574 | nstitute for Public Health | Laboratory for advanced genomics |
| EPI_ISL_454575 | Laboratory of virology, National Center of Expertise | Laboratory of molecular-genetic research, National Center for Expertise, Kazakhstan National Center for Biotechnology, Kazakhstan |
| EPI_ISL_454576 | Laboratory of virology, National Center of Expertise | Laboratory of molecular-genetic research, National Center of Expertise, Kazakhstan National Center for Biotechnology, Kazakhstan |
| EPI_ISL_454577 | Laboratory of virology, National Center of Expertise | Laboratory of molecular-genetic research, National Center of Expertise, Kazakhstan National Center for Biotechnology, Kazakhstan |
| EPI_ISL_454578 | University Hospital for Infectious Diseases "Dr. Fran MihaljeviÄ‡", Research Unit | University of Zagreb, Centre for research and knowledge transfer in biotechnology |

| EPI_Accession_ID | Originating Laboratory | Submitting Laboratory |
|---|---|---|
| EPI_ISL_454579 | Laboratory of virology, National Center of Expertise | Laboratory of molecular-genetic research, National Center of Expertise, Kazakhstan National Center for Biotechnology, Kazakhstan |
| EPI_ISL_454580 | Laboratory of virology, National Center of Expertise | Laboratory of molecular-genetic research, National Center of Expertise, Kazakhstan National Center for Biotechnology, Kazakhstan |
| EPI_ISL_454581 | University Hospital for Infectious Diseases "Dr. Fran MihaljeviÄ‡", Research Unit | University of Zagreb, Centre for research and knowledge transfer in biotechnology |
| EPI_ISL_454582 | Laboratory of virology, National Center of Expertise | Laboratory of molecular-genetic research, National Center of Expertise, Kazakhstan National Center for Biotechnology, Kazakhstan |
| EPI_ISL_454583 | University Hospital for Infectious Diseases "Dr. Fran MihaljeviÄ‡", Research Unit | University of Zagreb, Centre for research and knowledge transfer in biotechnology |
| EPI_ISL_454584 | Laboratory of virology, National Center of Expertise | Laboratory of molecular-genetic research, National Center for Expertise, Kazakhstan National Center for Biotechnology, Kazakhstan |
| EPI_ISL_454585 | Laboratory of virology, National Center of Expertise | Laboratory of molecular-genetic research, National Center for Expertise, Kazakhstan National Center for Biotechnology, Kazakhstan |
| EPI_ISL_454586 | Laboratory of virology, National Center of Expertise | Laboratory of molecular-genetic research, National Center for Expertise, Kazakhstan National Center for Biotechnology, Kazakhstan |
| EPI_ISL_454587 | Laboratory of virology, National Center of Expertise | Laboratory of molecular-genetic research, National Center of Expertise, Kazakhstan National Center for Biotechnology, Kazakhstan |
| EPI_ISL_454588 | University Hospital for Infectious Diseases "Dr. Fran MihaljeviÄ‡", Research Unit | University of Zagreb, Centre for research and knowledge transfer in biotechnology |
| EPI_ISL_454589 | Laboratory of virology, National Center of Expertise | Laboratory of molecular-genetic research, National Center for Expertise, Kazakhstan National Center for Biotechnology, Kazakhstan |
| EPI_ISL_454590 | Laboratory of virology, National Center of Expertise | Laboratory of molecular-genetic research, National Center of Expertise, Kazakhstan National Center for Biotechnology, Kazakhstan |

| EPI_Accession_ID | Originating Laboratory | Submitting Laboratory |
|---|---|---|
| EPI_ISL_454591 | Laboratory of virology, National Center of Expertise | Laboratory of molecular-genetic research, National Center of Expertise, Kazakhstan National Center for Biotechnology, Kazakhstan |
| EPI_ISL_454592 | University Hospital for Infectious Diseases "Dr. Fran MihaljeviÄ‡", Research Unit | University of Zagreb, Centre for research and knowledge transfer in biotechnology |
| EPI_ISL_454593 | Laboratory of virology, National Center of Expertise | Laboratory of molecular-genetic research, National Center of Expertise, Kazakhstan National Center for Biotechnology, Kazakhstan |
| EPI_ISL_454594 | Laboratory of virology, National Center of Expertise | Laboratory of molecular-genetic research, National Center of Expertise, Kazakhstan National Center for Biotechnology, Kazakhstan |
| EPI_ISL_454596 | Laboratory of virology, National Center of Expertise | Laboratory of molecular-genetic research, National Center for Expertise, Kazakhstan National Center for Biotechnology, Kazakhstan |
| EPI_ISL_454597 | Laboratory of virology, National Center of Expertise | Laboratory of molecular-genetic research, National Center for Expertise, Kazakhstan National Center for Biotechnology, Kazakhstan |
| EPI_ISL_454598 | Laboratory of virology, National Center of Expertise | Laboratory of molecular-genetic research, National Center of Expertise, Kazakhstan National Center for Biotechnology, Kazakhstan |
| EPI_ISL_454599 | Laboratory of virology, National Center of Expertise | Laboratory of molecular-genetic research, National Center for Expertise, Kazakhstan National Center for Biotechnology, Kazakhstan |
| EPI_ISL_454600 | Laboratory of virology, National Center of Expertise | Laboratory of molecular-genetic research, National Center for Expertise, Kazakhstan National Center for Biotechnology, Kazakhstan |
| EPI_ISL_454601 | Laboratory of virology, National Center of Expertise | Laboratory of molecular-genetic research, National Center of Expertise, Kazakhstan National Center for Biotechnology, Kazakhstan |
| EPI_ISL_454602 | Croatian Institute of Public Health | University of Zagreb, Centre for research and knowledge transfer in biotechnology |

| EPI_Accession_ID | Originating Laboratory | Submitting Laboratory |
|---|---|---|
| EPI_ISL_454603 | Laboratory of virology, National Center of Expertise | Laboratory of molecular-genetic research, National Center of Expertise, Kazakhstan National Center for Biotechnology, Kazakhstan |
| EPI_ISL_454604 | Laboratory of virology, National Center of Expertise | Laboratory of molecular-genetic research, National Center for Expertise, Kazakhstan National Center for Biotechnology, Kazakhstan |
| EPI_ISL_454605 | Institute for Public Health | Laboratory for advanced genomics |
| EPI_ISL_454606 | Institute for Public Health | Laboratory for advanced genomics |
| EPI_ISL_454607 | Alameda County Public Health Lab | Chan-Zuckerberg Biohub |
| EPI_ISL_454608 | Alameda County Public Health Lab | Chan-Zuckerberg Biohub |
| EPI_ISL_454609 | Alameda County Public Health Lab | Chan-Zuckerberg Biohub |
| EPI_ISL_454610 | Alameda County Public Health Lab | Chan-Zuckerberg Biohub |
| EPI_ISL_454611 | Alameda County Public Health Lab | Chan-Zuckerberg Biohub |
| EPI_ISL_454612 | Alameda County Public Health Lab | Chan-Zuckerberg Biohub |
| EPI_ISL_454613 | Alameda County Public Health Lab | Chan-Zuckerberg Biohub |
| EPI_ISL_454614 | UCSF Clinical Microbiology Laboratory | Chan-Zuckerberg Biohub |
| EPI_ISL_454615 | UCSF Clinical Microbiology Laboratory | Chan-Zuckerberg Biohub |
| EPI_ISL_454616 | UCSF Clinical Microbiology Laboratory | Chan-Zuckerberg Biohub |
| EPI_ISL_454617 | UCSF Clinical Microbiology Laboratory | Chan-Zuckerberg Biohub |
| EPI_ISL_454618 | UCSF Clinical Microbiology Laboratory | Chan-Zuckerberg Biohub |
| EPI_ISL_454619 | UCSF Clinical Microbiology Laboratory | Chan-Zuckerberg Biohub |
| EPI_ISL_454620 | UCSF Clinical Microbiology Laboratory | Chan-Zuckerberg Biohub |
| EPI_ISL_454621 | UCSF Clinical Microbiology Laboratory | Chan-Zuckerberg Biohub |
| EPI_ISL_454622 | UCSF Clinical Microbiology Laboratory | Chan-Zuckerberg Biohub |
| EPI_ISL_454623 | UCSF Clinical Microbiology Laboratory | Chan-Zuckerberg Biohub |
| EPI_ISL_454624 | UCSF Clinical Microbiology Laboratory | Chan-Zuckerberg Biohub |
| EPI_ISL_454625 | UCSF Clinical Microbiology Laboratory | Chan-Zuckerberg Biohub |
| EPI_ISL_454626 | UCSF Clinical Microbiology Laboratory | Chan-Zuckerberg Biohub |
| EPI_ISL_454627 | UCSF Clinical Microbiology Laboratory | Chan-Zuckerberg Biohub |
| EPI_ISL_454628 | UCSF Clinical Microbiology Laboratory | Chan-Zuckerberg Biohub |
| EPI_ISL_454629 | UCSF Clinical Microbiology Laboratory | Chan-Zuckerberg Biohub |
| EPI_ISL_454630 | UCSF Clinical Microbiology Laboratory | Chan-Zuckerberg Biohub |
| EPI_ISL_454631 | UCSF Clinical Microbiology Laboratory | Chan-Zuckerberg Biohub |

| EPI_Accession_ID | Originating Laboratory | Submitting Laboratory |
|---|---|---|
| EPI_ISL_454632 | UCSF Clinical Microbiology Laboratory | Chan-Zuckerberg Biohub |
| EPI_ISL_454633 | UCSF Clinical Microbiology Laboratory | Chan-Zuckerberg Biohub |
| EPI_ISL_454634 | UCSF Clinical Microbiology Laboratory | Chan-Zuckerberg Biohub |
| EPI_ISL_454635 | County Of San Luis Obispo Public Health Laboratory | Chan-Zuckerberg Biohub |
| EPI_ISL_454636 | Humboldt County Public Health Laboratory | Chan-Zuckerberg Biohub |
| EPI_ISL_454637 | Humboldt County Public Health Laboratory | Chan-Zuckerberg Biohub |
| EPI_ISL_454638 | Humboldt County Public Health Laboratory | Chan-Zuckerberg Biohub |
| EPI_ISL_454639 | Humboldt County Public Health Laboratory | Chan-Zuckerberg Biohub |
| EPI_ISL_454640 | Humboldt County Public Health Laboratory | Chan-Zuckerberg Biohub |
| EPI_ISL_454641 | Humboldt County Public Health Laboratory | Chan-Zuckerberg Biohub |
| EPI_ISL_454642 | CT-Dr. Katherine A. Kelley State Public Health Lab | Pathogen Discovery, Respiratory Viruses Branch, Division of Viral Diseases, Centers for Disease Control and Prevention |
| EPI_ISL_454643 | VI-US Virgin Islands Department of Health | Pathogen Discovery, Respiratory Viruses Branch, Division of Viral Diseases, Centers for Disease Control and Prevention |
| EPI_ISL_454644 | VI-US Virgin Islands Department of Health | Pathogen Discovery, Respiratory Viruses Branch, Division of Viral Diseases, Centers for Disease Control and Prevention |
| EPI_ISL_454645 | CT-Dr. Katherine A. Kelley State Public Health Lab | Pathogen Discovery, Respiratory Viruses Branch, Division of Viral Diseases, Centers for Disease Control and Prevention |
| EPI_ISL_454646 | City of El Paso Department of Public Health Laboratory | Pathogen Discovery, Respiratory Viruses Branch, Division of Viral Diseases, Centers for Disease Control and Prevention |
| EPI_ISL_454647 | VI-US Virgin Islands Department of Health | Pathogen Discovery, Respiratory Viruses Branch, Division of Viral Diseases, Centers for Disease Control and Prevention |
| EPI_ISL_454648 | VI-US Virgin Islands Department of Health | Pathogen Discovery, Respiratory Viruses Branch, Division of Viral Diseases, Centers for Disease Control and Prevention |

| EPI_Accession_ID | Originating Laboratory | Submitting Laboratory |
|---|---|---|
| EPI_ISL_454649 | VI-US Virgin Islands Department of Health | Pathogen Discovery, Respiratory Viruses Branch, Division of Viral Diseases, Centers for Disease Control and Prevention |
| EPI_ISL_454650 | VI-US Virgin Islands Department of Health | Pathogen Discovery, Respiratory Viruses Branch, Division of Viral Diseases, Centers for Disease Control and Prevention |
| EPI_ISL_454651 | VI-US Virgin Islands Department of Health | Pathogen Discovery, Respiratory Viruses Branch, Division of Viral Diseases, Centers for Disease Control and Prevention |
| EPI_ISL_454652 | VI-US Virgin Islands Department of Health | Pathogen Discovery, Respiratory Viruses Branch, Division of Viral Diseases, Centers for Disease Control and Prevention |
| EPI_ISL_454653 | County of Santa Clara Public Health Department | Chan-Zuckerberg Biohub |
| EPI_ISL_454654 | County of Santa Clara Public Health Department | Chan-Zuckerberg Biohub |
| EPI_ISL_454655 | County of Santa Clara Public Health Department | Chan-Zuckerberg Biohub |
| EPI_ISL_454656 | County of Santa Clara Public Health Department | Chan-Zuckerberg Biohub |
| EPI_ISL_454657 | County of Santa Clara Public Health Department | Chan-Zuckerberg Biohub |
| EPI_ISL_454658 | County of Santa Clara Public Health Department | Chan-Zuckerberg Biohub |
| EPI_ISL_454659 | County of Santa Clara Public Health Department | Chan-Zuckerberg Biohub |
| EPI_ISL_454660 | County of Santa Clara Public Health Department | Chan-Zuckerberg Biohub |
| EPI_ISL_454661 | County of Santa Clara Public Health Department | Chan-Zuckerberg Biohub |
| EPI_ISL_454662 | County of Santa Clara Public Health Department | Chan-Zuckerberg Biohub |
| EPI_ISL_454663 | County of Santa Clara Public Health Department | Chan-Zuckerberg Biohub |
| EPI_ISL_454664 | County of Santa Clara Public Health Department | Chan-Zuckerberg Biohub |
| EPI_ISL_454665 | County of Santa Clara Public Health Department | Chan-Zuckerberg Biohub |
| EPI_ISL_454666 | County of Santa Clara Public Health Department | Chan-Zuckerberg Biohub |
| EPI_ISL_454667 | County of Santa Clara Public Health Department | Chan-Zuckerberg Biohub |
| EPI_ISL_454668 | County of Santa Clara Public Health Department | Chan-Zuckerberg Biohub |
| EPI_ISL_454669 | County of Santa Clara Public Health Department | Chan-Zuckerberg Biohub |
| EPI_ISL_454670 | County of Santa Clara Public Health Department | Chan-Zuckerberg Biohub |
| EPI_ISL_454671 | County of Santa Clara Public Health Department | Chan-Zuckerberg Biohub |
| EPI_ISL_454672 | County of Santa Clara Public Health Department | Chan-Zuckerberg Biohub |
| EPI_ISL_454673 | County of Santa Clara Public Health Department | Chan-Zuckerberg Biohub |

| EPI_Accession_ID | Originating Laboratory | Submitting Laboratory |
|---|---|---|
| EPI_ISL_454674 | County of Santa Clara Public Health Department | Chan-Zuckerberg Biohub |
| EPI_ISL_454675 | County of Santa Clara Public Health Department | Chan-Zuckerberg Biohub |
| EPI_ISL_454676 | County of Santa Clara Public Health Department | Chan-Zuckerberg Biohub |
| EPI_ISL_454677 | County of Santa Clara Public Health Department | Chan-Zuckerberg Biohub |
| EPI_ISL_454678 | County of Santa Clara Public Health Department | Chan-Zuckerberg Biohub |
| EPI_ISL_454679 | County of Santa Clara Public Health Department | Chan-Zuckerberg Biohub |
| EPI_ISL_454680 | County of Santa Clara Public Health Department | Chan-Zuckerberg Biohub |
| EPI_ISL_454681 | County of Santa Clara Public Health Department | Chan-Zuckerberg Biohub |
| EPI_ISL_454682 | County of Santa Clara Public Health Department | Chan-Zuckerberg Biohub |
| EPI_ISL_454683 | County of Santa Clara Public Health Department | Chan-Zuckerberg Biohub |
| EPI_ISL_454684 | County of Santa Clara Public Health Department | Chan-Zuckerberg Biohub |
| EPI_ISL_454685 | County of Santa Clara Public Health Department | Chan-Zuckerberg Biohub |
| EPI_ISL_454686 | County of Santa Clara Public Health Department | Chan-Zuckerberg Biohub |
| EPI_ISL_454687 | County of Santa Clara Public Health Department | Chan-Zuckerberg Biohub |
| EPI_ISL_454688 | County of Santa Clara Public Health Department | Chan-Zuckerberg Biohub |
| EPI_ISL_454689 | County of Santa Clara Public Health Department | Chan-Zuckerberg Biohub |
| EPI_ISL_454690 | Emory Molecular Diagnostics Laboratory, Emory Healthcare | Piantadosi Lab, Emory Department of Pathology |
| EPI_ISL_454691 | Quest Diagnostics | Quest Diagnostics |
| EPI_ISL_454692 | Quest Diagnostics | Quest Diagnostics |
| EPI_ISL_454693 | Quest Diagnostics | Quest Diagnostics |
| EPI_ISL_454707 | Utah Public Health Laboratory | Utah Public Health Laboratory |
| EPI_ISL_454708 | Utah Public Health Laboratory | Utah Public Health Laboratory |
| EPI_ISL_454709 | Utah Public Health Laboratory | Utah Public Health Laboratory |
| EPI_ISL_454710 | Utah Public Health Laboratory | Utah Public Health Laboratory |
| EPI_ISL_454711 | Utah Public Health Laboratory | Utah Public Health Laboratory |
| EPI_ISL_454712 | Utah Public Health Laboratory | Utah Public Health Laboratory |
| EPI_ISL_454713 | Utah Public Health Laboratory | Utah Public Health Laboratory |
| EPI_ISL_454714 | Utah Public Health Laboratory | Utah Public Health Laboratory |
| EPI_ISL_454715 | Utah Public Health Laboratory | Utah Public Health Laboratory |
| EPI_ISL_454716 | Utah Public Health Laboratory | Utah Public Health Laboratory |
| EPI_ISL_454717 | Utah Public Health Laboratory | Utah Public Health Laboratory |
| EPI_ISL_454718 | Utah Public Health Laboratory | Utah Public Health Laboratory |

| EPI_Accession_ID | Originating Laboratory | Submitting Laboratory |
|---|---|---|
| EPI_ISL_454719 | Utah Public Health Laboratory | Utah Public Health Laboratory |
| EPI_ISL_454720 | Utah Public Health Laboratory | Utah Public Health Laboratory |
| EPI_ISL_454721 | Utah Public Health Laboratory | Utah Public Health Laboratory |
| EPI_ISL_454722 | Utah Public Health Laboratory | Utah Public Health Laboratory |
| EPI_ISL_454723 | Utah Public Health Laboratory | Utah Public Health Laboratory |
| EPI_ISL_454724 | Utah Public Health Laboratory | Utah Public Health Laboratory |
| EPI_ISL_454725 | Utah Public Health Laboratory | Utah Public Health Laboratory |
| EPI_ISL_454726 | Utah Public Health Laboratory | Utah Public Health Laboratory |
| EPI_ISL_454727 | Utah Public Health Laboratory | Utah Public Health Laboratory |
| EPI_ISL_454728 | Utah Public Health Laboratory | Utah Public Health Laboratory |
| EPI_ISL_454729 | Utah Public Health Laboratory | Utah Public Health Laboratory |
| EPI_ISL_454730 | Utah Public Health Laboratory | Utah Public Health Laboratory |
| EPI_ISL_454731 | Utah Public Health Laboratory | Utah Public Health Laboratory |
| EPI_ISL_454732 | Russian State Collection of Viruses | Pathogenic Microorganisms Variability Laboratory |
| EPI_ISL_454733 | unknown | Department of Medical |
| EPI_ISL_454749 | Japanese Quarantine Stations | Pathogen Genomics Center, National Institute of Infectious Diseases |
| EPI_ISL_454750 | Dutch COVID-19 response team | National Institute for Public Health and the Environment (RIVM) |
| EPI_ISL_454751 | Dutch COVID-19 response team | National Institute for Public Health and the Environment (RIVM) |
| EPI_ISL_454752 | Dutch COVID-19 response team | National Institute for Public Health and the Environment (RIVM) |
| EPI_ISL_454753 | Dutch COVID-19 response team | National Institute for Public Health and the Environment (RIVM) |
| EPI_ISL_454754 | Dutch COVID-19 response team | National Institute for Public Health and the Environment (RIVM) |
| EPI_ISL_454755 | Dutch COVID-19 response team | National Institute for Public Health and the Environment (RIVM) |
| EPI_ISL_454756 | Dutch COVID-19 response team | National Institute for Public Health and the Environment (RIVM) |
| EPI_ISL_454757 | Dutch COVID-19 response team | National Institute for Public Health and the Environment (RIVM) |

| EPI_Accession_ID | Originating Laboratory | Submitting Laboratory |
|---|---|---|
| EPI_ISL_454758 | Dutch COVID-19 response team | National Institute for Public Health and the Environment (RIVM) |
| EPI_ISL_454759 | Dutch COVID-19 response team | National Institute for Public Health and the Environment (RIVM) |
| EPI_ISL_454760 | Dutch COVID-19 response team | National Institute for Public Health and the Environment (RIVM) |
| EPI_ISL_454761 | Dutch COVID-19 response team | National Institute for Public Health and the Environment (RIVM) |
| EPI_ISL_454762 | Dutch COVID-19 response team | National Institute for Public Health and the Environment (RIVM) |
| EPI_ISL_454763 | Dutch COVID-19 response team | National Institute for Public Health and the Environment (RIVM) |
| EPI_ISL_454764 | Dutch COVID-19 response team | National Institute for Public Health and the Environment (RIVM) |
| EPI_ISL_454765 | Dutch COVID-19 response team | National Institute for Public Health and the Environment (RIVM) |
| EPI_ISL_454766 | Dutch COVID-19 response team | National Institute for Public Health and the Environment (RIVM) |
| EPI_ISL_454767 | Dutch COVID-19 response team | National Institute for Public Health and the Environment (RIVM) |
| EPI_ISL_454769 | Dutch COVID-19 response team | National Institute for Public Health and the Environment (RIVM) |
| EPI_ISL_454770 | Dutch COVID-19 response team | National Institute for Public Health and the Environment (RIVM) |
| EPI_ISL_454771 | Dutch COVID-19 response team | National Institute for Public Health and the Environment (RIVM) |
| EPI_ISL_454772 | Dutch COVID-19 response team | National Institute for Public Health and the Environment (RIVM) |
| EPI_ISL_454773 | Dutch COVID-19 response team | National Institute for Public Health and the Environment (RIVM) |
| EPI_ISL_454774 | Dutch COVID-19 response team | National Institute for Public Health and the Environment (RIVM) |

| EPI_Accession_ID | Originating Laboratory | Submitting Laboratory |
|---|---|---|
| EPI_ISL_454775 | Dutch COVID-19 response team | National Institute for Public Health and the Environment (RIVM) |
| EPI_ISL_454776 | Dutch COVID-19 response team | National Institute for Public Health and the Environment (RIVM) |
| EPI_ISL_454777 | Dutch COVID-19 response team | National Institute for Public Health and the Environment (RIVM) |
| EPI_ISL_454778 | Dutch COVID-19 response team | National Institute for Public Health and the Environment (RIVM) |
| EPI_ISL_454779 | Dutch COVID-19 response team | National Institute for Public Health and the Environment (RIVM) |
| EPI_ISL_454780 | Dutch COVID-19 response team | National Institute for Public Health and the Environment (RIVM) |
| EPI_ISL_454781 | Dutch COVID-19 response team | National Institute for Public Health and the Environment (RIVM) |
| EPI_ISL_454782 | Dutch COVID-19 response team | National Institute for Public Health and the Environment (RIVM) |
| EPI_ISL_454783 | Dutch COVID-19 response team | National Institute for Public Health and the Environment (RIVM) |
| EPI_ISL_454784 | Dutch COVID-19 response team | National Institute for Public Health and the Environment (RIVM) |
| EPI_ISL_454785 | Dutch COVID-19 response team | National Institute for Public Health and the Environment (RIVM) |
| EPI_ISL_454786 | Dutch COVID-19 response team | National Institute for Public Health and the Environment (RIVM) |
| EPI_ISL_454787 | Dutch COVID-19 response team | National Institute for Public Health and the Environment (RIVM) |
| EPI_ISL_454788 | Dutch COVID-19 response team | National Institute for Public Health and the Environment (RIVM) |
| EPI_ISL_454789 | Dutch COVID-19 response team | National Institute for Public Health and the Environment (RIVM) |
| EPI_ISL_454790 | Dutch COVID-19 response team | National Institute for Public Health and the Environment (RIVM) |

| EPI_Accession_ID | Originating Laboratory | Submitting Laboratory |
|---|---|---|
| EPI_ISL_454791 | Dutch COVID-19 response team | National Institute for Public Health and the Environment (RIVM) |
| EPI_ISL_454792 | Dutch COVID-19 response team | National Institute for Public Health and the Environment (RIVM) |
| EPI_ISL_454793 | Dutch COVID-19 response team | National Institute for Public Health and the Environment (RIVM) |
| EPI_ISL_454794 | Dutch COVID-19 response team | National Institute for Public Health and the Environment (RIVM) |
| EPI_ISL_454795 | Veterinary Specialized Institute Kraljevo | Veterinary Specialized Institute Kraljevo |
| EPI_ISL_454830 | SMS Medical College, Jaipur | CSIR Institute of Genomics and Integrative Biology |
| EPI_ISL_454831 | SMS Medical College, Jaipur | CSIR Institute of Genomics and Integrative Biology |
| EPI_ISL_454832 | SMS Medical College, Jaipur | CSIR Institute of Genomics and Integrative Biology |
| EPI_ISL_454833 | SMS Medical College, Jaipur | CSIR Institute of Genomics and Integrative Biology |
| EPI_ISL_454858 | Translational Health Science and Technology Institute -ESIC medical college and hospital, Faridabad | THSTI Bioassay laboratory |
| EPI_ISL_454859 | Translational Health Science and Technology Institute -ESIC medical college and hospital, Faridabad | THSTI Bioassay laboratory |
| EPI_ISL_454860 | Translational Health Science and Technology Institute -ESIC medical college and hospital, Faridabad | THSTI Bioassay laboratory |
| EPI_ISL_454861 | Translational Health Science and Technology Institute -ESIC medical college and hospital, Faridabad | THSTI Bioassay laboratory |
| EPI_ISL_454862 | Translational Health Science and Technology Institute -ESIC medical college and hospital, Faridabad | THSTI Bioassay laboratory |
| EPI_ISL_454863 | Translational Health Science and Technology Institute -ESIC medical college and hospital, Faridabad | THSTI Bioassay laboratory |
| EPI_ISL_454864 | Translational Health Science and Technology Institute -ESIC medical college and hospital, Faridabad | THSTI Bioassay laboratory |
| EPI_ISL_454865 | Translational Health Science and Technology Institute -ESIC medical college and hospital, Faridabad | THSTI Bioassay laboratory |
| EPI_ISL_454866 | Translational Health Science and Technology Institute -ESIC medical college and hospital, Faridabad | THSTI Bioassay laboratory |
| EPI_ISL_454867 | Translational Health Science and Technology Institute -ESIC medical college and hospital, Faridabad | THSTI Bioassay laboratory |

| EPI_Accession_ID | Originating Laboratory | Submitting Laboratory |
|---|---|---|
| EPI_ISL_454868 | Karolinska Universitetslaboratoriet | The Public Health Agency of Sweden |
| EPI_ISL_454869 | Karolinska Universitetslaboratoriet | The Public Health Agency of Sweden |
| EPI_ISL_454870 | Karolinska Universitetslaboratoriet | The Public Health Agency of Sweden |
| EPI_ISL_454871 | Karolinska Universitetslaboratoriet | The Public Health Agency of Sweden |
| EPI_ISL_454872 | Karolinska Universitetslaboratoriet | The Public Health Agency of Sweden |
| EPI_ISL_454873 | Karolinska Universitetslaboratoriet | The Public Health Agency of Sweden |
| EPI_ISL_454874 | Karolinska Universitetslaboratoriet | The Public Health Agency of Sweden |
| EPI_ISL_454875 | Karolinska Universitetslaboratoriet | The Public Health Agency of Sweden |
| EPI_ISL_454876 | Karolinska Universitetslaboratoriet | The Public Health Agency of Sweden |
| EPI_ISL_454877 | Karolinska Universitetslaboratoriet | The Public Health Agency of Sweden |
| EPI_ISL_454878 | Karolinska Universitetslaboratoriet | The Public Health Agency of Sweden |
| EPI_ISL_454879 | Karolinska Universitetslaboratoriet | The Public Health Agency of Sweden |
| EPI_ISL_454880 | Karolinska Universitetslaboratoriet | The Public Health Agency of Sweden |
| EPI_ISL_454881 | Karolinska Universitetslaboratoriet | The Public Health Agency of Sweden |
| EPI_ISL_454882 | Karolinska Universitetslaboratoriet | The Public Health Agency of Sweden |
| EPI_ISL_454883 | Karolinska Universitetslaboratoriet | The Public Health Agency of Sweden |
| EPI_ISL_454884 | Karolinska Universitetslaboratoriet | The Public Health Agency of Sweden |
| EPI_ISL_454885 | Karolinska Universitetslaboratoriet | The Public Health Agency of Sweden |
| EPI_ISL_454886 | Karolinska Universitetslaboratoriet | The Public Health Agency of Sweden |
| EPI_ISL_454887 | Karolinska Universitetslaboratoriet | The Public Health Agency of Sweden |
| EPI_ISL_454888 | Karolinska Universitetslaboratoriet | The Public Health Agency of Sweden |
| EPI_ISL_454889 | Karolinska Universitetslaboratoriet | The Public Health Agency of Sweden |
| EPI_ISL_454890 | Karolinska Universitetslaboratoriet | The Public Health Agency of Sweden |
| EPI_ISL_454891 | Karolinska Universitetslaboratoriet | The Public Health Agency of Sweden |
| EPI_ISL_454892 | Karolinska Universitetslaboratoriet | The Public Health Agency of Sweden |
| EPI_ISL_454893 | Karolinska Universitetslaboratoriet | The Public Health Agency of Sweden |
| EPI_ISL_454894 | Karolinska Universitetslaboratoriet | The Public Health Agency of Sweden |
| EPI_ISL_454895 | Karolinska Universitetslaboratoriet | The Public Health Agency of Sweden |
| EPI_ISL_454896 | Karolinska Universitetslaboratoriet | The Public Health Agency of Sweden |
| EPI_ISL_454897 | Karolinska Universitetslaboratoriet | The Public Health Agency of Sweden |
| EPI_ISL_454898 | Karolinska Universitetslaboratoriet | The Public Health Agency of Sweden |
| EPI_ISL_454899 | Karolinska Universitetslaboratoriet | The Public Health Agency of Sweden |
| EPI_ISL_454900 | Karolinska Universitetslaboratoriet | The Public Health Agency of Sweden |

| EPI_Accession_ID | Originating Laboratory | Submitting Laboratory |
|---|---|---|
| EPI_ISL_454901 | Karolinska Universitetslaboratoriet | The Public Health Agency of Sweden |
| EPI_ISL_454902 | Karolinska Universitetslaboratoriet | The Public Health Agency of Sweden |
| EPI_ISL_454903 | Klinisk mikrobiologi, UAS | The Public Health Agency of Sweden |
| EPI_ISL_454904 | Wuhan Chain Medical Labs (CMLabs) | State Key Laboratory of Biotherapy of Sichuan University |
| EPI_ISL_454905 | Wuhan Chain Medical Labs (CMLabs) | State Key Laboratory of Biotherapy of Sichuan University |
| EPI_ISL_454906 | Wuhan Chain Medical Labs (CMLabs) | State Key Laboratory of Biotherapy of Sichuan University |
| EPI_ISL_454907 | Wuhan Chain Medical Labs (CMLabs) | State Key Laboratory of Biotherapy of Sichuan University |
| EPI_ISL_454908 | Wuhan Chain Medical Labs (CMLabs) | State Key Laboratory of Biotherapy of Sichuan University |
| EPI_ISL_454909 | Wuhan Chain Medical Labs (CMLabs) | State Key Laboratory of Biotherapy of Sichuan University |
| EPI_ISL_454910 | Wuhan Chain Medical Labs (CMLabs) | State Key Laboratory of Biotherapy of Sichuan University |
| EPI_ISL_454911 | Wuhan Chain Medical Labs (CMLabs) | State Key Laboratory of Biotherapy of Sichuan University |
| EPI_ISL_454912 | Wuhan Chain Medical Labs (CMLabs) | State Key Laboratory of Biotherapy of Sichuan University |
| EPI_ISL_454913 | Wuhan Chain Medical Labs (CMLabs) | State Key Laboratory of Biotherapy of Sichuan University |
| EPI_ISL_454914 | Wuhan Chain Medical Labs (CMLabs) | State Key Laboratory of Biotherapy of Sichuan University |
| EPI_ISL_454915 | Wuhan Chain Medical Labs (CMLabs) | State Key Laboratory of Biotherapy of Sichuan University |
| EPI_ISL_454916 | Wuhan Chain Medical Labs (CMLabs) | State Key Laboratory of Biotherapy of Sichuan University |
| EPI_ISL_454917 | Wuhan Chain Medical Labs (CMLabs) | State Key Laboratory of Biotherapy of Sichuan University |
| EPI_ISL_454918 | Wuhan Chain Medical Labs (CMLabs) | State Key Laboratory of Biotherapy of Sichuan University |

| EPI_Accession_ID | Originating Laboratory | Submitting Laboratory |
|---|---|---|
| EPI_ISL_454919 | Wuhan Chain Medical Labs (CMLabs) | State Key Laboratory of Biotherapy of Sichuan University |
| EPI_ISL_454920 | Wuhan Chain Medical Labs (CMLabs) | State Key Laboratory of Biotherapy of Sichuan University |
| EPI_ISL_454921 | Wuhan Chain Medical Labs (CMLabs) | State Key Laboratory of Biotherapy of Sichuan University |
| EPI_ISL_454922 | Wuhan Chain Medical Labs (CMLabs) | State Key Laboratory of Biotherapy of Sichuan University |
| EPI_ISL_454923 | Wuhan Chain Medical Labs (CMLabs) | State Key Laboratory of Biotherapy of Sichuan University |
| EPI_ISL_454924 | Wuhan Chain Medical Labs (CMLabs) | State Key Laboratory of Biotherapy of Sichuan University |
| EPI_ISL_454925 | Wuhan Chain Medical Labs (CMLabs) | State Key Laboratory of Biotherapy of Sichuan University |
| EPI_ISL_454926 | Wuhan Chain Medical Labs (CMLabs) | State Key Laboratory of Biotherapy of Sichuan University |
| EPI_ISL_454927 | Wuhan Chain Medical Labs (CMLabs) | State Key Laboratory of Biotherapy of Sichuan University |
| EPI_ISL_454928 | Wuhan Chain Medical Labs (CMLabs) | State Key Laboratory of Biotherapy of Sichuan University |
| EPI_ISL_454929 | Wuhan Chain Medical Labs (CMLabs) | State Key Laboratory of Biotherapy of Sichuan University |
| EPI_ISL_454930 | Wuhan Chain Medical Labs (CMLabs) | State Key Laboratory of Biotherapy of Sichuan University |
| EPI_ISL_454931 | Wuhan Chain Medical Labs (CMLabs) | State Key Laboratory of Biotherapy of Sichuan University |
| EPI_ISL_454932 | Wuhan Chain Medical Labs (CMLabs) | State Key Laboratory of Biotherapy of Sichuan University |
| EPI_ISL_454933 | Wuhan Chain Medical Labs (CMLabs) | State Key Laboratory of Biotherapy of Sichuan University |
| EPI_ISL_454934 | Wuhan Chain Medical Labs (CMLabs) | State Key Laboratory of Biotherapy of Sichuan University |

| EPI_Accession_ID | Originating Laboratory | Submitting Laboratory |
|---|---|---|
| EPI_ISL_454935 | Wuhan Chain Medical Labs (CMLabs) | State Key Laboratory of Biotherapy of Sichuan University |
| EPI_ISL_454936 | Wuhan Chain Medical Labs (CMLabs) | State Key Laboratory of Biotherapy of Sichuan University |
| EPI_ISL_454937 | Wuhan Chain Medical Labs (CMLabs) | State Key Laboratory of Biotherapy of Sichuan University |
| EPI_ISL_454938 | Wuhan Chain Medical Labs (CMLabs) | State Key Laboratory of Biotherapy of Sichuan University |
| EPI_ISL_454939 | Wuhan Chain Medical Labs (CMLabs) | State Key Laboratory of Biotherapy of Sichuan University |
| EPI_ISL_454940 | Wuhan Chain Medical Labs (CMLabs) | State Key Laboratory of Biotherapy of Sichuan University |
| EPI_ISL_454941 | Wuhan Chain Medical Labs (CMLabs) | State Key Laboratory of Biotherapy of Sichuan University |
| EPI_ISL_454942 | Wuhan Chain Medical Labs (CMLabs) | State Key Laboratory of Biotherapy of Sichuan University |
| EPI_ISL_454943 | Wuhan Chain Medical Labs (CMLabs) | State Key Laboratory of Biotherapy of Sichuan University |
| EPI_ISL_454944 | Wuhan Chain Medical Labs (CMLabs) | State Key Laboratory of Biotherapy of Sichuan University |
| EPI_ISL_454945 | Wuhan Chain Medical Labs (CMLabs) | State Key Laboratory of Biotherapy of Sichuan University |
| EPI_ISL_454946 | Wuhan Chain Medical Labs (CMLabs) | State Key Laboratory of Biotherapy of Sichuan University |
| EPI_ISL_454947 | Wuhan Chain Medical Labs (CMLabs) | State Key Laboratory of Biotherapy of Sichuan University |
| EPI_ISL_454948 | Wuhan Chain Medical Labs (CMLabs) | State Key Laboratory of Biotherapy of Sichuan University |
| EPI_ISL_454949 | Wuhan Chain Medical Labs (CMLabs) | State Key Laboratory of Biotherapy of Sichuan University |
| EPI_ISL_454950 | Wuhan Chain Medical Labs (CMLabs) | State Key Laboratory of Biotherapy of Sichuan University |

| EPI_Accession_ID | Originating Laboratory | Submitting Laboratory |
| --- | --- | --- |
| EPI_ISL_454951 | Wuhan Chain Medical Labs (CMLabs) | State Key Laboratory of Biotherapy of Sichuan University |
| EPI_ISL_454952 | Wuhan Chain Medical Labs (CMLabs) | State Key Laboratory of Biotherapy of Sichuan University |
| EPI_ISL_454953 | Wuhan Chain Medical Labs (CMLabs) | State Key Laboratory of Biotherapy of Sichuan University |
| EPI_ISL_454954 | Wuhan Chain Medical Labs (CMLabs) | State Key Laboratory of Biotherapy of Sichuan University |
| EPI_ISL_454955 | Wuhan Chain Medical Labs (CMLabs) | State Key Laboratory of Biotherapy of Sichuan University |
| EPI_ISL_454956 | Wuhan Chain Medical Labs (CMLabs) | State Key Laboratory of Biotherapy of Sichuan University |
| EPI_ISL_454957 | Wuhan Chain Medical Labs (CMLabs) | State Key Laboratory of Biotherapy of Sichuan University |
| EPI_ISL_454958 | Wuhan Chain Medical Labs (CMLabs) | State Key Laboratory of Biotherapy of Sichuan University |
| EPI_ISL_454959 | Wuhan Chain Medical Labs (CMLabs) | State Key Laboratory of Biotherapy of Sichuan University |
| EPI_ISL_454960 | Wuhan Chain Medical Labs (CMLabs) | State Key Laboratory of Biotherapy of Sichuan University |
| EPI_ISL_454961 | Wuhan Chain Medical Labs (CMLabs) | State Key Laboratory of Biotherapy of Sichuan University |
| EPI_ISL_454962 | Wuhan Chain Medical Labs (CMLabs) | State Key Laboratory of Biotherapy of Sichuan University |
| EPI_ISL_454963 | Wuhan Chain Medical Labs (CMLabs) | State Key Laboratory of Biotherapy of Sichuan University |
| EPI_ISL_454964 | Wuhan Chain Medical Labs (CMLabs) | State Key Laboratory of Biotherapy of Sichuan University |
| EPI_ISL_454965 | Wuhan Chain Medical Labs (CMLabs) | State Key Laboratory of Biotherapy of Sichuan University |
| EPI_ISL_454966 | Wuhan Chain Medical Labs (CMLabs) | State Key Laboratory of Biotherapy of Sichuan University |

| EPI_Accession_ID | Originating Laboratory | Submitting Laboratory |
|---|---|---|
| EPI_ISL_454967 | Wuhan Chain Medical Labs (CMLabs) | State Key Laboratory of Biotherapy of Sichuan University |
| EPI_ISL_454968 | Wuhan Chain Medical Labs (CMLabs) | State Key Laboratory of Biotherapy of Sichuan University |
| EPI_ISL_454969 | Wuhan Chain Medical Labs (CMLabs) | State Key Laboratory of Biotherapy of Sichuan University |
| EPI_ISL_454970 | Wuhan Chain Medical Labs (CMLabs) | State Key Laboratory of Biotherapy of Sichuan University |
| EPI_ISL_454971 | Wuhan Chain Medical Labs (CMLabs) | State Key Laboratory of Biotherapy of Sichuan University |
| EPI_ISL_454972 | Wuhan Chain Medical Labs (CMLabs) | State Key Laboratory of Biotherapy of Sichuan University |
| EPI_ISL_454973 | Wuhan Chain Medical Labs (CMLabs) | State Key Laboratory of Biotherapy of Sichuan University |
| EPI_ISL_454974 | Wuhan Chain Medical Labs (CMLabs) | State Key Laboratory of Biotherapy of Sichuan University |
| EPI_ISL_454975 | Wuhan Chain Medical Labs (CMLabs) | State Key Laboratory of Biotherapy of Sichuan University |
| EPI_ISL_454976 | Wuhan Chain Medical Labs (CMLabs) | State Key Laboratory of Biotherapy of Sichuan University |
| EPI_ISL_454977 | Wuhan Chain Medical Labs (CMLabs) | State Key Laboratory of Biotherapy of Sichuan University |
| EPI_ISL_454978 | Wuhan Chain Medical Labs (CMLabs) | State Key Laboratory of Biotherapy of Sichuan University |
| EPI_ISL_454979 | Wuhan Chain Medical Labs (CMLabs) | State Key Laboratory of Biotherapy of Sichuan University |
| EPI_ISL_454980 | Wuhan Chain Medical Labs (CMLabs) | State Key Laboratory of Biotherapy of Sichuan University |
| EPI_ISL_454981 | Wuhan Chain Medical Labs (CMLabs) | State Key Laboratory of Biotherapy of Sichuan University |
| EPI_ISL_454982 | Wuhan Chain Medical Labs (CMLabs) | State Key Laboratory of Biotherapy of Sichuan University |

| EPI_Accession_ID | Originating Laboratory | Submitting Laboratory |
|---|---|---|
| EPI_ISL_454983 | Wuhan Chain Medical Labs (CMLabs) | State Key Laboratory of Biotherapy of Sichuan University |
| EPI_ISL_454984 | Wuhan Chain Medical Labs (CMLabs) | State Key Laboratory of Biotherapy of Sichuan University |
| EPI_ISL_454985 | Wuhan Chain Medical Labs (CMLabs) | State Key Laboratory of Biotherapy of Sichuan University |
| EPI_ISL_454986 | Wuhan Chain Medical Labs (CMLabs) | State Key Laboratory of Biotherapy of Sichuan University |
| EPI_ISL_454987 | Wuhan Chain Medical Labs (CMLabs) | State Key Laboratory of Biotherapy of Sichuan University |
| EPI_ISL_454988 | Wuhan Chain Medical Labs (CMLabs) | State Key Laboratory of Biotherapy of Sichuan University |
| EPI_ISL_454989 | Wuhan Chain Medical Labs (CMLabs) | State Key Laboratory of Biotherapy of Sichuan University |
| EPI_ISL_454990 | Wuhan Chain Medical Labs (CMLabs) | State Key Laboratory of Biotherapy of Sichuan University |
| EPI_ISL_454991 | Wuhan Chain Medical Labs (CMLabs) | State Key Laboratory of Biotherapy of Sichuan University |
| EPI_ISL_454992 | Wuhan Chain Medical Labs (CMLabs) | State Key Laboratory of Biotherapy of Sichuan University |
| EPI_ISL_454993 | Wuhan Chain Medical Labs (CMLabs) | State Key Laboratory of Biotherapy of Sichuan University |
| EPI_ISL_454994 | Wuhan Chain Medical Labs (CMLabs) | State Key Laboratory of Biotherapy of Sichuan University |
| EPI_ISL_454995 | Wuhan Chain Medical Labs (CMLabs) | State Key Laboratory of Biotherapy of Sichuan University |
| EPI_ISL_454996 | Wuhan Chain Medical Labs (CMLabs) | State Key Laboratory of Biotherapy of Sichuan University |
| EPI_ISL_454997 | Wuhan Chain Medical Labs (CMLabs) | State Key Laboratory of Biotherapy of Sichuan University |
| EPI_ISL_454998 | Wuhan Chain Medical Labs (CMLabs) | State Key Laboratory of Biotherapy of Sichuan University |

| EPI_Accession_ID | Originating Laboratory | Submitting Laboratory |
|---|---|---|
| EPI_ISL_454999 | Wuhan Chain Medical Labs (CMLabs) | State Key Laboratory of Biotherapy of Sichuan University |
| EPI_ISL_455000 | Wuhan Chain Medical Labs (CMLabs) | State Key Laboratory of Biotherapy of Sichuan University |
| EPI_ISL_455001 | Wuhan Chain Medical Labs (CMLabs) | State Key Laboratory of Biotherapy of Sichuan University |
| EPI_ISL_455002 | Wuhan Chain Medical Labs (CMLabs) | State Key Laboratory of Biotherapy of Sichuan University |
| EPI_ISL_455003 | Wuhan Chain Medical Labs (CMLabs) | State Key Laboratory of Biotherapy of Sichuan University |
| EPI_ISL_455004 | Wuhan Chain Medical Labs (CMLabs) | State Key Laboratory of Biotherapy of Sichuan University |
| EPI_ISL_455005 | Wuhan Chain Medical Labs (CMLabs) | State Key Laboratory of Biotherapy of Sichuan University |
| EPI_ISL_455006 | Wuhan Chain Medical Labs (CMLabs) | State Key Laboratory of Biotherapy of Sichuan University |
| EPI_ISL_455007 | Wuhan Chain Medical Labs (CMLabs) | State Key Laboratory of Biotherapy of Sichuan University |
| EPI_ISL_455008 | Wuhan Chain Medical Labs (CMLabs) | State Key Laboratory of Biotherapy of Sichuan University |
| EPI_ISL_455009 | Wuhan Chain Medical Labs (CMLabs) | State Key Laboratory of Biotherapy of Sichuan University |
| EPI_ISL_455010 | Wuhan Chain Medical Labs (CMLabs) | State Key Laboratory of Biotherapy of Sichuan University |
| EPI_ISL_455011 | Wuhan Chain Medical Labs (CMLabs) | State Key Laboratory of Biotherapy of Sichuan University |
| EPI_ISL_455012 | Wuhan Chain Medical Labs (CMLabs) | State Key Laboratory of Biotherapy of Sichuan University |
| EPI_ISL_455013 | Wuhan Chain Medical Labs (CMLabs) | State Key Laboratory of Biotherapy of Sichuan University |
| EPI_ISL_455014 | Wuhan Chain Medical Labs (CMLabs) | State Key Laboratory of Biotherapy of Sichuan University |

| EPI_Accession_ID | Originating Laboratory | Submitting Laboratory |
|---|---|---|
| EPI_ISL_455015 | Pandit Deendayal Upadhyay Government Medical College, Rajkot | Gujarat Biotechnology Research Centre |
| EPI_ISL_455016 | Pandit Deendayal Upadhyay Government Medical College, Rajkot | Gujarat Biotechnology Research Centre |
| EPI_ISL_455017 | Government Medical College, Vadodara | Gujarat Biotechnology Research Centre |
| EPI_ISL_455018 | Government Medical College, Vadodara | Gujarat Biotechnology Research Centre |
| EPI_ISL_455019 | Government Medical College, Vadodara | Gujarat Biotechnology Research Centre |
| EPI_ISL_455020 | Government Medical College, Vadodara | Gujarat Biotechnology Research Centre |
| EPI_ISL_455021 | Government Medical College, Vadodara | Gujarat Biotechnology Research Centre |
| EPI_ISL_455022 | Government Medical College, Vadodara | Gujarat Biotechnology Research Centre |
| EPI_ISL_455023 | Government Medical College, Vadodara | Gujarat Biotechnology Research Centre |
| EPI_ISL_455024 | Government Medical College, Vadodara | Gujarat Biotechnology Research Centre |
| EPI_ISL_455025 | Government Medical College, Vadodara | Gujarat Biotechnology Research Centre |
| EPI_ISL_455026 | Government Medical College, Vadodara | Gujarat Biotechnology Research Centre |
| EPI_ISL_455027 | Government Medical College, Vadodara | Gujarat Biotechnology Research Centre |
| EPI_ISL_455028 | Pathology West - NSW Health Pathology | NSW Health Pathology - Institute of Clinical Pathology and Medical Research; Westmead Hospital; University of Sydney |
| EPI_ISL_455029 | Pathology West - NSW Health Pathology | NSW Health Pathology - Institute of Clinical Pathology and Medical Research; Westmead Hospital; University of Sydney |
| EPI_ISL_455030 | Pathology West - NSW Health Pathology | NSW Health Pathology - Institute of Clinical Pathology and Medical Research; Westmead Hospital; University of Sydney |
| EPI_ISL_455031 | Pathology West - NSW Health Pathology | NSW Health Pathology - Institute of Clinical Pathology and Medical Research; Westmead Hospital; University of Sydney |
| EPI_ISL_455032 | Pathology West - NSW Health Pathology | NSW Health Pathology - Institute of Clinical Pathology and Medical Research; Westmead Hospital; University of Sydney |
| EPI_ISL_455033 | Pathology West - NSW Health Pathology | NSW Health Pathology - Institute of Clinical Pathology and Medical Research; Westmead Hospital; University of Sydney |

| EPI_Accession_ID | Originating Laboratory | Submitting Laboratory |
|---|---|---|
| EPI_ISL_455034 | Pathology West - NSW Health Pathology | NSW Health Pathology - Institute of Clinical Pathology and Medical Research; Westmead Hospital; University of Sydney |
| EPI_ISL_455035 | Pathology West - NSW Health Pathology | NSW Health Pathology - Institute of Clinical Pathology and Medical Research; Westmead Hospital; University of Sydney |
| EPI_ISL_455036 | South Eastern Area Laboratory Services | NSW Health Pathology - Institute of Clinical Pathology and Medical Research; Westmead Hospital; University of Sydney |
| EPI_ISL_455037 | Pathology Sydney South West - NSW Health Pathology | NSW Health Pathology - Institute of Clinical Pathology and Medical Research; Westmead Hospital; University of Sydney |
| EPI_ISL_455038 | Pathology West - NSW Health Pathology | NSW Health Pathology - Institute of Clinical Pathology and Medical Research; Westmead Hospital; University of Sydney |
| EPI_ISL_455039 | Pathology West - NSW Health Pathology | NSW Health Pathology - Institute of Clinical Pathology and Medical Research; Westmead Hospital; University of Sydney |
| EPI_ISL_455040 | Pathology West - NSW Health Pathology | NSW Health Pathology - Institute of Clinical Pathology and Medical Research; Westmead Hospital; University of Sydney |
| EPI_ISL_455041 | Laverty Pathology | NSW Health Pathology - Institute of Clinical Pathology and Medical Research; Westmead Hospital; University of Sydney |
| EPI_ISL_455042 | ACT Pathology | NSW Health Pathology - Institute of Clinical Pathology and Medical Research; Westmead Hospital; University of Sydney |
| EPI_ISL_455043 | ACT Pathology | NSW Health Pathology - Institute of Clinical Pathology and Medical Research; Westmead Hospital; University of Sydney |
| EPI_ISL_455044 | Pathology West - NSW Health Pathology | NSW Health Pathology - Institute of Clinical Pathology and Medical Research; Westmead Hospital; University of Sydney |

| EPI_Accession_ID | Originating Laboratory | Submitting Laboratory |
|---|---|---|
| EPI_ISL_455045 | Pathology West - NSW Health Pathology | NSW Health Pathology - Institute of Clinical Pathology and Medical Research; Westmead Hospital; University of Sydney |
| EPI_ISL_455046 | Pathology West - NSW Health Pathology | NSW Health Pathology - Institute of Clinical Pathology and Medical Research; Westmead Hospital; University of Sydney |
| EPI_ISL_455047 | Pathology West - NSW Health Pathology | NSW Health Pathology - Institute of Clinical Pathology and Medical Research; Westmead Hospital; University of Sydney |
| EPI_ISL_455048 | Laverty Pathology | NSW Health Pathology - Institute of Clinical Pathology and Medical Research; Westmead Hospital; University of Sydney |
| EPI_ISL_455049 | Pathology West - NSW Health Pathology | NSW Health Pathology - Institute of Clinical Pathology and Medical Research; Westmead Hospital; University of Sydney |
| EPI_ISL_455050 | ACT Pathology | NSW Health Pathology - Institute of Clinical Pathology and Medical Research; Westmead Hospital; University of Sydney |
| EPI_ISL_455051 | Douglas Hanly Moir Pathology | NSW Health Pathology - Institute of Clinical Pathology and Medical Research; Westmead Hospital; University of Sydney |
| EPI_ISL_455052 | Douglas Hanly Moir Pathology | NSW Health Pathology - Institute of Clinical Pathology and Medical Research; Westmead Hospital; University of Sydney |
| EPI_ISL_455053 | Douglas Hanly Moir Pathology | NSW Health Pathology - Institute of Clinical Pathology and Medical Research; Westmead Hospital; University of Sydney |
| EPI_ISL_455054 | South Eastern Area Laboratory Services | NSW Health Pathology - Institute of Clinical Pathology and Medical Research; Westmead Hospital; University of Sydney |
| EPI_ISL_455055 | Pathology West - NSW Health Pathology | NSW Health Pathology - Institute of Clinical Pathology and Medical Research; Westmead Hospital; University of Sydney |

| EPI_Accession_ID | Originating Laboratory | Submitting Laboratory |
|---|---|---|
| EPI_ISL_455056 | Pathology West - NSW Health Pathology | NSW Health Pathology - Institute of Clinical Pathology and Medical Research; Westmead Hospital; University of Sydney |
| EPI_ISL_455057 | Pathology West - NSW Health Pathology | NSW Health Pathology - Institute of Clinical Pathology and Medical Research; Westmead Hospital; University of Sydney |
| EPI_ISL_455058 | Pathology West - NSW Health Pathology | NSW Health Pathology - Institute of Clinical Pathology and Medical Research; Westmead Hospital; University of Sydney |
| EPI_ISL_455059 | Pathology West - NSW Health Pathology | NSW Health Pathology - Institute of Clinical Pathology and Medical Research; Westmead Hospital; University of Sydney |
| EPI_ISL_455060 | Pathology West - NSW Health Pathology | NSW Health Pathology - Institute of Clinical Pathology and Medical Research; Westmead Hospital; University of Sydney |
| EPI_ISL_455061 | Pathology West - NSW Health Pathology | NSW Health Pathology - Institute of Clinical Pathology and Medical Research; Westmead Hospital; University of Sydney |
| EPI_ISL_455062 | Pathology West - NSW Health Pathology | NSW Health Pathology - Institute of Clinical Pathology and Medical Research; Westmead Hospital; University of Sydney |
| EPI_ISL_455063 | Laverty Pathology | NSW Health Pathology - Institute of Clinical Pathology and Medical Research; Westmead Hospital; University of Sydney |
| EPI_ISL_455064 | Pathology West - NSW Health Pathology | NSW Health Pathology - Institute of Clinical Pathology and Medical Research; Westmead Hospital; University of Sydney |
| EPI_ISL_455065 | Douglas Hanly Moir Pathology | NSW Health Pathology - Institute of Clinical Pathology and Medical Research; Westmead Hospital; University of Sydney |
| EPI_ISL_455066 | Pathology West - NSW Health Pathology | NSW Health Pathology - Institute of Clinical Pathology and Medical Research; Westmead Hospital; University of Sydney |

| EPI_Accession_ID | Originating Laboratory | Submitting Laboratory |
|---|---|---|
| EPI_ISL_455067 | Pathology West - NSW Health Pathology | NSW Health Pathology - Institute of Clinical Pathology and Medical Research; Westmead Hospital; University of Sydney |
| EPI_ISL_455068 | Childrens Hospital Westmead | NSW Health Pathology - Institute of Clinical Pathology and Medical Research; Westmead Hospital; University of Sydney |
| EPI_ISL_455069 | Pathology West - NSW Health Pathology | NSW Health Pathology - Institute of Clinical Pathology and Medical Research; Westmead Hospital; University of Sydney |
| EPI_ISL_455070 | Pathology West - NSW Health Pathology | NSW Health Pathology - Institute of Clinical Pathology and Medical Research; Westmead Hospital; University of Sydney |
| EPI_ISL_455071 | Pathology Sydney South West - NSW Health Pathology | NSW Health Pathology - Institute of Clinical Pathology and Medical Research; Westmead Hospital; University of Sydney |
| EPI_ISL_455072 | Sullivan Nicolaides Pathology | NSW Health Pathology - Institute of Clinical Pathology and Medical Research; Westmead Hospital; University of Sydney |
| EPI_ISL_455073 | Sullivan Nicolaides Pathology | NSW Health Pathology - Institute of Clinical Pathology and Medical Research; Westmead Hospital; University of Sydney |
| EPI_ISL_455074 | ACT Pathology | NSW Health Pathology - Institute of Clinical Pathology and Medical Research; Westmead Hospital; University of Sydney |
| EPI_ISL_455075 | South Eastern Area Laboratory Services | NSW Health Pathology - Institute of Clinical Pathology and Medical Research; Westmead Hospital; University of Sydney |
| EPI_ISL_455076 | South Eastern Area Laboratory Services | NSW Health Pathology - Institute of Clinical Pathology and Medical Research; Westmead Hospital; University of Sydney |
| EPI_ISL_455077 | South Eastern Area Laboratory Services | NSW Health Pathology - Institute of Clinical Pathology and Medical Research; Westmead Hospital; University of Sydney |

| EPI_Accession_ID | Originating Laboratory | Submitting Laboratory |
|---|---|---|
| EPI_ISL_455078 | South Eastern Area Laboratory Services | NSW Health Pathology - Institute of Clinical Pathology and Medical Research; Westmead Hospital; University of Sydney |
| EPI_ISL_455079 | South Eastern Area Laboratory Services | NSW Health Pathology - Institute of Clinical Pathology and Medical Research; Westmead Hospital; University of Sydney |
| EPI_ISL_455080 | South Eastern Area Laboratory Services | NSW Health Pathology - Institute of Clinical Pathology and Medical Research; Westmead Hospital; University of Sydney |
| EPI_ISL_455081 | South Eastern Area Laboratory Services | NSW Health Pathology - Institute of Clinical Pathology and Medical Research; Westmead Hospital; University of Sydney |
| EPI_ISL_455082 | South Eastern Area Laboratory Services | NSW Health Pathology - Institute of Clinical Pathology and Medical Research; Westmead Hospital; University of Sydney |
| EPI_ISL_455083 | South Eastern Area Laboratory Services | NSW Health Pathology - Institute of Clinical Pathology and Medical Research; Westmead Hospital; University of Sydney |
| EPI_ISL_455084 | South Eastern Area Laboratory Services | NSW Health Pathology - Institute of Clinical Pathology and Medical Research; Westmead Hospital; University of Sydney |
| EPI_ISL_455085 | South Eastern Area Laboratory Services | NSW Health Pathology - Institute of Clinical Pathology and Medical Research; Westmead Hospital; University of Sydney |
| EPI_ISL_455086 | South Eastern Area Laboratory Services | NSW Health Pathology - Institute of Clinical Pathology and Medical Research; Westmead Hospital; University of Sydney |
| EPI_ISL_455087 | South Eastern Area Laboratory Services | NSW Health Pathology - Institute of Clinical Pathology and Medical Research; Westmead Hospital; University of Sydney |
| EPI_ISL_455088 | South Eastern Area Laboratory Services | NSW Health Pathology - Institute of Clinical Pathology and Medical Research; Westmead Hospital; University of Sydney |

| EPI_Accession_ID | Originating Laboratory | Submitting Laboratory |
|---|---|---|
| EPI_ISL_455089 | South Eastern Area Laboratory Services | NSW Health Pathology - Institute of Clinical Pathology and Medical Research; Westmead Hospital; University of Sydney |
| EPI_ISL_455090 | South Eastern Area Laboratory Services | NSW Health Pathology - Institute of Clinical Pathology and Medical Research; Westmead Hospital; University of Sydney |
| EPI_ISL_455091 | South Eastern Area Laboratory Services | NSW Health Pathology - Institute of Clinical Pathology and Medical Research; Westmead Hospital; University of Sydney |
| EPI_ISL_455092 | South Eastern Area Laboratory Services | NSW Health Pathology - Institute of Clinical Pathology and Medical Research; Westmead Hospital; University of Sydney |
| EPI_ISL_455093 | South Eastern Area Laboratory Services | NSW Health Pathology - Institute of Clinical Pathology and Medical Research; Westmead Hospital; University of Sydney |
| EPI_ISL_455094 | Pathology Sydney South West - NSW Health Pathology | NSW Health Pathology - Institute of Clinical Pathology and Medical Research; Westmead Hospital; University of Sydney |
| EPI_ISL_455095 | Pathology West - NSW Health Pathology | NSW Health Pathology - Institute of Clinical Pathology and Medical Research; Westmead Hospital; University of Sydney |
| EPI_ISL_455096 | South Eastern Area Laboratory Services | NSW Health Pathology - Institute of Clinical Pathology and Medical Research; Westmead Hospital; University of Sydney |
| EPI_ISL_455097 | South Eastern Area Laboratory Services | NSW Health Pathology - Institute of Clinical Pathology and Medical Research; Westmead Hospital; University of Sydney |
| EPI_ISL_455098 | South Eastern Area Laboratory Services | NSW Health Pathology - Institute of Clinical Pathology and Medical Research; Westmead Hospital; University of Sydney |
| EPI_ISL_455099 | South Eastern Area Laboratory Services | NSW Health Pathology - Institute of Clinical Pathology and Medical Research; Westmead Hospital; University of Sydney |

| EPI_Accession_ID | Originating Laboratory | Submitting Laboratory |
|---|---|---|
| EPI_ISL_455100 | Orsa VC | The Public Health Agency of Sweden |
| EPI_ISL_455101 | Jourcentralen | The Public Health Agency of Sweden |
| EPI_ISL_455102 | Kungsors VC | The Public Health Agency of Sweden |
| EPI_ISL_455103 | Huslakarna Varmbadhuset Varberg | The Public Health Agency of Sweden |
| EPI_ISL_455104 | Ulltuna Vardcentral | The Public Health Agency of Sweden |
| EPI_ISL_455105 | Wetterhalsan | The Public Health Agency of Sweden |
| EPI_ISL_455106 | Surbrunns VC | The Public Health Agency of Sweden |
| EPI_ISL_455107 | Narhalsan Mellerud | The Public Health Agency of Sweden |
| EPI_ISL_455108 | Smedby HC | The Public Health Agency of Sweden |
| EPI_ISL_455109 | Ulltuna Vardcentral | The Public Health Agency of Sweden |
| EPI_ISL_455110 | Scania Halsocenter, B288 | The Public Health Agency of Sweden |
| EPI_ISL_455111 | Olof Norrby | The Public Health Agency of Sweden |
| EPI_ISL_455112 | Dutch COVID-19 response team | Erasmus Medical Center |
| EPI_ISL_455113 | Dutch COVID-19 response team | Erasmus Medical Center |
| EPI_ISL_455114 | Dutch COVID-19 response team | Erasmus Medical Center |
| EPI_ISL_455115 | Dutch COVID-19 response team | Erasmus Medical Center |
| EPI_ISL_455116 | Dutch COVID-19 response team | Erasmus Medical Center |
| EPI_ISL_455117 | Dutch COVID-19 response team | Erasmus Medical Center |
| EPI_ISL_455118 | Dutch COVID-19 response team | Erasmus Medical Center |
| EPI_ISL_455119 | Dutch COVID-19 response team | Erasmus Medical Center |
| EPI_ISL_455120 | Dutch COVID-19 response team | Erasmus Medical Center |
| EPI_ISL_455121 | Dutch COVID-19 response team | Erasmus Medical Center |
| EPI_ISL_455122 | Dutch COVID-19 response team | Erasmus Medical Center |
| EPI_ISL_455123 | Dutch COVID-19 response team | Erasmus Medical Center |
| EPI_ISL_455124 | Dutch COVID-19 response team | Erasmus Medical Center |
| EPI_ISL_455125 | Dutch COVID-19 response team | Erasmus Medical Center |
| EPI_ISL_455126 | Dutch COVID-19 response team | Erasmus Medical Center |
| EPI_ISL_455127 | Dutch COVID-19 response team | Erasmus Medical Center |
| EPI_ISL_455128 | Dutch COVID-19 response team | Erasmus Medical Center |
| EPI_ISL_455129 | Dutch COVID-19 response team | Erasmus Medical Center |
| EPI_ISL_455130 | Dutch COVID-19 response team | Erasmus Medical Center |
| EPI_ISL_455131 | Dutch COVID-19 response team | Erasmus Medical Center |
| EPI_ISL_455132 | Dutch COVID-19 response team | Erasmus Medical Center |

| EPI_Accession_ID | Originating Laboratory | Submitting Laboratory |
|---|---|---|
| EPI_ISL_455133 | Dutch COVID-19 response team | Erasmus Medical Center |
| EPI_ISL_455134 | Dutch COVID-19 response team | Erasmus Medical Center |
| EPI_ISL_455135 | Dutch COVID-19 response team | Erasmus Medical Center |
| EPI_ISL_455136 | Dutch COVID-19 response team | Erasmus Medical Center |
| EPI_ISL_455137 | Dutch COVID-19 response team | Erasmus Medical Center |
| EPI_ISL_455138 | Dutch COVID-19 response team | Erasmus Medical Center |
| EPI_ISL_455139 | Dutch COVID-19 response team | Erasmus Medical Center |
| EPI_ISL_455140 | Dutch COVID-19 response team | Erasmus Medical Center |
| EPI_ISL_455141 | Dutch COVID-19 response team | Erasmus Medical Center |
| EPI_ISL_455142 | Dutch COVID-19 response team | Erasmus Medical Center |
| EPI_ISL_455143 | Dutch COVID-19 response team | Erasmus Medical Center |
| EPI_ISL_455144 | Dutch COVID-19 response team | Erasmus Medical Center |
| EPI_ISL_455145 | Dutch COVID-19 response team | Erasmus Medical Center |
| EPI_ISL_455146 | Dutch COVID-19 response team | Erasmus Medical Center |
| EPI_ISL_455147 | Dutch COVID-19 response team | Erasmus Medical Center |
| EPI_ISL_455148 | Dutch COVID-19 response team | Erasmus Medical Center |
| EPI_ISL_455149 | Dutch COVID-19 response team | Erasmus Medical Center |
| EPI_ISL_455150 | Dutch COVID-19 response team | Erasmus Medical Center |
| EPI_ISL_455151 | Dutch COVID-19 response team | Erasmus Medical Center |
| EPI_ISL_455152 | Dutch COVID-19 response team | Erasmus Medical Center |
| EPI_ISL_455153 | Dutch COVID-19 response team | Erasmus Medical Center |
| EPI_ISL_455154 | Dutch COVID-19 response team | Erasmus Medical Center |
| EPI_ISL_455155 | Dutch COVID-19 response team | Erasmus Medical Center |
| EPI_ISL_455156 | Dutch COVID-19 response team | Erasmus Medical Center |
| EPI_ISL_455157 | Dutch COVID-19 response team | Erasmus Medical Center |
| EPI_ISL_455158 | Dutch COVID-19 response team | Erasmus Medical Center |
| EPI_ISL_455159 | Dutch COVID-19 response team | Erasmus Medical Center |
| EPI_ISL_455160 | Dutch COVID-19 response team | Erasmus Medical Center |
| EPI_ISL_455161 | Dutch COVID-19 response team | Erasmus Medical Center |
| EPI_ISL_455162 | Dutch COVID-19 response team | Erasmus Medical Center |
| EPI_ISL_455163 | Dutch COVID-19 response team | Erasmus Medical Center |
| EPI_ISL_455164 | Dutch COVID-19 response team | Erasmus Medical Center |
| EPI_ISL_455165 | Dutch COVID-19 response team | Erasmus Medical Center |

| EPI_Accession_ID | Originating Laboratory | Submitting Laboratory |
|---|---|---|
| EPI_ISL_455166 | Dutch COVID-19 response team | Erasmus Medical Center |
| EPI_ISL_455167 | Dutch COVID-19 response team | Erasmus Medical Center |
| EPI_ISL_455168 | Dutch COVID-19 response team | Erasmus Medical Center |
| EPI_ISL_455169 | Dutch COVID-19 response team | Erasmus Medical Center |
| EPI_ISL_455170 | Dutch COVID-19 response team | Erasmus Medical Center |
| EPI_ISL_455171 | Dutch COVID-19 response team | Erasmus Medical Center |
| EPI_ISL_455172 | Dutch COVID-19 response team | Erasmus Medical Center |
| EPI_ISL_455173 | Dutch COVID-19 response team | Erasmus Medical Center |
| EPI_ISL_455174 | Dutch COVID-19 response team | Erasmus Medical Center |
| EPI_ISL_455175 | Dutch COVID-19 response team | Erasmus Medical Center |
| EPI_ISL_455176 | Dutch COVID-19 response team | Erasmus Medical Center |
| EPI_ISL_455177 | Dutch COVID-19 response team | Erasmus Medical Center |
| EPI_ISL_455178 | Dutch COVID-19 response team | Erasmus Medical Center |
| EPI_ISL_455179 | Dutch COVID-19 response team | Erasmus Medical Center |
| EPI_ISL_455180 | Dutch COVID-19 response team | Erasmus Medical Center |
| EPI_ISL_455194 | Dutch COVID-19 response team | Erasmus Medical Center |
| EPI_ISL_455195 | Dutch COVID-19 response team | Erasmus Medical Center |
| EPI_ISL_455196 | Dutch COVID-19 response team | Erasmus Medical Center |
| EPI_ISL_455197 | Dutch COVID-19 response team | Erasmus Medical Center |
| EPI_ISL_455198 | Dutch COVID-19 response team | Erasmus Medical Center |
| EPI_ISL_455199 | Dutch COVID-19 response team | Erasmus Medical Center |
| EPI_ISL_455200 | Dutch COVID-19 response team | Erasmus Medical Center |
| EPI_ISL_455202 | Dutch COVID-19 response team | Erasmus Medical Center |
| EPI_ISL_455203 | Dutch COVID-19 response team | Erasmus Medical Center |
| EPI_ISL_455204 | Dutch COVID-19 response team | Erasmus Medical Center |
| EPI_ISL_455205 | Dutch COVID-19 response team | Erasmus Medical Center |
| EPI_ISL_455206 | Dutch COVID-19 response team | Erasmus Medical Center |
| EPI_ISL_455207 | Dutch COVID-19 response team | Erasmus Medical Center |
| EPI_ISL_455208 | Dutch COVID-19 response team | Erasmus Medical Center |
| EPI_ISL_455209 | Dutch COVID-19 response team | Erasmus Medical Center |
| EPI_ISL_455210 | Dutch COVID-19 response team | Erasmus Medical Center |
| EPI_ISL_455211 | Dutch COVID-19 response team | Erasmus Medical Center |
| EPI_ISL_455212 | Dutch COVID-19 response team | Erasmus Medical Center |

| EPI_Accession_ID | Originating Laboratory | Submitting Laboratory |
|---|---|---|
| EPI_ISL_455213 | Dutch COVID-19 response team | Erasmus Medical Center |
| EPI_ISL_455214 | Dutch COVID-19 response team | Erasmus Medical Center |
| EPI_ISL_455215 | Dutch COVID-19 response team | Erasmus Medical Center |
| EPI_ISL_455216 | Dutch COVID-19 response team | Erasmus Medical Center |
| EPI_ISL_455217 | Dutch COVID-19 response team | Erasmus Medical Center |
| EPI_ISL_455218 | Dutch COVID-19 response team | Erasmus Medical Center |
| EPI_ISL_455219 | Dutch COVID-19 response team | Erasmus Medical Center |
| EPI_ISL_455220 | Dutch COVID-19 response team | Erasmus Medical Center |
| EPI_ISL_455221 | Dutch COVID-19 response team | Erasmus Medical Center |
| EPI_ISL_455222 | Dutch COVID-19 response team | Erasmus Medical Center |
| EPI_ISL_455226 | Dutch COVID-19 response team | Erasmus Medical Center |
| EPI_ISL_455227 | Dutch COVID-19 response team | Erasmus Medical Center |
| EPI_ISL_455228 | Dutch COVID-19 response team | Erasmus Medical Center |
| EPI_ISL_455229 | Dutch COVID-19 response team | Erasmus Medical Center |
| EPI_ISL_455230 | Dutch COVID-19 response team | Erasmus Medical Center |
| EPI_ISL_455231 | Dutch COVID-19 response team | Erasmus Medical Center |
| EPI_ISL_455232 | Dutch COVID-19 response team | Erasmus Medical Center |
| EPI_ISL_455233 | Dutch COVID-19 response team | Erasmus Medical Center |
| EPI_ISL_455234 | Dutch COVID-19 response team | Erasmus Medical Center |
| EPI_ISL_455235 | Dutch COVID-19 response team | Erasmus Medical Center |
| EPI_ISL_455236 | Dutch COVID-19 response team | Erasmus Medical Center |
| EPI_ISL_455237 | Dutch COVID-19 response team | Erasmus Medical Center |
| EPI_ISL_455238 | Dutch COVID-19 response team | Erasmus Medical Center |
| EPI_ISL_455239 | Dutch COVID-19 response team | Erasmus Medical Center |
| EPI_ISL_455240 | Dutch COVID-19 response team | Erasmus Medical Center |
| EPI_ISL_455241 | Dutch COVID-19 response team | Erasmus Medical Center |
| EPI_ISL_455242 | Dutch COVID-19 response team | Erasmus Medical Center |
| EPI_ISL_455243 | Dutch COVID-19 response team | Erasmus Medical Center |
| EPI_ISL_455244 | Dutch COVID-19 response team | Erasmus Medical Center |
| EPI_ISL_455245 | Dutch COVID-19 response team | Erasmus Medical Center |
| EPI_ISL_455246 | Dutch COVID-19 response team | Erasmus Medical Center |
| EPI_ISL_455247 | Dutch COVID-19 response team | Erasmus Medical Center |
| EPI_ISL_455248 | Dutch COVID-19 response team | Erasmus Medical Center |

| EPI_Accession_ID | Originating Laboratory | Submitting Laboratory |
|---|---|---|
| EPI_ISL_455249 | Dutch COVID-19 response team | Erasmus Medical Center |
| EPI_ISL_455250 | Dutch COVID-19 response team | Erasmus Medical Center |
| EPI_ISL_455251 | Dutch COVID-19 response team | Erasmus Medical Center |
| EPI_ISL_455252 | Dutch COVID-19 response team | Erasmus Medical Center |
| EPI_ISL_455253 | Dutch COVID-19 response team | Erasmus Medical Center |
| EPI_ISL_455254 | Dutch COVID-19 response team | Erasmus Medical Center |
| EPI_ISL_455255 | Dutch COVID-19 response team | Erasmus Medical Center |
| EPI_ISL_455256 | Dutch COVID-19 response team | Erasmus Medical Center |
| EPI_ISL_455257 | Dutch COVID-19 response team | Erasmus Medical Center |
| EPI_ISL_455258 | Dutch COVID-19 response team | Erasmus Medical Center |
| EPI_ISL_455261 | Dutch COVID-19 response team | Erasmus Medical Center |
| EPI_ISL_455263 | Dutch COVID-19 response team | Erasmus Medical Center |
| EPI_ISL_455264 | Dutch COVID-19 response team | Erasmus Medical Center |
| EPI_ISL_455265 | Dutch COVID-19 response team | Erasmus Medical Center |
| EPI_ISL_455266 | Dutch COVID-19 response team | Erasmus Medical Center |
| EPI_ISL_455267 | Dutch COVID-19 response team | Erasmus Medical Center |
| EPI_ISL_455268 | Dutch COVID-19 response team | Erasmus Medical Center |
| EPI_ISL_455269 | Dutch COVID-19 response team | Erasmus Medical Center |
| EPI_ISL_455270 | Dutch COVID-19 response team | Erasmus Medical Center |
| EPI_ISL_455271 | Dutch COVID-19 response team | Erasmus Medical Center |
| EPI_ISL_455272 | Dutch COVID-19 response team | Erasmus Medical Center |
| EPI_ISL_455273 | Dutch COVID-19 response team | Erasmus Medical Center |
| EPI_ISL_455274 | Dutch COVID-19 response team | Erasmus Medical Center |
| EPI_ISL_455275 | Dutch COVID-19 response team | Erasmus Medical Center |
| EPI_ISL_455276 | Dutch COVID-19 response team | Erasmus Medical Center |
| EPI_ISL_455277 | Dutch COVID-19 response team | Erasmus Medical Center |
| EPI_ISL_455278 | Dutch COVID-19 response team | Erasmus Medical Center |
| EPI_ISL_455279 | Dutch COVID-19 response team | Erasmus Medical Center |
| EPI_ISL_455280 | Dutch COVID-19 response team | Erasmus Medical Center |
| EPI_ISL_455281 | Dutch COVID-19 response team | Erasmus Medical Center |
| EPI_ISL_455282 | Dutch COVID-19 response team | Erasmus Medical Center |
| EPI_ISL_455283 | Dutch COVID-19 response team | Erasmus Medical Center |
| EPI_ISL_455284 | Dutch COVID-19 response team | Erasmus Medical Center |

| EPI_Accession_ID | Originating Laboratory | Submitting Laboratory |
|---|---|---|
| EPI_ISL_455285 | Dutch COVID-19 response team | Erasmus Medical Center |
| EPI_ISL_455286 | Dutch COVID-19 response team | Erasmus Medical Center |
| EPI_ISL_455287 | Dutch COVID-19 response team | Erasmus Medical Center |
| EPI_ISL_455288 | Dutch COVID-19 response team | Erasmus Medical Center |
| EPI_ISL_455289 | Dutch COVID-19 response team | Erasmus Medical Center |
| EPI_ISL_455290 | Dutch COVID-19 response team | Erasmus Medical Center |
| EPI_ISL_455291 | Dutch COVID-19 response team | Erasmus Medical Center |
| EPI_ISL_455292 | Dutch COVID-19 response team | Erasmus Medical Center |
| EPI_ISL_455293 | Dutch COVID-19 response team | Erasmus Medical Center |
| EPI_ISL_455294 | Dutch COVID-19 response team | Erasmus Medical Center |
| EPI_ISL_455295 | Dutch COVID-19 response team | Erasmus Medical Center |
| EPI_ISL_455296 | Dutch COVID-19 response team | Erasmus Medical Center |
| EPI_ISL_455297 | Dutch COVID-19 response team | Erasmus Medical Center |
| EPI_ISL_455298 | Dutch COVID-19 response team | Erasmus Medical Center |
| EPI_ISL_455299 | Dutch COVID-19 response team | Erasmus Medical Center |
| EPI_ISL_455300 | Dutch COVID-19 response team | Erasmus Medical Center |
| EPI_ISL_455301 | Dutch COVID-19 response team | Erasmus Medical Center |
| EPI_ISL_455302 | Dutch COVID-19 response team | Erasmus Medical Center |
| EPI_ISL_455303 | Dutch COVID-19 response team | Erasmus Medical Center |
| EPI_ISL_455304 | Dutch COVID-19 response team | Erasmus Medical Center |
| EPI_ISL_455305 | Dutch COVID-19 response team | Erasmus Medical Center |
| EPI_ISL_455306 | Dutch COVID-19 response team | Erasmus Medical Center |
| EPI_ISL_455307 | Dutch COVID-19 response team | Erasmus Medical Center |
| EPI_ISL_455309 | REGIONAL VRDL,ICMR-RMRC BBSR | Immunogenomics group, Institute of Life Sciences, Bhubaneswar |
| EPI_ISL_455311 | REGIONAL VRDL,ICMR-RMRC BBSR | Immunogenomics group, Institute of Life Sciences, Bhubaneswar |
| EPI_ISL_455312 | Microbiology Unit, Department of Pathology & Laboratory Medicine, IIUM Medical Centre | SEA Microbiome Unit, Faculty of Industrial Sciences & Technology, Universiti Malaysia Pahang |
| EPI_ISL_455313 | Microbiology Unit, Department of Pathology & Laboratory Medicine, IIUM Medical Centre | SEA Microbiome Unit, Faculty of Industrial Sciences & Technology, Universiti Malaysia Pahang |
| EPI_ISL_455314 | Hospital Virgen del Rocio | Instituto de Salud Carlos III |
| EPI_ISL_455315 | Hospital Virgen de las Nieves | Instituto de Salud Carlos III |

| EPI_Accession_ID | Originating Laboratory | Submitting Laboratory |
|---|---|---|
| EPI_ISL_455316 | Hospital Virgen de las Nieves | Instituto de Salud Carlos III |
| EPI_ISL_455317 | Hospital Virgen de las Nieves | Instituto de Salud Carlos III |
| EPI_ISL_455318 | Hospital Virgen de las Nieves | Instituto de Salud Carlos III |
| EPI_ISL_455319 | Hospital Virgen de las Nieves | Instituto de Salud Carlos III |
| EPI_ISL_455320 | Hospital Virgen de las Nieves | Instituto de Salud Carlos III |
| EPI_ISL_455321 | Hospital Virgen de las Nieves | Instituto de Salud Carlos III |
| EPI_ISL_455322 | Hospital Virgen de las Nieves | Instituto de Salud Carlos III |
| EPI_ISL_455324 | Hospital Virgen de las Nieves | Instituto de Salud Carlos III |
| EPI_ISL_455325 | Hospital Universitario de Canarias | Instituto de Salud Carlos III |
| EPI_ISL_455326 | Hospital Universitario Insular de Gran Canaria | Instituto de Salud Carlos III |
| EPI_ISL_455327 | Consejeria de Sanidad y Asuntos Sociales | Instituto de Salud Carlos III |
| EPI_ISL_455328 | Complejo Hospitalario Universitario La Coruna | Instituto de Salud Carlos III |
| EPI_ISL_455329 | Complejo Hospitalario Universitario La Coruna | Instituto de Salud Carlos III |
| EPI_ISL_455330 | Complejo Hospitalario Universitario La Coruna | Instituto de Salud Carlos III |
| EPI_ISL_455331 | Complejo Hospitalario Universitario La Coruna | Instituto de Salud Carlos III |
| EPI_ISL_455332 | Xerencia de XestiÃ³n Integrada de Pontevedra e o SalnÃ©s | Instituto de Salud Carlos III |
| EPI_ISL_455333 | Complejo Hospitalario Universitario de Santiago | Instituto de Salud Carlos III |
| EPI_ISL_455334 | Complejo Hospitalario de Orense | Instituto de Salud Carlos III |
| EPI_ISL_455335 | Complejo Hospitalario de Orense | Instituto de Salud Carlos III |
| EPI_ISL_455336 | Hospital San Pedro | Instituto de Salud Carlos III |
| EPI_ISL_455337 | Hospital San Pedro | Instituto de Salud Carlos III |
| EPI_ISL_455338 | Hospital San Pedro | Instituto de Salud Carlos III |
| EPI_ISL_455339 | Hospital San Pedro | Instituto de Salud Carlos III |
| EPI_ISL_455340 | Hospital San Pedro | Instituto de Salud Carlos III |
| EPI_ISL_455341 | Hospital San Pedro | Instituto de Salud Carlos III |
| EPI_ISL_455342 | Hospital San Pedro | Instituto de Salud Carlos III |
| EPI_ISL_455343 | Hospital San Pedro | Instituto de Salud Carlos III |
| EPI_ISL_455344 | Hospital Comarcal de Melilla | Instituto de Salud Carlos III |
| EPI_ISL_455345 | Hospital Comarcal de Melilla | Instituto de Salud Carlos III |
| EPI_ISL_455346 | Hospital Comarcal de Melilla | Instituto de Salud Carlos III |
| EPI_ISL_455347 | Hospital Comarcal de Melilla | Instituto de Salud Carlos III |
| EPI_ISL_455348 | Hospital Comarcal de Melilla | Instituto de Salud Carlos III |

| EPI_Accession_ID | Originating Laboratory | Submitting Laboratory |
|---|---|---|
| EPI_ISL_455349 | Hospital Comarcal de Melilla | Instituto de Salud Carlos III |
| EPI_ISL_455351 | Hospital Txagorritxu | Instituto de Salud Carlos III |
| EPI_ISL_455352 | Hospital de Cruces | Instituto de Salud Carlos III |
| EPI_ISL_455353 | Hospital de Cruces | Instituto de Salud Carlos III |
| EPI_ISL_455354 | Hospital de Cruces | Instituto de Salud Carlos III |
| EPI_ISL_455355 | Emory Molecular Diagnostics Laboratory, Emory Healthcare | Piantadosi Lab, Emory Department of Pathology |
| EPI_ISL_455356 | Emory Molecular Diagnostics Laboratory, Emory Healthcare | Piantadosi Lab, Emory Department of Pathology |
| EPI_ISL_455358 | Emory Molecular Diagnostics Laboratory, Emory Healthcare | Piantadosi Lab, Emory Department of Pathology |
| EPI_ISL_455359 | Emory Molecular Diagnostics Laboratory, Emory Healthcare | Piantadosi Lab, Emory Department of Pathology |
| EPI_ISL_455360 | Emory Molecular Diagnostics Laboratory, Emory Healthcare | Piantadosi Lab, Emory Department of Pathology |
| EPI_ISL_455361 | Emory Molecular Diagnostics Laboratory, Emory Healthcare | Piantadosi Lab, Emory Department of Pathology |
| EPI_ISL_455362 | Nigeria Centre for Disease Control (NCDC) | African Centre of Excellence for Genomics of Infectious Diseases (ACEGID), Redeemer's University, Ede, Osun State, Nigeria |
| EPI_ISL_455363 | Wuhan Chain Medical Labs (CMLabs) | State Key Laboratory of Biotherapy of Sichuan University |
| EPI_ISL_455364 | Wuhan Chain Medical Labs (CMLabs) | State Key Laboratory of Biotherapy of Sichuan University |
| EPI_ISL_455365 | Wuhan Chain Medical Labs (CMLabs) | State Key Laboratory of Biotherapy of Sichuan University |
| EPI_ISL_455366 | Wuhan Chain Medical Labs (CMLabs) | State Key Laboratory of Biotherapy of Sichuan University |
| EPI_ISL_455367 | Wuhan Chain Medical Labs (CMLabs) | State Key Laboratory of Biotherapy of Sichuan University |
| EPI_ISL_455368 | Wuhan Chain Medical Labs (CMLabs) | State Key Laboratory of Biotherapy of Sichuan University |

| EPI_Accession_ID | Originating Laboratory | Submitting Laboratory |
|---|---|---|
| EPI_ISL_455369 | Wuhan Chain Medical Labs (CMLabs) | State Key Laboratory of Biotherapy of Sichuan University |
| EPI_ISL_455370 | Wuhan Chain Medical Labs (CMLabs) | State Key Laboratory of Biotherapy of Sichuan University |
| EPI_ISL_455371 | Wuhan Chain Medical Labs (CMLabs) | State Key Laboratory of Biotherapy of Sichuan University |
| EPI_ISL_455372 | Wuhan Chain Medical Labs (CMLabs) | State Key Laboratory of Biotherapy of Sichuan University |
| EPI_ISL_455373 | Wuhan Chain Medical Labs (CMLabs) | State Key Laboratory of Biotherapy of Sichuan University |
| EPI_ISL_455374 | Wuhan Chain Medical Labs (CMLabs) | State Key Laboratory of Biotherapy of Sichuan University |
| EPI_ISL_455375 | Wuhan Chain Medical Labs (CMLabs) | State Key Laboratory of Biotherapy of Sichuan University |
| EPI_ISL_455376 | Wuhan Chain Medical Labs (CMLabs) | State Key Laboratory of Biotherapy of Sichuan University |
| EPI_ISL_455377 | Wuhan Chain Medical Labs (CMLabs) | State Key Laboratory of Biotherapy of Sichuan University |
| EPI_ISL_455378 | Wuhan Chain Medical Labs (CMLabs) | State Key Laboratory of Biotherapy of Sichuan University |
| EPI_ISL_455379 | Wuhan Chain Medical Labs (CMLabs) | State Key Laboratory of Biotherapy of Sichuan University |
| EPI_ISL_455380 | Wuhan Chain Medical Labs (CMLabs) | State Key Laboratory of Biotherapy of Sichuan University |
| EPI_ISL_455381 | Wuhan Chain Medical Labs (CMLabs) | State Key Laboratory of Biotherapy of Sichuan University |
| EPI_ISL_455382 | Wuhan Chain Medical Labs (CMLabs) | State Key Laboratory of Biotherapy of Sichuan University |
| EPI_ISL_455383 | Wuhan Chain Medical Labs (CMLabs) | State Key Laboratory of Biotherapy of Sichuan University |
| EPI_ISL_455384 | Wuhan Chain Medical Labs (CMLabs) | State Key Laboratory of Biotherapy of Sichuan University |

| EPI_Accession_ID | Originating Laboratory | Submitting Laboratory |
|---|---|---|
| EPI_ISL_455385 | Wuhan Chain Medical Labs (CMLabs) | State Key Laboratory of Biotherapy of Sichuan University |
| EPI_ISL_455386 | Wuhan Chain Medical Labs (CMLabs) | State Key Laboratory of Biotherapy of Sichuan University |
| EPI_ISL_455387 | Wuhan Chain Medical Labs (CMLabs) | State Key Laboratory of Biotherapy of Sichuan University |
| EPI_ISL_455388 | Wuhan Chain Medical Labs (CMLabs) | State Key Laboratory of Biotherapy of Sichuan University |
| EPI_ISL_455389 | Wuhan Chain Medical Labs (CMLabs) | State Key Laboratory of Biotherapy of Sichuan University |
| EPI_ISL_455390 | Wuhan Chain Medical Labs (CMLabs) | State Key Laboratory of Biotherapy of Sichuan University |
| EPI_ISL_455391 | Wuhan Chain Medical Labs (CMLabs) | State Key Laboratory of Biotherapy of Sichuan University |
| EPI_ISL_455392 | Wuhan Chain Medical Labs (CMLabs) | State Key Laboratory of Biotherapy of Sichuan University |
| EPI_ISL_455393 | Wuhan Chain Medical Labs (CMLabs) | State Key Laboratory of Biotherapy of Sichuan University |
| EPI_ISL_455394 | Wuhan Chain Medical Labs (CMLabs) | State Key Laboratory of Biotherapy of Sichuan University |
| EPI_ISL_455395 | Wuhan Chain Medical Labs (CMLabs) | State Key Laboratory of Biotherapy of Sichuan University |
| EPI_ISL_455396 | Wuhan Chain Medical Labs (CMLabs) | State Key Laboratory of Biotherapy of Sichuan University |
| EPI_ISL_455397 | Wuhan Chain Medical Labs (CMLabs) | State Key Laboratory of Biotherapy of Sichuan University |
| EPI_ISL_455398 | Wuhan Chain Medical Labs (CMLabs) | State Key Laboratory of Biotherapy of Sichuan University |
| EPI_ISL_455399 | Wuhan Chain Medical Labs (CMLabs) | State Key Laboratory of Biotherapy of Sichuan University |
| EPI_ISL_455400 | Wuhan Chain Medical Labs (CMLabs) | State Key Laboratory of Biotherapy of Sichuan University |

| EPI_Accession_ID | Originating Laboratory | Submitting Laboratory |
|---|---|---|
| EPI_ISL_455401 | Wuhan Chain Medical Labs (CMLabs) | State Key Laboratory of Biotherapy of Sichuan University |
| EPI_ISL_455402 | Wuhan Chain Medical Labs (CMLabs) | State Key Laboratory of Biotherapy of Sichuan University |
| EPI_ISL_455403 | Wuhan Chain Medical Labs (CMLabs) | State Key Laboratory of Biotherapy of Sichuan University |
| EPI_ISL_455404 | Wuhan Chain Medical Labs (CMLabs) | State Key Laboratory of Biotherapy of Sichuan University |
| EPI_ISL_455405 | Wuhan Chain Medical Labs (CMLabs) | State Key Laboratory of Biotherapy of Sichuan University |
| EPI_ISL_455406 | Wuhan Chain Medical Labs (CMLabs) | State Key Laboratory of Biotherapy of Sichuan University |
| EPI_ISL_455407 | Wuhan Chain Medical Labs (CMLabs) | State Key Laboratory of Biotherapy of Sichuan University |
| EPI_ISL_455408 | Wuhan Chain Medical Labs (CMLabs) | State Key Laboratory of Biotherapy of Sichuan University |
| EPI_ISL_455409 | Wuhan Chain Medical Labs (CMLabs) | State Key Laboratory of Biotherapy of Sichuan University |
| EPI_ISL_455410 | Wuhan Chain Medical Labs (CMLabs) | State Key Laboratory of Biotherapy of Sichuan University |
| EPI_ISL_455411 | Wuhan Chain Medical Labs (CMLabs) | State Key Laboratory of Biotherapy of Sichuan University |
| EPI_ISL_455412 | Nigeria Centre for Disease Control (NCDC) | African Centre of Excellence for Genomics of Infectious Diseases (ACEGID), Redeemer's University, Ede, Osun State, Nigeria |
| EPI_ISL_455413 | Nigeria Centre for Disease Control (NCDC) | African Centre of Excellence for Genomics of Infectious Diseases (ACEGID), Redeemer's University, Ede, Osun State, Nigeria |
| EPI_ISL_455415 | Nigeria Centre for Disease Control (NCDC) | African Centre of Excellence for Genomics of Infectious Diseases (ACEGID), Redeemer's University, Ede, Osun State, Nigeria |

| EPI_Accession_ID | Originating Laboratory | Submitting Laboratory |
| --- | --- | --- |
| EPI_ISL_455418 | Nigeria Centre for Disease Control (NCDC) | African Centre of Excellence for Genomics of Infectious Diseases (ACEGID), Redeemer's University, Ede, Osun State, Nigeria |
| EPI_ISL_455419 | Nigeria Centre for Disease Control (NCDC) | African Centre of Excellence for Genomics of Infectious Diseases (ACEGID), Redeemer's University, Ede, Osun State, Nigeria |
| EPI_ISL_455420 | National Institute of Laboratory Medicine and Referral Center | Genomic Research Lab, BCSIR |
| EPI_ISL_455422 | Nigeria Centre for Disease Control | African Centre of Excellence for Genomics of Infectious Diseases (ACEGID), Redeemer's University, Ede, Osun State, Nigeria |
| EPI_ISL_455423 | Nigeria Centre for Disease Control (NCDC) | African Centre of Excellence for Genomics of Infectious Diseases (ACEGID), Redeemer's University, Ede, Osun State, Nigeria |
| EPI_ISL_455424 | Nigeria Centre for Disease Control (NCDC) | African Centre of Excellence for Genomics of Infectious Diseases (ACEGID), Redeemer's University, Ede, Osun State, Nigeria |
| EPI_ISL_455425 | Nigeria Centre for Disease Control (NCDC) | African Centre of Excellence for Genomics of Infectious Diseases (ACEGID), Redeemer's University, Ede, Osun State, Nigeria |
| EPI_ISL_455426 | Nigeria Centre for Disease Control | African Centre of Excellence for Genomics of Infectious Diseases (ACEGID), Redeemer's University, Ede, Osun State, Nigeria |
| EPI_ISL_455427 | Nigeria Centre for Disease Control (NCDC) | African Centre of Excellence for Genomics of Infectious Diseases (ACEGID), Redeemer's University, Ede, Osun State, Nigeria |
| EPI_ISL_455429 | Nigeria Centre for Disease Control (NCDC) | African Centre of Excellence for Genomics of Infectious Diseases (ACEGID), Redeemer's University, Ede, Osun State, Nigeria |
| EPI_ISL_455431 | Nigeria Centre for Disease Control (NCDC) | African Centre of Excellence for Genomics of Infectious Diseases (ACEGID), Redeemer's University, Ede, Osun State, Nigeria |

| EPI_Accession_ID | Originating Laboratory | Submitting Laboratory |
|---|---|---|
| EPI_ISL_455432 | Instituto de Diagnostico y Referencia Epidemiologicos (INDRE) | Instituto de Diagnostico y Referencia Epidemiologicos (INDRE) |
| EPI_ISL_455434 | Instituto de Diagnostico y Referencia Epidemiologicos (INDRE) | Instituto de Diagnostico y Referencia Epidemiologicos (INDRE) |
| EPI_ISL_455435 | Instituto de Diagnostico y Referencia Epidemiologicos (INDRE) | Instituto de Diagnostico y Referencia Epidemiologicos (INDRE) |
| EPI_ISL_455436 | Instituto de Diagnostico y Referencia Epidemiologicos (INDRE) | Instituto de Diagnostico y Referencia Epidemiologicos (INDRE) |
| EPI_ISL_455437 | Instituto de Diagnostico y Referencia Epidemiologicos (INDRE) | Instituto de Diagnostico y Referencia Epidemiologicos (INDRE) |
| EPI_ISL_455438 | Instituto de Diagnostico y Referencia Epidemiologicos (INDRE) | Instituto de Diagnostico y Referencia Epidemiologicos (INDRE) |
| EPI_ISL_455439 | Instituto de Diagnostico y Referencia Epidemiologicos (INDRE) | Instituto de Diagnostico y Referencia Epidemiologicos (INDRE) |
| EPI_ISL_455440 | 1. ViroGenetics - BSL3 Laboratory of Virology, MaÅ‚opolska Centre of Biotechnology, Jagiellonian University; 2. II Department of Internal Medicine, Faculty of Medicine, Jagiellonian University Medical College; 3. Narodowy Instytut Zdrowia Publicznego â€“ PaÅ„stwowy ZakÅ‚ad Higieny (NIZP-PZH) | 1. ViroGenetics - BSL3 Laboratory of Virology, MaÅ‚opolska Centre of Biotechnology, Jagiellonian University; 2. II Department of Internal Medicine, Faculty of Medicine, Jagiellonian University Medical College; 3. Narodowy Instytut Zdrowia Publicznego â€“ PaÅ„stwowy ZakÅ‚ad Higieny (NIZP-PZH). |
| EPI_ISL_455441 | 1. ViroGenetics - BSL3 Laboratory of Virology, MaÅ‚opolska Centre of Biotechnology, Jagiellonian University; 2. II Department of Internal Medicine, Faculty of Medicine, Jagiellonian University Medical College; 3. Narodowy Instytut Zdrowia Publicznego â€“ PaÅ„stwowy ZakÅ‚ad Higieny (NIZP-PZH) | 1. ViroGenetics - BSL3 Laboratory of Virology, MaÅ‚opolska Centre of Biotechnology, Jagiellonian University; 2. II Department of Internal Medicine, Faculty of Medicine, Jagiellonian University Medical College; 3. Narodowy Instytut Zdrowia Publicznego â€“ PaÅ„stwowy ZakÅ‚ad Higieny (NIZP-PZH). |
| EPI_ISL_455442 | 1. ViroGenetics - BSL3 Laboratory of Virology, MaÅ‚opolska Centre of Biotechnology, Jagiellonian University; 2. II Department of Internal Medicine, Faculty of Medicine, Jagiellonian University Medical College; 3. Narodowy Instytut Zdrowia Publicznego â€“ PaÅ„stwowy ZakÅ‚ad Higieny (NIZP-PZH) | 1. ViroGenetics - BSL3 Laboratory of Virology, MaÅ‚opolska Centre of Biotechnology, Jagiellonian University; 2. II Department of Internal Medicine, Faculty of Medicine, Jagiellonian University Medical College; 3. Narodowy Instytut Zdrowia Publicznego â€“ PaÅ„stwowy ZakÅ‚ad Higieny (NIZP-PZH). |

| EPI_Accession_ID | Originating Laboratory | Submitting Laboratory |
|---|---|---|
| EPI_ISL_455443 | 1. ViroGenetics - BSL3 Laboratory of Virology, MaÅ‚opolska Centre of Biotechnology, Jagiellonian University; 2. II Department of Internal Medicine, Faculty of Medicine, Jagiellonian University Medical College; 3. Narodowy Instytut Zdrowia Publicznego â€“ PaÅ„stwowy ZakÅ‚ad Higieny (NIZP-PZH) | 1. ViroGenetics - BSL3 Laboratory of Virology, MaÅ‚opolska Centre of Biotechnology, Jagiellonian University; 2. II Department of Internal Medicine, Faculty of Medicine, Jagiellonian University Medical College; 3. Narodowy Instytut Zdrowia Publicznego â€“ PaÅ„stwowy ZakÅ‚ad Higieny (NIZP-PZH). |
| EPI_ISL_455444 | 1. ViroGenetics - BSL3 Laboratory of Virology, MaÅ‚opolska Centre of Biotechnology, Jagiellonian University; 2. II Department of Internal Medicine, Faculty of Medicine, Jagiellonian University Medical College; 3. Narodowy Instytut Zdrowia Publicznego â€“ PaÅ„stwowy ZakÅ‚ad Higieny (NIZP-PZH) | 1. ViroGenetics - BSL3 Laboratory of Virology, MaÅ‚opolska Centre of Biotechnology, Jagiellonian University; 2. II Department of Internal Medicine, Faculty of Medicine, Jagiellonian University Medical College; 3. Narodowy Instytut Zdrowia Publicznego â€“ PaÅ„stwowy ZakÅ‚ad Higieny (NIZP-PZH). |
| EPI_ISL_455445 | 1. ViroGenetics - BSL3 Laboratory of Virology, MaÅ‚opolska Centre of Biotechnology, Jagiellonian University; 2. II Department of Internal Medicine, Faculty of Medicine, Jagiellonian University Medical College; 3. Narodowy Instytut Zdrowia Publicznego â€“ PaÅ„stwowy ZakÅ‚ad Higieny (NIZP-PZH) | 1. ViroGenetics - BSL3 Laboratory of Virology, MaÅ‚opolska Centre of Biotechnology, Jagiellonian University; 2. II Department of Internal Medicine, Faculty of Medicine, Jagiellonian University Medical College; 3. Narodowy Instytut Zdrowia Publicznego â€“ PaÅ„stwowy ZakÅ‚ad Higieny (NIZP-PZH). |
| EPI_ISL_455446 | 1. ViroGenetics - BSL3 Laboratory of Virology, MaÅ‚opolska Centre of Biotechnology, Jagiellonian University; 2. II Department of Internal Medicine, Faculty of Medicine, Jagiellonian University Medical College; 3. Narodowy Instytut Zdrowia Publicznego â€“ PaÅ„stwowy ZakÅ‚ad Higieny (NIZP-PZH) | 1. ViroGenetics - BSL3 Laboratory of Virology, MaÅ‚opolska Centre of Biotechnology, Jagiellonian University; 2. II Department of Internal Medicine, Faculty of Medicine, Jagiellonian University Medical College; 3. Narodowy Instytut Zdrowia Publicznego â€“ PaÅ„stwowy ZakÅ‚ad Higieny (NIZP-PZH). |
| EPI_ISL_455447 | 1. ViroGenetics - BSL3 Laboratory of Virology, MaÅ‚opolska Centre of Biotechnology, Jagiellonian University; 2. II Department of Internal Medicine, Faculty of Medicine, Jagiellonian University Medical College; 3. Narodowy Instytut Zdrowia Publicznego â€“ PaÅ„stwowy ZakÅ‚ad Higieny (NIZP-PZH) | 1. ViroGenetics - BSL3 Laboratory of Virology, MaÅ‚opolska Centre of Biotechnology, Jagiellonian University; 2. II Department of Internal Medicine, Faculty of Medicine, Jagiellonian University Medical College; 3. Narodowy Instytut Zdrowia Publicznego â€“ PaÅ„stwowy ZakÅ‚ad Higieny (NIZP-PZH). |

| EPI_Accession_ID | Originating Laboratory | Submitting Laboratory |
|---|---|---|
| EPI_ISL_455448 | 1. ViroGenetics - BSL3 Laboratory of Virology, MaÅ‚opolska Centre of Biotechnology, Jagiellonian University; 2. II Department of Internal Medicine, Faculty of Medicine, Jagiellonian University Medical College; 3. Narodowy Instytut Zdrowia Publicznego â€“ PaÅ„stwowy ZakÅ‚ad Higieny (NIZP-PZH) | 1. ViroGenetics - BSL3 Laboratory of Virology, MaÅ‚opolska Centre of Biotechnology, Jagiellonian University; 2. II Department of Internal Medicine, Faculty of Medicine, Jagiellonian University Medical College; 3. Narodowy Instytut Zdrowia Publicznego â€“ PaÅ„stwowy ZakÅ‚ad Higieny (NIZP-PZH). |
| EPI_ISL_455449 | 1. ViroGenetics - BSL3 Laboratory of Virology, MaÅ‚opolska Centre of Biotechnology, Jagiellonian University; 2. II Department of Internal Medicine, Faculty of Medicine, Jagiellonian University Medical College; 3. Narodowy Instytut Zdrowia Publicznego â€“ PaÅ„stwowy ZakÅ‚ad Higieny (NIZP-PZH) | 1. ViroGenetics - BSL3 Laboratory of Virology, MaÅ‚opolska Centre of Biotechnology, Jagiellonian University; 2. II Department of Internal Medicine, Faculty of Medicine, Jagiellonian University Medical College; 3. Narodowy Instytut Zdrowia Publicznego â€“ PaÅ„stwowy ZakÅ‚ad Higieny (NIZP-PZH). |
| EPI_ISL_455450 | 1. ViroGenetics - BSL3 Laboratory of Virology, MaÅ‚opolska Centre of Biotechnology, Jagiellonian University; 2. II Department of Internal Medicine, Faculty of Medicine, Jagiellonian University Medical College; 3. Narodowy Instytut Zdrowia Publicznego â€“ PaÅ„stwowy ZakÅ‚ad Higieny (NIZP-PZH) | 1. ViroGenetics - BSL3 Laboratory of Virology, MaÅ‚opolska Centre of Biotechnology, Jagiellonian University; 2. II Department of Internal Medicine, Faculty of Medicine, Jagiellonian University Medical College; 3. Narodowy Instytut Zdrowia Publicznego â€“ PaÅ„stwowy ZakÅ‚ad Higieny (NIZP-PZH). |
| EPI_ISL_455451 | 1. ViroGenetics - BSL3 Laboratory of Virology, MaÅ‚opolska Centre of Biotechnology, Jagiellonian University; 2. II Department of Internal Medicine, Faculty of Medicine, Jagiellonian University Medical College; 3. Narodowy Instytut Zdrowia Publicznego â€“ PaÅ„stwowy ZakÅ‚ad Higieny (NIZP-PZH) | 1. ViroGenetics - BSL3 Laboratory of Virology, MaÅ‚opolska Centre of Biotechnology, Jagiellonian University; 2. II Department of Internal Medicine, Faculty of Medicine, Jagiellonian University Medical College; 3. Narodowy Instytut Zdrowia Publicznego â€“ PaÅ„stwowy ZakÅ‚ad Higieny (NIZP-PZH). |
| EPI_ISL_455452 | 1. ViroGenetics - BSL3 Laboratory of Virology, MaÅ‚opolska Centre of Biotechnology, Jagiellonian University; 2. II Department of Internal Medicine, Faculty of Medicine, Jagiellonian University Medical College; 3. Narodowy Instytut Zdrowia Publicznego â€“ PaÅ„stwowy ZakÅ‚ad Higieny (NIZP-PZH) | 1. ViroGenetics - BSL3 Laboratory of Virology, MaÅ‚opolska Centre of Biotechnology, Jagiellonian University; 2. II Department of Internal Medicine, Faculty of Medicine, Jagiellonian University Medical College; 3. Narodowy Instytut Zdrowia Publicznego â€“ PaÅ„stwowy ZakÅ‚ad Higieny (NIZP-PZH). |

| EPI_Accession_ID | Originating Laboratory | Submitting Laboratory |
|---|---|---|
| EPI_ISL_455453 | 1. ViroGenetics - BSL3 Laboratory of Virology, MaÅ‚opolska Centre of Biotechnology, Jagiellonian University; 2. II Department of Internal Medicine, Faculty of Medicine, Jagiellonian University Medical College; 3. Narodowy Instytut Zdrowia Publicznego â€“ PaÅ„stwowy ZakÅ‚ad Higieny (NIZP-PZH) | 1. ViroGenetics - BSL3 Laboratory of Virology, MaÅ‚opolska Centre of Biotechnology, Jagiellonian University; 2. II Department of Internal Medicine, Faculty of Medicine, Jagiellonian University Medical College; 3. Narodowy Instytut Zdrowia Publicznego â€“ PaÅ„stwowy ZakÅ‚ad Higieny (NIZP-PZH). |
| EPI_ISL_455454 | Instituto de Diagnostico y Referencia Epidemiologicos (INDRE) | Instituto de Diagnostico y Referencia Epidemiologicos (INDRE) |
| EPI_ISL_455455 | Instituto de Diagnostico y Referencia Epidemiologicos (INDRE) | Instituto de Diagnostico y Referencia Epidemiologicos (INDRE) |
| EPI_ISL_455456 | Instituto de Diagnostico y Referencia Epidemiologicos (INDRE) | Instituto de Diagnostico y Referencia Epidemiologicos (INDRE) |
| EPI_ISL_455458 | National Institute of Laboratory Medicine and Referral Center | Genomic Research Lab, BCSIR |
| EPI_ISL_455459 | National Institute of Laboratory Medicine and Referral Center | Genomic Research Lab, BCSIR |
| EPI_ISL_455460 | Jiangxi Province Center for Disease Control and Prevention | Jiangxi Province Center for Disease Control and Prevention |
| EPI_ISL_455461 | Jiangxi Province Center for Disease Control and Prevention | Jiangxi Province Center for Disease Control and Prevention |
| EPI_ISL_455462 | Jiangxi Province Center for Disease Control and Prevention | Jiangxi Province Center for Disease Control and Prevention |
| EPI_ISL_455463 | Jiangxi Province Center for Disease Control and Prevention | Jiangxi Province Center for Disease Control and Prevention |
| EPI_ISL_455464 | Jiangxi Province Center for Disease Control and Prevention | Jiangxi Province Center for Disease Control and Prevention |
| EPI_ISL_455465 | Jiangxi Province Center for Disease Control and Prevention | Jiangxi Province Center for Disease Control and Prevention |
| EPI_ISL_455466 | Jiangxi Province Center for Disease Control and Prevention | Jiangxi Province Center for Disease Control and Prevention |
| EPI_ISL_455467 | Jiangxi Province Center for Disease Control and Prevention | Jiangxi Province Center for Disease Control and Prevention |

| EPI_Accession_ID | Originating Laboratory | Submitting Laboratory |
|---|---|---|
| EPI_ISL_455468 | Laboratory for Respiratory Viruses, Cantacuzino National Military-Medical Institute for Research and Development | Cantacuzino Institute |
| EPI_ISL_455469 | Laboratory for Respiratory Viruses, Cantacuzino National Military-Medical Institute for Research and Development | Cantacuzino Institute |
| EPI_ISL_455470 | Laboratory for Respiratory Viruses, Cantacuzino National Military-Medical Institute for Research and Development | Cantacuzino Institute |
| EPI_ISL_455471 | Laboratory for Respiratory Viruses, Cantacuzino National Military-Medical Institute for Research and Development | Cantacuzino Institute |
| EPI_ISL_455472 | Laboratory for Respiratory Viruses, Cantacuzino National Military-Medical Institute for Research and Development | Cantacuzino Institute |
| EPI_ISL_455473 | Laboratory for Respiratory Viruses, Cantacuzino National Military-Medical Institute for Research and Development | Cantacuzino Institute |
| EPI_ISL_455474 | Laboratory for Respiratory Viruses, Cantacuzino National Military-Medical Institute for Research and Development | Cantacuzino Institute |
| EPI_ISL_455475 | Laboratory for Respiratory Viruses, Cantacuzino National Military-Medical Institute for Research and Development | Cantacuzino Institute |
| EPI_ISL_455476 | Laboratory for Respiratory Viruses, Cantacuzino National Military-Medical Institute for Research and Development | Cantacuzino Institute |
| EPI_ISL_455477 | Laboratory for Respiratory Viruses, Cantacuzino National Military-Medical Institute for Research and Development | Cantacuzino Institute |
| EPI_ISL_455478 | REGIONAL VRDL,ICMR-RMRC BBSR | Immunogenomics group, Institute of Life Sciences, Bhubaneswar |
| EPI_ISL_455480 | Veterinary Specialized Institute Kraljevo | Veterinary Specialized Institute Kraljevo |

| EPI_Accession_ID | Originating Laboratory | Submitting Laboratory |
|---|---|---|
| EPI_ISL_455566 | Institute for Public Health | Laboratory for advanced genomics |
| EPI_ISL_455567 | Institute for Public Health | Laboratory for advanced genomics |
| EPI_ISL_455568 | Gundersen Molecular Diagnostics Laboratory | Kabara Cancer Research Institute |
| EPI_ISL_455569 | Gundersen Molecular Diagnostics Laboratory | Kabara Cancer Research Institute |
| EPI_ISL_455570 | Gundersen Molecular Diagnostics Laboratory | Kabara Cancer Research Institute |
| EPI_ISL_455571 | Gundersen Molecular Diagnostics Laboratory | Kabara Cancer Research Institute |
| EPI_ISL_455572 | Gundersen Molecular Diagnostics Laboratory | Kabara Cancer Research Institute |
| EPI_ISL_455573 | Gundersen Clinical Microbiology Laboratory | Kabara Cancer Research Institute |
| EPI_ISL_455574 | Gundersen Clinical Microbiology Laboratory | Kabara Cancer Research Institute |
| EPI_ISL_455575 | Gundersen Molecular Diagnostics Laboratory | Kabara Cancer Research Institute |
| EPI_ISL_455576 | Gundersen Molecular Diagnostics Laboratory | Kabara Cancer Research Institute |
| EPI_ISL_455577 | Gundersen Molecular Diagnostics Laboratory | Kabara Cancer Research Institute |
| EPI_ISL_455578 | Gundersen Molecular Diagnostics Laboratory | Kabara Cancer Research Institute |
| EPI_ISL_455579 | Gundersen Molecular Diagnostics Laboratory | Kabara Cancer Research Institute |
| EPI_ISL_455580 | Gundersen Clinical Microbiology Laboratory | Kabara Cancer Research Institute |
| EPI_ISL_455581 | Gundersen Clinical Microbiology Laboratory | Kabara Cancer Research Institute |
| EPI_ISL_455582 | Gundersen Clinical Microbiology Laboratory | Kabara Cancer Research Institute |
| EPI_ISL_455583 | Central Chest Institute of Thailand | National Institute of Health. Department of medical Sciences, Ministry of Public Health, Thailand |
| EPI_ISL_455584 | unknown | National Institute of Health. Department of medical Sciences, Ministry of Public Health, Thailand |
| EPI_ISL_455585 | Phramongkutklao Hospital | National Institute of Health. Department of medical Sciences, Ministry of Public Health, Thailand |
| EPI_ISL_455586 | siriraj hospital | National Institute of Health. Department of medical Sciences, Ministry of Public Health, Thailand |
| EPI_ISL_455587 | RH Princess Maha Chakri Sirindhorn Medical Center-MSMC Hospital | National Institute of Health. Department of medical Sciences, Ministry of Public Health, Thailand |
| EPI_ISL_455588 | Trang hospital | National Institute of Health. Department of medical Sciences, Ministry of Public Health, Thailand |
| EPI_ISL_455589 | unknown | National Institute of Health. Department of medical Sciences, Ministry of Public Health, Thailand |
| EPI_ISL_455590 | unknown | National Institute of Health. Department of medical Sciences, Ministry of Public Health, Thailand |

| EPI_Accession_ID | Originating Laboratory | Submitting Laboratory |
|---|---|---|
| EPI_ISL_455591 | unknown | National Institute of Health. Department of medical Sciences, Ministry of Public Health, Thailand |
| EPI_ISL_455592 | unknown | National Institute of Health. Department of medical Sciences, Ministry of Public Health, Thailand |
| EPI_ISL_455593 | unknown | National Institute of Health. Department of medical Sciences, Ministry of Public Health, Thailand |
| EPI_ISL_455594 | Central Chest Institute of Thailand | National Institute of Health. Department of medical Sciences, Ministry of Public Health, Thailand |
| EPI_ISL_455595 | SA Pathology | VPRL |
| EPI_ISL_455596 | SA Pathology | VPRL |
| EPI_ISL_455597 | SA Pathology | VPRL |
| EPI_ISL_455598 | SA Pathology | VPRL |
| EPI_ISL_455599 | SA Pathology | VPRL |
| EPI_ISL_455600 | SA Pathology | VPRL |
| EPI_ISL_455601 | SA Pathology | VPRL |
| EPI_ISL_455602 | SA Pathology | VPRL |
| EPI_ISL_455603 | SA Pathology | VPRL |
| EPI_ISL_455604 | Ramkhamhaeng Hospital | National Institute of Health. Department of medical Sciences, Ministry of Public Health, Thailand |
| EPI_ISL_455605 | Panyananthaphikkhu Chonprathan Medical Center | National Institute of Health. Department of medical Sciences, Ministry of Public Health, Thailand |
| EPI_ISL_455606 | Praram 9 Hospital | National Institute of Health. Department of medical Sciences, Ministry of Public Health, Thailand |
| EPI_ISL_455607 | Praram 9 Hospital | National Institute of Health. Department of medical Sciences, Ministry of Public Health, Thailand |
| EPI_ISL_455608 | Phramongkutklao Hospital | National Institute of Health. Department of medical Sciences, Ministry of Public Health, Thailand |
| EPI_ISL_455609 | Ochsner Health | Bioinfoexperts, LLC |
| EPI_ISL_455610 | Ochsner Health | Bioinfoexperts, LLC |
| EPI_ISL_455611 | Ochsner Health | Bioinfoexperts, LLC |
| EPI_ISL_455612 | Ochsner Health | Bioinfoexperts, LLC |
| EPI_ISL_455613 | Ochsner Health | Bioinfoexperts, LLC |
| EPI_ISL_455614 | Ochsner Health | Bioinfoexperts, LLC |

| EPI_Accession_ID | Originating Laboratory | Submitting Laboratory |
|---|---|---|
| EPI_ISL_455615 | Ochsner Health | Bioinfoexperts, LLC |
| EPI_ISL_455616 | Ochsner Health | Bioinfoexperts, LLC |
| EPI_ISL_455617 | Ochsner Health | Bioinfoexperts, LLC |
| EPI_ISL_455618 | Ochsner Health | Bioinfoexperts, LLC |
| EPI_ISL_455619 | Ochsner Health | Bioinfoexperts, LLC |
| EPI_ISL_455620 | Ochsner Health | Bioinfoexperts, LLC |
| EPI_ISL_455621 | Ochsner Health | Bioinfoexperts, LLC |
| EPI_ISL_455622 | Ochsner Health | Bioinfoexperts, LLC |
| EPI_ISL_455623 | Ochsner Health | Bioinfoexperts, LLC |
| EPI_ISL_455624 | n/a | National Institute of Health. Department of medical Sciences, Ministry of Public Health, Thailand |
| EPI_ISL_455625 | unknown | Instituto Nacional de Saude (INSA) |
| EPI_ISL_455626 | unknown | Instituto Nacional de Saude (INSA) |
| EPI_ISL_455627 | unknown | Instituto Nacional de Saude (INSA) |
| EPI_ISL_455629 | KRISP, KZN Research Innovation and Sequencing Platform | KRISP, KZN Research Innovation and Sequencing Platform |
| EPI_ISL_455630 | KRISP, KZN Research Innovation and Sequencing Platform | KRISP, KZN Research Innovation and Sequencing Platform |
| EPI_ISL_455631 | KRISP, KZN Research Innovation and Sequencing Platform | KRISP, KZN Research Innovation and Sequencing Platform |
| EPI_ISL_455632 | KRISP, KZN Research Innovation and Sequencing Platform | KRISP, KZN Research Innovation and Sequencing Platform |
| EPI_ISL_455633 | KRISP, KZN Research Innovation and Sequencing Platform | KRISP, KZN Research Innovation and Sequencing Platform |
| EPI_ISL_455634 | KRISP, KZN Research Innovation and Sequencing Platform | KRISP, KZN Research Innovation and Sequencing Platform |
| EPI_ISL_455635 | KRISP, KZN Research Innovation and Sequencing Platform | KRISP, KZN Research Innovation and Sequencing Platform |
| EPI_ISL_455636 | KRISP, KZN Research Innovation and Sequencing Platform | KRISP, KZN Research Innovation and Sequencing Platform |
| EPI_ISL_455637 | KRISP, KZN Research Innovation and Sequencing Platform | KRISP, KZN Research Innovation and Sequencing Platform |

| EPI_Accession_ID | Originating Laboratory | Submitting Laboratory |
|---|---|---|
| EPI_ISL_455638 | KRISP, KZN Research Innovation and Sequencing Platform | KRISP, KZN Research Innovation and Sequencing Platform |
| EPI_ISL_455639 | KRISP, KZN Research Innovation and Sequencing Platform | KRISP, KZN Research Innovation and Sequencing Platform |
| EPI_ISL_455640 | ICMR-National Institute of Cholera and Enteric Diseases | National Institute of Biomedical Genomics |
| EPI_ISL_455641 | ICMR-National Institute of Cholera and Enteric Diseases | National Institute of Biomedical Genomics |
| EPI_ISL_455642 | ICMR-National Institute of Cholera and Enteric Diseases | National Institute of Biomedical Genomics |
| EPI_ISL_455643 | ICMR-National Institute of Cholera and Enteric Diseases | National Institute of Biomedical Genomics |
| EPI_ISL_455644 | ICMR-National Institute of Cholera and Enteric Diseases | National Institute of Biomedical Genomics |
| EPI_ISL_455645 | ICMR-National Institute of Cholera and Enteric Diseases | National Institute of Biomedical Genomics |
| EPI_ISL_455646 | ICMR-National Institute of Cholera and Enteric Diseases | National Institute of Biomedical Genomics |
| EPI_ISL_455647 | ICMR-National Institute of Cholera and Enteric Diseases | National Institute of Biomedical Genomics |
| EPI_ISL_455648 | ICMR-National Institute of Cholera and Enteric Diseases | National Institute of Biomedical Genomics |
| EPI_ISL_455649 | ICMR-National Institute of Cholera and Enteric Diseases | National Institute of Biomedical Genomics |
| EPI_ISL_455650 | ICMR-National Institute of Cholera and Enteric Diseases | National Institute of Biomedical Genomics |
| EPI_ISL_455651 | ICMR-National Institute of Cholera and Enteric Diseases | National Institute of Biomedical Genomics |
| EPI_ISL_455652 | ICMR-National Institute of Cholera and Enteric Diseases | National Institute of Biomedical Genomics |
| EPI_ISL_455653 | ICMR-National Institute of Cholera and Enteric Diseases | National Institute of Biomedical Genomics |

| EPI_Accession_ID | Originating Laboratory | Submitting Laboratory |
|---|---|---|
| EPI_ISL_455654 | ICMR-National Institute of Cholera and Enteric Diseases | National Institute of Biomedical Genomics |
| EPI_ISL_455655 | ICMR-National Institute of Cholera and Enteric Diseases | National Institute of Biomedical Genomics |
| EPI_ISL_455656 | ICMR-National Institute of Cholera and Enteric Diseases | National Institute of Biomedical Genomics |
| EPI_ISL_455657 | ICMR-National Institute of Cholera and Enteric Diseases | National Institute of Biomedical Genomics |
| EPI_ISL_455658 | ICMR-National Institute of Cholera and Enteric Diseases | National Institute of Biomedical Genomics |
| EPI_ISL_455659 | ICMR-National Institute of Cholera and Enteric Diseases | National Institute of Biomedical Genomics |
| EPI_ISL_455660 | ICMR-National Institute of Cholera and Enteric Diseases | National Institute of Biomedical Genomics |
| EPI_ISL_455661 | ICMR-National Institute of Cholera and Enteric Diseases | National Institute of Biomedical Genomics |
| EPI_ISL_455662 | ICMR-National Institute of Cholera and Enteric Diseases | National Institute of Biomedical Genomics |
| EPI_ISL_455663 | ICMR-National Institute of Cholera and Enteric Diseases | National Institute of Biomedical Genomics |
| EPI_ISL_455664 | ICMR-National Institute of Cholera and Enteric Diseases | National Institute of Biomedical Genomics |
| EPI_ISL_455665 | ICMR-National Institute of Cholera and Enteric Diseases | National Institute of Biomedical Genomics |
| EPI_ISL_455666 | ICMR-National Institute of Cholera and Enteric Diseases | National Institute of Biomedical Genomics |
| EPI_ISL_455667 | ICMR-National Institute of Cholera and Enteric Diseases | National Institute of Biomedical Genomics |
| EPI_ISL_455668 | ICMR-National Institute of Cholera and Enteric Diseases | National Institute of Biomedical Genomics |
| EPI_ISL_455669 | ICMR-National Institute of Cholera and Enteric Diseases | National Institute of Biomedical Genomics |

| EPI_Accession_ID | Originating Laboratory | Submitting Laboratory |
|---|---|---|
| EPI_ISL_455670 | ICMR-National Institute of Cholera and Enteric Diseases | National Institute of Biomedical Genomics |
| EPI_ISL_455671 | ICMR-National Institute of Cholera and Enteric Diseases | National Institute of Biomedical Genomics |
| EPI_ISL_455672 | ICMR-National Institute of Cholera and Enteric Diseases | National Institute of Biomedical Genomics |
| EPI_ISL_455673 | ICMR-National Institute of Cholera and Enteric Diseases | National Institute of Biomedical Genomics |
| EPI_ISL_455674 | ICMR-National Institute of Cholera and Enteric Diseases | National Institute of Biomedical Genomics |
| EPI_ISL_455675 | ICMR-National Institute of Cholera and Enteric Diseases | National Institute of Biomedical Genomics |
| EPI_ISL_455676 | ICMR-National Institute of Cholera and Enteric Diseases | National Institute of Biomedical Genomics |
| EPI_ISL_455677 | ICMR-National Institute of Cholera and Enteric Diseases | National Institute of Biomedical Genomics |
| EPI_ISL_455678 | ICMR-National Institute of Cholera and Enteric Diseases | National Institute of Biomedical Genomics |
| EPI_ISL_455679 | ICMR-National Institute of Cholera and Enteric Diseases | National Institute of Biomedical Genomics |
| EPI_ISL_455680 | unknown | Institute of pathogenic microbiology |
| EPI_ISL_455682 | unknown | Environmental and Global Health |
| EPI_ISL_455683 | unknown | Department of Microbiology |
| EPI_ISL_455684 | unknown | Department of Microbiology |
| EPI_ISL_455685 | unknown | Department of Microbiology |
| EPI_ISL_455686 | unknown | Department of Microbiology |
| EPI_ISL_455687 | unknown | Department of Microbiology |
| EPI_ISL_455688 | unknown | Department of Microbiology |
| EPI_ISL_455689 | unknown | Department of Microbiology |
| EPI_ISL_455690 | unknown | Department of Microbiology |
| EPI_ISL_455691 | unknown | Department of Microbiology |
| EPI_ISL_455692 | unknown | Department of Microbiology |
| EPI_ISL_455693 | unknown | Department of Microbiology |

| EPI_Accession_ID | Originating Laboratory | Submitting Laboratory |
|---|---|---|
| EPI_ISL_455694 | National Hospital of Tropical Diseases | Oxford University Clinical Research Unit, Hanoi, Vietnam |
| EPI_ISL_455695 | National Hospital of Tropical Diseases | Oxford University Clinical Research Unit, Hanoi, Vietnam |
| EPI_ISL_455696 | National Hospital of Tropical Diseases | Oxford University Clinical Research Unit, Hanoi, Vietnam |
| EPI_ISL_455697 | National Hospital of Tropical Diseases | Oxford University Clinical Research Unit, Hanoi, Vietnam |
| EPI_ISL_455698 | National Hospital of Tropical Diseases | Oxford University Clinical Research Unit, Hanoi, Vietnam |
| EPI_ISL_455699 | National Hospital of Tropical Diseases | Oxford University Clinical Research Unit, Hanoi, Vietnam |
| EPI_ISL_455700 | National Hospital of Tropical Diseases | Oxford University Clinical Research Unit, Hanoi, Vietnam |
| EPI_ISL_455701 | National Hospital of Tropical Diseases | Oxford University Clinical Research Unit, Hanoi, Vietnam |
| EPI_ISL_455702 | National Hospital of Tropical Diseases | Oxford University Clinical Research Unit, Hanoi, Vietnam |
| EPI_ISL_455703 | National Hospital of Tropical Diseases | Oxford University Clinical Research Unit, Hanoi, Vietnam |
| EPI_ISL_455704 | National Hospital of Tropical Diseases | Oxford University Clinical Research Unit, Hanoi, Vietnam |
| EPI_ISL_455705 | National Hospital of Tropical Diseases | Oxford University Clinical Research Unit, Hanoi, Vietnam |
| EPI_ISL_455706 | National Hospital of Tropical Diseases | Oxford University Clinical Research Unit, Hanoi, Vietnam |
| EPI_ISL_455707 | National Hospital of Tropical Diseases | Oxford University Clinical Research Unit, Hanoi, Vietnam |
| EPI_ISL_455708 | National Hospital of Tropical Diseases | Oxford University Clinical Research Unit, Hanoi, Vietnam |
| EPI_ISL_455709 | National Hospital of Tropical Diseases | Oxford University Clinical Research Unit, Hanoi, Vietnam |

| EPI_Accession_ID | Originating Laboratory | Submitting Laboratory |
|---|---|---|
| EPI_ISL_455710 | National Hospital of Tropical Diseases | Oxford University Clinical Research Unit, Hanoi, Vietnam |
| EPI_ISL_455711 | National Hospital of Tropical Diseases | Oxford University Clinical Research Unit, Hanoi, Vietnam |
| EPI_ISL_455712 | National Hospital of Tropical Diseases | Oxford University Clinical Research Unit, Hanoi, Vietnam |
| EPI_ISL_455713 | National Hospital of Tropical Diseases | Oxford University Clinical Research Unit, Hanoi, Vietnam |
| EPI_ISL_455714 | National Hospital of Tropical Diseases | Oxford University Clinical Research Unit, Hanoi, Vietnam |
| EPI_ISL_455715 | National Hospital of Tropical Diseases | Oxford University Clinical Research Unit, Hanoi, Vietnam |
| EPI_ISL_455716 | National Hospital of Tropical Diseases | Oxford University Clinical Research Unit, Hanoi, Vietnam |
| EPI_ISL_455717 | National Hospital of Tropical Diseases | Oxford University Clinical Research Unit, Hanoi, Vietnam |
| EPI_ISL_455718 | National Hospital of Tropical Diseases | Oxford University Clinical Research Unit, Hanoi, Vietnam |
| EPI_ISL_455719 | T.C. SaÄŸlÄ±k BakanlÄ±ÄŸÄ± AdÄ±yaman Ä°l SaÄŸlÄ±k MÃ¼dÃ¼rlÃ¼ÄŸÃ¼ AdÄ±yaman EÄŸitim Ve AraÅŸtÄ±rma Hastanesi | VETAL Animal Health Products Company, BSL3+ Production Laboratuary /Turkey |
| EPI_ISL_455721 | Servicio de Microbiologia. Hospital Clinico Universitario de Valencia | Sequencing and Bioinformatics Service and Molecular Epidemiology Research Group. FISABIO-Public Health, and SeqCOVID-Spain Consortium |
| EPI_ISL_455722 | Servicio de Microbiologia. Hospital Clinico Universitario de Valencia | Sequencing and Bioinformatics Service and Molecular Epidemiology Research Group. FISABIO-Public Health, and SeqCOVID-Spain Consortium |
| EPI_ISL_455723 | Servicio de Microbiologia. Hospital Clinico Universitario de Valencia | Sequencing and Bioinformatics Service and Molecular Epidemiology Research Group. FISABIO-Public Health, and SeqCOVID-Spain Consortium |
| EPI_ISL_455724 | Servicio de Microbiologia. Hospital Clinico Universitario de Valencia | Sequencing and Bioinformatics Service and Molecular Epidemiology Research Group. FISABIO-Public Health, and SeqCOVID-Spain Consortium |

| EPI_Accession_ID | Originating Laboratory | Submitting Laboratory |
|---|---|---|
| EPI_ISL_455725 | Servicio de Microbiologia. Hospital Clinico Universitario de Valencia | Sequencing and Bioinformatics Service and Molecular Epidemiology Research Group. FISABIO-Public Health, and SeqCOVID-Spain Consortium |
| EPI_ISL_455726 | Servicio de Microbiologia. Hospital Clinico Universitario de Valencia | Sequencing and Bioinformatics Service and Molecular Epidemiology Research Group. FISABIO-Public Health, and SeqCOVID-Spain Consortium |
| EPI_ISL_455727 | Servicio de Microbiologia. Hospital Clinico Universitario de Valencia | Sequencing and Bioinformatics Service and Molecular Epidemiology Research Group. FISABIO-Public Health, and SeqCOVID-Spain Consortium |
| EPI_ISL_455728 | Servicio de Microbiologia. Hospital Clinico Universitario de Valencia | Sequencing and Bioinformatics Service and Molecular Epidemiology Research Group. FISABIO-Public Health, and SeqCOVID-Spain Consortium |
| EPI_ISL_455729 | Servicio de Microbiologia. Hospital Clinico Universitario de Valencia | Sequencing and Bioinformatics Service and Molecular Epidemiology Research Group. FISABIO-Public Health, and SeqCOVID-Spain Consortium |
| EPI_ISL_455730 | Servicio de Microbiologia. Hospital Clinico Universitario de Valencia | Sequencing and Bioinformatics Service and Molecular Epidemiology Research Group. FISABIO-Public Health, and SeqCOVID-Spain Consortium |
| EPI_ISL_455731 | Servicio de Microbiologia. Hospital Clinico Universitario de Valencia | Sequencing and Bioinformatics Service and Molecular Epidemiology Research Group. FISABIO-Public Health, and SeqCOVID-Spain Consortium |
| EPI_ISL_455732 | Servicio de Microbiologia. Hospital Clinico Universitario de Valencia | Sequencing and Bioinformatics Service and Molecular Epidemiology Research Group. FISABIO-Public Health, and SeqCOVID-Spain Consortium |
| EPI_ISL_455733 | Servicio de Microbiologia. Hospital Clinico Universitario de Valencia | Sequencing and Bioinformatics Service and Molecular Epidemiology Research Group. FISABIO-Public Health, and SeqCOVID-Spain Consortium |
| EPI_ISL_455734 | Servicio de Microbiologia. Hospital Clinico Universitario de Valencia | Sequencing and Bioinformatics Service and Molecular Epidemiology Research Group. FISABIO-Public Health, and SeqCOVID-Spain Consortium |
| EPI_ISL_455735 | Servicio de Microbiologia. Hospital_Arnau_de_Vilanova | Sequencing and Bioinformatics Service and Molecular Epidemiology Research Group. FISABIO-Public Health, and SeqCOVID-Spain Consortium |

| EPI_Accession_ID | Originating Laboratory | Submitting Laboratory |
|---|---|---|
| EPI_ISL_455736 | Servicio de Microbiologia. Hospital_Arnau_de_Vilanova | Sequencing and Bioinformatics Service and Molecular Epidemiology Research Group. FISABIO-Public Health, and SeqCOVID-Spain Consortium |
| EPI_ISL_455737 | Servicio de Microbiologia. Hospital Clinico Universitario de Valencia | Sequencing and Bioinformatics Service and Molecular Epidemiology Research Group. FISABIO-Public Health, and SeqCOVID-Spain Consortium |
| EPI_ISL_455738 | Servicio de Microbiologia. Hospital Clinico Universitario de Valencia | Sequencing and Bioinformatics Service and Molecular Epidemiology Research Group. FISABIO-Public Health, and SeqCOVID-Spain Consortium |
| EPI_ISL_455739 | Servicio de Microbiologia. Hospital Clinico Universitario de Valencia | Sequencing and Bioinformatics Service and Molecular Epidemiology Research Group. FISABIO-Public Health, and SeqCOVID-Spain Consortium |
| EPI_ISL_455740 | Servicio de Microbiologia. Hospital Clinico Universitario de Valencia | Sequencing and Bioinformatics Service and Molecular Epidemiology Research Group. FISABIO-Public Health, and SeqCOVID-Spain Consortium |
| EPI_ISL_455741 | Servicio de Microbiologia. Hospital Clinico Universitario de Valencia | Sequencing and Bioinformatics Service and Molecular Epidemiology Research Group. FISABIO-Public Health, and SeqCOVID-Spain Consortium |
| EPI_ISL_455742 | Servicio de Microbiologia. Hospital Clinico Universitario de Valencia | Sequencing and Bioinformatics Service and Molecular Epidemiology Research Group. FISABIO-Public Health, and SeqCOVID-Spain Consortium |
| EPI_ISL_455743 | Servicio de Microbiologia. Hospital Clinico Universitario de Valencia | Sequencing and Bioinformatics Service and Molecular Epidemiology Research Group. FISABIO-Public Health, and SeqCOVID-Spain Consortium |
| EPI_ISL_455744 | Servicio de Microbiologia. Hospital Clinico Universitario de Valencia | Sequencing and Bioinformatics Service and Molecular Epidemiology Research Group. FISABIO-Public Health, and SeqCOVID-Spain Consortium |
| EPI_ISL_455749 | REGIONAL VRDL,ICMR-RMRC BBSR | Immunogenomics lab, Institute of Life Sciences, Bhubaneswar |
| EPI_ISL_455750 | REGIONAL VRDL,ICMR-RMRC BBSR | Immunogenomics lab, Institute of Life Sciences, Bhubaneswar |
| EPI_ISL_455751 | REGIONAL VRDL,ICMR-RMRC BBSR | Immunogenomics lab, Institute of Life Sciences, Bhubaneswar |

| EPI_Accession_ID | Originating Laboratory | Submitting Laboratory |
|---|---|---|
| EPI_ISL_455752 | REGIONAL VRDL,ICMR-RMRC BBSR | Immunogenomics lab, Institute of Life Sciences, Bhubaneswar |
| EPI_ISL_455753 | REGIONAL VRDL,ICMR-RMRC BBSR | Immunogenomics lab, Institute of Life Sciences, Bhubaneswar |
| EPI_ISL_455754 | REGIONAL VRDL,ICMR-RMRC BBSR | Immunogenomics lab, Institute of Life Sciences, Bhubaneswar |
| EPI_ISL_455755 | REGIONAL VRDL,ICMR-RMRC BBSR | Immunogenomics lab, Institute of Life Sciences, Bhubaneswar |
| EPI_ISL_455756 | REGIONAL VRDL,ICMR-RMRC BBSR | Immunogenomics lab, Institute of Life Sciences, Bhubaneswar |
| EPI_ISL_455757 | REGIONAL VRDL,ICMR-RMRC BBSR | Immunogenomics lab, Institute of Life Sciences, Bhubaneswar |
| EPI_ISL_455758 | REGIONAL VRDL,ICMR-RMRC BBSR | Immunogenomics lab, Institute of Life Sciences, Bhubaneswar |
| EPI_ISL_455759 | REGIONAL VRDL,ICMR-RMRC BBSR | Immunogenomics lab, Institute of Life Sciences, Bhubaneswar |
| EPI_ISL_455760 | REGIONAL VRDL,ICMR-RMRC BBSR | Immunogenomics lab, Institute of Life Sciences, Bhubaneswar |
| EPI_ISL_455761 | REGIONAL VRDL,ICMR-RMRC BBSR | Immunogenomics lab, Institute of Life Sciences, Bhubaneswar |
| EPI_ISL_455762 | REGIONAL VRDL,ICMR-RMRC BBSR | Immunogenomics lab, Institute of Life Sciences, Bhubaneswar |
| EPI_ISL_455763 | REGIONAL VRDL,ICMR-RMRC BBSR | Immunogenomics lab, Institute of Life Sciences, Bhubaneswar |
| EPI_ISL_455764 | REGIONAL VRDL,ICMR-RMRC BBSR | Immunogenomics lab, Institute of Life Sciences, Bhubaneswar |
| EPI_ISL_455765 | REGIONAL VRDL,ICMR-RMRC BBSR | Immunogenomics lab, Institute of Life Sciences, Bhubaneswar |
| EPI_ISL_455766 | REGIONAL VRDL,ICMR-RMRC BBSR | Immunogenomics lab, Institute of Life Sciences, Bhubaneswar |
| EPI_ISL_455767 | REGIONAL VRDL,ICMR-RMRC BBSR | Immunogenomics lab, Institute of Life Sciences, Bhubaneswar |

| EPI_Accession_ID | Originating Laboratory | Submitting Laboratory |
| --- | --- | --- |
| EPI_ISL_455768 | REGIONAL VRDL,ICMR-RMRC BBSR | Immunogenomics lab, Institute of Life Sciences, Bhubaneswar |
| EPI_ISL_455769 | REGIONAL VRDL,ICMR-RMRC BBSR | Immunogenomics lab, Institute of Life Sciences, Bhubaneswar |
| EPI_ISL_455770 | REGIONAL VRDL,ICMR-RMRC BBSR | Immunogenomics lab, Institute of Life Sciences, Bhubaneswar |
| EPI_ISL_455771 | REGIONAL VRDL,ICMR-RMRC BBSR | Immunogenomics lab, Institute of Life Sciences, Bhubaneswar |
| EPI_ISL_455772 | REGIONAL VRDL,ICMR-RMRC BBSR | Immunogenomics lab, Institute of Life Sciences, Bhubaneswar |
| EPI_ISL_455773 | REGIONAL VRDL,ICMR-RMRC BBSR | Immunogenomics lab, Institute of Life Sciences, Bhubaneswar |
| EPI_ISL_455774 | REGIONAL VRDL,ICMR-RMRC BBSR | Immunogenomics lab, Institute of Life Sciences, Bhubaneswar |
| EPI_ISL_455775 | REGIONAL VRDL,ICMR-RMRC BBSR | Immunogenomics lab, Institute of Life Sciences, Bhubaneswar |
| EPI_ISL_455776 | REGIONAL VRDL,ICMR-RMRC BBSR | Immunogenomics lab, Institute of Life Sciences, Bhubaneswar |
| EPI_ISL_455777 | REGIONAL VRDL,ICMR-RMRC BBSR | Immunogenomics lab, Institute of Life Sciences, Bhubaneswar |
| EPI_ISL_455778 | REGIONAL VRDL,ICMR-RMRC BBSR | Immunogenomics lab, Institute of Life Sciences, Bhubaneswar |
| EPI_ISL_455779 | REGIONAL VRDL,ICMR-RMRC BBSR | Immunogenomics lab, Institute of Life Sciences, Bhubaneswar |
| EPI_ISL_455780 | REGIONAL VRDL,ICMR-RMRC BBSR | Immunogenomics lab, Institute of Life Sciences, Bhubaneswar |
| EPI_ISL_455782 | REGIONAL VRDL,ICMR-RMRC BBSR | Immunogenomics lab, Institute of Life Sciences, Bhubaneswar |
| EPI_ISL_455783 | REGIONAL VRDL,ICMR-RMRC BBSR | Immunogenomics lab, Institute of Life Sciences, Bhubaneswar |
| EPI_ISL_455784 | REGIONAL VRDL,ICMR-RMRC BBSR | Immunogenomics lab, Institute of Life Sciences, Bhubaneswar |

| EPI_Accession_ID | Originating Laboratory | Submitting Laboratory |
|---|---|---|
| EPI_ISL_455786 | REGIONAL VRDL,ICMR-RMRC BBSR | Immunogenomics lab, Institute of Life Sciences, Bhubaneswar |
| EPI_ISL_455787 | REGIONAL VRDL,ICMR-RMRC BBSR | Immunogenomics lab, Institute of Life Sciences, Bhubaneswar |
| EPI_ISL_455790 | Institute for Medical Research, Infectious Disease Research Centre, National Institutes of Health, Ministry of Health Malaysia | Malaysia Genome Institute |
| EPI_ISL_455791 | Institute for Medical Research, Infectious Disease Research Centre, National Institutes of Health, Ministry of Health Malaysia | Malaysia Genome Institute |
| EPI_ISL_455792 | Institute for Medical Research, Infectious Disease Research Centre, National Institutes of Health, Ministry of Health Malaysia | Malaysia Genome Institute |
| EPI_ISL_455793 | Institute for Medical Research, Infectious Disease Research Centre, National Institutes of Health, Ministry of Health Malaysia | Malaysia Genome Institute |
| EPI_ISL_455840 | Orebro klinisk mikrobiologi | The Public Health Agency of Sweden |
| EPI_ISL_455841 | Orebro klinisk mikrobiologi | The Public Health Agency of Sweden |
| EPI_ISL_455842 | Skovde/Unilabs | The Public Health Agency of Sweden |
| EPI_ISL_455843 | Skovde/Unilabs | The Public Health Agency of Sweden |
| EPI_ISL_455844 | Skovde/Unilabs | The Public Health Agency of Sweden |
| EPI_ISL_455845 | Skovde/Unilabs | The Public Health Agency of Sweden |
| EPI_ISL_455846 | Karolinska Universitetslaboratoriet | The Public Health Agency of Sweden |
| EPI_ISL_455847 | Karolinska Universitetslaboratoriet | The Public Health Agency of Sweden |
| EPI_ISL_455848 | Karolinska Universitetslaboratoriet | The Public Health Agency of Sweden |
| EPI_ISL_455849 | Karolinska Universitetslaboratoriet | The Public Health Agency of Sweden |
| EPI_ISL_455850 | Karolinska Universitetslaboratoriet | The Public Health Agency of Sweden |
| EPI_ISL_455851 | Karolinska Universitetslaboratoriet | The Public Health Agency of Sweden |
| EPI_ISL_455852 | Karolinska Universitetslaboratoriet | The Public Health Agency of Sweden |
| EPI_ISL_455853 | Karolinska Universitetslaboratoriet | The Public Health Agency of Sweden |
| EPI_ISL_455854 | Karolinska Universitetslaboratoriet | The Public Health Agency of Sweden |
| EPI_ISL_455855 | Karolinska Universitetslaboratoriet | The Public Health Agency of Sweden |
| EPI_ISL_455856 | Karolinska Universitetslaboratoriet | The Public Health Agency of Sweden |

| EPI_Accession_ID | Originating Laboratory | Submitting Laboratory |
|---|---|---|
| EPI_ISL_455857 | Karolinska Universitetslaboratoriet | The Public Health Agency of Sweden |
| EPI_ISL_455858 | Karolinska Universitetslaboratoriet | The Public Health Agency of Sweden |
| EPI_ISL_455859 | Karolinska Universitetslaboratoriet | The Public Health Agency of Sweden |
| EPI_ISL_455860 | Karolinska Universitetslaboratoriet | The Public Health Agency of Sweden |
| EPI_ISL_455861 | Karolinska Universitetslaboratoriet | The Public Health Agency of Sweden |
| EPI_ISL_455862 | Karolinska Universitetslaboratoriet | The Public Health Agency of Sweden |
| EPI_ISL_455863 | Karolinska Universitetslaboratoriet | The Public Health Agency of Sweden |
| EPI_ISL_455864 | Karolinska Universitetslaboratoriet | The Public Health Agency of Sweden |
| EPI_ISL_455865 | Karolinska Universitetslaboratoriet | The Public Health Agency of Sweden |
| EPI_ISL_455866 | Karolinska Universitetslaboratoriet | The Public Health Agency of Sweden |
| EPI_ISL_455867 | Karolinska Universitetslaboratoriet | The Public Health Agency of Sweden |
| EPI_ISL_455868 | Karolinska Universitetslaboratoriet | The Public Health Agency of Sweden |
| EPI_ISL_455869 | Karolinska Universitetslaboratoriet | The Public Health Agency of Sweden |
| EPI_ISL_455870 | Karolinska Universitetslaboratoriet | The Public Health Agency of Sweden |
| EPI_ISL_455871 | Karolinska Universitetslaboratoriet | The Public Health Agency of Sweden |
| EPI_ISL_455872 | Karolinska Universitetslaboratoriet | The Public Health Agency of Sweden |
| EPI_ISL_455873 | Karolinska Universitetslaboratoriet | The Public Health Agency of Sweden |
| EPI_ISL_455874 | Karolinska Universitetslaboratoriet | The Public Health Agency of Sweden |
| EPI_ISL_455875 | Karolinska Universitetslaboratoriet | The Public Health Agency of Sweden |
| EPI_ISL_455876 | Karolinska Universitetslaboratoriet | The Public Health Agency of Sweden |
| EPI_ISL_455877 | Karolinska Universitetslaboratoriet | The Public Health Agency of Sweden |
| EPI_ISL_455878 | Karolinska Universitetslaboratoriet | The Public Health Agency of Sweden |
| EPI_ISL_455879 | Karolinska Universitetslaboratoriet | The Public Health Agency of Sweden |
| EPI_ISL_455880 | Karolinska Universitetslaboratoriet | The Public Health Agency of Sweden |
| EPI_ISL_455881 | Karolinska Universitetslaboratoriet | The Public Health Agency of Sweden |
| EPI_ISL_455882 | Karolinska Universitetslaboratoriet | The Public Health Agency of Sweden |
| EPI_ISL_455883 | Karolinska Universitetslaboratoriet | The Public Health Agency of Sweden |
| EPI_ISL_455884 | Karolinska Universitetslaboratoriet | The Public Health Agency of Sweden |
| EPI_ISL_455885 | Karolinska Universitetslaboratoriet | The Public Health Agency of Sweden |
| EPI_ISL_455886 | Karolinska Universitetslaboratoriet | The Public Health Agency of Sweden |
| EPI_ISL_455887 | Karolinska Universitetslaboratoriet | The Public Health Agency of Sweden |
| EPI_ISL_455888 | Karolinska Universitetslaboratoriet | The Public Health Agency of Sweden |
| EPI_ISL_455889 | Karolinska Universitetslaboratoriet | The Public Health Agency of Sweden |

| EPI_Accession_ID | Originating Laboratory | Submitting Laboratory |
|---|---|---|
| EPI_ISL_455890 | Karolinska Universitetslaboratoriet | The Public Health Agency of Sweden |
| EPI_ISL_455891 | Karolinska Universitetslaboratoriet | The Public Health Agency of Sweden |
| EPI_ISL_455892 | Karolinska Universitetslaboratoriet | The Public Health Agency of Sweden |
| EPI_ISL_455893 | Karolinska Universitetslaboratoriet | The Public Health Agency of Sweden |
| EPI_ISL_455894 | Karolinska Universitetslaboratoriet | The Public Health Agency of Sweden |
| EPI_ISL_455895 | Karolinska Universitetslaboratoriet | The Public Health Agency of Sweden |
| EPI_ISL_455896 | Karolinska Universitetslaboratoriet | The Public Health Agency of Sweden |
| EPI_ISL_455897 | Karolinska Universitetslaboratoriet | The Public Health Agency of Sweden |
| EPI_ISL_455898 | Karolinska Universitetslaboratoriet | The Public Health Agency of Sweden |
| EPI_ISL_455899 | Karolinska Universitetslaboratoriet | The Public Health Agency of Sweden |
| EPI_ISL_455900 | Karolinska Universitetslaboratoriet | The Public Health Agency of Sweden |
| EPI_ISL_455901 | Karolinska Universitetslaboratoriet | The Public Health Agency of Sweden |
| EPI_ISL_455902 | Halmstad klinisk mikrobiologi | The Public Health Agency of Sweden |
| EPI_ISL_455903 | Folkhalsomyndigheten | The Public Health Agency of Sweden |
| EPI_ISL_455904 | Klinisk mikrobiologi, UAS | The Public Health Agency of Sweden |
| EPI_ISL_455905 | Klinisk mikrobiologi, UAS | The Public Health Agency of Sweden |
| EPI_ISL_455906 | Klinisk mikrobiologi, UAS | The Public Health Agency of Sweden |
| EPI_ISL_455907 | Klinisk mikrobiologi, UAS | The Public Health Agency of Sweden |
| EPI_ISL_455908 | Ramathibodi Hospital | COVID-19 Network Investigations (CONI) Alliance |
| EPI_ISL_455909 | Ramathibodi Hospital | COVID-19 Network Investigations (CONI) Alliance |
| EPI_ISL_455910 | Ramathibodi Hospital | COVID-19 Network Investigations (CONI) Alliance |
| EPI_ISL_455911 | Ramathibodi Hospital | COVID-19 Network Investigations (CONI) Alliance |
| EPI_ISL_455912 | Ramathibodi Hospital | COVID-19 Network Investigations (CONI) Alliance |
| EPI_ISL_455913 | Ramathibodi Hospital | COVID-19 Network Investigations (CONI) Alliance |
| EPI_ISL_455914 | Ramathibodi Hospital | COVID-19 Network Investigations (CONI) Alliance |
| EPI_ISL_455915 | Ramathibodi Hospital | COVID-19 Network Investigations (CONI) Alliance |
| EPI_ISL_455916 | Ramathibodi Hospital | COVID-19 Network Investigations (CONI) Alliance |
| EPI_ISL_455917 | Ramathibodi Hospital | COVID-19 Network Investigations (CONI) Alliance |
| EPI_ISL_455918 | Ramathibodi Hospital | COVID-19 Network Investigations (CONI) Alliance |
| EPI_ISL_455919 | Ramathibodi Hospital | COVID-19 Network Investigations (CONI) Alliance |
| EPI_ISL_455920 | Ramathibodi Hospital | COVID-19 Network Investigations (CONI) Alliance |
| EPI_ISL_455921 | Ramathibodi Hospital | COVID-19 Network Investigations (CONI) Alliance |
| EPI_ISL_455922 | Ramathibodi Hospital | COVID-19 Network Investigations (CONI) Alliance |

| EPI_Accession_ID | Originating Laboratory | Submitting Laboratory |
|---|---|---|
| EPI_ISL_455923 | Ramathibodi Hospital | COVID-19 Network Investigations (CONI) Alliance |
| EPI_ISL_455924 | Ramathibodi Hospital | COVID-19 Network Investigations (CONI) Alliance |
| EPI_ISL_455925 | Ramathibodi Hospital | COVID-19 Network Investigations (CONI) Alliance |
| EPI_ISL_455926 | Ramathibodi Hospital | COVID-19 Network Investigations (CONI) Alliance |
| EPI_ISL_455927 | Ramathibodi Hospital | COVID-19 Network Investigations (CONI) Alliance |
| EPI_ISL_455928 | Ramathibodi Hospital | COVID-19 Network Investigations (CONI) Alliance |
| EPI_ISL_455929 | Ramathibodi Hospital | COVID-19 Network Investigations (CONI) Alliance |
| EPI_ISL_455930 | Ramathibodi Hospital | COVID-19 Network Investigations (CONI) Alliance |
| EPI_ISL_455931 | Ramathibodi Hospital | COVID-19 Network Investigations (CONI) Alliance |
| EPI_ISL_455932 | Ramathibodi Hospital | COVID-19 Network Investigations (CONI) Alliance |
| EPI_ISL_455933 | Ramathibodi Hospital | COVID-19 Network Investigations (CONI) Alliance |
| EPI_ISL_455934 | Ramathibodi Hospital | COVID-19 Network Investigations (CONI) Alliance |
| EPI_ISL_455935 | Ramathibodi Hospital | COVID-19 Network Investigations (CONI) Alliance |
| EPI_ISL_455936 | Ramathibodi Hospital | COVID-19 Network Investigations (CONI) Alliance |
| EPI_ISL_455937 | Ramathibodi Hospital | COVID-19 Network Investigations (CONI) Alliance |
| EPI_ISL_455938 | Ramathibodi Hospital | COVID-19 Network Investigations (CONI) Alliance |
| EPI_ISL_455939 | Ramathibodi Hospital | COVID-19 Network Investigations (CONI) Alliance |
| EPI_ISL_455940 | Ramathibodi Hospital | COVID-19 Network Investigations (CONI) Alliance |
| EPI_ISL_455941 | Ramathibodi Hospital | COVID-19 Network Investigations (CONI) Alliance |
| EPI_ISL_455942 | Ramathibodi Hospital | COVID-19 Network Investigations (CONI) Alliance |
| EPI_ISL_455943 | Ramathibodi Hospital | COVID-19 Network Investigations (CONI) Alliance |
| EPI_ISL_455944 | Ramathibodi Hospital | COVID-19 Network Investigations (CONI) Alliance |
| EPI_ISL_455945 | Ramathibodi Hospital | COVID-19 Network Investigations (CONI) Alliance |
| EPI_ISL_455946 | Ramathibodi Hospital | COVID-19 Network Investigations (CONI) Alliance |
| EPI_ISL_455947 | Ramathibodi Hospital | COVID-19 Network Investigations (CONI) Alliance |
| EPI_ISL_455948 | Ramathibodi Hospital | COVID-19 Network Investigations (CONI) Alliance |
| EPI_ISL_455958 | Department of Clinical Microbiology | GIGA Medical Genomics |
| EPI_ISL_455959 | Department of Clinical Microbiology | GIGA Medical Genomics |
| EPI_ISL_455960 | Department of Clinical Microbiology | GIGA Medical Genomics |
| EPI_ISL_455961 | Department of Clinical Microbiology | GIGA Medical Genomics |
| EPI_ISL_455962 | Department of Clinical Microbiology | GIGA Medical Genomics |
| EPI_ISL_455963 | Department of Clinical Microbiology | GIGA Medical Genomics |
| EPI_ISL_455964 | Department of Clinical Microbiology | GIGA Medical Genomics |

| EPI_Accession_ID | Originating Laboratory | Submitting Laboratory |
|---|---|---|
| EPI_ISL_455965 | Department of Clinical Microbiology | GIGA Medical Genomics |
| EPI_ISL_455966 | Department of Clinical Microbiology | GIGA Medical Genomics |
| EPI_ISL_455967 | Department of Clinical Microbiology | GIGA Medical Genomics |
| EPI_ISL_455968 | Department of Clinical Microbiology | GIGA Medical Genomics |
| EPI_ISL_455969 | Department of Clinical Microbiology | GIGA Medical Genomics |
| EPI_ISL_455970 | Department of Clinical Microbiology | GIGA Medical Genomics |
| EPI_ISL_455971 | Department of Clinical Microbiology | GIGA Medical Genomics |
| EPI_ISL_455972 | Department of Clinical Microbiology | GIGA Medical Genomics |
| EPI_ISL_455973 | Department of Clinical Microbiology | GIGA Medical Genomics |
| EPI_ISL_455974 | Department of Clinical Microbiology | GIGA Medical Genomics |
| EPI_ISL_455975 | Department of Clinical Microbiology | GIGA Medical Genomics |
| EPI_ISL_455976 | Department of Clinical Microbiology | GIGA Medical Genomics |
| EPI_ISL_455977 | Department of Clinical Microbiology | GIGA Medical Genomics |
| EPI_ISL_455978 | Department of Clinical Microbiology | GIGA Medical Genomics |
| EPI_ISL_455979 | Department of Clinical Microbiology | GIGA Medical Genomics |
| EPI_ISL_455980 | LSUHS Emerging Viral Threat Laboratory | Microbial Genome Sequencing Center |
| EPI_ISL_455981 | LSUHS Emerging Viral Threat Laboratory | Microbial Genome Sequencing Center |
| EPI_ISL_455982 | LSUHS Emerging Viral Threat Laboratory | Microbial Genome Sequencing Center |
| EPI_ISL_455983 | LSUHS Emerging Viral Threat Laboratory | Microbial Genome Sequencing Center |
| EPI_ISL_455984 | LSUHS Emerging Viral Threat Laboratory | Microbial Genome Sequencing Center |
| EPI_ISL_455985 | LSUHS Emerging Viral Threat Laboratory | Microbial Genome Sequencing Center |
| EPI_ISL_455986 | LSUHS Emerging Viral Threat Laboratory | Microbial Genome Sequencing Center |
| EPI_ISL_455987 | LSUHS Emerging Viral Threat Laboratory | Microbial Genome Sequencing Center |
| EPI_ISL_455988 | LSUHS Emerging Viral Threat Laboratory | Microbial Genome Sequencing Center |
| EPI_ISL_455989 | LSUHS Emerging Viral Threat Laboratory | Microbial Genome Sequencing Center |
| EPI_ISL_455990 | LSUHS Emerging Viral Threat Laboratory | Microbial Genome Sequencing Center |
| EPI_ISL_455991 | LSUHS Emerging Viral Threat Laboratory | Microbial Genome Sequencing Center |
| EPI_ISL_455992 | LSUHS Emerging Viral Threat Laboratory | Microbial Genome Sequencing Center |
| EPI_ISL_455993 | LSUHS Emerging Viral Threat Laboratory | Microbial Genome Sequencing Center |
| EPI_ISL_455994 | LSUHS Emerging Viral Threat Laboratory | Microbial Genome Sequencing Center |
| EPI_ISL_455995 | LSUHS Emerging Viral Threat Laboratory | Microbial Genome Sequencing Center |
| EPI_ISL_455996 | LSUHS Emerging Viral Threat Laboratory | Microbial Genome Sequencing Center |
| EPI_ISL_455997 | LSUHS Emerging Viral Threat Laboratory | Microbial Genome Sequencing Center |

| EPI_Accession_ID | Originating Laboratory | Submitting Laboratory |
|---|---|---|
| EPI_ISL_455998 | LSUHS Emerging Viral Threat Laboratory | Microbial Genome Sequencing Center |
| EPI_ISL_455999 | LSUHS Emerging Viral Threat Laboratory | Microbial Genome Sequencing Center |
| EPI_ISL_456000 | LSUHS Emerging Viral Threat Laboratory | Microbial Genome Sequencing Center |
| EPI_ISL_456001 | LSUHS Emerging Viral Threat Laboratory | Microbial Genome Sequencing Center |
| EPI_ISL_456002 | LSUHS Emerging Viral Threat Laboratory | Microbial Genome Sequencing Center |
| EPI_ISL_456003 | NYU Langone Health | Departments of Pathology and Medicine, New York University School of Medicine |
| EPI_ISL_456004 | NYU Langone Health | Departments of Pathology and Medicine, New York University School of Medicine |
| EPI_ISL_456005 | NYU Langone Health | Departments of Pathology and Medicine, New York University School of Medicine |
| EPI_ISL_456006 | NYU Langone Health | Departments of Pathology and Medicine, New York University School of Medicine |
| EPI_ISL_456007 | NYU Langone Health | Departments of Pathology and Medicine, New York University School of Medicine |
| EPI_ISL_456008 | NYU Langone Health | Departments of Pathology and Medicine, New York University School of Medicine |
| EPI_ISL_456009 | NYU Langone Health | Departments of Pathology and Medicine, New York University School of Medicine |
| EPI_ISL_456010 | NYU Langone Health | Departments of Pathology and Medicine, New York University School of Medicine |
| EPI_ISL_456011 | NYU Langone Health | Departments of Pathology and Medicine, New York University School of Medicine |
| EPI_ISL_456012 | NYU Langone Health | Departments of Pathology and Medicine, New York University School of Medicine |
| EPI_ISL_456013 | NYU Langone Health | Departments of Pathology and Medicine, New York University School of Medicine |
| EPI_ISL_456014 | NYU Langone Health | Departments of Pathology and Medicine, New York University School of Medicine |
| EPI_ISL_456015 | NYU Langone Health | Departments of Pathology and Medicine, New York University School of Medicine |
| EPI_ISL_456016 | NYU Langone Health | Departments of Pathology and Medicine, New York University School of Medicine |

| EPI_Accession_ID | Originating Laboratory | Submitting Laboratory |
|---|---|---|
| EPI_ISL_456017 | NYU Langone Health | Departments of Pathology and Medicine, New York University School of Medicine |
| EPI_ISL_456018 | NYU Langone Health | Departments of Pathology and Medicine, New York University School of Medicine |
| EPI_ISL_456019 | NYU Langone Health | Departments of Pathology and Medicine, New York University School of Medicine |
| EPI_ISL_456020 | NYU Langone Health | Departments of Pathology and Medicine, New York University School of Medicine |
| EPI_ISL_456021 | NYU Langone Health | Departments of Pathology and Medicine, New York University School of Medicine |
| EPI_ISL_456022 | NYU Langone Health | Departments of Pathology and Medicine, New York University School of Medicine |
| EPI_ISL_456023 | NYU Langone Health | Departments of Pathology and Medicine, New York University School of Medicine |
| EPI_ISL_456024 | NYU Langone Health | Departments of Pathology and Medicine, New York University School of Medicine |
| EPI_ISL_456025 | NYU Langone Health | Departments of Pathology and Medicine, New York University School of Medicine |
| EPI_ISL_456026 | NYU Langone Health | Departments of Pathology and Medicine, New York University School of Medicine |
| EPI_ISL_456027 | NYU Langone Health | Departments of Pathology and Medicine, New York University School of Medicine |
| EPI_ISL_456028 | NYU Langone Health | Departments of Pathology and Medicine, New York University School of Medicine |
| EPI_ISL_456029 | NYU Langone Health | Departments of Pathology and Medicine, New York University School of Medicine |
| EPI_ISL_456030 | NYU Langone Health | Departments of Pathology and Medicine, New York University School of Medicine |
| EPI_ISL_456031 | NYU Langone Health | Departments of Pathology and Medicine, New York University School of Medicine |
| EPI_ISL_456032 | NYU Langone Health | Departments of Pathology and Medicine, New York University School of Medicine |

| EPI_Accession_ID | Originating Laboratory | Submitting Laboratory |
|---|---|---|
| EPI_ISL_456033 | NYU Langone Health | Departments of Pathology and Medicine, New York University School of Medicine |
| EPI_ISL_456034 | NYU Langone Health | Departments of Pathology and Medicine, New York University School of Medicine |
| EPI_ISL_456035 | NYU Langone Health | Departments of Pathology and Medicine, New York University School of Medicine |
| EPI_ISL_456036 | NYU Langone Health | Departments of Pathology and Medicine, New York University School of Medicine |
| EPI_ISL_456037 | NYU Langone Health | Departments of Pathology and Medicine, New York University School of Medicine |
| EPI_ISL_456038 | NYU Langone Health | Departments of Pathology and Medicine, New York University School of Medicine |
| EPI_ISL_456039 | NYU Langone Health | Departments of Pathology and Medicine, New York University School of Medicine |
| EPI_ISL_456040 | NYU Langone Health | Departments of Pathology and Medicine, New York University School of Medicine |
| EPI_ISL_456041 | NYU Langone Health | Departments of Pathology and Medicine, New York University School of Medicine |
| EPI_ISL_456042 | NYU Langone Health | Departments of Pathology and Medicine, New York University School of Medicine |
| EPI_ISL_456043 | NYU Langone Health | Departments of Pathology and Medicine, New York University School of Medicine |
| EPI_ISL_456044 | NYU Langone Health | Departments of Pathology and Medicine, New York University School of Medicine |
| EPI_ISL_456045 | NYU Langone Health | Departments of Pathology and Medicine, New York University School of Medicine |
| EPI_ISL_456046 | NYU Langone Health | Departments of Pathology and Medicine, New York University School of Medicine |
| EPI_ISL_456047 | NYU Langone Health | Departments of Pathology and Medicine, New York University School of Medicine |
| EPI_ISL_456048 | NYU Langone Health | Departments of Pathology and Medicine, New York University School of Medicine |

| EPI_Accession_ID | Originating Laboratory | Submitting Laboratory |
|---|---|---|
| EPI_ISL_456049 | NYU Langone Health | Departments of Pathology and Medicine, New York University School of Medicine |
| EPI_ISL_456050 | NYU Langone Health | Departments of Pathology and Medicine, New York University School of Medicine |
| EPI_ISL_456051 | NYU Langone Health | Departments of Pathology and Medicine, New York University School of Medicine |
| EPI_ISL_456052 | NYU Langone Health | Departments of Pathology and Medicine, New York University School of Medicine |
| EPI_ISL_456053 | NYU Langone Health | Departments of Pathology and Medicine, New York University School of Medicine |
| EPI_ISL_456054 | NYU Langone Health | Departments of Pathology and Medicine, New York University School of Medicine |
| EPI_ISL_456055 | NYU Langone Health | Departments of Pathology and Medicine, New York University School of Medicine |
| EPI_ISL_456056 | NYU Langone Health | Departments of Pathology and Medicine, New York University School of Medicine |
| EPI_ISL_456057 | NYU Langone Health | Departments of Pathology and Medicine, New York University School of Medicine |
| EPI_ISL_456058 | NYU Langone Health | Departments of Pathology and Medicine, New York University School of Medicine |
| EPI_ISL_456059 | NYU Langone Health | Departments of Pathology and Medicine, New York University School of Medicine |
| EPI_ISL_456060 | NYU Langone Health | Departments of Pathology and Medicine, New York University School of Medicine |
| EPI_ISL_456061 | NYU Langone Health | Departments of Pathology and Medicine, New York University School of Medicine |
| EPI_ISL_456062 | NYU Langone Health | Departments of Pathology and Medicine, New York University School of Medicine |
| EPI_ISL_456063 | NYU Langone Health | Departments of Pathology and Medicine, New York University School of Medicine |
| EPI_ISL_456064 | NYU Langone Health | Departments of Pathology and Medicine, New York University School of Medicine |

| EPI_Accession_ID | Originating Laboratory | Submitting Laboratory |
|---|---|---|
| EPI_ISL_456065 | NYU Langone Health | Departments of Pathology and Medicine, New York University School of Medicine |
| EPI_ISL_456066 | NYU Langone Health | Departments of Pathology and Medicine, New York University School of Medicine |
| EPI_ISL_456067 | NYU Langone Health | Departments of Pathology and Medicine, New York University School of Medicine |
| EPI_ISL_456068 | NYU Langone Health | Departments of Pathology and Medicine, New York University School of Medicine |
| EPI_ISL_456069 | NYU Langone Health | Departments of Pathology and Medicine, New York University School of Medicine |
| EPI_ISL_456071 | Laboratory of Respiratory Viruses and Measles, Oswaldo Cruz Institute, FIOCRUZ | Laboratory of Respiratory Viruses and Measles, Oswaldo Cruz Institute, FIOCRUZ |
| EPI_ISL_456072 | Laboratory of Respiratory Viruses and Measles, Oswaldo Cruz Institute, FIOCRUZ | Laboratory of Respiratory Viruses and Measles, Oswaldo Cruz Institute, FIOCRUZ |
| EPI_ISL_456073 | Laboratory of Respiratory Viruses and Measles, Oswaldo Cruz Institute, FIOCRUZ | Laboratory of Respiratory Viruses and Measles, Oswaldo Cruz Institute, FIOCRUZ |
| EPI_ISL_456074 | Laboratory of Respiratory Viruses and Measles, Oswaldo Cruz Institute, FIOCRUZ | Laboratory of Respiratory Viruses and Measles, Oswaldo Cruz Institute, FIOCRUZ |
| EPI_ISL_456075 | Laboratory of Respiratory Viruses and Measles, Oswaldo Cruz Institute, FIOCRUZ | Laboratory of Respiratory Viruses and Measles, Oswaldo Cruz Institute, FIOCRUZ |
| EPI_ISL_456076 | LACEN RJ - LaboratÃ³rio Central de SaÃºde PÃºblica Noel Nutels | Laboratory of Respiratory Viruses and Measles, Oswaldo Cruz Institute, FIOCRUZ |
| EPI_ISL_456077 | LACEN RJ - LaboratÃ³rio Central de SaÃºde PÃºblica Noel Nutels | Laboratory of Respiratory Viruses and Measles, Oswaldo Cruz Institute, FIOCRUZ |
| EPI_ISL_456079 | Laboratory of Respiratory Viruses and Measles, Oswaldo Cruz Institute, FIOCRUZ | Laboratory of Respiratory Viruses and Measles, Oswaldo Cruz Institute, FIOCRUZ |
| EPI_ISL_456080 | Laboratory of Respiratory Viruses and Measles, Oswaldo Cruz Institute, FIOCRUZ | Laboratory of Respiratory Viruses and Measles, Oswaldo Cruz Institute, FIOCRUZ |
| EPI_ISL_456081 | Laboratory of Respiratory Viruses and Measles, Oswaldo Cruz Institute, FIOCRUZ | Laboratory of Respiratory Viruses and Measles, Oswaldo Cruz Institute, FIOCRUZ |
| EPI_ISL_456082 | LACEN RJ - LaboratÃ³rio Central de SaÃºde PÃºblica Noel Nutels | Laboratory of Respiratory Viruses and Measles, Oswaldo Cruz Institute, FIOCRUZ |

| EPI_Accession_ID | Originating Laboratory | Submitting Laboratory |
|---|---|---|
| EPI_ISL_456083 | LACEN RJ - LaboratÃ³rio Central de SaÃºde PÃºblica Noel Nutels | Laboratory of Respiratory Viruses and Measles, Oswaldo Cruz Institute, FIOCRUZ |
| EPI_ISL_456084 | Laboratory of Respiratory Viruses and Measles, Oswaldo Cruz Institute, FIOCRUZ | Laboratory of Respiratory Viruses and Measles, Oswaldo Cruz Institute, FIOCRUZ |
| EPI_ISL_456085 | Laboratory of Respiratory Viruses and Measles, Oswaldo Cruz Institute, FIOCRUZ | Laboratory of Respiratory Viruses and Measles, Oswaldo Cruz Institute, FIOCRUZ |
| EPI_ISL_456086 | Laboratory of Respiratory Viruses and Measles, Oswaldo Cruz Institute, FIOCRUZ | Laboratory of Respiratory Viruses and Measles, Oswaldo Cruz Institute, FIOCRUZ |
| EPI_ISL_456087 | Laboratory of Respiratory Viruses and Measles, Oswaldo Cruz Institute, FIOCRUZ | Laboratory of Respiratory Viruses and Measles, Oswaldo Cruz Institute, FIOCRUZ |
| EPI_ISL_456088 | LACEN RJ - LaboratÃ³rio Central de SaÃºde PÃºblica Noel Nutels | Laboratory of Respiratory Viruses and Measles, Oswaldo Cruz Institute, FIOCRUZ |
| EPI_ISL_456089 | Laboratory of Respiratory Viruses and Measles, Oswaldo Cruz Institute, FIOCRUZ | Laboratory of Respiratory Viruses and Measles, Oswaldo Cruz Institute, FIOCRUZ |
| EPI_ISL_456090 | Laboratory of Respiratory Viruses and Measles, Oswaldo Cruz Institute, FIOCRUZ | Laboratory of Respiratory Viruses and Measles, Oswaldo Cruz Institute, FIOCRUZ |
| EPI_ISL_456091 | Laboratory of Respiratory Viruses and Measles, Oswaldo Cruz Institute, FIOCRUZ | Laboratory of Respiratory Viruses and Measles, Oswaldo Cruz Institute, FIOCRUZ |
| EPI_ISL_456092 | Laboratory of Respiratory Viruses and Measles, Oswaldo Cruz Institute, FIOCRUZ | Laboratory of Respiratory Viruses and Measles, Oswaldo Cruz Institute, FIOCRUZ |
| EPI_ISL_456093 | Laboratory of Respiratory Viruses and Measles, Oswaldo Cruz Institute, FIOCRUZ | Laboratory of Respiratory Viruses and Measles, Oswaldo Cruz Institute, FIOCRUZ |
| EPI_ISL_456094 | Laboratory of Respiratory Viruses and Measles, Oswaldo Cruz Institute, FIOCRUZ | Laboratory of Respiratory Viruses and Measles, Oswaldo Cruz Institute, FIOCRUZ |
| EPI_ISL_456095 | Laboratory of Respiratory Viruses and Measles, Oswaldo Cruz Institute, FIOCRUZ | Laboratory of Respiratory Viruses and Measles, Oswaldo Cruz Institute, FIOCRUZ |
| EPI_ISL_456096 | Laboratory of Respiratory Viruses and Measles, Oswaldo Cruz Institute, FIOCRUZ | Laboratory of Respiratory Viruses and Measles, Oswaldo Cruz Institute, FIOCRUZ |
| EPI_ISL_456097 | Laboratory of Respiratory Viruses and Measles, Oswaldo Cruz Institute, FIOCRUZ | Laboratory of Respiratory Viruses and Measles, Oswaldo Cruz Institute, FIOCRUZ |
| EPI_ISL_456098 | Laboratory of Respiratory Viruses and Measles, Oswaldo Cruz Institute, FIOCRUZ | Laboratory of Respiratory Viruses and Measles, Oswaldo Cruz Institute, FIOCRUZ |

| EPI_Accession_ID | Originating Laboratory | Submitting Laboratory |
|---|---|---|
| EPI_ISL_456099 | Laboratory of Respiratory Viruses and Measles, Oswaldo Cruz Institute, FIOCRUZ | Laboratory of Respiratory Viruses and Measles, Oswaldo Cruz Institute, FIOCRUZ |
| EPI_ISL_456100 | Laboratory of Respiratory Viruses and Measles, Oswaldo Cruz Institute, FIOCRUZ | Laboratory of Respiratory Viruses and Measles, Oswaldo Cruz Institute, FIOCRUZ |
| EPI_ISL_456101 | Laboratory of Respiratory Viruses and Measles, Oswaldo Cruz Institute, FIOCRUZ | Laboratory of Respiratory Viruses and Measles, Oswaldo Cruz Institute, FIOCRUZ |
| EPI_ISL_456102 | Laboratory of Respiratory Viruses and Measles, Oswaldo Cruz Institute, FIOCRUZ | Laboratory of Respiratory Viruses and Measles, Oswaldo Cruz Institute, FIOCRUZ |
| EPI_ISL_456103 | Laboratory of Respiratory Viruses and Measles, Oswaldo Cruz Institute, FIOCRUZ | Laboratory of Respiratory Viruses and Measles, Oswaldo Cruz Institute, FIOCRUZ |
| EPI_ISL_456104 | Laboratory of Respiratory Viruses and Measles, Oswaldo Cruz Institute, FIOCRUZ | Laboratory of Respiratory Viruses and Measles, Oswaldo Cruz Institute, FIOCRUZ |
| EPI_ISL_456105 | Laboratory of Respiratory Viruses and Measles, Oswaldo Cruz Institute, FIOCRUZ | Laboratory of Respiratory Viruses and Measles, Oswaldo Cruz Institute, FIOCRUZ |
| EPI_ISL_456106 | Laboratory of Respiratory Viruses and Measles, Oswaldo Cruz Institute, FIOCRUZ | Laboratory of Respiratory Viruses and Measles, Oswaldo Cruz Institute, FIOCRUZ |
| EPI_ISL_456107 | NYU Langone Health | Departments of Pathology and Medicine, New York University School of Medicine |
| EPI_ISL_456108 | NYU Langone Health | Departments of Pathology and Medicine, New York University School of Medicine |
| EPI_ISL_456109 | NYU Langone Health | Departments of Pathology and Medicine, New York University School of Medicine |
| EPI_ISL_456111 | NYU Langone Health | Departments of Pathology and Medicine, New York University School of Medicine |
| EPI_ISL_456112 | NYU Langone Health | Departments of Pathology and Medicine, New York University School of Medicine |
| EPI_ISL_456113 | NYU Langone Health | Departments of Pathology and Medicine, New York University School of Medicine |
| EPI_ISL_456114 | NYU Langone Health | Departments of Pathology and Medicine, New York University School of Medicine |

| EPI_Accession_ID | Originating Laboratory | Submitting Laboratory |
|---|---|---|
| EPI_ISL_456116 | Instituto Nacional de Salud - Unidad de SecuenciaciÃ³n y AnÃ¡lisis GenÃ³mico | Instituto Nacional de Salud, Universidad Cooperativa de Colombia, Instituto Alexander von Humboldt, Imperial College-London, London School of Hygiene & Tropical Medicine |
| EPI_ISL_456117 | Instituto Nacional de Salud - Unidad de SecuenciaciÃ³n y AnÃ¡lisis GenÃ³mico | Instituto Nacional de Salud, Universidad Cooperativa de Colombia, Instituto Alexander von Humboldt, Imperial College-London, London School of Hygiene & Tropical Medicine |
| EPI_ISL_456118 | Instituto Nacional de Salud - Unidad de SecuenciaciÃ³n y AnÃ¡lisis GenÃ³mico | Instituto Nacional de Salud, Universidad Cooperativa de Colombia, Instituto Alexander von Humboldt, Imperial College-London, London School of Hygiene & Tropical Medicine |
| EPI_ISL_456119 | Instituto Nacional de Salud - Unidad de SecuenciaciÃ³n y AnÃ¡lisis GenÃ³mico | Instituto Nacional de Salud, Universidad Cooperativa de Colombia, Instituto Alexander von Humboldt, Imperial College-London, London School of Hygiene & Tropical Medicine |
| EPI_ISL_456120 | Instituto Nacional de Salud - Unidad de SecuenciaciÃ³n y AnÃ¡lisis GenÃ³mico | Instituto Nacional de Salud, Universidad Cooperativa de Colombia, Instituto Alexander von Humboldt, Imperial College-London, London School of Hygiene & Tropical Medicine |
| EPI_ISL_456121 | Instituto Nacional de Salud - Unidad de SecuenciaciÃ³n y AnÃ¡lisis GenÃ³mico | Instituto Nacional de Salud, Universidad Cooperativa de Colombia, Instituto Alexander von Humboldt, Imperial College-London, London School of Hygiene & Tropical Medicine |
| EPI_ISL_456122 | Instituto Nacional de Salud - Unidad de SecuenciaciÃ³n y AnÃ¡lisis GenÃ³mico | Instituto Nacional de Salud, Universidad Cooperativa de Colombia, Instituto Alexander von Humboldt, Imperial College-London, London School of Hygiene & Tropical Medicine |
| EPI_ISL_456123 | Instituto Nacional de Salud - Unidad de SecuenciaciÃ³n y AnÃ¡lisis GenÃ³mico | Instituto Nacional de Salud, Universidad Cooperativa de Colombia, Instituto Alexander von Humboldt, Imperial College-London, London School of Hygiene & Tropical Medicine |

| EPI_Accession_ID | Originating Laboratory | Submitting Laboratory |
|---|---|---|
| EPI_ISL_456124 | Instituto Nacional de Salud - Unidad de SecuenciaciÃ³n y AnÃ¡lisis GenÃ³mico | Instituto Nacional de Salud, Universidad Cooperativa de Colombia, Instituto Alexander von Humboldt, Imperial College-London, London School of Hygiene & Tropical Medicine |
| EPI_ISL_456125 | Instituto Nacional de Salud - Unidad de SecuenciaciÃ³n y AnÃ¡lisis GenÃ³mico | Instituto Nacional de Salud, Universidad Cooperativa de Colombia, Instituto Alexander von Humboldt, Imperial College-London, London School of Hygiene & Tropical Medicine |
| EPI_ISL_456126 | Instituto Nacional de Salud - Unidad de SecuenciaciÃ³n y AnÃ¡lisis GenÃ³mico | Instituto Nacional de Salud, Universidad Cooperativa de Colombia, Instituto Alexander von Humboldt, Imperial College-London, London School of Hygiene & Tropical Medicine |
| EPI_ISL_456127 | Instituto Nacional de Salud - Unidad de SecuenciaciÃ³n y AnÃ¡lisis GenÃ³mico | Instituto Nacional de Salud, Universidad Cooperativa de Colombia, Instituto Alexander von Humboldt, Imperial College-London, London School of Hygiene & Tropical Medicine |
| EPI_ISL_456128 | Instituto Nacional de Salud - Unidad de SecuenciaciÃ³n y AnÃ¡lisis GenÃ³mico | Instituto Nacional de Salud, Universidad Cooperativa de Colombia, Instituto Alexander von Humboldt, Imperial College-London, London School of Hygiene & Tropical Medicine |
| EPI_ISL_456129 | Instituto Nacional de Salud - Unidad de SecuenciaciÃ³n y AnÃ¡lisis GenÃ³mico | Instituto Nacional de Salud, Universidad Cooperativa de Colombia, Instituto Alexander von Humboldt, Imperial College-London, London School of Hygiene & Tropical Medicine |
| EPI_ISL_456130 | Instituto Nacional de Salud - Unidad de SecuenciaciÃ³n y AnÃ¡lisis GenÃ³mico | Instituto Nacional de Salud, Universidad Cooperativa de Colombia, Instituto Alexander von Humboldt, Imperial College-London, London School of Hygiene & Tropical Medicine |
| EPI_ISL_456131 | Instituto Nacional de Salud - Unidad de SecuenciaciÃ³n y AnÃ¡lisis GenÃ³mico | Instituto Nacional de Salud, Universidad Cooperativa de Colombia, Instituto Alexander von Humboldt, Imperial College-London, London School of Hygiene & Tropical Medicine |

| EPI_Accession_ID | Originating Laboratory | Submitting Laboratory |
| --- | --- | --- |
| EPI_ISL_456132 | Instituto Nacional de Salud - Unidad de SecuenciaciÃ³n y AnÃ¡lisis GenÃ³mico | Instituto Nacional de Salud, Universidad Cooperativa de Colombia, Instituto Alexander von Humboldt, Imperial College-London, London School of Hygiene & Tropical Medicine |
| EPI_ISL_456133 | Instituto Nacional de Salud - Unidad de SecuenciaciÃ³n y AnÃ¡lisis GenÃ³mico | Instituto Nacional de Salud, Universidad Cooperativa de Colombia, Instituto Alexander von Humboldt, Imperial College-London, London School of Hygiene & Tropical Medicine |
| EPI_ISL_456134 | Instituto Nacional de Salud - Unidad de SecuenciaciÃ³n y AnÃ¡lisis GenÃ³mico | Instituto Nacional de Salud, Universidad Cooperativa de Colombia, Instituto Alexander von Humboldt, Imperial College-London, London School of Hygiene & Tropical Medicine |
| EPI_ISL_456135 | Instituto Nacional de Salud - Unidad de SecuenciaciÃ³n y AnÃ¡lisis GenÃ³mico | Instituto Nacional de Salud, Universidad Cooperativa de Colombia, Instituto Alexander von Humboldt, Imperial College-London, London School of Hygiene & Tropical Medicine |
| EPI_ISL_456136 | Instituto Nacional de Salud - Unidad de SecuenciaciÃ³n y AnÃ¡lisis GenÃ³mico | Instituto Nacional de Salud, Universidad Cooperativa de Colombia, Instituto Alexander von Humboldt, Imperial College-London, London School of Hygiene & Tropical Medicine |
| EPI_ISL_456137 | Instituto Nacional de Salud - Unidad de SecuenciaciÃ³n y AnÃ¡lisis GenÃ³mico | Instituto Nacional de Salud, Universidad Cooperativa de Colombia, Instituto Alexander von Humboldt, Imperial College-London, London School of Hygiene & Tropical Medicine |
| EPI_ISL_456138 | Instituto Nacional de Salud - Unidad de SecuenciaciÃ³n y AnÃ¡lisis GenÃ³mico | Instituto Nacional de Salud, Universidad Cooperativa de Colombia, Instituto Alexander von Humboldt, Imperial College-London, London School of Hygiene & Tropical Medicine |
| EPI_ISL_456139 | Instituto Nacional de Salud - Unidad de SecuenciaciÃ³n y AnÃ¡lisis GenÃ³mico | Instituto Nacional de Salud, Universidad Cooperativa de Colombia, Instituto Alexander von Humboldt, Imperial College-London, London School of Hygiene & Tropical Medicine |

| EPI_Accession_ID | Originating Laboratory | Submitting Laboratory |
|---|---|---|
| EPI_ISL_456140 | Instituto Nacional de Salud - Unidad de SecuenciaciÃ³n y AnÃ¡lisis GenÃ³mico | Instituto Nacional de Salud, Universidad Cooperativa de Colombia, Instituto Alexander von Humboldt, Imperial College-London, London School of Hygiene & Tropical Medicine |
| EPI_ISL_456141 | Instituto Nacional de Salud - Unidad de SecuenciaciÃ³n y AnÃ¡lisis GenÃ³mico | Instituto Nacional de Salud, Universidad Cooperativa de Colombia, Instituto Alexander von Humboldt, Imperial College-London, London School of Hygiene & Tropical Medicine |
| EPI_ISL_456142 | Instituto Nacional de Salud - Unidad de SecuenciaciÃ³n y AnÃ¡lisis GenÃ³mico | Instituto Nacional de Salud, Universidad Cooperativa de Colombia, Instituto Alexander von Humboldt, Imperial College-London, London School of Hygiene & Tropical Medicine |
| EPI_ISL_456143 | Instituto Nacional de Salud - Unidad de SecuenciaciÃ³n y AnÃ¡lisis GenÃ³mico | Instituto Nacional de Salud, Universidad Cooperativa de Colombia, Instituto Alexander von Humboldt, Imperial College-London, London School of Hygiene & Tropical Medicine |
| EPI_ISL_456144 | Instituto Nacional de Salud - Unidad de SecuenciaciÃ³n y AnÃ¡lisis GenÃ³mico | Instituto Nacional de Salud, Universidad Cooperativa de Colombia, Instituto Alexander von Humboldt, Imperial College-London, London School of Hygiene & Tropical Medicine |
| EPI_ISL_456145 | Instituto Nacional de Salud - Unidad de SecuenciaciÃ³n y AnÃ¡lisis GenÃ³mico | Instituto Nacional de Salud, Universidad Cooperativa de Colombia, Instituto Alexander von Humboldt, Imperial College-London, London School of Hygiene & Tropical Medicine |
| EPI_ISL_456146 | Instituto Nacional de Salud - Unidad de SecuenciaciÃ³n y AnÃ¡lisis GenÃ³mico | Instituto Nacional de Salud, Universidad Cooperativa de Colombia, Instituto Alexander von Humboldt, Imperial College-London, London School of Hygiene & Tropical Medicine |
| EPI_ISL_456147 | Instituto Nacional de Salud - Unidad de SecuenciaciÃ³n y AnÃ¡lisis GenÃ³mico | Instituto Nacional de Salud, Universidad Cooperativa de Colombia, Instituto Alexander von Humboldt, Imperial College-London, London School of Hygiene & Tropical Medicine |

| EPI_Accession_ID | Originating Laboratory | Submitting Laboratory |
|---|---|---|
| EPI_ISL_456148 | Instituto Nacional de Salud - Unidad de SecuenciaciÃ³n y AnÃ¡lisis GenÃ³mico | Instituto Nacional de Salud, Universidad Cooperativa de Colombia, Instituto Alexander von Humboldt, Imperial College-London, London School of Hygiene & Tropical Medicine |
| EPI_ISL_456149 | Instituto Nacional de Salud - Unidad de SecuenciaciÃ³n y AnÃ¡lisis GenÃ³mico | Instituto Nacional de Salud, Universidad Cooperativa de Colombia, Instituto Alexander von Humboldt, Imperial College-London, London School of Hygiene & Tropical Medicine |
| EPI_ISL_456150 | Instituto Nacional de Salud - Unidad de SecuenciaciÃ³n y AnÃ¡lisis GenÃ³mico | Instituto Nacional de Salud, Universidad Cooperativa de Colombia, Instituto Alexander von Humboldt, Imperial College-London, London School of Hygiene & Tropical Medicine |
| EPI_ISL_456151 | Instituto Nacional de Salud - Unidad de SecuenciaciÃ³n y AnÃ¡lisis GenÃ³mico | Instituto Nacional de Salud, Universidad Cooperativa de Colombia, Instituto Alexander von Humboldt, Imperial College-London, London School of Hygiene & Tropical Medicine |
| EPI_ISL_456152 | Instituto Nacional de Salud - Unidad de SecuenciaciÃ³n y AnÃ¡lisis GenÃ³mico | Instituto Nacional de Salud, Universidad Cooperativa de Colombia, Instituto Alexander von Humboldt, Imperial College-London, London School of Hygiene & Tropical Medicine |
| EPI_ISL_456153 | Instituto Nacional de Salud - Unidad de SecuenciaciÃ³n y AnÃ¡lisis GenÃ³mico | Instituto Nacional de Salud, Universidad Cooperativa de Colombia, Instituto Alexander von Humboldt, Imperial College-London, London School of Hygiene & Tropical Medicine |
| EPI_ISL_456154 | Instituto Nacional de Salud - Unidad de SecuenciaciÃ³n y AnÃ¡lisis GenÃ³mico | Instituto Nacional de Salud, Universidad Cooperativa de Colombia, Instituto Alexander von Humboldt, Imperial College-London, London School of Hygiene & Tropical Medicine |
| EPI_ISL_456155 | Instituto Nacional de Salud - Unidad de SecuenciaciÃ³n y AnÃ¡lisis GenÃ³mico | Instituto Nacional de Salud, Universidad Cooperativa de Colombia, Instituto Alexander von Humboldt, Imperial College-London, London School of Hygiene & Tropical Medicine |

| EPI_Accession_ID | Originating Laboratory | Submitting Laboratory |
|---|---|---|
| EPI_ISL_456156 | Instituto Nacional de Salud - Unidad de SecuenciaciÃ³n y AnÃ¡lisis GenÃ³mico | Instituto Nacional de Salud, Universidad Cooperativa de Colombia, Instituto Alexander von Humboldt, Imperial College-London, London School of Hygiene & Tropical Medicine |
| EPI_ISL_456157 | LabPLUS | Institute of Environmental Science and Research (ESR) |
| EPI_ISL_456158 | Southern Community Labs Dunedin | Institute of Environmental Science and Research (ESR) |
| EPI_ISL_456159 | Southern Community Labs Dunedin | Institute of Environmental Science and Research (ESR) |
| EPI_ISL_456160 | Waikato Hospital | Institute of Environmental Science and Research (ESR) |
| EPI_ISL_456161 | Wellington SCL | Institute of Environmental Science and Research (ESR) |
| EPI_ISL_456162 | Southern Community Labs Dunedin | Institute of Environmental Science and Research (ESR) |
| EPI_ISL_456163 | PathLab Bay of Plenty | Institute of Environmental Science and Research (ESR) |
| EPI_ISL_456164 | Wellington SCL | Institute of Environmental Science and Research (ESR) |
| EPI_ISL_456165 | Wellington SCL | Institute of Environmental Science and Research (ESR) |
| EPI_ISL_456166 | Wellington SCL | Institute of Environmental Science and Research (ESR) |
| EPI_ISL_456167 | Southern Community Labs Dunedin | Institute of Environmental Science and Research (ESR) |
| EPI_ISL_456168 | Waikato Hospital | Institute of Environmental Science and Research (ESR) |
| EPI_ISL_456169 | Wellington SCL | Institute of Environmental Science and Research (ESR) |
| EPI_ISL_456170 | Wellington SCL | Institute of Environmental Science and Research (ESR) |
| EPI_ISL_456171 | PathLab Bay of Plenty | Institute of Environmental Science and Research (ESR) |
| EPI_ISL_456172 | LabPLUS | Institute of Environmental Science and Research (ESR) |
| EPI_ISL_456173 | LabPLUS | Institute of Environmental Science and Research (ESR) |
| EPI_ISL_456174 | LabPLUS | Institute of Environmental Science and Research (ESR) |
| EPI_ISL_456175 | LabPLUS | Institute of Environmental Science and Research (ESR) |
| EPI_ISL_456176 | LabPLUS | Institute of Environmental Science and Research (ESR) |
| EPI_ISL_456177 | LabPLUS | Institute of Environmental Science and Research (ESR) |
| EPI_ISL_456178 | LabPLUS | Institute of Environmental Science and Research (ESR) |
| EPI_ISL_456179 | LabPLUS | Institute of Environmental Science and Research (ESR) |
| EPI_ISL_456180 | LabPLUS | Institute of Environmental Science and Research (ESR) |
| EPI_ISL_456181 | Southern Community Labs Dunedin | Institute of Environmental Science and Research (ESR) |
| EPI_ISL_456182 | Wellington SCL | Institute of Environmental Science and Research (ESR) |
| EPI_ISL_456183 | Wellington SCL | Institute of Environmental Science and Research (ESR) |
| EPI_ISL_456184 | Wellington SCL | Institute of Environmental Science and Research (ESR) |
| EPI_ISL_456185 | Wellington SCL | Institute of Environmental Science and Research (ESR) |

| EPI_Accession_ID | Originating Laboratory | Submitting Laboratory |
|---|---|---|
| EPI_ISL_456186 | Wellington SCL | Institute of Environmental Science and Research (ESR) |
| EPI_ISL_456187 | Wellington SCL | Institute of Environmental Science and Research (ESR) |
| EPI_ISL_456188 | Wellington SCL | Institute of Environmental Science and Research (ESR) |
| EPI_ISL_456189 | Waikato Hospital | Institute of Environmental Science and Research (ESR) |
| EPI_ISL_456190 | Waikato Hospital | Institute of Environmental Science and Research (ESR) |
| EPI_ISL_456191 | Waikato Hospital | Institute of Environmental Science and Research (ESR) |
| EPI_ISL_456192 | Waikato Hospital | Institute of Environmental Science and Research (ESR) |
| EPI_ISL_456193 | Waikato Hospital | Institute of Environmental Science and Research (ESR) |
| EPI_ISL_456194 | PathLab Bay of Plenty | Institute of Environmental Science and Research (ESR) |
| EPI_ISL_456195 | PathLab Bay of Plenty | Institute of Environmental Science and Research (ESR) |
| EPI_ISL_456196 | LabPLUS | Institute of Environmental Science and Research (ESR) |
| EPI_ISL_456197 | LabPLUS | Institute of Environmental Science and Research (ESR) |
| EPI_ISL_456198 | LabPLUS | Institute of Environmental Science and Research (ESR) |
| EPI_ISL_456199 | LabPLUS | Institute of Environmental Science and Research (ESR) |
| EPI_ISL_456200 | LabPLUS | Institute of Environmental Science and Research (ESR) |
| EPI_ISL_456201 | LabPLUS | Institute of Environmental Science and Research (ESR) |
| EPI_ISL_456202 | LabPLUS | Institute of Environmental Science and Research (ESR) |
| EPI_ISL_456203 | LabPLUS | Institute of Environmental Science and Research (ESR) |
| EPI_ISL_456204 | LabPLUS | Institute of Environmental Science and Research (ESR) |
| EPI_ISL_456205 | LabPLUS | Institute of Environmental Science and Research (ESR) |
| EPI_ISL_456206 | LabPLUS | Institute of Environmental Science and Research (ESR) |
| EPI_ISL_456207 | LabPLUS | Institute of Environmental Science and Research (ESR) |
| EPI_ISL_456208 | LabPLUS | Institute of Environmental Science and Research (ESR) |
| EPI_ISL_456209 | LabPLUS | Institute of Environmental Science and Research (ESR) |
| EPI_ISL_456210 | LabPLUS | Institute of Environmental Science and Research (ESR) |
| EPI_ISL_456211 | LabPLUS | Institute of Environmental Science and Research (ESR) |
| EPI_ISL_456212 | LabPLUS | Institute of Environmental Science and Research (ESR) |
| EPI_ISL_456213 | Middlemore Hospital | Institute of Environmental Science and Research (ESR) |
| EPI_ISL_456214 | Middlemore Hospital | Institute of Environmental Science and Research (ESR) |
| EPI_ISL_456215 | PathLab Bay of Plenty | Institute of Environmental Science and Research (ESR) |
| EPI_ISL_456216 | Southern Community Labs Dunedin | Institute of Environmental Science and Research (ESR) |
| EPI_ISL_456217 | Southern Community Labs Dunedin | Institute of Environmental Science and Research (ESR) |
| EPI_ISL_456218 | Southern Community Labs Dunedin | Institute of Environmental Science and Research (ESR) |

| EPI_Accession_ID | Originating Laboratory | Submitting Laboratory |
|---|---|---|
| EPI_ISL_456219 | Southern Community Labs Dunedin | Institute of Environmental Science and Research (ESR) |
| EPI_ISL_456220 | Southern Community Labs Dunedin | Institute of Environmental Science and Research (ESR) |
| EPI_ISL_456221 | PathLab Bay of Plenty | Institute of Environmental Science and Research (ESR) |
| EPI_ISL_456222 | PathLab Bay of Plenty | Institute of Environmental Science and Research (ESR) |
| EPI_ISL_456223 | PathLab Bay of Plenty | Institute of Environmental Science and Research (ESR) |
| EPI_ISL_456224 | PathLab Bay of Plenty | Institute of Environmental Science and Research (ESR) |
| EPI_ISL_456225 | Southern Community Labs Dunedin | Institute of Environmental Science and Research (ESR) |
| EPI_ISL_456226 | Southern Community Labs Dunedin | Institute of Environmental Science and Research (ESR) |
| EPI_ISL_456227 | Southern Community Labs Dunedin | Institute of Environmental Science and Research (ESR) |
| EPI_ISL_456228 | Southern Community Labs Dunedin | Institute of Environmental Science and Research (ESR) |
| EPI_ISL_456229 | Southern Community Labs Dunedin | Institute of Environmental Science and Research (ESR) |
| EPI_ISL_456230 | Southern Community Labs Dunedin | Institute of Environmental Science and Research (ESR) |
| EPI_ISL_456231 | Southern Community Labs Dunedin | Institute of Environmental Science and Research (ESR) |
| EPI_ISL_456232 | Southern Community Labs Dunedin | Institute of Environmental Science and Research (ESR) |
| EPI_ISL_456233 | Southern Community Labs Dunedin | Institute of Environmental Science and Research (ESR) |
| EPI_ISL_456234 | Southern Community Labs Dunedin | Institute of Environmental Science and Research (ESR) |
| EPI_ISL_456235 | Southern Community Labs Dunedin | Institute of Environmental Science and Research (ESR) |
| EPI_ISL_456236 | Southern Community Labs Dunedin | Institute of Environmental Science and Research (ESR) |
| EPI_ISL_456237 | Southern Community Labs Dunedin | Institute of Environmental Science and Research (ESR) |
| EPI_ISL_456238 | Southern Community Labs Dunedin | Institute of Environmental Science and Research (ESR) |
| EPI_ISL_456239 | Southern Community Labs Dunedin | Institute of Environmental Science and Research (ESR) |
| EPI_ISL_456240 | Southern Community Labs Dunedin | Institute of Environmental Science and Research (ESR) |
| EPI_ISL_456241 | Waikato Hospital | Institute of Environmental Science and Research (ESR) |
| EPI_ISL_456242 | Southern Community Labs Dunedin | Institute of Environmental Science and Research (ESR) |
| EPI_ISL_456243 | Southern Community Labs Dunedin | Institute of Environmental Science and Research (ESR) |
| EPI_ISL_456244 | Southern Community Labs Dunedin | Institute of Environmental Science and Research (ESR) |
| EPI_ISL_456245 | Southern Community Labs Dunedin | Institute of Environmental Science and Research (ESR) |
| EPI_ISL_456246 | Southern Community Labs Dunedin | Institute of Environmental Science and Research (ESR) |
| EPI_ISL_456247 | Southern Community Labs Dunedin | Institute of Environmental Science and Research (ESR) |
| EPI_ISL_456248 | Southern Community Labs Dunedin | Institute of Environmental Science and Research (ESR) |
| EPI_ISL_456249 | Southern Community Labs Dunedin | Institute of Environmental Science and Research (ESR) |
| EPI_ISL_456250 | Southern Community Labs Dunedin | Institute of Environmental Science and Research (ESR) |
| EPI_ISL_456251 | Southern Community Labs Dunedin | Institute of Environmental Science and Research (ESR) |

| EPI_Accession_ID | Originating Laboratory | Submitting Laboratory |
|---|---|---|
| EPI_ISL_456252 | Southern Community Labs Dunedin | Institute of Environmental Science and Research (ESR) |
| EPI_ISL_456253 | Southern Community Labs Dunedin | Institute of Environmental Science and Research (ESR) |
| EPI_ISL_456254 | Southern Community Labs Dunedin | Institute of Environmental Science and Research (ESR) |
| EPI_ISL_456255 | Middlemore Hospital | Institute of Environmental Science and Research (ESR) |
| EPI_ISL_456256 | Middlemore Hospital | Institute of Environmental Science and Research (ESR) |
| EPI_ISL_456257 | Middlemore Hospital | Institute of Environmental Science and Research (ESR) |
| EPI_ISL_456258 | Middlemore Hospital | Institute of Environmental Science and Research (ESR) |
| EPI_ISL_456259 | Waikato Hospital | Institute of Environmental Science and Research (ESR) |
| EPI_ISL_456260 | Waikato Hospital | Institute of Environmental Science and Research (ESR) |
| EPI_ISL_456261 | Southern Community Labs Dunedin | Institute of Environmental Science and Research (ESR) |
| EPI_ISL_456262 | Southern Community Labs Dunedin | Institute of Environmental Science and Research (ESR) |
| EPI_ISL_456263 | Southern Community Labs Dunedin | Institute of Environmental Science and Research (ESR) |
| EPI_ISL_456264 | Southern Community Labs Dunedin | Institute of Environmental Science and Research (ESR) |
| EPI_ISL_456265 | Southern Community Labs Dunedin | Institute of Environmental Science and Research (ESR) |
| EPI_ISL_456266 | Southern Community Labs Dunedin | Institute of Environmental Science and Research (ESR) |
| EPI_ISL_456267 | Southern Community Labs Dunedin | Institute of Environmental Science and Research (ESR) |
| EPI_ISL_456268 | Southern Community Labs Dunedin | Institute of Environmental Science and Research (ESR) |
| EPI_ISL_456269 | Southern Community Labs Dunedin | Institute of Environmental Science and Research (ESR) |
| EPI_ISL_456270 | Southern Community Labs Dunedin | Institute of Environmental Science and Research (ESR) |
| EPI_ISL_456271 | Southern Community Labs Dunedin | Institute of Environmental Science and Research (ESR) |
| EPI_ISL_456272 | Southern Community Labs Dunedin | Institute of Environmental Science and Research (ESR) |
| EPI_ISL_456273 | Southern Community Labs Dunedin | Institute of Environmental Science and Research (ESR) |
| EPI_ISL_456274 | Southern Community Labs Dunedin | Institute of Environmental Science and Research (ESR) |
| EPI_ISL_456275 | Southern Community Labs Dunedin | Institute of Environmental Science and Research (ESR) |
| EPI_ISL_456276 | Southern Community Labs Dunedin | Institute of Environmental Science and Research (ESR) |
| EPI_ISL_456277 | Southern Community Labs Dunedin | Institute of Environmental Science and Research (ESR) |
| EPI_ISL_456278 | Southern Community Labs Dunedin | Institute of Environmental Science and Research (ESR) |
| EPI_ISL_456279 | Southern Community Labs Dunedin | Institute of Environmental Science and Research (ESR) |
| EPI_ISL_456280 | Southern Community Labs Dunedin | Institute of Environmental Science and Research (ESR) |
| EPI_ISL_456281 | Southern Community Labs Dunedin | Institute of Environmental Science and Research (ESR) |
| EPI_ISL_456282 | Southern Community Labs Dunedin | Institute of Environmental Science and Research (ESR) |
| EPI_ISL_456283 | Southern Community Labs Dunedin | Institute of Environmental Science and Research (ESR) |
| EPI_ISL_456284 | Southern Community Labs Dunedin | Institute of Environmental Science and Research (ESR) |

| EPI_Accession_ID | Originating Laboratory | Submitting Laboratory |
|---|---|---|
| EPI_ISL_456285 | Southern Community Labs Dunedin | Institute of Environmental Science and Research (ESR) |
| EPI_ISL_456286 | Southern Community Labs Dunedin | Institute of Environmental Science and Research (ESR) |
| EPI_ISL_456287 | Southern Community Labs Dunedin | Institute of Environmental Science and Research (ESR) |
| EPI_ISL_456288 | Southern Community Labs Dunedin | Institute of Environmental Science and Research (ESR) |
| EPI_ISL_456289 | Southern Community Labs Dunedin | Institute of Environmental Science and Research (ESR) |
| EPI_ISL_456290 | Southern Community Labs Dunedin | Institute of Environmental Science and Research (ESR) |
| EPI_ISL_456291 | Southern Community Labs Dunedin | Institute of Environmental Science and Research (ESR) |
| EPI_ISL_456292 | Southern Community Labs Dunedin | Institute of Environmental Science and Research (ESR) |
| EPI_ISL_456293 | Southern Community Labs Dunedin | Institute of Environmental Science and Research (ESR) |
| EPI_ISL_456294 | North Shore Hospital | Institute of Environmental Science and Research (ESR) |
| EPI_ISL_456295 | Southern Community Labs Dunedin | Institute of Environmental Science and Research (ESR) |
| EPI_ISL_456296 | Southern Community Labs Dunedin | Institute of Environmental Science and Research (ESR) |
| EPI_ISL_456297 | Southern Community Labs Dunedin | Institute of Environmental Science and Research (ESR) |
| EPI_ISL_456298 | Southern Community Labs Dunedin | Institute of Environmental Science and Research (ESR) |
| EPI_ISL_456299 | Southern Community Labs Dunedin | Institute of Environmental Science and Research (ESR) |
| EPI_ISL_456300 | LabPLUS | Institute of Environmental Science and Research (ESR) |
| EPI_ISL_456301 | PathLab Bay of Plenty | Institute of Environmental Science and Research (ESR) |
| EPI_ISL_456302 | Southern Community Labs Dunedin | Institute of Environmental Science and Research (ESR) |
| EPI_ISL_456303 | Southern Community Labs Dunedin | Institute of Environmental Science and Research (ESR) |
| EPI_ISL_456304 | Southern Community Labs Dunedin | Institute of Environmental Science and Research (ESR) |
| EPI_ISL_456305 | Southern Community Labs Dunedin | Institute of Environmental Science and Research (ESR) |
| EPI_ISL_456306 | Southern Community Labs Dunedin | Institute of Environmental Science and Research (ESR) |
| EPI_ISL_456307 | Southern Community Labs Dunedin | Institute of Environmental Science and Research (ESR) |
| EPI_ISL_456308 | Southern Community Labs Dunedin | Institute of Environmental Science and Research (ESR) |
| EPI_ISL_456309 | Southern Community Labs Dunedin | Institute of Environmental Science and Research (ESR) |
| EPI_ISL_456310 | Southern Community Labs Dunedin | Institute of Environmental Science and Research (ESR) |
| EPI_ISL_456311 | Southern Community Labs Dunedin | Institute of Environmental Science and Research (ESR) |
| EPI_ISL_456312 | Southern Community Labs Dunedin | Institute of Environmental Science and Research (ESR) |
| EPI_ISL_456313 | Southern Community Labs Dunedin | Institute of Environmental Science and Research (ESR) |
| EPI_ISL_456314 | Southern Community Labs Dunedin | Institute of Environmental Science and Research (ESR) |
| EPI_ISL_456315 | Southern Community Labs Dunedin | Institute of Environmental Science and Research (ESR) |
| EPI_ISL_456316 | Southern Community Labs Dunedin | Institute of Environmental Science and Research (ESR) |
| EPI_ISL_456317 | Southern Community Labs Dunedin | Institute of Environmental Science and Research (ESR) |

| EPI_Accession_ID | Originating Laboratory | Submitting Laboratory |
|---|---|---|
| EPI_ISL_456318 | Southern Community Labs Dunedin | Institute of Environmental Science and Research (ESR) |
| EPI_ISL_456319 | Wellington SCL | Institute of Environmental Science and Research (ESR) |
| EPI_ISL_456320 | Wellington SCL | Institute of Environmental Science and Research (ESR) |
| EPI_ISL_456321 | Canterbury Health Laboratories | Institute of Environmental Science and Research (ESR) |
| EPI_ISL_456322 | Canterbury Health Laboratories | Institute of Environmental Science and Research (ESR) |
| EPI_ISL_456323 | North Shore Hospital | Institute of Environmental Science and Research (ESR) |
| EPI_ISL_456324 | Wellington SCL | Institute of Environmental Science and Research (ESR) |
| EPI_ISL_456325 | Wellington SCL | Institute of Environmental Science and Research (ESR) |
| EPI_ISL_456326 | Wellington SCL | Institute of Environmental Science and Research (ESR) |
| EPI_ISL_456327 | Wellington SCL | Institute of Environmental Science and Research (ESR) |
| EPI_ISL_456328 | Wellington SCL | Institute of Environmental Science and Research (ESR) |
| EPI_ISL_456329 | Wellington SCL | Institute of Environmental Science and Research (ESR) |
| EPI_ISL_456330 | Wellington SCL | Institute of Environmental Science and Research (ESR) |
| EPI_ISL_456331 | Wellington SCL | Institute of Environmental Science and Research (ESR) |
| EPI_ISL_456332 | Wellington SCL | Institute of Environmental Science and Research (ESR) |
| EPI_ISL_456333 | Wellington SCL | Institute of Environmental Science and Research (ESR) |
| EPI_ISL_456334 | Wellington SCL | Institute of Environmental Science and Research (ESR) |
| EPI_ISL_456335 | Wellington SCL | Institute of Environmental Science and Research (ESR) |
| EPI_ISL_456336 | Wellington SCL | Institute of Environmental Science and Research (ESR) |
| EPI_ISL_456337 | Wellington SCL | Institute of Environmental Science and Research (ESR) |
| EPI_ISL_456338 | Wellington SCL | Institute of Environmental Science and Research (ESR) |
| EPI_ISL_456339 | Wellington SCL | Institute of Environmental Science and Research (ESR) |
| EPI_ISL_456340 | Wellington SCL | Institute of Environmental Science and Research (ESR) |
| EPI_ISL_456341 | Wellington SCL | Institute of Environmental Science and Research (ESR) |
| EPI_ISL_456342 | Wellington SCL | Institute of Environmental Science and Research (ESR) |
| EPI_ISL_456343 | Wellington SCL | Institute of Environmental Science and Research (ESR) |
| EPI_ISL_456344 | Southern Community Labs Dunedin | Institute of Environmental Science and Research (ESR) |
| EPI_ISL_456345 | Southern Community Labs Dunedin | Institute of Environmental Science and Research (ESR) |
| EPI_ISL_456346 | Southern Community Labs Dunedin | Institute of Environmental Science and Research (ESR) |
| EPI_ISL_456347 | Southern Community Labs Dunedin | Institute of Environmental Science and Research (ESR) |
| EPI_ISL_456348 | Canterbury Health Laboratories | Institute of Environmental Science and Research (ESR) |
| EPI_ISL_456349 | Canterbury Health Laboratories | Institute of Environmental Science and Research (ESR) |
| EPI_ISL_456350 | Canterbury Health Laboratories | Institute of Environmental Science and Research (ESR) |

| EPI_Accession_ID | Originating Laboratory | Submitting Laboratory |
|---|---|---|
| EPI_ISL_456351 | Canterbury Health Laboratories | Institute of Environmental Science and Research (ESR) |
| EPI_ISL_456352 | Canterbury Health Laboratories | Institute of Environmental Science and Research (ESR) |
| EPI_ISL_456353 | Canterbury Health Laboratories | Institute of Environmental Science and Research (ESR) |
| EPI_ISL_456354 | Canterbury Health Laboratories | Institute of Environmental Science and Research (ESR) |
| EPI_ISL_456355 | Canterbury Health Laboratories | Institute of Environmental Science and Research (ESR) |
| EPI_ISL_456356 | Canterbury Health Laboratories | Institute of Environmental Science and Research (ESR) |
| EPI_ISL_456357 | Canterbury Health Laboratories | Institute of Environmental Science and Research (ESR) |
| EPI_ISL_456358 | Canterbury Health Laboratories | Institute of Environmental Science and Research (ESR) |
| EPI_ISL_456359 | Canterbury Health Laboratories | Institute of Environmental Science and Research (ESR) |
| EPI_ISL_456360 | Canterbury Health Laboratories | Institute of Environmental Science and Research (ESR) |
| EPI_ISL_456361 | Canterbury Health Laboratories | Institute of Environmental Science and Research (ESR) |
| EPI_ISL_456362 | Canterbury Health Laboratories | Institute of Environmental Science and Research (ESR) |
| EPI_ISL_456363 | Canterbury Health Laboratories | Institute of Environmental Science and Research (ESR) |
| EPI_ISL_456364 | Canterbury Health Laboratories | Institute of Environmental Science and Research (ESR) |
| EPI_ISL_456365 | Canterbury Health Laboratories | Institute of Environmental Science and Research (ESR) |
| EPI_ISL_456366 | Canterbury Health Laboratories | Institute of Environmental Science and Research (ESR) |
| EPI_ISL_456367 | Canterbury Health Laboratories | Institute of Environmental Science and Research (ESR) |
| EPI_ISL_456368 | Canterbury Health Laboratories | Institute of Environmental Science and Research (ESR) |
| EPI_ISL_456369 | Canterbury Health Laboratories | Institute of Environmental Science and Research (ESR) |
| EPI_ISL_456370 | Canterbury Health Laboratories | Institute of Environmental Science and Research (ESR) |
| EPI_ISL_456371 | Canterbury Health Laboratories | Institute of Environmental Science and Research (ESR) |
| EPI_ISL_456372 | Canterbury Health Laboratories | Institute of Environmental Science and Research (ESR) |
| EPI_ISL_456373 | Canterbury Health Laboratories | Institute of Environmental Science and Research (ESR) |
| EPI_ISL_456374 | Canterbury Health Laboratories | Institute of Environmental Science and Research (ESR) |
| EPI_ISL_456375 | Canterbury Health Laboratories | Institute of Environmental Science and Research (ESR) |
| EPI_ISL_456376 | Middlemore Hospital | Institute of Environmental Science and Research (ESR) |
| EPI_ISL_456377 | MedLab Central Ltd | Institute of Environmental Science and Research (ESR) |
| EPI_ISL_456378 | MedLab Central Ltd | Institute of Environmental Science and Research (ESR) |
| EPI_ISL_456379 | LabPLUS | Institute of Environmental Science and Research (ESR) |
| EPI_ISL_456380 | LabPLUS | Institute of Environmental Science and Research (ESR) |
| EPI_ISL_456381 | LabPLUS | Institute of Environmental Science and Research (ESR) |
| EPI_ISL_456382 | LabPLUS | Institute of Environmental Science and Research (ESR) |
| EPI_ISL_456383 | LabPLUS | Institute of Environmental Science and Research (ESR) |

| EPI_Accession_ID | Originating Laboratory | Submitting Laboratory |
|---|---|---|
| EPI_ISL_456384 | LabPLUS | Institute of Environmental Science and Research (ESR) |
| EPI_ISL_456385 | Middlemore Hospital | Institute of Environmental Science and Research (ESR) |
| EPI_ISL_456386 | Middlemore Hospital | Institute of Environmental Science and Research (ESR) |
| EPI_ISL_456387 | Middlemore Hospital | Institute of Environmental Science and Research (ESR) |
| EPI_ISL_456388 | LabPLUS | Institute of Environmental Science and Research (ESR) |
| EPI_ISL_456389 | LabPLUS | Institute of Environmental Science and Research (ESR) |
| EPI_ISL_456390 | LabPLUS | Institute of Environmental Science and Research (ESR) |
| EPI_ISL_456391 | LabPLUS | Institute of Environmental Science and Research (ESR) |
| EPI_ISL_456392 | LabPLUS | Institute of Environmental Science and Research (ESR) |
| EPI_ISL_456393 | LabPLUS | Institute of Environmental Science and Research (ESR) |
| EPI_ISL_456394 | LabPLUS | Institute of Environmental Science and Research (ESR) |
| EPI_ISL_456395 | North Shore Hospital | Institute of Environmental Science and Research (ESR) |
| EPI_ISL_456396 | North Shore Hospital | Institute of Environmental Science and Research (ESR) |
| EPI_ISL_456397 | North Shore Hospital | Institute of Environmental Science and Research (ESR) |
| EPI_ISL_456398 | Wellington SCL | Institute of Environmental Science and Research (ESR) |
| EPI_ISL_456399 | Wellington SCL | Institute of Environmental Science and Research (ESR) |
| EPI_ISL_456400 | Wellington SCL | Institute of Environmental Science and Research (ESR) |
| EPI_ISL_456401 | Wellington SCL | Institute of Environmental Science and Research (ESR) |
| EPI_ISL_456402 | Wellington SCL | Institute of Environmental Science and Research (ESR) |
| EPI_ISL_456403 | Middlemore Hospital | Institute of Environmental Science and Research (ESR) |
| EPI_ISL_456410 | Victorian Infectious Diseases Reference Laboratory (VIDRL) | Microbiological Diagnostic Unit Public Health Laboratory and Victorian Infectious Diseases Reference Laboratory, Doherty Institute |
| EPI_ISL_456411 | Victorian Infectious Diseases Reference Laboratory (VIDRL) | Microbiological Diagnostic Unit Public Health Laboratory and Victorian Infectious Diseases Reference Laboratory, Doherty Institute |
| EPI_ISL_456413 | Victorian Infectious Diseases Reference Laboratory (VIDRL) | Microbiological Diagnostic Unit Public Health Laboratory and Victorian Infectious Diseases Reference Laboratory, Doherty Institute |
| EPI_ISL_456414 | Victorian Infectious Diseases Reference Laboratory (VIDRL) | Microbiological Diagnostic Unit Public Health Laboratory and Victorian Infectious Diseases Reference Laboratory, Doherty Institute |

| EPI_Accession_ID | Originating Laboratory | Submitting Laboratory |
|---|---|---|
| EPI_ISL_456415 | Victorian Infectious Diseases Reference Laboratory (VIDRL) | Microbiological Diagnostic Unit Public Health Laboratory and Victorian Infectious Diseases Reference Laboratory, Doherty Institute |
| EPI_ISL_456416 | Victorian Infectious Diseases Reference Laboratory (VIDRL) | Microbiological Diagnostic Unit Public Health Laboratory and Victorian Infectious Diseases Reference Laboratory, Doherty Institute |
| EPI_ISL_456417 | Victorian Infectious Diseases Reference Laboratory (VIDRL) | Microbiological Diagnostic Unit Public Health Laboratory and Victorian Infectious Diseases Reference Laboratory, Doherty Institute |
| EPI_ISL_456418 | Victorian Infectious Diseases Reference Laboratory (VIDRL) | Microbiological Diagnostic Unit Public Health Laboratory and Victorian Infectious Diseases Reference Laboratory, Doherty Institute |
| EPI_ISL_456419 | Victorian Infectious Diseases Reference Laboratory (VIDRL) | Microbiological Diagnostic Unit Public Health Laboratory and Victorian Infectious Diseases Reference Laboratory, Doherty Institute |
| EPI_ISL_456420 | Victorian Infectious Diseases Reference Laboratory (VIDRL) | Microbiological Diagnostic Unit Public Health Laboratory and Victorian Infectious Diseases Reference Laboratory, Doherty Institute |
| EPI_ISL_456421 | Victorian Infectious Diseases Reference Laboratory (VIDRL) | Microbiological Diagnostic Unit Public Health Laboratory and Victorian Infectious Diseases Reference Laboratory, Doherty Institute |
| EPI_ISL_456422 | Victorian Infectious Diseases Reference Laboratory (VIDRL) | Microbiological Diagnostic Unit Public Health Laboratory and Victorian Infectious Diseases Reference Laboratory, Doherty Institute |
| EPI_ISL_456423 | Victorian Infectious Diseases Reference Laboratory (VIDRL) | Microbiological Diagnostic Unit Public Health Laboratory and Victorian Infectious Diseases Reference Laboratory, Doherty Institute |
| EPI_ISL_456424 | Victorian Infectious Diseases Reference Laboratory (VIDRL) | Microbiological Diagnostic Unit Public Health Laboratory and Victorian Infectious Diseases Reference Laboratory, Doherty Institute |
| EPI_ISL_456425 | Victorian Infectious Diseases Reference Laboratory (VIDRL) | Microbiological Diagnostic Unit Public Health Laboratory and Victorian Infectious Diseases Reference Laboratory, Doherty Institute |

| EPI_Accession_ID | Originating Laboratory | Submitting Laboratory |
|---|---|---|
| EPI_ISL_456426 | Victorian Infectious Diseases Reference Laboratory (VIDRL) | Microbiological Diagnostic Unit Public Health Laboratory and Victorian Infectious Diseases Reference Laboratory, Doherty Institute |
| EPI_ISL_456427 | Victorian Infectious Diseases Reference Laboratory (VIDRL) | Microbiological Diagnostic Unit Public Health Laboratory and Victorian Infectious Diseases Reference Laboratory, Doherty Institute |
| EPI_ISL_456428 | Victorian Infectious Diseases Reference Laboratory (VIDRL) | Microbiological Diagnostic Unit Public Health Laboratory and Victorian Infectious Diseases Reference Laboratory, Doherty Institute |
| EPI_ISL_456429 | Victorian Infectious Diseases Reference Laboratory (VIDRL) | Microbiological Diagnostic Unit Public Health Laboratory and Victorian Infectious Diseases Reference Laboratory, Doherty Institute |
| EPI_ISL_456430 | Victorian Infectious Diseases Reference Laboratory (VIDRL) | Microbiological Diagnostic Unit Public Health Laboratory and Victorian Infectious Diseases Reference Laboratory, Doherty Institute |
| EPI_ISL_456431 | Victorian Infectious Diseases Reference Laboratory (VIDRL) | Microbiological Diagnostic Unit Public Health Laboratory and Victorian Infectious Diseases Reference Laboratory, Doherty Institute |
| EPI_ISL_456432 | Victorian Infectious Diseases Reference Laboratory (VIDRL) | Microbiological Diagnostic Unit Public Health Laboratory and Victorian Infectious Diseases Reference Laboratory, Doherty Institute |
| EPI_ISL_456433 | Victorian Infectious Diseases Reference Laboratory (VIDRL) | Microbiological Diagnostic Unit Public Health Laboratory and Victorian Infectious Diseases Reference Laboratory, Doherty Institute |
| EPI_ISL_456434 | Victorian Infectious Diseases Reference Laboratory (VIDRL) | Microbiological Diagnostic Unit Public Health Laboratory and Victorian Infectious Diseases Reference Laboratory, Doherty Institute |
| EPI_ISL_456435 | Victorian Infectious Diseases Reference Laboratory (VIDRL) | Microbiological Diagnostic Unit Public Health Laboratory and Victorian Infectious Diseases Reference Laboratory, Doherty Institute |
| EPI_ISL_456436 | Victorian Infectious Diseases Reference Laboratory (VIDRL) | Microbiological Diagnostic Unit Public Health Laboratory and Victorian Infectious Diseases Reference Laboratory, Doherty Institute |

| EPI_Accession_ID | Originating Laboratory | Submitting Laboratory |
|---|---|---|
| EPI_ISL_456437 | Victorian Infectious Diseases Reference Laboratory (VIDRL) | Microbiological Diagnostic Unit Public Health Laboratory and Victorian Infectious Diseases Reference Laboratory, Doherty Institute |
| EPI_ISL_456438 | Victorian Infectious Diseases Reference Laboratory (VIDRL) | Microbiological Diagnostic Unit Public Health Laboratory and Victorian Infectious Diseases Reference Laboratory, Doherty Institute |
| EPI_ISL_456439 | Victorian Infectious Diseases Reference Laboratory (VIDRL) | Microbiological Diagnostic Unit Public Health Laboratory and Victorian Infectious Diseases Reference Laboratory, Doherty Institute |
| EPI_ISL_456440 | Victorian Infectious Diseases Reference Laboratory (VIDRL) | Microbiological Diagnostic Unit Public Health Laboratory and Victorian Infectious Diseases Reference Laboratory, Doherty Institute |
| EPI_ISL_456441 | Victorian Infectious Diseases Reference Laboratory (VIDRL) | Microbiological Diagnostic Unit Public Health Laboratory and Victorian Infectious Diseases Reference Laboratory, Doherty Institute |
| EPI_ISL_456442 | Victorian Infectious Diseases Reference Laboratory (VIDRL) | Microbiological Diagnostic Unit Public Health Laboratory and Victorian Infectious Diseases Reference Laboratory, Doherty Institute |
| EPI_ISL_456443 | Victorian Infectious Diseases Reference Laboratory (VIDRL) | Microbiological Diagnostic Unit Public Health Laboratory and Victorian Infectious Diseases Reference Laboratory, Doherty Institute |
| EPI_ISL_456444 | Victorian Infectious Diseases Reference Laboratory (VIDRL) | Microbiological Diagnostic Unit Public Health Laboratory and Victorian Infectious Diseases Reference Laboratory, Doherty Institute |
| EPI_ISL_456445 | Victorian Infectious Diseases Reference Laboratory (VIDRL) | Microbiological Diagnostic Unit Public Health Laboratory and Victorian Infectious Diseases Reference Laboratory, Doherty Institute |
| EPI_ISL_456446 | Victorian Infectious Diseases Reference Laboratory (VIDRL) | Microbiological Diagnostic Unit Public Health Laboratory and Victorian Infectious Diseases Reference Laboratory, Doherty Institute |
| EPI_ISL_456447 | Victorian Infectious Diseases Reference Laboratory (VIDRL) | Microbiological Diagnostic Unit Public Health Laboratory and Victorian Infectious Diseases Reference Laboratory, Doherty Institute |

| EPI_Accession_ID | Originating Laboratory | Submitting Laboratory |
|---|---|---|
| EPI_ISL_456448 | Victorian Infectious Diseases Reference Laboratory (VIDRL) | Microbiological Diagnostic Unit Public Health Laboratory and Victorian Infectious Diseases Reference Laboratory, Doherty Institute |
| EPI_ISL_456449 | Victorian Infectious Diseases Reference Laboratory (VIDRL) | Microbiological Diagnostic Unit Public Health Laboratory and Victorian Infectious Diseases Reference Laboratory, Doherty Institute |
| EPI_ISL_456450 | Victorian Infectious Diseases Reference Laboratory (VIDRL) | Microbiological Diagnostic Unit Public Health Laboratory and Victorian Infectious Diseases Reference Laboratory, Doherty Institute |
| EPI_ISL_456451 | Victorian Infectious Diseases Reference Laboratory (VIDRL) | Microbiological Diagnostic Unit Public Health Laboratory and Victorian Infectious Diseases Reference Laboratory, Doherty Institute |
| EPI_ISL_456453 | Victorian Infectious Diseases Reference Laboratory (VIDRL) | Microbiological Diagnostic Unit Public Health Laboratory and Victorian Infectious Diseases Reference Laboratory, Doherty Institute |
| EPI_ISL_456454 | Victorian Infectious Diseases Reference Laboratory (VIDRL) | Microbiological Diagnostic Unit Public Health Laboratory and Victorian Infectious Diseases Reference Laboratory, Doherty Institute |
| EPI_ISL_456455 | Victorian Infectious Diseases Reference Laboratory (VIDRL) | Microbiological Diagnostic Unit Public Health Laboratory and Victorian Infectious Diseases Reference Laboratory, Doherty Institute |
| EPI_ISL_456456 | Victorian Infectious Diseases Reference Laboratory (VIDRL) | Microbiological Diagnostic Unit Public Health Laboratory and Victorian Infectious Diseases Reference Laboratory, Doherty Institute |
| EPI_ISL_456457 | Victorian Infectious Diseases Reference Laboratory (VIDRL) | Microbiological Diagnostic Unit Public Health Laboratory and Victorian Infectious Diseases Reference Laboratory, Doherty Institute |
| EPI_ISL_456458 | Victorian Infectious Diseases Reference Laboratory (VIDRL) | Microbiological Diagnostic Unit Public Health Laboratory and Victorian Infectious Diseases Reference Laboratory, Doherty Institute |
| EPI_ISL_456459 | Victorian Infectious Diseases Reference Laboratory (VIDRL) | Microbiological Diagnostic Unit Public Health Laboratory and Victorian Infectious Diseases Reference Laboratory, Doherty Institute |

| EPI_Accession_ID | Originating Laboratory | Submitting Laboratory |
|---|---|---|
| EPI_ISL_456460 | Victorian Infectious Diseases Reference Laboratory (VIDRL) | Microbiological Diagnostic Unit Public Health Laboratory and Victorian Infectious Diseases Reference Laboratory, Doherty Institute |
| EPI_ISL_456461 | Victorian Infectious Diseases Reference Laboratory (VIDRL) | Microbiological Diagnostic Unit Public Health Laboratory and Victorian Infectious Diseases Reference Laboratory, Doherty Institute |
| EPI_ISL_456462 | Victorian Infectious Diseases Reference Laboratory (VIDRL) | Microbiological Diagnostic Unit Public Health Laboratory and Victorian Infectious Diseases Reference Laboratory, Doherty Institute |
| EPI_ISL_456463 | Victorian Infectious Diseases Reference Laboratory (VIDRL) | Microbiological Diagnostic Unit Public Health Laboratory and Victorian Infectious Diseases Reference Laboratory, Doherty Institute |
| EPI_ISL_456464 | Victorian Infectious Diseases Reference Laboratory (VIDRL) | Microbiological Diagnostic Unit Public Health Laboratory and Victorian Infectious Diseases Reference Laboratory, Doherty Institute |
| EPI_ISL_456465 | Victorian Infectious Diseases Reference Laboratory (VIDRL) | Microbiological Diagnostic Unit Public Health Laboratory and Victorian Infectious Diseases Reference Laboratory, Doherty Institute |
| EPI_ISL_456466 | Victorian Infectious Diseases Reference Laboratory (VIDRL) | Microbiological Diagnostic Unit Public Health Laboratory and Victorian Infectious Diseases Reference Laboratory, Doherty Institute |
| EPI_ISL_456467 | Victorian Infectious Diseases Reference Laboratory (VIDRL) | Microbiological Diagnostic Unit Public Health Laboratory and Victorian Infectious Diseases Reference Laboratory, Doherty Institute |
| EPI_ISL_456468 | Victorian Infectious Diseases Reference Laboratory (VIDRL) | Microbiological Diagnostic Unit Public Health Laboratory and Victorian Infectious Diseases Reference Laboratory, Doherty Institute |
| EPI_ISL_456469 | Victorian Infectious Diseases Reference Laboratory (VIDRL) | Microbiological Diagnostic Unit Public Health Laboratory and Victorian Infectious Diseases Reference Laboratory, Doherty Institute |
| EPI_ISL_456470 | Victorian Infectious Diseases Reference Laboratory (VIDRL) | Microbiological Diagnostic Unit Public Health Laboratory and Victorian Infectious Diseases Reference Laboratory, Doherty Institute |

| EPI_Accession_ID | Originating Laboratory | Submitting Laboratory |
|---|---|---|
| EPI_ISL_456471 | Victorian Infectious Diseases Reference Laboratory (VIDRL) | Microbiological Diagnostic Unit Public Health Laboratory and Victorian Infectious Diseases Reference Laboratory, Doherty Institute |
| EPI_ISL_456472 | Victorian Infectious Diseases Reference Laboratory (VIDRL) | Microbiological Diagnostic Unit Public Health Laboratory and Victorian Infectious Diseases Reference Laboratory, Doherty Institute |
| EPI_ISL_456473 | Victorian Infectious Diseases Reference Laboratory (VIDRL) | Microbiological Diagnostic Unit Public Health Laboratory and Victorian Infectious Diseases Reference Laboratory, Doherty Institute |
| EPI_ISL_456474 | Victorian Infectious Diseases Reference Laboratory (VIDRL) | Microbiological Diagnostic Unit Public Health Laboratory and Victorian Infectious Diseases Reference Laboratory, Doherty Institute |
| EPI_ISL_456475 | Victorian Infectious Diseases Reference Laboratory (VIDRL) | Microbiological Diagnostic Unit Public Health Laboratory and Victorian Infectious Diseases Reference Laboratory, Doherty Institute |
| EPI_ISL_456476 | Victorian Infectious Diseases Reference Laboratory (VIDRL) | Microbiological Diagnostic Unit Public Health Laboratory and Victorian Infectious Diseases Reference Laboratory, Doherty Institute |
| EPI_ISL_456477 | Victorian Infectious Diseases Reference Laboratory (VIDRL) | Microbiological Diagnostic Unit Public Health Laboratory and Victorian Infectious Diseases Reference Laboratory, Doherty Institute |
| EPI_ISL_456478 | Microbiological Diagnostic Unit Public Health Laboratory | Microbiological Diagnostic Unit Public Health Laboratory, The Peter Doherty Institute for Infection and Immunity |
| EPI_ISL_456479 | Microbiological Diagnostic Unit Public Health Laboratory | Microbiological Diagnostic Unit Public Health Laboratory, The Peter Doherty Institute for Infection and Immunity |
| EPI_ISL_456480 | Victorian Infectious Diseases Reference Laboratory (VIDRL) | Microbiological Diagnostic Unit Public Health Laboratory and Victorian Infectious Diseases Reference Laboratory, Doherty Institute |
| EPI_ISL_456481 | Victorian Infectious Diseases Reference Laboratory (VIDRL) | Microbiological Diagnostic Unit Public Health Laboratory and Victorian Infectious Diseases Reference Laboratory, Doherty Institute |

| EPI_Accession_ID | Originating Laboratory | Submitting Laboratory |
|---|---|---|
| EPI_ISL_456482 | Victorian Infectious Diseases Reference Laboratory (VIDRL) | Microbiological Diagnostic Unit Public Health Laboratory and Victorian Infectious Diseases Reference Laboratory, Doherty Institute |
| EPI_ISL_456483 | Victorian Infectious Diseases Reference Laboratory (VIDRL) | Microbiological Diagnostic Unit Public Health Laboratory and Victorian Infectious Diseases Reference Laboratory, Doherty Institute |
| EPI_ISL_456484 | Victorian Infectious Diseases Reference Laboratory (VIDRL) | Microbiological Diagnostic Unit Public Health Laboratory and Victorian Infectious Diseases Reference Laboratory, Doherty Institute |
| EPI_ISL_456485 | Victorian Infectious Diseases Reference Laboratory (VIDRL) | Microbiological Diagnostic Unit Public Health Laboratory and Victorian Infectious Diseases Reference Laboratory, Doherty Institute |
| EPI_ISL_456486 | Victorian Infectious Diseases Reference Laboratory (VIDRL) | Microbiological Diagnostic Unit Public Health Laboratory and Victorian Infectious Diseases Reference Laboratory, Doherty Institute |
| EPI_ISL_456487 | Victorian Infectious Diseases Reference Laboratory (VIDRL) | Microbiological Diagnostic Unit Public Health Laboratory and Victorian Infectious Diseases Reference Laboratory, Doherty Institute |
| EPI_ISL_456488 | Victorian Infectious Diseases Reference Laboratory (VIDRL) | Microbiological Diagnostic Unit Public Health Laboratory and Victorian Infectious Diseases Reference Laboratory, Doherty Institute |
| EPI_ISL_456489 | Victorian Infectious Diseases Reference Laboratory (VIDRL) | Microbiological Diagnostic Unit Public Health Laboratory and Victorian Infectious Diseases Reference Laboratory, Doherty Institute |
| EPI_ISL_456490 | Victorian Infectious Diseases Reference Laboratory (VIDRL) | Microbiological Diagnostic Unit Public Health Laboratory and Victorian Infectious Diseases Reference Laboratory, Doherty Institute |
| EPI_ISL_456491 | Victorian Infectious Diseases Reference Laboratory (VIDRL) | Microbiological Diagnostic Unit Public Health Laboratory and Victorian Infectious Diseases Reference Laboratory, Doherty Institute |
| EPI_ISL_456492 | Victorian Infectious Diseases Reference Laboratory (VIDRL) | Microbiological Diagnostic Unit Public Health Laboratory and Victorian Infectious Diseases Reference Laboratory, Doherty Institute |

| EPI_Accession_ID | Originating Laboratory | Submitting Laboratory |
|---|---|---|
| EPI_ISL_456497 | Royal Darwin Hospital Pathology | Microbiological Diagnostic Unit Public Health Laboratory, The Peter Doherty Institute for Infection and Immunity |
| EPI_ISL_456498 | Royal Darwin Hospital Pathology | Microbiological Diagnostic Unit Public Health Laboratory, The Peter Doherty Institute for Infection and Immunity |
| EPI_ISL_456499 | Victorian Infectious Diseases Reference Laboratory (VIDRL) | Microbiological Diagnostic Unit Public Health Laboratory and Victorian Infectious Diseases Reference Laboratory, Doherty Institute |
| EPI_ISL_456500 | Victorian Infectious Diseases Reference Laboratory (VIDRL) | Microbiological Diagnostic Unit Public Health Laboratory and Victorian Infectious Diseases Reference Laboratory, Doherty Institute |
| EPI_ISL_456502 | Victorian Infectious Diseases Reference Laboratory (VIDRL) | Microbiological Diagnostic Unit Public Health Laboratory and Victorian Infectious Diseases Reference Laboratory, Doherty Institute |
| EPI_ISL_456503 | Victorian Infectious Diseases Reference Laboratory (VIDRL) | Microbiological Diagnostic Unit Public Health Laboratory and Victorian Infectious Diseases Reference Laboratory, Doherty Institute |
| EPI_ISL_456504 | Victorian Infectious Diseases Reference Laboratory (VIDRL) | Microbiological Diagnostic Unit Public Health Laboratory and Victorian Infectious Diseases Reference Laboratory, Doherty Institute |
| EPI_ISL_456505 | Victorian Infectious Diseases Reference Laboratory (VIDRL) | Microbiological Diagnostic Unit Public Health Laboratory and Victorian Infectious Diseases Reference Laboratory, Doherty Institute |
| EPI_ISL_456508 | Victorian Infectious Diseases Reference Laboratory (VIDRL) | Microbiological Diagnostic Unit Public Health Laboratory and Victorian Infectious Diseases Reference Laboratory, Doherty Institute |
| EPI_ISL_456509 | Victorian Infectious Diseases Reference Laboratory (VIDRL) | Microbiological Diagnostic Unit Public Health Laboratory and Victorian Infectious Diseases Reference Laboratory, Doherty Institute |
| EPI_ISL_456510 | Victorian Infectious Diseases Reference Laboratory (VIDRL) | Microbiological Diagnostic Unit Public Health Laboratory and Victorian Infectious Diseases Reference Laboratory, Doherty Institute |

| EPI_Accession_ID | Originating Laboratory | Submitting Laboratory |
|---|---|---|
| EPI_ISL_456511 | Victorian Infectious Diseases Reference Laboratory (VIDRL) | Microbiological Diagnostic Unit Public Health Laboratory and Victorian Infectious Diseases Reference Laboratory, Doherty Institute |
| EPI_ISL_456512 | Victorian Infectious Diseases Reference Laboratory (VIDRL) | Microbiological Diagnostic Unit Public Health Laboratory and Victorian Infectious Diseases Reference Laboratory, Doherty Institute |
| EPI_ISL_456513 | Victorian Infectious Diseases Reference Laboratory (VIDRL) | Microbiological Diagnostic Unit Public Health Laboratory and Victorian Infectious Diseases Reference Laboratory, Doherty Institute |
| EPI_ISL_456515 | Victorian Infectious Diseases Reference Laboratory (VIDRL) | Microbiological Diagnostic Unit Public Health Laboratory and Victorian Infectious Diseases Reference Laboratory, Doherty Institute |
| EPI_ISL_456516 | Victorian Infectious Diseases Reference Laboratory (VIDRL) | Microbiological Diagnostic Unit Public Health Laboratory and Victorian Infectious Diseases Reference Laboratory, Doherty Institute |
| EPI_ISL_456517 | Victorian Infectious Diseases Reference Laboratory (VIDRL) | Microbiological Diagnostic Unit Public Health Laboratory and Victorian Infectious Diseases Reference Laboratory, Doherty Institute |
| EPI_ISL_456519 | Victorian Infectious Diseases Reference Laboratory (VIDRL) | Microbiological Diagnostic Unit Public Health Laboratory and Victorian Infectious Diseases Reference Laboratory, Doherty Institute |
| EPI_ISL_456520 | Victorian Infectious Diseases Reference Laboratory (VIDRL) | Microbiological Diagnostic Unit Public Health Laboratory and Victorian Infectious Diseases Reference Laboratory, Doherty Institute |
| EPI_ISL_456521 | Victorian Infectious Diseases Reference Laboratory (VIDRL) | Microbiological Diagnostic Unit Public Health Laboratory and Victorian Infectious Diseases Reference Laboratory, Doherty Institute |
| EPI_ISL_456522 | Victorian Infectious Diseases Reference Laboratory (VIDRL) | Microbiological Diagnostic Unit Public Health Laboratory and Victorian Infectious Diseases Reference Laboratory, Doherty Institute |
| EPI_ISL_456523 | Victorian Infectious Diseases Reference Laboratory (VIDRL) | Microbiological Diagnostic Unit Public Health Laboratory and Victorian Infectious Diseases Reference Laboratory, Doherty Institute |

| EPI_Accession_ID | Originating Laboratory | Submitting Laboratory |
|---|---|---|
| EPI_ISL_456524 | Victorian Infectious Diseases Reference Laboratory (VIDRL) | Microbiological Diagnostic Unit Public Health Laboratory and Victorian Infectious Diseases Reference Laboratory, Doherty Institute |
| EPI_ISL_456525 | Victorian Infectious Diseases Reference Laboratory (VIDRL) | Microbiological Diagnostic Unit Public Health Laboratory and Victorian Infectious Diseases Reference Laboratory, Doherty Institute |
| EPI_ISL_456526 | Victorian Infectious Diseases Reference Laboratory (VIDRL) | Microbiological Diagnostic Unit Public Health Laboratory and Victorian Infectious Diseases Reference Laboratory, Doherty Institute |
| EPI_ISL_456527 | Victorian Infectious Diseases Reference Laboratory (VIDRL) | Microbiological Diagnostic Unit Public Health Laboratory and Victorian Infectious Diseases Reference Laboratory, Doherty Institute |
| EPI_ISL_456528 | Victorian Infectious Diseases Reference Laboratory (VIDRL) | Microbiological Diagnostic Unit Public Health Laboratory and Victorian Infectious Diseases Reference Laboratory, Doherty Institute |
| EPI_ISL_456529 | Victorian Infectious Diseases Reference Laboratory (VIDRL) | Microbiological Diagnostic Unit Public Health Laboratory and Victorian Infectious Diseases Reference Laboratory, Doherty Institute |
| EPI_ISL_456530 | Victorian Infectious Diseases Reference Laboratory (VIDRL) | Microbiological Diagnostic Unit Public Health Laboratory and Victorian Infectious Diseases Reference Laboratory, Doherty Institute |
| EPI_ISL_456531 | Victorian Infectious Diseases Reference Laboratory (VIDRL) | Microbiological Diagnostic Unit Public Health Laboratory and Victorian Infectious Diseases Reference Laboratory, Doherty Institute |
| EPI_ISL_456532 | Victorian Infectious Diseases Reference Laboratory (VIDRL) | Microbiological Diagnostic Unit Public Health Laboratory and Victorian Infectious Diseases Reference Laboratory, Doherty Institute |
| EPI_ISL_456533 | Victorian Infectious Diseases Reference Laboratory (VIDRL) | Microbiological Diagnostic Unit Public Health Laboratory and Victorian Infectious Diseases Reference Laboratory, Doherty Institute |
| EPI_ISL_456534 | Victorian Infectious Diseases Reference Laboratory (VIDRL) | Microbiological Diagnostic Unit Public Health Laboratory and Victorian Infectious Diseases Reference Laboratory, Doherty Institute |

| EPI_Accession_ID | Originating Laboratory | Submitting Laboratory |
|---|---|---|
| EPI_ISL_456535 | Victorian Infectious Diseases Reference Laboratory (VIDRL) | Microbiological Diagnostic Unit Public Health Laboratory and Victorian Infectious Diseases Reference Laboratory, Doherty Institute |
| EPI_ISL_456536 | Victorian Infectious Diseases Reference Laboratory (VIDRL) | Microbiological Diagnostic Unit Public Health Laboratory and Victorian Infectious Diseases Reference Laboratory, Doherty Institute |
| EPI_ISL_456537 | Victorian Infectious Diseases Reference Laboratory (VIDRL) | Microbiological Diagnostic Unit Public Health Laboratory and Victorian Infectious Diseases Reference Laboratory, Doherty Institute |
| EPI_ISL_456538 | Victorian Infectious Diseases Reference Laboratory (VIDRL) | Microbiological Diagnostic Unit Public Health Laboratory and Victorian Infectious Diseases Reference Laboratory, Doherty Institute |
| EPI_ISL_456539 | Victorian Infectious Diseases Reference Laboratory (VIDRL) | Microbiological Diagnostic Unit Public Health Laboratory and Victorian Infectious Diseases Reference Laboratory, Doherty Institute |
| EPI_ISL_456540 | Victorian Infectious Diseases Reference Laboratory (VIDRL) | Microbiological Diagnostic Unit Public Health Laboratory and Victorian Infectious Diseases Reference Laboratory, Doherty Institute |
| EPI_ISL_456541 | Victorian Infectious Diseases Reference Laboratory (VIDRL) | Microbiological Diagnostic Unit Public Health Laboratory and Victorian Infectious Diseases Reference Laboratory, Doherty Institute |
| EPI_ISL_456542 | Victorian Infectious Diseases Reference Laboratory (VIDRL) | Microbiological Diagnostic Unit Public Health Laboratory and Victorian Infectious Diseases Reference Laboratory, Doherty Institute |
| EPI_ISL_456543 | Victorian Infectious Diseases Reference Laboratory (VIDRL) | Microbiological Diagnostic Unit Public Health Laboratory and Victorian Infectious Diseases Reference Laboratory, Doherty Institute |
| EPI_ISL_456544 | Victorian Infectious Diseases Reference Laboratory (VIDRL) | Microbiological Diagnostic Unit Public Health Laboratory and Victorian Infectious Diseases Reference Laboratory, Doherty Institute |
| EPI_ISL_456545 | Victorian Infectious Diseases Reference Laboratory (VIDRL) | Microbiological Diagnostic Unit Public Health Laboratory and Victorian Infectious Diseases Reference Laboratory, Doherty Institute |

| EPI_Accession_ID | Originating Laboratory | Submitting Laboratory |
|---|---|---|
| EPI_ISL_456546 | Victorian Infectious Diseases Reference Laboratory (VIDRL) | Microbiological Diagnostic Unit Public Health Laboratory and Victorian Infectious Diseases Reference Laboratory, Doherty Institute |
| EPI_ISL_456548 | Victorian Infectious Diseases Reference Laboratory (VIDRL) | Microbiological Diagnostic Unit Public Health Laboratory and Victorian Infectious Diseases Reference Laboratory, Doherty Institute |
| EPI_ISL_456550 | Victorian Infectious Diseases Reference Laboratory (VIDRL) | Microbiological Diagnostic Unit Public Health Laboratory and Victorian Infectious Diseases Reference Laboratory, Doherty Institute |
| EPI_ISL_456551 | Victorian Infectious Diseases Reference Laboratory (VIDRL) | Microbiological Diagnostic Unit Public Health Laboratory and Victorian Infectious Diseases Reference Laboratory, Doherty Institute |
| EPI_ISL_456552 | Victorian Infectious Diseases Reference Laboratory (VIDRL) | Microbiological Diagnostic Unit Public Health Laboratory and Victorian Infectious Diseases Reference Laboratory, Doherty Institute |
| EPI_ISL_456553 | Victorian Infectious Diseases Reference Laboratory (VIDRL) | Microbiological Diagnostic Unit Public Health Laboratory and Victorian Infectious Diseases Reference Laboratory, Doherty Institute |
| EPI_ISL_456554 | Victorian Infectious Diseases Reference Laboratory (VIDRL) | Microbiological Diagnostic Unit Public Health Laboratory and Victorian Infectious Diseases Reference Laboratory, Doherty Institute |
| EPI_ISL_456555 | Victorian Infectious Diseases Reference Laboratory (VIDRL) | Microbiological Diagnostic Unit Public Health Laboratory and Victorian Infectious Diseases Reference Laboratory, Doherty Institute |
| EPI_ISL_456556 | Victorian Infectious Diseases Reference Laboratory (VIDRL) | Microbiological Diagnostic Unit Public Health Laboratory and Victorian Infectious Diseases Reference Laboratory, Doherty Institute |
| EPI_ISL_456557 | Victorian Infectious Diseases Reference Laboratory (VIDRL) | Microbiological Diagnostic Unit Public Health Laboratory and Victorian Infectious Diseases Reference Laboratory, Doherty Institute |
| EPI_ISL_456558 | Victorian Infectious Diseases Reference Laboratory (VIDRL) | Microbiological Diagnostic Unit Public Health Laboratory and Victorian Infectious Diseases Reference Laboratory, Doherty Institute |

| EPI_Accession_ID | Originating Laboratory | Submitting Laboratory |
|---|---|---|
| EPI_ISL_456559 | Victorian Infectious Diseases Reference Laboratory (VIDRL) | Microbiological Diagnostic Unit Public Health Laboratory and Victorian Infectious Diseases Reference Laboratory, Doherty Institute |
| EPI_ISL_456560 | Victorian Infectious Diseases Reference Laboratory (VIDRL) | Microbiological Diagnostic Unit Public Health Laboratory and Victorian Infectious Diseases Reference Laboratory, Doherty Institute |
| EPI_ISL_456561 | Victorian Infectious Diseases Reference Laboratory (VIDRL) | Microbiological Diagnostic Unit Public Health Laboratory and Victorian Infectious Diseases Reference Laboratory, Doherty Institute |
| EPI_ISL_456562 | Victorian Infectious Diseases Reference Laboratory (VIDRL) | Microbiological Diagnostic Unit Public Health Laboratory and Victorian Infectious Diseases Reference Laboratory, Doherty Institute |
| EPI_ISL_456563 | Victorian Infectious Diseases Reference Laboratory (VIDRL) | Microbiological Diagnostic Unit Public Health Laboratory and Victorian Infectious Diseases Reference Laboratory, Doherty Institute |
| EPI_ISL_456564 | Victorian Infectious Diseases Reference Laboratory (VIDRL) | Microbiological Diagnostic Unit Public Health Laboratory and Victorian Infectious Diseases Reference Laboratory, Doherty Institute |
| EPI_ISL_456565 | Victorian Infectious Diseases Reference Laboratory (VIDRL) | Microbiological Diagnostic Unit Public Health Laboratory and Victorian Infectious Diseases Reference Laboratory, Doherty Institute |
| EPI_ISL_456566 | Victorian Infectious Diseases Reference Laboratory (VIDRL) | Microbiological Diagnostic Unit Public Health Laboratory and Victorian Infectious Diseases Reference Laboratory, Doherty Institute |
| EPI_ISL_456567 | Victorian Infectious Diseases Reference Laboratory (VIDRL) | Microbiological Diagnostic Unit Public Health Laboratory and Victorian Infectious Diseases Reference Laboratory, Doherty Institute |
| EPI_ISL_456568 | Victorian Infectious Diseases Reference Laboratory (VIDRL) | Microbiological Diagnostic Unit Public Health Laboratory and Victorian Infectious Diseases Reference Laboratory, Doherty Institute |
| EPI_ISL_456569 | Victorian Infectious Diseases Reference Laboratory (VIDRL) | Microbiological Diagnostic Unit Public Health Laboratory and Victorian Infectious Diseases Reference Laboratory, Doherty Institute |

| EPI_Accession_ID | Originating Laboratory | Submitting Laboratory |
|---|---|---|
| EPI_ISL_456570 | Victorian Infectious Diseases Reference Laboratory (VIDRL) | Microbiological Diagnostic Unit Public Health Laboratory and Victorian Infectious Diseases Reference Laboratory, Doherty Institute |
| EPI_ISL_456571 | Victorian Infectious Diseases Reference Laboratory (VIDRL) | Microbiological Diagnostic Unit Public Health Laboratory and Victorian Infectious Diseases Reference Laboratory, Doherty Institute |
| EPI_ISL_456572 | Victorian Infectious Diseases Reference Laboratory (VIDRL) | Microbiological Diagnostic Unit Public Health Laboratory and Victorian Infectious Diseases Reference Laboratory, Doherty Institute |
| EPI_ISL_456573 | Victorian Infectious Diseases Reference Laboratory (VIDRL) | Microbiological Diagnostic Unit Public Health Laboratory and Victorian Infectious Diseases Reference Laboratory, Doherty Institute |
| EPI_ISL_456574 | Victorian Infectious Diseases Reference Laboratory (VIDRL) | Microbiological Diagnostic Unit Public Health Laboratory and Victorian Infectious Diseases Reference Laboratory, Doherty Institute |
| EPI_ISL_456575 | Victorian Infectious Diseases Reference Laboratory (VIDRL) | Microbiological Diagnostic Unit Public Health Laboratory and Victorian Infectious Diseases Reference Laboratory, Doherty Institute |
| EPI_ISL_456577 | Victorian Infectious Diseases Reference Laboratory (VIDRL) | Microbiological Diagnostic Unit Public Health Laboratory and Victorian Infectious Diseases Reference Laboratory, Doherty Institute |
| EPI_ISL_456578 | Victorian Infectious Diseases Reference Laboratory (VIDRL) | Microbiological Diagnostic Unit Public Health Laboratory and Victorian Infectious Diseases Reference Laboratory, Doherty Institute |
| EPI_ISL_456579 | Victorian Infectious Diseases Reference Laboratory (VIDRL) | Microbiological Diagnostic Unit Public Health Laboratory and Victorian Infectious Diseases Reference Laboratory, Doherty Institute |
| EPI_ISL_456580 | Victorian Infectious Diseases Reference Laboratory (VIDRL) | Microbiological Diagnostic Unit Public Health Laboratory and Victorian Infectious Diseases Reference Laboratory, Doherty Institute |
| EPI_ISL_456581 | Victorian Infectious Diseases Reference Laboratory (VIDRL) | Microbiological Diagnostic Unit Public Health Laboratory and Victorian Infectious Diseases Reference Laboratory, Doherty Institute |

| EPI_Accession_ID | Originating Laboratory | Submitting Laboratory |
|---|---|---|
| EPI_ISL_456582 | Victorian Infectious Diseases Reference Laboratory (VIDRL) | Microbiological Diagnostic Unit Public Health Laboratory and Victorian Infectious Diseases Reference Laboratory, Doherty Institute |
| EPI_ISL_456583 | Victorian Infectious Diseases Reference Laboratory (VIDRL) | Microbiological Diagnostic Unit Public Health Laboratory and Victorian Infectious Diseases Reference Laboratory, Doherty Institute |
| EPI_ISL_456584 | Victorian Infectious Diseases Reference Laboratory (VIDRL) | Microbiological Diagnostic Unit Public Health Laboratory and Victorian Infectious Diseases Reference Laboratory, Doherty Institute |
| EPI_ISL_456585 | Victorian Infectious Diseases Reference Laboratory (VIDRL) | Microbiological Diagnostic Unit Public Health Laboratory and Victorian Infectious Diseases Reference Laboratory, Doherty Institute |
| EPI_ISL_456586 | Victorian Infectious Diseases Reference Laboratory (VIDRL) | Microbiological Diagnostic Unit Public Health Laboratory and Victorian Infectious Diseases Reference Laboratory, Doherty Institute |
| EPI_ISL_456587 | Victorian Infectious Diseases Reference Laboratory (VIDRL) | Microbiological Diagnostic Unit Public Health Laboratory and Victorian Infectious Diseases Reference Laboratory, Doherty Institute |
| EPI_ISL_456588 | Victorian Infectious Diseases Reference Laboratory (VIDRL) | Microbiological Diagnostic Unit Public Health Laboratory and Victorian Infectious Diseases Reference Laboratory, Doherty Institute |
| EPI_ISL_456589 | Victorian Infectious Diseases Reference Laboratory (VIDRL) | Microbiological Diagnostic Unit Public Health Laboratory and Victorian Infectious Diseases Reference Laboratory, Doherty Institute |
| EPI_ISL_456590 | Victorian Infectious Diseases Reference Laboratory (VIDRL) | Microbiological Diagnostic Unit Public Health Laboratory and Victorian Infectious Diseases Reference Laboratory, Doherty Institute |
| EPI_ISL_456591 | Victorian Infectious Diseases Reference Laboratory (VIDRL) | Microbiological Diagnostic Unit Public Health Laboratory and Victorian Infectious Diseases Reference Laboratory, Doherty Institute |
| EPI_ISL_456592 | Victorian Infectious Diseases Reference Laboratory (VIDRL) | Microbiological Diagnostic Unit Public Health Laboratory and Victorian Infectious Diseases Reference Laboratory, Doherty Institute |

| EPI_Accession_ID | Originating Laboratory | Submitting Laboratory |
|---|---|---|
| EPI_ISL_456593 | Victorian Infectious Diseases Reference Laboratory (VIDRL) | Microbiological Diagnostic Unit Public Health Laboratory and Victorian Infectious Diseases Reference Laboratory, Doherty Institute |
| EPI_ISL_456594 | Microbiological Diagnostic Unit Public Health Laboratory | Microbiological Diagnostic Unit Public Health Laboratory, The Peter Doherty Institute for Infection and Immunity |
| EPI_ISL_456595 | Microbiological Diagnostic Unit Public Health Laboratory | Microbiological Diagnostic Unit Public Health Laboratory, The Peter Doherty Institute for Infection and Immunity |
| EPI_ISL_456597 | National Health Laboratory, Timor-Leste | Microbiological Diagnostic Unit Public Health Laboratory, The Peter Doherty Institute for Infection and Immunity |
| EPI_ISL_456598 | National Health Laboratory, Timor-Leste | Microbiological Diagnostic Unit Public Health Laboratory, The Peter Doherty Institute for Infection and Immunity |
| EPI_ISL_456599 | National Health Laboratory, Timor-Leste | Microbiological Diagnostic Unit Public Health Laboratory, The Peter Doherty Institute for Infection and Immunity |
| EPI_ISL_456600 | National Health Laboratory, Timor-Leste | Microbiological Diagnostic Unit Public Health Laboratory, The Peter Doherty Institute for Infection and Immunity |
| EPI_ISL_456601 | National Health Laboratory, Timor-Leste | Microbiological Diagnostic Unit Public Health Laboratory, The Peter Doherty Institute for Infection and Immunity |
| EPI_ISL_456602 | National Health Laboratory, Timor-Leste | Microbiological Diagnostic Unit Public Health Laboratory, The Peter Doherty Institute for Infection and Immunity |
| EPI_ISL_456603 | National Health Laboratory, Timor-Leste | Microbiological Diagnostic Unit Public Health Laboratory, The Peter Doherty Institute for Infection and Immunity |
| EPI_ISL_456604 | National Health Laboratory, Timor-Leste | Microbiological Diagnostic Unit Public Health Laboratory, The Peter Doherty Institute for Infection and Immunity |

| EPI_Accession_ID | Originating Laboratory | Submitting Laboratory |
|---|---|---|
| EPI_ISL_456605 | National Health Laboratory, Timor-Leste | Microbiological Diagnostic Unit Public Health Laboratory, The Peter Doherty Institute for Infection and Immunity |
| EPI_ISL_456606 | National Health Laboratory, Timor-Leste | Microbiological Diagnostic Unit Public Health Laboratory, The Peter Doherty Institute for Infection and Immunity |
| EPI_ISL_456607 | National Health Laboratory, Timor-Leste | Microbiological Diagnostic Unit Public Health Laboratory, The Peter Doherty Institute for Infection and Immunity |
| EPI_ISL_456608 | National Health Laboratory, Timor-Leste | Microbiological Diagnostic Unit Public Health Laboratory, The Peter Doherty Institute for Infection and Immunity |
| EPI_ISL_456609 | National Health Laboratory, Timor-Leste | Microbiological Diagnostic Unit Public Health Laboratory, The Peter Doherty Institute for Infection and Immunity |
| EPI_ISL_456610 | National Health Laboratory, Timor-Leste | Microbiological Diagnostic Unit Public Health Laboratory, The Peter Doherty Institute for Infection and Immunity |
| EPI_ISL_456611 | National Health Laboratory, Timor-Leste | Microbiological Diagnostic Unit Public Health Laboratory, The Peter Doherty Institute for Infection and Immunity |
| EPI_ISL_456612 | National Health Laboratory, Timor-Leste | Microbiological Diagnostic Unit Public Health Laboratory, The Peter Doherty Institute for Infection and Immunity |
| EPI_ISL_456613 | Victorian Infectious Diseases Reference Laboratory (VIDRL) | Microbiological Diagnostic Unit Public Health Laboratory and Victorian Infectious Diseases Reference Laboratory, Doherty Institute |
| EPI_ISL_456614 | Victorian Infectious Diseases Reference Laboratory (VIDRL) | Microbiological Diagnostic Unit Public Health Laboratory and Victorian Infectious Diseases Reference Laboratory, Doherty Institute |
| EPI_ISL_456615 | Victorian Infectious Diseases Reference Laboratory (VIDRL) | Microbiological Diagnostic Unit Public Health Laboratory and Victorian Infectious Diseases Reference Laboratory, Doherty Institute |

| EPI_Accession_ID | Originating Laboratory | Submitting Laboratory |
|---|---|---|
| EPI_ISL_456616 | Victorian Infectious Diseases Reference Laboratory (VIDRL) | Microbiological Diagnostic Unit Public Health Laboratory and Victorian Infectious Diseases Reference Laboratory, Doherty Institute |
| EPI_ISL_456617 | Victorian Infectious Diseases Reference Laboratory (VIDRL) | Microbiological Diagnostic Unit Public Health Laboratory and Victorian Infectious Diseases Reference Laboratory, Doherty Institute |
| EPI_ISL_456618 | Victorian Infectious Diseases Reference Laboratory (VIDRL) | Microbiological Diagnostic Unit Public Health Laboratory and Victorian Infectious Diseases Reference Laboratory, Doherty Institute |
| EPI_ISL_456619 | Victorian Infectious Diseases Reference Laboratory (VIDRL) | Microbiological Diagnostic Unit Public Health Laboratory and Victorian Infectious Diseases Reference Laboratory, Doherty Institute |
| EPI_ISL_456620 | Victorian Infectious Diseases Reference Laboratory (VIDRL) | Microbiological Diagnostic Unit Public Health Laboratory and Victorian Infectious Diseases Reference Laboratory, Doherty Institute |
| EPI_ISL_456621 | Victorian Infectious Diseases Reference Laboratory (VIDRL) | Microbiological Diagnostic Unit Public Health Laboratory and Victorian Infectious Diseases Reference Laboratory, Doherty Institute |
| EPI_ISL_456622 | Victorian Infectious Diseases Reference Laboratory (VIDRL) | Microbiological Diagnostic Unit Public Health Laboratory and Victorian Infectious Diseases Reference Laboratory, Doherty Institute |
| EPI_ISL_456625 | Victorian Infectious Diseases Reference Laboratory (VIDRL) | Microbiological Diagnostic Unit Public Health Laboratory and Victorian Infectious Diseases Reference Laboratory, Doherty Institute |
| EPI_ISL_456626 | Victorian Infectious Diseases Reference Laboratory (VIDRL) | Microbiological Diagnostic Unit Public Health Laboratory and Victorian Infectious Diseases Reference Laboratory, Doherty Institute |
| EPI_ISL_456627 | Victorian Infectious Diseases Reference Laboratory (VIDRL) | Microbiological Diagnostic Unit Public Health Laboratory and Victorian Infectious Diseases Reference Laboratory, Doherty Institute |
| EPI_ISL_456628 | Victorian Infectious Diseases Reference Laboratory (VIDRL) | Microbiological Diagnostic Unit Public Health Laboratory and Victorian Infectious Diseases Reference Laboratory, Doherty Institute |

| EPI_Accession_ID | Originating Laboratory | Submitting Laboratory |
|---|---|---|
| EPI_ISL_456629 | Victorian Infectious Diseases Reference Laboratory (VIDRL) | Microbiological Diagnostic Unit Public Health Laboratory and Victorian Infectious Diseases Reference Laboratory, Doherty Institute |
| EPI_ISL_456630 | Victorian Infectious Diseases Reference Laboratory (VIDRL) | Microbiological Diagnostic Unit Public Health Laboratory and Victorian Infectious Diseases Reference Laboratory, Doherty Institute |
| EPI_ISL_456631 | Victorian Infectious Diseases Reference Laboratory (VIDRL) | Microbiological Diagnostic Unit Public Health Laboratory and Victorian Infectious Diseases Reference Laboratory, Doherty Institute |
| EPI_ISL_456632 | Victorian Infectious Diseases Reference Laboratory (VIDRL) | Microbiological Diagnostic Unit Public Health Laboratory and Victorian Infectious Diseases Reference Laboratory, Doherty Institute |
| EPI_ISL_456633 | Victorian Infectious Diseases Reference Laboratory (VIDRL) | Microbiological Diagnostic Unit Public Health Laboratory and Victorian Infectious Diseases Reference Laboratory, Doherty Institute |
| EPI_ISL_456634 | Victorian Infectious Diseases Reference Laboratory (VIDRL) | Microbiological Diagnostic Unit Public Health Laboratory and Victorian Infectious Diseases Reference Laboratory, Doherty Institute |
| EPI_ISL_456635 | Victorian Infectious Diseases Reference Laboratory (VIDRL) | Microbiological Diagnostic Unit Public Health Laboratory and Victorian Infectious Diseases Reference Laboratory, Doherty Institute |
| EPI_ISL_456636 | Victorian Infectious Diseases Reference Laboratory (VIDRL) | Microbiological Diagnostic Unit Public Health Laboratory and Victorian Infectious Diseases Reference Laboratory, Doherty Institute |
| EPI_ISL_456637 | Victorian Infectious Diseases Reference Laboratory (VIDRL) | Microbiological Diagnostic Unit Public Health Laboratory and Victorian Infectious Diseases Reference Laboratory, Doherty Institute |
| EPI_ISL_456638 | Victorian Infectious Diseases Reference Laboratory (VIDRL) | Microbiological Diagnostic Unit Public Health Laboratory and Victorian Infectious Diseases Reference Laboratory, Doherty Institute |
| EPI_ISL_456639 | Victorian Infectious Diseases Reference Laboratory (VIDRL) | Microbiological Diagnostic Unit Public Health Laboratory and Victorian Infectious Diseases Reference Laboratory, Doherty Institute |

| EPI_Accession_ID | Originating Laboratory | Submitting Laboratory |
|---|---|---|
| EPI_ISL_456640 | Victorian Infectious Diseases Reference Laboratory (VIDRL) | Microbiological Diagnostic Unit Public Health Laboratory and Victorian Infectious Diseases Reference Laboratory, Doherty Institute |
| EPI_ISL_456641 | Victorian Infectious Diseases Reference Laboratory (VIDRL) | Microbiological Diagnostic Unit Public Health Laboratory and Victorian Infectious Diseases Reference Laboratory, Doherty Institute |
| EPI_ISL_456642 | Victorian Infectious Diseases Reference Laboratory (VIDRL) | Microbiological Diagnostic Unit Public Health Laboratory and Victorian Infectious Diseases Reference Laboratory, Doherty Institute |
| EPI_ISL_456643 | Victorian Infectious Diseases Reference Laboratory (VIDRL) | Microbiological Diagnostic Unit Public Health Laboratory and Victorian Infectious Diseases Reference Laboratory, Doherty Institute |
| EPI_ISL_456644 | Victorian Infectious Diseases Reference Laboratory (VIDRL) | Microbiological Diagnostic Unit Public Health Laboratory and Victorian Infectious Diseases Reference Laboratory, Doherty Institute |
| EPI_ISL_456645 | Victorian Infectious Diseases Reference Laboratory (VIDRL) | Microbiological Diagnostic Unit Public Health Laboratory and Victorian Infectious Diseases Reference Laboratory, Doherty Institute |
| EPI_ISL_456646 | Victorian Infectious Diseases Reference Laboratory (VIDRL) | Microbiological Diagnostic Unit Public Health Laboratory and Victorian Infectious Diseases Reference Laboratory, Doherty Institute |
| EPI_ISL_456647 | Victorian Infectious Diseases Reference Laboratory (VIDRL) | Microbiological Diagnostic Unit Public Health Laboratory and Victorian Infectious Diseases Reference Laboratory, Doherty Institute |
| EPI_ISL_456648 | Victorian Infectious Diseases Reference Laboratory (VIDRL) | Microbiological Diagnostic Unit Public Health Laboratory and Victorian Infectious Diseases Reference Laboratory, Doherty Institute |
| EPI_ISL_456649 | Victorian Infectious Diseases Reference Laboratory (VIDRL) | Microbiological Diagnostic Unit Public Health Laboratory and Victorian Infectious Diseases Reference Laboratory, Doherty Institute |
| EPI_ISL_456650 | Victorian Infectious Diseases Reference Laboratory (VIDRL) | Microbiological Diagnostic Unit Public Health Laboratory and Victorian Infectious Diseases Reference Laboratory, Doherty Institute |

| EPI_Accession_ID | Originating Laboratory | Submitting Laboratory |
|---|---|---|
| EPI_ISL_456651 | Victorian Infectious Diseases Reference Laboratory (VIDRL) | Microbiological Diagnostic Unit Public Health Laboratory and Victorian Infectious Diseases Reference Laboratory, Doherty Institute |
| EPI_ISL_456652 | Victorian Infectious Diseases Reference Laboratory (VIDRL) | Microbiological Diagnostic Unit Public Health Laboratory and Victorian Infectious Diseases Reference Laboratory, Doherty Institute |
| EPI_ISL_456653 | Victorian Infectious Diseases Reference Laboratory (VIDRL) | Microbiological Diagnostic Unit Public Health Laboratory and Victorian Infectious Diseases Reference Laboratory, Doherty Institute |
| EPI_ISL_456654 | Victorian Infectious Diseases Reference Laboratory (VIDRL) | Microbiological Diagnostic Unit Public Health Laboratory and Victorian Infectious Diseases Reference Laboratory, Doherty Institute |
| EPI_ISL_456655 | Victorian Infectious Diseases Reference Laboratory (VIDRL) | Microbiological Diagnostic Unit Public Health Laboratory and Victorian Infectious Diseases Reference Laboratory, Doherty Institute |
| EPI_ISL_456656 | Sequencing and Bioinformatics Center | Sequencing and Bioinformatics Center |
| EPI_ISL_456666 | University of Birmingham | COVID-19 Genomics UK (COG-UK) Consortium |
| EPI_ISL_456667 | University of Birmingham | COVID-19 Genomics UK (COG-UK) Consortium |
| EPI_ISL_456668 | University of Birmingham | COVID-19 Genomics UK (COG-UK) Consortium |
| EPI_ISL_456669 | University of Birmingham | COVID-19 Genomics UK (COG-UK) Consortium |
| EPI_ISL_456670 | University of Birmingham | COVID-19 Genomics UK (COG-UK) Consortium |
| EPI_ISL_456671 | University of Birmingham | COVID-19 Genomics UK (COG-UK) Consortium |
| EPI_ISL_456672 | University of Birmingham | COVID-19 Genomics UK (COG-UK) Consortium |
| EPI_ISL_456673 | University of Birmingham | COVID-19 Genomics UK (COG-UK) Consortium |
| EPI_ISL_456674 | University of Birmingham | COVID-19 Genomics UK (COG-UK) Consortium |
| EPI_ISL_456675 | University of Birmingham | COVID-19 Genomics UK (COG-UK) Consortium |
| EPI_ISL_456676 | University of Birmingham | COVID-19 Genomics UK (COG-UK) Consortium |
| EPI_ISL_456677 | Department of Pathology, University of Cambridge | COVID-19 Genomics UK (COG-UK) Consortium |
| EPI_ISL_456678 | Department of Pathology, University of Cambridge | COVID-19 Genomics UK (COG-UK) Consortium |
| EPI_ISL_456679 | Department of Pathology, University of Cambridge | COVID-19 Genomics UK (COG-UK) Consortium |
| EPI_ISL_456680 | Department of Pathology, University of Cambridge | COVID-19 Genomics UK (COG-UK) Consortium |
| EPI_ISL_456681 | Department of Pathology, University of Cambridge | COVID-19 Genomics UK (COG-UK) Consortium |
| EPI_ISL_456682 | Department of Pathology, University of Cambridge | COVID-19 Genomics UK (COG-UK) Consortium |

| EPI_Accession_ID | Originating Laboratory | Submitting Laboratory |
|---|---|---|
| EPI_ISL_456686 | Department of Pathology, University of Cambridge | COVID-19 Genomics UK (COG-UK) Consortium |
| EPI_ISL_456688 | Department of Pathology, University of Cambridge | COVID-19 Genomics UK (COG-UK) Consortium |
| EPI_ISL_456690 | Department of Pathology, University of Cambridge | COVID-19 Genomics UK (COG-UK) Consortium |
| EPI_ISL_456692 | Department of Pathology, University of Cambridge | COVID-19 Genomics UK (COG-UK) Consortium |
| EPI_ISL_456695 | Department of Pathology, University of Cambridge | COVID-19 Genomics UK (COG-UK) Consortium |
| EPI_ISL_456696 | Department of Pathology, University of Cambridge | COVID-19 Genomics UK (COG-UK) Consortium |
| EPI_ISL_456697 | Department of Pathology, University of Cambridge | COVID-19 Genomics UK (COG-UK) Consortium |
| EPI_ISL_456698 | Department of Pathology, University of Cambridge | COVID-19 Genomics UK (COG-UK) Consortium |
| EPI_ISL_456699 | Department of Pathology, University of Cambridge | COVID-19 Genomics UK (COG-UK) Consortium |
| EPI_ISL_456700 | Department of Pathology, University of Cambridge | COVID-19 Genomics UK (COG-UK) Consortium |
| EPI_ISL_456701 | Department of Pathology, University of Cambridge | COVID-19 Genomics UK (COG-UK) Consortium |
| EPI_ISL_456702 | Department of Pathology, University of Cambridge | COVID-19 Genomics UK (COG-UK) Consortium |
| EPI_ISL_456703 | Department of Pathology, University of Cambridge | COVID-19 Genomics UK (COG-UK) Consortium |
| EPI_ISL_456704 | Department of Pathology, University of Cambridge | COVID-19 Genomics UK (COG-UK) Consortium |
| EPI_ISL_456705 | Department of Pathology, University of Cambridge | COVID-19 Genomics UK (COG-UK) Consortium |
| EPI_ISL_456706 | Department of Pathology, University of Cambridge | COVID-19 Genomics UK (COG-UK) Consortium |
| EPI_ISL_456708 | Department of Pathology, University of Cambridge | COVID-19 Genomics UK (COG-UK) Consortium |
| EPI_ISL_456709 | Department of Pathology, University of Cambridge | COVID-19 Genomics UK (COG-UK) Consortium |
| EPI_ISL_456710 | Department of Pathology, University of Cambridge | COVID-19 Genomics UK (COG-UK) Consortium |
| EPI_ISL_456711 | Department of Pathology, University of Cambridge | COVID-19 Genomics UK (COG-UK) Consortium |
| EPI_ISL_456712 | Department of Pathology, University of Cambridge | COVID-19 Genomics UK (COG-UK) Consortium |
| EPI_ISL_456713 | Department of Pathology, University of Cambridge | COVID-19 Genomics UK (COG-UK) Consortium |
| EPI_ISL_456714 | Department of Pathology, University of Cambridge | COVID-19 Genomics UK (COG-UK) Consortium |
| EPI_ISL_456715 | Department of Pathology, University of Cambridge | COVID-19 Genomics UK (COG-UK) Consortium |
| EPI_ISL_456716 | Department of Pathology, University of Cambridge | COVID-19 Genomics UK (COG-UK) Consortium |
| EPI_ISL_456717 | Department of Pathology, University of Cambridge | COVID-19 Genomics UK (COG-UK) Consortium |
| EPI_ISL_456718 | Department of Pathology, University of Cambridge | COVID-19 Genomics UK (COG-UK) Consortium |
| EPI_ISL_456719 | Department of Pathology, University of Cambridge | COVID-19 Genomics UK (COG-UK) Consortium |
| EPI_ISL_456720 | Department of Pathology, University of Cambridge | COVID-19 Genomics UK (COG-UK) Consortium |
| EPI_ISL_456721 | Department of Pathology, University of Cambridge | COVID-19 Genomics UK (COG-UK) Consortium |
| EPI_ISL_456722 | Department of Pathology, University of Cambridge | COVID-19 Genomics UK (COG-UK) Consortium |
| EPI_ISL_456723 | Department of Pathology, University of Cambridge | COVID-19 Genomics UK (COG-UK) Consortium |
| EPI_ISL_456724 | Department of Pathology, University of Cambridge | COVID-19 Genomics UK (COG-UK) Consortium |

| EPI_Accession_ID | Originating Laboratory | Submitting Laboratory |
|---|---|---|
| EPI_ISL_456725 | Department of Pathology, University of Cambridge | COVID-19 Genomics UK (COG-UK) Consortium |
| EPI_ISL_456726 | Department of Pathology, University of Cambridge | COVID-19 Genomics UK (COG-UK) Consortium |
| EPI_ISL_456727 | Department of Pathology, University of Cambridge | COVID-19 Genomics UK (COG-UK) Consortium |
| EPI_ISL_456728 | Department of Pathology, University of Cambridge | COVID-19 Genomics UK (COG-UK) Consortium |
| EPI_ISL_456729 | Department of Pathology, University of Cambridge | COVID-19 Genomics UK (COG-UK) Consortium |
| EPI_ISL_456730 | Department of Pathology, University of Cambridge | COVID-19 Genomics UK (COG-UK) Consortium |
| EPI_ISL_456731 | Department of Pathology, University of Cambridge | COVID-19 Genomics UK (COG-UK) Consortium |
| EPI_ISL_456732 | Department of Pathology, University of Cambridge | COVID-19 Genomics UK (COG-UK) Consortium |
| EPI_ISL_456733 | Department of Pathology, University of Cambridge | COVID-19 Genomics UK (COG-UK) Consortium |
| EPI_ISL_456735 | Department of Pathology, University of Cambridge | COVID-19 Genomics UK (COG-UK) Consortium |
| EPI_ISL_456736 | Department of Pathology, University of Cambridge | COVID-19 Genomics UK (COG-UK) Consortium |
| EPI_ISL_456737 | Department of Pathology, University of Cambridge | COVID-19 Genomics UK (COG-UK) Consortium |
| EPI_ISL_456738 | Department of Pathology, University of Cambridge | COVID-19 Genomics UK (COG-UK) Consortium |
| EPI_ISL_456739 | Department of Pathology, University of Cambridge | COVID-19 Genomics UK (COG-UK) Consortium |
| EPI_ISL_456740 | Department of Pathology, University of Cambridge | COVID-19 Genomics UK (COG-UK) Consortium |
| EPI_ISL_456741 | Department of Pathology, University of Cambridge | COVID-19 Genomics UK (COG-UK) Consortium |
| EPI_ISL_456742 | Department of Pathology, University of Cambridge | COVID-19 Genomics UK (COG-UK) Consortium |
| EPI_ISL_456743 | Department of Pathology, University of Cambridge | COVID-19 Genomics UK (COG-UK) Consortium |
| EPI_ISL_456744 | Department of Pathology, University of Cambridge | COVID-19 Genomics UK (COG-UK) Consortium |
| EPI_ISL_456745 | Department of Pathology, University of Cambridge | COVID-19 Genomics UK (COG-UK) Consortium |
| EPI_ISL_456746 | Department of Pathology, University of Cambridge | COVID-19 Genomics UK (COG-UK) Consortium |
| EPI_ISL_456747 | Department of Pathology, University of Cambridge | COVID-19 Genomics UK (COG-UK) Consortium |
| EPI_ISL_456748 | Department of Pathology, University of Cambridge | COVID-19 Genomics UK (COG-UK) Consortium |
| EPI_ISL_456749 | Department of Pathology, University of Cambridge | COVID-19 Genomics UK (COG-UK) Consortium |
| EPI_ISL_456750 | Department of Pathology, University of Cambridge | COVID-19 Genomics UK (COG-UK) Consortium |
| EPI_ISL_456751 | Department of Pathology, University of Cambridge | COVID-19 Genomics UK (COG-UK) Consortium |
| EPI_ISL_456752 | Department of Pathology, University of Cambridge | COVID-19 Genomics UK (COG-UK) Consortium |
| EPI_ISL_456753 | Department of Pathology, University of Cambridge | COVID-19 Genomics UK (COG-UK) Consortium |
| EPI_ISL_456754 | Department of Pathology, University of Cambridge | COVID-19 Genomics UK (COG-UK) Consortium |
| EPI_ISL_456755 | Department of Pathology, University of Cambridge | COVID-19 Genomics UK (COG-UK) Consortium |
| EPI_ISL_456756 | West of Scotland Specialist Virology Centre, NHSGGC / MRC-University of Glasgow Centre for Virus Research | COVID-19 Genomics UK (COG-UK) Consortium |

| EPI_Accession_ID | Originating Laboratory | Submitting Laboratory |
|---|---|---|
| EPI_ISL_456757 | West of Scotland Specialist Virology Centre, NHSGGC / MRC-University of Glasgow Centre for Virus Research | COVID-19 Genomics UK (COG-UK) Consortium |
| EPI_ISL_456758 | West of Scotland Specialist Virology Centre, NHSGGC / MRC-University of Glasgow Centre for Virus Research | COVID-19 Genomics UK (COG-UK) Consortium |
| EPI_ISL_456759 | West of Scotland Specialist Virology Centre, NHSGGC / MRC-University of Glasgow Centre for Virus Research | COVID-19 Genomics UK (COG-UK) Consortium |
| EPI_ISL_456760 | West of Scotland Specialist Virology Centre, NHSGGC / MRC-University of Glasgow Centre for Virus Research | COVID-19 Genomics UK (COG-UK) Consortium |
| EPI_ISL_456761 | West of Scotland Specialist Virology Centre, NHSGGC / MRC-University of Glasgow Centre for Virus Research | COVID-19 Genomics UK (COG-UK) Consortium |
| EPI_ISL_456762 | West of Scotland Specialist Virology Centre, NHSGGC / MRC-University of Glasgow Centre for Virus Research | COVID-19 Genomics UK (COG-UK) Consortium |
| EPI_ISL_456763 | West of Scotland Specialist Virology Centre, NHSGGC / MRC-University of Glasgow Centre for Virus Research | COVID-19 Genomics UK (COG-UK) Consortium |
| EPI_ISL_456764 | West of Scotland Specialist Virology Centre, NHSGGC / MRC-University of Glasgow Centre for Virus Research | COVID-19 Genomics UK (COG-UK) Consortium |
| EPI_ISL_456765 | West of Scotland Specialist Virology Centre, NHSGGC / MRC-University of Glasgow Centre for Virus Research | COVID-19 Genomics UK (COG-UK) Consortium |
| EPI_ISL_456766 | West of Scotland Specialist Virology Centre, NHSGGC / MRC-University of Glasgow Centre for Virus Research | COVID-19 Genomics UK (COG-UK) Consortium |
| EPI_ISL_456767 | West of Scotland Specialist Virology Centre, NHSGGC / MRC-University of Glasgow Centre for Virus Research | COVID-19 Genomics UK (COG-UK) Consortium |
| EPI_ISL_456768 | West of Scotland Specialist Virology Centre, NHSGGC / MRC-University of Glasgow Centre for Virus Research | COVID-19 Genomics UK (COG-UK) Consortium |
| EPI_ISL_456769 | West of Scotland Specialist Virology Centre, NHSGGC / MRC-University of Glasgow Centre for Virus Research | COVID-19 Genomics UK (COG-UK) Consortium |
| EPI_ISL_456770 | West of Scotland Specialist Virology Centre, NHSGGC / MRC-University of Glasgow Centre for Virus Research | COVID-19 Genomics UK (COG-UK) Consortium |
| EPI_ISL_456771 | West of Scotland Specialist Virology Centre, NHSGGC / MRC-University of Glasgow Centre for Virus Research | COVID-19 Genomics UK (COG-UK) Consortium |
| EPI_ISL_456772 | West of Scotland Specialist Virology Centre, NHSGGC / MRC-University of Glasgow Centre for Virus Research | COVID-19 Genomics UK (COG-UK) Consortium |

| EPI_Accession_ID | Originating Laboratory | Submitting Laboratory |
|---|---|---|
| EPI_ISL_456773 | West of Scotland Specialist Virology Centre, NHSGGC / MRC-University of Glasgow Centre for Virus Research | COVID-19 Genomics UK (COG-UK) Consortium |
| EPI_ISL_456774 | West of Scotland Specialist Virology Centre, NHSGGC / MRC-University of Glasgow Centre for Virus Research | COVID-19 Genomics UK (COG-UK) Consortium |
| EPI_ISL_456775 | West of Scotland Specialist Virology Centre, NHSGGC / MRC-University of Glasgow Centre for Virus Research | COVID-19 Genomics UK (COG-UK) Consortium |
| EPI_ISL_456776 | West of Scotland Specialist Virology Centre, NHSGGC / MRC-University of Glasgow Centre for Virus Research | COVID-19 Genomics UK (COG-UK) Consortium |
| EPI_ISL_456777 | West of Scotland Specialist Virology Centre, NHSGGC / MRC-University of Glasgow Centre for Virus Research | COVID-19 Genomics UK (COG-UK) Consortium |
| EPI_ISL_456778 | West of Scotland Specialist Virology Centre, NHSGGC / MRC-University of Glasgow Centre for Virus Research | COVID-19 Genomics UK (COG-UK) Consortium |
| EPI_ISL_456779 | West of Scotland Specialist Virology Centre, NHSGGC / MRC-University of Glasgow Centre for Virus Research | COVID-19 Genomics UK (COG-UK) Consortium |
| EPI_ISL_456780 | West of Scotland Specialist Virology Centre, NHSGGC / MRC-University of Glasgow Centre for Virus Research | COVID-19 Genomics UK (COG-UK) Consortium |
| EPI_ISL_456781 | West of Scotland Specialist Virology Centre, NHSGGC / MRC-University of Glasgow Centre for Virus Research | COVID-19 Genomics UK (COG-UK) Consortium |
| EPI_ISL_456782 | West of Scotland Specialist Virology Centre, NHSGGC / MRC-University of Glasgow Centre for Virus Research | COVID-19 Genomics UK (COG-UK) Consortium |
| EPI_ISL_456783 | West of Scotland Specialist Virology Centre, NHSGGC / MRC-University of Glasgow Centre for Virus Research | COVID-19 Genomics UK (COG-UK) Consortium |
| EPI_ISL_456784 | West of Scotland Specialist Virology Centre, NHSGGC / MRC-University of Glasgow Centre for Virus Research | COVID-19 Genomics UK (COG-UK) Consortium |
| EPI_ISL_456785 | West of Scotland Specialist Virology Centre, NHSGGC / MRC-University of Glasgow Centre for Virus Research | COVID-19 Genomics UK (COG-UK) Consortium |
| EPI_ISL_456786 | West of Scotland Specialist Virology Centre, NHSGGC / MRC-University of Glasgow Centre for Virus Research | COVID-19 Genomics UK (COG-UK) Consortium |
| EPI_ISL_456787 | West of Scotland Specialist Virology Centre, NHSGGC / MRC-University of Glasgow Centre for Virus Research | COVID-19 Genomics UK (COG-UK) Consortium |
| EPI_ISL_456788 | West of Scotland Specialist Virology Centre, NHSGGC / MRC-University of Glasgow Centre for Virus Research | COVID-19 Genomics UK (COG-UK) Consortium |

| EPI_Accession_ID | Originating Laboratory | Submitting Laboratory |
|---|---|---|
| EPI_ISL_456789 | West of Scotland Specialist Virology Centre, NHSGGC / MRC-University of Glasgow Centre for Virus Research | COVID-19 Genomics UK (COG-UK) Consortium |
| EPI_ISL_456790 | West of Scotland Specialist Virology Centre, NHSGGC / MRC-University of Glasgow Centre for Virus Research | COVID-19 Genomics UK (COG-UK) Consortium |
| EPI_ISL_456791 | West of Scotland Specialist Virology Centre, NHSGGC / MRC-University of Glasgow Centre for Virus Research | COVID-19 Genomics UK (COG-UK) Consortium |
| EPI_ISL_456792 | West of Scotland Specialist Virology Centre, NHSGGC / MRC-University of Glasgow Centre for Virus Research | COVID-19 Genomics UK (COG-UK) Consortium |
| EPI_ISL_456793 | West of Scotland Specialist Virology Centre, NHSGGC / MRC-University of Glasgow Centre for Virus Research | COVID-19 Genomics UK (COG-UK) Consortium |
| EPI_ISL_456794 | West of Scotland Specialist Virology Centre, NHSGGC / MRC-University of Glasgow Centre for Virus Research | COVID-19 Genomics UK (COG-UK) Consortium |
| EPI_ISL_456795 | West of Scotland Specialist Virology Centre, NHSGGC / MRC-University of Glasgow Centre for Virus Research | COVID-19 Genomics UK (COG-UK) Consortium |
| EPI_ISL_456796 | West of Scotland Specialist Virology Centre, NHSGGC / MRC-University of Glasgow Centre for Virus Research | COVID-19 Genomics UK (COG-UK) Consortium |
| EPI_ISL_456797 | West of Scotland Specialist Virology Centre, NHSGGC / MRC-University of Glasgow Centre for Virus Research | COVID-19 Genomics UK (COG-UK) Consortium |
| EPI_ISL_456798 | West of Scotland Specialist Virology Centre, NHSGGC / MRC-University of Glasgow Centre for Virus Research | COVID-19 Genomics UK (COG-UK) Consortium |
| EPI_ISL_456799 | West of Scotland Specialist Virology Centre, NHSGGC / MRC-University of Glasgow Centre for Virus Research | COVID-19 Genomics UK (COG-UK) Consortium |
| EPI_ISL_456800 | West of Scotland Specialist Virology Centre, NHSGGC / MRC-University of Glasgow Centre for Virus Research | COVID-19 Genomics UK (COG-UK) Consortium |
| EPI_ISL_456801 | West of Scotland Specialist Virology Centre, NHSGGC / MRC-University of Glasgow Centre for Virus Research | COVID-19 Genomics UK (COG-UK) Consortium |
| EPI_ISL_456802 | West of Scotland Specialist Virology Centre, NHSGGC / MRC-University of Glasgow Centre for Virus Research | COVID-19 Genomics UK (COG-UK) Consortium |
| EPI_ISL_456803 | West of Scotland Specialist Virology Centre, NHSGGC / MRC-University of Glasgow Centre for Virus Research | COVID-19 Genomics UK (COG-UK) Consortium |
| EPI_ISL_456804 | West of Scotland Specialist Virology Centre, NHSGGC / MRC-University of Glasgow Centre for Virus Research | COVID-19 Genomics UK (COG-UK) Consortium |

| EPI_Accession_ID | Originating Laboratory | Submitting Laboratory |
|---|---|---|
| EPI_ISL_456805 | West of Scotland Specialist Virology Centre, NHSGGC / MRC-University of Glasgow Centre for Virus Research | COVID-19 Genomics UK (COG-UK) Consortium |
| EPI_ISL_456806 | West of Scotland Specialist Virology Centre, NHSGGC / MRC-University of Glasgow Centre for Virus Research | COVID-19 Genomics UK (COG-UK) Consortium |
| EPI_ISL_456807 | West of Scotland Specialist Virology Centre, NHSGGC / MRC-University of Glasgow Centre for Virus Research | COVID-19 Genomics UK (COG-UK) Consortium |
| EPI_ISL_456808 | West of Scotland Specialist Virology Centre, NHSGGC / MRC-University of Glasgow Centre for Virus Research | COVID-19 Genomics UK (COG-UK) Consortium |
| EPI_ISL_456809 | West of Scotland Specialist Virology Centre, NHSGGC / MRC-University of Glasgow Centre for Virus Research | COVID-19 Genomics UK (COG-UK) Consortium |
| EPI_ISL_456810 | West of Scotland Specialist Virology Centre, NHSGGC / MRC-University of Glasgow Centre for Virus Research | COVID-19 Genomics UK (COG-UK) Consortium |
| EPI_ISL_456811 | West of Scotland Specialist Virology Centre, NHSGGC / MRC-University of Glasgow Centre for Virus Research | COVID-19 Genomics UK (COG-UK) Consortium |
| EPI_ISL_456812 | West of Scotland Specialist Virology Centre, NHSGGC / MRC-University of Glasgow Centre for Virus Research | COVID-19 Genomics UK (COG-UK) Consortium |
| EPI_ISL_456813 | West of Scotland Specialist Virology Centre, NHSGGC / MRC-University of Glasgow Centre for Virus Research | COVID-19 Genomics UK (COG-UK) Consortium |
| EPI_ISL_456814 | West of Scotland Specialist Virology Centre, NHSGGC / MRC-University of Glasgow Centre for Virus Research | COVID-19 Genomics UK (COG-UK) Consortium |
| EPI_ISL_456815 | West of Scotland Specialist Virology Centre, NHSGGC / MRC-University of Glasgow Centre for Virus Research | COVID-19 Genomics UK (COG-UK) Consortium |
| EPI_ISL_456816 | West of Scotland Specialist Virology Centre, NHSGGC / MRC-University of Glasgow Centre for Virus Research | COVID-19 Genomics UK (COG-UK) Consortium |
| EPI_ISL_456817 | West of Scotland Specialist Virology Centre, NHSGGC / MRC-University of Glasgow Centre for Virus Research | COVID-19 Genomics UK (COG-UK) Consortium |
| EPI_ISL_456818 | West of Scotland Specialist Virology Centre, NHSGGC / MRC-University of Glasgow Centre for Virus Research | COVID-19 Genomics UK (COG-UK) Consortium |
| EPI_ISL_456819 | West of Scotland Specialist Virology Centre, NHSGGC / MRC-University of Glasgow Centre for Virus Research | COVID-19 Genomics UK (COG-UK) Consortium |
| EPI_ISL_456820 | West of Scotland Specialist Virology Centre, NHSGGC / MRC-University of Glasgow Centre for Virus Research | COVID-19 Genomics UK (COG-UK) Consortium |

| EPI_Accession_ID | Originating Laboratory | Submitting Laboratory |
|---|---|---|
| EPI_ISL_456821 | West of Scotland Specialist Virology Centre, NHSGGC / MRC-University of Glasgow Centre for Virus Research | COVID-19 Genomics UK (COG-UK) Consortium |
| EPI_ISL_456822 | West of Scotland Specialist Virology Centre, NHSGGC / MRC-University of Glasgow Centre for Virus Research | COVID-19 Genomics UK (COG-UK) Consortium |
| EPI_ISL_456823 | West of Scotland Specialist Virology Centre, NHSGGC / MRC-University of Glasgow Centre for Virus Research | COVID-19 Genomics UK (COG-UK) Consortium |
| EPI_ISL_456824 | West of Scotland Specialist Virology Centre, NHSGGC / MRC-University of Glasgow Centre for Virus Research | COVID-19 Genomics UK (COG-UK) Consortium |
| EPI_ISL_456825 | West of Scotland Specialist Virology Centre, NHSGGC / MRC-University of Glasgow Centre for Virus Research | COVID-19 Genomics UK (COG-UK) Consortium |
| EPI_ISL_456826 | West of Scotland Specialist Virology Centre, NHSGGC / MRC-University of Glasgow Centre for Virus Research | COVID-19 Genomics UK (COG-UK) Consortium |
| EPI_ISL_456827 | West of Scotland Specialist Virology Centre, NHSGGC / MRC-University of Glasgow Centre for Virus Research | COVID-19 Genomics UK (COG-UK) Consortium |
| EPI_ISL_456828 | West of Scotland Specialist Virology Centre, NHSGGC / MRC-University of Glasgow Centre for Virus Research | COVID-19 Genomics UK (COG-UK) Consortium |
| EPI_ISL_456829 | West of Scotland Specialist Virology Centre, NHSGGC / MRC-University of Glasgow Centre for Virus Research | COVID-19 Genomics UK (COG-UK) Consortium |
| EPI_ISL_456830 | West of Scotland Specialist Virology Centre, NHSGGC / MRC-University of Glasgow Centre for Virus Research | COVID-19 Genomics UK (COG-UK) Consortium |
| EPI_ISL_456831 | West of Scotland Specialist Virology Centre, NHSGGC / MRC-University of Glasgow Centre for Virus Research | COVID-19 Genomics UK (COG-UK) Consortium |
| EPI_ISL_456832 | West of Scotland Specialist Virology Centre, NHSGGC / MRC-University of Glasgow Centre for Virus Research | COVID-19 Genomics UK (COG-UK) Consortium |
| EPI_ISL_456833 | West of Scotland Specialist Virology Centre, NHSGGC / MRC-University of Glasgow Centre for Virus Research | COVID-19 Genomics UK (COG-UK) Consortium |
| EPI_ISL_456834 | West of Scotland Specialist Virology Centre, NHSGGC / MRC-University of Glasgow Centre for Virus Research | COVID-19 Genomics UK (COG-UK) Consortium |
| EPI_ISL_456835 | West of Scotland Specialist Virology Centre, NHSGGC / MRC-University of Glasgow Centre for Virus Research | COVID-19 Genomics UK (COG-UK) Consortium |
| EPI_ISL_456836 | West of Scotland Specialist Virology Centre, NHSGGC / MRC-University of Glasgow Centre for Virus Research | COVID-19 Genomics UK (COG-UK) Consortium |

| EPI_Accession_ID | Originating Laboratory | Submitting Laboratory |
|---|---|---|
| EPI_ISL_456837 | West of Scotland Specialist Virology Centre, NHSGGC / MRC-University of Glasgow Centre for Virus Research | COVID-19 Genomics UK (COG-UK) Consortium |
| EPI_ISL_456838 | West of Scotland Specialist Virology Centre, NHSGGC / MRC-University of Glasgow Centre for Virus Research | COVID-19 Genomics UK (COG-UK) Consortium |
| EPI_ISL_456839 | West of Scotland Specialist Virology Centre, NHSGGC / MRC-University of Glasgow Centre for Virus Research | COVID-19 Genomics UK (COG-UK) Consortium |
| EPI_ISL_456840 | West of Scotland Specialist Virology Centre, NHSGGC / MRC-University of Glasgow Centre for Virus Research | COVID-19 Genomics UK (COG-UK) Consortium |
| EPI_ISL_456841 | West of Scotland Specialist Virology Centre, NHSGGC / MRC-University of Glasgow Centre for Virus Research | COVID-19 Genomics UK (COG-UK) Consortium |
| EPI_ISL_456842 | West of Scotland Specialist Virology Centre, NHSGGC / MRC-University of Glasgow Centre for Virus Research | COVID-19 Genomics UK (COG-UK) Consortium |
| EPI_ISL_456843 | West of Scotland Specialist Virology Centre, NHSGGC / MRC-University of Glasgow Centre for Virus Research | COVID-19 Genomics UK (COG-UK) Consortium |
| EPI_ISL_456844 | West of Scotland Specialist Virology Centre, NHSGGC / MRC-University of Glasgow Centre for Virus Research | COVID-19 Genomics UK (COG-UK) Consortium |
| EPI_ISL_456845 | West of Scotland Specialist Virology Centre, NHSGGC / MRC-University of Glasgow Centre for Virus Research | COVID-19 Genomics UK (COG-UK) Consortium |
| EPI_ISL_456846 | West of Scotland Specialist Virology Centre, NHSGGC / MRC-University of Glasgow Centre for Virus Research | COVID-19 Genomics UK (COG-UK) Consortium |
| EPI_ISL_456847 | West of Scotland Specialist Virology Centre, NHSGGC / MRC-University of Glasgow Centre for Virus Research | COVID-19 Genomics UK (COG-UK) Consortium |
| EPI_ISL_456848 | West of Scotland Specialist Virology Centre, NHSGGC / MRC-University of Glasgow Centre for Virus Research | COVID-19 Genomics UK (COG-UK) Consortium |
| EPI_ISL_456849 | West of Scotland Specialist Virology Centre, NHSGGC / MRC-University of Glasgow Centre for Virus Research | COVID-19 Genomics UK (COG-UK) Consortium |
| EPI_ISL_456850 | West of Scotland Specialist Virology Centre, NHSGGC / MRC-University of Glasgow Centre for Virus Research | COVID-19 Genomics UK (COG-UK) Consortium |
| EPI_ISL_456851 | West of Scotland Specialist Virology Centre, NHSGGC / MRC-University of Glasgow Centre for Virus Research | COVID-19 Genomics UK (COG-UK) Consortium |
| EPI_ISL_456852 | West of Scotland Specialist Virology Centre, NHSGGC / MRC-University of Glasgow Centre for Virus Research | COVID-19 Genomics UK (COG-UK) Consortium |

| EPI_Accession_ID | Originating Laboratory | Submitting Laboratory |
|---|---|---|
| EPI_ISL_456853 | West of Scotland Specialist Virology Centre, NHSGGC / MRC-University of Glasgow Centre for Virus Research | COVID-19 Genomics UK (COG-UK) Consortium |
| EPI_ISL_456854 | West of Scotland Specialist Virology Centre, NHSGGC / MRC-University of Glasgow Centre for Virus Research | COVID-19 Genomics UK (COG-UK) Consortium |
| EPI_ISL_456855 | West of Scotland Specialist Virology Centre, NHSGGC / MRC-University of Glasgow Centre for Virus Research | COVID-19 Genomics UK (COG-UK) Consortium |
| EPI_ISL_456856 | West of Scotland Specialist Virology Centre, NHSGGC / MRC-University of Glasgow Centre for Virus Research | COVID-19 Genomics UK (COG-UK) Consortium |
| EPI_ISL_456857 | West of Scotland Specialist Virology Centre, NHSGGC / MRC-University of Glasgow Centre for Virus Research | COVID-19 Genomics UK (COG-UK) Consortium |
| EPI_ISL_456858 | West of Scotland Specialist Virology Centre, NHSGGC / MRC-University of Glasgow Centre for Virus Research | COVID-19 Genomics UK (COG-UK) Consortium |
| EPI_ISL_456859 | West of Scotland Specialist Virology Centre, NHSGGC / MRC-University of Glasgow Centre for Virus Research | COVID-19 Genomics UK (COG-UK) Consortium |
| EPI_ISL_456860 | West of Scotland Specialist Virology Centre, NHSGGC / MRC-University of Glasgow Centre for Virus Research | COVID-19 Genomics UK (COG-UK) Consortium |
| EPI_ISL_456861 | West of Scotland Specialist Virology Centre, NHSGGC / MRC-University of Glasgow Centre for Virus Research | COVID-19 Genomics UK (COG-UK) Consortium |
| EPI_ISL_456862 | West of Scotland Specialist Virology Centre, NHSGGC / MRC-University of Glasgow Centre for Virus Research | COVID-19 Genomics UK (COG-UK) Consortium |
| EPI_ISL_456863 | West of Scotland Specialist Virology Centre, NHSGGC / MRC-University of Glasgow Centre for Virus Research | COVID-19 Genomics UK (COG-UK) Consortium |
| EPI_ISL_456864 | West of Scotland Specialist Virology Centre, NHSGGC / MRC-University of Glasgow Centre for Virus Research | COVID-19 Genomics UK (COG-UK) Consortium |
| EPI_ISL_456865 | West of Scotland Specialist Virology Centre, NHSGGC / MRC-University of Glasgow Centre for Virus Research | COVID-19 Genomics UK (COG-UK) Consortium |
| EPI_ISL_456866 | West of Scotland Specialist Virology Centre, NHSGGC / MRC-University of Glasgow Centre for Virus Research | COVID-19 Genomics UK (COG-UK) Consortium |
| EPI_ISL_456867 | West of Scotland Specialist Virology Centre, NHSGGC / MRC-University of Glasgow Centre for Virus Research | COVID-19 Genomics UK (COG-UK) Consortium |
| EPI_ISL_456868 | West of Scotland Specialist Virology Centre, NHSGGC / MRC-University of Glasgow Centre for Virus Research | COVID-19 Genomics UK (COG-UK) Consortium |

| EPI_Accession_ID | Originating Laboratory | Submitting Laboratory |
|---|---|---|
| EPI_ISL_456869 | West of Scotland Specialist Virology Centre, NHSGGC / MRC-University of Glasgow Centre for Virus Research | COVID-19 Genomics UK (COG-UK) Consortium |
| EPI_ISL_456870 | West of Scotland Specialist Virology Centre, NHSGGC / MRC-University of Glasgow Centre for Virus Research | COVID-19 Genomics UK (COG-UK) Consortium |
| EPI_ISL_456871 | West of Scotland Specialist Virology Centre, NHSGGC / MRC-University of Glasgow Centre for Virus Research | COVID-19 Genomics UK (COG-UK) Consortium |
| EPI_ISL_456872 | West of Scotland Specialist Virology Centre, NHSGGC / MRC-University of Glasgow Centre for Virus Research | COVID-19 Genomics UK (COG-UK) Consortium |
| EPI_ISL_456873 | West of Scotland Specialist Virology Centre, NHSGGC / MRC-University of Glasgow Centre for Virus Research | COVID-19 Genomics UK (COG-UK) Consortium |
| EPI_ISL_456874 | West of Scotland Specialist Virology Centre, NHSGGC / MRC-University of Glasgow Centre for Virus Research | COVID-19 Genomics UK (COG-UK) Consortium |
| EPI_ISL_456875 | West of Scotland Specialist Virology Centre, NHSGGC / MRC-University of Glasgow Centre for Virus Research | COVID-19 Genomics UK (COG-UK) Consortium |
| EPI_ISL_456876 | West of Scotland Specialist Virology Centre, NHSGGC / MRC-University of Glasgow Centre for Virus Research | COVID-19 Genomics UK (COG-UK) Consortium |
| EPI_ISL_456877 | West of Scotland Specialist Virology Centre, NHSGGC / MRC-University of Glasgow Centre for Virus Research | COVID-19 Genomics UK (COG-UK) Consortium |
| EPI_ISL_456878 | West of Scotland Specialist Virology Centre, NHSGGC / MRC-University of Glasgow Centre for Virus Research | COVID-19 Genomics UK (COG-UK) Consortium |
| EPI_ISL_456879 | West of Scotland Specialist Virology Centre, NHSGGC / MRC-University of Glasgow Centre for Virus Research | COVID-19 Genomics UK (COG-UK) Consortium |
| EPI_ISL_456880 | West of Scotland Specialist Virology Centre, NHSGGC / MRC-University of Glasgow Centre for Virus Research | COVID-19 Genomics UK (COG-UK) Consortium |
| EPI_ISL_456881 | West of Scotland Specialist Virology Centre, NHSGGC / MRC-University of Glasgow Centre for Virus Research | COVID-19 Genomics UK (COG-UK) Consortium |
| EPI_ISL_456882 | West of Scotland Specialist Virology Centre, NHSGGC / MRC-University of Glasgow Centre for Virus Research | COVID-19 Genomics UK (COG-UK) Consortium |
| EPI_ISL_456883 | West of Scotland Specialist Virology Centre, NHSGGC / MRC-University of Glasgow Centre for Virus Research | COVID-19 Genomics UK (COG-UK) Consortium |
| EPI_ISL_456884 | West of Scotland Specialist Virology Centre, NHSGGC / MRC-University of Glasgow Centre for Virus Research | COVID-19 Genomics UK (COG-UK) Consortium |

| EPI_Accession_ID | Originating Laboratory | Submitting Laboratory |
|---|---|---|
| EPI_ISL_456885 | West of Scotland Specialist Virology Centre, NHSGGC / MRC-University of Glasgow Centre for Virus Research | COVID-19 Genomics UK (COG-UK) Consortium |
| EPI_ISL_456886 | West of Scotland Specialist Virology Centre, NHSGGC / MRC-University of Glasgow Centre for Virus Research | COVID-19 Genomics UK (COG-UK) Consortium |
| EPI_ISL_456887 | West of Scotland Specialist Virology Centre, NHSGGC / MRC-University of Glasgow Centre for Virus Research | COVID-19 Genomics UK (COG-UK) Consortium |
| EPI_ISL_456888 | West of Scotland Specialist Virology Centre, NHSGGC / MRC-University of Glasgow Centre for Virus Research | COVID-19 Genomics UK (COG-UK) Consortium |
| EPI_ISL_456889 | West of Scotland Specialist Virology Centre, NHSGGC / MRC-University of Glasgow Centre for Virus Research | COVID-19 Genomics UK (COG-UK) Consortium |
| EPI_ISL_456890 | Virology Department, Royal Infirmary of Edinburgh, NHS Lothian / School of Biological Sciences, University of Edinburgh / Institute of Genetics and Molecular Medicine, University of Edinburgh | COVID-19 Genomics UK (COG-UK) Consortium |
| EPI_ISL_456891 | Virology Department, Royal Infirmary of Edinburgh, NHS Lothian / School of Biological Sciences, University of Edinburgh / Institute of Genetics and Molecular Medicine, University of Edinburgh | COVID-19 Genomics UK (COG-UK) Consortium |
| EPI_ISL_456892 | Virology Department, Royal Infirmary of Edinburgh, NHS Lothian / School of Biological Sciences, University of Edinburgh / Institute of Genetics and Molecular Medicine, University of Edinburgh | COVID-19 Genomics UK (COG-UK) Consortium |
| EPI_ISL_456893 | Virology Department, Royal Infirmary of Edinburgh, NHS Lothian / School of Biological Sciences, University of Edinburgh / Institute of Genetics and Molecular Medicine, University of Edinburgh | COVID-19 Genomics UK (COG-UK) Consortium |
| EPI_ISL_456894 | Virology Department, Royal Infirmary of Edinburgh, NHS Lothian / School of Biological Sciences, University of Edinburgh / Institute of Genetics and Molecular Medicine, University of Edinburgh | COVID-19 Genomics UK (COG-UK) Consortium |

| EPI_Accession_ID | Originating Laboratory | Submitting Laboratory |
|---|---|---|
| EPI_ISL_456895 | Virology Department, Royal Infirmary of Edinburgh, NHS Lothian / School of Biological Sciences, University of Edinburgh / Institute of Genetics and Molecular Medicine, University of Edinburgh | COVID-19 Genomics UK (COG-UK) Consortium |
| EPI_ISL_456896 | Virology Department, Royal Infirmary of Edinburgh, NHS Lothian / School of Biological Sciences, University of Edinburgh / Institute of Genetics and Molecular Medicine, University of Edinburgh | COVID-19 Genomics UK (COG-UK) Consortium |
| EPI_ISL_456897 | Virology Department, Royal Infirmary of Edinburgh, NHS Lothian / School of Biological Sciences, University of Edinburgh / Institute of Genetics and Molecular Medicine, University of Edinburgh | COVID-19 Genomics UK (COG-UK) Consortium |
| EPI_ISL_456898 | Virology Department, Royal Infirmary of Edinburgh, NHS Lothian / School of Biological Sciences, University of Edinburgh / Institute of Genetics and Molecular Medicine, University of Edinburgh | COVID-19 Genomics UK (COG-UK) Consortium |
| EPI_ISL_456899 | Virology Department, Royal Infirmary of Edinburgh, NHS Lothian / School of Biological Sciences, University of Edinburgh / Institute of Genetics and Molecular Medicine, University of Edinburgh | COVID-19 Genomics UK (COG-UK) Consortium |
| EPI_ISL_456900 | Virology Department, Royal Infirmary of Edinburgh, NHS Lothian / School of Biological Sciences, University of Edinburgh / Institute of Genetics and Molecular Medicine, University of Edinburgh | COVID-19 Genomics UK (COG-UK) Consortium |
| EPI_ISL_456901 | Virology Department, Royal Infirmary of Edinburgh, NHS Lothian / School of Biological Sciences, University of Edinburgh / Institute of Genetics and Molecular Medicine, University of Edinburgh | COVID-19 Genomics UK (COG-UK) Consortium |
| EPI_ISL_456902 | Virology Department, Royal Infirmary of Edinburgh, NHS Lothian / School of Biological Sciences, University of Edinburgh / Institute of Genetics and Molecular Medicine, University of Edinburgh | COVID-19 Genomics UK (COG-UK) Consortium |

| EPI_Accession_ID | Originating Laboratory | Submitting Laboratory |
|---|---|---|
| EPI_ISL_456903 | Virology Department, Royal Infirmary of Edinburgh, NHS Lothian / School of Biological Sciences, University of Edinburgh / Institute of Genetics and Molecular Medicine, University of Edinburgh | COVID-19 Genomics UK (COG-UK) Consortium |
| EPI_ISL_456904 | Virology Department, Royal Infirmary of Edinburgh, NHS Lothian / School of Biological Sciences, University of Edinburgh / Institute of Genetics and Molecular Medicine, University of Edinburgh | COVID-19 Genomics UK (COG-UK) Consortium |
| EPI_ISL_456905 | Virology Department, Royal Infirmary of Edinburgh, NHS Lothian / School of Biological Sciences, University of Edinburgh / Institute of Genetics and Molecular Medicine, University of Edinburgh | COVID-19 Genomics UK (COG-UK) Consortium |
| EPI_ISL_456906 | Virology Department, Royal Infirmary of Edinburgh, NHS Lothian / School of Biological Sciences, University of Edinburgh / Institute of Genetics and Molecular Medicine, University of Edinburgh | COVID-19 Genomics UK (COG-UK) Consortium |
| EPI_ISL_456907 | Virology Department, Royal Infirmary of Edinburgh, NHS Lothian / School of Biological Sciences, University of Edinburgh / Institute of Genetics and Molecular Medicine, University of Edinburgh | COVID-19 Genomics UK (COG-UK) Consortium |
| EPI_ISL_456908 | Virology Department, Royal Infirmary of Edinburgh, NHS Lothian / School of Biological Sciences, University of Edinburgh / Institute of Genetics and Molecular Medicine, University of Edinburgh | COVID-19 Genomics UK (COG-UK) Consortium |
| EPI_ISL_456909 | Virology Department, Royal Infirmary of Edinburgh, NHS Lothian / School of Biological Sciences, University of Edinburgh / Institute of Genetics and Molecular Medicine, University of Edinburgh | COVID-19 Genomics UK (COG-UK) Consortium |
| EPI_ISL_456910 | Virology Department, Royal Infirmary of Edinburgh, NHS Lothian / School of Biological Sciences, University of Edinburgh / Institute of Genetics and Molecular Medicine, University of Edinburgh | COVID-19 Genomics UK (COG-UK) Consortium |

| EPI_Accession_ID | Originating Laboratory | Submitting Laboratory |
|---|---|---|
| EPI_ISL_456911 | Virology Department, Royal Infirmary of Edinburgh, NHS Lothian / School of Biological Sciences, University of Edinburgh / Institute of Genetics and Molecular Medicine, University of Edinburgh | COVID-19 Genomics UK (COG-UK) Consortium |
| EPI_ISL_456912 | Virology Department, Royal Infirmary of Edinburgh, NHS Lothian / School of Biological Sciences, University of Edinburgh / Institute of Genetics and Molecular Medicine, University of Edinburgh | COVID-19 Genomics UK (COG-UK) Consortium |
| EPI_ISL_456913 | Virology Department, Royal Infirmary of Edinburgh, NHS Lothian / School of Biological Sciences, University of Edinburgh / Institute of Genetics and Molecular Medicine, University of Edinburgh | COVID-19 Genomics UK (COG-UK) Consortium |
| EPI_ISL_456914 | Virology Department, Royal Infirmary of Edinburgh, NHS Lothian / School of Biological Sciences, University of Edinburgh / Institute of Genetics and Molecular Medicine, University of Edinburgh | COVID-19 Genomics UK (COG-UK) Consortium |
| EPI_ISL_456915 | Virology Department, Royal Infirmary of Edinburgh, NHS Lothian / School of Biological Sciences, University of Edinburgh / Institute of Genetics and Molecular Medicine, University of Edinburgh | COVID-19 Genomics UK (COG-UK) Consortium |
| EPI_ISL_456916 | Virology Department, Royal Infirmary of Edinburgh, NHS Lothian / School of Biological Sciences, University of Edinburgh / Institute of Genetics and Molecular Medicine, University of Edinburgh | COVID-19 Genomics UK (COG-UK) Consortium |
| EPI_ISL_456917 | Virology Department, Royal Infirmary of Edinburgh, NHS Lothian / School of Biological Sciences, University of Edinburgh / Institute of Genetics and Molecular Medicine, University of Edinburgh | COVID-19 Genomics UK (COG-UK) Consortium |
| EPI_ISL_456918 | Virology Department, Royal Infirmary of Edinburgh, NHS Lothian / School of Biological Sciences, University of Edinburgh / Institute of Genetics and Molecular Medicine, University of Edinburgh | COVID-19 Genomics UK (COG-UK) Consortium |

| EPI_Accession_ID | Originating Laboratory | Submitting Laboratory |
|---|---|---|
| EPI_ISL_456919 | Virology Department, Royal Infirmary of Edinburgh, NHS Lothian / School of Biological Sciences, University of Edinburgh / Institute of Genetics and Molecular Medicine, University of Edinburgh | COVID-19 Genomics UK (COG-UK) Consortium |
| EPI_ISL_456920 | Virology Department, Royal Infirmary of Edinburgh, NHS Lothian / School of Biological Sciences, University of Edinburgh / Institute of Genetics and Molecular Medicine, University of Edinburgh | COVID-19 Genomics UK (COG-UK) Consortium |
| EPI_ISL_456921 | Virology Department, Royal Infirmary of Edinburgh, NHS Lothian / School of Biological Sciences, University of Edinburgh / Institute of Genetics and Molecular Medicine, University of Edinburgh | COVID-19 Genomics UK (COG-UK) Consortium |
| EPI_ISL_456922 | Virology Department, Royal Infirmary of Edinburgh, NHS Lothian / School of Biological Sciences, University of Edinburgh / Institute of Genetics and Molecular Medicine, University of Edinburgh | COVID-19 Genomics UK (COG-UK) Consortium |
| EPI_ISL_456923 | Virology Department, Royal Infirmary of Edinburgh, NHS Lothian / School of Biological Sciences, University of Edinburgh / Institute of Genetics and Molecular Medicine, University of Edinburgh | COVID-19 Genomics UK (COG-UK) Consortium |
| EPI_ISL_456924 | Virology Department, Royal Infirmary of Edinburgh, NHS Lothian / School of Biological Sciences, University of Edinburgh / Institute of Genetics and Molecular Medicine, University of Edinburgh | COVID-19 Genomics UK (COG-UK) Consortium |
| EPI_ISL_456925 | Virology Department, Royal Infirmary of Edinburgh, NHS Lothian / School of Biological Sciences, University of Edinburgh / Institute of Genetics and Molecular Medicine, University of Edinburgh | COVID-19 Genomics UK (COG-UK) Consortium |
| EPI_ISL_456926 | Virology Department, Royal Infirmary of Edinburgh, NHS Lothian / School of Biological Sciences, University of Edinburgh / Institute of Genetics and Molecular Medicine, University of Edinburgh | COVID-19 Genomics UK (COG-UK) Consortium |

| EPI_Accession_ID | Originating Laboratory | Submitting Laboratory |
|---|---|---|
| EPI_ISL_456927 | Virology Department, Royal Infirmary of Edinburgh, NHS Lothian / School of Biological Sciences, University of Edinburgh / Institute of Genetics and Molecular Medicine, University of Edinburgh | COVID-19 Genomics UK (COG-UK) Consortium |
| EPI_ISL_456928 | Virology Department, Royal Infirmary of Edinburgh, NHS Lothian / School of Biological Sciences, University of Edinburgh / Institute of Genetics and Molecular Medicine, University of Edinburgh | COVID-19 Genomics UK (COG-UK) Consortium |
| EPI_ISL_456929 | Virology Department, Royal Infirmary of Edinburgh, NHS Lothian / School of Biological Sciences, University of Edinburgh / Institute of Genetics and Molecular Medicine, University of Edinburgh | COVID-19 Genomics UK (COG-UK) Consortium |
| EPI_ISL_456930 | Virology Department, Royal Infirmary of Edinburgh, NHS Lothian / School of Biological Sciences, University of Edinburgh / Institute of Genetics and Molecular Medicine, University of Edinburgh | COVID-19 Genomics UK (COG-UK) Consortium |
| EPI_ISL_456931 | Virology Department, Royal Infirmary of Edinburgh, NHS Lothian / School of Biological Sciences, University of Edinburgh / Institute of Genetics and Molecular Medicine, University of Edinburgh | COVID-19 Genomics UK (COG-UK) Consortium |
| EPI_ISL_456932 | Virology Department, Royal Infirmary of Edinburgh, NHS Lothian / School of Biological Sciences, University of Edinburgh / Institute of Genetics and Molecular Medicine, University of Edinburgh | COVID-19 Genomics UK (COG-UK) Consortium |
| EPI_ISL_456933 | Virology Department, Royal Infirmary of Edinburgh, NHS Lothian / School of Biological Sciences, University of Edinburgh / Institute of Genetics and Molecular Medicine, University of Edinburgh | COVID-19 Genomics UK (COG-UK) Consortium |
| EPI_ISL_456934 | Virology Department, Royal Infirmary of Edinburgh, NHS Lothian / School of Biological Sciences, University of Edinburgh / Institute of Genetics and Molecular Medicine, University of Edinburgh | COVID-19 Genomics UK (COG-UK) Consortium |

| EPI_Accession_ID | Originating Laboratory | Submitting Laboratory |
|---|---|---|
| EPI_ISL_456935 | Virology Department, Royal Infirmary of Edinburgh, NHS Lothian / School of Biological Sciences, University of Edinburgh / Institute of Genetics and Molecular Medicine, University of Edinburgh | COVID-19 Genomics UK (COG-UK) Consortium |
| EPI_ISL_456936 | Virology Department, Royal Infirmary of Edinburgh, NHS Lothian / School of Biological Sciences, University of Edinburgh / Institute of Genetics and Molecular Medicine, University of Edinburgh | COVID-19 Genomics UK (COG-UK) Consortium |
| EPI_ISL_456937 | Virology Department, Royal Infirmary of Edinburgh, NHS Lothian / School of Biological Sciences, University of Edinburgh / Institute of Genetics and Molecular Medicine, University of Edinburgh | COVID-19 Genomics UK (COG-UK) Consortium |
| EPI_ISL_456938 | Virology Department, Royal Infirmary of Edinburgh, NHS Lothian / School of Biological Sciences, University of Edinburgh / Institute of Genetics and Molecular Medicine, University of Edinburgh | COVID-19 Genomics UK (COG-UK) Consortium |
| EPI_ISL_456939 | Virology Department, Royal Infirmary of Edinburgh, NHS Lothian / School of Biological Sciences, University of Edinburgh / Institute of Genetics and Molecular Medicine, University of Edinburgh | COVID-19 Genomics UK (COG-UK) Consortium |
| EPI_ISL_456940 | Virology Department, Royal Infirmary of Edinburgh, NHS Lothian / School of Biological Sciences, University of Edinburgh / Institute of Genetics and Molecular Medicine, University of Edinburgh | COVID-19 Genomics UK (COG-UK) Consortium |
| EPI_ISL_456941 | Virology Department, Royal Infirmary of Edinburgh, NHS Lothian / School of Biological Sciences, University of Edinburgh / Institute of Genetics and Molecular Medicine, University of Edinburgh | COVID-19 Genomics UK (COG-UK) Consortium |
| EPI_ISL_456942 | Virology Department, Royal Infirmary of Edinburgh, NHS Lothian / School of Biological Sciences, University of Edinburgh / Institute of Genetics and Molecular Medicine, University of Edinburgh | COVID-19 Genomics UK (COG-UK) Consortium |

| EPI_Accession_ID | Originating Laboratory | Submitting Laboratory |
|---|---|---|
| EPI_ISL_456943 | Virology Department, Royal Infirmary of Edinburgh, NHS Lothian / School of Biological Sciences, University of Edinburgh / Institute of Genetics and Molecular Medicine, University of Edinburgh | COVID-19 Genomics UK (COG-UK) Consortium |
| EPI_ISL_456944 | Virology Department, Royal Infirmary of Edinburgh, NHS Lothian / School of Biological Sciences, University of Edinburgh / Institute of Genetics and Molecular Medicine, University of Edinburgh | COVID-19 Genomics UK (COG-UK) Consortium |
| EPI_ISL_456945 | Virology Department, Royal Infirmary of Edinburgh, NHS Lothian / School of Biological Sciences, University of Edinburgh / Institute of Genetics and Molecular Medicine, University of Edinburgh | COVID-19 Genomics UK (COG-UK) Consortium |
| EPI_ISL_456946 | Virology Department, Royal Infirmary of Edinburgh, NHS Lothian / School of Biological Sciences, University of Edinburgh / Institute of Genetics and Molecular Medicine, University of Edinburgh | COVID-19 Genomics UK (COG-UK) Consortium |
| EPI_ISL_456947 | Virology Department, Royal Infirmary of Edinburgh, NHS Lothian / School of Biological Sciences, University of Edinburgh / Institute of Genetics and Molecular Medicine, University of Edinburgh | COVID-19 Genomics UK (COG-UK) Consortium |
| EPI_ISL_456948 | Virology Department, Royal Infirmary of Edinburgh, NHS Lothian / School of Biological Sciences, University of Edinburgh / Institute of Genetics and Molecular Medicine, University of Edinburgh | COVID-19 Genomics UK (COG-UK) Consortium |
| EPI_ISL_456949 | Virology Department, Royal Infirmary of Edinburgh, NHS Lothian / School of Biological Sciences, University of Edinburgh / Institute of Genetics and Molecular Medicine, University of Edinburgh | COVID-19 Genomics UK (COG-UK) Consortium |
| EPI_ISL_456950 | Virology Department, Royal Infirmary of Edinburgh, NHS Lothian / School of Biological Sciences, University of Edinburgh / Institute of Genetics and Molecular Medicine, University of Edinburgh | COVID-19 Genomics UK (COG-UK) Consortium |

| EPI_Accession_ID | Originating Laboratory | Submitting Laboratory |
|---|---|---|
| EPI_ISL_456951 | Virology Department, Royal Infirmary of Edinburgh, NHS Lothian / School of Biological Sciences, University of Edinburgh / Institute of Genetics and Molecular Medicine, University of Edinburgh | COVID-19 Genomics UK (COG-UK) Consortium |
| EPI_ISL_456952 | Virology Department, Royal Infirmary of Edinburgh, NHS Lothian / School of Biological Sciences, University of Edinburgh / Institute of Genetics and Molecular Medicine, University of Edinburgh | COVID-19 Genomics UK (COG-UK) Consortium |
| EPI_ISL_456953 | Virology Department, Royal Infirmary of Edinburgh, NHS Lothian / School of Biological Sciences, University of Edinburgh / Institute of Genetics and Molecular Medicine, University of Edinburgh | COVID-19 Genomics UK (COG-UK) Consortium |
| EPI_ISL_456954 | Virology Department, Royal Infirmary of Edinburgh, NHS Lothian / School of Biological Sciences, University of Edinburgh / Institute of Genetics and Molecular Medicine, University of Edinburgh | COVID-19 Genomics UK (COG-UK) Consortium |
| EPI_ISL_456955 | Virology Department, Royal Infirmary of Edinburgh, NHS Lothian / School of Biological Sciences, University of Edinburgh / Institute of Genetics and Molecular Medicine, University of Edinburgh | COVID-19 Genomics UK (COG-UK) Consortium |
| EPI_ISL_456956 | Virology Department, Royal Infirmary of Edinburgh, NHS Lothian / School of Biological Sciences, University of Edinburgh / Institute of Genetics and Molecular Medicine, University of Edinburgh | COVID-19 Genomics UK (COG-UK) Consortium |
| EPI_ISL_456957 | Virology Department, Royal Infirmary of Edinburgh, NHS Lothian / School of Biological Sciences, University of Edinburgh / Institute of Genetics and Molecular Medicine, University of Edinburgh | COVID-19 Genomics UK (COG-UK) Consortium |
| EPI_ISL_456958 | Virology Department, Royal Infirmary of Edinburgh, NHS Lothian / School of Biological Sciences, University of Edinburgh / Institute of Genetics and Molecular Medicine, University of Edinburgh | COVID-19 Genomics UK (COG-UK) Consortium |

| EPI_Accession_ID | Originating Laboratory | Submitting Laboratory |
|---|---|---|
| EPI_ISL_456959 | Virology Department, Royal Infirmary of Edinburgh, NHS Lothian / School of Biological Sciences, University of Edinburgh / Institute of Genetics and Molecular Medicine, University of Edinburgh | COVID-19 Genomics UK (COG-UK) Consortium |
| EPI_ISL_456960 | Virology Department, Royal Infirmary of Edinburgh, NHS Lothian / School of Biological Sciences, University of Edinburgh / Institute of Genetics and Molecular Medicine, University of Edinburgh | COVID-19 Genomics UK (COG-UK) Consortium |
| EPI_ISL_456961 | Virology Department, Royal Infirmary of Edinburgh, NHS Lothian / School of Biological Sciences, University of Edinburgh / Institute of Genetics and Molecular Medicine, University of Edinburgh | COVID-19 Genomics UK (COG-UK) Consortium |
| EPI_ISL_456962 | Virology Department, Royal Infirmary of Edinburgh, NHS Lothian / School of Biological Sciences, University of Edinburgh / Institute of Genetics and Molecular Medicine, University of Edinburgh | COVID-19 Genomics UK (COG-UK) Consortium |
| EPI_ISL_456963 | Virology Department, Royal Infirmary of Edinburgh, NHS Lothian / School of Biological Sciences, University of Edinburgh / Institute of Genetics and Molecular Medicine, University of Edinburgh | COVID-19 Genomics UK (COG-UK) Consortium |
| EPI_ISL_456964 | Virology Department, Royal Infirmary of Edinburgh, NHS Lothian / School of Biological Sciences, University of Edinburgh / Institute of Genetics and Molecular Medicine, University of Edinburgh | COVID-19 Genomics UK (COG-UK) Consortium |
| EPI_ISL_456965 | Virology Department, Royal Infirmary of Edinburgh, NHS Lothian / School of Biological Sciences, University of Edinburgh / Institute of Genetics and Molecular Medicine, University of Edinburgh | COVID-19 Genomics UK (COG-UK) Consortium |
| EPI_ISL_456966 | Virology Department, Royal Infirmary of Edinburgh, NHS Lothian / School of Biological Sciences, University of Edinburgh / Institute of Genetics and Molecular Medicine, University of Edinburgh | COVID-19 Genomics UK (COG-UK) Consortium |

| EPI_Accession_ID | Originating Laboratory | Submitting Laboratory |
|---|---|---|
| EPI_ISL_456967 | Virology Department, Royal Infirmary of Edinburgh, NHS Lothian / School of Biological Sciences, University of Edinburgh / Institute of Genetics and Molecular Medicine, University of Edinburgh | COVID-19 Genomics UK (COG-UK) Consortium |
| EPI_ISL_456968 | Virology Department, Royal Infirmary of Edinburgh, NHS Lothian / School of Biological Sciences, University of Edinburgh / Institute of Genetics and Molecular Medicine, University of Edinburgh | COVID-19 Genomics UK (COG-UK) Consortium |
| EPI_ISL_456969 | Virology Department, Royal Infirmary of Edinburgh, NHS Lothian / School of Biological Sciences, University of Edinburgh / Institute of Genetics and Molecular Medicine, University of Edinburgh | COVID-19 Genomics UK (COG-UK) Consortium |
| EPI_ISL_456970 | Virology Department, Royal Infirmary of Edinburgh, NHS Lothian / School of Biological Sciences, University of Edinburgh / Institute of Genetics and Molecular Medicine, University of Edinburgh | COVID-19 Genomics UK (COG-UK) Consortium |
| EPI_ISL_456971 | Virology Department, Royal Infirmary of Edinburgh, NHS Lothian / School of Biological Sciences, University of Edinburgh / Institute of Genetics and Molecular Medicine, University of Edinburgh | COVID-19 Genomics UK (COG-UK) Consortium |
| EPI_ISL_456972 | Virology Department, Royal Infirmary of Edinburgh, NHS Lothian / School of Biological Sciences, University of Edinburgh / Institute of Genetics and Molecular Medicine, University of Edinburgh | COVID-19 Genomics UK (COG-UK) Consortium |
| EPI_ISL_456973 | Virology Department, Royal Infirmary of Edinburgh, NHS Lothian / School of Biological Sciences, University of Edinburgh / Institute of Genetics and Molecular Medicine, University of Edinburgh | COVID-19 Genomics UK (COG-UK) Consortium |
| EPI_ISL_456974 | Virology Department, Royal Infirmary of Edinburgh, NHS Lothian / School of Biological Sciences, University of Edinburgh / Institute of Genetics and Molecular Medicine, University of Edinburgh | COVID-19 Genomics UK (COG-UK) Consortium |

| EPI_Accession_ID | Originating Laboratory | Submitting Laboratory |
| --- | --- | --- |
| EPI_ISL_456975 | Virology Department, Royal Infirmary of Edinburgh, NHS Lothian / School of Biological Sciences, University of Edinburgh / Institute of Genetics and Molecular Medicine, University of Edinburgh | COVID-19 Genomics UK (COG-UK) Consortium |
| EPI_ISL_456976 | Virology Department, Royal Infirmary of Edinburgh, NHS Lothian / School of Biological Sciences, University of Edinburgh / Institute of Genetics and Molecular Medicine, University of Edinburgh | COVID-19 Genomics UK (COG-UK) Consortium |
| EPI_ISL_456977 | Virology Department, Royal Infirmary of Edinburgh, NHS Lothian / School of Biological Sciences, University of Edinburgh / Institute of Genetics and Molecular Medicine, University of Edinburgh | COVID-19 Genomics UK (COG-UK) Consortium |
| EPI_ISL_456978 | Virology Department, Royal Infirmary of Edinburgh, NHS Lothian / School of Biological Sciences, University of Edinburgh / Institute of Genetics and Molecular Medicine, University of Edinburgh | COVID-19 Genomics UK (COG-UK) Consortium |
| EPI_ISL_456979 | Virology Department, Royal Infirmary of Edinburgh, NHS Lothian / School of Biological Sciences, University of Edinburgh / Institute of Genetics and Molecular Medicine, University of Edinburgh | COVID-19 Genomics UK (COG-UK) Consortium |
| EPI_ISL_456980 | Virology Department, Royal Infirmary of Edinburgh, NHS Lothian / School of Biological Sciences, University of Edinburgh / Institute of Genetics and Molecular Medicine, University of Edinburgh | COVID-19 Genomics UK (COG-UK) Consortium |
| EPI_ISL_456981 | Virology Department, Royal Infirmary of Edinburgh, NHS Lothian / School of Biological Sciences, University of Edinburgh / Institute of Genetics and Molecular Medicine, University of Edinburgh | COVID-19 Genomics UK (COG-UK) Consortium |
| EPI_ISL_456982 | Virology Department, Royal Infirmary of Edinburgh, NHS Lothian / School of Biological Sciences, University of Edinburgh / Institute of Genetics and Molecular Medicine, University of Edinburgh | COVID-19 Genomics UK (COG-UK) Consortium |

| EPI_Accession_ID | Originating Laboratory | Submitting Laboratory |
|---|---|---|
| EPI_ISL_456983 | Virology Department, Royal Infirmary of Edinburgh, NHS Lothian / School of Biological Sciences, University of Edinburgh / Institute of Genetics and Molecular Medicine, University of Edinburgh | COVID-19 Genomics UK (COG-UK) Consortium |
| EPI_ISL_456984 | Virology Department, Royal Infirmary of Edinburgh, NHS Lothian / School of Biological Sciences, University of Edinburgh / Institute of Genetics and Molecular Medicine, University of Edinburgh | COVID-19 Genomics UK (COG-UK) Consortium |
| EPI_ISL_456985 | Virology Department, Royal Infirmary of Edinburgh, NHS Lothian / School of Biological Sciences, University of Edinburgh / Institute of Genetics and Molecular Medicine, University of Edinburgh | COVID-19 Genomics UK (COG-UK) Consortium |
| EPI_ISL_456986 | Virology Department, Royal Infirmary of Edinburgh, NHS Lothian / School of Biological Sciences, University of Edinburgh / Institute of Genetics and Molecular Medicine, University of Edinburgh | COVID-19 Genomics UK (COG-UK) Consortium |
| EPI_ISL_456987 | Virology Department, Royal Infirmary of Edinburgh, NHS Lothian / School of Biological Sciences, University of Edinburgh / Institute of Genetics and Molecular Medicine, University of Edinburgh | COVID-19 Genomics UK (COG-UK) Consortium |
| EPI_ISL_456988 | Virology Department, Royal Infirmary of Edinburgh, NHS Lothian / School of Biological Sciences, University of Edinburgh / Institute of Genetics and Molecular Medicine, University of Edinburgh | COVID-19 Genomics UK (COG-UK) Consortium |
| EPI_ISL_456989 | Virology Department, Royal Infirmary of Edinburgh, NHS Lothian / School of Biological Sciences, University of Edinburgh / Institute of Genetics and Molecular Medicine, University of Edinburgh | COVID-19 Genomics UK (COG-UK) Consortium |
| EPI_ISL_456990 | Virology Department, Royal Infirmary of Edinburgh, NHS Lothian / School of Biological Sciences, University of Edinburgh / Institute of Genetics and Molecular Medicine, University of Edinburgh | COVID-19 Genomics UK (COG-UK) Consortium |

| EPI_Accession_ID | Originating Laboratory | Submitting Laboratory |
| --- | --- | --- |
| EPI_ISL_456991 | Virology Department, Royal Infirmary of Edinburgh, NHS Lothian / School of Biological Sciences, University of Edinburgh / Institute of Genetics and Molecular Medicine, University of Edinburgh | COVID-19 Genomics UK (COG-UK) Consortium |
| EPI_ISL_456992 | Virology Department, Royal Infirmary of Edinburgh, NHS Lothian / School of Biological Sciences, University of Edinburgh / Institute of Genetics and Molecular Medicine, University of Edinburgh | COVID-19 Genomics UK (COG-UK) Consortium |
| EPI_ISL_456993 | Virology Department, Royal Infirmary of Edinburgh, NHS Lothian / School of Biological Sciences, University of Edinburgh / Institute of Genetics and Molecular Medicine, University of Edinburgh | COVID-19 Genomics UK (COG-UK) Consortium |
| EPI_ISL_456994 | Virology Department, Royal Infirmary of Edinburgh, NHS Lothian / School of Biological Sciences, University of Edinburgh / Institute of Genetics and Molecular Medicine, University of Edinburgh | COVID-19 Genomics UK (COG-UK) Consortium |
| EPI_ISL_456995 | Virology Department, Royal Infirmary of Edinburgh, NHS Lothian / School of Biological Sciences, University of Edinburgh / Institute of Genetics and Molecular Medicine, University of Edinburgh | COVID-19 Genomics UK (COG-UK) Consortium |
| EPI_ISL_456996 | Virology Department, Royal Infirmary of Edinburgh, NHS Lothian / School of Biological Sciences, University of Edinburgh / Institute of Genetics and Molecular Medicine, University of Edinburgh | COVID-19 Genomics UK (COG-UK) Consortium |
| EPI_ISL_456997 | Virology Department, Royal Infirmary of Edinburgh, NHS Lothian / School of Biological Sciences, University of Edinburgh / Institute of Genetics and Molecular Medicine, University of Edinburgh | COVID-19 Genomics UK (COG-UK) Consortium |
| EPI_ISL_456998 | Virology Department, Royal Infirmary of Edinburgh, NHS Lothian / School of Biological Sciences, University of Edinburgh / Institute of Genetics and Molecular Medicine, University of Edinburgh | COVID-19 Genomics UK (COG-UK) Consortium |

| EPI_Accession_ID | Originating Laboratory | Submitting Laboratory |
|---|---|---|
| EPI_ISL_456999 | Virology Department, Royal Infirmary of Edinburgh, NHS Lothian / School of Biological Sciences, University of Edinburgh / Institute of Genetics and Molecular Medicine, University of Edinburgh | COVID-19 Genomics UK (COG-UK) Consortium |
| EPI_ISL_457000 | Virology Department, Royal Infirmary of Edinburgh, NHS Lothian / School of Biological Sciences, University of Edinburgh / Institute of Genetics and Molecular Medicine, University of Edinburgh | COVID-19 Genomics UK (COG-UK) Consortium |
| EPI_ISL_457001 | Virology Department, Royal Infirmary of Edinburgh, NHS Lothian / School of Biological Sciences, University of Edinburgh / Institute of Genetics and Molecular Medicine, University of Edinburgh | COVID-19 Genomics UK (COG-UK) Consortium |
| EPI_ISL_457002 | Virology Department, Royal Infirmary of Edinburgh, NHS Lothian / School of Biological Sciences, University of Edinburgh / Institute of Genetics and Molecular Medicine, University of Edinburgh | COVID-19 Genomics UK (COG-UK) Consortium |
| EPI_ISL_457003 | Virology Department, Royal Infirmary of Edinburgh, NHS Lothian / School of Biological Sciences, University of Edinburgh / Institute of Genetics and Molecular Medicine, University of Edinburgh | COVID-19 Genomics UK (COG-UK) Consortium |
| EPI_ISL_457004 | Virology Department, Royal Infirmary of Edinburgh, NHS Lothian / School of Biological Sciences, University of Edinburgh / Institute of Genetics and Molecular Medicine, University of Edinburgh | COVID-19 Genomics UK (COG-UK) Consortium |
| EPI_ISL_457005 | Virology Department, Royal Infirmary of Edinburgh, NHS Lothian / School of Biological Sciences, University of Edinburgh / Institute of Genetics and Molecular Medicine, University of Edinburgh | COVID-19 Genomics UK (COG-UK) Consortium |
| EPI_ISL_457006 | Virology Department, Royal Infirmary of Edinburgh, NHS Lothian / School of Biological Sciences, University of Edinburgh / Institute of Genetics and Molecular Medicine, University of Edinburgh | COVID-19 Genomics UK (COG-UK) Consortium |

| EPI_Accession_ID | Originating Laboratory | Submitting Laboratory |
|---|---|---|
| EPI_ISL_457007 | Virology Department, Royal Infirmary of Edinburgh, NHS Lothian / School of Biological Sciences, University of Edinburgh / Institute of Genetics and Molecular Medicine, University of Edinburgh | COVID-19 Genomics UK (COG-UK) Consortium |
| EPI_ISL_457008 | Virology Department, Royal Infirmary of Edinburgh, NHS Lothian / School of Biological Sciences, University of Edinburgh / Institute of Genetics and Molecular Medicine, University of Edinburgh | COVID-19 Genomics UK (COG-UK) Consortium |
| EPI_ISL_457009 | Virology Department, Royal Infirmary of Edinburgh, NHS Lothian / School of Biological Sciences, University of Edinburgh / Institute of Genetics and Molecular Medicine, University of Edinburgh | COVID-19 Genomics UK (COG-UK) Consortium |
| EPI_ISL_457010 | Virology Department, Royal Infirmary of Edinburgh, NHS Lothian / School of Biological Sciences, University of Edinburgh / Institute of Genetics and Molecular Medicine, University of Edinburgh | COVID-19 Genomics UK (COG-UK) Consortium |
| EPI_ISL_457011 | Virology Department, Royal Infirmary of Edinburgh, NHS Lothian / School of Biological Sciences, University of Edinburgh / Institute of Genetics and Molecular Medicine, University of Edinburgh | COVID-19 Genomics UK (COG-UK) Consortium |
| EPI_ISL_457012 | Virology Department, Royal Infirmary of Edinburgh, NHS Lothian / School of Biological Sciences, University of Edinburgh / Institute of Genetics and Molecular Medicine, University of Edinburgh | COVID-19 Genomics UK (COG-UK) Consortium |
| EPI_ISL_457013 | Virology Department, Royal Infirmary of Edinburgh, NHS Lothian / School of Biological Sciences, University of Edinburgh / Institute of Genetics and Molecular Medicine, University of Edinburgh | COVID-19 Genomics UK (COG-UK) Consortium |
| EPI_ISL_457014 | Virology Department, Royal Infirmary of Edinburgh, NHS Lothian / School of Biological Sciences, University of Edinburgh / Institute of Genetics and Molecular Medicine, University of Edinburgh | COVID-19 Genomics UK (COG-UK) Consortium |

| EPI_Accession_ID | Originating Laboratory | Submitting Laboratory |
|---|---|---|
| EPI_ISL_457015 | Virology Department, Royal Infirmary of Edinburgh, NHS Lothian / School of Biological Sciences, University of Edinburgh / Institute of Genetics and Molecular Medicine, University of Edinburgh | COVID-19 Genomics UK (COG-UK) Consortium |
| EPI_ISL_457016 | Virology Department, Royal Infirmary of Edinburgh, NHS Lothian / School of Biological Sciences, University of Edinburgh / Institute of Genetics and Molecular Medicine, University of Edinburgh | COVID-19 Genomics UK (COG-UK) Consortium |
| EPI_ISL_457017 | Virology Department, Royal Infirmary of Edinburgh, NHS Lothian / School of Biological Sciences, University of Edinburgh / Institute of Genetics and Molecular Medicine, University of Edinburgh | COVID-19 Genomics UK (COG-UK) Consortium |
| EPI_ISL_457018 | Virology Department, Royal Infirmary of Edinburgh, NHS Lothian / School of Biological Sciences, University of Edinburgh / Institute of Genetics and Molecular Medicine, University of Edinburgh | COVID-19 Genomics UK (COG-UK) Consortium |
| EPI_ISL_457019 | Virology Department, Royal Infirmary of Edinburgh, NHS Lothian / School of Biological Sciences, University of Edinburgh / Institute of Genetics and Molecular Medicine, University of Edinburgh | COVID-19 Genomics UK (COG-UK) Consortium |
| EPI_ISL_457020 | Virology Department, Royal Infirmary of Edinburgh, NHS Lothian / School of Biological Sciences, University of Edinburgh / Institute of Genetics and Molecular Medicine, University of Edinburgh | COVID-19 Genomics UK (COG-UK) Consortium |
| EPI_ISL_457021 | Virology Department, Royal Infirmary of Edinburgh, NHS Lothian / School of Biological Sciences, University of Edinburgh / Institute of Genetics and Molecular Medicine, University of Edinburgh | COVID-19 Genomics UK (COG-UK) Consortium |
| EPI_ISL_457022 | Virology Department, Royal Infirmary of Edinburgh, NHS Lothian / School of Biological Sciences, University of Edinburgh / Institute of Genetics and Molecular Medicine, University of Edinburgh | COVID-19 Genomics UK (COG-UK) Consortium |

| EPI_Accession_ID | Originating Laboratory | Submitting Laboratory |
|---|---|---|
| EPI_ISL_457023 | Virology Department, Royal Infirmary of Edinburgh, NHS Lothian / School of Biological Sciences, University of Edinburgh / Institute of Genetics and Molecular Medicine, University of Edinburgh | COVID-19 Genomics UK (COG-UK) Consortium |
| EPI_ISL_457028 | University of Exeter | COVID-19 Genomics UK (COG-UK) Consortium |
| EPI_ISL_457029 | University of Exeter | COVID-19 Genomics UK (COG-UK) Consortium |
| EPI_ISL_457031 | University of Exeter | COVID-19 Genomics UK (COG-UK) Consortium |
| EPI_ISL_457032 | University of Exeter | COVID-19 Genomics UK (COG-UK) Consortium |
| EPI_ISL_457033 | University of Exeter | COVID-19 Genomics UK (COG-UK) Consortium |
| EPI_ISL_457034 | University of Exeter | COVID-19 Genomics UK (COG-UK) Consortium |
| EPI_ISL_457035 | University of Exeter | COVID-19 Genomics UK (COG-UK) Consortium |
| EPI_ISL_457036 | University of Exeter | COVID-19 Genomics UK (COG-UK) Consortium |
| EPI_ISL_457037 | University of Exeter | COVID-19 Genomics UK (COG-UK) Consortium |
| EPI_ISL_457038 | University of Exeter | COVID-19 Genomics UK (COG-UK) Consortium |
| EPI_ISL_457039 | University of Exeter | COVID-19 Genomics UK (COG-UK) Consortium |
| EPI_ISL_457040 | University of Exeter | COVID-19 Genomics UK (COG-UK) Consortium |
| EPI_ISL_457041 | University of Exeter | COVID-19 Genomics UK (COG-UK) Consortium |
| EPI_ISL_457042 | University of Exeter | COVID-19 Genomics UK (COG-UK) Consortium |
| EPI_ISL_457043 | University of Exeter | COVID-19 Genomics UK (COG-UK) Consortium |
| EPI_ISL_457044 | University of Exeter | COVID-19 Genomics UK (COG-UK) Consortium |
| EPI_ISL_457045 | University of Exeter | COVID-19 Genomics UK (COG-UK) Consortium |
| EPI_ISL_457046 | University of Exeter | COVID-19 Genomics UK (COG-UK) Consortium |
| EPI_ISL_457047 | University of Exeter | COVID-19 Genomics UK (COG-UK) Consortium |
| EPI_ISL_457048 | University of Exeter | COVID-19 Genomics UK (COG-UK) Consortium |
| EPI_ISL_457049 | University of Exeter | COVID-19 Genomics UK (COG-UK) Consortium |
| EPI_ISL_457050 | University of Exeter | COVID-19 Genomics UK (COG-UK) Consortium |
| EPI_ISL_457051 | University of Exeter | COVID-19 Genomics UK (COG-UK) Consortium |
| EPI_ISL_457052 | University of Exeter | COVID-19 Genomics UK (COG-UK) Consortium |
| EPI_ISL_457053 | University of Exeter | COVID-19 Genomics UK (COG-UK) Consortium |
| EPI_ISL_457054 | University of Exeter | COVID-19 Genomics UK (COG-UK) Consortium |
| EPI_ISL_457055 | University of Exeter | COVID-19 Genomics UK (COG-UK) Consortium |
| EPI_ISL_457056 | University of Exeter | COVID-19 Genomics UK (COG-UK) Consortium |
| EPI_ISL_457057 | University of Exeter | COVID-19 Genomics UK (COG-UK) Consortium |

| EPI_Accession_ID | Originating Laboratory | Submitting Laboratory |
|---|---|---|
| EPI_ISL_457058 | University of Exeter | COVID-19 Genomics UK (COG-UK) Consortium |
| EPI_ISL_457059 | University of Exeter | COVID-19 Genomics UK (COG-UK) Consortium |
| EPI_ISL_457060 | University of Exeter | COVID-19 Genomics UK (COG-UK) Consortium |
| EPI_ISL_457061 | University of Exeter | COVID-19 Genomics UK (COG-UK) Consortium |
| EPI_ISL_457062 | University of Exeter | COVID-19 Genomics UK (COG-UK) Consortium |
| EPI_ISL_457063 | University of Exeter | COVID-19 Genomics UK (COG-UK) Consortium |
| EPI_ISL_457064 | University of Exeter | COVID-19 Genomics UK (COG-UK) Consortium |
| EPI_ISL_457065 | University of Exeter | COVID-19 Genomics UK (COG-UK) Consortium |
| EPI_ISL_457066 | University of Exeter | COVID-19 Genomics UK (COG-UK) Consortium |
| EPI_ISL_457067 | University of Exeter | COVID-19 Genomics UK (COG-UK) Consortium |
| EPI_ISL_457068 | University of Exeter | COVID-19 Genomics UK (COG-UK) Consortium |
| EPI_ISL_457069 | University of Exeter | COVID-19 Genomics UK (COG-UK) Consortium |
| EPI_ISL_457070 | University of Exeter | COVID-19 Genomics UK (COG-UK) Consortium |
| EPI_ISL_457071 | University of Exeter | COVID-19 Genomics UK (COG-UK) Consortium |
| EPI_ISL_457072 | University of Exeter | COVID-19 Genomics UK (COG-UK) Consortium |
| EPI_ISL_457073 | University of Exeter | COVID-19 Genomics UK (COG-UK) Consortium |
| EPI_ISL_457074 | University of Exeter | COVID-19 Genomics UK (COG-UK) Consortium |
| EPI_ISL_457075 | University of Exeter | COVID-19 Genomics UK (COG-UK) Consortium |
| EPI_ISL_457076 | University of Exeter | COVID-19 Genomics UK (COG-UK) Consortium |
| EPI_ISL_457077 | University of Exeter | COVID-19 Genomics UK (COG-UK) Consortium |
| EPI_ISL_457078 | University of Exeter | COVID-19 Genomics UK (COG-UK) Consortium |
| EPI_ISL_457079 | University of Exeter | COVID-19 Genomics UK (COG-UK) Consortium |
| EPI_ISL_457080 | University of Exeter | COVID-19 Genomics UK (COG-UK) Consortium |
| EPI_ISL_457081 | University of Exeter | COVID-19 Genomics UK (COG-UK) Consortium |
| EPI_ISL_457082 | University of Exeter | COVID-19 Genomics UK (COG-UK) Consortium |
| EPI_ISL_457083 | University of Exeter | COVID-19 Genomics UK (COG-UK) Consortium |
| EPI_ISL_457084 | University of Exeter | COVID-19 Genomics UK (COG-UK) Consortium |
| EPI_ISL_457085 | University of Exeter | COVID-19 Genomics UK (COG-UK) Consortium |
| EPI_ISL_457086 | University of Exeter | COVID-19 Genomics UK (COG-UK) Consortium |
| EPI_ISL_457087 | University of Exeter | COVID-19 Genomics UK (COG-UK) Consortium |
| EPI_ISL_457088 | University of Exeter | COVID-19 Genomics UK (COG-UK) Consortium |
| EPI_ISL_457089 | University of Exeter | COVID-19 Genomics UK (COG-UK) Consortium |
| EPI_ISL_457090 | University of Exeter | COVID-19 Genomics UK (COG-UK) Consortium |

| EPI_Accession_ID | Originating Laboratory | Submitting Laboratory |
|---|---|---|
| EPI_ISL_457091 | University of Exeter | COVID-19 Genomics UK (COG-UK) Consortium |
| EPI_ISL_457092 | University of Exeter | COVID-19 Genomics UK (COG-UK) Consortium |
| EPI_ISL_457093 | University of Exeter | COVID-19 Genomics UK (COG-UK) Consortium |
| EPI_ISL_457094 | University of Exeter | COVID-19 Genomics UK (COG-UK) Consortium |
| EPI_ISL_457095 | University of Exeter | COVID-19 Genomics UK (COG-UK) Consortium |
| EPI_ISL_457096 | University of Exeter | COVID-19 Genomics UK (COG-UK) Consortium |
| EPI_ISL_457097 | University of Exeter | COVID-19 Genomics UK (COG-UK) Consortium |
| EPI_ISL_457098 | University of Exeter | COVID-19 Genomics UK (COG-UK) Consortium |
| EPI_ISL_457099 | University of Exeter | COVID-19 Genomics UK (COG-UK) Consortium |
| EPI_ISL_457100 | University of Exeter | COVID-19 Genomics UK (COG-UK) Consortium |
| EPI_ISL_457101 | University of Exeter | COVID-19 Genomics UK (COG-UK) Consortium |
| EPI_ISL_457102 | University of Exeter | COVID-19 Genomics UK (COG-UK) Consortium |
| EPI_ISL_457103 | University of Exeter | COVID-19 Genomics UK (COG-UK) Consortium |
| EPI_ISL_457104 | University of Exeter | COVID-19 Genomics UK (COG-UK) Consortium |
| EPI_ISL_457105 | University of Exeter | COVID-19 Genomics UK (COG-UK) Consortium |
| EPI_ISL_457106 | University of Exeter | COVID-19 Genomics UK (COG-UK) Consortium |
| EPI_ISL_457107 | University of Exeter | COVID-19 Genomics UK (COG-UK) Consortium |
| EPI_ISL_457108 | University of Exeter | COVID-19 Genomics UK (COG-UK) Consortium |
| EPI_ISL_457109 | University of Exeter | COVID-19 Genomics UK (COG-UK) Consortium |
| EPI_ISL_457110 | University of Exeter | COVID-19 Genomics UK (COG-UK) Consortium |
| EPI_ISL_457111 | University of Exeter | COVID-19 Genomics UK (COG-UK) Consortium |
| EPI_ISL_457112 | University of Exeter | COVID-19 Genomics UK (COG-UK) Consortium |
| EPI_ISL_457113 | University of Exeter | COVID-19 Genomics UK (COG-UK) Consortium |
| EPI_ISL_457114 | University of Exeter | COVID-19 Genomics UK (COG-UK) Consortium |
| EPI_ISL_457115 | University of Exeter | COVID-19 Genomics UK (COG-UK) Consortium |
| EPI_ISL_457116 | University of Exeter | COVID-19 Genomics UK (COG-UK) Consortium |
| EPI_ISL_457117 | University of Exeter | COVID-19 Genomics UK (COG-UK) Consortium |
| EPI_ISL_457118 | University of Exeter | COVID-19 Genomics UK (COG-UK) Consortium |
| EPI_ISL_457119 | University of Exeter | COVID-19 Genomics UK (COG-UK) Consortium |
| EPI_ISL_457120 | University of Exeter | COVID-19 Genomics UK (COG-UK) Consortium |
| EPI_ISL_457121 | University of Exeter | COVID-19 Genomics UK (COG-UK) Consortium |
| EPI_ISL_457122 | University of Exeter | COVID-19 Genomics UK (COG-UK) Consortium |
| EPI_ISL_457123 | University of Exeter | COVID-19 Genomics UK (COG-UK) Consortium |

| EPI_Accession_ID | Originating Laboratory | Submitting Laboratory |
|---|---|---|
| EPI_ISL_457124 | University of Exeter | COVID-19 Genomics UK (COG-UK) Consortium |
| EPI_ISL_457125 | University of Exeter | COVID-19 Genomics UK (COG-UK) Consortium |
| EPI_ISL_457126 | University of Exeter | COVID-19 Genomics UK (COG-UK) Consortium |
| EPI_ISL_457127 | University of Exeter | COVID-19 Genomics UK (COG-UK) Consortium |
| EPI_ISL_457128 | University of Exeter | COVID-19 Genomics UK (COG-UK) Consortium |
| EPI_ISL_457129 | University of Exeter | COVID-19 Genomics UK (COG-UK) Consortium |
| EPI_ISL_457130 | University of Exeter | COVID-19 Genomics UK (COG-UK) Consortium |
| EPI_ISL_457131 | University of Exeter | COVID-19 Genomics UK (COG-UK) Consortium |
| EPI_ISL_457132 | University of Exeter | COVID-19 Genomics UK (COG-UK) Consortium |
| EPI_ISL_457133 | University of Exeter | COVID-19 Genomics UK (COG-UK) Consortium |
| EPI_ISL_457134 | University of Exeter | COVID-19 Genomics UK (COG-UK) Consortium |
| EPI_ISL_457135 | University of Exeter | COVID-19 Genomics UK (COG-UK) Consortium |
| EPI_ISL_457136 | University of Exeter | COVID-19 Genomics UK (COG-UK) Consortium |
| EPI_ISL_457138 | University of Exeter | COVID-19 Genomics UK (COG-UK) Consortium |
| EPI_ISL_457139 | University of Exeter | COVID-19 Genomics UK (COG-UK) Consortium |
| EPI_ISL_457140 | University of Exeter | COVID-19 Genomics UK (COG-UK) Consortium |
| EPI_ISL_457141 | University of Exeter | COVID-19 Genomics UK (COG-UK) Consortium |
| EPI_ISL_457142 | University of Exeter | COVID-19 Genomics UK (COG-UK) Consortium |
| EPI_ISL_457143 | University of Exeter | COVID-19 Genomics UK (COG-UK) Consortium |
| EPI_ISL_457144 | University of Exeter | COVID-19 Genomics UK (COG-UK) Consortium |
| EPI_ISL_457145 | University of Exeter | COVID-19 Genomics UK (COG-UK) Consortium |
| EPI_ISL_457146 | University of Exeter | COVID-19 Genomics UK (COG-UK) Consortium |
| EPI_ISL_457147 | University of Exeter | COVID-19 Genomics UK (COG-UK) Consortium |
| EPI_ISL_457149 | University of Exeter | COVID-19 Genomics UK (COG-UK) Consortium |
| EPI_ISL_457150 | University of Exeter | COVID-19 Genomics UK (COG-UK) Consortium |
| EPI_ISL_457151 | University of Exeter | COVID-19 Genomics UK (COG-UK) Consortium |
| EPI_ISL_457153 | University of Exeter | COVID-19 Genomics UK (COG-UK) Consortium |
| EPI_ISL_457154 | University of Exeter | COVID-19 Genomics UK (COG-UK) Consortium |
| EPI_ISL_457155 | University of Exeter | COVID-19 Genomics UK (COG-UK) Consortium |
| EPI_ISL_457156 | University of Exeter | COVID-19 Genomics UK (COG-UK) Consortium |
| EPI_ISL_457157 | University of Exeter | COVID-19 Genomics UK (COG-UK) Consortium |
| EPI_ISL_457158 | University of Exeter | COVID-19 Genomics UK (COG-UK) Consortium |
| EPI_ISL_457159 | University of Exeter | COVID-19 Genomics UK (COG-UK) Consortium |

| EPI_Accession_ID | Originating Laboratory | Submitting Laboratory |
|---|---|---|
| EPI_ISL_457160 | University of Exeter | COVID-19 Genomics UK (COG-UK) Consortium |
| EPI_ISL_457161 | University of Exeter | COVID-19 Genomics UK (COG-UK) Consortium |
| EPI_ISL_457162 | University of Exeter | COVID-19 Genomics UK (COG-UK) Consortium |
| EPI_ISL_457163 | University of Exeter | COVID-19 Genomics UK (COG-UK) Consortium |
| EPI_ISL_457164 | University of Exeter | COVID-19 Genomics UK (COG-UK) Consortium |
| EPI_ISL_457165 | University of Exeter | COVID-19 Genomics UK (COG-UK) Consortium |
| EPI_ISL_457166 | University of Exeter | COVID-19 Genomics UK (COG-UK) Consortium |
| EPI_ISL_457167 | University of Exeter | COVID-19 Genomics UK (COG-UK) Consortium |
| EPI_ISL_457168 | University of Exeter | COVID-19 Genomics UK (COG-UK) Consortium |
| EPI_ISL_457169 | University of Exeter | COVID-19 Genomics UK (COG-UK) Consortium |
| EPI_ISL_457170 | University of Exeter | COVID-19 Genomics UK (COG-UK) Consortium |
| EPI_ISL_457171 | University of Exeter | COVID-19 Genomics UK (COG-UK) Consortium |
| EPI_ISL_457172 | University of Exeter | COVID-19 Genomics UK (COG-UK) Consortium |
| EPI_ISL_457173 | University of Exeter | COVID-19 Genomics UK (COG-UK) Consortium |
| EPI_ISL_457174 | University of Exeter | COVID-19 Genomics UK (COG-UK) Consortium |
| EPI_ISL_457175 | University of Exeter | COVID-19 Genomics UK (COG-UK) Consortium |
| EPI_ISL_457176 | University of Exeter | COVID-19 Genomics UK (COG-UK) Consortium |
| EPI_ISL_457177 | University of Exeter | COVID-19 Genomics UK (COG-UK) Consortium |
| EPI_ISL_457178 | University of Exeter | COVID-19 Genomics UK (COG-UK) Consortium |
| EPI_ISL_457179 | University of Exeter | COVID-19 Genomics UK (COG-UK) Consortium |
| EPI_ISL_457180 | University of Exeter | COVID-19 Genomics UK (COG-UK) Consortium |
| EPI_ISL_457181 | University of Exeter | COVID-19 Genomics UK (COG-UK) Consortium |
| EPI_ISL_457182 | University of Exeter | COVID-19 Genomics UK (COG-UK) Consortium |
| EPI_ISL_457183 | University of Exeter | COVID-19 Genomics UK (COG-UK) Consortium |
| EPI_ISL_457184 | University of Exeter | COVID-19 Genomics UK (COG-UK) Consortium |
| EPI_ISL_457185 | University of Exeter | COVID-19 Genomics UK (COG-UK) Consortium |
| EPI_ISL_457186 | University of Exeter | COVID-19 Genomics UK (COG-UK) Consortium |
| EPI_ISL_457187 | University of Exeter | COVID-19 Genomics UK (COG-UK) Consortium |
| EPI_ISL_457188 | University of Exeter | COVID-19 Genomics UK (COG-UK) Consortium |
| EPI_ISL_457189 | University of Exeter | COVID-19 Genomics UK (COG-UK) Consortium |
| EPI_ISL_457190 | University of Exeter | COVID-19 Genomics UK (COG-UK) Consortium |
| EPI_ISL_457191 | University of Exeter | COVID-19 Genomics UK (COG-UK) Consortium |
| EPI_ISL_457192 | University of Exeter | COVID-19 Genomics UK (COG-UK) Consortium |

| EPI_Accession_ID | Originating Laboratory | Submitting Laboratory |
|---|---|---|
| EPI_ISL_457193 | University of Exeter | COVID-19 Genomics UK (COG-UK) Consortium |
| EPI_ISL_457194 | University of Exeter | COVID-19 Genomics UK (COG-UK) Consortium |
| EPI_ISL_457195 | University of Exeter | COVID-19 Genomics UK (COG-UK) Consortium |
| EPI_ISL_457196 | University of Exeter | COVID-19 Genomics UK (COG-UK) Consortium |
| EPI_ISL_457197 | University of Exeter | COVID-19 Genomics UK (COG-UK) Consortium |
| EPI_ISL_457198 | University of Exeter | COVID-19 Genomics UK (COG-UK) Consortium |
| EPI_ISL_457199 | University of Exeter | COVID-19 Genomics UK (COG-UK) Consortium |
| EPI_ISL_457200 | University of Exeter | COVID-19 Genomics UK (COG-UK) Consortium |
| EPI_ISL_457201 | University of Exeter | COVID-19 Genomics UK (COG-UK) Consortium |
| EPI_ISL_457202 | University of Exeter | COVID-19 Genomics UK (COG-UK) Consortium |
| EPI_ISL_457203 | University of Exeter | COVID-19 Genomics UK (COG-UK) Consortium |
| EPI_ISL_457204 | University of Exeter | COVID-19 Genomics UK (COG-UK) Consortium |
| EPI_ISL_457205 | University of Exeter | COVID-19 Genomics UK (COG-UK) Consortium |
| EPI_ISL_457206 | University of Exeter | COVID-19 Genomics UK (COG-UK) Consortium |
| EPI_ISL_457207 | University of Exeter | COVID-19 Genomics UK (COG-UK) Consortium |
| EPI_ISL_457208 | University of Exeter | COVID-19 Genomics UK (COG-UK) Consortium |
| EPI_ISL_457209 | University of Exeter | COVID-19 Genomics UK (COG-UK) Consortium |
| EPI_ISL_457210 | University of Exeter | COVID-19 Genomics UK (COG-UK) Consortium |
| EPI_ISL_457211 | University of Exeter | COVID-19 Genomics UK (COG-UK) Consortium |
| EPI_ISL_457212 | University of Exeter | COVID-19 Genomics UK (COG-UK) Consortium |
| EPI_ISL_457213 | University of Exeter | COVID-19 Genomics UK (COG-UK) Consortium |
| EPI_ISL_457214 | University of Exeter | COVID-19 Genomics UK (COG-UK) Consortium |
| EPI_ISL_457215 | University of Exeter | COVID-19 Genomics UK (COG-UK) Consortium |
| EPI_ISL_457216 | University of Exeter | COVID-19 Genomics UK (COG-UK) Consortium |
| EPI_ISL_457217 | University of Exeter | COVID-19 Genomics UK (COG-UK) Consortium |
| EPI_ISL_457218 | University of Exeter | COVID-19 Genomics UK (COG-UK) Consortium |
| EPI_ISL_457219 | University of Exeter | COVID-19 Genomics UK (COG-UK) Consortium |
| EPI_ISL_457220 | University of Exeter | COVID-19 Genomics UK (COG-UK) Consortium |
| EPI_ISL_457221 | University of Exeter | COVID-19 Genomics UK (COG-UK) Consortium |
| EPI_ISL_457222 | University of Exeter | COVID-19 Genomics UK (COG-UK) Consortium |
| EPI_ISL_457223 | University of Exeter | COVID-19 Genomics UK (COG-UK) Consortium |
| EPI_ISL_457224 | University of Exeter | COVID-19 Genomics UK (COG-UK) Consortium |
| EPI_ISL_457225 | University of Exeter | COVID-19 Genomics UK (COG-UK) Consortium |

| EPI_Accession_ID | Originating Laboratory | Submitting Laboratory |
|---|---|---|
| EPI_ISL_457226 | University of Exeter | COVID-19 Genomics UK (COG-UK) Consortium |
| EPI_ISL_457227 | University of Exeter | COVID-19 Genomics UK (COG-UK) Consortium |
| EPI_ISL_457228 | University of Exeter | COVID-19 Genomics UK (COG-UK) Consortium |
| EPI_ISL_457229 | University of Exeter | COVID-19 Genomics UK (COG-UK) Consortium |
| EPI_ISL_457230 | University of Exeter | COVID-19 Genomics UK (COG-UK) Consortium |
| EPI_ISL_457231 | University of Exeter | COVID-19 Genomics UK (COG-UK) Consortium |
| EPI_ISL_457232 | University of Exeter | COVID-19 Genomics UK (COG-UK) Consortium |
| EPI_ISL_457233 | University of Exeter | COVID-19 Genomics UK (COG-UK) Consortium |
| EPI_ISL_457234 | University of Exeter | COVID-19 Genomics UK (COG-UK) Consortium |
| EPI_ISL_457235 | University of Exeter | COVID-19 Genomics UK (COG-UK) Consortium |
| EPI_ISL_457236 | University of Exeter | COVID-19 Genomics UK (COG-UK) Consortium |
| EPI_ISL_457237 | University of Exeter | COVID-19 Genomics UK (COG-UK) Consortium |
| EPI_ISL_457238 | University of Exeter | COVID-19 Genomics UK (COG-UK) Consortium |
| EPI_ISL_457239 | University of Exeter | COVID-19 Genomics UK (COG-UK) Consortium |
| EPI_ISL_457240 | University of Exeter | COVID-19 Genomics UK (COG-UK) Consortium |
| EPI_ISL_457241 | University of Exeter | COVID-19 Genomics UK (COG-UK) Consortium |
| EPI_ISL_457242 | University of Exeter | COVID-19 Genomics UK (COG-UK) Consortium |
| EPI_ISL_457243 | University of Exeter | COVID-19 Genomics UK (COG-UK) Consortium |
| EPI_ISL_457244 | University of Exeter | COVID-19 Genomics UK (COG-UK) Consortium |
| EPI_ISL_457245 | University of Exeter | COVID-19 Genomics UK (COG-UK) Consortium |
| EPI_ISL_457246 | University of Exeter | COVID-19 Genomics UK (COG-UK) Consortium |
| EPI_ISL_457247 | University of Exeter | COVID-19 Genomics UK (COG-UK) Consortium |
| EPI_ISL_457248 | University of Exeter | COVID-19 Genomics UK (COG-UK) Consortium |
| EPI_ISL_457249 | University of Exeter | COVID-19 Genomics UK (COG-UK) Consortium |
| EPI_ISL_457250 | University of Exeter | COVID-19 Genomics UK (COG-UK) Consortium |
| EPI_ISL_457251 | University of Exeter | COVID-19 Genomics UK (COG-UK) Consortium |
| EPI_ISL_457252 | University of Exeter | COVID-19 Genomics UK (COG-UK) Consortium |
| EPI_ISL_457253 | University of Exeter | COVID-19 Genomics UK (COG-UK) Consortium |
| EPI_ISL_457254 | University of Exeter | COVID-19 Genomics UK (COG-UK) Consortium |
| EPI_ISL_457255 | University of Exeter | COVID-19 Genomics UK (COG-UK) Consortium |
| EPI_ISL_457256 | University of Exeter | COVID-19 Genomics UK (COG-UK) Consortium |
| EPI_ISL_457257 | University of Exeter | COVID-19 Genomics UK (COG-UK) Consortium |
| EPI_ISL_457258 | University of Exeter | COVID-19 Genomics UK (COG-UK) Consortium |

| EPI_Accession_ID | Originating Laboratory | Submitting Laboratory |
|---|---|---|
| EPI_ISL_457259 | Liverpool Clinical Laboratories | COVID-19 Genomics UK (COG-UK) Consortium |
| EPI_ISL_457260 | Liverpool Clinical Laboratories | COVID-19 Genomics UK (COG-UK) Consortium |
| EPI_ISL_457261 | Liverpool Clinical Laboratories | COVID-19 Genomics UK (COG-UK) Consortium |
| EPI_ISL_457262 | Liverpool Clinical Laboratories | COVID-19 Genomics UK (COG-UK) Consortium |
| EPI_ISL_457263 | Liverpool Clinical Laboratories | COVID-19 Genomics UK (COG-UK) Consortium |
| EPI_ISL_457264 | Liverpool Clinical Laboratories | COVID-19 Genomics UK (COG-UK) Consortium |
| EPI_ISL_457265 | Liverpool Clinical Laboratories | COVID-19 Genomics UK (COG-UK) Consortium |
| EPI_ISL_457266 | Liverpool Clinical Laboratories | COVID-19 Genomics UK (COG-UK) Consortium |
| EPI_ISL_457267 | Liverpool Clinical Laboratories | COVID-19 Genomics UK (COG-UK) Consortium |
| EPI_ISL_457268 | Liverpool Clinical Laboratories | COVID-19 Genomics UK (COG-UK) Consortium |
| EPI_ISL_457269 | Liverpool Clinical Laboratories | COVID-19 Genomics UK (COG-UK) Consortium |
| EPI_ISL_457270 | University College London, Great Ormond Street Hospital for Children NHS Foundation Trust, Imperial College Healthcare NHS Trust | COVID-19 Genomics UK (COG-UK) Consortium |
| EPI_ISL_457271 | University College London, Great Ormond Street Hospital for Children NHS Foundation Trust, Imperial College Healthcare NHS Trust | COVID-19 Genomics UK (COG-UK) Consortium |
| EPI_ISL_457272 | University College London, Great Ormond Street Hospital for Children NHS Foundation Trust, Imperial College Healthcare NHS Trust | COVID-19 Genomics UK (COG-UK) Consortium |
| EPI_ISL_457273 | University College London, Great Ormond Street Hospital for Children NHS Foundation Trust, Imperial College Healthcare NHS Trust | COVID-19 Genomics UK (COG-UK) Consortium |
| EPI_ISL_457274 | University College London, Great Ormond Street Hospital for Children NHS Foundation Trust, Imperial College Healthcare NHS Trust | COVID-19 Genomics UK (COG-UK) Consortium |
| EPI_ISL_457275 | University College London, Great Ormond Street Hospital for Children NHS Foundation Trust, Imperial College Healthcare NHS Trust | COVID-19 Genomics UK (COG-UK) Consortium |
| EPI_ISL_457276 | University College London, Great Ormond Street Hospital for Children NHS Foundation Trust, Imperial College Healthcare NHS Trust | COVID-19 Genomics UK (COG-UK) Consortium |

| EPI_Accession_ID | Originating Laboratory | Submitting Laboratory |
|---|---|---|
| EPI_ISL_457277 | University College London, Great Ormond Street Hospital for Children NHS Foundation Trust, Imperial College Healthcare NHS Trust | COVID-19 Genomics UK (COG-UK) Consortium |
| EPI_ISL_457278 | University College London, Great Ormond Street Hospital for Children NHS Foundation Trust, Imperial College Healthcare NHS Trust | COVID-19 Genomics UK (COG-UK) Consortium |
| EPI_ISL_457279 | University College London, Great Ormond Street Hospital for Children NHS Foundation Trust, Imperial College Healthcare NHS Trust | COVID-19 Genomics UK (COG-UK) Consortium |
| EPI_ISL_457280 | University College London, Great Ormond Street Hospital for Children NHS Foundation Trust, Imperial College Healthcare NHS Trust | COVID-19 Genomics UK (COG-UK) Consortium |
| EPI_ISL_457281 | University College London, Great Ormond Street Hospital for Children NHS Foundation Trust, Imperial College Healthcare NHS Trust | COVID-19 Genomics UK (COG-UK) Consortium |
| EPI_ISL_457282 | University College London, Great Ormond Street Hospital for Children NHS Foundation Trust, Imperial College Healthcare NHS Trust | COVID-19 Genomics UK (COG-UK) Consortium |
| EPI_ISL_457283 | University College London, Great Ormond Street Hospital for Children NHS Foundation Trust, Imperial College Healthcare NHS Trust | COVID-19 Genomics UK (COG-UK) Consortium |
| EPI_ISL_457284 | University College London, Great Ormond Street Hospital for Children NHS Foundation Trust, Imperial College Healthcare NHS Trust | COVID-19 Genomics UK (COG-UK) Consortium |
| EPI_ISL_457285 | University College London, Great Ormond Street Hospital for Children NHS Foundation Trust, Imperial College Healthcare NHS Trust | COVID-19 Genomics UK (COG-UK) Consortium |
| EPI_ISL_457286 | University College London, Great Ormond Street Hospital for Children NHS Foundation Trust, Imperial College Healthcare NHS Trust | COVID-19 Genomics UK (COG-UK) Consortium |
| EPI_ISL_457287 | University College London, Great Ormond Street Hospital for Children NHS Foundation Trust, Imperial College Healthcare NHS Trust | COVID-19 Genomics UK (COG-UK) Consortium |

| EPI_Accession_ID | Originating Laboratory | Submitting Laboratory |
|---|---|---|
| EPI_ISL_457288 | University College London, Great Ormond Street Hospital for Children NHS Foundation Trust, Imperial College Healthcare NHS Trust | COVID-19 Genomics UK (COG-UK) Consortium |
| EPI_ISL_457289 | University College London, Great Ormond Street Hospital for Children NHS Foundation Trust, Imperial College Healthcare NHS Trust | COVID-19 Genomics UK (COG-UK) Consortium |
| EPI_ISL_457290 | University College London, Great Ormond Street Hospital for Children NHS Foundation Trust, Imperial College Healthcare NHS Trust | COVID-19 Genomics UK (COG-UK) Consortium |
| EPI_ISL_457291 | University College London, Great Ormond Street Hospital for Children NHS Foundation Trust, Imperial College Healthcare NHS Trust | COVID-19 Genomics UK (COG-UK) Consortium |
| EPI_ISL_457292 | University College London, Great Ormond Street Hospital for Children NHS Foundation Trust, Imperial College Healthcare NHS Trust | COVID-19 Genomics UK (COG-UK) Consortium |
| EPI_ISL_457293 | University College London, Great Ormond Street Hospital for Children NHS Foundation Trust, Imperial College Healthcare NHS Trust | COVID-19 Genomics UK (COG-UK) Consortium |
| EPI_ISL_457294 | University College London, Great Ormond Street Hospital for Children NHS Foundation Trust, Imperial College Healthcare NHS Trust | COVID-19 Genomics UK (COG-UK) Consortium |
| EPI_ISL_457295 | University College London, Great Ormond Street Hospital for Children NHS Foundation Trust, Imperial College Healthcare NHS Trust | COVID-19 Genomics UK (COG-UK) Consortium |
| EPI_ISL_457296 | University College London, Great Ormond Street Hospital for Children NHS Foundation Trust, Imperial College Healthcare NHS Trust | COVID-19 Genomics UK (COG-UK) Consortium |
| EPI_ISL_457297 | University College London, Great Ormond Street Hospital for Children NHS Foundation Trust, Imperial College Healthcare NHS Trust | COVID-19 Genomics UK (COG-UK) Consortium |
| EPI_ISL_457298 | University College London, Great Ormond Street Hospital for Children NHS Foundation Trust, Imperial College Healthcare NHS Trust | COVID-19 Genomics UK (COG-UK) Consortium |

| EPI_Accession_ID | Originating Laboratory | Submitting Laboratory |
|---|---|---|
| EPI_ISL_457299 | University College London, Great Ormond Street Hospital for Children NHS Foundation Trust, Imperial College Healthcare NHS Trust | COVID-19 Genomics UK (COG-UK) Consortium |
| EPI_ISL_457300 | University College London, Great Ormond Street Hospital for Children NHS Foundation Trust, Imperial College Healthcare NHS Trust | COVID-19 Genomics UK (COG-UK) Consortium |
| EPI_ISL_457301 | University College London, Great Ormond Street Hospital for Children NHS Foundation Trust, Imperial College Healthcare NHS Trust | COVID-19 Genomics UK (COG-UK) Consortium |
| EPI_ISL_457302 | University College London, Great Ormond Street Hospital for Children NHS Foundation Trust, Imperial College Healthcare NHS Trust | COVID-19 Genomics UK (COG-UK) Consortium |
| EPI_ISL_457303 | Northumbria University / South Tees Hospitals NHS Foundation Trust / North Cumbria Integrated Care NHS Foundation Trust / North Tees and Hartlepool NHS Foundation Trust / Newcastle Hospitals NHS Foundation Trust | COVID-19 Genomics UK (COG-UK) Consortium |
| EPI_ISL_457304 | Northumbria University / South Tees Hospitals NHS Foundation Trust / North Cumbria Integrated Care NHS Foundation Trust / North Tees and Hartlepool NHS Foundation Trust / Newcastle Hospitals NHS Foundation Trust | COVID-19 Genomics UK (COG-UK) Consortium |
| EPI_ISL_457305 | Northumbria University / South Tees Hospitals NHS Foundation Trust / North Cumbria Integrated Care NHS Foundation Trust / North Tees and Hartlepool NHS Foundation Trust / Newcastle Hospitals NHS Foundation Trust | COVID-19 Genomics UK (COG-UK) Consortium |
| EPI_ISL_457306 | Northumbria University / South Tees Hospitals NHS Foundation Trust / North Cumbria Integrated Care NHS Foundation Trust / North Tees and Hartlepool NHS Foundation Trust / Newcastle Hospitals NHS Foundation Trust | COVID-19 Genomics UK (COG-UK) Consortium |

| EPI_Accession_ID | Originating Laboratory | Submitting Laboratory |
|---|---|---|
| EPI_ISL_457307 | Northumbria University / South Tees Hospitals NHS Foundation Trust / North Cumbria Integrated Care NHS Foundation Trust / North Tees and Hartlepool NHS Foundation Trust / Newcastle Hospitals NHS Foundation Trust | COVID-19 Genomics UK (COG-UK) Consortium |
| EPI_ISL_457308 | Northumbria University / South Tees Hospitals NHS Foundation Trust / North Cumbria Integrated Care NHS Foundation Trust / North Tees and Hartlepool NHS Foundation Trust / Newcastle Hospitals NHS Foundation Trust | COVID-19 Genomics UK (COG-UK) Consortium |
| EPI_ISL_457309 | Northumbria University / South Tees Hospitals NHS Foundation Trust / North Cumbria Integrated Care NHS Foundation Trust / North Tees and Hartlepool NHS Foundation Trust / Newcastle Hospitals NHS Foundation Trust | COVID-19 Genomics UK (COG-UK) Consortium |
| EPI_ISL_457310 | Northumbria University / South Tees Hospitals NHS Foundation Trust / North Cumbria Integrated Care NHS Foundation Trust / North Tees and Hartlepool NHS Foundation Trust / Newcastle Hospitals NHS Foundation Trust | COVID-19 Genomics UK (COG-UK) Consortium |
| EPI_ISL_457311 | Northumbria University / South Tees Hospitals NHS Foundation Trust / North Cumbria Integrated Care NHS Foundation Trust / North Tees and Hartlepool NHS Foundation Trust / Newcastle Hospitals NHS Foundation Trust | COVID-19 Genomics UK (COG-UK) Consortium |
| EPI_ISL_457312 | Northumbria University / South Tees Hospitals NHS Foundation Trust / North Cumbria Integrated Care NHS Foundation Trust / North Tees and Hartlepool NHS Foundation Trust / Newcastle Hospitals NHS Foundation Trust | COVID-19 Genomics UK (COG-UK) Consortium |

| EPI_Accession_ID | Originating Laboratory | Submitting Laboratory |
|---|---|---|
| EPI_ISL_457313 | Northumbria University / South Tees Hospitals NHS Foundation Trust / North Cumbria Integrated Care NHS Foundation Trust / North Tees and Hartlepool NHS Foundation Trust / Newcastle Hospitals NHS Foundation Trust | COVID-19 Genomics UK (COG-UK) Consortium |
| EPI_ISL_457314 | Northumbria University / South Tees Hospitals NHS Foundation Trust / North Cumbria Integrated Care NHS Foundation Trust / North Tees and Hartlepool NHS Foundation Trust / Newcastle Hospitals NHS Foundation Trust | COVID-19 Genomics UK (COG-UK) Consortium |
| EPI_ISL_457315 | Northumbria University / South Tees Hospitals NHS Foundation Trust / North Cumbria Integrated Care NHS Foundation Trust / North Tees and Hartlepool NHS Foundation Trust / Newcastle Hospitals NHS Foundation Trust | COVID-19 Genomics UK (COG-UK) Consortium |
| EPI_ISL_457316 | Northumbria University / South Tees Hospitals NHS Foundation Trust / North Cumbria Integrated Care NHS Foundation Trust / North Tees and Hartlepool NHS Foundation Trust / Newcastle Hospitals NHS Foundation Trust | COVID-19 Genomics UK (COG-UK) Consortium |
| EPI_ISL_457317 | Northumbria University / South Tees Hospitals NHS Foundation Trust / North Cumbria Integrated Care NHS Foundation Trust / North Tees and Hartlepool NHS Foundation Trust / Newcastle Hospitals NHS Foundation Trust | COVID-19 Genomics UK (COG-UK) Consortium |
| EPI_ISL_457318 | Northumbria University / South Tees Hospitals NHS Foundation Trust / North Cumbria Integrated Care NHS Foundation Trust / North Tees and Hartlepool NHS Foundation Trust / Newcastle Hospitals NHS Foundation Trust | COVID-19 Genomics UK (COG-UK) Consortium |

| EPI_Accession_ID | Originating Laboratory | Submitting Laboratory |
|---|---|---|
| EPI_ISL_457319 | Northumbria University / South Tees Hospitals NHS Foundation Trust / North Cumbria Integrated Care NHS Foundation Trust / North Tees and Hartlepool NHS Foundation Trust / Newcastle Hospitals NHS Foundation Trust | COVID-19 Genomics UK (COG-UK) Consortium |
| EPI_ISL_457320 | Northumbria University / South Tees Hospitals NHS Foundation Trust / North Cumbria Integrated Care NHS Foundation Trust / North Tees and Hartlepool NHS Foundation Trust / Newcastle Hospitals NHS Foundation Trust | COVID-19 Genomics UK (COG-UK) Consortium |
| EPI_ISL_457321 | Northumbria University / South Tees Hospitals NHS Foundation Trust / North Cumbria Integrated Care NHS Foundation Trust / North Tees and Hartlepool NHS Foundation Trust / Newcastle Hospitals NHS Foundation Trust | COVID-19 Genomics UK (COG-UK) Consortium |
| EPI_ISL_457322 | Northumbria University / South Tees Hospitals NHS Foundation Trust / North Cumbria Integrated Care NHS Foundation Trust / North Tees and Hartlepool NHS Foundation Trust / Newcastle Hospitals NHS Foundation Trust | COVID-19 Genomics UK (COG-UK) Consortium |
| EPI_ISL_457323 | Northumbria University / South Tees Hospitals NHS Foundation Trust / North Cumbria Integrated Care NHS Foundation Trust / North Tees and Hartlepool NHS Foundation Trust / Newcastle Hospitals NHS Foundation Trust | COVID-19 Genomics UK (COG-UK) Consortium |
| EPI_ISL_457324 | Northumbria University / South Tees Hospitals NHS Foundation Trust / North Cumbria Integrated Care NHS Foundation Trust / North Tees and Hartlepool NHS Foundation Trust / Newcastle Hospitals NHS Foundation Trust | COVID-19 Genomics UK (COG-UK) Consortium |

| EPI_Accession_ID | Originating Laboratory | Submitting Laboratory |
|---|---|---|
| EPI_ISL_457325 | Northumbria University / South Tees Hospitals NHS Foundation Trust / North Cumbria Integrated Care NHS Foundation Trust / North Tees and Hartlepool NHS Foundation Trust / Newcastle Hospitals NHS Foundation Trust | COVID-19 Genomics UK (COG-UK) Consortium |
| EPI_ISL_457326 | Quadram Institute Bioscience | COVID-19 Genomics UK (COG-UK) Consortium |
| EPI_ISL_457327 | Quadram Institute Bioscience | COVID-19 Genomics UK (COG-UK) Consortium |
| EPI_ISL_457328 | Quadram Institute Bioscience | COVID-19 Genomics UK (COG-UK) Consortium |
| EPI_ISL_457329 | Quadram Institute Bioscience | COVID-19 Genomics UK (COG-UK) Consortium |
| EPI_ISL_457330 | Quadram Institute Bioscience | COVID-19 Genomics UK (COG-UK) Consortium |
| EPI_ISL_457331 | Quadram Institute Bioscience | COVID-19 Genomics UK (COG-UK) Consortium |
| EPI_ISL_457332 | Quadram Institute Bioscience | COVID-19 Genomics UK (COG-UK) Consortium |
| EPI_ISL_457333 | Quadram Institute Bioscience | COVID-19 Genomics UK (COG-UK) Consortium |
| EPI_ISL_457334 | Quadram Institute Bioscience | COVID-19 Genomics UK (COG-UK) Consortium |
| EPI_ISL_457335 | Quadram Institute Bioscience | COVID-19 Genomics UK (COG-UK) Consortium |
| EPI_ISL_457336 | Quadram Institute Bioscience | COVID-19 Genomics UK (COG-UK) Consortium |
| EPI_ISL_457337 | Quadram Institute Bioscience | COVID-19 Genomics UK (COG-UK) Consortium |
| EPI_ISL_457338 | Quadram Institute Bioscience | COVID-19 Genomics UK (COG-UK) Consortium |
| EPI_ISL_457339 | Quadram Institute Bioscience | COVID-19 Genomics UK (COG-UK) Consortium |
| EPI_ISL_457340 | Quadram Institute Bioscience | COVID-19 Genomics UK (COG-UK) Consortium |
| EPI_ISL_457341 | Quadram Institute Bioscience | COVID-19 Genomics UK (COG-UK) Consortium |
| EPI_ISL_457342 | Quadram Institute Bioscience | COVID-19 Genomics UK (COG-UK) Consortium |
| EPI_ISL_457343 | Quadram Institute Bioscience | COVID-19 Genomics UK (COG-UK) Consortium |
| EPI_ISL_457344 | Quadram Institute Bioscience | COVID-19 Genomics UK (COG-UK) Consortium |
| EPI_ISL_457345 | Quadram Institute Bioscience | COVID-19 Genomics UK (COG-UK) Consortium |
| EPI_ISL_457346 | Quadram Institute Bioscience | COVID-19 Genomics UK (COG-UK) Consortium |
| EPI_ISL_457347 | Quadram Institute Bioscience | COVID-19 Genomics UK (COG-UK) Consortium |
| EPI_ISL_457348 | Quadram Institute Bioscience | COVID-19 Genomics UK (COG-UK) Consortium |
| EPI_ISL_457349 | Quadram Institute Bioscience | COVID-19 Genomics UK (COG-UK) Consortium |
| EPI_ISL_457350 | Quadram Institute Bioscience | COVID-19 Genomics UK (COG-UK) Consortium |
| EPI_ISL_457351 | Quadram Institute Bioscience | COVID-19 Genomics UK (COG-UK) Consortium |
| EPI_ISL_457352 | Quadram Institute Bioscience | COVID-19 Genomics UK (COG-UK) Consortium |
| EPI_ISL_457353 | Quadram Institute Bioscience | COVID-19 Genomics UK (COG-UK) Consortium |

| EPI_Accession_ID | Originating Laboratory | Submitting Laboratory |
|---|---|---|
| EPI_ISL_457354 | Quadram Institute Bioscience | COVID-19 Genomics UK (COG-UK) Consortium |
| EPI_ISL_457355 | Quadram Institute Bioscience | COVID-19 Genomics UK (COG-UK) Consortium |
| EPI_ISL_457356 | Quadram Institute Bioscience | COVID-19 Genomics UK (COG-UK) Consortium |
| EPI_ISL_457357 | Quadram Institute Bioscience | COVID-19 Genomics UK (COG-UK) Consortium |
| EPI_ISL_457358 | Quadram Institute Bioscience | COVID-19 Genomics UK (COG-UK) Consortium |
| EPI_ISL_457359 | Quadram Institute Bioscience | COVID-19 Genomics UK (COG-UK) Consortium |
| EPI_ISL_457360 | Quadram Institute Bioscience | COVID-19 Genomics UK (COG-UK) Consortium |
| EPI_ISL_457361 | Quadram Institute Bioscience | COVID-19 Genomics UK (COG-UK) Consortium |
| EPI_ISL_457362 | Quadram Institute Bioscience | COVID-19 Genomics UK (COG-UK) Consortium |
| EPI_ISL_457363 | Quadram Institute Bioscience | COVID-19 Genomics UK (COG-UK) Consortium |
| EPI_ISL_457364 | Quadram Institute Bioscience | COVID-19 Genomics UK (COG-UK) Consortium |
| EPI_ISL_457365 | Quadram Institute Bioscience | COVID-19 Genomics UK (COG-UK) Consortium |
| EPI_ISL_457366 | Quadram Institute Bioscience | COVID-19 Genomics UK (COG-UK) Consortium |
| EPI_ISL_457367 | Quadram Institute Bioscience | COVID-19 Genomics UK (COG-UK) Consortium |
| EPI_ISL_457368 | Quadram Institute Bioscience | COVID-19 Genomics UK (COG-UK) Consortium |
| EPI_ISL_457369 | Quadram Institute Bioscience | COVID-19 Genomics UK (COG-UK) Consortium |
| EPI_ISL_457370 | Quadram Institute Bioscience | COVID-19 Genomics UK (COG-UK) Consortium |
| EPI_ISL_457371 | Quadram Institute Bioscience | COVID-19 Genomics UK (COG-UK) Consortium |
| EPI_ISL_457372 | Quadram Institute Bioscience | COVID-19 Genomics UK (COG-UK) Consortium |
| EPI_ISL_457373 | Quadram Institute Bioscience | COVID-19 Genomics UK (COG-UK) Consortium |
| EPI_ISL_457374 | Quadram Institute Bioscience | COVID-19 Genomics UK (COG-UK) Consortium |
| EPI_ISL_457375 | Quadram Institute Bioscience | COVID-19 Genomics UK (COG-UK) Consortium |
| EPI_ISL_457376 | Quadram Institute Bioscience | COVID-19 Genomics UK (COG-UK) Consortium |
| EPI_ISL_457377 | Quadram Institute Bioscience | COVID-19 Genomics UK (COG-UK) Consortium |
| EPI_ISL_457378 | Quadram Institute Bioscience | COVID-19 Genomics UK (COG-UK) Consortium |
| EPI_ISL_457379 | Quadram Institute Bioscience | COVID-19 Genomics UK (COG-UK) Consortium |
| EPI_ISL_457380 | Quadram Institute Bioscience | COVID-19 Genomics UK (COG-UK) Consortium |
| EPI_ISL_457381 | Quadram Institute Bioscience | COVID-19 Genomics UK (COG-UK) Consortium |
| EPI_ISL_457382 | Quadram Institute Bioscience | COVID-19 Genomics UK (COG-UK) Consortium |
| EPI_ISL_457383 | Quadram Institute Bioscience | COVID-19 Genomics UK (COG-UK) Consortium |
| EPI_ISL_457384 | Quadram Institute Bioscience | COVID-19 Genomics UK (COG-UK) Consortium |
| EPI_ISL_457385 | Quadram Institute Bioscience | COVID-19 Genomics UK (COG-UK) Consortium |
| EPI_ISL_457386 | Quadram Institute Bioscience | COVID-19 Genomics UK (COG-UK) Consortium |

| EPI_Accession_ID | Originating Laboratory | Submitting Laboratory |
|---|---|---|
| EPI_ISL_457387 | Quadram Institute Bioscience | COVID-19 Genomics UK (COG-UK) Consortium |
| EPI_ISL_457388 | Quadram Institute Bioscience | COVID-19 Genomics UK (COG-UK) Consortium |
| EPI_ISL_457389 | Quadram Institute Bioscience | COVID-19 Genomics UK (COG-UK) Consortium |
| EPI_ISL_457390 | Quadram Institute Bioscience | COVID-19 Genomics UK (COG-UK) Consortium |
| EPI_ISL_457391 | Quadram Institute Bioscience | COVID-19 Genomics UK (COG-UK) Consortium |
| EPI_ISL_457392 | Quadram Institute Bioscience | COVID-19 Genomics UK (COG-UK) Consortium |
| EPI_ISL_457393 | Quadram Institute Bioscience | COVID-19 Genomics UK (COG-UK) Consortium |
| EPI_ISL_457394 | Quadram Institute Bioscience | COVID-19 Genomics UK (COG-UK) Consortium |
| EPI_ISL_457395 | Quadram Institute Bioscience | COVID-19 Genomics UK (COG-UK) Consortium |
| EPI_ISL_457396 | Quadram Institute Bioscience | COVID-19 Genomics UK (COG-UK) Consortium |
| EPI_ISL_457397 | Quadram Institute Bioscience | COVID-19 Genomics UK (COG-UK) Consortium |
| EPI_ISL_457398 | Quadram Institute Bioscience | COVID-19 Genomics UK (COG-UK) Consortium |
| EPI_ISL_457399 | Quadram Institute Bioscience | COVID-19 Genomics UK (COG-UK) Consortium |
| EPI_ISL_457400 | Quadram Institute Bioscience | COVID-19 Genomics UK (COG-UK) Consortium |
| EPI_ISL_457401 | Quadram Institute Bioscience | COVID-19 Genomics UK (COG-UK) Consortium |
| EPI_ISL_457402 | Quadram Institute Bioscience | COVID-19 Genomics UK (COG-UK) Consortium |
| EPI_ISL_457403 | Quadram Institute Bioscience | COVID-19 Genomics UK (COG-UK) Consortium |
| EPI_ISL_457404 | Quadram Institute Bioscience | COVID-19 Genomics UK (COG-UK) Consortium |
| EPI_ISL_457405 | Quadram Institute Bioscience | COVID-19 Genomics UK (COG-UK) Consortium |
| EPI_ISL_457406 | Quadram Institute Bioscience | COVID-19 Genomics UK (COG-UK) Consortium |
| EPI_ISL_457407 | Quadram Institute Bioscience | COVID-19 Genomics UK (COG-UK) Consortium |
| EPI_ISL_457408 | Quadram Institute Bioscience | COVID-19 Genomics UK (COG-UK) Consortium |
| EPI_ISL_457409 | Quadram Institute Bioscience | COVID-19 Genomics UK (COG-UK) Consortium |
| EPI_ISL_457410 | Quadram Institute Bioscience | COVID-19 Genomics UK (COG-UK) Consortium |
| EPI_ISL_457411 | Quadram Institute Bioscience | COVID-19 Genomics UK (COG-UK) Consortium |
| EPI_ISL_457412 | Quadram Institute Bioscience | COVID-19 Genomics UK (COG-UK) Consortium |
| EPI_ISL_457413 | Quadram Institute Bioscience | COVID-19 Genomics UK (COG-UK) Consortium |
| EPI_ISL_457414 | Quadram Institute Bioscience | COVID-19 Genomics UK (COG-UK) Consortium |
| EPI_ISL_457415 | Quadram Institute Bioscience | COVID-19 Genomics UK (COG-UK) Consortium |
| EPI_ISL_457416 | Quadram Institute Bioscience | COVID-19 Genomics UK (COG-UK) Consortium |
| EPI_ISL_457417 | Quadram Institute Bioscience | COVID-19 Genomics UK (COG-UK) Consortium |
| EPI_ISL_457418 | Quadram Institute Bioscience | COVID-19 Genomics UK (COG-UK) Consortium |
| EPI_ISL_457419 | Quadram Institute Bioscience | COVID-19 Genomics UK (COG-UK) Consortium |

| EPI_Accession_ID | Originating Laboratory | Submitting Laboratory |
|---|---|---|
| EPI_ISL_457420 | Quadram Institute Bioscience | COVID-19 Genomics UK (COG-UK) Consortium |
| EPI_ISL_457421 | Quadram Institute Bioscience | COVID-19 Genomics UK (COG-UK) Consortium |
| EPI_ISL_457422 | Quadram Institute Bioscience | COVID-19 Genomics UK (COG-UK) Consortium |
| EPI_ISL_457423 | Quadram Institute Bioscience | COVID-19 Genomics UK (COG-UK) Consortium |
| EPI_ISL_457424 | Quadram Institute Bioscience | COVID-19 Genomics UK (COG-UK) Consortium |
| EPI_ISL_457425 | Quadram Institute Bioscience | COVID-19 Genomics UK (COG-UK) Consortium |
| EPI_ISL_457426 | Quadram Institute Bioscience | COVID-19 Genomics UK (COG-UK) Consortium |
| EPI_ISL_457427 | Quadram Institute Bioscience | COVID-19 Genomics UK (COG-UK) Consortium |
| EPI_ISL_457428 | Quadram Institute Bioscience | COVID-19 Genomics UK (COG-UK) Consortium |
| EPI_ISL_457429 | Quadram Institute Bioscience | COVID-19 Genomics UK (COG-UK) Consortium |
| EPI_ISL_457430 | Quadram Institute Bioscience | COVID-19 Genomics UK (COG-UK) Consortium |
| EPI_ISL_457431 | Quadram Institute Bioscience | COVID-19 Genomics UK (COG-UK) Consortium |
| EPI_ISL_457432 | Quadram Institute Bioscience | COVID-19 Genomics UK (COG-UK) Consortium |
| EPI_ISL_457433 | Quadram Institute Bioscience | COVID-19 Genomics UK (COG-UK) Consortium |
| EPI_ISL_457434 | Quadram Institute Bioscience | COVID-19 Genomics UK (COG-UK) Consortium |
| EPI_ISL_457435 | Quadram Institute Bioscience | COVID-19 Genomics UK (COG-UK) Consortium |
| EPI_ISL_457436 | Quadram Institute Bioscience | COVID-19 Genomics UK (COG-UK) Consortium |
| EPI_ISL_457437 | Quadram Institute Bioscience | COVID-19 Genomics UK (COG-UK) Consortium |
| EPI_ISL_457438 | Quadram Institute Bioscience | COVID-19 Genomics UK (COG-UK) Consortium |
| EPI_ISL_457439 | Quadram Institute Bioscience | COVID-19 Genomics UK (COG-UK) Consortium |
| EPI_ISL_457440 | Quadram Institute Bioscience | COVID-19 Genomics UK (COG-UK) Consortium |
| EPI_ISL_457441 | Quadram Institute Bioscience | COVID-19 Genomics UK (COG-UK) Consortium |
| EPI_ISL_457442 | Quadram Institute Bioscience | COVID-19 Genomics UK (COG-UK) Consortium |
| EPI_ISL_457443 | Quadram Institute Bioscience | COVID-19 Genomics UK (COG-UK) Consortium |
| EPI_ISL_457444 | Quadram Institute Bioscience | COVID-19 Genomics UK (COG-UK) Consortium |
| EPI_ISL_457445 | Quadram Institute Bioscience | COVID-19 Genomics UK (COG-UK) Consortium |
| EPI_ISL_457446 | Quadram Institute Bioscience | COVID-19 Genomics UK (COG-UK) Consortium |
| EPI_ISL_457447 | Quadram Institute Bioscience | COVID-19 Genomics UK (COG-UK) Consortium |
| EPI_ISL_457448 | Quadram Institute Bioscience | COVID-19 Genomics UK (COG-UK) Consortium |
| EPI_ISL_457449 | Quadram Institute Bioscience | COVID-19 Genomics UK (COG-UK) Consortium |
| EPI_ISL_457450 | Quadram Institute Bioscience | COVID-19 Genomics UK (COG-UK) Consortium |
| EPI_ISL_457451 | Quadram Institute Bioscience | COVID-19 Genomics UK (COG-UK) Consortium |
| EPI_ISL_457452 | Quadram Institute Bioscience | COVID-19 Genomics UK (COG-UK) Consortium |

| EPI_Accession_ID | Originating Laboratory | Submitting Laboratory |
|---|---|---|
| EPI_ISL_457453 | Quadram Institute Bioscience | COVID-19 Genomics UK (COG-UK) Consortium |
| EPI_ISL_457454 | Quadram Institute Bioscience | COVID-19 Genomics UK (COG-UK) Consortium |
| EPI_ISL_457455 | Quadram Institute Bioscience | COVID-19 Genomics UK (COG-UK) Consortium |
| EPI_ISL_457456 | Quadram Institute Bioscience | COVID-19 Genomics UK (COG-UK) Consortium |
| EPI_ISL_457457 | Quadram Institute Bioscience | COVID-19 Genomics UK (COG-UK) Consortium |
| EPI_ISL_457458 | Quadram Institute Bioscience | COVID-19 Genomics UK (COG-UK) Consortium |
| EPI_ISL_457459 | Quadram Institute Bioscience | COVID-19 Genomics UK (COG-UK) Consortium |
| EPI_ISL_457460 | Quadram Institute Bioscience | COVID-19 Genomics UK (COG-UK) Consortium |
| EPI_ISL_457461 | Quadram Institute Bioscience | COVID-19 Genomics UK (COG-UK) Consortium |
| EPI_ISL_457462 | Quadram Institute Bioscience | COVID-19 Genomics UK (COG-UK) Consortium |
| EPI_ISL_457463 | Quadram Institute Bioscience | COVID-19 Genomics UK (COG-UK) Consortium |
| EPI_ISL_457464 | Quadram Institute Bioscience | COVID-19 Genomics UK (COG-UK) Consortium |
| EPI_ISL_457465 | Quadram Institute Bioscience | COVID-19 Genomics UK (COG-UK) Consortium |
| EPI_ISL_457466 | Quadram Institute Bioscience | COVID-19 Genomics UK (COG-UK) Consortium |
| EPI_ISL_457467 | Quadram Institute Bioscience | COVID-19 Genomics UK (COG-UK) Consortium |
| EPI_ISL_457468 | Quadram Institute Bioscience | COVID-19 Genomics UK (COG-UK) Consortium |
| EPI_ISL_457469 | Quadram Institute Bioscience | COVID-19 Genomics UK (COG-UK) Consortium |
| EPI_ISL_457470 | Quadram Institute Bioscience | COVID-19 Genomics UK (COG-UK) Consortium |
| EPI_ISL_457471 | Quadram Institute Bioscience | COVID-19 Genomics UK (COG-UK) Consortium |
| EPI_ISL_457472 | Quadram Institute Bioscience | COVID-19 Genomics UK (COG-UK) Consortium |
| EPI_ISL_457473 | Quadram Institute Bioscience | COVID-19 Genomics UK (COG-UK) Consortium |
| EPI_ISL_457474 | Quadram Institute Bioscience | COVID-19 Genomics UK (COG-UK) Consortium |
| EPI_ISL_457475 | Quadram Institute Bioscience | COVID-19 Genomics UK (COG-UK) Consortium |
| EPI_ISL_457476 | Quadram Institute Bioscience | COVID-19 Genomics UK (COG-UK) Consortium |
| EPI_ISL_457477 | Quadram Institute Bioscience | COVID-19 Genomics UK (COG-UK) Consortium |
| EPI_ISL_457478 | Quadram Institute Bioscience | COVID-19 Genomics UK (COG-UK) Consortium |
| EPI_ISL_457479 | Quadram Institute Bioscience | COVID-19 Genomics UK (COG-UK) Consortium |
| EPI_ISL_457480 | Quadram Institute Bioscience | COVID-19 Genomics UK (COG-UK) Consortium |
| EPI_ISL_457481 | Quadram Institute Bioscience | COVID-19 Genomics UK (COG-UK) Consortium |
| EPI_ISL_457482 | Quadram Institute Bioscience | COVID-19 Genomics UK (COG-UK) Consortium |
| EPI_ISL_457483 | Quadram Institute Bioscience | COVID-19 Genomics UK (COG-UK) Consortium |
| EPI_ISL_457484 | Quadram Institute Bioscience | COVID-19 Genomics UK (COG-UK) Consortium |
| EPI_ISL_457485 | Quadram Institute Bioscience | COVID-19 Genomics UK (COG-UK) Consortium |

| EPI_Accession_ID | Originating Laboratory | Submitting Laboratory |
|---|---|---|
| EPI_ISL_457486 | Quadram Institute Bioscience | COVID-19 Genomics UK (COG-UK) Consortium |
| EPI_ISL_457487 | Quadram Institute Bioscience | COVID-19 Genomics UK (COG-UK) Consortium |
| EPI_ISL_457488 | Quadram Institute Bioscience | COVID-19 Genomics UK (COG-UK) Consortium |
| EPI_ISL_457489 | Quadram Institute Bioscience | COVID-19 Genomics UK (COG-UK) Consortium |
| EPI_ISL_457490 | Quadram Institute Bioscience | COVID-19 Genomics UK (COG-UK) Consortium |
| EPI_ISL_457491 | Quadram Institute Bioscience | COVID-19 Genomics UK (COG-UK) Consortium |
| EPI_ISL_457492 | Quadram Institute Bioscience | COVID-19 Genomics UK (COG-UK) Consortium |
| EPI_ISL_457493 | Quadram Institute Bioscience | COVID-19 Genomics UK (COG-UK) Consortium |
| EPI_ISL_457494 | Quadram Institute Bioscience | COVID-19 Genomics UK (COG-UK) Consortium |
| EPI_ISL_457495 | Quadram Institute Bioscience | COVID-19 Genomics UK (COG-UK) Consortium |
| EPI_ISL_457496 | Quadram Institute Bioscience | COVID-19 Genomics UK (COG-UK) Consortium |
| EPI_ISL_457497 | Quadram Institute Bioscience | COVID-19 Genomics UK (COG-UK) Consortium |
| EPI_ISL_457498 | Quadram Institute Bioscience | COVID-19 Genomics UK (COG-UK) Consortium |
| EPI_ISL_457499 | Quadram Institute Bioscience | COVID-19 Genomics UK (COG-UK) Consortium |
| EPI_ISL_457500 | Quadram Institute Bioscience | COVID-19 Genomics UK (COG-UK) Consortium |
| EPI_ISL_457501 | Quadram Institute Bioscience | COVID-19 Genomics UK (COG-UK) Consortium |
| EPI_ISL_457502 | Quadram Institute Bioscience | COVID-19 Genomics UK (COG-UK) Consortium |
| EPI_ISL_457503 | Quadram Institute Bioscience | COVID-19 Genomics UK (COG-UK) Consortium |
| EPI_ISL_457504 | Quadram Institute Bioscience | COVID-19 Genomics UK (COG-UK) Consortium |
| EPI_ISL_457505 | Quadram Institute Bioscience | COVID-19 Genomics UK (COG-UK) Consortium |
| EPI_ISL_457506 | Quadram Institute Bioscience | COVID-19 Genomics UK (COG-UK) Consortium |
| EPI_ISL_457507 | Quadram Institute Bioscience | COVID-19 Genomics UK (COG-UK) Consortium |
| EPI_ISL_457508 | Quadram Institute Bioscience | COVID-19 Genomics UK (COG-UK) Consortium |
| EPI_ISL_457509 | Quadram Institute Bioscience | COVID-19 Genomics UK (COG-UK) Consortium |
| EPI_ISL_457510 | Quadram Institute Bioscience | COVID-19 Genomics UK (COG-UK) Consortium |
| EPI_ISL_457511 | Quadram Institute Bioscience | COVID-19 Genomics UK (COG-UK) Consortium |
| EPI_ISL_457512 | Quadram Institute Bioscience | COVID-19 Genomics UK (COG-UK) Consortium |
| EPI_ISL_457513 | Quadram Institute Bioscience | COVID-19 Genomics UK (COG-UK) Consortium |
| EPI_ISL_457514 | Quadram Institute Bioscience | COVID-19 Genomics UK (COG-UK) Consortium |
| EPI_ISL_457515 | Quadram Institute Bioscience | COVID-19 Genomics UK (COG-UK) Consortium |
| EPI_ISL_457516 | Quadram Institute Bioscience | COVID-19 Genomics UK (COG-UK) Consortium |
| EPI_ISL_457517 | Quadram Institute Bioscience | COVID-19 Genomics UK (COG-UK) Consortium |
| EPI_ISL_457518 | Quadram Institute Bioscience | COVID-19 Genomics UK (COG-UK) Consortium |

| EPI_Accession_ID | Originating Laboratory | Submitting Laboratory |
|---|---|---|
| EPI_ISL_457519 | Quadram Institute Bioscience | COVID-19 Genomics UK (COG-UK) Consortium |
| EPI_ISL_457520 | Quadram Institute Bioscience | COVID-19 Genomics UK (COG-UK) Consortium |
| EPI_ISL_457521 | Quadram Institute Bioscience | COVID-19 Genomics UK (COG-UK) Consortium |
| EPI_ISL_457522 | Quadram Institute Bioscience | COVID-19 Genomics UK (COG-UK) Consortium |
| EPI_ISL_457523 | Quadram Institute Bioscience | COVID-19 Genomics UK (COG-UK) Consortium |
| EPI_ISL_457524 | Quadram Institute Bioscience | COVID-19 Genomics UK (COG-UK) Consortium |
| EPI_ISL_457525 | Quadram Institute Bioscience | COVID-19 Genomics UK (COG-UK) Consortium |
| EPI_ISL_457526 | Quadram Institute Bioscience | COVID-19 Genomics UK (COG-UK) Consortium |
| EPI_ISL_457527 | Quadram Institute Bioscience | COVID-19 Genomics UK (COG-UK) Consortium |
| EPI_ISL_457528 | Quadram Institute Bioscience | COVID-19 Genomics UK (COG-UK) Consortium |
| EPI_ISL_457529 | Quadram Institute Bioscience | COVID-19 Genomics UK (COG-UK) Consortium |
| EPI_ISL_457530 | Quadram Institute Bioscience | COVID-19 Genomics UK (COG-UK) Consortium |
| EPI_ISL_457531 | Quadram Institute Bioscience | COVID-19 Genomics UK (COG-UK) Consortium |
| EPI_ISL_457532 | Quadram Institute Bioscience | COVID-19 Genomics UK (COG-UK) Consortium |
| EPI_ISL_457533 | Quadram Institute Bioscience | COVID-19 Genomics UK (COG-UK) Consortium |
| EPI_ISL_457534 | Quadram Institute Bioscience | COVID-19 Genomics UK (COG-UK) Consortium |
| EPI_ISL_457535 | Quadram Institute Bioscience | COVID-19 Genomics UK (COG-UK) Consortium |
| EPI_ISL_457536 | Quadram Institute Bioscience | COVID-19 Genomics UK (COG-UK) Consortium |
| EPI_ISL_457537 | Quadram Institute Bioscience | COVID-19 Genomics UK (COG-UK) Consortium |
| EPI_ISL_457538 | Quadram Institute Bioscience | COVID-19 Genomics UK (COG-UK) Consortium |
| EPI_ISL_457539 | Quadram Institute Bioscience | COVID-19 Genomics UK (COG-UK) Consortium |
| EPI_ISL_457540 | Quadram Institute Bioscience | COVID-19 Genomics UK (COG-UK) Consortium |
| EPI_ISL_457541 | Quadram Institute Bioscience | COVID-19 Genomics UK (COG-UK) Consortium |
| EPI_ISL_457542 | Quadram Institute Bioscience | COVID-19 Genomics UK (COG-UK) Consortium |
| EPI_ISL_457543 | Quadram Institute Bioscience | COVID-19 Genomics UK (COG-UK) Consortium |
| EPI_ISL_457544 | Quadram Institute Bioscience | COVID-19 Genomics UK (COG-UK) Consortium |
| EPI_ISL_457545 | Quadram Institute Bioscience | COVID-19 Genomics UK (COG-UK) Consortium |
| EPI_ISL_457546 | Quadram Institute Bioscience | COVID-19 Genomics UK (COG-UK) Consortium |
| EPI_ISL_457547 | Quadram Institute Bioscience | COVID-19 Genomics UK (COG-UK) Consortium |
| EPI_ISL_457548 | Quadram Institute Bioscience | COVID-19 Genomics UK (COG-UK) Consortium |
| EPI_ISL_457549 | Quadram Institute Bioscience | COVID-19 Genomics UK (COG-UK) Consortium |
| EPI_ISL_457550 | Quadram Institute Bioscience | COVID-19 Genomics UK (COG-UK) Consortium |
| EPI_ISL_457551 | Quadram Institute Bioscience | COVID-19 Genomics UK (COG-UK) Consortium |

| EPI_Accession_ID | Originating Laboratory | Submitting Laboratory |
|---|---|---|
| EPI_ISL_457552 | Quadram Institute Bioscience | COVID-19 Genomics UK (COG-UK) Consortium |
| EPI_ISL_457553 | Quadram Institute Bioscience | COVID-19 Genomics UK (COG-UK) Consortium |
| EPI_ISL_457554 | Quadram Institute Bioscience | COVID-19 Genomics UK (COG-UK) Consortium |
| EPI_ISL_457555 | Quadram Institute Bioscience | COVID-19 Genomics UK (COG-UK) Consortium |
| EPI_ISL_457556 | Quadram Institute Bioscience | COVID-19 Genomics UK (COG-UK) Consortium |
| EPI_ISL_457557 | Quadram Institute Bioscience | COVID-19 Genomics UK (COG-UK) Consortium |
| EPI_ISL_457558 | Quadram Institute Bioscience | COVID-19 Genomics UK (COG-UK) Consortium |
| EPI_ISL_457559 | Quadram Institute Bioscience | COVID-19 Genomics UK (COG-UK) Consortium |
| EPI_ISL_457560 | Quadram Institute Bioscience | COVID-19 Genomics UK (COG-UK) Consortium |
| EPI_ISL_457561 | Quadram Institute Bioscience | COVID-19 Genomics UK (COG-UK) Consortium |
| EPI_ISL_457562 | Quadram Institute Bioscience | COVID-19 Genomics UK (COG-UK) Consortium |
| EPI_ISL_457563 | Quadram Institute Bioscience | COVID-19 Genomics UK (COG-UK) Consortium |
| EPI_ISL_457564 | Quadram Institute Bioscience | COVID-19 Genomics UK (COG-UK) Consortium |
| EPI_ISL_457565 | Quadram Institute Bioscience | COVID-19 Genomics UK (COG-UK) Consortium |
| EPI_ISL_457566 | Quadram Institute Bioscience | COVID-19 Genomics UK (COG-UK) Consortium |
| EPI_ISL_457567 | Quadram Institute Bioscience | COVID-19 Genomics UK (COG-UK) Consortium |
| EPI_ISL_457568 | Quadram Institute Bioscience | COVID-19 Genomics UK (COG-UK) Consortium |
| EPI_ISL_457569 | Quadram Institute Bioscience | COVID-19 Genomics UK (COG-UK) Consortium |
| EPI_ISL_457570 | Quadram Institute Bioscience | COVID-19 Genomics UK (COG-UK) Consortium |
| EPI_ISL_457571 | Quadram Institute Bioscience | COVID-19 Genomics UK (COG-UK) Consortium |
| EPI_ISL_457572 | Quadram Institute Bioscience | COVID-19 Genomics UK (COG-UK) Consortium |
| EPI_ISL_457573 | Queens Medical Centre, Clinical Microbiology Department / DeepSeq Nottingham | COVID-19 Genomics UK (COG-UK) Consortium |
| EPI_ISL_457574 | Queens Medical Centre, Clinical Microbiology Department / DeepSeq Nottingham | COVID-19 Genomics UK (COG-UK) Consortium |
| EPI_ISL_457575 | Queens Medical Centre, Clinical Microbiology Department / DeepSeq Nottingham | COVID-19 Genomics UK (COG-UK) Consortium |
| EPI_ISL_457576 | Queens Medical Centre, Clinical Microbiology Department / DeepSeq Nottingham | COVID-19 Genomics UK (COG-UK) Consortium |
| EPI_ISL_457577 | Queens Medical Centre, Clinical Microbiology Department / DeepSeq Nottingham | COVID-19 Genomics UK (COG-UK) Consortium |
| EPI_ISL_457578 | Queens Medical Centre, Clinical Microbiology Department / DeepSeq Nottingham | COVID-19 Genomics UK (COG-UK) Consortium |

| EPI_Accession_ID | Originating Laboratory | Submitting Laboratory |
| --- | --- | --- |
| EPI_ISL_457579 | Queens Medical Centre, Clinical Microbiology Department / DeepSeq Nottingham | COVID-19 Genomics UK (COG-UK) Consortium |
| EPI_ISL_457580 | Queens Medical Centre, Clinical Microbiology Department / DeepSeq Nottingham | COVID-19 Genomics UK (COG-UK) Consortium |
| EPI_ISL_457581 | Virology Department, Sheffield Teaching Hospitals NHS Foundation Trust/Department of Infection, Immunity and Cardiovascular Disease, The Medical School, University of Sheffield | COVID-19 Genomics UK (COG-UK) Consortium |
| EPI_ISL_457582 | Virology Department, Sheffield Teaching Hospitals NHS Foundation Trust/Department of Infection, Immunity and Cardiovascular Disease, The Medical School, University of Sheffield | COVID-19 Genomics UK (COG-UK) Consortium |
| EPI_ISL_457583 | Virology Department, Sheffield Teaching Hospitals NHS Foundation Trust/Department of Infection, Immunity and Cardiovascular Disease, The Medical School, University of Sheffield | COVID-19 Genomics UK (COG-UK) Consortium |
| EPI_ISL_457584 | Virology Department, Sheffield Teaching Hospitals NHS Foundation Trust/Department of Infection, Immunity and Cardiovascular Disease, The Medical School, University of Sheffield | COVID-19 Genomics UK (COG-UK) Consortium |
| EPI_ISL_457585 | Virology Department, Sheffield Teaching Hospitals NHS Foundation Trust/Department of Infection, Immunity and Cardiovascular Disease, The Medical School, University of Sheffield | COVID-19 Genomics UK (COG-UK) Consortium |
| EPI_ISL_457586 | Virology Department, Sheffield Teaching Hospitals NHS Foundation Trust/Department of Infection, Immunity and Cardiovascular Disease, The Medical School, University of Sheffield | COVID-19 Genomics UK (COG-UK) Consortium |
| EPI_ISL_457587 | Virology Department, Sheffield Teaching Hospitals NHS Foundation Trust/Department of Infection, Immunity and Cardiovascular Disease, The Medical School, University of Sheffield | COVID-19 Genomics UK (COG-UK) Consortium |

| EPI_Accession_ID | Originating Laboratory | Submitting Laboratory |
|---|---|---|
| EPI_ISL_457588 | Virology Department, Sheffield Teaching Hospitals NHS Foundation Trust/Department of Infection, Immunity and Cardiovascular Disease, The Medical School, University of Sheffield | COVID-19 Genomics UK (COG-UK) Consortium |
| EPI_ISL_457589 | Virology Department, Sheffield Teaching Hospitals NHS Foundation Trust/Department of Infection, Immunity and Cardiovascular Disease, The Medical School, University of Sheffield | COVID-19 Genomics UK (COG-UK) Consortium |
| EPI_ISL_457590 | Virology Department, Sheffield Teaching Hospitals NHS Foundation Trust/Department of Infection, Immunity and Cardiovascular Disease, The Medical School, University of Sheffield | COVID-19 Genomics UK (COG-UK) Consortium |
| EPI_ISL_457591 | Virology Department, Sheffield Teaching Hospitals NHS Foundation Trust/Department of Infection, Immunity and Cardiovascular Disease, The Medical School, University of Sheffield | COVID-19 Genomics UK (COG-UK) Consortium |
| EPI_ISL_457592 | Virology Department, Sheffield Teaching Hospitals NHS Foundation Trust/Department of Infection, Immunity and Cardiovascular Disease, The Medical School, University of Sheffield | COVID-19 Genomics UK (COG-UK) Consortium |
| EPI_ISL_457593 | Virology Department, Sheffield Teaching Hospitals NHS Foundation Trust/Department of Infection, Immunity and Cardiovascular Disease, The Medical School, University of Sheffield | COVID-19 Genomics UK (COG-UK) Consortium |
| EPI_ISL_457594 | Virology Department, Sheffield Teaching Hospitals NHS Foundation Trust/Department of Infection, Immunity and Cardiovascular Disease, The Medical School, University of Sheffield | COVID-19 Genomics UK (COG-UK) Consortium |
| EPI_ISL_457595 | Virology Department, Sheffield Teaching Hospitals NHS Foundation Trust/Department of Infection, Immunity and Cardiovascular Disease, The Medical School, University of Sheffield | COVID-19 Genomics UK (COG-UK) Consortium |

| EPI_Accession_ID | Originating Laboratory | Submitting Laboratory |
| --- | --- | --- |
| EPI_ISL_457596 | Virology Department, Sheffield Teaching Hospitals NHS Foundation Trust/Department of Infection, Immunity and Cardiovascular Disease, The Medical School, University of Sheffield | COVID-19 Genomics UK (COG-UK) Consortium |
| EPI_ISL_457597 | Virology Department, Sheffield Teaching Hospitals NHS Foundation Trust/Department of Infection, Immunity and Cardiovascular Disease, The Medical School, University of Sheffield | COVID-19 Genomics UK (COG-UK) Consortium |
| EPI_ISL_457598 | Virology Department, Sheffield Teaching Hospitals NHS Foundation Trust/Department of Infection, Immunity and Cardiovascular Disease, The Medical School, University of Sheffield | COVID-19 Genomics UK (COG-UK) Consortium |
| EPI_ISL_457599 | Virology Department, Sheffield Teaching Hospitals NHS Foundation Trust/Department of Infection, Immunity and Cardiovascular Disease, The Medical School, University of Sheffield | COVID-19 Genomics UK (COG-UK) Consortium |
| EPI_ISL_457600 | Virology Department, Sheffield Teaching Hospitals NHS Foundation Trust/Department of Infection, Immunity and Cardiovascular Disease, The Medical School, University of Sheffield | COVID-19 Genomics UK (COG-UK) Consortium |
| EPI_ISL_457601 | Virology Department, Sheffield Teaching Hospitals NHS Foundation Trust/Department of Infection, Immunity and Cardiovascular Disease, The Medical School, University of Sheffield | COVID-19 Genomics UK (COG-UK) Consortium |
| EPI_ISL_457602 | Virology Department, Sheffield Teaching Hospitals NHS Foundation Trust/Department of Infection, Immunity and Cardiovascular Disease, The Medical School, University of Sheffield | COVID-19 Genomics UK (COG-UK) Consortium |
| EPI_ISL_457603 | Virology Department, Sheffield Teaching Hospitals NHS Foundation Trust/Department of Infection, Immunity and Cardiovascular Disease, The Medical School, University of Sheffield | COVID-19 Genomics UK (COG-UK) Consortium |

| EPI_Accession_ID | Originating Laboratory | Submitting Laboratory |
|---|---|---|
| EPI_ISL_457604 | Virology Department, Sheffield Teaching Hospitals NHS Foundation Trust/Department of Infection, Immunity and Cardiovascular Disease, The Medical School, University of Sheffield | COVID-19 Genomics UK (COG-UK) Consortium |
| EPI_ISL_457605 | Virology Department, Sheffield Teaching Hospitals NHS Foundation Trust/Department of Infection, Immunity and Cardiovascular Disease, The Medical School, University of Sheffield | COVID-19 Genomics UK (COG-UK) Consortium |
| EPI_ISL_457606 | Virology Department, Sheffield Teaching Hospitals NHS Foundation Trust/Department of Infection, Immunity and Cardiovascular Disease, The Medical School, University of Sheffield | COVID-19 Genomics UK (COG-UK) Consortium |
| EPI_ISL_457607 | Virology Department, Sheffield Teaching Hospitals NHS Foundation Trust/Department of Infection, Immunity and Cardiovascular Disease, The Medical School, University of Sheffield | COVID-19 Genomics UK (COG-UK) Consortium |
| EPI_ISL_457608 | Virology Department, Sheffield Teaching Hospitals NHS Foundation Trust/Department of Infection, Immunity and Cardiovascular Disease, The Medical School, University of Sheffield | COVID-19 Genomics UK (COG-UK) Consortium |
| EPI_ISL_457609 | Virology Department, Sheffield Teaching Hospitals NHS Foundation Trust/Department of Infection, Immunity and Cardiovascular Disease, The Medical School, University of Sheffield | COVID-19 Genomics UK (COG-UK) Consortium |
| EPI_ISL_457610 | Virology Department, Sheffield Teaching Hospitals NHS Foundation Trust/Department of Infection, Immunity and Cardiovascular Disease, The Medical School, University of Sheffield | COVID-19 Genomics UK (COG-UK) Consortium |
| EPI_ISL_457611 | Virology Department, Sheffield Teaching Hospitals NHS Foundation Trust/Department of Infection, Immunity and Cardiovascular Disease, The Medical School, University of Sheffield | COVID-19 Genomics UK (COG-UK) Consortium |

| EPI_Accession_ID | Originating Laboratory | Submitting Laboratory |
|---|---|---|
| EPI_ISL_457612 | Virology Department, Sheffield Teaching Hospitals NHS Foundation Trust/Department of Infection, Immunity and Cardiovascular Disease, The Medical School, University of Sheffield | COVID-19 Genomics UK (COG-UK) Consortium |
| EPI_ISL_457613 | Virology Department, Sheffield Teaching Hospitals NHS Foundation Trust/Department of Infection, Immunity and Cardiovascular Disease, The Medical School, University of Sheffield | COVID-19 Genomics UK (COG-UK) Consortium |
| EPI_ISL_457614 | Virology Department, Sheffield Teaching Hospitals NHS Foundation Trust/Department of Infection, Immunity and Cardiovascular Disease, The Medical School, University of Sheffield | COVID-19 Genomics UK (COG-UK) Consortium |
| EPI_ISL_457615 | Virology Department, Sheffield Teaching Hospitals NHS Foundation Trust/Department of Infection, Immunity and Cardiovascular Disease, The Medical School, University of Sheffield | COVID-19 Genomics UK (COG-UK) Consortium |
| EPI_ISL_457616 | Virology Department, Sheffield Teaching Hospitals NHS Foundation Trust/Department of Infection, Immunity and Cardiovascular Disease, The Medical School, University of Sheffield | COVID-19 Genomics UK (COG-UK) Consortium |
| EPI_ISL_457617 | Virology Department, Sheffield Teaching Hospitals NHS Foundation Trust/Department of Infection, Immunity and Cardiovascular Disease, The Medical School, University of Sheffield | COVID-19 Genomics UK (COG-UK) Consortium |
| EPI_ISL_457618 | Virology Department, Sheffield Teaching Hospitals NHS Foundation Trust/Department of Infection, Immunity and Cardiovascular Disease, The Medical School, University of Sheffield | COVID-19 Genomics UK (COG-UK) Consortium |
| EPI_ISL_457619 | Virology Department, Sheffield Teaching Hospitals NHS Foundation Trust/Department of Infection, Immunity and Cardiovascular Disease, The Medical School, University of Sheffield | COVID-19 Genomics UK (COG-UK) Consortium |

| EPI_Accession_ID | Originating Laboratory | Submitting Laboratory |
|---|---|---|
| EPI_ISL_457620 | Virology Department, Sheffield Teaching Hospitals NHS Foundation Trust/Department of Infection, Immunity and Cardiovascular Disease, The Medical School, University of Sheffield | COVID-19 Genomics UK (COG-UK) Consortium |
| EPI_ISL_457621 | Virology Department, Sheffield Teaching Hospitals NHS Foundation Trust/Department of Infection, Immunity and Cardiovascular Disease, The Medical School, University of Sheffield | COVID-19 Genomics UK (COG-UK) Consortium |
| EPI_ISL_457622 | Virology Department, Sheffield Teaching Hospitals NHS Foundation Trust/Department of Infection, Immunity and Cardiovascular Disease, The Medical School, University of Sheffield | COVID-19 Genomics UK (COG-UK) Consortium |
| EPI_ISL_457623 | Virology Department, Sheffield Teaching Hospitals NHS Foundation Trust/Department of Infection, Immunity and Cardiovascular Disease, The Medical School, University of Sheffield | COVID-19 Genomics UK (COG-UK) Consortium |
| EPI_ISL_457624 | Virology Department, Sheffield Teaching Hospitals NHS Foundation Trust/Department of Infection, Immunity and Cardiovascular Disease, The Medical School, University of Sheffield | COVID-19 Genomics UK (COG-UK) Consortium |
| EPI_ISL_457625 | Virology Department, Sheffield Teaching Hospitals NHS Foundation Trust/Department of Infection, Immunity and Cardiovascular Disease, The Medical School, University of Sheffield | COVID-19 Genomics UK (COG-UK) Consortium |
| EPI_ISL_457626 | Virology Department, Sheffield Teaching Hospitals NHS Foundation Trust/Department of Infection, Immunity and Cardiovascular Disease, The Medical School, University of Sheffield | COVID-19 Genomics UK (COG-UK) Consortium |
| EPI_ISL_457627 | Virology Department, Sheffield Teaching Hospitals NHS Foundation Trust/Department of Infection, Immunity and Cardiovascular Disease, The Medical School, University of Sheffield | COVID-19 Genomics UK (COG-UK) Consortium |

| EPI_Accession_ID | Originating Laboratory | Submitting Laboratory |
|---|---|---|
| EPI_ISL_457628 | Virology Department, Sheffield Teaching Hospitals NHS Foundation Trust/Department of Infection, Immunity and Cardiovascular Disease, The Medical School, University of Sheffield | COVID-19 Genomics UK (COG-UK) Consortium |
| EPI_ISL_457629 | Virology Department, Sheffield Teaching Hospitals NHS Foundation Trust/Department of Infection, Immunity and Cardiovascular Disease, The Medical School, University of Sheffield | COVID-19 Genomics UK (COG-UK) Consortium |
| EPI_ISL_457630 | Virology Department, Sheffield Teaching Hospitals NHS Foundation Trust/Department of Infection, Immunity and Cardiovascular Disease, The Medical School, University of Sheffield | COVID-19 Genomics UK (COG-UK) Consortium |
| EPI_ISL_457631 | Virology Department, Sheffield Teaching Hospitals NHS Foundation Trust/Department of Infection, Immunity and Cardiovascular Disease, The Medical School, University of Sheffield | COVID-19 Genomics UK (COG-UK) Consortium |
| EPI_ISL_457632 | Virology Department, Sheffield Teaching Hospitals NHS Foundation Trust/Department of Infection, Immunity and Cardiovascular Disease, The Medical School, University of Sheffield | COVID-19 Genomics UK (COG-UK) Consortium |
| EPI_ISL_457633 | Virology Department, Sheffield Teaching Hospitals NHS Foundation Trust/Department of Infection, Immunity and Cardiovascular Disease, The Medical School, University of Sheffield | COVID-19 Genomics UK (COG-UK) Consortium |
| EPI_ISL_457634 | Virology Department, Sheffield Teaching Hospitals NHS Foundation Trust/Department of Infection, Immunity and Cardiovascular Disease, The Medical School, University of Sheffield | COVID-19 Genomics UK (COG-UK) Consortium |
| EPI_ISL_457635 | Virology Department, Sheffield Teaching Hospitals NHS Foundation Trust/Department of Infection, Immunity and Cardiovascular Disease, The Medical School, University of Sheffield | COVID-19 Genomics UK (COG-UK) Consortium |

| EPI_Accession_ID | Originating Laboratory | Submitting Laboratory |
|---|---|---|
| EPI_ISL_457636 | Virology Department, Sheffield Teaching Hospitals NHS Foundation Trust/Department of Infection, Immunity and Cardiovascular Disease, The Medical School, University of Sheffield | COVID-19 Genomics UK (COG-UK) Consortium |
| EPI_ISL_457637 | Virology Department, Sheffield Teaching Hospitals NHS Foundation Trust/Department of Infection, Immunity and Cardiovascular Disease, The Medical School, University of Sheffield | COVID-19 Genomics UK (COG-UK) Consortium |
| EPI_ISL_457638 | Virology Department, Sheffield Teaching Hospitals NHS Foundation Trust/Department of Infection, Immunity and Cardiovascular Disease, The Medical School, University of Sheffield | COVID-19 Genomics UK (COG-UK) Consortium |
| EPI_ISL_457639 | Virology Department, Sheffield Teaching Hospitals NHS Foundation Trust/Department of Infection, Immunity and Cardiovascular Disease, The Medical School, University of Sheffield | COVID-19 Genomics UK (COG-UK) Consortium |
| EPI_ISL_457640 | Virology Department, Sheffield Teaching Hospitals NHS Foundation Trust/Department of Infection, Immunity and Cardiovascular Disease, The Medical School, University of Sheffield | COVID-19 Genomics UK (COG-UK) Consortium |
| EPI_ISL_457641 | Virology Department, Sheffield Teaching Hospitals NHS Foundation Trust/Department of Infection, Immunity and Cardiovascular Disease, The Medical School, University of Sheffield | COVID-19 Genomics UK (COG-UK) Consortium |
| EPI_ISL_457642 | Virology Department, Sheffield Teaching Hospitals NHS Foundation Trust/Department of Infection, Immunity and Cardiovascular Disease, The Medical School, University of Sheffield | COVID-19 Genomics UK (COG-UK) Consortium |
| EPI_ISL_457643 | Virology Department, Sheffield Teaching Hospitals NHS Foundation Trust/Department of Infection, Immunity and Cardiovascular Disease, The Medical School, University of Sheffield | COVID-19 Genomics UK (COG-UK) Consortium |

| EPI_Accession_ID | Originating Laboratory | Submitting Laboratory |
|---|---|---|
| EPI_ISL_457644 | Virology Department, Sheffield Teaching Hospitals NHS Foundation Trust/Department of Infection, Immunity and Cardiovascular Disease, The Medical School, University of Sheffield | COVID-19 Genomics UK (COG-UK) Consortium |
| EPI_ISL_457645 | Virology Department, Sheffield Teaching Hospitals NHS Foundation Trust/Department of Infection, Immunity and Cardiovascular Disease, The Medical School, University of Sheffield | COVID-19 Genomics UK (COG-UK) Consortium |
| EPI_ISL_457646 | Virology Department, Sheffield Teaching Hospitals NHS Foundation Trust/Department of Infection, Immunity and Cardiovascular Disease, The Medical School, University of Sheffield | COVID-19 Genomics UK (COG-UK) Consortium |
| EPI_ISL_457647 | Virology Department, Sheffield Teaching Hospitals NHS Foundation Trust/Department of Infection, Immunity and Cardiovascular Disease, The Medical School, University of Sheffield | COVID-19 Genomics UK (COG-UK) Consortium |
| EPI_ISL_457648 | Virology Department, Sheffield Teaching Hospitals NHS Foundation Trust/Department of Infection, Immunity and Cardiovascular Disease, The Medical School, University of Sheffield | COVID-19 Genomics UK (COG-UK) Consortium |
| EPI_ISL_457649 | Virology Department, Sheffield Teaching Hospitals NHS Foundation Trust/Department of Infection, Immunity and Cardiovascular Disease, The Medical School, University of Sheffield | COVID-19 Genomics UK (COG-UK) Consortium |
| EPI_ISL_457650 | Virology Department, Sheffield Teaching Hospitals NHS Foundation Trust/Department of Infection, Immunity and Cardiovascular Disease, The Medical School, University of Sheffield | COVID-19 Genomics UK (COG-UK) Consortium |
| EPI_ISL_457651 | Virology Department, Sheffield Teaching Hospitals NHS Foundation Trust/Department of Infection, Immunity and Cardiovascular Disease, The Medical School, University of Sheffield | COVID-19 Genomics UK (COG-UK) Consortium |

| EPI_Accession_ID | Originating Laboratory | Submitting Laboratory |
|---|---|---|
| EPI_ISL_457652 | Virology Department, Sheffield Teaching Hospitals NHS Foundation Trust/Department of Infection, Immunity and Cardiovascular Disease, The Medical School, University of Sheffield | COVID-19 Genomics UK (COG-UK) Consortium |
| EPI_ISL_457653 | Virology Department, Sheffield Teaching Hospitals NHS Foundation Trust/Department of Infection, Immunity and Cardiovascular Disease, The Medical School, University of Sheffield | COVID-19 Genomics UK (COG-UK) Consortium |
| EPI_ISL_457654 | Virology Department, Sheffield Teaching Hospitals NHS Foundation Trust/Department of Infection, Immunity and Cardiovascular Disease, The Medical School, University of Sheffield | COVID-19 Genomics UK (COG-UK) Consortium |
| EPI_ISL_457655 | Virology Department, Sheffield Teaching Hospitals NHS Foundation Trust/Department of Infection, Immunity and Cardiovascular Disease, The Medical School, University of Sheffield | COVID-19 Genomics UK (COG-UK) Consortium |
| EPI_ISL_457656 | Virology Department, Sheffield Teaching Hospitals NHS Foundation Trust/Department of Infection, Immunity and Cardiovascular Disease, The Medical School, University of Sheffield | COVID-19 Genomics UK (COG-UK) Consortium |
| EPI_ISL_457657 | Virology Department, Sheffield Teaching Hospitals NHS Foundation Trust/Department of Infection, Immunity and Cardiovascular Disease, The Medical School, University of Sheffield | COVID-19 Genomics UK (COG-UK) Consortium |
| EPI_ISL_457658 | Virology Department, Sheffield Teaching Hospitals NHS Foundation Trust/Department of Infection, Immunity and Cardiovascular Disease, The Medical School, University of Sheffield | COVID-19 Genomics UK (COG-UK) Consortium |
| EPI_ISL_457659 | Virology Department, Sheffield Teaching Hospitals NHS Foundation Trust/Department of Infection, Immunity and Cardiovascular Disease, The Medical School, University of Sheffield | COVID-19 Genomics UK (COG-UK) Consortium |

| EPI_Accession_ID | Originating Laboratory | Submitting Laboratory |
|---|---|---|
| EPI_ISL_457660 | Virology Department, Sheffield Teaching Hospitals NHS Foundation Trust/Department of Infection, Immunity and Cardiovascular Disease, The Medical School, University of Sheffield | COVID-19 Genomics UK (COG-UK) Consortium |
| EPI_ISL_457661 | Virology Department, Sheffield Teaching Hospitals NHS Foundation Trust/Department of Infection, Immunity and Cardiovascular Disease, The Medical School, University of Sheffield | COVID-19 Genomics UK (COG-UK) Consortium |
| EPI_ISL_457662 | Virology Department, Sheffield Teaching Hospitals NHS Foundation Trust/Department of Infection, Immunity and Cardiovascular Disease, The Medical School, University of Sheffield | COVID-19 Genomics UK (COG-UK) Consortium |
| EPI_ISL_457663 | Virology Department, Sheffield Teaching Hospitals NHS Foundation Trust/Department of Infection, Immunity and Cardiovascular Disease, The Medical School, University of Sheffield | COVID-19 Genomics UK (COG-UK) Consortium |
| EPI_ISL_457664 | Virology Department, Sheffield Teaching Hospitals NHS Foundation Trust/Department of Infection, Immunity and Cardiovascular Disease, The Medical School, University of Sheffield | COVID-19 Genomics UK (COG-UK) Consortium |
| EPI_ISL_457665 | Virology Department, Sheffield Teaching Hospitals NHS Foundation Trust/Department of Infection, Immunity and Cardiovascular Disease, The Medical School, University of Sheffield | COVID-19 Genomics UK (COG-UK) Consortium |
| EPI_ISL_457666 | Virology Department, Sheffield Teaching Hospitals NHS Foundation Trust/Department of Infection, Immunity and Cardiovascular Disease, The Medical School, University of Sheffield | COVID-19 Genomics UK (COG-UK) Consortium |
| EPI_ISL_457667 | Virology Department, Sheffield Teaching Hospitals NHS Foundation Trust/Department of Infection, Immunity and Cardiovascular Disease, The Medical School, University of Sheffield | COVID-19 Genomics UK (COG-UK) Consortium |

| EPI_Accession_ID | Originating Laboratory | Submitting Laboratory |
| --- | --- | --- |
| EPI_ISL_457668 | Virology Department, Sheffield Teaching Hospitals NHS Foundation Trust/Department of Infection, Immunity and Cardiovascular Disease, The Medical School, University of Sheffield | COVID-19 Genomics UK (COG-UK) Consortium |
| EPI_ISL_457669 | Virology Department, Sheffield Teaching Hospitals NHS Foundation Trust/Department of Infection, Immunity and Cardiovascular Disease, The Medical School, University of Sheffield | COVID-19 Genomics UK (COG-UK) Consortium |
| EPI_ISL_457670 | Virology Department, Sheffield Teaching Hospitals NHS Foundation Trust/Department of Infection, Immunity and Cardiovascular Disease, The Medical School, University of Sheffield | COVID-19 Genomics UK (COG-UK) Consortium |
| EPI_ISL_457671 | Virology Department, Sheffield Teaching Hospitals NHS Foundation Trust/Department of Infection, Immunity and Cardiovascular Disease, The Medical School, University of Sheffield | COVID-19 Genomics UK (COG-UK) Consortium |
| EPI_ISL_457672 | Virology Department, Sheffield Teaching Hospitals NHS Foundation Trust/Department of Infection, Immunity and Cardiovascular Disease, The Medical School, University of Sheffield | COVID-19 Genomics UK (COG-UK) Consortium |
| EPI_ISL_457673 | Virology Department, Sheffield Teaching Hospitals NHS Foundation Trust/Department of Infection, Immunity and Cardiovascular Disease, The Medical School, University of Sheffield | COVID-19 Genomics UK (COG-UK) Consortium |
| EPI_ISL_457674 | Virology Department, Sheffield Teaching Hospitals NHS Foundation Trust/Department of Infection, Immunity and Cardiovascular Disease, The Medical School, University of Sheffield | COVID-19 Genomics UK (COG-UK) Consortium |
| EPI_ISL_457675 | Virology Department, Sheffield Teaching Hospitals NHS Foundation Trust/Department of Infection, Immunity and Cardiovascular Disease, The Medical School, University of Sheffield | COVID-19 Genomics UK (COG-UK) Consortium |

| EPI_Accession_ID | Originating Laboratory | Submitting Laboratory |
|---|---|---|
| EPI_ISL_457676 | Virology Department, Sheffield Teaching Hospitals NHS Foundation Trust/Department of Infection, Immunity and Cardiovascular Disease, The Medical School, University of Sheffield | COVID-19 Genomics UK (COG-UK) Consortium |
| EPI_ISL_457677 | Virology Department, Sheffield Teaching Hospitals NHS Foundation Trust/Department of Infection, Immunity and Cardiovascular Disease, The Medical School, University of Sheffield | COVID-19 Genomics UK (COG-UK) Consortium |
| EPI_ISL_457678 | Virology Department, Sheffield Teaching Hospitals NHS Foundation Trust/Department of Infection, Immunity and Cardiovascular Disease, The Medical School, University of Sheffield | COVID-19 Genomics UK (COG-UK) Consortium |
| EPI_ISL_457679 | Virology Department, Sheffield Teaching Hospitals NHS Foundation Trust/Department of Infection, Immunity and Cardiovascular Disease, The Medical School, University of Sheffield | COVID-19 Genomics UK (COG-UK) Consortium |
| EPI_ISL_457680 | Virology Department, Sheffield Teaching Hospitals NHS Foundation Trust/Department of Infection, Immunity and Cardiovascular Disease, The Medical School, University of Sheffield | COVID-19 Genomics UK (COG-UK) Consortium |
| EPI_ISL_457681 | Virology Department, Sheffield Teaching Hospitals NHS Foundation Trust/Department of Infection, Immunity and Cardiovascular Disease, The Medical School, University of Sheffield | COVID-19 Genomics UK (COG-UK) Consortium |
| EPI_ISL_457682 | Virology Department, Sheffield Teaching Hospitals NHS Foundation Trust/Department of Infection, Immunity and Cardiovascular Disease, The Medical School, University of Sheffield | COVID-19 Genomics UK (COG-UK) Consortium |
| EPI_ISL_457683 | Virology Department, Sheffield Teaching Hospitals NHS Foundation Trust/Department of Infection, Immunity and Cardiovascular Disease, The Medical School, University of Sheffield | COVID-19 Genomics UK (COG-UK) Consortium |

| EPI_Accession_ID | Originating Laboratory | Submitting Laboratory |
|---|---|---|
| EPI_ISL_457684 | Virology Department, Sheffield Teaching Hospitals NHS Foundation Trust/Department of Infection, Immunity and Cardiovascular Disease, The Medical School, University of Sheffield | COVID-19 Genomics UK (COG-UK) Consortium |
| EPI_ISL_457685 | Virology Department, Sheffield Teaching Hospitals NHS Foundation Trust/Department of Infection, Immunity and Cardiovascular Disease, The Medical School, University of Sheffield | COVID-19 Genomics UK (COG-UK) Consortium |
| EPI_ISL_457686 | Virology Department, Sheffield Teaching Hospitals NHS Foundation Trust/Department of Infection, Immunity and Cardiovascular Disease, The Medical School, University of Sheffield | COVID-19 Genomics UK (COG-UK) Consortium |
| EPI_ISL_457687 | The First Affiliated Hospital of Guangzhou Medical University, Guangzhou, China | BGI-shenzhen & The First Affiliated Hospital of Guangzhou Medical University |
| EPI_ISL_457688 | The First Affiliated Hospital of Guangzhou Medical University, Guangzhou, China | BGI-shenzhen & The First Affiliated Hospital of Guangzhou Medical University |
| EPI_ISL_457689 | The First Affiliated Hospital of Guangzhou Medical University, Guangzhou, China | BGI-shenzhen & The First Affiliated Hospital of Guangzhou Medical University |
| EPI_ISL_457690 | The First Affiliated Hospital of Guangzhou Medical University, Guangzhou, China | BGI-shenzhen & The First Affiliated Hospital of Guangzhou Medical University |
| EPI_ISL_457691 | The First Affiliated Hospital of Guangzhou Medical University, Guangzhou, China | BGI-shenzhen & The First Affiliated Hospital of Guangzhou Medical University |
| EPI_ISL_457692 | The First Affiliated Hospital of Guangzhou Medical University, Guangzhou, China | BGI-shenzhen & The First Affiliated Hospital of Guangzhou Medical University |
| EPI_ISL_457693 | The First Affiliated Hospital of Guangzhou Medical University, Guangzhou, China | BGI-shenzhen & The First Affiliated Hospital of Guangzhou Medical University |
| EPI_ISL_457694 | The First Affiliated Hospital of Guangzhou Medical University, Guangzhou, China | BGI-shenzhen & The First Affiliated Hospital of Guangzhou Medical University |
| EPI_ISL_457695 | The First Affiliated Hospital of Guangzhou Medical University, Guangzhou, China | BGI-shenzhen & The First Affiliated Hospital of Guangzhou Medical University |
| EPI_ISL_457696 | The First Affiliated Hospital of Guangzhou Medical University, Guangzhou, China | BGI-shenzhen & The First Affiliated Hospital of Guangzhou Medical University |

| EPI_Accession_ID | Originating Laboratory | Submitting Laboratory |
|---|---|---|
| EPI_ISL_457697 | The First Affiliated Hospital of Guangzhou Medical University, Guangzhou, China | BGI-shenzhen & The First Affiliated Hospital of Guangzhou Medical University |
| EPI_ISL_457698 | The First Affiliated Hospital of Guangzhou Medical University, Guangzhou, China | BGI-shenzhen & The First Affiliated Hospital of Guangzhou Medical University |
| EPI_ISL_457699 | Department of Infectious Diseases, Istituto Superiore di SanitÃ , Roma , Italy | Army Medical and Veterinary Research Center |
| EPI_ISL_457700 | Department of Infectious Diseases, Istituto Superiore di SanitÃ , Roma , Italy | Army Medical and Veterinary Research Center |
| EPI_ISL_457701 | Oman-NIC | Oman-NIC |
| EPI_ISL_457702 | Oman-NIC | Microbiology laboratory- Sultan Qaboos University Hospital |
| EPI_ISL_457703 | Oman-NIC | Department of Microbiology and Immunology- SQUH |
| EPI_ISL_457704 | Oman-NIC | Oman-NIC |
| EPI_ISL_457705 | OMAN-NIC | Department of Microbiology and Immunology- SQUH |
| EPI_ISL_457706 | Oman-NIC | Oman-NIC |
| EPI_ISL_457707 | Oman-NIC | Department of Microbiology and Immunology- SQUH |
| EPI_ISL_457721 | Department of Infectious Diseases, Istituto Superiore di SanitÃ , Roma , Italy | Army Medical and Veterinary Research Center |
| EPI_ISL_457724 | Department of Infectious Diseases, Istituto Superiore di SanitÃ , Roma , Italy | Army Medical and Veterinary Research Center |
| EPI_ISL_457726 | TSGH-CP molecular lab | TSGH-CP molecular lab |
| EPI_ISL_457728 | Department of Infectious Diseases, Istituto Superiore di SanitÃ , Roma , Italy | Army Medical and Veterinary Research Center |
| EPI_ISL_457730 | TSGH-CP molecular lab | TSGH-CP molecular lab |
| EPI_ISL_457732 | Department of Infectious Diseases, Istituto Superiore di SanitÃ , Roma , Italy | Army Medical and Veterinary Research Center |
| EPI_ISL_457733 | TSGH-CP molecular lab | TSGH-CP molecular lab |
| EPI_ISL_457736 | Department of Infectious Diseases, Istituto Superiore di SanitÃ , Roma , Italy | Army Medical and Veterinary Research Center |
| EPI_ISL_457749 | Department of Infectious Diseases, Istituto Superiore di SanitÃ , Roma , Italy | Army Medical and Veterinary Research Center |
| EPI_ISL_457750 | Centogene AG | Centogene AG |
| EPI_ISL_457751 | Johns Hopkins Hospital Department of Pathology | Johns Hopkins Hospital Department of Pathology |

| EPI_Accession_ID | Originating Laboratory | Submitting Laboratory |
|---|---|---|
| EPI_ISL_457752 | Johns Hopkins Hospital Department of Pathology | Johns Hopkins Hospital Department of Pathology |
| EPI_ISL_457753 | Johns Hopkins Hospital Department of Pathology | Johns Hopkins Hospital Department of Pathology |
| EPI_ISL_457754 | Johns Hopkins Hospital Department of Pathology | Johns Hopkins Hospital Department of Pathology |
| EPI_ISL_457755 | Johns Hopkins Hospital Department of Pathology | Johns Hopkins Hospital Department of Pathology |
| EPI_ISL_457756 | Johns Hopkins Hospital Department of Pathology | Johns Hopkins Hospital Department of Pathology |
| EPI_ISL_457757 | Johns Hopkins Hospital Department of Pathology | Johns Hopkins Hospital Department of Pathology |
| EPI_ISL_457758 | Johns Hopkins Hospital Department of Pathology | Johns Hopkins Hospital Department of Pathology |
| EPI_ISL_457759 | Johns Hopkins Hospital Department of Pathology | Johns Hopkins Hospital Department of Pathology |
| EPI_ISL_457760 | Johns Hopkins Hospital Department of Pathology | Johns Hopkins Hospital Department of Pathology |
| EPI_ISL_457761 | Johns Hopkins Hospital Department of Pathology | Johns Hopkins Hospital Department of Pathology |
| EPI_ISL_457762 | Johns Hopkins Hospital Department of Pathology | Johns Hopkins Hospital Department of Pathology |
| EPI_ISL_457763 | Johns Hopkins Hospital Department of Pathology | Johns Hopkins Hospital Department of Pathology |
| EPI_ISL_457764 | Johns Hopkins Hospital Department of Pathology | Johns Hopkins Hospital Department of Pathology |
| EPI_ISL_457765 | Johns Hopkins Hospital Department of Pathology | Johns Hopkins Hospital Department of Pathology |
| EPI_ISL_457766 | Johns Hopkins Hospital Department of Pathology | Johns Hopkins Hospital Department of Pathology |
| EPI_ISL_457767 | Johns Hopkins Hospital Department of Pathology | Johns Hopkins Hospital Department of Pathology |
| EPI_ISL_457768 | Johns Hopkins Hospital Department of Pathology | Johns Hopkins Hospital Department of Pathology |
| EPI_ISL_457769 | Johns Hopkins Hospital Department of Pathology | Johns Hopkins Hospital Department of Pathology |
| EPI_ISL_457770 | Johns Hopkins Hospital Department of Pathology | Johns Hopkins Hospital Department of Pathology |
| EPI_ISL_457771 | Johns Hopkins Hospital Department of Pathology | Johns Hopkins Hospital Department of Pathology |
| EPI_ISL_457772 | Johns Hopkins Hospital Department of Pathology | Johns Hopkins Hospital Department of Pathology |
| EPI_ISL_457773 | Johns Hopkins Hospital Department of Pathology | Johns Hopkins Hospital Department of Pathology |
| EPI_ISL_457774 | Johns Hopkins Hospital Department of Pathology | Johns Hopkins Hospital Department of Pathology |
| EPI_ISL_457775 | Johns Hopkins Hospital Department of Pathology | Johns Hopkins Hospital Department of Pathology |
| EPI_ISL_457776 | Johns Hopkins Hospital Department of Pathology | Johns Hopkins Hospital Department of Pathology |
| EPI_ISL_457777 | Johns Hopkins Hospital Department of Pathology | Johns Hopkins Hospital Department of Pathology |
| EPI_ISL_457778 | Johns Hopkins Hospital Department of Pathology | Johns Hopkins Hospital Department of Pathology |
| EPI_ISL_457779 | Johns Hopkins Hospital Department of Pathology | Johns Hopkins Hospital Department of Pathology |
| EPI_ISL_457780 | Johns Hopkins Hospital Department of Pathology | Johns Hopkins Hospital Department of Pathology |
| EPI_ISL_457781 | Johns Hopkins Hospital Department of Pathology | Johns Hopkins Hospital Department of Pathology |
| EPI_ISL_457782 | Johns Hopkins Hospital Department of Pathology | Johns Hopkins Hospital Department of Pathology |
| EPI_ISL_457783 | Johns Hopkins Hospital Department of Pathology | Johns Hopkins Hospital Department of Pathology |
| EPI_ISL_457784 | Johns Hopkins Hospital Department of Pathology | Johns Hopkins Hospital Department of Pathology |

| EPI_Accession_ID | Originating Laboratory | Submitting Laboratory |
|---|---|---|
| EPI_ISL_457785 | Johns Hopkins Hospital Department of Pathology | Johns Hopkins Hospital Department of Pathology |
| EPI_ISL_457786 | Johns Hopkins Hospital Department of Pathology | Johns Hopkins Hospital Department of Pathology |
| EPI_ISL_457787 | Johns Hopkins Hospital Department of Pathology | Johns Hopkins Hospital Department of Pathology |
| EPI_ISL_457788 | Johns Hopkins Hospital Department of Pathology | Johns Hopkins Hospital Department of Pathology |
| EPI_ISL_457789 | Johns Hopkins Hospital Department of Pathology | Johns Hopkins Hospital Department of Pathology |
| EPI_ISL_457790 | Johns Hopkins Hospital Department of Pathology | Johns Hopkins Hospital Department of Pathology |
| EPI_ISL_457791 | Johns Hopkins Hospital Department of Pathology | Johns Hopkins Hospital Department of Pathology |
| EPI_ISL_457792 | Johns Hopkins Hospital Department of Pathology | Johns Hopkins Hospital Department of Pathology |
| EPI_ISL_457793 | Johns Hopkins Hospital Department of Pathology | Johns Hopkins Hospital Department of Pathology |
| EPI_ISL_457794 | Johns Hopkins Hospital Department of Pathology | Johns Hopkins Hospital Department of Pathology |
| EPI_ISL_457795 | Johns Hopkins Hospital Department of Pathology | Johns Hopkins Hospital Department of Pathology |
| EPI_ISL_457796 | Johns Hopkins Hospital Department of Pathology | Johns Hopkins Hospital Department of Pathology |
| EPI_ISL_457797 | Johns Hopkins Hospital Department of Pathology | Johns Hopkins Hospital Department of Pathology |
| EPI_ISL_457798 | Johns Hopkins Hospital Department of Pathology | Johns Hopkins Hospital Department of Pathology |
| EPI_ISL_457799 | Johns Hopkins Hospital Department of Pathology | Johns Hopkins Hospital Department of Pathology |
| EPI_ISL_457800 | Johns Hopkins Hospital Department of Pathology | Johns Hopkins Hospital Department of Pathology |
| EPI_ISL_457801 | Johns Hopkins Hospital Department of Pathology | Johns Hopkins Hospital Department of Pathology |
| EPI_ISL_457802 | Johns Hopkins Hospital Department of Pathology | Johns Hopkins Hospital Department of Pathology |
| EPI_ISL_457803 | Johns Hopkins Hospital Department of Pathology | Johns Hopkins Hospital Department of Pathology |
| EPI_ISL_457804 | Johns Hopkins Hospital Department of Pathology | Johns Hopkins Hospital Department of Pathology |
| EPI_ISL_457805 | Johns Hopkins Hospital Department of Pathology | Johns Hopkins Hospital Department of Pathology |
| EPI_ISL_457806 | Johns Hopkins Hospital Department of Pathology | Johns Hopkins Hospital Department of Pathology |
| EPI_ISL_457807 | Johns Hopkins Hospital Department of Pathology | Johns Hopkins Hospital Department of Pathology |
| EPI_ISL_457808 | Johns Hopkins Hospital Department of Pathology | Johns Hopkins Hospital Department of Pathology |
| EPI_ISL_457809 | Johns Hopkins Hospital Department of Pathology | Johns Hopkins Hospital Department of Pathology |
| EPI_ISL_457810 | Johns Hopkins Hospital Department of Pathology | Johns Hopkins Hospital Department of Pathology |
| EPI_ISL_457811 | Johns Hopkins Hospital Department of Pathology | Johns Hopkins Hospital Department of Pathology |
| EPI_ISL_457812 | Johns Hopkins Hospital Department of Pathology | Johns Hopkins Hospital Department of Pathology |
| EPI_ISL_457813 | Johns Hopkins Hospital Department of Pathology | Johns Hopkins Hospital Department of Pathology |
| EPI_ISL_457814 | Johns Hopkins Hospital Department of Pathology | Johns Hopkins Hospital Department of Pathology |
| EPI_ISL_457815 | Johns Hopkins Hospital Department of Pathology | Johns Hopkins Hospital Department of Pathology |
| EPI_ISL_457816 | Johns Hopkins Hospital Department of Pathology | Johns Hopkins Hospital Department of Pathology |
| EPI_ISL_457817 | Johns Hopkins Hospital Department of Pathology | Johns Hopkins Hospital Department of Pathology |

| EPI_Accession_ID | Originating Laboratory | Submitting Laboratory |
|---|---|---|
| EPI_ISL_457818 | Johns Hopkins Hospital Department of Pathology | Johns Hopkins Hospital Department of Pathology |
| EPI_ISL_457819 | Johns Hopkins Hospital Department of Pathology | Johns Hopkins Hospital Department of Pathology |
| EPI_ISL_457820 | Johns Hopkins Hospital Department of Pathology | Johns Hopkins Hospital Department of Pathology |
| EPI_ISL_457821 | Johns Hopkins Hospital Department of Pathology | Johns Hopkins Hospital Department of Pathology |
| EPI_ISL_457822 | Johns Hopkins Hospital Department of Pathology | Johns Hopkins Hospital Department of Pathology |
| EPI_ISL_457823 | Johns Hopkins Hospital Department of Pathology | Johns Hopkins Hospital Department of Pathology |
| EPI_ISL_457824 | Bezmialem Vakif University, Dept Microbiology, Medical School, Fatih, Istanbul, Turkey | Bezmialem Vakif University, Medical School & Beykoz Institute of Life Sciences & Biotechnology |
| EPI_ISL_457825 | Army Medical Research Center - Scientific Department | Army Medical and Veterinary Research Center |
| EPI_ISL_457826 | Army Medical Center - Scientific Department | Army Medical and Veterinary Research Center |
| EPI_ISL_457937 | Oman-NIC | Oman-NIC |
| EPI_ISL_457938 | Oman-NIC | Oman-NIC |
| EPI_ISL_457939 | Oman-NIC | Oman-NIC |
| EPI_ISL_457940 | Laboratorio de BiologÃ-a Molecular AsociaciÃ³n EspaÃ±ola Primera en Salud | Departments of Pathology and Medicine, New York University School of Medicine |
| EPI_ISL_457941 | Laboratorio de BiologÃ-a Molecular AsociaciÃ³n EspaÃ±ola Primera en Salud | Departments of Pathology and Medicine, New York University School of Medicine |
| EPI_ISL_457942 | Laboratorio de BiologÃ-a Molecular AsociaciÃ³n EspaÃ±ola Primera en Salud | Departments of Pathology and Medicine, New York University School of Medicine |
| EPI_ISL_457943 | Laboratorio de BiologÃ-a Molecular AsociaciÃ³n EspaÃ±ola Primera en Salud | Departments of Pathology and Medicine, New York University School of Medicine |
| EPI_ISL_457944 | Laboratorio de BiologÃ-a Molecular AsociaciÃ³n EspaÃ±ola Primera en Salud | Departments of Pathology and Medicine, New York University School of Medicine |
| EPI_ISL_457946 | Laboratorio de BiologÃ-a Molecular AsociaciÃ³n EspaÃ±ola Primera en Salud | Departments of Pathology and Medicine, New York University School of Medicine |
| EPI_ISL_457947 | Laboratorio de BiologÃ-a Molecular AsociaciÃ³n EspaÃ±ola Primera en Salud | Departments of Pathology and Medicine, New York University School of Medicine |
| EPI_ISL_457948 | Laboratorio de BiologÃ-a Molecular AsociaciÃ³n EspaÃ±ola Primera en Salud | Departments of Pathology and Medicine, New York University School of Medicine |
| EPI_ISL_457949 | Laboratorio de BiologÃ-a Molecular AsociaciÃ³n EspaÃ±ola Primera en Salud | Departments of Pathology and Medicine, New York University School of Medicine |
| EPI_ISL_457950 | Laboratorio de BiologÃ-a Molecular AsociaciÃ³n EspaÃ±ola Primera en Salud | Departments of Pathology and Medicine, New York University School of Medicine |

| EPI_Accession_ID | Originating Laboratory | Submitting Laboratory |
| --- | --- | --- |
| EPI_ISL_457951 | Laboratorio de BiologÃ-a Molecular AsociaciÃ³n EspaÃ±ola Primera en Salud | Departments of Pathology and Medicine, New York University School of Medicine |
| EPI_ISL_457952 | Laboratorio de BiologÃ-a Molecular AsociaciÃ³n EspaÃ±ola Primera en Salud | Departments of Pathology and Medicine, New York University School of Medicine |
| EPI_ISL_457953 | Laboratorio de BiologÃ-a Molecular AsociaciÃ³n EspaÃ±ola Primera en Salud | Departments of Pathology and Medicine, New York University School of Medicine |
| EPI_ISL_457954 | Laboratorio de BiologÃ-a Molecular AsociaciÃ³n EspaÃ±ola Primera en Salud | Departments of Pathology and Medicine, New York University School of Medicine |
| EPI_ISL_457955 | Laboratorio de BiologÃ-a Molecular AsociaciÃ³n EspaÃ±ola Primera en Salud | Departments of Pathology and Medicine, New York University School of Medicine |
| EPI_ISL_457956 | Laboratorio de BiologÃ-a Molecular AsociaciÃ³n EspaÃ±ola Primera en Salud | Departments of Pathology and Medicine, New York University School of Medicine |
| EPI_ISL_457957 | Laboratorio de BiologÃ-a Molecular AsociaciÃ³n EspaÃ±ola Primera en Salud | Departments of Pathology and Medicine, New York University School of Medicine |
| EPI_ISL_457958 | Laboratorio de BiologÃ-a Molecular AsociaciÃ³n EspaÃ±ola Primera en Salud | Departments of Pathology and Medicine, New York University School of Medicine |
| EPI_ISL_457959 | Laboratorio de BiologÃ-a Molecular AsociaciÃ³n EspaÃ±ola Primera en Salud | Departments of Pathology and Medicine, New York University School of Medicine |
| EPI_ISL_457960 | Laboratorio de BiologÃ-a Molecular AsociaciÃ³n EspaÃ±ola Primera en Salud | Departments of Pathology and Medicine, New York University School of Medicine |
| EPI_ISL_457961 | Laboratorio de BiologÃ-a Molecular AsociaciÃ³n EspaÃ±ola Primera en Salud | Departments of Pathology and Medicine, New York University School of Medicine |
| EPI_ISL_457962 | Laboratorio de BiologÃ-a Molecular AsociaciÃ³n EspaÃ±ola Primera en Salud | Departments of Pathology and Medicine, New York University School of Medicine |
| EPI_ISL_457963 | Laboratorio de BiologÃ-a Molecular AsociaciÃ³n EspaÃ±ola Primera en Salud | Departments of Pathology and Medicine, New York University School of Medicine |
| EPI_ISL_457964 | Laboratorio de BiologÃ-a Molecular AsociaciÃ³n EspaÃ±ola Primera en Salud | Departments of Pathology and Medicine, New York University School of Medicine |
| EPI_ISL_457965 | Laboratorio de BiologÃ-a Molecular AsociaciÃ³n EspaÃ±ola Primera en Salud | Departments of Pathology and Medicine, New York University School of Medicine |
| EPI_ISL_457966 | Laboratorio de BiologÃ-a Molecular AsociaciÃ³n EspaÃ±ola Primera en Salud | Departments of Pathology and Medicine, New York University School of Medicine |

| EPI_Accession_ID | Originating Laboratory | Submitting Laboratory |
|---|---|---|
| EPI_ISL_457967 | Laboratorio de BiologÃ-a Molecular AsociaciÃ³n EspaÃ±ola Primera en Salud | Departments of Pathology and Medicine, New York University School of Medicine |
| EPI_ISL_457968 | Laboratorio de BiologÃ-a Molecular AsociaciÃ³n EspaÃ±ola Primera en Salud | Departments of Pathology and Medicine, New York University School of Medicine |
| EPI_ISL_457969 | Laboratorio de BiologÃ-a Molecular AsociaciÃ³n EspaÃ±ola Primera en Salud | Departments of Pathology and Medicine, New York University School of Medicine |
| EPI_ISL_457970 | Laboratorio de BiologÃ-a Molecular AsociaciÃ³n EspaÃ±ola Primera en Salud | Departments of Pathology and Medicine, New York University School of Medicine |
| EPI_ISL_457971 | Laboratorio de BiologÃ-a Molecular AsociaciÃ³n EspaÃ±ola Primera en Salud | Departments of Pathology and Medicine, New York University School of Medicine |
| EPI_ISL_457972 | Laboratorio de BiologÃ-a Molecular AsociaciÃ³n EspaÃ±ola Primera en Salud | Departments of Pathology and Medicine, New York University School of Medicine |
| EPI_ISL_457973 | Laboratorio de BiologÃ-a Molecular AsociaciÃ³n EspaÃ±ola Primera en Salud | Departments of Pathology and Medicine, New York University School of Medicine |
| EPI_ISL_457974 | Oman-NIC | Oman-NIC |
| EPI_ISL_457975 | Oman-NIC | Oman-NIC |
| EPI_ISL_457976 | Oman-NIC | Oman-NIC |
| EPI_ISL_457977 | Oman-NIC | Oman-NIC |
| EPI_ISL_457978 | Oman-NIC | Oman-NIC |
| EPI_ISL_457979 | Oman-NIC | Oman-NIC |
| EPI_ISL_457980 | Oman-NIC | Oman-NIC |
| EPI_ISL_457981 | Oman-NIC | Department of Microbiology and Immunology-SQUH |
| EPI_ISL_457982 | Oman-NIC | Department of Microbiology and Immunology-SQUH |
| EPI_ISL_457983 | Oman-NIC | Department of Microbiology and Immunology-SQUH |
| EPI_ISL_457984 | Oman-NIC | Department of Microbiology and Immunology-SQUH |
| EPI_ISL_457985 | Oman-NIC | Oman-NIC |
| EPI_ISL_457986 | Oman-NIC | Oman-NIC |
| EPI_ISL_457987 | Oman-NIC | Oman-NIC |
| EPI_ISL_457988 | Oman-NIC | Oman-NIC |
| EPI_ISL_457989 | Oman-NIC | Oman-NIC |
| EPI_ISL_457990 | Oman-NIC | Oman-NIC |
| EPI_ISL_457991 | Oman-NIC | Oman-NIC |
| EPI_ISL_457992 | Oman-NIC | Oman-NIC |

| EPI_Accession_ID | Originating Laboratory | Submitting Laboratory |
|---|---|---|
| EPI_ISL_457993 | Oman-NIC | Oman-NIC |
| EPI_ISL_457994 | Oman-NIC | Oman-NIC |
| EPI_ISL_457995 | Oman-NIC | Oman-NIC |
| EPI_ISL_457996 | Oman-NIC | Oman-NIC |
| EPI_ISL_457997 | Oman-NIC | Oman-NIC |
| EPI_ISL_457998 | Oman-NIC | Oman-NIC |
| EPI_ISL_457999 | unknown | Centre For Biotechnology Research and Development |
| EPI_ISL_458000 | unknown | Centre For Biotechnology Research and Development |
| EPI_ISL_458017 | NYU Langone Health | Departments of Pathology and Medicine, New York University School of Medicine |
| EPI_ISL_458018 | NYU Langone Health | Departments of Pathology and Medicine, New York University School of Medicine |
| EPI_ISL_458019 | NYU Langone Health | Departments of Pathology and Medicine, New York University School of Medicine |
| EPI_ISL_458020 | NYU Langone Health | Departments of Pathology and Medicine, New York University School of Medicine |
| EPI_ISL_458021 | NYU Langone Health | Departments of Pathology and Medicine, New York University School of Medicine |
| EPI_ISL_458022 | NYU Langone Health | Departments of Pathology and Medicine, New York University School of Medicine |
| EPI_ISL_458023 | Hospital for Tropical Diseases | COVID-19 Network Investigations (CONI) Alliance |
| EPI_ISL_458024 | Hospital for Tropical Diseases | COVID-19 Network Investigations (CONI) Alliance |
| EPI_ISL_458025 | Hospital for Tropical Diseases | COVID-19 Network Investigations (CONI) Alliance |
| EPI_ISL_458026 | Hospital for Tropical Diseases | COVID-19 Network Investigations (CONI) Alliance |
| EPI_ISL_458027 | Hospital for Tropical Diseases | COVID-19 Network Investigations (CONI) Alliance |
| EPI_ISL_458028 | Hospital for Tropical Diseases | COVID-19 Network Investigations (CONI) Alliance |
| EPI_ISL_458029 | TSGH-CP molecular lab | TSGH-CP molecular lab |
| EPI_ISL_458030 | King Institute of Preventive Medicine & Research | CSIR-Centre for Cellular and Molecular Biology |
| EPI_ISL_458031 | King Institute of Preventive Medicine & Research | CSIR-Centre for Cellular and Molecular Biology |
| EPI_ISL_458032 | King Institute of Preventive Medicine & Research | CSIR-Centre for Cellular and Molecular Biology |
| EPI_ISL_458033 | King Institute of Preventive Medicine & Research | CSIR-Centre for Cellular and Molecular Biology |
| EPI_ISL_458034 | King Institute of Preventive Medicine & Research | CSIR-Centre for Cellular and Molecular Biology |
| EPI_ISL_458035 | King Institute of Preventive Medicine & Research | CSIR-Centre for Cellular and Molecular Biology |

| EPI_Accession_ID | Originating Laboratory | Submitting Laboratory |
|---|---|---|
| EPI_ISL_458036 | King Institute of Preventive Medicine & Research | CSIR-Centre for Cellular and Molecular Biology |
| EPI_ISL_458037 | King Institute of Preventive Medicine & Research | CSIR-Centre for Cellular and Molecular Biology |
| EPI_ISL_458038 | King Institute of Preventive Medicine & Research | CSIR-Centre for Cellular and Molecular Biology |
| EPI_ISL_458039 | King Institute of Preventive Medicine & Research | CSIR-Centre for Cellular and Molecular Biology |
| EPI_ISL_458040 | King Institute of Preventive Medicine & Research | CSIR-Centre for Cellular and Molecular Biology |
| EPI_ISL_458041 | King Institute of Preventive Medicine & Research | CSIR-Centre for Cellular and Molecular Biology |
| EPI_ISL_458042 | King Institute of Preventive Medicine & Research | CSIR-Centre for Cellular and Molecular Biology |
| EPI_ISL_458043 | King Institute of Preventive Medicine & Research | CSIR-Centre for Cellular and Molecular Biology |
| EPI_ISL_458044 | King Institute of Preventive Medicine & Research | CSIR-Centre for Cellular and Molecular Biology |
| EPI_ISL_458045 | CSIR-Centre for Cellular and Molecular Biology | CSIR-Centre for Cellular and Molecular Biology |
| EPI_ISL_458046 | CSIR-Centre for Cellular and Molecular Biology | CSIR-Centre for Cellular and Molecular Biology |
| EPI_ISL_458047 | CSIR-Centre for Cellular and Molecular Biology | CSIR-Centre for Cellular and Molecular Biology |
| EPI_ISL_458048 | CSIR-Centre for Cellular and Molecular Biology | CSIR-Centre for Cellular and Molecular Biology |
| EPI_ISL_458049 | CSIR-Centre for Cellular and Molecular Biology | CSIR-Centre for Cellular and Molecular Biology |
| EPI_ISL_458050 | CSIR-Centre for Cellular and Molecular Biology | CSIR-Centre for Cellular and Molecular Biology |
| EPI_ISL_458051 | CSIR-Centre for Cellular and Molecular Biology | CSIR-Centre for Cellular and Molecular Biology |
| EPI_ISL_458052 | CSIR-Centre for Cellular and Molecular Biology | CSIR-Centre for Cellular and Molecular Biology |
| EPI_ISL_458053 | CSIR-Centre for Cellular and Molecular Biology | CSIR-Centre for Cellular and Molecular Biology |
| EPI_ISL_458054 | CSIR-Centre for Cellular and Molecular Biology | CSIR-Centre for Cellular and Molecular Biology |
| EPI_ISL_458055 | CSIR-Centre for Cellular and Molecular Biology | CSIR-Centre for Cellular and Molecular Biology |
| EPI_ISL_458056 | CSIR-Centre for Cellular and Molecular Biology | CSIR-Centre for Cellular and Molecular Biology |
| EPI_ISL_458057 | CSIR-Centre for Cellular and Molecular Biology | CSIR-Centre for Cellular and Molecular Biology |
| EPI_ISL_458058 | CSIR-Centre for Cellular and Molecular Biology | CSIR-Centre for Cellular and Molecular Biology |
| EPI_ISL_458059 | CSIR-Centre for Cellular and Molecular Biology | CSIR-Centre for Cellular and Molecular Biology |
| EPI_ISL_458060 | CSIR-Centre for Cellular and Molecular Biology | CSIR-Centre for Cellular and Molecular Biology |
| EPI_ISL_458061 | CSIR-Centre for Cellular and Molecular Biology | CSIR-Centre for Cellular and Molecular Biology |
| EPI_ISL_458062 | CSIR-Centre for Cellular and Molecular Biology | CSIR-Centre for Cellular and Molecular Biology |
| EPI_ISL_458063 | CSIR-Centre for Cellular and Molecular Biology | CSIR-Centre for Cellular and Molecular Biology |
| EPI_ISL_458064 | CSIR-Centre for Cellular and Molecular Biology | CSIR-Centre for Cellular and Molecular Biology |
| EPI_ISL_458065 | CSIR-Centre for Cellular and Molecular Biology | CSIR-Centre for Cellular and Molecular Biology |
| EPI_ISL_458066 | Osmania Medical College | CSIR-Centre for Cellular and Molecular Biology |
| EPI_ISL_458067 | Osmania Medical College | CSIR-Centre for Cellular and Molecular Biology |
| EPI_ISL_458068 | Osmania Medical College | CSIR-Centre for Cellular and Molecular Biology |

| EPI_Accession_ID | Originating Laboratory | Submitting Laboratory |
|---|---|---|
| EPI_ISL_458069 | Osmania Medical College | CSIR-Centre for Cellular and Molecular Biology |
| EPI_ISL_458070 | CSIR-Centre for Cellular and Molecular Biology | CSIR-Centre for Cellular and Molecular Biology |
| EPI_ISL_458071 | CSIR-Centre for Cellular and Molecular Biology | CSIR-Centre for Cellular and Molecular Biology |
| EPI_ISL_458072 | CSIR-Centre for Cellular and Molecular Biology | CSIR-Centre for Cellular and Molecular Biology |
| EPI_ISL_458073 | CSIR-Centre for Cellular and Molecular Biology | CSIR-Centre for Cellular and Molecular Biology |
| EPI_ISL_458074 | CSIR-Centre for Cellular and Molecular Biology | CSIR-Centre for Cellular and Molecular Biology |
| EPI_ISL_458075 | CSIR-Centre for Cellular and Molecular Biology | CSIR-Centre for Cellular and Molecular Biology |
| EPI_ISL_458076 | CSIR-Centre for Cellular and Molecular Biology | CSIR-Centre for Cellular and Molecular Biology |
| EPI_ISL_458077 | CSIR-Centre for Cellular and Molecular Biology | CSIR-Centre for Cellular and Molecular Biology |
| EPI_ISL_458079 | Mitra Keluarga Hospital Kenjeran | Institute of Tropical Disease, Universitas Airlangga |
| EPI_ISL_458080 | CSIR-Centre for Cellular and Molecular Biology | CSIR-Centre for Cellular and Molecular Biology |
| EPI_ISL_458081 | RSUD Bangil Pasuruan | Institute of Tropical Disease, Universitas Airlangga |
| EPI_ISL_458083 | Adi Husada Undaan Hospital | Institute of Tropical Disease, Universitas Airlangga |
| EPI_ISL_458084 | Laboratorio Biologia Molecolare Sars Cov2 - UOC Laboratorio Analisi - Servizio Medicina di Laboratorio, Ospedale "San Francesco" - ATS-ASSL Nuoro | Laboratorio specialistico UOC Ematologia - Ospedale "San Francesco" - ATS-ASSL Nuoro |
| EPI_ISL_458085 | Laboratorio Biologia Molecolare Sars Cov2 - UOC Laboratorio Analisi - Servizio Medicina di Laboratorio , Ospedale "San Francesco" - ATS- ASSL Nuoro | Laboratorio specialistico UOC Ematologia - Ospedale "San Francesco"- ATS-ASSL Nuoro |
| EPI_ISL_458086 | B.J. Medical College and Civil hospital | Gujarat Biotechnology Research Centre |
| EPI_ISL_458087 | B.J. Medical College and Civil hospital | Gujarat Biotechnology Research Centre |
| EPI_ISL_458088 | B.J. Medical College and Civil hospital | Gujarat Biotechnology Research Centre |
| EPI_ISL_458089 | B.J. Medical College and Civil hospital | Gujarat Biotechnology Research Centre |
| EPI_ISL_458090 | B.J. Medical College and Civil hospital | Gujarat Biotechnology Research Centre |
| EPI_ISL_458091 | B.J. Medical College and Civil hospital | Gujarat Biotechnology Research Centre |
| EPI_ISL_458092 | B.J. Medical College and Civil hospital | Gujarat Biotechnology Research Centre |
| EPI_ISL_458093 | B.J. Medical College and Civil hospital | Gujarat Biotechnology Research Centre |
| EPI_ISL_458094 | B.J. Medical College and Civil hospital | Gujarat Biotechnology Research Centre |
| EPI_ISL_458095 | B.J. Medical College and Civil hospital | Gujarat Biotechnology Research Centre |
| EPI_ISL_458096 | B.J. Medical College and Civil hospital | Gujarat Biotechnology Research Centre |
| EPI_ISL_458097 | B.J. Medical College and Civil hospital | Gujarat Biotechnology Research Centre |
| EPI_ISL_458098 | B.J. Medical College and Civil hospital | Gujarat Biotechnology Research Centre |
| EPI_ISL_458099 | B.J. Medical College and Civil hospital | Gujarat Biotechnology Research Centre |

| EPI_Accession_ID | Originating Laboratory | Submitting Laboratory |
|---|---|---|
| EPI_ISL_458100 | B.J. Medical College and Civil hospital | Gujarat Biotechnology Research Centre |
| EPI_ISL_458101 | B.J. Medical College and Civil hospital | Gujarat Biotechnology Research Centre |
| EPI_ISL_458102 | B.J. Medical College and Civil hospital | Gujarat Biotechnology Research Centre |
| EPI_ISL_458103 | Gujarat Biotechnology Research Centre | Gujarat Biotechnology Research Centre |
| EPI_ISL_458104 | Gujarat Biotechnology Research Centre | Gujarat Biotechnology Research Centre |
| EPI_ISL_458105 | Gujarat Biotechnology Research Centre | Gujarat Biotechnology Research Centre |
| EPI_ISL_458106 | Gujarat Biotechnology Research Centre | Gujarat Biotechnology Research Centre |
| EPI_ISL_458107 | Gujarat Biotechnology Research Centre | Gujarat Biotechnology Research Centre |
| EPI_ISL_458108 | Gujarat Biotechnology Research Centre | Gujarat Biotechnology Research Centre |
| EPI_ISL_458109 | Gujarat Biotechnology Research Centre | Gujarat Biotechnology Research Centre |
| EPI_ISL_458110 | Gujarat Biotechnology Research Centre | Gujarat Biotechnology Research Centre |
| EPI_ISL_458111 | Gujarat Biotechnology Research Centre | Gujarat Biotechnology Research Centre |
| EPI_ISL_458112 | Gujarat Biotechnology Research Centre | Gujarat Biotechnology Research Centre |
| EPI_ISL_458113 | Gujarat Biotechnology Research Centre | Gujarat Biotechnology Research Centre |
| EPI_ISL_458116 | Oman National Influenza Centre | Department of Microbiology and Immunology-SQUH |
| EPI_ISL_458117 | Oman National Influenza Centre | Department of Microbiology and Immunology-SQUH |
| EPI_ISL_458118 | Oman National Influenza Centre | Department of Microbiology and Immunology-SQUH |
| EPI_ISL_458119 | Oman National Influenza Centre | Department of Microbiology and Immunology-SQUH |
| EPI_ISL_458120 | Oman National Influenza Centre | Department of Microbiology and Immunology-SQUH |
| EPI_ISL_458121 | Oman National Influenza Centre | Department of Microbiology and Immunology-SQUH |
| EPI_ISL_458122 | Oman National Influenza Centre | Department of Microbiology and Immunology-SQUH |
| EPI_ISL_458123 | Oman National Influenza Centre | Department of Microbiology and Immunology-SQUH |
| EPI_ISL_458124 | Oman National Influenza Centre | Department of Microbiology and Immunology-SQUH |
| EPI_ISL_458125 | Oman National Influenza Centre | Department of Microbiology and Immunology-SQUH |
| EPI_ISL_458126 | Oman National Influenza Centre | Department of Microbiology and Immunology-SQUH |
| EPI_ISL_458127 | Oman National Influenza Centre | Department of Microbiology and Immunology-SQUH |
| EPI_ISL_458128 | Oman National Influenza Centre | Department of Microbiology and Immunology-SQUH |
| EPI_ISL_458130 | Hospital Universitari Vall d'Hebron - Vall d'Hebron Institut de Recerca | Hospital Universitari Vall d'Hebron |
| EPI_ISL_458131 | Hospital Universitari Vall d'Hebron - Vall d'Hebron Institut de Recerca | Hospital Universitari Vall d'Hebron |
| EPI_ISL_458132 | Hospital Universitari Vall d'Hebron - Vall d'Hebron Institut de Rercerca | Hospital Universitari Vall d'Hebron |

| EPI_Accession_ID | Originating Laboratory | Submitting Laboratory |
|---|---|---|
| EPI_ISL_458133 | National Institute of Biotechnology | Bioinformatics Division, National Institute of Biotechnology |
| EPI_ISL_458137 | Oman National Influenza Centre | Department of Microbiology and Immunology |
| EPI_ISL_458138 | ï»¿Evandro Chagas Institute | ï»¿Evandro Chagas Institute |
| EPI_ISL_458139 | ï»¿Evandro Chagas Institute | ï»¿Evandro Chagas Institute |
| EPI_ISL_458140 | ï»¿Evandro Chagas Institute | ï»¿Evandro Chagas Institute |
| EPI_ISL_458141 | ï»¿Evandro Chagas Institute | ï»¿Evandro Chagas Institute |
| EPI_ISL_458142 | ï»¿Evandro Chagas Institute | ï»¿Evandro Chagas Institute |
| EPI_ISL_458143 | ï»¿Evandro Chagas Institute | ï»¿Evandro Chagas Institute |
| EPI_ISL_458144 | ï»¿Evandro Chagas Institute | ï»¿Evandro Chagas Institute |
| EPI_ISL_458145 | ï»¿Evandro Chagas Institute | ï»¿Evandro Chagas Institute |
| EPI_ISL_458146 | ï»¿Evandro Chagas Institute | ï»¿Evandro Chagas Institute |
| EPI_ISL_458147 | ï»¿Evandro Chagas Institute | ï»¿Evandro Chagas Institute |
| EPI_ISL_458148 | ï»¿Evandro Chagas Institute | ï»¿Evandro Chagas Institute |
| EPI_ISL_458149 | ï»¿Evandro Chagas Institute | ï»¿Evandro Chagas Institute |
| EPI_ISL_458150 | ANOUAL | ANOUAL |
| EPI_ISL_458156 | KU Leuven, Rega Institute, Clinical and Epidemiological Virology | KU Leuven, Rega Institute, Clinical and Epidemiological Virology |
| EPI_ISL_458157 | KU Leuven, Rega Institute, Clinical and Epidemiological Virology | KU Leuven, Rega Institute, Clinical and Epidemiological Virology |
| EPI_ISL_458158 | KU Leuven, Rega Institute, Clinical and Epidemiological Virology | KU Leuven, Rega Institute, Clinical and Epidemiological Virology |
| EPI_ISL_458159 | KU Leuven, Rega Institute, Clinical and Epidemiological Virology | KU Leuven, Rega Institute, Clinical and Epidemiological Virology |
| EPI_ISL_458160 | KU Leuven, Rega Institute, Clinical and Epidemiological Virology | KU Leuven, Rega Institute, Clinical and Epidemiological Virology |
| EPI_ISL_458161 | KU Leuven, Rega Institute, Clinical and Epidemiological Virology | KU Leuven, Rega Institute, Clinical and Epidemiological Virology |
| EPI_ISL_458162 | KU Leuven, Rega Institute, Clinical and Epidemiological Virology | KU Leuven, Rega Institute, Clinical and Epidemiological Virology |
| EPI_ISL_458163 | KU Leuven, Rega Institute, Clinical and Epidemiological Virology | KU Leuven, Rega Institute, Clinical and Epidemiological Virology |

| EPI_Accession_ID | Originating Laboratory | Submitting Laboratory |
|---|---|---|
| EPI_ISL_458164 | KU Leuven, Rega Institute, Clinical and Epidemiological Virology | KU Leuven, Rega Institute, Clinical and Epidemiological Virology |
| EPI_ISL_458165 | KU Leuven, Rega Institute, Clinical and Epidemiological Virology | KU Leuven, Rega Institute, Clinical and Epidemiological Virology |
| EPI_ISL_458166 | KU Leuven, Rega Institute, Clinical and Epidemiological Virology | KU Leuven, Rega Institute, Clinical and Epidemiological Virology |
| EPI_ISL_458167 | KU Leuven, Rega Institute, Clinical and Epidemiological Virology | KU Leuven, Rega Institute, Clinical and Epidemiological Virology |
| EPI_ISL_458168 | KU Leuven, Rega Institute, Clinical and Epidemiological Virology | KU Leuven, Rega Institute, Clinical and Epidemiological Virology |
| EPI_ISL_458169 | KU Leuven, Rega Institute, Clinical and Epidemiological Virology | KU Leuven, Rega Institute, Clinical and Epidemiological Virology |
| EPI_ISL_458170 | KU Leuven, Rega Institute, Clinical and Epidemiological Virology | KU Leuven, Rega Institute, Clinical and Epidemiological Virology |
| EPI_ISL_458171 | KU Leuven, Rega Institute, Clinical and Epidemiological Virology | KU Leuven, Rega Institute, Clinical and Epidemiological Virology |
| EPI_ISL_458172 | KU Leuven, Rega Institute, Clinical and Epidemiological Virology | KU Leuven, Rega Institute, Clinical and Epidemiological Virology |
| EPI_ISL_458173 | KU Leuven, Rega Institute, Clinical and Epidemiological Virology | KU Leuven, Rega Institute, Clinical and Epidemiological Virology |
| EPI_ISL_458174 | KU Leuven, Rega Institute, Clinical and Epidemiological Virology | KU Leuven, Rega Institute, Clinical and Epidemiological Virology |
| EPI_ISL_458175 | KU Leuven, Rega Institute, Clinical and Epidemiological Virology | KU Leuven, Rega Institute, Clinical and Epidemiological Virology |
| EPI_ISL_458176 | KU Leuven, Rega Institute, Clinical and Epidemiological Virology | KU Leuven, Rega Institute, Clinical and Epidemiological Virology |
| EPI_ISL_458177 | KU Leuven, Rega Institute, Clinical and Epidemiological Virology | KU Leuven, Rega Institute, Clinical and Epidemiological Virology |
| EPI_ISL_458178 | KU Leuven, Rega Institute, Clinical and Epidemiological Virology | KU Leuven, Rega Institute, Clinical and Epidemiological Virology |
| EPI_ISL_458179 | KU Leuven, Rega Institute, Clinical and Epidemiological Virology | KU Leuven, Rega Institute, Clinical and Epidemiological Virology |

| EPI_Accession_ID | Originating Laboratory | Submitting Laboratory |
|---|---|---|
| EPI_ISL_458180 | KU Leuven, Rega Institute, Clinical and Epidemiological Virology | KU Leuven, Rega Institute, Clinical and Epidemiological Virology |
| EPI_ISL_458181 | KU Leuven, Rega Institute, Clinical and Epidemiological Virology | KU Leuven, Rega Institute, Clinical and Epidemiological Virology |
| EPI_ISL_458182 | KU Leuven, Rega Institute, Clinical and Epidemiological Virology | KU Leuven, Rega Institute, Clinical and Epidemiological Virology |
| EPI_ISL_458183 | KU Leuven, Rega Institute, Clinical and Epidemiological Virology | KU Leuven, Rega Institute, Clinical and Epidemiological Virology |
| EPI_ISL_458184 | KU Leuven, Rega Institute, Clinical and Epidemiological Virology | KU Leuven, Rega Institute, Clinical and Epidemiological Virology |
| EPI_ISL_458185 | KU Leuven, Rega Institute, Clinical and Epidemiological Virology | KU Leuven, Rega Institute, Clinical and Epidemiological Virology |
| EPI_ISL_458186 | KU Leuven, Rega Institute, Clinical and Epidemiological Virology | KU Leuven, Rega Institute, Clinical and Epidemiological Virology |
| EPI_ISL_458187 | KU Leuven, Rega Institute, Clinical and Epidemiological Virology | KU Leuven, Rega Institute, Clinical and Epidemiological Virology |
| EPI_ISL_458188 | KU Leuven, Rega Institute, Clinical and Epidemiological Virology | KU Leuven, Rega Institute, Clinical and Epidemiological Virology |
| EPI_ISL_458189 | KU Leuven, Rega Institute, Clinical and Epidemiological Virology | KU Leuven, Rega Institute, Clinical and Epidemiological Virology |
| EPI_ISL_458190 | KU Leuven, Rega Institute, Clinical and Epidemiological Virology | KU Leuven, Rega Institute, Clinical and Epidemiological Virology |
| EPI_ISL_458191 | KU Leuven, Rega Institute, Clinical and Epidemiological Virology | KU Leuven, Rega Institute, Clinical and Epidemiological Virology |
| EPI_ISL_458192 | KU Leuven, Rega Institute, Clinical and Epidemiological Virology | KU Leuven, Rega Institute, Clinical and Epidemiological Virology |
| EPI_ISL_458193 | KU Leuven, Rega Institute, Clinical and Epidemiological Virology | KU Leuven, Rega Institute, Clinical and Epidemiological Virology |
| EPI_ISL_458194 | KU Leuven, Rega Institute, Clinical and Epidemiological Virology | KU Leuven, Rega Institute, Clinical and Epidemiological Virology |
| EPI_ISL_458195 | KU Leuven, Rega Institute, Clinical and Epidemiological Virology | KU Leuven, Rega Institute, Clinical and Epidemiological Virology |

| EPI_Accession_ID | Originating Laboratory | Submitting Laboratory |
|---|---|---|
| EPI_ISL_458196 | KU Leuven, Rega Institute, Clinical and Epidemiological Virology | KU Leuven, Rega Institute, Clinical and Epidemiological Virology |
| EPI_ISL_458197 | KU Leuven, Rega Institute, Clinical and Epidemiological Virology | KU Leuven, Rega Institute, Clinical and Epidemiological Virology |
| EPI_ISL_458198 | KU Leuven, Rega Institute, Clinical and Epidemiological Virology | KU Leuven, Rega Institute, Clinical and Epidemiological Virology |
| EPI_ISL_458199 | KU Leuven, Rega Institute, Clinical and Epidemiological Virology | KU Leuven, Rega Institute, Clinical and Epidemiological Virology |
| EPI_ISL_458200 | KU Leuven, Rega Institute, Clinical and Epidemiological Virology | KU Leuven, Rega Institute, Clinical and Epidemiological Virology |
| EPI_ISL_458201 | KU Leuven, Rega Institute, Clinical and Epidemiological Virology | KU Leuven, Rega Institute, Clinical and Epidemiological Virology |
| EPI_ISL_458202 | KU Leuven, Rega Institute, Clinical and Epidemiological Virology | KU Leuven, Rega Institute, Clinical and Epidemiological Virology |
| EPI_ISL_458203 | KU Leuven, Rega Institute, Clinical and Epidemiological Virology | KU Leuven, Rega Institute, Clinical and Epidemiological Virology |
| EPI_ISL_458204 | KU Leuven, Rega Institute, Clinical and Epidemiological Virology | KU Leuven, Rega Institute, Clinical and Epidemiological Virology |
| EPI_ISL_458205 | KU Leuven, Rega Institute, Clinical and Epidemiological Virology | KU Leuven, Rega Institute, Clinical and Epidemiological Virology |
| EPI_ISL_458206 | KU Leuven, Rega Institute, Clinical and Epidemiological Virology | KU Leuven, Rega Institute, Clinical and Epidemiological Virology |
| EPI_ISL_458207 | KU Leuven, Rega Institute, Clinical and Epidemiological Virology | KU Leuven, Rega Institute, Clinical and Epidemiological Virology |
| EPI_ISL_458208 | KU Leuven, Rega Institute, Clinical and Epidemiological Virology | KU Leuven, Rega Institute, Clinical and Epidemiological Virology |
| EPI_ISL_458209 | KU Leuven, Rega Institute, Clinical and Epidemiological Virology | KU Leuven, Rega Institute, Clinical and Epidemiological Virology |
| EPI_ISL_458210 | KU Leuven, Rega Institute, Clinical and Epidemiological Virology | KU Leuven, Rega Institute, Clinical and Epidemiological Virology |
| EPI_ISL_458211 | KU Leuven, Rega Institute, Clinical and Epidemiological Virology | KU Leuven, Rega Institute, Clinical and Epidemiological Virology |

| EPI_Accession_ID | Originating Laboratory | Submitting Laboratory |
|---|---|---|
| EPI_ISL_458212 | KU Leuven, Rega Institute, Clinical and Epidemiological Virology | KU Leuven, Rega Institute, Clinical and Epidemiological Virology |
| EPI_ISL_458213 | KU Leuven, Rega Institute, Clinical and Epidemiological Virology | KU Leuven, Rega Institute, Clinical and Epidemiological Virology |
| EPI_ISL_458214 | KU Leuven, Rega Institute, Clinical and Epidemiological Virology | KU Leuven, Rega Institute, Clinical and Epidemiological Virology |
| EPI_ISL_458215 | KU Leuven, Rega Institute, Clinical and Epidemiological Virology | KU Leuven, Rega Institute, Clinical and Epidemiological Virology |
| EPI_ISL_458216 | KU Leuven, Rega Institute, Clinical and Epidemiological Virology | KU Leuven, Rega Institute, Clinical and Epidemiological Virology |
| EPI_ISL_458217 | KU Leuven, Rega Institute, Clinical and Epidemiological Virology | KU Leuven, Rega Institute, Clinical and Epidemiological Virology |
| EPI_ISL_458218 | KU Leuven, Rega Institute, Clinical and Epidemiological Virology | KU Leuven, Rega Institute, Clinical and Epidemiological Virology |
| EPI_ISL_458219 | KU Leuven, Rega Institute, Clinical and Epidemiological Virology | KU Leuven, Rega Institute, Clinical and Epidemiological Virology |
| EPI_ISL_458220 | KU Leuven, Rega Institute, Clinical and Epidemiological Virology | KU Leuven, Rega Institute, Clinical and Epidemiological Virology |
| EPI_ISL_458221 | KU Leuven, Rega Institute, Clinical and Epidemiological Virology | KU Leuven, Rega Institute, Clinical and Epidemiological Virology |
| EPI_ISL_458222 | KU Leuven, Rega Institute, Clinical and Epidemiological Virology | KU Leuven, Rega Institute, Clinical and Epidemiological Virology |
| EPI_ISL_458223 | KU Leuven, Rega Institute, Clinical and Epidemiological Virology | KU Leuven, Rega Institute, Clinical and Epidemiological Virology |
| EPI_ISL_458224 | KU Leuven, Rega Institute, Clinical and Epidemiological Virology | KU Leuven, Rega Institute, Clinical and Epidemiological Virology |
| EPI_ISL_458225 | KU Leuven, Rega Institute, Clinical and Epidemiological Virology | KU Leuven, Rega Institute, Clinical and Epidemiological Virology |
| EPI_ISL_458226 | KU Leuven, Rega Institute, Clinical and Epidemiological Virology | KU Leuven, Rega Institute, Clinical and Epidemiological Virology |
| EPI_ISL_458227 | KU Leuven, Rega Institute, Clinical and Epidemiological Virology | KU Leuven, Rega Institute, Clinical and Epidemiological Virology |

| EPI_Accession_ID | Originating Laboratory | Submitting Laboratory |
|---|---|---|
| EPI_ISL_458228 | KU Leuven, Rega Institute, Clinical and Epidemiological Virology | KU Leuven, Rega Institute, Clinical and Epidemiological Virology |
| EPI_ISL_458229 | KU Leuven, Rega Institute, Clinical and Epidemiological Virology | KU Leuven, Rega Institute, Clinical and Epidemiological Virology |
| EPI_ISL_458230 | KU Leuven, Rega Institute, Clinical and Epidemiological Virology | KU Leuven, Rega Institute, Clinical and Epidemiological Virology |
| EPI_ISL_458231 | KU Leuven, Rega Institute, Clinical and Epidemiological Virology | KU Leuven, Rega Institute, Clinical and Epidemiological Virology |
| EPI_ISL_458232 | KU Leuven, Rega Institute, Clinical and Epidemiological Virology | KU Leuven, Rega Institute, Clinical and Epidemiological Virology |
| EPI_ISL_458233 | KU Leuven, Rega Institute, Clinical and Epidemiological Virology | KU Leuven, Rega Institute, Clinical and Epidemiological Virology |
| EPI_ISL_458234 | KU Leuven, Rega Institute, Clinical and Epidemiological Virology | KU Leuven, Rega Institute, Clinical and Epidemiological Virology |
| EPI_ISL_458235 | KU Leuven, Rega Institute, Clinical and Epidemiological Virology | KU Leuven, Rega Institute, Clinical and Epidemiological Virology |
| EPI_ISL_458236 | Hospital Mexico | CharitÃ© Virology-University of Costa Rica |
| EPI_ISL_458237 | Scripps Medical Laboratory | Andersen lab at Scripps Research |
| EPI_ISL_458238 | Scripps Medical Laboratory | Andersen lab at Scripps Research |
| EPI_ISL_458239 | Scripps Medical Laboratory | Andersen lab at Scripps Research |
| EPI_ISL_458240 | Scripps Medical Laboratory | Andersen lab at Scripps Research |
| EPI_ISL_458241 | Scripps Medical Laboratory | Andersen lab at Scripps Research |
| EPI_ISL_458242 | Scripps Medical Laboratory | Andersen lab at Scripps Research |
| EPI_ISL_458243 | Scripps Medical Laboratory | Andersen lab at Scripps Research |
| EPI_ISL_458244 | Scripps Medical Laboratory | Andersen lab at Scripps Research |
| EPI_ISL_458245 | Scripps Medical Laboratory | Andersen lab at Scripps Research |
| EPI_ISL_458246 | Scripps Medical Laboratory | Andersen lab at Scripps Research |
| EPI_ISL_458247 | Scripps Medical Laboratory | Andersen lab at Scripps Research |
| EPI_ISL_458248 | Scripps Medical Laboratory | Andersen lab at Scripps Research |
| EPI_ISL_458249 | Scripps Medical Laboratory | Andersen lab at Scripps Research |
| EPI_ISL_458250 | Scripps Medical Laboratory | Andersen lab at Scripps Research |
| EPI_ISL_458251 | Scripps Medical Laboratory | Andersen lab at Scripps Research |
| EPI_ISL_458252 | Scripps Medical Laboratory | Andersen lab at Scripps Research |

| EPI_Accession_ID | Originating Laboratory | Submitting Laboratory |
|---|---|---|
| EPI_ISL_458253 | Scripps Medical Laboratory | Andersen lab at Scripps Research |
| EPI_ISL_458254 | Scripps Medical Laboratory | Andersen lab at Scripps Research |
| EPI_ISL_458255 | Scripps Medical Laboratory | Andersen lab at Scripps Research |
| EPI_ISL_458256 | Scripps Medical Laboratory | Andersen lab at Scripps Research |
| EPI_ISL_458257 | Scripps Medical Laboratory | Andersen lab at Scripps Research |
| EPI_ISL_458258 | Scripps Medical Laboratory | Andersen lab at Scripps Research |
| EPI_ISL_458259 | Scripps Medical Laboratory | Andersen lab at Scripps Research |
| EPI_ISL_458260 | Scripps Medical Laboratory | Andersen lab at Scripps Research |
| EPI_ISL_458261 | Scripps Medical Laboratory | Andersen lab at Scripps Research |
| EPI_ISL_458262 | Scripps Medical Laboratory | Andersen lab at Scripps Research |
| EPI_ISL_458263 | Scripps Medical Laboratory | Andersen lab at Scripps Research |
| EPI_ISL_458264 | Scripps Medical Laboratory | Andersen lab at Scripps Research |
| EPI_ISL_458265 | Scripps Medical Laboratory | Andersen lab at Scripps Research |
| EPI_ISL_458266 | Scripps Medical Laboratory | Andersen lab at Scripps Research |
| EPI_ISL_458267 | Scripps Medical Laboratory | Andersen lab at Scripps Research |
| EPI_ISL_458268 | Scripps Medical Laboratory | Andersen lab at Scripps Research |
| EPI_ISL_458269 | Scripps Medical Laboratory | Andersen lab at Scripps Research |
| EPI_ISL_458270 | Scripps Medical Laboratory | Andersen lab at Scripps Research |
| EPI_ISL_458271 | Scripps Medical Laboratory | Andersen lab at Scripps Research |
| EPI_ISL_458272 | Scripps Medical Laboratory | Andersen lab at Scripps Research |
| EPI_ISL_458273 | Scripps Medical Laboratory | Andersen lab at Scripps Research |
| EPI_ISL_458274 | Scripps Medical Laboratory | Andersen lab at Scripps Research |
| EPI_ISL_458275 | Scripps Medical Laboratory | Andersen lab at Scripps Research |
| EPI_ISL_458276 | Scripps Medical Laboratory | Andersen lab at Scripps Research |
| EPI_ISL_458277 | Scripps Medical Laboratory | Andersen lab at Scripps Research |
| EPI_ISL_458278 | Scripps Medical Laboratory | Andersen lab at Scripps Research |
| EPI_ISL_458279 | Scripps Medical Laboratory | Andersen lab at Scripps Research |
| EPI_ISL_458280 | Scripps Medical Laboratory | Andersen lab at Scripps Research |
| EPI_ISL_458281 | Scripps Medical Laboratory | Andersen lab at Scripps Research |
| EPI_ISL_458282 | Scripps Medical Laboratory | Andersen lab at Scripps Research |
| EPI_ISL_458283 | Scripps Medical Laboratory | Andersen lab at Scripps Research |
| EPI_ISL_458284 | Scripps Medical Laboratory | Andersen lab at Scripps Research |
| EPI_ISL_458285 | unknown | Bundeswehr Institute of Microbiology |

| EPI_Accession_ID | Originating Laboratory | Submitting Laboratory |
|---|---|---|
| EPI_ISL_458286 | unknown | Bundeswehr Institute of Microbiology |
| EPI_ISL_458298 | CSIR-Centre for Cellular and Molecular Biology | CSIR-Centre for Cellular and Molecular Biology |
| EPI_ISL_458299 | PHE South West Regional Laboratory, National Infection Service | Wellcome Sanger Institute for the COVID-19 Genomics UK Consortium |
| EPI_ISL_458300 | PHE South West Regional Laboratory, National Infection Service | Wellcome Sanger Institute for the COVID-19 Genomics UK Consortium |
| EPI_ISL_458301 | PHE South West Regional Laboratory, National Infection Service | Wellcome Sanger Institute for the COVID-19 Genomics UK Consortium |
| EPI_ISL_458302 | PHE South West Regional Laboratory, National Infection Service | Wellcome Sanger Institute for the COVID-19 Genomics UK Consortium |
| EPI_ISL_458303 | PHE South West Regional Laboratory, National Infection Service | Wellcome Sanger Institute for the COVID-19 Genomics UK Consortium |
| EPI_ISL_458304 | PHE South West Regional Laboratory, National Infection Service | Wellcome Sanger Institute for the COVID-19 Genomics UK Consortium |
| EPI_ISL_458305 | PHE South West Regional Laboratory, National Infection Service | Wellcome Sanger Institute for the COVID-19 Genomics UK Consortium |
| EPI_ISL_458306 | PHE South West Regional Laboratory, National Infection Service | Wellcome Sanger Institute for the COVID-19 Genomics UK Consortium |
| EPI_ISL_458307 | PHE South West Regional Laboratory, National Infection Service | Wellcome Sanger Institute for the COVID-19 Genomics UK Consortium |
| EPI_ISL_458308 | PHE South West Regional Laboratory, National Infection Service | Wellcome Sanger Institute for the COVID-19 Genomics UK Consortium |
| EPI_ISL_458309 | PHE South West Regional Laboratory, National Infection Service | Wellcome Sanger Institute for the COVID-19 Genomics UK Consortium |
| EPI_ISL_458310 | PHE South West Regional Laboratory, National Infection Service | Wellcome Sanger Institute for the COVID-19 Genomics UK Consortium |
| EPI_ISL_458311 | PHE South West Regional Laboratory, National Infection Service | Wellcome Sanger Institute for the COVID-19 Genomics UK Consortium |
| EPI_ISL_458312 | PHE South West Regional Laboratory, National Infection Service | Wellcome Sanger Institute for the COVID-19 Genomics UK Consortium |
| EPI_ISL_458313 | PHE South West Regional Laboratory, National Infection Service | Wellcome Sanger Institute for the COVID-19 Genomics UK Consortium |

| EPI_Accession_ID | Originating Laboratory | Submitting Laboratory |
|---|---|---|
| EPI_ISL_458314 | PHE South West Regional Laboratory, National Infection Service | Wellcome Sanger Institute for the COVID-19 Genomics UK Consortium |
| EPI_ISL_458315 | PHE South West Regional Laboratory, National Infection Service | Wellcome Sanger Institute for the COVID-19 Genomics UK Consortium |
| EPI_ISL_458316 | PHE South West Regional Laboratory, National Infection Service | Wellcome Sanger Institute for the COVID-19 Genomics UK Consortium |
| EPI_ISL_458317 | PHE South West Regional Laboratory, National Infection Service | Wellcome Sanger Institute for the COVID-19 Genomics UK Consortium |
| EPI_ISL_458318 | PHE South West Regional Laboratory, National Infection Service | Wellcome Sanger Institute for the COVID-19 Genomics UK Consortium |
| EPI_ISL_458319 | PHE South West Regional Laboratory, National Infection Service | Wellcome Sanger Institute for the COVID-19 Genomics UK Consortium |
| EPI_ISL_458320 | PHE South West Regional Laboratory, National Infection Service | Wellcome Sanger Institute for the COVID-19 Genomics UK Consortium |
| EPI_ISL_458321 | PHE South West Regional Laboratory, National Infection Service | Wellcome Sanger Institute for the COVID-19 Genomics UK Consortium |
| EPI_ISL_458322 | PHE South West Regional Laboratory, National Infection Service | Wellcome Sanger Institute for the COVID-19 Genomics UK Consortium |
| EPI_ISL_458323 | PHE South West Regional Laboratory, National Infection Service | Wellcome Sanger Institute for the COVID-19 Genomics UK Consortium |
| EPI_ISL_458324 | PHE South West Regional Laboratory, National Infection Service | Wellcome Sanger Institute for the COVID-19 Genomics UK Consortium |
| EPI_ISL_458325 | PHE South West Regional Laboratory, National Infection Service | Wellcome Sanger Institute for the COVID-19 Genomics UK Consortium |
| EPI_ISL_458326 | PHE South West Regional Laboratory, National Infection Service | Wellcome Sanger Institute for the COVID-19 Genomics UK Consortium |
| EPI_ISL_458327 | PHE South West Regional Laboratory, National Infection Service | Wellcome Sanger Institute for the COVID-19 Genomics UK Consortium |
| EPI_ISL_458328 | PHE South West Regional Laboratory, National Infection Service | Wellcome Sanger Institute for the COVID-19 Genomics UK Consortium |
| EPI_ISL_458329 | PHE South West Regional Laboratory, National Infection Service | Wellcome Sanger Institute for the COVID-19 Genomics UK Consortium |

| EPI_Accession_ID | Originating Laboratory | Submitting Laboratory |
|---|---|---|
| EPI_ISL_458330 | PHE South West Regional Laboratory, National Infection Service | Wellcome Sanger Institute for the COVID-19 Genomics UK Consortium |
| EPI_ISL_458331 | PHE South West Regional Laboratory, National Infection Service | Wellcome Sanger Institute for the COVID-19 Genomics UK Consortium |
| EPI_ISL_458332 | PHE South West Regional Laboratory, National Infection Service | Wellcome Sanger Institute for the COVID-19 Genomics UK Consortium |
| EPI_ISL_458333 | PHE South West Regional Laboratory, National Infection Service | Wellcome Sanger Institute for the COVID-19 Genomics UK Consortium |
| EPI_ISL_458334 | PHE South West Regional Laboratory, National Infection Service | Wellcome Sanger Institute for the COVID-19 Genomics UK Consortium |
| EPI_ISL_458335 | PHE South West Regional Laboratory, National Infection Service | Wellcome Sanger Institute for the COVID-19 Genomics UK Consortium |
| EPI_ISL_458336 | PHE South West Regional Laboratory, National Infection Service | Wellcome Sanger Institute for the COVID-19 Genomics UK Consortium |
| EPI_ISL_458337 | PHE South West Regional Laboratory, National Infection Service | Wellcome Sanger Institute for the COVID-19 Genomics UK Consortium |
| EPI_ISL_458338 | PHE South West Regional Laboratory, National Infection Service | Wellcome Sanger Institute for the COVID-19 Genomics UK Consortium |
| EPI_ISL_458339 | PHE South West Regional Laboratory, National Infection Service | Wellcome Sanger Institute for the COVID-19 Genomics UK Consortium |
| EPI_ISL_458340 | PHE South West Regional Laboratory, National Infection Service | Wellcome Sanger Institute for the COVID-19 Genomics UK Consortium |
| EPI_ISL_458341 | PHE South West Regional Laboratory, National Infection Service | Wellcome Sanger Institute for the COVID-19 Genomics UK Consortium |
| EPI_ISL_458342 | PHE South West Regional Laboratory, National Infection Service | Wellcome Sanger Institute for the COVID-19 Genomics UK Consortium |
| EPI_ISL_458343 | PHE South West Regional Laboratory, National Infection Service | Wellcome Sanger Institute for the COVID-19 Genomics UK Consortium |
| EPI_ISL_458344 | PHE South West Regional Laboratory, National Infection Service | Wellcome Sanger Institute for the COVID-19 Genomics UK Consortium |
| EPI_ISL_458345 | PHE South West Regional Laboratory, National Infection Service | Wellcome Sanger Institute for the COVID-19 Genomics UK Consortium |

| EPI_Accession_ID | Originating Laboratory | Submitting Laboratory |
|---|---|---|
| EPI_ISL_458346 | PHE South West Regional Laboratory, National Infection Service | Wellcome Sanger Institute for the COVID-19 Genomics UK Consortium |
| EPI_ISL_458347 | PHE South West Regional Laboratory, National Infection Service | Wellcome Sanger Institute for the COVID-19 Genomics UK Consortium |
| EPI_ISL_458348 | PHE South West Regional Laboratory, National Infection Service | Wellcome Sanger Institute for the COVID-19 Genomics UK Consortium |
| EPI_ISL_458349 | PHE South West Regional Laboratory, National Infection Service | Wellcome Sanger Institute for the COVID-19 Genomics UK Consortium |
| EPI_ISL_458350 | PHE South West Regional Laboratory, National Infection Service | Wellcome Sanger Institute for the COVID-19 Genomics UK Consortium |
| EPI_ISL_458351 | PHE South West Regional Laboratory, National Infection Service | Wellcome Sanger Institute for the COVID-19 Genomics UK Consortium |
| EPI_ISL_458352 | PHE South West Regional Laboratory, National Infection Service | Wellcome Sanger Institute for the COVID-19 Genomics UK Consortium |
| EPI_ISL_458353 | PHE South West Regional Laboratory, National Infection Service | Wellcome Sanger Institute for the COVID-19 Genomics UK Consortium |
| EPI_ISL_458354 | PHE South West Regional Laboratory, National Infection Service | Wellcome Sanger Institute for the COVID-19 Genomics UK Consortium |
| EPI_ISL_458355 | PHE South West Regional Laboratory, National Infection Service | Wellcome Sanger Institute for the COVID-19 Genomics UK Consortium |
| EPI_ISL_458356 | PHE South West Regional Laboratory, National Infection Service | Wellcome Sanger Institute for the COVID-19 Genomics UK Consortium |
| EPI_ISL_458357 | PHE South West Regional Laboratory, National Infection Service | Wellcome Sanger Institute for the COVID-19 Genomics UK Consortium |
| EPI_ISL_458358 | PHE South West Regional Laboratory, National Infection Service | Wellcome Sanger Institute for the COVID-19 Genomics UK Consortium |
| EPI_ISL_458359 | PHE South West Regional Laboratory, National Infection Service | Wellcome Sanger Institute for the COVID-19 Genomics UK Consortium |
| EPI_ISL_458360 | PHE South West Regional Laboratory, National Infection Service | Wellcome Sanger Institute for the COVID-19 Genomics UK Consortium |
| EPI_ISL_458361 | PHE South West Regional Laboratory, National Infection Service | Wellcome Sanger Institute for the COVID-19 Genomics UK Consortium |

| EPI_Accession_ID | Originating Laboratory | Submitting Laboratory |
| --- | --- | --- |
| EPI_ISL_458362 | PHE South West Regional Laboratory, National Infection Service | Wellcome Sanger Institute for the COVID-19 Genomics UK Consortium |
| EPI_ISL_458363 | PHE South West Regional Laboratory, National Infection Service | Wellcome Sanger Institute for the COVID-19 Genomics UK Consortium |
| EPI_ISL_458364 | PHE South West Regional Laboratory, National Infection Service | Wellcome Sanger Institute for the COVID-19 Genomics UK Consortium |
| EPI_ISL_458365 | PHE South West Regional Laboratory, National Infection Service | Wellcome Sanger Institute for the COVID-19 Genomics UK Consortium |
| EPI_ISL_458366 | PHE South West Regional Laboratory, National Infection Service | Wellcome Sanger Institute for the COVID-19 Genomics UK Consortium |
| EPI_ISL_458367 | PHE South West Regional Laboratory, National Infection Service | Wellcome Sanger Institute for the COVID-19 Genomics UK Consortium |
| EPI_ISL_458368 | PHE South West Regional Laboratory, National Infection Service | Wellcome Sanger Institute for the COVID-19 Genomics UK Consortium |
| EPI_ISL_458369 | PHE South West Regional Laboratory, National Infection Service | Wellcome Sanger Institute for the COVID-19 Genomics UK Consortium |
| EPI_ISL_458370 | PHE South West Regional Laboratory, National Infection Service | Wellcome Sanger Institute for the COVID-19 Genomics UK Consortium |
| EPI_ISL_458371 | PHE South West Regional Laboratory, National Infection Service | Wellcome Sanger Institute for the COVID-19 Genomics UK Consortium |
| EPI_ISL_458372 | PHE South West Regional Laboratory, National Infection Service | Wellcome Sanger Institute for the COVID-19 Genomics UK Consortium |
| EPI_ISL_458373 | PHE South West Regional Laboratory, National Infection Service | Wellcome Sanger Institute for the COVID-19 Genomics UK Consortium |
| EPI_ISL_458374 | PHE South West Regional Laboratory, National Infection Service | Wellcome Sanger Institute for the COVID-19 Genomics UK Consortium |
| EPI_ISL_458375 | PHE South West Regional Laboratory, National Infection Service | Wellcome Sanger Institute for the COVID-19 Genomics UK Consortium |
| EPI_ISL_458376 | PHE South West Regional Laboratory, National Infection Service | Wellcome Sanger Institute for the COVID-19 Genomics UK Consortium |
| EPI_ISL_458377 | PHE South West Regional Laboratory, National Infection Service | Wellcome Sanger Institute for the COVID-19 Genomics UK Consortium |

| EPI_Accession_ID | Originating Laboratory | Submitting Laboratory |
|---|---|---|
| EPI_ISL_458378 | PHE South West Regional Laboratory, National Infection Service | Wellcome Sanger Institute for the COVID-19 Genomics UK Consortium |
| EPI_ISL_458379 | PHE South West Regional Laboratory, National Infection Service | Wellcome Sanger Institute for the COVID-19 Genomics UK Consortium |
| EPI_ISL_458380 | PHE South West Regional Laboratory, National Infection Service | Wellcome Sanger Institute for the COVID-19 Genomics UK Consortium |
| EPI_ISL_458381 | PHE South West Regional Laboratory, National Infection Service | Wellcome Sanger Institute for the COVID-19 Genomics UK Consortium |
| EPI_ISL_458382 | PHE South West Regional Laboratory, National Infection Service | Wellcome Sanger Institute for the COVID-19 Genomics UK Consortium |
| EPI_ISL_458383 | PHE South West Regional Laboratory, National Infection Service | Wellcome Sanger Institute for the COVID-19 Genomics UK Consortium |
| EPI_ISL_458384 | PHE South West Regional Laboratory, National Infection Service | Wellcome Sanger Institute for the COVID-19 Genomics UK Consortium |
| EPI_ISL_458385 | PHE South West Regional Laboratory, National Infection Service | Wellcome Sanger Institute for the COVID-19 Genomics UK Consortium |
| EPI_ISL_458386 | PHE South West Regional Laboratory, National Infection Service | Wellcome Sanger Institute for the COVID-19 Genomics UK Consortium |
| EPI_ISL_458387 | PHE South West Regional Laboratory, National Infection Service | Wellcome Sanger Institute for the COVID-19 Genomics UK Consortium |
| EPI_ISL_458388 | PHE South West Regional Laboratory, National Infection Service | Wellcome Sanger Institute for the COVID-19 Genomics UK Consortium |
| EPI_ISL_458389 | PHE South West Regional Laboratory, National Infection Service | Wellcome Sanger Institute for the COVID-19 Genomics UK Consortium |
| EPI_ISL_458390 | PHE South West Regional Laboratory, National Infection Service | Wellcome Sanger Institute for the COVID-19 Genomics UK Consortium |
| EPI_ISL_458391 | PHE South West Regional Laboratory, National Infection Service | Wellcome Sanger Institute for the COVID-19 Genomics UK Consortium |
| EPI_ISL_458392 | PHE South West Regional Laboratory, National Infection Service | Wellcome Sanger Institute for the COVID-19 Genomics UK Consortium |
| EPI_ISL_458393 | PHE South West Regional Laboratory, National Infection Service | Wellcome Sanger Institute for the COVID-19 Genomics UK Consortium |

| EPI_Accession_ID | Originating Laboratory | Submitting Laboratory |
|---|---|---|
| EPI_ISL_458394 | PHE South West Regional Laboratory, National Infection Service | Wellcome Sanger Institute for the COVID-19 Genomics UK Consortium |
| EPI_ISL_458395 | PHE South West Regional Laboratory, National Infection Service | Wellcome Sanger Institute for the COVID-19 Genomics UK Consortium |
| EPI_ISL_458396 | PHE South West Regional Laboratory, National Infection Service | Wellcome Sanger Institute for the COVID-19 Genomics UK Consortium |
| EPI_ISL_458397 | PHE South West Regional Laboratory, National Infection Service | Wellcome Sanger Institute for the COVID-19 Genomics UK Consortium |
| EPI_ISL_458398 | PHE South West Regional Laboratory, National Infection Service | Wellcome Sanger Institute for the COVID-19 Genomics UK Consortium |
| EPI_ISL_458399 | PHE South West Regional Laboratory, National Infection Service | Wellcome Sanger Institute for the COVID-19 Genomics UK Consortium |
| EPI_ISL_458400 | PHE South West Regional Laboratory, National Infection Service | Wellcome Sanger Institute for the COVID-19 Genomics UK Consortium |
| EPI_ISL_458401 | PHE South West Regional Laboratory, National Infection Service | Wellcome Sanger Institute for the COVID-19 Genomics UK Consortium |
| EPI_ISL_458402 | PHE South West Regional Laboratory, National Infection Service | Wellcome Sanger Institute for the COVID-19 Genomics UK Consortium |
| EPI_ISL_458403 | PHE South West Regional Laboratory, National Infection Service | Wellcome Sanger Institute for the COVID-19 Genomics UK Consortium |
| EPI_ISL_458404 | PHE South West Regional Laboratory, National Infection Service | Wellcome Sanger Institute for the COVID-19 Genomics UK Consortium |
| EPI_ISL_458405 | PHE South West Regional Laboratory, National Infection Service | Wellcome Sanger Institute for the COVID-19 Genomics UK Consortium |
| EPI_ISL_458406 | PHE South West Regional Laboratory, National Infection Service | Wellcome Sanger Institute for the COVID-19 Genomics UK Consortium |
| EPI_ISL_458407 | PHE South West Regional Laboratory, National Infection Service | Wellcome Sanger Institute for the COVID-19 Genomics UK Consortium |
| EPI_ISL_458408 | PHE South West Regional Laboratory, National Infection Service | Wellcome Sanger Institute for the COVID-19 Genomics UK Consortium |
| EPI_ISL_458409 | PHE South West Regional Laboratory, National Infection Service | Wellcome Sanger Institute for the COVID-19 Genomics UK Consortium |

| EPI_Accession_ID | Originating Laboratory | Submitting Laboratory |
|---|---|---|
| EPI_ISL_458410 | PHE South West Regional Laboratory, National Infection Service | Wellcome Sanger Institute for the COVID-19 Genomics UK Consortium |
| EPI_ISL_458411 | PHE South West Regional Laboratory, National Infection Service | Wellcome Sanger Institute for the COVID-19 Genomics UK Consortium |
| EPI_ISL_458412 | PHE South West Regional Laboratory, National Infection Service | Wellcome Sanger Institute for the COVID-19 Genomics UK Consortium |
| EPI_ISL_458413 | PHE South West Regional Laboratory, National Infection Service | Wellcome Sanger Institute for the COVID-19 Genomics UK Consortium |
| EPI_ISL_458414 | PHE South West Regional Laboratory, National Infection Service | Wellcome Sanger Institute for the COVID-19 Genomics UK Consortium |
| EPI_ISL_458415 | PHE South West Regional Laboratory, National Infection Service | Wellcome Sanger Institute for the COVID-19 Genomics UK Consortium |
| EPI_ISL_458416 | PHE South West Regional Laboratory, National Infection Service | Wellcome Sanger Institute for the COVID-19 Genomics UK Consortium |
| EPI_ISL_458417 | PHE South West Regional Laboratory, National Infection Service | Wellcome Sanger Institute for the COVID-19 Genomics UK Consortium |
| EPI_ISL_458418 | PHE South West Regional Laboratory, National Infection Service | Wellcome Sanger Institute for the COVID-19 Genomics UK Consortium |
| EPI_ISL_458419 | PHE South West Regional Laboratory, National Infection Service | Wellcome Sanger Institute for the COVID-19 Genomics UK Consortium |
| EPI_ISL_458420 | PHE South West Regional Laboratory, National Infection Service | Wellcome Sanger Institute for the COVID-19 Genomics UK Consortium |
| EPI_ISL_458421 | PHE South West Regional Laboratory, National Infection Service | Wellcome Sanger Institute for the COVID-19 Genomics UK Consortium |
| EPI_ISL_458422 | PHE South West Regional Laboratory, National Infection Service | Wellcome Sanger Institute for the COVID-19 Genomics UK Consortium |
| EPI_ISL_458423 | PHE South West Regional Laboratory, National Infection Service | Wellcome Sanger Institute for the COVID-19 Genomics UK Consortium |
| EPI_ISL_458424 | PHE South West Regional Laboratory, National Infection Service | Wellcome Sanger Institute for the COVID-19 Genomics UK Consortium |
| EPI_ISL_458425 | PHE South West Regional Laboratory, National Infection Service | Wellcome Sanger Institute for the COVID-19 Genomics UK Consortium |

| EPI_Accession_ID | Originating Laboratory | Submitting Laboratory |
|---|---|---|
| EPI_ISL_458426 | PHE South West Regional Laboratory, National Infection Service | Wellcome Sanger Institute for the COVID-19 Genomics UK Consortium |
| EPI_ISL_458427 | PHE South West Regional Laboratory, National Infection Service | Wellcome Sanger Institute for the COVID-19 Genomics UK Consortium |
| EPI_ISL_458428 | PHE South West Regional Laboratory, National Infection Service | Wellcome Sanger Institute for the COVID-19 Genomics UK Consortium |
| EPI_ISL_458429 | PHE South West Regional Laboratory, National Infection Service | Wellcome Sanger Institute for the COVID-19 Genomics UK Consortium |
| EPI_ISL_458430 | PHE South West Regional Laboratory, National Infection Service | Wellcome Sanger Institute for the COVID-19 Genomics UK Consortium |
| EPI_ISL_458431 | PHE South West Regional Laboratory, National Infection Service | Wellcome Sanger Institute for the COVID-19 Genomics UK Consortium |
| EPI_ISL_458432 | PHE South West Regional Laboratory, National Infection Service | Wellcome Sanger Institute for the COVID-19 Genomics UK Consortium |
| EPI_ISL_458433 | PHE South West Regional Laboratory, National Infection Service | Wellcome Sanger Institute for the COVID-19 Genomics UK Consortium |
| EPI_ISL_458434 | PHE South West Regional Laboratory, National Infection Service | Wellcome Sanger Institute for the COVID-19 Genomics UK Consortium |
| EPI_ISL_458435 | PHE South West Regional Laboratory, National Infection Service | Wellcome Sanger Institute for the COVID-19 Genomics UK Consortium |
| EPI_ISL_458436 | PHE South West Regional Laboratory, National Infection Service | Wellcome Sanger Institute for the COVID-19 Genomics UK Consortium |
| EPI_ISL_458437 | PHE South West Regional Laboratory, National Infection Service | Wellcome Sanger Institute for the COVID-19 Genomics UK Consortium |
| EPI_ISL_458438 | PHE South West Regional Laboratory, National Infection Service | Wellcome Sanger Institute for the COVID-19 Genomics UK Consortium |
| EPI_ISL_458439 | PHE South West Regional Laboratory, National Infection Service | Wellcome Sanger Institute for the COVID-19 Genomics UK Consortium |
| EPI_ISL_458440 | PHE South West Regional Laboratory, National Infection Service | Wellcome Sanger Institute for the COVID-19 Genomics UK Consortium |
| EPI_ISL_458441 | PHE South West Regional Laboratory, National Infection Service | Wellcome Sanger Institute for the COVID-19 Genomics UK Consortium |

| EPI_Accession_ID | Originating Laboratory | Submitting Laboratory |
|---|---|---|
| EPI_ISL_458442 | PHE South West Regional Laboratory, National Infection Service | Wellcome Sanger Institute for the COVID-19 Genomics UK Consortium |
| EPI_ISL_458443 | PHE South West Regional Laboratory, National Infection Service | Wellcome Sanger Institute for the COVID-19 Genomics UK Consortium |
| EPI_ISL_458444 | PHE South West Regional Laboratory, National Infection Service | Wellcome Sanger Institute for the COVID-19 Genomics UK Consortium |
| EPI_ISL_458445 | PHE South West Regional Laboratory, National Infection Service | Wellcome Sanger Institute for the COVID-19 Genomics UK Consortium |
| EPI_ISL_458446 | PHE South West Regional Laboratory, National Infection Service | Wellcome Sanger Institute for the COVID-19 Genomics UK Consortium |
| EPI_ISL_458447 | PHE South West Regional Laboratory, National Infection Service | Wellcome Sanger Institute for the COVID-19 Genomics UK Consortium |
| EPI_ISL_458448 | PHE South West Regional Laboratory, National Infection Service | Wellcome Sanger Institute for the COVID-19 Genomics UK Consortium |
| EPI_ISL_458449 | PHE South West Regional Laboratory, National Infection Service | Wellcome Sanger Institute for the COVID-19 Genomics UK Consortium |
| EPI_ISL_458450 | PHE South West Regional Laboratory, National Infection Service | Wellcome Sanger Institute for the COVID-19 Genomics UK Consortium |
| EPI_ISL_458451 | PHE South West Regional Laboratory, National Infection Service | Wellcome Sanger Institute for the COVID-19 Genomics UK Consortium |
| EPI_ISL_458452 | PHE South West Regional Laboratory, National Infection Service | Wellcome Sanger Institute for the COVID-19 Genomics UK Consortium |
| EPI_ISL_458453 | PHE South West Regional Laboratory, National Infection Service | Wellcome Sanger Institute for the COVID-19 Genomics UK Consortium |
| EPI_ISL_458454 | PHE South West Regional Laboratory, National Infection Service | Wellcome Sanger Institute for the COVID-19 Genomics UK Consortium |
| EPI_ISL_458455 | PHE South West Regional Laboratory, National Infection Service | Wellcome Sanger Institute for the COVID-19 Genomics UK Consortium |
| EPI_ISL_458456 | PHE South West Regional Laboratory, National Infection Service | Wellcome Sanger Institute for the COVID-19 Genomics UK Consortium |
| EPI_ISL_458457 | PHE South West Regional Laboratory, National Infection Service | Wellcome Sanger Institute for the COVID-19 Genomics UK Consortium |

| EPI_Accession_ID | Originating Laboratory | Submitting Laboratory |
|---|---|---|
| EPI_ISL_458458 | PHE South West Regional Laboratory, National Infection Service | Wellcome Sanger Institute for the COVID-19 Genomics UK Consortium |
| EPI_ISL_458459 | PHE South West Regional Laboratory, National Infection Service | Wellcome Sanger Institute for the COVID-19 Genomics UK Consortium |
| EPI_ISL_458460 | PHE South West Regional Laboratory, National Infection Service | Wellcome Sanger Institute for the COVID-19 Genomics UK Consortium |
| EPI_ISL_458461 | PHE South West Regional Laboratory, National Infection Service | Wellcome Sanger Institute for the COVID-19 Genomics UK Consortium |
| EPI_ISL_458462 | PHE South West Regional Laboratory, National Infection Service | Wellcome Sanger Institute for the COVID-19 Genomics UK Consortium |
| EPI_ISL_458463 | PHE South West Regional Laboratory, National Infection Service | Wellcome Sanger Institute for the COVID-19 Genomics UK Consortium |
| EPI_ISL_458464 | PHE South West Regional Laboratory, National Infection Service | Wellcome Sanger Institute for the COVID-19 Genomics UK Consortium |
| EPI_ISL_458465 | PHE South West Regional Laboratory, National Infection Service | Wellcome Sanger Institute for the COVID-19 Genomics UK Consortium |
| EPI_ISL_458466 | PHE South West Regional Laboratory, National Infection Service | Wellcome Sanger Institute for the COVID-19 Genomics UK Consortium |
| EPI_ISL_458467 | PHE South West Regional Laboratory, National Infection Service | Wellcome Sanger Institute for the COVID-19 Genomics UK Consortium |
| EPI_ISL_458468 | PHE South West Regional Laboratory, National Infection Service | Wellcome Sanger Institute for the COVID-19 Genomics UK Consortium |
| EPI_ISL_458469 | PHE South West Regional Laboratory, National Infection Service | Wellcome Sanger Institute for the COVID-19 Genomics UK Consortium |
| EPI_ISL_458470 | PHE South West Regional Laboratory, National Infection Service | Wellcome Sanger Institute for the COVID-19 Genomics UK Consortium |
| EPI_ISL_458471 | PHE South West Regional Laboratory, National Infection Service | Wellcome Sanger Institute for the COVID-19 Genomics UK Consortium |
| EPI_ISL_458472 | PHE South West Regional Laboratory, National Infection Service | Wellcome Sanger Institute for the COVID-19 Genomics UK Consortium |
| EPI_ISL_458473 | PHE South West Regional Laboratory, National Infection Service | Wellcome Sanger Institute for the COVID-19 Genomics UK Consortium |

| EPI_Accession_ID | Originating Laboratory | Submitting Laboratory |
|---|---|---|
| EPI_ISL_458474 | PHE South West Regional Laboratory, National Infection Service | Wellcome Sanger Institute for the COVID-19 Genomics UK Consortium |
| EPI_ISL_458475 | PHE South West Regional Laboratory, National Infection Service | Wellcome Sanger Institute for the COVID-19 Genomics UK Consortium |
| EPI_ISL_458476 | PHE South West Regional Laboratory, National Infection Service | Wellcome Sanger Institute for the COVID-19 Genomics UK Consortium |
| EPI_ISL_458477 | PHE South West Regional Laboratory, National Infection Service | Wellcome Sanger Institute for the COVID-19 Genomics UK Consortium |
| EPI_ISL_458478 | PHE South West Regional Laboratory, National Infection Service | Wellcome Sanger Institute for the COVID-19 Genomics UK Consortium |
| EPI_ISL_458479 | PHE South West Regional Laboratory, National Infection Service | Wellcome Sanger Institute for the COVID-19 Genomics UK Consortium |
| EPI_ISL_458480 | PHE South West Regional Laboratory, National Infection Service | Wellcome Sanger Institute for the COVID-19 Genomics UK Consortium |
| EPI_ISL_458481 | PHE South West Regional Laboratory, National Infection Service | Wellcome Sanger Institute for the COVID-19 Genomics UK Consortium |
| EPI_ISL_458482 | PHE South West Regional Laboratory, National Infection Service | Wellcome Sanger Institute for the COVID-19 Genomics UK Consortium |
| EPI_ISL_458483 | PHE South West Regional Laboratory, National Infection Service | Wellcome Sanger Institute for the COVID-19 Genomics UK Consortium |
| EPI_ISL_458484 | PHE South West Regional Laboratory, National Infection Service | Wellcome Sanger Institute for the COVID-19 Genomics UK Consortium |
| EPI_ISL_458485 | PHE South West Regional Laboratory, National Infection Service | Wellcome Sanger Institute for the COVID-19 Genomics UK Consortium |
| EPI_ISL_458486 | PHE South West Regional Laboratory, National Infection Service | Wellcome Sanger Institute for the COVID-19 Genomics UK Consortium |
| EPI_ISL_458487 | PHE South West Regional Laboratory, National Infection Service | Wellcome Sanger Institute for the COVID-19 Genomics UK Consortium |
| EPI_ISL_458488 | PHE South West Regional Laboratory, National Infection Service | Wellcome Sanger Institute for the COVID-19 Genomics UK Consortium |
| EPI_ISL_458489 | PHE South West Regional Laboratory, National Infection Service | Wellcome Sanger Institute for the COVID-19 Genomics UK Consortium |

| EPI_Accession_ID | Originating Laboratory | Submitting Laboratory |
|---|---|---|
| EPI_ISL_458490 | PHE South West Regional Laboratory, National Infection Service | Wellcome Sanger Institute for the COVID-19 Genomics UK Consortium |
| EPI_ISL_458491 | PHE South West Regional Laboratory, National Infection Service | Wellcome Sanger Institute for the COVID-19 Genomics UK Consortium |
| EPI_ISL_458492 | PHE South West Regional Laboratory, National Infection Service | Wellcome Sanger Institute for the COVID-19 Genomics UK Consortium |
| EPI_ISL_458493 | PHE South West Regional Laboratory, National Infection Service | Wellcome Sanger Institute for the COVID-19 Genomics UK Consortium |
| EPI_ISL_458494 | PHE South West Regional Laboratory, National Infection Service | Wellcome Sanger Institute for the COVID-19 Genomics UK Consortium |
| EPI_ISL_458495 | PHE South West Regional Laboratory, National Infection Service | Wellcome Sanger Institute for the COVID-19 Genomics UK Consortium |
| EPI_ISL_458496 | PHE South West Regional Laboratory, National Infection Service | Wellcome Sanger Institute for the COVID-19 Genomics UK Consortium |
| EPI_ISL_458497 | PHE South West Regional Laboratory, National Infection Service | Wellcome Sanger Institute for the COVID-19 Genomics UK Consortium |
| EPI_ISL_458498 | PHE South West Regional Laboratory, National Infection Service | Wellcome Sanger Institute for the COVID-19 Genomics UK Consortium |
| EPI_ISL_458499 | PHE South West Regional Laboratory, National Infection Service | Wellcome Sanger Institute for the COVID-19 Genomics UK Consortium |
| EPI_ISL_458500 | PHE South West Regional Laboratory, National Infection Service | Wellcome Sanger Institute for the COVID-19 Genomics UK Consortium |
| EPI_ISL_458501 | PHE South West Regional Laboratory, National Infection Service | Wellcome Sanger Institute for the COVID-19 Genomics UK Consortium |
| EPI_ISL_458502 | PHE South West Regional Laboratory, National Infection Service | Wellcome Sanger Institute for the COVID-19 Genomics UK Consortium |
| EPI_ISL_458503 | PHE South West Regional Laboratory, National Infection Service | Wellcome Sanger Institute for the COVID-19 Genomics UK Consortium |
| EPI_ISL_458504 | PHE South West Regional Laboratory, National Infection Service | Wellcome Sanger Institute for the COVID-19 Genomics UK Consortium |
| EPI_ISL_458505 | PHE South West Regional Laboratory, National Infection Service | Wellcome Sanger Institute for the COVID-19 Genomics UK Consortium |

| EPI_Accession_ID | Originating Laboratory | Submitting Laboratory |
|---|---|---|
| EPI_ISL_458506 | PHE South West Regional Laboratory, National Infection Service | Wellcome Sanger Institute for the COVID-19 Genomics UK Consortium |
| EPI_ISL_458507 | PHE South West Regional Laboratory, National Infection Service | Wellcome Sanger Institute for the COVID-19 Genomics UK Consortium |
| EPI_ISL_458508 | PHE South West Regional Laboratory, National Infection Service | Wellcome Sanger Institute for the COVID-19 Genomics UK Consortium |
| EPI_ISL_458509 | PHE South West Regional Laboratory, National Infection Service | Wellcome Sanger Institute for the COVID-19 Genomics UK Consortium |
| EPI_ISL_458510 | PHE South West Regional Laboratory, National Infection Service | Wellcome Sanger Institute for the COVID-19 Genomics UK Consortium |
| EPI_ISL_458511 | PHE South West Regional Laboratory, National Infection Service | Wellcome Sanger Institute for the COVID-19 Genomics UK Consortium |
| EPI_ISL_458512 | PHE South West Regional Laboratory, National Infection Service | Wellcome Sanger Institute for the COVID-19 Genomics UK Consortium |
| EPI_ISL_458513 | PHE South West Regional Laboratory, National Infection Service | Wellcome Sanger Institute for the COVID-19 Genomics UK Consortium |
| EPI_ISL_458514 | PHE South West Regional Laboratory, National Infection Service | Wellcome Sanger Institute for the COVID-19 Genomics UK Consortium |
| EPI_ISL_458515 | PHE South West Regional Laboratory, National Infection Service | Wellcome Sanger Institute for the COVID-19 Genomics UK Consortium |
| EPI_ISL_458516 | Department of Pathology, University of Cambridge | Wellcome Sanger Institute for the COVID-19 Genomics UK Consortium |
| EPI_ISL_458517 | Department of Pathology, University of Cambridge | Wellcome Sanger Institute for the COVID-19 Genomics UK Consortium |
| EPI_ISL_458518 | Department of Pathology, University of Cambridge | Wellcome Sanger Institute for the COVID-19 Genomics UK Consortium |
| EPI_ISL_458519 | Department of Pathology, University of Cambridge | Wellcome Sanger Institute for the COVID-19 Genomics UK Consortium |
| EPI_ISL_458520 | Department of Pathology, University of Cambridge | Wellcome Sanger Institute for the COVID-19 Genomics UK Consortium |
| EPI_ISL_458521 | Department of Pathology, University of Cambridge | Wellcome Sanger Institute for the COVID-19 Genomics UK Consortium |

| EPI_Accession_ID | Originating Laboratory | Submitting Laboratory |
|---|---|---|
| EPI_ISL_458522 | Department of Pathology, University of Cambridge | Wellcome Sanger Institute for the COVID-19 Genomics UK Consortium |
| EPI_ISL_458523 | Department of Pathology, University of Cambridge | Wellcome Sanger Institute for the COVID-19 Genomics UK Consortium |
| EPI_ISL_458524 | Department of Pathology, University of Cambridge | Wellcome Sanger Institute for the COVID-19 Genomics UK Consortium |
| EPI_ISL_458525 | Department of Pathology, University of Cambridge | Wellcome Sanger Institute for the COVID-19 Genomics UK Consortium |
| EPI_ISL_458526 | Department of Pathology, University of Cambridge | Wellcome Sanger Institute for the COVID-19 Genomics UK Consortium |
| EPI_ISL_458527 | Department of Pathology, University of Cambridge | Wellcome Sanger Institute for the COVID-19 Genomics UK Consortium |
| EPI_ISL_458528 | Department of Pathology, University of Cambridge | Wellcome Sanger Institute for the COVID-19 Genomics UK Consortium |
| EPI_ISL_458529 | Department of Pathology, University of Cambridge | Wellcome Sanger Institute for the COVID-19 Genomics UK Consortium |
| EPI_ISL_458530 | Department of Pathology, University of Cambridge | Wellcome Sanger Institute for the COVID-19 Genomics UK Consortium |
| EPI_ISL_458531 | Department of Pathology, University of Cambridge | Wellcome Sanger Institute for the COVID-19 Genomics UK Consortium |
| EPI_ISL_458532 | Department of Pathology, University of Cambridge | Wellcome Sanger Institute for the COVID-19 Genomics UK Consortium |
| EPI_ISL_458533 | Department of Pathology, University of Cambridge | Wellcome Sanger Institute for the COVID-19 Genomics UK Consortium |
| EPI_ISL_458534 | Department of Pathology, University of Cambridge | Wellcome Sanger Institute for the COVID-19 Genomics UK Consortium |
| EPI_ISL_458535 | Department of Pathology, University of Cambridge | Wellcome Sanger Institute for the COVID-19 Genomics UK Consortium |
| EPI_ISL_458536 | Department of Pathology, University of Cambridge | Wellcome Sanger Institute for the COVID-19 Genomics UK Consortium |
| EPI_ISL_458537 | Department of Pathology, University of Cambridge | Wellcome Sanger Institute for the COVID-19 Genomics UK Consortium |

| EPI_Accession_ID | Originating Laboratory | Submitting Laboratory |
|---|---|---|
| EPI_ISL_458538 | Department of Pathology, University of Cambridge | Wellcome Sanger Institute for the COVID-19 Genomics UK Consortium |
| EPI_ISL_458539 | Department of Pathology, University of Cambridge | Wellcome Sanger Institute for the COVID-19 Genomics UK Consortium |
| EPI_ISL_458540 | Department of Pathology, University of Cambridge | Wellcome Sanger Institute for the COVID-19 Genomics UK Consortium |
| EPI_ISL_458541 | Department of Pathology, University of Cambridge | Wellcome Sanger Institute for the COVID-19 Genomics UK Consortium |
| EPI_ISL_458542 | Department of Pathology, University of Cambridge | Wellcome Sanger Institute for the COVID-19 Genomics UK Consortium |
| EPI_ISL_458543 | Department of Pathology, University of Cambridge | Wellcome Sanger Institute for the COVID-19 Genomics UK Consortium |
| EPI_ISL_458544 | Department of Pathology, University of Cambridge | Wellcome Sanger Institute for the COVID-19 Genomics UK Consortium |
| EPI_ISL_458545 | Department of Pathology, University of Cambridge | Wellcome Sanger Institute for the COVID-19 Genomics UK Consortium |
| EPI_ISL_458546 | Department of Pathology, University of Cambridge | Wellcome Sanger Institute for the COVID-19 Genomics UK Consortium |
| EPI_ISL_458547 | Department of Pathology, University of Cambridge | Wellcome Sanger Institute for the COVID-19 Genomics UK Consortium |
| EPI_ISL_458548 | Department of Pathology, University of Cambridge | Wellcome Sanger Institute for the COVID-19 Genomics UK Consortium |
| EPI_ISL_458549 | Department of Pathology, University of Cambridge | Wellcome Sanger Institute for the COVID-19 Genomics UK Consortium |
| EPI_ISL_458550 | Department of Pathology, University of Cambridge | Wellcome Sanger Institute for the COVID-19 Genomics UK Consortium |
| EPI_ISL_458551 | Department of Pathology, University of Cambridge | Wellcome Sanger Institute for the COVID-19 Genomics UK Consortium |
| EPI_ISL_458552 | Department of Pathology, University of Cambridge | Wellcome Sanger Institute for the COVID-19 Genomics UK Consortium |
| EPI_ISL_458553 | Department of Pathology, University of Cambridge | Wellcome Sanger Institute for the COVID-19 Genomics UK Consortium |

| EPI_Accession_ID | Originating Laboratory | Submitting Laboratory |
|---|---|---|
| EPI_ISL_458554 | Department of Pathology, University of Cambridge | Wellcome Sanger Institute for the COVID-19 Genomics UK Consortium |
| EPI_ISL_458555 | Department of Pathology, University of Cambridge | Wellcome Sanger Institute for the COVID-19 Genomics UK Consortium |
| EPI_ISL_458556 | Department of Pathology, University of Cambridge | Wellcome Sanger Institute for the COVID-19 Genomics UK Consortium |
| EPI_ISL_458557 | Department of Pathology, University of Cambridge | Wellcome Sanger Institute for the COVID-19 Genomics UK Consortium |
| EPI_ISL_458558 | Department of Pathology, University of Cambridge | Wellcome Sanger Institute for the COVID-19 Genomics UK Consortium |
| EPI_ISL_458559 | Department of Pathology, University of Cambridge | Wellcome Sanger Institute for the COVID-19 Genomics UK Consortium |
| EPI_ISL_458560 | Department of Pathology, University of Cambridge | Wellcome Sanger Institute for the COVID-19 Genomics UK Consortium |
| EPI_ISL_458561 | Department of Pathology, University of Cambridge | Wellcome Sanger Institute for the COVID-19 Genomics UK Consortium |
| EPI_ISL_458562 | Department of Pathology, University of Cambridge | Wellcome Sanger Institute for the COVID-19 Genomics UK Consortium |
| EPI_ISL_458563 | Department of Pathology, University of Cambridge | Wellcome Sanger Institute for the COVID-19 Genomics UK Consortium |
| EPI_ISL_458564 | Department of Pathology, University of Cambridge | Wellcome Sanger Institute for the COVID-19 Genomics UK Consortium |
| EPI_ISL_458565 | Department of Pathology, University of Cambridge | Wellcome Sanger Institute for the COVID-19 Genomics UK Consortium |
| EPI_ISL_458566 | Department of Pathology, University of Cambridge | Wellcome Sanger Institute for the COVID-19 Genomics UK Consortium |
| EPI_ISL_458567 | Department of Pathology, University of Cambridge | Wellcome Sanger Institute for the COVID-19 Genomics UK Consortium |
| EPI_ISL_458568 | Department of Pathology, University of Cambridge | Wellcome Sanger Institute for the COVID-19 Genomics UK Consortium |
| EPI_ISL_458569 | Department of Pathology, University of Cambridge | Wellcome Sanger Institute for the COVID-19 Genomics UK Consortium |

| EPI_Accession_ID | Originating Laboratory | Submitting Laboratory |
|---|---|---|
| EPI_ISL_458570 | Department of Pathology, University of Cambridge | Wellcome Sanger Institute for the COVID-19 Genomics UK Consortium |
| EPI_ISL_458571 | Department of Pathology, University of Cambridge | Wellcome Sanger Institute for the COVID-19 Genomics UK Consortium |
| EPI_ISL_458572 | Department of Pathology, University of Cambridge | Wellcome Sanger Institute for the COVID-19 Genomics UK Consortium |
| EPI_ISL_458573 | Department of Pathology, University of Cambridge | Wellcome Sanger Institute for the COVID-19 Genomics UK Consortium |
| EPI_ISL_458574 | Department of Pathology, University of Cambridge | Wellcome Sanger Institute for the COVID-19 Genomics UK Consortium |
| EPI_ISL_458575 | Department of Pathology, University of Cambridge | Wellcome Sanger Institute for the COVID-19 Genomics UK Consortium |
| EPI_ISL_458576 | Department of Pathology, University of Cambridge | Wellcome Sanger Institute for the COVID-19 Genomics UK Consortium |
| EPI_ISL_458577 | Department of Pathology, University of Cambridge | Wellcome Sanger Institute for the COVID-19 Genomics UK Consortium |
| EPI_ISL_458578 | NU-OMICS DNA Sequencing research facility, Northumbria University | Wellcome Sanger Institute for the COVID-19 Genomics UK Consortium |
| EPI_ISL_458579 | NU-OMICS DNA Sequencing research facility, Northumbria University | Wellcome Sanger Institute for the COVID-19 Genomics UK Consortium |
| EPI_ISL_458580 | NU-OMICS DNA Sequencing research facility, Northumbria University | Wellcome Sanger Institute for the COVID-19 Genomics UK Consortium |
| EPI_ISL_458581 | NU-OMICS DNA Sequencing research facility, Northumbria University | Wellcome Sanger Institute for the COVID-19 Genomics UK Consortium |
| EPI_ISL_458582 | NU-OMICS DNA Sequencing research facility, Northumbria University | Wellcome Sanger Institute for the COVID-19 Genomics UK Consortium |
| EPI_ISL_458583 | NU-OMICS DNA Sequencing research facility, Northumbria University | Wellcome Sanger Institute for the COVID-19 Genomics UK Consortium |
| EPI_ISL_458584 | NU-OMICS DNA Sequencing research facility, Northumbria University | Wellcome Sanger Institute for the COVID-19 Genomics UK Consortium |
| EPI_ISL_458585 | NU-OMICS DNA Sequencing research facility, Northumbria University | Wellcome Sanger Institute for the COVID-19 Genomics UK Consortium |

| EPI_Accession_ID | Originating Laboratory | Submitting Laboratory |
|---|---|---|
| EPI_ISL_458586 | NU-OMICS DNA Sequencing research facility, Northumbria University | Wellcome Sanger Institute for the COVID-19 Genomics UK Consortium |
| EPI_ISL_458587 | NU-OMICS DNA Sequencing research facility, Northumbria University | Wellcome Sanger Institute for the COVID-19 Genomics UK Consortium |
| EPI_ISL_458588 | NU-OMICS DNA Sequencing research facility, Northumbria University | Wellcome Sanger Institute for the COVID-19 Genomics UK Consortium |
| EPI_ISL_458589 | NU-OMICS DNA Sequencing research facility, Northumbria University | Wellcome Sanger Institute for the COVID-19 Genomics UK Consortium |
| EPI_ISL_458590 | NU-OMICS DNA Sequencing research facility, Northumbria University | Wellcome Sanger Institute for the COVID-19 Genomics UK Consortium |
| EPI_ISL_458591 | NU-OMICS DNA Sequencing research facility, Northumbria University | Wellcome Sanger Institute for the COVID-19 Genomics UK Consortium |
| EPI_ISL_458592 | NU-OMICS DNA Sequencing research facility, Northumbria University | Wellcome Sanger Institute for the COVID-19 Genomics UK Consortium |
| EPI_ISL_458593 | NU-OMICS DNA Sequencing research facility, Northumbria University | Wellcome Sanger Institute for the COVID-19 Genomics UK Consortium |
| EPI_ISL_458594 | NU-OMICS DNA Sequencing research facility, Northumbria University | Wellcome Sanger Institute for the COVID-19 Genomics UK Consortium |
| EPI_ISL_458595 | NU-OMICS DNA Sequencing research facility, Northumbria University | Wellcome Sanger Institute for the COVID-19 Genomics UK Consortium |
| EPI_ISL_458596 | NU-OMICS DNA Sequencing research facility, Northumbria University | Wellcome Sanger Institute for the COVID-19 Genomics UK Consortium |
| EPI_ISL_458597 | NU-OMICS DNA Sequencing research facility, Northumbria University | Wellcome Sanger Institute for the COVID-19 Genomics UK Consortium |
| EPI_ISL_458598 | NU-OMICS DNA Sequencing research facility, Northumbria University | Wellcome Sanger Institute for the COVID-19 Genomics UK Consortium |
| EPI_ISL_458599 | NU-OMICS DNA Sequencing research facility, Northumbria University | Wellcome Sanger Institute for the COVID-19 Genomics UK Consortium |
| EPI_ISL_458600 | NU-OMICS DNA Sequencing research facility, Northumbria University | Wellcome Sanger Institute for the COVID-19 Genomics UK Consortium |
| EPI_ISL_458601 | NU-OMICS DNA Sequencing research facility, Northumbria University | Wellcome Sanger Institute for the COVID-19 Genomics UK Consortium |

| EPI_Accession_ID | Originating Laboratory | Submitting Laboratory |
|---|---|---|
| EPI_ISL_458602 | NU-OMICS DNA Sequencing research facility, Northumbria University | Wellcome Sanger Institute for the COVID-19 Genomics UK Consortium |
| EPI_ISL_458603 | NU-OMICS DNA Sequencing research facility, Northumbria University | Wellcome Sanger Institute for the COVID-19 Genomics UK Consortium |
| EPI_ISL_458604 | NU-OMICS DNA Sequencing research facility, Northumbria University | Wellcome Sanger Institute for the COVID-19 Genomics UK Consortium |
| EPI_ISL_458605 | NU-OMICS DNA Sequencing research facility, Northumbria University | Wellcome Sanger Institute for the COVID-19 Genomics UK Consortium |
| EPI_ISL_458606 | NU-OMICS DNA Sequencing research facility, Northumbria University | Wellcome Sanger Institute for the COVID-19 Genomics UK Consortium |
| EPI_ISL_458607 | NU-OMICS DNA Sequencing research facility, Northumbria University | Wellcome Sanger Institute for the COVID-19 Genomics UK Consortium |
| EPI_ISL_458608 | NU-OMICS DNA Sequencing research facility, Northumbria University | Wellcome Sanger Institute for the COVID-19 Genomics UK Consortium |
| EPI_ISL_458609 | NU-OMICS DNA Sequencing research facility, Northumbria University | Wellcome Sanger Institute for the COVID-19 Genomics UK Consortium |
| EPI_ISL_458610 | NU-OMICS DNA Sequencing research facility, Northumbria University | Wellcome Sanger Institute for the COVID-19 Genomics UK Consortium |
| EPI_ISL_458611 | NU-OMICS DNA Sequencing research facility, Northumbria University | Wellcome Sanger Institute for the COVID-19 Genomics UK Consortium |
| EPI_ISL_458612 | NU-OMICS DNA Sequencing research facility, Northumbria University | Wellcome Sanger Institute for the COVID-19 Genomics UK Consortium |
| EPI_ISL_458613 | NU-OMICS DNA Sequencing research facility, Northumbria University | Wellcome Sanger Institute for the COVID-19 Genomics UK Consortium |
| EPI_ISL_458614 | NU-OMICS DNA Sequencing research facility, Northumbria University | Wellcome Sanger Institute for the COVID-19 Genomics UK Consortium |
| EPI_ISL_458615 | NU-OMICS DNA Sequencing research facility, Northumbria University | Wellcome Sanger Institute for the COVID-19 Genomics UK Consortium |
| EPI_ISL_458616 | NU-OMICS DNA Sequencing research facility, Northumbria University | Wellcome Sanger Institute for the COVID-19 Genomics UK Consortium |
| EPI_ISL_458617 | NU-OMICS DNA Sequencing research facility, Northumbria University | Wellcome Sanger Institute for the COVID-19 Genomics UK Consortium |

| EPI_Accession_ID | Originating Laboratory | Submitting Laboratory |
|---|---|---|
| EPI_ISL_458618 | NU-OMICS DNA Sequencing research facility, Northumbria University | Wellcome Sanger Institute for the COVID-19 Genomics UK Consortium |
| EPI_ISL_458619 | NU-OMICS DNA Sequencing research facility, Northumbria University | Wellcome Sanger Institute for the COVID-19 Genomics UK Consortium |
| EPI_ISL_458620 | NU-OMICS DNA Sequencing research facility, Northumbria University | Wellcome Sanger Institute for the COVID-19 Genomics UK Consortium |
| EPI_ISL_458621 | NU-OMICS DNA Sequencing research facility, Northumbria University | Wellcome Sanger Institute for the COVID-19 Genomics UK Consortium |
| EPI_ISL_458622 | NU-OMICS DNA Sequencing research facility, Northumbria University | Wellcome Sanger Institute for the COVID-19 Genomics UK Consortium |
| EPI_ISL_458623 | NU-OMICS DNA Sequencing research facility, Northumbria University | Wellcome Sanger Institute for the COVID-19 Genomics UK Consortium |
| EPI_ISL_458624 | NU-OMICS DNA Sequencing research facility, Northumbria University | Wellcome Sanger Institute for the COVID-19 Genomics UK Consortium |
| EPI_ISL_458625 | NU-OMICS DNA Sequencing research facility, Northumbria University | Wellcome Sanger Institute for the COVID-19 Genomics UK Consortium |
| EPI_ISL_458626 | NU-OMICS DNA Sequencing research facility, Northumbria University | Wellcome Sanger Institute for the COVID-19 Genomics UK Consortium |
| EPI_ISL_458627 | NU-OMICS DNA Sequencing research facility, Northumbria University | Wellcome Sanger Institute for the COVID-19 Genomics UK Consortium |
| EPI_ISL_458628 | NU-OMICS DNA Sequencing research facility, Northumbria University | Wellcome Sanger Institute for the COVID-19 Genomics UK Consortium |
| EPI_ISL_458629 | NU-OMICS DNA Sequencing research facility, Northumbria University | Wellcome Sanger Institute for the COVID-19 Genomics UK Consortium |
| EPI_ISL_458630 | NU-OMICS DNA Sequencing research facility, Northumbria University | Wellcome Sanger Institute for the COVID-19 Genomics UK Consortium |
| EPI_ISL_458631 | NU-OMICS DNA Sequencing research facility, Northumbria University | Wellcome Sanger Institute for the COVID-19 Genomics UK Consortium |
| EPI_ISL_458632 | NU-OMICS DNA Sequencing research facility, Northumbria University | Wellcome Sanger Institute for the COVID-19 Genomics UK Consortium |
| EPI_ISL_458633 | NU-OMICS DNA Sequencing research facility, Northumbria University | Wellcome Sanger Institute for the COVID-19 Genomics UK Consortium |

| EPI_Accession_ID | Originating Laboratory | Submitting Laboratory |
|---|---|---|
| EPI_ISL_458634 | NU-OMICS DNA Sequencing research facility, Northumbria University | Wellcome Sanger Institute for the COVID-19 Genomics UK Consortium |
| EPI_ISL_458635 | NU-OMICS DNA Sequencing research facility, Northumbria University | Wellcome Sanger Institute for the COVID-19 Genomics UK Consortium |
| EPI_ISL_458636 | NU-OMICS DNA Sequencing research facility, Northumbria University | Wellcome Sanger Institute for the COVID-19 Genomics UK Consortium |
| EPI_ISL_458637 | NU-OMICS DNA Sequencing research facility, Northumbria University | Wellcome Sanger Institute for the COVID-19 Genomics UK Consortium |
| EPI_ISL_458638 | NU-OMICS DNA Sequencing research facility, Northumbria University | Wellcome Sanger Institute for the COVID-19 Genomics UK Consortium |
| EPI_ISL_458639 | NU-OMICS DNA Sequencing research facility, Northumbria University | Wellcome Sanger Institute for the COVID-19 Genomics UK Consortium |
| EPI_ISL_458640 | NU-OMICS DNA Sequencing research facility, Northumbria University | Wellcome Sanger Institute for the COVID-19 Genomics UK Consortium |
| EPI_ISL_458641 | NU-OMICS DNA Sequencing research facility, Northumbria University | Wellcome Sanger Institute for the COVID-19 Genomics UK Consortium |
| EPI_ISL_458642 | NU-OMICS DNA Sequencing research facility, Northumbria University | Wellcome Sanger Institute for the COVID-19 Genomics UK Consortium |
| EPI_ISL_458643 | NU-OMICS DNA Sequencing research facility, Northumbria University | Wellcome Sanger Institute for the COVID-19 Genomics UK Consortium |
| EPI_ISL_458644 | NU-OMICS DNA Sequencing research facility, Northumbria University | Wellcome Sanger Institute for the COVID-19 Genomics UK Consortium |
| EPI_ISL_458645 | NU-OMICS DNA Sequencing research facility, Northumbria University | Wellcome Sanger Institute for the COVID-19 Genomics UK Consortium |
| EPI_ISL_458646 | NU-OMICS DNA Sequencing research facility, Northumbria University | Wellcome Sanger Institute for the COVID-19 Genomics UK Consortium |
| EPI_ISL_458647 | NU-OMICS DNA Sequencing research facility, Northumbria University | Wellcome Sanger Institute for the COVID-19 Genomics UK Consortium |
| EPI_ISL_458648 | NU-OMICS DNA Sequencing research facility, Northumbria University | Wellcome Sanger Institute for the COVID-19 Genomics UK Consortium |
| EPI_ISL_458649 | NU-OMICS DNA Sequencing research facility, Northumbria University | Wellcome Sanger Institute for the COVID-19 Genomics UK Consortium |

| EPI_Accession_ID | Originating Laboratory | Submitting Laboratory |
|---|---|---|
| EPI_ISL_458650 | NU-OMICS DNA Sequencing research facility, Northumbria University | Wellcome Sanger Institute for the COVID-19 Genomics UK Consortium |
| EPI_ISL_458651 | NU-OMICS DNA Sequencing research facility, Northumbria University | Wellcome Sanger Institute for the COVID-19 Genomics UK Consortium |
| EPI_ISL_458652 | NU-OMICS DNA Sequencing research facility, Northumbria University | Wellcome Sanger Institute for the COVID-19 Genomics UK Consortium |
| EPI_ISL_458653 | NU-OMICS DNA Sequencing research facility, Northumbria University | Wellcome Sanger Institute for the COVID-19 Genomics UK Consortium |
| EPI_ISL_458654 | NU-OMICS DNA Sequencing research facility, Northumbria University | Wellcome Sanger Institute for the COVID-19 Genomics UK Consortium |
| EPI_ISL_458655 | NU-OMICS DNA Sequencing research facility, Northumbria University | Wellcome Sanger Institute for the COVID-19 Genomics UK Consortium |
| EPI_ISL_458656 | NU-OMICS DNA Sequencing research facility, Northumbria University | Wellcome Sanger Institute for the COVID-19 Genomics UK Consortium |
| EPI_ISL_458657 | NU-OMICS DNA Sequencing research facility, Northumbria University | Wellcome Sanger Institute for the COVID-19 Genomics UK Consortium |
| EPI_ISL_458658 | NU-OMICS DNA Sequencing research facility, Northumbria University | Wellcome Sanger Institute for the COVID-19 Genomics UK Consortium |
| EPI_ISL_458659 | NU-OMICS DNA Sequencing research facility, Northumbria University | Wellcome Sanger Institute for the COVID-19 Genomics UK Consortium |
| EPI_ISL_458660 | NU-OMICS DNA Sequencing research facility, Northumbria University | Wellcome Sanger Institute for the COVID-19 Genomics UK Consortium |
| EPI_ISL_458661 | NU-OMICS DNA Sequencing research facility, Northumbria University | Wellcome Sanger Institute for the COVID-19 Genomics UK Consortium |
| EPI_ISL_458662 | NU-OMICS DNA Sequencing research facility, Northumbria University | Wellcome Sanger Institute for the COVID-19 Genomics UK Consortium |
| EPI_ISL_458663 | NU-OMICS DNA Sequencing research facility, Northumbria University | Wellcome Sanger Institute for the COVID-19 Genomics UK Consortium |
| EPI_ISL_458664 | NU-OMICS DNA Sequencing research facility, Northumbria University | Wellcome Sanger Institute for the COVID-19 Genomics UK Consortium |
| EPI_ISL_458665 | NU-OMICS DNA Sequencing research facility, Northumbria University | Wellcome Sanger Institute for the COVID-19 Genomics UK Consortium |

| EPI_Accession_ID | Originating Laboratory | Submitting Laboratory |
|---|---|---|
| EPI_ISL_458666 | NU-OMICS DNA Sequencing research facility, Northumbria University | Wellcome Sanger Institute for the COVID-19 Genomics UK Consortium |
| EPI_ISL_458667 | NU-OMICS DNA Sequencing research facility, Northumbria University | Wellcome Sanger Institute for the COVID-19 Genomics UK Consortium |
| EPI_ISL_458668 | NU-OMICS DNA Sequencing research facility, Northumbria University | Wellcome Sanger Institute for the COVID-19 Genomics UK Consortium |
| EPI_ISL_458669 | NU-OMICS DNA Sequencing research facility, Northumbria University | Wellcome Sanger Institute for the COVID-19 Genomics UK Consortium |
| EPI_ISL_458670 | NU-OMICS DNA Sequencing research facility, Northumbria University | Wellcome Sanger Institute for the COVID-19 Genomics UK Consortium |
| EPI_ISL_458671 | NU-OMICS DNA Sequencing research facility, Northumbria University | Wellcome Sanger Institute for the COVID-19 Genomics UK Consortium |
| EPI_ISL_458672 | NU-OMICS DNA Sequencing research facility, Northumbria University | Wellcome Sanger Institute for the COVID-19 Genomics UK Consortium |
| EPI_ISL_458673 | NU-OMICS DNA Sequencing research facility, Northumbria University | Wellcome Sanger Institute for the COVID-19 Genomics UK Consortium |
| EPI_ISL_458674 | NU-OMICS DNA Sequencing research facility, Northumbria University | Wellcome Sanger Institute for the COVID-19 Genomics UK Consortium |
| EPI_ISL_458675 | NU-OMICS DNA Sequencing research facility, Northumbria University | Wellcome Sanger Institute for the COVID-19 Genomics UK Consortium |
| EPI_ISL_458676 | NU-OMICS DNA Sequencing research facility, Northumbria University | Wellcome Sanger Institute for the COVID-19 Genomics UK Consortium |
| EPI_ISL_458677 | NU-OMICS DNA Sequencing research facility, Northumbria University | Wellcome Sanger Institute for the COVID-19 Genomics UK Consortium |
| EPI_ISL_458678 | NU-OMICS DNA Sequencing research facility, Northumbria University | Wellcome Sanger Institute for the COVID-19 Genomics UK Consortium |
| EPI_ISL_458679 | NU-OMICS DNA Sequencing research facility, Northumbria University | Wellcome Sanger Institute for the COVID-19 Genomics UK Consortium |
| EPI_ISL_458680 | NU-OMICS DNA Sequencing research facility, Northumbria University | Wellcome Sanger Institute for the COVID-19 Genomics UK Consortium |
| EPI_ISL_458681 | NU-OMICS DNA Sequencing research facility, Northumbria University | Wellcome Sanger Institute for the COVID-19 Genomics UK Consortium |

| EPI_Accession_ID | Originating Laboratory | Submitting Laboratory |
|---|---|---|
| EPI_ISL_458682 | NU-OMICS DNA Sequencing research facility, Northumbria University | Wellcome Sanger Institute for the COVID-19 Genomics UK Consortium |
| EPI_ISL_458683 | NU-OMICS DNA Sequencing research facility, Northumbria University | Wellcome Sanger Institute for the COVID-19 Genomics UK Consortium |
| EPI_ISL_458684 | NU-OMICS DNA Sequencing research facility, Northumbria University | Wellcome Sanger Institute for the COVID-19 Genomics UK Consortium |
| EPI_ISL_458685 | NU-OMICS DNA Sequencing research facility, Northumbria University | Wellcome Sanger Institute for the COVID-19 Genomics UK Consortium |
| EPI_ISL_458686 | NU-OMICS DNA Sequencing research facility, Northumbria University | Wellcome Sanger Institute for the COVID-19 Genomics UK Consortium |
| EPI_ISL_458687 | NU-OMICS DNA Sequencing research facility, Northumbria University | Wellcome Sanger Institute for the COVID-19 Genomics UK Consortium |
| EPI_ISL_458688 | NU-OMICS DNA Sequencing research facility, Northumbria University | Wellcome Sanger Institute for the COVID-19 Genomics UK Consortium |
| EPI_ISL_458689 | NU-OMICS DNA Sequencing research facility, Northumbria University | Wellcome Sanger Institute for the COVID-19 Genomics UK Consortium |
| EPI_ISL_458690 | NU-OMICS DNA Sequencing research facility, Northumbria University | Wellcome Sanger Institute for the COVID-19 Genomics UK Consortium |
| EPI_ISL_458691 | NU-OMICS DNA Sequencing research facility, Northumbria University | Wellcome Sanger Institute for the COVID-19 Genomics UK Consortium |
| EPI_ISL_458692 | NU-OMICS DNA Sequencing research facility, Northumbria University | Wellcome Sanger Institute for the COVID-19 Genomics UK Consortium |
| EPI_ISL_458693 | NU-OMICS DNA Sequencing research facility, Northumbria University | Wellcome Sanger Institute for the COVID-19 Genomics UK Consortium |
| EPI_ISL_458694 | NU-OMICS DNA Sequencing research facility, Northumbria University | Wellcome Sanger Institute for the COVID-19 Genomics UK Consortium |
| EPI_ISL_458695 | NU-OMICS DNA Sequencing research facility, Northumbria University | Wellcome Sanger Institute for the COVID-19 Genomics UK Consortium |
| EPI_ISL_458696 | NU-OMICS DNA Sequencing research facility, Northumbria University | Wellcome Sanger Institute for the COVID-19 Genomics UK Consortium |
| EPI_ISL_458697 | NU-OMICS DNA Sequencing research facility, Northumbria University | Wellcome Sanger Institute for the COVID-19 Genomics UK Consortium |

| EPI_Accession_ID | Originating Laboratory | Submitting Laboratory |
|---|---|---|
| EPI_ISL_458698 | NU-OMICS DNA Sequencing research facility, Northumbria University | Wellcome Sanger Institute for the COVID-19 Genomics UK Consortium |
| EPI_ISL_458699 | NU-OMICS DNA Sequencing research facility, Northumbria University | Wellcome Sanger Institute for the COVID-19 Genomics UK Consortium |
| EPI_ISL_458700 | NU-OMICS DNA Sequencing research facility, Northumbria University | Wellcome Sanger Institute for the COVID-19 Genomics UK Consortium |
| EPI_ISL_458701 | NU-OMICS DNA Sequencing research facility, Northumbria University | Wellcome Sanger Institute for the COVID-19 Genomics UK Consortium |
| EPI_ISL_458702 | NU-OMICS DNA Sequencing research facility, Northumbria University | Wellcome Sanger Institute for the COVID-19 Genomics UK Consortium |
| EPI_ISL_458703 | NU-OMICS DNA Sequencing research facility, Northumbria University | Wellcome Sanger Institute for the COVID-19 Genomics UK Consortium |
| EPI_ISL_458704 | NU-OMICS DNA Sequencing research facility, Northumbria University | Wellcome Sanger Institute for the COVID-19 Genomics UK Consortium |
| EPI_ISL_458705 | NU-OMICS DNA Sequencing research facility, Northumbria University | Wellcome Sanger Institute for the COVID-19 Genomics UK Consortium |
| EPI_ISL_458706 | NU-OMICS DNA Sequencing research facility, Northumbria University | Wellcome Sanger Institute for the COVID-19 Genomics UK Consortium |
| EPI_ISL_458707 | NU-OMICS DNA Sequencing research facility, Northumbria University | Wellcome Sanger Institute for the COVID-19 Genomics UK Consortium |
| EPI_ISL_458708 | NU-OMICS DNA Sequencing research facility, Northumbria University | Wellcome Sanger Institute for the COVID-19 Genomics UK Consortium |
| EPI_ISL_458709 | NU-OMICS DNA Sequencing research facility, Northumbria University | Wellcome Sanger Institute for the COVID-19 Genomics UK Consortium |
| EPI_ISL_458710 | NU-OMICS DNA Sequencing research facility, Northumbria University | Wellcome Sanger Institute for the COVID-19 Genomics UK Consortium |
| EPI_ISL_458711 | NU-OMICS DNA Sequencing research facility, Northumbria University | Wellcome Sanger Institute for the COVID-19 Genomics UK Consortium |
| EPI_ISL_458712 | NU-OMICS DNA Sequencing research facility, Northumbria University | Wellcome Sanger Institute for the COVID-19 Genomics UK Consortium |
| EPI_ISL_458713 | NU-OMICS DNA Sequencing research facility, Northumbria University | Wellcome Sanger Institute for the COVID-19 Genomics UK Consortium |

| EPI_Accession_ID | Originating Laboratory | Submitting Laboratory |
|---|---|---|
| EPI_ISL_458714 | NU-OMICS DNA Sequencing research facility, Northumbria University | Wellcome Sanger Institute for the COVID-19 Genomics UK Consortium |
| EPI_ISL_458715 | NU-OMICS DNA Sequencing research facility, Northumbria University | Wellcome Sanger Institute for the COVID-19 Genomics UK Consortium |
| EPI_ISL_458716 | NU-OMICS DNA Sequencing research facility, Northumbria University | Wellcome Sanger Institute for the COVID-19 Genomics UK Consortium |
| EPI_ISL_458717 | NU-OMICS DNA Sequencing research facility, Northumbria University | Wellcome Sanger Institute for the COVID-19 Genomics UK Consortium |
| EPI_ISL_458718 | NU-OMICS DNA Sequencing research facility, Northumbria University | Wellcome Sanger Institute for the COVID-19 Genomics UK Consortium |
| EPI_ISL_458719 | PHE South West Regional Laboratory, National Infection Service | Wellcome Sanger Institute for the COVID-19 Genomics UK Consortium |
| EPI_ISL_458720 | PHE South West Regional Laboratory, National Infection Service | Wellcome Sanger Institute for the COVID-19 Genomics UK Consortium |
| EPI_ISL_458721 | PHE South West Regional Laboratory, National Infection Service | Wellcome Sanger Institute for the COVID-19 Genomics UK Consortium |
| EPI_ISL_458722 | PHE South West Regional Laboratory, National Infection Service | Wellcome Sanger Institute for the COVID-19 Genomics UK Consortium |
| EPI_ISL_458723 | PHE South West Regional Laboratory, National Infection Service | Wellcome Sanger Institute for the COVID-19 Genomics UK Consortium |
| EPI_ISL_458724 | PHE South West Regional Laboratory, National Infection Service | Wellcome Sanger Institute for the COVID-19 Genomics UK Consortium |
| EPI_ISL_458725 | PHE South West Regional Laboratory, National Infection Service | Wellcome Sanger Institute for the COVID-19 Genomics UK Consortium |
| EPI_ISL_458726 | PHE South West Regional Laboratory, National Infection Service | Wellcome Sanger Institute for the COVID-19 Genomics UK Consortium |
| EPI_ISL_458727 | PHE South West Regional Laboratory, National Infection Service | Wellcome Sanger Institute for the COVID-19 Genomics UK Consortium |
| EPI_ISL_458728 | PHE South West Regional Laboratory, National Infection Service | Wellcome Sanger Institute for the COVID-19 Genomics UK Consortium |
| EPI_ISL_458729 | PHE South West Regional Laboratory, National Infection Service | Wellcome Sanger Institute for the COVID-19 Genomics UK Consortium |

| EPI_Accession_ID | Originating Laboratory | Submitting Laboratory |
|---|---|---|
| EPI_ISL_458730 | PHE South West Regional Laboratory, National Infection Service | Wellcome Sanger Institute for the COVID-19 Genomics UK Consortium |
| EPI_ISL_458731 | PHE South West Regional Laboratory, National Infection Service | Wellcome Sanger Institute for the COVID-19 Genomics UK Consortium |
| EPI_ISL_458732 | PHE South West Regional Laboratory, National Infection Service | Wellcome Sanger Institute for the COVID-19 Genomics UK Consortium |
| EPI_ISL_458733 | PHE South West Regional Laboratory, National Infection Service | Wellcome Sanger Institute for the COVID-19 Genomics UK Consortium |
| EPI_ISL_458734 | PHE South West Regional Laboratory, National Infection Service | Wellcome Sanger Institute for the COVID-19 Genomics UK Consortium |
| EPI_ISL_458735 | PHE South West Regional Laboratory, National Infection Service | Wellcome Sanger Institute for the COVID-19 Genomics UK Consortium |
| EPI_ISL_458736 | PHE South West Regional Laboratory, National Infection Service | Wellcome Sanger Institute for the COVID-19 Genomics UK Consortium |
| EPI_ISL_458737 | PHE South West Regional Laboratory, National Infection Service | Wellcome Sanger Institute for the COVID-19 Genomics UK Consortium |
| EPI_ISL_458738 | PHE South West Regional Laboratory, National Infection Service | Wellcome Sanger Institute for the COVID-19 Genomics UK Consortium |
| EPI_ISL_458739 | PHE South West Regional Laboratory, National Infection Service | Wellcome Sanger Institute for the COVID-19 Genomics UK Consortium |
| EPI_ISL_458740 | PHE South West Regional Laboratory, National Infection Service | Wellcome Sanger Institute for the COVID-19 Genomics UK Consortium |
| EPI_ISL_458741 | PHE South West Regional Laboratory, National Infection Service | Wellcome Sanger Institute for the COVID-19 Genomics UK Consortium |
| EPI_ISL_458742 | PHE South West Regional Laboratory, National Infection Service | Wellcome Sanger Institute for the COVID-19 Genomics UK Consortium |
| EPI_ISL_458743 | PHE South West Regional Laboratory, National Infection Service | Wellcome Sanger Institute for the COVID-19 Genomics UK Consortium |
| EPI_ISL_458744 | PHE South West Regional Laboratory, National Infection Service | Wellcome Sanger Institute for the COVID-19 Genomics UK Consortium |
| EPI_ISL_458745 | PHE South West Regional Laboratory, National Infection Service | Wellcome Sanger Institute for the COVID-19 Genomics UK Consortium |

| EPI_Accession_ID | Originating Laboratory | Submitting Laboratory |
| --- | --- | --- |
| EPI_ISL_458746 | PHE South West Regional Laboratory, National Infection Service | Wellcome Sanger Institute for the COVID-19 Genomics UK Consortium |
| EPI_ISL_458747 | PHE South West Regional Laboratory, National Infection Service | Wellcome Sanger Institute for the COVID-19 Genomics UK Consortium |
| EPI_ISL_458748 | PHE South West Regional Laboratory, National Infection Service | Wellcome Sanger Institute for the COVID-19 Genomics UK Consortium |
| EPI_ISL_458749 | PHE South West Regional Laboratory, National Infection Service | Wellcome Sanger Institute for the COVID-19 Genomics UK Consortium |
| EPI_ISL_458750 | PHE South West Regional Laboratory, National Infection Service | Wellcome Sanger Institute for the COVID-19 Genomics UK Consortium |
| EPI_ISL_458751 | PHE South West Regional Laboratory, National Infection Service | Wellcome Sanger Institute for the COVID-19 Genomics UK Consortium |
| EPI_ISL_458752 | PHE South West Regional Laboratory, National Infection Service | Wellcome Sanger Institute for the COVID-19 Genomics UK Consortium |
| EPI_ISL_458753 | PHE South West Regional Laboratory, National Infection Service | Wellcome Sanger Institute for the COVID-19 Genomics UK Consortium |
| EPI_ISL_458754 | PHE South West Regional Laboratory, National Infection Service | Wellcome Sanger Institute for the COVID-19 Genomics UK Consortium |
| EPI_ISL_458755 | PHE South West Regional Laboratory, National Infection Service | Wellcome Sanger Institute for the COVID-19 Genomics UK Consortium |
| EPI_ISL_458756 | PHE South West Regional Laboratory, National Infection Service | Wellcome Sanger Institute for the COVID-19 Genomics UK Consortium |
| EPI_ISL_458757 | PHE South West Regional Laboratory, National Infection Service | Wellcome Sanger Institute for the COVID-19 Genomics UK Consortium |
| EPI_ISL_458758 | PHE South West Regional Laboratory, National Infection Service | Wellcome Sanger Institute for the COVID-19 Genomics UK Consortium |
| EPI_ISL_458759 | PHE South West Regional Laboratory, National Infection Service | Wellcome Sanger Institute for the COVID-19 Genomics UK Consortium |
| EPI_ISL_458760 | PHE South West Regional Laboratory, National Infection Service | Wellcome Sanger Institute for the COVID-19 Genomics UK Consortium |
| EPI_ISL_458761 | PHE South West Regional Laboratory, National Infection Service | Wellcome Sanger Institute for the COVID-19 Genomics UK Consortium |

| EPI_Accession_ID | Originating Laboratory | Submitting Laboratory |
|---|---|---|
| EPI_ISL_458762 | PHE South West Regional Laboratory, National Infection Service | Wellcome Sanger Institute for the COVID-19 Genomics UK Consortium |
| EPI_ISL_458763 | PHE South West Regional Laboratory, National Infection Service | Wellcome Sanger Institute for the COVID-19 Genomics UK Consortium |
| EPI_ISL_458764 | PHE South West Regional Laboratory, National Infection Service | Wellcome Sanger Institute for the COVID-19 Genomics UK Consortium |
| EPI_ISL_458765 | PHE South West Regional Laboratory, National Infection Service | Wellcome Sanger Institute for the COVID-19 Genomics UK Consortium |
| EPI_ISL_458766 | PHE South West Regional Laboratory, National Infection Service | Wellcome Sanger Institute for the COVID-19 Genomics UK Consortium |
| EPI_ISL_458767 | PHE South West Regional Laboratory, National Infection Service | Wellcome Sanger Institute for the COVID-19 Genomics UK Consortium |
| EPI_ISL_458768 | PHE South West Regional Laboratory, National Infection Service | Wellcome Sanger Institute for the COVID-19 Genomics UK Consortium |
| EPI_ISL_458769 | PHE South West Regional Laboratory, National Infection Service | Wellcome Sanger Institute for the COVID-19 Genomics UK Consortium |
| EPI_ISL_458770 | PHE South West Regional Laboratory, National Infection Service | Wellcome Sanger Institute for the COVID-19 Genomics UK Consortium |
| EPI_ISL_458771 | PHE South West Regional Laboratory, National Infection Service | Wellcome Sanger Institute for the COVID-19 Genomics UK Consortium |
| EPI_ISL_458772 | PHE South West Regional Laboratory, National Infection Service | Wellcome Sanger Institute for the COVID-19 Genomics UK Consortium |
| EPI_ISL_458773 | PHE South West Regional Laboratory, National Infection Service | Wellcome Sanger Institute for the COVID-19 Genomics UK Consortium |
| EPI_ISL_458774 | PHE South West Regional Laboratory, National Infection Service | Wellcome Sanger Institute for the COVID-19 Genomics UK Consortium |
| EPI_ISL_458775 | PHE South West Regional Laboratory, National Infection Service | Wellcome Sanger Institute for the COVID-19 Genomics UK Consortium |
| EPI_ISL_458776 | PHE South West Regional Laboratory, National Infection Service | Wellcome Sanger Institute for the COVID-19 Genomics UK Consortium |
| EPI_ISL_458777 | PHE South West Regional Laboratory, National Infection Service | Wellcome Sanger Institute for the COVID-19 Genomics UK Consortium |

| EPI_Accession_ID | Originating Laboratory | Submitting Laboratory |
|---|---|---|
| EPI_ISL_458778 | PHE South West Regional Laboratory, National Infection Service | Wellcome Sanger Institute for the COVID-19 Genomics UK Consortium |
| EPI_ISL_458779 | PHE South West Regional Laboratory, National Infection Service | Wellcome Sanger Institute for the COVID-19 Genomics UK Consortium |
| EPI_ISL_458780 | PHE South West Regional Laboratory, National Infection Service | Wellcome Sanger Institute for the COVID-19 Genomics UK Consortium |
| EPI_ISL_458781 | PHE South West Regional Laboratory, National Infection Service | Wellcome Sanger Institute for the COVID-19 Genomics UK Consortium |
| EPI_ISL_458782 | PHE South West Regional Laboratory, National Infection Service | Wellcome Sanger Institute for the COVID-19 Genomics UK Consortium |
| EPI_ISL_458783 | PHE South West Regional Laboratory, National Infection Service | Wellcome Sanger Institute for the COVID-19 Genomics UK Consortium |
| EPI_ISL_458784 | PHE South West Regional Laboratory, National Infection Service | Wellcome Sanger Institute for the COVID-19 Genomics UK Consortium |
| EPI_ISL_458785 | PHE South West Regional Laboratory, National Infection Service | Wellcome Sanger Institute for the COVID-19 Genomics UK Consortium |
| EPI_ISL_458786 | PHE South West Regional Laboratory, National Infection Service | Wellcome Sanger Institute for the COVID-19 Genomics UK Consortium |
| EPI_ISL_458787 | PHE South West Regional Laboratory, National Infection Service | Wellcome Sanger Institute for the COVID-19 Genomics UK Consortium |
| EPI_ISL_458788 | PHE South West Regional Laboratory, National Infection Service | Wellcome Sanger Institute for the COVID-19 Genomics UK Consortium |
| EPI_ISL_458789 | PHE South West Regional Laboratory, National Infection Service | Wellcome Sanger Institute for the COVID-19 Genomics UK Consortium |
| EPI_ISL_458790 | PHE South West Regional Laboratory, National Infection Service | Wellcome Sanger Institute for the COVID-19 Genomics UK Consortium |
| EPI_ISL_458791 | PHE South West Regional Laboratory, National Infection Service | Wellcome Sanger Institute for the COVID-19 Genomics UK Consortium |
| EPI_ISL_458792 | PHE South West Regional Laboratory, National Infection Service | Wellcome Sanger Institute for the COVID-19 Genomics UK Consortium |
| EPI_ISL_458793 | PHE South West Regional Laboratory, National Infection Service | Wellcome Sanger Institute for the COVID-19 Genomics UK Consortium |

| EPI_Accession_ID | Originating Laboratory | Submitting Laboratory |
|---|---|---|
| EPI_ISL_458794 | PHE South West Regional Laboratory, National Infection Service | Wellcome Sanger Institute for the COVID-19 Genomics UK Consortium |
| EPI_ISL_458795 | PHE South West Regional Laboratory, National Infection Service | Wellcome Sanger Institute for the COVID-19 Genomics UK Consortium |
| EPI_ISL_458796 | PHE South West Regional Laboratory, National Infection Service | Wellcome Sanger Institute for the COVID-19 Genomics UK Consortium |
| EPI_ISL_458797 | PHE South West Regional Laboratory, National Infection Service | Wellcome Sanger Institute for the COVID-19 Genomics UK Consortium |
| EPI_ISL_458798 | PHE South West Regional Laboratory, National Infection Service | Wellcome Sanger Institute for the COVID-19 Genomics UK Consortium |
| EPI_ISL_458799 | PHE South West Regional Laboratory, National Infection Service | Wellcome Sanger Institute for the COVID-19 Genomics UK Consortium |
| EPI_ISL_458800 | PHE South West Regional Laboratory, National Infection Service | Wellcome Sanger Institute for the COVID-19 Genomics UK Consortium |
| EPI_ISL_458801 | PHE South West Regional Laboratory, National Infection Service | Wellcome Sanger Institute for the COVID-19 Genomics UK Consortium |
| EPI_ISL_458802 | PHE South West Regional Laboratory, National Infection Service | Wellcome Sanger Institute for the COVID-19 Genomics UK Consortium |
| EPI_ISL_458803 | PHE South West Regional Laboratory, National Infection Service | Wellcome Sanger Institute for the COVID-19 Genomics UK Consortium |
| EPI_ISL_458804 | PHE South West Regional Laboratory, National Infection Service | Wellcome Sanger Institute for the COVID-19 Genomics UK Consortium |
| EPI_ISL_458805 | PHE South West Regional Laboratory, National Infection Service | Wellcome Sanger Institute for the COVID-19 Genomics UK Consortium |
| EPI_ISL_458806 | PHE South West Regional Laboratory, National Infection Service | Wellcome Sanger Institute for the COVID-19 Genomics UK Consortium |
| EPI_ISL_458807 | PHE South West Regional Laboratory, National Infection Service | Wellcome Sanger Institute for the COVID-19 Genomics UK Consortium |
| EPI_ISL_458808 | PHE South West Regional Laboratory, National Infection Service | Wellcome Sanger Institute for the COVID-19 Genomics UK Consortium |
| EPI_ISL_458809 | PHE South West Regional Laboratory, National Infection Service | Wellcome Sanger Institute for the COVID-19 Genomics UK Consortium |

| EPI_Accession_ID | Originating Laboratory | Submitting Laboratory |
|---|---|---|
| EPI_ISL_458810 | PHE South West Regional Laboratory, National Infection Service | Wellcome Sanger Institute for the COVID-19 Genomics UK Consortium |
| EPI_ISL_458811 | PHE South West Regional Laboratory, National Infection Service | Wellcome Sanger Institute for the COVID-19 Genomics UK Consortium |
| EPI_ISL_458812 | PHE South West Regional Laboratory, National Infection Service | Wellcome Sanger Institute for the COVID-19 Genomics UK Consortium |
| EPI_ISL_458813 | PHE South West Regional Laboratory, National Infection Service | Wellcome Sanger Institute for the COVID-19 Genomics UK Consortium |
| EPI_ISL_458814 | PHE South West Regional Laboratory, National Infection Service | Wellcome Sanger Institute for the COVID-19 Genomics UK Consortium |
| EPI_ISL_458815 | PHE South West Regional Laboratory, National Infection Service | Wellcome Sanger Institute for the COVID-19 Genomics UK Consortium |
| EPI_ISL_458816 | PHE South West Regional Laboratory, National Infection Service | Wellcome Sanger Institute for the COVID-19 Genomics UK Consortium |
| EPI_ISL_458817 | PHE South West Regional Laboratory, National Infection Service | Wellcome Sanger Institute for the COVID-19 Genomics UK Consortium |
| EPI_ISL_458818 | PHE South West Regional Laboratory, National Infection Service | Wellcome Sanger Institute for the COVID-19 Genomics UK Consortium |
| EPI_ISL_458819 | PHE South West Regional Laboratory, National Infection Service | Wellcome Sanger Institute for the COVID-19 Genomics UK Consortium |
| EPI_ISL_458820 | PHE South West Regional Laboratory, National Infection Service | Wellcome Sanger Institute for the COVID-19 Genomics UK Consortium |
| EPI_ISL_458821 | PHE South West Regional Laboratory, National Infection Service | Wellcome Sanger Institute for the COVID-19 Genomics UK Consortium |
| EPI_ISL_458822 | PHE South West Regional Laboratory, National Infection Service | Wellcome Sanger Institute for the COVID-19 Genomics UK Consortium |
| EPI_ISL_458823 | PHE South West Regional Laboratory, National Infection Service | Wellcome Sanger Institute for the COVID-19 Genomics UK Consortium |
| EPI_ISL_458824 | PHE South West Regional Laboratory, National Infection Service | Wellcome Sanger Institute for the COVID-19 Genomics UK Consortium |
| EPI_ISL_458825 | PHE South West Regional Laboratory, National Infection Service | Wellcome Sanger Institute for the COVID-19 Genomics UK Consortium |

| EPI_Accession_ID | Originating Laboratory | Submitting Laboratory |
|---|---|---|
| EPI_ISL_458826 | PHE South West Regional Laboratory, National Infection Service | Wellcome Sanger Institute for the COVID-19 Genomics UK Consortium |
| EPI_ISL_458827 | PHE South West Regional Laboratory, National Infection Service | Wellcome Sanger Institute for the COVID-19 Genomics UK Consortium |
| EPI_ISL_458828 | PHE South West Regional Laboratory, National Infection Service | Wellcome Sanger Institute for the COVID-19 Genomics UK Consortium |
| EPI_ISL_458829 | PHE South West Regional Laboratory, National Infection Service | Wellcome Sanger Institute for the COVID-19 Genomics UK Consortium |
| EPI_ISL_458830 | PHE South West Regional Laboratory, National Infection Service | Wellcome Sanger Institute for the COVID-19 Genomics UK Consortium |
| EPI_ISL_458831 | PHE South West Regional Laboratory, National Infection Service | Wellcome Sanger Institute for the COVID-19 Genomics UK Consortium |
| EPI_ISL_458832 | PHE South West Regional Laboratory, National Infection Service | Wellcome Sanger Institute for the COVID-19 Genomics UK Consortium |
| EPI_ISL_458833 | PHE South West Regional Laboratory, National Infection Service | Wellcome Sanger Institute for the COVID-19 Genomics UK Consortium |
| EPI_ISL_458834 | PHE South West Regional Laboratory, National Infection Service | Wellcome Sanger Institute for the COVID-19 Genomics UK Consortium |
| EPI_ISL_458835 | PHE South West Regional Laboratory, National Infection Service | Wellcome Sanger Institute for the COVID-19 Genomics UK Consortium |
| EPI_ISL_458836 | PHE South West Regional Laboratory, National Infection Service | Wellcome Sanger Institute for the COVID-19 Genomics UK Consortium |
| EPI_ISL_458837 | PHE South West Regional Laboratory, National Infection Service | Wellcome Sanger Institute for the COVID-19 Genomics UK Consortium |
| EPI_ISL_458838 | PHE South West Regional Laboratory, National Infection Service | Wellcome Sanger Institute for the COVID-19 Genomics UK Consortium |
| EPI_ISL_458839 | PHE South West Regional Laboratory, National Infection Service | Wellcome Sanger Institute for the COVID-19 Genomics UK Consortium |
| EPI_ISL_458840 | PHE South West Regional Laboratory, National Infection Service | Wellcome Sanger Institute for the COVID-19 Genomics UK Consortium |
| EPI_ISL_458841 | PHE South West Regional Laboratory, National Infection Service | Wellcome Sanger Institute for the COVID-19 Genomics UK Consortium |

| EPI_Accession_ID | Originating Laboratory | Submitting Laboratory |
|---|---|---|
| EPI_ISL_458842 | PHE South West Regional Laboratory, National Infection Service | Wellcome Sanger Institute for the COVID-19 Genomics UK Consortium |
| EPI_ISL_458843 | PHE South West Regional Laboratory, National Infection Service | Wellcome Sanger Institute for the COVID-19 Genomics UK Consortium |
| EPI_ISL_458844 | PHE South West Regional Laboratory, National Infection Service | Wellcome Sanger Institute for the COVID-19 Genomics UK Consortium |
| EPI_ISL_458845 | PHE South West Regional Laboratory, National Infection Service | Wellcome Sanger Institute for the COVID-19 Genomics UK Consortium |
| EPI_ISL_458846 | PHE South West Regional Laboratory, National Infection Service | Wellcome Sanger Institute for the COVID-19 Genomics UK Consortium |
| EPI_ISL_458847 | PHE South West Regional Laboratory, National Infection Service | Wellcome Sanger Institute for the COVID-19 Genomics UK Consortium |
| EPI_ISL_458848 | PHE South West Regional Laboratory, National Infection Service | Wellcome Sanger Institute for the COVID-19 Genomics UK Consortium |
| EPI_ISL_458849 | PHE South West Regional Laboratory, National Infection Service | Wellcome Sanger Institute for the COVID-19 Genomics UK Consortium |
| EPI_ISL_458850 | PHE South West Regional Laboratory, National Infection Service | Wellcome Sanger Institute for the COVID-19 Genomics UK Consortium |
| EPI_ISL_458851 | PHE South West Regional Laboratory, National Infection Service | Wellcome Sanger Institute for the COVID-19 Genomics UK Consortium |
| EPI_ISL_458852 | PHE South West Regional Laboratory, National Infection Service | Wellcome Sanger Institute for the COVID-19 Genomics UK Consortium |
| EPI_ISL_458853 | PHE South West Regional Laboratory, National Infection Service | Wellcome Sanger Institute for the COVID-19 Genomics UK Consortium |
| EPI_ISL_458854 | PHE South West Regional Laboratory, National Infection Service | Wellcome Sanger Institute for the COVID-19 Genomics UK Consortium |
| EPI_ISL_458855 | PHE South West Regional Laboratory, National Infection Service | Wellcome Sanger Institute for the COVID-19 Genomics UK Consortium |
| EPI_ISL_458856 | PHE South West Regional Laboratory, National Infection Service | Wellcome Sanger Institute for the COVID-19 Genomics UK Consortium |
| EPI_ISL_458857 | PHE South West Regional Laboratory, National Infection Service | Wellcome Sanger Institute for the COVID-19 Genomics UK Consortium |

| EPI_Accession_ID | Originating Laboratory | Submitting Laboratory |
|---|---|---|
| EPI_ISL_458858 | PHE South West Regional Laboratory, National Infection Service | Wellcome Sanger Institute for the COVID-19 Genomics UK Consortium |
| EPI_ISL_458859 | PHE South West Regional Laboratory, National Infection Service | Wellcome Sanger Institute for the COVID-19 Genomics UK Consortium |
| EPI_ISL_458860 | PHE South West Regional Laboratory, National Infection Service | Wellcome Sanger Institute for the COVID-19 Genomics UK Consortium |
| EPI_ISL_458861 | PHE South West Regional Laboratory, National Infection Service | Wellcome Sanger Institute for the COVID-19 Genomics UK Consortium |
| EPI_ISL_458862 | PHE South West Regional Laboratory, National Infection Service | Wellcome Sanger Institute for the COVID-19 Genomics UK Consortium |
| EPI_ISL_458863 | PHE South West Regional Laboratory, National Infection Service | Wellcome Sanger Institute for the COVID-19 Genomics UK Consortium |
| EPI_ISL_458864 | PHE South West Regional Laboratory, National Infection Service | Wellcome Sanger Institute for the COVID-19 Genomics UK Consortium |
| EPI_ISL_458865 | PHE South West Regional Laboratory, National Infection Service | Wellcome Sanger Institute for the COVID-19 Genomics UK Consortium |
| EPI_ISL_458866 | PHE South West Regional Laboratory, National Infection Service | Wellcome Sanger Institute for the COVID-19 Genomics UK Consortium |
| EPI_ISL_458867 | PHE South West Regional Laboratory, National Infection Service | Wellcome Sanger Institute for the COVID-19 Genomics UK Consortium |
| EPI_ISL_458868 | PHE South West Regional Laboratory, National Infection Service | Wellcome Sanger Institute for the COVID-19 Genomics UK Consortium |
| EPI_ISL_458869 | PHE South West Regional Laboratory, National Infection Service | Wellcome Sanger Institute for the COVID-19 Genomics UK Consortium |
| EPI_ISL_458870 | PHE South West Regional Laboratory, National Infection Service | Wellcome Sanger Institute for the COVID-19 Genomics UK Consortium |
| EPI_ISL_458871 | PHE South West Regional Laboratory, National Infection Service | Wellcome Sanger Institute for the COVID-19 Genomics UK Consortium |
| EPI_ISL_458872 | PHE South West Regional Laboratory, National Infection Service | Wellcome Sanger Institute for the COVID-19 Genomics UK Consortium |
| EPI_ISL_458873 | PHE South West Regional Laboratory, National Infection Service | Wellcome Sanger Institute for the COVID-19 Genomics UK Consortium |

| EPI_Accession_ID | Originating Laboratory | Submitting Laboratory |
|---|---|---|
| EPI_ISL_458874 | PHE South West Regional Laboratory, National Infection Service | Wellcome Sanger Institute for the COVID-19 Genomics UK Consortium |
| EPI_ISL_458875 | PHE South West Regional Laboratory, National Infection Service | Wellcome Sanger Institute for the COVID-19 Genomics UK Consortium |
| EPI_ISL_458876 | PHE South West Regional Laboratory, National Infection Service | Wellcome Sanger Institute for the COVID-19 Genomics UK Consortium |
| EPI_ISL_458877 | PHE South West Regional Laboratory, National Infection Service | Wellcome Sanger Institute for the COVID-19 Genomics UK Consortium |
| EPI_ISL_458878 | PHE South West Regional Laboratory, National Infection Service | Wellcome Sanger Institute for the COVID-19 Genomics UK Consortium |
| EPI_ISL_458879 | PHE South West Regional Laboratory, National Infection Service | Wellcome Sanger Institute for the COVID-19 Genomics UK Consortium |
| EPI_ISL_458880 | PHE South West Regional Laboratory, National Infection Service | Wellcome Sanger Institute for the COVID-19 Genomics UK Consortium |
| EPI_ISL_458881 | PHE South West Regional Laboratory, National Infection Service | Wellcome Sanger Institute for the COVID-19 Genomics UK Consortium |
| EPI_ISL_458882 | PHE South West Regional Laboratory, National Infection Service | Wellcome Sanger Institute for the COVID-19 Genomics UK Consortium |
| EPI_ISL_458883 | PHE South West Regional Laboratory, National Infection Service | Wellcome Sanger Institute for the COVID-19 Genomics UK Consortium |
| EPI_ISL_458884 | PHE South West Regional Laboratory, National Infection Service | Wellcome Sanger Institute for the COVID-19 Genomics UK Consortium |
| EPI_ISL_458885 | PHE South West Regional Laboratory, National Infection Service | Wellcome Sanger Institute for the COVID-19 Genomics UK Consortium |
| EPI_ISL_458886 | PHE South West Regional Laboratory, National Infection Service | Wellcome Sanger Institute for the COVID-19 Genomics UK Consortium |
| EPI_ISL_458887 | PHE South West Regional Laboratory, National Infection Service | Wellcome Sanger Institute for the COVID-19 Genomics UK Consortium |
| EPI_ISL_458888 | PHE South West Regional Laboratory, National Infection Service | Wellcome Sanger Institute for the COVID-19 Genomics UK Consortium |
| EPI_ISL_458889 | PHE South West Regional Laboratory, National Infection Service | Wellcome Sanger Institute for the COVID-19 Genomics UK Consortium |

| EPI_Accession_ID | Originating Laboratory | Submitting Laboratory |
| --- | --- | --- |
| EPI_ISL_458890 | PHE South West Regional Laboratory, National Infection Service | Wellcome Sanger Institute for the COVID-19 Genomics UK Consortium |
| EPI_ISL_458891 | PHE South West Regional Laboratory, National Infection Service | Wellcome Sanger Institute for the COVID-19 Genomics UK Consortium |
| EPI_ISL_458892 | PHE South West Regional Laboratory, National Infection Service | Wellcome Sanger Institute for the COVID-19 Genomics UK Consortium |
| EPI_ISL_458893 | PHE South West Regional Laboratory, National Infection Service | Wellcome Sanger Institute for the COVID-19 Genomics UK Consortium |
| EPI_ISL_458894 | PHE South West Regional Laboratory, National Infection Service | Wellcome Sanger Institute for the COVID-19 Genomics UK Consortium |
| EPI_ISL_458895 | PHE South West Regional Laboratory, National Infection Service | Wellcome Sanger Institute for the COVID-19 Genomics UK Consortium |
| EPI_ISL_458896 | PHE South West Regional Laboratory, National Infection Service | Wellcome Sanger Institute for the COVID-19 Genomics UK Consortium |
| EPI_ISL_458897 | PHE South West Regional Laboratory, National Infection Service | Wellcome Sanger Institute for the COVID-19 Genomics UK Consortium |
| EPI_ISL_458898 | PHE South West Regional Laboratory, National Infection Service | Wellcome Sanger Institute for the COVID-19 Genomics UK Consortium |
| EPI_ISL_458899 | PHE South West Regional Laboratory, National Infection Service | Wellcome Sanger Institute for the COVID-19 Genomics UK Consortium |
| EPI_ISL_458900 | PHE South West Regional Laboratory, National Infection Service | Wellcome Sanger Institute for the COVID-19 Genomics UK Consortium |
| EPI_ISL_458901 | PHE South West Regional Laboratory, National Infection Service | Wellcome Sanger Institute for the COVID-19 Genomics UK Consortium |
| EPI_ISL_458902 | PHE South West Regional Laboratory, National Infection Service | Wellcome Sanger Institute for the COVID-19 Genomics UK Consortium |
| EPI_ISL_458903 | PHE South West Regional Laboratory, National Infection Service | Wellcome Sanger Institute for the COVID-19 Genomics UK Consortium |
| EPI_ISL_458904 | PHE South West Regional Laboratory, National Infection Service | Wellcome Sanger Institute for the COVID-19 Genomics UK Consortium |
| EPI_ISL_458905 | PHE South West Regional Laboratory, National Infection Service | Wellcome Sanger Institute for the COVID-19 Genomics UK Consortium |

| EPI_Accession_ID | Originating Laboratory | Submitting Laboratory |
|---|---|---|
| EPI_ISL_458906 | PHE South West Regional Laboratory, National Infection Service | Wellcome Sanger Institute for the COVID-19 Genomics UK Consortium |
| EPI_ISL_458907 | PHE South West Regional Laboratory, National Infection Service | Wellcome Sanger Institute for the COVID-19 Genomics UK Consortium |
| EPI_ISL_458908 | PHE South West Regional Laboratory, National Infection Service | Wellcome Sanger Institute for the COVID-19 Genomics UK Consortium |
| EPI_ISL_458909 | PHE South West Regional Laboratory, National Infection Service | Wellcome Sanger Institute for the COVID-19 Genomics UK Consortium |
| EPI_ISL_458910 | PHE South West Regional Laboratory, National Infection Service | Wellcome Sanger Institute for the COVID-19 Genomics UK Consortium |
| EPI_ISL_458911 | PHE South West Regional Laboratory, National Infection Service | Wellcome Sanger Institute for the COVID-19 Genomics UK Consortium |
| EPI_ISL_458912 | PHE South West Regional Laboratory, National Infection Service | Wellcome Sanger Institute for the COVID-19 Genomics UK Consortium |
| EPI_ISL_458913 | PHE South West Regional Laboratory, National Infection Service | Wellcome Sanger Institute for the COVID-19 Genomics UK Consortium |
| EPI_ISL_458914 | PHE South West Regional Laboratory, National Infection Service | Wellcome Sanger Institute for the COVID-19 Genomics UK Consortium |
| EPI_ISL_458915 | PHE South West Regional Laboratory, National Infection Service | Wellcome Sanger Institute for the COVID-19 Genomics UK Consortium |
| EPI_ISL_458916 | PHE South West Regional Laboratory, National Infection Service | Wellcome Sanger Institute for the COVID-19 Genomics UK Consortium |
| EPI_ISL_458917 | PHE South West Regional Laboratory, National Infection Service | Wellcome Sanger Institute for the COVID-19 Genomics UK Consortium |
| EPI_ISL_458918 | PHE South West Regional Laboratory, National Infection Service | Wellcome Sanger Institute for the COVID-19 Genomics UK Consortium |
| EPI_ISL_458919 | PHE South West Regional Laboratory, National Infection Service | Wellcome Sanger Institute for the COVID-19 Genomics UK Consortium |
| EPI_ISL_458920 | PHE South West Regional Laboratory, National Infection Service | Wellcome Sanger Institute for the COVID-19 Genomics UK Consortium |
| EPI_ISL_458921 | PHE South West Regional Laboratory, National Infection Service | Wellcome Sanger Institute for the COVID-19 Genomics UK Consortium |

| EPI_Accession_ID | Originating Laboratory | Submitting Laboratory |
|---|---|---|
| EPI_ISL_458922 | PHE South West Regional Laboratory, National Infection Service | Wellcome Sanger Institute for the COVID-19 Genomics UK Consortium |
| EPI_ISL_458923 | PHE South West Regional Laboratory, National Infection Service | Wellcome Sanger Institute for the COVID-19 Genomics UK Consortium |
| EPI_ISL_458924 | PHE South West Regional Laboratory, National Infection Service | Wellcome Sanger Institute for the COVID-19 Genomics UK Consortium |
| EPI_ISL_458925 | PHE South West Regional Laboratory, National Infection Service | Wellcome Sanger Institute for the COVID-19 Genomics UK Consortium |
| EPI_ISL_458926 | PHE South West Regional Laboratory, National Infection Service | Wellcome Sanger Institute for the COVID-19 Genomics UK Consortium |
| EPI_ISL_458927 | PHE South West Regional Laboratory, National Infection Service | Wellcome Sanger Institute for the COVID-19 Genomics UK Consortium |
| EPI_ISL_458928 | PHE South West Regional Laboratory, National Infection Service | Wellcome Sanger Institute for the COVID-19 Genomics UK Consortium |
| EPI_ISL_458929 | PHE South West Regional Laboratory, National Infection Service | Wellcome Sanger Institute for the COVID-19 Genomics UK Consortium |
| EPI_ISL_458930 | PHE South West Regional Laboratory, National Infection Service | Wellcome Sanger Institute for the COVID-19 Genomics UK Consortium |
| EPI_ISL_458931 | PHE South West Regional Laboratory, National Infection Service | Wellcome Sanger Institute for the COVID-19 Genomics UK Consortium |
| EPI_ISL_458932 | PHE South West Regional Laboratory, National Infection Service | Wellcome Sanger Institute for the COVID-19 Genomics UK Consortium |
| EPI_ISL_458933 | PHE South West Regional Laboratory, National Infection Service | Wellcome Sanger Institute for the COVID-19 Genomics UK Consortium |
| EPI_ISL_458934 | PHE South West Regional Laboratory, National Infection Service | Wellcome Sanger Institute for the COVID-19 Genomics UK Consortium |
| EPI_ISL_458935 | PHE South West Regional Laboratory, National Infection Service | Wellcome Sanger Institute for the COVID-19 Genomics UK Consortium |
| EPI_ISL_458936 | PHE South West Regional Laboratory, National Infection Service | Wellcome Sanger Institute for the COVID-19 Genomics UK Consortium |
| EPI_ISL_458937 | PHE South West Regional Laboratory, National Infection Service | Wellcome Sanger Institute for the COVID-19 Genomics UK Consortium |

| EPI_Accession_ID | Originating Laboratory | Submitting Laboratory |
|---|---|---|
| EPI_ISL_458938 | PHE South West Regional Laboratory, National Infection Service | Wellcome Sanger Institute for the COVID-19 Genomics UK Consortium |
| EPI_ISL_458939 | PHE South West Regional Laboratory, National Infection Service | Wellcome Sanger Institute for the COVID-19 Genomics UK Consortium |
| EPI_ISL_458940 | PHE South West Regional Laboratory, National Infection Service | Wellcome Sanger Institute for the COVID-19 Genomics UK Consortium |
| EPI_ISL_458941 | PHE South West Regional Laboratory, National Infection Service | Wellcome Sanger Institute for the COVID-19 Genomics UK Consortium |
| EPI_ISL_458942 | PHE South West Regional Laboratory, National Infection Service | Wellcome Sanger Institute for the COVID-19 Genomics UK Consortium |
| EPI_ISL_458943 | PHE South West Regional Laboratory, National Infection Service | Wellcome Sanger Institute for the COVID-19 Genomics UK Consortium |
| EPI_ISL_458944 | PHE South West Regional Laboratory, National Infection Service | Wellcome Sanger Institute for the COVID-19 Genomics UK Consortium |
| EPI_ISL_458945 | PHE South West Regional Laboratory, National Infection Service | Wellcome Sanger Institute for the COVID-19 Genomics UK Consortium |
| EPI_ISL_458946 | PHE South West Regional Laboratory, National Infection Service | Wellcome Sanger Institute for the COVID-19 Genomics UK Consortium |
| EPI_ISL_458947 | PHE South West Regional Laboratory, National Infection Service | Wellcome Sanger Institute for the COVID-19 Genomics UK Consortium |
| EPI_ISL_458948 | PHE South West Regional Laboratory, National Infection Service | Wellcome Sanger Institute for the COVID-19 Genomics UK Consortium |
| EPI_ISL_458949 | PHE South West Regional Laboratory, National Infection Service | Wellcome Sanger Institute for the COVID-19 Genomics UK Consortium |
| EPI_ISL_458950 | PHE South West Regional Laboratory, National Infection Service | Wellcome Sanger Institute for the COVID-19 Genomics UK Consortium |
| EPI_ISL_458951 | PHE South West Regional Laboratory, National Infection Service | Wellcome Sanger Institute for the COVID-19 Genomics UK Consortium |
| EPI_ISL_458952 | PHE South West Regional Laboratory, National Infection Service | Wellcome Sanger Institute for the COVID-19 Genomics UK Consortium |
| EPI_ISL_458953 | PHE South West Regional Laboratory, National Infection Service | Wellcome Sanger Institute for the COVID-19 Genomics UK Consortium |

| EPI_Accession_ID | Originating Laboratory | Submitting Laboratory |
|---|---|---|
| EPI_ISL_458954 | PHE South West Regional Laboratory, National Infection Service | Wellcome Sanger Institute for the COVID-19 Genomics UK Consortium |
| EPI_ISL_458955 | PHE South West Regional Laboratory, National Infection Service | Wellcome Sanger Institute for the COVID-19 Genomics UK Consortium |
| EPI_ISL_458956 | PHE South West Regional Laboratory, National Infection Service | Wellcome Sanger Institute for the COVID-19 Genomics UK Consortium |
| EPI_ISL_458957 | PHE South West Regional Laboratory, National Infection Service | Wellcome Sanger Institute for the COVID-19 Genomics UK Consortium |
| EPI_ISL_458958 | PHE South West Regional Laboratory, National Infection Service | Wellcome Sanger Institute for the COVID-19 Genomics UK Consortium |
| EPI_ISL_458959 | PHE South West Regional Laboratory, National Infection Service | Wellcome Sanger Institute for the COVID-19 Genomics UK Consortium |
| EPI_ISL_458960 | PHE South West Regional Laboratory, National Infection Service | Wellcome Sanger Institute for the COVID-19 Genomics UK Consortium |
| EPI_ISL_458961 | PHE South West Regional Laboratory, National Infection Service | Wellcome Sanger Institute for the COVID-19 Genomics UK Consortium |
| EPI_ISL_458962 | PHE South West Regional Laboratory, National Infection Service | Wellcome Sanger Institute for the COVID-19 Genomics UK Consortium |
| EPI_ISL_458963 | PHE South West Regional Laboratory, National Infection Service | Wellcome Sanger Institute for the COVID-19 Genomics UK Consortium |
| EPI_ISL_458964 | PHE South West Regional Laboratory, National Infection Service | Wellcome Sanger Institute for the COVID-19 Genomics UK Consortium |
| EPI_ISL_458965 | PHE South West Regional Laboratory, National Infection Service | Wellcome Sanger Institute for the COVID-19 Genomics UK Consortium |
| EPI_ISL_458966 | PHE South West Regional Laboratory, National Infection Service | Wellcome Sanger Institute for the COVID-19 Genomics UK Consortium |
| EPI_ISL_458967 | PHE South West Regional Laboratory, National Infection Service | Wellcome Sanger Institute for the COVID-19 Genomics UK Consortium |
| EPI_ISL_458968 | PHE South West Regional Laboratory, National Infection Service | Wellcome Sanger Institute for the COVID-19 Genomics UK Consortium |
| EPI_ISL_458969 | PHE South West Regional Laboratory, National Infection Service | Wellcome Sanger Institute for the COVID-19 Genomics UK Consortium |

| EPI_Accession_ID | Originating Laboratory | Submitting Laboratory |
|---|---|---|
| EPI_ISL_458970 | PHE South West Regional Laboratory, National Infection Service | Wellcome Sanger Institute for the COVID-19 Genomics UK Consortium |
| EPI_ISL_458971 | PHE South West Regional Laboratory, National Infection Service | Wellcome Sanger Institute for the COVID-19 Genomics UK Consortium |
| EPI_ISL_458972 | PHE South West Regional Laboratory, National Infection Service | Wellcome Sanger Institute for the COVID-19 Genomics UK Consortium |
| EPI_ISL_458973 | PHE South West Regional Laboratory, National Infection Service | Wellcome Sanger Institute for the COVID-19 Genomics UK Consortium |
| EPI_ISL_458974 | PHE South West Regional Laboratory, National Infection Service | Wellcome Sanger Institute for the COVID-19 Genomics UK Consortium |
| EPI_ISL_458975 | PHE South West Regional Laboratory, National Infection Service | Wellcome Sanger Institute for the COVID-19 Genomics UK Consortium |
| EPI_ISL_458976 | PHE South West Regional Laboratory, National Infection Service | Wellcome Sanger Institute for the COVID-19 Genomics UK Consortium |
| EPI_ISL_458977 | PHE South West Regional Laboratory, National Infection Service | Wellcome Sanger Institute for the COVID-19 Genomics UK Consortium |
| EPI_ISL_458978 | PHE South West Regional Laboratory, National Infection Service | Wellcome Sanger Institute for the COVID-19 Genomics UK Consortium |
| EPI_ISL_458979 | PHE South West Regional Laboratory, National Infection Service | Wellcome Sanger Institute for the COVID-19 Genomics UK Consortium |
| EPI_ISL_458980 | PHE South West Regional Laboratory, National Infection Service | Wellcome Sanger Institute for the COVID-19 Genomics UK Consortium |
| EPI_ISL_458981 | PHE South West Regional Laboratory, National Infection Service | Wellcome Sanger Institute for the COVID-19 Genomics UK Consortium |
| EPI_ISL_458982 | PHE South West Regional Laboratory, National Infection Service | Wellcome Sanger Institute for the COVID-19 Genomics UK Consortium |
| EPI_ISL_458983 | PHE South West Regional Laboratory, National Infection Service | Wellcome Sanger Institute for the COVID-19 Genomics UK Consortium |
| EPI_ISL_458984 | PHE South West Regional Laboratory, National Infection Service | Wellcome Sanger Institute for the COVID-19 Genomics UK Consortium |
| EPI_ISL_458985 | PHE South West Regional Laboratory, National Infection Service | Wellcome Sanger Institute for the COVID-19 Genomics UK Consortium |

| EPI_Accession_ID | Originating Laboratory | Submitting Laboratory |
| --- | --- | --- |
| EPI_ISL_458986 | PHE South West Regional Laboratory, National Infection Service | Wellcome Sanger Institute for the COVID-19 Genomics UK Consortium |
| EPI_ISL_458987 | PHE South West Regional Laboratory, National Infection Service | Wellcome Sanger Institute for the COVID-19 Genomics UK Consortium |
| EPI_ISL_458988 | PHE South West Regional Laboratory, National Infection Service | Wellcome Sanger Institute for the COVID-19 Genomics UK Consortium |
| EPI_ISL_458989 | PHE South West Regional Laboratory, National Infection Service | Wellcome Sanger Institute for the COVID-19 Genomics UK Consortium |
| EPI_ISL_458990 | PHE South West Regional Laboratory, National Infection Service | Wellcome Sanger Institute for the COVID-19 Genomics UK Consortium |
| EPI_ISL_458991 | PHE South West Regional Laboratory, National Infection Service | Wellcome Sanger Institute for the COVID-19 Genomics UK Consortium |
| EPI_ISL_458992 | PHE South West Regional Laboratory, National Infection Service | Wellcome Sanger Institute for the COVID-19 Genomics UK Consortium |
| EPI_ISL_458993 | PHE South West Regional Laboratory, National Infection Service | Wellcome Sanger Institute for the COVID-19 Genomics UK Consortium |
| EPI_ISL_458994 | PHE South West Regional Laboratory, National Infection Service | Wellcome Sanger Institute for the COVID-19 Genomics UK Consortium |
| EPI_ISL_458995 | PHE South West Regional Laboratory, National Infection Service | Wellcome Sanger Institute for the COVID-19 Genomics UK Consortium |
| EPI_ISL_458996 | PHE South West Regional Laboratory, National Infection Service | Wellcome Sanger Institute for the COVID-19 Genomics UK Consortium |
| EPI_ISL_458997 | PHE South West Regional Laboratory, National Infection Service | Wellcome Sanger Institute for the COVID-19 Genomics UK Consortium |
| EPI_ISL_458998 | PHE South West Regional Laboratory, National Infection Service | Wellcome Sanger Institute for the COVID-19 Genomics UK Consortium |
| EPI_ISL_458999 | PHE South West Regional Laboratory, National Infection Service | Wellcome Sanger Institute for the COVID-19 Genomics UK Consortium |
| EPI_ISL_459000 | PHE South West Regional Laboratory, National Infection Service | Wellcome Sanger Institute for the COVID-19 Genomics UK Consortium |
| EPI_ISL_459001 | PHE South West Regional Laboratory, National Infection Service | Wellcome Sanger Institute for the COVID-19 Genomics UK Consortium |

| EPI_Accession_ID | Originating Laboratory | Submitting Laboratory |
|---|---|---|
| EPI_ISL_459002 | PHE South West Regional Laboratory, National Infection Service | Wellcome Sanger Institute for the COVID-19 Genomics UK Consortium |
| EPI_ISL_459003 | PHE South West Regional Laboratory, National Infection Service | Wellcome Sanger Institute for the COVID-19 Genomics UK Consortium |
| EPI_ISL_459004 | PHE South West Regional Laboratory, National Infection Service | Wellcome Sanger Institute for the COVID-19 Genomics UK Consortium |
| EPI_ISL_459005 | PHE South West Regional Laboratory, National Infection Service | Wellcome Sanger Institute for the COVID-19 Genomics UK Consortium |
| EPI_ISL_459006 | PHE South West Regional Laboratory, National Infection Service | Wellcome Sanger Institute for the COVID-19 Genomics UK Consortium |
| EPI_ISL_459007 | PHE South West Regional Laboratory, National Infection Service | Wellcome Sanger Institute for the COVID-19 Genomics UK Consortium |
| EPI_ISL_459008 | PHE South West Regional Laboratory, National Infection Service | Wellcome Sanger Institute for the COVID-19 Genomics UK Consortium |
| EPI_ISL_459009 | PHE South West Regional Laboratory, National Infection Service | Wellcome Sanger Institute for the COVID-19 Genomics UK Consortium |
| EPI_ISL_459010 | PHE South West Regional Laboratory, National Infection Service | Wellcome Sanger Institute for the COVID-19 Genomics UK Consortium |
| EPI_ISL_459011 | PHE South West Regional Laboratory, National Infection Service | Wellcome Sanger Institute for the COVID-19 Genomics UK Consortium |
| EPI_ISL_459012 | PHE South West Regional Laboratory, National Infection Service | Wellcome Sanger Institute for the COVID-19 Genomics UK Consortium |
| EPI_ISL_459013 | PHE South West Regional Laboratory, National Infection Service | Wellcome Sanger Institute for the COVID-19 Genomics UK Consortium |
| EPI_ISL_459014 | PHE South West Regional Laboratory, National Infection Service | Wellcome Sanger Institute for the COVID-19 Genomics UK Consortium |
| EPI_ISL_459015 | PHE South West Regional Laboratory, National Infection Service | Wellcome Sanger Institute for the COVID-19 Genomics UK Consortium |
| EPI_ISL_459016 | PHE South West Regional Laboratory, National Infection Service | Wellcome Sanger Institute for the COVID-19 Genomics UK Consortium |
| EPI_ISL_459017 | PHE South West Regional Laboratory, National Infection Service | Wellcome Sanger Institute for the COVID-19 Genomics UK Consortium |

| EPI_Accession_ID | Originating Laboratory | Submitting Laboratory |
|---|---|---|
| EPI_ISL_459018 | PHE South West Regional Laboratory, National Infection Service | Wellcome Sanger Institute for the COVID-19 Genomics UK Consortium |
| EPI_ISL_459019 | PHE South West Regional Laboratory, National Infection Service | Wellcome Sanger Institute for the COVID-19 Genomics UK Consortium |
| EPI_ISL_459020 | PHE South West Regional Laboratory, National Infection Service | Wellcome Sanger Institute for the COVID-19 Genomics UK Consortium |
| EPI_ISL_459021 | PHE South West Regional Laboratory, National Infection Service | Wellcome Sanger Institute for the COVID-19 Genomics UK Consortium |
| EPI_ISL_459022 | PHE South West Regional Laboratory, National Infection Service | Wellcome Sanger Institute for the COVID-19 Genomics UK Consortium |
| EPI_ISL_459023 | PHE South West Regional Laboratory, National Infection Service | Wellcome Sanger Institute for the COVID-19 Genomics UK Consortium |
| EPI_ISL_459024 | PHE South West Regional Laboratory, National Infection Service | Wellcome Sanger Institute for the COVID-19 Genomics UK Consortium |
| EPI_ISL_459025 | PHE South West Regional Laboratory, National Infection Service | Wellcome Sanger Institute for the COVID-19 Genomics UK Consortium |
| EPI_ISL_459026 | PHE South West Regional Laboratory, National Infection Service | Wellcome Sanger Institute for the COVID-19 Genomics UK Consortium |
| EPI_ISL_459027 | PHE South West Regional Laboratory, National Infection Service | Wellcome Sanger Institute for the COVID-19 Genomics UK Consortium |
| EPI_ISL_459028 | PHE South West Regional Laboratory, National Infection Service | Wellcome Sanger Institute for the COVID-19 Genomics UK Consortium |
| EPI_ISL_459029 | PHE South West Regional Laboratory, National Infection Service | Wellcome Sanger Institute for the COVID-19 Genomics UK Consortium |
| EPI_ISL_459030 | PHE South West Regional Laboratory, National Infection Service | Wellcome Sanger Institute for the COVID-19 Genomics UK Consortium |
| EPI_ISL_459031 | PHE South West Regional Laboratory, National Infection Service | Wellcome Sanger Institute for the COVID-19 Genomics UK Consortium |
| EPI_ISL_459032 | PHE South West Regional Laboratory, National Infection Service | Wellcome Sanger Institute for the COVID-19 Genomics UK Consortium |
| EPI_ISL_459033 | PHE South West Regional Laboratory, National Infection Service | Wellcome Sanger Institute for the COVID-19 Genomics UK Consortium |

| EPI_Accession_ID | Originating Laboratory | Submitting Laboratory |
|---|---|---|
| EPI_ISL_459034 | PHE South West Regional Laboratory, National Infection Service | Wellcome Sanger Institute for the COVID-19 Genomics UK Consortium |
| EPI_ISL_459035 | PHE South West Regional Laboratory, National Infection Service | Wellcome Sanger Institute for the COVID-19 Genomics UK Consortium |
| EPI_ISL_459036 | PHE South West Regional Laboratory, National Infection Service | Wellcome Sanger Institute for the COVID-19 Genomics UK Consortium |
| EPI_ISL_459037 | PHE South West Regional Laboratory, National Infection Service | Wellcome Sanger Institute for the COVID-19 Genomics UK Consortium |
| EPI_ISL_459038 | PHE South West Regional Laboratory, National Infection Service | Wellcome Sanger Institute for the COVID-19 Genomics UK Consortium |
| EPI_ISL_459039 | PHE South West Regional Laboratory, National Infection Service | Wellcome Sanger Institute for the COVID-19 Genomics UK Consortium |
| EPI_ISL_459040 | PHE South West Regional Laboratory, National Infection Service | Wellcome Sanger Institute for the COVID-19 Genomics UK Consortium |
| EPI_ISL_459041 | PHE South West Regional Laboratory, National Infection Service | Wellcome Sanger Institute for the COVID-19 Genomics UK Consortium |
| EPI_ISL_459042 | PHE South West Regional Laboratory, National Infection Service | Wellcome Sanger Institute for the COVID-19 Genomics UK Consortium |
| EPI_ISL_459043 | PHE South West Regional Laboratory, National Infection Service | Wellcome Sanger Institute for the COVID-19 Genomics UK Consortium |
| EPI_ISL_459044 | PHE South West Regional Laboratory, National Infection Service | Wellcome Sanger Institute for the COVID-19 Genomics UK Consortium |
| EPI_ISL_459045 | PHE South West Regional Laboratory, National Infection Service | Wellcome Sanger Institute for the COVID-19 Genomics UK Consortium |
| EPI_ISL_459046 | PHE South West Regional Laboratory, National Infection Service | Wellcome Sanger Institute for the COVID-19 Genomics UK Consortium |
| EPI_ISL_459047 | PHE South West Regional Laboratory, National Infection Service | Wellcome Sanger Institute for the COVID-19 Genomics UK Consortium |
| EPI_ISL_459048 | PHE South West Regional Laboratory, National Infection Service | Wellcome Sanger Institute for the COVID-19 Genomics UK Consortium |
| EPI_ISL_459049 | PHE South West Regional Laboratory, National Infection Service | Wellcome Sanger Institute for the COVID-19 Genomics UK Consortium |

| EPI_Accession_ID | Originating Laboratory | Submitting Laboratory |
|---|---|---|
| EPI_ISL_459050 | PHE South West Regional Laboratory, National Infection Service | Wellcome Sanger Institute for the COVID-19 Genomics UK Consortium |
| EPI_ISL_459051 | PHE South West Regional Laboratory, National Infection Service | Wellcome Sanger Institute for the COVID-19 Genomics UK Consortium |
| EPI_ISL_459052 | PHE South West Regional Laboratory, National Infection Service | Wellcome Sanger Institute for the COVID-19 Genomics UK Consortium |
| EPI_ISL_459053 | PHE South West Regional Laboratory, National Infection Service | Wellcome Sanger Institute for the COVID-19 Genomics UK Consortium |
| EPI_ISL_459054 | PHE South West Regional Laboratory, National Infection Service | Wellcome Sanger Institute for the COVID-19 Genomics UK Consortium |
| EPI_ISL_459055 | PHE South West Regional Laboratory, National Infection Service | Wellcome Sanger Institute for the COVID-19 Genomics UK Consortium |
| EPI_ISL_459056 | PHE South West Regional Laboratory, National Infection Service | Wellcome Sanger Institute for the COVID-19 Genomics UK Consortium |
| EPI_ISL_459057 | PHE South West Regional Laboratory, National Infection Service | Wellcome Sanger Institute for the COVID-19 Genomics UK Consortium |
| EPI_ISL_459058 | PHE South West Regional Laboratory, National Infection Service | Wellcome Sanger Institute for the COVID-19 Genomics UK Consortium |
| EPI_ISL_459059 | PHE South West Regional Laboratory, National Infection Service | Wellcome Sanger Institute for the COVID-19 Genomics UK Consortium |
| EPI_ISL_459060 | PHE South West Regional Laboratory, National Infection Service | Wellcome Sanger Institute for the COVID-19 Genomics UK Consortium |
| EPI_ISL_459061 | PHE South West Regional Laboratory, National Infection Service | Wellcome Sanger Institute for the COVID-19 Genomics UK Consortium |
| EPI_ISL_459062 | PHE South West Regional Laboratory, National Infection Service | Wellcome Sanger Institute for the COVID-19 Genomics UK Consortium |
| EPI_ISL_459063 | PHE South West Regional Laboratory, National Infection Service | Wellcome Sanger Institute for the COVID-19 Genomics UK Consortium |
| EPI_ISL_459064 | PHE South West Regional Laboratory, National Infection Service | Wellcome Sanger Institute for the COVID-19 Genomics UK Consortium |
| EPI_ISL_459065 | PHE South West Regional Laboratory, National Infection Service | Wellcome Sanger Institute for the COVID-19 Genomics UK Consortium |

| EPI_Accession_ID | Originating Laboratory | Submitting Laboratory |
|---|---|---|
| EPI_ISL_459066 | PHE South West Regional Laboratory, National Infection Service | Wellcome Sanger Institute for the COVID-19 Genomics UK Consortium |
| EPI_ISL_459067 | PHE South West Regional Laboratory, National Infection Service | Wellcome Sanger Institute for the COVID-19 Genomics UK Consortium |
| EPI_ISL_459068 | PHE South West Regional Laboratory, National Infection Service | Wellcome Sanger Institute for the COVID-19 Genomics UK Consortium |
| EPI_ISL_459069 | PHE South West Regional Laboratory, National Infection Service | Wellcome Sanger Institute for the COVID-19 Genomics UK Consortium |
| EPI_ISL_459070 | PHE South West Regional Laboratory, National Infection Service | Wellcome Sanger Institute for the COVID-19 Genomics UK Consortium |
| EPI_ISL_459071 | PHE South West Regional Laboratory, National Infection Service | Wellcome Sanger Institute for the COVID-19 Genomics UK Consortium |
| EPI_ISL_459072 | PHE South West Regional Laboratory, National Infection Service | Wellcome Sanger Institute for the COVID-19 Genomics UK Consortium |
| EPI_ISL_459073 | PHE South West Regional Laboratory, National Infection Service | Wellcome Sanger Institute for the COVID-19 Genomics UK Consortium |
| EPI_ISL_459074 | PHE South West Regional Laboratory, National Infection Service | Wellcome Sanger Institute for the COVID-19 Genomics UK Consortium |
| EPI_ISL_459075 | PHE South West Regional Laboratory, National Infection Service | Wellcome Sanger Institute for the COVID-19 Genomics UK Consortium |
| EPI_ISL_459076 | PHE South West Regional Laboratory, National Infection Service | Wellcome Sanger Institute for the COVID-19 Genomics UK Consortium |
| EPI_ISL_459077 | PHE South West Regional Laboratory, National Infection Service | Wellcome Sanger Institute for the COVID-19 Genomics UK Consortium |
| EPI_ISL_459078 | PHE South West Regional Laboratory, National Infection Service | Wellcome Sanger Institute for the COVID-19 Genomics UK Consortium |
| EPI_ISL_459079 | PHE South West Regional Laboratory, National Infection Service | Wellcome Sanger Institute for the COVID-19 Genomics UK Consortium |
| EPI_ISL_459080 | PHE South West Regional Laboratory, National Infection Service | Wellcome Sanger Institute for the COVID-19 Genomics UK Consortium |
| EPI_ISL_459081 | PHE South West Regional Laboratory, National Infection Service | Wellcome Sanger Institute for the COVID-19 Genomics UK Consortium |

| EPI_Accession_ID | Originating Laboratory | Submitting Laboratory |
|---|---|---|
| EPI_ISL_459082 | PHE South West Regional Laboratory, National Infection Service | Wellcome Sanger Institute for the COVID-19 Genomics UK Consortium |
| EPI_ISL_459083 | PHE South West Regional Laboratory, National Infection Service | Wellcome Sanger Institute for the COVID-19 Genomics UK Consortium |
| EPI_ISL_459084 | PHE South West Regional Laboratory, National Infection Service | Wellcome Sanger Institute for the COVID-19 Genomics UK Consortium |
| EPI_ISL_459085 | PHE South West Regional Laboratory, National Infection Service | Wellcome Sanger Institute for the COVID-19 Genomics UK Consortium |
| EPI_ISL_459086 | PHE South West Regional Laboratory, National Infection Service | Wellcome Sanger Institute for the COVID-19 Genomics UK Consortium |
| EPI_ISL_459087 | PHE South West Regional Laboratory, National Infection Service | Wellcome Sanger Institute for the COVID-19 Genomics UK Consortium |
| EPI_ISL_459088 | PHE South West Regional Laboratory, National Infection Service | Wellcome Sanger Institute for the COVID-19 Genomics UK Consortium |
| EPI_ISL_459089 | PHE South West Regional Laboratory, National Infection Service | Wellcome Sanger Institute for the COVID-19 Genomics UK Consortium |
| EPI_ISL_459090 | PHE South West Regional Laboratory, National Infection Service | Wellcome Sanger Institute for the COVID-19 Genomics UK Consortium |
| EPI_ISL_459091 | PHE South West Regional Laboratory, National Infection Service | Wellcome Sanger Institute for the COVID-19 Genomics UK Consortium |
| EPI_ISL_459092 | PHE South West Regional Laboratory, National Infection Service | Wellcome Sanger Institute for the COVID-19 Genomics UK Consortium |
| EPI_ISL_459093 | PHE South West Regional Laboratory, National Infection Service | Wellcome Sanger Institute for the COVID-19 Genomics UK Consortium |
| EPI_ISL_459094 | PHE South West Regional Laboratory, National Infection Service | Wellcome Sanger Institute for the COVID-19 Genomics UK Consortium |
| EPI_ISL_459095 | NHSGGC West of Scotland Specialist Virology Centre / MRC-University of Glasgow Centre for Virus Research | Wellcome Sanger Institute for the COVID-19 Genomics UK Consortium |
| EPI_ISL_459096 | NHSGGC West of Scotland Specialist Virology Centre / MRC-University of Glasgow Centre for Virus Research | Wellcome Sanger Institute for the COVID-19 Genomics UK Consortium |
| EPI_ISL_459097 | NHSGGC West of Scotland Specialist Virology Centre / MRC-University of Glasgow Centre for Virus Research | Wellcome Sanger Institute for the COVID-19 Genomics UK Consortium |

| EPI_Accession_ID | Originating Laboratory | Submitting Laboratory |
|---|---|---|
| EPI_ISL_459098 | NHSGGC West of Scotland Specialist Virology Centre / MRC-University of Glasgow Centre for Virus Research | Wellcome Sanger Institute for the COVID-19 Genomics UK Consortium |
| EPI_ISL_459099 | NHSGGC West of Scotland Specialist Virology Centre / MRC-University of Glasgow Centre for Virus Research | Wellcome Sanger Institute for the COVID-19 Genomics UK Consortium |
| EPI_ISL_459100 | NHSGGC West of Scotland Specialist Virology Centre / MRC-University of Glasgow Centre for Virus Research | Wellcome Sanger Institute for the COVID-19 Genomics UK Consortium |
| EPI_ISL_459101 | NHSGGC West of Scotland Specialist Virology Centre / MRC-University of Glasgow Centre for Virus Research | Wellcome Sanger Institute for the COVID-19 Genomics UK Consortium |
| EPI_ISL_459102 | NHSGGC West of Scotland Specialist Virology Centre / MRC-University of Glasgow Centre for Virus Research | Wellcome Sanger Institute for the COVID-19 Genomics UK Consortium |
| EPI_ISL_459103 | NHSGGC West of Scotland Specialist Virology Centre / MRC-University of Glasgow Centre for Virus Research | Wellcome Sanger Institute for the COVID-19 Genomics UK Consortium |
| EPI_ISL_459104 | NHSGGC West of Scotland Specialist Virology Centre / MRC-University of Glasgow Centre for Virus Research | Wellcome Sanger Institute for the COVID-19 Genomics UK Consortium |
| EPI_ISL_459105 | NHSGGC West of Scotland Specialist Virology Centre / MRC-University of Glasgow Centre for Virus Research | Wellcome Sanger Institute for the COVID-19 Genomics UK Consortium |
| EPI_ISL_459106 | NHSGGC West of Scotland Specialist Virology Centre / MRC-University of Glasgow Centre for Virus Research | Wellcome Sanger Institute for the COVID-19 Genomics UK Consortium |
| EPI_ISL_459107 | NHSGGC West of Scotland Specialist Virology Centre / MRC-University of Glasgow Centre for Virus Research | Wellcome Sanger Institute for the COVID-19 Genomics UK Consortium |
| EPI_ISL_459108 | NHSGGC West of Scotland Specialist Virology Centre / MRC-University of Glasgow Centre for Virus Research | Wellcome Sanger Institute for the COVID-19 Genomics UK Consortium |
| EPI_ISL_459109 | NHSGGC West of Scotland Specialist Virology Centre / MRC-University of Glasgow Centre for Virus Research | Wellcome Sanger Institute for the COVID-19 Genomics UK Consortium |
| EPI_ISL_459110 | NHSGGC West of Scotland Specialist Virology Centre / MRC-University of Glasgow Centre for Virus Research | Wellcome Sanger Institute for the COVID-19 Genomics UK Consortium |
| EPI_ISL_459111 | NHSGGC West of Scotland Specialist Virology Centre / MRC-University of Glasgow Centre for Virus Research | Wellcome Sanger Institute for the COVID-19 Genomics UK Consortium |
| EPI_ISL_459112 | NHSGGC West of Scotland Specialist Virology Centre / MRC-University of Glasgow Centre for Virus Research | Wellcome Sanger Institute for the COVID-19 Genomics UK Consortium |
| EPI_ISL_459113 | NHSGGC West of Scotland Specialist Virology Centre / MRC-University of Glasgow Centre for Virus Research | Wellcome Sanger Institute for the COVID-19 Genomics UK Consortium |

| EPI_Accession_ID | Originating Laboratory | Submitting Laboratory |
|---|---|---|
| EPI_ISL_459114 | NHSGGC West of Scotland Specialist Virology Centre / MRC-University of Glasgow Centre for Virus Research | Wellcome Sanger Institute for the COVID-19 Genomics UK Consortium |
| EPI_ISL_459115 | NHSGGC West of Scotland Specialist Virology Centre / MRC-University of Glasgow Centre for Virus Research | Wellcome Sanger Institute for the COVID-19 Genomics UK Consortium |
| EPI_ISL_459116 | NHSGGC West of Scotland Specialist Virology Centre / MRC-University of Glasgow Centre for Virus Research | Wellcome Sanger Institute for the COVID-19 Genomics UK Consortium |
| EPI_ISL_459117 | NHSGGC West of Scotland Specialist Virology Centre / MRC-University of Glasgow Centre for Virus Research | Wellcome Sanger Institute for the COVID-19 Genomics UK Consortium |
| EPI_ISL_459118 | NHSGGC West of Scotland Specialist Virology Centre / MRC-University of Glasgow Centre for Virus Research | Wellcome Sanger Institute for the COVID-19 Genomics UK Consortium |
| EPI_ISL_459119 | NHSGGC West of Scotland Specialist Virology Centre / MRC-University of Glasgow Centre for Virus Research | Wellcome Sanger Institute for the COVID-19 Genomics UK Consortium |
| EPI_ISL_459120 | NHSGGC West of Scotland Specialist Virology Centre / MRC-University of Glasgow Centre for Virus Research | Wellcome Sanger Institute for the COVID-19 Genomics UK Consortium |
| EPI_ISL_459121 | NHSGGC West of Scotland Specialist Virology Centre / MRC-University of Glasgow Centre for Virus Research | Wellcome Sanger Institute for the COVID-19 Genomics UK Consortium |
| EPI_ISL_459122 | NHSGGC West of Scotland Specialist Virology Centre / MRC-University of Glasgow Centre for Virus Research | Wellcome Sanger Institute for the COVID-19 Genomics UK Consortium |
| EPI_ISL_459123 | NHSGGC West of Scotland Specialist Virology Centre / MRC-University of Glasgow Centre for Virus Research | Wellcome Sanger Institute for the COVID-19 Genomics UK Consortium |
| EPI_ISL_459124 | NHSGGC West of Scotland Specialist Virology Centre / MRC-University of Glasgow Centre for Virus Research | Wellcome Sanger Institute for the COVID-19 Genomics UK Consortium |
| EPI_ISL_459125 | NHSGGC West of Scotland Specialist Virology Centre / MRC-University of Glasgow Centre for Virus Research | Wellcome Sanger Institute for the COVID-19 Genomics UK Consortium |
| EPI_ISL_459126 | NHSGGC West of Scotland Specialist Virology Centre / MRC-University of Glasgow Centre for Virus Research | Wellcome Sanger Institute for the COVID-19 Genomics UK Consortium |
| EPI_ISL_459127 | NHSGGC West of Scotland Specialist Virology Centre / MRC-University of Glasgow Centre for Virus Research | Wellcome Sanger Institute for the COVID-19 Genomics UK Consortium |
| EPI_ISL_459128 | NHSGGC West of Scotland Specialist Virology Centre / MRC-University of Glasgow Centre for Virus Research | Wellcome Sanger Institute for the COVID-19 Genomics UK Consortium |
| EPI_ISL_459129 | NHSGGC West of Scotland Specialist Virology Centre / MRC-University of Glasgow Centre for Virus Research | Wellcome Sanger Institute for the COVID-19 Genomics UK Consortium |

| EPI_Accession_ID | Originating Laboratory | Submitting Laboratory |
|---|---|---|
| EPI_ISL_459130 | NHSGGC West of Scotland Specialist Virology Centre / MRC-University of Glasgow Centre for Virus Research | Wellcome Sanger Institute for the COVID-19 Genomics UK Consortium |
| EPI_ISL_459131 | NHSGGC West of Scotland Specialist Virology Centre / MRC-University of Glasgow Centre for Virus Research | Wellcome Sanger Institute for the COVID-19 Genomics UK Consortium |
| EPI_ISL_459132 | NHSGGC West of Scotland Specialist Virology Centre / MRC-University of Glasgow Centre for Virus Research | Wellcome Sanger Institute for the COVID-19 Genomics UK Consortium |
| EPI_ISL_459133 | NHSGGC West of Scotland Specialist Virology Centre / MRC-University of Glasgow Centre for Virus Research | Wellcome Sanger Institute for the COVID-19 Genomics UK Consortium |
| EPI_ISL_459134 | NHSGGC West of Scotland Specialist Virology Centre / MRC-University of Glasgow Centre for Virus Research | Wellcome Sanger Institute for the COVID-19 Genomics UK Consortium |
| EPI_ISL_459135 | NHSGGC West of Scotland Specialist Virology Centre / MRC-University of Glasgow Centre for Virus Research | Wellcome Sanger Institute for the COVID-19 Genomics UK Consortium |
| EPI_ISL_459136 | NHSGGC West of Scotland Specialist Virology Centre / MRC-University of Glasgow Centre for Virus Research | Wellcome Sanger Institute for the COVID-19 Genomics UK Consortium |
| EPI_ISL_459137 | NHSGGC West of Scotland Specialist Virology Centre / MRC-University of Glasgow Centre for Virus Research | Wellcome Sanger Institute for the COVID-19 Genomics UK Consortium |
| EPI_ISL_459138 | NHSGGC West of Scotland Specialist Virology Centre / MRC-University of Glasgow Centre for Virus Research | Wellcome Sanger Institute for the COVID-19 Genomics UK Consortium |
| EPI_ISL_459139 | NHSGGC West of Scotland Specialist Virology Centre / MRC-University of Glasgow Centre for Virus Research | Wellcome Sanger Institute for the COVID-19 Genomics UK Consortium |
| EPI_ISL_459140 | NHSGGC West of Scotland Specialist Virology Centre / MRC-University of Glasgow Centre for Virus Research | Wellcome Sanger Institute for the COVID-19 Genomics UK Consortium |
| EPI_ISL_459141 | NHSGGC West of Scotland Specialist Virology Centre / MRC-University of Glasgow Centre for Virus Research | Wellcome Sanger Institute for the COVID-19 Genomics UK Consortium |
| EPI_ISL_459142 | NHSGGC West of Scotland Specialist Virology Centre / MRC-University of Glasgow Centre for Virus Research | Wellcome Sanger Institute for the COVID-19 Genomics UK Consortium |
| EPI_ISL_459143 | NHSGGC West of Scotland Specialist Virology Centre / MRC-University of Glasgow Centre for Virus Research | Wellcome Sanger Institute for the COVID-19 Genomics UK Consortium |
| EPI_ISL_459144 | NHSGGC West of Scotland Specialist Virology Centre / MRC-University of Glasgow Centre for Virus Research | Wellcome Sanger Institute for the COVID-19 Genomics UK Consortium |
| EPI_ISL_459145 | NHSGGC West of Scotland Specialist Virology Centre / MRC-University of Glasgow Centre for Virus Research | Wellcome Sanger Institute for the COVID-19 Genomics UK Consortium |

| EPI_Accession_ID | Originating Laboratory | Submitting Laboratory |
|---|---|---|
| EPI_ISL_459146 | NHSGGC West of Scotland Specialist Virology Centre / MRC-University of Glasgow Centre for Virus Research | Wellcome Sanger Institute for the COVID-19 Genomics UK Consortium |
| EPI_ISL_459147 | NHSGGC West of Scotland Specialist Virology Centre / MRC-University of Glasgow Centre for Virus Research | Wellcome Sanger Institute for the COVID-19 Genomics UK Consortium |
| EPI_ISL_459148 | NHSGGC West of Scotland Specialist Virology Centre / MRC-University of Glasgow Centre for Virus Research | Wellcome Sanger Institute for the COVID-19 Genomics UK Consortium |
| EPI_ISL_459149 | NHSGGC West of Scotland Specialist Virology Centre / MRC-University of Glasgow Centre for Virus Research | Wellcome Sanger Institute for the COVID-19 Genomics UK Consortium |
| EPI_ISL_459150 | NHSGGC West of Scotland Specialist Virology Centre / MRC-University of Glasgow Centre for Virus Research | Wellcome Sanger Institute for the COVID-19 Genomics UK Consortium |
| EPI_ISL_459151 | NHSGGC West of Scotland Specialist Virology Centre / MRC-University of Glasgow Centre for Virus Research | Wellcome Sanger Institute for the COVID-19 Genomics UK Consortium |
| EPI_ISL_459152 | NHSGGC West of Scotland Specialist Virology Centre / MRC-University of Glasgow Centre for Virus Research | Wellcome Sanger Institute for the COVID-19 Genomics UK Consortium |
| EPI_ISL_459153 | NHSGGC West of Scotland Specialist Virology Centre / MRC-University of Glasgow Centre for Virus Research | Wellcome Sanger Institute for the COVID-19 Genomics UK Consortium |
| EPI_ISL_459154 | NHSGGC West of Scotland Specialist Virology Centre / MRC-University of Glasgow Centre for Virus Research | Wellcome Sanger Institute for the COVID-19 Genomics UK Consortium |
| EPI_ISL_459155 | NHSGGC West of Scotland Specialist Virology Centre / MRC-University of Glasgow Centre for Virus Research | Wellcome Sanger Institute for the COVID-19 Genomics UK Consortium |
| EPI_ISL_459156 | NHSGGC West of Scotland Specialist Virology Centre / MRC-University of Glasgow Centre for Virus Research | Wellcome Sanger Institute for the COVID-19 Genomics UK Consortium |
| EPI_ISL_459157 | NHSGGC West of Scotland Specialist Virology Centre / MRC-University of Glasgow Centre for Virus Research | Wellcome Sanger Institute for the COVID-19 Genomics UK Consortium |
| EPI_ISL_459158 | NHSGGC West of Scotland Specialist Virology Centre / MRC-University of Glasgow Centre for Virus Research | Wellcome Sanger Institute for the COVID-19 Genomics UK Consortium |
| EPI_ISL_459159 | NHSGGC West of Scotland Specialist Virology Centre / MRC-University of Glasgow Centre for Virus Research | Wellcome Sanger Institute for the COVID-19 Genomics UK Consortium |
| EPI_ISL_459160 | NHSGGC West of Scotland Specialist Virology Centre / MRC-University of Glasgow Centre for Virus Research | Wellcome Sanger Institute for the COVID-19 Genomics UK Consortium |
| EPI_ISL_459161 | NHSGGC West of Scotland Specialist Virology Centre / MRC-University of Glasgow Centre for Virus Research | Wellcome Sanger Institute for the COVID-19 Genomics UK Consortium |

| EPI_Accession_ID | Originating Laboratory | Submitting Laboratory |
|---|---|---|
| EPI_ISL_459162 | NHSGGC West of Scotland Specialist Virology Centre / MRC-University of Glasgow Centre for Virus Research | Wellcome Sanger Institute for the COVID-19 Genomics UK Consortium |
| EPI_ISL_459163 | NHSGGC West of Scotland Specialist Virology Centre / MRC-University of Glasgow Centre for Virus Research | Wellcome Sanger Institute for the COVID-19 Genomics UK Consortium |
| EPI_ISL_459164 | NHSGGC West of Scotland Specialist Virology Centre / MRC-University of Glasgow Centre for Virus Research | Wellcome Sanger Institute for the COVID-19 Genomics UK Consortium |
| EPI_ISL_459165 | NHSGGC West of Scotland Specialist Virology Centre / MRC-University of Glasgow Centre for Virus Research | Wellcome Sanger Institute for the COVID-19 Genomics UK Consortium |
| EPI_ISL_459166 | Department of Pathology, University of Cambridge | Wellcome Sanger Institute for the COVID-19 Genomics UK Consortium |
| EPI_ISL_459167 | Department of Pathology, University of Cambridge | Wellcome Sanger Institute for the COVID-19 Genomics UK Consortium |
| EPI_ISL_459168 | Department of Pathology, University of Cambridge | Wellcome Sanger Institute for the COVID-19 Genomics UK Consortium |
| EPI_ISL_459169 | Department of Pathology, University of Cambridge | Wellcome Sanger Institute for the COVID-19 Genomics UK Consortium |
| EPI_ISL_459170 | Department of Pathology, University of Cambridge | Wellcome Sanger Institute for the COVID-19 Genomics UK Consortium |
| EPI_ISL_459171 | Department of Pathology, University of Cambridge | Wellcome Sanger Institute for the COVID-19 Genomics UK Consortium |
| EPI_ISL_459172 | Department of Pathology, University of Cambridge | Wellcome Sanger Institute for the COVID-19 Genomics UK Consortium |
| EPI_ISL_459173 | Department of Pathology, University of Cambridge | Wellcome Sanger Institute for the COVID-19 Genomics UK Consortium |
| EPI_ISL_459174 | Department of Pathology, University of Cambridge | Wellcome Sanger Institute for the COVID-19 Genomics UK Consortium |
| EPI_ISL_459175 | Department of Pathology, University of Cambridge | Wellcome Sanger Institute for the COVID-19 Genomics UK Consortium |
| EPI_ISL_459176 | Department of Pathology, University of Cambridge | Wellcome Sanger Institute for the COVID-19 Genomics UK Consortium |
| EPI_ISL_459177 | Department of Pathology, University of Cambridge | Wellcome Sanger Institute for the COVID-19 Genomics UK Consortium |

| EPI_Accession_ID | Originating Laboratory | Submitting Laboratory |
|---|---|---|
| EPI_ISL_459178 | Department of Pathology, University of Cambridge | Wellcome Sanger Institute for the COVID-19 Genomics UK Consortium |
| EPI_ISL_459179 | Department of Pathology, University of Cambridge | Wellcome Sanger Institute for the COVID-19 Genomics UK Consortium |
| EPI_ISL_459180 | Department of Pathology, University of Cambridge | Wellcome Sanger Institute for the COVID-19 Genomics UK Consortium |
| EPI_ISL_459181 | Department of Pathology, University of Cambridge | Wellcome Sanger Institute for the COVID-19 Genomics UK Consortium |
| EPI_ISL_459182 | Department of Pathology, University of Cambridge | Wellcome Sanger Institute for the COVID-19 Genomics UK Consortium |
| EPI_ISL_459183 | Department of Pathology, University of Cambridge | Wellcome Sanger Institute for the COVID-19 Genomics UK Consortium |
| EPI_ISL_459184 | Department of Pathology, University of Cambridge | Wellcome Sanger Institute for the COVID-19 Genomics UK Consortium |
| EPI_ISL_459185 | Department of Pathology, University of Cambridge | Wellcome Sanger Institute for the COVID-19 Genomics UK Consortium |
| EPI_ISL_459186 | Department of Pathology, University of Cambridge | Wellcome Sanger Institute for the COVID-19 Genomics UK Consortium |
| EPI_ISL_459187 | Department of Pathology, University of Cambridge | Wellcome Sanger Institute for the COVID-19 Genomics UK Consortium |
| EPI_ISL_459188 | Department of Pathology, University of Cambridge | Wellcome Sanger Institute for the COVID-19 Genomics UK Consortium |
| EPI_ISL_459189 | Department of Pathology, University of Cambridge | Wellcome Sanger Institute for the COVID-19 Genomics UK Consortium |
| EPI_ISL_459190 | Department of Pathology, University of Cambridge | Wellcome Sanger Institute for the COVID-19 Genomics UK Consortium |
| EPI_ISL_459191 | Department of Pathology, University of Cambridge | Wellcome Sanger Institute for the COVID-19 Genomics UK Consortium |
| EPI_ISL_459192 | Department of Pathology, University of Cambridge | Wellcome Sanger Institute for the COVID-19 Genomics UK Consortium |
| EPI_ISL_459193 | Department of Pathology, University of Cambridge | Wellcome Sanger Institute for the COVID-19 Genomics UK Consortium |

| EPI_Accession_ID | Originating Laboratory | Submitting Laboratory |
|---|---|---|
| EPI_ISL_459194 | Department of Pathology, University of Cambridge | Wellcome Sanger Institute for the COVID-19 Genomics UK Consortium |
| EPI_ISL_459195 | Department of Pathology, University of Cambridge | Wellcome Sanger Institute for the COVID-19 Genomics UK Consortium |
| EPI_ISL_459196 | Department of Pathology, University of Cambridge | Wellcome Sanger Institute for the COVID-19 Genomics UK Consortium |
| EPI_ISL_459197 | Department of Pathology, University of Cambridge | Wellcome Sanger Institute for the COVID-19 Genomics UK Consortium |
| EPI_ISL_459198 | Department of Pathology, University of Cambridge | Wellcome Sanger Institute for the COVID-19 Genomics UK Consortium |
| EPI_ISL_459199 | Department of Pathology, University of Cambridge | Wellcome Sanger Institute for the COVID-19 Genomics UK Consortium |
| EPI_ISL_459200 | Department of Pathology, University of Cambridge | Wellcome Sanger Institute for the COVID-19 Genomics UK Consortium |
| EPI_ISL_459201 | Department of Pathology, University of Cambridge | Wellcome Sanger Institute for the COVID-19 Genomics UK Consortium |
| EPI_ISL_459202 | Department of Pathology, University of Cambridge | Wellcome Sanger Institute for the COVID-19 Genomics UK Consortium |
| EPI_ISL_459203 | Department of Pathology, University of Cambridge | Wellcome Sanger Institute for the COVID-19 Genomics UK Consortium |
| EPI_ISL_459204 | Department of Pathology, University of Cambridge | Wellcome Sanger Institute for the COVID-19 Genomics UK Consortium |
| EPI_ISL_459205 | Department of Pathology, University of Cambridge | Wellcome Sanger Institute for the COVID-19 Genomics UK Consortium |
| EPI_ISL_459206 | Department of Pathology, University of Cambridge | Wellcome Sanger Institute for the COVID-19 Genomics UK Consortium |
| EPI_ISL_459207 | Department of Pathology, University of Cambridge | Wellcome Sanger Institute for the COVID-19 Genomics UK Consortium |
| EPI_ISL_459208 | Department of Pathology, University of Cambridge | Wellcome Sanger Institute for the COVID-19 Genomics UK Consortium |
| EPI_ISL_459209 | Department of Pathology, University of Cambridge | Wellcome Sanger Institute for the COVID-19 Genomics UK Consortium |

| EPI_Accession_ID | Originating Laboratory | Submitting Laboratory |
|---|---|---|
| EPI_ISL_459210 | Department of Pathology, University of Cambridge | Wellcome Sanger Institute for the COVID-19 Genomics UK Consortium |
| EPI_ISL_459211 | Department of Pathology, University of Cambridge | Wellcome Sanger Institute for the COVID-19 Genomics UK Consortium |
| EPI_ISL_459212 | Department of Pathology, University of Cambridge | Wellcome Sanger Institute for the COVID-19 Genomics UK Consortium |
| EPI_ISL_459213 | Department of Pathology, University of Cambridge | Wellcome Sanger Institute for the COVID-19 Genomics UK Consortium |
| EPI_ISL_459214 | Department of Pathology, University of Cambridge | Wellcome Sanger Institute for the COVID-19 Genomics UK Consortium |
| EPI_ISL_459215 | Department of Pathology, University of Cambridge | Wellcome Sanger Institute for the COVID-19 Genomics UK Consortium |
| EPI_ISL_459216 | Department of Pathology, University of Cambridge | Wellcome Sanger Institute for the COVID-19 Genomics UK Consortium |
| EPI_ISL_459217 | Department of Pathology, University of Cambridge | Wellcome Sanger Institute for the COVID-19 Genomics UK Consortium |
| EPI_ISL_459218 | Department of Pathology, University of Cambridge | Wellcome Sanger Institute for the COVID-19 Genomics UK Consortium |
| EPI_ISL_459219 | Department of Pathology, University of Cambridge | Wellcome Sanger Institute for the COVID-19 Genomics UK Consortium |
| EPI_ISL_459220 | Department of Pathology, University of Cambridge | Wellcome Sanger Institute for the COVID-19 Genomics UK Consortium |
| EPI_ISL_459221 | Department of Pathology, University of Cambridge | Wellcome Sanger Institute for the COVID-19 Genomics UK Consortium |
| EPI_ISL_459222 | Department of Pathology, University of Cambridge | Wellcome Sanger Institute for the COVID-19 Genomics UK Consortium |
| EPI_ISL_459223 | Department of Pathology, University of Cambridge | Wellcome Sanger Institute for the COVID-19 Genomics UK Consortium |
| EPI_ISL_459224 | Department of Pathology, University of Cambridge | Wellcome Sanger Institute for the COVID-19 Genomics UK Consortium |
| EPI_ISL_459225 | Department of Pathology, University of Cambridge | Wellcome Sanger Institute for the COVID-19 Genomics UK Consortium |

| EPI_Accession_ID | Originating Laboratory | Submitting Laboratory |
|---|---|---|
| EPI_ISL_459226 | Department of Pathology, University of Cambridge | Wellcome Sanger Institute for the COVID-19 Genomics UK Consortium |
| EPI_ISL_459227 | Department of Pathology, University of Cambridge | Wellcome Sanger Institute for the COVID-19 Genomics UK Consortium |
| EPI_ISL_459228 | Department of Pathology, University of Cambridge | Wellcome Sanger Institute for the COVID-19 Genomics UK Consortium |
| EPI_ISL_459229 | Department of Pathology, University of Cambridge | Wellcome Sanger Institute for the COVID-19 Genomics UK Consortium |
| EPI_ISL_459230 | Department of Pathology, University of Cambridge | Wellcome Sanger Institute for the COVID-19 Genomics UK Consortium |
| EPI_ISL_459231 | Department of Pathology, University of Cambridge | Wellcome Sanger Institute for the COVID-19 Genomics UK Consortium |
| EPI_ISL_459232 | Department of Pathology, University of Cambridge | Wellcome Sanger Institute for the COVID-19 Genomics UK Consortium |
| EPI_ISL_459233 | Department of Pathology, University of Cambridge | Wellcome Sanger Institute for the COVID-19 Genomics UK Consortium |
| EPI_ISL_459234 | Department of Pathology, University of Cambridge | Wellcome Sanger Institute for the COVID-19 Genomics UK Consortium |
| EPI_ISL_459235 | Department of Pathology, University of Cambridge | Wellcome Sanger Institute for the COVID-19 Genomics UK Consortium |
| EPI_ISL_459236 | Department of Pathology, University of Cambridge | Wellcome Sanger Institute for the COVID-19 Genomics UK Consortium |
| EPI_ISL_459237 | Department of Pathology, University of Cambridge | Wellcome Sanger Institute for the COVID-19 Genomics UK Consortium |
| EPI_ISL_459238 | Department of Pathology, University of Cambridge | Wellcome Sanger Institute for the COVID-19 Genomics UK Consortium |
| EPI_ISL_459239 | Department of Pathology, University of Cambridge | Wellcome Sanger Institute for the COVID-19 Genomics UK Consortium |
| EPI_ISL_459240 | Department of Pathology, University of Cambridge | Wellcome Sanger Institute for the COVID-19 Genomics UK Consortium |
| EPI_ISL_459241 | Department of Pathology, University of Cambridge | Wellcome Sanger Institute for the COVID-19 Genomics UK Consortium |

| EPI_Accession_ID | Originating Laboratory | Submitting Laboratory |
|---|---|---|
| EPI_ISL_459242 | Department of Pathology, University of Cambridge | Wellcome Sanger Institute for the COVID-19 Genomics UK Consortium |
| EPI_ISL_459243 | Department of Pathology, University of Cambridge | Wellcome Sanger Institute for the COVID-19 Genomics UK Consortium |
| EPI_ISL_459244 | Department of Pathology, University of Cambridge | Wellcome Sanger Institute for the COVID-19 Genomics UK Consortium |
| EPI_ISL_459245 | Department of Pathology, University of Cambridge | Wellcome Sanger Institute for the COVID-19 Genomics UK Consortium |
| EPI_ISL_459246 | Department of Pathology, University of Cambridge | Wellcome Sanger Institute for the COVID-19 Genomics UK Consortium |
| EPI_ISL_459247 | Department of Pathology, University of Cambridge | Wellcome Sanger Institute for the COVID-19 Genomics UK Consortium |
| EPI_ISL_459248 | Department of Pathology, University of Cambridge | Wellcome Sanger Institute for the COVID-19 Genomics UK Consortium |
| EPI_ISL_459249 | Department of Pathology, University of Cambridge | Wellcome Sanger Institute for the COVID-19 Genomics UK Consortium |
| EPI_ISL_459250 | Department of Pathology, University of Cambridge | Wellcome Sanger Institute for the COVID-19 Genomics UK Consortium |
| EPI_ISL_459251 | Department of Pathology, University of Cambridge | Wellcome Sanger Institute for the COVID-19 Genomics UK Consortium |
| EPI_ISL_459252 | Department of Pathology, University of Cambridge | Wellcome Sanger Institute for the COVID-19 Genomics UK Consortium |
| EPI_ISL_459253 | Department of Pathology, University of Cambridge | Wellcome Sanger Institute for the COVID-19 Genomics UK Consortium |
| EPI_ISL_459254 | Department of Pathology, University of Cambridge | Wellcome Sanger Institute for the COVID-19 Genomics UK Consortium |
| EPI_ISL_459255 | Department of Pathology, University of Cambridge | Wellcome Sanger Institute for the COVID-19 Genomics UK Consortium |
| EPI_ISL_459256 | Department of Pathology, University of Cambridge | Wellcome Sanger Institute for the COVID-19 Genomics UK Consortium |
| EPI_ISL_459257 | Department of Pathology, University of Cambridge | Wellcome Sanger Institute for the COVID-19 Genomics UK Consortium |

| EPI_Accession_ID | Originating Laboratory | Submitting Laboratory |
|---|---|---|
| EPI_ISL_459258 | Department of Pathology, University of Cambridge | Wellcome Sanger Institute for the COVID-19 Genomics UK Consortium |
| EPI_ISL_459259 | Department of Pathology, University of Cambridge | Wellcome Sanger Institute for the COVID-19 Genomics UK Consortium |
| EPI_ISL_459260 | Department of Pathology, University of Cambridge | Wellcome Sanger Institute for the COVID-19 Genomics UK Consortium |
| EPI_ISL_459261 | Department of Pathology, University of Cambridge | Wellcome Sanger Institute for the COVID-19 Genomics UK Consortium |
| EPI_ISL_459262 | Department of Pathology, University of Cambridge | Wellcome Sanger Institute for the COVID-19 Genomics UK Consortium |
| EPI_ISL_459263 | Department of Pathology, University of Cambridge | Wellcome Sanger Institute for the COVID-19 Genomics UK Consortium |
| EPI_ISL_459264 | Department of Pathology, University of Cambridge | Wellcome Sanger Institute for the COVID-19 Genomics UK Consortium |
| EPI_ISL_459265 | Department of Pathology, University of Cambridge | Wellcome Sanger Institute for the COVID-19 Genomics UK Consortium |
| EPI_ISL_459266 | Department of Pathology, University of Cambridge | Wellcome Sanger Institute for the COVID-19 Genomics UK Consortium |
| EPI_ISL_459267 | Department of Pathology, University of Cambridge | Wellcome Sanger Institute for the COVID-19 Genomics UK Consortium |
| EPI_ISL_459268 | Department of Pathology, University of Cambridge | Wellcome Sanger Institute for the COVID-19 Genomics UK Consortium |
| EPI_ISL_459269 | Department of Pathology, University of Cambridge | Wellcome Sanger Institute for the COVID-19 Genomics UK Consortium |
| EPI_ISL_459270 | Department of Pathology, University of Cambridge | Wellcome Sanger Institute for the COVID-19 Genomics UK Consortium |
| EPI_ISL_459271 | Department of Pathology, University of Cambridge | Wellcome Sanger Institute for the COVID-19 Genomics UK Consortium |
| EPI_ISL_459272 | Department of Pathology, University of Cambridge | Wellcome Sanger Institute for the COVID-19 Genomics UK Consortium |
| EPI_ISL_459273 | Department of Pathology, University of Cambridge | Wellcome Sanger Institute for the COVID-19 Genomics UK Consortium |

| EPI_Accession_ID | Originating Laboratory | Submitting Laboratory |
|---|---|---|
| EPI_ISL_459274 | Department of Pathology, University of Cambridge | Wellcome Sanger Institute for the COVID-19 Genomics UK Consortium |
| EPI_ISL_459275 | Department of Pathology, University of Cambridge | Wellcome Sanger Institute for the COVID-19 Genomics UK Consortium |
| EPI_ISL_459276 | Department of Pathology, University of Cambridge | Wellcome Sanger Institute for the COVID-19 Genomics UK Consortium |
| EPI_ISL_459277 | Department of Pathology, University of Cambridge | Wellcome Sanger Institute for the COVID-19 Genomics UK Consortium |
| EPI_ISL_459278 | Department of Pathology, University of Cambridge | Wellcome Sanger Institute for the COVID-19 Genomics UK Consortium |
| EPI_ISL_459279 | Department of Pathology, University of Cambridge | Wellcome Sanger Institute for the COVID-19 Genomics UK Consortium |
| EPI_ISL_459280 | Department of Pathology, University of Cambridge | Wellcome Sanger Institute for the COVID-19 Genomics UK Consortium |
| EPI_ISL_459281 | Department of Pathology, University of Cambridge | Wellcome Sanger Institute for the COVID-19 Genomics UK Consortium |
| EPI_ISL_459282 | Department of Pathology, University of Cambridge | Wellcome Sanger Institute for the COVID-19 Genomics UK Consortium |
| EPI_ISL_459283 | Department of Pathology, University of Cambridge | Wellcome Sanger Institute for the COVID-19 Genomics UK Consortium |
| EPI_ISL_459284 | Department of Pathology, University of Cambridge | Wellcome Sanger Institute for the COVID-19 Genomics UK Consortium |
| EPI_ISL_459285 | Department of Pathology, University of Cambridge | Wellcome Sanger Institute for the COVID-19 Genomics UK Consortium |
| EPI_ISL_459286 | Department of Pathology, University of Cambridge | Wellcome Sanger Institute for the COVID-19 Genomics UK Consortium |
| EPI_ISL_459287 | Department of Pathology, University of Cambridge | Wellcome Sanger Institute for the COVID-19 Genomics UK Consortium |
| EPI_ISL_459288 | Department of Pathology, University of Cambridge | Wellcome Sanger Institute for the COVID-19 Genomics UK Consortium |
| EPI_ISL_459289 | Department of Pathology, University of Cambridge | Wellcome Sanger Institute for the COVID-19 Genomics UK Consortium |

| EPI_Accession_ID | Originating Laboratory | Submitting Laboratory |
|---|---|---|
| EPI_ISL_459290 | Department of Pathology, University of Cambridge | Wellcome Sanger Institute for the COVID-19 Genomics UK Consortium |
| EPI_ISL_459291 | Department of Pathology, University of Cambridge | Wellcome Sanger Institute for the COVID-19 Genomics UK Consortium |
| EPI_ISL_459292 | Department of Pathology, University of Cambridge | Wellcome Sanger Institute for the COVID-19 Genomics UK Consortium |
| EPI_ISL_459293 | Department of Pathology, University of Cambridge | Wellcome Sanger Institute for the COVID-19 Genomics UK Consortium |
| EPI_ISL_459294 | Department of Pathology, University of Cambridge | Wellcome Sanger Institute for the COVID-19 Genomics UK Consortium |
| EPI_ISL_459295 | Department of Pathology, University of Cambridge | Wellcome Sanger Institute for the COVID-19 Genomics UK Consortium |
| EPI_ISL_459296 | Department of Pathology, University of Cambridge | Wellcome Sanger Institute for the COVID-19 Genomics UK Consortium |
| EPI_ISL_459297 | Department of Pathology, University of Cambridge | Wellcome Sanger Institute for the COVID-19 Genomics UK Consortium |
| EPI_ISL_459304 | Department of Pathology, University of Cambridge | Wellcome Sanger Institute for the COVID-19 Genomics UK Consortium |
| EPI_ISL_459305 | Department of Pathology, University of Cambridge | Wellcome Sanger Institute for the COVID-19 Genomics UK Consortium |
| EPI_ISL_459306 | Department of Pathology, University of Cambridge | Wellcome Sanger Institute for the COVID-19 Genomics UK Consortium |
| EPI_ISL_459307 | Department of Pathology, University of Cambridge | Wellcome Sanger Institute for the COVID-19 Genomics UK Consortium |
| EPI_ISL_459308 | Department of Pathology, University of Cambridge | Wellcome Sanger Institute for the COVID-19 Genomics UK Consortium |
| EPI_ISL_459309 | Department of Pathology, University of Cambridge | Wellcome Sanger Institute for the COVID-19 Genomics UK Consortium |
| EPI_ISL_459310 | Department of Pathology, University of Cambridge | Wellcome Sanger Institute for the COVID-19 Genomics UK Consortium |
| EPI_ISL_459311 | Department of Pathology, University of Cambridge | Wellcome Sanger Institute for the COVID-19 Genomics UK Consortium |

| EPI_Accession_ID | Originating Laboratory | Submitting Laboratory |
|---|---|---|
| EPI_ISL_459312 | Department of Pathology, University of Cambridge | Wellcome Sanger Institute for the COVID-19 Genomics UK Consortium |
| EPI_ISL_459313 | Department of Pathology, University of Cambridge | Wellcome Sanger Institute for the COVID-19 Genomics UK Consortium |
| EPI_ISL_459314 | Department of Pathology, University of Cambridge | Wellcome Sanger Institute for the COVID-19 Genomics UK Consortium |
| EPI_ISL_459315 | Department of Pathology, University of Cambridge | Wellcome Sanger Institute for the COVID-19 Genomics UK Consortium |
| EPI_ISL_459316 | Department of Pathology, University of Cambridge | Wellcome Sanger Institute for the COVID-19 Genomics UK Consortium |
| EPI_ISL_459317 | Department of Pathology, University of Cambridge | Wellcome Sanger Institute for the COVID-19 Genomics UK Consortium |
| EPI_ISL_459318 | Department of Pathology, University of Cambridge | Wellcome Sanger Institute for the COVID-19 Genomics UK Consortium |
| EPI_ISL_459319 | Department of Pathology, University of Cambridge | Wellcome Sanger Institute for the COVID-19 Genomics UK Consortium |
| EPI_ISL_459320 | Department of Pathology, University of Cambridge | Wellcome Sanger Institute for the COVID-19 Genomics UK Consortium |
| EPI_ISL_459321 | Department of Pathology, University of Cambridge | Wellcome Sanger Institute for the COVID-19 Genomics UK Consortium |
| EPI_ISL_459322 | Department of Pathology, University of Cambridge | Wellcome Sanger Institute for the COVID-19 Genomics UK Consortium |
| EPI_ISL_459323 | Department of Pathology, University of Cambridge | Wellcome Sanger Institute for the COVID-19 Genomics UK Consortium |
| EPI_ISL_459324 | Regional Virus Laboratory, Belfast Health and Social Care Trust | Wellcome Sanger Institute for the COVID-19 Genomics UK Consortium |
| EPI_ISL_459325 | Regional Virus Laboratory, Belfast Health and Social Care Trust | Wellcome Sanger Institute for the COVID-19 Genomics UK Consortium |
| EPI_ISL_459326 | Regional Virus Laboratory, Belfast Health and Social Care Trust | Wellcome Sanger Institute for the COVID-19 Genomics UK Consortium |
| EPI_ISL_459327 | Regional Virus Laboratory, Belfast Health and Social Care Trust | Wellcome Sanger Institute for the COVID-19 Genomics UK Consortium |

| EPI_Accession_ID | Originating Laboratory | Submitting Laboratory |
|---|---|---|
| EPI_ISL_459328 | Regional Virus Laboratory, Belfast Health and Social Care Trust | Wellcome Sanger Institute for the COVID-19 Genomics UK Consortium |
| EPI_ISL_459329 | Regional Virus Laboratory, Belfast Health and Social Care Trust | Wellcome Sanger Institute for the COVID-19 Genomics UK Consortium |
| EPI_ISL_459330 | Regional Virus Laboratory, Belfast Health and Social Care Trust | Wellcome Sanger Institute for the COVID-19 Genomics UK Consortium |
| EPI_ISL_459331 | Regional Virus Laboratory, Belfast Health and Social Care Trust | Wellcome Sanger Institute for the COVID-19 Genomics UK Consortium |
| EPI_ISL_459332 | Regional Virus Laboratory, Belfast Health and Social Care Trust | Wellcome Sanger Institute for the COVID-19 Genomics UK Consortium |
| EPI_ISL_459333 | Regional Virus Laboratory, Belfast Health and Social Care Trust | Wellcome Sanger Institute for the COVID-19 Genomics UK Consortium |
| EPI_ISL_459334 | Regional Virus Laboratory, Belfast Health and Social Care Trust | Wellcome Sanger Institute for the COVID-19 Genomics UK Consortium |
| EPI_ISL_459335 | Regional Virus Laboratory, Belfast Health and Social Care Trust | Wellcome Sanger Institute for the COVID-19 Genomics UK Consortium |
| EPI_ISL_459336 | Regional Virus Laboratory, Belfast Health and Social Care Trust | Wellcome Sanger Institute for the COVID-19 Genomics UK Consortium |
| EPI_ISL_459337 | Regional Virus Laboratory, Belfast Health and Social Care Trust | Wellcome Sanger Institute for the COVID-19 Genomics UK Consortium |
| EPI_ISL_459338 | Regional Virus Laboratory, Belfast Health and Social Care Trust | Wellcome Sanger Institute for the COVID-19 Genomics UK Consortium |
| EPI_ISL_459339 | Regional Virus Laboratory, Belfast Health and Social Care Trust | Wellcome Sanger Institute for the COVID-19 Genomics UK Consortium |
| EPI_ISL_459340 | Regional Virus Laboratory, Belfast Health and Social Care Trust | Wellcome Sanger Institute for the COVID-19 Genomics UK Consortium |
| EPI_ISL_459341 | Regional Virus Laboratory, Belfast Health and Social Care Trust | Wellcome Sanger Institute for the COVID-19 Genomics UK Consortium |
| EPI_ISL_459342 | Regional Virus Laboratory, Belfast Health and Social Care Trust | Wellcome Sanger Institute for the COVID-19 Genomics UK Consortium |
| EPI_ISL_459343 | Regional Virus Laboratory, Belfast Health and Social Care Trust | Wellcome Sanger Institute for the COVID-19 Genomics UK Consortium |

| EPI_Accession_ID | Originating Laboratory | Submitting Laboratory |
|---|---|---|
| EPI_ISL_459344 | Regional Virus Laboratory, Belfast Health and Social Care Trust | Wellcome Sanger Institute for the COVID-19 Genomics UK Consortium |
| EPI_ISL_459345 | Regional Virus Laboratory, Belfast Health and Social Care Trust | Wellcome Sanger Institute for the COVID-19 Genomics UK Consortium |
| EPI_ISL_459346 | Regional Virus Laboratory, Belfast Health and Social Care Trust | Wellcome Sanger Institute for the COVID-19 Genomics UK Consortium |
| EPI_ISL_459347 | Regional Virus Laboratory, Belfast Health and Social Care Trust | Wellcome Sanger Institute for the COVID-19 Genomics UK Consortium |
| EPI_ISL_459348 | Regional Virus Laboratory, Belfast Health and Social Care Trust | Wellcome Sanger Institute for the COVID-19 Genomics UK Consortium |
| EPI_ISL_459349 | Regional Virus Laboratory, Belfast Health and Social Care Trust | Wellcome Sanger Institute for the COVID-19 Genomics UK Consortium |
| EPI_ISL_459350 | Regional Virus Laboratory, Belfast Health and Social Care Trust | Wellcome Sanger Institute for the COVID-19 Genomics UK Consortium |
| EPI_ISL_459351 | Regional Virus Laboratory, Belfast Health and Social Care Trust | Wellcome Sanger Institute for the COVID-19 Genomics UK Consortium |
| EPI_ISL_459352 | Regional Virus Laboratory, Belfast Health and Social Care Trust | Wellcome Sanger Institute for the COVID-19 Genomics UK Consortium |
| EPI_ISL_459353 | Regional Virus Laboratory, Belfast Health and Social Care Trust | Wellcome Sanger Institute for the COVID-19 Genomics UK Consortium |
| EPI_ISL_459354 | Regional Virus Laboratory, Belfast Health and Social Care Trust | Wellcome Sanger Institute for the COVID-19 Genomics UK Consortium |
| EPI_ISL_459355 | Regional Virus Laboratory, Belfast Health and Social Care Trust | Wellcome Sanger Institute for the COVID-19 Genomics UK Consortium |
| EPI_ISL_459356 | Regional Virus Laboratory, Belfast Health and Social Care Trust | Wellcome Sanger Institute for the COVID-19 Genomics UK Consortium |
| EPI_ISL_459357 | Regional Virus Laboratory, Belfast Health and Social Care Trust | Wellcome Sanger Institute for the COVID-19 Genomics UK Consortium |
| EPI_ISL_459358 | Regional Virus Laboratory, Belfast Health and Social Care Trust | Wellcome Sanger Institute for the COVID-19 Genomics UK Consortium |
| EPI_ISL_459359 | Regional Virus Laboratory, Belfast Health and Social Care Trust | Wellcome Sanger Institute for the COVID-19 Genomics UK Consortium |

| EPI_Accession_ID | Originating Laboratory | Submitting Laboratory |
|---|---|---|
| EPI_ISL_459360 | Regional Virus Laboratory, Belfast Health and Social Care Trust | Wellcome Sanger Institute for the COVID-19 Genomics UK Consortium |
| EPI_ISL_459361 | Regional Virus Laboratory, Belfast Health and Social Care Trust | Wellcome Sanger Institute for the COVID-19 Genomics UK Consortium |
| EPI_ISL_459362 | Regional Virus Laboratory, Belfast Health and Social Care Trust | Wellcome Sanger Institute for the COVID-19 Genomics UK Consortium |
| EPI_ISL_459363 | Regional Virus Laboratory, Belfast Health and Social Care Trust | Wellcome Sanger Institute for the COVID-19 Genomics UK Consortium |
| EPI_ISL_459364 | Regional Virus Laboratory, Belfast Health and Social Care Trust | Wellcome Sanger Institute for the COVID-19 Genomics UK Consortium |
| EPI_ISL_459365 | Regional Virus Laboratory, Belfast Health and Social Care Trust | Wellcome Sanger Institute for the COVID-19 Genomics UK Consortium |
| EPI_ISL_459366 | Regional Virus Laboratory, Belfast Health and Social Care Trust | Wellcome Sanger Institute for the COVID-19 Genomics UK Consortium |
| EPI_ISL_459367 | Regional Virus Laboratory, Belfast Health and Social Care Trust | Wellcome Sanger Institute for the COVID-19 Genomics UK Consortium |
| EPI_ISL_459368 | Regional Virus Laboratory, Belfast Health and Social Care Trust | Wellcome Sanger Institute for the COVID-19 Genomics UK Consortium |
| EPI_ISL_459369 | Regional Virus Laboratory, Belfast Health and Social Care Trust | Wellcome Sanger Institute for the COVID-19 Genomics UK Consortium |
| EPI_ISL_459370 | Regional Virus Laboratory, Belfast Health and Social Care Trust | Wellcome Sanger Institute for the COVID-19 Genomics UK Consortium |
| EPI_ISL_459371 | Regional Virus Laboratory, Belfast Health and Social Care Trust | Wellcome Sanger Institute for the COVID-19 Genomics UK Consortium |
| EPI_ISL_459372 | Regional Virus Laboratory, Belfast Health and Social Care Trust | Wellcome Sanger Institute for the COVID-19 Genomics UK Consortium |
| EPI_ISL_459373 | Regional Virus Laboratory, Belfast Health and Social Care Trust | Wellcome Sanger Institute for the COVID-19 Genomics UK Consortium |
| EPI_ISL_459374 | Regional Virus Laboratory, Belfast Health and Social Care Trust | Wellcome Sanger Institute for the COVID-19 Genomics UK Consortium |
| EPI_ISL_459375 | Regional Virus Laboratory, Belfast Health and Social Care Trust | Wellcome Sanger Institute for the COVID-19 Genomics UK Consortium |

| EPI_Accession_ID | Originating Laboratory | Submitting Laboratory |
|---|---|---|
| EPI_ISL_459376 | Regional Virus Laboratory, Belfast Health and Social Care Trust | Wellcome Sanger Institute for the COVID-19 Genomics UK Consortium |
| EPI_ISL_459377 | Regional Virus Laboratory, Belfast Health and Social Care Trust | Wellcome Sanger Institute for the COVID-19 Genomics UK Consortium |
| EPI_ISL_459378 | Regional Virus Laboratory, Belfast Health and Social Care Trust | Wellcome Sanger Institute for the COVID-19 Genomics UK Consortium |
| EPI_ISL_459379 | Regional Virus Laboratory, Belfast Health and Social Care Trust | Wellcome Sanger Institute for the COVID-19 Genomics UK Consortium |
| EPI_ISL_459380 | Regional Virus Laboratory, Belfast Health and Social Care Trust | Wellcome Sanger Institute for the COVID-19 Genomics UK Consortium |
| EPI_ISL_459381 | Regional Virus Laboratory, Belfast Health and Social Care Trust | Wellcome Sanger Institute for the COVID-19 Genomics UK Consortium |
| EPI_ISL_459382 | Regional Virus Laboratory, Belfast Health and Social Care Trust | Wellcome Sanger Institute for the COVID-19 Genomics UK Consortium |
| EPI_ISL_459383 | Regional Virus Laboratory, Belfast Health and Social Care Trust | Wellcome Sanger Institute for the COVID-19 Genomics UK Consortium |
| EPI_ISL_459384 | Regional Virus Laboratory, Belfast Health and Social Care Trust | Wellcome Sanger Institute for the COVID-19 Genomics UK Consortium |
| EPI_ISL_459385 | Regional Virus Laboratory, Belfast Health and Social Care Trust | Wellcome Sanger Institute for the COVID-19 Genomics UK Consortium |
| EPI_ISL_459386 | Regional Virus Laboratory, Belfast Health and Social Care Trust | Wellcome Sanger Institute for the COVID-19 Genomics UK Consortium |
| EPI_ISL_459387 | Regional Virus Laboratory, Belfast Health and Social Care Trust | Wellcome Sanger Institute for the COVID-19 Genomics UK Consortium |
| EPI_ISL_459388 | Regional Virus Laboratory, Belfast Health and Social Care Trust | Wellcome Sanger Institute for the COVID-19 Genomics UK Consortium |
| EPI_ISL_459389 | Regional Virus Laboratory, Belfast Health and Social Care Trust | Wellcome Sanger Institute for the COVID-19 Genomics UK Consortium |
| EPI_ISL_459390 | Regional Virus Laboratory, Belfast Health and Social Care Trust | Wellcome Sanger Institute for the COVID-19 Genomics UK Consortium |
| EPI_ISL_459391 | Regional Virus Laboratory, Belfast Health and Social Care Trust | Wellcome Sanger Institute for the COVID-19 Genomics UK Consortium |

| EPI_Accession_ID | Originating Laboratory | Submitting Laboratory |
|---|---|---|
| EPI_ISL_459392 | Regional Virus Laboratory, Belfast Health and Social Care Trust | Wellcome Sanger Institute for the COVID-19 Genomics UK Consortium |
| EPI_ISL_459393 | Regional Virus Laboratory, Belfast Health and Social Care Trust | Wellcome Sanger Institute for the COVID-19 Genomics UK Consortium |
| EPI_ISL_459394 | Regional Virus Laboratory, Belfast Health and Social Care Trust | Wellcome Sanger Institute for the COVID-19 Genomics UK Consortium |
| EPI_ISL_459395 | Regional Virus Laboratory, Belfast Health and Social Care Trust | Wellcome Sanger Institute for the COVID-19 Genomics UK Consortium |
| EPI_ISL_459396 | Regional Virus Laboratory, Belfast Health and Social Care Trust | Wellcome Sanger Institute for the COVID-19 Genomics UK Consortium |
| EPI_ISL_459397 | Regional Virus Laboratory, Belfast Health and Social Care Trust | Wellcome Sanger Institute for the COVID-19 Genomics UK Consortium |
| EPI_ISL_459398 | Regional Virus Laboratory, Belfast Health and Social Care Trust | Wellcome Sanger Institute for the COVID-19 Genomics UK Consortium |
| EPI_ISL_459399 | Regional Virus Laboratory, Belfast Health and Social Care Trust | Wellcome Sanger Institute for the COVID-19 Genomics UK Consortium |
| EPI_ISL_459400 | Regional Virus Laboratory, Belfast Health and Social Care Trust | Wellcome Sanger Institute for the COVID-19 Genomics UK Consortium |
| EPI_ISL_459401 | Regional Virus Laboratory, Belfast Health and Social Care Trust | Wellcome Sanger Institute for the COVID-19 Genomics UK Consortium |
| EPI_ISL_459402 | Regional Virus Laboratory, Belfast Health and Social Care Trust | Wellcome Sanger Institute for the COVID-19 Genomics UK Consortium |
| EPI_ISL_459403 | Regional Virus Laboratory, Belfast Health and Social Care Trust | Wellcome Sanger Institute for the COVID-19 Genomics UK Consortium |
| EPI_ISL_459404 | Regional Virus Laboratory, Belfast Health and Social Care Trust | Wellcome Sanger Institute for the COVID-19 Genomics UK Consortium |
| EPI_ISL_459405 | Regional Virus Laboratory, Belfast Health and Social Care Trust | Wellcome Sanger Institute for the COVID-19 Genomics UK Consortium |
| EPI_ISL_459406 | Regional Virus Laboratory, Belfast Health and Social Care Trust | Wellcome Sanger Institute for the COVID-19 Genomics UK Consortium |
| EPI_ISL_459407 | Regional Virus Laboratory, Belfast Health and Social Care Trust | Wellcome Sanger Institute for the COVID-19 Genomics UK Consortium |

| EPI_Accession_ID | Originating Laboratory | Submitting Laboratory |
|---|---|---|
| EPI_ISL_459408 | Regional Virus Laboratory, Belfast Health and Social Care Trust | Wellcome Sanger Institute for the COVID-19 Genomics UK Consortium |
| EPI_ISL_459409 | Regional Virus Laboratory, Belfast Health and Social Care Trust | Wellcome Sanger Institute for the COVID-19 Genomics UK Consortium |
| EPI_ISL_459410 | Department of Pathology, University of Cambridge | Wellcome Sanger Institute for the COVID-19 Genomics UK Consortium |
| EPI_ISL_459411 | Department of Pathology, University of Cambridge | Wellcome Sanger Institute for the COVID-19 Genomics UK Consortium |
| EPI_ISL_459412 | Department of Pathology, University of Cambridge | Wellcome Sanger Institute for the COVID-19 Genomics UK Consortium |
| EPI_ISL_459413 | Department of Pathology, University of Cambridge | Wellcome Sanger Institute for the COVID-19 Genomics UK Consortium |
| EPI_ISL_459414 | Department of Pathology, University of Cambridge | Wellcome Sanger Institute for the COVID-19 Genomics UK Consortium |
| EPI_ISL_459415 | Department of Pathology, University of Cambridge | Wellcome Sanger Institute for the COVID-19 Genomics UK Consortium |
| EPI_ISL_459416 | Department of Pathology, University of Cambridge | Wellcome Sanger Institute for the COVID-19 Genomics UK Consortium |
| EPI_ISL_459417 | Department of Pathology, University of Cambridge | Wellcome Sanger Institute for the COVID-19 Genomics UK Consortium |
| EPI_ISL_459418 | Department of Pathology, University of Cambridge | Wellcome Sanger Institute for the COVID-19 Genomics UK Consortium |
| EPI_ISL_459419 | Department of Pathology, University of Cambridge | Wellcome Sanger Institute for the COVID-19 Genomics UK Consortium |
| EPI_ISL_459420 | Department of Pathology, University of Cambridge | Wellcome Sanger Institute for the COVID-19 Genomics UK Consortium |
| EPI_ISL_459421 | Department of Pathology, University of Cambridge | Wellcome Sanger Institute for the COVID-19 Genomics UK Consortium |
| EPI_ISL_459422 | Department of Pathology, University of Cambridge | Wellcome Sanger Institute for the COVID-19 Genomics UK Consortium |
| EPI_ISL_459423 | Department of Pathology, University of Cambridge | Wellcome Sanger Institute for the COVID-19 Genomics UK Consortium |

| EPI_Accession_ID | Originating Laboratory | Submitting Laboratory |
|---|---|---|
| EPI_ISL_459424 | Department of Pathology, University of Cambridge | Wellcome Sanger Institute for the COVID-19 Genomics UK Consortium |
| EPI_ISL_459425 | Department of Pathology, University of Cambridge | Wellcome Sanger Institute for the COVID-19 Genomics UK Consortium |
| EPI_ISL_459426 | Department of Pathology, University of Cambridge | Wellcome Sanger Institute for the COVID-19 Genomics UK Consortium |
| EPI_ISL_459427 | Department of Pathology, University of Cambridge | Wellcome Sanger Institute for the COVID-19 Genomics UK Consortium |
| EPI_ISL_459428 | Department of Pathology, University of Cambridge | Wellcome Sanger Institute for the COVID-19 Genomics UK Consortium |
| EPI_ISL_459429 | Department of Pathology, University of Cambridge | Wellcome Sanger Institute for the COVID-19 Genomics UK Consortium |
| EPI_ISL_459430 | Department of Pathology, University of Cambridge | Wellcome Sanger Institute for the COVID-19 Genomics UK Consortium |
| EPI_ISL_459431 | Department of Pathology, University of Cambridge | Wellcome Sanger Institute for the COVID-19 Genomics UK Consortium |
| EPI_ISL_459432 | Department of Pathology, University of Cambridge | Wellcome Sanger Institute for the COVID-19 Genomics UK Consortium |
| EPI_ISL_459433 | Department of Pathology, University of Cambridge | Wellcome Sanger Institute for the COVID-19 Genomics UK Consortium |
| EPI_ISL_459434 | Department of Pathology, University of Cambridge | Wellcome Sanger Institute for the COVID-19 Genomics UK Consortium |
| EPI_ISL_459435 | Department of Pathology, University of Cambridge | Wellcome Sanger Institute for the COVID-19 Genomics UK Consortium |
| EPI_ISL_459436 | Department of Pathology, University of Cambridge | Wellcome Sanger Institute for the COVID-19 Genomics UK Consortium |
| EPI_ISL_459437 | Department of Pathology, University of Cambridge | Wellcome Sanger Institute for the COVID-19 Genomics UK Consortium |
| EPI_ISL_459438 | Department of Pathology, University of Cambridge | Wellcome Sanger Institute for the COVID-19 Genomics UK Consortium |
| EPI_ISL_459439 | Department of Pathology, University of Cambridge | Wellcome Sanger Institute for the COVID-19 Genomics UK Consortium |

| EPI_Accession_ID | Originating Laboratory | Submitting Laboratory |
|---|---|---|
| EPI_ISL_459440 | Department of Pathology, University of Cambridge | Wellcome Sanger Institute for the COVID-19 Genomics UK Consortium |
| EPI_ISL_459441 | Department of Pathology, University of Cambridge | Wellcome Sanger Institute for the COVID-19 Genomics UK Consortium |
| EPI_ISL_459442 | Department of Pathology, University of Cambridge | Wellcome Sanger Institute for the COVID-19 Genomics UK Consortium |
| EPI_ISL_459443 | Department of Pathology, University of Cambridge | Wellcome Sanger Institute for the COVID-19 Genomics UK Consortium |
| EPI_ISL_459444 | Department of Pathology, University of Cambridge | Wellcome Sanger Institute for the COVID-19 Genomics UK Consortium |
| EPI_ISL_459445 | Department of Pathology, University of Cambridge | Wellcome Sanger Institute for the COVID-19 Genomics UK Consortium |
| EPI_ISL_459446 | Department of Pathology, University of Cambridge | Wellcome Sanger Institute for the COVID-19 Genomics UK Consortium |
| EPI_ISL_459447 | Department of Pathology, University of Cambridge | Wellcome Sanger Institute for the COVID-19 Genomics UK Consortium |
| EPI_ISL_459448 | Department of Pathology, University of Cambridge | Wellcome Sanger Institute for the COVID-19 Genomics UK Consortium |
| EPI_ISL_459449 | Department of Pathology, University of Cambridge | Wellcome Sanger Institute for the COVID-19 Genomics UK Consortium |
| EPI_ISL_459450 | Department of Pathology, University of Cambridge | Wellcome Sanger Institute for the COVID-19 Genomics UK Consortium |
| EPI_ISL_459451 | Department of Pathology, University of Cambridge | Wellcome Sanger Institute for the COVID-19 Genomics UK Consortium |
| EPI_ISL_459452 | Department of Pathology, University of Cambridge | Wellcome Sanger Institute for the COVID-19 Genomics UK Consortium |
| EPI_ISL_459453 | Department of Pathology, University of Cambridge | Wellcome Sanger Institute for the COVID-19 Genomics UK Consortium |
| EPI_ISL_459454 | Department of Pathology, University of Cambridge | Wellcome Sanger Institute for the COVID-19 Genomics UK Consortium |
| EPI_ISL_459455 | Department of Pathology, University of Cambridge | Wellcome Sanger Institute for the COVID-19 Genomics UK Consortium |

| EPI_Accession_ID | Originating Laboratory | Submitting Laboratory |
|---|---|---|
| EPI_ISL_459456 | Department of Pathology, University of Cambridge | Wellcome Sanger Institute for the COVID-19 Genomics UK Consortium |
| EPI_ISL_459457 | Department of Pathology, University of Cambridge | Wellcome Sanger Institute for the COVID-19 Genomics UK Consortium |
| EPI_ISL_459458 | Department of Pathology, University of Cambridge | Wellcome Sanger Institute for the COVID-19 Genomics UK Consortium |
| EPI_ISL_459459 | Department of Pathology, University of Cambridge | Wellcome Sanger Institute for the COVID-19 Genomics UK Consortium |
| EPI_ISL_459460 | Department of Pathology, University of Cambridge | Wellcome Sanger Institute for the COVID-19 Genomics UK Consortium |
| EPI_ISL_459461 | Department of Pathology, University of Cambridge | Wellcome Sanger Institute for the COVID-19 Genomics UK Consortium |
| EPI_ISL_459462 | Department of Pathology, University of Cambridge | Wellcome Sanger Institute for the COVID-19 Genomics UK Consortium |
| EPI_ISL_459463 | Department of Pathology, University of Cambridge | Wellcome Sanger Institute for the COVID-19 Genomics UK Consortium |
| EPI_ISL_459464 | Department of Pathology, University of Cambridge | Wellcome Sanger Institute for the COVID-19 Genomics UK Consortium |
| EPI_ISL_459465 | Department of Pathology, University of Cambridge | Wellcome Sanger Institute for the COVID-19 Genomics UK Consortium |
| EPI_ISL_459466 | Department of Pathology, University of Cambridge | Wellcome Sanger Institute for the COVID-19 Genomics UK Consortium |
| EPI_ISL_459467 | Department of Pathology, University of Cambridge | Wellcome Sanger Institute for the COVID-19 Genomics UK Consortium |
| EPI_ISL_459468 | Department of Pathology, University of Cambridge | Wellcome Sanger Institute for the COVID-19 Genomics UK Consortium |
| EPI_ISL_459469 | Department of Pathology, University of Cambridge | Wellcome Sanger Institute for the COVID-19 Genomics UK Consortium |
| EPI_ISL_459470 | Department of Pathology, University of Cambridge | Wellcome Sanger Institute for the COVID-19 Genomics UK Consortium |
| EPI_ISL_459471 | Department of Pathology, University of Cambridge | Wellcome Sanger Institute for the COVID-19 Genomics UK Consortium |

| EPI_Accession_ID | Originating Laboratory | Submitting Laboratory |
| --- | --- | --- |
| EPI_ISL_459472 | Department of Pathology, University of Cambridge | Wellcome Sanger Institute for the COVID-19 Genomics UK Consortium |
| EPI_ISL_459473 | Department of Pathology, University of Cambridge | Wellcome Sanger Institute for the COVID-19 Genomics UK Consortium |
| EPI_ISL_459474 | Department of Pathology, University of Cambridge | Wellcome Sanger Institute for the COVID-19 Genomics UK Consortium |
| EPI_ISL_459475 | Department of Pathology, University of Cambridge | Wellcome Sanger Institute for the COVID-19 Genomics UK Consortium |
| EPI_ISL_459476 | Department of Pathology, University of Cambridge | Wellcome Sanger Institute for the COVID-19 Genomics UK Consortium |
| EPI_ISL_459477 | Department of Pathology, University of Cambridge | Wellcome Sanger Institute for the COVID-19 Genomics UK Consortium |
| EPI_ISL_459478 | Department of Pathology, University of Cambridge | Wellcome Sanger Institute for the COVID-19 Genomics UK Consortium |
| EPI_ISL_459479 | Department of Pathology, University of Cambridge | Wellcome Sanger Institute for the COVID-19 Genomics UK Consortium |
| EPI_ISL_459480 | Department of Pathology, University of Cambridge | Wellcome Sanger Institute for the COVID-19 Genomics UK Consortium |
| EPI_ISL_459481 | Department of Pathology, University of Cambridge | Wellcome Sanger Institute for the COVID-19 Genomics UK Consortium |
| EPI_ISL_459482 | Department of Pathology, University of Cambridge | Wellcome Sanger Institute for the COVID-19 Genomics UK Consortium |
| EPI_ISL_459483 | Department of Pathology, University of Cambridge | Wellcome Sanger Institute for the COVID-19 Genomics UK Consortium |
| EPI_ISL_459484 | Department of Pathology, University of Cambridge | Wellcome Sanger Institute for the COVID-19 Genomics UK Consortium |
| EPI_ISL_459485 | Department of Pathology, University of Cambridge | Wellcome Sanger Institute for the COVID-19 Genomics UK Consortium |
| EPI_ISL_459486 | Department of Pathology, University of Cambridge | Wellcome Sanger Institute for the COVID-19 Genomics UK Consortium |
| EPI_ISL_459487 | Department of Pathology, University of Cambridge | Wellcome Sanger Institute for the COVID-19 Genomics UK Consortium |

| EPI_Accession_ID | Originating Laboratory | Submitting Laboratory |
|---|---|---|
| EPI_ISL_459488 | Department of Pathology, University of Cambridge | Wellcome Sanger Institute for the COVID-19 Genomics UK Consortium |
| EPI_ISL_459489 | Department of Pathology, University of Cambridge | Wellcome Sanger Institute for the COVID-19 Genomics UK Consortium |
| EPI_ISL_459490 | Department of Pathology, University of Cambridge | Wellcome Sanger Institute for the COVID-19 Genomics UK Consortium |
| EPI_ISL_459491 | Department of Pathology, University of Cambridge | Wellcome Sanger Institute for the COVID-19 Genomics UK Consortium |
| EPI_ISL_459492 | Department of Pathology, University of Cambridge | Wellcome Sanger Institute for the COVID-19 Genomics UK Consortium |
| EPI_ISL_459493 | Department of Pathology, University of Cambridge | Wellcome Sanger Institute for the COVID-19 Genomics UK Consortium |
| EPI_ISL_459494 | Department of Pathology, University of Cambridge | Wellcome Sanger Institute for the COVID-19 Genomics UK Consortium |
| EPI_ISL_459495 | Department of Pathology, University of Cambridge | Wellcome Sanger Institute for the COVID-19 Genomics UK Consortium |
| EPI_ISL_459496 | Department of Pathology, University of Cambridge | Wellcome Sanger Institute for the COVID-19 Genomics UK Consortium |
| EPI_ISL_459497 | Department of Pathology, University of Cambridge | Wellcome Sanger Institute for the COVID-19 Genomics UK Consortium |
| EPI_ISL_459498 | Department of Pathology, University of Cambridge | Wellcome Sanger Institute for the COVID-19 Genomics UK Consortium |
| EPI_ISL_459499 | Department of Pathology, University of Cambridge | Wellcome Sanger Institute for the COVID-19 Genomics UK Consortium |
| EPI_ISL_459500 | Department of Pathology, University of Cambridge | Wellcome Sanger Institute for the COVID-19 Genomics UK Consortium |
| EPI_ISL_459501 | Department of Pathology, University of Cambridge | Wellcome Sanger Institute for the COVID-19 Genomics UK Consortium |
| EPI_ISL_459502 | Department of Pathology, University of Cambridge | Wellcome Sanger Institute for the COVID-19 Genomics UK Consortium |
| EPI_ISL_459503 | Department of Pathology, University of Cambridge | Wellcome Sanger Institute for the COVID-19 Genomics UK Consortium |

| EPI_Accession_ID | Originating Laboratory | Submitting Laboratory |
|---|---|---|
| EPI_ISL_459504 | Department of Pathology, University of Cambridge | Wellcome Sanger Institute for the COVID-19 Genomics UK Consortium |
| EPI_ISL_459505 | NHSGGC West of Scotland Specialist Virology Centre / MRC-University of Glasgow Centre for Virus Research | Wellcome Sanger Institute for the COVID-19 Genomics UK Consortium |
| EPI_ISL_459506 | NHSGGC West of Scotland Specialist Virology Centre / MRC-University of Glasgow Centre for Virus Research | Wellcome Sanger Institute for the COVID-19 Genomics UK Consortium |
| EPI_ISL_459507 | NHSGGC West of Scotland Specialist Virology Centre / MRC-University of Glasgow Centre for Virus Research | Wellcome Sanger Institute for the COVID-19 Genomics UK Consortium |
| EPI_ISL_459508 | NHSGGC West of Scotland Specialist Virology Centre / MRC-University of Glasgow Centre for Virus Research | Wellcome Sanger Institute for the COVID-19 Genomics UK Consortium |
| EPI_ISL_459509 | NHSGGC West of Scotland Specialist Virology Centre / MRC-University of Glasgow Centre for Virus Research | Wellcome Sanger Institute for the COVID-19 Genomics UK Consortium |
| EPI_ISL_459510 | NHSGGC West of Scotland Specialist Virology Centre / MRC-University of Glasgow Centre for Virus Research | Wellcome Sanger Institute for the COVID-19 Genomics UK Consortium |
| EPI_ISL_459511 | NHSGGC West of Scotland Specialist Virology Centre / MRC-University of Glasgow Centre for Virus Research | Wellcome Sanger Institute for the COVID-19 Genomics UK Consortium |
| EPI_ISL_459512 | NHSGGC West of Scotland Specialist Virology Centre / MRC-University of Glasgow Centre for Virus Research | Wellcome Sanger Institute for the COVID-19 Genomics UK Consortium |
| EPI_ISL_459513 | NHSGGC West of Scotland Specialist Virology Centre / MRC-University of Glasgow Centre for Virus Research | Wellcome Sanger Institute for the COVID-19 Genomics UK Consortium |
| EPI_ISL_459514 | NHSGGC West of Scotland Specialist Virology Centre / MRC-University of Glasgow Centre for Virus Research | Wellcome Sanger Institute for the COVID-19 Genomics UK Consortium |
| EPI_ISL_459515 | NHSGGC West of Scotland Specialist Virology Centre / MRC-University of Glasgow Centre for Virus Research | Wellcome Sanger Institute for the COVID-19 Genomics UK Consortium |
| EPI_ISL_459516 | NHSGGC West of Scotland Specialist Virology Centre / MRC-University of Glasgow Centre for Virus Research | Wellcome Sanger Institute for the COVID-19 Genomics UK Consortium |
| EPI_ISL_459517 | NHSGGC West of Scotland Specialist Virology Centre / MRC-University of Glasgow Centre for Virus Research | Wellcome Sanger Institute for the COVID-19 Genomics UK Consortium |
| EPI_ISL_459518 | NHSGGC West of Scotland Specialist Virology Centre / MRC-University of Glasgow Centre for Virus Research | Wellcome Sanger Institute for the COVID-19 Genomics UK Consortium |
| EPI_ISL_459519 | NHSGGC West of Scotland Specialist Virology Centre / MRC-University of Glasgow Centre for Virus Research | Wellcome Sanger Institute for the COVID-19 Genomics UK Consortium |

| EPI_Accession_ID | Originating Laboratory | Submitting Laboratory |
|---|---|---|
| EPI_ISL_459520 | NHSGGC West of Scotland Specialist Virology Centre / MRC-University of Glasgow Centre for Virus Research | Wellcome Sanger Institute for the COVID-19 Genomics UK Consortium |
| EPI_ISL_459521 | NHSGGC West of Scotland Specialist Virology Centre / MRC-University of Glasgow Centre for Virus Research | Wellcome Sanger Institute for the COVID-19 Genomics UK Consortium |
| EPI_ISL_459522 | NHSGGC West of Scotland Specialist Virology Centre / MRC-University of Glasgow Centre for Virus Research | Wellcome Sanger Institute for the COVID-19 Genomics UK Consortium |
| EPI_ISL_459523 | NHSGGC West of Scotland Specialist Virology Centre / MRC-University of Glasgow Centre for Virus Research | Wellcome Sanger Institute for the COVID-19 Genomics UK Consortium |
| EPI_ISL_459524 | NHSGGC West of Scotland Specialist Virology Centre / MRC-University of Glasgow Centre for Virus Research | Wellcome Sanger Institute for the COVID-19 Genomics UK Consortium |
| EPI_ISL_459525 | NHSGGC West of Scotland Specialist Virology Centre / MRC-University of Glasgow Centre for Virus Research | Wellcome Sanger Institute for the COVID-19 Genomics UK Consortium |
| EPI_ISL_459526 | NHSGGC West of Scotland Specialist Virology Centre / MRC-University of Glasgow Centre for Virus Research | Wellcome Sanger Institute for the COVID-19 Genomics UK Consortium |
| EPI_ISL_459527 | NHSGGC West of Scotland Specialist Virology Centre / MRC-University of Glasgow Centre for Virus Research | Wellcome Sanger Institute for the COVID-19 Genomics UK Consortium |
| EPI_ISL_459528 | NHSGGC West of Scotland Specialist Virology Centre / MRC-University of Glasgow Centre for Virus Research | Wellcome Sanger Institute for the COVID-19 Genomics UK Consortium |
| EPI_ISL_459529 | NHSGGC West of Scotland Specialist Virology Centre / MRC-University of Glasgow Centre for Virus Research | Wellcome Sanger Institute for the COVID-19 Genomics UK Consortium |
| EPI_ISL_459530 | NHSGGC West of Scotland Specialist Virology Centre / MRC-University of Glasgow Centre for Virus Research | Wellcome Sanger Institute for the COVID-19 Genomics UK Consortium |
| EPI_ISL_459531 | NHSGGC West of Scotland Specialist Virology Centre / MRC-University of Glasgow Centre for Virus Research | Wellcome Sanger Institute for the COVID-19 Genomics UK Consortium |
| EPI_ISL_459532 | NHSGGC West of Scotland Specialist Virology Centre / MRC-University of Glasgow Centre for Virus Research | Wellcome Sanger Institute for the COVID-19 Genomics UK Consortium |
| EPI_ISL_459533 | NHSGGC West of Scotland Specialist Virology Centre / MRC-University of Glasgow Centre for Virus Research | Wellcome Sanger Institute for the COVID-19 Genomics UK Consortium |
| EPI_ISL_459534 | NHSGGC West of Scotland Specialist Virology Centre / MRC-University of Glasgow Centre for Virus Research | Wellcome Sanger Institute for the COVID-19 Genomics UK Consortium |
| EPI_ISL_459535 | NHSGGC West of Scotland Specialist Virology Centre / MRC-University of Glasgow Centre for Virus Research | Wellcome Sanger Institute for the COVID-19 Genomics UK Consortium |

| EPI_Accession_ID | Originating Laboratory | Submitting Laboratory |
|---|---|---|
| EPI_ISL_459536 | NHSGGC West of Scotland Specialist Virology Centre / MRC-University of Glasgow Centre for Virus Research | Wellcome Sanger Institute for the COVID-19 Genomics UK Consortium |
| EPI_ISL_459537 | NHSGGC West of Scotland Specialist Virology Centre / MRC-University of Glasgow Centre for Virus Research | Wellcome Sanger Institute for the COVID-19 Genomics UK Consortium |
| EPI_ISL_459538 | NHSGGC West of Scotland Specialist Virology Centre / MRC-University of Glasgow Centre for Virus Research | Wellcome Sanger Institute for the COVID-19 Genomics UK Consortium |
| EPI_ISL_459539 | NHSGGC West of Scotland Specialist Virology Centre / MRC-University of Glasgow Centre for Virus Research | Wellcome Sanger Institute for the COVID-19 Genomics UK Consortium |
| EPI_ISL_459540 | NHSGGC West of Scotland Specialist Virology Centre / MRC-University of Glasgow Centre for Virus Research | Wellcome Sanger Institute for the COVID-19 Genomics UK Consortium |
| EPI_ISL_459541 | NHSGGC West of Scotland Specialist Virology Centre / MRC-University of Glasgow Centre for Virus Research | Wellcome Sanger Institute for the COVID-19 Genomics UK Consortium |
| EPI_ISL_459542 | NHSGGC West of Scotland Specialist Virology Centre / MRC-University of Glasgow Centre for Virus Research | Wellcome Sanger Institute for the COVID-19 Genomics UK Consortium |
| EPI_ISL_459543 | NHSGGC West of Scotland Specialist Virology Centre / MRC-University of Glasgow Centre for Virus Research | Wellcome Sanger Institute for the COVID-19 Genomics UK Consortium |
| EPI_ISL_459544 | NHSGGC West of Scotland Specialist Virology Centre / MRC-University of Glasgow Centre for Virus Research | Wellcome Sanger Institute for the COVID-19 Genomics UK Consortium |
| EPI_ISL_459545 | NHSGGC West of Scotland Specialist Virology Centre / MRC-University of Glasgow Centre for Virus Research | Wellcome Sanger Institute for the COVID-19 Genomics UK Consortium |
| EPI_ISL_459546 | NHSGGC West of Scotland Specialist Virology Centre / MRC-University of Glasgow Centre for Virus Research | Wellcome Sanger Institute for the COVID-19 Genomics UK Consortium |
| EPI_ISL_459547 | NHSGGC West of Scotland Specialist Virology Centre / MRC-University of Glasgow Centre for Virus Research | Wellcome Sanger Institute for the COVID-19 Genomics UK Consortium |
| EPI_ISL_459548 | NHSGGC West of Scotland Specialist Virology Centre / MRC-University of Glasgow Centre for Virus Research | Wellcome Sanger Institute for the COVID-19 Genomics UK Consortium |
| EPI_ISL_459549 | NHSGGC West of Scotland Specialist Virology Centre / MRC-University of Glasgow Centre for Virus Research | Wellcome Sanger Institute for the COVID-19 Genomics UK Consortium |
| EPI_ISL_459550 | NHSGGC West of Scotland Specialist Virology Centre / MRC-University of Glasgow Centre for Virus Research | Wellcome Sanger Institute for the COVID-19 Genomics UK Consortium |
| EPI_ISL_459551 | NHSGGC West of Scotland Specialist Virology Centre / MRC-University of Glasgow Centre for Virus Research | Wellcome Sanger Institute for the COVID-19 Genomics UK Consortium |

| EPI_Accession_ID | Originating Laboratory | Submitting Laboratory |
|---|---|---|
| EPI_ISL_459552 | NHSGGC West of Scotland Specialist Virology Centre / MRC-University of Glasgow Centre for Virus Research | Wellcome Sanger Institute for the COVID-19 Genomics UK Consortium |
| EPI_ISL_459553 | NHSGGC West of Scotland Specialist Virology Centre / MRC-University of Glasgow Centre for Virus Research | Wellcome Sanger Institute for the COVID-19 Genomics UK Consortium |
| EPI_ISL_459554 | NHSGGC West of Scotland Specialist Virology Centre / MRC-University of Glasgow Centre for Virus Research | Wellcome Sanger Institute for the COVID-19 Genomics UK Consortium |
| EPI_ISL_459555 | NHSGGC West of Scotland Specialist Virology Centre / MRC-University of Glasgow Centre for Virus Research | Wellcome Sanger Institute for the COVID-19 Genomics UK Consortium |
| EPI_ISL_459556 | NHSGGC West of Scotland Specialist Virology Centre / MRC-University of Glasgow Centre for Virus Research | Wellcome Sanger Institute for the COVID-19 Genomics UK Consortium |
| EPI_ISL_459557 | NHSGGC West of Scotland Specialist Virology Centre / MRC-University of Glasgow Centre for Virus Research | Wellcome Sanger Institute for the COVID-19 Genomics UK Consortium |
| EPI_ISL_459558 | NHSGGC West of Scotland Specialist Virology Centre / MRC-University of Glasgow Centre for Virus Research | Wellcome Sanger Institute for the COVID-19 Genomics UK Consortium |
| EPI_ISL_459559 | NHSGGC West of Scotland Specialist Virology Centre / MRC-University of Glasgow Centre for Virus Research | Wellcome Sanger Institute for the COVID-19 Genomics UK Consortium |
| EPI_ISL_459560 | NHSGGC West of Scotland Specialist Virology Centre / MRC-University of Glasgow Centre for Virus Research | Wellcome Sanger Institute for the COVID-19 Genomics UK Consortium |
| EPI_ISL_459561 | NHSGGC West of Scotland Specialist Virology Centre / MRC-University of Glasgow Centre for Virus Research | Wellcome Sanger Institute for the COVID-19 Genomics UK Consortium |
| EPI_ISL_459562 | NHSGGC West of Scotland Specialist Virology Centre / MRC-University of Glasgow Centre for Virus Research | Wellcome Sanger Institute for the COVID-19 Genomics UK Consortium |
| EPI_ISL_459563 | NHSGGC West of Scotland Specialist Virology Centre / MRC-University of Glasgow Centre for Virus Research | Wellcome Sanger Institute for the COVID-19 Genomics UK Consortium |
| EPI_ISL_459564 | NHSGGC West of Scotland Specialist Virology Centre / MRC-University of Glasgow Centre for Virus Research | Wellcome Sanger Institute for the COVID-19 Genomics UK Consortium |
| EPI_ISL_459565 | NHSGGC West of Scotland Specialist Virology Centre / MRC-University of Glasgow Centre for Virus Research | Wellcome Sanger Institute for the COVID-19 Genomics UK Consortium |
| EPI_ISL_459566 | NHSGGC West of Scotland Specialist Virology Centre / MRC-University of Glasgow Centre for Virus Research | Wellcome Sanger Institute for the COVID-19 Genomics UK Consortium |
| EPI_ISL_459567 | NHSGGC West of Scotland Specialist Virology Centre / MRC-University of Glasgow Centre for Virus Research | Wellcome Sanger Institute for the COVID-19 Genomics UK Consortium |

| EPI_Accession_ID | Originating Laboratory | Submitting Laboratory |
|---|---|---|
| EPI_ISL_459568 | NHSGGC West of Scotland Specialist Virology Centre / MRC-University of Glasgow Centre for Virus Research | Wellcome Sanger Institute for the COVID-19 Genomics UK Consortium |
| EPI_ISL_459569 | NHSGGC West of Scotland Specialist Virology Centre / MRC-University of Glasgow Centre for Virus Research | Wellcome Sanger Institute for the COVID-19 Genomics UK Consortium |
| EPI_ISL_459570 | NHSGGC West of Scotland Specialist Virology Centre / MRC-University of Glasgow Centre for Virus Research | Wellcome Sanger Institute for the COVID-19 Genomics UK Consortium |
| EPI_ISL_459571 | NHSGGC West of Scotland Specialist Virology Centre / MRC-University of Glasgow Centre for Virus Research | Wellcome Sanger Institute for the COVID-19 Genomics UK Consortium |
| EPI_ISL_459572 | NHSGGC West of Scotland Specialist Virology Centre / MRC-University of Glasgow Centre for Virus Research | Wellcome Sanger Institute for the COVID-19 Genomics UK Consortium |
| EPI_ISL_459573 | NHSGGC West of Scotland Specialist Virology Centre / MRC-University of Glasgow Centre for Virus Research | Wellcome Sanger Institute for the COVID-19 Genomics UK Consortium |
| EPI_ISL_459574 | NHSGGC West of Scotland Specialist Virology Centre / MRC-University of Glasgow Centre for Virus Research | Wellcome Sanger Institute for the COVID-19 Genomics UK Consortium |
| EPI_ISL_459575 | NHSGGC West of Scotland Specialist Virology Centre / MRC-University of Glasgow Centre for Virus Research | Wellcome Sanger Institute for the COVID-19 Genomics UK Consortium |
| EPI_ISL_459576 | NHSGGC West of Scotland Specialist Virology Centre / MRC-University of Glasgow Centre for Virus Research | Wellcome Sanger Institute for the COVID-19 Genomics UK Consortium |
| EPI_ISL_459577 | NHSGGC West of Scotland Specialist Virology Centre / MRC-University of Glasgow Centre for Virus Research | Wellcome Sanger Institute for the COVID-19 Genomics UK Consortium |
| EPI_ISL_459578 | NHSGGC West of Scotland Specialist Virology Centre / MRC-University of Glasgow Centre for Virus Research | Wellcome Sanger Institute for the COVID-19 Genomics UK Consortium |
| EPI_ISL_459579 | NHSGGC West of Scotland Specialist Virology Centre / MRC-University of Glasgow Centre for Virus Research | Wellcome Sanger Institute for the COVID-19 Genomics UK Consortium |
| EPI_ISL_459580 | NHSGGC West of Scotland Specialist Virology Centre / MRC-University of Glasgow Centre for Virus Research | Wellcome Sanger Institute for the COVID-19 Genomics UK Consortium |
| EPI_ISL_459581 | NHSGGC West of Scotland Specialist Virology Centre / MRC-University of Glasgow Centre for Virus Research | Wellcome Sanger Institute for the COVID-19 Genomics UK Consortium |
| EPI_ISL_459582 | NHSGGC West of Scotland Specialist Virology Centre / MRC-University of Glasgow Centre for Virus Research | Wellcome Sanger Institute for the COVID-19 Genomics UK Consortium |
| EPI_ISL_459583 | NHSGGC West of Scotland Specialist Virology Centre / MRC-University of Glasgow Centre for Virus Research | Wellcome Sanger Institute for the COVID-19 Genomics UK Consortium |

| EPI_Accession_ID | Originating Laboratory | Submitting Laboratory |
|---|---|---|
| EPI_ISL_459584 | NHSGGC West of Scotland Specialist Virology Centre / MRC-University of Glasgow Centre for Virus Research | Wellcome Sanger Institute for the COVID-19 Genomics UK Consortium |
| EPI_ISL_459585 | NHSGGC West of Scotland Specialist Virology Centre / MRC-University of Glasgow Centre for Virus Research | Wellcome Sanger Institute for the COVID-19 Genomics UK Consortium |
| EPI_ISL_459586 | NHSGGC West of Scotland Specialist Virology Centre / MRC-University of Glasgow Centre for Virus Research | Wellcome Sanger Institute for the COVID-19 Genomics UK Consortium |
| EPI_ISL_459587 | NHSGGC West of Scotland Specialist Virology Centre / MRC-University of Glasgow Centre for Virus Research | Wellcome Sanger Institute for the COVID-19 Genomics UK Consortium |
| EPI_ISL_459588 | NHSGGC West of Scotland Specialist Virology Centre / MRC-University of Glasgow Centre for Virus Research | Wellcome Sanger Institute for the COVID-19 Genomics UK Consortium |
| EPI_ISL_459589 | NHSGGC West of Scotland Specialist Virology Centre / MRC-University of Glasgow Centre for Virus Research | Wellcome Sanger Institute for the COVID-19 Genomics UK Consortium |
| EPI_ISL_459590 | NHSGGC West of Scotland Specialist Virology Centre / MRC-University of Glasgow Centre for Virus Research | Wellcome Sanger Institute for the COVID-19 Genomics UK Consortium |
| EPI_ISL_459591 | NHSGGC West of Scotland Specialist Virology Centre / MRC-University of Glasgow Centre for Virus Research | Wellcome Sanger Institute for the COVID-19 Genomics UK Consortium |
| EPI_ISL_459592 | NHSGGC West of Scotland Specialist Virology Centre / MRC-University of Glasgow Centre for Virus Research | Wellcome Sanger Institute for the COVID-19 Genomics UK Consortium |
| EPI_ISL_459593 | NHSGGC West of Scotland Specialist Virology Centre / MRC-University of Glasgow Centre for Virus Research | Wellcome Sanger Institute for the COVID-19 Genomics UK Consortium |
| EPI_ISL_459594 | NHSGGC West of Scotland Specialist Virology Centre / MRC-University of Glasgow Centre for Virus Research | Wellcome Sanger Institute for the COVID-19 Genomics UK Consortium |
| EPI_ISL_459595 | NHSGGC West of Scotland Specialist Virology Centre / MRC-University of Glasgow Centre for Virus Research | Wellcome Sanger Institute for the COVID-19 Genomics UK Consortium |
| EPI_ISL_459596 | NHSGGC West of Scotland Specialist Virology Centre / MRC-University of Glasgow Centre for Virus Research | Wellcome Sanger Institute for the COVID-19 Genomics UK Consortium |
| EPI_ISL_459597 | NHSGGC West of Scotland Specialist Virology Centre / MRC-University of Glasgow Centre for Virus Research | Wellcome Sanger Institute for the COVID-19 Genomics UK Consortium |
| EPI_ISL_459598 | NHSGGC West of Scotland Specialist Virology Centre / MRC-University of Glasgow Centre for Virus Research | Wellcome Sanger Institute for the COVID-19 Genomics UK Consortium |
| EPI_ISL_459599 | NHSGGC West of Scotland Specialist Virology Centre / MRC-University of Glasgow Centre for Virus Research | Wellcome Sanger Institute for the COVID-19 Genomics UK Consortium |

| EPI_Accession_ID | Originating Laboratory | Submitting Laboratory |
|---|---|---|
| EPI_ISL_459600 | NHSGGC West of Scotland Specialist Virology Centre / MRC-University of Glasgow Centre for Virus Research | Wellcome Sanger Institute for the COVID-19 Genomics UK Consortium |
| EPI_ISL_459601 | NHSGGC West of Scotland Specialist Virology Centre / MRC-University of Glasgow Centre for Virus Research | Wellcome Sanger Institute for the COVID-19 Genomics UK Consortium |
| EPI_ISL_459602 | NHSGGC West of Scotland Specialist Virology Centre / MRC-University of Glasgow Centre for Virus Research | Wellcome Sanger Institute for the COVID-19 Genomics UK Consortium |
| EPI_ISL_459603 | NHSGGC West of Scotland Specialist Virology Centre / MRC-University of Glasgow Centre for Virus Research | Wellcome Sanger Institute for the COVID-19 Genomics UK Consortium |
| EPI_ISL_459604 | NHSGGC West of Scotland Specialist Virology Centre / MRC-University of Glasgow Centre for Virus Research | Wellcome Sanger Institute for the COVID-19 Genomics UK Consortium |
| EPI_ISL_459605 | NHSGGC West of Scotland Specialist Virology Centre / MRC-University of Glasgow Centre for Virus Research | Wellcome Sanger Institute for the COVID-19 Genomics UK Consortium |
| EPI_ISL_459606 | NHSGGC West of Scotland Specialist Virology Centre / MRC-University of Glasgow Centre for Virus Research | Wellcome Sanger Institute for the COVID-19 Genomics UK Consortium |
| EPI_ISL_459607 | NHSGGC West of Scotland Specialist Virology Centre / MRC-University of Glasgow Centre for Virus Research | Wellcome Sanger Institute for the COVID-19 Genomics UK Consortium |
| EPI_ISL_459608 | NHSGGC West of Scotland Specialist Virology Centre / MRC-University of Glasgow Centre for Virus Research | Wellcome Sanger Institute for the COVID-19 Genomics UK Consortium |
| EPI_ISL_459609 | NHSGGC West of Scotland Specialist Virology Centre / MRC-University of Glasgow Centre for Virus Research | Wellcome Sanger Institute for the COVID-19 Genomics UK Consortium |
| EPI_ISL_459610 | NHSGGC West of Scotland Specialist Virology Centre / MRC-University of Glasgow Centre for Virus Research | Wellcome Sanger Institute for the COVID-19 Genomics UK Consortium |
| EPI_ISL_459611 | NHSGGC West of Scotland Specialist Virology Centre / MRC-University of Glasgow Centre for Virus Research | Wellcome Sanger Institute for the COVID-19 Genomics UK Consortium |
| EPI_ISL_459612 | NHSGGC West of Scotland Specialist Virology Centre / MRC-University of Glasgow Centre for Virus Research | Wellcome Sanger Institute for the COVID-19 Genomics UK Consortium |
| EPI_ISL_459613 | NHSGGC West of Scotland Specialist Virology Centre / MRC-University of Glasgow Centre for Virus Research | Wellcome Sanger Institute for the COVID-19 Genomics UK Consortium |
| EPI_ISL_459614 | NHSGGC West of Scotland Specialist Virology Centre / MRC-University of Glasgow Centre for Virus Research | Wellcome Sanger Institute for the COVID-19 Genomics UK Consortium |
| EPI_ISL_459615 | NHSGGC West of Scotland Specialist Virology Centre / MRC-University of Glasgow Centre for Virus Research | Wellcome Sanger Institute for the COVID-19 Genomics UK Consortium |

| EPI_Accession_ID | Originating Laboratory | Submitting Laboratory |
|---|---|---|
| EPI_ISL_459616 | NHSGGC West of Scotland Specialist Virology Centre / MRC-University of Glasgow Centre for Virus Research | Wellcome Sanger Institute for the COVID-19 Genomics UK Consortium |
| EPI_ISL_459617 | NHSGGC West of Scotland Specialist Virology Centre / MRC-University of Glasgow Centre for Virus Research | Wellcome Sanger Institute for the COVID-19 Genomics UK Consortium |
| EPI_ISL_459618 | NHSGGC West of Scotland Specialist Virology Centre / MRC-University of Glasgow Centre for Virus Research | Wellcome Sanger Institute for the COVID-19 Genomics UK Consortium |
| EPI_ISL_459619 | NHSGGC West of Scotland Specialist Virology Centre / MRC-University of Glasgow Centre for Virus Research | Wellcome Sanger Institute for the COVID-19 Genomics UK Consortium |
| EPI_ISL_459620 | NHSGGC West of Scotland Specialist Virology Centre / MRC-University of Glasgow Centre for Virus Research | Wellcome Sanger Institute for the COVID-19 Genomics UK Consortium |
| EPI_ISL_459621 | NHSGGC West of Scotland Specialist Virology Centre / MRC-University of Glasgow Centre for Virus Research | Wellcome Sanger Institute for the COVID-19 Genomics UK Consortium |
| EPI_ISL_459622 | NHSGGC West of Scotland Specialist Virology Centre / MRC-University of Glasgow Centre for Virus Research | Wellcome Sanger Institute for the COVID-19 Genomics UK Consortium |
| EPI_ISL_459623 | NHSGGC West of Scotland Specialist Virology Centre / MRC-University of Glasgow Centre for Virus Research | Wellcome Sanger Institute for the COVID-19 Genomics UK Consortium |
| EPI_ISL_459624 | NHSGGC West of Scotland Specialist Virology Centre / MRC-University of Glasgow Centre for Virus Research | Wellcome Sanger Institute for the COVID-19 Genomics UK Consortium |
| EPI_ISL_459625 | NHSGGC West of Scotland Specialist Virology Centre / MRC-University of Glasgow Centre for Virus Research | Wellcome Sanger Institute for the COVID-19 Genomics UK Consortium |
| EPI_ISL_459626 | NHSGGC West of Scotland Specialist Virology Centre / MRC-University of Glasgow Centre for Virus Research | Wellcome Sanger Institute for the COVID-19 Genomics UK Consortium |
| EPI_ISL_459627 | NHSGGC West of Scotland Specialist Virology Centre / MRC-University of Glasgow Centre for Virus Research | Wellcome Sanger Institute for the COVID-19 Genomics UK Consortium |
| EPI_ISL_459628 | NHSGGC West of Scotland Specialist Virology Centre / MRC-University of Glasgow Centre for Virus Research | Wellcome Sanger Institute for the COVID-19 Genomics UK Consortium |
| EPI_ISL_459629 | NHSGGC West of Scotland Specialist Virology Centre / MRC-University of Glasgow Centre for Virus Research | Wellcome Sanger Institute for the COVID-19 Genomics UK Consortium |
| EPI_ISL_459630 | NHSGGC West of Scotland Specialist Virology Centre / MRC-University of Glasgow Centre for Virus Research | Wellcome Sanger Institute for the COVID-19 Genomics UK Consortium |
| EPI_ISL_459631 | NHSGGC West of Scotland Specialist Virology Centre / MRC-University of Glasgow Centre for Virus Research | Wellcome Sanger Institute for the COVID-19 Genomics UK Consortium |

| EPI_Accession_ID | Originating Laboratory | Submitting Laboratory |
|---|---|---|
| EPI_ISL_459632 | NHSGGC West of Scotland Specialist Virology Centre / MRC-University of Glasgow Centre for Virus Research | Wellcome Sanger Institute for the COVID-19 Genomics UK Consortium |
| EPI_ISL_459633 | NHSGGC West of Scotland Specialist Virology Centre / MRC-University of Glasgow Centre for Virus Research | Wellcome Sanger Institute for the COVID-19 Genomics UK Consortium |
| EPI_ISL_459634 | NHSGGC West of Scotland Specialist Virology Centre / MRC-University of Glasgow Centre for Virus Research | Wellcome Sanger Institute for the COVID-19 Genomics UK Consortium |
| EPI_ISL_459635 | NHSGGC West of Scotland Specialist Virology Centre / MRC-University of Glasgow Centre for Virus Research | Wellcome Sanger Institute for the COVID-19 Genomics UK Consortium |
| EPI_ISL_459636 | NHSGGC West of Scotland Specialist Virology Centre / MRC-University of Glasgow Centre for Virus Research | Wellcome Sanger Institute for the COVID-19 Genomics UK Consortium |
| EPI_ISL_459637 | NHSGGC West of Scotland Specialist Virology Centre / MRC-University of Glasgow Centre for Virus Research | Wellcome Sanger Institute for the COVID-19 Genomics UK Consortium |
| EPI_ISL_459638 | NHSGGC West of Scotland Specialist Virology Centre / MRC-University of Glasgow Centre for Virus Research | Wellcome Sanger Institute for the COVID-19 Genomics UK Consortium |
| EPI_ISL_459639 | NHSGGC West of Scotland Specialist Virology Centre / MRC-University of Glasgow Centre for Virus Research | Wellcome Sanger Institute for the COVID-19 Genomics UK Consortium |
| EPI_ISL_459640 | NHSGGC West of Scotland Specialist Virology Centre / MRC-University of Glasgow Centre for Virus Research | Wellcome Sanger Institute for the COVID-19 Genomics UK Consortium |
| EPI_ISL_459641 | NHSGGC West of Scotland Specialist Virology Centre / MRC-University of Glasgow Centre for Virus Research | Wellcome Sanger Institute for the COVID-19 Genomics UK Consortium |
| EPI_ISL_459642 | NHSGGC West of Scotland Specialist Virology Centre / MRC-University of Glasgow Centre for Virus Research | Wellcome Sanger Institute for the COVID-19 Genomics UK Consortium |
| EPI_ISL_459643 | NHSGGC West of Scotland Specialist Virology Centre / MRC-University of Glasgow Centre for Virus Research | Wellcome Sanger Institute for the COVID-19 Genomics UK Consortium |
| EPI_ISL_459644 | NHSGGC West of Scotland Specialist Virology Centre / MRC-University of Glasgow Centre for Virus Research | Wellcome Sanger Institute for the COVID-19 Genomics UK Consortium |
| EPI_ISL_459645 | NHSGGC West of Scotland Specialist Virology Centre / MRC-University of Glasgow Centre for Virus Research | Wellcome Sanger Institute for the COVID-19 Genomics UK Consortium |
| EPI_ISL_459646 | NHSGGC West of Scotland Specialist Virology Centre / MRC-University of Glasgow Centre for Virus Research | Wellcome Sanger Institute for the COVID-19 Genomics UK Consortium |
| EPI_ISL_459647 | NHSGGC West of Scotland Specialist Virology Centre / MRC-University of Glasgow Centre for Virus Research | Wellcome Sanger Institute for the COVID-19 Genomics UK Consortium |

| EPI_Accession_ID | Originating Laboratory | Submitting Laboratory |
|---|---|---|
| EPI_ISL_459648 | NHSGGC West of Scotland Specialist Virology Centre / MRC-University of Glasgow Centre for Virus Research | Wellcome Sanger Institute for the COVID-19 Genomics UK Consortium |
| EPI_ISL_459649 | NHSGGC West of Scotland Specialist Virology Centre / MRC-University of Glasgow Centre for Virus Research | Wellcome Sanger Institute for the COVID-19 Genomics UK Consortium |
| EPI_ISL_459650 | NHSGGC West of Scotland Specialist Virology Centre / MRC-University of Glasgow Centre for Virus Research | Wellcome Sanger Institute for the COVID-19 Genomics UK Consortium |
| EPI_ISL_459651 | NHSGGC West of Scotland Specialist Virology Centre / MRC-University of Glasgow Centre for Virus Research | Wellcome Sanger Institute for the COVID-19 Genomics UK Consortium |
| EPI_ISL_459652 | NHSGGC West of Scotland Specialist Virology Centre / MRC-University of Glasgow Centre for Virus Research | Wellcome Sanger Institute for the COVID-19 Genomics UK Consortium |
| EPI_ISL_459653 | NHSGGC West of Scotland Specialist Virology Centre / MRC-University of Glasgow Centre for Virus Research | Wellcome Sanger Institute for the COVID-19 Genomics UK Consortium |
| EPI_ISL_459654 | NHSGGC West of Scotland Specialist Virology Centre / MRC-University of Glasgow Centre for Virus Research | Wellcome Sanger Institute for the COVID-19 Genomics UK Consortium |
| EPI_ISL_459655 | NHSGGC West of Scotland Specialist Virology Centre / MRC-University of Glasgow Centre for Virus Research | Wellcome Sanger Institute for the COVID-19 Genomics UK Consortium |
| EPI_ISL_459656 | NHSGGC West of Scotland Specialist Virology Centre / MRC-University of Glasgow Centre for Virus Research | Wellcome Sanger Institute for the COVID-19 Genomics UK Consortium |
| EPI_ISL_459657 | NHSGGC West of Scotland Specialist Virology Centre / MRC-University of Glasgow Centre for Virus Research | Wellcome Sanger Institute for the COVID-19 Genomics UK Consortium |
| EPI_ISL_459658 | NHSGGC West of Scotland Specialist Virology Centre / MRC-University of Glasgow Centre for Virus Research | Wellcome Sanger Institute for the COVID-19 Genomics UK Consortium |
| EPI_ISL_459659 | NHSGGC West of Scotland Specialist Virology Centre / MRC-University of Glasgow Centre for Virus Research | Wellcome Sanger Institute for the COVID-19 Genomics UK Consortium |
| EPI_ISL_459660 | NHSGGC West of Scotland Specialist Virology Centre / MRC-University of Glasgow Centre for Virus Research | Wellcome Sanger Institute for the COVID-19 Genomics UK Consortium |
| EPI_ISL_459661 | NHSGGC West of Scotland Specialist Virology Centre / MRC-University of Glasgow Centre for Virus Research | Wellcome Sanger Institute for the COVID-19 Genomics UK Consortium |
| EPI_ISL_459662 | NHSGGC West of Scotland Specialist Virology Centre / MRC-University of Glasgow Centre for Virus Research | Wellcome Sanger Institute for the COVID-19 Genomics UK Consortium |
| EPI_ISL_459663 | NHSGGC West of Scotland Specialist Virology Centre / MRC-University of Glasgow Centre for Virus Research | Wellcome Sanger Institute for the COVID-19 Genomics UK Consortium |

| EPI_Accession_ID | Originating Laboratory | Submitting Laboratory |
|---|---|---|
| EPI_ISL_459664 | NHSGGC West of Scotland Specialist Virology Centre / MRC-University of Glasgow Centre for Virus Research | Wellcome Sanger Institute for the COVID-19 Genomics UK Consortium |
| EPI_ISL_459665 | NHSGGC West of Scotland Specialist Virology Centre / MRC-University of Glasgow Centre for Virus Research | Wellcome Sanger Institute for the COVID-19 Genomics UK Consortium |
| EPI_ISL_459666 | NHSGGC West of Scotland Specialist Virology Centre / MRC-University of Glasgow Centre for Virus Research | Wellcome Sanger Institute for the COVID-19 Genomics UK Consortium |
| EPI_ISL_459667 | NHSGGC West of Scotland Specialist Virology Centre / MRC-University of Glasgow Centre for Virus Research | Wellcome Sanger Institute for the COVID-19 Genomics UK Consortium |
| EPI_ISL_459668 | NHSGGC West of Scotland Specialist Virology Centre / MRC-University of Glasgow Centre for Virus Research | Wellcome Sanger Institute for the COVID-19 Genomics UK Consortium |
| EPI_ISL_459669 | NHSGGC West of Scotland Specialist Virology Centre / MRC-University of Glasgow Centre for Virus Research | Wellcome Sanger Institute for the COVID-19 Genomics UK Consortium |
| EPI_ISL_459670 | NHSGGC West of Scotland Specialist Virology Centre / MRC-University of Glasgow Centre for Virus Research | Wellcome Sanger Institute for the COVID-19 Genomics UK Consortium |
| EPI_ISL_459671 | NHSGGC West of Scotland Specialist Virology Centre / MRC-University of Glasgow Centre for Virus Research | Wellcome Sanger Institute for the COVID-19 Genomics UK Consortium |
| EPI_ISL_459672 | NHSGGC West of Scotland Specialist Virology Centre / MRC-University of Glasgow Centre for Virus Research | Wellcome Sanger Institute for the COVID-19 Genomics UK Consortium |
| EPI_ISL_459673 | NHSGGC West of Scotland Specialist Virology Centre / MRC-University of Glasgow Centre for Virus Research | Wellcome Sanger Institute for the COVID-19 Genomics UK Consortium |
| EPI_ISL_459674 | NHSGGC West of Scotland Specialist Virology Centre / MRC-University of Glasgow Centre for Virus Research | Wellcome Sanger Institute for the COVID-19 Genomics UK Consortium |
| EPI_ISL_459675 | NHSGGC West of Scotland Specialist Virology Centre / MRC-University of Glasgow Centre for Virus Research | Wellcome Sanger Institute for the COVID-19 Genomics UK Consortium |
| EPI_ISL_459676 | NHSGGC West of Scotland Specialist Virology Centre / MRC-University of Glasgow Centre for Virus Research | Wellcome Sanger Institute for the COVID-19 Genomics UK Consortium |
| EPI_ISL_459677 | NHSGGC West of Scotland Specialist Virology Centre / MRC-University of Glasgow Centre for Virus Research | Wellcome Sanger Institute for the COVID-19 Genomics UK Consortium |
| EPI_ISL_459678 | NHSGGC West of Scotland Specialist Virology Centre / MRC-University of Glasgow Centre for Virus Research | Wellcome Sanger Institute for the COVID-19 Genomics UK Consortium |
| EPI_ISL_459679 | NHSGGC West of Scotland Specialist Virology Centre / MRC-University of Glasgow Centre for Virus Research | Wellcome Sanger Institute for the COVID-19 Genomics UK Consortium |

| EPI_Accession_ID | Originating Laboratory | Submitting Laboratory |
|---|---|---|
| EPI_ISL_459680 | NHSGGC West of Scotland Specialist Virology Centre / MRC-University of Glasgow Centre for Virus Research | Wellcome Sanger Institute for the COVID-19 Genomics UK Consortium |
| EPI_ISL_459681 | NHSGGC West of Scotland Specialist Virology Centre / MRC-University of Glasgow Centre for Virus Research | Wellcome Sanger Institute for the COVID-19 Genomics UK Consortium |
| EPI_ISL_459682 | NHSGGC West of Scotland Specialist Virology Centre / MRC-University of Glasgow Centre for Virus Research | Wellcome Sanger Institute for the COVID-19 Genomics UK Consortium |
| EPI_ISL_459683 | NHSGGC West of Scotland Specialist Virology Centre / MRC-University of Glasgow Centre for Virus Research | Wellcome Sanger Institute for the COVID-19 Genomics UK Consortium |
| EPI_ISL_459684 | NHSGGC West of Scotland Specialist Virology Centre / MRC-University of Glasgow Centre for Virus Research | Wellcome Sanger Institute for the COVID-19 Genomics UK Consortium |
| EPI_ISL_459685 | NHSGGC West of Scotland Specialist Virology Centre / MRC-University of Glasgow Centre for Virus Research | Wellcome Sanger Institute for the COVID-19 Genomics UK Consortium |
| EPI_ISL_459686 | NHSGGC West of Scotland Specialist Virology Centre / MRC-University of Glasgow Centre for Virus Research | Wellcome Sanger Institute for the COVID-19 Genomics UK Consortium |
| EPI_ISL_459687 | NHSGGC West of Scotland Specialist Virology Centre / MRC-University of Glasgow Centre for Virus Research | Wellcome Sanger Institute for the COVID-19 Genomics UK Consortium |
| EPI_ISL_459688 | NHSGGC West of Scotland Specialist Virology Centre / MRC-University of Glasgow Centre for Virus Research | Wellcome Sanger Institute for the COVID-19 Genomics UK Consortium |
| EPI_ISL_459689 | NHSGGC West of Scotland Specialist Virology Centre / MRC-University of Glasgow Centre for Virus Research | Wellcome Sanger Institute for the COVID-19 Genomics UK Consortium |
| EPI_ISL_459690 | NHSGGC West of Scotland Specialist Virology Centre / MRC-University of Glasgow Centre for Virus Research | Wellcome Sanger Institute for the COVID-19 Genomics UK Consortium |
| EPI_ISL_459691 | NHSGGC West of Scotland Specialist Virology Centre / MRC-University of Glasgow Centre for Virus Research | Wellcome Sanger Institute for the COVID-19 Genomics UK Consortium |
| EPI_ISL_459692 | NHSGGC West of Scotland Specialist Virology Centre / MRC-University of Glasgow Centre for Virus Research | Wellcome Sanger Institute for the COVID-19 Genomics UK Consortium |
| EPI_ISL_459693 | NHSGGC West of Scotland Specialist Virology Centre / MRC-University of Glasgow Centre for Virus Research | Wellcome Sanger Institute for the COVID-19 Genomics UK Consortium |
| EPI_ISL_459694 | NHSGGC West of Scotland Specialist Virology Centre / MRC-University of Glasgow Centre for Virus Research | Wellcome Sanger Institute for the COVID-19 Genomics UK Consortium |
| EPI_ISL_459695 | NHSGGC West of Scotland Specialist Virology Centre / MRC-University of Glasgow Centre for Virus Research | Wellcome Sanger Institute for the COVID-19 Genomics UK Consortium |

| EPI_Accession_ID | Originating Laboratory | Submitting Laboratory |
|---|---|---|
| EPI_ISL_459696 | NHSGGC West of Scotland Specialist Virology Centre / MRC-University of Glasgow Centre for Virus Research | Wellcome Sanger Institute for the COVID-19 Genomics UK Consortium |
| EPI_ISL_459697 | NHSGGC West of Scotland Specialist Virology Centre / MRC-University of Glasgow Centre for Virus Research | Wellcome Sanger Institute for the COVID-19 Genomics UK Consortium |
| EPI_ISL_459698 | NHSGGC West of Scotland Specialist Virology Centre / MRC-University of Glasgow Centre for Virus Research | Wellcome Sanger Institute for the COVID-19 Genomics UK Consortium |
| EPI_ISL_459699 | NHSGGC West of Scotland Specialist Virology Centre / MRC-University of Glasgow Centre for Virus Research | Wellcome Sanger Institute for the COVID-19 Genomics UK Consortium |
| EPI_ISL_459700 | NHSGGC West of Scotland Specialist Virology Centre / MRC-University of Glasgow Centre for Virus Research | Wellcome Sanger Institute for the COVID-19 Genomics UK Consortium |
| EPI_ISL_459701 | NHSGGC West of Scotland Specialist Virology Centre / MRC-University of Glasgow Centre for Virus Research | Wellcome Sanger Institute for the COVID-19 Genomics UK Consortium |
| EPI_ISL_459702 | NHSGGC West of Scotland Specialist Virology Centre / MRC-University of Glasgow Centre for Virus Research | Wellcome Sanger Institute for the COVID-19 Genomics UK Consortium |
| EPI_ISL_459703 | NHSGGC West of Scotland Specialist Virology Centre / MRC-University of Glasgow Centre for Virus Research | Wellcome Sanger Institute for the COVID-19 Genomics UK Consortium |
| EPI_ISL_459704 | NHSGGC West of Scotland Specialist Virology Centre / MRC-University of Glasgow Centre for Virus Research | Wellcome Sanger Institute for the COVID-19 Genomics UK Consortium |
| EPI_ISL_459705 | NHSGGC West of Scotland Specialist Virology Centre / MRC-University of Glasgow Centre for Virus Research | Wellcome Sanger Institute for the COVID-19 Genomics UK Consortium |
| EPI_ISL_459706 | NHSGGC West of Scotland Specialist Virology Centre / MRC-University of Glasgow Centre for Virus Research | Wellcome Sanger Institute for the COVID-19 Genomics UK Consortium |
| EPI_ISL_459707 | NHSGGC West of Scotland Specialist Virology Centre / MRC-University of Glasgow Centre for Virus Research | Wellcome Sanger Institute for the COVID-19 Genomics UK Consortium |
| EPI_ISL_459708 | NHSGGC West of Scotland Specialist Virology Centre / MRC-University of Glasgow Centre for Virus Research | Wellcome Sanger Institute for the COVID-19 Genomics UK Consortium |
| EPI_ISL_459709 | NHSGGC West of Scotland Specialist Virology Centre / MRC-University of Glasgow Centre for Virus Research | Wellcome Sanger Institute for the COVID-19 Genomics UK Consortium |
| EPI_ISL_459710 | NHSGGC West of Scotland Specialist Virology Centre / MRC-University of Glasgow Centre for Virus Research | Wellcome Sanger Institute for the COVID-19 Genomics UK Consortium |
| EPI_ISL_459711 | NHSGGC West of Scotland Specialist Virology Centre / MRC-University of Glasgow Centre for Virus Research | Wellcome Sanger Institute for the COVID-19 Genomics UK Consortium |

| EPI_Accession_ID | Originating Laboratory | Submitting Laboratory |
|---|---|---|
| EPI_ISL_459712 | NHSGGC West of Scotland Specialist Virology Centre / MRC-University of Glasgow Centre for Virus Research | Wellcome Sanger Institute for the COVID-19 Genomics UK Consortium |
| EPI_ISL_459713 | NHSGGC West of Scotland Specialist Virology Centre / MRC-University of Glasgow Centre for Virus Research | Wellcome Sanger Institute for the COVID-19 Genomics UK Consortium |
| EPI_ISL_459714 | NHSGGC West of Scotland Specialist Virology Centre / MRC-University of Glasgow Centre for Virus Research | Wellcome Sanger Institute for the COVID-19 Genomics UK Consortium |
| EPI_ISL_459715 | NHSGGC West of Scotland Specialist Virology Centre / MRC-University of Glasgow Centre for Virus Research | Wellcome Sanger Institute for the COVID-19 Genomics UK Consortium |
| EPI_ISL_459716 | NHSGGC West of Scotland Specialist Virology Centre / MRC-University of Glasgow Centre for Virus Research | Wellcome Sanger Institute for the COVID-19 Genomics UK Consortium |
| EPI_ISL_459717 | NHSGGC West of Scotland Specialist Virology Centre / MRC-University of Glasgow Centre for Virus Research | Wellcome Sanger Institute for the COVID-19 Genomics UK Consortium |
| EPI_ISL_459718 | NHSGGC West of Scotland Specialist Virology Centre / MRC-University of Glasgow Centre for Virus Research | Wellcome Sanger Institute for the COVID-19 Genomics UK Consortium |
| EPI_ISL_459719 | NHSGGC West of Scotland Specialist Virology Centre / MRC-University of Glasgow Centre for Virus Research | Wellcome Sanger Institute for the COVID-19 Genomics UK Consortium |
| EPI_ISL_459720 | NHSGGC West of Scotland Specialist Virology Centre / MRC-University of Glasgow Centre for Virus Research | Wellcome Sanger Institute for the COVID-19 Genomics UK Consortium |
| EPI_ISL_459721 | NHSGGC West of Scotland Specialist Virology Centre / MRC-University of Glasgow Centre for Virus Research | Wellcome Sanger Institute for the COVID-19 Genomics UK Consortium |
| EPI_ISL_459722 | NHSGGC West of Scotland Specialist Virology Centre / MRC-University of Glasgow Centre for Virus Research | Wellcome Sanger Institute for the COVID-19 Genomics UK Consortium |
| EPI_ISL_459723 | NHSGGC West of Scotland Specialist Virology Centre / MRC-University of Glasgow Centre for Virus Research | Wellcome Sanger Institute for the COVID-19 Genomics UK Consortium |
| EPI_ISL_459724 | NHSGGC West of Scotland Specialist Virology Centre / MRC-University of Glasgow Centre for Virus Research | Wellcome Sanger Institute for the COVID-19 Genomics UK Consortium |
| EPI_ISL_459725 | PHE South West Regional Laboratory, National Infection Service | Wellcome Sanger Institute for the COVID-19 Genomics UK Consortium |
| EPI_ISL_459726 | PHE South West Regional Laboratory, National Infection Service | Wellcome Sanger Institute for the COVID-19 Genomics UK Consortium |
| EPI_ISL_459727 | PHE South West Regional Laboratory, National Infection Service | Wellcome Sanger Institute for the COVID-19 Genomics UK Consortium |

| EPI_Accession_ID | Originating Laboratory | Submitting Laboratory |
|---|---|---|
| EPI_ISL_459856 | Center for Genome Regulation (CRG) | Center for Mathematical Modeling and Center for Genome Regulation. Santiago, Chile |
| EPI_ISL_459857 | Center for Genome Regulation (CRG) | Center for Mathematical Modeling and Center for Genome Regulation. Santiago, Chile |
| EPI_ISL_459858 | Center for Genome Regulation (CRG) | Center for Mathematical Modeling and Center for Genome Regulation. Santiago, Chile |
| EPI_ISL_459859 | Center for Genome Regulation (CRG) | Center for Mathematical Modeling and Center for Genome Regulation. Santiago, Chile |
| EPI_ISL_459860 | Center for Genome Regulation (CRG) | Center for Mathematical Modeling and Center for Genome Regulation. Santiago, Chile |
| EPI_ISL_459861 | Center for Genome Regulation (CRG) | Center for Mathematical Modeling and Center for Genome Regulation. Santiago, Chile |
| EPI_ISL_459862 | Center for Genome Regulation (CRG) | Center for Mathematical Modeling and Center for Genome Regulation. Santiago, Chile |
| EPI_ISL_459863 | Center for Genome Regulation (CRG) | Center for Mathematical Modeling and Center for Genome Regulation. Santiago, Chile |
| EPI_ISL_459864 | Center for Genome Regulation (CRG) | Center for Mathematical Modeling and Center for Genome Regulation. Santiago, Chile |
| EPI_ISL_459866 | Kingston Health Sciences Center | Queen's Genomics Lab at Ongwanada (Q-GLO) |
| EPI_ISL_459867 | Kingston Health Sciences Center | Queen's Genomics Lab at Ongwanada (Q-GLO) |
| EPI_ISL_459868 | Kingston Health Sciences Center | Queen's Genomics Lab at Ongwanada (Q-GLO) |
| EPI_ISL_459869 | Kingston Health Sciences Center | Queen's Genomics Lab at Ongwanada (Q-GLO) |
| EPI_ISL_459871 | Kingston Health Sciences Center | Queen's Genomics Lab at Ongwanada (Q-GLO) |
| EPI_ISL_459872 | Kingston Health Sciences Center | Queen's Genomics Lab at Ongwanada (Q-GLO) |
| EPI_ISL_459873 | Kingston Health Sciences Center | Queen's Genomics Lab at Ongwanada (Q-GLO) |
| EPI_ISL_459874 | Kingston Health Sciences Center | Queen's Genomics Lab at Ongwanada (Q-GLO) |
| EPI_ISL_459875 | Kingston Health Sciences Center | Queen's Genomics Lab at Ongwanada (Q-GLO) |
| EPI_ISL_459879 | Kingston Health Sciences Center | Queen's Genomics Lab at Ongwanada (Q-GLO) |
| EPI_ISL_459880 | Kingston Health Sciences Center | Queen's Genomics Lab at Ongwanada (Q-GLO) |
| EPI_ISL_459881 | Kingston Health Sciences Center | Queen's Genomics Lab at Ongwanada (Q-GLO) |
| EPI_ISL_459882 | Kingston Health Sciences Center | Queen's Genomics Lab at Ongwanada (Q-GLO) |
| EPI_ISL_459883 | Kingston Health Sciences Center | Queen's Genomics Lab at Ongwanada (Q-GLO) |
| EPI_ISL_459884 | Kingston Health Sciences Center | Queen's Genomics Lab at Ongwanada (Q-GLO) |

| EPI_Accession_ID | Originating Laboratory | Submitting Laboratory |
|---|---|---|
| EPI_ISL_459885 | Kingston Health Sciences Center | Queen's Genomics Lab at Ongwanada (Q-GLO) |
| EPI_ISL_459886 | Kingston Health Sciences Center | Queen's Genomics Lab at Ongwanada (Q-GLO) |
| EPI_ISL_459887 | Kingston Health Sciences Center | Queen's Genomics Lab at Ongwanada (Q-GLO) |
| EPI_ISL_459888 | Kingston Health Sciences Center | Queen's Genomics Lab at Ongwanada (Q-GLO) |
| EPI_ISL_459889 | Kingston Health Sciences Center | Queen's Genomics Lab at Ongwanada (Q-GLO) |
| EPI_ISL_459890 | Kingston Health Sciences Center | Queen's Genomics Lab at Ongwanada (Q-GLO) |
| EPI_ISL_459891 | Kingston Health Sciences Center | Queen's Genomics Lab at Ongwanada (Q-GLO) |
| EPI_ISL_459892 | Kingston Health Sciences Center | Queen's Genomics Lab at Ongwanada (Q-GLO) |
| EPI_ISL_459893 | Laboratoire National de Sante, Microbiology, Virology | Laboratoire National de Sante, Microbiology, Epidemiology and Microbial Genomics |
| EPI_ISL_459894 | Laboratoire National de Sante, Microbiology, Virology | Laboratoire National de Sante, Microbiology, Epidemiology and Microbial Genomics |
| EPI_ISL_459895 | Laboratoire National de Sante, Microbiology, Virology | Laboratoire National de Sante, Microbiology, Epidemiology and Microbial Genomics |
| EPI_ISL_459896 | Laboratoire National de Sante, Microbiology, Virology | Laboratoire National de Sante, Microbiology, Epidemiology and Microbial Genomics |
| EPI_ISL_459897 | Laboratoire National de Sante, Microbiology, Virology | Laboratoire National de Sante, Microbiology, Epidemiology and Microbial Genomics |
| EPI_ISL_459898 | Laboratoire National de Sante, Microbiology, Virology | Laboratoire National de Sante, Microbiology, Epidemiology and Microbial Genomics |
| EPI_ISL_459899 | Laboratoire National de Sante, Microbiology, Virology | Laboratoire National de Sante, Microbiology, Epidemiology and Microbial Genomics |
| EPI_ISL_459900 | Laboratoire National de Sante, Microbiology, Virology | Laboratoire National de Sante, Microbiology, Epidemiology and Microbial Genomics |
| EPI_ISL_459901 | Laboratoire National de Sante, Microbiology, Virology | Laboratoire National de Sante, Microbiology, Epidemiology and Microbial Genomics |
| EPI_ISL_459902 | Laboratoire National de Sante, Microbiology, Virology | Laboratoire National de Sante, Microbiology, Epidemiology and Microbial Genomics |
| EPI_ISL_459903 | Laboratoire National de Sante, Microbiology, Virology | Laboratoire National de Sante, Microbiology, Epidemiology and Microbial Genomics |
| EPI_ISL_459904 | Laboratoire National de Sante, Microbiology, Virology | Laboratoire National de Sante, Microbiology, Epidemiology and Microbial Genomics |

| EPI_Accession_ID | Originating Laboratory | Submitting Laboratory |
|---|---|---|
| EPI_ISL_459905 | Laboratoire National de Sante, Microbiology, Virology | Laboratoire National de Sante, Microbiology, Epidemiology and Microbial Genomics |
| EPI_ISL_459906 | Laboratoire National de Sante, Microbiology, Virology | Laboratoire National de Sante, Microbiology, Epidemiology and Microbial Genomics |
| EPI_ISL_459909 | Zoonotic and Exotic infection Diseases Division, Harbin Veterinary Research Institute, CAAS | Zoonotic and Exotic infection Diseases Division, Harbin Veterinary Research Institute, CAAS |
| EPI_ISL_459910 | Zoonotic and Exotic infection Diseases Division, Harbin Veterinary Research Institute, CAAS | Zoonotic and Exotic infection Diseases Division, Harbin Veterinary Resrarch Institute, CAAS |
| EPI_ISL_459911 | Devki Devi Foundation, a unit of Max Healthcare | CSIR-IGIB/Max |
| EPI_ISL_459912 | Devki Devi Foundation, a unit of Max Healthcare | CSIR-IGIB/Max |
| EPI_ISL_459913 | Devki Devi Foundation, a unit of Max Healthcare | CSIR-IGIB/Max |
| EPI_ISL_459914 | Devki Devi Foundation, a unit of Max Healthcare | CSIR-IGIB/Max |
| EPI_ISL_459915 | Devki Devi Foundation, a unit of Max Healthcare | CSIR-IGIB/Max |
| EPI_ISL_459916 | Devki Devi Foundation, a unit of Max Healthcare | CSIR-IGIB/Max |
| EPI_ISL_459917 | Devki Devi Foundation, a unit of Max Healthcare | CSIR-IGIB/Max |
| EPI_ISL_459918 | Devki Devi Foundation, a unit of Max Healthcare | CSIR-IGIB/Max |
| EPI_ISL_459919 | Devki Devi Foundation, a unit of Max Healthcare | CSIR-IGIB/Max |
| EPI_ISL_459920 | Devki Devi Foundation, a unit of Max Healthcare | CSIR-IGIB/Max |
| EPI_ISL_459921 | Devki Devi Foundation, a unit of Max Healthcare | CSIR-IGIB/Max |
| EPI_ISL_459922 | Devki Devi Foundation, a unit of Max Healthcare | CSIR-IGIB/Max |
| EPI_ISL_459923 | Devki Devi Foundation, a unit of Max Healthcare | CSIR-IGIB/Max |
| EPI_ISL_459924 | Devki Devi Foundation, a unit of Max Healthcare | CSIR-IGIB/Max |
| EPI_ISL_459925 | Devki Devi Foundation, a unit of Max Healthcare | CSIR-IGIB/Max |
| EPI_ISL_459931 | Devki Devi Foundation, a unit of Max Healthcare | CSIR-IGIB/Max |
| EPI_ISL_459932 | Devki Devi Foundation, a unit of Max Healthcare | CSIR-IGIB/Max |
| EPI_ISL_459933 | Devki Devi Foundation, a unit of Max Healthcare | CSIR-IGIB/Max |
| EPI_ISL_459934 | Devki Devi Foundation, a unit of Max Healthcare | CSIR-IGIB/Max |
| EPI_ISL_459935 | Devki Devi Foundation, a unit of Max Healthcare | CSIR-IGIB/Max |
| EPI_ISL_459936 | Devki Devi Foundation, a unit of Max Healthcare | CSIR-IGIB/Max |
| EPI_ISL_459937 | Devki Devi Foundation, a unit of Max Healthcare | CSIR-IGIB/Max |
| EPI_ISL_459938 | Devki Devi Foundation, a unit of Max Healthcare | CSIR-IGIB/Max |
| EPI_ISL_459939 | Devki Devi Foundation, a unit of Max Healthcare | CSIR-IGIB/Max |
| EPI_ISL_459940 | Devki Devi Foundation, a unit of Max Healthcare | CSIR-IGIB/Max |

| EPI_Accession_ID | Originating Laboratory | Submitting Laboratory |
|---|---|---|
| EPI_ISL_459941 | Devki Devi Foundation, a unit of Max Healthcare | CSIR-IGIB/Max |
| EPI_ISL_459942 | Devki Devi Foundation, a unit of Max Healthcare | CSIR-IGIB/Max |
| EPI_ISL_459943 | Devki Devi Foundation, a unit of Max Healthcare | CSIR-IGIB/Max |
| EPI_ISL_459944 | Devki Devi Foundation, a unit of Max Healthcare | CSIR-IGIB/Max |
| EPI_ISL_459945 | Devki Devi Foundation, a unit of Max Healthcare | CSIR-IGIB/Max |
| EPI_ISL_459946 | Devki Devi Foundation, a unit of Max Healthcare | CSIR-IGIB/Max |
| EPI_ISL_459947 | Devki Devi Foundation, a unit of Max Healthcare | CSIR-IGIB/Max |
| EPI_ISL_459948 | Devki Devi Foundation, a unit of Max Healthcare | CSIR-IGIB/Max |
| EPI_ISL_459949 | Devki Devi Foundation, a unit of Max Healthcare | CSIR-IGIB/Max |
| EPI_ISL_459950 | Devki Devi Foundation, a unit of Max Healthcare | CSIR-IGIB/Max |
| EPI_ISL_459951 | Devki Devi Foundation, a unit of Max Healthcare | CSIR-IGIB/Max |
| EPI_ISL_459952 | Devki Devi Foundation, a unit of Max Healthcare | CSIR-IGIB/Max |
| EPI_ISL_459953 | Institute for Medical Research, Infectious Disease Research Centre, National Institutes of Health, Ministry of Health Malaysia | Institute for Medical Research Infectious Disease Research Centre, National Institutes of Health, Ministry of Health Malaysia |
| EPI_ISL_459954 | Institute for Medical Research, Infectious Disease Research Centre, National Institutes of Health, Ministry of Health Malaysia | Institute for Medical Research, Infectious Disease Research Centre, National Institutes of Health, Ministry of Health Malaysia |
| EPI_ISL_459955 | Institute for Medical Research, Infectious Disease Research Centre, National Institutes of Health, Ministry of Health Malaysia | Institute for Medical Research, Infectious Disease Research Centre, National Institutes of Health, Ministry of Health Malaysia |
| EPI_ISL_459956 | Institute for Medical Research, Infectious Disease Research Centre, National Institutes of Health, Ministry of Health Malaysia | Institute for Medical Research, Infectious Disease Research Centre, National Institutes of Health, Ministry of Health Malaysia |
| EPI_ISL_459957 | Institute for Medical Research, Infectious Disease Research Centre, National Institutes of Health, Minis | Institute for Medical Research, Infectious Disease Research Centre, National Institutes of Health, Minis |
| EPI_ISL_459958 | Respiratory Virus Unit, Microbiology Services Colindale, Public Health England | Respiratory Virus Unit, Microbiology Services Colindale, Public Health England |
| EPI_ISL_459959 | Respiratory Virus Unit, Microbiology Services Colindale, Public Health England | Respiratory Virus Unit, Microbiology Services Colindale, Public Health England |
| EPI_ISL_459960 | Respiratory Virus Unit, Microbiology Services Colindale, Public Health England | Respiratory Virus Unit, Microbiology Services Colindale, Public Health England |

| EPI_Accession_ID | Originating Laboratory | Submitting Laboratory |
|---|---|---|
| EPI_ISL_459961 | Respiratory Virus Unit, Microbiology Services Colindale, Public Health England | Respiratory Virus Unit, Microbiology Services Colindale, Public Health England |
| EPI_ISL_459962 | Centogene AG | Centogene AG |
| EPI_ISL_459963 | Centogene AG | Centogene AG |
| EPI_ISL_459964 | Centogene AG | Centogene AG |
| EPI_ISL_459965 | Institut Pasteur du Maroc | Institut Pasteur du Maroc |
| EPI_ISL_459966 | Institut Pasteur du Maroc | Institut Pasteur du Maroc |
| EPI_ISL_459967 | Institut Pasteur du Maroc | Institut Pasteur du Maroc |
| EPI_ISL_459968 | Institut Pasteur du Maroc | Institut Pasteur du Maroc |
| EPI_ISL_459969 | Institut Pasteur du Maroc | Institut Pasteur du Maroc |
| EPI_ISL_459970 | Institut Pasteur du Maroc | Institut Pasteur du Maroc |
| EPI_ISL_459971 | Institut Pasteur du Maroc | Institut Pasteur du Maroc |
| EPI_ISL_459972 | Institut Pasteur du Maroc | Institut Pasteur du Maroc |
| EPI_ISL_459973 | Institut Pasteur du Maroc | Institut Pasteur du Maroc |
| EPI_ISL_459974 | Institut Pasteur du Maroc | Institut Pasteur du Maroc |
| EPI_ISL_459975 | Institut Pasteur du Maroc | Institut Pasteur du Maroc |
| EPI_ISL_459976 | Institut Pasteur du Maroc | Institut Pasteur du Maroc |
| EPI_ISL_459977 | Institut Pasteur du Maroc | Institut Pasteur du Maroc |
| EPI_ISL_459978 | Institut Pasteur du Maroc | Institut Pasteur du Maroc |
| EPI_ISL_459979 | Institut Pasteur du Maroc | Institut Pasteur du Maroc |
| EPI_ISL_459980 | Institut Pasteur du Maroc | Institut Pasteur du Maroc |
| EPI_ISL_459981 | Institut Pasteur du Maroc | Institut Pasteur du Maroc |
| EPI_ISL_459982 | Institut Pasteur du Maroc | Institut Pasteur du Maroc |
| EPI_ISL_459983 | Institut Pasteur du Maroc | Institut Pasteur du Maroc |
| EPI_ISL_459984 | Institut Pasteur du Maroc | Institut Pasteur du Maroc |
| EPI_ISL_459992 | Michigan Department of Health and Human Services, Bureau of Laboratories | Michigan Department of Health and Human Services, Bureau of Laboratories |
| EPI_ISL_459993 | Michigan Department of Health and Human Services, Bureau of Laboratories | Michigan Department of Health and Human Services, Bureau of Laboratories |
| EPI_ISL_459994 | Michigan Department of Health and Human Services, Bureau of Laboratories | Michigan Department of Health and Human Services, Bureau of Laboratories |
| EPI_ISL_459995 | Michigan Department of Health and Human Services, Bureau of Laboratories | Michigan Department of Health and Human Services, Bureau of Laboratories |

| EPI_Accession_ID | Originating Laboratory | Submitting Laboratory |
|---|---|---|
| EPI_ISL_459996 | Michigan Department of Health and Human Services, Bureau of Laboratories | Michigan Department of Health and Human Services, Bureau of Laboratories |
| EPI_ISL_459997 | Michigan Department of Health and Human Services, Bureau of Laboratories | Michigan Department of Health and Human Services, Bureau of Laboratories |
| EPI_ISL_459998 | Michigan Department of Health and Human Services, Bureau of Laboratories | Michigan Department of Health and Human Services, Bureau of Laboratories |
| EPI_ISL_459999 | Michigan Department of Health and Human Services, Bureau of Laboratories | Michigan Department of Health and Human Services, Bureau of Laboratories |
| EPI_ISL_460000 | Michigan Department of Health and Human Services, Bureau of Laboratories | Michigan Department of Health and Human Services, Bureau of Laboratories |
| EPI_ISL_460001 | Michigan Department of Health and Human Services, Bureau of Laboratories | Michigan Department of Health and Human Services, Bureau of Laboratories |
| EPI_ISL_460002 | Michigan Department of Health and Human Services, Bureau of Laboratories | Michigan Department of Health and Human Services, Bureau of Laboratories |
| EPI_ISL_460003 | Michigan Department of Health and Human Services, Bureau of Laboratories | Michigan Department of Health and Human Services, Bureau of Laboratories |
| EPI_ISL_460004 | Michigan Department of Health and Human Services, Bureau of Laboratories | Michigan Department of Health and Human Services, Bureau of Laboratories |
| EPI_ISL_460005 | Michigan Department of Health and Human Services, Bureau of Laboratories | Michigan Department of Health and Human Services, Bureau of Laboratories |
| EPI_ISL_460006 | Michigan Department of Health and Human Services, Bureau of Laboratories | Michigan Department of Health and Human Services, Bureau of Laboratories |
| EPI_ISL_460007 | Michigan Department of Health and Human Services, Bureau of Laboratories | Michigan Department of Health and Human Services, Bureau of Laboratories |
| EPI_ISL_460008 | Michigan Department of Health and Human Services, Bureau of Laboratories | Michigan Department of Health and Human Services, Bureau of Laboratories |
| EPI_ISL_460009 | Michigan Department of Health and Human Services, Bureau of Laboratories | Michigan Department of Health and Human Services, Bureau of Laboratories |
| EPI_ISL_460010 | Michigan Department of Health and Human Services, Bureau of Laboratories | Michigan Department of Health and Human Services, Bureau of Laboratories |
| EPI_ISL_460011 | Michigan Department of Health and Human Services, Bureau of Laboratories | Michigan Department of Health and Human Services, Bureau of Laboratories |

| EPI_Accession_ID | Originating Laboratory | Submitting Laboratory |
|---|---|---|
| EPI_ISL_460012 | Michigan Department of Health and Human Services, Bureau of Laboratories | Michigan Department of Health and Human Services, Bureau of Laboratories |
| EPI_ISL_460013 | Michigan Department of Health and Human Services, Bureau of Laboratories | Michigan Department of Health and Human Services, Bureau of Laboratories |
| EPI_ISL_460014 | Michigan Department of Health and Human Services, Bureau of Laboratories | Michigan Department of Health and Human Services, Bureau of Laboratories |
| EPI_ISL_460015 | Michigan Department of Health and Human Services, Bureau of Laboratories | Michigan Department of Health and Human Services, Bureau of Laboratories |
| EPI_ISL_460016 | Michigan Department of Health and Human Services, Bureau of Laboratories | Michigan Department of Health and Human Services, Bureau of Laboratories |
| EPI_ISL_460017 | Michigan Department of Health and Human Services, Bureau of Laboratories | Michigan Department of Health and Human Services, Bureau of Laboratories |
| EPI_ISL_460018 | Michigan Department of Health and Human Services, Bureau of Laboratories | Michigan Department of Health and Human Services, Bureau of Laboratories |
| EPI_ISL_460019 | Michigan Department of Health and Human Services, Bureau of Laboratories | Michigan Department of Health and Human Services, Bureau of Laboratories |
| EPI_ISL_460020 | Michigan Department of Health and Human Services, Bureau of Laboratories | Michigan Department of Health and Human Services, Bureau of Laboratories |
| EPI_ISL_460021 | Michigan Department of Health and Human Services, Bureau of Laboratories | Michigan Department of Health and Human Services, Bureau of Laboratories |
| EPI_ISL_460022 | Michigan Department of Health and Human Services, Bureau of Laboratories | Michigan Department of Health and Human Services, Bureau of Laboratories |
| EPI_ISL_460023 | Michigan Department of Health and Human Services, Bureau of Laboratories | Michigan Department of Health and Human Services, Bureau of Laboratories |
| EPI_ISL_460024 | Michigan Department of Health and Human Services, Bureau of Laboratories | Michigan Department of Health and Human Services, Bureau of Laboratories |
| EPI_ISL_460025 | Michigan Department of Health and Human Services, Bureau of Laboratories | Michigan Department of Health and Human Services, Bureau of Laboratories |
| EPI_ISL_460026 | Michigan Department of Health and Human Services, Bureau of Laboratories | Michigan Department of Health and Human Services, Bureau of Laboratories |
| EPI_ISL_460027 | Michigan Department of Health and Human Services, Bureau of Laboratories | Michigan Department of Health and Human Services, Bureau of Laboratories |

| EPI_Accession_ID | Originating Laboratory | Submitting Laboratory |
|---|---|---|
| EPI_ISL_460028 | Michigan Department of Health and Human Services, Bureau of Laboratories | Michigan Department of Health and Human Services, Bureau of Laboratories |
| EPI_ISL_460029 | Michigan Department of Health and Human Services, Bureau of Laboratories | Michigan Department of Health and Human Services, Bureau of Laboratories |
| EPI_ISL_460030 | Michigan Department of Health and Human Services, Bureau of Laboratories | Michigan Department of Health and Human Services, Bureau of Laboratories |
| EPI_ISL_460031 | Michigan Department of Health and Human Services, Bureau of Laboratories | Michigan Department of Health and Human Services, Bureau of Laboratories |
| EPI_ISL_460032 | Michigan Department of Health and Human Services, Bureau of Laboratories | Michigan Department of Health and Human Services, Bureau of Laboratories |
| EPI_ISL_460033 | Michigan Department of Health and Human Services, Bureau of Laboratories | Michigan Department of Health and Human Services, Bureau of Laboratories |
| EPI_ISL_460034 | Michigan Department of Health and Human Services, Bureau of Laboratories | Michigan Department of Health and Human Services, Bureau of Laboratories |
| EPI_ISL_460035 | Michigan Department of Health and Human Services, Bureau of Laboratories | Michigan Department of Health and Human Services, Bureau of Laboratories |
| EPI_ISL_460036 | Michigan Department of Health and Human Services, Bureau of Laboratories | Michigan Department of Health and Human Services, Bureau of Laboratories |
| EPI_ISL_460037 | Michigan Department of Health and Human Services, Bureau of Laboratories | Michigan Department of Health and Human Services, Bureau of Laboratories |
| EPI_ISL_460038 | Michigan Department of Health and Human Services, Bureau of Laboratories | Michigan Department of Health and Human Services, Bureau of Laboratories |
| EPI_ISL_460039 | Michigan Department of Health and Human Services, Bureau of Laboratories | Michigan Department of Health and Human Services, Bureau of Laboratories |
| EPI_ISL_460040 | Michigan Department of Health and Human Services, Bureau of Laboratories | Michigan Department of Health and Human Services, Bureau of Laboratories |
| EPI_ISL_460041 | Michigan Department of Health and Human Services, Bureau of Laboratories | Michigan Department of Health and Human Services, Bureau of Laboratories |
| EPI_ISL_460042 | Michigan Department of Health and Human Services, Bureau of Laboratories | Michigan Department of Health and Human Services, Bureau of Laboratories |
| EPI_ISL_460043 | Michigan Department of Health and Human Services, Bureau of Laboratories | Michigan Department of Health and Human Services, Bureau of Laboratories |

| EPI_Accession_ID | Originating Laboratory | Submitting Laboratory |
|---|---|---|
| EPI_ISL_460045 | Minnesota Department of Health, Public Health Laboratory | Minnesota Department of Health, Public Health Laboratory |
| EPI_ISL_460046 | Minnesota Department of Health, Public Health Laboratory | Minnesota Department of Health, Public Health Laboratory |
| EPI_ISL_460047 | Minnesota Department of Health, Public Health Laboratory | Minnesota Department of Health, Public Health Laboratory |
| EPI_ISL_460048 | Minnesota Department of Health, Public Health Laboratory | Minnesota Department of Health, Public Health Laboratory |
| EPI_ISL_460049 | Minnesota Department of Health, Public Health Laboratory | Minnesota Department of Health, Public Health Laboratory |
| EPI_ISL_460050 | Minnesota Department of Health, Public Health Laboratory | Minnesota Department of Health, Public Health Laboratory |
| EPI_ISL_460051 | Minnesota Department of Health, Public Health Laboratory | Minnesota Department of Health, Public Health Laboratory |
| EPI_ISL_460052 | Minnesota Department of Health, Public Health Laboratory | Minnesota Department of Health, Public Health Laboratory |
| EPI_ISL_460053 | Minnesota Department of Health, Public Health Laboratory | Minnesota Department of Health, Public Health Laboratory |
| EPI_ISL_460054 | Minnesota Department of Health, Public Health Laboratory | Minnesota Department of Health, Public Health Laboratory |
| EPI_ISL_460055 | Minnesota Department of Health, Public Health Laboratory | Minnesota Department of Health, Public Health Laboratory |
| EPI_ISL_460056 | Minnesota Department of Health, Public Health Laboratory | Minnesota Department of Health, Public Health Laboratory |
| EPI_ISL_460057 | Minnesota Department of Health, Public Health Laboratory | Minnesota Department of Health, Public Health Laboratory |
| EPI_ISL_460058 | Minnesota Department of Health, Public Health Laboratory | Minnesota Department of Health, Public Health Laboratory |
| EPI_ISL_460059 | Minnesota Department of Health, Public Health Laboratory | Minnesota Department of Health, Public Health Laboratory |
| EPI_ISL_460060 | Minnesota Department of Health, Public Health Laboratory | Minnesota Department of Health, Public Health Laboratory |

| EPI_Accession_ID | Originating Laboratory | Submitting Laboratory |
| --- | --- | --- |
| EPI_ISL_460061 | Minnesota Department of Health, Public Health Laboratory | Minnesota Department of Health, Public Health Laboratory |
| EPI_ISL_460062 | Minnesota Department of Health, Public Health Laboratory | Minnesota Department of Health, Public Health Laboratory |
| EPI_ISL_460063 | Minnesota Department of Health, Public Health Laboratory | Minnesota Department of Health, Public Health Laboratory |
| EPI_ISL_460064 | Minnesota Department of Health, Public Health Laboratory | Minnesota Department of Health, Public Health Laboratory |
| EPI_ISL_460065 | Minnesota Department of Health, Public Health Laboratory | Minnesota Department of Health, Public Health Laboratory |
| EPI_ISL_460066 | Minnesota Department of Health, Public Health Laboratory | Minnesota Department of Health, Public Health Laboratory |
| EPI_ISL_460067 | Minnesota Department of Health, Public Health Laboratory | Minnesota Department of Health, Public Health Laboratory |
| EPI_ISL_460068 | Minnesota Department of Health, Public Health Laboratory | Minnesota Department of Health, Public Health Laboratory |
| EPI_ISL_460069 | Minnesota Department of Health, Public Health Laboratory | Minnesota Department of Health, Public Health Laboratory |
| EPI_ISL_460070 | Minnesota Department of Health, Public Health Laboratory | Minnesota Department of Health, Public Health Laboratory |
| EPI_ISL_460071 | Minnesota Department of Health, Public Health Laboratory | Minnesota Department of Health, Public Health Laboratory |
| EPI_ISL_460072 | Minnesota Department of Health, Public Health Laboratory | Minnesota Department of Health, Public Health Laboratory |
| EPI_ISL_460073 | Minnesota Department of Health, Public Health Laboratory | Minnesota Department of Health, Public Health Laboratory |
| EPI_ISL_460074 | Minnesota Department of Health, Public Health Laboratory | Minnesota Department of Health, Public Health Laboratory |
| EPI_ISL_460075 | Minnesota Department of Health, Public Health Laboratory | Minnesota Department of Health, Public Health Laboratory |
| EPI_ISL_460076 | Minnesota Department of Health, Public Health Laboratory | Minnesota Department of Health, Public Health Laboratory |

| EPI_Accession_ID | Originating Laboratory | Submitting Laboratory |
|---|---|---|
| EPI_ISL_460077 | Minnesota Department of Health, Public Health Laboratory | Minnesota Department of Health, Public Health Laboratory |
| EPI_ISL_460078 | Minnesota Department of Health, Public Health Laboratory | Minnesota Department of Health, Public Health Laboratory |
| EPI_ISL_460079 | Molecular Virology Unit, Fondazione IRCCS Policlinico San Matteo , Pavia | Laboratory of Virology, INMI Lazzaro Spallanzani IRCCS |
| EPI_ISL_460080 | Molecular Virology Unit, Fondazione IRCCS Policlinico San Matteo , Pavia | Laboratory of Virology, INMI Lazzaro Spallanzani IRCCS |
| EPI_ISL_460081 | Molecular Virology Unit, Fondazione IRCCS Policlinico San Matteo , Pavia | Laboratory of Virology, INMI Lazzaro Spallanzani IRCCS |
| EPI_ISL_460082 | Molecular Virology Unit, Fondazione IRCCS Policlinico San Matteo , Pavia | Laboratory of Virology, INMI Lazzaro Spallanzani IRCCS |
| EPI_ISL_460083 | Molecular Virology Unit, Fondazione IRCCS Policlinico San Matteo , Pavia | Laboratory of Virology, INMI Lazzaro Spallanzani IRCCS |
| EPI_ISL_460084 | Molecular Virology Unit, Fondazione IRCCS Policlinico San Matteo , Pavia | Laboratory of Virology, INMI Lazzaro Spallanzani IRCCS |
| EPI_ISL_460085 | Molecular Virology Unit, Fondazione IRCCS Policlinico San Matteo , Pavia | Laboratory of Virology, INMI Lazzaro Spallanzani IRCCS |
| EPI_ISL_460086 | Molecular Virology Unit, Fondazione IRCCS Policlinico San Matteo , Pavia | Laboratory of Virology, INMI Lazzaro Spallanzani IRCCS |
| EPI_ISL_460087 | Molecular Virology Unit, Fondazione IRCCS Policlinico San Matteo , Pavia | Laboratory of Virology, INMI Lazzaro Spallanzani IRCCS |
| EPI_ISL_460088 | Molecular Virology Unit, Fondazione IRCCS Policlinico San Matteo , Pavia | Laboratory of Virology, INMI Lazzaro Spallanzani IRCCS |
| EPI_ISL_460089 | Molecular Virology Unit, Fondazione IRCCS Policlinico San Matteo , Pavia | Laboratory of Virology, INMI Lazzaro Spallanzani IRCCS |
| EPI_ISL_460090 | Molecular Virology Unit, Fondazione IRCCS Policlinico San Matteo , Pavia | Laboratory of Virology, INMI Lazzaro Spallanzani IRCCS |
| EPI_ISL_460091 | Molecular Virology Unit, Fondazione IRCCS Policlinico San Matteo , Pavia | Laboratory of Virology, INMI Lazzaro Spallanzani IRCCS |
| EPI_ISL_460092 | Molecular Virology Unit, Fondazione IRCCS Policlinico San Matteo , Pavia | Laboratory of Virology, INMI Lazzaro Spallanzani IRCCS |

| EPI_Accession_ID | Originating Laboratory | Submitting Laboratory |
|---|---|---|
| EPI_ISL_460093 | Molecular Virology Unit, Fondazione IRCCS Policlinico San Matteo , Pavia | Laboratory of Virology, INMI Lazzaro Spallanzani IRCCS |
| EPI_ISL_460094 | Molecular Virology Unit, Fondazione IRCCS Policlinico San Matteo , Pavia | Laboratory of Virology, INMI Lazzaro Spallanzani IRCCS |
| EPI_ISL_460095 | Molecular Virology Unit, Fondazione IRCCS Policlinico San Matteo , Pavia | Laboratory of Virology, INMI Lazzaro Spallanzani IRCCS |
| EPI_ISL_460096 | Molecular diagnostic laboratory of Federal Budget Institution of Science "Central Research Institute of Epidemiology" of The Federal Service on Customers' Rights Protection and Human Well-being Surveillance | Group of Genomics and Postgenomic Technologies of Central Research Institute of Epidemilology |
| EPI_ISL_460097 | Massachusetts General Hospital | Infectious Disease Program, Broad Institute of Harvard and MIT |
| EPI_ISL_460098 | Massachusetts General Hospital | Infectious Disease Program, Broad Institute of Harvard and MIT |
| EPI_ISL_460099 | Massachusetts General Hospital | Infectious Disease Program, Broad Institute of Harvard and MIT |
| EPI_ISL_460100 | Massachusetts General Hospital | Infectious Disease Program, Broad Institute of Harvard and MIT |
| EPI_ISL_460101 | Massachusetts General Hospital | Infectious Disease Program, Broad Institute of Harvard and MIT |
| EPI_ISL_460102 | Massachusetts General Hospital | Infectious Disease Program, Broad Institute of Harvard and MIT |
| EPI_ISL_460103 | Massachusetts General Hospital | Infectious Disease Program, Broad Institute of Harvard and MIT |
| EPI_ISL_460104 | Massachusetts General Hospital | Infectious Disease Program, Broad Institute of Harvard and MIT |
| EPI_ISL_460105 | Massachusetts General Hospital | Infectious Disease Program, Broad Institute of Harvard and MIT |
| EPI_ISL_460106 | Massachusetts General Hospital | Infectious Disease Program, Broad Institute of Harvard and MIT |
| EPI_ISL_460107 | Massachusetts General Hospital | Infectious Disease Program, Broad Institute of Harvard and MIT |

| EPI_Accession_ID | Originating Laboratory | Submitting Laboratory |
|---|---|---|
| EPI_ISL_460108 | Massachusetts General Hospital | Infectious Disease Program, Broad Institute of Harvard and MIT |
| EPI_ISL_460109 | Massachusetts General Hospital | Infectious Disease Program, Broad Institute of Harvard and MIT |
| EPI_ISL_460110 | Massachusetts General Hospital | Infectious Disease Program, Broad Institute of Harvard and MIT |
| EPI_ISL_460111 | Massachusetts General Hospital | Infectious Disease Program, Broad Institute of Harvard and MIT |
| EPI_ISL_460112 | Massachusetts General Hospital | Infectious Disease Program, Broad Institute of Harvard and MIT |
| EPI_ISL_460113 | Massachusetts General Hospital | Infectious Disease Program, Broad Institute of Harvard and MIT |
| EPI_ISL_460114 | Massachusetts General Hospital | Infectious Disease Program, Broad Institute of Harvard and MIT |
| EPI_ISL_460115 | Massachusetts General Hospital | Infectious Disease Program, Broad Institute of Harvard and MIT |
| EPI_ISL_460116 | Massachusetts General Hospital | Infectious Disease Program, Broad Institute of Harvard and MIT |
| EPI_ISL_460117 | Massachusetts General Hospital | Infectious Disease Program, Broad Institute of Harvard and MIT |
| EPI_ISL_460118 | Massachusetts General Hospital | Infectious Disease Program, Broad Institute of Harvard and MIT |
| EPI_ISL_460119 | Massachusetts General Hospital | Infectious Disease Program, Broad Institute of Harvard and MIT |
| EPI_ISL_460120 | Massachusetts General Hospital | Infectious Disease Program, Broad Institute of Harvard and MIT |
| EPI_ISL_460121 | Massachusetts General Hospital | Infectious Disease Program, Broad Institute of Harvard and MIT |
| EPI_ISL_460122 | unknown | Infectious Disease Program, Broad Institute of Harvard and MIT |
| EPI_ISL_460123 | Massachusetts General Hospital | Infectious Disease Program, Broad Institute of Harvard and MIT |

| EPI_Accession_ID | Originating Laboratory | Submitting Laboratory |
|---|---|---|
| EPI_ISL_460124 | Massachusetts General Hospital | Infectious Disease Program, Broad Institute of Harvard and MIT |
| EPI_ISL_460125 | Massachusetts General Hospital | Infectious Disease Program, Broad Institute of Harvard and MIT |
| EPI_ISL_460126 | Massachusetts General Hospital | Infectious Disease Program, Broad Institute of Harvard and MIT |
| EPI_ISL_460127 | Massachusetts General Hospital | Infectious Disease Program, Broad Institute of Harvard and MIT |
| EPI_ISL_460128 | Massachusetts General Hospital | Infectious Disease Program, Broad Institute of Harvard and MIT |
| EPI_ISL_460129 | Massachusetts General Hospital | Infectious Disease Program, Broad Institute of Harvard and MIT |
| EPI_ISL_460130 | Massachusetts General Hospital | Infectious Disease Program, Broad Institute of Harvard and MIT |
| EPI_ISL_460131 | Massachusetts General Hospital | Infectious Disease Program, Broad Institute of Harvard and MIT |
| EPI_ISL_460132 | Massachusetts General Hospital | Infectious Disease Program, Broad Institute of Harvard and MIT |
| EPI_ISL_460133 | Massachusetts General Hospital | Infectious Disease Program, Broad Institute of Harvard and MIT |
| EPI_ISL_460134 | Massachusetts General Hospital | Infectious Disease Program, Broad Institute of Harvard and MIT |
| EPI_ISL_460135 | Massachusetts General Hospital | Infectious Disease Program, Broad Institute of Harvard and MIT |
| EPI_ISL_460136 | Massachusetts General Hospital | Infectious Disease Program, Broad Institute of Harvard and MIT |
| EPI_ISL_460137 | Massachusetts General Hospital | Infectious Disease Program, Broad Institute of Harvard and MIT |
| EPI_ISL_460138 | Massachusetts General Hospital | Infectious Disease Program, Broad Institute of Harvard and MIT |
| EPI_ISL_460139 | Massachusetts General Hospital | Infectious Disease Program, Broad Institute of Harvard and MIT |

| EPI_Accession_ID | Originating Laboratory | Submitting Laboratory |
|---|---|---|
| EPI_ISL_460140 | Massachusetts General Hospital | Infectious Disease Program, Broad Institute of Harvard and MIT |
| EPI_ISL_460141 | Massachusetts General Hospital | Infectious Disease Program, Broad Institute of Harvard and MIT |
| EPI_ISL_460142 | Massachusetts General Hospital | Infectious Disease Program, Broad Institute of Harvard and MIT |
| EPI_ISL_460143 | Massachusetts General Hospital | Infectious Disease Program, Broad Institute of Harvard and MIT |
| EPI_ISL_460144 | Massachusetts General Hospital | Infectious Disease Program, Broad Institute of Harvard and MIT |
| EPI_ISL_460145 | Massachusetts General Hospital | Infectious Disease Program, Broad Institute of Harvard and MIT |
| EPI_ISL_460146 | Massachusetts General Hospital | Infectious Disease Program, Broad Institute of Harvard and MIT |
| EPI_ISL_460147 | Massachusetts General Hospital | Infectious Disease Program, Broad Institute of Harvard and MIT |
| EPI_ISL_460148 | Massachusetts General Hospital | Infectious Disease Program, Broad Institute of Harvard and MIT |
| EPI_ISL_460149 | Massachusetts General Hospital | Infectious Disease Program, Broad Institute of Harvard and MIT |
| EPI_ISL_460150 | Massachusetts General Hospital | Infectious Disease Program, Broad Institute of Harvard and MIT |
| EPI_ISL_460151 | Massachusetts General Hospital | Infectious Disease Program, Broad Institute of Harvard and MIT |
| EPI_ISL_460152 | Massachusetts General Hospital | Infectious Disease Program, Broad Institute of Harvard and MIT |
| EPI_ISL_460153 | Massachusetts General Hospital | Infectious Disease Program, Broad Institute of Harvard and MIT |
| EPI_ISL_460154 | Massachusetts General Hospital | Infectious Disease Program, Broad Institute of Harvard and MIT |
| EPI_ISL_460155 | Massachusetts General Hospital | Infectious Disease Program, Broad Institute of Harvard and MIT |

| EPI_Accession_ID | Originating Laboratory | Submitting Laboratory |
|---|---|---|
| EPI_ISL_460156 | Massachusetts General Hospital | Infectious Disease Program, Broad Institute of Harvard and MIT |
| EPI_ISL_460157 | Massachusetts General Hospital | Infectious Disease Program, Broad Institute of Harvard and MIT |
| EPI_ISL_460158 | Massachusetts General Hospital | Infectious Disease Program, Broad Institute of Harvard and MIT |
| EPI_ISL_460159 | Massachusetts General Hospital | Infectious Disease Program, Broad Institute of Harvard and MIT |
| EPI_ISL_460160 | Massachusetts General Hospital | Infectious Disease Program, Broad Institute of Harvard and MIT |
| EPI_ISL_460161 | Massachusetts General Hospital | Infectious Disease Program, Broad Institute of Harvard and MIT |
| EPI_ISL_460162 | Massachusetts General Hospital | Infectious Disease Program, Broad Institute of Harvard and MIT |
| EPI_ISL_460163 | Massachusetts General Hospital | Infectious Disease Program, Broad Institute of Harvard and MIT |
| EPI_ISL_460164 | Massachusetts General Hospital | Infectious Disease Program, Broad Institute of Harvard and MIT |
| EPI_ISL_460165 | Massachusetts General Hospital | Infectious Disease Program, Broad Institute of Harvard and MIT |
| EPI_ISL_460166 | Massachusetts General Hospital | Infectious Disease Program, Broad Institute of Harvard and MIT |
| EPI_ISL_460167 | Massachusetts General Hospital | Infectious Disease Program, Broad Institute of Harvard and MIT |
| EPI_ISL_460168 | Massachusetts General Hospital | Infectious Disease Program, Broad Institute of Harvard and MIT |
| EPI_ISL_460169 | Massachusetts General Hospital | Infectious Disease Program, Broad Institute of Harvard and MIT |
| EPI_ISL_460170 | Massachusetts General Hospital | Infectious Disease Program, Broad Institute of Harvard and MIT |
| EPI_ISL_460171 | Massachusetts General Hospital | Infectious Disease Program, Broad Institute of Harvard and MIT |

| EPI_Accession_ID | Originating Laboratory | Submitting Laboratory |
|---|---|---|
| EPI_ISL_460172 | Massachusetts General Hospital | Infectious Disease Program, Broad Institute of Harvard and MIT |
| EPI_ISL_460173 | Massachusetts General Hospital | Infectious Disease Program, Broad Institute of Harvard and MIT |
| EPI_ISL_460174 | Massachusetts General Hospital | Infectious Disease Program, Broad Institute of Harvard and MIT |
| EPI_ISL_460175 | Massachusetts General Hospital | Infectious Disease Program, Broad Institute of Harvard and MIT |
| EPI_ISL_460176 | Massachusetts General Hospital | Infectious Disease Program, Broad Institute of Harvard and MIT |
| EPI_ISL_460177 | Massachusetts General Hospital | Infectious Disease Program, Broad Institute of Harvard and MIT |
| EPI_ISL_460178 | Massachusetts General Hospital | Infectious Disease Program, Broad Institute of Harvard and MIT |
| EPI_ISL_460179 | Massachusetts General Hospital | Infectious Disease Program, Broad Institute of Harvard and MIT |
| EPI_ISL_460180 | Massachusetts General Hospital | Infectious Disease Program, Broad Institute of Harvard and MIT |
| EPI_ISL_460181 | Massachusetts General Hospital | Infectious Disease Program, Broad Institute of Harvard and MIT |
| EPI_ISL_460182 | Massachusetts General Hospital | Infectious Disease Program, Broad Institute of Harvard and MIT |
| EPI_ISL_460183 | Massachusetts General Hospital | Infectious Disease Program, Broad Institute of Harvard and MIT |
| EPI_ISL_460184 | Massachusetts General Hospital | Infectious Disease Program, Broad Institute of Harvard and MIT |
| EPI_ISL_460185 | Massachusetts General Hospital | Infectious Disease Program, Broad Institute of Harvard and MIT |
| EPI_ISL_460186 | Massachusetts General Hospital | Infectious Disease Program, Broad Institute of Harvard and MIT |
| EPI_ISL_460187 | Massachusetts General Hospital | Infectious Disease Program, Broad Institute of Harvard and MIT |

| EPI_Accession_ID | Originating Laboratory | Submitting Laboratory |
|---|---|---|
| EPI_ISL_460188 | Massachusetts General Hospital | Infectious Disease Program, Broad Institute of Harvard and MIT |
| EPI_ISL_460189 | Massachusetts General Hospital | Infectious Disease Program, Broad Institute of Harvard and MIT |
| EPI_ISL_460190 | Massachusetts General Hospital | Infectious Disease Program, Broad Institute of Harvard and MIT |
| EPI_ISL_460191 | Massachusetts General Hospital | Infectious Disease Program, Broad Institute of Harvard and MIT |
| EPI_ISL_460192 | Massachusetts General Hospital | Infectious Disease Program, Broad Institute of Harvard and MIT |
| EPI_ISL_460193 | Massachusetts General Hospital | Infectious Disease Program, Broad Institute of Harvard and MIT |
| EPI_ISL_460194 | Massachusetts General Hospital | Infectious Disease Program, Broad Institute of Harvard and MIT |
| EPI_ISL_460195 | Massachusetts General Hospital | Infectious Disease Program, Broad Institute of Harvard and MIT |
| EPI_ISL_460196 | Massachusetts General Hospital | Infectious Disease Program, Broad Institute of Harvard and MIT |
| EPI_ISL_460197 | Massachusetts General Hospital | Infectious Disease Program, Broad Institute of Harvard and MIT |
| EPI_ISL_460198 | Massachusetts General Hospital | Infectious Disease Program, Broad Institute of Harvard and MIT |
| EPI_ISL_460199 | Massachusetts General Hospital | Infectious Disease Program, Broad Institute of Harvard and MIT |
| EPI_ISL_460200 | Massachusetts General Hospital | Infectious Disease Program, Broad Institute of Harvard and MIT |
| EPI_ISL_460201 | Massachusetts General Hospital | Infectious Disease Program, Broad Institute of Harvard and MIT |
| EPI_ISL_460202 | Massachusetts General Hospital | Infectious Disease Program, Broad Institute of Harvard and MIT |
| EPI_ISL_460203 | Massachusetts General Hospital | Infectious Disease Program, Broad Institute of Harvard and MIT |

| EPI_Accession_ID | Originating Laboratory | Submitting Laboratory |
|---|---|---|
| EPI_ISL_460204 | Massachusetts General Hospital | Infectious Disease Program, Broad Institute of Harvard and MIT |
| EPI_ISL_460205 | Massachusetts General Hospital | Infectious Disease Program, Broad Institute of Harvard and MIT |
| EPI_ISL_460206 | Massachusetts General Hospital | Infectious Disease Program, Broad Institute of Harvard and MIT |
| EPI_ISL_460207 | Massachusetts General Hospital | Infectious Disease Program, Broad Institute of Harvard and MIT |
| EPI_ISL_460208 | Massachusetts General Hospital | Infectious Disease Program, Broad Institute of Harvard and MIT |
| EPI_ISL_460209 | Massachusetts General Hospital | Infectious Disease Program, Broad Institute of Harvard and MIT |
| EPI_ISL_460210 | Massachusetts General Hospital | Infectious Disease Program, Broad Institute of Harvard and MIT |
| EPI_ISL_460211 | Massachusetts General Hospital | Infectious Disease Program, Broad Institute of Harvard and MIT |
| EPI_ISL_460212 | Massachusetts General Hospital | Infectious Disease Program, Broad Institute of Harvard and MIT |
| EPI_ISL_460213 | Massachusetts General Hospital | Infectious Disease Program, Broad Institute of Harvard and MIT |
| EPI_ISL_460214 | Massachusetts General Hospital | Infectious Disease Program, Broad Institute of Harvard and MIT |
| EPI_ISL_460215 | Massachusetts General Hospital | Infectious Disease Program, Broad Institute of Harvard and MIT |
| EPI_ISL_460216 | Massachusetts General Hospital | Infectious Disease Program, Broad Institute of Harvard and MIT |
| EPI_ISL_460217 | Massachusetts General Hospital | Infectious Disease Program, Broad Institute of Harvard and MIT |
| EPI_ISL_460218 | Massachusetts General Hospital | Infectious Disease Program, Broad Institute of Harvard and MIT |
| EPI_ISL_460219 | Massachusetts General Hospital | Infectious Disease Program, Broad Institute of Harvard and MIT |

| EPI_Accession_ID | Originating Laboratory | Submitting Laboratory |
|---|---|---|
| EPI_ISL_460220 | Massachusetts General Hospital | Infectious Disease Program, Broad Institute of Harvard and MIT |
| EPI_ISL_460221 | Massachusetts General Hospital | Infectious Disease Program, Broad Institute of Harvard and MIT |
| EPI_ISL_460223 | Massachusetts General Hospital | Infectious Disease Program, Broad Institute of Harvard and MIT |
| EPI_ISL_460224 | Massachusetts General Hospital | Infectious Disease Program, Broad Institute of Harvard and MIT |
| EPI_ISL_460225 | Massachusetts General Hospital | Infectious Disease Program, Broad Institute of Harvard and MIT |
| EPI_ISL_460226 | Massachusetts General Hospital | Infectious Disease Program, Broad Institute of Harvard and MIT |
| EPI_ISL_460227 | Massachusetts General Hospital | Infectious Disease Program, Broad Institute of Harvard and MIT |
| EPI_ISL_460228 | Massachusetts General Hospital | Infectious Disease Program, Broad Institute of Harvard and MIT |
| EPI_ISL_460229 | Massachusetts General Hospital | Infectious Disease Program, Broad Institute of Harvard and MIT |
| EPI_ISL_460230 | Massachusetts General Hospital | Infectious Disease Program, Broad Institute of Harvard and MIT |
| EPI_ISL_460231 | Massachusetts General Hospital | Infectious Disease Program, Broad Institute of Harvard and MIT |
| EPI_ISL_460232 | Massachusetts General Hospital | Infectious Disease Program, Broad Institute of Harvard and MIT |
| EPI_ISL_460233 | Massachusetts General Hospital | Infectious Disease Program, Broad Institute of Harvard and MIT |
| EPI_ISL_460234 | Massachusetts General Hospital | Infectious Disease Program, Broad Institute of Harvard and MIT |
| EPI_ISL_460235 | Massachusetts General Hospital | Infectious Disease Program, Broad Institute of Harvard and MIT |
| EPI_ISL_460236 | Massachusetts General Hospital | Infectious Disease Program, Broad Institute of Harvard and MIT |

| EPI_Accession_ID | Originating Laboratory | Submitting Laboratory |
|---|---|---|
| EPI_ISL_460237 | Massachusetts General Hospital | Infectious Disease Program, Broad Institute of Harvard and MIT |
| EPI_ISL_460238 | Massachusetts General Hospital | Infectious Disease Program, Broad Institute of Harvard and MIT |
| EPI_ISL_460239 | Massachusetts General Hospital | Infectious Disease Program, Broad Institute of Harvard and MIT |
| EPI_ISL_460240 | Massachusetts General Hospital | Infectious Disease Program, Broad Institute of Harvard and MIT |
| EPI_ISL_460241 | Massachusetts General Hospital | Infectious Disease Program, Broad Institute of Harvard and MIT |
| EPI_ISL_460242 | Massachusetts General Hospital | Infectious Disease Program, Broad Institute of Harvard and MIT |
| EPI_ISL_460243 | Massachusetts General Hospital | Infectious Disease Program, Broad Institute of Harvard and MIT |
| EPI_ISL_460244 | Massachusetts General Hospital | Infectious Disease Program, Broad Institute of Harvard and MIT |
| EPI_ISL_460245 | Massachusetts General Hospital | Infectious Disease Program, Broad Institute of Harvard and MIT |
| EPI_ISL_460246 | Massachusetts General Hospital | Infectious Disease Program, Broad Institute of Harvard and MIT |
| EPI_ISL_460247 | Massachusetts General Hospital | Infectious Disease Program, Broad Institute of Harvard and MIT |
| EPI_ISL_460248 | Massachusetts General Hospital | Infectious Disease Program, Broad Institute of Harvard and MIT |
| EPI_ISL_460249 | Massachusetts General Hospital | Infectious Disease Program, Broad Institute of Harvard and MIT |
| EPI_ISL_460250 | Massachusetts General Hospital | Infectious Disease Program, Broad Institute of Harvard and MIT |
| EPI_ISL_460251 | Massachusetts General Hospital | Infectious Disease Program, Broad Institute of Harvard and MIT |
| EPI_ISL_460252 | Massachusetts General Hospital | Infectious Disease Program, Broad Institute of Harvard and MIT |

| EPI_Accession_ID | Originating Laboratory | Submitting Laboratory |
|---|---|---|
| EPI_ISL_460253 | Massachusetts General Hospital | Infectious Disease Program, Broad Institute of Harvard and MIT |
| EPI_ISL_460254 | Massachusetts General Hospital | Infectious Disease Program, Broad Institute of Harvard and MIT |
| EPI_ISL_460255 | Massachusetts General Hospital | Infectious Disease Program, Broad Institute of Harvard and MIT |
| EPI_ISL_460256 | Massachusetts General Hospital | Infectious Disease Program, Broad Institute of Harvard and MIT |
| EPI_ISL_460257 | Massachusetts General Hospital | Infectious Disease Program, Broad Institute of Harvard and MIT |
| EPI_ISL_460258 | Massachusetts General Hospital | Infectious Disease Program, Broad Institute of Harvard and MIT |
| EPI_ISL_460259 | Massachusetts General Hospital | Infectious Disease Program, Broad Institute of Harvard and MIT |
| EPI_ISL_460260 | Massachusetts General Hospital | Infectious Disease Program, Broad Institute of Harvard and MIT |
| EPI_ISL_460261 | Massachusetts General Hospital | Infectious Disease Program, Broad Institute of Harvard and MIT |
| EPI_ISL_460262 | Massachusetts General Hospital | Infectious Disease Program, Broad Institute of Harvard and MIT |
| EPI_ISL_460263 | Massachusetts General Hospital | Infectious Disease Program, Broad Institute of Harvard and MIT |
| EPI_ISL_460264 | Massachusetts General Hospital | Infectious Disease Program, Broad Institute of Harvard and MIT |
| EPI_ISL_460265 | Massachusetts General Hospital | Infectious Disease Program, Broad Institute of Harvard and MIT |
| EPI_ISL_460266 | Massachusetts General Hospital | Infectious Disease Program, Broad Institute of Harvard and MIT |
| EPI_ISL_460267 | Massachusetts General Hospital | Infectious Disease Program, Broad Institute of Harvard and MIT |
| EPI_ISL_460268 | Massachusetts General Hospital | Infectious Disease Program, Broad Institute of Harvard and MIT |

| EPI_Accession_ID | Originating Laboratory | Submitting Laboratory |
|---|---|---|
| EPI_ISL_460269 | Massachusetts General Hospital | Infectious Disease Program, Broad Institute of Harvard and MIT |
| EPI_ISL_460270 | Massachusetts General Hospital | Infectious Disease Program, Broad Institute of Harvard and MIT |
| EPI_ISL_460271 | Massachusetts General Hospital | Infectious Disease Program, Broad Institute of Harvard and MIT |
| EPI_ISL_460272 | Massachusetts General Hospital | Infectious Disease Program, Broad Institute of Harvard and MIT |
| EPI_ISL_460273 | Massachusetts General Hospital | Infectious Disease Program, Broad Institute of Harvard and MIT |
| EPI_ISL_460274 | Massachusetts General Hospital | Infectious Disease Program, Broad Institute of Harvard and MIT |
| EPI_ISL_460275 | Massachusetts General Hospital | Infectious Disease Program, Broad Institute of Harvard and MIT |
| EPI_ISL_460276 | Massachusetts General Hospital | Infectious Disease Program, Broad Institute of Harvard and MIT |
| EPI_ISL_460277 | Massachusetts General Hospital | Infectious Disease Program, Broad Institute of Harvard and MIT |
| EPI_ISL_460278 | Massachusetts General Hospital | Infectious Disease Program, Broad Institute of Harvard and MIT |
| EPI_ISL_460279 | Massachusetts General Hospital | Infectious Disease Program, Broad Institute of Harvard and MIT |
| EPI_ISL_460280 | Massachusetts General Hospital | Infectious Disease Program, Broad Institute of Harvard and MIT |
| EPI_ISL_460281 | Massachusetts General Hospital | Infectious Disease Program, Broad Institute of Harvard and MIT |
| EPI_ISL_460282 | Massachusetts General Hospital | Infectious Disease Program, Broad Institute of Harvard and MIT |
| EPI_ISL_460283 | Massachusetts General Hospital | Infectious Disease Program, Broad Institute of Harvard and MIT |
| EPI_ISL_460284 | Massachusetts General Hospital | Infectious Disease Program, Broad Institute of Harvard and MIT |

| EPI_Accession_ID | Originating Laboratory | Submitting Laboratory |
|---|---|---|
| EPI_ISL_460285 | Massachusetts General Hospital | Infectious Disease Program, Broad Institute of Harvard and MIT |
| EPI_ISL_460286 | Massachusetts General Hospital | Infectious Disease Program, Broad Institute of Harvard and MIT |
| EPI_ISL_460287 | Massachusetts General Hospital | Infectious Disease Program, Broad Institute of Harvard and MIT |
| EPI_ISL_460288 | Massachusetts General Hospital | Infectious Disease Program, Broad Institute of Harvard and MIT |
| EPI_ISL_460289 | Massachusetts General Hospital | Infectious Disease Program, Broad Institute of Harvard and MIT |
| EPI_ISL_460290 | Massachusetts General Hospital | Infectious Disease Program, Broad Institute of Harvard and MIT |
| EPI_ISL_460291 | Massachusetts General Hospital | Infectious Disease Program, Broad Institute of Harvard and MIT |
| EPI_ISL_460292 | Massachusetts General Hospital | Infectious Disease Program, Broad Institute of Harvard and MIT |
| EPI_ISL_460293 | Massachusetts General Hospital | Infectious Disease Program, Broad Institute of Harvard and MIT |
| EPI_ISL_460294 | Massachusetts General Hospital | Infectious Disease Program, Broad Institute of Harvard and MIT |
| EPI_ISL_460295 | Massachusetts General Hospital | Infectious Disease Program, Broad Institute of Harvard and MIT |
| EPI_ISL_460296 | Massachusetts General Hospital | Infectious Disease Program, Broad Institute of Harvard and MIT |
| EPI_ISL_460297 | Massachusetts General Hospital | Infectious Disease Program, Broad Institute of Harvard and MIT |
| EPI_ISL_460298 | Massachusetts General Hospital | Infectious Disease Program, Broad Institute of Harvard and MIT |
| EPI_ISL_460299 | Massachusetts General Hospital | Infectious Disease Program, Broad Institute of Harvard and MIT |
| EPI_ISL_460300 | Massachusetts General Hospital | Infectious Disease Program, Broad Institute of Harvard and MIT |

| EPI_Accession_ID | Originating Laboratory | Submitting Laboratory |
|---|---|---|
| EPI_ISL_460301 | Massachusetts General Hospital | Infectious Disease Program, Broad Institute of Harvard and MIT |
| EPI_ISL_460302 | Massachusetts General Hospital | Infectious Disease Program, Broad Institute of Harvard and MIT |
| EPI_ISL_460303 | Massachusetts General Hospital | Infectious Disease Program, Broad Institute of Harvard and MIT |
| EPI_ISL_460304 | Massachusetts General Hospital | Infectious Disease Program, Broad Institute of Harvard and MIT |
| EPI_ISL_460305 | Massachusetts General Hospital | Infectious Disease Program, Broad Institute of Harvard and MIT |
| EPI_ISL_460306 | Massachusetts General Hospital | Infectious Disease Program, Broad Institute of Harvard and MIT |
| EPI_ISL_460307 | Massachusetts General Hospital | Infectious Disease Program, Broad Institute of Harvard and MIT |
| EPI_ISL_460308 | Massachusetts General Hospital | Infectious Disease Program, Broad Institute of Harvard and MIT |
| EPI_ISL_460309 | Massachusetts General Hospital | Infectious Disease Program, Broad Institute of Harvard and MIT |
| EPI_ISL_460310 | Massachusetts General Hospital | Infectious Disease Program, Broad Institute of Harvard and MIT |
| EPI_ISL_460311 | Massachusetts General Hospital | Infectious Disease Program, Broad Institute of Harvard and MIT |
| EPI_ISL_460312 | Massachusetts General Hospital | Infectious Disease Program, Broad Institute of Harvard and MIT |
| EPI_ISL_460313 | Massachusetts General Hospital | Infectious Disease Program, Broad Institute of Harvard and MIT |
| EPI_ISL_460314 | Massachusetts General Hospital | Infectious Disease Program, Broad Institute of Harvard and MIT |
| EPI_ISL_460315 | Massachusetts General Hospital | Infectious Disease Program, Broad Institute of Harvard and MIT |
| EPI_ISL_460316 | Massachusetts General Hospital | Infectious Disease Program, Broad Institute of Harvard and MIT |

| EPI_Accession_ID | Originating Laboratory | Submitting Laboratory |
|---|---|---|
| EPI_ISL_460317 | Massachusetts General Hospital | Infectious Disease Program, Broad Institute of Harvard and MIT |
| EPI_ISL_460319 | Massachusetts General Hospital | Infectious Disease Program, Broad Institute of Harvard and MIT |
| EPI_ISL_460320 | Massachusetts General Hospital | Infectious Disease Program, Broad Institute of Harvard and MIT |
| EPI_ISL_460321 | Massachusetts General Hospital | Infectious Disease Program, Broad Institute of Harvard and MIT |
| EPI_ISL_460322 | Massachusetts General Hospital | Infectious Disease Program, Broad Institute of Harvard and MIT |
| EPI_ISL_460323 | Massachusetts General Hospital | Infectious Disease Program, Broad Institute of Harvard and MIT |
| EPI_ISL_460324 | Massachusetts General Hospital | Infectious Disease Program, Broad Institute of Harvard and MIT |
| EPI_ISL_460325 | Massachusetts General Hospital | Infectious Disease Program, Broad Institute of Harvard and MIT |
| EPI_ISL_460326 | Massachusetts General Hospital | Infectious Disease Program, Broad Institute of Harvard and MIT |
| EPI_ISL_460327 | Massachusetts General Hospital | Infectious Disease Program, Broad Institute of Harvard and MIT |
| EPI_ISL_460328 | Massachusetts General Hospital | Infectious Disease Program, Broad Institute of Harvard and MIT |
| EPI_ISL_460329 | Massachusetts General Hospital | Infectious Disease Program, Broad Institute of Harvard and MIT |
| EPI_ISL_460330 | Massachusetts General Hospital | Infectious Disease Program, Broad Institute of Harvard and MIT |
| EPI_ISL_460331 | Massachusetts General Hospital | Infectious Disease Program, Broad Institute of Harvard and MIT |
| EPI_ISL_460332 | Massachusetts General Hospital | Infectious Disease Program, Broad Institute of Harvard and MIT |
| EPI_ISL_460333 | Massachusetts General Hospital | Infectious Disease Program, Broad Institute of Harvard and MIT |

| EPI_Accession_ID | Originating Laboratory | Submitting Laboratory |
|---|---|---|
| EPI_ISL_460334 | Massachusetts General Hospital | Infectious Disease Program, Broad Institute of Harvard and MIT |
| EPI_ISL_460335 | Massachusetts General Hospital | Infectious Disease Program, Broad Institute of Harvard and MIT |
| EPI_ISL_460336 | Massachusetts General Hospital | Infectious Disease Program, Broad Institute of Harvard and MIT |
| EPI_ISL_460337 | Massachusetts General Hospital | Infectious Disease Program, Broad Institute of Harvard and MIT |
| EPI_ISL_460338 | Massachusetts General Hospital | Infectious Disease Program, Broad Institute of Harvard and MIT |
| EPI_ISL_460339 | Massachusetts General Hospital | Infectious Disease Program, Broad Institute of Harvard and MIT |
| EPI_ISL_460340 | Massachusetts General Hospital | Infectious Disease Program, Broad Institute of Harvard and MIT |
| EPI_ISL_460341 | Massachusetts General Hospital | Infectious Disease Program, Broad Institute of Harvard and MIT |
| EPI_ISL_460342 | Massachusetts General Hospital | Infectious Disease Program, Broad Institute of Harvard and MIT |
| EPI_ISL_460343 | Massachusetts General Hospital | Infectious Disease Program, Broad Institute of Harvard and MIT |
| EPI_ISL_460344 | Massachusetts General Hospital | Infectious Disease Program, Broad Institute of Harvard and MIT |
| EPI_ISL_460345 | Massachusetts General Hospital | Infectious Disease Program, Broad Institute of Harvard and MIT |
| EPI_ISL_460346 | Massachusetts General Hospital | Infectious Disease Program, Broad Institute of Harvard and MIT |
| EPI_ISL_460347 | Massachusetts General Hospital | Infectious Disease Program, Broad Institute of Harvard and MIT |
| EPI_ISL_460348 | Massachusetts General Hospital | Infectious Disease Program, Broad Institute of Harvard and MIT |
| EPI_ISL_460349 | Massachusetts General Hospital | Infectious Disease Program, Broad Institute of Harvard and MIT |

| EPI_Accession_ID | Originating Laboratory | Submitting Laboratory |
|---|---|---|
| EPI_ISL_460350 | Massachusetts General Hospital | Infectious Disease Program, Broad Institute of Harvard and MIT |
| EPI_ISL_460351 | Massachusetts General Hospital | Infectious Disease Program, Broad Institute of Harvard and MIT |
| EPI_ISL_460352 | Massachusetts General Hospital | Infectious Disease Program, Broad Institute of Harvard and MIT |
| EPI_ISL_460353 | Massachusetts General Hospital | Infectious Disease Program, Broad Institute of Harvard and MIT |
| EPI_ISL_460354 | Massachusetts General Hospital | Infectious Disease Program, Broad Institute of Harvard and MIT |
| EPI_ISL_460355 | Massachusetts General Hospital | Infectious Disease Program, Broad Institute of Harvard and MIT |
| EPI_ISL_460356 | Massachusetts General Hospital | Infectious Disease Program, Broad Institute of Harvard and MIT |
| EPI_ISL_460357 | Massachusetts General Hospital | Infectious Disease Program, Broad Institute of Harvard and MIT |
| EPI_ISL_460358 | Massachusetts General Hospital | Infectious Disease Program, Broad Institute of Harvard and MIT |
| EPI_ISL_460359 | Massachusetts General Hospital | Infectious Disease Program, Broad Institute of Harvard and MIT |
| EPI_ISL_460360 | Massachusetts General Hospital | Infectious Disease Program, Broad Institute of Harvard and MIT |
| EPI_ISL_460361 | Massachusetts General Hospital | Infectious Disease Program, Broad Institute of Harvard and MIT |
| EPI_ISL_460362 | Massachusetts General Hospital | Infectious Disease Program, Broad Institute of Harvard and MIT |
| EPI_ISL_460363 | Massachusetts General Hospital | Infectious Disease Program, Broad Institute of Harvard and MIT |
| EPI_ISL_460364 | Massachusetts General Hospital | Infectious Disease Program, Broad Institute of Harvard and MIT |
| EPI_ISL_460365 | Massachusetts General Hospital | Infectious Disease Program, Broad Institute of Harvard and MIT |

| EPI_Accession_ID | Originating Laboratory | Submitting Laboratory |
|---|---|---|
| EPI_ISL_460366 | Massachusetts General Hospital | Infectious Disease Program, Broad Institute of Harvard and MIT |
| EPI_ISL_460367 | Massachusetts General Hospital | Infectious Disease Program, Broad Institute of Harvard and MIT |
| EPI_ISL_460368 | Massachusetts General Hospital | Infectious Disease Program, Broad Institute of Harvard and MIT |
| EPI_ISL_460369 | Massachusetts General Hospital | Infectious Disease Program, Broad Institute of Harvard and MIT |
| EPI_ISL_460370 | Massachusetts General Hospital | Infectious Disease Program, Broad Institute of Harvard and MIT |
| EPI_ISL_460371 | Massachusetts General Hospital | Infectious Disease Program, Broad Institute of Harvard and MIT |
| EPI_ISL_460372 | Massachusetts General Hospital | Infectious Disease Program, Broad Institute of Harvard and MIT |
| EPI_ISL_460373 | Massachusetts General Hospital | Infectious Disease Program, Broad Institute of Harvard and MIT |
| EPI_ISL_460374 | Massachusetts General Hospital | Infectious Disease Program, Broad Institute of Harvard and MIT |
| EPI_ISL_460375 | Massachusetts General Hospital | Infectious Disease Program, Broad Institute of Harvard and MIT |
| EPI_ISL_460376 | Massachusetts General Hospital | Infectious Disease Program, Broad Institute of Harvard and MIT |
| EPI_ISL_460377 | Massachusetts General Hospital | Infectious Disease Program, Broad Institute of Harvard and MIT |
| EPI_ISL_460378 | Massachusetts General Hospital | Infectious Disease Program, Broad Institute of Harvard and MIT |
| EPI_ISL_460379 | Massachusetts General Hospital | Infectious Disease Program, Broad Institute of Harvard and MIT |
| EPI_ISL_460380 | Massachusetts General Hospital | Infectious Disease Program, Broad Institute of Harvard and MIT |
| EPI_ISL_460381 | Massachusetts General Hospital | Infectious Disease Program, Broad Institute of Harvard and MIT |

| EPI_Accession_ID | Originating Laboratory | Submitting Laboratory |
|---|---|---|
| EPI_ISL_460382 | Massachusetts General Hospital | Infectious Disease Program, Broad Institute of Harvard and MIT |
| EPI_ISL_460383 | Massachusetts General Hospital | Infectious Disease Program, Broad Institute of Harvard and MIT |
| EPI_ISL_460384 | Massachusetts General Hospital | Infectious Disease Program, Broad Institute of Harvard and MIT |
| EPI_ISL_460385 | Massachusetts General Hospital | Infectious Disease Program, Broad Institute of Harvard and MIT |
| EPI_ISL_460386 | Massachusetts General Hospital | Infectious Disease Program, Broad Institute of Harvard and MIT |
| EPI_ISL_460387 | Massachusetts General Hospital | Infectious Disease Program, Broad Institute of Harvard and MIT |
| EPI_ISL_460388 | Massachusetts General Hospital | Infectious Disease Program, Broad Institute of Harvard and MIT |
| EPI_ISL_460389 | Massachusetts General Hospital | Infectious Disease Program, Broad Institute of Harvard and MIT |
| EPI_ISL_460390 | Massachusetts General Hospital | Infectious Disease Program, Broad Institute of Harvard and MIT |
| EPI_ISL_460391 | Massachusetts General Hospital | Infectious Disease Program, Broad Institute of Harvard and MIT |
| EPI_ISL_460392 | Massachusetts General Hospital | Infectious Disease Program, Broad Institute of Harvard and MIT |
| EPI_ISL_460393 | Massachusetts General Hospital | Infectious Disease Program, Broad Institute of Harvard and MIT |
| EPI_ISL_460394 | Massachusetts General Hospital | Infectious Disease Program, Broad Institute of Harvard and MIT |
| EPI_ISL_460395 | Massachusetts General Hospital | Infectious Disease Program, Broad Institute of Harvard and MIT |
| EPI_ISL_460396 | Massachusetts General Hospital | Infectious Disease Program, Broad Institute of Harvard and MIT |
| EPI_ISL_460397 | Massachusetts General Hospital | Infectious Disease Program, Broad Institute of Harvard and MIT |

| EPI_Accession_ID | Originating Laboratory | Submitting Laboratory |
|---|---|---|
| EPI_ISL_460398 | Massachusetts General Hospital | Infectious Disease Program, Broad Institute of Harvard and MIT |
| EPI_ISL_460399 | Massachusetts General Hospital | Infectious Disease Program, Broad Institute of Harvard and MIT |
| EPI_ISL_460400 | Massachusetts General Hospital | Infectious Disease Program, Broad Institute of Harvard and MIT |
| EPI_ISL_460401 | Massachusetts General Hospital | Infectious Disease Program, Broad Institute of Harvard and MIT |
| EPI_ISL_460402 | Massachusetts General Hospital | Infectious Disease Program, Broad Institute of Harvard and MIT |
| EPI_ISL_460403 | Massachusetts General Hospital | Infectious Disease Program, Broad Institute of Harvard and MIT |
| EPI_ISL_460404 | Massachusetts General Hospital | Infectious Disease Program, Broad Institute of Harvard and MIT |
| EPI_ISL_460405 | Massachusetts General Hospital | Infectious Disease Program, Broad Institute of Harvard and MIT |
| EPI_ISL_460406 | Massachusetts General Hospital | Infectious Disease Program, Broad Institute of Harvard and MIT |
| EPI_ISL_460407 | Massachusetts General Hospital | Infectious Disease Program, Broad Institute of Harvard and MIT |
| EPI_ISL_460408 | Massachusetts General Hospital | Infectious Disease Program, Broad Institute of Harvard and MIT |
| EPI_ISL_460409 | Massachusetts General Hospital | Infectious Disease Program, Broad Institute of Harvard and MIT |
| EPI_ISL_460410 | Massachusetts General Hospital | Infectious Disease Program, Broad Institute of Harvard and MIT |
| EPI_ISL_460411 | Massachusetts General Hospital | Infectious Disease Program, Broad Institute of Harvard and MIT |
| EPI_ISL_460412 | Massachusetts General Hospital | Infectious Disease Program, Broad Institute of Harvard and MIT |
| EPI_ISL_460413 | Massachusetts General Hospital | Infectious Disease Program, Broad Institute of Harvard and MIT |

| EPI_Accession_ID | Originating Laboratory | Submitting Laboratory |
|---|---|---|
| EPI_ISL_460414 | Massachusetts General Hospital | Infectious Disease Program, Broad Institute of Harvard and MIT |
| EPI_ISL_460415 | Massachusetts General Hospital | Infectious Disease Program, Broad Institute of Harvard and MIT |
| EPI_ISL_460417 | Massachusetts General Hospital | Infectious Disease Program, Broad Institute of Harvard and MIT |
| EPI_ISL_460418 | Massachusetts General Hospital | Infectious Disease Program, Broad Institute of Harvard and MIT |
| EPI_ISL_460419 | Massachusetts General Hospital | Infectious Disease Program, Broad Institute of Harvard and MIT |
| EPI_ISL_460420 | Massachusetts General Hospital | Infectious Disease Program, Broad Institute of Harvard and MIT |
| EPI_ISL_460421 | Massachusetts General Hospital | Infectious Disease Program, Broad Institute of Harvard and MIT |
| EPI_ISL_460422 | Massachusetts General Hospital | Infectious Disease Program, Broad Institute of Harvard and MIT |
| EPI_ISL_460423 | Massachusetts General Hospital | Infectious Disease Program, Broad Institute of Harvard and MIT |
| EPI_ISL_460424 | Massachusetts General Hospital | Infectious Disease Program, Broad Institute of Harvard and MIT |
| EPI_ISL_460425 | Massachusetts General Hospital | Infectious Disease Program, Broad Institute of Harvard and MIT |
| EPI_ISL_460426 | Massachusetts General Hospital | Infectious Disease Program, Broad Institute of Harvard and MIT |
| EPI_ISL_460427 | Massachusetts General Hospital | Infectious Disease Program, Broad Institute of Harvard and MIT |
| EPI_ISL_460428 | Massachusetts General Hospital | Infectious Disease Program, Broad Institute of Harvard and MIT |
| EPI_ISL_460429 | Massachusetts General Hospital | Infectious Disease Program, Broad Institute of Harvard and MIT |
| EPI_ISL_460430 | Massachusetts General Hospital | Infectious Disease Program, Broad Institute of Harvard and MIT |

| EPI_Accession_ID | Originating Laboratory | Submitting Laboratory |
|---|---|---|
| EPI_ISL_460431 | Massachusetts General Hospital | Infectious Disease Program, Broad Institute of Harvard and MIT |
| EPI_ISL_460432 | Massachusetts General Hospital | Infectious Disease Program, Broad Institute of Harvard and MIT |
| EPI_ISL_460433 | Massachusetts General Hospital | Infectious Disease Program, Broad Institute of Harvard and MIT |
| EPI_ISL_460434 | Massachusetts General Hospital | Infectious Disease Program, Broad Institute of Harvard and MIT |
| EPI_ISL_460435 | Massachusetts General Hospital | Infectious Disease Program, Broad Institute of Harvard and MIT |
| EPI_ISL_460436 | Massachusetts General Hospital | Infectious Disease Program, Broad Institute of Harvard and MIT |
| EPI_ISL_460437 | Massachusetts General Hospital | Infectious Disease Program, Broad Institute of Harvard and MIT |
| EPI_ISL_460438 | Massachusetts General Hospital | Infectious Disease Program, Broad Institute of Harvard and MIT |
| EPI_ISL_460439 | Massachusetts General Hospital | Infectious Disease Program, Broad Institute of Harvard and MIT |
| EPI_ISL_460440 | Massachusetts General Hospital | Infectious Disease Program, Broad Institute of Harvard and MIT |
| EPI_ISL_460441 | Massachusetts General Hospital | Infectious Disease Program, Broad Institute of Harvard and MIT |
| EPI_ISL_460442 | Massachusetts General Hospital | Infectious Disease Program, Broad Institute of Harvard and MIT |
| EPI_ISL_460443 | Massachusetts General Hospital | Infectious Disease Program, Broad Institute of Harvard and MIT |
| EPI_ISL_460444 | Massachusetts General Hospital | Infectious Disease Program, Broad Institute of Harvard and MIT |
| EPI_ISL_460445 | Massachusetts General Hospital | Infectious Disease Program, Broad Institute of Harvard and MIT |
| EPI_ISL_460446 | Massachusetts General Hospital | Infectious Disease Program, Broad Institute of Harvard and MIT |

| EPI_Accession_ID | Originating Laboratory | Submitting Laboratory |
|---|---|---|
| EPI_ISL_460447 | Massachusetts General Hospital | Infectious Disease Program, Broad Institute of Harvard and MIT |
| EPI_ISL_460448 | Massachusetts General Hospital | Infectious Disease Program, Broad Institute of Harvard and MIT |
| EPI_ISL_460450 | Massachusetts General Hospital | Infectious Disease Program, Broad Institute of Harvard and MIT |
| EPI_ISL_460451 | Massachusetts General Hospital | Infectious Disease Program, Broad Institute of Harvard and MIT |
| EPI_ISL_460452 | Massachusetts General Hospital | Infectious Disease Program, Broad Institute of Harvard and MIT |
| EPI_ISL_460453 | Massachusetts General Hospital | Infectious Disease Program, Broad Institute of Harvard and MIT |
| EPI_ISL_460454 | Massachusetts General Hospital | Infectious Disease Program, Broad Institute of Harvard and MIT |
| EPI_ISL_460455 | Massachusetts General Hospital | Infectious Disease Program, Broad Institute of Harvard and MIT |
| EPI_ISL_460456 | Massachusetts General Hospital | Infectious Disease Program, Broad Institute of Harvard and MIT |
| EPI_ISL_460457 | Massachusetts General Hospital | Infectious Disease Program, Broad Institute of Harvard and MIT |
| EPI_ISL_460458 | Massachusetts General Hospital | Infectious Disease Program, Broad Institute of Harvard and MIT |
| EPI_ISL_460459 | Massachusetts General Hospital | Infectious Disease Program, Broad Institute of Harvard and MIT |
| EPI_ISL_460460 | Massachusetts General Hospital | Infectious Disease Program, Broad Institute of Harvard and MIT |
| EPI_ISL_460461 | Massachusetts General Hospital | Infectious Disease Program, Broad Institute of Harvard and MIT |
| EPI_ISL_460462 | Massachusetts General Hospital | Infectious Disease Program, Broad Institute of Harvard and MIT |
| EPI_ISL_460463 | Massachusetts General Hospital | Infectious Disease Program, Broad Institute of Harvard and MIT |

| EPI_Accession_ID | Originating Laboratory | Submitting Laboratory |
|---|---|---|
| EPI_ISL_460464 | Massachusetts General Hospital | Infectious Disease Program, Broad Institute of Harvard and MIT |
| EPI_ISL_460465 | Massachusetts General Hospital | Infectious Disease Program, Broad Institute of Harvard and MIT |
| EPI_ISL_460466 | Massachusetts General Hospital | Infectious Disease Program, Broad Institute of Harvard and MIT |
| EPI_ISL_460467 | Massachusetts General Hospital | Infectious Disease Program, Broad Institute of Harvard and MIT |
| EPI_ISL_460468 | Massachusetts General Hospital | Infectious Disease Program, Broad Institute of Harvard and MIT |
| EPI_ISL_460469 | Massachusetts General Hospital | Infectious Disease Program, Broad Institute of Harvard and MIT |
| EPI_ISL_460470 | Massachusetts General Hospital | Infectious Disease Program, Broad Institute of Harvard and MIT |
| EPI_ISL_460471 | Massachusetts General Hospital | Infectious Disease Program, Broad Institute of Harvard and MIT |
| EPI_ISL_460472 | Massachusetts General Hospital | Infectious Disease Program, Broad Institute of Harvard and MIT |
| EPI_ISL_460554 | Michigan Department of Health and Human Services, Bureau of Laboratories | Michigan Department of Health and Human Services, Bureau of Laboratories |
| EPI_ISL_460555 | Michigan Department of Health and Human Services, Bureau of Laboratories | Michigan Department of Health and Human Services, Bureau of Laboratories |
| EPI_ISL_460556 | Michigan Department of Health and Human Services, Bureau of Laboratories | Michigan Department of Health and Human Services, Bureau of Laboratories |
| EPI_ISL_460557 | Michigan Department of Health and Human Services, Bureau of Laboratories | Michigan Department of Health and Human Services, Bureau of Laboratories |
| EPI_ISL_460558 | Michigan Department of Health and Human Services, Bureau of Laboratories | Michigan Department of Health and Human Services, Bureau of Laboratories |
| EPI_ISL_460559 | Michigan Department of Health and Human Services, Bureau of Laboratories | Michigan Department of Health and Human Services, Bureau of Laboratories |
| EPI_ISL_460560 | Michigan Department of Health and Human Services, Bureau of Laboratories | Michigan Department of Health and Human Services, Bureau of Laboratories |

| EPI_Accession_ID | Originating Laboratory | Submitting Laboratory |
|---|---|---|
| EPI_ISL_460561 | Michigan Department of Health and Human Services, Bureau of Laboratories | Michigan Department of Health and Human Services, Bureau of Laboratories |
| EPI_ISL_460562 | Michigan Department of Health and Human Services, Bureau of Laboratories | Michigan Department of Health and Human Services, Bureau of Laboratories |
| EPI_ISL_460563 | Michigan Department of Health and Human Services, Bureau of Laboratories | Michigan Department of Health and Human Services, Bureau of Laboratories |
| EPI_ISL_460564 | Michigan Department of Health and Human Services, Bureau of Laboratories | Michigan Department of Health and Human Services, Bureau of Laboratories |
| EPI_ISL_460565 | Michigan Department of Health and Human Services, Bureau of Laboratories | Michigan Department of Health and Human Services, Bureau of Laboratories |
| EPI_ISL_460566 | Michigan Department of Health and Human Services, Bureau of Laboratories | Michigan Department of Health and Human Services, Bureau of Laboratories |
| EPI_ISL_460567 | Michigan Department of Health and Human Services, Bureau of Laboratories | Michigan Department of Health and Human Services, Bureau of Laboratories |
| EPI_ISL_460568 | Michigan Department of Health and Human Services, Bureau of Laboratories | Michigan Department of Health and Human Services, Bureau of Laboratories |
| EPI_ISL_460569 | Michigan Department of Health and Human Services, Bureau of Laboratories | Michigan Department of Health and Human Services, Bureau of Laboratories |
| EPI_ISL_460570 | Michigan Department of Health and Human Services, Bureau of Laboratories | Michigan Department of Health and Human Services, Bureau of Laboratories |
| EPI_ISL_460571 | Michigan Department of Health and Human Services, Bureau of Laboratories | Michigan Department of Health and Human Services, Bureau of Laboratories |
| EPI_ISL_460572 | Michigan Department of Health and Human Services, Bureau of Laboratories | Michigan Department of Health and Human Services, Bureau of Laboratories |
| EPI_ISL_460573 | Michigan Department of Health and Human Services, Bureau of Laboratories | Michigan Department of Health and Human Services, Bureau of Laboratories |
| EPI_ISL_460574 | Michigan Department of Health and Human Services, Bureau of Laboratories | Michigan Department of Health and Human Services, Bureau of Laboratories |
| EPI_ISL_460575 | Michigan Department of Health and Human Services, Bureau of Laboratories | Michigan Department of Health and Human Services, Bureau of Laboratories |
| EPI_ISL_460576 | Michigan Department of Health and Human Services, Bureau of Laboratories | Michigan Department of Health and Human Services, Bureau of Laboratories |

| EPI_Accession_ID | Originating Laboratory | Submitting Laboratory |
|---|---|---|
| EPI_ISL_460577 | Michigan Department of Health and Human Services, Bureau of Laboratories | Michigan Department of Health and Human Services, Bureau of Laboratories |
| EPI_ISL_460578 | Michigan Department of Health and Human Services, Bureau of Laboratories | Michigan Department of Health and Human Services, Bureau of Laboratories |
| EPI_ISL_460579 | Michigan Department of Health and Human Services, Bureau of Laboratories | Michigan Department of Health and Human Services, Bureau of Laboratories |
| EPI_ISL_460580 | Michigan Department of Health and Human Services, Bureau of Laboratories | Michigan Department of Health and Human Services, Bureau of Laboratories |
| EPI_ISL_460581 | Michigan Department of Health and Human Services, Bureau of Laboratories | Michigan Department of Health and Human Services, Bureau of Laboratories |
| EPI_ISL_460582 | Michigan Department of Health and Human Services, Bureau of Laboratories | Michigan Department of Health and Human Services, Bureau of Laboratories |
| EPI_ISL_460583 | Michigan Department of Health and Human Services, Bureau of Laboratories | Michigan Department of Health and Human Services, Bureau of Laboratories |
| EPI_ISL_460584 | Michigan Department of Health and Human Services, Bureau of Laboratories | Michigan Department of Health and Human Services, Bureau of Laboratories |
| EPI_ISL_460585 | Michigan Department of Health and Human Services, Bureau of Laboratories | Michigan Department of Health and Human Services, Bureau of Laboratories |
| EPI_ISL_460586 | Michigan Department of Health and Human Services, Bureau of Laboratories | Michigan Department of Health and Human Services, Bureau of Laboratories |
| EPI_ISL_460587 | Michigan Department of Health and Human Services, Bureau of Laboratories | Michigan Department of Health and Human Services, Bureau of Laboratories |
| EPI_ISL_460588 | Michigan Department of Health and Human Services, Bureau of Laboratories | Michigan Department of Health and Human Services, Bureau of Laboratories |
| EPI_ISL_460590 | Michigan Department of Health and Human Services, Bureau of Laboratories | Michigan Department of Health and Human Services, Bureau of Laboratories |
| EPI_ISL_460592 | Michigan Department of Health and Human Services, Bureau of Laboratories | Michigan Department of Health and Human Services, Bureau of Laboratories |
| EPI_ISL_460593 | Michigan Department of Health and Human Services, Bureau of Laboratories | Michigan Department of Health and Human Services, Bureau of Laboratories |
| EPI_ISL_460594 | Michigan Department of Health and Human Services, Bureau of Laboratories | Michigan Department of Health and Human Services, Bureau of Laboratories |

| EPI_Accession_ID | Originating Laboratory | Submitting Laboratory |
|---|---|---|
| EPI_ISL_460595 | Michigan Department of Health and Human Services, Bureau of Laboratories | Michigan Department of Health and Human Services, Bureau of Laboratories |
| EPI_ISL_460596 | Michigan Department of Health and Human Services, Bureau of Laboratories | Michigan Department of Health and Human Services, Bureau of Laboratories |
| EPI_ISL_460597 | Michigan Department of Health and Human Services, Bureau of Laboratories | Michigan Department of Health and Human Services, Bureau of Laboratories |
| EPI_ISL_460598 | Michigan Department of Health and Human Services, Bureau of Laboratories | Michigan Department of Health and Human Services, Bureau of Laboratories |
| EPI_ISL_460599 | Michigan Department of Health and Human Services, Bureau of Laboratories | Michigan Department of Health and Human Services, Bureau of Laboratories |
| EPI_ISL_460600 | Michigan Department of Health and Human Services, Bureau of Laboratories | Michigan Department of Health and Human Services, Bureau of Laboratories |
| EPI_ISL_460601 | Michigan Department of Health and Human Services, Bureau of Laboratories | Michigan Department of Health and Human Services, Bureau of Laboratories |
| EPI_ISL_460602 | Michigan Department of Health and Human Services, Bureau of Laboratories | Michigan Department of Health and Human Services, Bureau of Laboratories |
| EPI_ISL_460603 | NYU Langone Health | Departments of Pathology and Medicine, New York University School of Medicine |
| EPI_ISL_460604 | Molecular diagnostic laboratory of Federal Budget Institution of Science "Central Research Institute of Epidemiology" of The Federal Service on Customers' Rights Protection and Human Well-being Surveillance | Group of Genomics and Postgenomic Technologies of Central Research Institute of Epidemilology |
| EPI_ISL_460605 | Molecular diagnostic laboratory of Federal Budget Institution of Science "Central Research Institute of Epidemiology" of The Federal Service on Customers' Rights Protection and Human Well-being Surveillance | Group of Genomics and Postgenomic Technologies of Central Research Institute of Epidemilology |
| EPI_ISL_460617 | unknown | Physiology |
| EPI_ISL_460618 | unknown | Physiology |
| EPI_ISL_460619 | unknown | Physiology |
| EPI_ISL_460621 | UW Virology Lab | UW Virology Lab |
| EPI_ISL_460622 | UW Virology Lab | UW Virology Lab |
| EPI_ISL_460623 | UW Virology Lab | UW Virology Lab |
| EPI_ISL_460624 | UW Virology Lab | UW Virology Lab |

| EPI_Accession_ID | Originating Laboratory | Submitting Laboratory |
|---|---|---|
| EPI_ISL_460625 | UW Virology Lab | UW Virology Lab |
| EPI_ISL_460626 | UW Virology Lab | UW Virology Lab |
| EPI_ISL_460627 | UW Virology Lab | UW Virology Lab |
| EPI_ISL_460628 | UW Virology Lab | UW Virology Lab |
| EPI_ISL_460629 | UW Virology Lab | UW Virology Lab |
| EPI_ISL_460630 | UW Virology Lab | UW Virology Lab |
| EPI_ISL_460631 | UW Virology Lab | UW Virology Lab |
| EPI_ISL_460632 | UW Virology Lab | UW Virology Lab |
| EPI_ISL_460633 | UW Virology Lab | UW Virology Lab |
| EPI_ISL_460634 | UW Virology Lab | UW Virology Lab |
| EPI_ISL_460635 | UHCW Pathology / University of Warwick | University of Warwick, for the COVID-19 Genomics (COG) UK Consortium |
| EPI_ISL_460636 | Dutch COVID-19 response team | Erasmus Medical Center |
| EPI_ISL_460637 | Dutch COVID-19 response team | Erasmus Medical Center |
| EPI_ISL_460638 | Dutch COVID-19 response team | Erasmus Medical Center |
| EPI_ISL_460639 | Dutch COVID-19 response team | Erasmus Medical Center |
| EPI_ISL_460640 | Dutch COVID-19 response team | Erasmus Medical Center |
| EPI_ISL_460641 | Dutch COVID-19 response team | Erasmus Medical Center |
| EPI_ISL_460642 | Dutch COVID-19 response team | Erasmus Medical Center |
| EPI_ISL_460643 | Dutch COVID-19 response team | Erasmus Medical Center |
| EPI_ISL_460644 | Dutch COVID-19 response team | Erasmus Medical Center |
| EPI_ISL_460645 | Dutch COVID-19 response team | Erasmus Medical Center |
| EPI_ISL_460646 | Dutch COVID-19 response team | Erasmus Medical Center |
| EPI_ISL_460647 | Dutch COVID-19 response team | Erasmus Medical Center |
| EPI_ISL_460648 | Dutch COVID-19 response team | Erasmus Medical Center |
| EPI_ISL_460649 | Dutch COVID-19 response team | Erasmus Medical Center |
| EPI_ISL_460650 | Dutch COVID-19 response team | Erasmus Medical Center |
| EPI_ISL_460651 | Dutch COVID-19 response team | Erasmus Medical Center |
| EPI_ISL_460652 | Dutch COVID-19 response team | Erasmus Medical Center |
| EPI_ISL_460653 | Dutch COVID-19 response team | Erasmus Medical Center |
| EPI_ISL_460654 | Dutch COVID-19 response team | Erasmus Medical Center |
| EPI_ISL_460655 | Dutch COVID-19 response team | Erasmus Medical Center |
| EPI_ISL_460656 | Dutch COVID-19 response team | Erasmus Medical Center |

| EPI_Accession_ID | Originating Laboratory | Submitting Laboratory |
|---|---|---|
| EPI_ISL_460657 | Dutch COVID-19 response team | Erasmus Medical Center |
| EPI_ISL_460658 | Dutch COVID-19 response team | Erasmus Medical Center |
| EPI_ISL_460659 | Dutch COVID-19 response team | Erasmus Medical Center |
| EPI_ISL_460660 | Dutch COVID-19 response team | Erasmus Medical Center |
| EPI_ISL_460661 | Dutch COVID-19 response team | Erasmus Medical Center |
| EPI_ISL_460662 | Dutch COVID-19 response team | Erasmus Medical Center |
| EPI_ISL_460663 | Dutch COVID-19 response team | Erasmus Medical Center |
| EPI_ISL_460664 | Dutch COVID-19 response team | Erasmus Medical Center |
| EPI_ISL_460665 | Dutch COVID-19 response team | Erasmus Medical Center |
| EPI_ISL_460666 | Dutch COVID-19 response team | Erasmus Medical Center |
| EPI_ISL_460667 | Dutch COVID-19 response team | Erasmus Medical Center |
| EPI_ISL_460668 | Dutch COVID-19 response team | Erasmus Medical Center |
| EPI_ISL_460669 | Dutch COVID-19 response team | Erasmus Medical Center |
| EPI_ISL_460670 | Dutch COVID-19 response team | Erasmus Medical Center |
| EPI_ISL_460671 | Dutch COVID-19 response team | Erasmus Medical Center |
| EPI_ISL_460672 | Dutch COVID-19 response team | Erasmus Medical Center |
| EPI_ISL_460673 | Dutch COVID-19 response team | Erasmus Medical Center |
| EPI_ISL_460674 | Dutch COVID-19 response team | Erasmus Medical Center |
| EPI_ISL_460675 | Dutch COVID-19 response team | Erasmus Medical Center |
| EPI_ISL_460676 | Dutch COVID-19 response team | Erasmus Medical Center |
| EPI_ISL_460677 | Dutch COVID-19 response team | Erasmus Medical Center |
| EPI_ISL_460678 | Dutch COVID-19 response team | Erasmus Medical Center |
| EPI_ISL_460679 | Dutch COVID-19 response team | Erasmus Medical Center |
| EPI_ISL_460680 | Dutch COVID-19 response team | Erasmus Medical Center |
| EPI_ISL_460681 | Dutch COVID-19 response team | Erasmus Medical Center |
| EPI_ISL_460682 | Dutch COVID-19 response team | Erasmus Medical Center |
| EPI_ISL_460683 | Dutch COVID-19 response team | Erasmus Medical Center |
| EPI_ISL_460684 | Dutch COVID-19 response team | Erasmus Medical Center |
| EPI_ISL_460685 | Dutch COVID-19 response team | Erasmus Medical Center |
| EPI_ISL_460686 | Dutch COVID-19 response team | Erasmus Medical Center |
| EPI_ISL_460687 | Dutch COVID-19 response team | Erasmus Medical Center |
| EPI_ISL_460688 | Dutch COVID-19 response team | Erasmus Medical Center |
| EPI_ISL_460689 | Dutch COVID-19 response team | Erasmus Medical Center |

| EPI_Accession_ID | Originating Laboratory | Submitting Laboratory |
|---|---|---|
| EPI_ISL_460690 | Dutch COVID-19 response team | Erasmus Medical Center |
| EPI_ISL_460691 | Dutch COVID-19 response team | Erasmus Medical Center |
| EPI_ISL_460692 | Dutch COVID-19 response team | Erasmus Medical Center |
| EPI_ISL_460693 | Dutch COVID-19 response team | Erasmus Medical Center |
| EPI_ISL_460694 | Dutch COVID-19 response team | Erasmus Medical Center |
| EPI_ISL_460695 | Dutch COVID-19 response team | Erasmus Medical Center |
| EPI_ISL_460696 | Dutch COVID-19 response team | Erasmus Medical Center |
| EPI_ISL_460697 | Dutch COVID-19 response team | Erasmus Medical Center |
| EPI_ISL_460698 | Dutch COVID-19 response team | Erasmus Medical Center |
| EPI_ISL_460699 | Dutch COVID-19 response team | Erasmus Medical Center |
| EPI_ISL_460700 | Dutch COVID-19 response team | Erasmus Medical Center |
| EPI_ISL_460701 | Dutch COVID-19 response team | Erasmus Medical Center |
| EPI_ISL_460702 | Dutch COVID-19 response team | Erasmus Medical Center |
| EPI_ISL_460703 | Dutch COVID-19 response team | Erasmus Medical Center |
| EPI_ISL_460704 | Dutch COVID-19 response team | Erasmus Medical Center |
| EPI_ISL_460705 | Dutch COVID-19 response team | Erasmus Medical Center |
| EPI_ISL_460714 | Dutch COVID-19 response team | Erasmus Medical Center |
| EPI_ISL_460715 | Dutch COVID-19 response team | Erasmus Medical Center |
| EPI_ISL_460716 | Dutch COVID-19 response team | Erasmus Medical Center |
| EPI_ISL_460717 | Dutch COVID-19 response team | Erasmus Medical Center |
| EPI_ISL_460718 | Dutch COVID-19 response team | Erasmus Medical Center |
| EPI_ISL_460719 | Dutch COVID-19 response team | Erasmus Medical Center |
| EPI_ISL_460720 | Dutch COVID-19 response team | Erasmus Medical Center |
| EPI_ISL_460721 | Dutch COVID-19 response team | Erasmus Medical Center |
| EPI_ISL_460723 | Dutch COVID-19 response team | Erasmus Medical Center |
| EPI_ISL_460724 | Dutch COVID-19 response team | Erasmus Medical Center |
| EPI_ISL_460725 | Dutch COVID-19 response team | Erasmus Medical Center |
| EPI_ISL_460726 | Dutch COVID-19 response team | Erasmus Medical Center |
| EPI_ISL_460727 | Dutch COVID-19 response team | Erasmus Medical Center |
| EPI_ISL_460728 | Dutch COVID-19 response team | Erasmus Medical Center |
| EPI_ISL_460729 | Dutch COVID-19 response team | Erasmus Medical Center |
| EPI_ISL_460730 | Dutch COVID-19 response team | Erasmus Medical Center |
| EPI_ISL_460731 | Dutch COVID-19 response team | Erasmus Medical Center |

| EPI_Accession_ID | Originating Laboratory | Submitting Laboratory |
|---|---|---|
| EPI_ISL_460732 | Dutch COVID-19 response team | Erasmus Medical Center |
| EPI_ISL_460733 | Dutch COVID-19 response team | Erasmus Medical Center |
| EPI_ISL_460734 | Dutch COVID-19 response team | Erasmus Medical Center |
| EPI_ISL_460735 | Dutch COVID-19 response team | Erasmus Medical Center |
| EPI_ISL_460736 | Dutch COVID-19 response team | Erasmus Medical Center |
| EPI_ISL_460737 | Dutch COVID-19 response team | Erasmus Medical Center |
| EPI_ISL_460738 | Dutch COVID-19 response team | Erasmus Medical Center |
| EPI_ISL_460753 | Dutch COVID-19 response team | Erasmus Medical Center |
| EPI_ISL_460754 | Dutch COVID-19 response team | Erasmus Medical Center |
| EPI_ISL_460755 | Dutch COVID-19 response team | Erasmus Medical Center |
| EPI_ISL_460756 | Dutch COVID-19 response team | Erasmus Medical Center |
| EPI_ISL_460757 | Dutch COVID-19 response team | Erasmus Medical Center |
| EPI_ISL_460758 | Dutch COVID-19 response team | Erasmus Medical Center |
| EPI_ISL_460759 | Dutch COVID-19 response team | Erasmus Medical Center |
| EPI_ISL_460760 | Dutch COVID-19 response team | Erasmus Medical Center |
| EPI_ISL_460761 | Dutch COVID-19 response team | Erasmus Medical Center |
| EPI_ISL_460762 | Dutch COVID-19 response team | Erasmus Medical Center |
| EPI_ISL_460763 | Dutch COVID-19 response team | Erasmus Medical Center |
| EPI_ISL_460764 | Dutch COVID-19 response team | Erasmus Medical Center |
| EPI_ISL_460765 | Dutch COVID-19 response team | Erasmus Medical Center |
| EPI_ISL_460766 | Dutch COVID-19 response team | Erasmus Medical Center |
| EPI_ISL_460767 | Dutch COVID-19 response team | Erasmus Medical Center |
| EPI_ISL_460768 | Dutch COVID-19 response team | Erasmus Medical Center |
| EPI_ISL_460770 | Dutch COVID-19 response team | Erasmus Medical Center |
| EPI_ISL_460771 | Dutch COVID-19 response team | Erasmus Medical Center |
| EPI_ISL_460773 | Dutch COVID-19 response team | Erasmus Medical Center |
| EPI_ISL_460774 | Dutch COVID-19 response team | Erasmus Medical Center |
| EPI_ISL_460775 | Dutch COVID-19 response team | Erasmus Medical Center |
| EPI_ISL_460776 | Dutch COVID-19 response team | Erasmus Medical Center |
| EPI_ISL_460777 | Dutch COVID-19 response team | Erasmus Medical Center |
| EPI_ISL_460778 | Dutch COVID-19 response team | Erasmus Medical Center |
| EPI_ISL_460779 | Dutch COVID-19 response team | Erasmus Medical Center |
| EPI_ISL_460780 | Dutch COVID-19 response team | Erasmus Medical Center |

| EPI_Accession_ID | Originating Laboratory | Submitting Laboratory |
|---|---|---|
| EPI_ISL_460781 | Dutch COVID-19 response team | Erasmus Medical Center |
| EPI_ISL_460782 | Dutch COVID-19 response team | Erasmus Medical Center |
| EPI_ISL_460783 | Dutch COVID-19 response team | Erasmus Medical Center |
| EPI_ISL_460784 | Dutch COVID-19 response team | Erasmus Medical Center |
| EPI_ISL_460785 | Dutch COVID-19 response team | Erasmus Medical Center |
| EPI_ISL_460786 | Dutch COVID-19 response team | Erasmus Medical Center |
| EPI_ISL_460787 | Dutch COVID-19 response team | Erasmus Medical Center |
| EPI_ISL_460788 | Dutch COVID-19 response team | Erasmus Medical Center |
| EPI_ISL_460789 | Dutch COVID-19 response team | Erasmus Medical Center |
| EPI_ISL_460790 | Dutch COVID-19 response team | Erasmus Medical Center |
| EPI_ISL_460791 | Dutch COVID-19 response team | Erasmus Medical Center |
| EPI_ISL_460792 | Dutch COVID-19 response team | Erasmus Medical Center |
| EPI_ISL_460793 | Dutch COVID-19 response team | Erasmus Medical Center |
| EPI_ISL_460794 | Dutch COVID-19 response team | Erasmus Medical Center |
| EPI_ISL_460795 | Dutch COVID-19 response team | Erasmus Medical Center |
| EPI_ISL_460796 | Dutch COVID-19 response team | Erasmus Medical Center |
| EPI_ISL_460797 | Dutch COVID-19 response team | Erasmus Medical Center |
| EPI_ISL_460798 | Dutch COVID-19 response team | Erasmus Medical Center |
| EPI_ISL_460799 | Dutch COVID-19 response team | Erasmus Medical Center |
| EPI_ISL_460800 | Dutch COVID-19 response team | Erasmus Medical Center |
| EPI_ISL_460801 | Dutch COVID-19 response team | Erasmus Medical Center |
| EPI_ISL_460802 | Dutch COVID-19 response team | Erasmus Medical Center |
| EPI_ISL_460803 | Dutch COVID-19 response team | Erasmus Medical Center |
| EPI_ISL_460804 | Dutch COVID-19 response team | Erasmus Medical Center |
| EPI_ISL_460805 | Dutch COVID-19 response team | Erasmus Medical Center |
| EPI_ISL_460806 | Dutch COVID-19 response team | Erasmus Medical Center |
| EPI_ISL_460807 | Dutch COVID-19 response team | Erasmus Medical Center |
| EPI_ISL_460808 | Dutch COVID-19 response team | Erasmus Medical Center |
| EPI_ISL_460809 | Dutch COVID-19 response team | Erasmus Medical Center |
| EPI_ISL_460810 | Dutch COVID-19 response team | Erasmus Medical Center |
| EPI_ISL_460811 | Dutch COVID-19 response team | Erasmus Medical Center |
| EPI_ISL_460812 | Dutch COVID-19 response team | Erasmus Medical Center |
| EPI_ISL_460813 | Dutch COVID-19 response team | Erasmus Medical Center |

| EPI_Accession_ID | Originating Laboratory | Submitting Laboratory |
|---|---|---|
| EPI_ISL_460814 | Dutch COVID-19 response team | Erasmus Medical Center |
| EPI_ISL_460815 | Dutch COVID-19 response team | Erasmus Medical Center |
| EPI_ISL_460816 | Dutch COVID-19 response team | Erasmus Medical Center |
| EPI_ISL_460817 | Dutch COVID-19 response team | Erasmus Medical Center |
| EPI_ISL_460818 | Dutch COVID-19 response team | Erasmus Medical Center |
| EPI_ISL_460819 | Dutch COVID-19 response team | Erasmus Medical Center |
| EPI_ISL_460820 | Dutch COVID-19 response team | Erasmus Medical Center |
| EPI_ISL_460821 | Dutch COVID-19 response team | Erasmus Medical Center |
| EPI_ISL_460823 | Dutch COVID-19 response team | Erasmus Medical Center |
| EPI_ISL_460824 | Dutch COVID-19 response team | Erasmus Medical Center |
| EPI_ISL_460825 | Dutch COVID-19 response team | Erasmus Medical Center |
| EPI_ISL_460826 | Dutch COVID-19 response team | Erasmus Medical Center |
| EPI_ISL_460827 | Dutch COVID-19 response team | Erasmus Medical Center |
| EPI_ISL_460828 | Dutch COVID-19 response team | Erasmus Medical Center |
| EPI_ISL_460829 | Dutch COVID-19 response team | Erasmus Medical Center |
| EPI_ISL_460830 | Dutch COVID-19 response team | Erasmus Medical Center |
| EPI_ISL_460831 | Dutch COVID-19 response team | Erasmus Medical Center |
| EPI_ISL_460832 | Dutch COVID-19 response team | Erasmus Medical Center |
| EPI_ISL_460833 | Dutch COVID-19 response team | Erasmus Medical Center |
| EPI_ISL_460834 | Dutch COVID-19 response team | Erasmus Medical Center |
| EPI_ISL_460835 | Dutch COVID-19 response team | Erasmus Medical Center |
| EPI_ISL_460836 | Dutch COVID-19 response team | Erasmus Medical Center |
| EPI_ISL_460837 | Dutch COVID-19 response team | Erasmus Medical Center |
| EPI_ISL_460838 | Dutch COVID-19 response team | Erasmus Medical Center |
| EPI_ISL_460839 | Dutch COVID-19 response team | Erasmus Medical Center |
| EPI_ISL_460840 | Dutch COVID-19 response team | Erasmus Medical Center |
| EPI_ISL_460841 | Dutch COVID-19 response team | Erasmus Medical Center |
| EPI_ISL_460842 | Dutch COVID-19 response team | Erasmus Medical Center |
| EPI_ISL_460843 | Dutch COVID-19 response team | Erasmus Medical Center |
| EPI_ISL_460844 | Dutch COVID-19 response team | Erasmus Medical Center |
| EPI_ISL_460845 | Dutch COVID-19 response team | Erasmus Medical Center |
| EPI_ISL_460846 | Dutch COVID-19 response team | Erasmus Medical Center |
| EPI_ISL_460847 | Dutch COVID-19 response team | Erasmus Medical Center |

| EPI_Accession_ID | Originating Laboratory | Submitting Laboratory |
|---|---|---|
| EPI_ISL_460848 | Dutch COVID-19 response team | Erasmus Medical Center |
| EPI_ISL_460849 | Dutch COVID-19 response team | Erasmus Medical Center |
| EPI_ISL_460850 | Dutch COVID-19 response team | Erasmus Medical Center |
| EPI_ISL_460851 | Dutch COVID-19 response team | Erasmus Medical Center |
| EPI_ISL_460852 | Dutch COVID-19 response team | Erasmus Medical Center |
| EPI_ISL_460853 | Dutch COVID-19 response team | Erasmus Medical Center |
| EPI_ISL_460854 | Dutch COVID-19 response team | Erasmus Medical Center |
| EPI_ISL_460855 | Dutch COVID-19 response team | Erasmus Medical Center |
| EPI_ISL_460856 | Dutch COVID-19 response team | Erasmus Medical Center |
| EPI_ISL_460857 | Dutch COVID-19 response team | Erasmus Medical Center |
| EPI_ISL_460858 | Dutch COVID-19 response team | Erasmus Medical Center |
| EPI_ISL_460859 | Dutch COVID-19 response team | Erasmus Medical Center |
| EPI_ISL_460860 | Dutch COVID-19 response team | Erasmus Medical Center |
| EPI_ISL_460861 | Dutch COVID-19 response team | Erasmus Medical Center |
| EPI_ISL_460862 | Dutch COVID-19 response team | Erasmus Medical Center |
| EPI_ISL_460863 | Dutch COVID-19 response team | Erasmus Medical Center |
| EPI_ISL_460864 | Dutch COVID-19 response team | Erasmus Medical Center |
| EPI_ISL_460865 | Dutch COVID-19 response team | Erasmus Medical Center |
| EPI_ISL_460866 | Dutch COVID-19 response team | Erasmus Medical Center |
| EPI_ISL_460867 | Dutch COVID-19 response team | Erasmus Medical Center |
| EPI_ISL_460868 | Dutch COVID-19 response team | Erasmus Medical Center |
| EPI_ISL_460869 | Dutch COVID-19 response team | Erasmus Medical Center |
| EPI_ISL_460870 | Dutch COVID-19 response team | Erasmus Medical Center |
| EPI_ISL_460871 | Dutch COVID-19 response team | Erasmus Medical Center |
| EPI_ISL_460872 | Dutch COVID-19 response team | Erasmus Medical Center |
| EPI_ISL_460873 | Dutch COVID-19 response team | Erasmus Medical Center |
| EPI_ISL_460874 | Dutch COVID-19 response team | Erasmus Medical Center |
| EPI_ISL_460875 | Dutch COVID-19 response team | Erasmus Medical Center |
| EPI_ISL_460876 | Dutch COVID-19 response team | Erasmus Medical Center |
| EPI_ISL_460877 | Dutch COVID-19 response team | Erasmus Medical Center |
| EPI_ISL_460878 | Dutch COVID-19 response team | Erasmus Medical Center |
| EPI_ISL_460879 | Dutch COVID-19 response team | Erasmus Medical Center |
| EPI_ISL_460880 | Dutch COVID-19 response team | Erasmus Medical Center |

| EPI_Accession_ID | Originating Laboratory | Submitting Laboratory |
|---|---|---|
| EPI_ISL_460881 | Dutch COVID-19 response team | Erasmus Medical Center |
| EPI_ISL_460882 | Dutch COVID-19 response team | Erasmus Medical Center |
| EPI_ISL_460883 | Dutch COVID-19 response team | Erasmus Medical Center |
| EPI_ISL_460884 | Dutch COVID-19 response team | Erasmus Medical Center |
| EPI_ISL_460885 | Dutch COVID-19 response team | Erasmus Medical Center |
| EPI_ISL_460886 | Dutch COVID-19 response team | Erasmus Medical Center |
| EPI_ISL_460887 | Dutch COVID-19 response team | Erasmus Medical Center |
| EPI_ISL_460888 | Dutch COVID-19 response team | Erasmus Medical Center |
| EPI_ISL_460889 | Dutch COVID-19 response team | Erasmus Medical Center |
| EPI_ISL_460890 | Dutch COVID-19 response team | Erasmus Medical Center |
| EPI_ISL_460892 | Dutch COVID-19 response team | Erasmus Medical Center |
| EPI_ISL_460893 | Dutch COVID-19 response team | Erasmus Medical Center |
| EPI_ISL_460894 | Dutch COVID-19 response team | Erasmus Medical Center |
| EPI_ISL_460895 | Dutch COVID-19 response team | Erasmus Medical Center |
| EPI_ISL_460896 | Dutch COVID-19 response team | Erasmus Medical Center |
| EPI_ISL_460897 | Dutch COVID-19 response team | Erasmus Medical Center |
| EPI_ISL_460898 | Dutch COVID-19 response team | Erasmus Medical Center |
| EPI_ISL_460899 | Dutch COVID-19 response team | Erasmus Medical Center |
| EPI_ISL_460900 | Dutch COVID-19 response team | Erasmus Medical Center |
| EPI_ISL_460901 | Dutch COVID-19 response team | Erasmus Medical Center |
| EPI_ISL_460902 | Dutch COVID-19 response team | Erasmus Medical Center |
| EPI_ISL_460903 | Dutch COVID-19 response team | Erasmus Medical Center |
| EPI_ISL_460904 | Dutch COVID-19 response team | Erasmus Medical Center |
| EPI_ISL_460905 | Dutch COVID-19 response team | Erasmus Medical Center |
| EPI_ISL_460906 | Dutch COVID-19 response team | Erasmus Medical Center |
| EPI_ISL_460907 | Dutch COVID-19 response team | Erasmus Medical Center |
| EPI_ISL_460908 | Dutch COVID-19 response team | Erasmus Medical Center |
| EPI_ISL_460909 | Dutch COVID-19 response team | Erasmus Medical Center |
| EPI_ISL_460910 | Dutch COVID-19 response team | Erasmus Medical Center |
| EPI_ISL_460911 | Dutch COVID-19 response team | Erasmus Medical Center |
| EPI_ISL_460912 | Dutch COVID-19 response team | Erasmus Medical Center |
| EPI_ISL_460913 | Dutch COVID-19 response team | Erasmus Medical Center |
| EPI_ISL_460914 | Dutch COVID-19 response team | Erasmus Medical Center |

| EPI_Accession_ID | Originating Laboratory | Submitting Laboratory |
|---|---|---|
| EPI_ISL_460915 | Dutch COVID-19 response team | Erasmus Medical Center |
| EPI_ISL_460916 | Dutch COVID-19 response team | Erasmus Medical Center |
| EPI_ISL_460917 | Dutch COVID-19 response team | Erasmus Medical Center |
| EPI_ISL_460918 | Dutch COVID-19 response team | Erasmus Medical Center |
| EPI_ISL_460919 | Dutch COVID-19 response team | Erasmus Medical Center |
| EPI_ISL_460920 | Dutch COVID-19 response team | Erasmus Medical Center |
| EPI_ISL_460921 | Dutch COVID-19 response team | Erasmus Medical Center |
| EPI_ISL_460922 | Dutch COVID-19 response team | Erasmus Medical Center |
| EPI_ISL_460923 | Dutch COVID-19 response team | Erasmus Medical Center |
| EPI_ISL_460924 | Dutch COVID-19 response team | Erasmus Medical Center |
| EPI_ISL_460925 | Dutch COVID-19 response team | Erasmus Medical Center |
| EPI_ISL_460926 | Dutch COVID-19 response team | Erasmus Medical Center |
| EPI_ISL_460927 | Dutch COVID-19 response team | Erasmus Medical Center |
| EPI_ISL_460928 | Dutch COVID-19 response team | Erasmus Medical Center |
| EPI_ISL_460929 | Dutch COVID-19 response team | Erasmus Medical Center |
| EPI_ISL_460930 | Dutch COVID-19 response team | Erasmus Medical Center |
| EPI_ISL_460931 | Dutch COVID-19 response team | Erasmus Medical Center |
| EPI_ISL_460932 | Dutch COVID-19 response team | Erasmus Medical Center |
| EPI_ISL_460933 | Dutch COVID-19 response team | Erasmus Medical Center |
| EPI_ISL_460934 | Dutch COVID-19 response team | Erasmus Medical Center |
| EPI_ISL_460935 | Dutch COVID-19 response team | Erasmus Medical Center |
| EPI_ISL_460936 | Dutch COVID-19 response team | Erasmus Medical Center |
| EPI_ISL_460937 | Dutch COVID-19 response team | Erasmus Medical Center |
| EPI_ISL_460938 | Dutch COVID-19 response team | Erasmus Medical Center |
| EPI_ISL_460939 | Dutch COVID-19 response team | Erasmus Medical Center |
| EPI_ISL_460940 | Dutch COVID-19 response team | Erasmus Medical Center |
| EPI_ISL_460941 | Dutch COVID-19 response team | Erasmus Medical Center |
| EPI_ISL_460942 | Dutch COVID-19 response team | Erasmus Medical Center |
| EPI_ISL_460943 | Dutch COVID-19 response team | Erasmus Medical Center |
| EPI_ISL_460944 | Dutch COVID-19 response team | Erasmus Medical Center |
| EPI_ISL_460945 | Dutch COVID-19 response team | Erasmus Medical Center |
| EPI_ISL_460946 | Dutch COVID-19 response team | Erasmus Medical Center |
| EPI_ISL_460947 | Dutch COVID-19 response team | Erasmus Medical Center |

| EPI_Accession_ID | Originating Laboratory | Submitting Laboratory |
|---|---|---|
| EPI_ISL_460948 | Dutch COVID-19 response team | Erasmus Medical Center |
| EPI_ISL_460949 | Dutch COVID-19 response team | Erasmus Medical Center |
| EPI_ISL_460950 | Dutch COVID-19 response team | Erasmus Medical Center |
| EPI_ISL_460951 | Dutch COVID-19 response team | Erasmus Medical Center |
| EPI_ISL_460952 | Dutch COVID-19 response team | Erasmus Medical Center |
| EPI_ISL_460953 | Dutch COVID-19 response team | Erasmus Medical Center |
| EPI_ISL_460954 | Dutch COVID-19 response team | Erasmus Medical Center |
| EPI_ISL_460955 | Dutch COVID-19 response team | Erasmus Medical Center |
| EPI_ISL_460956 | Dutch COVID-19 response team | Erasmus Medical Center |
| EPI_ISL_460957 | Dutch COVID-19 response team | Erasmus Medical Center |
| EPI_ISL_460958 | Dutch COVID-19 response team | Erasmus Medical Center |
| EPI_ISL_460959 | Dutch COVID-19 response team | Erasmus Medical Center |
| EPI_ISL_460960 | Dutch COVID-19 response team | Erasmus Medical Center |
| EPI_ISL_460961 | Dutch COVID-19 response team | Erasmus Medical Center |
| EPI_ISL_460962 | Dutch COVID-19 response team | Erasmus Medical Center |
| EPI_ISL_460963 | Dutch COVID-19 response team | Erasmus Medical Center |
| EPI_ISL_460964 | Dutch COVID-19 response team | Erasmus Medical Center |
| EPI_ISL_460965 | Dutch COVID-19 response team | Erasmus Medical Center |
| EPI_ISL_460966 | Dutch COVID-19 response team | Erasmus Medical Center |
| EPI_ISL_460967 | Dutch COVID-19 response team | Erasmus Medical Center |
| EPI_ISL_460968 | Dutch COVID-19 response team | Erasmus Medical Center |
| EPI_ISL_460969 | Dutch COVID-19 response team | Erasmus Medical Center |
| EPI_ISL_460970 | Dutch COVID-19 response team | Erasmus Medical Center |
| EPI_ISL_460971 | Dutch COVID-19 response team | Erasmus Medical Center |
| EPI_ISL_460972 | Dutch COVID-19 response team | Erasmus Medical Center |
| EPI_ISL_460973 | Dutch COVID-19 response team | Erasmus Medical Center |
| EPI_ISL_460974 | Dutch COVID-19 response team | Erasmus Medical Center |
| EPI_ISL_460975 | Dutch COVID-19 response team | Erasmus Medical Center |
| EPI_ISL_460976 | Dutch COVID-19 response team | Erasmus Medical Center |
| EPI_ISL_460977 | Dutch COVID-19 response team | Erasmus Medical Center |
| EPI_ISL_460978 | Dutch COVID-19 response team | Erasmus Medical Center |
| EPI_ISL_460979 | Dutch COVID-19 response team | Erasmus Medical Center |
| EPI_ISL_460980 | Dutch COVID-19 response team | Erasmus Medical Center |

| EPI_Accession_ID | Originating Laboratory | Submitting Laboratory |
|---|---|---|
| EPI_ISL_460981 | Dutch COVID-19 response team | Erasmus Medical Center |
| EPI_ISL_460982 | Dutch COVID-19 response team | Erasmus Medical Center |
| EPI_ISL_460983 | Dutch COVID-19 response team | Erasmus Medical Center |
| EPI_ISL_460984 | Dutch COVID-19 response team | Erasmus Medical Center |
| EPI_ISL_460985 | Dutch COVID-19 response team | Erasmus Medical Center |
| EPI_ISL_460986 | Dutch COVID-19 response team | Erasmus Medical Center |
| EPI_ISL_460987 | Dutch COVID-19 response team | Erasmus Medical Center |
| EPI_ISL_460988 | Dutch COVID-19 response team | Erasmus Medical Center |
| EPI_ISL_460989 | Dutch COVID-19 response team | Erasmus Medical Center |
| EPI_ISL_460990 | Dutch COVID-19 response team | Erasmus Medical Center |
| EPI_ISL_460991 | Dutch COVID-19 response team | Erasmus Medical Center |
| EPI_ISL_460992 | Dutch COVID-19 response team | Erasmus Medical Center |
| EPI_ISL_460993 | Dutch COVID-19 response team | Erasmus Medical Center |
| EPI_ISL_460994 | Dutch COVID-19 response team | Erasmus Medical Center |
| EPI_ISL_460995 | Dutch COVID-19 response team | Erasmus Medical Center |
| EPI_ISL_460996 | Dutch COVID-19 response team | Erasmus Medical Center |
| EPI_ISL_460997 | Dutch COVID-19 response team | Erasmus Medical Center |
| EPI_ISL_460998 | Dutch COVID-19 response team | Erasmus Medical Center |
| EPI_ISL_460999 | Dutch COVID-19 response team | Erasmus Medical Center |
| EPI_ISL_461000 | Dutch COVID-19 response team | Erasmus Medical Center |
| EPI_ISL_461001 | Dutch COVID-19 response team | Erasmus Medical Center |
| EPI_ISL_461002 | Dutch COVID-19 response team | Erasmus Medical Center |
| EPI_ISL_461003 | Dutch COVID-19 response team | Erasmus Medical Center |
| EPI_ISL_461004 | Dutch COVID-19 response team | Erasmus Medical Center |
| EPI_ISL_461005 | Dutch COVID-19 response team | Erasmus Medical Center |
| EPI_ISL_461006 | Dutch COVID-19 response team | Erasmus Medical Center |
| EPI_ISL_461007 | Dutch COVID-19 response team | Erasmus Medical Center |
| EPI_ISL_461008 | Dutch COVID-19 response team | Erasmus Medical Center |
| EPI_ISL_461009 | Dutch COVID-19 response team | Erasmus Medical Center |
| EPI_ISL_461010 | Dutch COVID-19 response team | Erasmus Medical Center |
| EPI_ISL_461011 | Dutch COVID-19 response team | Erasmus Medical Center |
| EPI_ISL_461012 | Dutch COVID-19 response team | Erasmus Medical Center |
| EPI_ISL_461013 | Dutch COVID-19 response team | Erasmus Medical Center |

| EPI_Accession_ID | Originating Laboratory | Submitting Laboratory |
|---|---|---|
| EPI_ISL_461014 | Dutch COVID-19 response team | Erasmus Medical Center |
| EPI_ISL_461015 | Dutch COVID-19 response team | Erasmus Medical Center |
| EPI_ISL_461016 | Dutch COVID-19 response team | Erasmus Medical Center |
| EPI_ISL_461017 | Dutch COVID-19 response team | Erasmus Medical Center |
| EPI_ISL_461018 | Dutch COVID-19 response team | Erasmus Medical Center |
| EPI_ISL_461019 | Dutch COVID-19 response team | Erasmus Medical Center |
| EPI_ISL_461020 | Dutch COVID-19 response team | Erasmus Medical Center |
| EPI_ISL_461021 | Dutch COVID-19 response team | Erasmus Medical Center |
| EPI_ISL_461022 | Dutch COVID-19 response team | Erasmus Medical Center |
| EPI_ISL_461023 | Dutch COVID-19 response team | Erasmus Medical Center |
| EPI_ISL_461024 | Dutch COVID-19 response team | Erasmus Medical Center |
| EPI_ISL_461025 | Dutch COVID-19 response team | Erasmus Medical Center |
| EPI_ISL_461026 | Dutch COVID-19 response team | Erasmus Medical Center |
| EPI_ISL_461027 | Dutch COVID-19 response team | Erasmus Medical Center |
| EPI_ISL_461028 | Dutch COVID-19 response team | Erasmus Medical Center |
| EPI_ISL_461029 | Dutch COVID-19 response team | Erasmus Medical Center |
| EPI_ISL_461030 | Dutch COVID-19 response team | Erasmus Medical Center |
| EPI_ISL_461031 | Dutch COVID-19 response team | Erasmus Medical Center |
| EPI_ISL_461032 | Dutch COVID-19 response team | Erasmus Medical Center |
| EPI_ISL_461033 | Dutch COVID-19 response team | Erasmus Medical Center |
| EPI_ISL_461034 | Dutch COVID-19 response team | Erasmus Medical Center |
| EPI_ISL_461035 | Dutch COVID-19 response team | Erasmus Medical Center |
| EPI_ISL_461036 | Dutch COVID-19 response team | Erasmus Medical Center |
| EPI_ISL_461037 | Dutch COVID-19 response team | Erasmus Medical Center |
| EPI_ISL_461038 | Dutch COVID-19 response team | Erasmus Medical Center |
| EPI_ISL_461039 | Dutch COVID-19 response team | Erasmus Medical Center |
| EPI_ISL_461040 | Dutch COVID-19 response team | Erasmus Medical Center |
| EPI_ISL_461041 | Dutch COVID-19 response team | Erasmus Medical Center |
| EPI_ISL_461042 | Dutch COVID-19 response team | Erasmus Medical Center |
| EPI_ISL_461043 | Dutch COVID-19 response team | Erasmus Medical Center |
| EPI_ISL_461044 | Dutch COVID-19 response team | Erasmus Medical Center |
| EPI_ISL_461045 | Dutch COVID-19 response team | Erasmus Medical Center |
| EPI_ISL_461046 | Dutch COVID-19 response team | Erasmus Medical Center |

| EPI_Accession_ID | Originating Laboratory | Submitting Laboratory |
|---|---|---|
| EPI_ISL_461047 | Dutch COVID-19 response team | Erasmus Medical Center |
| EPI_ISL_461048 | Dutch COVID-19 response team | Erasmus Medical Center |
| EPI_ISL_461049 | Dutch COVID-19 response team | Erasmus Medical Center |
| EPI_ISL_461050 | Dutch COVID-19 response team | Erasmus Medical Center |
| EPI_ISL_461061 | Dutch COVID-19 response team | Erasmus Medical Center |
| EPI_ISL_461062 | Dutch COVID-19 response team | Erasmus Medical Center |
| EPI_ISL_461063 | Dutch COVID-19 response team | Erasmus Medical Center |
| EPI_ISL_461064 | Dutch COVID-19 response team | Erasmus Medical Center |
| EPI_ISL_461065 | Dutch COVID-19 response team | Erasmus Medical Center |
| EPI_ISL_461066 | Dutch COVID-19 response team | Erasmus Medical Center |
| EPI_ISL_461067 | Dutch COVID-19 response team | Erasmus Medical Center |
| EPI_ISL_461068 | Dutch COVID-19 response team | Erasmus Medical Center |
| EPI_ISL_461069 | Dutch COVID-19 response team | Erasmus Medical Center |
| EPI_ISL_461070 | Dutch COVID-19 response team | Erasmus Medical Center |
| EPI_ISL_461071 | Dutch COVID-19 response team | Erasmus Medical Center |
| EPI_ISL_461072 | Dutch COVID-19 response team | Erasmus Medical Center |
| EPI_ISL_461073 | Dutch COVID-19 response team | Erasmus Medical Center |
| EPI_ISL_461074 | Dutch COVID-19 response team | Erasmus Medical Center |
| EPI_ISL_461075 | Dutch COVID-19 response team | Erasmus Medical Center |
| EPI_ISL_461076 | Dutch COVID-19 response team | Erasmus Medical Center |
| EPI_ISL_461077 | Dutch COVID-19 response team | Erasmus Medical Center |
| EPI_ISL_461078 | Dutch COVID-19 response team | Erasmus Medical Center |
| EPI_ISL_461079 | Dutch COVID-19 response team | Erasmus Medical Center |
| EPI_ISL_461080 | Dutch COVID-19 response team | Erasmus Medical Center |
| EPI_ISL_461081 | Dutch COVID-19 response team | Erasmus Medical Center |
| EPI_ISL_461082 | Dutch COVID-19 response team | Erasmus Medical Center |
| EPI_ISL_461083 | Dutch COVID-19 response team | Erasmus Medical Center |
| EPI_ISL_461084 | Dutch COVID-19 response team | Erasmus Medical Center |
| EPI_ISL_461085 | Dutch COVID-19 response team | Erasmus Medical Center |
| EPI_ISL_461086 | Dutch COVID-19 response team | Erasmus Medical Center |
| EPI_ISL_461087 | Dutch COVID-19 response team | Erasmus Medical Center |
| EPI_ISL_461101 | Dutch COVID-19 response team | Erasmus Medical Center |
| EPI_ISL_461102 | Dutch COVID-19 response team | Erasmus Medical Center |

| EPI_Accession_ID | Originating Laboratory | Submitting Laboratory |
|---|---|---|
| EPI_ISL_461103 | Dutch COVID-19 response team | Erasmus Medical Center |
| EPI_ISL_461104 | Dutch COVID-19 response team | Erasmus Medical Center |
| EPI_ISL_461105 | Dutch COVID-19 response team | Erasmus Medical Center |
| EPI_ISL_461106 | Dutch COVID-19 response team | Erasmus Medical Center |
| EPI_ISL_461107 | Dutch COVID-19 response team | Erasmus Medical Center |
| EPI_ISL_461108 | Dutch COVID-19 response team | Erasmus Medical Center |
| EPI_ISL_461109 | Dutch COVID-19 response team | Erasmus Medical Center |
| EPI_ISL_461110 | Dutch COVID-19 response team | Erasmus Medical Center |
| EPI_ISL_461111 | Dutch COVID-19 response team | Erasmus Medical Center |
| EPI_ISL_461113 | Dutch COVID-19 response team | Erasmus Medical Center |
| EPI_ISL_461114 | Dutch COVID-19 response team | Erasmus Medical Center |
| EPI_ISL_461115 | Dutch COVID-19 response team | Erasmus Medical Center |
| EPI_ISL_461116 | Dutch COVID-19 response team | Erasmus Medical Center |
| EPI_ISL_461117 | Dutch COVID-19 response team | Erasmus Medical Center |
| EPI_ISL_461118 | Dutch COVID-19 response team | Erasmus Medical Center |
| EPI_ISL_461119 | Dutch COVID-19 response team | Erasmus Medical Center |
| EPI_ISL_461120 | Dutch COVID-19 response team | Erasmus Medical Center |
| EPI_ISL_461121 | Dutch COVID-19 response team | Erasmus Medical Center |
| EPI_ISL_461122 | Dutch COVID-19 response team | Erasmus Medical Center |
| EPI_ISL_461123 | Dutch COVID-19 response team | Erasmus Medical Center |
| EPI_ISL_461124 | Dutch COVID-19 response team | Erasmus Medical Center |
| EPI_ISL_461125 | Dutch COVID-19 response team | Erasmus Medical Center |
| EPI_ISL_461126 | Dutch COVID-19 response team | Erasmus Medical Center |
| EPI_ISL_461127 | Dutch COVID-19 response team | Erasmus Medical Center |
| EPI_ISL_461128 | Dutch COVID-19 response team | Erasmus Medical Center |
| EPI_ISL_461129 | Dutch COVID-19 response team | Erasmus Medical Center |
| EPI_ISL_461130 | Dutch COVID-19 response team | Erasmus Medical Center |
| EPI_ISL_461131 | Dutch COVID-19 response team | Erasmus Medical Center |
| EPI_ISL_461132 | Dutch COVID-19 response team | Erasmus Medical Center |
| EPI_ISL_461133 | Dutch COVID-19 response team | Erasmus Medical Center |
| EPI_ISL_461134 | Dutch COVID-19 response team | Erasmus Medical Center |
| EPI_ISL_461135 | Dutch COVID-19 response team | Erasmus Medical Center |
| EPI_ISL_461136 | Dutch COVID-19 response team | Erasmus Medical Center |

| EPI_Accession_ID | Originating Laboratory | Submitting Laboratory |
|---|---|---|
| EPI_ISL_461137 | Dutch COVID-19 response team | Erasmus Medical Center |
| EPI_ISL_461138 | Dutch COVID-19 response team | Erasmus Medical Center |
| EPI_ISL_461139 | Dutch COVID-19 response team | Erasmus Medical Center |
| EPI_ISL_461140 | Dutch COVID-19 response team | Erasmus Medical Center |
| EPI_ISL_461141 | Dutch COVID-19 response team | Erasmus Medical Center |
| EPI_ISL_461142 | Dutch COVID-19 response team | Erasmus Medical Center |
| EPI_ISL_461143 | Dutch COVID-19 response team | Erasmus Medical Center |
| EPI_ISL_461144 | Dutch COVID-19 response team | Erasmus Medical Center |
| EPI_ISL_461145 | Dutch COVID-19 response team | Erasmus Medical Center |
| EPI_ISL_461146 | Dutch COVID-19 response team | Erasmus Medical Center |
| EPI_ISL_461147 | Dutch COVID-19 response team | Erasmus Medical Center |
| EPI_ISL_461148 | Dutch COVID-19 response team | Erasmus Medical Center |
| EPI_ISL_461149 | Dutch COVID-19 response team | Erasmus Medical Center |
| EPI_ISL_461150 | Dutch COVID-19 response team | Erasmus Medical Center |
| EPI_ISL_461151 | Dutch COVID-19 response team | Erasmus Medical Center |
| EPI_ISL_461152 | Dutch COVID-19 response team | Erasmus Medical Center |
| EPI_ISL_461153 | Dutch COVID-19 response team | Erasmus Medical Center |
| EPI_ISL_461154 | Dutch COVID-19 response team | Erasmus Medical Center |
| EPI_ISL_461155 | Dutch COVID-19 response team | Erasmus Medical Center |
| EPI_ISL_461156 | Dutch COVID-19 response team | Erasmus Medical Center |
| EPI_ISL_461157 | Dutch COVID-19 response team | Erasmus Medical Center |
| EPI_ISL_461158 | Dutch COVID-19 response team | Erasmus Medical Center |
| EPI_ISL_461159 | Dutch COVID-19 response team | Erasmus Medical Center |
| EPI_ISL_461161 | Dutch COVID-19 response team | Erasmus Medical Center |
| EPI_ISL_461163 | Dutch COVID-19 response team | Erasmus Medical Center |
| EPI_ISL_461164 | Dutch COVID-19 response team | Erasmus Medical Center |
| EPI_ISL_461165 | Dutch COVID-19 response team | Erasmus Medical Center |
| EPI_ISL_461166 | Dutch COVID-19 response team | Erasmus Medical Center |
| EPI_ISL_461167 | Dutch COVID-19 response team | Erasmus Medical Center |
| EPI_ISL_461172 | Dutch COVID-19 response team | Erasmus Medical Center |
| EPI_ISL_461177 | Dutch COVID-19 response team | Erasmus Medical Center |
| EPI_ISL_461179 | Dutch COVID-19 response team | Erasmus Medical Center |
| EPI_ISL_461181 | Dutch COVID-19 response team | Erasmus Medical Center |

| EPI_Accession_ID | Originating Laboratory | Submitting Laboratory |
|---|---|---|
| EPI_ISL_461182 | Dutch COVID-19 response team | Erasmus Medical Center |
| EPI_ISL_461183 | Dutch COVID-19 response team | Erasmus Medical Center |
| EPI_ISL_461184 | Dutch COVID-19 response team | Erasmus Medical Center |
| EPI_ISL_461185 | Dutch COVID-19 response team | Erasmus Medical Center |
| EPI_ISL_461186 | Dutch COVID-19 response team | Erasmus Medical Center |
| EPI_ISL_461187 | Dutch COVID-19 response team | Erasmus Medical Center |
| EPI_ISL_461188 | Dutch COVID-19 response team | Erasmus Medical Center |
| EPI_ISL_461189 | Dutch COVID-19 response team | Erasmus Medical Center |
| EPI_ISL_461193 | Dutch COVID-19 response team | Erasmus Medical Center |
| EPI_ISL_461194 | Dutch COVID-19 response team | Erasmus Medical Center |
| EPI_ISL_461195 | Dutch COVID-19 response team | Erasmus Medical Center |
| EPI_ISL_461196 | Dutch COVID-19 response team | Erasmus Medical Center |
| EPI_ISL_461197 | Dutch COVID-19 response team | Erasmus Medical Center |
| EPI_ISL_461198 | Dutch COVID-19 response team | Erasmus Medical Center |
| EPI_ISL_461199 | Dutch COVID-19 response team | Erasmus Medical Center |
| EPI_ISL_461201 | Dutch COVID-19 response team | Erasmus Medical Center |
| EPI_ISL_461202 | Dutch COVID-19 response team | Erasmus Medical Center |
| EPI_ISL_461204 | Dutch COVID-19 response team | Erasmus Medical Center |
| EPI_ISL_461205 | Dutch COVID-19 response team | Erasmus Medical Center |
| EPI_ISL_461206 | Dutch COVID-19 response team | Erasmus Medical Center |
| EPI_ISL_461207 | Dutch COVID-19 response team | Erasmus Medical Center |
| EPI_ISL_461208 | Dutch COVID-19 response team | Erasmus Medical Center |
| EPI_ISL_461209 | Dutch COVID-19 response team | Erasmus Medical Center |
| EPI_ISL_461210 | Dutch COVID-19 response team | Erasmus Medical Center |
| EPI_ISL_461211 | Dutch COVID-19 response team | Erasmus Medical Center |
| EPI_ISL_461212 | Dutch COVID-19 response team | Erasmus Medical Center |
| EPI_ISL_461213 | Dutch COVID-19 response team | Erasmus Medical Center |
| EPI_ISL_461214 | Dutch COVID-19 response team | Erasmus Medical Center |
| EPI_ISL_461215 | Dutch COVID-19 response team | Erasmus Medical Center |
| EPI_ISL_461216 | Dutch COVID-19 response team | Erasmus Medical Center |
| EPI_ISL_461217 | Dutch COVID-19 response team | Erasmus Medical Center |
| EPI_ISL_461218 | Dutch COVID-19 response team | Erasmus Medical Center |
| EPI_ISL_461219 | Dutch COVID-19 response team | Erasmus Medical Center |

| EPI_Accession_ID | Originating Laboratory | Submitting Laboratory |
|---|---|---|
| EPI_ISL_461220 | Dutch COVID-19 response team | Erasmus Medical Center |
| EPI_ISL_461221 | Dutch COVID-19 response team | Erasmus Medical Center |
| EPI_ISL_461222 | Dutch COVID-19 response team | Erasmus Medical Center |
| EPI_ISL_461223 | Dutch COVID-19 response team | Erasmus Medical Center |
| EPI_ISL_461224 | Dutch COVID-19 response team | Erasmus Medical Center |
| EPI_ISL_461225 | Dutch COVID-19 response team | Erasmus Medical Center |
| EPI_ISL_461226 | Dutch COVID-19 response team | Erasmus Medical Center |
| EPI_ISL_461227 | Dutch COVID-19 response team | Erasmus Medical Center |
| EPI_ISL_461228 | Dutch COVID-19 response team | Erasmus Medical Center |
| EPI_ISL_461229 | Dutch COVID-19 response team | Erasmus Medical Center |
| EPI_ISL_461230 | Dutch COVID-19 response team | Erasmus Medical Center |
| EPI_ISL_461231 | Dutch COVID-19 response team | Erasmus Medical Center |
| EPI_ISL_461232 | Dutch COVID-19 response team | Erasmus Medical Center |
| EPI_ISL_461234 | Dutch COVID-19 response team | Erasmus Medical Center |
| EPI_ISL_461235 | Dutch COVID-19 response team | Erasmus Medical Center |
| EPI_ISL_461236 | Dutch COVID-19 response team | Erasmus Medical Center |
| EPI_ISL_461237 | Dutch COVID-19 response team | Erasmus Medical Center |
| EPI_ISL_461238 | Dutch COVID-19 response team | Erasmus Medical Center |
| EPI_ISL_461239 | Dutch COVID-19 response team | Erasmus Medical Center |
| EPI_ISL_461240 | Dutch COVID-19 response team | Erasmus Medical Center |
| EPI_ISL_461241 | Dutch COVID-19 response team | Erasmus Medical Center |
| EPI_ISL_461242 | Dutch COVID-19 response team | Erasmus Medical Center |
| EPI_ISL_461243 | Dutch COVID-19 response team | Erasmus Medical Center |
| EPI_ISL_461244 | Dutch COVID-19 response team | Erasmus Medical Center |
| EPI_ISL_461245 | Dutch COVID-19 response team | Erasmus Medical Center |
| EPI_ISL_461246 | Dutch COVID-19 response team | Erasmus Medical Center |
| EPI_ISL_461247 | Dutch COVID-19 response team | Erasmus Medical Center |
| EPI_ISL_461248 | Dutch COVID-19 response team | Erasmus Medical Center |
| EPI_ISL_461249 | Dutch COVID-19 response team | Erasmus Medical Center |
| EPI_ISL_461250 | Dutch COVID-19 response team | Erasmus Medical Center |
| EPI_ISL_461251 | Dutch COVID-19 response team | Erasmus Medical Center |
| EPI_ISL_461252 | Dutch COVID-19 response team | Erasmus Medical Center |
| EPI_ISL_461253 | Dutch COVID-19 response team | Erasmus Medical Center |

| EPI_Accession_ID | Originating Laboratory | Submitting Laboratory |
|---|---|---|
| EPI_ISL_461254 | Dutch COVID-19 response team | Erasmus Medical Center |
| EPI_ISL_461255 | Dutch COVID-19 response team | Erasmus Medical Center |
| EPI_ISL_461256 | Dutch COVID-19 response team | Erasmus Medical Center |
| EPI_ISL_461257 | Dutch COVID-19 response team | Erasmus Medical Center |
| EPI_ISL_461258 | Dutch COVID-19 response team | Erasmus Medical Center |
| EPI_ISL_461259 | Dutch COVID-19 response team | Erasmus Medical Center |
| EPI_ISL_461260 | Dutch COVID-19 response team | Erasmus Medical Center |
| EPI_ISL_461261 | Dutch COVID-19 response team | Erasmus Medical Center |
| EPI_ISL_461262 | Dutch COVID-19 response team | Erasmus Medical Center |
| EPI_ISL_461263 | Dutch COVID-19 response team | Erasmus Medical Center |
| EPI_ISL_461264 | Dutch COVID-19 response team | Erasmus Medical Center |
| EPI_ISL_461265 | Dutch COVID-19 response team | Erasmus Medical Center |
| EPI_ISL_461266 | Dutch COVID-19 response team | Erasmus Medical Center |
| EPI_ISL_461267 | Dutch COVID-19 response team | Erasmus Medical Center |
| EPI_ISL_461268 | Dutch COVID-19 response team | Erasmus Medical Center |
| EPI_ISL_461269 | Dutch COVID-19 response team | Erasmus Medical Center |
| EPI_ISL_461270 | Dutch COVID-19 response team | Erasmus Medical Center |
| EPI_ISL_461271 | Dutch COVID-19 response team | Erasmus Medical Center |
| EPI_ISL_461272 | Dutch COVID-19 response team | Erasmus Medical Center |
| EPI_ISL_461273 | Dutch COVID-19 response team | Erasmus Medical Center |
| EPI_ISL_461274 | Dutch COVID-19 response team | Erasmus Medical Center |
| EPI_ISL_461275 | Dutch COVID-19 response team | Erasmus Medical Center |
| EPI_ISL_461276 | Dutch COVID-19 response team | Erasmus Medical Center |
| EPI_ISL_461277 | Dutch COVID-19 response team | Erasmus Medical Center |
| EPI_ISL_461278 | Dutch COVID-19 response team | Erasmus Medical Center |
| EPI_ISL_461279 | Dutch COVID-19 response team | Erasmus Medical Center |
| EPI_ISL_461280 | Dutch COVID-19 response team | Erasmus Medical Center |
| EPI_ISL_461281 | Dutch COVID-19 response team | Erasmus Medical Center |
| EPI_ISL_461282 | Dutch COVID-19 response team | Erasmus Medical Center |
| EPI_ISL_461283 | Dutch COVID-19 response team | Erasmus Medical Center |
| EPI_ISL_461284 | Dutch COVID-19 response team | Erasmus Medical Center |
| EPI_ISL_461285 | Dutch COVID-19 response team | Erasmus Medical Center |
| EPI_ISL_461286 | Dutch COVID-19 response team | Erasmus Medical Center |

| EPI_Accession_ID | Originating Laboratory | Submitting Laboratory |
| --- | --- | --- |
| EPI_ISL_461287 | Dutch COVID-19 response team | Erasmus Medical Center |
| EPI_ISL_461288 | Dutch COVID-19 response team | Erasmus Medical Center |
| EPI_ISL_461289 | Dutch COVID-19 response team | Erasmus Medical Center |
| EPI_ISL_461290 | Dutch COVID-19 response team | Erasmus Medical Center |
| EPI_ISL_461291 | Dutch COVID-19 response team | Erasmus Medical Center |
| EPI_ISL_461292 | Dutch COVID-19 response team | Erasmus Medical Center |
| EPI_ISL_461293 | Dutch COVID-19 response team | Erasmus Medical Center |
| EPI_ISL_461294 | Dutch COVID-19 response team | Erasmus Medical Center |
| EPI_ISL_461295 | Dutch COVID-19 response team | Erasmus Medical Center |
| EPI_ISL_461296 | Dutch COVID-19 response team | Erasmus Medical Center |
| EPI_ISL_461297 | Dutch COVID-19 response team | Erasmus Medical Center |
| EPI_ISL_461298 | Dutch COVID-19 response team | Erasmus Medical Center |
| EPI_ISL_461299 | Dutch COVID-19 response team | Erasmus Medical Center |
| EPI_ISL_461300 | Dutch COVID-19 response team | Erasmus Medical Center |
| EPI_ISL_461301 | Dutch COVID-19 response team | Erasmus Medical Center |
| EPI_ISL_461302 | Dutch COVID-19 response team | Erasmus Medical Center |
| EPI_ISL_461303 | Dutch COVID-19 response team | Erasmus Medical Center |
| EPI_ISL_461304 | Dutch COVID-19 response team | Erasmus Medical Center |
| EPI_ISL_461305 | Dutch COVID-19 response team | Erasmus Medical Center |
| EPI_ISL_461306 | Dutch COVID-19 response team | Erasmus Medical Center |
| EPI_ISL_461307 | Dutch COVID-19 response team | Erasmus Medical Center |
| EPI_ISL_461308 | Dutch COVID-19 response team | Erasmus Medical Center |
| EPI_ISL_461309 | Dutch COVID-19 response team | Erasmus Medical Center |
| EPI_ISL_461310 | Dutch COVID-19 response team | Erasmus Medical Center |
| EPI_ISL_461311 | Dutch COVID-19 response team | Erasmus Medical Center |
| EPI_ISL_461312 | Dutch COVID-19 response team | Erasmus Medical Center |
| EPI_ISL_461313 | Dutch COVID-19 response team | Erasmus Medical Center |
| EPI_ISL_461314 | Dutch COVID-19 response team | Erasmus Medical Center |
| EPI_ISL_461315 | Dutch COVID-19 response team | Erasmus Medical Center |
| EPI_ISL_461316 | Dutch COVID-19 response team | Erasmus Medical Center |
| EPI_ISL_461317 | Dutch COVID-19 response team | Erasmus Medical Center |
| EPI_ISL_461318 | Dutch COVID-19 response team | Erasmus Medical Center |
| EPI_ISL_461319 | Dutch COVID-19 response team | Erasmus Medical Center |

| EPI_Accession_ID | Originating Laboratory | Submitting Laboratory |
|---|---|---|
| EPI_ISL_461320 | Dutch COVID-19 response team | Erasmus Medical Center |
| EPI_ISL_461321 | Dutch COVID-19 response team | Erasmus Medical Center |
| EPI_ISL_461322 | Dutch COVID-19 response team | Erasmus Medical Center |
| EPI_ISL_461323 | Dutch COVID-19 response team | Erasmus Medical Center |
| EPI_ISL_461324 | Dutch COVID-19 response team | Erasmus Medical Center |
| EPI_ISL_461325 | Dutch COVID-19 response team | Erasmus Medical Center |
| EPI_ISL_461326 | Dutch COVID-19 response team | Erasmus Medical Center |
| EPI_ISL_461327 | Dutch COVID-19 response team | Erasmus Medical Center |
| EPI_ISL_461328 | Dutch COVID-19 response team | Erasmus Medical Center |
| EPI_ISL_461329 | Dutch COVID-19 response team | Erasmus Medical Center |
| EPI_ISL_461330 | Dutch COVID-19 response team | Erasmus Medical Center |
| EPI_ISL_461331 | Dutch COVID-19 response team | Erasmus Medical Center |
| EPI_ISL_461332 | Dutch COVID-19 response team | Erasmus Medical Center |
| EPI_ISL_461333 | Dutch COVID-19 response team | Erasmus Medical Center |
| EPI_ISL_461334 | Dutch COVID-19 response team | Erasmus Medical Center |
| EPI_ISL_461335 | Dutch COVID-19 response team | Erasmus Medical Center |
| EPI_ISL_461336 | Dutch COVID-19 response team | Erasmus Medical Center |
| EPI_ISL_461337 | Dutch COVID-19 response team | Erasmus Medical Center |
| EPI_ISL_461338 | Dutch COVID-19 response team | Erasmus Medical Center |
| EPI_ISL_461339 | Dutch COVID-19 response team | Erasmus Medical Center |
| EPI_ISL_461340 | Dutch COVID-19 response team | Erasmus Medical Center |
| EPI_ISL_461341 | Dutch COVID-19 response team | Erasmus Medical Center |
| EPI_ISL_461342 | Dutch COVID-19 response team | Erasmus Medical Center |
| EPI_ISL_461343 | Dutch COVID-19 response team | Erasmus Medical Center |
| EPI_ISL_461344 | Dutch COVID-19 response team | Erasmus Medical Center |
| EPI_ISL_461345 | Dutch COVID-19 response team | Erasmus Medical Center |
| EPI_ISL_461346 | Dutch COVID-19 response team | Erasmus Medical Center |
| EPI_ISL_461347 | Dutch COVID-19 response team | Erasmus Medical Center |
| EPI_ISL_461348 | Dutch COVID-19 response team | Erasmus Medical Center |
| EPI_ISL_461349 | Dutch COVID-19 response team | Erasmus Medical Center |
| EPI_ISL_461350 | Dutch COVID-19 response team | Erasmus Medical Center |
| EPI_ISL_461351 | Dutch COVID-19 response team | Erasmus Medical Center |
| EPI_ISL_461352 | Dutch COVID-19 response team | Erasmus Medical Center |

| EPI_Accession_ID | Originating Laboratory | Submitting Laboratory |
|---|---|---|
| EPI_ISL_461353 | Dutch COVID-19 response team | Erasmus Medical Center |
| EPI_ISL_461354 | Dutch COVID-19 response team | Erasmus Medical Center |
| EPI_ISL_461355 | Dutch COVID-19 response team | Erasmus Medical Center |
| EPI_ISL_461356 | Dutch COVID-19 response team | Erasmus Medical Center |
| EPI_ISL_461357 | Dutch COVID-19 response team | Erasmus Medical Center |
| EPI_ISL_461358 | Dutch COVID-19 response team | Erasmus Medical Center |
| EPI_ISL_461359 | Dutch COVID-19 response team | Erasmus Medical Center |
| EPI_ISL_461360 | Dutch COVID-19 response team | Erasmus Medical Center |
| EPI_ISL_461361 | Dutch COVID-19 response team | Erasmus Medical Center |
| EPI_ISL_461362 | Dutch COVID-19 response team | Erasmus Medical Center |
| EPI_ISL_461363 | Dutch COVID-19 response team | Erasmus Medical Center |
| EPI_ISL_461364 | Dutch COVID-19 response team | Erasmus Medical Center |
| EPI_ISL_461365 | Dutch COVID-19 response team | Erasmus Medical Center |
| EPI_ISL_461366 | Dutch COVID-19 response team | Erasmus Medical Center |
| EPI_ISL_461367 | Dutch COVID-19 response team | Erasmus Medical Center |
| EPI_ISL_461368 | Dutch COVID-19 response team | Erasmus Medical Center |
| EPI_ISL_461369 | Dutch COVID-19 response team | Erasmus Medical Center |
| EPI_ISL_461370 | Dutch COVID-19 response team | Erasmus Medical Center |
| EPI_ISL_461371 | Dutch COVID-19 response team | Erasmus Medical Center |
| EPI_ISL_461372 | Dutch COVID-19 response team | Erasmus Medical Center |
| EPI_ISL_461373 | Dutch COVID-19 response team | Erasmus Medical Center |
| EPI_ISL_461375 | Dutch COVID-19 response team | Erasmus Medical Center |
| EPI_ISL_461376 | Dutch COVID-19 response team | Erasmus Medical Center |
| EPI_ISL_461377 | Dutch COVID-19 response team | Erasmus Medical Center |
| EPI_ISL_461378 | Dutch COVID-19 response team | Erasmus Medical Center |
| EPI_ISL_461379 | Dutch COVID-19 response team | Erasmus Medical Center |
| EPI_ISL_461380 | Dutch COVID-19 response team | Erasmus Medical Center |
| EPI_ISL_461381 | Dutch COVID-19 response team | Erasmus Medical Center |
| EPI_ISL_461383 | Dutch COVID-19 response team | Erasmus Medical Center |
| EPI_ISL_461384 | Dutch COVID-19 response team | Erasmus Medical Center |
| EPI_ISL_461385 | Dutch COVID-19 response team | Erasmus Medical Center |
| EPI_ISL_461386 | Dutch COVID-19 response team | Erasmus Medical Center |
| EPI_ISL_461387 | Dutch COVID-19 response team | Erasmus Medical Center |

| EPI_Accession_ID | Originating Laboratory | Submitting Laboratory |
|---|---|---|
| EPI_ISL_461388 | Dutch COVID-19 response team | Erasmus Medical Center |
| EPI_ISL_461389 | Dutch COVID-19 response team | Erasmus Medical Center |
| EPI_ISL_461390 | Dutch COVID-19 response team | Erasmus Medical Center |
| EPI_ISL_461391 | Dutch COVID-19 response team | Erasmus Medical Center |
| EPI_ISL_461394 | Dutch COVID-19 response team | Erasmus Medical Center |
| EPI_ISL_461396 | Dutch COVID-19 response team | Erasmus Medical Center |
| EPI_ISL_461397 | Dutch COVID-19 response team | Erasmus Medical Center |
| EPI_ISL_461398 | Dutch COVID-19 response team | Erasmus Medical Center |
| EPI_ISL_461399 | UW Virology Lab | UW Virology Lab |
| EPI_ISL_461400 | UW Virology Lab | UW Virology Lab |
| EPI_ISL_461401 | UW Virology Lab | UW Virology Lab |
| EPI_ISL_461402 | UW Virology Lab | UW Virology Lab |
| EPI_ISL_461403 | UW Virology Lab | UW Virology Lab |
| EPI_ISL_461404 | UW Virology Lab | UW Virology Lab |
| EPI_ISL_461405 | UW Virology Lab | UW Virology Lab |
| EPI_ISL_461406 | UW Virology Lab | UW Virology Lab |
| EPI_ISL_461407 | UW Virology Lab | UW Virology Lab |
| EPI_ISL_461408 | UW Virology Lab | UW Virology Lab |
| EPI_ISL_461409 | UW Virology Lab | UW Virology Lab |
| EPI_ISL_461410 | UW Virology Lab | UW Virology Lab |
| EPI_ISL_461411 | UW Virology Lab | UW Virology Lab |
| EPI_ISL_461412 | UW Virology Lab | UW Virology Lab |
| EPI_ISL_461413 | UW Virology Lab | UW Virology Lab |
| EPI_ISL_461414 | UW Virology Lab | UW Virology Lab |
| EPI_ISL_461415 | UW Virology Lab | UW Virology Lab |
| EPI_ISL_461416 | UW Virology Lab | UW Virology Lab |
| EPI_ISL_461417 | UW Virology Lab | UW Virology Lab |
| EPI_ISL_461418 | UW Virology Lab | UW Virology Lab |
| EPI_ISL_461419 | UW Virology Lab | UW Virology Lab |
| EPI_ISL_461420 | UW Virology Lab | UW Virology Lab |
| EPI_ISL_461421 | UW Virology Lab | UW Virology Lab |
| EPI_ISL_461422 | UW Virology Lab | UW Virology Lab |
| EPI_ISL_461423 | UW Virology Lab | UW Virology Lab |

| EPI_Accession_ID | Originating Laboratory | Submitting Laboratory |
|---|---|---|
| EPI_ISL_461424 | UW Virology Lab | UW Virology Lab |
| EPI_ISL_461425 | UW Virology Lab | UW Virology Lab |
| EPI_ISL_461426 | UW Virology Lab | UW Virology Lab |
| EPI_ISL_461427 | UW Virology Lab | UW Virology Lab |
| EPI_ISL_461428 | UW Virology Lab | UW Virology Lab |
| EPI_ISL_461429 | UW Virology Lab | UW Virology Lab |
| EPI_ISL_461430 | UW Virology Lab | UW Virology Lab |
| EPI_ISL_461431 | UW Virology Lab | UW Virology Lab |
| EPI_ISL_461432 | UW Virology Lab | UW Virology Lab |
| EPI_ISL_461433 | UW Virology Lab | UW Virology Lab |
| EPI_ISL_461434 | UW Virology Lab | UW Virology Lab |
| EPI_ISL_461435 | UW Virology Lab | UW Virology Lab |
| EPI_ISL_461436 | UW Virology Lab | UW Virology Lab |
| EPI_ISL_461437 | UW Virology Lab | UW Virology Lab |
| EPI_ISL_461438 | UW Virology Lab | UW Virology Lab |
| EPI_ISL_461439 | UW Virology Lab | UW Virology Lab |
| EPI_ISL_461440 | UW Virology Lab | UW Virology Lab |
| EPI_ISL_461441 | UW Virology Lab | UW Virology Lab |
| EPI_ISL_461442 | UW Virology Lab | UW Virology Lab |
| EPI_ISL_461443 | UW Virology Lab | UW Virology Lab |
| EPI_ISL_461444 | UW Virology Lab | UW Virology Lab |
| EPI_ISL_461445 | UW Virology Lab | UW Virology Lab |
| EPI_ISL_461446 | UW Virology Lab | UW Virology Lab |
| EPI_ISL_461447 | UW Virology Lab | UW Virology Lab |
| EPI_ISL_461448 | UW Virology Lab | UW Virology Lab |
| EPI_ISL_461449 | UW Virology Lab | UW Virology Lab |
| EPI_ISL_461450 | UW Virology Lab | UW Virology Lab |
| EPI_ISL_461451 | UW Virology Lab | UW Virology Lab |
| EPI_ISL_461452 | UW Virology Lab | UW Virology Lab |
| EPI_ISL_461453 | UW Virology Lab | UW Virology Lab |
| EPI_ISL_461454 | UW Virology Lab | UW Virology Lab |
| EPI_ISL_461455 | UW Virology Lab | UW Virology Lab |
| EPI_ISL_461456 | UW Virology Lab | UW Virology Lab |

| EPI_Accession_ID | Originating Laboratory | Submitting Laboratory |
|---|---|---|
| EPI_ISL_461457 | UW Virology Lab | UW Virology Lab |
| EPI_ISL_461458 | UW Virology Lab | UW Virology Lab |
| EPI_ISL_461459 | UW Virology Lab | UW Virology Lab |
| EPI_ISL_461460 | UW Virology Lab | UW Virology Lab |
| EPI_ISL_461461 | UW Virology Lab | UW Virology Lab |
| EPI_ISL_461462 | UW Virology Lab | UW Virology Lab |
| EPI_ISL_461463 | UW Virology Lab | UW Virology Lab |
| EPI_ISL_461464 | UW Virology Lab | UW Virology Lab |
| EPI_ISL_461465 | UW Virology Lab | UW Virology Lab |
| EPI_ISL_461466 | UW Virology Lab | UW Virology Lab |
| EPI_ISL_461467 | UW Virology Lab | UW Virology Lab |
| EPI_ISL_461468 | UW Virology Lab | UW Virology Lab |
| EPI_ISL_461469 | UW Virology Lab | UW Virology Lab |
| EPI_ISL_461470 | UW Virology Lab | UW Virology Lab |
| EPI_ISL_461471 | UW Virology Lab | UW Virology Lab |
| EPI_ISL_461472 | UW Virology Lab | UW Virology Lab |
| EPI_ISL_461473 | UW Virology Lab | UW Virology Lab |
| EPI_ISL_461474 | UW Virology Lab | UW Virology Lab |
| EPI_ISL_461475 | UW Virology Lab | UW Virology Lab |
| EPI_ISL_461476 | UW Virology Lab | UW Virology Lab |
| EPI_ISL_461477 | UW Virology Lab | UW Virology Lab |
| EPI_ISL_461478 | Government Medical College, Vadodara | Gujarat Biotechnology Research Centre |
| EPI_ISL_461479 | Government Medical College, Vadodara | Gujarat Biotechnology Research Centre |
| EPI_ISL_461480 | Government Medical College, Vadodara | Gujarat Biotechnology Research Centre |
| EPI_ISL_461481 | Pandit Deendayal Upadhyay Government Medical College, Rajkot | Gujarat Biotechnology Research Centre |
| EPI_ISL_461482 | Pandit Deendayal Upadhyay Government Medical College, Rajkot | Gujarat Biotechnology Research Centre |
| EPI_ISL_461483 | B.J. Medical College and Civil hospital | Gujarat Biotechnology Research Centre |
| EPI_ISL_461484 | B.J. Medical College and Civil hospital | Gujarat Biotechnology Research Centre |
| EPI_ISL_461485 | B.J. Medical College and Civil hospital | Gujarat Biotechnology Research Centre |
| EPI_ISL_461486 | B.J. Medical College and Civil hospital | Gujarat Biotechnology Research Centre |
| EPI_ISL_461487 | B.J. Medical College and Civil hospital | Gujarat Biotechnology Research Centre |

| EPI_Accession_ID | Originating Laboratory | Submitting Laboratory |
|---|---|---|
| EPI_ISL_461488 | B.J. Medical College and Civil hospital | Gujarat Biotechnology Research Centre |
| EPI_ISL_461489 | B.J. Medical College and Civil hospital | Gujarat Biotechnology Research Centre |
| EPI_ISL_461490 | B.J. Medical College and Civil hospital | Gujarat Biotechnology Research Centre |
| EPI_ISL_461491 | B.J. Medical College and Civil hospital | Gujarat Biotechnology Research Centre |
| EPI_ISL_461492 | B.J. Medical College and Civil hospital | Gujarat Biotechnology Research Centre |
| EPI_ISL_461493 | B.J. Medical College and Civil hospital | Gujarat Biotechnology Research Centre |
| EPI_ISL_461494 | B.J. Medical College and Civil hospital | Gujarat Biotechnology Research Centre |
| EPI_ISL_461495 | B.J. Medical College and Civil hospital | Gujarat Biotechnology Research Centre |
| EPI_ISL_461496 | B.J. Medical College and Civil hospital | Gujarat Biotechnology Research Centre |
| EPI_ISL_461497 | B.J. Medical College and Civil hospital | Gujarat Biotechnology Research Centre |
| EPI_ISL_461498 | B.J. Medical College and Civil hospital | Gujarat Biotechnology Research Centre |
| EPI_ISL_461499 | B.J. Medical College and Civil hospital | Gujarat Biotechnology Research Centre |
| EPI_ISL_461500 | B.J. Medical College and Civil hospital | Gujarat Biotechnology Research Centre |
| EPI_ISL_461501 | B.J. Medical College and Civil hospital | Gujarat Biotechnology Research Centre |
| EPI_ISL_461502 | B.J. Medical College and Civil hospital | Gujarat Biotechnology Research Centre |
| EPI_ISL_461503 | B.J. Medical College and Civil hospital | Gujarat Biotechnology Research Centre |
| EPI_ISL_461504 | B.J. Medical College and Civil hospital | Gujarat Biotechnology Research Centre |
| EPI_ISL_461505 | B.J. Medical College and Civil hospital | Gujarat Biotechnology Research Centre |
| EPI_ISL_461506 | B.J. Medical College and Civil hospital | Gujarat Biotechnology Research Centre |
| EPI_ISL_461546 | Department of Pathology, University of Cambridge | COVID-19 Genomics UK (COG-UK) Consortium |
| EPI_ISL_461547 | Department of Pathology, University of Cambridge | COVID-19 Genomics UK (COG-UK) Consortium |
| EPI_ISL_461548 | Department of Pathology, University of Cambridge | COVID-19 Genomics UK (COG-UK) Consortium |
| EPI_ISL_461549 | Department of Pathology, University of Cambridge | COVID-19 Genomics UK (COG-UK) Consortium |
| EPI_ISL_461550 | Department of Pathology, University of Cambridge | COVID-19 Genomics UK (COG-UK) Consortium |
| EPI_ISL_461551 | Department of Pathology, University of Cambridge | COVID-19 Genomics UK (COG-UK) Consortium |
| EPI_ISL_461552 | Department of Pathology, University of Cambridge | COVID-19 Genomics UK (COG-UK) Consortium |
| EPI_ISL_461553 | Department of Pathology, University of Cambridge | COVID-19 Genomics UK (COG-UK) Consortium |
| EPI_ISL_461554 | Department of Pathology, University of Cambridge | COVID-19 Genomics UK (COG-UK) Consortium |
| EPI_ISL_461555 | Department of Pathology, University of Cambridge | COVID-19 Genomics UK (COG-UK) Consortium |
| EPI_ISL_461556 | Department of Pathology, University of Cambridge | COVID-19 Genomics UK (COG-UK) Consortium |
| EPI_ISL_461557 | Department of Pathology, University of Cambridge | COVID-19 Genomics UK (COG-UK) Consortium |
| EPI_ISL_461558 | Department of Pathology, University of Cambridge | COVID-19 Genomics UK (COG-UK) Consortium |
| EPI_ISL_461559 | Department of Pathology, University of Cambridge | COVID-19 Genomics UK (COG-UK) Consortium |

| EPI_Accession_ID | Originating Laboratory | Submitting Laboratory |
|---|---|---|
| EPI_ISL_461560 | Department of Pathology, University of Cambridge | COVID-19 Genomics UK (COG-UK) Consortium |
| EPI_ISL_461561 | Department of Pathology, University of Cambridge | COVID-19 Genomics UK (COG-UK) Consortium |
| EPI_ISL_461562 | Department of Pathology, University of Cambridge | COVID-19 Genomics UK (COG-UK) Consortium |
| EPI_ISL_461563 | Department of Pathology, University of Cambridge | COVID-19 Genomics UK (COG-UK) Consortium |
| EPI_ISL_461565 | Department of Pathology, University of Cambridge | COVID-19 Genomics UK (COG-UK) Consortium |
| EPI_ISL_461566 | Department of Pathology, University of Cambridge | COVID-19 Genomics UK (COG-UK) Consortium |
| EPI_ISL_461567 | Department of Pathology, University of Cambridge | COVID-19 Genomics UK (COG-UK) Consortium |
| EPI_ISL_461568 | Department of Pathology, University of Cambridge | COVID-19 Genomics UK (COG-UK) Consortium |
| EPI_ISL_461569 | Department of Pathology, University of Cambridge | COVID-19 Genomics UK (COG-UK) Consortium |
| EPI_ISL_461570 | Department of Pathology, University of Cambridge | COVID-19 Genomics UK (COG-UK) Consortium |
| EPI_ISL_461571 | Department of Pathology, University of Cambridge | COVID-19 Genomics UK (COG-UK) Consortium |
| EPI_ISL_461572 | Department of Pathology, University of Cambridge | COVID-19 Genomics UK (COG-UK) Consortium |
| EPI_ISL_461573 | Department of Pathology, University of Cambridge | COVID-19 Genomics UK (COG-UK) Consortium |
| EPI_ISL_461574 | Department of Pathology, University of Cambridge | COVID-19 Genomics UK (COG-UK) Consortium |
| EPI_ISL_461575 | Department of Pathology, University of Cambridge | COVID-19 Genomics UK (COG-UK) Consortium |
| EPI_ISL_461576 | Department of Pathology, University of Cambridge | COVID-19 Genomics UK (COG-UK) Consortium |
| EPI_ISL_461577 | Department of Pathology, University of Cambridge | COVID-19 Genomics UK (COG-UK) Consortium |
| EPI_ISL_461578 | Department of Pathology, University of Cambridge | COVID-19 Genomics UK (COG-UK) Consortium |
| EPI_ISL_461579 | Department of Pathology, University of Cambridge | COVID-19 Genomics UK (COG-UK) Consortium |
| EPI_ISL_461580 | Department of Pathology, University of Cambridge | COVID-19 Genomics UK (COG-UK) Consortium |
| EPI_ISL_461581 | Department of Pathology, University of Cambridge | COVID-19 Genomics UK (COG-UK) Consortium |
| EPI_ISL_461582 | Department of Pathology, University of Cambridge | COVID-19 Genomics UK (COG-UK) Consortium |
| EPI_ISL_461583 | Department of Pathology, University of Cambridge | COVID-19 Genomics UK (COG-UK) Consortium |
| EPI_ISL_461584 | Department of Pathology, University of Cambridge | COVID-19 Genomics UK (COG-UK) Consortium |
| EPI_ISL_461585 | Department of Pathology, University of Cambridge | COVID-19 Genomics UK (COG-UK) Consortium |
| EPI_ISL_461586 | Department of Pathology, University of Cambridge | COVID-19 Genomics UK (COG-UK) Consortium |
| EPI_ISL_461587 | Department of Pathology, University of Cambridge | COVID-19 Genomics UK (COG-UK) Consortium |
| EPI_ISL_461588 | Department of Pathology, University of Cambridge | COVID-19 Genomics UK (COG-UK) Consortium |
| EPI_ISL_461589 | West of Scotland Specialist Virology Centre, NHSGGC / MRC-University of Glasgow Centre for Virus Research | COVID-19 Genomics UK (COG-UK) Consortium |
| EPI_ISL_461590 | West of Scotland Specialist Virology Centre, NHSGGC / MRC-University of Glasgow Centre for Virus Research | COVID-19 Genomics UK (COG-UK) Consortium |

| EPI_Accession_ID | Originating Laboratory | Submitting Laboratory |
|---|---|---|
| EPI_ISL_461591 | West of Scotland Specialist Virology Centre, NHSGGC / MRC-University of Glasgow Centre for Virus Research | COVID-19 Genomics UK (COG-UK) Consortium |
| EPI_ISL_461592 | West of Scotland Specialist Virology Centre, NHSGGC / MRC-University of Glasgow Centre for Virus Research | COVID-19 Genomics UK (COG-UK) Consortium |
| EPI_ISL_461593 | West of Scotland Specialist Virology Centre, NHSGGC / MRC-University of Glasgow Centre for Virus Research | COVID-19 Genomics UK (COG-UK) Consortium |
| EPI_ISL_461594 | West of Scotland Specialist Virology Centre, NHSGGC / MRC-University of Glasgow Centre for Virus Research | COVID-19 Genomics UK (COG-UK) Consortium |
| EPI_ISL_461595 | West of Scotland Specialist Virology Centre, NHSGGC / MRC-University of Glasgow Centre for Virus Research | COVID-19 Genomics UK (COG-UK) Consortium |
| EPI_ISL_461596 | West of Scotland Specialist Virology Centre, NHSGGC / MRC-University of Glasgow Centre for Virus Research | COVID-19 Genomics UK (COG-UK) Consortium |
| EPI_ISL_461597 | West of Scotland Specialist Virology Centre, NHSGGC / MRC-University of Glasgow Centre for Virus Research | COVID-19 Genomics UK (COG-UK) Consortium |
| EPI_ISL_461598 | West of Scotland Specialist Virology Centre, NHSGGC / MRC-University of Glasgow Centre for Virus Research | COVID-19 Genomics UK (COG-UK) Consortium |
| EPI_ISL_461599 | West of Scotland Specialist Virology Centre, NHSGGC / MRC-University of Glasgow Centre for Virus Research | COVID-19 Genomics UK (COG-UK) Consortium |
| EPI_ISL_461600 | West of Scotland Specialist Virology Centre, NHSGGC / MRC-University of Glasgow Centre for Virus Research | COVID-19 Genomics UK (COG-UK) Consortium |
| EPI_ISL_461601 | West of Scotland Specialist Virology Centre, NHSGGC / MRC-University of Glasgow Centre for Virus Research | COVID-19 Genomics UK (COG-UK) Consortium |
| EPI_ISL_461602 | West of Scotland Specialist Virology Centre, NHSGGC / MRC-University of Glasgow Centre for Virus Research | COVID-19 Genomics UK (COG-UK) Consortium |
| EPI_ISL_461603 | West of Scotland Specialist Virology Centre, NHSGGC / MRC-University of Glasgow Centre for Virus Research | COVID-19 Genomics UK (COG-UK) Consortium |
| EPI_ISL_461604 | West of Scotland Specialist Virology Centre, NHSGGC / MRC-University of Glasgow Centre for Virus Research | COVID-19 Genomics UK (COG-UK) Consortium |
| EPI_ISL_461605 | West of Scotland Specialist Virology Centre, NHSGGC / MRC-University of Glasgow Centre for Virus Research | COVID-19 Genomics UK (COG-UK) Consortium |
| EPI_ISL_461606 | West of Scotland Specialist Virology Centre, NHSGGC / MRC-University of Glasgow Centre for Virus Research | COVID-19 Genomics UK (COG-UK) Consortium |

| EPI_Accession_ID | Originating Laboratory | Submitting Laboratory |
|---|---|---|
| EPI_ISL_461607 | West of Scotland Specialist Virology Centre, NHSGGC / MRC-University of Glasgow Centre for Virus Research | COVID-19 Genomics UK (COG-UK) Consortium |
| EPI_ISL_461608 | West of Scotland Specialist Virology Centre, NHSGGC / MRC-University of Glasgow Centre for Virus Research | COVID-19 Genomics UK (COG-UK) Consortium |
| EPI_ISL_461609 | West of Scotland Specialist Virology Centre, NHSGGC / MRC-University of Glasgow Centre for Virus Research | COVID-19 Genomics UK (COG-UK) Consortium |
| EPI_ISL_461610 | West of Scotland Specialist Virology Centre, NHSGGC / MRC-University of Glasgow Centre for Virus Research | COVID-19 Genomics UK (COG-UK) Consortium |
| EPI_ISL_461611 | West of Scotland Specialist Virology Centre, NHSGGC / MRC-University of Glasgow Centre for Virus Research | COVID-19 Genomics UK (COG-UK) Consortium |
| EPI_ISL_461612 | West of Scotland Specialist Virology Centre, NHSGGC / MRC-University of Glasgow Centre for Virus Research | COVID-19 Genomics UK (COG-UK) Consortium |
| EPI_ISL_461613 | West of Scotland Specialist Virology Centre, NHSGGC / MRC-University of Glasgow Centre for Virus Research | COVID-19 Genomics UK (COG-UK) Consortium |
| EPI_ISL_461614 | West of Scotland Specialist Virology Centre, NHSGGC / MRC-University of Glasgow Centre for Virus Research | COVID-19 Genomics UK (COG-UK) Consortium |
| EPI_ISL_461615 | West of Scotland Specialist Virology Centre, NHSGGC / MRC-University of Glasgow Centre for Virus Research | COVID-19 Genomics UK (COG-UK) Consortium |
| EPI_ISL_461616 | West of Scotland Specialist Virology Centre, NHSGGC / MRC-University of Glasgow Centre for Virus Research | COVID-19 Genomics UK (COG-UK) Consortium |
| EPI_ISL_461617 | West of Scotland Specialist Virology Centre, NHSGGC / MRC-University of Glasgow Centre for Virus Research | COVID-19 Genomics UK (COG-UK) Consortium |
| EPI_ISL_461618 | West of Scotland Specialist Virology Centre, NHSGGC / MRC-University of Glasgow Centre for Virus Research | COVID-19 Genomics UK (COG-UK) Consortium |
| EPI_ISL_461619 | West of Scotland Specialist Virology Centre, NHSGGC / MRC-University of Glasgow Centre for Virus Research | COVID-19 Genomics UK (COG-UK) Consortium |
| EPI_ISL_461620 | West of Scotland Specialist Virology Centre, NHSGGC / MRC-University of Glasgow Centre for Virus Research | COVID-19 Genomics UK (COG-UK) Consortium |
| EPI_ISL_461621 | West of Scotland Specialist Virology Centre, NHSGGC / MRC-University of Glasgow Centre for Virus Research | COVID-19 Genomics UK (COG-UK) Consortium |
| EPI_ISL_461622 | West of Scotland Specialist Virology Centre, NHSGGC / MRC-University of Glasgow Centre for Virus Research | COVID-19 Genomics UK (COG-UK) Consortium |

| EPI_Accession_ID | Originating Laboratory | Submitting Laboratory |
|---|---|---|
| EPI_ISL_461623 | West of Scotland Specialist Virology Centre, NHSGGC / MRC-University of Glasgow Centre for Virus Research | COVID-19 Genomics UK (COG-UK) Consortium |
| EPI_ISL_461624 | West of Scotland Specialist Virology Centre, NHSGGC / MRC-University of Glasgow Centre for Virus Research | COVID-19 Genomics UK (COG-UK) Consortium |
| EPI_ISL_461625 | West of Scotland Specialist Virology Centre, NHSGGC / MRC-University of Glasgow Centre for Virus Research | COVID-19 Genomics UK (COG-UK) Consortium |
| EPI_ISL_461626 | West of Scotland Specialist Virology Centre, NHSGGC / MRC-University of Glasgow Centre for Virus Research | COVID-19 Genomics UK (COG-UK) Consortium |
| EPI_ISL_461627 | West of Scotland Specialist Virology Centre, NHSGGC / MRC-University of Glasgow Centre for Virus Research | COVID-19 Genomics UK (COG-UK) Consortium |
| EPI_ISL_461628 | West of Scotland Specialist Virology Centre, NHSGGC / MRC-University of Glasgow Centre for Virus Research | COVID-19 Genomics UK (COG-UK) Consortium |
| EPI_ISL_461629 | West of Scotland Specialist Virology Centre, NHSGGC / MRC-University of Glasgow Centre for Virus Research | COVID-19 Genomics UK (COG-UK) Consortium |
| EPI_ISL_461630 | West of Scotland Specialist Virology Centre, NHSGGC / MRC-University of Glasgow Centre for Virus Research | COVID-19 Genomics UK (COG-UK) Consortium |
| EPI_ISL_461631 | West of Scotland Specialist Virology Centre, NHSGGC / MRC-University of Glasgow Centre for Virus Research | COVID-19 Genomics UK (COG-UK) Consortium |
| EPI_ISL_461632 | West of Scotland Specialist Virology Centre, NHSGGC / MRC-University of Glasgow Centre for Virus Research | COVID-19 Genomics UK (COG-UK) Consortium |
| EPI_ISL_461633 | West of Scotland Specialist Virology Centre, NHSGGC / MRC-University of Glasgow Centre for Virus Research | COVID-19 Genomics UK (COG-UK) Consortium |
| EPI_ISL_461634 | West of Scotland Specialist Virology Centre, NHSGGC / MRC-University of Glasgow Centre for Virus Research | COVID-19 Genomics UK (COG-UK) Consortium |
| EPI_ISL_461635 | West of Scotland Specialist Virology Centre, NHSGGC / MRC-University of Glasgow Centre for Virus Research | COVID-19 Genomics UK (COG-UK) Consortium |
| EPI_ISL_461636 | West of Scotland Specialist Virology Centre, NHSGGC / MRC-University of Glasgow Centre for Virus Research | COVID-19 Genomics UK (COG-UK) Consortium |
| EPI_ISL_461637 | West of Scotland Specialist Virology Centre, NHSGGC / MRC-University of Glasgow Centre for Virus Research | COVID-19 Genomics UK (COG-UK) Consortium |
| EPI_ISL_461638 | West of Scotland Specialist Virology Centre, NHSGGC / MRC-University of Glasgow Centre for Virus Research | COVID-19 Genomics UK (COG-UK) Consortium |

| EPI_Accession_ID | Originating Laboratory | Submitting Laboratory |
|---|---|---|
| EPI_ISL_461639 | West of Scotland Specialist Virology Centre, NHSGGC / MRC-University of Glasgow Centre for Virus Research | COVID-19 Genomics UK (COG-UK) Consortium |
| EPI_ISL_461640 | West of Scotland Specialist Virology Centre, NHSGGC / MRC-University of Glasgow Centre for Virus Research | COVID-19 Genomics UK (COG-UK) Consortium |
| EPI_ISL_461641 | West of Scotland Specialist Virology Centre, NHSGGC / MRC-University of Glasgow Centre for Virus Research | COVID-19 Genomics UK (COG-UK) Consortium |
| EPI_ISL_461642 | West of Scotland Specialist Virology Centre, NHSGGC / MRC-University of Glasgow Centre for Virus Research | COVID-19 Genomics UK (COG-UK) Consortium |
| EPI_ISL_461643 | West of Scotland Specialist Virology Centre, NHSGGC / MRC-University of Glasgow Centre for Virus Research | COVID-19 Genomics UK (COG-UK) Consortium |
| EPI_ISL_461644 | West of Scotland Specialist Virology Centre, NHSGGC / MRC-University of Glasgow Centre for Virus Research | COVID-19 Genomics UK (COG-UK) Consortium |
| EPI_ISL_461645 | West of Scotland Specialist Virology Centre, NHSGGC / MRC-University of Glasgow Centre for Virus Research | COVID-19 Genomics UK (COG-UK) Consortium |
| EPI_ISL_461646 | West of Scotland Specialist Virology Centre, NHSGGC / MRC-University of Glasgow Centre for Virus Research | COVID-19 Genomics UK (COG-UK) Consortium |
| EPI_ISL_461647 | West of Scotland Specialist Virology Centre, NHSGGC / MRC-University of Glasgow Centre for Virus Research | COVID-19 Genomics UK (COG-UK) Consortium |
| EPI_ISL_461648 | West of Scotland Specialist Virology Centre, NHSGGC / MRC-University of Glasgow Centre for Virus Research | COVID-19 Genomics UK (COG-UK) Consortium |
| EPI_ISL_461649 | West of Scotland Specialist Virology Centre, NHSGGC / MRC-University of Glasgow Centre for Virus Research | COVID-19 Genomics UK (COG-UK) Consortium |
| EPI_ISL_461650 | West of Scotland Specialist Virology Centre, NHSGGC / MRC-University of Glasgow Centre for Virus Research | COVID-19 Genomics UK (COG-UK) Consortium |
| EPI_ISL_461651 | West of Scotland Specialist Virology Centre, NHSGGC / MRC-University of Glasgow Centre for Virus Research | COVID-19 Genomics UK (COG-UK) Consortium |
| EPI_ISL_461652 | West of Scotland Specialist Virology Centre, NHSGGC / MRC-University of Glasgow Centre for Virus Research | COVID-19 Genomics UK (COG-UK) Consortium |
| EPI_ISL_461653 | West of Scotland Specialist Virology Centre, NHSGGC / MRC-University of Glasgow Centre for Virus Research | COVID-19 Genomics UK (COG-UK) Consortium |
| EPI_ISL_461654 | West of Scotland Specialist Virology Centre, NHSGGC / MRC-University of Glasgow Centre for Virus Research | COVID-19 Genomics UK (COG-UK) Consortium |

| EPI_Accession_ID | Originating Laboratory | Submitting Laboratory |
|---|---|---|
| EPI_ISL_461655 | West of Scotland Specialist Virology Centre, NHSGGC / MRC-University of Glasgow Centre for Virus Research | COVID-19 Genomics UK (COG-UK) Consortium |
| EPI_ISL_461656 | West of Scotland Specialist Virology Centre, NHSGGC / MRC-University of Glasgow Centre for Virus Research | COVID-19 Genomics UK (COG-UK) Consortium |
| EPI_ISL_461657 | West of Scotland Specialist Virology Centre, NHSGGC / MRC-University of Glasgow Centre for Virus Research | COVID-19 Genomics UK (COG-UK) Consortium |
| EPI_ISL_461658 | West of Scotland Specialist Virology Centre, NHSGGC / MRC-University of Glasgow Centre for Virus Research | COVID-19 Genomics UK (COG-UK) Consortium |
| EPI_ISL_461659 | West of Scotland Specialist Virology Centre, NHSGGC / MRC-University of Glasgow Centre for Virus Research | COVID-19 Genomics UK (COG-UK) Consortium |
| EPI_ISL_461660 | West of Scotland Specialist Virology Centre, NHSGGC / MRC-University of Glasgow Centre for Virus Research | COVID-19 Genomics UK (COG-UK) Consortium |
| EPI_ISL_461661 | West of Scotland Specialist Virology Centre, NHSGGC / MRC-University of Glasgow Centre for Virus Research | COVID-19 Genomics UK (COG-UK) Consortium |
| EPI_ISL_461662 | West of Scotland Specialist Virology Centre, NHSGGC / MRC-University of Glasgow Centre for Virus Research | COVID-19 Genomics UK (COG-UK) Consortium |
| EPI_ISL_461663 | West of Scotland Specialist Virology Centre, NHSGGC / MRC-University of Glasgow Centre for Virus Research | COVID-19 Genomics UK (COG-UK) Consortium |
| EPI_ISL_461664 | West of Scotland Specialist Virology Centre, NHSGGC / MRC-University of Glasgow Centre for Virus Research | COVID-19 Genomics UK (COG-UK) Consortium |
| EPI_ISL_461665 | West of Scotland Specialist Virology Centre, NHSGGC / MRC-University of Glasgow Centre for Virus Research | COVID-19 Genomics UK (COG-UK) Consortium |
| EPI_ISL_461666 | West of Scotland Specialist Virology Centre, NHSGGC / MRC-University of Glasgow Centre for Virus Research | COVID-19 Genomics UK (COG-UK) Consortium |
| EPI_ISL_461667 | West of Scotland Specialist Virology Centre, NHSGGC / MRC-University of Glasgow Centre for Virus Research | COVID-19 Genomics UK (COG-UK) Consortium |
| EPI_ISL_461668 | West of Scotland Specialist Virology Centre, NHSGGC / MRC-University of Glasgow Centre for Virus Research | COVID-19 Genomics UK (COG-UK) Consortium |
| EPI_ISL_461669 | West of Scotland Specialist Virology Centre, NHSGGC / MRC-University of Glasgow Centre for Virus Research | COVID-19 Genomics UK (COG-UK) Consortium |
| EPI_ISL_461670 | West of Scotland Specialist Virology Centre, NHSGGC / MRC-University of Glasgow Centre for Virus Research | COVID-19 Genomics UK (COG-UK) Consortium |

| EPI_Accession_ID | Originating Laboratory | Submitting Laboratory |
|---|---|---|
| EPI_ISL_461671 | West of Scotland Specialist Virology Centre, NHSGGC / MRC-University of Glasgow Centre for Virus Research | COVID-19 Genomics UK (COG-UK) Consortium |
| EPI_ISL_461672 | West of Scotland Specialist Virology Centre, NHSGGC / MRC-University of Glasgow Centre for Virus Research | COVID-19 Genomics UK (COG-UK) Consortium |
| EPI_ISL_461673 | West of Scotland Specialist Virology Centre, NHSGGC / MRC-University of Glasgow Centre for Virus Research | COVID-19 Genomics UK (COG-UK) Consortium |
| EPI_ISL_461674 | West of Scotland Specialist Virology Centre, NHSGGC / MRC-University of Glasgow Centre for Virus Research | COVID-19 Genomics UK (COG-UK) Consortium |
| EPI_ISL_461675 | West of Scotland Specialist Virology Centre, NHSGGC / MRC-University of Glasgow Centre for Virus Research | COVID-19 Genomics UK (COG-UK) Consortium |
| EPI_ISL_461676 | West of Scotland Specialist Virology Centre, NHSGGC / MRC-University of Glasgow Centre for Virus Research | COVID-19 Genomics UK (COG-UK) Consortium |
| EPI_ISL_461677 | West of Scotland Specialist Virology Centre, NHSGGC / MRC-University of Glasgow Centre for Virus Research | COVID-19 Genomics UK (COG-UK) Consortium |
| EPI_ISL_461678 | West of Scotland Specialist Virology Centre, NHSGGC / MRC-University of Glasgow Centre for Virus Research | COVID-19 Genomics UK (COG-UK) Consortium |
| EPI_ISL_461679 | West of Scotland Specialist Virology Centre, NHSGGC / MRC-University of Glasgow Centre for Virus Research | COVID-19 Genomics UK (COG-UK) Consortium |
| EPI_ISL_461680 | West of Scotland Specialist Virology Centre, NHSGGC / MRC-University of Glasgow Centre for Virus Research | COVID-19 Genomics UK (COG-UK) Consortium |
| EPI_ISL_461681 | West of Scotland Specialist Virology Centre, NHSGGC / MRC-University of Glasgow Centre for Virus Research | COVID-19 Genomics UK (COG-UK) Consortium |
| EPI_ISL_461682 | West of Scotland Specialist Virology Centre, NHSGGC / MRC-University of Glasgow Centre for Virus Research | COVID-19 Genomics UK (COG-UK) Consortium |
| EPI_ISL_461683 | West of Scotland Specialist Virology Centre, NHSGGC / MRC-University of Glasgow Centre for Virus Research | COVID-19 Genomics UK (COG-UK) Consortium |
| EPI_ISL_461684 | West of Scotland Specialist Virology Centre, NHSGGC / MRC-University of Glasgow Centre for Virus Research | COVID-19 Genomics UK (COG-UK) Consortium |
| EPI_ISL_461685 | West of Scotland Specialist Virology Centre, NHSGGC / MRC-University of Glasgow Centre for Virus Research | COVID-19 Genomics UK (COG-UK) Consortium |
| EPI_ISL_461686 | West of Scotland Specialist Virology Centre, NHSGGC / MRC-University of Glasgow Centre for Virus Research | COVID-19 Genomics UK (COG-UK) Consortium |

| EPI_Accession_ID | Originating Laboratory | Submitting Laboratory |
|---|---|---|
| EPI_ISL_461687 | West of Scotland Specialist Virology Centre, NHSGGC / MRC-University of Glasgow Centre for Virus Research | COVID-19 Genomics UK (COG-UK) Consortium |
| EPI_ISL_461688 | West of Scotland Specialist Virology Centre, NHSGGC / MRC-University of Glasgow Centre for Virus Research | COVID-19 Genomics UK (COG-UK) Consortium |
| EPI_ISL_461689 | West of Scotland Specialist Virology Centre, NHSGGC / MRC-University of Glasgow Centre for Virus Research | COVID-19 Genomics UK (COG-UK) Consortium |
| EPI_ISL_461690 | West of Scotland Specialist Virology Centre, NHSGGC / MRC-University of Glasgow Centre for Virus Research | COVID-19 Genomics UK (COG-UK) Consortium |
| EPI_ISL_461691 | West of Scotland Specialist Virology Centre, NHSGGC / MRC-University of Glasgow Centre for Virus Research | COVID-19 Genomics UK (COG-UK) Consortium |
| EPI_ISL_461692 | West of Scotland Specialist Virology Centre, NHSGGC / MRC-University of Glasgow Centre for Virus Research | COVID-19 Genomics UK (COG-UK) Consortium |
| EPI_ISL_461693 | West of Scotland Specialist Virology Centre, NHSGGC / MRC-University of Glasgow Centre for Virus Research | COVID-19 Genomics UK (COG-UK) Consortium |
| EPI_ISL_461694 | West of Scotland Specialist Virology Centre, NHSGGC / MRC-University of Glasgow Centre for Virus Research | COVID-19 Genomics UK (COG-UK) Consortium |
| EPI_ISL_461695 | West of Scotland Specialist Virology Centre, NHSGGC / MRC-University of Glasgow Centre for Virus Research | COVID-19 Genomics UK (COG-UK) Consortium |
| EPI_ISL_461696 | West of Scotland Specialist Virology Centre, NHSGGC / MRC-University of Glasgow Centre for Virus Research | COVID-19 Genomics UK (COG-UK) Consortium |
| EPI_ISL_461697 | West of Scotland Specialist Virology Centre, NHSGGC / MRC-University of Glasgow Centre for Virus Research | COVID-19 Genomics UK (COG-UK) Consortium |
| EPI_ISL_461698 | West of Scotland Specialist Virology Centre, NHSGGC / MRC-University of Glasgow Centre for Virus Research | COVID-19 Genomics UK (COG-UK) Consortium |
| EPI_ISL_461699 | West of Scotland Specialist Virology Centre, NHSGGC / MRC-University of Glasgow Centre for Virus Research | COVID-19 Genomics UK (COG-UK) Consortium |
| EPI_ISL_461700 | West of Scotland Specialist Virology Centre, NHSGGC / MRC-University of Glasgow Centre for Virus Research | COVID-19 Genomics UK (COG-UK) Consortium |
| EPI_ISL_461701 | West of Scotland Specialist Virology Centre, NHSGGC / MRC-University of Glasgow Centre for Virus Research | COVID-19 Genomics UK (COG-UK) Consortium |
| EPI_ISL_461702 | West of Scotland Specialist Virology Centre, NHSGGC / MRC-University of Glasgow Centre for Virus Research | COVID-19 Genomics UK (COG-UK) Consortium |

| EPI_Accession_ID | Originating Laboratory | Submitting Laboratory |
|---|---|---|
| EPI_ISL_461703 | West of Scotland Specialist Virology Centre, NHSGGC / MRC-University of Glasgow Centre for Virus Research | COVID-19 Genomics UK (COG-UK) Consortium |
| EPI_ISL_461704 | West of Scotland Specialist Virology Centre, NHSGGC / MRC-University of Glasgow Centre for Virus Research | COVID-19 Genomics UK (COG-UK) Consortium |
| EPI_ISL_461705 | West of Scotland Specialist Virology Centre, NHSGGC / MRC-University of Glasgow Centre for Virus Research | COVID-19 Genomics UK (COG-UK) Consortium |
| EPI_ISL_461706 | Virology Department, Royal Infirmary of Edinburgh, NHS Lothian / School of Biological Sciences, University of Edinburgh / Institute of Genetics and Molecular Medicine, University of Edinburgh | COVID-19 Genomics UK (COG-UK) Consortium |
| EPI_ISL_461707 | Virology Department, Royal Infirmary of Edinburgh, NHS Lothian / School of Biological Sciences, University of Edinburgh / Institute of Genetics and Molecular Medicine, University of Edinburgh | COVID-19 Genomics UK (COG-UK) Consortium |
| EPI_ISL_461708 | Virology Department, Royal Infirmary of Edinburgh, NHS Lothian / School of Biological Sciences, University of Edinburgh / Institute of Genetics and Molecular Medicine, University of Edinburgh | COVID-19 Genomics UK (COG-UK) Consortium |
| EPI_ISL_461709 | Virology Department, Royal Infirmary of Edinburgh, NHS Lothian / School of Biological Sciences, University of Edinburgh / Institute of Genetics and Molecular Medicine, University of Edinburgh | COVID-19 Genomics UK (COG-UK) Consortium |
| EPI_ISL_461710 | Virology Department, Royal Infirmary of Edinburgh, NHS Lothian / School of Biological Sciences, University of Edinburgh / Institute of Genetics and Molecular Medicine, University of Edinburgh | COVID-19 Genomics UK (COG-UK) Consortium |
| EPI_ISL_461711 | Virology Department, Royal Infirmary of Edinburgh, NHS Lothian / School of Biological Sciences, University of Edinburgh / Institute of Genetics and Molecular Medicine, University of Edinburgh | COVID-19 Genomics UK (COG-UK) Consortium |

| EPI_Accession_ID | Originating Laboratory | Submitting Laboratory |
|---|---|---|
| EPI_ISL_461712 | Virology Department, Royal Infirmary of Edinburgh, NHS Lothian / School of Biological Sciences, University of Edinburgh / Institute of Genetics and Molecular Medicine, University of Edinburgh | COVID-19 Genomics UK (COG-UK) Consortium |
| EPI_ISL_461713 | Virology Department, Royal Infirmary of Edinburgh, NHS Lothian / School of Biological Sciences, University of Edinburgh / Institute of Genetics and Molecular Medicine, University of Edinburgh | COVID-19 Genomics UK (COG-UK) Consortium |
| EPI_ISL_461714 | Virology Department, Royal Infirmary of Edinburgh, NHS Lothian / School of Biological Sciences, University of Edinburgh / Institute of Genetics and Molecular Medicine, University of Edinburgh | COVID-19 Genomics UK (COG-UK) Consortium |
| EPI_ISL_461715 | Virology Department, Royal Infirmary of Edinburgh, NHS Lothian / School of Biological Sciences, University of Edinburgh / Institute of Genetics and Molecular Medicine, University of Edinburgh | COVID-19 Genomics UK (COG-UK) Consortium |
| EPI_ISL_461716 | Virology Department, Royal Infirmary of Edinburgh, NHS Lothian / School of Biological Sciences, University of Edinburgh / Institute of Genetics and Molecular Medicine, University of Edinburgh | COVID-19 Genomics UK (COG-UK) Consortium |
| EPI_ISL_461717 | Virology Department, Royal Infirmary of Edinburgh, NHS Lothian / School of Biological Sciences, University of Edinburgh / Institute of Genetics and Molecular Medicine, University of Edinburgh | COVID-19 Genomics UK (COG-UK) Consortium |
| EPI_ISL_461718 | Virology Department, Royal Infirmary of Edinburgh, NHS Lothian / School of Biological Sciences, University of Edinburgh / Institute of Genetics and Molecular Medicine, University of Edinburgh | COVID-19 Genomics UK (COG-UK) Consortium |
| EPI_ISL_461719 | Virology Department, Royal Infirmary of Edinburgh, NHS Lothian / School of Biological Sciences, University of Edinburgh / Institute of Genetics and Molecular Medicine, University of Edinburgh | COVID-19 Genomics UK (COG-UK) Consortium |

| EPI_Accession_ID | Originating Laboratory | Submitting Laboratory |
|---|---|---|
| EPI_ISL_461720 | Virology Department, Royal Infirmary of Edinburgh, NHS Lothian / School of Biological Sciences, University of Edinburgh / Institute of Genetics and Molecular Medicine, University of Edinburgh | COVID-19 Genomics UK (COG-UK) Consortium |
| EPI_ISL_461721 | Virology Department, Royal Infirmary of Edinburgh, NHS Lothian / School of Biological Sciences, University of Edinburgh / Institute of Genetics and Molecular Medicine, University of Edinburgh | COVID-19 Genomics UK (COG-UK) Consortium |
| EPI_ISL_461722 | Virology Department, Royal Infirmary of Edinburgh, NHS Lothian / School of Biological Sciences, University of Edinburgh / Institute of Genetics and Molecular Medicine, University of Edinburgh | COVID-19 Genomics UK (COG-UK) Consortium |
| EPI_ISL_461723 | Virology Department, Royal Infirmary of Edinburgh, NHS Lothian / School of Biological Sciences, University of Edinburgh / Institute of Genetics and Molecular Medicine, University of Edinburgh | COVID-19 Genomics UK (COG-UK) Consortium |
| EPI_ISL_461724 | Virology Department, Royal Infirmary of Edinburgh, NHS Lothian / School of Biological Sciences, University of Edinburgh / Institute of Genetics and Molecular Medicine, University of Edinburgh | COVID-19 Genomics UK (COG-UK) Consortium |
| EPI_ISL_461725 | Virology Department, Royal Infirmary of Edinburgh, NHS Lothian / School of Biological Sciences, University of Edinburgh / Institute of Genetics and Molecular Medicine, University of Edinburgh | COVID-19 Genomics UK (COG-UK) Consortium |
| EPI_ISL_461726 | Virology Department, Royal Infirmary of Edinburgh, NHS Lothian / School of Biological Sciences, University of Edinburgh / Institute of Genetics and Molecular Medicine, University of Edinburgh | COVID-19 Genomics UK (COG-UK) Consortium |
| EPI_ISL_461727 | Virology Department, Royal Infirmary of Edinburgh, NHS Lothian / School of Biological Sciences, University of Edinburgh / Institute of Genetics and Molecular Medicine, University of Edinburgh | COVID-19 Genomics UK (COG-UK) Consortium |

| EPI_Accession_ID | Originating Laboratory | Submitting Laboratory |
|---|---|---|
| EPI_ISL_461728 | Virology Department, Royal Infirmary of Edinburgh, NHS Lothian / School of Biological Sciences, University of Edinburgh / Institute of Genetics and Molecular Medicine, University of Edinburgh | COVID-19 Genomics UK (COG-UK) Consortium |
| EPI_ISL_461729 | Virology Department, Royal Infirmary of Edinburgh, NHS Lothian / School of Biological Sciences, University of Edinburgh / Institute of Genetics and Molecular Medicine, University of Edinburgh | COVID-19 Genomics UK (COG-UK) Consortium |
| EPI_ISL_461730 | Virology Department, Royal Infirmary of Edinburgh, NHS Lothian / School of Biological Sciences, University of Edinburgh / Institute of Genetics and Molecular Medicine, University of Edinburgh | COVID-19 Genomics UK (COG-UK) Consortium |
| EPI_ISL_461731 | Virology Department, Royal Infirmary of Edinburgh, NHS Lothian / School of Biological Sciences, University of Edinburgh / Institute of Genetics and Molecular Medicine, University of Edinburgh | COVID-19 Genomics UK (COG-UK) Consortium |
| EPI_ISL_461732 | Virology Department, Royal Infirmary of Edinburgh, NHS Lothian / School of Biological Sciences, University of Edinburgh / Institute of Genetics and Molecular Medicine, University of Edinburgh | COVID-19 Genomics UK (COG-UK) Consortium |
| EPI_ISL_461733 | Virology Department, Royal Infirmary of Edinburgh, NHS Lothian / School of Biological Sciences, University of Edinburgh / Institute of Genetics and Molecular Medicine, University of Edinburgh | COVID-19 Genomics UK (COG-UK) Consortium |
| EPI_ISL_461734 | Virology Department, Royal Infirmary of Edinburgh, NHS Lothian / School of Biological Sciences, University of Edinburgh / Institute of Genetics and Molecular Medicine, University of Edinburgh | COVID-19 Genomics UK (COG-UK) Consortium |
| EPI_ISL_461735 | Virology Department, Royal Infirmary of Edinburgh, NHS Lothian / School of Biological Sciences, University of Edinburgh / Institute of Genetics and Molecular Medicine, University of Edinburgh | COVID-19 Genomics UK (COG-UK) Consortium |

| EPI_Accession_ID | Originating Laboratory | Submitting Laboratory |
|---|---|---|
| EPI_ISL_461736 | Virology Department, Royal Infirmary of Edinburgh, NHS Lothian / School of Biological Sciences, University of Edinburgh / Institute of Genetics and Molecular Medicine, University of Edinburgh | COVID-19 Genomics UK (COG-UK) Consortium |
| EPI_ISL_461737 | Virology Department, Royal Infirmary of Edinburgh, NHS Lothian / School of Biological Sciences, University of Edinburgh / Institute of Genetics and Molecular Medicine, University of Edinburgh | COVID-19 Genomics UK (COG-UK) Consortium |
| EPI_ISL_461738 | Virology Department, Royal Infirmary of Edinburgh, NHS Lothian / School of Biological Sciences, University of Edinburgh / Institute of Genetics and Molecular Medicine, University of Edinburgh | COVID-19 Genomics UK (COG-UK) Consortium |
| EPI_ISL_461739 | Virology Department, Royal Infirmary of Edinburgh, NHS Lothian / School of Biological Sciences, University of Edinburgh / Institute of Genetics and Molecular Medicine, University of Edinburgh | COVID-19 Genomics UK (COG-UK) Consortium |
| EPI_ISL_461740 | Virology Department, Royal Infirmary of Edinburgh, NHS Lothian / School of Biological Sciences, University of Edinburgh / Institute of Genetics and Molecular Medicine, University of Edinburgh | COVID-19 Genomics UK (COG-UK) Consortium |
| EPI_ISL_461741 | Virology Department, Royal Infirmary of Edinburgh, NHS Lothian / School of Biological Sciences, University of Edinburgh / Institute of Genetics and Molecular Medicine, University of Edinburgh | COVID-19 Genomics UK (COG-UK) Consortium |
| EPI_ISL_461742 | Virology Department, Royal Infirmary of Edinburgh, NHS Lothian / School of Biological Sciences, University of Edinburgh / Institute of Genetics and Molecular Medicine, University of Edinburgh | COVID-19 Genomics UK (COG-UK) Consortium |
| EPI_ISL_461743 | Virology Department, Royal Infirmary of Edinburgh, NHS Lothian / School of Biological Sciences, University of Edinburgh / Institute of Genetics and Molecular Medicine, University of Edinburgh | COVID-19 Genomics UK (COG-UK) Consortium |

| EPI_Accession_ID | Originating Laboratory | Submitting Laboratory |
|---|---|---|
| EPI_ISL_461744 | Virology Department, Royal Infirmary of Edinburgh, NHS Lothian / School of Biological Sciences, University of Edinburgh / Institute of Genetics and Molecular Medicine, University of Edinburgh | COVID-19 Genomics UK (COG-UK) Consortium |
| EPI_ISL_461745 | Virology Department, Royal Infirmary of Edinburgh, NHS Lothian / School of Biological Sciences, University of Edinburgh / Institute of Genetics and Molecular Medicine, University of Edinburgh | COVID-19 Genomics UK (COG-UK) Consortium |
| EPI_ISL_461746 | Virology Department, Royal Infirmary of Edinburgh, NHS Lothian / School of Biological Sciences, University of Edinburgh / Institute of Genetics and Molecular Medicine, University of Edinburgh | COVID-19 Genomics UK (COG-UK) Consortium |
| EPI_ISL_461747 | Virology Department, Royal Infirmary of Edinburgh, NHS Lothian / School of Biological Sciences, University of Edinburgh / Institute of Genetics and Molecular Medicine, University of Edinburgh | COVID-19 Genomics UK (COG-UK) Consortium |
| EPI_ISL_461748 | Virology Department, Royal Infirmary of Edinburgh, NHS Lothian / School of Biological Sciences, University of Edinburgh / Institute of Genetics and Molecular Medicine, University of Edinburgh | COVID-19 Genomics UK (COG-UK) Consortium |
| EPI_ISL_461749 | Virology Department, Royal Infirmary of Edinburgh, NHS Lothian / School of Biological Sciences, University of Edinburgh / Institute of Genetics and Molecular Medicine, University of Edinburgh | COVID-19 Genomics UK (COG-UK) Consortium |
| EPI_ISL_461750 | Virology Department, Royal Infirmary of Edinburgh, NHS Lothian / School of Biological Sciences, University of Edinburgh / Institute of Genetics and Molecular Medicine, University of Edinburgh | COVID-19 Genomics UK (COG-UK) Consortium |
| EPI_ISL_461751 | Virology Department, Royal Infirmary of Edinburgh, NHS Lothian / School of Biological Sciences, University of Edinburgh / Institute of Genetics and Molecular Medicine, University of Edinburgh | COVID-19 Genomics UK (COG-UK) Consortium |

| EPI_Accession_ID | Originating Laboratory | Submitting Laboratory |
|---|---|---|
| EPI_ISL_461752 | Virology Department, Royal Infirmary of Edinburgh, NHS Lothian / School of Biological Sciences, University of Edinburgh / Institute of Genetics and Molecular Medicine, University of Edinburgh | COVID-19 Genomics UK (COG-UK) Consortium |
| EPI_ISL_461753 | Virology Department, Royal Infirmary of Edinburgh, NHS Lothian / School of Biological Sciences, University of Edinburgh / Institute of Genetics and Molecular Medicine, University of Edinburgh | COVID-19 Genomics UK (COG-UK) Consortium |
| EPI_ISL_461754 | Virology Department, Royal Infirmary of Edinburgh, NHS Lothian / School of Biological Sciences, University of Edinburgh / Institute of Genetics and Molecular Medicine, University of Edinburgh | COVID-19 Genomics UK (COG-UK) Consortium |
| EPI_ISL_461755 | Virology Department, Royal Infirmary of Edinburgh, NHS Lothian / School of Biological Sciences, University of Edinburgh / Institute of Genetics and Molecular Medicine, University of Edinburgh | COVID-19 Genomics UK (COG-UK) Consortium |
| EPI_ISL_461756 | Virology Department, Royal Infirmary of Edinburgh, NHS Lothian / School of Biological Sciences, University of Edinburgh / Institute of Genetics and Molecular Medicine, University of Edinburgh | COVID-19 Genomics UK (COG-UK) Consortium |
| EPI_ISL_461757 | Virology Department, Royal Infirmary of Edinburgh, NHS Lothian / School of Biological Sciences, University of Edinburgh / Institute of Genetics and Molecular Medicine, University of Edinburgh | COVID-19 Genomics UK (COG-UK) Consortium |
| EPI_ISL_461758 | Virology Department, Royal Infirmary of Edinburgh, NHS Lothian / School of Biological Sciences, University of Edinburgh / Institute of Genetics and Molecular Medicine, University of Edinburgh | COVID-19 Genomics UK (COG-UK) Consortium |
| EPI_ISL_461759 | Virology Department, Royal Infirmary of Edinburgh, NHS Lothian / School of Biological Sciences, University of Edinburgh / Institute of Genetics and Molecular Medicine, University of Edinburgh | COVID-19 Genomics UK (COG-UK) Consortium |

| EPI_Accession_ID | Originating Laboratory | Submitting Laboratory |
|---|---|---|
| EPI_ISL_461760 | Virology Department, Royal Infirmary of Edinburgh, NHS Lothian / School of Biological Sciences, University of Edinburgh / Institute of Genetics and Molecular Medicine, University of Edinburgh | COVID-19 Genomics UK (COG-UK) Consortium |
| EPI_ISL_461761 | Virology Department, Royal Infirmary of Edinburgh, NHS Lothian / School of Biological Sciences, University of Edinburgh / Institute of Genetics and Molecular Medicine, University of Edinburgh | COVID-19 Genomics UK (COG-UK) Consortium |
| EPI_ISL_461762 | Virology Department, Royal Infirmary of Edinburgh, NHS Lothian / School of Biological Sciences, University of Edinburgh / Institute of Genetics and Molecular Medicine, University of Edinburgh | COVID-19 Genomics UK (COG-UK) Consortium |
| EPI_ISL_461763 | University College London, Great Ormond Street Hospital for Children NHS Foundation Trust, Imperial College Healthcare NHS Trust | COVID-19 Genomics UK (COG-UK) Consortium |
| EPI_ISL_461764 | University College London, Great Ormond Street Hospital for Children NHS Foundation Trust, Imperial College Healthcare NHS Trust | COVID-19 Genomics UK (COG-UK) Consortium |
| EPI_ISL_461765 | University College London, Great Ormond Street Hospital for Children NHS Foundation Trust, Imperial College Healthcare NHS Trust | COVID-19 Genomics UK (COG-UK) Consortium |
| EPI_ISL_461766 | University College London, Great Ormond Street Hospital for Children NHS Foundation Trust, Imperial College Healthcare NHS Trust | COVID-19 Genomics UK (COG-UK) Consortium |
| EPI_ISL_461767 | University College London, Great Ormond Street Hospital for Children NHS Foundation Trust, Imperial College Healthcare NHS Trust | COVID-19 Genomics UK (COG-UK) Consortium |
| EPI_ISL_461768 | University College London, Great Ormond Street Hospital for Children NHS Foundation Trust, Imperial College Healthcare NHS Trust | COVID-19 Genomics UK (COG-UK) Consortium |
| EPI_ISL_461769 | University College London, Great Ormond Street Hospital for Children NHS Foundation Trust, Imperial College Healthcare NHS Trust | COVID-19 Genomics UK (COG-UK) Consortium |

| EPI_Accession_ID | Originating Laboratory | Submitting Laboratory |
|---|---|---|
| EPI_ISL_461770 | University College London, Great Ormond Street Hospital for Children NHS Foundation Trust, Imperial College Healthcare NHS Trust | COVID-19 Genomics UK (COG-UK) Consortium |
| EPI_ISL_461771 | University College London, Great Ormond Street Hospital for Children NHS Foundation Trust, Imperial College Healthcare NHS Trust | COVID-19 Genomics UK (COG-UK) Consortium |
| EPI_ISL_461772 | University College London, Great Ormond Street Hospital for Children NHS Foundation Trust, Imperial College Healthcare NHS Trust | COVID-19 Genomics UK (COG-UK) Consortium |
| EPI_ISL_461773 | Regional Virus Laboratory, Belfast Health and Social Care Trust | COVID-19 Genomics UK (COG-UK) Consortium |
| EPI_ISL_461774 | Regional Virus Laboratory, Belfast Health and Social Care Trust | COVID-19 Genomics UK (COG-UK) Consortium |
| EPI_ISL_461775 | Regional Virus Laboratory, Belfast Health and Social Care Trust | COVID-19 Genomics UK (COG-UK) Consortium |
| EPI_ISL_461776 | Regional Virus Laboratory, Belfast Health and Social Care Trust | COVID-19 Genomics UK (COG-UK) Consortium |
| EPI_ISL_461777 | Regional Virus Laboratory, Belfast Health and Social Care Trust | COVID-19 Genomics UK (COG-UK) Consortium |
| EPI_ISL_461778 | Regional Virus Laboratory, Belfast Health and Social Care Trust | COVID-19 Genomics UK (COG-UK) Consortium |
| EPI_ISL_461779 | Regional Virus Laboratory, Belfast Health and Social Care Trust | COVID-19 Genomics UK (COG-UK) Consortium |
| EPI_ISL_461780 | Regional Virus Laboratory, Belfast Health and Social Care Trust | COVID-19 Genomics UK (COG-UK) Consortium |
| EPI_ISL_461781 | Regional Virus Laboratory, Belfast Health and Social Care Trust | COVID-19 Genomics UK (COG-UK) Consortium |
| EPI_ISL_461782 | Regional Virus Laboratory, Belfast Health and Social Care Trust | COVID-19 Genomics UK (COG-UK) Consortium |
| EPI_ISL_461783 | Regional Virus Laboratory, Belfast Health and Social Care Trust | COVID-19 Genomics UK (COG-UK) Consortium |
| EPI_ISL_461784 | Regional Virus Laboratory, Belfast Health and Social Care Trust | COVID-19 Genomics UK (COG-UK) Consortium |

| EPI_Accession_ID | Originating Laboratory | Submitting Laboratory |
|---|---|---|
| EPI_ISL_461785 | Regional Virus Laboratory, Belfast Health and Social Care Trust | COVID-19 Genomics UK (COG-UK) Consortium |
| EPI_ISL_461786 | Regional Virus Laboratory, Belfast Health and Social Care Trust | COVID-19 Genomics UK (COG-UK) Consortium |
| EPI_ISL_461787 | Regional Virus Laboratory, Belfast Health and Social Care Trust | COVID-19 Genomics UK (COG-UK) Consortium |
| EPI_ISL_461788 | Regional Virus Laboratory, Belfast Health and Social Care Trust | COVID-19 Genomics UK (COG-UK) Consortium |
| EPI_ISL_461789 | Regional Virus Laboratory, Belfast Health and Social Care Trust | COVID-19 Genomics UK (COG-UK) Consortium |
| EPI_ISL_461790 | Regional Virus Laboratory, Belfast Health and Social Care Trust | COVID-19 Genomics UK (COG-UK) Consortium |
| EPI_ISL_461791 | Northumbria University / South Tees Hospitals NHS Foundation Trust / North Cumbria Integrated Care NHS Foundation Trust / North Tees and Hartlepool NHS Foundation Trust / Newcastle Hospitals NHS Foundation Trust | COVID-19 Genomics UK (COG-UK) Consortium |
| EPI_ISL_461792 | Northumbria University / South Tees Hospitals NHS Foundation Trust / North Cumbria Integrated Care NHS Foundation Trust / North Tees and Hartlepool NHS Foundation Trust / Newcastle Hospitals NHS Foundation Trust | COVID-19 Genomics UK (COG-UK) Consortium |
| EPI_ISL_461793 | Northumbria University / South Tees Hospitals NHS Foundation Trust / North Cumbria Integrated Care NHS Foundation Trust / North Tees and Hartlepool NHS Foundation Trust / Newcastle Hospitals NHS Foundation Trust | COVID-19 Genomics UK (COG-UK) Consortium |
| EPI_ISL_461794 | Northumbria University / South Tees Hospitals NHS Foundation Trust / North Cumbria Integrated Care NHS Foundation Trust / North Tees and Hartlepool NHS Foundation Trust / Newcastle Hospitals NHS Foundation Trust | COVID-19 Genomics UK (COG-UK) Consortium |

| EPI_Accession_ID | Originating Laboratory | Submitting Laboratory |
|---|---|---|
| EPI_ISL_461795 | Northumbria University / South Tees Hospitals NHS Foundation Trust / North Cumbria Integrated Care NHS Foundation Trust / North Tees and Hartlepool NHS Foundation Trust / Newcastle Hospitals NHS Foundation Trust | COVID-19 Genomics UK (COG-UK) Consortium |
| EPI_ISL_461796 | Northumbria University / South Tees Hospitals NHS Foundation Trust / North Cumbria Integrated Care NHS Foundation Trust / North Tees and Hartlepool NHS Foundation Trust / Newcastle Hospitals NHS Foundation Trust | COVID-19 Genomics UK (COG-UK) Consortium |
| EPI_ISL_461797 | Northumbria University / South Tees Hospitals NHS Foundation Trust / North Cumbria Integrated Care NHS Foundation Trust / North Tees and Hartlepool NHS Foundation Trust / Newcastle Hospitals NHS Foundation Trust | COVID-19 Genomics UK (COG-UK) Consortium |
| EPI_ISL_461798 | Northumbria University / South Tees Hospitals NHS Foundation Trust / North Cumbria Integrated Care NHS Foundation Trust / North Tees and Hartlepool NHS Foundation Trust / Newcastle Hospitals NHS Foundation Trust | COVID-19 Genomics UK (COG-UK) Consortium |
| EPI_ISL_461799 | Northumbria University / South Tees Hospitals NHS Foundation Trust / North Cumbria Integrated Care NHS Foundation Trust / North Tees and Hartlepool NHS Foundation Trust / Newcastle Hospitals NHS Foundation Trust | COVID-19 Genomics UK (COG-UK) Consortium |
| EPI_ISL_461800 | Northumbria University / South Tees Hospitals NHS Foundation Trust / North Cumbria Integrated Care NHS Foundation Trust / North Tees and Hartlepool NHS Foundation Trust / Newcastle Hospitals NHS Foundation Trust | COVID-19 Genomics UK (COG-UK) Consortium |

| EPI_Accession_ID | Originating Laboratory | Submitting Laboratory |
|---|---|---|
| EPI_ISL_461801 | Northumbria University / South Tees Hospitals NHS Foundation Trust / North Cumbria Integrated Care NHS Foundation Trust / North Tees and Hartlepool NHS Foundation Trust / Newcastle Hospitals NHS Foundation Trust | COVID-19 Genomics UK (COG-UK) Consortium |
| EPI_ISL_461802 | Northumbria University / South Tees Hospitals NHS Foundation Trust / North Cumbria Integrated Care NHS Foundation Trust / North Tees and Hartlepool NHS Foundation Trust / Newcastle Hospitals NHS Foundation Trust | COVID-19 Genomics UK (COG-UK) Consortium |
| EPI_ISL_461803 | Northumbria University / South Tees Hospitals NHS Foundation Trust / North Cumbria Integrated Care NHS Foundation Trust / North Tees and Hartlepool NHS Foundation Trust / Newcastle Hospitals NHS Foundation Trust | COVID-19 Genomics UK (COG-UK) Consortium |
| EPI_ISL_461804 | Northumbria University / South Tees Hospitals NHS Foundation Trust / North Cumbria Integrated Care NHS Foundation Trust / North Tees and Hartlepool NHS Foundation Trust / Newcastle Hospitals NHS Foundation Trust | COVID-19 Genomics UK (COG-UK) Consortium |
| EPI_ISL_461805 | Quadram Institute Bioscience | COVID-19 Genomics UK (COG-UK) Consortium |
| EPI_ISL_461806 | Quadram Institute Bioscience | COVID-19 Genomics UK (COG-UK) Consortium |
| EPI_ISL_461807 | Quadram Institute Bioscience | COVID-19 Genomics UK (COG-UK) Consortium |
| EPI_ISL_461808 | Quadram Institute Bioscience | COVID-19 Genomics UK (COG-UK) Consortium |
| EPI_ISL_461809 | Quadram Institute Bioscience | COVID-19 Genomics UK (COG-UK) Consortium |
| EPI_ISL_461810 | Quadram Institute Bioscience | COVID-19 Genomics UK (COG-UK) Consortium |
| EPI_ISL_461811 | Quadram Institute Bioscience | COVID-19 Genomics UK (COG-UK) Consortium |
| EPI_ISL_461812 | Quadram Institute Bioscience | COVID-19 Genomics UK (COG-UK) Consortium |
| EPI_ISL_461813 | Quadram Institute Bioscience | COVID-19 Genomics UK (COG-UK) Consortium |
| EPI_ISL_461814 | Quadram Institute Bioscience | COVID-19 Genomics UK (COG-UK) Consortium |
| EPI_ISL_461815 | Quadram Institute Bioscience | COVID-19 Genomics UK (COG-UK) Consortium |
| EPI_ISL_461816 | Quadram Institute Bioscience | COVID-19 Genomics UK (COG-UK) Consortium |
| EPI_ISL_461817 | Quadram Institute Bioscience | COVID-19 Genomics UK (COG-UK) Consortium |

| EPI_Accession_ID | Originating Laboratory | Submitting Laboratory |
|---|---|---|
| EPI_ISL_461818 | Quadram Institute Bioscience | COVID-19 Genomics UK (COG-UK) Consortium |
| EPI_ISL_461819 | Quadram Institute Bioscience | COVID-19 Genomics UK (COG-UK) Consortium |
| EPI_ISL_461820 | Quadram Institute Bioscience | COVID-19 Genomics UK (COG-UK) Consortium |
| EPI_ISL_461821 | Quadram Institute Bioscience | COVID-19 Genomics UK (COG-UK) Consortium |
| EPI_ISL_461822 | Quadram Institute Bioscience | COVID-19 Genomics UK (COG-UK) Consortium |
| EPI_ISL_461823 | Quadram Institute Bioscience | COVID-19 Genomics UK (COG-UK) Consortium |
| EPI_ISL_461824 | Quadram Institute Bioscience | COVID-19 Genomics UK (COG-UK) Consortium |
| EPI_ISL_461825 | Quadram Institute Bioscience | COVID-19 Genomics UK (COG-UK) Consortium |
| EPI_ISL_461826 | Quadram Institute Bioscience | COVID-19 Genomics UK (COG-UK) Consortium |
| EPI_ISL_461827 | Quadram Institute Bioscience | COVID-19 Genomics UK (COG-UK) Consortium |
| EPI_ISL_461828 | Quadram Institute Bioscience | COVID-19 Genomics UK (COG-UK) Consortium |
| EPI_ISL_461829 | Quadram Institute Bioscience | COVID-19 Genomics UK (COG-UK) Consortium |
| EPI_ISL_461830 | Quadram Institute Bioscience | COVID-19 Genomics UK (COG-UK) Consortium |
| EPI_ISL_461831 | Quadram Institute Bioscience | COVID-19 Genomics UK (COG-UK) Consortium |
| EPI_ISL_461832 | Quadram Institute Bioscience | COVID-19 Genomics UK (COG-UK) Consortium |
| EPI_ISL_461833 | Quadram Institute Bioscience | COVID-19 Genomics UK (COG-UK) Consortium |
| EPI_ISL_461834 | Quadram Institute Bioscience | COVID-19 Genomics UK (COG-UK) Consortium |
| EPI_ISL_461835 | Quadram Institute Bioscience | COVID-19 Genomics UK (COG-UK) Consortium |
| EPI_ISL_461836 | Quadram Institute Bioscience | COVID-19 Genomics UK (COG-UK) Consortium |
| EPI_ISL_461837 | Quadram Institute Bioscience | COVID-19 Genomics UK (COG-UK) Consortium |
| EPI_ISL_461838 | Quadram Institute Bioscience | COVID-19 Genomics UK (COG-UK) Consortium |
| EPI_ISL_461839 | Quadram Institute Bioscience | COVID-19 Genomics UK (COG-UK) Consortium |
| EPI_ISL_461840 | Quadram Institute Bioscience | COVID-19 Genomics UK (COG-UK) Consortium |
| EPI_ISL_461841 | Quadram Institute Bioscience | COVID-19 Genomics UK (COG-UK) Consortium |
| EPI_ISL_461842 | Quadram Institute Bioscience | COVID-19 Genomics UK (COG-UK) Consortium |
| EPI_ISL_461843 | Quadram Institute Bioscience | COVID-19 Genomics UK (COG-UK) Consortium |
| EPI_ISL_461844 | Quadram Institute Bioscience | COVID-19 Genomics UK (COG-UK) Consortium |
| EPI_ISL_461845 | Quadram Institute Bioscience | COVID-19 Genomics UK (COG-UK) Consortium |
| EPI_ISL_461846 | Quadram Institute Bioscience | COVID-19 Genomics UK (COG-UK) Consortium |
| EPI_ISL_461847 | Quadram Institute Bioscience | COVID-19 Genomics UK (COG-UK) Consortium |
| EPI_ISL_461848 | Quadram Institute Bioscience | COVID-19 Genomics UK (COG-UK) Consortium |
| EPI_ISL_461849 | Quadram Institute Bioscience | COVID-19 Genomics UK (COG-UK) Consortium |
| EPI_ISL_461850 | Quadram Institute Bioscience | COVID-19 Genomics UK (COG-UK) Consortium |

| EPI_Accession_ID | Originating Laboratory | Submitting Laboratory |
|---|---|---|
| EPI_ISL_461851 | Quadram Institute Bioscience | COVID-19 Genomics UK (COG-UK) Consortium |
| EPI_ISL_461852 | Quadram Institute Bioscience | COVID-19 Genomics UK (COG-UK) Consortium |
| EPI_ISL_461853 | Quadram Institute Bioscience | COVID-19 Genomics UK (COG-UK) Consortium |
| EPI_ISL_461854 | Quadram Institute Bioscience | COVID-19 Genomics UK (COG-UK) Consortium |
| EPI_ISL_461855 | Quadram Institute Bioscience | COVID-19 Genomics UK (COG-UK) Consortium |
| EPI_ISL_461856 | Quadram Institute Bioscience | COVID-19 Genomics UK (COG-UK) Consortium |
| EPI_ISL_461857 | Quadram Institute Bioscience | COVID-19 Genomics UK (COG-UK) Consortium |
| EPI_ISL_461858 | Quadram Institute Bioscience | COVID-19 Genomics UK (COG-UK) Consortium |
| EPI_ISL_461859 | Quadram Institute Bioscience | COVID-19 Genomics UK (COG-UK) Consortium |
| EPI_ISL_461860 | Quadram Institute Bioscience | COVID-19 Genomics UK (COG-UK) Consortium |
| EPI_ISL_461861 | Quadram Institute Bioscience | COVID-19 Genomics UK (COG-UK) Consortium |
| EPI_ISL_461862 | Quadram Institute Bioscience | COVID-19 Genomics UK (COG-UK) Consortium |
| EPI_ISL_461863 | Quadram Institute Bioscience | COVID-19 Genomics UK (COG-UK) Consortium |
| EPI_ISL_461864 | Quadram Institute Bioscience | COVID-19 Genomics UK (COG-UK) Consortium |
| EPI_ISL_461865 | Quadram Institute Bioscience | COVID-19 Genomics UK (COG-UK) Consortium |
| EPI_ISL_461866 | Quadram Institute Bioscience | COVID-19 Genomics UK (COG-UK) Consortium |
| EPI_ISL_461867 | Quadram Institute Bioscience | COVID-19 Genomics UK (COG-UK) Consortium |
| EPI_ISL_461868 | Quadram Institute Bioscience | COVID-19 Genomics UK (COG-UK) Consortium |
| EPI_ISL_461869 | Quadram Institute Bioscience | COVID-19 Genomics UK (COG-UK) Consortium |
| EPI_ISL_461870 | Quadram Institute Bioscience | COVID-19 Genomics UK (COG-UK) Consortium |
| EPI_ISL_461871 | Quadram Institute Bioscience | COVID-19 Genomics UK (COG-UK) Consortium |
| EPI_ISL_461872 | Quadram Institute Bioscience | COVID-19 Genomics UK (COG-UK) Consortium |
| EPI_ISL_461873 | Quadram Institute Bioscience | COVID-19 Genomics UK (COG-UK) Consortium |
| EPI_ISL_461874 | Quadram Institute Bioscience | COVID-19 Genomics UK (COG-UK) Consortium |
| EPI_ISL_461875 | Quadram Institute Bioscience | COVID-19 Genomics UK (COG-UK) Consortium |
| EPI_ISL_461876 | Quadram Institute Bioscience | COVID-19 Genomics UK (COG-UK) Consortium |
| EPI_ISL_461877 | Quadram Institute Bioscience | COVID-19 Genomics UK (COG-UK) Consortium |
| EPI_ISL_461878 | Quadram Institute Bioscience | COVID-19 Genomics UK (COG-UK) Consortium |
| EPI_ISL_461879 | Quadram Institute Bioscience | COVID-19 Genomics UK (COG-UK) Consortium |
| EPI_ISL_461880 | Quadram Institute Bioscience | COVID-19 Genomics UK (COG-UK) Consortium |
| EPI_ISL_461881 | Quadram Institute Bioscience | COVID-19 Genomics UK (COG-UK) Consortium |
| EPI_ISL_461882 | Quadram Institute Bioscience | COVID-19 Genomics UK (COG-UK) Consortium |
| EPI_ISL_461883 | Quadram Institute Bioscience | COVID-19 Genomics UK (COG-UK) Consortium |

| EPI_Accession_ID | Originating Laboratory | Submitting Laboratory |
|---|---|---|
| EPI_ISL_461884 | Quadram Institute Bioscience | COVID-19 Genomics UK (COG-UK) Consortium |
| EPI_ISL_461885 | Quadram Institute Bioscience | COVID-19 Genomics UK (COG-UK) Consortium |
| EPI_ISL_461886 | Quadram Institute Bioscience | COVID-19 Genomics UK (COG-UK) Consortium |
| EPI_ISL_461887 | Quadram Institute Bioscience | COVID-19 Genomics UK (COG-UK) Consortium |
| EPI_ISL_461888 | Quadram Institute Bioscience | COVID-19 Genomics UK (COG-UK) Consortium |
| EPI_ISL_461889 | Quadram Institute Bioscience | COVID-19 Genomics UK (COG-UK) Consortium |
| EPI_ISL_461890 | Quadram Institute Bioscience | COVID-19 Genomics UK (COG-UK) Consortium |
| EPI_ISL_461891 | Quadram Institute Bioscience | COVID-19 Genomics UK (COG-UK) Consortium |
| EPI_ISL_461892 | Quadram Institute Bioscience | COVID-19 Genomics UK (COG-UK) Consortium |
| EPI_ISL_461893 | Quadram Institute Bioscience | COVID-19 Genomics UK (COG-UK) Consortium |
| EPI_ISL_461894 | Queens Medical Centre, Clinical Microbiology Department / DeepSeq Nottingham | COVID-19 Genomics UK (COG-UK) Consortium |
| EPI_ISL_461895 | Queens Medical Centre, Clinical Microbiology Department / DeepSeq Nottingham | COVID-19 Genomics UK (COG-UK) Consortium |
| EPI_ISL_461896 | Queens Medical Centre, Clinical Microbiology Department / DeepSeq Nottingham | COVID-19 Genomics UK (COG-UK) Consortium |
| EPI_ISL_461897 | Queens Medical Centre, Clinical Microbiology Department / DeepSeq Nottingham | COVID-19 Genomics UK (COG-UK) Consortium |
| EPI_ISL_461898 | Queens Medical Centre, Clinical Microbiology Department / DeepSeq Nottingham | COVID-19 Genomics UK (COG-UK) Consortium |
| EPI_ISL_461899 | Queens Medical Centre, Clinical Microbiology Department / DeepSeq Nottingham | COVID-19 Genomics UK (COG-UK) Consortium |
| EPI_ISL_461900 | Queens Medical Centre, Clinical Microbiology Department / DeepSeq Nottingham | COVID-19 Genomics UK (COG-UK) Consortium |
| EPI_ISL_461901 | Queens Medical Centre, Clinical Microbiology Department / DeepSeq Nottingham | COVID-19 Genomics UK (COG-UK) Consortium |
| EPI_ISL_461902 | Queens Medical Centre, Clinical Microbiology Department / DeepSeq Nottingham | COVID-19 Genomics UK (COG-UK) Consortium |
| EPI_ISL_461903 | Queens Medical Centre, Clinical Microbiology Department / DeepSeq Nottingham | COVID-19 Genomics UK (COG-UK) Consortium |
| EPI_ISL_461904 | Queens Medical Centre, Clinical Microbiology Department / DeepSeq Nottingham | COVID-19 Genomics UK (COG-UK) Consortium |

| EPI_Accession_ID | Originating Laboratory | Submitting Laboratory |
|---|---|---|
| EPI_ISL_461905 | Queens Medical Centre, Clinical Microbiology Department / DeepSeq Nottingham | COVID-19 Genomics UK (COG-UK) Consortium |
| EPI_ISL_461906 | Queens Medical Centre, Clinical Microbiology Department / DeepSeq Nottingham | COVID-19 Genomics UK (COG-UK) Consortium |
| EPI_ISL_461907 | Queens Medical Centre, Clinical Microbiology Department / DeepSeq Nottingham | COVID-19 Genomics UK (COG-UK) Consortium |
| EPI_ISL_461908 | Queens Medical Centre, Clinical Microbiology Department / DeepSeq Nottingham | COVID-19 Genomics UK (COG-UK) Consortium |
| EPI_ISL_461909 | Queens Medical Centre, Clinical Microbiology Department / DeepSeq Nottingham | COVID-19 Genomics UK (COG-UK) Consortium |
| EPI_ISL_461910 | Queens Medical Centre, Clinical Microbiology Department / DeepSeq Nottingham | COVID-19 Genomics UK (COG-UK) Consortium |
| EPI_ISL_461911 | Queens Medical Centre, Clinical Microbiology Department / DeepSeq Nottingham | COVID-19 Genomics UK (COG-UK) Consortium |
| EPI_ISL_461912 | Queens Medical Centre, Clinical Microbiology Department / DeepSeq Nottingham | COVID-19 Genomics UK (COG-UK) Consortium |
| EPI_ISL_461913 | Queens Medical Centre, Clinical Microbiology Department / DeepSeq Nottingham | COVID-19 Genomics UK (COG-UK) Consortium |
| EPI_ISL_461914 | Queens Medical Centre, Clinical Microbiology Department / DeepSeq Nottingham | COVID-19 Genomics UK (COG-UK) Consortium |
| EPI_ISL_461915 | Queens Medical Centre, Clinical Microbiology Department / DeepSeq Nottingham | COVID-19 Genomics UK (COG-UK) Consortium |
| EPI_ISL_461916 | Queens Medical Centre, Clinical Microbiology Department / DeepSeq Nottingham | COVID-19 Genomics UK (COG-UK) Consortium |
| EPI_ISL_461917 | Queens Medical Centre, Clinical Microbiology Department / DeepSeq Nottingham | COVID-19 Genomics UK (COG-UK) Consortium |
| EPI_ISL_461918 | Queens Medical Centre, Clinical Microbiology Department / DeepSeq Nottingham | COVID-19 Genomics UK (COG-UK) Consortium |
| EPI_ISL_461919 | Queens Medical Centre, Clinical Microbiology Department / DeepSeq Nottingham | COVID-19 Genomics UK (COG-UK) Consortium |
| EPI_ISL_461920 | Queens Medical Centre, Clinical Microbiology Department / DeepSeq Nottingham | COVID-19 Genomics UK (COG-UK) Consortium |

| EPI_Accession_ID | Originating Laboratory | Submitting Laboratory |
|---|---|---|
| EPI_ISL_461921 | Queens Medical Centre, Clinical Microbiology Department / DeepSeq Nottingham | COVID-19 Genomics UK (COG-UK) Consortium |
| EPI_ISL_461922 | Queens Medical Centre, Clinical Microbiology Department / DeepSeq Nottingham | COVID-19 Genomics UK (COG-UK) Consortium |
| EPI_ISL_461923 | Queens Medical Centre, Clinical Microbiology Department / DeepSeq Nottingham | COVID-19 Genomics UK (COG-UK) Consortium |
| EPI_ISL_461924 | Queens Medical Centre, Clinical Microbiology Department / DeepSeq Nottingham | COVID-19 Genomics UK (COG-UK) Consortium |
| EPI_ISL_461925 | Queens Medical Centre, Clinical Microbiology Department / DeepSeq Nottingham | COVID-19 Genomics UK (COG-UK) Consortium |
| EPI_ISL_461926 | Queens Medical Centre, Clinical Microbiology Department / DeepSeq Nottingham | COVID-19 Genomics UK (COG-UK) Consortium |
| EPI_ISL_461927 | Queens Medical Centre, Clinical Microbiology Department / DeepSeq Nottingham | COVID-19 Genomics UK (COG-UK) Consortium |
| EPI_ISL_461928 | Queens Medical Centre, Clinical Microbiology Department / DeepSeq Nottingham | COVID-19 Genomics UK (COG-UK) Consortium |
| EPI_ISL_461929 | Queens Medical Centre, Clinical Microbiology Department / DeepSeq Nottingham | COVID-19 Genomics UK (COG-UK) Consortium |
| EPI_ISL_461930 | Queens Medical Centre, Clinical Microbiology Department / DeepSeq Nottingham | COVID-19 Genomics UK (COG-UK) Consortium |
| EPI_ISL_461931 | Queens Medical Centre, Clinical Microbiology Department / DeepSeq Nottingham | COVID-19 Genomics UK (COG-UK) Consortium |
| EPI_ISL_461932 | Queens Medical Centre, Clinical Microbiology Department / DeepSeq Nottingham | COVID-19 Genomics UK (COG-UK) Consortium |
| EPI_ISL_461933 | Queens Medical Centre, Clinical Microbiology Department / DeepSeq Nottingham | COVID-19 Genomics UK (COG-UK) Consortium |
| EPI_ISL_461934 | Queens Medical Centre, Clinical Microbiology Department / DeepSeq Nottingham | COVID-19 Genomics UK (COG-UK) Consortium |
| EPI_ISL_461935 | Queens Medical Centre, Clinical Microbiology Department / DeepSeq Nottingham | COVID-19 Genomics UK (COG-UK) Consortium |
| EPI_ISL_461936 | Queens Medical Centre, Clinical Microbiology Department / DeepSeq Nottingham | COVID-19 Genomics UK (COG-UK) Consortium |

| EPI_Accession_ID | Originating Laboratory | Submitting Laboratory |
| --- | --- | --- |
| EPI_ISL_461937 | Queens Medical Centre, Clinical Microbiology Department / DeepSeq Nottingham | COVID-19 Genomics UK (COG-UK) Consortium |
| EPI_ISL_461938 | Queens Medical Centre, Clinical Microbiology Department / DeepSeq Nottingham | COVID-19 Genomics UK (COG-UK) Consortium |
| EPI_ISL_461939 | Queens Medical Centre, Clinical Microbiology Department / DeepSeq Nottingham | COVID-19 Genomics UK (COG-UK) Consortium |
| EPI_ISL_461940 | Queens Medical Centre, Clinical Microbiology Department / DeepSeq Nottingham | COVID-19 Genomics UK (COG-UK) Consortium |
| EPI_ISL_461941 | Queens Medical Centre, Clinical Microbiology Department / DeepSeq Nottingham | COVID-19 Genomics UK (COG-UK) Consortium |
| EPI_ISL_461942 | Queens Medical Centre, Clinical Microbiology Department / DeepSeq Nottingham | COVID-19 Genomics UK (COG-UK) Consortium |
| EPI_ISL_461943 | Queens Medical Centre, Clinical Microbiology Department / DeepSeq Nottingham | COVID-19 Genomics UK (COG-UK) Consortium |
| EPI_ISL_461944 | Queens Medical Centre, Clinical Microbiology Department / DeepSeq Nottingham | COVID-19 Genomics UK (COG-UK) Consortium |
| EPI_ISL_461945 | Queens Medical Centre, Clinical Microbiology Department / DeepSeq Nottingham | COVID-19 Genomics UK (COG-UK) Consortium |
| EPI_ISL_461946 | Queens Medical Centre, Clinical Microbiology Department / DeepSeq Nottingham | COVID-19 Genomics UK (COG-UK) Consortium |
| EPI_ISL_461947 | Queens Medical Centre, Clinical Microbiology Department / DeepSeq Nottingham | COVID-19 Genomics UK (COG-UK) Consortium |
| EPI_ISL_461948 | Queens Medical Centre, Clinical Microbiology Department / DeepSeq Nottingham | COVID-19 Genomics UK (COG-UK) Consortium |
| EPI_ISL_461949 | Queens Medical Centre, Clinical Microbiology Department / DeepSeq Nottingham | COVID-19 Genomics UK (COG-UK) Consortium |
| EPI_ISL_461950 | Queens Medical Centre, Clinical Microbiology Department / DeepSeq Nottingham | COVID-19 Genomics UK (COG-UK) Consortium |
| EPI_ISL_461951 | Queens Medical Centre, Clinical Microbiology Department / DeepSeq Nottingham | COVID-19 Genomics UK (COG-UK) Consortium |
| EPI_ISL_461952 | Queens Medical Centre, Clinical Microbiology Department / DeepSeq Nottingham | COVID-19 Genomics UK (COG-UK) Consortium |

| EPI_Accession_ID | Originating Laboratory | Submitting Laboratory |
|---|---|---|
| EPI_ISL_461953 | Queens Medical Centre, Clinical Microbiology Department / DeepSeq Nottingham | COVID-19 Genomics UK (COG-UK) Consortium |
| EPI_ISL_461954 | Queens Medical Centre, Clinical Microbiology Department / DeepSeq Nottingham | COVID-19 Genomics UK (COG-UK) Consortium |
| EPI_ISL_461955 | Queens Medical Centre, Clinical Microbiology Department / DeepSeq Nottingham | COVID-19 Genomics UK (COG-UK) Consortium |
| EPI_ISL_461956 | Queens Medical Centre, Clinical Microbiology Department / DeepSeq Nottingham | COVID-19 Genomics UK (COG-UK) Consortium |
| EPI_ISL_461957 | Queens Medical Centre, Clinical Microbiology Department / DeepSeq Nottingham | COVID-19 Genomics UK (COG-UK) Consortium |
| EPI_ISL_461958 | Queens Medical Centre, Clinical Microbiology Department / DeepSeq Nottingham | COVID-19 Genomics UK (COG-UK) Consortium |
| EPI_ISL_461959 | Queens Medical Centre, Clinical Microbiology Department / DeepSeq Nottingham | COVID-19 Genomics UK (COG-UK) Consortium |
| EPI_ISL_461960 | Queens Medical Centre, Clinical Microbiology Department / DeepSeq Nottingham | COVID-19 Genomics UK (COG-UK) Consortium |
| EPI_ISL_461961 | Queens Medical Centre, Clinical Microbiology Department / DeepSeq Nottingham | COVID-19 Genomics UK (COG-UK) Consortium |
| EPI_ISL_461962 | Queens Medical Centre, Clinical Microbiology Department / DeepSeq Nottingham | COVID-19 Genomics UK (COG-UK) Consortium |
| EPI_ISL_461963 | Queens Medical Centre, Clinical Microbiology Department / DeepSeq Nottingham | COVID-19 Genomics UK (COG-UK) Consortium |
| EPI_ISL_461964 | Queens Medical Centre, Clinical Microbiology Department / DeepSeq Nottingham | COVID-19 Genomics UK (COG-UK) Consortium |
| EPI_ISL_461965 | Queens Medical Centre, Clinical Microbiology Department / DeepSeq Nottingham | COVID-19 Genomics UK (COG-UK) Consortium |
| EPI_ISL_461966 | Queens Medical Centre, Clinical Microbiology Department / DeepSeq Nottingham | COVID-19 Genomics UK (COG-UK) Consortium |
| EPI_ISL_461967 | Queens Medical Centre, Clinical Microbiology Department / DeepSeq Nottingham | COVID-19 Genomics UK (COG-UK) Consortium |
| EPI_ISL_461968 | Queens Medical Centre, Clinical Microbiology Department / DeepSeq Nottingham | COVID-19 Genomics UK (COG-UK) Consortium |

| EPI_Accession_ID | Originating Laboratory | Submitting Laboratory |
|---|---|---|
| EPI_ISL_461969 | Queens Medical Centre, Clinical Microbiology Department / DeepSeq Nottingham | COVID-19 Genomics UK (COG-UK) Consortium |
| EPI_ISL_461970 | Centre for Enzyme Innovation, University of Portsmouth / Translational Research Laboratory, Portsmouth Hospitals NHS Trust | COVID-19 Genomics UK (COG-UK) Consortium |
| EPI_ISL_461971 | Centre for Enzyme Innovation, University of Portsmouth / Translational Research Laboratory, Portsmouth Hospitals NHS Trust | COVID-19 Genomics UK (COG-UK) Consortium |
| EPI_ISL_461972 | Centre for Enzyme Innovation, University of Portsmouth / Translational Research Laboratory, Portsmouth Hospitals NHS Trust | COVID-19 Genomics UK (COG-UK) Consortium |
| EPI_ISL_461973 | Centre for Enzyme Innovation, University of Portsmouth / Translational Research Laboratory, Portsmouth Hospitals NHS Trust | COVID-19 Genomics UK (COG-UK) Consortium |
| EPI_ISL_461974 | Centre for Enzyme Innovation, University of Portsmouth / Translational Research Laboratory, Portsmouth Hospitals NHS Trust | COVID-19 Genomics UK (COG-UK) Consortium |
| EPI_ISL_461975 | Centre for Enzyme Innovation, University of Portsmouth / Translational Research Laboratory, Portsmouth Hospitals NHS Trust | COVID-19 Genomics UK (COG-UK) Consortium |
| EPI_ISL_461976 | Centre for Enzyme Innovation, University of Portsmouth / Translational Research Laboratory, Portsmouth Hospitals NHS Trust | COVID-19 Genomics UK (COG-UK) Consortium |
| EPI_ISL_461977 | Centre for Enzyme Innovation, University of Portsmouth / Translational Research Laboratory, Portsmouth Hospitals NHS Trust | COVID-19 Genomics UK (COG-UK) Consortium |
| EPI_ISL_461978 | Centre for Enzyme Innovation, University of Portsmouth / Translational Research Laboratory, Portsmouth Hospitals NHS Trust | COVID-19 Genomics UK (COG-UK) Consortium |
| EPI_ISL_461979 | Centre for Enzyme Innovation, University of Portsmouth / Translational Research Laboratory, Portsmouth Hospitals NHS Trust | COVID-19 Genomics UK (COG-UK) Consortium |

| EPI_Accession_ID | Originating Laboratory | Submitting Laboratory |
|---|---|---|
| EPI_ISL_461980 | Centre for Enzyme Innovation, University of Portsmouth / Translational Research Laboratory, Portsmouth Hospitals NHS Trust | COVID-19 Genomics UK (COG-UK) Consortium |
| EPI_ISL_461981 | Centre for Enzyme Innovation, University of Portsmouth / Translational Research Laboratory, Portsmouth Hospitals NHS Trust | COVID-19 Genomics UK (COG-UK) Consortium |
| EPI_ISL_461983 | Centre for Enzyme Innovation, University of Portsmouth / Translational Research Laboratory, Portsmouth Hospitals NHS Trust | COVID-19 Genomics UK (COG-UK) Consortium |
| EPI_ISL_461984 | Centre for Enzyme Innovation, University of Portsmouth / Translational Research Laboratory, Portsmouth Hospitals NHS Trust | COVID-19 Genomics UK (COG-UK) Consortium |
| EPI_ISL_461985 | Centre for Enzyme Innovation, University of Portsmouth / Translational Research Laboratory, Portsmouth Hospitals NHS Trust | COVID-19 Genomics UK (COG-UK) Consortium |
| EPI_ISL_461986 | Centre for Enzyme Innovation, University of Portsmouth / Translational Research Laboratory, Portsmouth Hospitals NHS Trust | COVID-19 Genomics UK (COG-UK) Consortium |
| EPI_ISL_461987 | Centre for Enzyme Innovation, University of Portsmouth / Translational Research Laboratory, Portsmouth Hospitals NHS Trust | COVID-19 Genomics UK (COG-UK) Consortium |
| EPI_ISL_461988 | Centre for Enzyme Innovation, University of Portsmouth / Translational Research Laboratory, Portsmouth Hospitals NHS Trust | COVID-19 Genomics UK (COG-UK) Consortium |
| EPI_ISL_461989 | Centre for Enzyme Innovation, University of Portsmouth / Translational Research Laboratory, Portsmouth Hospitals NHS Trust | COVID-19 Genomics UK (COG-UK) Consortium |
| EPI_ISL_461990 | Centre for Enzyme Innovation, University of Portsmouth / Translational Research Laboratory, Portsmouth Hospitals NHS Trust | COVID-19 Genomics UK (COG-UK) Consortium |
| EPI_ISL_461991 | Centre for Enzyme Innovation, University of Portsmouth / Translational Research Laboratory, Portsmouth Hospitals NHS Trust | COVID-19 Genomics UK (COG-UK) Consortium |

| EPI_Accession_ID | Originating Laboratory | Submitting Laboratory |
|---|---|---|
| EPI_ISL_461992 | Centre for Enzyme Innovation, University of Portsmouth / Translational Research Laboratory, Portsmouth Hospitals NHS Trust | COVID-19 Genomics UK (COG-UK) Consortium |
| EPI_ISL_461993 | Centre for Enzyme Innovation, University of Portsmouth / Translational Research Laboratory, Portsmouth Hospitals NHS Trust | COVID-19 Genomics UK (COG-UK) Consortium |
| EPI_ISL_461994 | Centre for Enzyme Innovation, University of Portsmouth / Translational Research Laboratory, Portsmouth Hospitals NHS Trust | COVID-19 Genomics UK (COG-UK) Consortium |
| EPI_ISL_461995 | Centre for Enzyme Innovation, University of Portsmouth / Translational Research Laboratory, Portsmouth Hospitals NHS Trust | COVID-19 Genomics UK (COG-UK) Consortium |
| EPI_ISL_461996 | Centre for Enzyme Innovation, University of Portsmouth / Translational Research Laboratory, Portsmouth Hospitals NHS Trust | COVID-19 Genomics UK (COG-UK) Consortium |
| EPI_ISL_461997 | Centre for Enzyme Innovation, University of Portsmouth / Translational Research Laboratory, Portsmouth Hospitals NHS Trust | COVID-19 Genomics UK (COG-UK) Consortium |
| EPI_ISL_461999 | Virology Department, Sheffield Teaching Hospitals NHS Foundation Trust/Department of Infection, Immunity and Cardiovascular Disease, The Medical School, University of Sheffield | COVID-19 Genomics UK (COG-UK) Consortium |
| EPI_ISL_462000 | Virology Department, Sheffield Teaching Hospitals NHS Foundation Trust/Department of Infection, Immunity and Cardiovascular Disease, The Medical School, University of Sheffield | COVID-19 Genomics UK (COG-UK) Consortium |
| EPI_ISL_462001 | Virology Department, Sheffield Teaching Hospitals NHS Foundation Trust/Department of Infection, Immunity and Cardiovascular Disease, The Medical School, University of Sheffield | COVID-19 Genomics UK (COG-UK) Consortium |

| EPI_Accession_ID | Originating Laboratory | Submitting Laboratory |
|---|---|---|
| EPI_ISL_462002 | Virology Department, Sheffield Teaching Hospitals NHS Foundation Trust/Department of Infection, Immunity and Cardiovascular Disease, The Medical School, University of Sheffield | COVID-19 Genomics UK (COG-UK) Consortium |
| EPI_ISL_462003 | Virology Department, Sheffield Teaching Hospitals NHS Foundation Trust/Department of Infection, Immunity and Cardiovascular Disease, The Medical School, University of Sheffield | COVID-19 Genomics UK (COG-UK) Consortium |
| EPI_ISL_462004 | Virology Department, Sheffield Teaching Hospitals NHS Foundation Trust/Department of Infection, Immunity and Cardiovascular Disease, The Medical School, University of Sheffield | COVID-19 Genomics UK (COG-UK) Consortium |
| EPI_ISL_462005 | Virology Department, Sheffield Teaching Hospitals NHS Foundation Trust/Department of Infection, Immunity and Cardiovascular Disease, The Medical School, University of Sheffield | COVID-19 Genomics UK (COG-UK) Consortium |
| EPI_ISL_462006 | Virology Department, Sheffield Teaching Hospitals NHS Foundation Trust/Department of Infection, Immunity and Cardiovascular Disease, The Medical School, University of Sheffield | COVID-19 Genomics UK (COG-UK) Consortium |
| EPI_ISL_462007 | Virology Department, Sheffield Teaching Hospitals NHS Foundation Trust/Department of Infection, Immunity and Cardiovascular Disease, The Medical School, University of Sheffield | COVID-19 Genomics UK (COG-UK) Consortium |
| EPI_ISL_462008 | Virology Department, Sheffield Teaching Hospitals NHS Foundation Trust/Department of Infection, Immunity and Cardiovascular Disease, The Medical School, University of Sheffield | COVID-19 Genomics UK (COG-UK) Consortium |
| EPI_ISL_462009 | Virology Department, Sheffield Teaching Hospitals NHS Foundation Trust/Department of Infection, Immunity and Cardiovascular Disease, The Medical School, University of Sheffield | COVID-19 Genomics UK (COG-UK) Consortium |

| EPI_Accession_ID | Originating Laboratory | Submitting Laboratory |
|---|---|---|
| EPI_ISL_462010 | Virology Department, Sheffield Teaching Hospitals NHS Foundation Trust/Department of Infection, Immunity and Cardiovascular Disease, The Medical School, University of Sheffield | COVID-19 Genomics UK (COG-UK) Consortium |
| EPI_ISL_462011 | Virology Department, Sheffield Teaching Hospitals NHS Foundation Trust/Department of Infection, Immunity and Cardiovascular Disease, The Medical School, University of Sheffield | COVID-19 Genomics UK (COG-UK) Consortium |
| EPI_ISL_462012 | Virology Department, Sheffield Teaching Hospitals NHS Foundation Trust/Department of Infection, Immunity and Cardiovascular Disease, The Medical School, University of Sheffield | COVID-19 Genomics UK (COG-UK) Consortium |
| EPI_ISL_462013 | Virology Department, Sheffield Teaching Hospitals NHS Foundation Trust/Department of Infection, Immunity and Cardiovascular Disease, The Medical School, University of Sheffield | COVID-19 Genomics UK (COG-UK) Consortium |
| EPI_ISL_462014 | Virology Department, Sheffield Teaching Hospitals NHS Foundation Trust/Department of Infection, Immunity and Cardiovascular Disease, The Medical School, University of Sheffield | COVID-19 Genomics UK (COG-UK) Consortium |
| EPI_ISL_462015 | Virology Department, Sheffield Teaching Hospitals NHS Foundation Trust/Department of Infection, Immunity and Cardiovascular Disease, The Medical School, University of Sheffield | COVID-19 Genomics UK (COG-UK) Consortium |
| EPI_ISL_462016 | Virology Department, Sheffield Teaching Hospitals NHS Foundation Trust/Department of Infection, Immunity and Cardiovascular Disease, The Medical School, University of Sheffield | COVID-19 Genomics UK (COG-UK) Consortium |
| EPI_ISL_462017 | Virology Department, Sheffield Teaching Hospitals NHS Foundation Trust/Department of Infection, Immunity and Cardiovascular Disease, The Medical School, University of Sheffield | COVID-19 Genomics UK (COG-UK) Consortium |

| EPI_Accession_ID | Originating Laboratory | Submitting Laboratory |
| --- | --- | --- |
| EPI_ISL_462018 | Virology Department, Sheffield Teaching Hospitals NHS Foundation Trust/Department of Infection, Immunity and Cardiovascular Disease, The Medical School, University of Sheffield | COVID-19 Genomics UK (COG-UK) Consortium |
| EPI_ISL_462019 | Virology Department, Sheffield Teaching Hospitals NHS Foundation Trust/Department of Infection, Immunity and Cardiovascular Disease, The Medical School, University of Sheffield | COVID-19 Genomics UK (COG-UK) Consortium |
| EPI_ISL_462020 | Virology Department, Sheffield Teaching Hospitals NHS Foundation Trust/Department of Infection, Immunity and Cardiovascular Disease, The Medical School, University of Sheffield | COVID-19 Genomics UK (COG-UK) Consortium |
| EPI_ISL_462021 | Virology Department, Sheffield Teaching Hospitals NHS Foundation Trust/Department of Infection, Immunity and Cardiovascular Disease, The Medical School, University of Sheffield | COVID-19 Genomics UK (COG-UK) Consortium |
| EPI_ISL_462022 | Virology Department, Sheffield Teaching Hospitals NHS Foundation Trust/Department of Infection, Immunity and Cardiovascular Disease, The Medical School, University of Sheffield | COVID-19 Genomics UK (COG-UK) Consortium |
| EPI_ISL_462023 | Virology Department, Sheffield Teaching Hospitals NHS Foundation Trust/Department of Infection, Immunity and Cardiovascular Disease, The Medical School, University of Sheffield | COVID-19 Genomics UK (COG-UK) Consortium |
| EPI_ISL_462024 | Virology Department, Sheffield Teaching Hospitals NHS Foundation Trust/Department of Infection, Immunity and Cardiovascular Disease, The Medical School, University of Sheffield | COVID-19 Genomics UK (COG-UK) Consortium |
| EPI_ISL_462025 | Virology Department, Sheffield Teaching Hospitals NHS Foundation Trust/Department of Infection, Immunity and Cardiovascular Disease, The Medical School, University of Sheffield | COVID-19 Genomics UK (COG-UK) Consortium |

| EPI_Accession_ID | Originating Laboratory | Submitting Laboratory |
| --- | --- | --- |
| EPI_ISL_462026 | Virology Department, Sheffield Teaching Hospitals NHS Foundation Trust/Department of Infection, Immunity and Cardiovascular Disease, The Medical School, University of Sheffield | COVID-19 Genomics UK (COG-UK) Consortium |
| EPI_ISL_462027 | Virology Department, Sheffield Teaching Hospitals NHS Foundation Trust/Department of Infection, Immunity and Cardiovascular Disease, The Medical School, University of Sheffield | COVID-19 Genomics UK (COG-UK) Consortium |
| EPI_ISL_462028 | Virology Department, Sheffield Teaching Hospitals NHS Foundation Trust/Department of Infection, Immunity and Cardiovascular Disease, The Medical School, University of Sheffield | COVID-19 Genomics UK (COG-UK) Consortium |
| EPI_ISL_462029 | Virology Department, Sheffield Teaching Hospitals NHS Foundation Trust/Department of Infection, Immunity and Cardiovascular Disease, The Medical School, University of Sheffield | COVID-19 Genomics UK (COG-UK) Consortium |
| EPI_ISL_462030 | Virology Department, Sheffield Teaching Hospitals NHS Foundation Trust/Department of Infection, Immunity and Cardiovascular Disease, The Medical School, University of Sheffield | COVID-19 Genomics UK (COG-UK) Consortium |
| EPI_ISL_462031 | Virology Department, Sheffield Teaching Hospitals NHS Foundation Trust/Department of Infection, Immunity and Cardiovascular Disease, The Medical School, University of Sheffield | COVID-19 Genomics UK (COG-UK) Consortium |
| EPI_ISL_462032 | Virology Department, Sheffield Teaching Hospitals NHS Foundation Trust/Department of Infection, Immunity and Cardiovascular Disease, The Medical School, University of Sheffield | COVID-19 Genomics UK (COG-UK) Consortium |
| EPI_ISL_462033 | Virology Department, Sheffield Teaching Hospitals NHS Foundation Trust/Department of Infection, Immunity and Cardiovascular Disease, The Medical School, University of Sheffield | COVID-19 Genomics UK (COG-UK) Consortium |

| EPI_Accession_ID | Originating Laboratory | Submitting Laboratory |
| --- | --- | --- |
| EPI_ISL_462034 | Virology Department, Sheffield Teaching Hospitals NHS Foundation Trust/Department of Infection, Immunity and Cardiovascular Disease, The Medical School, University of Sheffield | COVID-19 Genomics UK (COG-UK) Consortium |
| EPI_ISL_462035 | Virology Department, Sheffield Teaching Hospitals NHS Foundation Trust/Department of Infection, Immunity and Cardiovascular Disease, The Medical School, University of Sheffield | COVID-19 Genomics UK (COG-UK) Consortium |
| EPI_ISL_462036 | Virology Department, Sheffield Teaching Hospitals NHS Foundation Trust/Department of Infection, Immunity and Cardiovascular Disease, The Medical School, University of Sheffield | COVID-19 Genomics UK (COG-UK) Consortium |
| EPI_ISL_462037 | Virology Department, Sheffield Teaching Hospitals NHS Foundation Trust/Department of Infection, Immunity and Cardiovascular Disease, The Medical School, University of Sheffield | COVID-19 Genomics UK (COG-UK) Consortium |
| EPI_ISL_462038 | Virology Department, Sheffield Teaching Hospitals NHS Foundation Trust/Department of Infection, Immunity and Cardiovascular Disease, The Medical School, University of Sheffield | COVID-19 Genomics UK (COG-UK) Consortium |
| EPI_ISL_462039 | Virology Department, Sheffield Teaching Hospitals NHS Foundation Trust/Department of Infection, Immunity and Cardiovascular Disease, The Medical School, University of Sheffield | COVID-19 Genomics UK (COG-UK) Consortium |
| EPI_ISL_462040 | Virology Department, Sheffield Teaching Hospitals NHS Foundation Trust/Department of Infection, Immunity and Cardiovascular Disease, The Medical School, University of Sheffield | COVID-19 Genomics UK (COG-UK) Consortium |
| EPI_ISL_462041 | Virology Department, Sheffield Teaching Hospitals NHS Foundation Trust/Department of Infection, Immunity and Cardiovascular Disease, The Medical School, University of Sheffield | COVID-19 Genomics UK (COG-UK) Consortium |

| EPI_Accession_ID | Originating Laboratory | Submitting Laboratory |
| --- | --- | --- |
| EPI_ISL_462042 | Virology Department, Sheffield Teaching Hospitals NHS Foundation Trust/Department of Infection, Immunity and Cardiovascular Disease, The Medical School, University of Sheffield | COVID-19 Genomics UK (COG-UK) Consortium |
| EPI_ISL_462043 | Virology Department, Sheffield Teaching Hospitals NHS Foundation Trust/Department of Infection, Immunity and Cardiovascular Disease, The Medical School, University of Sheffield | COVID-19 Genomics UK (COG-UK) Consortium |
| EPI_ISL_462044 | Virology Department, Sheffield Teaching Hospitals NHS Foundation Trust/Department of Infection, Immunity and Cardiovascular Disease, The Medical School, University of Sheffield | COVID-19 Genomics UK (COG-UK) Consortium |
| EPI_ISL_462045 | Virology Department, Sheffield Teaching Hospitals NHS Foundation Trust/Department of Infection, Immunity and Cardiovascular Disease, The Medical School, University of Sheffield | COVID-19 Genomics UK (COG-UK) Consortium |
| EPI_ISL_462046 | Virology Department, Sheffield Teaching Hospitals NHS Foundation Trust/Department of Infection, Immunity and Cardiovascular Disease, The Medical School, University of Sheffield | COVID-19 Genomics UK (COG-UK) Consortium |
| EPI_ISL_462047 | Virology Department, Sheffield Teaching Hospitals NHS Foundation Trust/Department of Infection, Immunity and Cardiovascular Disease, The Medical School, University of Sheffield | COVID-19 Genomics UK (COG-UK) Consortium |
| EPI_ISL_462048 | Virology Department, Sheffield Teaching Hospitals NHS Foundation Trust/Department of Infection, Immunity and Cardiovascular Disease, The Medical School, University of Sheffield | COVID-19 Genomics UK (COG-UK) Consortium |
| EPI_ISL_462049 | Virology Department, Sheffield Teaching Hospitals NHS Foundation Trust/Department of Infection, Immunity and Cardiovascular Disease, The Medical School, University of Sheffield | COVID-19 Genomics UK (COG-UK) Consortium |

| EPI_Accession_ID | Originating Laboratory | Submitting Laboratory |
|---|---|---|
| EPI_ISL_462050 | Virology Department, Sheffield Teaching Hospitals NHS Foundation Trust/Department of Infection, Immunity and Cardiovascular Disease, The Medical School, University of Sheffield | COVID-19 Genomics UK (COG-UK) Consortium |
| EPI_ISL_462051 | Virology Department, Sheffield Teaching Hospitals NHS Foundation Trust/Department of Infection, Immunity and Cardiovascular Disease, The Medical School, University of Sheffield | COVID-19 Genomics UK (COG-UK) Consortium |
| EPI_ISL_462052 | Virology Department, Sheffield Teaching Hospitals NHS Foundation Trust/Department of Infection, Immunity and Cardiovascular Disease, The Medical School, University of Sheffield | COVID-19 Genomics UK (COG-UK) Consortium |
| EPI_ISL_462053 | Virology Department, Sheffield Teaching Hospitals NHS Foundation Trust/Department of Infection, Immunity and Cardiovascular Disease, The Medical School, University of Sheffield | COVID-19 Genomics UK (COG-UK) Consortium |
| EPI_ISL_462054 | Virology Department, Sheffield Teaching Hospitals NHS Foundation Trust/Department of Infection, Immunity and Cardiovascular Disease, The Medical School, University of Sheffield | COVID-19 Genomics UK (COG-UK) Consortium |
| EPI_ISL_462055 | Virology Department, Sheffield Teaching Hospitals NHS Foundation Trust/Department of Infection, Immunity and Cardiovascular Disease, The Medical School, University of Sheffield | COVID-19 Genomics UK (COG-UK) Consortium |
| EPI_ISL_462056 | Virology Department, Sheffield Teaching Hospitals NHS Foundation Trust/Department of Infection, Immunity and Cardiovascular Disease, The Medical School, University of Sheffield | COVID-19 Genomics UK (COG-UK) Consortium |
| EPI_ISL_462057 | Virology Department, Sheffield Teaching Hospitals NHS Foundation Trust/Department of Infection, Immunity and Cardiovascular Disease, The Medical School, University of Sheffield | COVID-19 Genomics UK (COG-UK) Consortium |

| EPI_Accession_ID | Originating Laboratory | Submitting Laboratory |
|---|---|---|
| EPI_ISL_462058 | Virology Department, Sheffield Teaching Hospitals NHS Foundation Trust/Department of Infection, Immunity and Cardiovascular Disease, The Medical School, University of Sheffield | COVID-19 Genomics UK (COG-UK) Consortium |
| EPI_ISL_462059 | Virology Department, Sheffield Teaching Hospitals NHS Foundation Trust/Department of Infection, Immunity and Cardiovascular Disease, The Medical School, University of Sheffield | COVID-19 Genomics UK (COG-UK) Consortium |
| EPI_ISL_462060 | Virology Department, Sheffield Teaching Hospitals NHS Foundation Trust/Department of Infection, Immunity and Cardiovascular Disease, The Medical School, University of Sheffield | COVID-19 Genomics UK (COG-UK) Consortium |
| EPI_ISL_462061 | Virology Department, Sheffield Teaching Hospitals NHS Foundation Trust/Department of Infection, Immunity and Cardiovascular Disease, The Medical School, University of Sheffield | COVID-19 Genomics UK (COG-UK) Consortium |
| EPI_ISL_462062 | Virology Department, Sheffield Teaching Hospitals NHS Foundation Trust/Department of Infection, Immunity and Cardiovascular Disease, The Medical School, University of Sheffield | COVID-19 Genomics UK (COG-UK) Consortium |
| EPI_ISL_462063 | Virology Department, Sheffield Teaching Hospitals NHS Foundation Trust/Department of Infection, Immunity and Cardiovascular Disease, The Medical School, University of Sheffield | COVID-19 Genomics UK (COG-UK) Consortium |
| EPI_ISL_462064 | Virology Department, Sheffield Teaching Hospitals NHS Foundation Trust/Department of Infection, Immunity and Cardiovascular Disease, The Medical School, University of Sheffield | COVID-19 Genomics UK (COG-UK) Consortium |
| EPI_ISL_462065 | Virology Department, Sheffield Teaching Hospitals NHS Foundation Trust/Department of Infection, Immunity and Cardiovascular Disease, The Medical School, University of Sheffield | COVID-19 Genomics UK (COG-UK) Consortium |

| EPI_Accession_ID | Originating Laboratory | Submitting Laboratory |
|---|---|---|
| EPI_ISL_462066 | Virology Department, Sheffield Teaching Hospitals NHS Foundation Trust/Department of Infection, Immunity and Cardiovascular Disease, The Medical School, University of Sheffield | COVID-19 Genomics UK (COG-UK) Consortium |
| EPI_ISL_462067 | Virology Department, Sheffield Teaching Hospitals NHS Foundation Trust/Department of Infection, Immunity and Cardiovascular Disease, The Medical School, University of Sheffield | COVID-19 Genomics UK (COG-UK) Consortium |
| EPI_ISL_462068 | Virology Department, Sheffield Teaching Hospitals NHS Foundation Trust/Department of Infection, Immunity and Cardiovascular Disease, The Medical School, University of Sheffield | COVID-19 Genomics UK (COG-UK) Consortium |
| EPI_ISL_462069 | Virology Department, Sheffield Teaching Hospitals NHS Foundation Trust/Department of Infection, Immunity and Cardiovascular Disease, The Medical School, University of Sheffield | COVID-19 Genomics UK (COG-UK) Consortium |
| EPI_ISL_462070 | Virology Department, Sheffield Teaching Hospitals NHS Foundation Trust/Department of Infection, Immunity and Cardiovascular Disease, The Medical School, University of Sheffield | COVID-19 Genomics UK (COG-UK) Consortium |
| EPI_ISL_462071 | Virology Department, Sheffield Teaching Hospitals NHS Foundation Trust/Department of Infection, Immunity and Cardiovascular Disease, The Medical School, University of Sheffield | COVID-19 Genomics UK (COG-UK) Consortium |
| EPI_ISL_462072 | Virology Department, Sheffield Teaching Hospitals NHS Foundation Trust/Department of Infection, Immunity and Cardiovascular Disease, The Medical School, University of Sheffield | COVID-19 Genomics UK (COG-UK) Consortium |
| EPI_ISL_462073 | Virology Department, Sheffield Teaching Hospitals NHS Foundation Trust/Department of Infection, Immunity and Cardiovascular Disease, The Medical School, University of Sheffield | COVID-19 Genomics UK (COG-UK) Consortium |

| EPI_Accession_ID | Originating Laboratory | Submitting Laboratory |
|---|---|---|
| EPI_ISL_462074 | Virology Department, Sheffield Teaching Hospitals NHS Foundation Trust/Department of Infection, Immunity and Cardiovascular Disease, The Medical School, University of Sheffield | COVID-19 Genomics UK (COG-UK) Consortium |
| EPI_ISL_462075 | Virology Department, Sheffield Teaching Hospitals NHS Foundation Trust/Department of Infection, Immunity and Cardiovascular Disease, The Medical School, University of Sheffield | COVID-19 Genomics UK (COG-UK) Consortium |
| EPI_ISL_462076 | Virology Department, Sheffield Teaching Hospitals NHS Foundation Trust/Department of Infection, Immunity and Cardiovascular Disease, The Medical School, University of Sheffield | COVID-19 Genomics UK (COG-UK) Consortium |
| EPI_ISL_462077 | Virology Department, Sheffield Teaching Hospitals NHS Foundation Trust/Department of Infection, Immunity and Cardiovascular Disease, The Medical School, University of Sheffield | COVID-19 Genomics UK (COG-UK) Consortium |
| EPI_ISL_462078 | Virology Department, Sheffield Teaching Hospitals NHS Foundation Trust/Department of Infection, Immunity and Cardiovascular Disease, The Medical School, University of Sheffield | COVID-19 Genomics UK (COG-UK) Consortium |
| EPI_ISL_462079 | Virology Department, Sheffield Teaching Hospitals NHS Foundation Trust/Department of Infection, Immunity and Cardiovascular Disease, The Medical School, University of Sheffield | COVID-19 Genomics UK (COG-UK) Consortium |
| EPI_ISL_462080 | Virology Department, Sheffield Teaching Hospitals NHS Foundation Trust/Department of Infection, Immunity and Cardiovascular Disease, The Medical School, University of Sheffield | COVID-19 Genomics UK (COG-UK) Consortium |
| EPI_ISL_462081 | Virology Department, Sheffield Teaching Hospitals NHS Foundation Trust/Department of Infection, Immunity and Cardiovascular Disease, The Medical School, University of Sheffield | COVID-19 Genomics UK (COG-UK) Consortium |

| EPI_Accession_ID | Originating Laboratory | Submitting Laboratory |
|---|---|---|
| EPI_ISL_462082 | Virology Department, Sheffield Teaching Hospitals NHS Foundation Trust/Department of Infection, Immunity and Cardiovascular Disease, The Medical School, University of Sheffield | COVID-19 Genomics UK (COG-UK) Consortium |
| EPI_ISL_462083 | Virology Department, Sheffield Teaching Hospitals NHS Foundation Trust/Department of Infection, Immunity and Cardiovascular Disease, The Medical School, University of Sheffield | COVID-19 Genomics UK (COG-UK) Consortium |
| EPI_ISL_462084 | Virology Department, Sheffield Teaching Hospitals NHS Foundation Trust/Department of Infection, Immunity and Cardiovascular Disease, The Medical School, University of Sheffield | COVID-19 Genomics UK (COG-UK) Consortium |
| EPI_ISL_462085 | Singapore General Hospital | Department of Microbiology |
| EPI_ISL_462086 | Singapore General Hospital | Department of Microbiology |
| EPI_ISL_462087 | Singapore General Hospital | Department of Microbiology |
| EPI_ISL_462088 | Singapore General Hospital | Department of Microbiology |
| EPI_ISL_462089 | Singapore General Hospital | Department of Microbiology |
| EPI_ISL_462090 | National Institute of Laboratory Medicine and Referral Center | Genomic Research Lab, BCSIR |
| EPI_ISL_462091 | National Institute of Laboratory Medicine and Referral Center | Genomic Research Lab, BCSIR |
| EPI_ISL_462092 | National Institute of Laboratory Medicine and Referral Center | Genomic Research Lab, BCSIR |
| EPI_ISL_462093 | National Institute of Laboratory Medicine and Referral Center | Genomic Research Lab, BCSIR |
| EPI_ISL_462094 | National Institute of Laboratory Medicine and Referral Center | Genomic Research Lab, BCSIR |
| EPI_ISL_462095 | National Institute of Laboratory Medicine and Referral Center | Genomic Research Lab, BCSIR |
| EPI_ISL_462096 | National Institute of Laboratory Medicine and Referral Center | Genomic Research Lab, BCSIR |
| EPI_ISL_462097 | National Institute of Laboratory Medicine and Referral Center | Genomic Research Lab, BCSIR |

| EPI_Accession_ID | Originating Laboratory | Submitting Laboratory |
|---|---|---|
| EPI_ISL_462098 | National Institute of Laboratory Medicine and Referral Center | Genomic Research Lab, BCSIR |
| EPI_ISL_462150 | Molecular diagnostic laboratory of Federal Budget Institution of Science "Central Research Institute of Epidemiology" of The Federal Service on Customers' Rights Protection and Human Well-being Surveillance | Group of Genomics and Postgenomic Technologies of Central Research Institute of Epidemilology |
| EPI_ISL_462151 | KU Leuven, Rega Institute, Clinical and Epidemiological Virology | KU Leuven, Rega Institute, Clinical and Epidemiological Virology |
| EPI_ISL_462152 | KU Leuven, Rega Institute, Clinical and Epidemiological Virology | KU Leuven, Rega Institute, Clinical and Epidemiological Virology |
| EPI_ISL_462153 | KU Leuven, Rega Institute, Clinical and Epidemiological Virology | KU Leuven, Rega Institute, Clinical and Epidemiological Virology |
| EPI_ISL_462154 | KU Leuven, Rega Institute, Clinical and Epidemiological Virology | KU Leuven, Rega Institute, Clinical and Epidemiological Virology |
| EPI_ISL_462155 | KU Leuven, Rega Institute, Clinical and Epidemiological Virology | KU Leuven, Rega Institute, Clinical and Epidemiological Virology |
| EPI_ISL_462156 | KU Leuven, Rega Institute, Clinical and Epidemiological Virology | KU Leuven, Rega Institute, Clinical and Epidemiological Virology |
| EPI_ISL_462157 | KU Leuven, Rega Institute, Clinical and Epidemiological Virology | KU Leuven, Rega Institute, Clinical and Epidemiological Virology |
| EPI_ISL_462158 | KU Leuven, Rega Institute, Clinical and Epidemiological Virology | KU Leuven, Rega Institute, Clinical and Epidemiological Virology |
| EPI_ISL_462159 | KU Leuven, Rega Institute, Clinical and Epidemiological Virology | KU Leuven, Rega Institute, Clinical and Epidemiological Virology |
| EPI_ISL_462160 | KU Leuven, Rega Institute, Clinical and Epidemiological Virology | KU Leuven, Rega Institute, Clinical and Epidemiological Virology |
| EPI_ISL_462161 | KU Leuven, Rega Institute, Clinical and Epidemiological Virology | KU Leuven, Rega Institute, Clinical and Epidemiological Virology |
| EPI_ISL_462162 | KU Leuven, Rega Institute, Clinical and Epidemiological Virology | KU Leuven, Rega Institute, Clinical and Epidemiological Virology |
| EPI_ISL_462163 | KU Leuven, Rega Institute, Clinical and Epidemiological Virology | KU Leuven, Rega Institute, Clinical and Epidemiological Virology |

| EPI_Accession_ID | Originating Laboratory | Submitting Laboratory |
|---|---|---|
| EPI_ISL_462164 | KU Leuven, Rega Institute, Clinical and Epidemiological Virology | KU Leuven, Rega Institute, Clinical and Epidemiological Virology |
| EPI_ISL_462165 | KU Leuven, Rega Institute, Clinical and Epidemiological Virology | KU Leuven, Rega Institute, Clinical and Epidemiological Virology |
| EPI_ISL_462166 | KU Leuven, Rega Institute, Clinical and Epidemiological Virology | KU Leuven, Rega Institute, Clinical and Epidemiological Virology |
| EPI_ISL_462167 | KU Leuven, Rega Institute, Clinical and Epidemiological Virology | KU Leuven, Rega Institute, Clinical and Epidemiological Virology |
| EPI_ISL_462168 | KU Leuven, Rega Institute, Clinical and Epidemiological Virology | KU Leuven, Rega Institute, Clinical and Epidemiological Virology |
| EPI_ISL_462169 | KU Leuven, Rega Institute, Clinical and Epidemiological Virology | KU Leuven, Rega Institute, Clinical and Epidemiological Virology |
| EPI_ISL_462170 | KU Leuven, Rega Institute, Clinical and Epidemiological Virology | KU Leuven, Rega Institute, Clinical and Epidemiological Virology |
| EPI_ISL_462171 | KU Leuven, Rega Institute, Clinical and Epidemiological Virology | KU Leuven, Rega Institute, Clinical and Epidemiological Virology |
| EPI_ISL_462172 | KU Leuven, Rega Institute, Clinical and Epidemiological Virology | KU Leuven, Rega Institute, Clinical and Epidemiological Virology |
| EPI_ISL_462173 | KU Leuven, Rega Institute, Clinical and Epidemiological Virology | KU Leuven, Rega Institute, Clinical and Epidemiological Virology |
| EPI_ISL_462174 | KU Leuven, Rega Institute, Clinical and Epidemiological Virology | KU Leuven, Rega Institute, Clinical and Epidemiological Virology |
| EPI_ISL_462175 | KU Leuven, Rega Institute, Clinical and Epidemiological Virology | KU Leuven, Rega Institute, Clinical and Epidemiological Virology |
| EPI_ISL_462176 | KU Leuven, Rega Institute, Clinical and Epidemiological Virology | KU Leuven, Rega Institute, Clinical and Epidemiological Virology |
| EPI_ISL_462177 | KU Leuven, Rega Institute, Clinical and Epidemiological Virology | KU Leuven, Rega Institute, Clinical and Epidemiological Virology |
| EPI_ISL_462178 | KU Leuven, Rega Institute, Clinical and Epidemiological Virology | KU Leuven, Rega Institute, Clinical and Epidemiological Virology |
| EPI_ISL_462179 | KU Leuven, Rega Institute, Clinical and Epidemiological Virology | KU Leuven, Rega Institute, Clinical and Epidemiological Virology |

| EPI_Accession_ID | Originating Laboratory | Submitting Laboratory |
|---|---|---|
| EPI_ISL_462180 | KU Leuven, Rega Institute, Clinical and Epidemiological Virology | KU Leuven, Rega Institute, Clinical and Epidemiological Virology |
| EPI_ISL_462181 | KU Leuven, Rega Institute, Clinical and Epidemiological Virology | KU Leuven, Rega Institute, Clinical and Epidemiological Virology |
| EPI_ISL_462182 | KU Leuven, Rega Institute, Clinical and Epidemiological Virology | KU Leuven, Rega Institute, Clinical and Epidemiological Virology |
| EPI_ISL_462183 | KU Leuven, Rega Institute, Clinical and Epidemiological Virology | KU Leuven, Rega Institute, Clinical and Epidemiological Virology |
| EPI_ISL_462184 | KU Leuven, Rega Institute, Clinical and Epidemiological Virology | KU Leuven, Rega Institute, Clinical and Epidemiological Virology |
| EPI_ISL_462185 | KU Leuven, Rega Institute, Clinical and Epidemiological Virology | KU Leuven, Rega Institute, Clinical and Epidemiological Virology |
| EPI_ISL_462186 | KU Leuven, Rega Institute, Clinical and Epidemiological Virology | KU Leuven, Rega Institute, Clinical and Epidemiological Virology |
| EPI_ISL_462187 | KU Leuven, Rega Institute, Clinical and Epidemiological Virology | KU Leuven, Rega Institute, Clinical and Epidemiological Virology |
| EPI_ISL_462188 | KU Leuven, Rega Institute, Clinical and Epidemiological Virology | KU Leuven, Rega Institute, Clinical and Epidemiological Virology |
| EPI_ISL_462189 | KU Leuven, Rega Institute, Clinical and Epidemiological Virology | KU Leuven, Rega Institute, Clinical and Epidemiological Virology |
| EPI_ISL_462190 | KU Leuven, Rega Institute, Clinical and Epidemiological Virology | KU Leuven, Rega Institute, Clinical and Epidemiological Virology |
| EPI_ISL_462191 | KU Leuven, Rega Institute, Clinical and Epidemiological Virology | KU Leuven, Rega Institute, Clinical and Epidemiological Virology |
| EPI_ISL_462192 | KU Leuven, Rega Institute, Clinical and Epidemiological Virology | KU Leuven, Rega Institute, Clinical and Epidemiological Virology |
| EPI_ISL_462193 | KU Leuven, Rega Institute, Clinical and Epidemiological Virology | KU Leuven, Rega Institute, Clinical and Epidemiological Virology |
| EPI_ISL_462194 | KU Leuven, Rega Institute, Clinical and Epidemiological Virology | KU Leuven, Rega Institute, Clinical and Epidemiological Virology |
| EPI_ISL_462195 | KU Leuven, Rega Institute, Clinical and Epidemiological Virology | KU Leuven, Rega Institute, Clinical and Epidemiological Virology |

| EPI_Accession_ID | Originating Laboratory | Submitting Laboratory |
|---|---|---|
| EPI_ISL_462196 | KU Leuven, Rega Institute, Clinical and Epidemiological Virology | KU Leuven, Rega Institute, Clinical and Epidemiological Virology |
| EPI_ISL_462197 | KU Leuven, Rega Institute, Clinical and Epidemiological Virology | KU Leuven, Rega Institute, Clinical and Epidemiological Virology |
| EPI_ISL_462198 | KU Leuven, Rega Institute, Clinical and Epidemiological Virology | KU Leuven, Rega Institute, Clinical and Epidemiological Virology |
| EPI_ISL_462199 | KU Leuven, Rega Institute, Clinical and Epidemiological Virology | KU Leuven, Rega Institute, Clinical and Epidemiological Virology |
| EPI_ISL_462200 | KU Leuven, Rega Institute, Clinical and Epidemiological Virology | KU Leuven, Rega Institute, Clinical and Epidemiological Virology |
| EPI_ISL_462201 | KU Leuven, Rega Institute, Clinical and Epidemiological Virology | KU Leuven, Rega Institute, Clinical and Epidemiological Virology |
| EPI_ISL_462202 | KU Leuven, Rega Institute, Clinical and Epidemiological Virology | KU Leuven, Rega Institute, Clinical and Epidemiological Virology |
| EPI_ISL_462203 | KU Leuven, Rega Institute, Clinical and Epidemiological Virology | KU Leuven, Rega Institute, Clinical and Epidemiological Virology |
| EPI_ISL_462204 | KU Leuven, Rega Institute, Clinical and Epidemiological Virology | KU Leuven, Rega Institute, Clinical and Epidemiological Virology |
| EPI_ISL_462205 | KU Leuven, Rega Institute, Clinical and Epidemiological Virology | KU Leuven, Rega Institute, Clinical and Epidemiological Virology |
| EPI_ISL_462206 | KU Leuven, Rega Institute, Clinical and Epidemiological Virology | KU Leuven, Rega Institute, Clinical and Epidemiological Virology |
| EPI_ISL_462207 | KU Leuven, Rega Institute, Clinical and Epidemiological Virology | KU Leuven, Rega Institute, Clinical and Epidemiological Virology |
| EPI_ISL_462208 | KU Leuven, Rega Institute, Clinical and Epidemiological Virology | KU Leuven, Rega Institute, Clinical and Epidemiological Virology |
| EPI_ISL_462209 | KU Leuven, Rega Institute, Clinical and Epidemiological Virology | KU Leuven, Rega Institute, Clinical and Epidemiological Virology |
| EPI_ISL_462210 | KU Leuven, Rega Institute, Clinical and Epidemiological Virology | KU Leuven, Rega Institute, Clinical and Epidemiological Virology |
| EPI_ISL_462211 | KU Leuven, Rega Institute, Clinical and Epidemiological Virology | KU Leuven, Rega Institute, Clinical and Epidemiological Virology |

| EPI_Accession_ID | Originating Laboratory | Submitting Laboratory |
|---|---|---|
| EPI_ISL_462212 | KU Leuven, Rega Institute, Clinical and Epidemiological Virology | KU Leuven, Rega Institute, Clinical and Epidemiological Virology |
| EPI_ISL_462213 | KU Leuven, Rega Institute, Clinical and Epidemiological Virology | KU Leuven, Rega Institute, Clinical and Epidemiological Virology |
| EPI_ISL_462214 | KU Leuven, Rega Institute, Clinical and Epidemiological Virology | KU Leuven, Rega Institute, Clinical and Epidemiological Virology |
| EPI_ISL_462215 | KU Leuven, Rega Institute, Clinical and Epidemiological Virology | KU Leuven, Rega Institute, Clinical and Epidemiological Virology |
| EPI_ISL_462216 | KU Leuven, Rega Institute, Clinical and Epidemiological Virology | KU Leuven, Rega Institute, Clinical and Epidemiological Virology |
| EPI_ISL_462217 | KU Leuven, Rega Institute, Clinical and Epidemiological Virology | KU Leuven, Rega Institute, Clinical and Epidemiological Virology |
| EPI_ISL_462218 | KU Leuven, Rega Institute, Clinical and Epidemiological Virology | KU Leuven, Rega Institute, Clinical and Epidemiological Virology |
| EPI_ISL_462219 | KU Leuven, Rega Institute, Clinical and Epidemiological Virology | KU Leuven, Rega Institute, Clinical and Epidemiological Virology |
| EPI_ISL_462220 | KU Leuven, Rega Institute, Clinical and Epidemiological Virology | KU Leuven, Rega Institute, Clinical and Epidemiological Virology |
| EPI_ISL_462221 | KU Leuven, Rega Institute, Clinical and Epidemiological Virology | KU Leuven, Rega Institute, Clinical and Epidemiological Virology |
| EPI_ISL_462222 | KU Leuven, Rega Institute, Clinical and Epidemiological Virology | KU Leuven, Rega Institute, Clinical and Epidemiological Virology |
| EPI_ISL_462223 | KU Leuven, Rega Institute, Clinical and Epidemiological Virology | KU Leuven, Rega Institute, Clinical and Epidemiological Virology |
| EPI_ISL_462224 | KU Leuven, Rega Institute, Clinical and Epidemiological Virology | KU Leuven, Rega Institute, Clinical and Epidemiological Virology |
| EPI_ISL_462225 | KU Leuven, Rega Institute, Clinical and Epidemiological Virology | KU Leuven, Rega Institute, Clinical and Epidemiological Virology |
| EPI_ISL_462226 | KU Leuven, Rega Institute, Clinical and Epidemiological Virology | KU Leuven, Rega Institute, Clinical and Epidemiological Virology |
| EPI_ISL_462227 | KU Leuven, Rega Institute, Clinical and Epidemiological Virology | KU Leuven, Rega Institute, Clinical and Epidemiological Virology |

| EPI_Accession_ID | Originating Laboratory | Submitting Laboratory |
|---|---|---|
| EPI_ISL_462228 | KU Leuven, Rega Institute, Clinical and Epidemiological Virology | KU Leuven, Rega Institute, Clinical and Epidemiological Virology |
| EPI_ISL_462229 | KU Leuven, Rega Institute, Clinical and Epidemiological Virology | KU Leuven, Rega Institute, Clinical and Epidemiological Virology |
| EPI_ISL_462230 | KU Leuven, Rega Institute, Clinical and Epidemiological Virology | KU Leuven, Rega Institute, Clinical and Epidemiological Virology |
| EPI_ISL_462231 | KU Leuven, Rega Institute, Clinical and Epidemiological Virology | KU Leuven, Rega Institute, Clinical and Epidemiological Virology |
| EPI_ISL_462232 | KU Leuven, Rega Institute, Clinical and Epidemiological Virology | KU Leuven, Rega Institute, Clinical and Epidemiological Virology |
| EPI_ISL_462233 | KU Leuven, Rega Institute, Clinical and Epidemiological Virology | KU Leuven, Rega Institute, Clinical and Epidemiological Virology |
| EPI_ISL_462234 | KU Leuven, Rega Institute, Clinical and Epidemiological Virology | KU Leuven, Rega Institute, Clinical and Epidemiological Virology |
| EPI_ISL_462235 | KU Leuven, Rega Institute, Clinical and Epidemiological Virology | KU Leuven, Rega Institute, Clinical and Epidemiological Virology |
| EPI_ISL_462236 | KU Leuven, Rega Institute, Clinical and Epidemiological Virology | KU Leuven, Rega Institute, Clinical and Epidemiological Virology |
| EPI_ISL_462237 | KU Leuven, Rega Institute, Clinical and Epidemiological Virology | KU Leuven, Rega Institute, Clinical and Epidemiological Virology |
| EPI_ISL_462238 | KU Leuven, Rega Institute, Clinical and Epidemiological Virology | KU Leuven, Rega Institute, Clinical and Epidemiological Virology |
| EPI_ISL_462239 | KU Leuven, Rega Institute, Clinical and Epidemiological Virology | KU Leuven, Rega Institute, Clinical and Epidemiological Virology |
| EPI_ISL_462240 | KU Leuven, Rega Institute, Clinical and Epidemiological Virology | KU Leuven, Rega Institute, Clinical and Epidemiological Virology |
| EPI_ISL_462241 | KU Leuven, Rega Institute, Clinical and Epidemiological Virology | KU Leuven, Rega Institute, Clinical and Epidemiological Virology |
| EPI_ISL_462242 | KU Leuven, Rega Institute, Clinical and Epidemiological Virology | KU Leuven, Rega Institute, Clinical and Epidemiological Virology |
| EPI_ISL_462243 | KU Leuven, Rega Institute, Clinical and Epidemiological Virology | KU Leuven, Rega Institute, Clinical and Epidemiological Virology |

| EPI_Accession_ID | Originating Laboratory | Submitting Laboratory |
|---|---|---|
| EPI_ISL_462244 | KU Leuven, Rega Institute, Clinical and Epidemiological Virology | KU Leuven, Rega Institute, Clinical and Epidemiological Virology |
| EPI_ISL_462245 | KU Leuven, Rega Institute, Clinical and Epidemiological Virology | KU Leuven, Rega Institute, Clinical and Epidemiological Virology |
| EPI_ISL_462246 | KU Leuven, Rega Institute, Clinical and Epidemiological Virology | KU Leuven, Rega Institute, Clinical and Epidemiological Virology |
| EPI_ISL_462247 | KU Leuven, Rega Institute, Clinical and Epidemiological Virology | KU Leuven, Rega Institute, Clinical and Epidemiological Virology |
| EPI_ISL_462248 | KU Leuven, Rega Institute, Clinical and Epidemiological Virology | KU Leuven, Rega Institute, Clinical and Epidemiological Virology |
| EPI_ISL_462249 | KU Leuven, Rega Institute, Clinical and Epidemiological Virology | KU Leuven, Rega Institute, Clinical and Epidemiological Virology |
| EPI_ISL_462250 | KU Leuven, Rega Institute, Clinical and Epidemiological Virology | KU Leuven, Rega Institute, Clinical and Epidemiological Virology |
| EPI_ISL_462251 | KU Leuven, Rega Institute, Clinical and Epidemiological Virology | KU Leuven, Rega Institute, Clinical and Epidemiological Virology |
| EPI_ISL_462252 | KU Leuven, Rega Institute, Clinical and Epidemiological Virology | KU Leuven, Rega Institute, Clinical and Epidemiological Virology |
| EPI_ISL_462253 | KU Leuven, Rega Institute, Clinical and Epidemiological Virology | KU Leuven, Rega Institute, Clinical and Epidemiological Virology |
| EPI_ISL_462254 | KU Leuven, Rega Institute, Clinical and Epidemiological Virology | KU Leuven, Rega Institute, Clinical and Epidemiological Virology |
| EPI_ISL_462255 | KU Leuven, Rega Institute, Clinical and Epidemiological Virology | KU Leuven, Rega Institute, Clinical and Epidemiological Virology |
| EPI_ISL_462256 | KU Leuven, Rega Institute, Clinical and Epidemiological Virology | KU Leuven, Rega Institute, Clinical and Epidemiological Virology |
| EPI_ISL_462257 | KU Leuven, Rega Institute, Clinical and Epidemiological Virology | KU Leuven, Rega Institute, Clinical and Epidemiological Virology |
| EPI_ISL_462258 | KU Leuven, Rega Institute, Clinical and Epidemiological Virology | KU Leuven, Rega Institute, Clinical and Epidemiological Virology |
| EPI_ISL_462259 | KU Leuven, Rega Institute, Clinical and Epidemiological Virology | KU Leuven, Rega Institute, Clinical and Epidemiological Virology |

| EPI_Accession_ID | Originating Laboratory | Submitting Laboratory |
|---|---|---|
| EPI_ISL_462260 | KU Leuven, Rega Institute, Clinical and Epidemiological Virology | KU Leuven, Rega Institute, Clinical and Epidemiological Virology |
| EPI_ISL_462261 | KU Leuven, Rega Institute, Clinical and Epidemiological Virology | KU Leuven, Rega Institute, Clinical and Epidemiological Virology |
| EPI_ISL_462262 | KU Leuven, Rega Institute, Clinical and Epidemiological Virology | KU Leuven, Rega Institute, Clinical and Epidemiological Virology |
| EPI_ISL_462263 | KU Leuven, Rega Institute, Clinical and Epidemiological Virology | KU Leuven, Rega Institute, Clinical and Epidemiological Virology |
| EPI_ISL_462264 | KU Leuven, Rega Institute, Clinical and Epidemiological Virology | KU Leuven, Rega Institute, Clinical and Epidemiological Virology |
| EPI_ISL_462265 | KU Leuven, Rega Institute, Clinical and Epidemiological Virology | KU Leuven, Rega Institute, Clinical and Epidemiological Virology |
| EPI_ISL_462266 | KU Leuven, Rega Institute, Clinical and Epidemiological Virology | KU Leuven, Rega Institute, Clinical and Epidemiological Virology |
| EPI_ISL_462267 | KU Leuven, Rega Institute, Clinical and Epidemiological Virology | KU Leuven, Rega Institute, Clinical and Epidemiological Virology |
| EPI_ISL_462268 | KU Leuven, Rega Institute, Clinical and Epidemiological Virology | KU Leuven, Rega Institute, Clinical and Epidemiological Virology |
| EPI_ISL_462269 | KU Leuven, Rega Institute, Clinical and Epidemiological Virology | KU Leuven, Rega Institute, Clinical and Epidemiological Virology |
| EPI_ISL_462270 | KU Leuven, Rega Institute, Clinical and Epidemiological Virology | KU Leuven, Rega Institute, Clinical and Epidemiological Virology |
| EPI_ISL_462271 | KU Leuven, Rega Institute, Clinical and Epidemiological Virology | KU Leuven, Rega Institute, Clinical and Epidemiological Virology |
| EPI_ISL_462272 | KU Leuven, Rega Institute, Clinical and Epidemiological Virology | KU Leuven, Rega Institute, Clinical and Epidemiological Virology |
| EPI_ISL_462273 | KU Leuven, Rega Institute, Clinical and Epidemiological Virology | KU Leuven, Rega Institute, Clinical and Epidemiological Virology |
| EPI_ISL_462274 | KU Leuven, Rega Institute, Clinical and Epidemiological Virology | KU Leuven, Rega Institute, Clinical and Epidemiological Virology |
| EPI_ISL_462275 | KU Leuven, Rega Institute, Clinical and Epidemiological Virology | KU Leuven, Rega Institute, Clinical and Epidemiological Virology |

| EPI_Accession_ID | Originating Laboratory | Submitting Laboratory |
|---|---|---|
| EPI_ISL_462276 | National Public Health Laboratory, National Centre for Infectious Diseases | National Public Health Laboratory, National Centre for Infectious Diseases |
| EPI_ISL_462277 | National Public Health Laboratory, National Centre for Infectious Diseases | National Public Health Laboratory, National Centre for Infectious Diseases |
| EPI_ISL_462278 | National Public Health Laboratory, National Centre for Infectious Diseases | National Public Health Laboratory, National Centre for Infectious Diseases |
| EPI_ISL_462279 | National Public Health Laboratory, National Centre for Infectious Diseases | National Public Health Laboratory, National Centre for Infectious Diseases |
| EPI_ISL_462280 | National Public Health Laboratory, National Centre for Infectious Diseases | National Public Health Laboratory, National Centre for Infectious Diseases |
| EPI_ISL_462281 | National Public Health Laboratory, National Centre for Infectious Diseases | National Public Health Laboratory, National Centre for Infectious Diseases |
| EPI_ISL_462282 | National Public Health Laboratory, National Centre for Infectious Diseases | National Public Health Laboratory, National Centre for Infectious Diseases |
| EPI_ISL_462283 | National Public Health Laboratory, National Centre for Infectious Diseases | National Public Health Laboratory, National Centre for Infectious Diseases |
| EPI_ISL_462284 | National Public Health Laboratory, National Centre for Infectious Diseases | National Public Health Laboratory, National Centre for Infectious Diseases |
| EPI_ISL_462285 | National Public Health Laboratory, National Centre for Infectious Diseases | National Public Health Laboratory, National Centre for Infectious Diseases |
| EPI_ISL_462286 | National Public Health Laboratory, National Centre for Infectious Diseases | National Public Health Laboratory, National Centre for Infectious Diseases |
| EPI_ISL_462287 | National Public Health Laboratory, National Centre for Infectious Diseases | National Public Health Laboratory, National Centre for Infectious Diseases |
| EPI_ISL_462288 | National Public Health Laboratory, National Centre for Infectious Diseases | National Public Health Laboratory, National Centre for Infectious Diseases |
| EPI_ISL_462289 | National Public Health Laboratory, National Centre for Infectious Diseases | National Public Health Laboratory, National Centre for Infectious Diseases |
| EPI_ISL_462290 | National Public Health Laboratory, National Centre for Infectious Diseases | National Public Health Laboratory, National Centre for Infectious Diseases |
| EPI_ISL_462291 | National Public Health Laboratory, National Centre for Infectious Diseases | National Public Health Laboratory, National Centre for Infectious Diseases |

| EPI_Accession_ID | Originating Laboratory | Submitting Laboratory |
|---|---|---|
| EPI_ISL_462292 | National Public Health Laboratory, National Centre for Infectious Diseases | National Public Health Laboratory, National Centre for Infectious Diseases |
| EPI_ISL_462293 | National Public Health Laboratory, National Centre for Infectious Diseases | National Public Health Laboratory, National Centre for Infectious Diseases |
| EPI_ISL_462294 | National Public Health Laboratory, National Centre for Infectious Diseases | National Public Health Laboratory, National Centre for Infectious Diseases |
| EPI_ISL_462295 | National Public Health Laboratory, National Centre for Infectious Diseases | National Public Health Laboratory, National Centre for Infectious Diseases |
| EPI_ISL_462296 | National Public Health Laboratory, National Centre for Infectious Diseases | National Public Health Laboratory, National Centre for Infectious Diseases |
| EPI_ISL_462297 | National Public Health Laboratory, National Centre for Infectious Diseases | National Public Health Laboratory, National Centre for Infectious Diseases |
| EPI_ISL_462298 | National Public Health Laboratory, National Centre for Infectious Diseases | National Public Health Laboratory, National Centre for Infectious Diseases |
| EPI_ISL_462299 | National Public Health Laboratory, National Centre for Infectious Diseases | National Public Health Laboratory, National Centre for Infectious Diseases |
| EPI_ISL_462300 | National Public Health Laboratory, National Centre for Infectious Diseases | National Public Health Laboratory, National Centre for Infectious Diseases |
| EPI_ISL_462301 | National Public Health Laboratory, National Centre for Infectious Diseases | National Public Health Laboratory, National Centre for Infectious Diseases |
| EPI_ISL_462302 | National Public Health Laboratory, National Centre for Infectious Diseases | National Public Health Laboratory, National Centre for Infectious Diseases |
| EPI_ISL_462303 | National Public Health Laboratory, National Centre for Infectious Diseases | National Public Health Laboratory, National Centre for Infectious Diseases |
| EPI_ISL_462304 | National Public Health Laboratory, National Centre for Infectious Diseases | National Public Health Laboratory, National Centre for Infectious Diseases |
| EPI_ISL_462305 | National Public Health Laboratory, National Centre for Infectious Diseases | National Public Health Laboratory, National Centre for Infectious Diseases |
| EPI_ISL_462306 | National Public Health Laboratory, National Centre for Infectious Diseases | National Public Health Laboratory, National Centre for Infectious Diseases |
| EPI_ISL_462307 | National Public Health Laboratory, National Centre for Infectious Diseases | National Public Health Laboratory, National Centre for Infectious Diseases |

| EPI_Accession_ID | Originating Laboratory | Submitting Laboratory |
|---|---|---|
| EPI_ISL_462308 | National Public Health Laboratory, National Centre for Infectious Diseases | National Public Health Laboratory, National Centre for Infectious Diseases |
| EPI_ISL_462309 | National Public Health Laboratory, National Centre for Infectious Diseases | National Public Health Laboratory, National Centre for Infectious Diseases |
| EPI_ISL_462310 | National Public Health Laboratory, National Centre for Infectious Diseases | National Public Health Laboratory, National Centre for Infectious Diseases |
| EPI_ISL_462311 | National Public Health Laboratory, National Centre for Infectious Diseases | National Public Health Laboratory, National Centre for Infectious Diseases |
| EPI_ISL_462312 | National Public Health Laboratory, National Centre for Infectious Diseases | National Public Health Laboratory, National Centre for Infectious Diseases |
| EPI_ISL_462313 | National Public Health Laboratory, National Centre for Infectious Diseases | National Public Health Laboratory, National Centre for Infectious Diseases |
| EPI_ISL_462314 | National Public Health Laboratory, National Centre for Infectious Diseases | National Public Health Laboratory, National Centre for Infectious Diseases |
| EPI_ISL_462315 | National Public Health Laboratory, National Centre for Infectious Diseases | National Public Health Laboratory, National Centre for Infectious Diseases |
| EPI_ISL_462316 | National Public Health Laboratory, National Centre for Infectious Diseases | National Public Health Laboratory, National Centre for Infectious Diseases |
| EPI_ISL_462317 | National Public Health Laboratory, National Centre for Infectious Diseases | National Public Health Laboratory, National Centre for Infectious Diseases |
| EPI_ISL_462318 | National Public Health Laboratory, National Centre for Infectious Diseases | National Public Health Laboratory, National Centre for Infectious Diseases |
| EPI_ISL_462319 | National Public Health Laboratory, National Centre for Infectious Diseases | National Public Health Laboratory, National Centre for Infectious Diseases |
| EPI_ISL_462320 | National Public Health Laboratory, National Centre for Infectious Diseases | National Public Health Laboratory, National Centre for Infectious Diseases |
| EPI_ISL_462321 | National Public Health Laboratory, National Centre for Infectious Diseases | National Public Health Laboratory, National Centre for Infectious Diseases |
| EPI_ISL_462322 | National Public Health Laboratory, National Centre for Infectious Diseases | National Public Health Laboratory, National Centre for Infectious Diseases |
| EPI_ISL_462323 | National Public Health Laboratory, National Centre for Infectious Diseases | National Public Health Laboratory, National Centre for Infectious Diseases |

| EPI_Accession_ID | Originating Laboratory | Submitting Laboratory |
|---|---|---|
| EPI_ISL_462324 | National Public Health Laboratory, National Centre for Infectious Diseases | National Public Health Laboratory, National Centre for Infectious Diseases |
| EPI_ISL_462325 | National Public Health Laboratory, National Centre for Infectious Diseases | National Public Health Laboratory, National Centre for Infectious Diseases |
| EPI_ISL_462326 | National Public Health Laboratory, National Centre for Infectious Diseases | National Public Health Laboratory, National Centre for Infectious Diseases |
| EPI_ISL_462327 | National Public Health Laboratory, National Centre for Infectious Diseases | National Public Health Laboratory, National Centre for Infectious Diseases |
| EPI_ISL_462328 | National Public Health Laboratory, National Centre for Infectious Diseases | National Public Health Laboratory, National Centre for Infectious Diseases |
| EPI_ISL_462329 | National Public Health Laboratory, National Centre for Infectious Diseases | National Public Health Laboratory, National Centre for Infectious Diseases |
| EPI_ISL_462330 | National Public Health Laboratory, National Centre for Infectious Diseases | National Public Health Laboratory, National Centre for Infectious Diseases |
| EPI_ISL_462331 | National Public Health Laboratory, National Centre for Infectious Diseases | National Public Health Laboratory, National Centre for Infectious Diseases |
| EPI_ISL_462332 | National Public Health Laboratory, National Centre for Infectious Diseases | National Public Health Laboratory, National Centre for Infectious Diseases |
| EPI_ISL_462333 | National Public Health Laboratory, National Centre for Infectious Diseases | National Public Health Laboratory, National Centre for Infectious Diseases |
| EPI_ISL_462334 | National Public Health Laboratory, National Centre for Infectious Diseases | National Public Health Laboratory, National Centre for Infectious Diseases |
| EPI_ISL_462335 | National Public Health Laboratory, National Centre for Infectious Diseases | National Public Health Laboratory, National Centre for Infectious Diseases |
| EPI_ISL_462336 | National Public Health Laboratory, National Centre for Infectious Diseases | National Public Health Laboratory, National Centre for Infectious Diseases |
| EPI_ISL_462337 | National Public Health Laboratory, National Centre for Infectious Diseases | National Public Health Laboratory, National Centre for Infectious Diseases |
| EPI_ISL_462338 | National Public Health Laboratory, National Centre for Infectious Diseases | National Public Health Laboratory, National Centre for Infectious Diseases |
| EPI_ISL_462339 | National Public Health Laboratory, National Centre for Infectious Diseases | National Public Health Laboratory, National Centre for Infectious Diseases |

| EPI_Accession_ID | Originating Laboratory | Submitting Laboratory |
|---|---|---|
| EPI_ISL_462340 | National Public Health Laboratory, National Centre for Infectious Diseases | National Public Health Laboratory, National Centre for Infectious Diseases |
| EPI_ISL_462341 | National Public Health Laboratory, National Centre for Infectious Diseases | National Public Health Laboratory, National Centre for Infectious Diseases |
| EPI_ISL_462342 | National Public Health Laboratory, National Centre for Infectious Diseases | National Public Health Laboratory, National Centre for Infectious Diseases |
| EPI_ISL_462343 | National Public Health Laboratory, National Centre for Infectious Diseases | National Public Health Laboratory, National Centre for Infectious Diseases |
| EPI_ISL_462344 | National Public Health Laboratory, National Centre for Infectious Diseases | National Public Health Laboratory, National Centre for Infectious Diseases |
| EPI_ISL_462345 | National Public Health Laboratory, National Centre for Infectious Diseases | National Public Health Laboratory, National Centre for Infectious Diseases |
| EPI_ISL_462346 | National Public Health Laboratory, National Centre for Infectious Diseases | National Public Health Laboratory, National Centre for Infectious Diseases |
| EPI_ISL_462347 | National Public Health Laboratory, National Centre for Infectious Diseases | National Public Health Laboratory, National Centre for Infectious Diseases |
| EPI_ISL_462348 | National Public Health Laboratory, National Centre for Infectious Diseases | National Public Health Laboratory, National Centre for Infectious Diseases |
| EPI_ISL_462349 | National Public Health Laboratory, National Centre for Infectious Diseases | National Public Health Laboratory, National Centre for Infectious Diseases |
| EPI_ISL_462350 | National Public Health Laboratory, National Centre for Infectious Diseases | National Public Health Laboratory, National Centre for Infectious Diseases |
| EPI_ISL_462351 | National Public Health Laboratory, National Centre for Infectious Diseases | National Public Health Laboratory, National Centre for Infectious Diseases |
| EPI_ISL_462352 | National Public Health Laboratory, National Centre for Infectious Diseases | National Public Health Laboratory, National Centre for Infectious Diseases |
| EPI_ISL_462353 | National Public Health Laboratory, National Centre for Infectious Diseases | National Public Health Laboratory, National Centre for Infectious Diseases |
| EPI_ISL_462354 | National Public Health Laboratory, National Centre for Infectious Diseases | National Public Health Laboratory, National Centre for Infectious Diseases |
| EPI_ISL_462355 | National Public Health Laboratory, National Centre for Infectious Diseases | National Public Health Laboratory, National Centre for Infectious Diseases |

| EPI_Accession_ID | Originating Laboratory | Submitting Laboratory |
|---|---|---|
| EPI_ISL_462356 | National Public Health Laboratory, National Centre for Infectious Diseases | National Public Health Laboratory, National Centre for Infectious Diseases |
| EPI_ISL_462357 | National Public Health Laboratory, National Centre for Infectious Diseases | National Public Health Laboratory, National Centre for Infectious Diseases |
| EPI_ISL_462358 | National Public Health Laboratory, National Centre for Infectious Diseases | National Public Health Laboratory, National Centre for Infectious Diseases |
| EPI_ISL_462359 | National Public Health Laboratory, National Centre for Infectious Diseases | National Public Health Laboratory, National Centre for Infectious Diseases |
| EPI_ISL_462360 | National Public Health Laboratory, National Centre for Infectious Diseases | National Public Health Laboratory, National Centre for Infectious Diseases |
| EPI_ISL_462361 | National Public Health Laboratory, National Centre for Infectious Diseases | National Public Health Laboratory, National Centre for Infectious Diseases |
| EPI_ISL_462362 | National Public Health Laboratory, National Centre for Infectious Diseases | National Public Health Laboratory, National Centre for Infectious Diseases |
| EPI_ISL_462363 | National Public Health Laboratory, National Centre for Infectious Diseases | National Public Health Laboratory, National Centre for Infectious Diseases |
| EPI_ISL_462364 | National Public Health Laboratory, National Centre for Infectious Diseases | National Public Health Laboratory, National Centre for Infectious Diseases |
| EPI_ISL_462365 | National Public Health Laboratory, National Centre for Infectious Diseases | National Public Health Laboratory, National Centre for Infectious Diseases |
| EPI_ISL_462366 | National Public Health Laboratory, National Centre for Infectious Diseases | National Public Health Laboratory, National Centre for Infectious Diseases |
| EPI_ISL_462367 | National Public Health Laboratory, National Centre for Infectious Diseases | National Public Health Laboratory, National Centre for Infectious Diseases |
| EPI_ISL_462368 | National Public Health Laboratory, National Centre for Infectious Diseases | National Public Health Laboratory, National Centre for Infectious Diseases |
| EPI_ISL_462369 | National Public Health Laboratory, National Centre for Infectious Diseases | National Public Health Laboratory, National Centre for Infectious Diseases |
| EPI_ISL_462370 | National Public Health Laboratory, National Centre for Infectious Diseases | National Public Health Laboratory, National Centre for Infectious Diseases |
| EPI_ISL_462371 | National Public Health Laboratory, National Centre for Infectious Diseases | National Public Health Laboratory, National Centre for Infectious Diseases |

| EPI_Accession_ID | Originating Laboratory | Submitting Laboratory |
|---|---|---|
| EPI_ISL_462372 | National Public Health Laboratory, National Centre for Infectious Diseases | National Public Health Laboratory, National Centre for Infectious Diseases |
| EPI_ISL_462373 | National Public Health Laboratory, National Centre for Infectious Diseases | National Public Health Laboratory, National Centre for Infectious Diseases |
| EPI_ISL_462374 | National Public Health Laboratory, National Centre for Infectious Diseases | National Public Health Laboratory, National Centre for Infectious Diseases |
| EPI_ISL_462375 | National Public Health Laboratory, National Centre for Infectious Diseases | National Public Health Laboratory, National Centre for Infectious Diseases |
| EPI_ISL_462376 | National Public Health Laboratory, National Centre for Infectious Diseases | National Public Health Laboratory, National Centre for Infectious Diseases |
| EPI_ISL_462377 | National Public Health Laboratory, National Centre for Infectious Diseases | National Public Health Laboratory, National Centre for Infectious Diseases |
| EPI_ISL_462378 | National Public Health Laboratory, National Centre for Infectious Diseases | National Public Health Laboratory, National Centre for Infectious Diseases |
| EPI_ISL_462379 | National Public Health Laboratory, National Centre for Infectious Diseases | National Public Health Laboratory, National Centre for Infectious Diseases |
| EPI_ISL_462380 | National Public Health Laboratory, National Centre for Infectious Diseases | National Public Health Laboratory, National Centre for Infectious Diseases |
| EPI_ISL_462381 | National Public Health Laboratory, National Centre for Infectious Diseases | National Public Health Laboratory, National Centre for Infectious Diseases |
| EPI_ISL_462382 | National Public Health Laboratory, National Centre for Infectious Diseases | National Public Health Laboratory, National Centre for Infectious Diseases |
| EPI_ISL_462383 | National Public Health Laboratory, National Centre for Infectious Diseases | National Public Health Laboratory, National Centre for Infectious Diseases |
| EPI_ISL_462384 | National Public Health Laboratory, National Centre for Infectious Diseases | National Public Health Laboratory, National Centre for Infectious Diseases |
| EPI_ISL_462385 | National Public Health Laboratory, National Centre for Infectious Diseases | National Public Health Laboratory, National Centre for Infectious Diseases |
| EPI_ISL_462386 | National Public Health Laboratory, National Centre for Infectious Diseases | National Public Health Laboratory, National Centre for Infectious Diseases |
| EPI_ISL_462387 | National Public Health Laboratory, National Centre for Infectious Diseases | National Public Health Laboratory, National Centre for Infectious Diseases |

| EPI_Accession_ID | Originating Laboratory | Submitting Laboratory |
|---|---|---|
| EPI_ISL_462388 | National Public Health Laboratory, National Centre for Infectious Diseases | National Public Health Laboratory, National Centre for Infectious Diseases |
| EPI_ISL_462389 | National Public Health Laboratory, National Centre for Infectious Diseases | National Public Health Laboratory, National Centre for Infectious Diseases |
| EPI_ISL_462390 | National Public Health Laboratory, National Centre for Infectious Diseases | National Public Health Laboratory, National Centre for Infectious Diseases |
| EPI_ISL_462391 | National Public Health Laboratory, National Centre for Infectious Diseases | National Public Health Laboratory, National Centre for Infectious Diseases |
| EPI_ISL_462392 | National Public Health Laboratory, National Centre for Infectious Diseases | National Public Health Laboratory, National Centre for Infectious Diseases |
| EPI_ISL_462393 | National Public Health Laboratory, National Centre for Infectious Diseases | National Public Health Laboratory, National Centre for Infectious Diseases |
| EPI_ISL_462394 | National Public Health Laboratory, National Centre for Infectious Diseases | National Public Health Laboratory, National Centre for Infectious Diseases |
| EPI_ISL_462395 | National Public Health Laboratory, National Centre for Infectious Diseases | National Public Health Laboratory, National Centre for Infectious Diseases |
| EPI_ISL_462396 | National Public Health Laboratory, National Centre for Infectious Diseases | National Public Health Laboratory, National Centre for Infectious Diseases |
| EPI_ISL_462397 | National Public Health Laboratory, National Centre for Infectious Diseases | National Public Health Laboratory, National Centre for Infectious Diseases |
| EPI_ISL_462398 | National Public Health Laboratory, National Centre for Infectious Diseases | National Public Health Laboratory, National Centre for Infectious Diseases |
| EPI_ISL_462399 | National Public Health Laboratory, National Centre for Infectious Diseases | National Public Health Laboratory, National Centre for Infectious Diseases |
| EPI_ISL_462400 | National Public Health Laboratory, National Centre for Infectious Diseases | National Public Health Laboratory, National Centre for Infectious Diseases |
| EPI_ISL_462401 | National Public Health Laboratory, National Centre for Infectious Diseases | National Public Health Laboratory, National Centre for Infectious Diseases |
| EPI_ISL_462402 | National Public Health Laboratory, National Centre for Infectious Diseases | National Public Health Laboratory, National Centre for Infectious Diseases |
| EPI_ISL_462403 | National Public Health Laboratory, National Centre for Infectious Diseases | National Public Health Laboratory, National Centre for Infectious Diseases |

| EPI_Accession_ID | Originating Laboratory | Submitting Laboratory |
|---|---|---|
| EPI_ISL_462404 | National Public Health Laboratory, National Centre for Infectious Diseases | National Public Health Laboratory, National Centre for Infectious Diseases |
| EPI_ISL_462405 | National Public Health Laboratory, National Centre for Infectious Diseases | National Public Health Laboratory, National Centre for Infectious Diseases |
| EPI_ISL_462406 | National Public Health Laboratory, National Centre for Infectious Diseases | National Public Health Laboratory, National Centre for Infectious Diseases |
| EPI_ISL_462407 | National Public Health Laboratory, National Centre for Infectious Diseases | National Public Health Laboratory, National Centre for Infectious Diseases |
| EPI_ISL_462408 | National Public Health Laboratory, National Centre for Infectious Diseases | National Public Health Laboratory, National Centre for Infectious Diseases |
| EPI_ISL_462409 | National Public Health Laboratory, National Centre for Infectious Diseases | National Public Health Laboratory, National Centre for Infectious Diseases |
| EPI_ISL_462410 | National Public Health Laboratory, National Centre for Infectious Diseases | National Public Health Laboratory, National Centre for Infectious Diseases |
| EPI_ISL_462411 | National Public Health Laboratory, National Centre for Infectious Diseases | National Public Health Laboratory, National Centre for Infectious Diseases |
| EPI_ISL_462412 | National Public Health Laboratory, National Centre for Infectious Diseases | National Public Health Laboratory, National Centre for Infectious Diseases |
| EPI_ISL_462413 | National Public Health Laboratory, National Centre for Infectious Diseases | National Public Health Laboratory, National Centre for Infectious Diseases |
| EPI_ISL_462414 | National Public Health Laboratory, National Centre for Infectious Diseases | National Public Health Laboratory, National Centre for Infectious Diseases |
| EPI_ISL_462415 | National Public Health Laboratory, National Centre for Infectious Diseases | National Public Health Laboratory, National Centre for Infectious Diseases |
| EPI_ISL_462416 | National Public Health Laboratory, National Centre for Infectious Diseases | National Public Health Laboratory, National Centre for Infectious Diseases |
| EPI_ISL_462417 | National Public Health Laboratory, National Centre for Infectious Diseases | National Public Health Laboratory, National Centre for Infectious Diseases |
| EPI_ISL_462418 | National Public Health Laboratory, National Centre for Infectious Diseases | National Public Health Laboratory, National Centre for Infectious Diseases |
| EPI_ISL_462419 | National Public Health Laboratory, National Centre for Infectious Diseases | National Public Health Laboratory, National Centre for Infectious Diseases |

| EPI_Accession_ID | Originating Laboratory | Submitting Laboratory |
|---|---|---|
| EPI_ISL_462420 | National Public Health Laboratory, National Centre for Infectious Diseases | National Public Health Laboratory, National Centre for Infectious Diseases |
| EPI_ISL_462421 | National Public Health Laboratory, National Centre for Infectious Diseases | National Public Health Laboratory, National Centre for Infectious Diseases |
| EPI_ISL_462422 | National Public Health Laboratory, National Centre for Infectious Diseases | National Public Health Laboratory, National Centre for Infectious Diseases |
| EPI_ISL_462423 | National Public Health Laboratory, National Centre for Infectious Diseases | National Public Health Laboratory, National Centre for Infectious Diseases |
| EPI_ISL_462424 | National Public Health Laboratory, National Centre for Infectious Diseases | National Public Health Laboratory, National Centre for Infectious Diseases |
| EPI_ISL_462425 | National Public Health Laboratory, National Centre for Infectious Diseases | National Public Health Laboratory, National Centre for Infectious Diseases |
| EPI_ISL_462426 | National Public Health Laboratory, National Centre for Infectious Diseases | National Public Health Laboratory, National Centre for Infectious Diseases |
| EPI_ISL_462427 | National Public Health Laboratory, National Centre for Infectious Diseases | National Public Health Laboratory, National Centre for Infectious Diseases |
| EPI_ISL_462428 | National Public Health Laboratory, National Centre for Infectious Diseases | National Public Health Laboratory, National Centre for Infectious Diseases |
| EPI_ISL_462429 | National Public Health Laboratory, National Centre for Infectious Diseases | National Public Health Laboratory, National Centre for Infectious Diseases |
| EPI_ISL_462430 | National Public Health Laboratory, National Centre for Infectious Diseases | National Public Health Laboratory, National Centre for Infectious Diseases |
| EPI_ISL_462431 | National Public Health Laboratory, National Centre for Infectious Diseases | National Public Health Laboratory, National Centre for Infectious Diseases |
| EPI_ISL_462432 | National Public Health Laboratory, National Centre for Infectious Diseases | National Public Health Laboratory, National Centre for Infectious Diseases |
| EPI_ISL_462433 | National Public Health Laboratory, National Centre for Infectious Diseases | National Public Health Laboratory, National Centre for Infectious Diseases |
| EPI_ISL_462434 | unknown | Laboratory Diagnostic |
| EPI_ISL_462435 | unknown | Laboratory Diagnostic |
| EPI_ISL_462436 | unknown | Laboratory Diagnostic |
| EPI_ISL_462437 | unknown | Laboratory Diagnostic |
| EPI_ISL_462438 | unknown | Laboratory Diagnostic |

| EPI_Accession_ID | Originating Laboratory | Submitting Laboratory |
|---|---|---|
| EPI_ISL_462447 | FundaciÃ³ Lluita contra la SIDA (FLSida)/Hospital Universitari Germans Trias i Pujol | IrsiCaixa AIDS Research Lab |
| EPI_ISL_462448 | FundaciÃ³ Lluita contra la SIDA (FLSida)/Hospital Universitari Germans Trias i Pujol | IrsiCaixa AIDS Research Lab |
| EPI_ISL_462449 | FundaciÃ³ Lluita contra la SIDA (FLSida)/Hospital Universitari Germans Trias i Pujol | IrsiCaixa AIDS Research Lab |
| EPI_ISL_462450 | Clinical Center, University of Sarajevo | Charite Universitatsmedizin Berlin, Institute of Virology |
| EPI_ISL_462451 | Clinical Center, University of Sarajevo | Charite Universitatsmedizin Berlin, Institute of Virology |
| EPI_ISL_462452 | Clinical Center, University of Sarajevo | Charite Universitatsmedizin Berlin, Institute of Virology |
| EPI_ISL_462453 | Clinical Center, University of Sarajevo | Charite Universitatsmedizin Berlin, Institute of Virology |
| EPI_ISL_462454 | Clinical Center, University of Sarajevo | Charite Universitatsmedizin Berlin, Institute of Virology |
| EPI_ISL_462455 | Clinical Center, University of Sarajevo | Charite Universitatsmedizin Berlin, Institute of Virology |
| EPI_ISL_462456 | Clinical Center, University of Sarajevo | Charite Universitatsmedizin Berlin, Institute of Virology |
| EPI_ISL_462457 | Clinical Center, University of Sarajevo | Charite Universitatsmedizin Berlin, Institute of Virology |
| EPI_ISL_462458 | Clinical Center, University of Sarajevo | Charite Universitatsmedizin Berlin, Institute of Virology |
| EPI_ISL_462459 | Clinical Center, University of Sarajevo | Charite Universitatsmedizin Berlin, Institute of Virology |
| EPI_ISL_462460 | Clinical Center, University of Sarajevo | Charite Universitatsmedizin Berlin, Institute of Virology |
| EPI_ISL_462461 | Clinical Center, University of Sarajevo | Charite Universitatsmedizin Berlin, Institute of Virology |
| EPI_ISL_462462 | Clinical Center, University of Sarajevo | Charite Universitatsmedizin Berlin, Institute of Virology |

| EPI_Accession_ID | Originating Laboratory | Submitting Laboratory |
|---|---|---|
| EPI_ISL_462463 | Clinical Center, University of Sarajevo | Charite Universitatsmedizin Berlin, Institute of Virology |
| EPI_ISL_462464 | Clinical Center, University of Sarajevo | Charite Universitatsmedizin Berlin, Institute of Virology |
| EPI_ISL_462465 | Clinical Center, University of Sarajevo | Charite Universitatsmedizin Berlin, Institute of Virology |
| EPI_ISL_462466 | Clinical Center, University of Sarajevo | Charite Universitatsmedizin Berlin, Institute of Virology |
| EPI_ISL_462467 | Clinical Center, University of Sarajevo | Charite Universitatsmedizin Berlin, Institute of Virology |
| EPI_ISL_462468 | Clinical Center, University of Sarajevo | Charite Universitatsmedizin Berlin, Institute of Virology |
| EPI_ISL_462469 | Clinical Center, University of Sarajevo | Charite Universitatsmedizin Berlin, Institute of Virology |
| EPI_ISL_462470 | Clinical Center, University of Sarajevo | Charite Universitatsmedizin Berlin, Institute of Virology |
| EPI_ISL_462471 | Clinical Center, University of Sarajevo | Charite Universitatsmedizin Berlin, Institute of Virology |
| EPI_ISL_462472 | Clinical Center, University of Sarajevo | Charite Universitatsmedizin Berlin, Institute of Virology |
| EPI_ISL_462473 | Clinical Center, University of Sarajevo | Charite Universitatsmedizin Berlin, Institute of Virology |
| EPI_ISL_462474 | Clinical Center, University of Sarajevo | Charite Universitatsmedizin Berlin, Institute of Virology |
| EPI_ISL_462475 | Clinical Center, University of Sarajevo | Charite Universitatsmedizin Berlin, Institute of Virology |
| EPI_ISL_462476 | Clinical Center, University of Sarajevo | Charite Universitatsmedizin Berlin, Institute of Virology |
| EPI_ISL_462477 | Hospital Costa del Sol | Instituto de Salud Carlos III |
| EPI_ISL_462480 | Institute of Human Genetics, Polish Academy of Sciences | Institute of Human Genetics, Polish Academy of Sciences |
| EPI_ISL_462636 | Michigan Department of Health and Human Services, Bureau of Laboratories | Michigan Department of Health and Human Services, Bureau of Laboratories |

| EPI_Accession_ID | Originating Laboratory | Submitting Laboratory |
|---|---|---|
| EPI_ISL_462637 | Michigan Department of Health and Human Services, Bureau of Laboratories | Michigan Department of Health and Human Services, Bureau of Laboratories |
| EPI_ISL_462638 | Michigan Department of Health and Human Services, Bureau of Laboratories | Michigan Department of Health and Human Services, Bureau of Laboratories |
| EPI_ISL_462639 | Michigan Department of Health and Human Services, Bureau of Laboratories | Michigan Department of Health and Human Services, Bureau of Laboratories |
| EPI_ISL_462640 | Michigan Department of Health and Human Services, Bureau of Laboratories | Michigan Department of Health and Human Services, Bureau of Laboratories |
| EPI_ISL_462641 | Michigan Department of Health and Human Services, Bureau of Laboratories | Michigan Department of Health and Human Services, Bureau of Laboratories |
| EPI_ISL_462642 | Michigan Department of Health and Human Services, Bureau of Laboratories | Michigan Department of Health and Human Services, Bureau of Laboratories |
| EPI_ISL_462643 | Michigan Department of Health and Human Services, Bureau of Laboratories | Michigan Department of Health and Human Services, Bureau of Laboratories |
| EPI_ISL_462644 | Michigan Department of Health and Human Services, Bureau of Laboratories | Michigan Department of Health and Human Services, Bureau of Laboratories |
| EPI_ISL_462645 | Michigan Department of Health and Human Services, Bureau of Laboratories | Michigan Department of Health and Human Services, Bureau of Laboratories |
| EPI_ISL_462646 | Michigan Department of Health and Human Services, Bureau of Laboratories | Michigan Department of Health and Human Services, Bureau of Laboratories |
| EPI_ISL_462647 | Michigan Department of Health and Human Services, Bureau of Laboratories | Michigan Department of Health and Human Services, Bureau of Laboratories |
| EPI_ISL_462648 | Michigan Department of Health and Human Services, Bureau of Laboratories | Michigan Department of Health and Human Services, Bureau of Laboratories |
| EPI_ISL_462649 | Michigan Department of Health and Human Services, Bureau of Laboratories | Michigan Department of Health and Human Services, Bureau of Laboratories |
| EPI_ISL_462650 | Michigan Department of Health and Human Services, Bureau of Laboratories | Michigan Department of Health and Human Services, Bureau of Laboratories |
| EPI_ISL_462651 | Michigan Department of Health and Human Services, Bureau of Laboratories | Michigan Department of Health and Human Services, Bureau of Laboratories |
| EPI_ISL_462652 | Michigan Department of Health and Human Services, Bureau of Laboratories | Michigan Department of Health and Human Services, Bureau of Laboratories |

| EPI_Accession_ID | Originating Laboratory | Submitting Laboratory |
|---|---|---|
| EPI_ISL_462653 | Michigan Department of Health and Human Services, Bureau of Laboratories | Michigan Department of Health and Human Services, Bureau of Laboratories |
| EPI_ISL_462654 | Michigan Department of Health and Human Services, Bureau of Laboratories | Michigan Department of Health and Human Services, Bureau of Laboratories |
| EPI_ISL_462655 | Michigan Department of Health and Human Services, Bureau of Laboratories | Michigan Department of Health and Human Services, Bureau of Laboratories |
| EPI_ISL_462656 | Michigan Department of Health and Human Services, Bureau of Laboratories | Michigan Department of Health and Human Services, Bureau of Laboratories |
| EPI_ISL_462657 | Michigan Department of Health and Human Services, Bureau of Laboratories | Michigan Department of Health and Human Services, Bureau of Laboratories |
| EPI_ISL_462658 | Michigan Department of Health and Human Services, Bureau of Laboratories | Michigan Department of Health and Human Services, Bureau of Laboratories |
| EPI_ISL_462659 | Michigan Department of Health and Human Services, Bureau of Laboratories | Michigan Department of Health and Human Services, Bureau of Laboratories |
| EPI_ISL_462660 | Michigan Department of Health and Human Services, Bureau of Laboratories | Michigan Department of Health and Human Services, Bureau of Laboratories |
| EPI_ISL_462661 | Michigan Department of Health and Human Services, Bureau of Laboratories | Michigan Department of Health and Human Services, Bureau of Laboratories |
| EPI_ISL_462662 | Michigan Department of Health and Human Services, Bureau of Laboratories | Michigan Department of Health and Human Services, Bureau of Laboratories |
| EPI_ISL_462663 | Michigan Department of Health and Human Services, Bureau of Laboratories | Michigan Department of Health and Human Services, Bureau of Laboratories |
| EPI_ISL_462664 | Michigan Department of Health and Human Services, Bureau of Laboratories | Michigan Department of Health and Human Services, Bureau of Laboratories |
| EPI_ISL_462665 | Michigan Department of Health and Human Services, Bureau of Laboratories | Michigan Department of Health and Human Services, Bureau of Laboratories |
| EPI_ISL_462666 | Michigan Department of Health and Human Services, Bureau of Laboratories | Michigan Department of Health and Human Services, Bureau of Laboratories |
| EPI_ISL_462667 | Michigan Department of Health and Human Services, Bureau of Laboratories | Michigan Department of Health and Human Services, Bureau of Laboratories |
| EPI_ISL_462668 | Michigan Department of Health and Human Services, Bureau of Laboratories | Michigan Department of Health and Human Services, Bureau of Laboratories |

| EPI_Accession_ID | Originating Laboratory | Submitting Laboratory |
|---|---|---|
| EPI_ISL_462669 | Michigan Department of Health and Human Services, Bureau of Laboratories | Michigan Department of Health and Human Services, Bureau of Laboratories |
| EPI_ISL_462670 | Michigan Department of Health and Human Services, Bureau of Laboratories | Michigan Department of Health and Human Services, Bureau of Laboratories |
| EPI_ISL_462671 | Michigan Department of Health and Human Services, Bureau of Laboratories | Michigan Department of Health and Human Services, Bureau of Laboratories |
| EPI_ISL_462672 | Michigan Department of Health and Human Services, Bureau of Laboratories | Michigan Department of Health and Human Services, Bureau of Laboratories |
| EPI_ISL_462673 | Michigan Department of Health and Human Services, Bureau of Laboratories | Michigan Department of Health and Human Services, Bureau of Laboratories |
| EPI_ISL_462674 | Michigan Department of Health and Human Services, Bureau of Laboratories | Michigan Department of Health and Human Services, Bureau of Laboratories |
| EPI_ISL_462675 | Michigan Department of Health and Human Services, Bureau of Laboratories | Michigan Department of Health and Human Services, Bureau of Laboratories |
| EPI_ISL_462676 | Michigan Department of Health and Human Services, Bureau of Laboratories | Michigan Department of Health and Human Services, Bureau of Laboratories |
| EPI_ISL_462677 | Michigan Department of Health and Human Services, Bureau of Laboratories | Michigan Department of Health and Human Services, Bureau of Laboratories |
| EPI_ISL_462678 | Michigan Department of Health and Human Services, Bureau of Laboratories | Michigan Department of Health and Human Services, Bureau of Laboratories |
| EPI_ISL_462679 | Michigan Department of Health and Human Services, Bureau of Laboratories | Michigan Department of Health and Human Services, Bureau of Laboratories |
| EPI_ISL_462680 | Michigan Department of Health and Human Services, Bureau of Laboratories | Michigan Department of Health and Human Services, Bureau of Laboratories |
| EPI_ISL_462681 | Michigan Department of Health and Human Services, Bureau of Laboratories | Michigan Department of Health and Human Services, Bureau of Laboratories |
| EPI_ISL_462682 | Michigan Department of Health and Human Services, Bureau of Laboratories | Michigan Department of Health and Human Services, Bureau of Laboratories |
| EPI_ISL_462683 | Michigan Department of Health and Human Services, Bureau of Laboratories | Michigan Department of Health and Human Services, Bureau of Laboratories |
| EPI_ISL_462684 | Michigan Department of Health and Human Services, Bureau of Laboratories | Michigan Department of Health and Human Services, Bureau of Laboratories |

| EPI_Accession_ID | Originating Laboratory | Submitting Laboratory |
|---|---|---|
| EPI_ISL_462685 | Michigan Department of Health and Human Services, Bureau of Laboratories | Michigan Department of Health and Human Services, Bureau of Laboratories |
| EPI_ISL_462686 | Michigan Department of Health and Human Services, Bureau of Laboratories | Michigan Department of Health and Human Services, Bureau of Laboratories |
| EPI_ISL_462687 | Michigan Department of Health and Human Services, Bureau of Laboratories | Michigan Department of Health and Human Services, Bureau of Laboratories |
| EPI_ISL_462688 | Michigan Department of Health and Human Services, Bureau of Laboratories | Michigan Department of Health and Human Services, Bureau of Laboratories |
| EPI_ISL_462689 | Michigan Department of Health and Human Services, Bureau of Laboratories | Michigan Department of Health and Human Services, Bureau of Laboratories |
| EPI_ISL_462690 | Michigan Department of Health and Human Services, Bureau of Laboratories | Michigan Department of Health and Human Services, Bureau of Laboratories |
| EPI_ISL_462691 | Michigan Department of Health and Human Services, Bureau of Laboratories | Michigan Department of Health and Human Services, Bureau of Laboratories |
| EPI_ISL_462692 | Michigan Department of Health and Human Services, Bureau of Laboratories | Michigan Department of Health and Human Services, Bureau of Laboratories |
| EPI_ISL_462693 | Michigan Department of Health and Human Services, Bureau of Laboratories | Michigan Department of Health and Human Services, Bureau of Laboratories |
| EPI_ISL_462694 | Michigan Department of Health and Human Services, Bureau of Laboratories | Michigan Department of Health and Human Services, Bureau of Laboratories |
| EPI_ISL_462695 | Michigan Department of Health and Human Services, Bureau of Laboratories | Michigan Department of Health and Human Services, Bureau of Laboratories |
| EPI_ISL_462696 | Michigan Department of Health and Human Services, Bureau of Laboratories | Michigan Department of Health and Human Services, Bureau of Laboratories |
| EPI_ISL_462697 | Michigan Department of Health and Human Services, Bureau of Laboratories | Michigan Department of Health and Human Services, Bureau of Laboratories |
| EPI_ISL_462698 | Michigan Department of Health and Human Services, Bureau of Laboratories | Michigan Department of Health and Human Services, Bureau of Laboratories |
| EPI_ISL_462699 | Michigan Department of Health and Human Services, Bureau of Laboratories | Michigan Department of Health and Human Services, Bureau of Laboratories |
| EPI_ISL_462700 | Michigan Department of Health and Human Services, Bureau of Laboratories | Michigan Department of Health and Human Services, Bureau of Laboratories |

| EPI_Accession_ID | Originating Laboratory | Submitting Laboratory |
|---|---|---|
| EPI_ISL_462701 | Michigan Department of Health and Human Services, Bureau of Laboratories | Michigan Department of Health and Human Services, Bureau of Laboratories |
| EPI_ISL_462702 | Michigan Department of Health and Human Services, Bureau of Laboratories | Michigan Department of Health and Human Services, Bureau of Laboratories |
| EPI_ISL_462703 | Michigan Department of Health and Human Services, Bureau of Laboratories | Michigan Department of Health and Human Services, Bureau of Laboratories |
| EPI_ISL_462704 | Michigan Department of Health and Human Services, Bureau of Laboratories | Michigan Department of Health and Human Services, Bureau of Laboratories |
| EPI_ISL_462705 | Michigan Department of Health and Human Services, Bureau of Laboratories | Michigan Department of Health and Human Services, Bureau of Laboratories |
| EPI_ISL_462706 | Michigan Department of Health and Human Services, Bureau of Laboratories | Michigan Department of Health and Human Services, Bureau of Laboratories |
| EPI_ISL_462707 | Michigan Department of Health and Human Services, Bureau of Laboratories | Michigan Department of Health and Human Services, Bureau of Laboratories |
| EPI_ISL_462708 | Michigan Department of Health and Human Services, Bureau of Laboratories | Michigan Department of Health and Human Services, Bureau of Laboratories |
| EPI_ISL_462709 | Michigan Department of Health and Human Services, Bureau of Laboratories | Michigan Department of Health and Human Services, Bureau of Laboratories |
| EPI_ISL_462710 | Michigan Department of Health and Human Services, Bureau of Laboratories | Michigan Department of Health and Human Services, Bureau of Laboratories |
| EPI_ISL_462711 | Michigan Department of Health and Human Services, Bureau of Laboratories | Michigan Department of Health and Human Services, Bureau of Laboratories |
| EPI_ISL_462712 | Michigan Department of Health and Human Services, Bureau of Laboratories | Michigan Department of Health and Human Services, Bureau of Laboratories |
| EPI_ISL_462713 | Michigan Department of Health and Human Services, Bureau of Laboratories | Michigan Department of Health and Human Services, Bureau of Laboratories |
| EPI_ISL_462714 | Michigan Department of Health and Human Services, Bureau of Laboratories | Michigan Department of Health and Human Services, Bureau of Laboratories |
| EPI_ISL_462715 | Michigan Department of Health and Human Services, Bureau of Laboratories | Michigan Department of Health and Human Services, Bureau of Laboratories |
| EPI_ISL_462716 | Michigan Department of Health and Human Services, Bureau of Laboratories | Michigan Department of Health and Human Services, Bureau of Laboratories |

| EPI_Accession_ID | Originating Laboratory | Submitting Laboratory |
|---|---|---|
| EPI_ISL_462717 | Michigan Department of Health and Human Services, Bureau of Laboratories | Michigan Department of Health and Human Services, Bureau of Laboratories |
| EPI_ISL_462718 | Michigan Department of Health and Human Services, Bureau of Laboratories | Michigan Department of Health and Human Services, Bureau of Laboratories |
| EPI_ISL_462719 | Michigan Department of Health and Human Services, Bureau of Laboratories | Michigan Department of Health and Human Services, Bureau of Laboratories |
| EPI_ISL_462720 | Michigan Department of Health and Human Services, Bureau of Laboratories | Michigan Department of Health and Human Services, Bureau of Laboratories |
| EPI_ISL_462721 | Michigan Department of Health and Human Services, Bureau of Laboratories | Michigan Department of Health and Human Services, Bureau of Laboratories |
| EPI_ISL_462722 | Michigan Department of Health and Human Services, Bureau of Laboratories | Michigan Department of Health and Human Services, Bureau of Laboratories |
| EPI_ISL_462723 | Michigan Department of Health and Human Services, Bureau of Laboratories | Michigan Department of Health and Human Services, Bureau of Laboratories |
| EPI_ISL_462724 | Michigan Department of Health and Human Services, Bureau of Laboratories | Michigan Department of Health and Human Services, Bureau of Laboratories |
| EPI_ISL_462725 | Michigan Department of Health and Human Services, Bureau of Laboratories | Michigan Department of Health and Human Services, Bureau of Laboratories |
| EPI_ISL_462726 | Michigan Department of Health and Human Services, Bureau of Laboratories | Michigan Department of Health and Human Services, Bureau of Laboratories |
| EPI_ISL_462727 | Michigan Department of Health and Human Services, Bureau of Laboratories | Michigan Department of Health and Human Services, Bureau of Laboratories |
| EPI_ISL_462728 | Michigan Department of Health and Human Services, Bureau of Laboratories | Michigan Department of Health and Human Services, Bureau of Laboratories |
| EPI_ISL_462729 | Michigan Department of Health and Human Services, Bureau of Laboratories | Michigan Department of Health and Human Services, Bureau of Laboratories |
| EPI_ISL_462730 | Michigan Department of Health and Human Services, Bureau of Laboratories | Michigan Department of Health and Human Services, Bureau of Laboratories |
| EPI_ISL_462731 | Michigan Department of Health and Human Services, Bureau of Laboratories | Michigan Department of Health and Human Services, Bureau of Laboratories |
| EPI_ISL_462732 | Michigan Department of Health and Human Services, Bureau of Laboratories | Michigan Department of Health and Human Services, Bureau of Laboratories |

| EPI_Accession_ID | Originating Laboratory | Submitting Laboratory |
|---|---|---|
| EPI_ISL_462733 | Michigan Department of Health and Human Services, Bureau of Laboratories | Michigan Department of Health and Human Services, Bureau of Laboratories |
| EPI_ISL_462734 | Michigan Department of Health and Human Services, Bureau of Laboratories | Michigan Department of Health and Human Services, Bureau of Laboratories |
| EPI_ISL_462735 | Michigan Department of Health and Human Services, Bureau of Laboratories | Michigan Department of Health and Human Services, Bureau of Laboratories |
| EPI_ISL_462736 | Michigan Department of Health and Human Services, Bureau of Laboratories | Michigan Department of Health and Human Services, Bureau of Laboratories |
| EPI_ISL_462753 | University Clinical Hospital of Mostar | University of Sarajevo Veterinary Faculty |
| EPI_ISL_462845 | Minnesota Department of Health, Public Health Laboratory | Minnesota Department of Health, Public Health Laboratory |
| EPI_ISL_462846 | Minnesota Department of Health, Public Health Laboratory | Minnesota Department of Health, Public Health Laboratory |
| EPI_ISL_462847 | Minnesota Department of Health, Public Health Laboratory | Minnesota Department of Health, Public Health Laboratory |
| EPI_ISL_462848 | Minnesota Department of Health, Public Health Laboratory | Minnesota Department of Health, Public Health Laboratory |
| EPI_ISL_462849 | Minnesota Department of Health, Public Health Laboratory | Minnesota Department of Health, Public Health Laboratory |
| EPI_ISL_462850 | Minnesota Department of Health, Public Health Laboratory | Minnesota Department of Health, Public Health Laboratory |
| EPI_ISL_462851 | Minnesota Department of Health, Public Health Laboratory | Minnesota Department of Health, Public Health Laboratory |
| EPI_ISL_462852 | Minnesota Department of Health, Public Health Laboratory | Minnesota Department of Health, Public Health Laboratory |
| EPI_ISL_462853 | Minnesota Department of Health, Public Health Laboratory | Minnesota Department of Health, Public Health Laboratory |
| EPI_ISL_462854 | Minnesota Department of Health, Public Health Laboratory | Minnesota Department of Health, Public Health Laboratory |
| EPI_ISL_462855 | Minnesota Department of Health, Public Health Laboratory | Minnesota Department of Health, Public Health Laboratory |
| EPI_ISL_462856 | Minnesota Department of Health, Public Health Laboratory | Minnesota Department of Health, Public Health Laboratory |

| EPI_Accession_ID | Originating Laboratory | Submitting Laboratory |
|---|---|---|
| EPI_ISL_462857 | Minnesota Department of Health, Public Health Laboratory | Minnesota Department of Health, Public Health Laboratory |
| EPI_ISL_462858 | Minnesota Department of Health, Public Health Laboratory | Minnesota Department of Health, Public Health Laboratory |
| EPI_ISL_462859 | Minnesota Department of Health, Public Health Laboratory | Minnesota Department of Health, Public Health Laboratory |
| EPI_ISL_462860 | Minnesota Department of Health, Public Health Laboratory | Minnesota Department of Health, Public Health Laboratory |
| EPI_ISL_462861 | Minnesota Department of Health, Public Health Laboratory | Minnesota Department of Health, Public Health Laboratory |
| EPI_ISL_462862 | Minnesota Department of Health, Public Health Laboratory | Minnesota Department of Health, Public Health Laboratory |
| EPI_ISL_462863 | Minnesota Department of Health, Public Health Laboratory | Minnesota Department of Health, Public Health Laboratory |
| EPI_ISL_462864 | Minnesota Department of Health, Public Health Laboratory | Minnesota Department of Health, Public Health Laboratory |
| EPI_ISL_462865 | Minnesota Department of Health, Public Health Laboratory | Minnesota Department of Health, Public Health Laboratory |
| EPI_ISL_462866 | Minnesota Department of Health, Public Health Laboratory | Minnesota Department of Health, Public Health Laboratory |
| EPI_ISL_462867 | Minnesota Department of Health, Public Health Laboratory | Minnesota Department of Health, Public Health Laboratory |
| EPI_ISL_462868 | Minnesota Department of Health, Public Health Laboratory | Minnesota Department of Health, Public Health Laboratory |
| EPI_ISL_462869 | Minnesota Department of Health, Public Health Laboratory | Minnesota Department of Health, Public Health Laboratory |
| EPI_ISL_462870 | Minnesota Department of Health, Public Health Laboratory | Minnesota Department of Health, Public Health Laboratory |
| EPI_ISL_462871 | Minnesota Department of Health, Public Health Laboratory | Minnesota Department of Health, Public Health Laboratory |
| EPI_ISL_462872 | Minnesota Department of Health, Public Health Laboratory | Minnesota Department of Health, Public Health Laboratory |

| EPI_Accession_ID | Originating Laboratory | Submitting Laboratory |
|---|---|---|
| EPI_ISL_462873 | Minnesota Department of Health, Public Health Laboratory | Minnesota Department of Health, Public Health Laboratory |
| EPI_ISL_462874 | Minnesota Department of Health, Public Health Laboratory | Minnesota Department of Health, Public Health Laboratory |
| EPI_ISL_462875 | Minnesota Department of Health, Public Health Laboratory | Minnesota Department of Health, Public Health Laboratory |
| EPI_ISL_462876 | Minnesota Department of Health, Public Health Laboratory | Minnesota Department of Health, Public Health Laboratory |
| EPI_ISL_462877 | Minnesota Department of Health, Public Health Laboratory | Minnesota Department of Health, Public Health Laboratory |
| EPI_ISL_462878 | Minnesota Department of Health, Public Health Laboratory | Minnesota Department of Health, Public Health Laboratory |
| EPI_ISL_462879 | Minnesota Department of Health, Public Health Laboratory | Minnesota Department of Health, Public Health Laboratory |
| EPI_ISL_462880 | Minnesota Department of Health, Public Health Laboratory | Minnesota Department of Health, Public Health Laboratory |
| EPI_ISL_462881 | Minnesota Department of Health, Public Health Laboratory | Minnesota Department of Health, Public Health Laboratory |
| EPI_ISL_462882 | Minnesota Department of Health, Public Health Laboratory | Minnesota Department of Health, Public Health Laboratory |
| EPI_ISL_462883 | Minnesota Department of Health, Public Health Laboratory | Minnesota Department of Health, Public Health Laboratory |
| EPI_ISL_462884 | Minnesota Department of Health, Public Health Laboratory | Minnesota Department of Health, Public Health Laboratory |
| EPI_ISL_462885 | Minnesota Department of Health, Public Health Laboratory | Minnesota Department of Health, Public Health Laboratory |
| EPI_ISL_462886 | Minnesota Department of Health, Public Health Laboratory | Minnesota Department of Health, Public Health Laboratory |
| EPI_ISL_462887 | Minnesota Department of Health, Public Health Laboratory | Minnesota Department of Health, Public Health Laboratory |
| EPI_ISL_462888 | Minnesota Department of Health, Public Health Laboratory | Minnesota Department of Health, Public Health Laboratory |

| EPI_Accession_ID | Originating Laboratory | Submitting Laboratory |
|---|---|---|
| EPI_ISL_462889 | Minnesota Department of Health, Public Health Laboratory | Minnesota Department of Health, Public Health Laboratory |
| EPI_ISL_462890 | Minnesota Department of Health, Public Health Laboratory | Minnesota Department of Health, Public Health Laboratory |
| EPI_ISL_462891 | Minnesota Department of Health, Public Health Laboratory | Minnesota Department of Health, Public Health Laboratory |
| EPI_ISL_462892 | Minnesota Department of Health, Public Health Laboratory | Minnesota Department of Health, Public Health Laboratory |
| EPI_ISL_462893 | Minnesota Department of Health, Public Health Laboratory | Minnesota Department of Health, Public Health Laboratory |
| EPI_ISL_462894 | Minnesota Department of Health, Public Health Laboratory | Minnesota Department of Health, Public Health Laboratory |
| EPI_ISL_462895 | Minnesota Department of Health, Public Health Laboratory | Minnesota Department of Health, Public Health Laboratory |
| EPI_ISL_462896 | Minnesota Department of Health, Public Health Laboratory | Minnesota Department of Health, Public Health Laboratory |
| EPI_ISL_462897 | Minnesota Department of Health, Public Health Laboratory | Minnesota Department of Health, Public Health Laboratory |
| EPI_ISL_462898 | Minnesota Department of Health, Public Health Laboratory | Minnesota Department of Health, Public Health Laboratory |
| EPI_ISL_462899 | Minnesota Department of Health, Public Health Laboratory | Minnesota Department of Health, Public Health Laboratory |
| EPI_ISL_462900 | Minnesota Department of Health, Public Health Laboratory | Minnesota Department of Health, Public Health Laboratory |
| EPI_ISL_462901 | Minnesota Department of Health, Public Health Laboratory | Minnesota Department of Health, Public Health Laboratory |
| EPI_ISL_462902 | Minnesota Department of Health, Public Health Laboratory | Minnesota Department of Health, Public Health Laboratory |
| EPI_ISL_462903 | Minnesota Department of Health, Public Health Laboratory | Minnesota Department of Health, Public Health Laboratory |
| EPI_ISL_462904 | Minnesota Department of Health, Public Health Laboratory | Minnesota Department of Health, Public Health Laboratory |

| EPI_Accession_ID | Originating Laboratory | Submitting Laboratory |
|---|---|---|
| EPI_ISL_462905 | Minnesota Department of Health, Public Health Laboratory | Minnesota Department of Health, Public Health Laboratory |
| EPI_ISL_462906 | Minnesota Department of Health, Public Health Laboratory | Minnesota Department of Health, Public Health Laboratory |
| EPI_ISL_462907 | Minnesota Department of Health, Public Health Laboratory | Minnesota Department of Health, Public Health Laboratory |
| EPI_ISL_462908 | Minnesota Department of Health, Public Health Laboratory | Minnesota Department of Health, Public Health Laboratory |
| EPI_ISL_462909 | Minnesota Department of Health, Public Health Laboratory | Minnesota Department of Health, Public Health Laboratory |
| EPI_ISL_462910 | Minnesota Department of Health, Public Health Laboratory | Minnesota Department of Health, Public Health Laboratory |
| EPI_ISL_462911 | Minnesota Department of Health, Public Health Laboratory | Minnesota Department of Health, Public Health Laboratory |
| EPI_ISL_462912 | Wyoming Public Health Laboratory | Center for Global Health, University of New Mexico Health Sciences Center |
| EPI_ISL_462913 | Wyoming Public Health Laboratory | Center for Global Health, University of New Mexico Health Sciences Center |
| EPI_ISL_462914 | Wyoming Public Health Laboratory | Center for Global Health, University of New Mexico Health Sciences Center |
| EPI_ISL_462915 | Wyoming Public Health Laboratory | Center for Global Health, University of New Mexico Health Sciences Center |
| EPI_ISL_462916 | Wyoming Public Health Laboratory | Center for Global Health, University of New Mexico Health Sciences Center |
| EPI_ISL_462917 | Wyoming Public Health Laboratory | Center for Global Health, University of New Mexico Health Sciences Center |
| EPI_ISL_462918 | Wyoming Public Health Laboratory | Center for Global Health, University of New Mexico Health Sciences Center |
| EPI_ISL_462919 | Wyoming Public Health Laboratory | Center for Global Health, University of New Mexico Health Sciences Center |
| EPI_ISL_462920 | Wyoming Public Health Laboratory | Center for Global Health, University of New Mexico Health Sciences Center |

| EPI_Accession_ID | Originating Laboratory | Submitting Laboratory |
|---|---|---|
| EPI_ISL_462921 | Wyoming Public Health Laboratory | Center for Global Health, University of New Mexico Health Sciences Center |
| EPI_ISL_462922 | Wyoming Public Health Laboratory | Center for Global Health, University of New Mexico Health Sciences Center |
| EPI_ISL_462923 | Wyoming Public Health Laboratory | Center for Global Health, University of New Mexico Health Sciences Center |
| EPI_ISL_462924 | Wyoming Public Health Laboratory | Center for Global Health, University of New Mexico Health Sciences Center |
| EPI_ISL_462925 | Wyoming Public Health Laboratory | Center for Global Health, University of New Mexico Health Sciences Center |
| EPI_ISL_462926 | Wyoming Public Health Laboratory | Center for Global Health, University of New Mexico Health Sciences Center |
| EPI_ISL_462927 | Wyoming Public Health Laboratory | Center for Global Health, University of New Mexico Health Sciences Center |
| EPI_ISL_462928 | Wyoming Public Health Laboratory | Center for Global Health, University of New Mexico Health Sciences Center |
| EPI_ISL_462929 | Wyoming Public Health Laboratory | Center for Global Health, University of New Mexico Health Sciences Center |
| EPI_ISL_462930 | Wyoming Public Health Laboratory | Center for Global Health, University of New Mexico Health Sciences Center |
| EPI_ISL_462931 | Wyoming Public Health Laboratory | Center for Global Health, University of New Mexico Health Sciences Center |
| EPI_ISL_462932 | Wyoming Public Health Laboratory | Center for Global Health, University of New Mexico Health Sciences Center |
| EPI_ISL_462933 | Wyoming Public Health Laboratory | Center for Global Health, University of New Mexico Health Sciences Center |
| EPI_ISL_462934 | Wyoming Public Health Laboratory | Center for Global Health, University of New Mexico Health Sciences Center |
| EPI_ISL_462935 | Wyoming Public Health Laboratory | Center for Global Health, University of New Mexico Health Sciences Center |
| EPI_ISL_462936 | Wyoming Public Health Laboratory | Center for Global Health, University of New Mexico Health Sciences Center |

| EPI_Accession_ID | Originating Laboratory | Submitting Laboratory |
|---|---|---|
| EPI_ISL_462937 | Wyoming Public Health Laboratory | Center for Global Health, University of New Mexico Health Sciences Center |
| EPI_ISL_462938 | Wyoming Public Health Laboratory | Center for Global Health, University of New Mexico Health Sciences Center |
| EPI_ISL_462939 | Wyoming Public Health Laboratory | Center for Global Health, University of New Mexico Health Sciences Center |
| EPI_ISL_462940 | Wyoming Public Health Laboratory | Center for Global Health, University of New Mexico Health Sciences Center |
| EPI_ISL_462941 | Wyoming Public Health Laboratory | Center for Global Health, University of New Mexico Health Sciences Center |
| EPI_ISL_462943 | Wyoming Public Health Laboratory | Center for Global Health, University of New Mexico Health Sciences Center |
| EPI_ISL_462944 | Wyoming Public Health Laboratory | Center for Global Health, University of New Mexico Health Sciences Center |
| EPI_ISL_462945 | Wyoming Public Health Laboratory | Center for Global Health, University of New Mexico Health Sciences Center |
| EPI_ISL_462946 | Wyoming Public Health Laboratory | Center for Global Health, University of New Mexico Health Sciences Center |
| EPI_ISL_462947 | Wyoming Public Health Laboratory | Center for Global Health, University of New Mexico Health Sciences Center |
| EPI_ISL_462948 | Wyoming Public Health Laboratory | Center for Global Health, University of New Mexico Health Sciences Center |
| EPI_ISL_462949 | Wyoming Public Health Laboratory | Center for Global Health, University of New Mexico Health Sciences Center |
| EPI_ISL_462950 | Wyoming Public Health Laboratory | Center for Global Health, University of New Mexico Health Sciences Center |
| EPI_ISL_462951 | Wyoming Public Health Laboratory | Center for Global Health, University of New Mexico Health Sciences Center |
| EPI_ISL_462952 | Wyoming Public Health Laboratory | Center for Global Health, University of New Mexico Health Sciences Center |
| EPI_ISL_462953 | Wyoming Public Health Laboratory | Center for Global Health, University of New Mexico Health Sciences Center |

| EPI_Accession_ID | Originating Laboratory | Submitting Laboratory |
|---|---|---|
| EPI_ISL_462954 | Wyoming Public Health Laboratory | Center for Global Health, University of New Mexico Health Sciences Center |
| EPI_ISL_462955 | Wyoming Public Health Laboratory | Center for Global Health, University of New Mexico Health Sciences Center |
| EPI_ISL_462956 | Wyoming Public Health Laboratory | Center for Global Health, University of New Mexico Health Sciences Center |
| EPI_ISL_462957 | Wyoming Public Health Laboratory | Center for Global Health, University of New Mexico Health Sciences Center |
| EPI_ISL_462958 | Wyoming Public Health Laboratory | Center for Global Health, University of New Mexico Health Sciences Center |
| EPI_ISL_462959 | Wyoming Public Health Laboratory | Center for Global Health, University of New Mexico Health Sciences Center |
| EPI_ISL_462960 | Wyoming Public Health Laboratory | Center for Global Health, University of New Mexico Health Sciences Center |
| EPI_ISL_462961 | Wyoming Public Health Laboratory | Center for Global Health, University of New Mexico Health Sciences Center |
| EPI_ISL_462962 | Wyoming Public Health Laboratory | Center for Global Health, University of New Mexico Health Sciences Center |
| EPI_ISL_462963 | Wyoming Public Health Laboratory | Center for Global Health, University of New Mexico Health Sciences Center |
| EPI_ISL_462964 | Wyoming Public Health Laboratory | Center for Global Health, University of New Mexico Health Sciences Center |
| EPI_ISL_462965 | Wyoming Public Health Laboratory | Center for Global Health, University of New Mexico Health Sciences Center |
| EPI_ISL_462966 | Wyoming Public Health Laboratory | Center for Global Health, University of New Mexico Health Sciences Center |
| EPI_ISL_462967 | Wyoming Public Health Laboratory | Center for Global Health, University of New Mexico Health Sciences Center |
| EPI_ISL_462968 | Wyoming Public Health Laboratory | Center for Global Health, University of New Mexico Health Sciences Center |
| EPI_ISL_462969 | Wyoming Public Health Laboratory | Center for Global Health, University of New Mexico Health Sciences Center |

| EPI_Accession_ID | Originating Laboratory | Submitting Laboratory |
|---|---|---|
| EPI_ISL_462970 | Wyoming Public Health Laboratory | Center for Global Health, University of New Mexico Health Sciences Center |
| EPI_ISL_462971 | Wyoming Public Health Laboratory | Center for Global Health, University of New Mexico Health Sciences Center |
| EPI_ISL_462972 | Wyoming Public Health Laboratory | Center for Global Health, University of New Mexico Health Sciences Center |
| EPI_ISL_462973 | Wyoming Public Health Laboratory | Center for Global Health, University of New Mexico Health Sciences Center |
| EPI_ISL_462975 | Wyoming Public Health Laboratory | Center for Global Health, University of New Mexico Health Sciences Center |
| EPI_ISL_462976 | Wyoming Public Health Laboratory | Center for Global Health, University of New Mexico Health Sciences Center |
| EPI_ISL_462977 | Wyoming Public Health Laboratory | Center for Global Health, University of New Mexico Health Sciences Center |
| EPI_ISL_462979 | Wyoming Public Health Laboratory | Center for Global Health, University of New Mexico Health Sciences Center |
| EPI_ISL_462980 | Wyoming Public Health Laboratory | Center for Global Health, University of New Mexico Health Sciences Center |
| EPI_ISL_462982 | Wyoming Public Health Laboratory | Center for Global Health, University of New Mexico Health Sciences Center |
| EPI_ISL_462983 | Wyoming Public Health Laboratory | Center for Global Health, University of New Mexico Health Sciences Center |
| EPI_ISL_462984 | Wyoming Public Health Laboratory | Center for Global Health, University of New Mexico Health Sciences Center |
| EPI_ISL_462987 | Wyoming Public Health Laboratory | Center for Global Health, University of New Mexico Health Sciences Center |
| EPI_ISL_462988 | Wyoming Public Health Laboratory | Center for Global Health, University of New Mexico Health Sciences Center |
| EPI_ISL_462989 | Wyoming Public Health Laboratory | Center for Global Health, University of New Mexico Health Sciences Center |
| EPI_ISL_462990 | University Clinical Centre of the Republic of Srpska | University of Sarajevo, Veterinary Faculty |
| EPI_ISL_462991 | unknown | Microbiology Division |
| EPI_ISL_462995 | unknown | Molecular Genetics |

| EPI_Accession_ID | Originating Laboratory | Submitting Laboratory |
| --- | --- | --- |
| EPI_ISL_462996 | unknown | Molecular Genetics |
| EPI_ISL_462997 | unknown | Molecular Genetics |
| EPI_ISL_462998 | unknown | Molecular Genetics |
| EPI_ISL_462999 | unknown | Molecular Genetics |
| EPI_ISL_463000 | unknown | Molecular Genetics |
| EPI_ISL_463001 | unknown | Clinical virology |
| EPI_ISL_463002 | unknown | Clinical virology |
| EPI_ISL_463003 | unknown | Clinical virology |
| EPI_ISL_463004 | unknown | Clinical virology |
| EPI_ISL_463005 | unknown | Clinical virology |
| EPI_ISL_463006 | unknown | Clinical virology |
| EPI_ISL_463007 | Department of Laboratory Medicine, National Taiwan University Hospital | Microbial Genomics Core Lab, National Taiwan University Centers of Genomic and Precision Medicine |
| EPI_ISL_463008 | Inst. of Molecular Virology | Inst. Hygiene |
| EPI_ISL_463010 | Institute of Life Sciences, Bhubaneswar | Immunogenomics lab, Institute of Life Sciences, Bhubaneswar |
| EPI_ISL_463011 | Institute of Life Sciences, Bhubaneswar | Immunogenomics lab, Institute of Life Sciences, Bhubaneswar |
| EPI_ISL_463013 | Institute of Life Sciences, Bhubaneswar | Immunogenomics lab, Institute of Life Sciences, Bhubaneswar |
| EPI_ISL_463014 | Institute of Life Sciences, Bhubaneswar | Immunogenomics lab, Institute of Life Sciences, Bhubaneswar |
| EPI_ISL_463015 | Institute of Life Sciences, Bhubaneswar | Immunogenomics lab, Institute of Life Sciences, Bhubaneswar |
| EPI_ISL_463016 | Institute of Life Sciences, Bhubaneswar | Immunogenomics lab, Institute of Life Sciences, Bhubaneswar |
| EPI_ISL_463017 | Institute of Life Sciences, Bhubaneswar | Immunogenomics lab, Institute of Life Sciences, Bhubaneswar |
| EPI_ISL_463018 | Institute of Life Sciences, Bhubaneswar | Immunogenomics lab, Institute of Life Sciences, Bhubaneswar |
| EPI_ISL_463019 | Institute of Life Sciences, Bhubaneswar | Immunogenomics lab, Institute of Life Sciences, Bhubaneswar |

| EPI_Accession_ID | Originating Laboratory | Submitting Laboratory |
|---|---|---|
| EPI_ISL_463020 | Institute of Life Sciences, Bhubaneswar | Immunogenomics lab, Institute of Life Sciences, Bhubaneswar |
| EPI_ISL_463021 | Institute of Life Sciences, Bhubaneswar | Immunogenomics lab, Institute of Life Sciences, Bhubaneswar |
| EPI_ISL_463022 | Institute of Life Sciences, Bhubaneswar | Immunogenomics lab, Institute of Life Sciences, Bhubaneswar |
| EPI_ISL_463023 | Institute of Life Sciences, Bhubaneswar | Immunogenomics lab, Institute of Life Sciences, Bhubaneswar |
| EPI_ISL_463024 | Institute of Life Sciences, Bhubaneswar | Immunogenomics lab, Institute of Life Sciences, Bhubaneswar |
| EPI_ISL_463025 | Institute of Life Sciences, Bhubaneswar | Immunogenomics lab, Institute of Life Sciences, Bhubaneswar |
| EPI_ISL_463026 | Institute of Life Sciences, Bhubaneswar | Immunogenomics lab, Institute of Life Sciences, Bhubaneswar |
| EPI_ISL_463027 | Institute of Life Sciences, Bhubaneswar | Immunogenomics lab, Institute of Life Sciences, Bhubaneswar |
| EPI_ISL_463028 | Institute of Life Sciences, Bhubaneswar | Immunogenomics lab, Institute of Life Sciences, Bhubaneswar |
| EPI_ISL_463029 | Institute of Life Sciences, Bhubaneswar | Immunogenomics lab, Institute of Life Sciences, Bhubaneswar |
| EPI_ISL_463030 | Institute of Life Sciences, Bhubaneswar | Immunogenomics lab, Institute of Life Sciences, Bhubaneswar |
| EPI_ISL_463031 | Institute of Life Sciences, Bhubaneswar | Immunogenomics lab, Institute of Life Sciences, Bhubaneswar |
| EPI_ISL_463032 | Institute of Life Sciences, Bhubaneswar | Immunogenomics lab, Institute of Life Sciences, Bhubaneswar |
| EPI_ISL_463033 | Institute of Life Sciences, Bhubaneswar | Immunogenomics lab, Institute of Life Sciences, Bhubaneswar |
| EPI_ISL_463034 | Institute of Life Sciences, Bhubaneswar | Immunogenomics lab, Institute of Life Sciences, Bhubaneswar |
| EPI_ISL_463035 | Institute of Life Sciences, Bhubaneswar | Immunogenomics lab, Institute of Life Sciences, Bhubaneswar |

| EPI_Accession_ID | Originating Laboratory | Submitting Laboratory |
|---|---|---|
| EPI_ISL_463036 | Institute of Life Sciences, Bhubaneswar | Immunogenomics lab, Institute of Life Sciences, Bhubaneswar |
| EPI_ISL_463037 | Institute of Life Sciences, Bhubaneswar | Immunogenomics lab, Institute of Life Sciences, Bhubaneswar |
| EPI_ISL_463038 | Institute of Life Sciences, Bhubaneswar | Immunogenomics lab, Institute of Life Sciences, Bhubaneswar |
| EPI_ISL_463039 | Institute of Life Sciences, Bhubaneswar | Immunogenomics lab, Institute of Life Sciences, Bhubaneswar |
| EPI_ISL_463040 | Institute of Life Sciences, Bhubaneswar | Immunogenomics lab, Institute of Life Sciences, Bhubaneswar |
| EPI_ISL_463041 | Institute of Life Sciences, Bhubaneswar | Immunogenomics lab, Institute of Life Sciences, Bhubaneswar |
| EPI_ISL_463042 | Institute of Life Sciences, Bhubaneswar | Immunogenomics lab, Institute of Life Sciences, Bhubaneswar |
| EPI_ISL_463043 | Institute of Life Sciences, Bhubaneswar | Immunogenomics lab, Institute of Life Sciences, Bhubaneswar |
| EPI_ISL_463044 | Institute of Life Sciences, Bhubaneswar | Immunogenomics lab, Institute of Life Sciences, Bhubaneswar |
| EPI_ISL_463045 | Institute of Life Sciences, Bhubaneswar | Immunogenomics lab, Institute of Life Sciences, Bhubaneswar |
| EPI_ISL_463046 | Institute of Life Sciences, Bhubaneswar | Immunogenomics lab, Institute of Life Sciences, Bhubaneswar |
| EPI_ISL_463047 | Institute of Life Sciences, Bhubaneswar | Immunogenomics lab, Institute of Life Sciences, Bhubaneswar |
| EPI_ISL_463048 | Institute of Life Sciences, Bhubaneswar | Immunogenomics lab, Institute of Life Sciences, Bhubaneswar |
| EPI_ISL_463049 | Institute of Life Sciences, Bhubaneswar | Immunogenomics lab, Institute of Life Sciences, Bhubaneswar |
| EPI_ISL_463050 | Institute of Life Sciences, Bhubaneswar | Immunogenomics lab, Institute of Life Sciences, Bhubaneswar |
| EPI_ISL_463051 | Institute of Life Sciences, Bhubaneswar | Immunogenomics lab, Institute of Life Sciences, Bhubaneswar |

| EPI_Accession_ID | Originating Laboratory | Submitting Laboratory |
|---|---|---|
| EPI_ISL_463052 | Institute of Life Sciences, Bhubaneswar | Immunogenomics lab, Institute of Life Sciences, Bhubaneswar |
| EPI_ISL_463053 | Institute of Life Sciences, Bhubaneswar | Immunogenomics lab, Institute of Life Sciences, Bhubaneswar |
| EPI_ISL_463054 | Institute of Life Sciences, Bhubaneswar | Immunogenomics lab, Institute of Life Sciences, Bhubaneswar |
| EPI_ISL_463055 | Institute of Life Sciences, Bhubaneswar | Immunogenomics lab, Institute of Life Sciences, Bhubaneswar |
| EPI_ISL_463057 | Institute of Life Sciences, Bhubaneswar | Immunogenomics lab, Institute of Life Sciences, Bhubaneswar |
| EPI_ISL_463058 | Institute of Life Sciences, Bhubaneswar | Immunogenomics lab, Institute of Life Sciences, Bhubaneswar |
| EPI_ISL_463060 | Institute of Life Sciences, Bhubaneswar | Immunogenomics lab, Institute of Life Sciences, Bhubaneswar |
| EPI_ISL_463061 | Institute of Life Sciences, Bhubaneswar | Immunogenomics lab, Institute of Life Sciences, Bhubaneswar |
| EPI_ISL_463062 | Institute of Life Sciences, Bhubaneswar | Immunogenomics lab, Institute of Life Sciences, Bhubaneswar |
| EPI_ISL_463063 | Institute of Life Sciences, Bhubaneswar | Immunogenomics lab, Institute of Life Sciences, Bhubaneswar |
| EPI_ISL_463064 | Institute of Life Sciences, Bhubaneswar | Immunogenomics lab, Institute of Life Sciences, Bhubaneswar |
| EPI_ISL_463065 | Institute of Life Sciences, Bhubaneswar | Immunogenomics lab, Institute of Life Sciences, Bhubaneswar |
| EPI_ISL_463066 | Institute of Life Sciences, Bhubaneswar | Immunogenomics lab, Institute of Life Sciences, Bhubaneswar |
| EPI_ISL_463067 | Institute of Life Sciences, Bhubaneswar | Immunogenomics lab, Institute of Life Sciences, Bhubaneswar |
| EPI_ISL_463068 | Institute of Life Sciences, Bhubaneswar | Immunogenomics lab, Institute of Life Sciences, Bhubaneswar |
| EPI_ISL_463069 | Institute of Life Sciences, Bhubaneswar | Immunogenomics lab, Institute of Life Sciences, Bhubaneswar |

| EPI_Accession_ID | Originating Laboratory | Submitting Laboratory |
|---|---|---|
| EPI_ISL_463070 | Institute of Life Sciences, Bhubaneswar | Immunogenomics lab, Institute of Life Sciences, Bhubaneswar |
| EPI_ISL_463071 | Institute of Life Sciences, Bhubaneswar | Immunogenomics lab, Institute of Life Sciences, Bhubaneswar |
| EPI_ISL_463072 | Institute of Life Sciences, Bhubaneswar | Immunogenomics lab, Institute of Life Sciences, Bhubaneswar |
| EPI_ISL_463073 | Institute of Life Sciences, Bhubaneswar | Immunogenomics lab, Institute of Life Sciences, Bhubaneswar |
| EPI_ISL_463074 | Institute of Life Sciences, Bhubaneswar | Immunogenomics lab, Institute of Life Sciences, Bhubaneswar |
| EPI_ISL_463075 | Institute of Life Sciences, Bhubaneswar | Immunogenomics lab, Institute of Life Sciences, Bhubaneswar |
| EPI_ISL_463076 | Institute of Life Sciences, Bhubaneswar | Immunogenomics lab, Institute of Life Sciences, Bhubaneswar |
| EPI_ISL_463077 | Institute of Life Sciences, Bhubaneswar | Immunogenomics lab, Institute of Life Sciences, Bhubaneswar |
| EPI_ISL_463078 | Institute of Life Sciences, Bhubaneswar | Immunogenomics lab, Institute of Life Sciences, Bhubaneswar |
| EPI_ISL_463079 | Institute of Life Sciences, Bhubaneswar | Immunogenomics lab, Institute of Life Sciences, Bhubaneswar |
| EPI_ISL_463080 | Institute of Life Sciences, Bhubaneswar | Immunogenomics lab, Institute of Life Sciences, Bhubaneswar |
| EPI_ISL_463081 | Institute of Life Sciences, Bhubaneswar | Immunogenomics lab, Institute of Life Sciences, Bhubaneswar |
| EPI_ISL_463082 | Institute of Life Sciences, Bhubaneswar | Immunogenomics lab, Institute of Life Sciences, Bhubaneswar |
| EPI_ISL_463083 | Institute of Life Sciences, Bhubaneswar | Immunogenomics lab, Institute of Life Sciences, Bhubaneswar |
| EPI_ISL_463084 | Institute of Life Sciences, Bhubaneswar | Immunogenomics lab, Institute of Life Sciences, Bhubaneswar |
| EPI_ISL_463085 | Institute of Life Sciences, Bhubaneswar | Immunogenomics lab, Institute of Life Sciences, Bhubaneswar |

| EPI_Accession_ID | Originating Laboratory | Submitting Laboratory |
|---|---|---|
| EPI_ISL_463087 | Institute of Life Sciences, Bhubaneswar | Immunogenomics lab, Institute of Life Sciences, Bhubaneswar |
| EPI_ISL_463088 | Institute of Life Sciences, Bhubaneswar | Immunogenomics lab, Institute of Life Sciences, Bhubaneswar |
| EPI_ISL_463089 | Institute of Life Sciences, Bhubaneswar | Immunogenomics lab, Institute of Life Sciences, Bhubaneswar |
| EPI_ISL_463092 | Institute of Life Sciences, Bhubaneswar | Immunogenomics lab, Institute of Life Sciences, Bhubaneswar |
| EPI_ISL_463093 | Institute of Life Sciences, Bhubaneswar | Immunogenomics lab, Institute of Life Sciences, Bhubaneswar |
| EPI_ISL_463094 | Virginia DCLS | Virginia DCLS |
| EPI_ISL_463095 | Virginia DCLS | Virginia DCLS |
| EPI_ISL_463096 | Virginia DCLS | Virginia DCLS |
| EPI_ISL_463097 | Virginia DCLS | Virginia DCLS |
| EPI_ISL_463098 | Virginia DCLS | Virginia DCLS |
| EPI_ISL_463099 | Virginia DCLS | Virginia DCLS |
| EPI_ISL_463100 | Virginia DCLS | Virginia DCLS |
| EPI_ISL_463101 | Virginia DCLS | Virginia DCLS |
| EPI_ISL_463102 | Virginia DCLS | Virginia DCLS |
| EPI_ISL_463103 | Virginia DCLS | Virginia DCLS |
| EPI_ISL_463104 | Virginia DCLS | Virginia DCLS |
| EPI_ISL_463105 | Virginia DCLS | Virginia DCLS |
| EPI_ISL_463106 | Virginia DCLS | Virginia DCLS |
| EPI_ISL_463107 | Virginia DCLS | Virginia DCLS |
| EPI_ISL_463108 | Virginia DCLS | Virginia DCLS |
| EPI_ISL_463109 | Virginia DCLS | Virginia DCLS |
| EPI_ISL_463110 | Virginia DCLS | Virginia DCLS |
| EPI_ISL_463111 | Virginia DCLS | Virginia DCLS |
| EPI_ISL_463112 | Virginia DCLS | Virginia DCLS |
| EPI_ISL_463113 | Virginia DCLS | Virginia DCLS |
| EPI_ISL_463114 | Virginia DCLS | Virginia DCLS |
| EPI_ISL_463115 | Virginia DCLS | Virginia DCLS |
| EPI_ISL_463116 | Virginia DCLS | Virginia DCLS |

| EPI_Accession_ID | Originating Laboratory | Submitting Laboratory |
|---|---|---|
| EPI_ISL_463117 | Virginia DCLS | Virginia DCLS |
| EPI_ISL_463118 | Virginia DCLS | Virginia DCLS |
| EPI_ISL_463119 | Virginia DCLS | Virginia DCLS |
| EPI_ISL_463120 | Virginia DCLS | Virginia DCLS |
| EPI_ISL_463121 | Virginia DCLS | Virginia DCLS |
| EPI_ISL_463122 | Virginia DCLS | Virginia DCLS |
| EPI_ISL_463123 | Virginia DCLS | Virginia DCLS |
| EPI_ISL_463124 | Virginia DCLS | Virginia DCLS |
| EPI_ISL_463125 | Virginia DCLS | Virginia DCLS |
| EPI_ISL_463126 | Virginia DCLS | Virginia DCLS |
| EPI_ISL_463127 | Virginia DCLS | Virginia DCLS |
| EPI_ISL_463128 | Virginia DCLS | Virginia DCLS |
| EPI_ISL_463129 | Virginia DCLS | Virginia DCLS |
| EPI_ISL_463130 | Virginia DCLS | Virginia DCLS |
| EPI_ISL_463131 | Virginia DCLS | Virginia DCLS |
| EPI_ISL_463132 | Virginia DCLS | Virginia DCLS |
| EPI_ISL_463133 | Virginia DCLS | Virginia DCLS |
| EPI_ISL_463134 | Virginia DCLS | Virginia DCLS |
| EPI_ISL_463135 | Virginia DCLS | Virginia DCLS |
| EPI_ISL_463136 | Virginia DCLS | Virginia DCLS |
| EPI_ISL_463137 | Virginia DCLS | Virginia DCLS |
| EPI_ISL_463138 | Yale Clinical Virology Laboratory | Grubaugh Lab - Yale School of Public Health |
| EPI_ISL_463139 | Yale Clinical Virology Laboratory | Grubaugh Lab - Yale School of Public Health |
| EPI_ISL_463140 | Yale Clinical Virology Laboratory | Grubaugh Lab - Yale School of Public Health |
| EPI_ISL_463141 | Yale Clinical Virology Laboratory | Grubaugh Lab - Yale School of Public Health |
| EPI_ISL_463142 | Yale Clinical Virology Laboratory | Grubaugh Lab - Yale School of Public Health |
| EPI_ISL_463143 | Yale Clinical Virology Laboratory | Grubaugh Lab - Yale School of Public Health |
| EPI_ISL_463144 | Yale Clinical Virology Laboratory | Grubaugh Lab - Yale School of Public Health |
| EPI_ISL_463145 | Yale Clinical Virology Laboratory | Grubaugh Lab - Yale School of Public Health |
| EPI_ISL_463146 | Yale Clinical Virology Laboratory | Grubaugh Lab - Yale School of Public Health |
| EPI_ISL_463147 | Yale Clinical Virology Laboratory | Grubaugh Lab - Yale School of Public Health |
| EPI_ISL_463148 | Yale Clinical Virology Laboratory | Grubaugh Lab - Yale School of Public Health |
| EPI_ISL_463149 | Yale Clinical Virology Laboratory | Grubaugh Lab - Yale School of Public Health |

| EPI_Accession_ID | Originating Laboratory | Submitting Laboratory |
|---|---|---|
| EPI_ISL_463150 | Yale Clinical Virology Laboratory | Grubaugh Lab - Yale School of Public Health |
| EPI_ISL_463151 | Yale Clinical Virology Laboratory | Grubaugh Lab - Yale School of Public Health |
| EPI_ISL_463152 | Yale Clinical Virology Laboratory | Grubaugh Lab - Yale School of Public Health |
| EPI_ISL_463153 | Yale Clinical Virology Laboratory | Grubaugh Lab - Yale School of Public Health |
| EPI_ISL_463154 | Yale Clinical Virology Laboratory | Grubaugh Lab - Yale School of Public Health |
| EPI_ISL_463155 | Yale Clinical Virology Laboratory | Grubaugh Lab - Yale School of Public Health |
| EPI_ISL_463156 | Yale Clinical Virology Laboratory | Grubaugh Lab - Yale School of Public Health |
| EPI_ISL_463157 | Yale Clinical Virology Laboratory | Grubaugh Lab - Yale School of Public Health |
| EPI_ISL_463158 | Yale Clinical Virology Laboratory | Grubaugh Lab - Yale School of Public Health |
| EPI_ISL_463159 | Yale Clinical Virology Laboratory | Grubaugh Lab - Yale School of Public Health |
| EPI_ISL_463160 | Yale Clinical Virology Laboratory | Grubaugh Lab - Yale School of Public Health |
| EPI_ISL_463161 | Yale Clinical Virology Laboratory | Grubaugh Lab - Yale School of Public Health |
| EPI_ISL_463162 | Yale Clinical Virology Laboratory | Grubaugh Lab - Yale School of Public Health |
| EPI_ISL_463163 | Yale Clinical Virology Laboratory | Grubaugh Lab - Yale School of Public Health |
| EPI_ISL_463164 | Yale Clinical Virology Laboratory | Grubaugh Lab - Yale School of Public Health |
| EPI_ISL_463165 | Yale Clinical Virology Laboratory | Grubaugh Lab - Yale School of Public Health |
| EPI_ISL_463166 | Yale Clinical Virology Laboratory | Grubaugh Lab - Yale School of Public Health |
| EPI_ISL_463167 | Yale Clinical Virology Laboratory | Grubaugh Lab - Yale School of Public Health |
| EPI_ISL_463168 | Yale Clinical Virology Laboratory | Grubaugh Lab - Yale School of Public Health |
| EPI_ISL_463169 | Yale Clinical Virology Laboratory | Grubaugh Lab - Yale School of Public Health |
| EPI_ISL_463170 | Yale Clinical Virology Laboratory | Grubaugh Lab - Yale School of Public Health |
| EPI_ISL_463171 | Yale Clinical Virology Laboratory | Grubaugh Lab - Yale School of Public Health |
| EPI_ISL_463172 | Yale Clinical Virology Laboratory | Grubaugh Lab - Yale School of Public Health |
| EPI_ISL_463173 | Yale Clinical Virology Laboratory | Grubaugh Lab - Yale School of Public Health |
| EPI_ISL_463277 | Ochsner Health | Bioinfoexperts, LLC |
| EPI_ISL_463278 | Ochsner Health | Bioinfoexperts, LLC |
| EPI_ISL_463279 | Ochsner Health | Bioinfoexperts, LLC |
| EPI_ISL_463280 | Ochsner Health | Bioinfoexperts, LLC |
| EPI_ISL_463281 | Ochsner Health | Bioinfoexperts, LLC |
| EPI_ISL_463282 | Ochsner Health | Bioinfoexperts, LLC |
| EPI_ISL_463283 | Ochsner Health | Bioinfoexperts, LLC |
| EPI_ISL_463284 | Ochsner Health | Bioinfoexperts, LLC |
| EPI_ISL_463285 | Ochsner Health | Bioinfoexperts, LLC |

| EPI_Accession_ID | Originating Laboratory | Submitting Laboratory |
|---|---|---|
| EPI_ISL_463286 | Ochsner Health | Bioinfoexperts, LLC |
| EPI_ISL_463287 | Ochsner Health | Bioinfoexperts, LLC |
| EPI_ISL_463288 | Ochsner Health | Bioinfoexperts, LLC |
| EPI_ISL_463289 | Ochsner Health | Bioinfoexperts, LLC |
| EPI_ISL_463290 | Ochsner Health | Bioinfoexperts, LLC |
| EPI_ISL_463291 | Ochsner Health | Bioinfoexperts, LLC |
| EPI_ISL_463292 | Ochsner Health | Bioinfoexperts, LLC |
| EPI_ISL_463293 | Ochsner Health | Bioinfoexperts, LLC |
| EPI_ISL_463294 | Ochsner Health | Bioinfoexperts, LLC |
| EPI_ISL_463295 | Ochsner Health | Bioinfoexperts, LLC |
| EPI_ISL_463296 | Ochsner Health | Bioinfoexperts, LLC |
| EPI_ISL_463297 | Ochsner Health | Bioinfoexperts, LLC |
| EPI_ISL_463298 | Ochsner Health | Bioinfoexperts, LLC |
| EPI_ISL_463299 | Ochsner Health | Bioinfoexperts, LLC |
| EPI_ISL_463300 | Ochsner Health | Bioinfoexperts, LLC |
